# Genome wide association study meta-analysis of neuropathologic lesions of Alzheimer’s disease and related dementias in a multi-site autopsy cohort

**DOI:** 10.64898/2026.01.22.26344634

**Authors:** Brenna Cholerton, Dana Godrich, Jeremy Pasteris, Joe Rivero, Eden R. Martin, Brian W. Kunkle, Adam C. Naj, Kara L. Hamilton-Nelson, Hui Wang, Wan-Ping Lee, Logan Dumitrescu, Timothy J. Hohman, Richard Mayeux, Eric B. Larson, Paul K. Crane, C. Dirk Keene, Caitlin S. Latimer, Shubhabrata Mukherjee, Julia K. Kofler, M. Ilyas Kamboh, David A. Bennett, Laura Molina-Porcel, Michael Cuccaro, Margaret A. Pericak-Vance, Tatjana Rundek, William K. Scott, Walter Kukull, Gerard Schellenberg, Alzheimer’s Disease Genetics Consortium, Gary W. Beecham, Thomas J. Montine

**Affiliations:** Department of Pathology, Stanford University School of Medicine, Stanford, CA, USA; Dr. John T Macdonald Foundation Department of Human Genetics, Miller School of Medicine, University of Miami, Miami, FL USA; John P. Hussman Institute for Human Genomics, Miller School of Medicine, University of Miami, Miami, FL USA; Department of Biostatistics and Data Science, Wake Forest University School of Medicine, Winston-Salem, NC, USA; Department of Pathology and Laboratory Medicine, Perelman School of Medicine, University of Pennsylvania, Philadelphia, Pennsylvania, USA; Penn Neurodegeneration Genomics Center, Perelman School of Medicine, University of Pennsylvania, Philadelphia, Pennsylvania, USA; Department of Biostatistics, Epidemiology, and Informatics, Perelman School of Medicine, University of Pennsylvania, Philadelphia, Pennsylvania, USA; Vanderbilt Memory & Alzheimer’s Center, Vanderbilt University Medical Center, Nashville, Tennessee, USA; Vanderbilt Genetics Institute, Vanderbilt University Medical Center, Nashville, Tennessee, USA; Department of Neurology, Vanderbilt University Medical Center, Nashville, TN, USA; Taub Institute on Alzheimer’s Disease and the Aging Brain, Department of Neurology, Columbia University, New York, NY, USA; Gertrude H. Sergievsky Center, Columbia University, New York, NY, USA; Department of Neurology, College of Physicians and Surgeons, Columbia University and the New York Presbyterian Hospital, New York, New York, USA; Department of Epidemiology, Mailman School of Public Health, Columbia University, New York, New York, USA; Kaiser Permanente Washington Health Research Institute, Seattle, Washington, USA; Department of Medicine, Division of General Internal Medicine, University of Washington, Harborview Medical Center, Seattle, Washington, USA; Department of Laboratory Medicine and Pathology, University of Washington, Seattle, Washington, USA; Division of General Internal Medicine, Department of Medicine, University of Washington, Seattle, USA; Department of Pathology, University of Pittsburgh, Pittsburgh, PA, USA; Department of Human Genetics, University of Pittsburgh School of Public Health, Pittsburgh, Pennsylvania, USA; Department of Psychiatry, School of Medicine, University of Pittsburgh, Pittsburgh, Pennsylvania, USA; Department of Epidemiology, University of Pittsburgh School of Public Health, Pittsburgh, Pennsylvania, USA; Rush Alzheimer’s Disease Center, Rush University Medical Center, Chicago, IL, USA; Alzheimer’s Disease and Other Cognitive Disorders Unit, Neurology Service, Hospital Clínic, Fundació Recerca Clínic Barcelona (FRCB), Institut d’Investigacions Biomediques August Pi I Sunyer (IDIBAPS), University of Barcelona, Barcelona, Spain; Neurological Tissue Bank of the Biobanc-Hospital Clínic-IDIBAPS, Barcelona, Spain; Department of Neurology, Evelyn F. McKnight Brain Institute, University of Miami Miller School of Medicine, Miami FL, USA; Department of Epidemiology, University of Washington, Seattle, WA, USA

## Abstract

Understanding the genetic foundations of dementia is critical to unraveling its complex molecular basis. Given that a clinical diagnosis of Alzheimer’s disease (AD) dementia often results from interplay between multiple underlying neuropathologic co-morbidities, previous genome-wide association studies (GWAS) of clinically diagnosed AD are restricted in their ability to translate genetic associations to potential targeted therapeutics. The current study seeks to address these limitations by presenting the largest GWAS to date (n=12,509) of neuropathologic hallmarks of AD and AD related dementias (ADRDs). We further performed a candidate-variant analysis using loci previously identified in GWAS of clinically diagnosed AD dementia and Parkinson’s disease (PD). Finally, we conducted heritability and genetic correlation analyses using linkage disequilibrium (LD) score regression. We found broad genome-wide significant associations with *APOE* across AD and ADRDs but not cerebrovascular disease and vascular brain injury. We further identified 12 significant loci across 10 neuropathologic phenotypes, including 5 loci previously implicated in GWAS of clinical AD and ADRDs (variants on *BIN1, PICALM / EED, TMEM106B, GRN,* and *SNCA / SNCA-AS1*) and 7 novel genome-wide associations (variants on *EPHA5, PSMG1, LINC00276, VAPA, LINC00290, DOCK4* and *SLAIN2 / SLC10A4*). Our analysis of AD and PD clinical candidate variants demonstrated several that were associated with AD neuropathologic change and Lewy body disease, as well as substantial overlap with neuropathologic lesions other than the primary neuropathologic hallmarks of these diseases. Heritability analyses demonstrated heritability that was high for amyloid plaques (78%) relative to prior clinical AD heritability analyses, intermediate for TDP-43 inclusions (41%), and low for remaining AD and ADRD pathologic features. This study underscores the importance of investigating the underlying neuropathologic hallmarks of AD and ADRDs as a step toward refining the translation of genetic associations to biomarker interpretation and development of targeted therapeutics.

**Author Summary:** Clinically diagnosed Alzheimer’s disease (AD) dementia is commonly associated with its hallmark pathologic changes plus neuropathologic features of prevalent co-morbid diseases such as cerebrovascular disease, Lewy body disease, and more recently discovered abnormalities in protein called TDP-43 (collectively, AD related dementias; ADRD). As a result, previous studies that associated clinical diagnosis of AD with specific genes may not tell us the whole story. For this study, we gathered autopsy and genetic data to identify relationships between genes and dementia-associated brain changes. We found some relationships between these diseases and genes that had been previously identified as contributing to clinical dementia, as well as some new relationships that had been previously unknown. We also found that some genes that had previously been identified in relation to AD were associated with different dementia-associated brain lesions. Finally, we found that the various brain lesions differ in the proportion that can be attributed to genetic vs. environmental differences. These results support that the pathway to a diagnosis of dementia can be caused by multiple factors and are an important step in beginning to identify individually based dementia treatments.

## Introduction

Although rare early onset dementia is commonly caused by an aggressive form of a single disease, data from multiple cohorts throughout the world strongly support that much more common late onset dementia is a complex interplay among multiple diseases. Indeed, there is substantial evidence that some mix of multiple neuropathologies, rather than Alzheimer’s disease (AD) only, is the most prevalent pathologic scenario among people clinically diagnosed with AD dementia.^1–7^ These commonly co-morbid AD related dementias (ADRDs) include cerebrovascular disease (CBVD) with resulting vascular brain injury (VBI), Lewy body disease (LBD), hippocampal sclerosis (HS), and limbic-predominant age-related TDP-43 encephalopathy (LATE). Importantly, dementia risk in older individuals, already a looming public health crisis, increases geometrically with increasing co-morbidity among AD and ADRDs.^2,7,8^

Understanding the genetic architecture of dementia is a critical component to unraveling the molecular basis for dementia. Many of the previous genome-wide association studies (GWAS) for AD focused on genes that correlated with a clinical diagnosis of AD dementia.^9–13^ Given the widely validated evidence for multiple neuropathologic processes underlying the clinical diagnosis of AD dementia,^7,14,15^ unaccounted co-morbidity may significantly undermine the translatability of clinical AD GWAS discoveries because it is unclear which of multiple co-morbidities are actually associated with the identified genetic locus.^16^ Translatability is further confounded by AD itself being a complex, multifaceted disease. For example, genetic loci identified by GWAS of AD dementia could operate through amyloid plaque pathways, neurofibrillary tangle pathways, other AD-related neuropathologic changes (e.g., neuroinflammation or vascular contributors, et cetera), or a combination of factors. GWAS of AD CSF biomarkers and amyloid PET may provide additional insights into genes that associate with risk, presence, and progression of AD, including possible comorbid conditions. However, these are limited in scope by their focus on specific proteinopathies. These are all potentially important distinctions when considering translation of genetic associations to therapeutics and interpretation of biomarkers.^17–20^

Given these potential limitations of clinical AD GWAS, the inclusion of neuropathologic hallmarks of AD and ADRDs may be potentially useful endophenotypes. However, only a few studies have attempted to include these pathologic endophenotypes in GWAS to disentangle genetic associations with specific diseases rather than their combined impact on clinical expression.^21–25^ A particular challenge to focusing on pathologic endophenotypes is limited sample sizes sufficient to power GWAS. Further complicating the effort is historical variation in how neuropathologic data were collected and described.

To address these challenges, we amassed the largest sample to date (n=12,509) of harmonized neuropathologic and genetic data gathered from the AD Genetics Consortium (ADGC). We hypothesized that using consensus methods for AD and ADRD hallmark lesions within a large sample would permit identification of genetic features that align with each clinic-pathologic entity rather than with the syndromic, functional endpoint of dementia that most commonly derives from multiple co-morbidities. Herein, we (1) present the largest GWAS meta-analysis conducted thus far of the commonly co-morbid neuropathologic lesions of AD and ADRDs assessed by current consensus protocols, (2) analyze previously identified clinical AD and PD GWAS candidates for their associations with AD neuropathologic change (ADNC) and ADRD neuropathologic lesions, and (3) present SNP heritability analysis of ADNC and ADRD lesions in the sample.

## Results

### Dataset characteristics and pathologic phenotype correlations

After exclusions and quality control, the final dataset consisted of 12,509 individuals. A description of contributing data sets and their demographics are included in **Table 1**. The dataset was mostly female (53.5%) with a mean age at death of 81.7 (±9.8). The prevalence of *APOE* ε4 allele was 37% ε4 heterozygotes and 9% ε4 homozygotes.

**Table 1.** Description of contributing data sets.

| **Cohort** | **N** | **Genotyping Array** | **Sex**<br>**% Female** | **Age at Death**<br>**Mean (SD)** | ***APOE***<br>**\*\*/\*$\epsilon 4$/$\epsilon 4\epsilon 4$** | **Dementia**<br>**(%)** |
| --- | --- | --- | --- | --- | --- | --- |
| **ACT** | **672** |  | 56.3 | 86.6 (4.6) | 0.73/0.25/0.02 | 25.2% |
| ACT1 | 557 | Illumina 660 |  |  |  |  |
| ACT2 | 115 | Illumina 660 |  |  |  |  |
| **ADC** | **7,103** |  | 48.9 | 80.6 (9.6) | 0.50/0.40/0.10 | 91.4% |
| ADC1 | 2,124 | Illumina 660 |  |  |  |  |
| ADC2 | 479 | Illumina 660 |  |  |  |  |
| ADC3-8 | 2,359 | Illumina OmniExpress |  |  |  |  |
| ADC9-12 | 1,078 | Illumina GSA MD v1-2 |  |  |  |  |
| ADC13,15 | 146 | Illumina GSA MD v3 |  |  |  |  |
| ADC14 | 917 | Illumina GSA MD v2 |  |  |  |  |
| **IDIBPAS** | **832** | Illumina GSA NBA | 57.5 | 80.2 (8.4) | 0.56/0.36/0.08 | -- |
| **NIA-LOAD** | **311** | Illumina 610 | 62.4 | 84.1 (7.3) | 0.35/0.50/0.15 | 95.6% |
| **TGEN** | **996** | Affymetrix 1M | 59.4 | 80.9 (9.8) | 0.50/0.40/0.10 | 64.7% |
| **MAYO** | **434** | Illumina HapMap300 | 43.5 | 72.6 (5.5) | 0.56/0.35/0.09 | 50.4% |
| **ROSMAP** | **1,576** |  | 67.3 | 89.2 (6.6) | 0.76/0.23/0.01 | 69.1% |
| ROSMAP1 | 1,297 | Affymetrix 6 |  |  |  |  |
| ROSMAP2 | 279 | Illumina OmniExpress |  |  |  |  |
| **UM** | **365** |  | 56.5 | 81.8 (10.2) | 0.62/0.29/0.09 | 74.8% |
| UM1 | 283 | Illumina GSA MD v2 |  |  |  |  |
| UM2 | 82 | Illumina 660 |  |  |  |  |
| **PITT** | **220** | Illumina Omni-Quad | 55.9 | 71 (6.6) | 0.37/0.50/0.13 | 99.1% |
| **Meta-analysis** | **12,509** |  | 53.5 | 81.7 (9.8) | 0.54/0.37/0.09 | 81.6% |

*APOE* column denotes frequency of no copies of the $\epsilon 4$ allele (\*\*), one copy (\* $\epsilon 4$ ), and two copies ( $\epsilon 4\epsilon 4$ ).
*Abbreviations:* **ACT**, Adult Changes in Thought Study; **ADC**, (NIH-funded) Alzheimer's Disease Research Center; **APOE**, apolipoprotein E; **IDIBAPS**, Instituto de Investigaciones Biomédicas August Pi i Sunyer; **MAF**, minor allele frequency; **MAYO**, Mayo Clinic Alzheimer's Disease Research Center; **NIA-LOAD**, National Institute on Aging Late-Onset Alzheimer's Disease Family Study; **ROSMAP**, Religious Orders Study/Memory and Aging Project; **SD**, standard deviation; **TGEN**, Translational Genomics Research Institute; **UM**, University of Miami Hussman Institute for Human Genomics; **PITT**, University of Pittsburgh Alzheimer's Disease Research Center

Consensus scores for ADNC and ADRD lesions were correlated as previously described,^26^ validating the pathologic assessments in our study. ADNC, including amyloid plaques (APs; assessed by Thal phase or its derivative National Institute on Aging and Alzheimer’s Association [NIA-AA] A score), neurofibrillary tangles (NFTs; assessed by Braak stage or its derivative NIA-AA B score), neuritic plaques (NPs; assessed by Consortium to Establish a Registry for AD [CERAD] NP score or its derivative NIA-AA C score), and the combined NIA-AA ADNC ABC score^27^ were strongly positively correlated with each other (r^2^=0.70 to 0.87) and had moderate correlations with *APOE* ε4, cerebral amyloid angiopathy (CAA), and LBD (r^2^=0.14 to 0.51, **Fig. S1**). Two measures of CBVD correlated moderately (r^2^=0.31 for atherosclerosis and arteriolosclerosis) while CAA was weakly correlated with these measures of CBVD (r^2^=0.05 to 0.07 for CAA with atherosclerosis or arteriolosclerosis). Two types of VBI (infarcts/lacunes and microinfarcts) also correlated positively but moderately with each other (r^2^=0.28). TDP-43 proteinopathy score and HS were correlated moderately (r^2^=0.32). In terms of frequency, the AD pathologies and CBVD measures were the most frequent (**Table S1**). Approximately 85% of the sample set was positive for amyloid at some level, and 81% showed moderate to high neurofibrillary tangles (Braak); when assessed, 90% of the samples were positive for one or more CBVD measure (atherosclerosis, arteriolosclerosis, or CAA).

### Genome-wide significant *APOE* associations with neuropathologic lesions

We observed genome-wide significant (GWS) *APOE* associations for NFTs, NPs, APs, CAA, LBD, HS, and TDP-43 proteinopathy but not CBVD or VBI, after controlling for age at death and sex (**Table S2**).

### GWS associations with ADNC

All GWAS were well-controlled for type-1 error (**Fig. S2**). As expected, we found strong GWS associations in the *APOE* locus for the dual proteinopathies of AD (**Fig. 1, Table S2**). Other (non-*APOE*) genome-wide significant variants are found in **Table 2**. GWAS of extent of APs (A score; **Fig. S3A**), extent of NFTs (B score, **Fig. S3B**), and NPs (C score **Fig. S3C**) identified GWS signal in the *BIN1* locus (top SNP: rs6733839; minimum *P_C_* = 1.6×10^−13^ and minimum *P_B_* = 1.4×10^−16^; **Fig. S4**). Effect sizes of top variants on *BIN1* were similar for NPs and NFTs (**Table 2, Table S3**). We also found a GWS signal with NPs (C score) on the *PICALM/EED* locus (top SNP: rs3851179; minimum *P* = 4.0×10^−8^), another previously identified AD region (**Fig. S5**). Additional ADNC variables include presence/absence of any amyloid plaques (NPs and/or APs), Thal phase,^28^ AD Braak stage, and NIA-AA ADNC ABC score. For the presence of APs, we identified 2 novel GWS associations: one near the Ephrin Receptor A5 (*EPHA5)* gene / the long intergenic non-protein coding RNA 2232 (*LINC02232*) and (rs144823952; *P* = 2.5×10^−8^) and another around the Proteasome Assembly Chaperone 1 gene (*PSMG1,* top SNP: rs2836880; minimum *P* = 2.4×10^−8^) which is also known as Down syndrome critical region 2 (**Fig. S6**). As expected, Thal phase and Braak stage showed similar results to their derived NIA-AA A and B scores (**Fig. S7**). Excepting the *EPHA5* locus, there was no evidence of effect-size heterogeneity at these loci; the *EPHA5* locus showed nominal evidence of heterogeneity (Cochran’s Q p-value=0.0448; Table 2).

**Figure 1:**
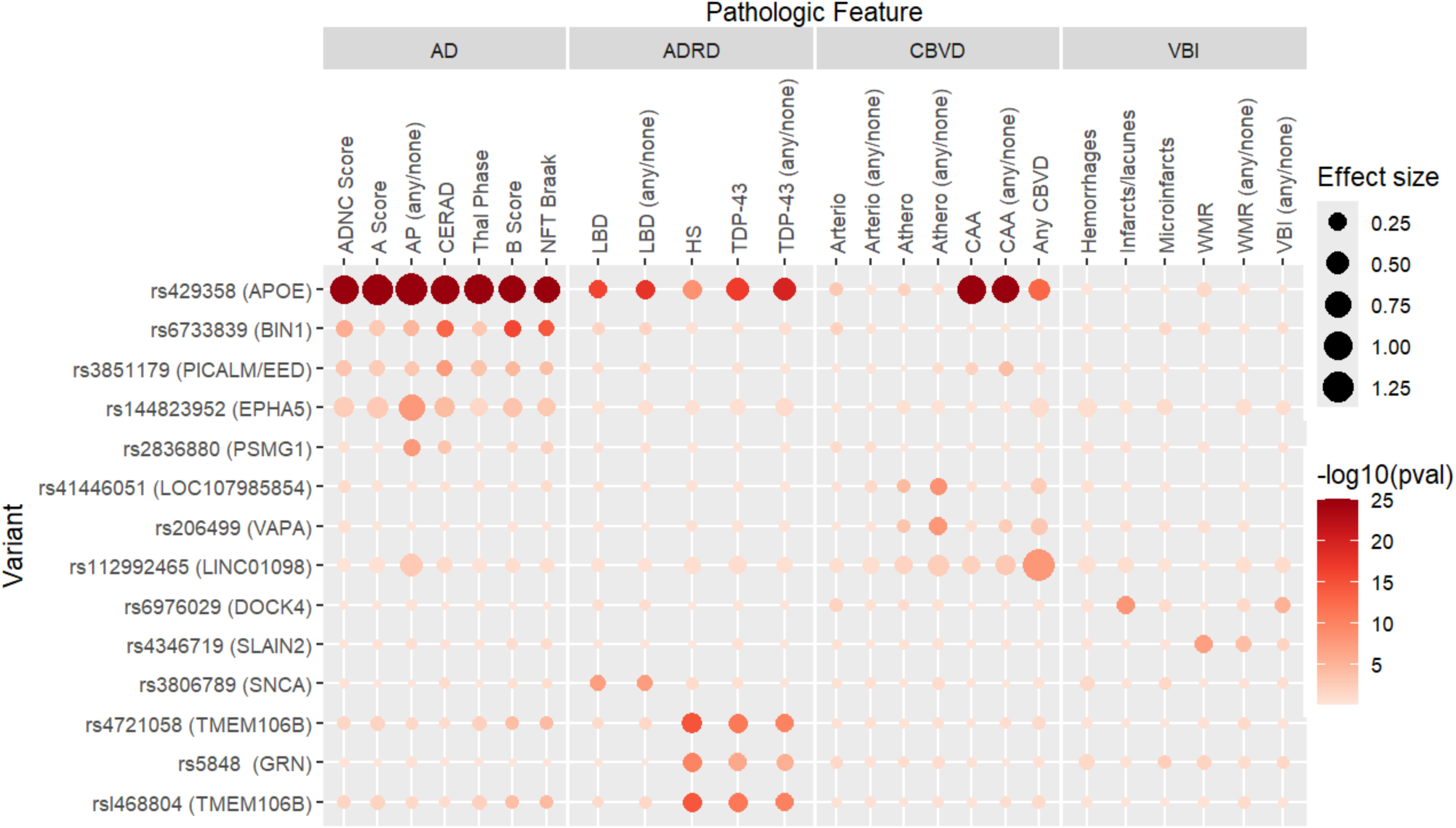
Variants associated with neuropathological lesions (genome-wide significant) Color denotes −log10(pval), capped at 25; size of points denotes absolute value of the effect size from the logistic regression models (e.g., “betas”).

**Table 2.**
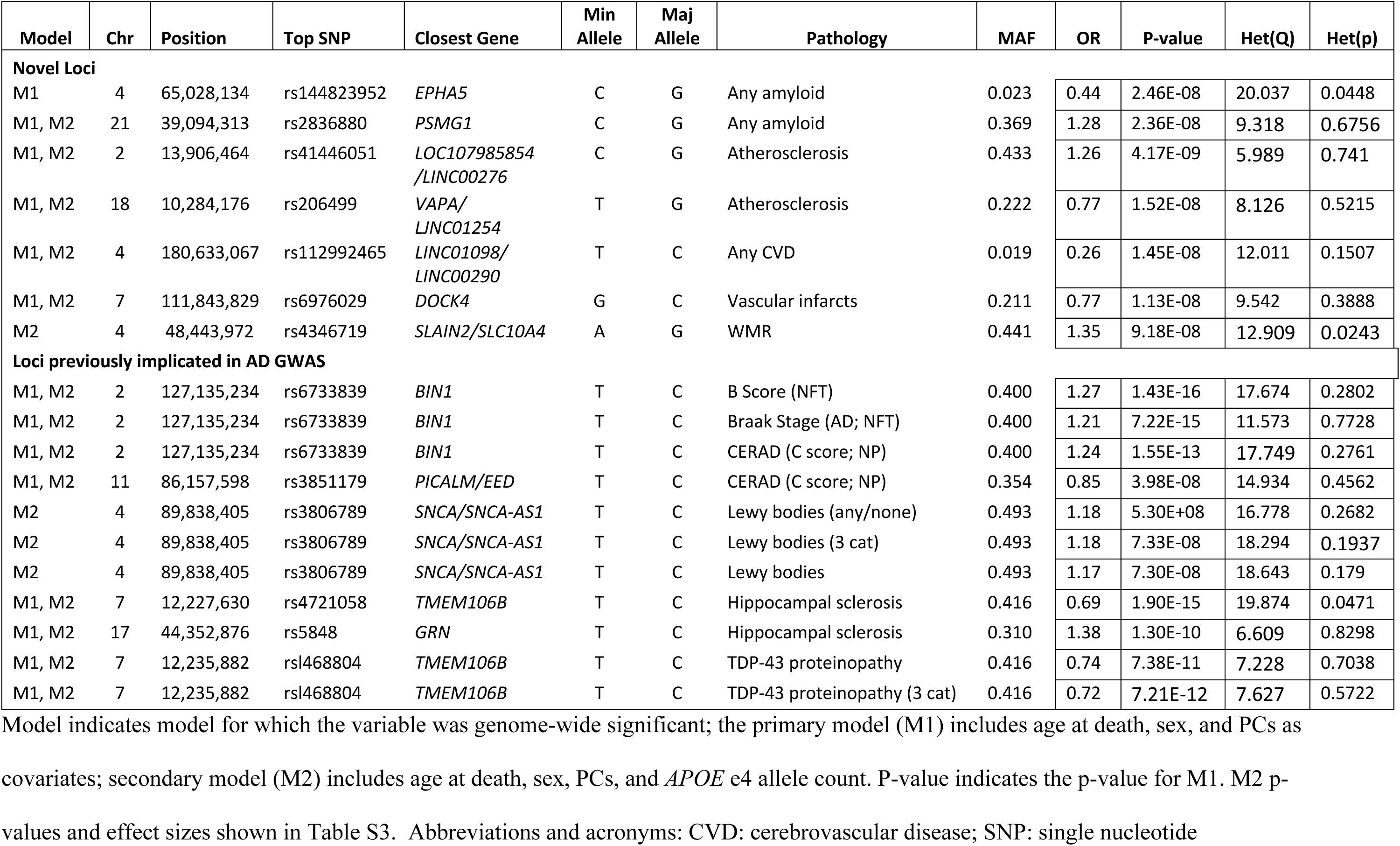
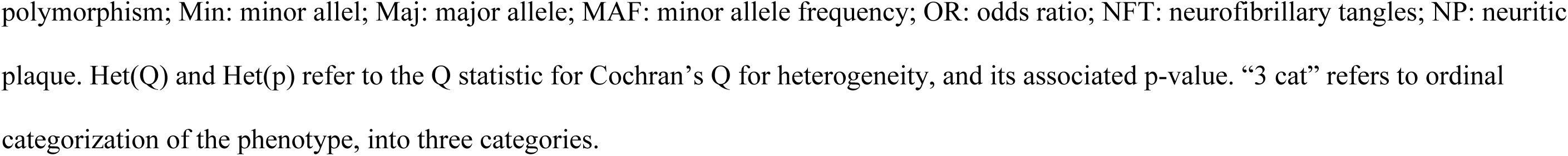
Genome-wide significant loci for AD and ADRD neuropathologies.

| Model | Chr | Position | Top SNP | Closest Gene | Min Allele | Maj Allele | Pathology | MAF | OR | P-value | Het(Q) | Het(p) |
| --- | --- | --- | --- | --- | --- | --- | --- | --- | --- | --- | --- | --- |
| **Novel Loci** |  |  |  |  |  |  |  |  |  |  |  |  |
| M1 | 4 | 65,028,134 | rs144823952 | *EPHA5* | C | G | Any amyloid | 0.023 | 0.44 | 2.46E-08 | 20.037 | 0.0448 |
| M1, M2 | 21 | 39,094,313 | rs2836880 | *PSMG1* | C | G | Any amyloid | 0.369 | 1.28 | 2.36E-08 | 9.318 | 0.6756 |
| M1, M2 | 2 | 13,906,464 | rs41446051 | *LOC107985854*<br>*/LINC00276* | C | G | Atherosclerosis | 0.433 | 1.26 | 4.17E-09 | 5.989 | 0.741 |
| M1, M2 | 18 | 10,284,176 | rs206499 | *VAPA*/<br>*LINC01254* | T | G | Atherosclerosis | 0.222 | 0.77 | 1.52E-08 | 8.126 | 0.5215 |
| M1, M2 | 4 | 180,633,067 | rs112992465 | *LINC01098*/<br>*LINC00290* | T | C | Any CVD | 0.019 | 0.26 | 1.45E-08 | 12.011 | 0.1507 |
| M1, M2 | 7 | 111,843,829 | rs6976029 | *DOCK4* | G | C | Vascular infarcts | 0.211 | 0.77 | 1.13E-08 | 9.542 | 0.3888 |
| M2 | 4 | 48,443,972 | rs4346719 | *SLAIN2/SLC10A4* | A | G | WMR | 0.441 | 1.35 | 9.18E-08 | 12.909 | 0.0243 |
| **Loci previously implicated in AD GWAS** |  |  |  |  |  |  |  |  |  |  |  |  |
| M1, M2 | 2 | 127,135,234 | rs6733839 | *BIN1* | T | C | B Score (NFT) | 0.400 | 1.27 | 1.43E-16 | 17.674 | 0.2802 |
| M1, M2 | 2 | 127,135,234 | rs6733839 | *BIN1* | T | C | Braak Stage (AD; NFT) | 0.400 | 1.21 | 7.22E-15 | 11.573 | 0.7728 |
| M1, M2 | 2 | 127,135,234 | rs6733839 | *BIN1* | T | C | CERAD (C score; NP) | 0.400 | 1.24 | 1.55E-13 | 17.749 | 0.2761 |
| M1, M2 | 11 | 86,157,598 | rs3851179 | *PICALM/EED* | T | C | CERAD (C score; NP) | 0.354 | 0.85 | 3.98E-08 | 14.934 | 0.4562 |
| M2 | 4 | 89,838,405 | rs3806789 | *SNCA/SNCA-AS1* | T | C | Lewy bodies (any/none) | 0.493 | 1.18 | 5.30E+08 | 16.778 | 0.2682 |
| M2 | 4 | 89,838,405 | rs3806789 | *SNCA/SNCA-AS1* | T | C | Lewy bodies (3 cat) | 0.493 | 1.18 | 7.33E-08 | 18.294 | 0.1937 |
| M2 | 4 | 89,838,405 | rs3806789 | *SNCA/SNCA-AS1* | T | C | Lewy bodies | 0.493 | 1.17 | 7.30E-08 | 18.643 | 0.179 |
| M1, M2 | 7 | 12,227,630 | rs4721058 | *TMEM106B* | T | C | Hippocampal sclerosis | 0.416 | 0.69 | 1.90E-15 | 19.874 | 0.0471 |
| M1, M2 | 17 | 44,352,876 | rs5848 | *GRN* | T | C | Hippocampal sclerosis | 0.310 | 1.38 | 1.30E-10 | 6.609 | 0.8298 |
| M1, M2 | 7 | 12,235,882 | rs1468804 | *TMEM106B* | T | C | TDP-43 proteinopathy | 0.416 | 0.74 | 7.38E-11 | 7.228 | 0.7038 |
| M1, M2 | 7 | 12,235,882 | rs1468804 | *TMEM106B* | T | C | TDP-43 proteinopathy (3 cat) | 0.416 | 0.72 | 7.21E-12 | 7.627 | 0.5722 |

polymorphism; Min: minor allele; Maj: major allele; MAF: minor allele frequency; OR: odds ratio; NFT: neurofibrillary tangles; NP: neuritic plaque. Het(Q) and Het(p) refer to the Q statistic for Cochran's Q for heterogeneity, and its associated p-value. "3 cat" refers to ordinal categorization of the phenotype, into three categories.

### GWS associations with ADRD lesions

#### CBVD

We identified three GWS signals among CBVD traits (**Fig. S8**): two for atherosclerosis and one for presence of CBVD (any atherosclerosis, arteriolosclerosis, or CAA). The atherosclerosis variants include a locus on chromosome 2 (*LINC00276*, top SNP = rs41446051, *P* = 4.2×10^−9^) and one variant near the Vesicle-Associated Membrane Protein (VAMP) Associated Protein A (*VAPA*, rs206499, *P* = 1.5×10^−8^) (**Figs. S9**, **Table 2**). The CBVD locus resides in an intergenic region of chromosome 4 (rs112992465, *P* = 1.4×10^−8^). There were no genome-wide significant associations with CAA, apart from *APOE* (**Fig. S10A**). There was, however, one nominally associated variant on chromosome 17 (rs17669645, *P* = 9.3×10^−7^) near the Cytochrome C Oxidase Assembly Factor Heme A:Farnesyltransferase (*COX10*) gene (**Fig. S10B; Table S3**).

#### VBI

GWAS of presence of infarcts/lacunes (**Fig. S11**) showed a GWS signal on the Dedicator of Cytokinesis 4 (*DOCK4*) gene (top SNP = rs6976029, *P* = 1.1×10^−8^) (**Fig. S11**, **Table 2**). Additional variables related to VBI, such as microinfarcts and to some extent white matter rarefaction, are presented in supplemental material (**Fig S12, Table S3**).

#### LBD

All primary models for each LBD categorization revealed GWS association with the *APOE* region with the same top SNP (rs429358; LBD 5-group, *P* = 9.2×10^−17^; LBD 3-group, *P* = 5.7×10^−18^; LBD 2-group, *P* = 1.9×10^−18^) (**Table S2**; **Fig. S13, Fig. S14**). None showed further GWS associations in the primary model. However, in the secondary model in which *APOE* ε4 count was a covariate, all three LBD groupings showed GWS associations with the alpha-synuclein gene and its antisense transcript (*SNCA / SNCA-AS1*) locus with the same top SNP (rs3806789; LBD 5-group, *P* = 4.3×10^−8^; LBD 3-group, *P* = 4.1×10^−8^; LBD 2-group, *P* = 2.8×10^−8^) (**Fig. 13B**). In primary models, top signals in the region reached suggestive significance (rs3806789; LBD 5-group, *P* = 7.3×10^−8^; LBD 3-group, *P* = 7.3×10^−8^; LBD 2-group, *P* = 5.3×10^−8^) (**Fig. S14**).

#### TDP-43 proteinopathy and HS

TDP-43 proteinopathy and HS are correlated (**Fig. S1**) and are commonly observed in FTLD-TDP, LATE-NC, and ADNC. Three loci achieved GWS with HS: *APOE* (lead SNP = rs429358, minimum *P* = 4.1×10−9), transmembrane protein 106B (*TMEM106B*; lead SNP = rs4721058, minimum *P* = 1.9×10−15), and granulin precursor gene (*GRN*; lead SNP = rs5848, minimum *P* = 1.3×10−10) (**Figs S15, S16**). There was nominal evidence of effect-size heterogeneity for the *TMEM106B* locus with the HS phenotype (Cochran’s Q p-value=0.0471). Primary results for these phenotypes have been previously reported elsewhere and are more fully discussed there.^29^

### Candidate variant associations

#### AD-candidate genetic variant associations with ADNC and ADRD lesions

Given the phenotypic correlations (**Fig S1**) and cooccurrence of neuropathological lesions, we assessed candidate variants from clinic-based AD and PD association studies for pleiotropic effects across the spectrum of neuropathological lesions. Of the 78 variants in our data that overlap with the Bellenguez *et al*.^30^ clinical AD GWAS, 20 were significantly associated (*P* < 0.05) with A score for ordinal ranking of APs, 30 with B score for ordinal ranking of NFTs, and 21 with C score for ordinal ranking of NPs. Variants with nominal significant in A, B, or C score are shown in **Table 3**. Of these, 11 associations are shared across all three scores; 9 were shared by ranking for NFTs and APs (B Score plus A and/or C Scores); eight variants were associated with ranking for APs alone (A and/or C Scores); and ten variants were associated with ranking NFTs alone (B Score) (**Table 3**). Of the 78 variants, 40 were not associated with any of the three ADNC scores.

**Table 3.**
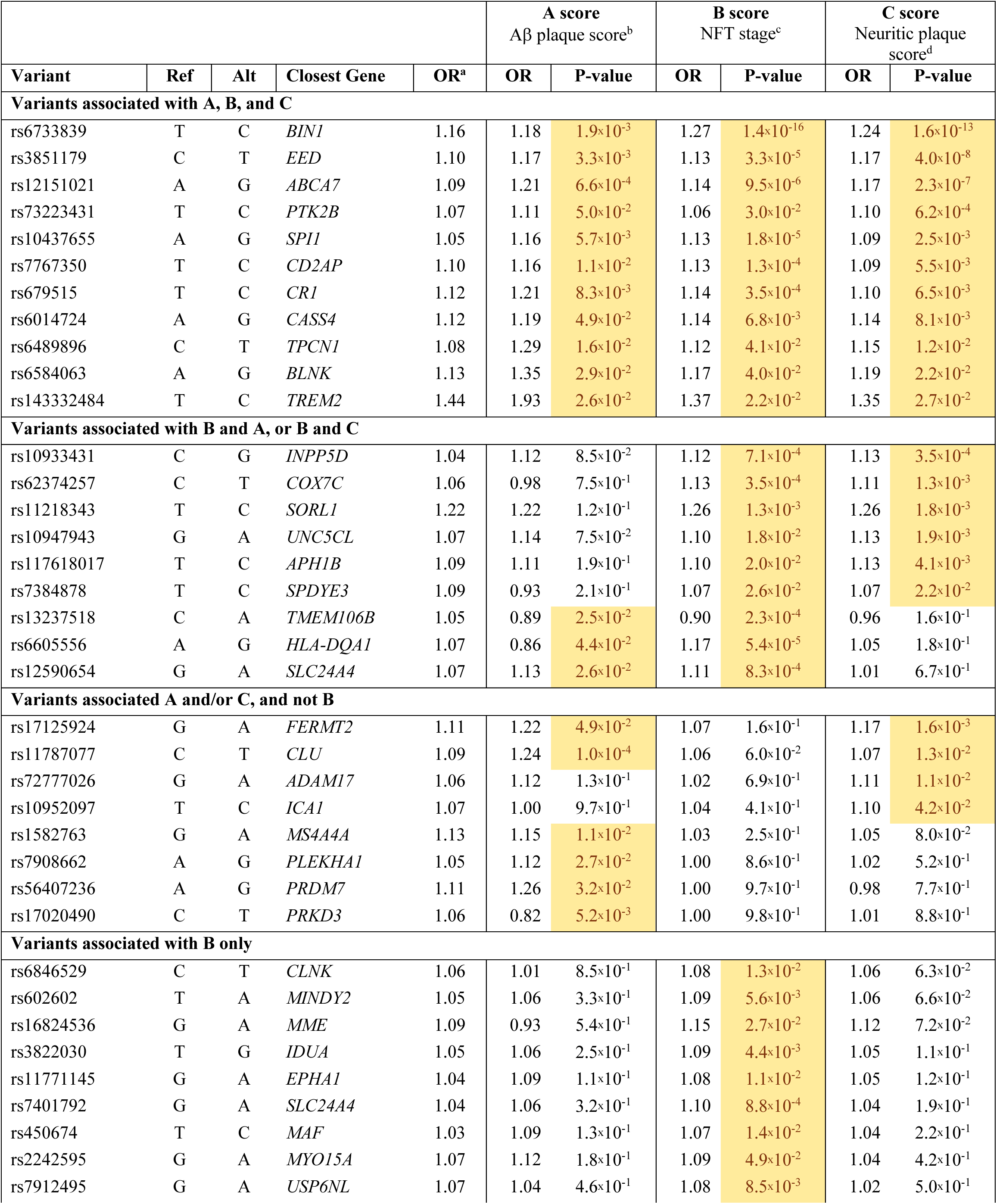
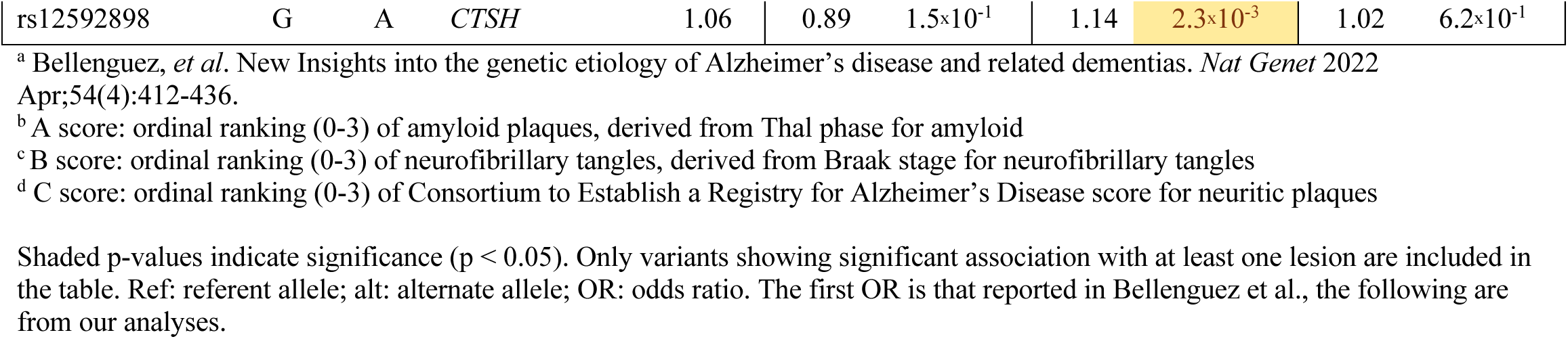
Summary statistics from Bellenguez *et al*.^a^ clinical AD candidate-variant analysis for AD pathologic phenotypes.

We extended this candidate variant analysis to ADRD lesions and again observed shared genetic risk with ADNC, although not as strong as *APOE*. Of note, the majority of candidate variants associated with the neuropathologic changes assessed were not associated with ADNC alone; indeed, 24 variants associated with ADNC were also associated with one or more ADRDs, and 24 variants were associated with ADRDs but not ADNC (**Figure 2**). Eight variants were associated with LBD presence or severity (*SORL1*, *SC1MP*, *IGH* gene cluster, *ADAM17*, *BIN1*, *SPI1*, *TMEM106B*, and *USP6NL*), 3 with CAA severity (*TPCN1*, *MINDY2*, and *EED*) and an additional 7 with presence of CAA (*SORL1*, *COX7C*, *IGH* gene cluster, *CLU*, *MAF*, *SLC2A4RG*, and *ACE*). Additional nominal association signals were observed with HS, TDP-43, CBVD, and VBI (**Tables S4, S5**).

**Fig. 2.**
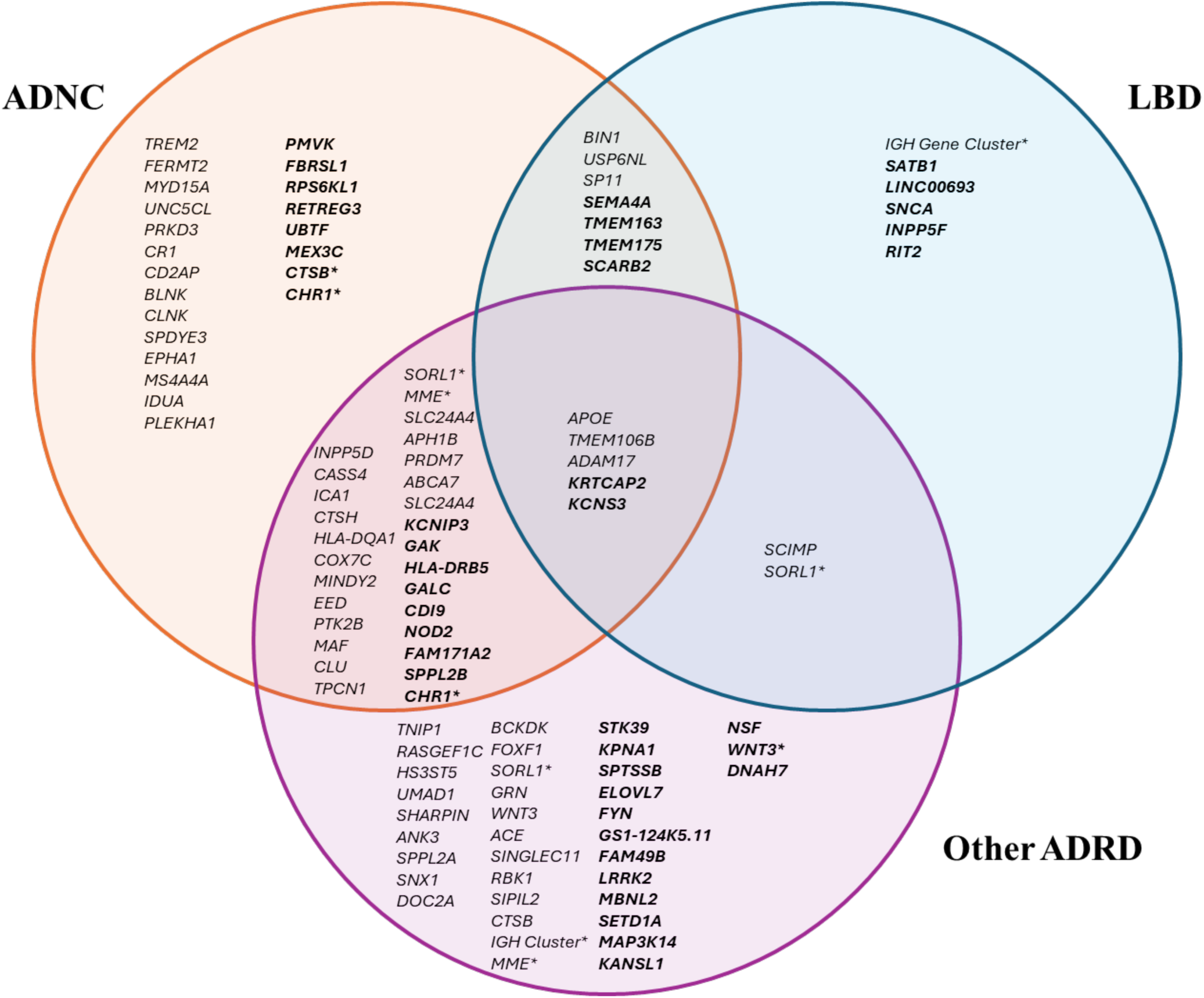
Venn diagram of clinical AD and PD candidate variant analyses. Plain text = AD candidate genes from Bellenguez et al. (2022) GWAS. Bold text = PD candidate genes from Nalls et al. (2019) GWAS. Other ADRD = HS, CAA, CBVD, VBI, or TDP-43 proteinopathy. *Genes with multiple variants represented within the diagram Abbreviations: AD, Alzheimer’s disease; ADRD, AD related dementias; CAA, cerebral amyloid angiopathy; CBVD, cerebrovascular disease; PD, Parkinson’s disease; VBI, vascular brain injury

#### PD-candidate genetic variant associations with ADNC and ADRD lesions

Mounting clinical, pathological, and biochemical data support some degree of overlap between AD and PD. For this reason, we examined the 90 previously identified PD clinical variants by Nalls *et al.*^31^ for association with the pathologic endophenotypes for AD and ADRD. Of these, 11 were associated with LBD (*INPP5F*, *LINC00693, SEMA4A*, *TMEM163*, *SNCA*, *RIT2*, *SCARB2, TMEM175*, *KCNS3*, *SATB1,* and *KRTCAP2*) in our analyses; most of these signals were robust against changes in how LBD was categorized (e.g., 5 ordinal ranks, 3 ordinal ranks, or presence/absence) (**Table S6**). Several PD-candidate variants also were associated with ADNC (**Fig.2, Table S7**), including 8 associated with A score, 10 associated with B score, 11 associated with C score, and 4 associated with ADNC ABC score. The PD candidate variants also were associated with other ADRD pathologic features, including CAA severity (5 variants), CAA presence (9 variants), HS (10 variants), TDP-43 proteinopathy presence (4 variants), and TDP-43 severity (9 variants) (**Table S8**).

### SNP heritability of ADNC and ADRD pathologic features

#### Heritability and genetic correlations

We performed SNP heritability analyses using linkage disequilibrium (LD) score regression (LDSC) on the summary statistics from the above GWAS analyses. AP severity by Thal phasing showed the highest SNP heritability (h^2^: 0.78; **Table 4**), neuritic plaque severity by CERAD score had intermediate heritability (h^2^: 0.24), and NFT severity by Braak staging had lowest heritability (h^2^: 0.19) among ADNC. TDP-43 proteinopathy measures (h^2^:0.21-0.41) and presence of CAA (h^2^ = 0.26) also had intermediate heritability. Other ADRD pathologic features (LBD, CBVD, and VBI) tended to have lower heritability (h^2^ < 0.20). While most of the heritability estimates were significantly larger than zero, many had large standard errors and wide confidence intervals (**Table 4**; full results in **Table S9**). Phenotypic correlations (**Fig S1**) motivated the assessment of genetic correlations among traits with significant heritability. These analyses suggested shared genetic architecture of CAA and presence of AP (*P* = 0.04 for presence of CAA and presence of AP; *P* = 0.044 for severity of CAA and presence of AP). However, overall, the genetic correlation estimates tended to be unstable (i.e., high variance on correlation estimates, leading to wide confidence intervals), suggesting the need for analysis with larger sample sizes (**Table S10**). Additional heritability analyses are described in the supplemental material (Supplemental Text; **Fig S17, S18; Tables S11 - S14**); there, we describe the assessment of cell and tissue-specific polygenic effects for neuropathologic changes.^32^ That is, we assess whether the SNP heritability estimates noted above are concentrated or enriched in specific cell or tissue types. Referent cell types were generated by a genome-wide study of tissue-specific gene expression in humans (GTEx), and a study of immune cell types in mice (ImmGen).^32,33^

**Table 4.** SNP heritability analyses.

| Category | Phenotype | h<sup>2</sup> | SE | CI | P-value |
| --- | --- | --- | --- | --- | --- |
| ADNC | ADNC ABC Score | 0.554 | 0.165 | 0.231, 0.877 | 7.7 × 10<sup>-4</sup> |
|  | CERAD NP Score | 0.242 | 0.059 | 0.126, 0.358 | 4.4 × 10<sup>-5</sup> |
|  | Thal Phase | 0.788 | 0.219 | 0.358, 1.218 | 3.3 × 10<sup>-4</sup> |
|  | NFT Braak Stage | 0.192 | 0.054 | 0.086, 0.298 | 4.0 × 10<sup>-4</sup> |
| CBVD | Arteriolosclerosis (any) | 0.163 | 0.075 | 0.017, 0.309 | 2.9 × 10<sup>-2</sup> |
|  | Atherosclerosis (any) | 0.189 | 0.073 | 0.046, 0.332 | 9.5 × 10<sup>-3</sup> |
|  | CAA (any) | 0.256 | 0.058 | 0.143, 0.37 | 9.6 × 10<sup>-6</sup> |
|  | Any CBVD | 0.202 | 0.127 | -0.047, 0.451 | 1.1 × 10<sup>-1</sup> |
| VBI | Hemorrhages | 0.251 | 0.203 | -0.147, 0.649 | 2.2 × 10<sup>-1</sup> |
|  | Infarcts/lacunes | 0.131 | 0.065 | 0.005, 0.258 | 4.2 × 10<sup>-2</sup> |
|  | Microinfarcts | 0.111 | 0.069 | -0.023, 0.246 | 1.1 × 10<sup>-1</sup> |
|  | VBI (any) | 0.046 | 0.045 | -0.043, 0.134 | 3.1 × 10<sup>-1</sup> |
|  | WMR (any) | 0.212 | 0.103 | 0.01, 0.414 | 4.0 × 10<sup>-2</sup> |
| LBD | LBD (any/none) | 0.131 | 0.045 | 0.043, 0.219 | 3.6 × 10<sup>-3</sup> |
|  | TDP-43 (any) | 0.410 | 0.128 | 0.159, 0.66 | 1.4 × 10<sup>-3</sup> |
| HS | HS (any) | 0.123 | 0.083 | -0.04, 0.285 | 1.4 × 10<sup>-1</sup> |

Analyses used Model 2 throughout, excluding *APOE* region; liability scale; intercept constrained to 1.
Shaded p-values indicate significance ( $p < 0.05$ ).
Abbreviations: **AD**, Alzheimer's disease; **ADNC**, AD neuropathologic change; **CAA**, cerebral amyloid angiopathy; **CI**, confidence interval; **CERAD**, Consortium to Establish a Registry for AD neuritic plaque score; **CBVD**, cerebrovascular disease; **HS**, hippocampal sclerosis; **LBD**, Lewy body disease; **SE**, standard error; **TDP-43**, TAR DNA-binding protein 43; **VBI**, vascular brain injury; **WMR**, white matter rarefaction

### Pathway and set-based tests

Pathway and set-based analyses were performed using MAGMA enrichment analysis (**Tables S15-S17**). The pathway analyses (GO, Biocarta, etc) yielded results overlapping with known AD pathways (**Table S15**). Top pathways for AD-related neuropathology phenotypes were dominated by immunity-related pathways (e.g., association between the Reactome pathway “Activation of RAS in B cells” and Thal Phase; p-value = 6.1 ^x^ 10^-7^). Other known AD pathways (cell signaling, amyloid, lipid metabolism) were also present, though with more nominal associations. Additional tissue-specific analyses are also included, based on MAGMA rather than LDSC (**Tables S16, S17**).

## Discussion

We present results from the largest GWAS of AD and ADRD neuropathologic lesions conducted to date, using data from several cohorts that collected research quality clinical, neuropathologic, and genetic data. We confirmed significant association of *APOE* with both hallmark lesions of AD as well as with all other ADRDs excepting cerebrovascular arteriolosclerosis, cerebrovascular atherosclerosis, and VBI. Furthermore, we mapped ten significant loci to ten disease-specific hallmark lesions, including five loci previously associated with AD dementia (*BIN1, PICALM/EED, TMEM106B, GRN,* and *SNCA/SNCA-AS1*) and five novel loci (*EPHA5, PSMG1, LINC00276, VAPA,* and *DOCK4*). While the largest study of its type, we recognize that the size of our cohort is still modest for GWAS. For this reason, we also analyzed clinical AD and PD GWAS candidates and found substantial overlap of candidate variants with multiple ADNC and ADRD lesions. Finally, we found variable heritability among ADNC and ADRD lesions.

### GWS associations with neuropathologic hallmarks

The *APOE* region was significantly associated with all five proteinopathies – AP, CAA, NFT, LBD, and TDP-43 inclusions - validating a broad role for *APOE* variants in AD and commonly comorbid ADRD.^25,34–36^ Indeed, associations between *APOE* and clinical AD GWAS may be especially strong because its association with pathologic features of both ADNC and ADRD. Many proposals, spanning from co-seeding of protein aggregates to shared influences of the biology of aging, have been offered to explain why LBD and TDP-43 inclusions co-occur with ADNC more commonly than expected by chance alone.^37–39^ Our data support that the increased likelihood of co-occurrence among ADNC, LBD, and TDP-43 inclusions is at least in part because these distinct pathologic features share genetic risk at the *APOE* locus. Although functional validation prior to drawing any conclusions as to the clinical implications of these findings will be vital, these results suggest that explorations into potential treatments targeting ApoE variants may be appropriate not only in AD but also multiple ADRDs. Interestingly, other than CAA, our results show only nominal association between the *APOE* locus and CBVD, and no association with forms of VBI. This could suggest a difference in the influence of ApoE isoforms in central vs. peripheral circulations. ^40^

Consistent with previous smaller autopsy GWAS, we observed strong GWS association between *BIN1* variants and NFT measures but weak GWS associations between *BIN1* and *PICALM / EED* variants with NP measures.^21,25,41^ We also identified two novel GWS variants associated with presence of APs on *PSMG1* and *EPHA5*. Variants on *PSMG1* (also known as Down Syndrome Critical Region Gene 2) have been implicated in several cardiometabolic phenotypes.^42–48^ Such associations, together with appropriate functional genetic follow-up, may provide a biological context for exploring the connection between *PSMG1* and AD/ADRD, and may help to identify future potential vascular therapeutic targets. Variants in *EPHA5* have been associated with general cognitive ability and are in the same ephrin receptor family as *EPHA1*,^49,50^ a known clinical AD-associated genetic risk factor.^51,52^ Given the role of ephrin receptors in cell and axon guidance and in synaptic development and plasticity^53,54^, *EPHA5* represents a biologically plausible target through which modulation of ephrin receptor pathways could support synaptic resilience in AD/ADRD. It is important to note that these associations require replication, and functional validation is needed to confirm mechanistic relevance and inform potential future therapeutic development.

Our data suggest that *VAPA* and two long noncoding RNA (*LINC00276*, *LINC00290*) may be associated with CBVD. Variants in *LINC00276* are associated with multiple cardiometabolic, hematological, and immune measures, including platelet, monocyte, and lymphocyte counts, waist-to-hip ratio.^55^ *LINC00290*, similarly, has been associated with immune phenotypes, as well as age of onset for AD.^56^ For macroinfarcts, our GWAS identified a significant association with variants at *DOCK4*, which participates in the transport of low-density lipoproteins.^57^ *DOCK4* has also been associated with multiple immune and cardiovascular phenotypes. ^50,55,57–62^

### Clinical candidate variants

Our candidate-variant approach based on clinical AD GWAS^30^ revealed 38 of 78 candidate variants associated with one or multiple ADNC features. Once validated, these may represent targets for translating these genetic risk associations into therapeutics. For example, *CLU* and *FERMT2* were both associated with APs but not NFTs, and their encoded proteins alter Aβ aggregation and clearance^63^ as well as APP metabolism and Aβ peptide production.^64^ Of note, 11 of the clinical AD GWAS candidate variants did not associate with any ADNC but instead were associated with one or more ADRDs, and 19 clinical AD GWAS variants that were associated with one or more ADNCs also were associated with one or more ADRD pathologic endophenotypes. These results provide compelling support for the heterogeneous nature of clinical AD, such that multiple pathways may lead to similar clinical outcomes. Interestingly, 40 candidate variants were not associated with any of the ADNC lesions. This last group is especially important because they may provide novel insights into largely unexplored hallmark-independent potential therapeutic approaches as well as resilience factors that suppress clinical expression of disease without altering pathologic hallmarks.

We do note some overlap between the current study and prior CSF and imaging GWAS. For example, *BIN1* has been reportedly associated with both CSF Aβ42 and tau, but not with amyloid PET.^18,19^ This suggests that *BIN1* may act early in the disease process to influence both Aβ42 and tau, and over time contribute to the accrual of plaques and tangles, while mid-stage markers may not capture the phase at which *BIN1* exerts its strongest effects. Alternatively, we report associations with amyloid and *FERMT,* a finding that is supported by amyloid PET, but not CSF, studies, suggesting that the variant on *FERMT* may influence more moderate to late stages of amyloid accumulation. Additional studies to determine stage-dependent genetic architecture of AD and genetic overlap between biomarker modalities will ultimately be needed to further understand these relationships and facilitate identification of treatment targets.

Similarly, a candidate variant approach for clinical PD^31^ found significant candidate associations with LBD, but also with ADNC, CAA, HS, and TDP-43. Of the 103 clinical PD candidate variants assessed 11 were nominally associated with LBD. However, 23 were associated with ADNC and 29 with ADRD lesions (CAA, HS, TDP-43 proteinopathy). For example, *KRTCAP2* was associated with LBD, but also nominally associated with measures of AP, NFT, and TDP-43 proteinopathy. These results further underscore how ADNC and ADRD lesions share partial genetic overlap with clinically diagnosed AD and PD.

### Heritability of neuropathology

Our heritability analyses suggest a strong polygenic effect for AP severity with high SNP heritability (up to 78%) relative to GWAS of clinical AD dementia (e.g., 9-30%).^65,66^ Heritability estimates for presence of TDP-43 inclusions was intermediate (41%) while NFT severity and CBVD measures showed relatively low heritability (11–26%) with presence of CAA, another form of amyloidosis, being the highest. This is consistent with moderate heritability of cardiovascular traits, and the higher heritability of amyloid measures.

### Limitations

There are several potential limitations to our GWAS of ADNC and ADRD lesions. First, while this study represents the largest brain autopsy AD GWAS to date, it is still limited in size compared to the clinical AD GWAS that surpasses one million individuals. Because of this, we cannot resolve whether the lack of association of some candidate variants to neuropathologic hallmarks is due to low statistical power or to the variants associated with hallmark-independent processes. Second, effect-size heterogeneity is of some concern for this study, with sample sets coming from a variety of sources and genotyping arrays. Meta-analyses such as these tend to be robust against false positives due to heterogeneity. However, such issues could contribute to reduced statistical power. Third, this study lacks racial and ethnic diversity among participants. Genetic variants may have different frequencies and/or relevance in other populations, and thus risk prediction based on these studies may not translate to non-European populations. The lack of generalizability to wider populations highlights the need for improved outreach and diversity in brain autopsy programs. Fourth, our explorations into AD and PD clinical candidate variants revealed several nominal associations, raising questions about multiple testing correction, and appropriate modeling. The correlated nature of some of the neuropathological phenotypes (**Figure S1**) may allow for improved efficiency and power with more complex multivariate modeling (e.g., MANCOVA), at least for some ensemble hypotheses; this will be the subject of future studies. Similarly, the multiple-testing correction of the candidate variants is worthy of consideration. Typically, multiple-testing corrections are relaxed when there is strong *a priori* evidence for the hypothesis. In this case, we have strong evidence for those loci from the clinical AD and PD GWAS, reducing the need for correction, for this subset of analyses. It is important to note here that these analyses are by nature exploratory and require independent replication due to the relatively small sample size in the context of GWAS. The wide confidence intervals noted in many of the heritability and genetic correlation estimates are likewise likely related to relatively small sample size, again highlighting the need for additional confirmation studies. Fifth, it is of note that white matter phenotypes (rarefaction), while commonly reported, do not yet have consensus protocols and reporting procedures. As such, we largely relegated analyses related of these phenotypes to supplemental material. Finally, while additional replication of our GWS associations is needed, we note that the meta-analysis methodology used here serves as a form of replication.

## Conclusion

Clinical AD GWAS are well-powered to identify candidate genes yet leave unaccounted the impact of common co-morbidities that potentially limit the translatability of findings. By offering greater pathologic specificity, GWAS-identified loci of neuropathologic endophenotypes, once validated, may provide important insights into potential disease-relevant pathways and enable the nomination of genetic and molecular biomarkers that may serve as targets for disease detection and intervention. Our study underscores a spectrum of genetic risk that is partially shared, most notably for *APOE* and *TMEM106B* variants, and partially distinct across pathologically verified AD and ADRD, providing a potential explanation for how these classically distinct clinical entities share neuropathologic features that co-occur more commonly than by chance alone. Although autopsy studies are limited by sample size, and thus genes not associated with any pathologic endophenotype might be underpowered for discovery in our cohort, the associations revealed here provide an important basis for further discovery. Alternatively, genes not identified here may be associated with neuropathologic hallmark-independent or yet to be discovered disease mechanisms, an important caveat for ongoing investigations into the genetic architecture of dementia.

## Methods

The methodology of the study is summarized and outlined in **Figure 3**.

**Fig. 3.**
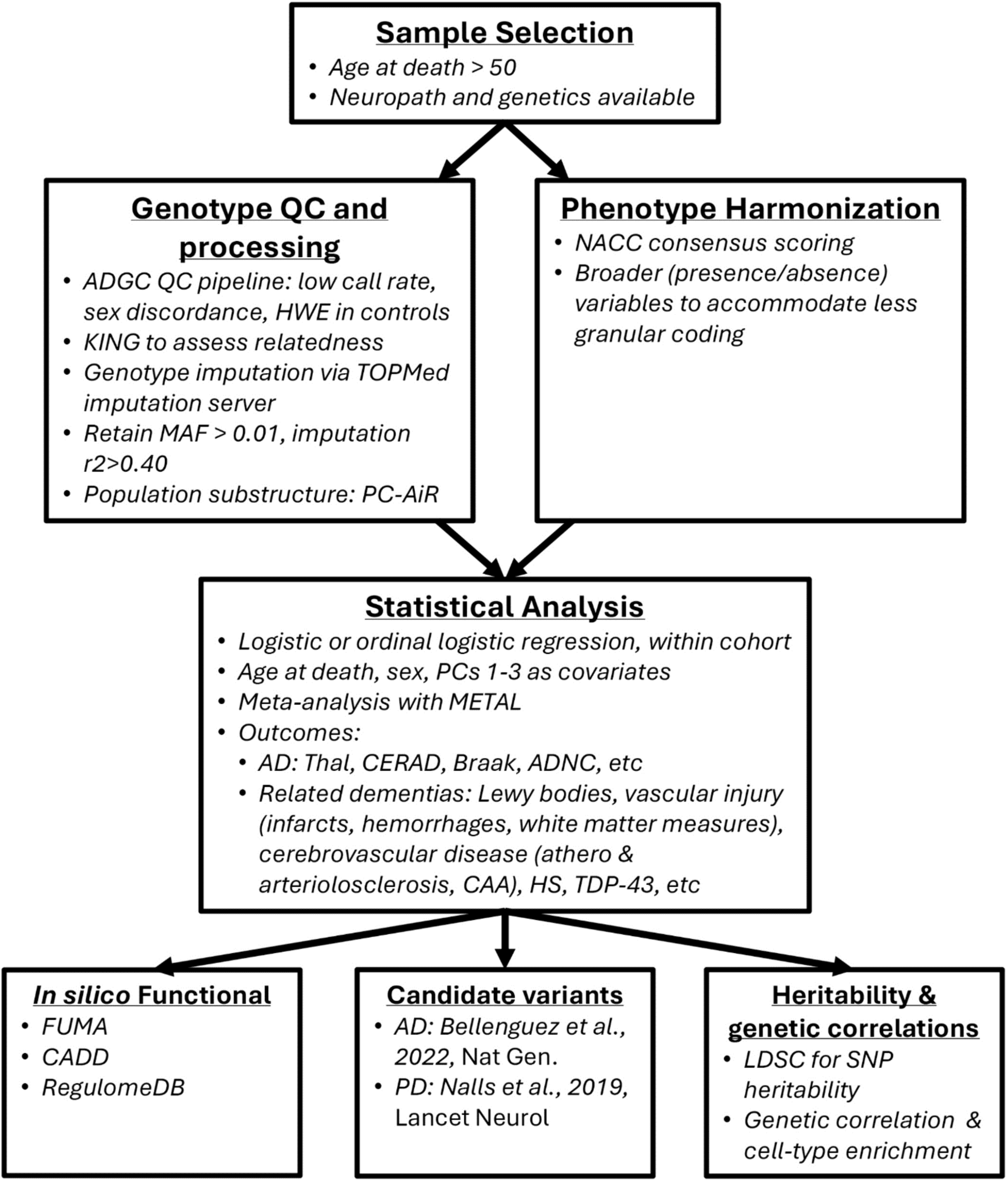
Overview of Primary Analyses.

### Sample acquisition and selection

#### Participant selection

Participants were selected from the ADGC (G.D. Schellenberg, PI) and affiliated studies. Contributing studies include the National Institute on Aging (NIA) funded AD Research Centers (ADRCs), the Adult Changes in Thought Study (ACT; L.K. McEvoy, P. Crane, A.Z. LaCroix, PIs), the Religious Orders Study/Memory and Aging Project (ROSMAP; David Bennett, PI), NIA Late-Onset AD Family Study (NIA-LOAD; Richard Mayeux, PI), Translational Genomics Research Institute (TGen; Eric Reiman, MD), the University of Pittsburgh ADRC (Ilyas Kamboh, PI), the Neurological Tissue Bank at the Biobank-Hospital Clínic, Instituto de Investigaciones Biomédicas August Pi i Sunyer, Barcelona (IDIBAPS; Laura Molina Porcel, PI), the Mayo Clinic ADRC (R.C. Petersen, PI), and the University of Miami Hussman Institute for Human Genomics (AD, Margaret Pericak-Vance, PI; Parkinson’s Disease, Jeffrey Vance, PI). The samples were predominantly from clinical dementia patients (Table 1), mostly with AD as primary suspected etiology. This reflects both the focus of contributing studies (largely AD dementia and cognitive aging) as well as bias in participation in autopsy programs. IDIBAPS is an autopsy-based study with no clinical data on participating samples (beyond sex and age at death). ADRC phenotypic data were obtained through the National Alzheimer’s Coordinating Centers (NACC; Walter Kukull, PI). All participants (or representatives) provided written informed consent; all protocols and assessments were performed with approval by the appropriate institutional internal review boards of the contributing studies. The study here reported is a secondary data analysis study conducted with approval of the Wake Forest University School of Medicine IRB.

#### Inclusion criteria

ADGC participants were included if age at death was greater than 50 years and both neuropathologic data and genetic array data were available. Participants with dementia whose primary dementia etiology was determined to be non-AD or non-ADRD (e.g., traumatic brain injury, chronic drug/alcohol use, etc.) were excluded.

### Genotyping and quality control

Genotyping of the ADC sets was performed at the Children’s Hospital of Pennsylvania, but genotyping chips differed across ADCs. For smaller sample sets we combined like chips into batches after initial QC and imputation (**Table 1**). Genotyping of ADGC-collaborating sets was performed on a variety of genotyping platforms and is described in elsewhere.^11,52^ IDIBAPS participant samples underwent genotyping with the Illumina NeuroBooster array at the University of Miami. Imputation quality (r2) for genome-wide significant loci are noted in the supplementary material (**Table S18**).

The standardized ADGC quality control pipeline was performed on the sample and variant level, detailed elsewhere.^11,52^ Briefly, samples or variants with low call rates (sample missingness > 2%; variant missingness > 5%), sex discordance, or deviations from Hardy-Weinberg Equilibrium (*P*_HWE_<10^-6^ among controls) were dropped. Relatedness checks were performed with the KING algorithm from the SNPRelate package.^67,68^ The analysis revealed identical pairs (kinship ≥ 0.480) that were subsequently dropped. For pairs with lower kinship (0.177 ≤ kinship < 0.480), only one individual from each related pair was kept, whichever belonged to the larger dataset. If that was accounted for, the sample with the most complete (non-missing) neuropathology phenotype information was kept. The samples were imputed with the Trans-Omics for Precision Medicine program server (r2).^69^ Genetic variants with minor allele frequency (MAF) ≥ 0.01 and imputation quality score R2 ≥ 0.40 were used for analysis. When like sets were combined, MAF and R2 scores were weighted by comprising sets. After imputation, principal components analysis was conducted using PC-AiR to assess and account for population substructure.^70^ Outliers for genetic ancestry (>6 standard deviations from mean within any of the first ten PCs) were dropped.

### Phenotype harmonization

To the extent possible, we used consensus methods for neuropathologic lesion assessment and scoring,^27,71,72^ as we have done previously (**Table S19**).^21^ When scoring methods were incompatible with consensus methods, we first restricted categorization to achieve compatibility or, when that was not achievable, we created less granular categorizations to accommodate the incompatible coding or less distinct codings.^21^ For example, LBD was categorized in three different ways: grouped into five categories (0 = none, 1 = olfactory or unspecific region, 2 = brainstem predominant, 3 = limbic, and 4 = neocortical), grouped into 3 categories (0 = none, 1 = olfactory, unspecific region, or brainstem predominant, and 2 = limbic/neocortical), and binary to insure compatibility across datasets while retaining the largest possible sample set. Meta-analyses were performed on seven AD phenotypes and 19 ADRD phenotypes, with some phenotypes representing derivations of others (primarily ordinal phenotypes reduced to an any/none binary phenotype) (**Table S19**).

### Statistical analyses

Unless noted, statistical analyses were conducted in R version 4.2.2.^73^ Spearman correlation and the corrgram R package were used to assess correlations.^74^

#### Single variant GWAS meta-analysis

GWAS was conducted with either logistic or ordinal logistic regression model testing for effects of dosages of imputed genotypes and adjusting for age at death, sex, and the first three principal components within dataset/batch. Regression for GWAS was performed using RVtests for binary endpoints or the “ordinal” package in R for ordinal endpoints, on each of the available datasets.^75,76^ Batch-specific results were then combined across datasets in a fixed-effect meta-analysis with an inverse-variance weighted approach, as implemented in METAL, excluding genetic variants appearing in less than 30% of datasets.^77^ QQ plots were generated along with genomic inflation factors. As a sensitivity analysis, we also conducted GWAS adjusting for *APOE* ε4 count. These results were very similar to the primary analyses, besides the lack of genome-wide signal from variants in *APOE*.

QQ plots and genomic inflation factor (*λ*) were generated in R using the “qqman” package to assess possible inflation from false positives and excluded the *APOE* region (chr19:44-46Mb).^78^ Manhattan plots to visualize the GWAS meta-analyses were created using the “ggplot2” package in R.^79^ Figures for regional association signals were created with LocusZoom.^80^

#### Functional analysis

Top signals from GWAS meta-analyses were followed up for functional assessment using scoring and eQTL databases via in Functional Mapping and Annotation (FUMA; v1.5.2),^81^ combined annotation-dependent depletion,^82^ and RegulomeDB.^83^ MAGMA, also implemented in FUMA, was utilized for set-based analysis of biochemical pathways and ontology sets, as well as GTEx-based tissue-based sets.

#### AD-candidate variants analysis

We leveraged summary statistics from a recent clinical AD GWAS meta-analysis by Bellenguez *et al.* totaling 111,326 clinically diagnosed or proxy AD cases and 677,663 controls that identified 83 independent GWS lead variants (excluding *APOE*).^30^ We located these variants within our dataset and found 78 variants, dropping 5 variants with either MAF < 0.01, not imputed on our reference panel, or imputed with R^2^ < 0.40. The AD-candidate variants dropped from analysis were on *TREM2, SORT1, TREML2, NCK2,* and *PLCG2.* Additionally, we did not consider any variants on *APOE* for this analysis. Using these 78 AD-candidate variant dosages in our dataset, we tested for association with both AD (A score, B score, C score, and ADNC) and comorbid pathologies (LBD, CAA, HS, and LATE-NC) using either logistic or ordinal logistic regression.

#### PD-candidate variants analysis

A recent meta-GWAS of Parkinson’s disease (PD) by Nalls *et al*. including 37.7K cases, 18.6K UK Biobank proxy-cases (having a first degree relative with PD), and 1.4M controls revealed 107 lead variants with independent GWS associations with PD, of which 90 passed quality control.^31^ We were able to identify 100 of these 107 variants in our dataset, again excluding rare variants (MAF < 0.01) that were not imputed on our reference panel or variants that did not impute well (R^2^ < 0.40), including variants on *GXYLT1*, *FGD4*, *GBA*, *LRRK2*, *PMVK*, *SEMA4A*, and *DPM3*. For each of these 100 variants, we tested for association with three categorizations of LBD. We also tested AD lesion scores (A score, B score, C score, and ADNC) and other common comorbid diseases (CAA, HS, and LATE-NC) for association with PD-candidate variants.

#### Heritability and genetic correlation analyses

We performed heritability and genetic correlation analyses using LD Score Regression (LDSR)^84^ using the LDSC software (https://github.com/bulik/ldsc/). To calculate heritability, we used the *APOE* adjusted summary statistics, with the *APOE* region removed (+/- 500kb of *APOE* coding region). Reference LD scores were computed using the 1000 Genomes European subset (obtained via https://github.com/bulik/ldsc/wiki/, January 2025). The H^2^ intercept was constrained to 1 (--intercept-h2). Prevalences were included (--samp_prev, --pop_prev), with sample prevalence based on our dataset. Since population prevalences for pathology are not widely available, we estimated the pathology prevalences within dementia cases and cognitively intact subsets (using NACC, ROSMAP, and ACT subsets). These dementia/intact-specific prevalences were then weighted by the population-prevalences of dementia (0.13 for this age-range) to estimate the population prevalence of neuropathological lesions. Genetic correlations were also estimated using LDSR (--rg) with the above noted references and prevalences, but without constraining H^2^; the intercept for rg was not constrained to account for sample overlap. Cell-specific enrichment of heritability methodology is reported in supplemental material, together with the results.

## Data availability statement

The authors are unable to share genotype or phenotype data from NACC, ADGC, ROSMAP or ACT due to data use restrictions. ROSMAP data can be requested at https://www.radc.rush.edu and https://www.synapse.org. ADGC data can be requested from NIAGADS at https://www.niagads.org/resources/related-projects/alzheimers-disease-genetics-consortium-adgc-collection. NACC neuropathology data can be requested at https://naccdata.org/. ACT data can be requested at https://actagingresearch.org/. Harmonized neuropathology data are available through NIAGADS at https://dss.niagads.org/datasets/ng00067/. IDIBAPS data can be requested at https://www.clinicbarcelona.org/en/idibaps. TGen data can be requested at https://www.tgen.org/. NIA-LOAD data can be requested at https://www.ncbi.nlm.nih.gov/projects/gap/cgi-bin/dataset

## Acknowledgements

This work was supported in part by the National Institutes for Health (NIH/NIA grant number R01 AG062695).

The Adult Changes in Thought (ACT) study notes support from NIH (U19 AG066567).

The Alzheimer Disease Genetics Consortium notes the following acknowledgements: The National Institutes of Health, National Institute on Aging (NIH-NIA) supported this work through the following grants: ADGC, U01 AG032984, RC2 AG036528; Samples from the National Cell Repository for Alzheimer’s Disease (NCRAD), which receives government support under a cooperative agreement grant (U24 AG21886) awarded by the National Institute on Aging (NIA), were used in this study.

We thank contributors who collected samples used in this study, as well as patients and their families, whose help and participation made this work possible; Data for this study were prepared, archived, and distributed by the National Institute on Aging Alzheimer’s Disease Data Storage Site (NIAGADS) at the University of Pennsylvania (U24-AG041689-01); NACC, U01 AG016976; NIA LOAD, U24 AG026395, R01AG041797; Banner Sun Health Research Institute P30 AG019610; Boston University, P30 AG013846, U01 AG10483, R01 CA129769, R01 MH080295, R01 AG017173, R01 AG025259, R01AG33193; Columbia University, P50 AG008702, R37 AG015473; Duke University, P30 AG028377, AG05128; Emory University, AG025688; Group Health Research Institute, UO1 AG006781, UO1 HG004610, UO1 HG006375; Indiana University, P30 AG10133; Johns Hopkins University, P50 AG005146, R01 AG020688; Massachusetts General Hospital, P50 AG005134; Mayo Clinic, P50 AG016574; Mount Sinai School of Medicine, P50 AG005138, P01 AG002219; New York University, P30 AG08051, UL1 RR029893, 5R01AG012101, 5R01AG022374, 5R01AG013616, 1RC2AG036502, 1R01AG035137; Northwestern University, P30 AG013854; Oregon Health & Science University, P30 AG008017, R01 AG026916; Rush University, P30 AG010161, R01 AG019085, R01 AG15819, R01 AG17917, R01 AG30146; TGen, R01 NS059873; University of Alabama at Birmingham, P50 AG016582; University of Arizona, R01 AG031581; University of California, Davis, P30 AG010129; University of California, Irvine, P50 AG016573; University of California, Los Angeles, P50 AG016570; University of California, San Diego, P50 AG005131; University of California, San Francisco, P50 AG023501, P01 AG019724; University of Kentucky, P30 AG028383, AG05144; University of Michigan, P50 AG008671; University of Pennsylvania, P30 AG010124; University of Pittsburgh, P50 AG0066468, AG0064877, AG007562, AG023651; University of Southern California, P50 AG005142; University of Texas Southwestern, P30 AG012300; University of Miami, R01 AG027944, AG010491, AG027944, AG021547, AG019757; University of Washington, P50 AG005136, P30 AG066509; University of Wisconsin, P50 AG033514; Vanderbilt University, R01 AG019085; and Washington University, P50 AG005681, P01 AG03991.

The Kathleen Price Bryan Brain Bank at Duke University Medical Center is funded by NINDS grant # NS39764, NIMH MH60451 and by Glaxo Smith Kline. Genotyping of the TGEN2 cohort was supported by Kronos Science. The TGen series was also funded by NIA grant AG041232 to AJM and MJH, The Banner Alzheimer’s Foundation, The Johnnie B. Byrd Sr. Alzheimer’s Institute, the Medical Research Council, and the state of Arizona and also includes samples from the following sites: Newcastle Brain Tissue Resource (funding via the Medical Research Council, local NHS trusts and Newcastle University), MRC London Brain Bank for Neurodegenerative Diseases (funding via the Medical Research Council),South West Dementia Brain Bank (funding via numerous sources including the Higher Education Funding Council for England (HEFCE), Alzheimer’s Research Trust (ART), BRACE as well as North Bristol NHS Trust Research and Innovation Department and DeNDRoN), The Netherlands Brain Bank (funding via numerous sources including Stichting MS Research, Brain Net Europe, Hersenstichting Nederland Breinbrekend Werk, International Parkinson Fonds, Internationale Stiching Alzheimer Onderzoek), Institut de Neuropatologia, Servei Anatomia Patologica, Universitat de Barcelona.

ADNI data collection and sharing was funded by the National Institutes of Health Grant U01 AG024904 and Department of Defense award number W81XWH-12-2-0012. ADNI is funded by the National Institute on Aging, the National Institute of Biomedical Imaging and Bioengineering, and through generous contributions from the following: AbbVie, Alzheimer’s Association; Alzheimer’s Drug Discovery Foundation; Araclon Biotech; BioClinica, Inc.; Biogen; Bristol-Myers Squibb Company; CereSpir, Inc.; Eisai Inc.; Elan Pharmaceuticals, Inc.; Eli Lilly and Company; EuroImmun; F. Hoffmann-La Roche Ltd and its affiliated company Genentech, Inc.; Fujirebio; GE Healthcare; IXICO Ltd.; Janssen Alzheimer Immunotherapy Research & Development, LLC.; Johnson & Johnson Pharmaceutical Research & Development LLC.; Lumosity; Lundbeck; Merck & Co., Inc.; Meso Scale Diagnostics, LLC.; NeuroRx Research; Neurotrack Technologies; Novartis Pharmaceuticals Corporation; Pfizer Inc.; Piramal Imaging; Servier; Takeda Pharmaceutical Company; and Transition Therapeutics.

The Canadian Institutes of Health Research is providing funds to support ADNI clinical sites in Canada. Private sector contributions are facilitated by the Foundation for the National Institutes of Health (www.fnih.org).

The grantee organization is the Northern California Institute for Research and Education, and the study is coordinated by the Alzheimer’s Disease Cooperative Study at the University of California, San Diego. ADNI data are disseminated by the Laboratory for Neuro Imaging at the University of Southern California. We thank Drs. D. Stephen Snyder and Marilyn Miller from NIA who are *ex-officio* ADGC members.

Support was also from the Alzheimer’s Association (LAF, IIRG-08-89720; MP-V, IIRG-05-14147) and the US Department of Veterans Affairs Administration, Office of Research and Development, Biomedical Laboratory Research Program. P.S.G.-H. is supported by Wellcome Trust, Howard Hughes Medical Institute, and the Canadian Institute of Health Research.

## Collaborators

Alzheimer’s Disease Genetics Consortium (ADGC): James D Bowen, Paul K Crane, Gail P Jarvik, C Dirk Keene, Eric B Larson, Wayne C McCormick, Susan M McCurry, Shubhabrata Mukherjee, Neil W Kowall, Ann C McKee, Robert A Stern, Clinton T Baldwin, Lindsay A Farrer, Gyungah Jun, Kathryn L Lunetta, Lawrence S Honig, Jean Paul Vonsattel, Jennifer Williamson, Scott Small, Sandra Barral, Christiane Reitz, Badri N Vardarajan, Richard Mayeux, James R Burke, Christine M Hulette, Kathleen A Welsh-Bohmer, Marla Gearing, James J Lah, Allan I Levey, Thomas S Wingo, Liana G Apostolova, Martin R Farlow, Bernardino Ghetti, Andrew J Saykin, Salvatore Spina, Kelley M Faber, Tatiana M Foroud, Marilyn S Albert, Constantine G Lyketsos, Juan C Troncoso, Matthew P Frosch, Robert C Green, John H Growdon, Bradley T Hyman, Rudolph E Tanzi, Huntington Potter, Dennis W Dickson, Nilufer Ertekin-Taner, Neill R Graff-Radford, Joseph E Parisi, Ronald C Petersen, Bradley F Boeve, Mariet Allen, Minerva M Carrasquillo, Steven G Younkin, Ranjan Duara, Joseph D Buxbaum, Alison M Goate, Mary Sano, Arjun V Masurkar, Thomas Wisniewski, Eileen H Bigio, Marsel Mesulam, Sandra Weintraub, Robert Vassar, Jeffrey A Kaye, Joseph F Quinn, Randall L Woltjer, Lisa L Barnes, Lei Yu, Denis A Evans, Victor Henderson, Kenneth B Fallon, Lindy E Harrell, Daniel C Marson, Erik D Roberson, Charles DeCarli, Lee-Way Jin, John M Olichney, Ronald Kim, Frank M LaFerla, Edwin Monuki, Elizabeth Head, David Sultzer, Daniel H Geschwind, Harry V Vinters, Marie-Francoise Chesselet, Douglas R Galasko, James B Brewer, Adam Boxer, Anna Karydas, Joel H Kramer, Bruce L Miller, Howard J Rosen, William W Seeley, Jeffrey M Burns, Russell H Swerdlow, Linda J Van Eldik, Roger L Albin, Andrew P Lieberman, Henry L Paulson, Steven E Arnold, John Q Trojanowski, Vivianna M Van Deerlin, Laura B Cantwell, Amanda P Kuzma, John Malamon, Adam C Naj, Liming Qu, Gerard D Schellenberg, Otto Valladares, Li-San Wang, Yi Zhao, Ronald L Hamilton, M Ilyas Kamboh, Oscar L Lopez, James T Becker, Chuanhai Cao, Ashok Raj, Amanda G Smith, Helena C Chui, Carol A Miller, John M Ringman, Lon S Schneider, Thomas D Bird, Joshua A Sonnen, Chang-En Yu, Thomas Grabowsk, Elaine Peskind, Murray Raskind, Ge Li, Debby W Tsuang, Sanjay Asthana, Craig S Atwood, Cynthia M Carlsson, Mark A Sager, Nathaniel A Chin, Suzanne Craft, Nigel J Cairns, John C Morris, Carlos Cruchaga, Stephen Strittmatter, Eric M Reiman, Thomas G Beach, Matthew J Huentelman, John Hardy, John S K Kauwe, Hakon Hakonarson, Deborah Blacker, Thomas J Montine, William S Bush, Jonathan L Haines, Alan J Lerner, Xiongwei Zhou, Gary W Beecham, Regina M Carney, Michael L Cuccaro, John R Gilbert, Kara L Hamilton-Nelson, Brian W Kunkle, Eden R Martin, Margaret A Pericak-Vance, Jeffery M Vance, Amanda J Myers, James B Leverenz, Philip L De Jager, Mindy J Katz, Richard B Lipton, Valory Pavlik, Paul Massman, Eveleen Darby, Monica Rodriguear, Aisha Khaleeq, Donald R Royall, Alan Stevens, Marcia Ory, John C DeToledo, Henrick Wilms, Kim Johnson, Victoria Perez, Michelle Hernandez, Kirk C Wilhelmsen, Jeffrey Tilson, Scott Chasse, Robert C Barber, Thomas J Fairchild, Sid E O’Bryant, Janice Knebl, James R Hall, Leigh Johnson, Douglas Mains, Lisa Alvarez, Adriana Gamboa, David Paydarfar, John Bertelson, Martin Woon, Gayle Ayres, Alyssa Aguirre, Raymond Palmer, Marsha Polk, Perrie M Adams, Ryan M Huebinger, Joan S Reisch, Roger N Rosenberg, Munro Cullum, Benjamin Williams, Mary Quiceno, Linda Hynan, Janet Smith, Barb Davis, Trung Nguyen, Ekaterina Rogaeva, Peter St George-Hyslop

## Supporting Information Titles and Legends

### Supplemental Text

**Supplemental Text.** The Supplemental Text includes details on additional SNP heritability analyses, including cell and tissue-specific enrichment analyses. Text includes results, discussion, and methodology specific to SNP heritability analyses.

### Supplemental Figures

**Figure S1:**
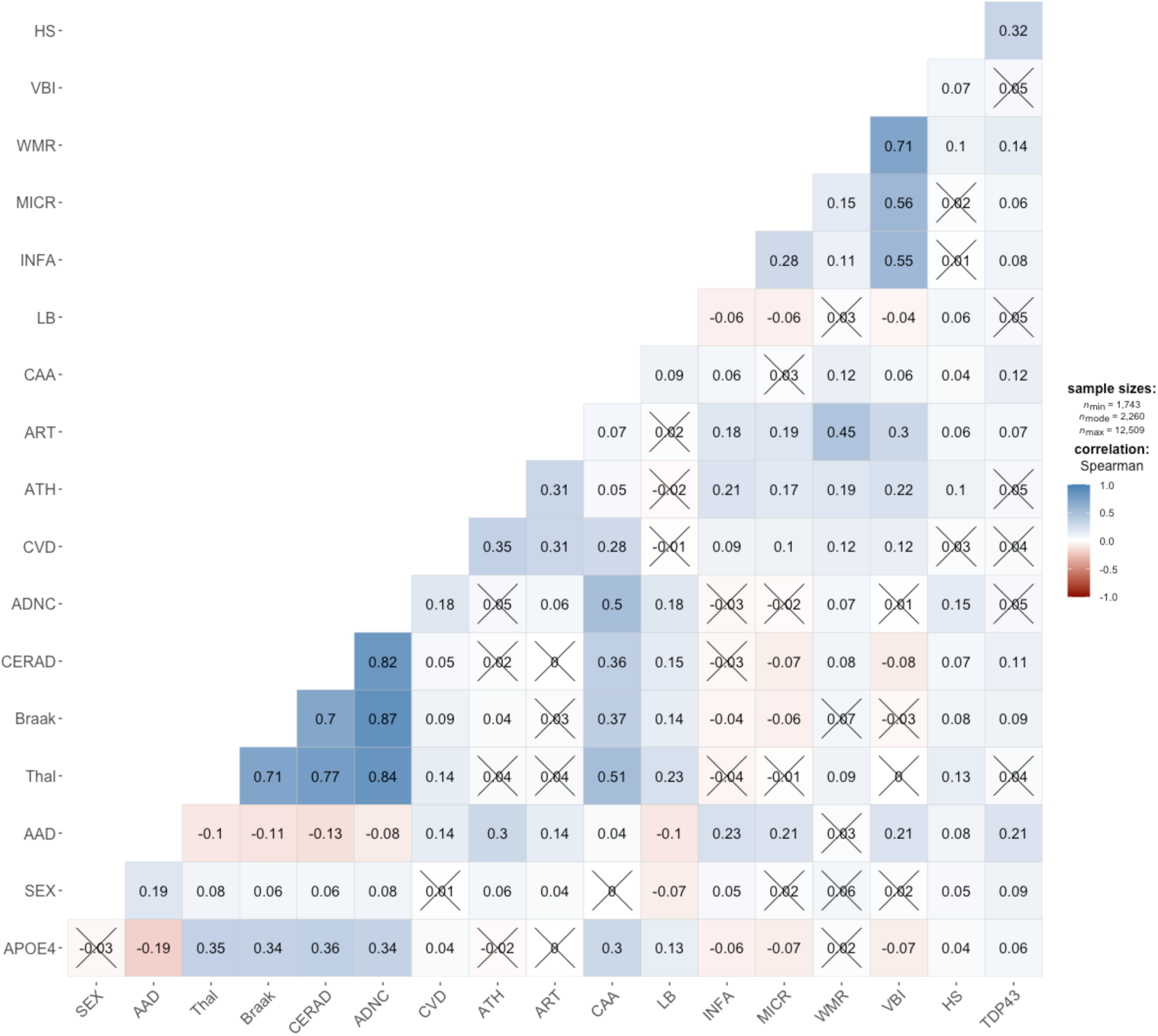
Correlation plot of neuropathology and related variables. Spearman correlation of neuropathology phenotypes and relevant covariates. Acronyms and abbreviations: APOE4, count of e4 allele; AAD, age at death; Thal, Thal Phase; Braak, NFT Braak stage; CERAD, CERAD NP score; ADNC, ADNC score (ABC score); ATH, cerebrovascular atherosclerosis; ART, cerebrovascular arteriolosclerosis; CAA, cerebral amyloid angiopathy; LBD, Lewy body disease; INFA, macroinfarcts/lacunes; MICR, microinfarcts; WMR, white matter rarefaction; VBI, vascular brain injury; HS, hippocampal sclerosis; TDP43, TDP-43 proteinopathy.

**Figure S2:**
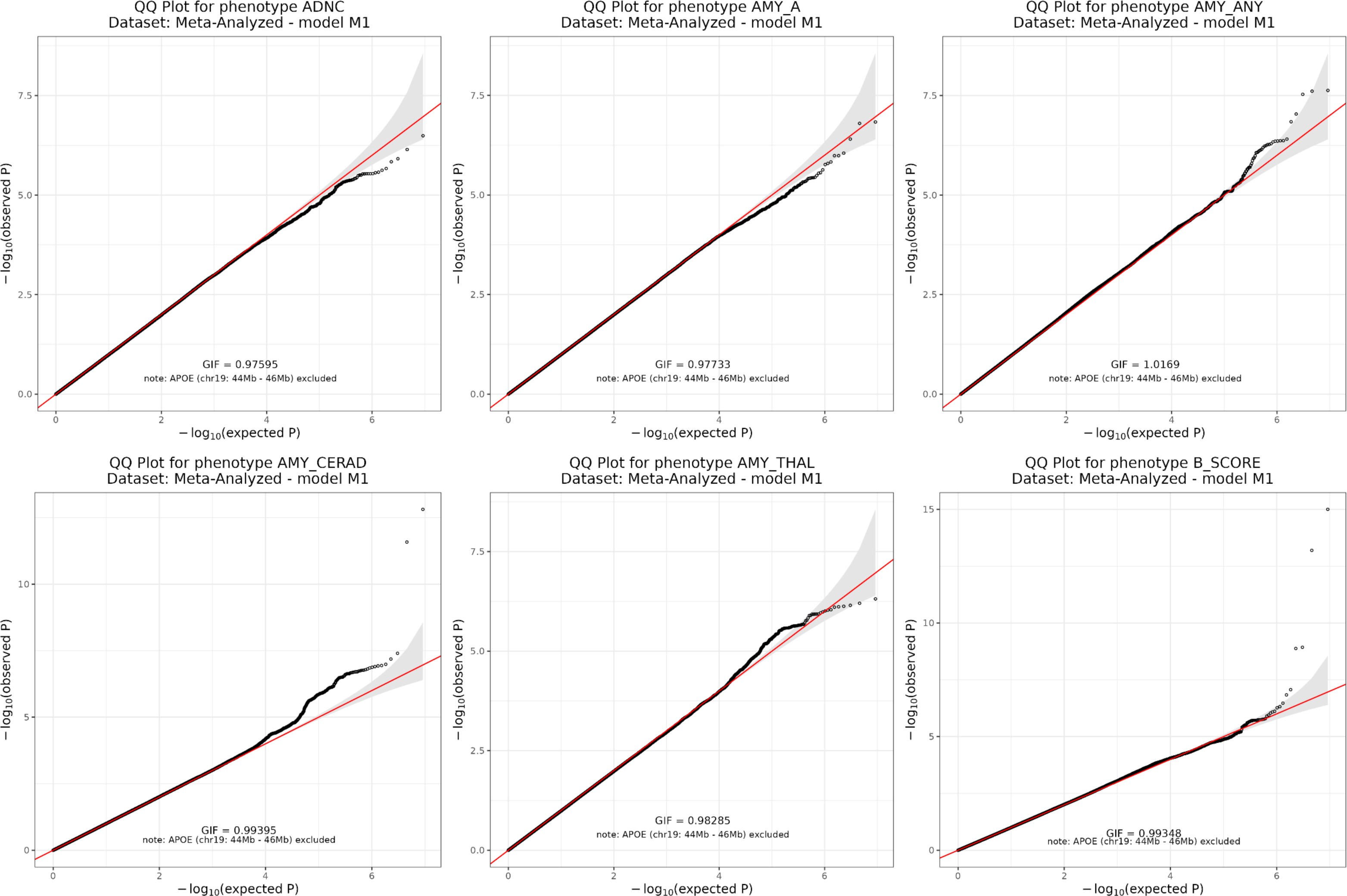
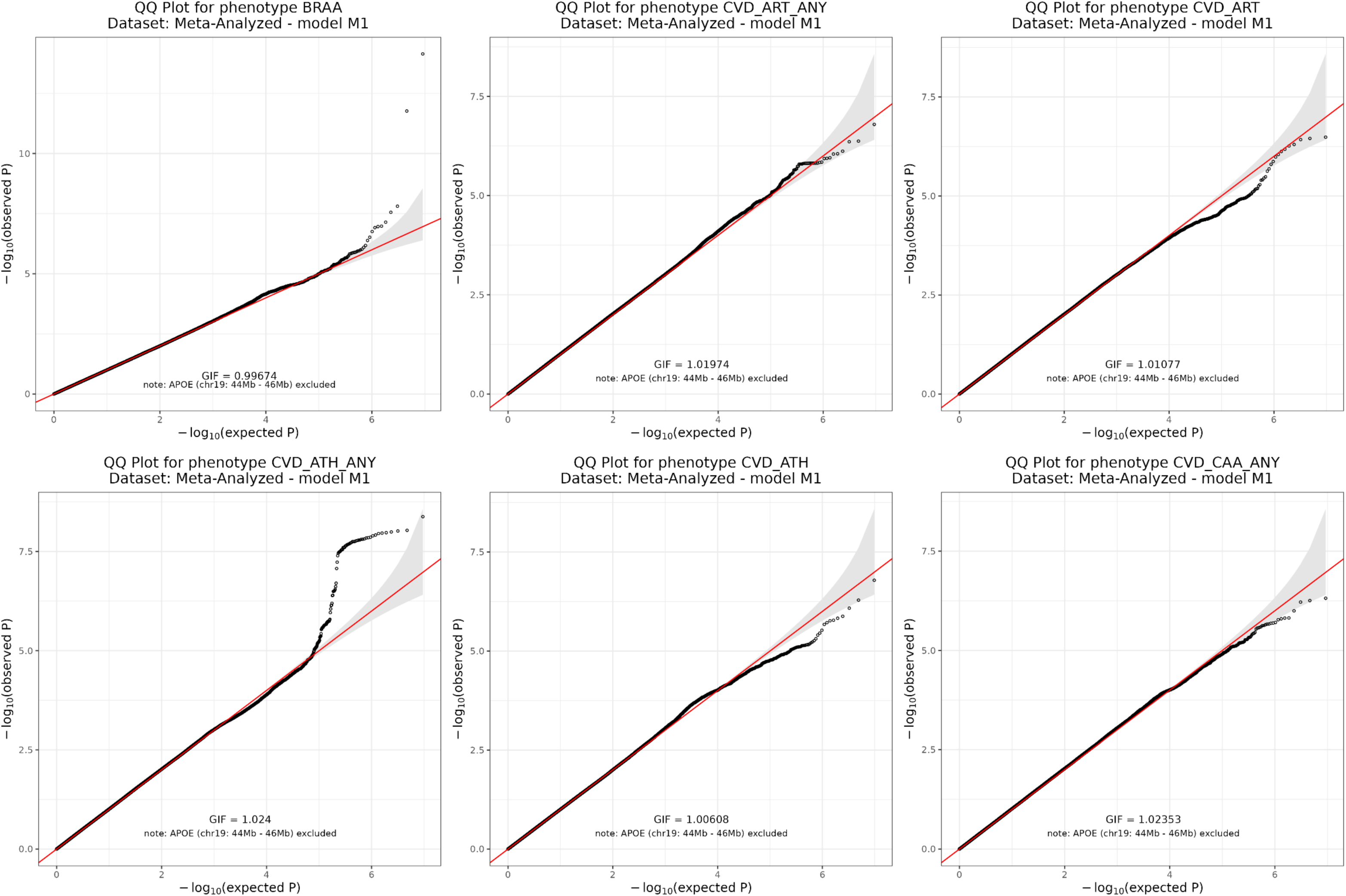
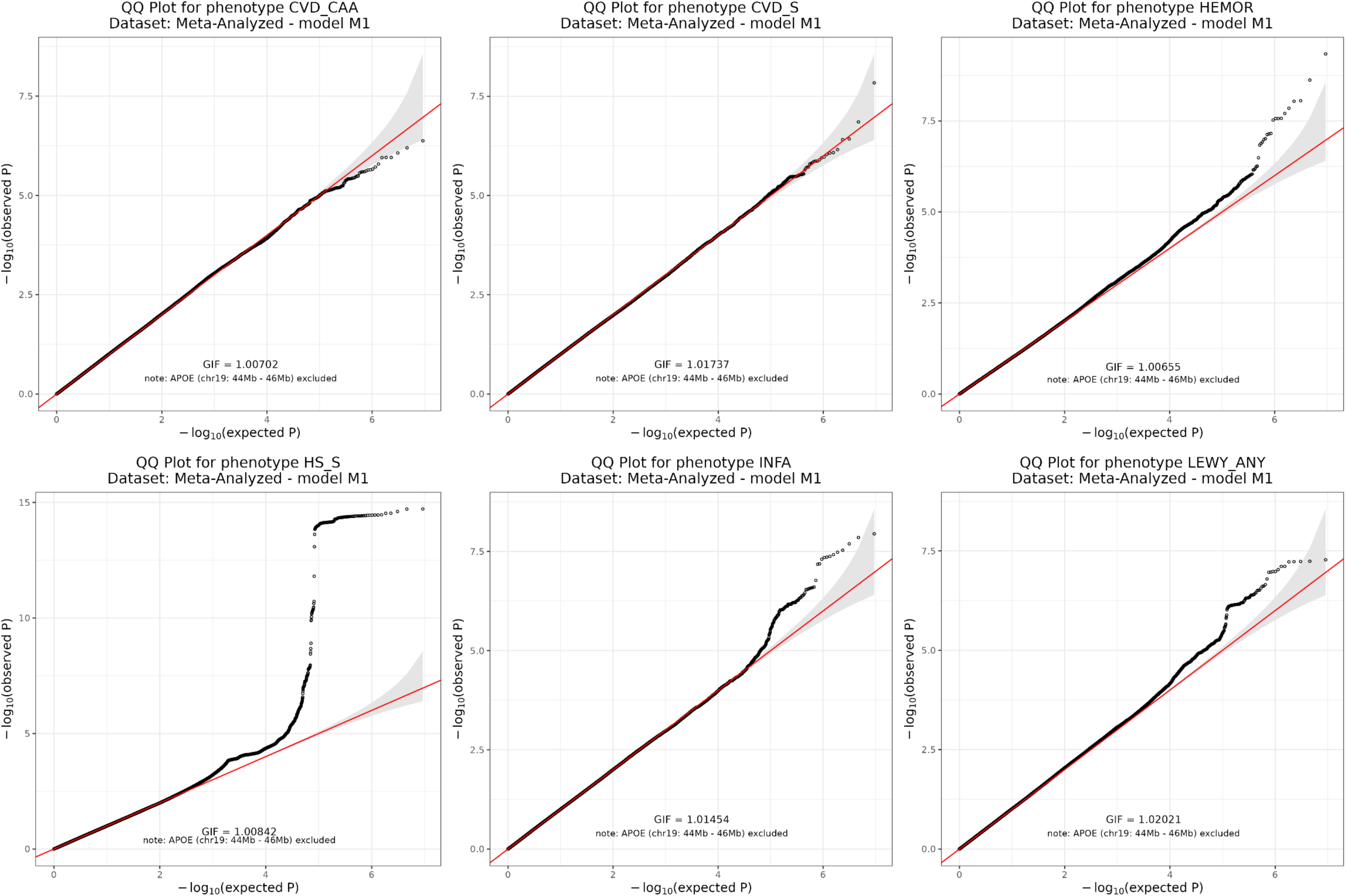
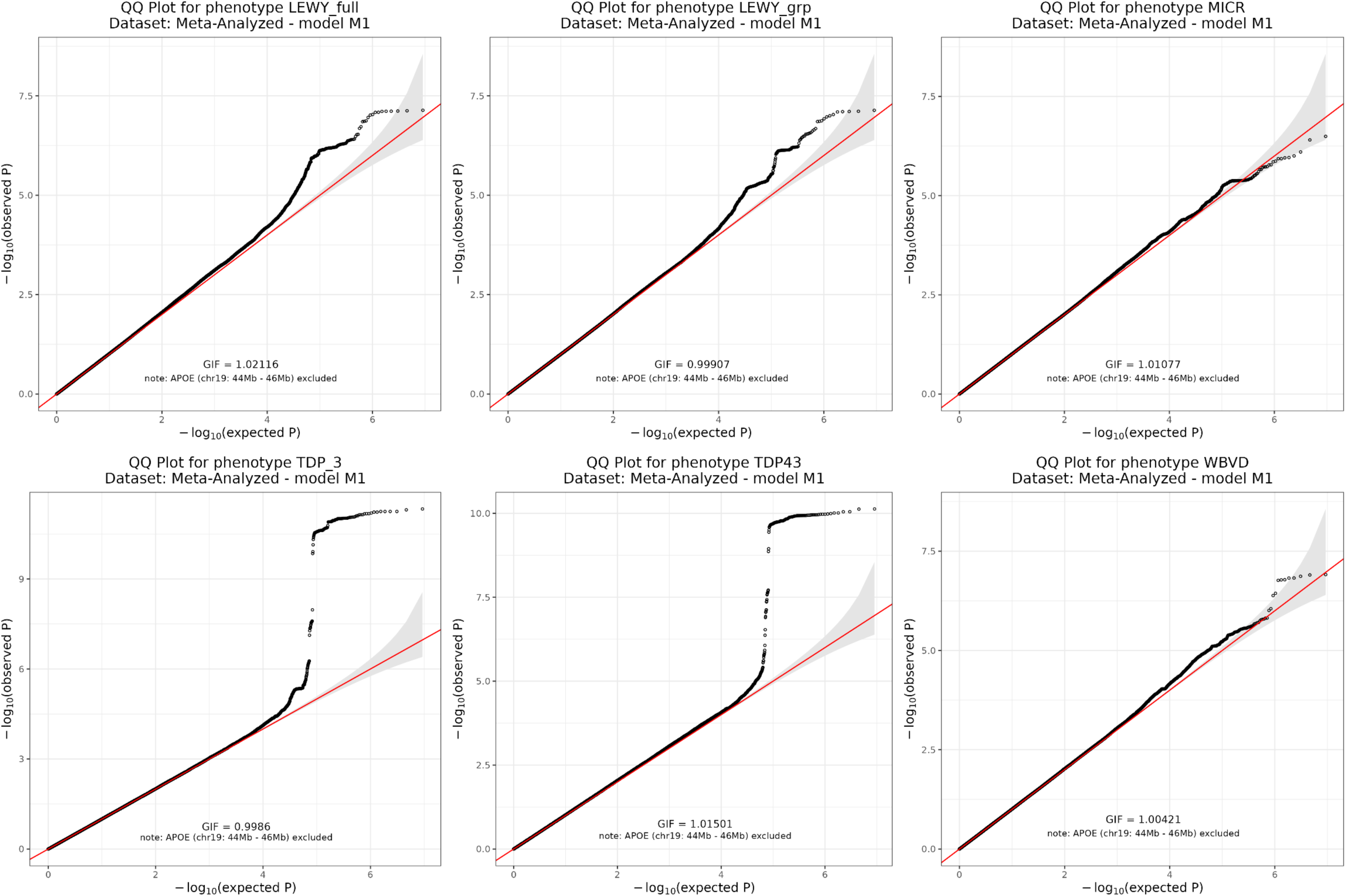
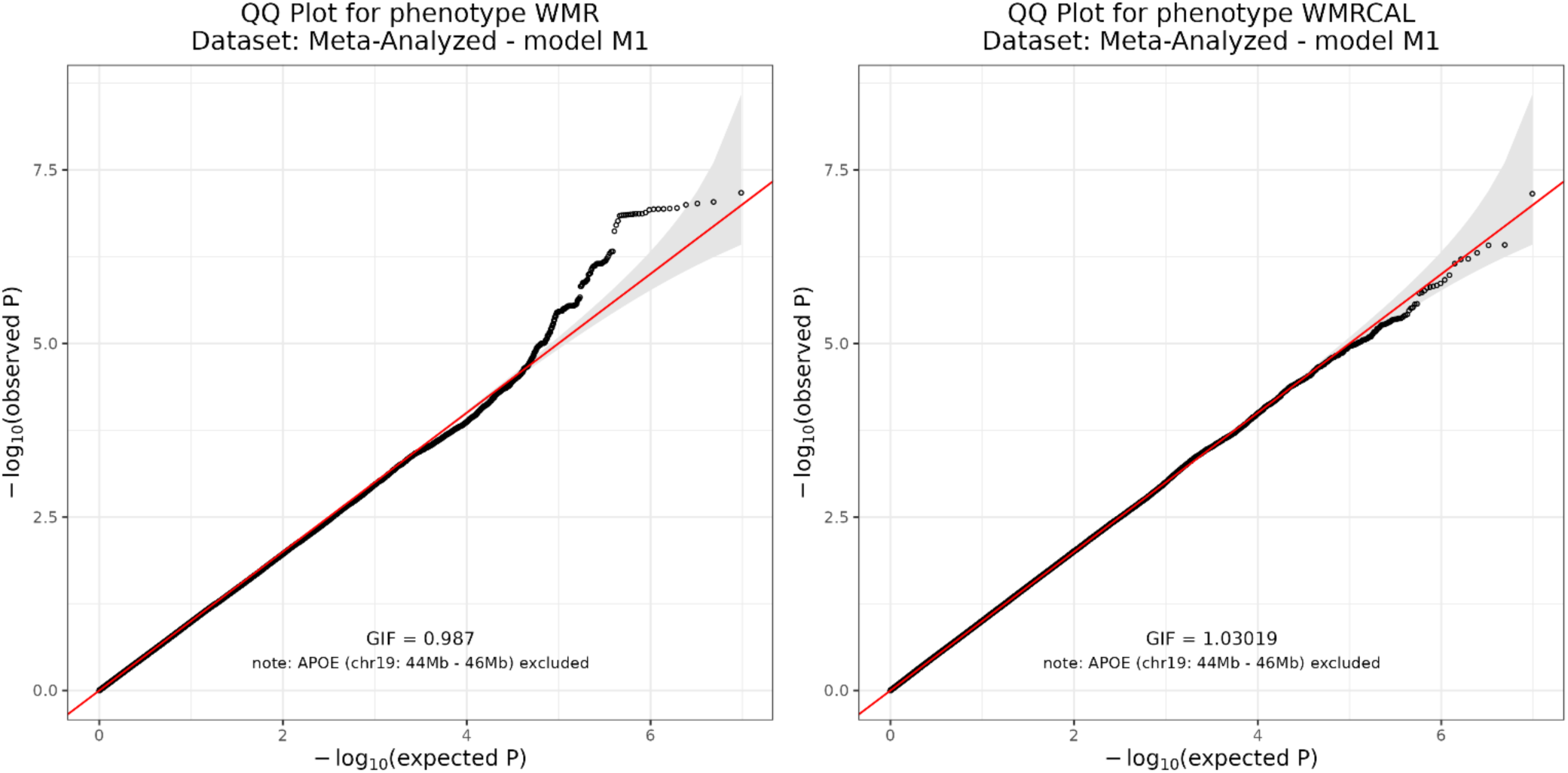
QQ plots for genome-wide association analyses. Quantile-quantile plots for each genome-wide association study. λ denotes the genomic inflation factor for the study.

**Figure S3:**
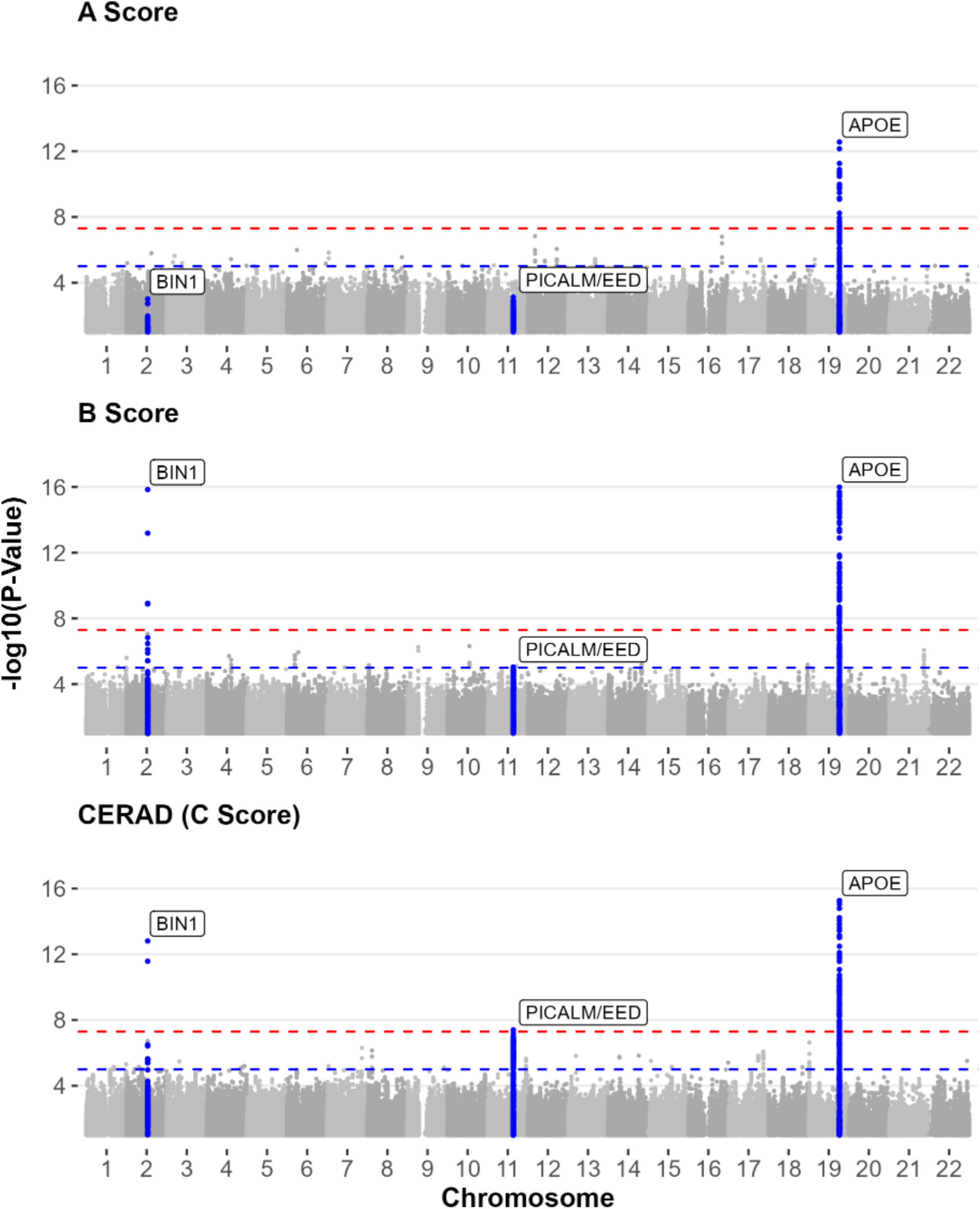
P-value by genomic position for A score, B score, and C score. Genome-wide association results for AD hallmark pathologies. P-values reported on the −log(10) scale. Variants at *APOE* with −log10(p-value) greater than 17 were censored to improve readability.

**Figure S4:**
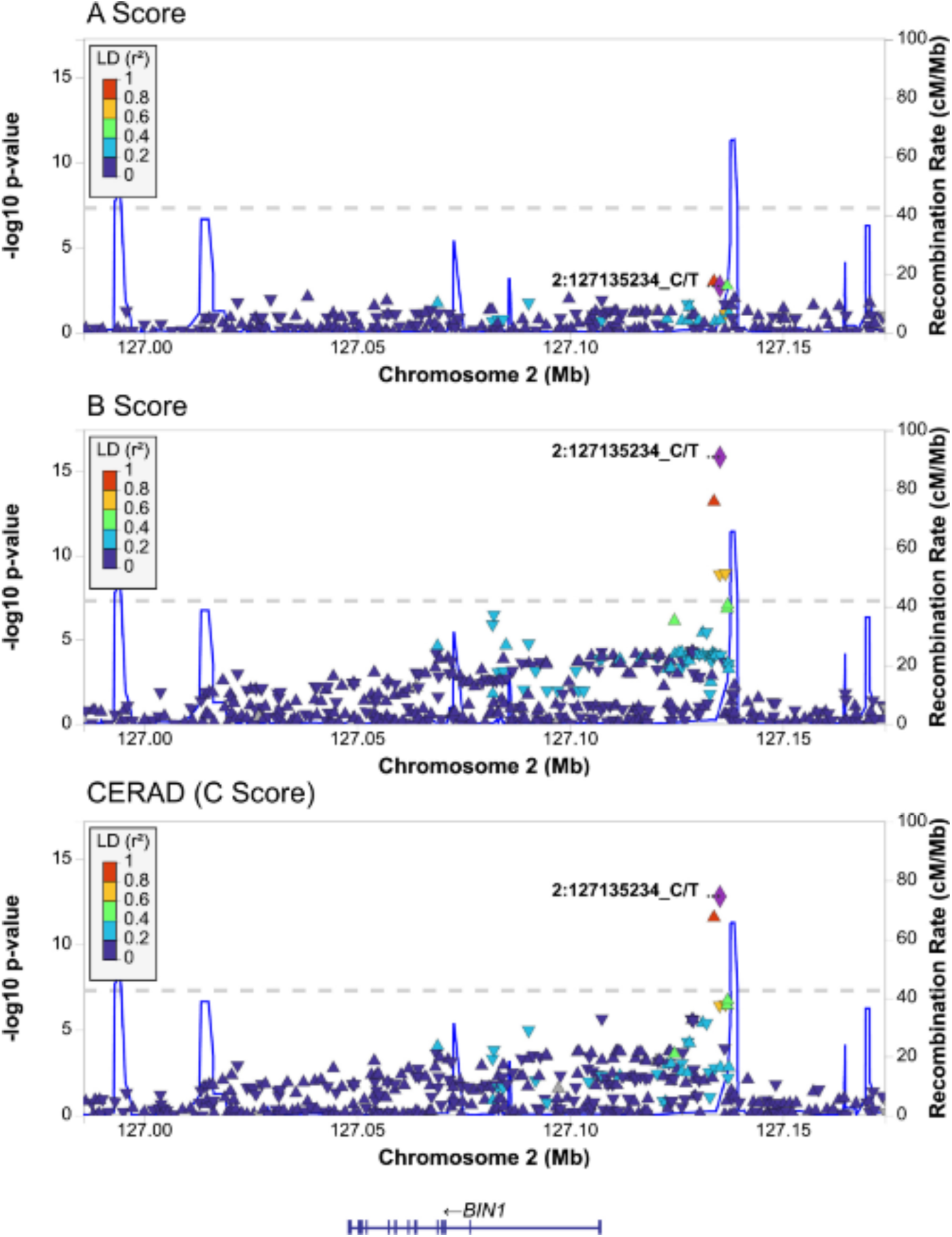
Regional association plot for the BIN1 locus, for A Score (NP/Thal), B Score (NFT Braak), and C Score (CERAD) Regional association plots for the *BIN1* locus for AD hallmark pathologies. P-values reported on the −log(10) scale.

**Figure S5:**
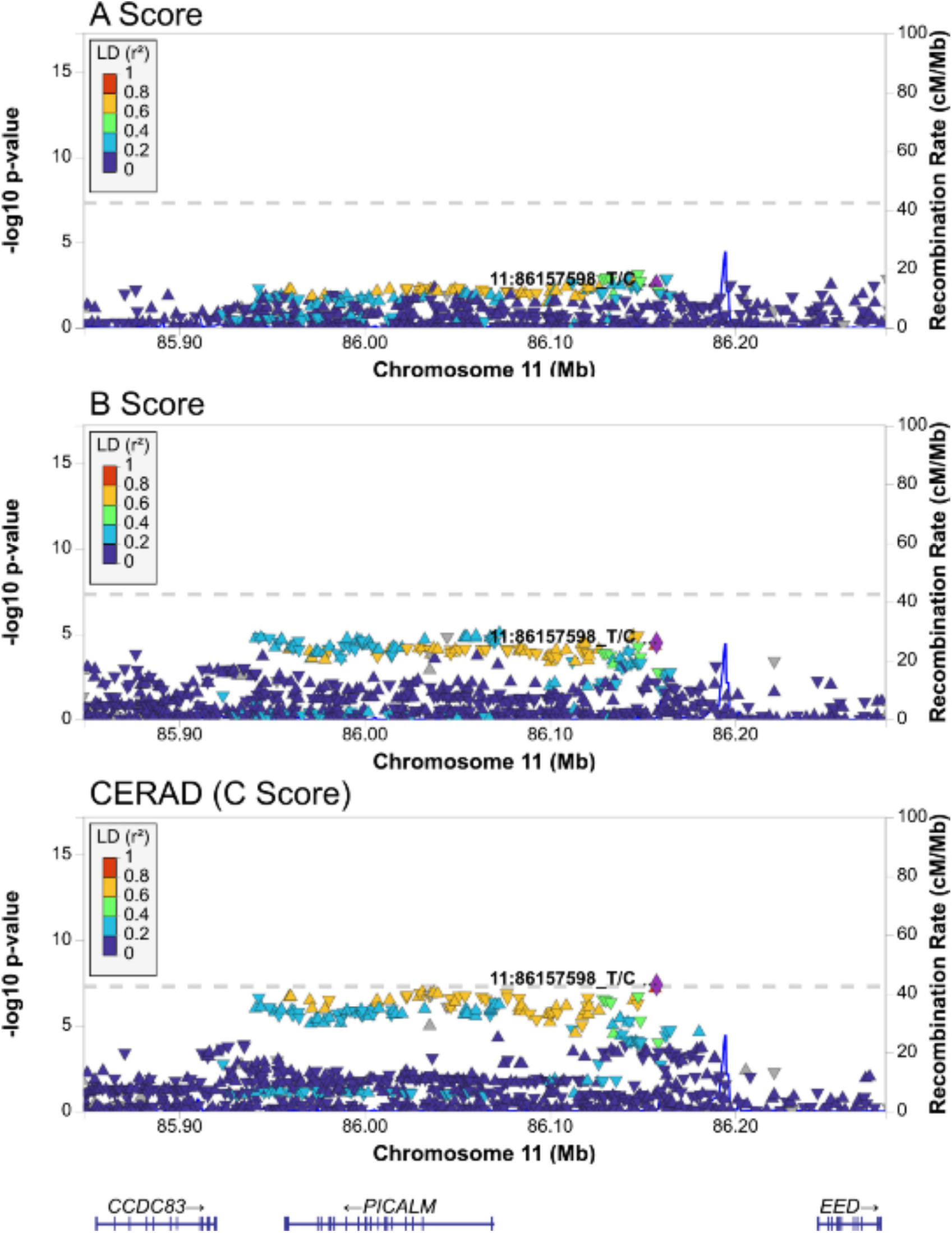
Regional association plot for the PICALM/EED locus, for A Score (NP/Thal), B Score (NFT Braak), and C Score (CERAD) Regional association plots for the *PICALM* locus for AD hallmark pathologies. P-values reported on the −log(10) scale.

**Figure S6:**
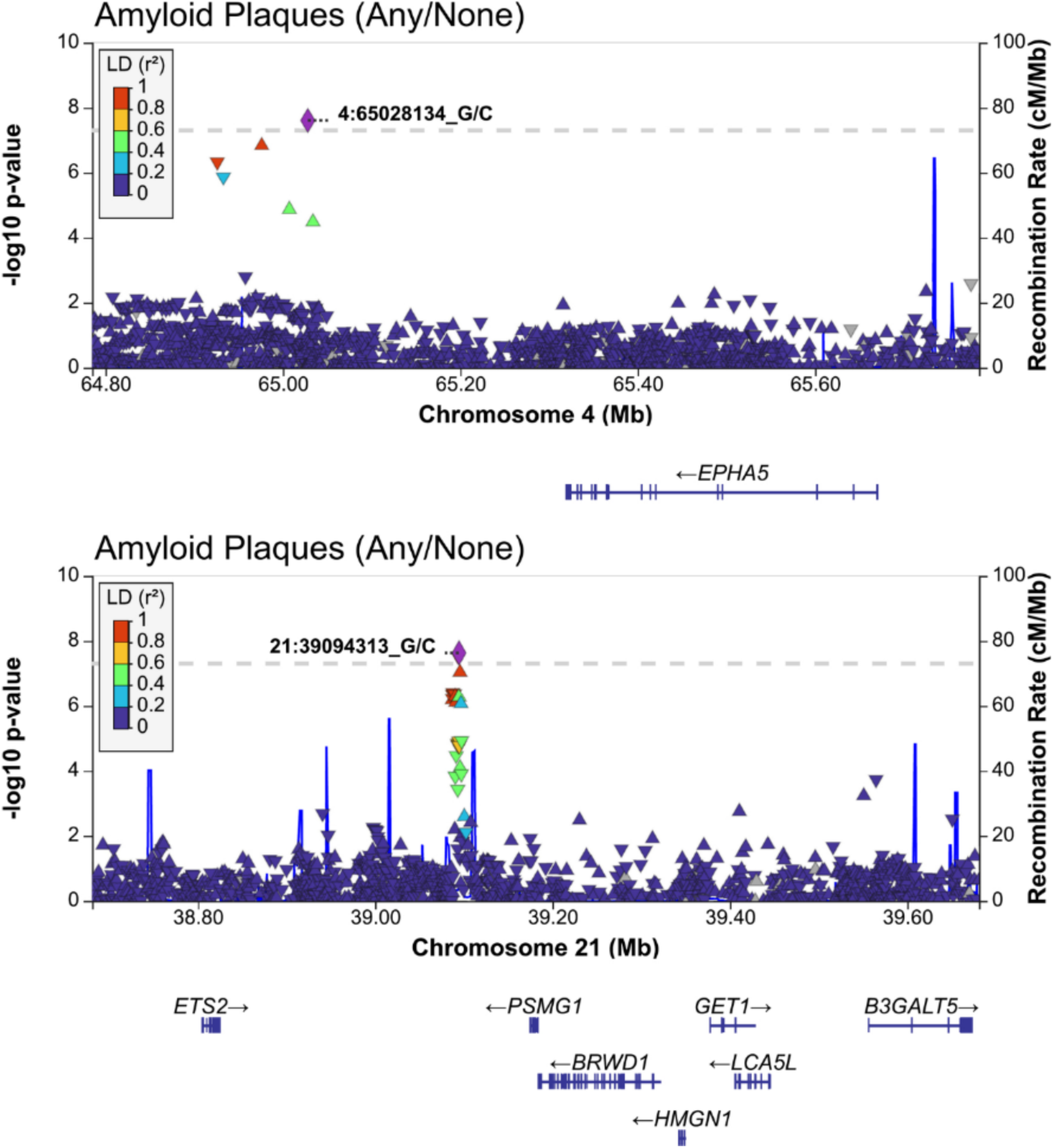
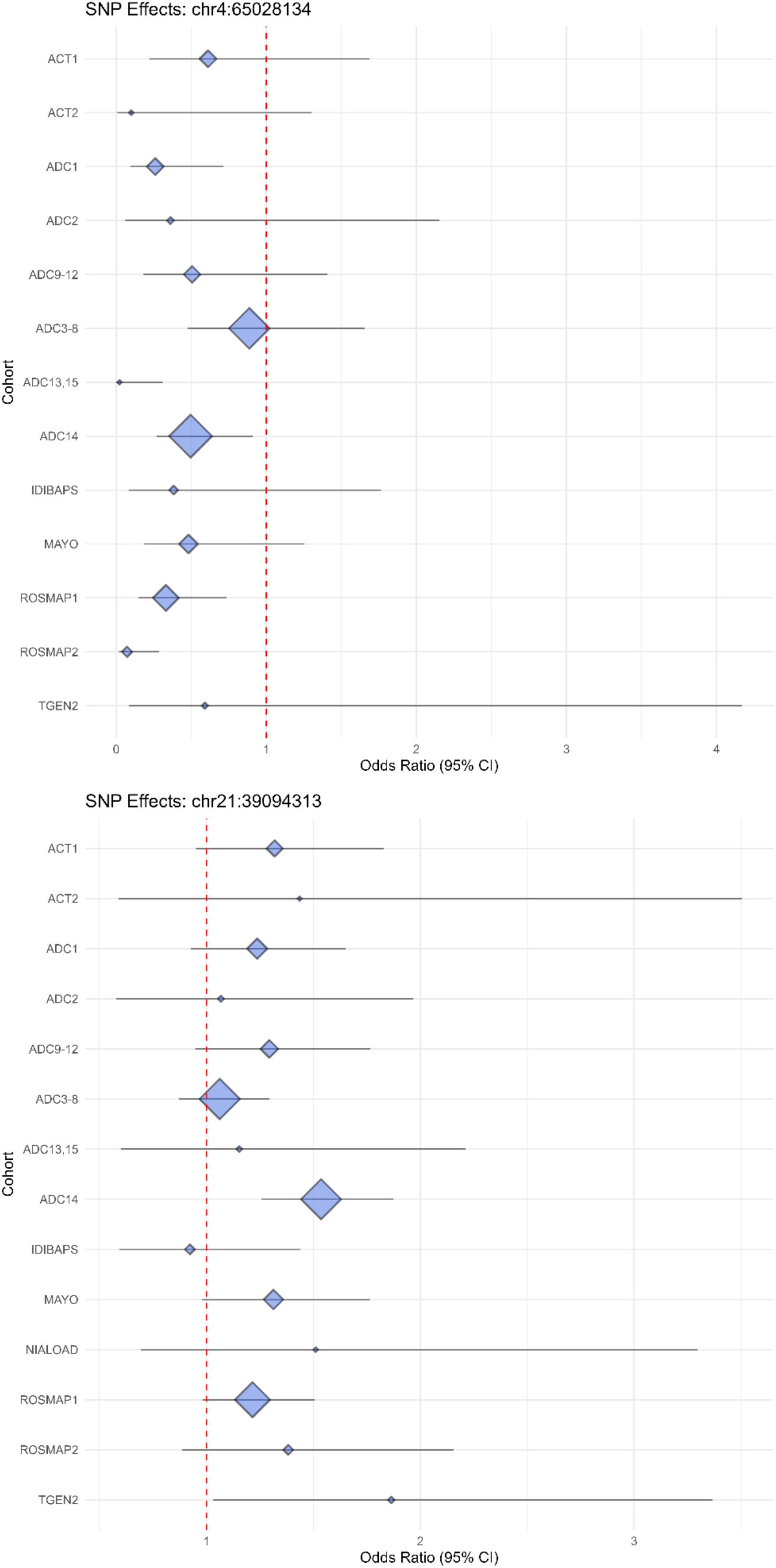
Regional association plots and forest plots for genome-wide significant variants from the amyloid plaque (presence/absence) analysis. Regional association and forest plots for the *EPHA5* and *PSMG1* loci for presence/absence of amyloid plaques. P-values reported on the −log(10) scale.

**Figure S7:**
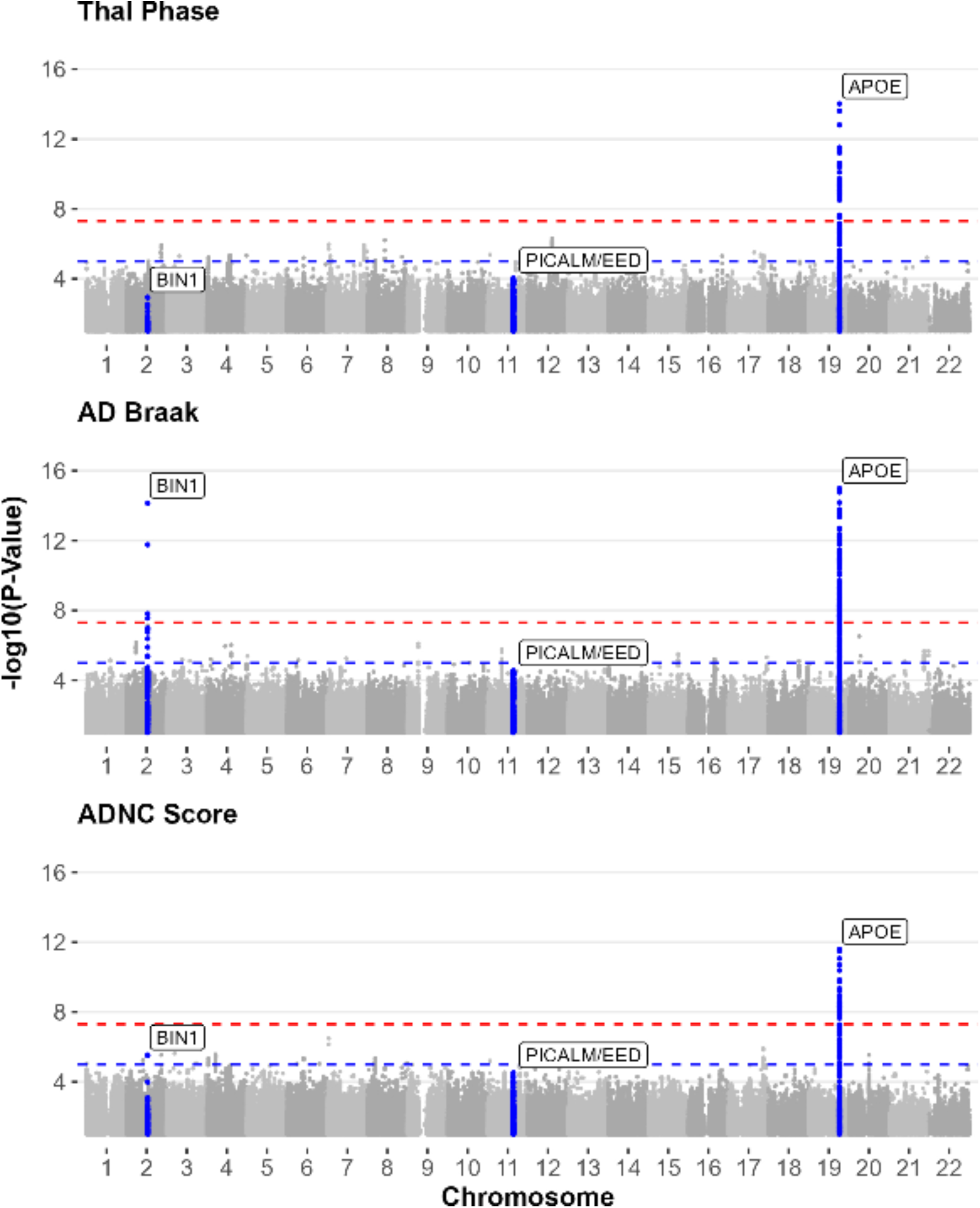
P-value by genomic position for association with Thal phase, NFT Braak, and ADNC (ABC) score. Genome-wide association results for Thal Phase, Braak (NFT), and ADNC composite score. P-value was capped at 1e-15. The minimum p-values for *APOE* were: p-value(THAL) = 6.379e-63, p-value(NFT BRAAK) = 8.06e-147, and p-value(ADNC) = 1.916e-55.

**Figure S8:**
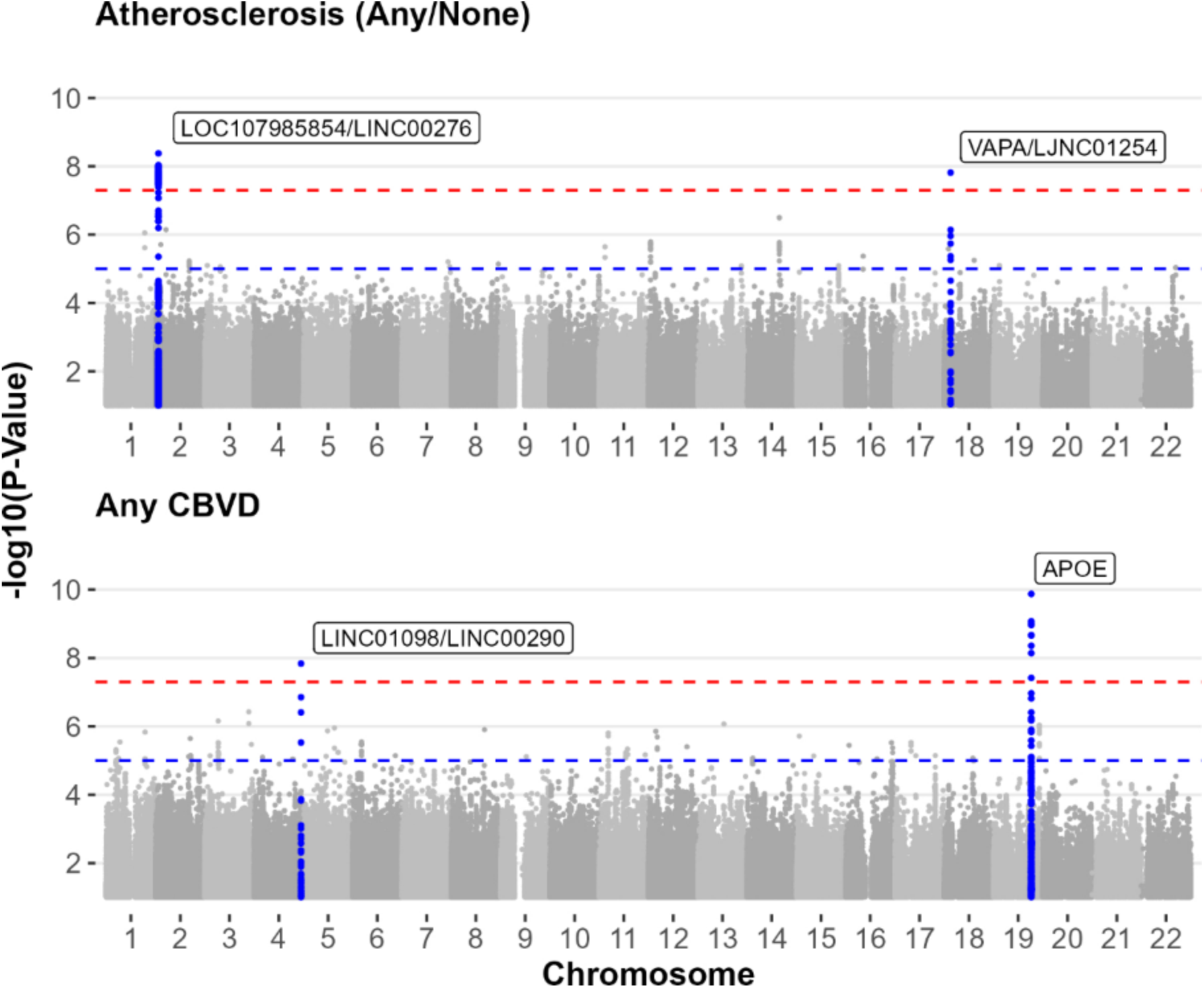
P-value by genomic position for cerebral atherosclerosis (any/none) and cerebrovascular disease (any/none) Genome-wide association results for atherosclerosis (any/none) pathology, and cerebrovascular disease (any/none).

**Figure S9:**
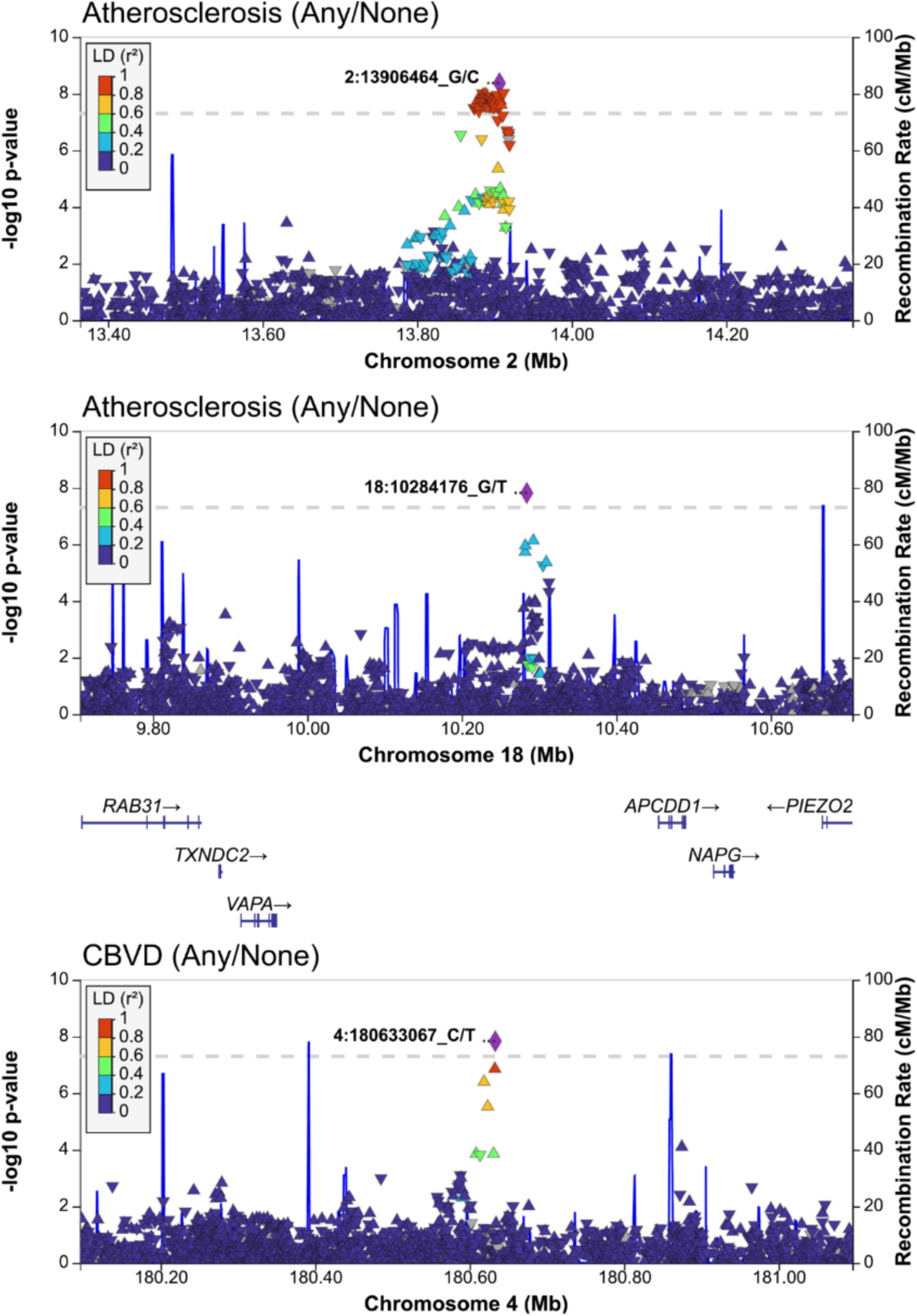
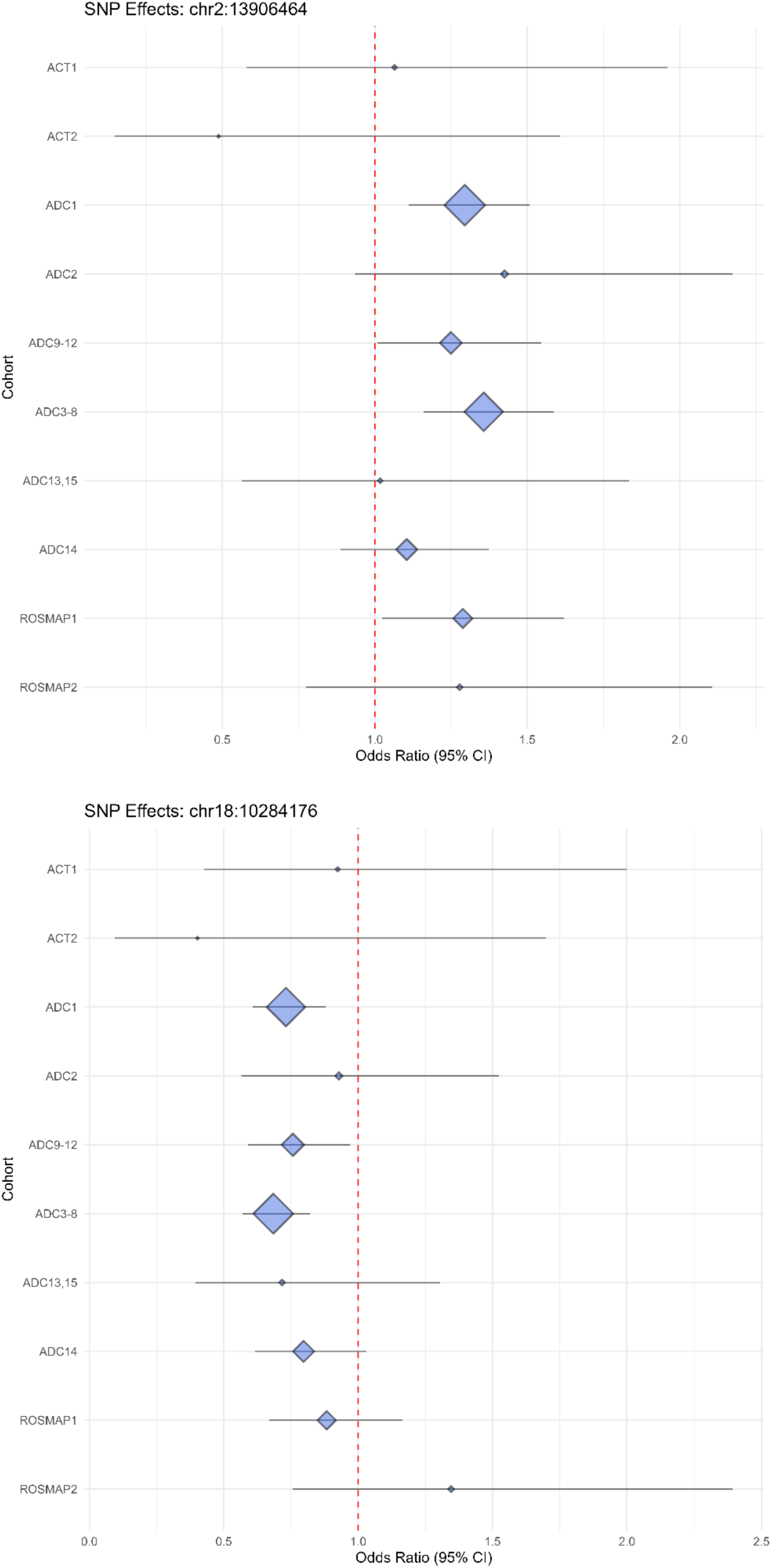
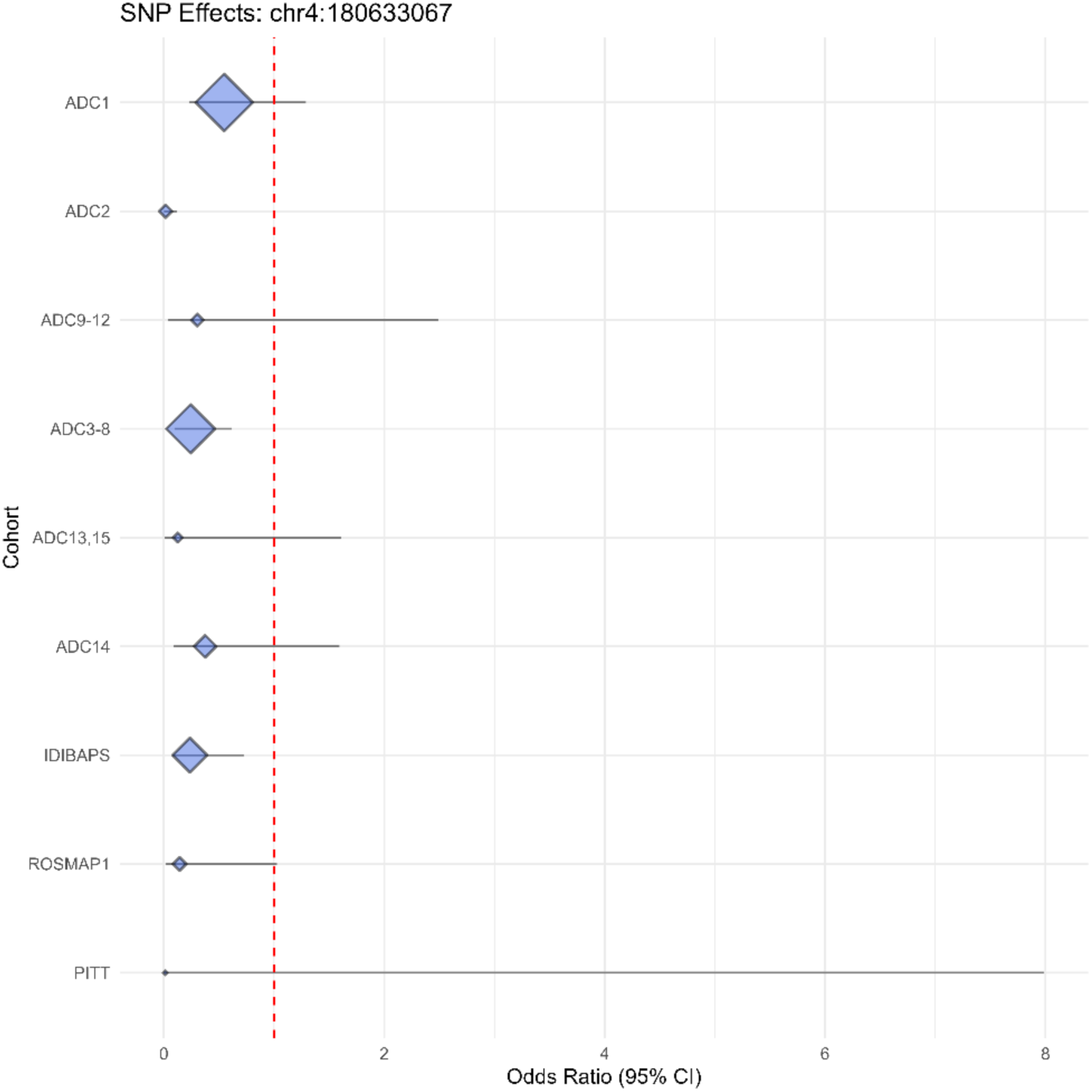
Regional association plots and forest plots for genome-wide significant variants from cerebral atherosclerosis (any/none) and cerebrovascular disease (any/none) analyses. Regional association and forest plots for the significant chromosome 2 locus (atherosclerosis any/none), *VAPA* (atherosclerosis any/none), and the significant chromosome 4 locus (CBVD any/none).

**Figure S10:**
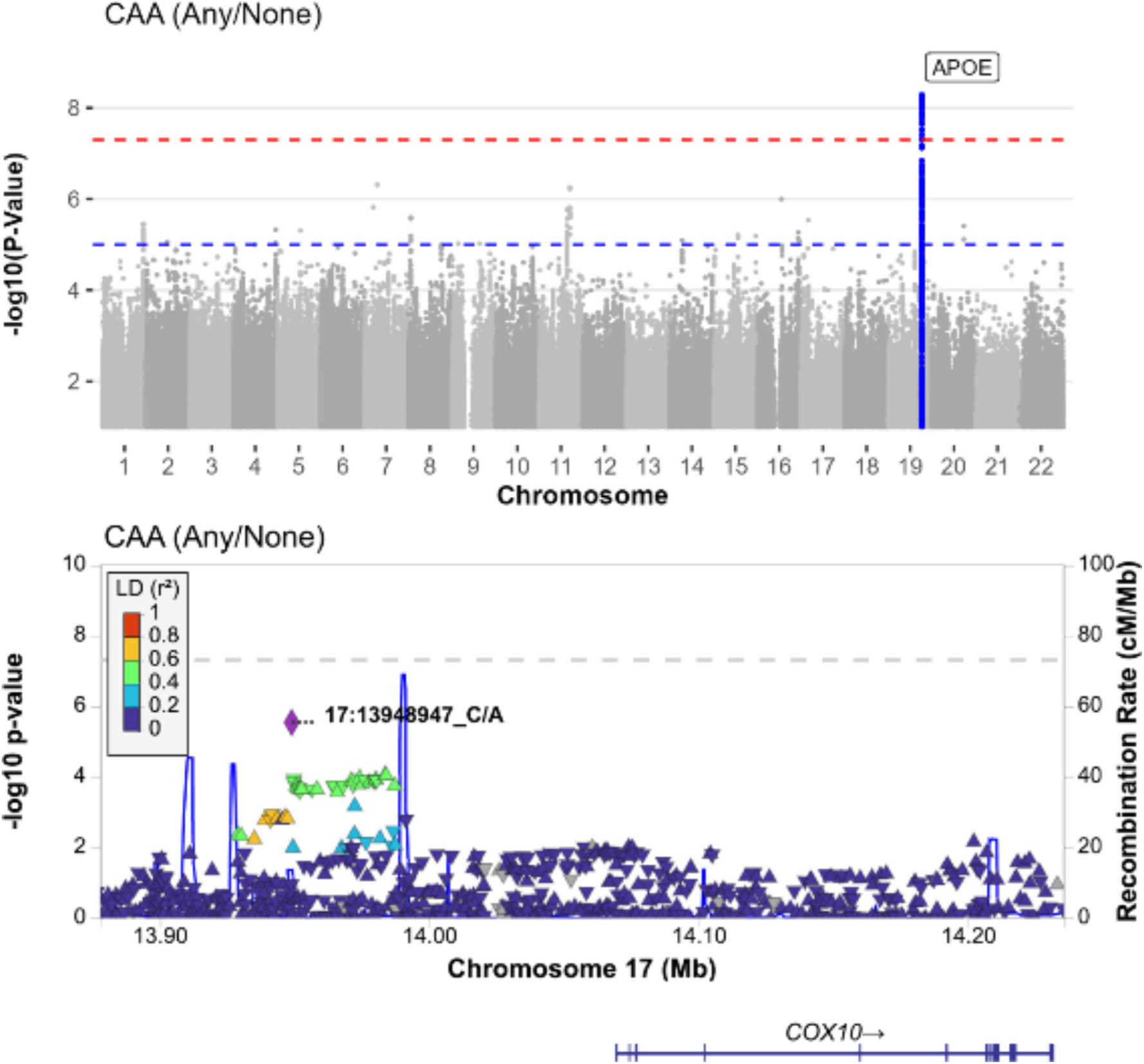
P-value by genomic position for association with cerebral amyloid angiopathy (CAA) analysis, and regional association analysis of the *COX10* region. Genome-wide association results for cerebral amyloid angiopathy, and regional association plot for the *COX10* region. P-values reported on the −log(10) scale.

**Figure S11:**
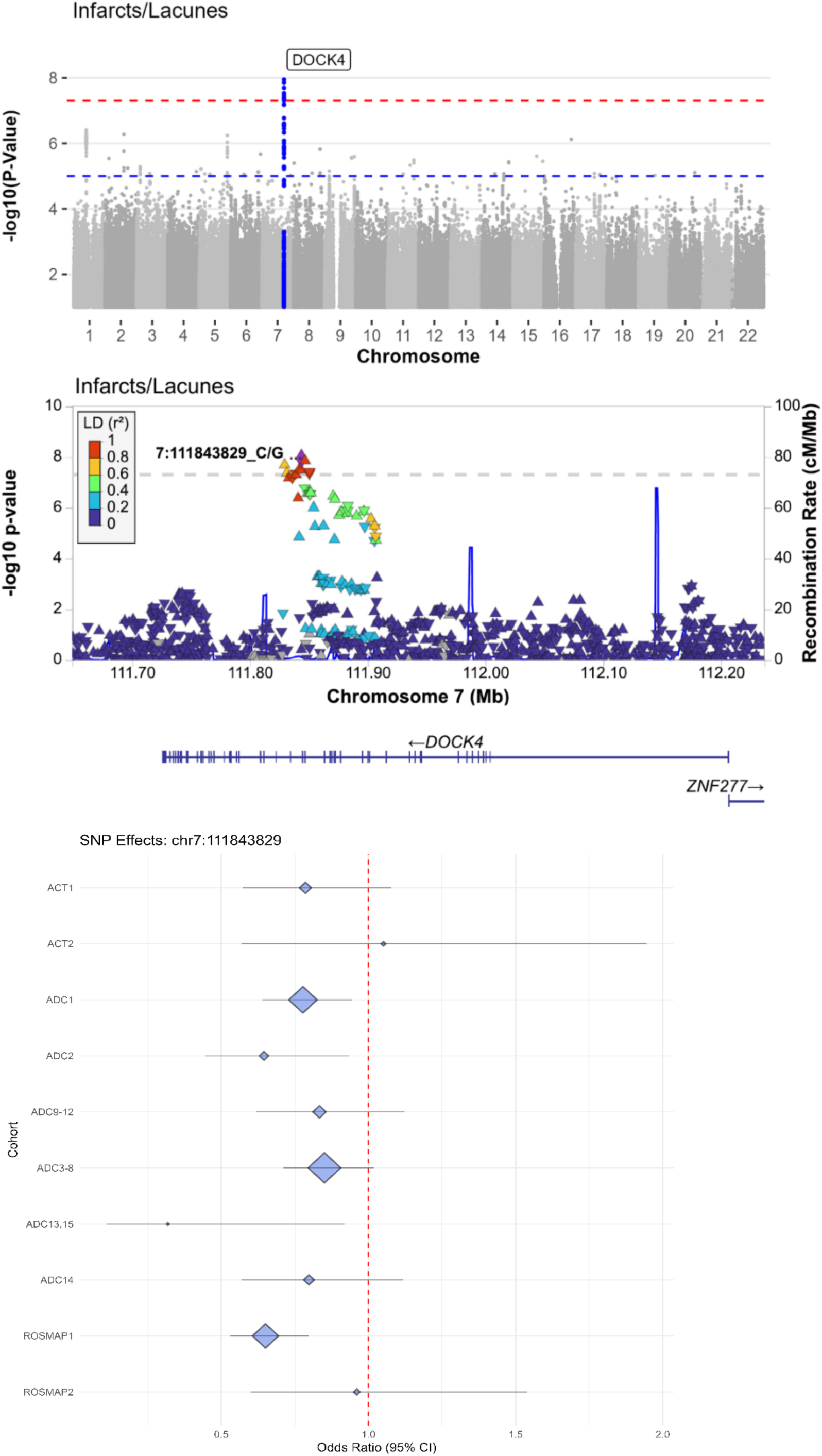
P-value by genomic position for association with infarcts/lacunes analyses, and regional association analysis of the *DOCK4* region, and forest plot for the index variant. Genome-wide association results for infarcts/lacunes, and regional association plot for the *DOCK4* region. P-values reported on the −log(10) scale.

**Figure S12:**
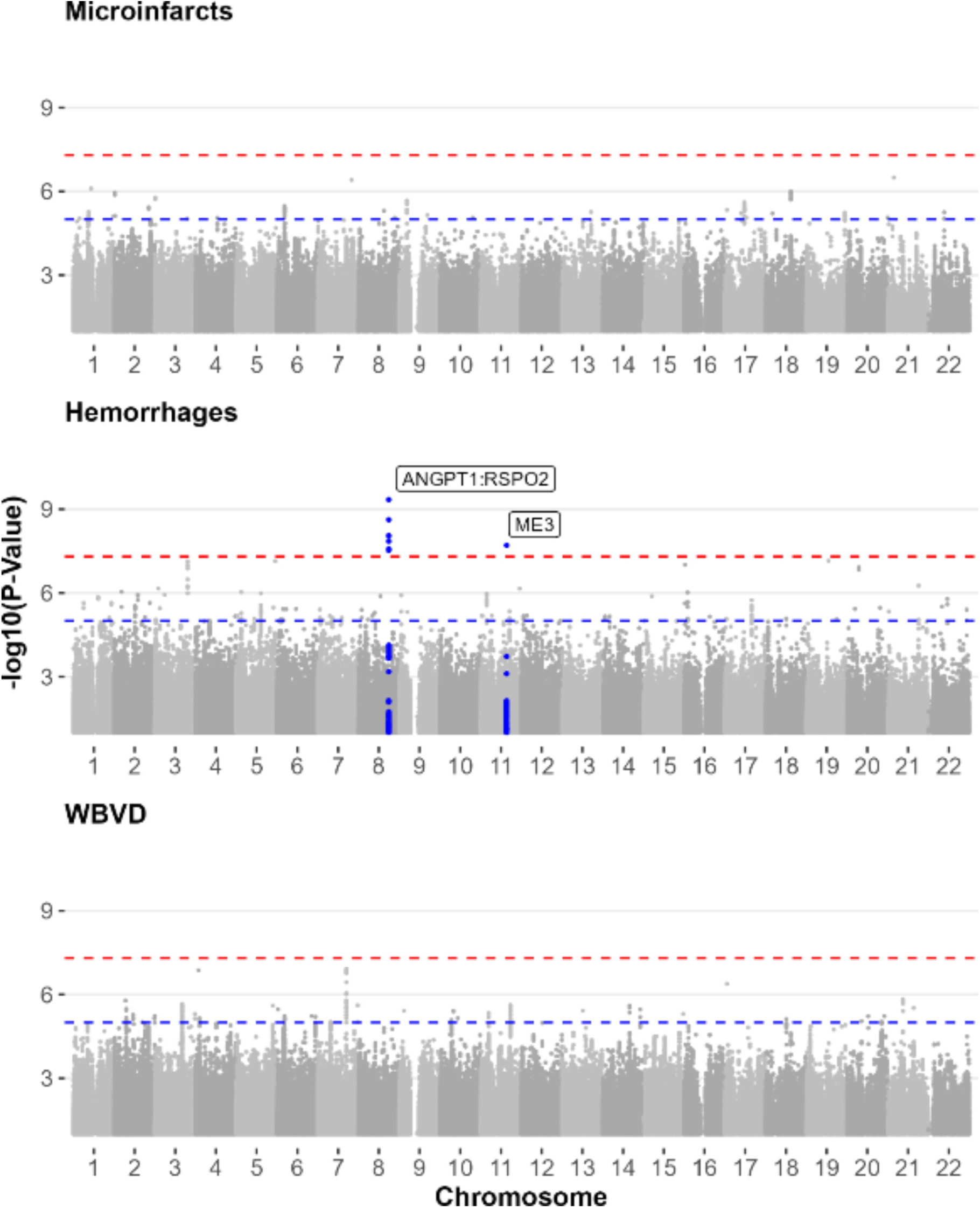
P-value by genomic position for association with microinfarcts, hemorrhages, and whole brain vascular disease (WBVD) Genome-wide association results for microinfarcts, hemorrhages, and whole brain vascular disease pathologies. P-values reported on the −log(10) scale.

**Figure S13:**
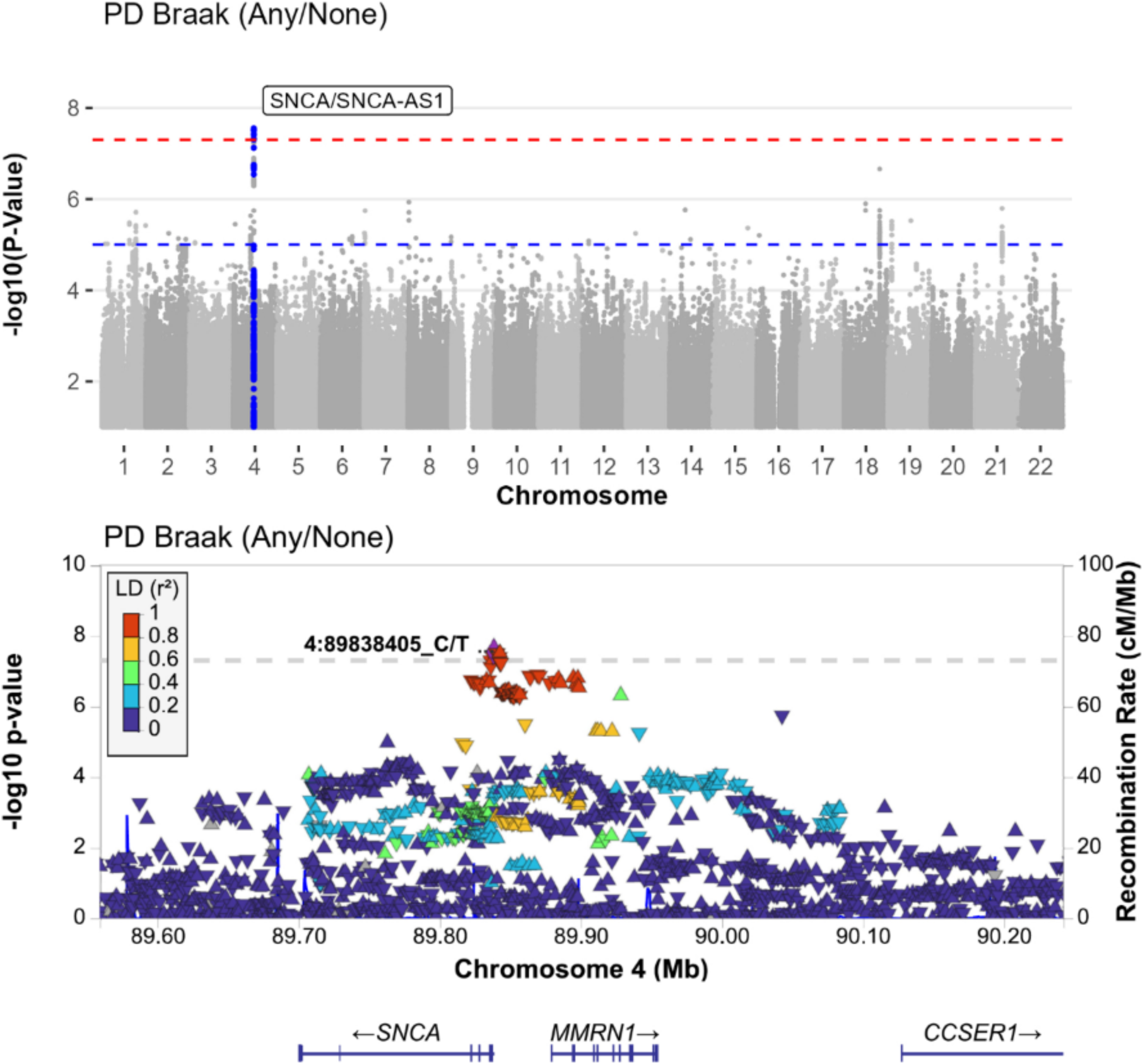
P-value by genomic position for association with lewy body (PD Braak) analyses, and regional association analysis of the *SNCA*/*MMRN1* region. Genome-wide association results for Lewy body pathology (any/none), and regional association plot for the *SNCA/MMRN1* region. P-values reported on the −log(10) scale.

**Figure S14:**
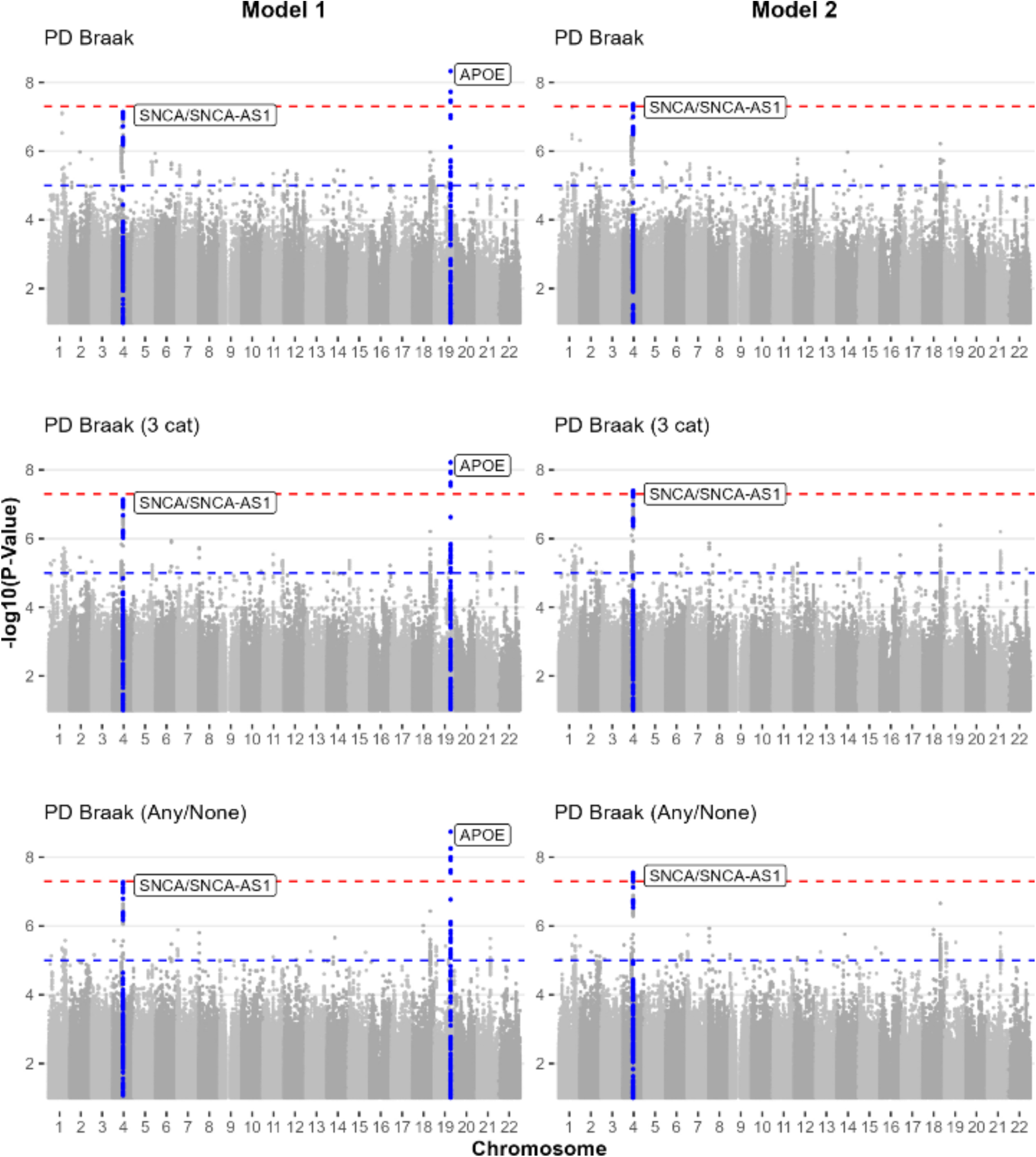
P-value by genomic position for association with lewy bodies. Model 1 includes age, sex and principal components in the model; model 2 includes *APOE* e4 allele count as well as age, sex, and principal components. Three different parameterizations of lewy bodies were included: PD Braak is the standard 5 category Braak staging for severity of LBD; the “3 cat(egory)” parameterization collapses these into three groups; “Any/None” parameterizes LBD to presence/absence.

**Figure S15:**
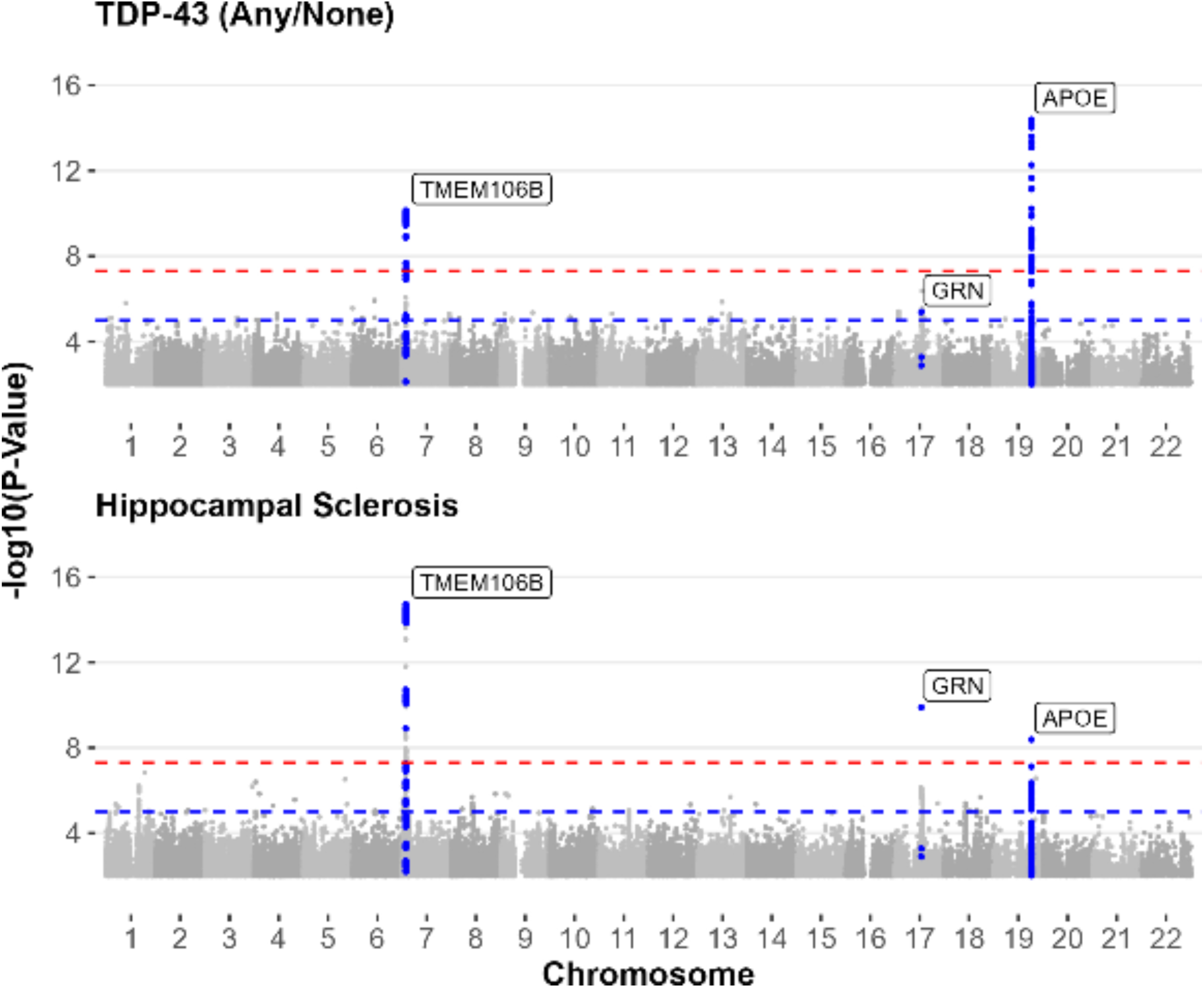
P-value by genomic position for association with TDP-43 proteinopathy and hippocampal sclerosis. Genome-wide association results for TDP-43 proteinopathy (any/none) and hippocampal sclerosis. P-values reported on the −log(10) scale.

**Figure S16:**
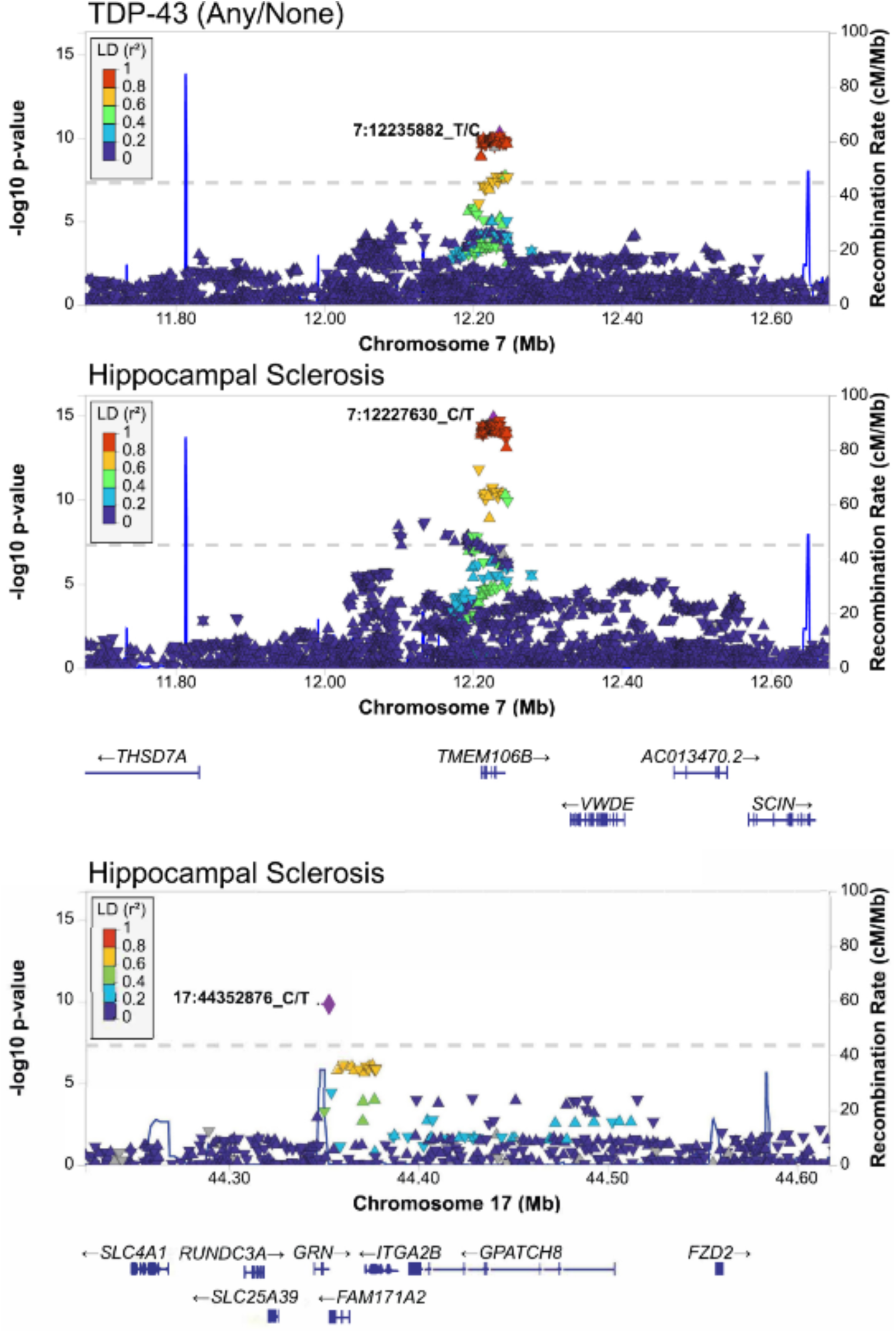
Regional association plot for the *TMEM106B* locus, for TDP-43 proteinopathy and hippocampal sclerosis, and the *GRN* locus for hippocampal sclerosis. Regional association plot for the *TMEM106B* locus, for TDP-43 proteinopathy (presence/absence) and hippocampal sclerosis, and the *GRN* locus for hippocampal sclerosis.

**Figure S17:**
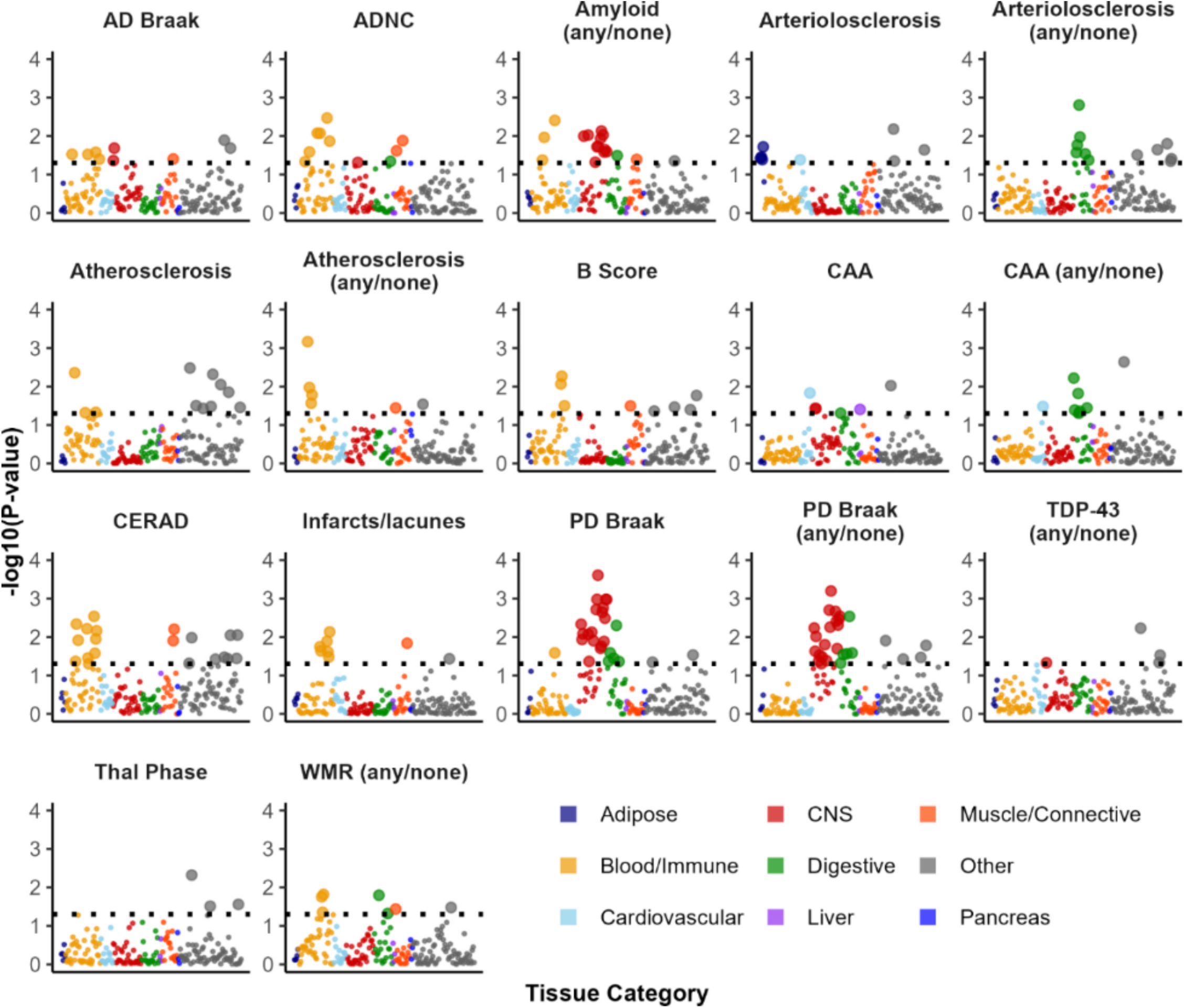
Cell-specific heritability enrichment results (multi-tissue GTEx) Results from cell-specific heritability enrichment analyses. Y axis denotes −log10(p-value) of the enrichment score. The dashed line denotes nominal association. Study-wide significance by false discovery rate (FDR) would be at −log10(p-value) = 2.75. Variant sets are derived from human gene expression data from the GTEx study, as in Finucane et al. See Supplemental Text for more details.

**Figure S18:**
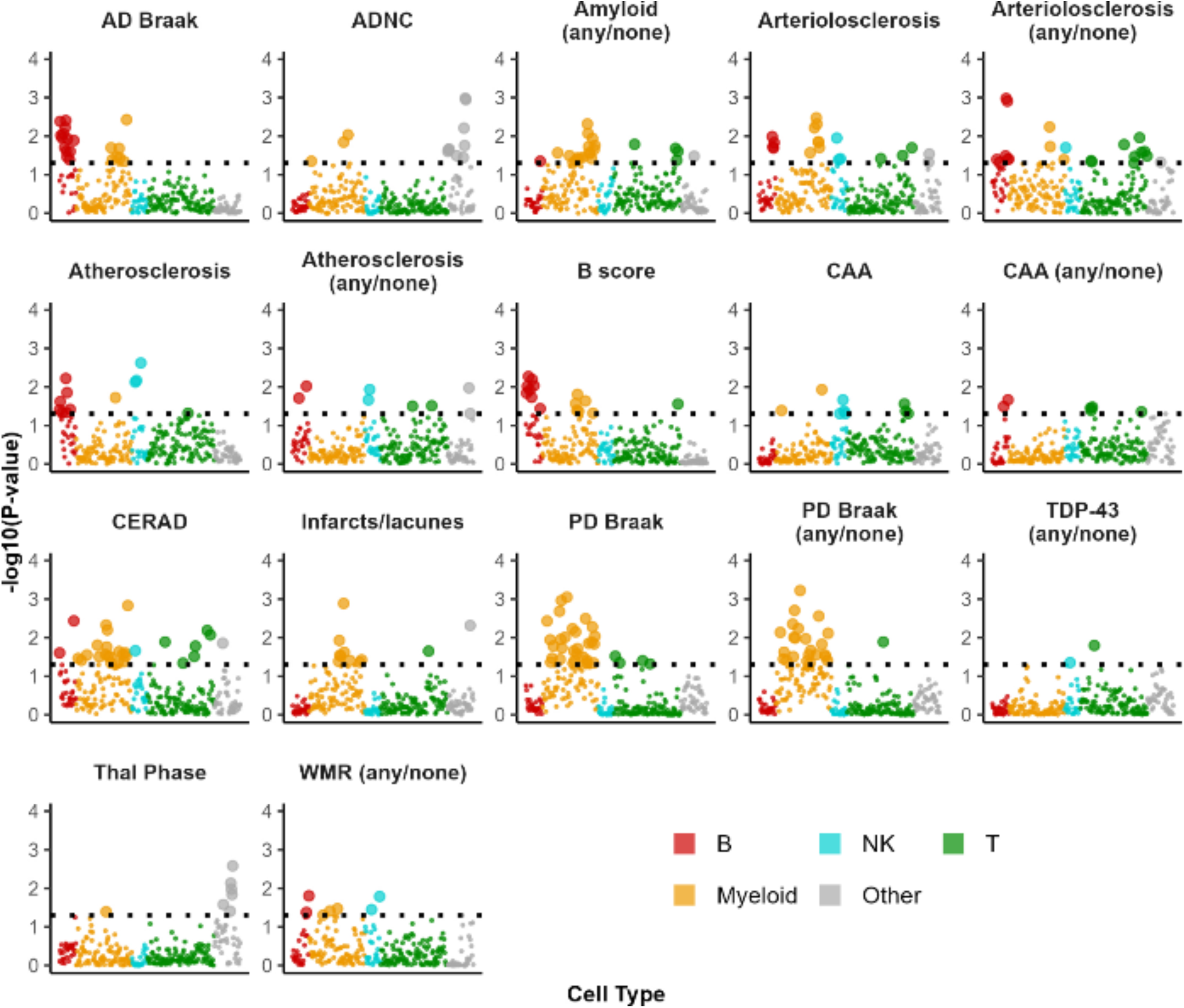
Cell-specific heritability enrichment results (ImmGen) Results from cell-specific heritability enrichment analyses. Y axis denotes −log10(p-value) of the enrichment score. The dashed line denotes nominal association. Study-wide significance by false discovery rate (FDR) would be at −log10(p-value) = 3.03. Variant sets are derived from mouse derived immune gene expression data from the GTEx study, as in Finucane et al. See Supplemental Text for more details.

### Supplemental Tables

**Table S1: Frequency and proportion of neuropathological lesions**

Abbreviations: AP, amyloid plaques; CAA, cerebral amyloid angiopathy; CBVD, cerebrovascular disease; PD Parkinson disease (e.g., Lewy bodies); HS, hippocampal sclerosis; VBI, vascular brain injury; WBVD, whole brain vascular disease; WMR, white matter rarefaction

**Table S2: Association of APOE e4, across neuropathology phenotypes (rs429358; chr19:44,908,684)**

Abbreviations: AP, amyloid plaques; CAA, cerebral amyloid angiopathy; CBVD, cerebrovascular disease; RD, (AD) related dementias; PD Parkinson disease (e.g., Lewy bodies); HS, hippocampal sclerosis; VBI, vascular brain injury; WBVD, whole brain vascular disease; SE, standard error WMR, white matter rarefaction Het Q, Het df and Hetp-value note the heterogeneity test statistic, degrees of freedom, and p-value, respectively.

**Table S3: All genome-wide association results with p < 0.0001**

**Table S4: Association of Bellenguez et al AD risk loci with AD related dementia pathologies** Odds ratios (OR) refer to the risk allele (RA) from Bellenguez et al. GWAS. Highlighted and bolded ORs indicate significance (P < 0.05). Only variants showing significant association with at least one lesion is included in the table. Abbreviations: RA, risk allele; OA, other allele; OR, odds ratio.

**Table S5: Association of Bellenguez et al AD risk loci with cerebrovascular and vascular brain injury pathologies**

**Table S6: Association of Nalls et al PD risk loci with lewy body pathology**

**Table S7: Association of Nalls et al PD risk loci with AD hallmark pathologies**

**Table S8: Association of Nalls et al PD risk loci with related dementia pathologies**

**Table S9. Estimated SNP heritability across all AD/RD pathologies**

Analyses used Model 2 throughout, excluding APOE region; liability scale; intercept constrained to 1. Shaded p-values indicate significance (p < 0.05). Abbreviations: AD, Alzheimer’s disease; ADNC, AD neuropathologic change; CAA, cerebral amyloid angiopathy; CI, confidence interval; CERAD, Consortium to Establish a Registry for AD neuritic plaque score; CBVD, cerebrovascular disease; HS, hippocampal sclerosis; LBD, Lewy body disease; SE, standard error; TDP-43, TAR DNA-binding protein 43; VBI, vascular brain injury; WMR, white matter rarefaction

**Table S10. Estimated SNP-based genetic correlation (shared heritability) across between neuropathological phenotypes.**

Genetic correlations only calculated when individual SNP heritabilities were significantly greater than zero (i.e., p < 0.05 in Table S8). Some comparisons not included due to poor model convergence.

**Table S11: Cell/Tissue-specific heritability enrichment p-values (GTEx, multi tissue)**

See Supplemental Text for methodology.

**Table S12: Cell/Tissue-specific heritability enrichment coefficients and p-values (GTEx, multi tissue)**

This table contains the same pvalues as Table S10, but with coefficients and in “long” format. See Supplemental Text for methodology.

**Table S13: Cell/Tissue-specific heritability enrichment p-values (ImmGen)**

See Supplemental Text for methodology.

**Table S14: Cell/Tissue-specific heritability enrichment coefficients and p-values (ImmGen)**

This table contains the same pvalues as Table S12, but with coefficients and in “long” format. See Supplemental Text for methodology.

**Table S15: MAGMA pathway analysis results**

**Table S16: MAGMA tissue-specific associations**

**Table S17: MAGMA tissue-specific associations, general tissues**

**Table S18: Genotype Imputation Quality (R2) by batch, for genome-wide associated variants**

**Table S19: Neuropathology phenotype coding**

## REFERENCES

1. Brenowitz WD, Hubbard RA, Keene CD, Hawes SE, Longstreth WT Jr, Woltjer RL. Mixed neuropathologies and estimated rates of clinical progression in a large autopsy sample. Alzheimers Dement. 2017;13(6):654–662.

2. Kapasi A, DeCarli C, Schneider JA. Impact of multiple pathologies on the threshold for clinically overt dementia. Acta Neuropathol. 2017;134(2):171–186. doi:10.1007/s00401-017-1717-7

3. Kawas CH, Kim RC, Sonnen JA, al et. Multiple pathologies are common and related to dementia in the oldest-old: The 90+ Study. Neurology. 2015;85(6):535–542. doi:10.1212/WNL.0000000000001831

4. Montine TJ, Sonnen JA, Montine KS, al et. Adult Changes in Thought study: dementia is an individually varying convergent syndrome with prevalent clinically silent diseases that may be modified by some commonly used therapeutics. Curr Alzheimer Res. 2012;9(6):718–723. doi:10.2174/156720512801322555

5. Rahimi J, Kovacs GG. Prevalence of mixed pathologies in the aging brain. Alzheimers Res Ther. 2014;6(9):82. doi:10.1186/s13195-014-0082-1

6. Schneider JA, Aggarwal NT, Barnes L, al et. The neuropathology of older persons with and without dementia from community versus clinic cohorts. J Alzheimers Dis. 2009;18(3):691–701. doi:10.3233/JAD-2009-1227

7. White LR, Edland SD, Hemmy LS, al et. Neuropathologic comorbidity and cognitive impairment in the Nun and Honolulu-Asia Aging Studies. Neurology. 2016;86(11):1000–1008. doi:10.1212/WNL.0000000000002480

8. Bennett DA, Buchman AS, Boyle PA, al et. Religious Orders Study and Rush Memory and Aging Project. J Alzheimers Dis. 2018;64(s1):S161-S189. doi:10.3233/JAD-179939

9. Escott-Price V, Hardy J. Genome-wide association studies for Alzheimer’s disease: bigger is not always better. Brain Commun. 2022;4(3).

10. Jansen IE, Savage JE, Watanabe K, Bryois J, Williams DM, Steinberg S. Genome-wide meta-analysis identifies new loci and functional pathways influencing Alzheimer’s disease risk. Nat Genet. 2019;51(3):404–413.

11. Kunkle BW, Grenier-Boley B, Sims R, al et. Genetic meta-analysis of diagnosed Alzheimer’s disease identifies new risk loci and implicates Aβ, Tau, and immunity. Nature Genet. 2019;51:414–430. doi:10.1038/s41588-019-0358-2

12. Wightman DP, Jansen IE, Savage JE, Shadrin AA, Bahrami S, Holland D. A genome-wide association study with 1,126,563 individuals identifies new risk loci for Alzheimer’s disease. Nat Genet. 2021;53(9):1276–1282.

13. Willett JDS, Waqas M, Choi Y, Ngai T, Mullin K, Tanzi RE. Identification of 16 novel Alzheimer’s disease loci using multi-ancestry meta-analyses. Alzheimers Dement. 2025;21(2).

14. Nelson PT, Alafuzoff I, Bigio EH, Bouras C, Braak H, Cairns NJ. Correlation of Alzheimer disease neuropathologic changes with cognitive status: a review of the literature. J Neuropathol Exp Neurol. 2012;71(5):362–381.

15. White LR, Corrada MM, Kawas CH, Cholerton BA, Edland SE, Flanagan ME. Neuropathologic Changes of Alzheimer’s Disease and Related Dementias: Relevance to Future Prevention. J Alzheimers Dis. 2023;95(1):307–316.

16. Xavier C, Pinto N. Navigating the blurred boundary: Neuropathologic changes versus clinical symptoms in Alzheimer’s disease, and its consequences for research in genetics. J Alzheimers Dis. 2025(13872877251317544).

17. Jansen IE, Lee SJ, Gomez-Fonseca D. Genome-wide meta-analysis for Alzheimer’s disease cerebrospinal fluid biomarkers. Acta Neuropathol. 2022;144:821–842. doi:10.1007/s00401-022-02454-z

18. Cruchaga C, Timsina J, Jiang C. GWAS meta-analysis of CSF Alzheimer’s disease biomarkers 18,948 individuals reveal novel loci and genes regulating lipid metabolism, brain volume and autophagy. Published online May 21, 2025.

19. Amanullah AA, Mirbod M, Pandey A, Singh SB, Gandhi OH, Ayubcha C. The Genetics of Amyloid Deposition: A Systematic Review of Genome-Wide Association Studies Using Amyloid PET Imaging in Alzheimer’s Disease. Journal of Imaging. 2025;11(8). doi:10.3390/jimaging11080280

20. Raghavan NS, Dumitrescu L, Mormino E. Association Between Common Variants in RBFOX1, an RNA-Binding Protein, and Brain Amyloidosis in Early and Preclinical Alzheimer Disease. JAMA Neurol. 2020;77(10):1288–1298. doi:10.1001/jamaneurol.2020.1760

21. Beecham GW, Hamilton K, Naj AC, al et. Genome-wide association meta-analysis of neuropathologic features of Alzheimer’s disease and related dementias. JAMA Neurol. 2014;71(5):686–694. doi:10.1001/jamaneurol.2014.235

22. Chibnik LB, White CC, Mukherjee S, al et. Susceptibility to neurofibrillary tangles: role of the PTPRD locus and limited pleiotropy with other neuropathologies. Mol Psychiatry. 2018;23(6):1521–1529. doi:10.1038/mp.2017.20

23. Guerreiro R, Ross OA, Kun-Rodrigues C, al et. Investigating the genetic architecture of dementia with Lewy bodies: a two-stage genome-wide association study. Lancet Neurol. 2018;17(1):64–74. doi:10.1016/S1474-4422(17)30400-3

24. Nelson PT, Estus S, Abner EL, al et. ABCC9 gene polymorphism is associated with hippocampal sclerosis of aging pathology. Acta Neuropathol. 2014;127(6):825–843. doi:10.1007/s00401-014-1282-2

25. Shade LMP, Katsumata Y, Abner EL, al et. GWAS of multiple neuropathology endophenotypes identifies new risk loci and provides insights into the genetic risk of dementia. Nat Genet. 2024;56(11):2407–2421. doi:10.1038/s41588-024-01939-9

26. Godrich D, Martin ER, Schellenberg G, Pericak-Vance MA, Cuccaro M, Scott WK. Neuropathological lesions and their contribution to dementia and cognitive impairment in a heterogeneous clinical population. Alzheimers Dement. 2022;18(12):2403–2412.

27. Hyman BT, Phelps CH, Beach TG, Bigio EH, Cairns NJ, Carrillo MC. National Institute on Aging-Alzheimer’s Association guidelines for the neuropathologic assessment of Alzheimer’s disease. Alzheimers Dement. 2012;8(1):1–13.

28. Thal DR, Rub U, Orantes M, Braak H. Phases of A beta-deposition in the human brain and its relevance for the development of AD. Neurology. 2002;58(12):1791–1800.

29. Godrich D, Pasteris J, Martin ER, et al. Genome-wide association studies of TDP-43 proteinopathy and hippocampal sclerosis reveal shared genetic associations with APOE and TMEM106B. Alzheimer’s & Dementia. 2025;21(11):e70760. doi:10.1002/alz.70760

30. Bellenguez C, Kucukali F, Jansen IE, others. New insights into the genetic etiology of Alzheimer’s disease and related dementias. Nature Genetics. 2022;54(4):412–436. doi:10.1038/s41588-022-01024-z

31. Nalls MA, Blauwendraat C, Vallerga CL, others. Identification of novel risk loci, causal insights, and heritable risk for Parkinson’s disease: a meta-analysis of genome-wide association studies. Lancet Neurology. 2019;18(12):1091–1102. doi:10.1016/S1474-4422(19)30320-5

32. Finucane HK, Reshef YA, Anttila V, et al. Heritability enrichment of specifically expressed genes identifies disease-relevant tissues and cell types. Nat Genet. 2018;50(4):621–629. doi:10.1038/s41588-018-0081-4

33. Heng TS, Painter MW. Immunological Genome Project C. The Immunological Genome Project: networks of gene expression in immune cells. Nat Immunol. 2008;9(10):1091–1094.

34. Jin Y, Li F, Sonoustoun B, others. APOE4 exacerbates alpha-synuclein seeding activity and contributes to neurotoxicity in Alzheimer’s disease with Lewy body pathology. Acta Neuropathologica. 2022;143(6):641–662. doi:10.1007/s00401-022-02421-8

35. Tsuang D, Leverenz JB, Lopez OL, others. APOE epsilon4 increases risk for dementia in pure synucleinopathies. JAMA Neurology. 2013;70(2):223–228. doi:10.1001/jamaneurol.2013.600

36. Yang HS, Yu L, White CC, others. Evaluation of TDP-43 proteinopathy and hippocampal sclerosis in relation to APOE epsilon4 haplotype status: a community-based cohort study. Lancet Neurology. 2018;17(9):773–781. doi:10.1016/S1474-4422(18)30251-5

37. Ferencz B, Gerritsen L. Genetics and underlying pathology of dementia. Neuropsychol Rev. 2015;25(1):113–124.

38. Spires-Jones TL, Attems J, Thal DR. Interactions of pathological proteins in neurodegenerative diseases. Acta Neuropathol. 2017;134(2):187–205.

39. Tome SO, Tsaka G, Ronisz A, Ospitalieri S, Gawor K, Gomes LA. TDP-43 pathology is associated with increased tau burdens and seeding. Mol Neurodegener. 2023;18(1).

40. Rasmussen KL, Luo J, Nordestgaard BG, Tybjaerg-Hansen A, Frikke-Schmidt R. APOE and vascular disease: Sequencing and genotyping in general population cohorts. Atherosclerosis. 2023;385:117218. doi:10.1016/j.atherosclerosis.2023.117218

41. Katsumata Y, Shade LM, Hohman TJ, Schneider JA, Bennett DA, Farfel JM. Multiple gene variants linked to Alzheimer’s-type clinical dementia via GWAS are also associated with non-Alzheimer’s neuropathologic entities. Neurobiol Dis. 2022;174(105880).

42. Ahola-Olli AV, Wurtz P, Havulinna AS, Aalto K, Pitkanen N, Lehtimaki T. Genome-wide Association Study Identifies 27 Loci Influencing Concentrations of Circulating Cytokines and Growth Factors. Am J Hum Genet. 2017;100(1):40–50.

43. Borges MC, Haycock PC, Zheng J, Hemani G, Holmes MV, Davey Smith G. Role of circulating polyunsaturated fatty acids on cardiovascular diseases risk: analysis using Mendelian randomization and fatty acid genetic association data from over 114,000 UK Biobank participants. BMC Med. 2022;20(1).

44. Chen VL, Du X, Chen Y, Kuppa A, Handelman SK, Vohnoutka RB. Genome-wide association study of serum liver enzymes implicates diverse metabolic and liver pathology. Nat Commun. 2021;12(1).

45. Graham SE, Clarke SL, Wu KHH, et al. The power of genetic diversity in genome-wide association studies of lipids. Nature. 2021;600(7890):675–679. doi:10.1038/s41586-021-04064-3

46. Richardson TG, Leyden GM, Wang Q, Bell JA, Elsworth B, Davey Smith G. Characterising metabolomic signatures of lipid-modifying therapies through drug target mendelian randomisation. PLoS Biol. 2022;20(2).

47. Richardson TG, Sanderson E, Palmer TM, Ala-Korpela M, Ference BA, Davey Smith G. Evaluating the relationship between circulating lipoprotein lipids and apolipoproteins with risk of coronary heart disease: A multivariable Mendelian randomisation analysis. PLoS Med. 2020;17(3).

48. Schoeler T, Speed D, Porcu E, Pirastu N, Pingault JB, Kutalik Z. Participation bias in the UK Biobank distorts genetic associations and downstream analyses. Nat Hum Behav. 2023;7(7):1216–1227.

49. Davies G, Lam M, Harris SE. Study of 300,486 individuals identifies 148 independent genetic loci influencing general cognitive function. Nat Commun. 2018;2018;9(1):2098. doi:10.1038/s41467-018-04362-x

50. Lee JJ, Wedow R, Okbay A, Kong E, Maghzian O, Zacher M. Gene discovery and polygenic prediction from a genome-wide association study of educational attainment in 1.1 million individuals. Nat Genet. 2018;50(8):1112–1121.

51. Hollingworth P, Harold D, Sims R, others. Common variants at ABCA7, MS4A6A/MS4A4E, EPHA1, CD33 and CD2AP are associated with Alzheimer’s disease. Nature Genetics. 2011;43(5):429–435. doi:10.1038/ng.803

52. Naj AC, Jun G, Beecham GW, al et. Common variants at MS4A4/MS4A6E, CD2AP, CD33 and EPHA1 are associated with late-onset Alzheimer’s disease. Nat Genet. 2011;43(5):436–441. doi:10.1038/ng.801

53. Martinez A, Soriano E. Functions of ephrin/Eph interactions in the development of the nervous system: emphasis on the hippocampal system. Brain Research Reviews. 2005;49(2):211–226. doi:10.1016/j.brainresrev.2005.02.001

54. Lai KO, Ip N. Synapse development and plasticity: roles of ephrin/Eph receptor signaling. Current Opinion in Neurobiology. 2009;19(3):275–283. doi:10.1016/j.conb.2009.04.009

55. Cerezo M, Sollis E, Ji Y, et al. The NHGRI-EBI GWAS Catalog: standards for reusability, sustainability and diversity. Nucleic Acids Res. 2025;53(D1):D998–D1005. doi:10.1093/nar/gkae1070

56. Li YJ, Nuytemans K, La JO, Jiang R, Slifer SH, Sun S. Identification of novel genes for age-at-onset of Alzheimer’s disease by combining quantitative and survival trait analyses. Alzheimers Dement. 2023;19(7):3148–3157.

57. Huang L, Chambliss KL, Gao X, others. SR-B1 drives endothelial cell LDL transcytosis via DOCK4 to promote atherosclerosis. Nature. 2019;569(7757):565–569. doi:10.1038/s41586-019-1140-4

58. Bone WP, Siewert KM, Jha A, Klarin D, Damrauer SM, Program VAMV. Multi-trait association studies discover pleiotropic loci between Alzheimer’s disease and cardiometabolic traits. Alzheimers Res Ther. 2021;13(1).

59. Xie X, Lu Y, Wang B, Yin X, Chen J. DOCK4 is a Novel Prognostic Biomarker and Correlated with Immune Infiltrates in Colon Adenocarcinoma. Comb Chem High Throughput Screen. 2024;27(8):1119–1130.

60. Huang J, Huffman JE, Huang Y, Do Valle I, Assimes TL, Raghavan S. Genomics and phenomics of body mass index reveals a complex disease network. Nat Commun. 2022;13(1).

61. Kong M, Lee C. Genetic associations with C-reactive protein level and white blood cell count in the KARE study. Int J Immunogenet. 2013;40(2):120–125.

62. Surendran P, Feofanova EV, Lahrouchi N, Ntalla I, Karthikeyan S, Cook J. Discovery of rare variants associated with blood pressure regulation through meta-analysis of 1.3 million individuals. Nat Genet. 2020;52(12):1314–1332.

63. Foster EM, Dangla-Valls A, Lovestone S, Ribe EM, Buckley NJ. Clusterin in Alzheimer’s Disease: Mechanisms, Genetics, and Lessons From Other Pathologies. Front Neurosci. 2019;13:164. doi:10.3389/fnins.2019.00164

64. Chapuis J, Flaig A, Grenier-Boley B, others. Genome-wide, high-content siRNA screening identifies the Alzheimer’s genetic risk factor FERMT2 as a major modulator of APP metabolism. Acta Neuropathol. 2017;133(6):955–966. doi:10.1007/s00401-016-1652-z

65. Wang H, Lo MT, Rosenthal SB, Makowski C, Andreassen OA, Salem RM. Similar Genetic Architecture of Alzheimer’s Disease and Differential APOE Effect Between Sexes. Front Aging Neurosci. 2021;13(674318).

66. Zhang Q, Sidorenko J, Couvy-Duchesne B, Marioni RE, Wright MJ, Goate AM. Risk prediction of late-onset Alzheimer’s disease implies an oligogenic architecture. Nat Commun. 2020;11(1).

67. Manichaikul A, Mychaleckyj JC, Rich SS, Daly K, Sale M, Chen WM. Robust relationship inference in genome-wide association studies. Bioinformatics. 2010;26(22):2867–2873. doi:10.1093/bioinformatics/btq559

68. Zheng X, Levine D, Shen J, Gogarten SM, Laurie C, Weir BS. A high-performance computing toolset for relatedness and principal component analysis of SNP data. Bioinformatics. 2012;28(24):3326–3328. doi:10.1093/bioinformatics/bts606

69. Taliun D, Harris DN, Kessler MD, al et. Sequencing of 53,831 diverse genomes from the NHLBI TOPMed Program. Nature. 2021;590(7845):290–299. doi:10.1038/s41586-021-03205-y

70. Conomos MP, Miller MB, Thornton TA. Robust inference of population structure for ancestry prediction and correction of stratification in the presence of relatedness. Genet Epidemiol. 2015;39(4):276–293. doi:10.1002/gepi.21896

71. Montine TJ, Phelps CH, Beach TG, al et. National Institute on Aging-Alzheimer’s Association guidelines for the neuropathologic assessment of Alzheimer’s disease: a practical approach. Acta Neuropathol. 2012;123(1):1–11. doi:10.1007/s00401-011-0910-3

72. Vonsattel JP, Myers RH, Hedley-Whyte ET, Ropper AH, Bird ED, Richardson EPJr. Cerebral amyloid angiopathy without and with cerebral hemorrhages: a comparative histological study. Ann Neurol. 1991;30(5):637–649. doi:10.1002/ana.410300503

73. R Core Team. R: A Language and Environment for Statistical Computing. Published online 2025. https://www.R-project.org

74. Friendly M. Corrgrams. The American Statistician. 2002;56(4):316–324. doi:10.1198/000313002533

75. Zhan X, Hu Y, Li B, Abecasis GR, Liu D. RVTESTS: An efficient and comprehensive tool for rare variant association analysis using sequence data. Bioinformatics. 2016;32(9):1423–1426. doi:10.1093/bioinformatics/btw079

76. Christensen RH. Cumulative Link Models for Ordinal Regression with the R Package ordinal. Published online 2023.

77. Willer CJ, Li Y, Abecasis GR. METAL: fast and efficient meta-analysis of genomewide association scans. Bioinformatics. 2010;26(17):2190–2191. doi:10.1093/bioinformatics/btq340

78. Turner SD. qqman: an R package for visualizing GWAS results using Q-Q and manhattan plots. Journal of Open Source Software. 2018;3(25):731. doi:10.21105/joss.00731

79. Wickham H. Ggplot2: Elegant Graphics for Data. Springer; 2009.

80. Pruim RJ, Welch RP, Sanna S, others. LocusZoom: regional visualization of genome-wide association scan results. Bioinformatics. 2010;26(18):2336–2337. doi:10.1093/bioinformatics/btq419

81. Watanabe K, Taskesen E, van Bochoven A, Posthuma D. Functional mapping and annotation of genetic associations with FUMA. Nature Communications. 2017;8(1):1826. doi:10.1038/s41467-017-01261-5

82. Kircher M, Witten DM, Jain P, O’Roak BJ, Cooper GM, Shendure J. A general framework for estimating the relative pathogenicity of human genetic variants. Nature Genetics. 2014;46(3):310–315. doi:10.1038/ng.2892

83. Boyle EA, Hong EL, Hariharan M, others. Annotation of functional variation in personal genomes using RegulomeDB. Genome Research. 2012;22(9):1790–1797. doi:10.1101/gr.137323.112

84. Bulik-Sullivan BK, Loh PR, Finucane HK, et al. LD Score regression distinguishes confounding from polygenicity in genome-wide association studies. Nat Genet. 2015;47(3):291–295. doi:10.1038/ng.3211

