## Supplemental Tables for "Genome wide association study meta-analysis of neuropathologic lesions of Alzheimer’s disease and related dementias in a multi-site autopsy cohort"

Table and Figure List

**Supplemental Tables**

**Title**

Table S1: Frequency and proportions of neuropathology phenotypes
Table S2: Association of APOE e4, across neuropathology phenotypes (rs429358; chr19:44,908,684)
Table S3: All genome-wide association results with $p < 0.0001$
Table S4: Association of Bellenguez et al AD risk loci with AD related dementia pathologies
Table S5: Association of Bellenguez et al AD risk loci with cerebrovascular and vascular brain injury patho
Table S6: Association of Nalls et al PD risk loci with lewy body pathology
Table S7: Association of Nalls et al PD risk loci with AD hallmark pathologies
Table S8: Association of Nalls et al PD risk loci with related dementia pathologies
Table S9. Estimated SNP heritability across all AD/RD pathologies
Table S10. Estimated SNP-based genetic correlation (shared heritability) across between neuropathologi
Table S11: Cell/Tissue-specific heritability enrichment p-values (GTEx, multi tissue)
Table S12: Cell/Tissue-specific heritability enrichment coefficients and p-values (GTEx, multi tissue)
Table S13: Cell/Tissue-specific heritability enrichment p-values (ImmGen)
Table S14: Cell/Tissue-specific heritability enrichment coefficients and p-values (ImmGen)
Table S15: MAGMA pathway analysis results
Table S16: MAGMA tissue-specific associations
Table S17: MAGMA tissue-specific associations, general tissues
Table S18: Genotype Imputation Quality (R2) by batch, for genome-wide associated variants
Table S19: Neuropathology phenotype coding

**Supplemental Figures**

**Title**

Figure S1: Correlation plot of neuropathology and related variables
Figure S2:QQ plots for genome-wide association analyses
Figure S3: P-value by genomic position for A score, B score, and C score
Figure S4: Regional association plot for genome-wide significant variants from the amyloid plaque (prese
Figure S5: P-value by genomic position for cerebral atherosclerosis (any/none) and cerebrovascular dise
Figure S6: Regional association plot for genome-wide significant variants from cerebral atherosclerosis (a
Figure S7: P-value by genomic position for association with infarcts/lacunes analyses, and regional associ
Figure S8: P-value by genomic position for association with lewy body (PD Braak) analyses, and regional a
Figure S9: Regional association plot for the *BIN1* locus, for A Score (NP/Thal), B Score (NFT Braak), and C
Figure S10: Regional association plot for the *PICALM/EED* locus, for A Score (NP/Thal), B Score (NFT Braa
Figure S11: P-value by genomic position for association with Thal phase, NFT Braak, and ADNC (ABC) scor
Figure S12: P-value by genomic position for association with cerebral amyloid angiopathy (CAA) analysis,
Figure S13: P-value by genomic position for association with microinfarcts, hemorrhages, and whole brai
Figure S14: P-value by genomic position for association with lewy bodies
Figure S15: P-value by genomic position for association with TDP-43 proteinopathy and hippocampal scle
Figure S16: Regional association plot for the TMEM106B locus, for TDP-43 proteinopathy and hippocamp
Figure S17: Cell-specific heritability enrichment results (multi-tissue GTEx)
Figure S18: Cell-specific heritability enrichment results (ImmGen)

| Type | Number |
|---|---|
| Table | S1 |
| Table | S2 |
| Table | S3 |
| Table | S4 |
| Table | S5 |
| Table | S6 |
| Table | S7 |
| Table | S8 |
| Table | S9 |
| Table | S10 |
| Table | S11 |
| Table | S12 |
| Table | S13 |
| Table | S14 |
| Table | S15 |
| Table | S16 |
| Table | S17 |
| Table | S18 |
| Table | S19 |

| Type | Number |
|---|---|
| Figure | S1 |
| Figure | S2 |
| Figure | S3 |
| Figure | S4 |
| Figure | S5 |
| Figure | S6 |
| Figure | S7 |
| Figure | S8 |
| Figure | S9 |
| Figure | S10 |
| Figure | S11 |
| Figure | S12 |
| Figure | S13 |
| Figure | S14 |
| Figure | S15 |
| Figure | S16 |
| Figure | S17 |
| Figure | S18 |

**Table S1: Frequency and proportion of neuropathological lesions**

| Thal Phase | | Freq | Prop |
|---|---|---|---|
| | 0 | 412 | 0.114 |
| | 1 | 266 | 0.073 |
| | 2 | 202 | 0.056 |
| | 3 | 397 | 0.11 |
| | 4 | 647 | 0.179 |
| | 5 | 1700 | 0.469 |
| Total | | 3624 | 1 |

| A Score | | Freq | Prop |
|---|---|---|---|
| | 0 | 412 | 0.112 |
| | 1 | 470 | 0.128 |
| | 2 | 403 | 0.11 |
| | 3 | 2389 | 0.65 |
| Total | | 3674 | 1 |

| CERAD (C Score) | | Freq | Prop |
|---|---|---|---|
| | 0 | 1959 | 0.176 |
| | 1 | 1061 | 0.095 |
| | 2 | 2111 | 0.189 |
| | 3 | 6009 | 0.539 |
| Total | | 11140 | 1 |

| AP (any/none) | | Freq | Prop |
|---|---|---|---|
| | 0 | 1627 | 0.146 |
| | 1 | 9518 | 0.854 |
| Total | | 11145 | 1 |

| AD Braak | | Freq | Prop |
|---|---|---|---|
| | 0 | 418 | 0.036 |
| | 1 | 722 | 0.062 |
| | 2 | 1051 | 0.091 |
| | 3 | 1297 | 0.112 |

| | | | |
|---|---|---|---|
| | 4 | 1791 | 0.155 |
| | 5 | 2735 | 0.236 |
| | 6 | 3566 | 0.308 |
| Total | | 11580 | 1 |

| B Score | | Freq | Prop |
|---|---|---|---|
| | 0 | 418 | 0.036 |
| | 1 | 1776 | 0.153 |
| | 2 | 3088 | 0.267 |
| | 3 | 6301 | 0.544 |
| Total | | 11583 | 1 |

| ADNC Score | | Freq | Prop |
|---|---|---|---|
| | 0 | 402 | 0.111 |
| | 1 | 618 | 0.17 |
| | 2 | 787 | 0.216 |
| | 3 | 1829 | 0.503 |
| Total | | 3636 | 1 |

| PD Braak (any/none) | | Freq | Prop |
|---|---|---|---|
| | 0 | 6914 | 0.659 |
| | 1 | 3578 | 0.341 |
| Total | | 10492 | 1 |

| PD Braak (3 cat) | | Freq | Prop |
|---|---|---|---|
| | 0 | 6914 | 0.659 |
| | 1 | 346 | 0.033 |
| | 2 | 3232 | 0.308 |
| Total | | 10492 | 1 |

| PD Braak | | Freq | Prop |
|---|---|---|---|
| | 0 | 6914 | 0.659 |
| | 1 | 346 | 0.033 |
| | 2 | 1390 | 0.132 |

| | | | |
|---|---|---|---|
| | 3 | 1434 | 0.137 |
| | 4 | 408 | 0.039 |
| Total | | 10492 | 1 |

| Infarcts/lacunes | | Freq | Prop |
|---|---|---|---|
| | 0 | 6649 | 0.755 |
| | 1 | 2153 | 0.245 |
| Total | | 8802 | 1 |

| Hemorrhages | | Freq | Prop |
|---|---|---|---|
| | 0 | 6316 | 0.942 |
| | 1 | 386 | 0.058 |
| Total | | 6702 | 1 |

| Microinfarcts | | Freq | Prop |
|---|---|---|---|
| | 0 | 6647 | 0.748 |
| | 1 | 2238 | 0.252 |
| Total | | 8885 | 1 |

| WMR (any/none) | | Freq | Prop |
|---|---|---|---|
| | 0 | 4811 | 0.729 |
| | 1 | 1791 | 0.271 |
| Total | | 6602 | 1 |

| WMR | | Freq | Prop |
|---|---|---|---|
| | 0 | 928 | 0.408 |
| | 1 | 667 | 0.294 |
| | 2 | 455 | 0.2 |
| | 3 | 222 | 0.098 |
| Total | | 2272 | 1 |

| WBVD | | Freq | Prop |
|---|---|---|---|
| | 0 | 4956 | 0.491 |
| | 1 | 5144 | 0.509 |

| | | |
|---|---|---|
| Total | 10100 | 1 |

| HS | Freq | Prop |
|---|---|---|
| 0 | 8337 | 0.873 |
| 1 | 1208 | 0.127 |
| Total | 9545 | 1 |

| TDP-43(any/none) | Freq | Prop |
|---|---|---|
| 0 | 2871 | 0.613 |
| 1 | 1811 | 0.387 |
| Total | 4682 | 1 |

| TDP-43 (3 cat) | Freq | Prop |
|---|---|---|
| 0 | 2299 | 0.597 |
| 1 | 464 | 0.12 |
| 2 | 586 | 0.152 |
| 3 | 504 | 0.131 |
| Total | 3853 | 1 |

| CBVD (any/none) | Freq | Prop |
|---|---|---|
| 0 | 771 | 0.076 |
| 1 | 9337 | 0.924 |
| Total | 10108 | 1 |

| Atherosclerosis | Freq | Prop |
|---|---|---|
| 0 | 1856 | 0.212 |
| 1 | 3394 | 0.387 |
| 2 | 2571 | 0.293 |
| 3 | 945 | 0.108 |
| Total | 8766 | 1 |

| Atherosclerosis (any/none) | Freq | Prop |
|---|---|---|
| 0 | 1856 | 0.212 |
| 1 | 6910 | 0.788 |

| | | |
|---|---|---|
| Total | 8766 | 1 |

| Arteriolosclerosis | Freq | Prop |
|---|---|---|
| 0 | 1883 | 0.236 |
| 1 | 2552 | 0.32 |
| 2 | 2460 | 0.309 |
| 3 | 1070 | 0.134 |
| Total | 7965 | 1 |

| Arteriolosclerosis (any/none) | Freq | Prop |
|---|---|---|
| 0 | 1883 | 0.236 |
| 1 | 6082 | 0.764 |
| Total | 7965 | 1 |

| CAA | Freq | Prop |
|---|---|---|
| 0 | 3318 | 0.377 |
| 1 | 2583 | 0.293 |
| 2 | 1899 | 0.216 |
| 3 | 1008 | 0.114 |
| Total | 8808 | 1 |

| CAA (any/none) | Freq | Prop |
|---|---|---|
| 0 | 3484 | 0.356 |
| 1 | 6313 | 0.644 |
| Total | 9797 | 1 |

Abbreviations: AP, amyloid plaques; CAA, cerebral amyloid angiopathy; CBVD, cerebrovascular disease; PD Parkinson disease (e.g., Lewy bodies); HS, hippocampal sclerosis; VBI, vascular brain injury; WBVD, whole brain vascular disease; WMR, white matter rarefaction

**Table S2: Association of APOE e4, across neuropathology phenotypes (rs429358; chr19:44,908,684)**

| Category | Phenotype | Beta | SE | p-value | Het (Q) sta | Het df | Het p-value |
|---|---|---|---|---|---|---|---|
| ADNC | ADNC Score | 0.9873 | 0.0629 | **1.92E-55** | 7.513 | 9 | 5.84E-01 |
| | A Score | 1.2414 | 0.0779 | **3.68E-57** | 6.453 | 8 | 5.97E-01 |
| | AP (any/none) | 1.2751 | 0.0528 | **9.88E-129** | 8.071 | 12 | 7.80E-01 |
| | CERAD (C Score) | 1.0294 | 0.037 | **3.89E-170** | 28.821 | 15 | **1.70E-02** |
| | Thal Phase | 1.0461 | 0.0625 | **6.38E-63** | 8.342 | 8 | 4.01E-01 |
| | B Score | 0.9129 | 0.0364 | **1.95E-138** | 27.805 | 15 | **2.28E-02** |
| | AD Braak | 0.7616 | 0.0295 | **8.06E-147** | 45.18 | 16 | **1.30E-04** |
| CBVD | Arteriolosclerosis | 0.1118 | 0.0348 | **1.34E-03** | 22.058 | 9 | **8.70E-03** |
| | Arteriolosclerosis (any/none) | -0.0286 | 0.0457 | 5.32E-01 | 13.045 | 7 | 7.10E-02 |
| | Atherosclerosis | 0.0837 | 0.0333 | **1.20E-02** | 7.594 | 9 | 5.76E-01 |
| | Atherosclerosis (any/none) | 0.0572 | 0.0445 | 1.98E-01 | 6.188 | 9 | 7.21E-01 |
| | CAA | 1.0072 | 0.0344 | **3.14E-188** | 30.488 | 10 | **7.13E-04** |
| | CAA (any/none) | 0.9239 | 0.037 | **1.35E-137** | 24.293 | 11 | **1.16E-02** |
| | Any CBVD | 0.4542 | 0.0614 | **1.41E-13** | 33.667 | 8 | **4.67E-05** |
| Other RD | PD Braak (any/none) | 0.3068 | 0.035 | **1.90E-18** | 53.195 | 14 | **1.76E-06** |
| | PD Braak | 0.2755 | 0.0331 | **9.17E-17** | 58.675 | 14 | **2.00E-07** |
| | PD Braak (3 cat) | 0.2939 | 0.034 | **5.73E-18** | 60.466 | 14 | **9.72E-08** |
| | HS | 0.3186 | 0.0542 | **4.16E-09** | 8.775 | 11 | 6.43E-01 |
| | TDP-43 (3 cat) | 0.5306 | 0.0624 | **1.88E-17** | 8.786 | 9 | 4.57E-01 |
| | TDP-43 (any/none) | 0.5378 | 0.0581 | **2.02E-20** | 6.54 | 10 | 7.68E-01 |
| VBI | Hemorrhages | 0.0394 | 0.0851 | 6.43E-01 | 3.853 | 4 | 4.26E-01 |
| | Infarcts/lacunes | 0.0111 | 0.0455 | 8.07E-01 | 8.897 | 9 | 4.47E-01 |
| | Microinfarcts | 0.0086 | 0.0446 | 8.47E-01 | 7.978 | 9 | 5.36E-01 |
| | WBVD | 0.0079 | 0.0346 | 8.19E-01 | 26.75 | 12 | **8.39E-03** |
| | WMR | 0.1248 | 0.0644 | 5.26E-02 | 5.557 | 5 | 3.52E-01 |
| | WMR (any/none) | 0.0479 | 0.0473 | 3.11E-01 | 7.413 | 5 | 1.92E-01 |

Abbreviations: AP, amyloid plaques; CAA, cerebral amyloid angiopathy; CBVD, cerebrovascular disease; RD, (AD) related dementias; PD Parkinson disease (e.g., Lewy bodies); HS, hippocampal sclerosis; VBI, vascular brain injury; WBVD, whole brain vascular disease; SE, standard error WMR, white matter rarefaction

Het Q, Het df and Hetp-value note the heterogeneity test statistic, degrees of freedom, and p-value, respectively.

Referant/alternate alleles: T/C
SE: standard error

**Table S3: All genome-wide association results with p < 0.0001**

| Phenotype | Chrm | Position | Variant | A1 | A2 | Beta | SE | p-value | Direction | Het (Q) sta | Het df | Het p-value |
|---|---|---|---|---|---|---|---|---|---|---|---|---|
| A Score | chr1 | 1,672,155 | chr1:1672155:G:A | a | g | -0.3056 | 0.0749 | 4.48E-05 | ----+--+- | 9.478 | 8 | 0.3036 |
| A Score | chr1 | 1,675,132 | chr1:1675132:A:G | a | g | -0.2838 | 0.0714 | 7.10E-05 | ----+---- | 4.736 | 8 | 0.7854 |
| A Score | chr1 | 4,465,096 | chr1:4465096:G:A | a | g | 1.0889 | 0.2691 | 5.19E-05 | ??++?+?+? | 2.244 | 3 | 0.5233 |
| A Score | chr1 | 7,476,178 | chr1:7476178:G:A | a | g | 0.2076 | 0.0531 | 9.29E-05 | +++++++++ | 3.965 | 8 | 0.8603 |
| A Score | chr1 | 53,078,925 | chr1:53078925:T:C | t | c | 0.3122 | 0.0793 | 8.19E-05 | +-+++++-+ | 5.247 | 8 | 0.7309 |
| A Score | chr1 | 53,101,146 | chr1:53101146:C:T | t | c | -0.3118 | 0.0788 | 7.54E-05 | -+-----+- | 5.419 | 8 | 0.712 |
| A Score | chr1 | 58,668,935 | chr1:58668935:C:CAG | cag | c | -0.6025 | 0.1528 | 8.08E-05 | ----+---- | 9.415 | 8 | 0.3085 |
| A Score | chr1 | 79,271,598 | chr1:79271598:T:C | t | c | -0.2134 | 0.0542 | 8.24E-05 | ----+---- | 6.665 | 8 | 0.5732 |
| A Score | chr1 | 79,275,797 | chr1:79275797:G:C | c | g | 0.2148 | 0.0544 | 7.97E-05 | ++++-++++ | 7.113 | 8 | 0.5245 |
| A Score | chr1 | 79,276,059 | chr1:79276059:A:G | a | g | -0.214 | 0.0539 | 7.25E-05 | ----+---- | 6.581 | 8 | 0.5824 |
| A Score | chr1 | 104,534,131 | chr1:104534131:T:A | a | t | 0.2627 | 0.0652 | 5.61E-05 | ++++-++-+ | 3.824 | 8 | 0.8726 |
| A Score | chr1 | 104,534,188 | chr1:104534188:C:T | t | c | 0.2592 | 0.0651 | 6.82E-05 | ++++-++-+ | 4.222 | 8 | 0.8365 |
| A Score | chr1 | 110,523,406 | chr1:110523406:G:T | t | g | -0.2277 | 0.0562 | 5.05E-05 | +-------- | 2.46 | 8 | 0.9636 |
| A Score | chr1 | 111,609,639 | chr1:111609639:T:G | t | g | -0.4809 | 0.1226 | 8.74E-05 | +-------- | 8.338 | 8 | 0.4012 |
| A Score | chr1 | 111,620,705 | chr1:111620705:T:C | t | c | -0.5233 | 0.1322 | 7.58E-05 | ?-------- | 5.227 | 7 | 0.6323 |
| A Score | chr1 | 111,627,740 | chr1:111627740:CA:C | ca | c | -0.6163 | 0.1565 | 8.25E-05 | ?----+--- | 6.718 | 7 | 0.4588 |
| A Score | chr1 | 111,660,750 | chr1:111660750:C:T | t | c | 0.5327 | 0.1333 | 6.41E-05 | ?++++++++ | 4.414 | 7 | 0.731 |
| A Score | chr1 | 165,444,943 | chr1:165444943:C:T | t | c | 0.8034 | 0.1931 | 3.16E-05 | -++++++-- | 14.87 | 8 | 0.06173 |
| A Score | chr1 | 181,837,497 | chr1:181837497:C:T | t | c | 0.9818 | 0.2509 | 9.12E-05 | --++??+?? | 3.291 | 4 | 0.5104 |
| A Score | chr1 | 183,911,830 | chr1:183911830:C:T | t | c | 0.3632 | 0.0872 | 3.13E-05 | +++++++++ | 8.828 | 8 | 0.357 |
| A Score | chr1 | 199,098,150 | chr1:199098150:T:C | t | c | -0.2115 | 0.0529 | 6.41E-05 | -------+- | 3.822 | 8 | 0.8728 |
| A Score | chr1 | 199,099,485 | chr1:199099485:A:G | a | g | -0.2109 | 0.0528 | 6.53E-05 | +------+- | 4.095 | 8 | 0.8485 |
| A Score | chr1 | 199,101,730 | chr1:199101730:C:T | t | c | 0.2149 | 0.0528 | 4.76E-05 | +++++++-+ | 3.755 | 8 | 0.8786 |
| A Score | chr1 | 199,105,262 | chr1:199105262:C:T | t | c | 0.2137 | 0.0534 | 6.18E-05 | +++++++-+ | 3.503 | 8 | 0.899 |
| A Score | chr1 | 199,127,383 | chr1:199127383:G:A | a | g | 0.2134 | 0.0533 | 6.23E-05 | ++++-++-+ | 4.142 | 8 | 0.8441 |
| A Score | chr1 | 199,135,585 | chr1:199135585:T:G | t | g | -0.2261 | 0.0541 | 2.87E-05 | -------+- | 3.24 | 8 | 0.9184 |
| A Score | chr1 | 199,144,761 | chr1:199144761:G:A | a | g | 0.3007 | 0.0763 | 8.14E-05 | ++++-++-+ | 6.039 | 8 | 0.6428 |
| A Score | chr1 | 199,144,762 | chr1:199144762:T:A | a | t | 0.2998 | 0.0763 | 8.59E-05 | ++++-++-+ | 6.027 | 8 | 0.6442 |
| A Score | chr1 | 199,148,095 | chr1:199148095:T:C | t | c | -0.2984 | 0.0764 | 9.43E-05 | ----+--+- | 6.086 | 8 | 0.6376 |
| A Score | chr1 | 199,148,459 | chr1:199148459:G:A | a | g | 0.2985 | 0.0764 | 9.35E-05 | ++++-++-+ | 6.079 | 8 | 0.6384 |
| A Score | chr1 | 222,503,444 | chr1:222503444:C:T | t | c | 0.7389 | 0.1727 | 1.88E-05 | +++++++++ | 2.755 | 8 | 0.9488 |
| A Score | chr1 | 222,514,632 | chr1:222514632:AAAAAAC:A | a | aaaaaac | 0.6322 | 0.1614 | 9.00E-05 | +++++++++ | 4.727 | 8 | 0.7864 |
| A Score | chr1 | 222,533,805 | chr1:222533805:C:T | t | c | 0.643 | 0.16 | 5.82E-05 | +++++++++ | 4.661 | 8 | 0.7931 |
| A Score | chr1 | 223,556,772 | chr1:223556772:G:A | a | g | 0.4565 | 0.1096 | 3.11E-05 | +-++?+++- | 7.048 | 7 | 0.4239 |
| A Score | chr1 | 242,462,207 | chr1:242462207:T:A | a | t | -0.5251 | 0.1331 | 8.00E-05 | --------- | 7.239 | 8 | 0.5111 |
| A Score | chr10 | 2,225,597 | chr10:2225597:C:T | t | c | 0.5583 | 0.1317 | 2.24E-05 | ++++--+++ | 9.872 | 8 | 0.2741 |
| A Score | chr10 | 2,245,630 | chr10:2245630:T:C | t | c | -0.5338 | 0.1279 | 3.01E-05 | ----++--- | 11.002 | 8 | 0.2016 |
| A Score | chr10 | 4,792,575 | chr10:4792575:G:A | a | g | -0.3627 | 0.0884 | 4.05E-05 | --------- | 9.051 | 8 | 0.338 |
| A Score | chr10 | 4,795,384 | chr10:4795384:A:G | a | g | 0.3565 | 0.0884 | 5.55E-05 | +++++++++ | 9.545 | 8 | 0.2984 |
| A Score | chr10 | 4,798,824 | chr10:4798824:G:T | t | g | -0.3635 | 0.0887 | 4.20E-05 | --------- | 9.415 | 8 | 0.3085 |
| A Score | chr10 | 4,798,825 | chr10:4798825:C:T | t | c | -0.3634 | 0.0887 | 4.21E-05 | --------- | 9.409 | 8 | 0.309 |
| A Score | chr10 | 25,665,405 | chr10:25665405:G:A | a | g | 0.5907 | 0.1511 | 9.29E-05 | +++++++-+ | 8.875 | 8 | 0.3529 |
| A Score | chr10 | 49,069,916 | chr10:49069916:G:A | a | g | 1.9131 | 0.4311 | 9.09E-06 | ++?????++ | 4.841 | 3 | 0.1838 |
| A Score | chr10 | 60,036,585 | chr10:60036585:G:A | a | g | 0.7439 | 0.1891 | 8.33E-05 | +-++++--+ | 10.43 | 8 | 0.2361 |

| | | | | | | | | | | | | |
|---|---|---|---|---|---|---|---|---|---|---|---|---|
| A Score | chr10 | 91,055,348 | chr10:91055348:G:A | a | g | -0.4208 | 0.0994 | 2.31E-05 | -+--?---- | 2.858 | 7 | 0.8978 |
| A Score | chr10 | 91,101,459 | chr10:91101459:C:A | a | c | -0.3654 | 0.0902 | 5.15E-05 | -+------- | 2.35 | 8 | 0.9684 |
| A Score | chr10 | 91,109,797 | chr10:91109797:G:C | c | g | -0.3505 | 0.0896 | 9.22E-05 | -+------- | 2.777 | 8 | 0.9475 |
| A Score | chr10 | 91,111,470 | chr10:91111470:G:C | c | g | -0.3497 | 0.0899 | 9.96E-05 | -+------- | 2.882 | 8 | 0.9415 |
| A Score | chr10 | 96,255,657 | chr10:96255657:G:T | t | g | 0.2221 | 0.0553 | 5.93E-05 | +-++++++- | 18.674 | 8 | 0.0167 |
| A Score | chr10 | 110,201,432 | chr10:110201432:G:A | a | g | 0.7797 | 0.1999 | 9.60E-05 | ++++-++-+ | 5.467 | 8 | 0.7067 |
| A Score | chr10 | 112,961,123 | chr10:112961123:C:T | t | c | 0.791 | 0.1924 | 3.96E-05 | -++++++++ | 6.272 | 8 | 0.6168 |
| A Score | chr10 | 115,267,830 | chr10:115267830:G:A | a | g | 0.7573 | 0.194 | 9.46E-05 | ?-++?+--+ | 15.577 | 6 | 0.01621 |
| A Score | chr10 | 115,433,397 | chr10:115433397:T:C | t | c | -0.8449 | 0.2154 | 8.74E-05 | ?+--?-++? | 10.048 | 5 | 0.07389 |
| A Score | chr10 | 115,569,206 | chr10:115569206:A:G | a | g | -0.8371 | 0.2138 | 9.01E-05 | ++--?-++? | 11.022 | 6 | 0.08771 |
| A Score | chr10 | 115,634,954 | chr10:115634954:T:C | t | c | -0.8007 | 0.1968 | 4.74E-05 | ++--+-++- | 14.176 | 8 | 0.07728 |
| A Score | chr10 | 115,660,835 | chr10:115660835:A:G | a | g | -0.8401 | 0.2136 | 8.39E-05 | ++--?-++? | 11.199 | 6 | 0.0824 |
| A Score | chr10 | 115,678,361 | chr10:115678361:A:G | a | g | -0.8045 | 0.1948 | 3.62E-05 | ++--+-++- | 13.51 | 8 | 0.09548 |
| A Score | chr10 | 115,726,027 | chr10:115726027:T:A | a | t | 0.8606 | 0.1995 | 1.60E-05 | --++-+--+ | 12.875 | 8 | 0.1162 |
| A Score | chr10 | 115,791,502 | chr10:115791502:A:G | a | g | -0.8409 | 0.213 | 7.87E-05 | ++--+-++? | 11.994 | 7 | 0.1008 |
| A Score | chr10 | 115,805,170 | chr10:115805170:A:G | a | g | -0.834 | 0.2129 | 8.97E-05 | ++--?-++? | 11.341 | 6 | 0.07838 |
| A Score | chr11 | 586,185 | chr11:586185:G:A | a | g | 0.7739 | 0.1951 | 7.28E-05 | -?+++++-? | 4.042 | 6 | 0.671 |
| A Score | chr11 | 596,102 | chr11:596102:G:A | a | g | 0.7739 | 0.1951 | 7.28E-05 | -?+++++-? | 4.042 | 6 | 0.671 |
| A Score | chr11 | 624,674 | chr11:624674:C:T | t | c | 0.7915 | 0.2009 | 8.17E-05 | -?+++++-? | 4.016 | 6 | 0.6745 |
| A Score | chr11 | 629,639 | chr11:629639:G:A | a | g | 0.7719 | 0.1942 | 7.07E-05 | -?+++++-? | 3.808 | 6 | 0.7027 |
| A Score | chr11 | 2,847,638 | chr11:2847638:G:A | a | g | 0.4551 | 0.1075 | 2.31E-05 | ++++-++++ | 3.5 | 8 | 0.8992 |
| A Score | chr11 | 7,617,805 | chr11:7617805:C:G | c | g | -0.2408 | 0.0555 | 1.46E-05 | +---+--+- | 5.02 | 8 | 0.7554 |
| A Score | chr11 | 21,094,713 | chr11:21094713:G:A | a | g | 1.0012 | 0.2252 | 8.78E-06 | +?++-++-+ | 3.251 | 7 | 0.8608 |
| A Score | chr11 | 21,147,168 | chr11:21147168:G:A | a | g | 0.8591 | 0.2155 | 6.72E-05 | +?++-++-+ | 4.5 | 7 | 0.7207 |
| A Score | chr11 | 23,987,715 | chr11:23987715:A:AAAGTGAG | a | aaagtgag | -0.3124 | 0.0777 | 5.83E-05 | ++------- | 5.057 | 8 | 0.7514 |
| A Score | chr11 | 34,484,379 | chr11:34484379:C:G | c | g | 0.2246 | 0.0528 | 2.12E-05 | ++++++-++ | 9.857 | 8 | 0.2752 |
| A Score | chr11 | 34,484,791 | chr11:34484791:T:G | t | g | 0.221 | 0.0528 | 2.85E-05 | ++++++-++ | 9.837 | 8 | 0.2767 |
| A Score | chr11 | 34,489,994 | chr11:34489994:C:A | a | c | 0.214 | 0.054 | 7.30E-05 | ++++++-++ | 7.025 | 8 | 0.5339 |
| A Score | chr11 | 34,490,562 | chr11:34490562:T:C | t | c | -0.2173 | 0.054 | 5.63E-05 | ------+-- | 7.137 | 8 | 0.5219 |
| A Score | chr11 | 34,490,630 | chr11:34490630:T:C | t | c | -0.2169 | 0.054 | 5.95E-05 | ------+-- | 7.175 | 8 | 0.5179 |
| A Score | chr11 | 34,491,015 | chr11:34491015:C:G | c | g | -0.2178 | 0.054 | 5.45E-05 | ------+-- | 7.122 | 8 | 0.5235 |
| A Score | chr11 | 34,491,883 | chr11:34491883:T:A | a | t | 0.2152 | 0.054 | 6.64E-05 | ++++++-++ | 6.944 | 8 | 0.5426 |
| A Score | chr11 | 34,492,724 | chr11:34492724:C:G | c | g | -0.2133 | 0.0539 | 7.64E-05 | ------+-- | 7.035 | 8 | 0.5329 |
| A Score | chr11 | 34,493,394 | chr11:34493394:T:G | t | g | -0.2122 | 0.0538 | 8.05E-05 | ------+-- | 7.043 | 8 | 0.532 |
| A Score | chr11 | 34,495,437 | chr11:34495437:G:A | a | g | 0.2262 | 0.0541 | 2.93E-05 | +-++++-++ | 8.001 | 8 | 0.4334 |
| A Score | chr11 | 61,083,746 | chr11:61083746:G:A | a | g | 1.2353 | 0.2958 | 2.97E-05 | ?-?+??-++ | 3.752 | 4 | 0.4406 |
| A Score | chr11 | 81,986,583 | chr11:81986583:G:A | a | g | 0.7157 | 0.1838 | 9.83E-05 | ++++-+-++ | 4.187 | 8 | 0.8398 |
| A Score | chr11 | 94,388,385 | chr11:94388385:C:T | t | c | 0.5884 | 0.1497 | 8.43E-05 | -++++++-+ | 3.757 | 8 | 0.8784 |
| A Score | chr11 | 132,640,520 | chr11:132640520:T:C | t | c | -0.4122 | 0.1039 | 7.30E-05 | ++--+--+- | 7.835 | 8 | 0.4498 |
| A Score | chr11 | 132,650,657 | chr11:132650657:T:G | t | g | -0.4879 | 0.1195 | 4.46E-05 | +---+-++- | 7.987 | 8 | 0.4348 |
| A Score | chr11 | 132,653,701 | chr11:132653701:G:A | a | g | 0.6083 | 0.1545 | 8.20E-05 | ?-++-++-+ | 7.735 | 7 | 0.3565 |
| A Score | chr11 | 132,655,547 | chr11:132655547:A:G | a | g | -0.4735 | 0.1208 | 8.91E-05 | +---+-++- | 7.514 | 8 | 0.4823 |
| A Score | chr11 | 132,656,430 | chr11:132656430:G:A | a | g | 0.4715 | 0.1207 | 9.36E-05 | -+++-+--+ | 7.075 | 8 | 0.5286 |
| A Score | chr11 | 132,667,983 | chr11:132667983:C:G | c | g | -0.6557 | 0.1572 | 3.02E-05 | ++--+--+- | 8.902 | 8 | 0.3506 |
| A Score | chr12 | 11,330,036 | chr12:11330036:C:T | t | c | 0.7264 | 0.1801 | 5.47E-05 | ?-++-++++ | 4.275 | 7 | 0.7476 |
| A Score | chr12 | 11,341,029 | chr12:11341029:C:T | t | c | 0.729 | 0.1801 | 5.18E-05 | ??++-++++ | 3.78 | 6 | 0.7064 |

| | | | | | | | | | | | | |
|---|---|---|---|---|---|---|---|---|---|---|---|---|
| A Score | chr12 | 11,385,828 | chr12:11385828:A:G | a | g | -0.6842 | 0.1737 | 8.17E-05 | +?----+-- | 5.787 | 7 | 0.5648 |
| A Score | chr12 | 11,417,453 | chr12:11417453:T:A | a | t | 0.741 | 0.1835 | 5.41E-05 | -?++-+-++ | 4.132 | 7 | 0.7645 |
| A Score | chr12 | 12,427,249 | chr12:12427249:C:T | t | c | 0.7153 | 0.1729 | 3.53E-05 | ++++-++-+ | 2.421 | 8 | 0.9653 |
| A Score | chr12 | 23,951,596 | chr12:23951596:T:A | a | t | -0.3042 | 0.0636 | 1.73E-06 | ----+---- | 6.075 | 8 | 0.6388 |
| A Score | chr12 | 23,954,482 | chr12:23954482:G:A | a | g | -0.3283 | 0.0625 | 1.47E-07 | --------- | 5.333 | 8 | 0.7214 |
| A Score | chr12 | 23,955,303 | chr12:23955303:A:G | a | g | 0.2633 | 0.0611 | 1.62E-05 | -++++++++ | 7.694 | 8 | 0.4639 |
| A Score | chr12 | 23,956,276 | chr12:23956276:G:GA | g | ga | 0.2824 | 0.0616 | 4.53E-06 | -++++++++ | 7.519 | 8 | 0.4818 |
| A Score | chr12 | 23,956,311 | chr12:23956311:T:C | t | c | 0.2969 | 0.0608 | 1.03E-06 | -++++++++ | 7.306 | 8 | 0.504 |
| A Score | chr12 | 23,956,707 | chr12:23956707:A:T | a | t | 0.2803 | 0.0615 | 5.16E-06 | -++++++++ | 7.48 | 8 | 0.4859 |
| A Score | chr12 | 56,285,425 | chr12:56285425:G:C | c | g | 1.347 | 0.294 | 4.61E-06 | ?+++??+-? | 5.242 | 4 | 0.2633 |
| A Score | chr12 | 56,343,467 | chr12:56343467:C:A | a | c | 1.3423 | 0.2944 | 5.14E-06 | ?+++??+-? | 5.422 | 4 | 0.2467 |
| A Score | chr12 | 71,965,900 | chr12:71965900:A:G | a | g | 0.5302 | 0.133 | 6.72E-05 | +-+++++++ | 5.427 | 8 | 0.7112 |
| A Score | chr12 | 71,985,632 | chr12:71985632:C:T | t | c | -0.5285 | 0.133 | 7.05E-05 | -+------- | 5.545 | 8 | 0.698 |
| A Score | chr12 | 72,001,546 | chr12:72001546:C:T | t | c | -0.5238 | 0.1324 | 7.58E-05 | -+------- | 5.074 | 8 | 0.7497 |
| A Score | chr12 | 73,868,601 | chr12:73868601:C:G | c | g | -0.9346 | 0.2375 | 8.32E-05 | -+--?--?? | 1.944 | 5 | 0.8568 |
| A Score | chr12 | 89,004,612 | chr12:89004612:C:T | t | c | 0.545 | 0.1398 | 9.64E-05 | -+++-+--+ | 11.001 | 8 | 0.2016 |
| A Score | chr12 | 89,009,210 | chr12:89009210:C:A | a | c | 0.5453 | 0.1398 | 9.62E-05 | -+++-+-?+ | 8.155 | 7 | 0.3191 |
| A Score | chr12 | 89,014,807 | chr12:89014807:C:A | a | c | 0.5638 | 0.1387 | 4.82E-05 | -+++?+-?+ | 7.13 | 6 | 0.309 |
| A Score | chr12 | 89,017,961 | chr12:89017961:G:C | c | g | 0.5633 | 0.1392 | 5.22E-05 | -+++?+-?+ | 7.125 | 6 | 0.3094 |
| A Score | chr12 | 89,354,271 | chr12:89354271:T:C | t | c | -0.9245 | 0.2114 | 1.23E-05 | ----?---- | 5.13 | 7 | 0.6441 |
| A Score | chr12 | 89,357,308 | chr12:89357308:CA:C | ca | c | -0.9232 | 0.2113 | 1.25E-05 | ----?---- | 5.205 | 7 | 0.635 |
| A Score | chr12 | 89,371,458 | chr12:89371458:G:A | a | g | 0.9483 | 0.2209 | 1.77E-05 | +++++++++ | 3.242 | 8 | 0.9183 |
| A Score | chr12 | 96,068,260 | chr12:96068260:G:A | a | g | 0.4508 | 0.0985 | 4.72E-06 | +++++++++ | 6.63 | 8 | 0.577 |
| A Score | chr12 | 96,071,652 | chr12:96071652:C:T | t | c | 0.4558 | 0.0986 | 3.82E-06 | +++++++++ | 6.582 | 8 | 0.5824 |
| A Score | chr12 | 96,075,266 | chr12:96075266:C:T | t | c | 0.5539 | 0.1127 | 8.90E-07 | +++++++++ | 9.279 | 8 | 0.3193 |
| A Score | chr12 | 96,079,327 | chr12:96079327:T:TC | t | tc | -0.2932 | 0.0698 | 2.70E-05 | --------- | 8.702 | 8 | 0.368 |
| A Score | chr12 | 96,083,084 | chr12:96083084:G:GAGAAC | g | gagaac | -0.4404 | 0.0994 | 9.47E-06 | --------- | 6.226 | 8 | 0.622 |
| A Score | chr12 | 96,083,440 | chr12:96083440:G:A | a | g | 0.4417 | 0.0992 | 8.49E-06 | +++++++++ | 5.77 | 8 | 0.673 |
| A Score | chr12 | 97,372,162 | chr12:97372162:C:T | t | c | 0.5868 | 0.1505 | 9.66E-05 | +?+++++++ | 4.207 | 7 | 0.7557 |
| A Score | chr12 | 97,373,976 | chr12:97373976:C:A | a | c | 0.6098 | 0.1494 | 4.49E-05 | +?+++++++ | 3.763 | 7 | 0.8067 |
| A Score | chr12 | 97,374,625 | chr12:97374625:C:G | c | g | -0.6103 | 0.1495 | 4.43E-05 | -?------- | 3.769 | 7 | 0.806 |
| A Score | chr12 | 97,378,632 | chr12:97378632:C:A | a | c | 0.6122 | 0.1494 | 4.16E-05 | +?+++++++ | 3.8 | 7 | 0.8026 |
| A Score | chr12 | 97,382,085 | chr12:97382085:A:C | a | c | -0.6131 | 0.1494 | 4.07E-05 | -?------- | 3.814 | 7 | 0.801 |
| A Score | chr12 | 97,384,101 | chr12:97384101:A:G | a | g | -0.6009 | 0.1484 | 5.12E-05 | -?------- | 3.086 | 7 | 0.8769 |
| A Score | chr12 | 97,385,041 | chr12:97385041:G:C | c | g | 0.5986 | 0.1502 | 6.74E-05 | +?+++++++ | 4.4 | 7 | 0.7328 |
| A Score | chr12 | 97,387,023 | chr12:97387023:G:A | a | g | 0.6122 | 0.1495 | 4.23E-05 | +?+++++++ | 3.833 | 7 | 0.7988 |
| A Score | chr12 | 97,391,828 | chr12:97391828:A:G | a | g | -0.5991 | 0.1502 | 6.66E-05 | -?------- | 4.408 | 7 | 0.7317 |
| A Score | chr12 | 97,393,154 | chr12:97393154:C:T | t | c | 0.599 | 0.1502 | 6.68E-05 | +?+++++++ | 4.408 | 7 | 0.7318 |
| A Score | chr12 | 125,515,616 | chr12:125515616:TCA:T | t | tca | 0.4445 | 0.1103 | 5.58E-05 | +++++++++ | 5.408 | 8 | 0.7133 |
| A Score | chr12 | 125,520,208 | chr12:125520208:G:T | t | g | 0.428 | 0.1093 | 9.02E-05 | +++++++++ | 4.863 | 8 | 0.7721 |
| A Score | chr12 | 127,656,440 | chr12:127656440:C:T | t | c | 0.8373 | 0.1919 | 1.28E-05 | -+++-++-+ | 5.107 | 8 | 0.7461 |
| A Score | chr13 | 19,645,556 | chr13:19645556:G:C | c | g | 1.0791 | 0.274 | 8.22E-05 | ?-++?+-+- | 5.835 | 6 | 0.442 |
| A Score | chr13 | 19,654,162 | chr13:19654162:T:C | t | c | -1.0874 | 0.2747 | 7.54E-05 | ?+--?-+-+ | 5.833 | 6 | 0.4422 |
| A Score | chr13 | 19,657,497 | chr13:19657497:G:C | c | g | 1.0787 | 0.2739 | 8.22E-05 | ?-++?+-+- | 5.829 | 6 | 0.4426 |
| A Score | chr13 | 19,757,723 | chr13:19757723:G:A | a | g | 1.1073 | 0.2796 | 7.49E-05 | ?-++?+-+- | 5.722 | 6 | 0.455 |
| A Score | chr13 | 19,897,546 | chr13:19897546:C:T | t | c | 1.1064 | 0.2753 | 5.84E-05 | ?-++?+-+- | 4.633 | 6 | 0.5916 |

| | | | | | | | | | | | | |
|---|---|---|---|---|---|---|---|---|---|---|---|---|
| A Score | chr13 | 21,052,013 | chr13:21052013:G:A | a | g | 0.4799 | 0.1158 | 3.43E-05 | +-+++++-+ | 7.45 | 8 | 0.4889 |
| A Score | chr13 | 21,065,330 | chr13:21065330:A:G | a | g | -0.4707 | 0.115 | 4.26E-05 | -+-----+- | 7.314 | 8 | 0.5032 |
| A Score | chr13 | 35,679,539 | chr13:35679539:G:T | t | g | 1.1531 | 0.2927 | 8.19E-05 | -++???+++ | 1.655 | 5 | 0.8945 |
| A Score | chr13 | 40,631,152 | chr13:40631152:G:A | a | g | 0.2686 | 0.0689 | 9.68E-05 | --+++++++ | 5.656 | 8 | 0.6857 |
| A Score | chr13 | 49,737,526 | chr13:49737526:CTGTTT:C | ctgttt | c | -1.0563 | 0.2661 | 7.21E-05 | ?---??-?? | 6.223 | 3 | 0.1012 |
| A Score | chr13 | 54,523,454 | chr13:54523454:T:C | t | c | -0.7786 | 0.1939 | 5.91E-05 | +?--?---- | 2.011 | 6 | 0.9187 |
| A Score | chr13 | 60,757,552 | chr13:60757552:C:T | t | c | 1.2664 | 0.3205 | 7.79E-05 | ?++???+?? | 0.443 | 2 | 0.8011 |
| A Score | chr13 | 60,759,576 | chr13:60759576:G:A | a | g | 1.2658 | 0.3205 | 7.83E-05 | ?++???+?? | 0.444 | 2 | 0.8008 |
| A Score | chr13 | 60,804,966 | chr13:60804966:T:A | a | t | 1.196 | 0.3003 | 6.80E-05 | ?++???++? | 0.81 | 3 | 0.8471 |
| A Score | chr13 | 60,810,290 | chr13:60810290:G:T | t | g | 1.2498 | 0.3193 | 9.05E-05 | ?++???+?? | 0.399 | 2 | 0.8191 |
| A Score | chr13 | 73,296,506 | chr13:73296506:C:T | t | c | 0.784 | 0.1784 | 1.12E-05 | -+++-+-++ | 10.396 | 8 | 0.2383 |
| A Score | chr13 | 73,337,529 | chr13:73337529:C:T | t | c | 1.0328 | 0.2346 | 1.08E-05 | ?-++?-?++ | 7.924 | 5 | 0.1605 |
| A Score | chr13 | 83,460,017 | chr13:83460017:C:T | t | c | 0.4454 | 0.1073 | 3.30E-05 | +-+++++-+ | 11.851 | 8 | 0.158 |
| A Score | chr13 | 83,462,492 | chr13:83462492:T:C | t | c | -0.4504 | 0.1072 | 2.65E-05 | -+-----+- | 11.64 | 8 | 0.168 |
| A Score | chr13 | 83,462,512 | chr13:83462512:A:T | a | t | -0.4504 | 0.1072 | 2.65E-05 | -+-----+- | 11.641 | 8 | 0.168 |
| A Score | chr13 | 83,465,499 | chr13:83465499:G:A | a | g | 0.4766 | 0.1089 | 1.22E-05 | +-+-+++++ | 12.092 | 8 | 0.1471 |
| A Score | chr13 | 83,466,668 | chr13:83466668:C:T | t | c | 0.4783 | 0.109 | 1.13E-05 | +-+-+++++ | 12.181 | 8 | 0.1433 |
| A Score | chr13 | 83,466,730 | chr13:83466730:A:G | a | g | -0.4523 | 0.1072 | 2.45E-05 | -+-----+- | 11.763 | 8 | 0.1621 |
| A Score | chr13 | 83,467,436 | chr13:83467436:G:A | a | g | 0.4969 | 0.1095 | 5.70E-06 | +-+-+++++ | 12.748 | 8 | 0.1208 |
| A Score | chr13 | 83,471,207 | chr13:83471207:C:T | t | c | 0.355 | 0.0808 | 1.13E-05 | +-+++++-+ | 9.729 | 8 | 0.2846 |
| A Score | chr13 | 83,471,582 | chr13:83471582:C:T | t | c | 0.3557 | 0.0808 | 1.09E-05 | +-+++++-+ | 9.685 | 8 | 0.2878 |
| A Score | chr13 | 83,471,678 | chr13:83471678:A:G | a | g | -0.3551 | 0.0808 | 1.12E-05 | -+-----+- | 9.725 | 8 | 0.2849 |
| A Score | chr13 | 83,471,778 | chr13:83471778:A:C | a | c | -0.3549 | 0.0808 | 1.12E-05 | -+-----+- | 9.704 | 8 | 0.2864 |
| A Score | chr13 | 83,472,862 | chr13:83472862:T:A | a | t | 0.5012 | 0.109 | 4.26E-06 | +-+-+++++ | 13.067 | 8 | 0.1096 |
| A Score | chr13 | 83,473,401 | chr13:83473401:T:C | t | c | -0.3551 | 0.0809 | 1.12E-05 | -+-----+- | 9.729 | 8 | 0.2846 |
| A Score | chr13 | 83,475,199 | chr13:83475199:G:A | a | g | 0.3519 | 0.0808 | 1.35E-05 | +-+++++-+ | 9.694 | 8 | 0.2871 |
| A Score | chr13 | 83,475,913 | chr13:83475913:G:A | a | g | 0.3551 | 0.0808 | 1.12E-05 | +-+++++-+ | 9.747 | 8 | 0.2832 |
| A Score | chr13 | 83,476,759 | chr13:83476759:C:T | t | c | 0.4855 | 0.1084 | 7.59E-06 | +-+-+++++ | 12.28 | 8 | 0.1391 |
| A Score | chr13 | 83,476,778 | chr13:83476778:A:G | a | g | -0.4855 | 0.1084 | 7.59E-06 | -+-+----- | 12.28 | 8 | 0.1391 |
| A Score | chr13 | 83,476,869 | chr13:83476869:G:A | a | g | 0.4836 | 0.1082 | 7.79E-06 | +-+-+++++ | 11.758 | 8 | 0.1623 |
| A Score | chr13 | 83,478,327 | chr13:83478327:G:A | a | g | 0.4437 | 0.1057 | 2.69E-05 | +-+-+++-+ | 11.94 | 8 | 0.1539 |
| A Score | chr13 | 83,483,499 | chr13:83483499:G:T | t | g | 0.3414 | 0.0805 | 2.25E-05 | +-+++++-+ | 8.839 | 8 | 0.3561 |
| A Score | chr13 | 83,485,304 | chr13:83485304:G:A | a | g | 0.4856 | 0.1085 | 7.57E-06 | +-+-+++++ | 12.237 | 8 | 0.1409 |
| A Score | chr13 | 83,485,362 | chr13:83485362:A:G | a | g | -0.4852 | 0.1084 | 7.69E-06 | -+-+----- | 12.268 | 8 | 0.1397 |
| A Score | chr13 | 83,486,754 | chr13:83486754:T:C | t | c | -0.4896 | 0.1081 | 5.96E-06 | -+-+----- | 12.547 | 8 | 0.1284 |
| A Score | chr13 | 83,487,103 | chr13:83487103:A:G | a | g | -0.5035 | 0.1087 | 3.60E-06 | -+-+----- | 13.065 | 8 | 0.1096 |
| A Score | chr13 | 83,487,297 | chr13:83487297:G:A | a | g | 0.3459 | 0.0799 | 1.51E-05 | +-+++++-+ | 9.364 | 8 | 0.3125 |
| A Score | chr13 | 83,488,580 | chr13:83488580:T:C | t | c | -0.4866 | 0.1081 | 6.76E-06 | -+-+----- | 12.222 | 8 | 0.1416 |
| A Score | chr13 | 83,491,071 | chr13:83491071:A:T | a | t | -0.4913 | 0.1081 | 5.49E-06 | -+-+----- | 12.652 | 8 | 0.1244 |
| A Score | chr13 | 83,491,344 | chr13:83491344:A:G | a | g | -0.3448 | 0.08 | 1.62E-05 | -+-----+- | 9.37 | 8 | 0.312 |
| A Score | chr13 | 83,492,117 | chr13:83492117:C:T | t | c | 0.345 | 0.08 | 1.61E-05 | +-+++++-+ | 9.359 | 8 | 0.3129 |
| A Score | chr13 | 83,492,926 | chr13:83492926:T:C | t | c | -0.3448 | 0.08 | 1.62E-05 | -+-----+- | 9.375 | 8 | 0.3116 |
| A Score | chr13 | 83,493,292 | chr13:83493292:T:G | t | g | -0.5016 | 0.1086 | 3.91E-06 | -+-+----- | 12.977 | 8 | 0.1126 |
| A Score | chr13 | 83,493,885 | chr13:83493885:C:T | t | c | 0.4877 | 0.1081 | 6.45E-06 | +-+-+++++ | 12.184 | 8 | 0.1432 |
| A Score | chr13 | 84,410,781 | chr13:84410781:G:C | c | g | 0.8266 | 0.1945 | 2.15E-05 | +?++?++++ | 5.803 | 6 | 0.4456 |
| A Score | chr13 | 90,270,206 | chr13:90270206:C:T | t | c | 1.7274 | 0.4351 | 7.20E-05 | ?+?+???+? | 1.623 | 2 | 0.4442 |

| | | | | | | | | | | | | |
|---|---|---|---|---|---|---|---|---|---|---|---|---|
| A Score | chr13 | 91,641,144 | chr13:91641144:G:A | a | g | 0.2266 | 0.0573 | 7.57E-05 | -++++++-+ | 6.135 | 8 | 0.6322 |
| A Score | chr13 | 102,580,521 | chr13:102580521:G:A | a | g | 0.4495 | 0.1135 | 7.47E-05 | +++++++++ | 5.793 | 8 | 0.6705 |
| A Score | chr13 | 105,497,778 | chr13:105497778:G:T | t | g | 0.5263 | 0.1336 | 8.20E-05 | ++++++-++ | 9.168 | 8 | 0.3283 |
| A Score | chr13 | 105,594,775 | chr13:105594775:C:T | t | c | -0.2412 | 0.0608 | 7.25E-05 | -------+- | 5.473 | 8 | 0.706 |
| A Score | chr14 | 28,376,433 | chr14:28376433:A:G | a | g | -0.2154 | 0.054 | 6.60E-05 | -+--+---- | 6.181 | 8 | 0.6269 |
| A Score | chr14 | 28,393,077 | chr14:28393077:A:G | a | g | -0.2143 | 0.0542 | 7.70E-05 | -+--+---- | 5.935 | 8 | 0.6545 |
| A Score | chr14 | 28,395,472 | chr14:28395472:G:A | a | g | 0.2152 | 0.0542 | 7.22E-05 | +-++-++++ | 5.903 | 8 | 0.6581 |
| A Score | chr14 | 28,417,968 | chr14:28417968:G:A | a | g | 0.2114 | 0.0542 | 9.54E-05 | +-++-++++ | 4.906 | 8 | 0.7676 |
| A Score | chr14 | 30,681,736 | chr14:30681736:T:A | a | t | 0.8865 | 0.2192 | 5.26E-05 | +-+++++-+ | 7.705 | 8 | 0.4628 |
| A Score | chr14 | 31,857,854 | chr14:31857854:C:T | t | c | 0.5661 | 0.1432 | 7.71E-05 | +?++?++?+ | 1.829 | 5 | 0.8723 |
| A Score | chr14 | 31,858,120 | chr14:31858120:C:T | t | c | 0.565 | 0.1432 | 7.99E-05 | +?++?++?+ | 1.817 | 5 | 0.8738 |
| A Score | chr14 | 32,076,110 | chr14:32076110:C:A | a | c | 0.8693 | 0.222 | 8.99E-05 | ++++?+++- | 7.469 | 7 | 0.3818 |
| A Score | chr14 | 32,132,034 | chr14:32132034:G:T | t | g | 0.8805 | 0.2198 | 6.17E-05 | ++++?+++- | 7.714 | 7 | 0.3585 |
| A Score | chr14 | 33,806,830 | chr14:33806830:A:G | a | g | -0.3243 | 0.0824 | 8.37E-05 | ----+---- | 5.586 | 8 | 0.6935 |
| A Score | chr14 | 36,354,877 | chr14:36354877:G:A | a | g | 1.0328 | 0.2528 | 4.40E-05 | +?++?+-++ | 3.818 | 6 | 0.7012 |
| A Score | chr14 | 47,758,444 | chr14:47758444:A:G | a | g | 0.3526 | 0.0901 | 9.01E-05 | ++++-+-++ | 13.318 | 8 | 0.1014 |
| A Score | chr14 | 47,763,626 | chr14:47763626:T:C | t | c | 0.3546 | 0.0898 | 7.81E-05 | ++++-+-++ | 13.627 | 8 | 0.09202 |
| A Score | chr14 | 47,768,006 | chr14:47768006:A:C | a | c | 0.3577 | 0.0899 | 6.86E-05 | ++++-+-++ | 13.308 | 8 | 0.1017 |
| A Score | chr14 | 47,786,248 | chr14:47786248:G:T | t | g | -0.3509 | 0.0896 | 9.01E-05 | ----+-+-- | 13.131 | 8 | 0.1074 |
| A Score | chr14 | 47,789,212 | chr14:47789212:T:C | t | c | 0.3537 | 0.09 | 8.45E-05 | ++++-+-++ | 12.844 | 8 | 0.1173 |
| A Score | chr14 | 47,789,293 | chr14:47789293:G:C | c | g | -0.3563 | 0.0899 | 7.45E-05 | ----+-+-- | 12.815 | 8 | 0.1184 |
| A Score | chr14 | 47,790,731 | chr14:47790731:G:C | c | g | -0.3563 | 0.0899 | 7.42E-05 | ----+-+-- | 12.79 | 8 | 0.1193 |
| A Score | chr14 | 47,791,731 | chr14:47791731:T:C | t | c | 0.3549 | 0.0898 | 7.81E-05 | ++++-+-++ | 12.791 | 8 | 0.1192 |
| A Score | chr14 | 47,791,870 | chr14:47791870:C:T | t | c | -0.3552 | 0.0898 | 7.66E-05 | ----+-+-- | 12.838 | 8 | 0.1176 |
| A Score | chr14 | 47,794,242 | chr14:47794242:T:C | t | c | 0.3556 | 0.0898 | 7.54E-05 | ++++-+-++ | 12.768 | 8 | 0.1201 |
| A Score | chr14 | 47,796,141 | chr14:47796141:C:T | t | c | -0.3531 | 0.0903 | 9.16E-05 | ----+-+-- | 12.625 | 8 | 0.1254 |
| A Score | chr14 | 47,796,637 | chr14:47796637:A:C | a | c | 0.3555 | 0.0898 | 7.56E-05 | ++++-+-++ | 12.79 | 8 | 0.1193 |
| A Score | chr14 | 47,798,549 | chr14:47798549:A:T | a | t | 0.3542 | 0.0899 | 8.10E-05 | ++++-+-++ | 12.727 | 8 | 0.1216 |
| A Score | chr14 | 47,799,006 | chr14:47799006:T:A | a | t | -0.3542 | 0.0897 | 7.85E-05 | ----+-+-- | 12.042 | 8 | 0.1493 |
| A Score | chr14 | 47,799,016 | chr14:47799016:T:C | t | c | 0.3529 | 0.0899 | 8.72E-05 | ++++-+-++ | 12.65 | 8 | 0.1245 |
| A Score | chr14 | 47,799,516 | chr14:47799516:G:A | a | g | -0.3536 | 0.0898 | 8.27E-05 | ----+-+-- | 12.555 | 8 | 0.1281 |
| A Score | chr14 | 47,800,615 | chr14:47800615:T:G | t | g | 0.3542 | 0.0899 | 8.12E-05 | ++++-+-++ | 12.688 | 8 | 0.123 |
| A Score | chr14 | 47,802,806 | chr14:47802806:AG:A | a | ag | -0.3568 | 0.0896 | 6.83E-05 | ----+-+-- | 12.095 | 8 | 0.147 |
| A Score | chr14 | 47,804,107 | chr14:47804107:T:G | t | g | 0.3505 | 0.0895 | 9.03E-05 | ++++-+-++ | 12.74 | 8 | 0.1211 |
| A Score | chr14 | 47,805,190 | chr14:47805190:C:A | a | c | -0.3498 | 0.0895 | 9.28E-05 | ----+-+-- | 12.732 | 8 | 0.1214 |
| A Score | chr14 | 47,806,283 | chr14:47806283:G:C | c | g | -0.3508 | 0.0893 | 8.54E-05 | ----+-+-- | 12.011 | 8 | 0.1507 |
| A Score | chr14 | 47,806,865 | chr14:47806865:C:T | t | c | -0.3491 | 0.0895 | 9.64E-05 | ----+-+-- | 12.688 | 8 | 0.123 |
| A Score | chr14 | 47,807,104 | chr14:47807104:C:T | t | c | -0.3509 | 0.0893 | 8.54E-05 | ----+-+-- | 12.015 | 8 | 0.1506 |
| A Score | chr14 | 47,807,217 | chr14:47807217:T:C | t | c | 0.3509 | 0.0893 | 8.53E-05 | ++++-+-++ | 12.013 | 8 | 0.1506 |
| A Score | chr14 | 47,837,392 | chr14:47837392:G:A | a | g | -0.3896 | 0.0909 | 1.80E-05 | ----+-+-- | 12.675 | 8 | 0.1235 |
| A Score | chr14 | 59,751,824 | chr14:59751824:C:A | a | c | 1.0584 | 0.2621 | 5.40E-05 | ?-++??++? | 0.258 | 4 | 0.9924 |
| A Score | chr14 | 59,769,597 | chr14:59769597:T:C | t | c | -1.069 | 0.2592 | 3.73E-05 | ?+--??--? | 0.208 | 4 | 0.995 |
| A Score | chr14 | 59,819,858 | chr14:59819858:TC:T | t | tc | 0.9589 | 0.2454 | 9.36E-05 | ?+++??++? | 0.563 | 4 | 0.9671 |
| A Score | chr14 | 59,843,625 | chr14:59843625:A:T | a | t | -0.9675 | 0.2396 | 5.39E-05 | ----??--? | 0.514 | 5 | 0.9916 |
| A Score | chr14 | 60,127,461 | chr14:60127461:C:T | t | c | 0.9556 | 0.2409 | 7.26E-05 | ++++??++? | 0.819 | 5 | 0.9758 |
| A Score | chr14 | 60,157,443 | chr14:60157443:C:T | t | c | 1.1015 | 0.2575 | 1.89E-05 | +-++??++? | 0.833 | 5 | 0.9749 |

| | | | | | | | | | | | | |
|---|---|---|---|---|---|---|---|---|---|---|---|---|
| A Score | chr14 | 60,194,016 | chr14:60194016:T:C | t | c | -1.1158 | 0.2578 | 1.51E-05 | -+--??--? | 0.865 | 5 | 0.9727 |
| A Score | chr14 | 65,205,194 | chr14:65205194:A:G | a | g | 0.2155 | 0.0549 | 8.69E-05 | +-+++++++ | 6.733 | 8 | 0.5657 |
| A Score | chr14 | 65,209,416 | chr14:65209416:T:G | t | g | 0.2211 | 0.0551 | 6.10E-05 | +-+++++++ | 6.817 | 8 | 0.5565 |
| A Score | chr14 | 65,209,577 | chr14:65209577:C:T | t | c | -0.2171 | 0.0556 | 9.50E-05 | -+------- | 8.705 | 8 | 0.3677 |
| A Score | chr14 | 69,614,312 | chr14:69614312:C:T | t | c | -0.223 | 0.0545 | 4.32E-05 | +-------- | 10.045 | 8 | 0.2619 |
| A Score | chr14 | 69,637,403 | chr14:69637403:C:T | t | c | -0.2197 | 0.0543 | 5.27E-05 | +-------- | 10.265 | 8 | 0.2469 |
| A Score | chr14 | 69,656,659 | chr14:69656659:CCAGGCACTTTG | ccaggcactt | c | 0.2207 | 0.0544 | 5.01E-05 | -++++++++ | 10.456 | 8 | 0.2345 |
| A Score | chr14 | 69,656,933 | chr14:69656933:G:T | t | g | -0.2207 | 0.0544 | 4.99E-05 | +-------- | 10.463 | 8 | 0.234 |
| A Score | chr14 | 69,680,276 | chr14:69680276:C:T | t | c | -0.2222 | 0.0545 | 4.57E-05 | +---+---- | 10.762 | 8 | 0.2155 |
| A Score | chr14 | 69,684,678 | chr14:69684678:C:T | t | c | -0.2197 | 0.0544 | 5.41E-05 | +-------- | 9.995 | 8 | 0.2654 |
| A Score | chr14 | 69,691,981 | chr14:69691981:G:T | t | g | -0.22 | 0.0545 | 5.43E-05 | +---+---- | 9.954 | 8 | 0.2683 |
| A Score | chr14 | 69,708,200 | chr14:69708200:T:C | t | c | 0.2218 | 0.0547 | 5.06E-05 | -++++++++ | 9.797 | 8 | 0.2796 |
| A Score | chr14 | 79,550,248 | chr14:79550248:C:A | a | c | 0.3155 | 0.0785 | 5.90E-05 | --+++++-+ | 6.52 | 8 | 0.5892 |
| A Score | chr14 | 79,571,815 | chr14:79571815:T:C | t | c | -0.3069 | 0.078 | 8.38E-05 | -+-----+- | 7.631 | 8 | 0.4704 |
| A Score | chr14 | 94,406,382 | chr14:94406382:T:G | t | g | -0.5774 | 0.1426 | 5.12E-05 | ------+-- | 6.029 | 8 | 0.644 |
| A Score | chr14 | 94,411,215 | chr14:94411215:G:C | c | g | 0.6222 | 0.1518 | 4.13E-05 | ++++++-++ | 8.331 | 8 | 0.4018 |
| A Score | chr14 | 94,412,711 | chr14:94412711:T:C | t | c | -0.6133 | 0.1507 | 4.70E-05 | ------+-- | 6.635 | 8 | 0.5765 |
| A Score | chr14 | 97,148,901 | chr14:97148901:T:C | t | c | -0.8717 | 0.2198 | 7.31E-05 | +---?-+-- | 10.958 | 7 | 0.1404 |
| A Score | chr14 | 98,586,716 | chr14:98586716:T:C | t | c | -0.2255 | 0.0559 | 5.39E-05 | -------+- | 2.931 | 8 | 0.9386 |
| A Score | chr14 | 98,587,627 | chr14:98587627:A:G | a | g | -0.2286 | 0.056 | 4.51E-05 | -------+- | 3.131 | 8 | 0.9259 |
| A Score | chr14 | 103,844,345 | chr14:103844345:G:T | t | g | 0.6748 | 0.1541 | 1.19E-05 | +-+++++-+ | 13.075 | 8 | 0.1093 |
| A Score | chr15 | 33,341,839 | chr15:33341839:G:A | a | g | 0.6196 | 0.1568 | 7.80E-05 | +++++++-+ | 4.307 | 8 | 0.8284 |
| A Score | chr15 | 44,787,809 | chr15:44787809:G:C | c | g | 1.6707 | 0.4161 | 5.94E-05 | ?+????+++ | 3.645 | 3 | 0.3024 |
| A Score | chr15 | 44,798,670 | chr15:44798670:G:A | a | g | 1.6908 | 0.4183 | 5.29E-05 | ?+????+++ | 3.562 | 3 | 0.3128 |
| A Score | chr15 | 69,871,287 | chr15:69871287:G:A | a | g | 0.227 | 0.0567 | 6.17E-05 | +++++++-+ | 5.841 | 8 | 0.6651 |
| A Score | chr15 | 69,873,075 | chr15:69873075:G:A | a | g | 0.2251 | 0.0561 | 5.95E-05 | +++++++-+ | 6.104 | 8 | 0.6355 |
| A Score | chr15 | 69,885,639 | chr15:69885639:TC:T | t | tc | 0.2412 | 0.0601 | 5.99E-05 | +-+++++-- | 7.214 | 8 | 0.5137 |
| A Score | chr15 | 69,886,596 | chr15:69886596:C:A | a | c | 0.2349 | 0.0601 | 9.33E-05 | +-++++++- | 7.606 | 8 | 0.4729 |
| A Score | chr15 | 69,896,334 | chr15:69896334:T:C | t | c | -0.2612 | 0.0639 | 4.35E-05 | -+-----++ | 6.816 | 8 | 0.5567 |
| A Score | chr15 | 88,947,924 | chr15:88947924:G:A | a | g | 1.0444 | 0.2642 | 7.70E-05 | +++++?++? | 4.404 | 6 | 0.6222 |
| A Score | chr15 | 92,134,287 | chr15:92134287:A:G | a | g | 0.2351 | 0.0577 | 4.57E-05 | -++++++++ | 2.872 | 8 | 0.9421 |
| A Score | chr15 | 92,143,115 | chr15:92143115:A:C | a | c | 0.2177 | 0.0557 | 9.31E-05 | +++++++++ | 6.566 | 8 | 0.5841 |
| A Score | chr15 | 92,144,165 | chr15:92144165:C:G | c | g | 0.2518 | 0.0583 | 1.58E-05 | +++++++++ | 3.572 | 8 | 0.8935 |
| A Score | chr15 | 92,144,326 | chr15:92144326:T:C | t | c | 0.2516 | 0.0583 | 1.58E-05 | +++++++++ | 3.543 | 8 | 0.8958 |
| A Score | chr15 | 92,144,553 | chr15:92144553:G:A | a | g | -0.2491 | 0.0584 | 2.01E-05 | --------- | 3.322 | 8 | 0.9125 |
| A Score | chr15 | 92,144,897 | chr15:92144897:A:T | a | t | 0.2419 | 0.0582 | 3.27E-05 | +++++++++ | 3.493 | 8 | 0.8997 |
| A Score | chr15 | 92,145,423 | chr15:92145423:T:TTC | t | ttc | 0.247 | 0.0582 | 2.21E-05 | +++++++++ | 3.379 | 8 | 0.9084 |
| A Score | chr15 | 92,145,728 | chr15:92145728:A:G | a | g | 0.244 | 0.0581 | 2.72E-05 | +++++++++ | 3.319 | 8 | 0.9127 |
| A Score | chr15 | 92,145,751 | chr15:92145751:C:T | t | c | -0.2397 | 0.0576 | 3.11E-05 | --------- | 3.521 | 8 | 0.8975 |
| A Score | chr15 | 92,145,962 | chr15:92145962:C:CA | ca | c | -0.2407 | 0.0582 | 3.50E-05 | --------- | 3.163 | 8 | 0.9237 |
| A Score | chr15 | 92,145,999 | chr15:92145999:A:G | a | g | 0.245 | 0.0581 | 2.49E-05 | +++++++++ | 3.355 | 8 | 0.9102 |
| A Score | chr15 | 92,146,465 | chr15:92146465:C:G | c | g | 0.2492 | 0.0583 | 1.91E-05 | +++++++++ | 2.87 | 8 | 0.9422 |
| A Score | chr15 | 92,147,318 | chr15:92147318:A:G | a | g | 0.2498 | 0.0581 | 1.71E-05 | +++++++++ | 3.252 | 8 | 0.9176 |
| A Score | chr15 | 92,147,786 | chr15:92147786:T:C | t | c | 0.2175 | 0.0556 | 9.19E-05 | +++++++++ | 6.423 | 8 | 0.6 |
| A Score | chr15 | 92,147,988 | chr15:92147988:T:G | t | g | 0.2262 | 0.0554 | 4.45E-05 | +++++++++ | 5.529 | 8 | 0.6999 |
| A Score | chr15 | 92,150,196 | chr15:92150196:T:G | t | g | 0.2336 | 0.057 | 4.24E-05 | -+++-++++ | 2.595 | 8 | 0.9572 |

| | | | | | | | | | | | | |
|---|---|---|---|---|---|---|---|---|---|---|---|---|
| A Score | chr15 | 93,265,804 | chr15:93265804:A:G | a | g | -0.3515 | 0.0853 | 3.80E-05 | ----+--+- | 7.861 | 8 | 0.4472 |
| A Score | chr15 | 93,276,039 | chr15:93276039:C:T | t | c | 0.3403 | 0.0855 | 6.90E-05 | ++++-++-+ | 6.852 | 8 | 0.5527 |
| A Score | chr16 | 26,461,059 | chr16:26461059:G:C | c | g | 1.6023 | 0.4118 | 9.97E-05 | ?+????+?+ | 3.945 | 2 | 0.1391 |
| A Score | chr16 | 27,847,613 | chr16:27847613:T:C | t | c | -0.8436 | 0.2042 | 3.62E-05 | +?--?---- | 5.329 | 6 | 0.5023 |
| A Score | chr16 | 29,171,909 | chr16:29171909:A:G | a | g | -0.2079 | 0.053 | 8.81E-05 | -+----+-- | 7.944 | 8 | 0.439 |
| A Score | chr16 | 29,174,870 | chr16:29174870:T:C | t | c | -0.2064 | 0.053 | 9.86E-05 | -+----+-- | 8.298 | 8 | 0.4049 |
| A Score | chr16 | 29,175,043 | chr16:29175043:A:G | a | g | -0.2079 | 0.053 | 8.77E-05 | -+----+-- | 7.93 | 8 | 0.4404 |
| A Score | chr16 | 29,177,973 | chr16:29177973:A:G | a | g | -0.2078 | 0.053 | 8.91E-05 | -+----+-- | 8.116 | 8 | 0.4222 |
| A Score | chr16 | 29,203,953 | chr16:29203953:A:G | a | g | -0.2065 | 0.0528 | 9.32E-05 | -+----+-- | 10.563 | 8 | 0.2277 |
| A Score | chr16 | 29,206,284 | chr16:29206284:A:G | a | g | -0.2078 | 0.0528 | 8.42E-05 | -+----+-- | 10.717 | 8 | 0.2182 |
| A Score | chr16 | 29,209,997 | chr16:29209997:T:G | t | g | -0.2089 | 0.0521 | 5.99E-05 | -+----+-- | 11.198 | 8 | 0.1907 |
| A Score | chr16 | 29,211,121 | chr16:29211121:T:C | t | c | -0.2058 | 0.0528 | 9.61E-05 | -+----+-- | 13.03 | 8 | 0.1108 |
| A Score | chr16 | 29,213,143 | chr16:29213143:A:G | a | g | -0.2071 | 0.0528 | 8.72E-05 | -+----+-- | 13.134 | 8 | 0.1073 |
| A Score | chr16 | 29,214,110 | chr16:29214110:G:A | a | g | 0.2123 | 0.0524 | 5.09E-05 | +-++++-++ | 11.424 | 8 | 0.1788 |
| A Score | chr16 | 29,214,168 | chr16:29214168:T:C | t | c | -0.2097 | 0.0524 | 6.24E-05 | -+----+-- | 11.398 | 8 | 0.1801 |
| A Score | chr16 | 29,220,052 | chr16:29220052:G:A | a | g | 0.2203 | 0.0525 | 2.72E-05 | +-++++-++ | 9.875 | 8 | 0.2739 |
| A Score | chr16 | 29,220,386 | chr16:29220386:A:G | a | g | -0.2209 | 0.0525 | 2.56E-05 | -+----+-- | 9.917 | 8 | 0.2709 |
| A Score | chr16 | 29,220,649 | chr16:29220649:C:A | a | c | 0.2204 | 0.0527 | 2.86E-05 | +-++++-++ | 10.391 | 8 | 0.2387 |
| A Score | chr16 | 29,220,650 | chr16:29220650:T:G | t | g | -0.2228 | 0.0526 | 2.27E-05 | -+----+-- | 9.947 | 8 | 0.2687 |
| A Score | chr16 | 29,221,356 | chr16:29221356:A:G | a | g | -0.2089 | 0.0529 | 7.98E-05 | -+----+-- | 9.995 | 8 | 0.2654 |
| A Score | chr16 | 29,221,646 | chr16:29221646:G:A | a | g | 0.2102 | 0.0531 | 7.52E-05 | +-++++-++ | 11.67 | 8 | 0.1666 |
| A Score | chr16 | 29,223,261 | chr16:29223261:G:C | c | g | 0.2144 | 0.0533 | 5.67E-05 | +-++++-++ | 11.717 | 8 | 0.1643 |
| A Score | chr16 | 29,223,807 | chr16:29223807:A:G | a | g | -0.2139 | 0.0533 | 5.93E-05 | -+----+-- | 11.762 | 8 | 0.1621 |
| A Score | chr16 | 29,224,212 | chr16:29224212:G:C | c | g | 0.2166 | 0.0532 | 4.71E-05 | +-++++-++ | 11.022 | 8 | 0.2005 |
| A Score | chr16 | 49,847,794 | chr16:49847794:G:A | a | g | 0.816 | 0.1914 | 2.01E-05 | ++++?++++ | 8.128 | 7 | 0.3215 |
| A Score | chr16 | 51,679,111 | chr16:51679111:C:A | a | c | -0.2145 | 0.0539 | 6.99E-05 | --------- | 3.155 | 8 | 0.9242 |
| A Score | chr16 | 51,684,425 | chr16:51684425:C:T | t | c | -0.214 | 0.054 | 7.51E-05 | --------- | 3.22 | 8 | 0.9198 |
| A Score | chr16 | 51,706,684 | chr16:51706684:C:T | t | c | -0.2108 | 0.054 | 9.51E-05 | --------- | 3.164 | 8 | 0.9236 |
| A Score | chr16 | 74,164,722 | chr16:74164722:A:C | a | c | -0.7747 | 0.1931 | 6.01E-05 | ----+-+?- | 7.992 | 7 | 0.3333 |
| A Score | chr16 | 75,517,054 | chr16:75517054:G:A | a | g | 1.2834 | 0.2449 | 1.59E-07 | --++??-?+ | 12.48 | 5 | 0.02878 |
| A Score | chr16 | 75,683,943 | chr16:75683943:C:T | t | c | 1.1146 | 0.2198 | 3.96E-07 | -?+++--++ | 15.995 | 7 | 0.02516 |
| A Score | chr16 | 75,686,202 | chr16:75686202:C:G | c | g | -0.7507 | 0.1662 | 6.31E-06 | ++--+++-- | 11.851 | 8 | 0.158 |
| A Score | chr16 | 75,717,066 | chr16:75717066:T:C | t | c | -0.6025 | 0.1515 | 6.98E-05 | ++--+-+-- | 9.668 | 8 | 0.2891 |
| A Score | chr16 | 75,837,636 | chr16:75837636:C:A | a | c | 1.0773 | 0.2301 | 2.85E-06 | -?++?--?+ | 10.897 | 5 | 0.05347 |
| A Score | chr16 | 75,932,311 | chr16:75932311:G:A | a | g | 0.7462 | 0.1863 | 6.19E-05 | +-+++++?+ | 5.153 | 7 | 0.6413 |
| A Score | chr16 | 84,541,196 | chr16:84541196:T:C | t | c | 0.3632 | 0.0927 | 8.86E-05 | -+++-++-+ | 14.006 | 8 | 0.0816 |
| A Score | chr16 | 84,541,672 | chr16:84541672:T:C | t | c | 0.3627 | 0.0927 | 9.16E-05 | -+++-++-+ | 14.012 | 8 | 0.08146 |
| A Score | chr16 | 84,541,826 | chr16:84541826:A:G | a | g | 0.3624 | 0.0927 | 9.24E-05 | -+++-++-+ | 13.998 | 8 | 0.08181 |
| A Score | chr16 | 84,541,935 | chr16:84541935:T:C | t | c | 0.3627 | 0.0931 | 9.78E-05 | -+++-++-+ | 13.988 | 8 | 0.08207 |
| A Score | chr16 | 84,542,217 | chr16:84542217:A:C | a | c | 0.3627 | 0.0931 | 9.75E-05 | -+++-++-+ | 13.99 | 8 | 0.08202 |
| A Score | chr16 | 84,545,755 | chr16:84545755:T:C | t | c | -0.3833 | 0.0969 | 7.66E-05 | +---+--+- | 14.297 | 8 | 0.07434 |
| A Score | chr16 | 84,545,866 | chr16:84545866:C:G | c | g | -0.3829 | 0.0973 | 8.31E-05 | +---+--+- | 14.009 | 8 | 0.08152 |
| A Score | chr16 | 84,546,565 | chr16:84546565:G:A | a | g | 0.3765 | 0.0955 | 8.09E-05 | --++-++-+ | 14.714 | 8 | 0.06496 |
| A Score | chr16 | 86,649,116 | chr16:86649116:C:A | a | c | -0.2107 | 0.054 | 9.47E-05 | --+-+---- | 17.013 | 8 | 0.02997 |
| A Score | chr16 | 86,650,436 | chr16:86650436:G:A | a | g | -0.2186 | 0.0538 | 4.79E-05 | --+-+---- | 15.66 | 8 | 0.04751 |
| A Score | chr16 | 86,650,496 | chr16:86650496:A:C | a | c | 0.2178 | 0.0538 | 5.09E-05 | ++-+-++++ | 15.802 | 8 | 0.0453 |

| | | | | | | | | | | | | |
|---|---|---|---|---|---|---|---|---|---|---|---|---|
| A Score | chr16 | 86,650,526 | chr16:86650526:G:A | a | g | -0.2163 | 0.0537 | 5.65E-05 | --+-+---- | 16.091 | 8 | 0.0411 |
| A Score | chr16 | 86,650,570 | chr16:86650570:A:G | a | g | 0.2208 | 0.0538 | 4.04E-05 | ++-+-++++ | 16.133 | 8 | 0.04051 |
| A Score | chr16 | 86,650,928 | chr16:86650928:A:G | a | g | 0.2204 | 0.0538 | 4.15E-05 | ++-+-++++ | 15.799 | 8 | 0.04534 |
| A Score | chr16 | 86,651,272 | chr16:86651272:T:C | t | c | 0.2183 | 0.0538 | 4.92E-05 | ++-+-++++ | 16.17 | 8 | 0.04001 |
| A Score | chr16 | 86,651,541 | chr16:86651541:G:A | a | g | -0.2171 | 0.0538 | 5.44E-05 | --+-+---- | 16.089 | 8 | 0.04112 |
| A Score | chr16 | 86,855,643 | chr16:86855643:C:T | t | c | -0.2242 | 0.0546 | 4.04E-05 | ----+---- | 6.854 | 8 | 0.5525 |
| A Score | chr16 | 86,858,411 | chr16:86858411:G:C | c | g | 0.2279 | 0.0552 | 3.68E-05 | ++++-++++ | 7.897 | 8 | 0.4436 |
| A Score | chr17 | 8,814,614 | chr17:8814614:C:T | t | c | 1.0368 | 0.2449 | 2.30E-05 | ?-++++-+? | 5.078 | 6 | 0.5339 |
| A Score | chr17 | 39,698,623 | chr17:39698623:T:TG | t | tg | -0.2312 | 0.0546 | 2.31E-05 | +-----+-- | 17.118 | 8 | 0.0289 |
| A Score | chr17 | 53,286,835 | chr17:53286835:G:A | a | g | 0.7696 | 0.1968 | 9.21E-05 | ??+++++++ | 2.774 | 6 | 0.8367 |
| A Score | chr17 | 53,287,727 | chr17:53287727:C:T | t | c | 0.7772 | 0.1885 | 3.74E-05 | ??+++++++ | 2.821 | 6 | 0.831 |
| A Score | chr17 | 66,918,973 | chr17:66918973:G:A | a | g | 0.5525 | 0.1194 | 3.70E-06 | -+++?+-++ | 16.015 | 7 | 0.02498 |
| A Score | chr17 | 66,929,858 | chr17:66929858:T:C | t | c | -0.5461 | 0.1193 | 4.74E-06 | +---?-+-- | 15.9 | 7 | 0.02605 |
| A Score | chr17 | 72,291,383 | chr17:72291383:G:C | c | g | -0.2485 | 0.0583 | 2.04E-05 | ----+---- | 6.443 | 8 | 0.5978 |
| A Score | chr17 | 72,293,175 | chr17:72293175:A:G | a | g | 0.2278 | 0.0578 | 8.19E-05 | ++++-++++ | 4.985 | 8 | 0.7592 |
| A Score | chr17 | 72,294,754 | chr17:72294754:AT:A | a | at | -0.2282 | 0.0579 | 8.10E-05 | ----+---- | 4.888 | 8 | 0.7694 |
| A Score | chr17 | 72,295,015 | chr17:72295015:C:A | a | c | -0.2302 | 0.0578 | 6.90E-05 | ----+---- | 5.044 | 8 | 0.7528 |
| A Score | chr17 | 72,297,185 | chr17:72297185:G:A | a | g | -0.2303 | 0.0578 | 6.80E-05 | ----+---- | 5.385 | 8 | 0.7158 |
| A Score | chr17 | 72,297,729 | chr17:72297729:G:A | a | g | -0.2314 | 0.0578 | 6.32E-05 | ----+---- | 5.256 | 8 | 0.7299 |
| A Score | chr17 | 72,297,812 | chr17:72297812:G:A | a | g | -0.2341 | 0.0578 | 5.17E-05 | ----+---- | 5.227 | 8 | 0.7331 |
| A Score | chr17 | 72,299,461 | chr17:72299461:A:G | a | g | 0.231 | 0.0578 | 6.34E-05 | ++++-++++ | 5.384 | 8 | 0.7159 |
| A Score | chr17 | 72,300,105 | chr17:72300105:A:G | a | g | 0.248 | 0.0588 | 2.49E-05 | ++++-++++ | 7.484 | 8 | 0.4855 |
| A Score | chr17 | 72,574,209 | chr17:72574209:G:A | a | g | 1.6389 | 0.3723 | 1.07E-05 | ?+????+?+ | 2.07 | 2 | 0.3551 |
| A Score | chr17 | 74,058,585 | chr17:74058585:C:T | t | c | 0.2833 | 0.0678 | 2.91E-05 | -++++++++ | 4.97 | 8 | 0.7608 |
| A Score | chr18 | 5,115,215 | chr18:5115215:GTTT:G | g | gttt | 0.2874 | 0.0732 | 8.72E-05 | --++-++++ | 7.1 | 8 | 0.5259 |
| A Score | chr18 | 5,121,672 | chr18:5121672:C:T | t | c | 0.2858 | 0.0731 | 9.25E-05 | --+++++++ | 7.156 | 8 | 0.5199 |
| A Score | chr18 | 14,132,511 | chr18:14132511:C:T | t | c | 1.0575 | 0.2583 | 4.25E-05 | +-++?++-? | 3.33 | 6 | 0.7665 |
| A Score | chr18 | 40,359,422 | chr18:40359422:T:G | t | g | 0.3127 | 0.0787 | 7.09E-05 | +++++++-+ | 7.61 | 8 | 0.4725 |
| A Score | chr18 | 45,565,808 | chr18:45565808:T:C | t | c | 0.443 | 0.1087 | 4.61E-05 | --++-+-++ | 8.082 | 8 | 0.4255 |
| A Score | chr18 | 45,971,556 | chr18:45971556:G:A | a | g | -1.2098 | 0.3024 | 6.31E-05 | ++--?---- | 6.029 | 7 | 0.5363 |
| A Score | chr18 | 50,906,489 | chr18:50906489:T:G | t | g | -0.3147 | 0.0769 | 4.30E-05 | -------+- | 6.039 | 8 | 0.6428 |
| A Score | chr18 | 50,923,320 | chr18:50923320:CAA:C | caa | c | -0.3249 | 0.0774 | 2.72E-05 | -------+- | 6.435 | 8 | 0.5986 |
| A Score | chr18 | 61,308,114 | chr18:61308114:T:C | t | c | -0.7058 | 0.1797 | 8.61E-05 | --+----+- | 12.47 | 8 | 0.1315 |
| A Score | chr18 | 62,346,885 | chr18:62346885:G:A | a | g | 0.2317 | 0.0594 | 9.66E-05 | ++++-++-+ | 4.037 | 8 | 0.8537 |
| A Score | chr18 | 62,355,440 | chr18:62355440:G:A | a | g | 0.2348 | 0.06 | 9.08E-05 | ++++-++-+ | 5.106 | 8 | 0.7462 |
| A Score | chr18 | 62,355,724 | chr18:62355724:ACTT:A | a | actt | 0.2351 | 0.06 | 8.90E-05 | ++++-++-+ | 5.111 | 8 | 0.7457 |
| A Score | chr18 | 62,359,460 | chr18:62359460:C:G | c | g | 0.2139 | 0.0536 | 6.67E-05 | +++++++++ | 3.627 | 8 | 0.8891 |
| A Score | chr18 | 62,359,485 | chr18:62359485:T:A | a | t | 0.2566 | 0.0644 | 6.70E-05 | +++++++-+ | 9.909 | 8 | 0.2715 |
| A Score | chr18 | 62,359,589 | chr18:62359589:G:A | a | g | 0.2564 | 0.0644 | 6.79E-05 | +++++++-+ | 9.906 | 8 | 0.2717 |
| A Score | chr18 | 73,739,641 | chr18:73739641:G:A | a | g | 0.5895 | 0.1448 | 4.66E-05 | +-+++++++ | 9.604 | 8 | 0.2939 |
| A Score | chr18 | 78,776,972 | chr18:78776972:C:T | t | c | 0.374 | 0.0871 | 1.74E-05 | ++++-++++ | 4.047 | 8 | 0.8529 |
| A Score | chr19 | 348,404 | chr19:348404:GA:G | g | ga | -0.2309 | 0.0588 | 8.66E-05 | ------+-- | 3.032 | 8 | 0.9323 |
| A Score | chr19 | 352,700 | chr19:352700:T:C | t | c | 0.2355 | 0.0589 | 6.42E-05 | ++++++-++ | 3.254 | 8 | 0.9175 |
| A Score | chr19 | 1,803,583 | chr19:1803583:G:T | t | g | -0.3047 | 0.0734 | 3.32E-05 | --------- | 7.572 | 8 | 0.4763 |
| A Score | chr19 | 4,665,213 | chr19:4665213:CTGTT:C | ctgtt | c | -0.3385 | 0.079 | 1.84E-05 | ----+---+ | 10.933 | 8 | 0.2055 |
| A Score | chr19 | 8,923,770 | chr19:8923770:T:C | t | c | 0.3109 | 0.0784 | 7.27E-05 | --+++++++ | 12.279 | 8 | 0.1392 |

| | | | | | | | | | | | | |
|---|---|---|---|---|---|---|---|---|---|---|---|---|
| A Score | chr19 | 8,924,496 | chr19:8924496:C:T | t | c | -0.3115 | 0.0781 | 6.60E-05 | ++------- | 12.202 | 8 | 0.1424 |
| A Score | chr19 | 8,957,698 | chr19:8957698:A:G | a | g | -0.653 | 0.1463 | 8.10E-06 | ++------- | 7.184 | 8 | 0.5169 |
| A Score | chr19 | 8,960,886 | chr19:8960886:C:T | t | c | 0.6767 | 0.1463 | 3.76E-06 | --+++++++ | 7.527 | 8 | 0.481 |
| A Score | chr19 | 14,799,628 | chr19:14799628:G:A | a | g | 0.4549 | 0.1155 | 8.17E-05 | ++++-++++ | 6.955 | 8 | 0.5415 |
| A Score | chr19 | 44,746,404 | chr19:44746404:C:T | t | c | -0.2469 | 0.0621 | 6.97E-05 | --------- | 4.65 | 8 | 0.7943 |
| A Score | chr19 | 44,816,374 | chr19:44816374:G:A | a | g | 0.6065 | 0.1503 | 5.48E-05 | +-++-++-+ | 13.428 | 8 | 0.09795 |
| A Score | chr19 | 44,820,881 | chr19:44820881:G:A | a | g | -0.2567 | 0.0572 | 7.14E-06 | +---+-+-- | 14.149 | 8 | 0.07796 |
| A Score | chr19 | 44,834,661 | chr19:44834661:G:A | a | g | -0.6782 | 0.1479 | 4.51E-06 | --------- | 1.681 | 8 | 0.9893 |
| A Score | chr19 | 44,842,530 | chr19:44842530:A:T | a | t | -0.4875 | 0.1069 | 5.07E-06 | -+--+++-- | 23.926 | 8 | 0.002358 |
| A Score | chr19 | 44,848,259 | chr19:44848259:C:G | c | g | 0.5474 | 0.1334 | 4.08E-05 | -++++++++ | 4.051 | 8 | 0.8525 |
| A Score | chr19 | 44,852,464 | chr19:44852464:C:G | c | g | 0.2457 | 0.0602 | 4.47E-05 | +++++++++ | 2.539 | 8 | 0.9599 |
| A Score | chr19 | 44,853,207 | chr19:44853207:T:A | a | t | -0.2456 | 0.0605 | 4.86E-05 | --------- | 2.109 | 8 | 0.9775 |
| A Score | chr19 | 44,853,746 | chr19:44853746:C:G | c | g | 0.2392 | 0.0596 | 5.93E-05 | ++++++-++ | 3.128 | 8 | 0.9261 |
| A Score | chr19 | 44,854,034 | chr19:44854034:G:C | c | g | -0.2499 | 0.0603 | 3.42E-05 | --------- | 2.315 | 8 | 0.9698 |
| A Score | chr19 | 44,854,120 | chr19:44854120:T:C | t | c | 0.2498 | 0.0605 | 3.69E-05 | +++++++++ | 1.729 | 8 | 0.9882 |
| A Score | chr19 | 44,856,329 | chr19:44856329:C:T | t | c | -0.2464 | 0.0604 | 4.56E-05 | --------- | 1.916 | 8 | 0.9835 |
| A Score | chr19 | 44,856,449 | chr19:44856449:G:A | a | g | -0.2496 | 0.0605 | 3.66E-05 | --------- | 2.105 | 8 | 0.9776 |
| A Score | chr19 | 44,865,946 | chr19:44865946:G:A | a | g | 0.6853 | 0.1481 | 3.71E-06 | ++++-+-++ | 14.282 | 8 | 0.0747 |
| A Score | chr19 | 44,878,777 | chr19:44878777:A:G | a | g | -0.3097 | 0.0532 | 5.87E-09 | ----+---- | 7.599 | 8 | 0.4736 |
| A Score | chr19 | 44,879,418 | chr19:44879418:G:A | a | g | 0.5922 | 0.1389 | 2.00E-05 | ++++-++-+ | 4.78 | 8 | 0.7808 |
| A Score | chr19 | 44,881,148 | chr19:44881148:T:C | t | c | -0.3128 | 0.078 | 6.01E-05 | ----+-++- | 6.482 | 8 | 0.5934 |
| A Score | chr19 | 44,882,502 | chr19:44882502:G:C | c | g | 0.2164 | 0.054 | 6.16E-05 | --+++++++ | 10.842 | 8 | 0.2108 |
| A Score | chr19 | 44,883,210 | chr19:44883210:G:GTAA | g | gtaa | 0.8949 | 0.0782 | 2.72E-30 | +++++++++ | 8.199 | 8 | 0.4143 |
| A Score | chr19 | 44,883,377 | chr19:44883377:C:T | t | c | -0.7136 | 0.1448 | 8.34E-07 | ?-------- | 3.75 | 7 | 0.8081 |
| A Score | chr19 | 44,884,202 | chr19:44884202:C:G | c | g | 0.8932 | 0.078 | 2.36E-30 | +++++++++ | 8.282 | 8 | 0.4064 |
| A Score | chr19 | 44,884,339 | chr19:44884339:G:A | a | g | -0.8919 | 0.0778 | 2.13E-30 | --------- | 8.035 | 8 | 0.43 |
| A Score | chr19 | 44,884,873 | chr19:44884873:G:A | a | g | -0.8629 | 0.0768 | 2.89E-29 | --------- | 8.559 | 8 | 0.3809 |
| A Score | chr19 | 44,885,243 | chr19:44885243:A:G | a | g | 0.7216 | 0.0642 | 2.39E-29 | +++++++++ | 3.652 | 8 | 0.8871 |
| A Score | chr19 | 44,887,076 | chr19:44887076:A:G | a | g | 0.7347 | 0.064 | 1.55E-30 | +++++++++ | 3.092 | 8 | 0.9285 |
| A Score | chr19 | 44,888,997 | chr19:44888997:C:T | t | c | -0.9813 | 0.0725 | 8.99E-42 | --------- | 5.429 | 8 | 0.7108 |
| A Score | chr19 | 44,889,415 | chr19:44889415:AT:A | a | at | 0.6743 | 0.1506 | 7.53E-06 | ++++-+-++ | 14.918 | 8 | 0.06076 |
| A Score | chr19 | 44,891,079 | chr19:44891079:T:C | t | c | 0.8918 | 0.0773 | 8.47E-31 | +++++++++ | 8.693 | 8 | 0.3689 |
| A Score | chr19 | 44,891,712 | chr19:44891712:T:G | t | g | 0.7411 | 0.0642 | 7.18E-31 | +++++++++ | 3.1 | 8 | 0.9279 |
| A Score | chr19 | 44,892,009 | chr19:44892009:G:A | a | g | -0.3538 | 0.0534 | 3.37E-11 | -------+- | 15.347 | 8 | 0.05274 |
| A Score | chr19 | 44,892,073 | chr19:44892073:A:G | a | g | -0.2318 | 0.0544 | 2.00E-05 | +++------ | 14.291 | 8 | 0.07449 |
| A Score | chr19 | 44,892,362 | chr19:44892362:A:G | a | g | 0.9161 | 0.0777 | 4.37E-32 | +++++++++ | 7.846 | 8 | 0.4486 |
| A Score | chr19 | 44,892,457 | chr19:44892457:T:C | t | c | 0.7452 | 0.0641 | 2.82E-31 | +++++++++ | 2.981 | 8 | 0.9355 |
| A Score | chr19 | 44,892,587 | chr19:44892587:G:A | a | g | -0.7946 | 0.0851 | 1.03E-20 | --------- | 5.156 | 8 | 0.7408 |
| A Score | chr19 | 44,892,652 | chr19:44892652:C:G | c | g | 0.9213 | 0.078 | 3.11E-32 | +++++++++ | 8.408 | 8 | 0.3947 |
| A Score | chr19 | 44,892,887 | chr19:44892887:C:T | t | c | -0.9255 | 0.078 | 1.85E-32 | --------- | 7.977 | 8 | 0.4358 |
| A Score | chr19 | 44,892,962 | chr19:44892962:C:T | t | c | -0.7328 | 0.0639 | 2.01E-30 | --------- | 3.097 | 8 | 0.9281 |
| A Score | chr19 | 44,893,408 | chr19:44893408:G:T | t | g | -0.8049 | 0.0657 | 1.49E-34 | --------- | 1.633 | 8 | 0.9903 |
| A Score | chr19 | 44,893,716 | chr19:44893716:G:A | a | g | -0.7125 | 0.1443 | 7.86E-07 | ----?---- | 4.273 | 7 | 0.7478 |
| A Score | chr19 | 44,893,972 | chr19:44893972:G:A | a | g | 0.7108 | 0.1517 | 2.79E-06 | ++++-+-++ | 12.761 | 8 | 0.1203 |
| A Score | chr19 | 44,894,050 | chr19:44894050:C:T | t | c | -0.7355 | 0.1452 | 4.07E-07 | ----?---- | 3.674 | 7 | 0.8165 |
| A Score | chr19 | 44,894,695 | chr19:44894695:T:C | t | c | 0.7069 | 0.1436 | 8.47E-07 | ++++?++++ | 4.617 | 7 | 0.7065 |

| | | | | | | | | | | | | |
|---|---|---|---|---|---|---|---|---|---|---|---|---|
| A Score | chr19 | 44,895,376 | chr19:44895376:G:C | c | g | 0.3398 | 0.0716 | 2.06E-06 | -+++++--+ | 8.953 | 8 | 0.3463 |
| A Score | chr19 | 44,895,528 | chr19:44895528:C:T | t | c | -0.7474 | 0.1458 | 2.95E-07 | ----?---- | 3.737 | 7 | 0.8095 |
| A Score | chr19 | 44,895,560 | chr19:44895560:C:T | t | c | 0.2653 | 0.0539 | 8.59E-07 | --+++++++ | 14.812 | 8 | 0.06291 |
| A Score | chr19 | 44,896,087 | chr19:44896087:G:T | t | g | -0.7445 | 0.1456 | 3.16E-07 | ----?---- | 3.741 | 7 | 0.8091 |
| A Score | chr19 | 44,896,639 | chr19:44896639:G:A | a | g | -0.7455 | 0.1457 | 3.11E-07 | ----?---- | 3.766 | 7 | 0.8063 |
| A Score | chr19 | 44,897,227 | chr19:44897227:T:TG | t | tg | 0.7495 | 0.1462 | 2.93E-07 | ++++?++++ | 3.778 | 7 | 0.805 |
| A Score | chr19 | 44,897,468 | chr19:44897468:C:T | t | c | -0.7445 | 0.1456 | 3.16E-07 | ----?---- | 3.741 | 7 | 0.8091 |
| A Score | chr19 | 44,897,490 | chr19:44897490:T:A | a | t | 0.7039 | 0.1514 | 3.35E-06 | ++++-+-++ | 12.531 | 8 | 0.129 |
| A Score | chr19 | 44,898,409 | chr19:44898409:A:G | a | g | -0.2832 | 0.0535 | 1.19E-07 | ++------- | 15.365 | 8 | 0.05243 |
| A Score | chr19 | 44,898,611 | chr19:44898611:T:C | t | c | 0.5288 | 0.1241 | 2.05E-05 | +++++++++ | 2.234 | 8 | 0.973 |
| A Score | chr19 | 44,899,005 | chr19:44899005:T:G | t | g | -0.701 | 0.1515 | 3.71E-06 | ----+-+-- | 12.589 | 8 | 0.1268 |
| A Score | chr19 | 44,899,220 | chr19:44899220:C:T | t | c | 0.2769 | 0.0539 | 2.80E-07 | --+++++++ | 14.507 | 8 | 0.06947 |
| A Score | chr19 | 44,899,959 | chr19:44899959:C:T | t | c | -0.7563 | 0.1409 | 7.93E-08 | --------- | 3.043 | 8 | 0.9316 |
| A Score | chr19 | 44,900,155 | chr19:44900155:C:T | t | c | 0.3357 | 0.0527 | 1.84E-10 | +++++++++ | 19.893 | 8 | 0.01075 |
| A Score | chr19 | 44,900,601 | chr19:44900601:A:G | a | g | -0.3376 | 0.0529 | 1.76E-10 | --------- | 20.075 | 8 | 0.01005 |
| A Score | chr19 | 44,901,174 | chr19:44901174:T:C | t | c | -0.3386 | 0.053 | 1.66E-10 | --------- | 20.106 | 8 | 0.009944 |
| A Score | chr19 | 44,901,434 | chr19:44901434:A:G | a | g | 0.2828 | 0.0587 | 1.44E-06 | +++++++-+ | 7.424 | 8 | 0.4916 |
| A Score | chr19 | 44,901,600 | chr19:44901600:T:C | t | c | 0.7689 | 0.1408 | 4.74E-08 | +++++++++ | 3.764 | 8 | 0.8777 |
| A Score | chr19 | 44,901,715 | chr19:44901715:T:C | t | c | -0.3422 | 0.053 | 1.11E-10 | --------- | 19.948 | 8 | 0.01053 |
| A Score | chr19 | 44,901,805 | chr19:44901805:A:G | a | g | -0.3423 | 0.053 | 1.03E-10 | --------- | 20.063 | 8 | 0.0101 |
| A Score | chr19 | 44,902,264 | chr19:44902264:G:C | c | g | 0.2776 | 0.0539 | 2.64E-07 | --+++++++ | 14.26 | 8 | 0.07522 |
| A Score | chr19 | 44,903,281 | chr19:44903281:C:CG | cg | c | 0.3413 | 0.0534 | 1.58E-10 | +-+++++++ | 18.135 | 8 | 0.02024 |
| A Score | chr19 | 44,903,416 | chr19:44903416:G:A | a | g | -0.6195 | 0.0583 | 2.09E-26 | --------- | 7.606 | 8 | 0.4729 |
| A Score | chr19 | 44,904,531 | chr19:44904531:G:A | a | g | 0.3541 | 0.053 | 2.46E-11 | +++++++++ | 19.976 | 8 | 0.01043 |
| A Score | chr19 | 44,905,307 | chr19:44905307:A:T | a | t | -0.3343 | 0.0733 | 5.05E-06 | ------+-- | 4.757 | 8 | 0.7832 |
| A Score | chr19 | 44,905,579 | chr19:44905579:T:G | t | g | -0.2989 | 0.0524 | 1.16E-08 | -+------- | 14.338 | 8 | 0.07336 |
| A Score | chr19 | 44,905,910 | chr19:44905910:C:G | c | g | 0.286 | 0.0547 | 1.70E-07 | -++++++++ | 13.907 | 8 | 0.08422 |
| A Score | chr19 | 44,906,745 | chr19:44906745:G:A | a | g | -1.1887 | 0.086 | 2.02E-43 | --------- | 5.905 | 8 | 0.6578 |
| A Score | chr19 | 44,907,187 | chr19:44907187:G:A | a | g | 0.306 | 0.0544 | 1.86E-08 | -++++++++ | 16.153 | 8 | 0.04024 |
| A Score | chr19 | 44,908,684 | chr19:44908684:T:C | t | c | 1.2414 | 0.0779 | 3.68E-57 | +++++++++ | 6.453 | 8 | 0.5967 |
| A Score | chr19 | 44,908,822 | chr19:44908822:C:T | t | c | 0.7647 | 0.1047 | 2.74E-13 | ++++-++++ | 9.882 | 8 | 0.2734 |
| A Score | chr19 | 44,909,698 | chr19:44909698:A:C | a | c | 0.7894 | 0.1411 | 2.21E-08 | +++++++++ | 3.513 | 8 | 0.8982 |
| A Score | chr19 | 44,909,976 | chr19:44909976:G:T | t | g | 0.7537 | 0.105 | 7.07E-13 | ++++-++++ | 9.524 | 8 | 0.3 |
| A Score | chr19 | 44,910,319 | chr19:44910319:C:T | t | c | -0.5994 | 0.067 | 3.90E-19 | +-------- | 13.024 | 8 | 0.111 |
| A Score | chr19 | 44,911,194 | chr19:44911194:T:C | t | c | 0.2203 | 0.0544 | 5.16E-05 | -+++++--+ | 16.877 | 8 | 0.03142 |
| A Score | chr19 | 44,912,456 | chr19:44912456:G:A | a | g | -1.2052 | 0.0861 | 1.54E-44 | --------- | 5.313 | 8 | 0.7237 |
| A Score | chr19 | 44,912,678 | chr19:44912678:G:T | t | g | -1.2061 | 0.0861 | 1.43E-44 | --------- | 5.291 | 8 | 0.7261 |
| A Score | chr19 | 44,912,921 | chr19:44912921:G:T | t | g | -0.7201 | 0.0637 | 1.19E-29 | +-------- | 11.17 | 8 | 0.1922 |
| A Score | chr19 | 44,913,034 | chr19:44913034:C:T | t | c | 0.373 | 0.0551 | 1.31E-11 | +++++++++ | 11.303 | 8 | 0.1851 |
| A Score | chr19 | 44,913,221 | chr19:44913221:A:G | a | g | 0.2287 | 0.0549 | 3.11E-05 | -+++++--+ | 17.998 | 8 | 0.02124 |
| A Score | chr19 | 44,913,484 | chr19:44913484:C:T | t | c | -0.7246 | 0.0637 | 5.22E-30 | +-------- | 11.008 | 8 | 0.2012 |
| A Score | chr19 | 44,915,533 | chr19:44915533:T:C | t | c | 0.7353 | 0.064 | 1.42E-30 | -++++++++ | 10.978 | 8 | 0.2029 |
| A Score | chr19 | 44,915,704 | chr19:44915704:T:C | t | c | 0.2231 | 0.0549 | 4.79E-05 | -+++++--+ | 16.254 | 8 | 0.03889 |
| A Score | chr19 | 44,916,825 | chr19:44916825:A:C | a | c | 1.1455 | 0.0832 | 3.74E-43 | -++++++++ | 10.51 | 8 | 0.231 |
| A Score | chr19 | 44,917,843 | chr19:44917843:G:A | a | g | 0.3814 | 0.0553 | 5.56E-12 | -++++++++ | 10.711 | 8 | 0.2186 |
| A Score | chr19 | 44,917,947 | chr19:44917947:C:T | t | c | -0.7574 | 0.1404 | 6.86E-08 | -+------- | 3.867 | 8 | 0.8689 |

| | | | | | | | | | | | | |
|---|---|---|---|---|---|---|---|---|---|---|---|---|
| A Score | chr19 | 44,917,997 | chr19:44917997:G:A | a | g | -0.984 | 0.0769 | 1.72E-37 | +-------- | 12.122 | 8 | 0.1459 |
| A Score | chr19 | 44,918,393 | chr19:44918393:G:A | a | g | -0.7633 | 0.1389 | 3.95E-08 | --------- | 3.684 | 8 | 0.8845 |
| A Score | chr19 | 44,918,487 | chr19:44918487:G:T | t | g | 0.3538 | 0.0574 | 7.08E-10 | -+++-++++ | 16.747 | 8 | 0.03286 |
| A Score | chr19 | 44,918,620 | chr19:44918620:A:G | a | g | 0.2863 | 0.0544 | 1.46E-07 | -+++++--+ | 16.52 | 8 | 0.03551 |
| A Score | chr19 | 44,918,715 | chr19:44918715:AG:A | a | ag | 0.3537 | 0.0575 | 7.62E-10 | -+++-++++ | 16.978 | 8 | 0.03034 |
| A Score | chr19 | 44,918,903 | chr19:44918903:C:G | c | g | 1.0566 | 0.0713 | 1.01E-49 | +++++++++ | 7.265 | 8 | 0.5083 |
| A Score | chr19 | 44,919,589 | chr19:44919589:G:A | a | g | -1.0528 | 0.0712 | 1.84E-49 | --------- | 7.115 | 8 | 0.5243 |
| A Score | chr19 | 44,919,689 | chr19:44919689:A:G | a | g | 1.0544 | 0.0709 | 4.63E-50 | +++++++++ | 7.093 | 8 | 0.5266 |
| A Score | chr19 | 44,920,379 | chr19:44920379:G:A | a | g | 0.3537 | 0.0576 | 8.31E-10 | -+++-++++ | 15.561 | 8 | 0.04911 |
| A Score | chr19 | 44,921,809 | chr19:44921809:G:A | a | g | -0.7766 | 0.1459 | 1.03E-07 | ----?---- | 2.821 | 7 | 0.901 |
| A Score | chr19 | 44,921,918 | chr19:44921918:T:G | t | g | 0.2839 | 0.055 | 2.50E-07 | -+++++--+ | 16.104 | 8 | 0.04092 |
| A Score | chr19 | 44,921,921 | chr19:44921921:G:A | a | g | 0.6746 | 0.1244 | 5.82E-08 | +-+++++-+ | 13.945 | 8 | 0.08322 |
| A Score | chr19 | 44,922,203 | chr19:44922203:A:G | a | g | 0.2824 | 0.0552 | 3.15E-07 | -+++++--+ | 15.814 | 8 | 0.04512 |
| A Score | chr19 | 44,923,535 | chr19:44923535:G:A | a | g | 0.6667 | 0.123 | 5.89E-08 | +-+++++-+ | 13.822 | 8 | 0.08652 |
| A Score | chr19 | 44,923,868 | chr19:44923868:T:A | a | t | -0.9989 | 0.0776 | 6.92E-38 | +-------- | 12.558 | 8 | 0.128 |
| A Score | chr19 | 44,924,096 | chr19:44924096:C:G | c | g | -0.3829 | 0.057 | 1.87E-11 | +---+---- | 17.268 | 8 | 0.02743 |
| A Score | chr19 | 44,924,977 | chr19:44924977:G:A | a | g | -0.9925 | 0.0777 | 2.23E-37 | +-------- | 12.665 | 8 | 0.1239 |
| A Score | chr19 | 44,925,202 | chr19:44925202:C:T | t | c | 0.3688 | 0.0587 | 3.29E-10 | -+++-++++ | 15.397 | 8 | 0.05187 |
| A Score | chr19 | 44,926,286 | chr19:44926286:C:T | t | c | 0.2631 | 0.0602 | 1.26E-05 | -+++-++++ | 10.225 | 8 | 0.2496 |
| A Score | chr19 | 44,926,451 | chr19:44926451:G:C | c | g | -0.5081 | 0.0999 | 3.64E-07 | +---+---- | 7.17 | 8 | 0.5185 |
| A Score | chr19 | 44,935,297 | chr19:44935297:C:T | t | c | -0.3427 | 0.062 | 3.34E-08 | +------+- | 7.206 | 8 | 0.5146 |
| A Score | chr19 | 45,207,018 | chr19:45207018:A:G | a | g | 0.2878 | 0.0739 | 9.77E-05 | +++++++++ | 2.254 | 8 | 0.9722 |
| A Score | chr19 | 45,207,281 | chr19:45207281:C:T | t | c | -0.2919 | 0.0741 | 8.14E-05 | --------- | 2.253 | 8 | 0.9722 |
| A Score | chr19 | 45,207,605 | chr19:45207605:C:T | t | c | -0.2884 | 0.0739 | 9.48E-05 | --------- | 2.204 | 8 | 0.9741 |
| A Score | chr2 | 1,609,532 | chr2:1609532:C:T | t | c | -0.2835 | 0.0669 | 2.29E-05 | ----+---- | 11.805 | 8 | 0.1601 |
| A Score | chr2 | 1,610,323 | chr2:1610323:G:A | a | g | -0.2789 | 0.0668 | 3.01E-05 | ----+---+ | 12.376 | 8 | 0.1352 |
| A Score | chr2 | 1,610,610 | chr2:1610610:G:A | a | g | -0.2795 | 0.0665 | 2.63E-05 | ----+---+ | 12.528 | 8 | 0.1292 |
| A Score | chr2 | 1,613,837 | chr2:1613837:G:A | a | g | -0.2874 | 0.0713 | 5.61E-05 | --------- | 11.696 | 8 | 0.1653 |
| A Score | chr2 | 1,613,905 | chr2:1613905:C:T | t | c | -0.261 | 0.0644 | 5.03E-05 | ----+---- | 7.906 | 8 | 0.4427 |
| A Score | chr2 | 1,615,831 | chr2:1615831:G:C | c | g | -0.2913 | 0.0713 | 4.37E-05 | --------- | 11.195 | 8 | 0.1909 |
| A Score | chr2 | 1,616,545 | chr2:1616545:T:C | t | c | 0.2912 | 0.0713 | 4.39E-05 | +++++++++ | 11.196 | 8 | 0.1909 |
| A Score | chr2 | 1,617,372 | chr2:1617372:C:T | t | c | -0.2913 | 0.0712 | 4.33E-05 | --------- | 11.135 | 8 | 0.1941 |
| A Score | chr2 | 1,617,787 | chr2:1617787:A:AC | a | ac | 0.2866 | 0.0713 | 5.83E-05 | +++++++++ | 11.333 | 8 | 0.1835 |
| A Score | chr2 | 1,618,368 | chr2:1618368:G:T | t | g | -0.2913 | 0.0712 | 4.32E-05 | --------- | 10.916 | 8 | 0.2065 |
| A Score | chr2 | 1,618,677 | chr2:1618677:T:C | t | c | 0.2922 | 0.0715 | 4.36E-05 | +++++++++ | 11.041 | 8 | 0.1994 |
| A Score | chr2 | 2,314,360 | chr2:2314360:T:C | t | c | -0.7025 | 0.1763 | 6.75E-05 | ----+--+- | 9.925 | 8 | 0.2703 |
| A Score | chr2 | 2,450,834 | chr2:2450834:C:T | t | c | 1.3265 | 0.3216 | 3.72E-05 | -????+??+ | 1.602 | 2 | 0.4489 |
| A Score | chr2 | 3,944,159 | chr2:3944159:C:T | t | c | 0.9092 | 0.2015 | 6.44E-06 | -+++-++-+ | 7.136 | 8 | 0.522 |
| A Score | chr2 | 16,993,519 | chr2:16993519:C:T | t | c | 1.1319 | 0.2728 | 3.34E-05 | --++??+?+ | 4.972 | 5 | 0.4193 |
| A Score | chr2 | 20,400,873 | chr2:20400873:CA:C | ca | c | -0.5625 | 0.1444 | 9.75E-05 | -?--+-+-- | 9.823 | 7 | 0.1988 |
| A Score | chr2 | 25,035,528 | chr2:25035528:T:C | t | c | -0.5442 | 0.1397 | 9.78E-05 | -+----+-- | 4.446 | 8 | 0.8148 |
| A Score | chr2 | 25,654,829 | chr2:25654829:C:T | t | c | 0.683 | 0.1705 | 6.16E-05 | +-++-++++ | 5.119 | 8 | 0.7448 |
| A Score | chr2 | 36,886,271 | chr2:36886271:T:C | t | c | -0.8919 | 0.2261 | 7.99E-05 | -+--+-+-- | 13.117 | 8 | 0.1079 |
| A Score | chr2 | 36,892,709 | chr2:36892709:T:C | t | c | -0.8942 | 0.2262 | 7.72E-05 | -+--+-+-- | 13.268 | 8 | 0.103 |
| A Score | chr2 | 41,299,229 | chr2:41299229:C:G | c | g | -0.4823 | 0.1217 | 7.40E-05 | +--++---- | 15.793 | 8 | 0.04544 |
| A Score | chr2 | 47,669,420 | chr2:47669420:C:G | c | g | -0.2597 | 0.0644 | 5.51E-05 | +---+--+- | 9.265 | 8 | 0.3205 |

| | | | | | | | | | | | | |
|---|---|---|---|---|---|---|---|---|---|---|---|---|
| A Score | chr2 | 47,670,117 | chr2:47670117:G:C | c | g | 0.251 | 0.0635 | 7.76E-05 | -+++-++++ | 6.676 | 8 | 0.572 |
| A Score | chr2 | 47,674,582 | chr2:47674582:A:G | a | g | -0.2546 | 0.0641 | 7.19E-05 | +---+---- | 6.621 | 8 | 0.578 |
| A Score | chr2 | 61,956,978 | chr2:61956978:T:C | t | c | -0.6929 | 0.1774 | 9.40E-05 | -+------- | 1.585 | 8 | 0.9912 |
| A Score | chr2 | 78,825,371 | chr2:78825371:G:T | t | g | 1.0758 | 0.2465 | 1.28E-05 | --+++++++ | 5.975 | 8 | 0.65 |
| A Score | chr2 | 98,742,324 | chr2:98742324:G:A | a | g | 1.3561 | 0.3466 | 9.12E-05 | ?++?+???? | 1.067 | 2 | 0.5864 |
| A Score | chr2 | 131,017,950 | chr2:131017950:T:G | t | g | 0.6422 | 0.1546 | 3.25E-05 | +++++++++ | 4.887 | 8 | 0.7696 |
| A Score | chr2 | 131,019,656 | chr2:131019656:T:C | t | c | 0.6495 | 0.1622 | 6.18E-05 | +++++++++ | 5.96 | 8 | 0.6517 |
| A Score | chr2 | 131,019,718 | chr2:131019718:C:T | t | c | -0.6459 | 0.1515 | 2.00E-05 | --------- | 5.132 | 8 | 0.7434 |
| A Score | chr2 | 131,020,202 | chr2:131020202:A:G | a | g | 0.6335 | 0.1516 | 2.93E-05 | +++++++++ | 5.546 | 8 | 0.698 |
| A Score | chr2 | 131,021,265 | chr2:131021265:T:C | t | c | 0.6562 | 0.1614 | 4.82E-05 | +++++++++ | 6.038 | 8 | 0.643 |
| A Score | chr2 | 131,022,854 | chr2:131022854:T:C | t | c | 0.6297 | 0.1519 | 3.39E-05 | +++++++++ | 5.545 | 8 | 0.6981 |
| A Score | chr2 | 131,023,250 | chr2:131023250:G:A | a | g | -0.5694 | 0.1463 | 9.93E-05 | --------- | 7.262 | 8 | 0.5087 |
| A Score | chr2 | 131,024,322 | chr2:131024322:T:C | t | c | 0.6534 | 0.162 | 5.47E-05 | +++++++++ | 5.893 | 8 | 0.6593 |
| A Score | chr2 | 131,024,413 | chr2:131024413:C:CT | ct | c | -0.6482 | 0.1545 | 2.71E-05 | --------- | 5.148 | 8 | 0.7416 |
| A Score | chr2 | 131,024,681 | chr2:131024681:A:G | a | g | 0.6395 | 0.1538 | 3.22E-05 | +++++++++ | 5.367 | 8 | 0.7178 |
| A Score | chr2 | 131,024,700 | chr2:131024700:C:T | t | c | -0.6534 | 0.162 | 5.48E-05 | --------- | 5.894 | 8 | 0.6591 |
| A Score | chr2 | 131,025,254 | chr2:131025254:T:C | t | c | 0.657 | 0.162 | 5.03E-05 | +++++++++ | 6.045 | 8 | 0.6422 |
| A Score | chr2 | 131,025,506 | chr2:131025506:C:T | t | c | -0.644 | 0.1523 | 2.34E-05 | --------- | 5.408 | 8 | 0.7132 |
| A Score | chr2 | 131,025,813 | chr2:131025813:G:A | a | g | -0.651 | 0.1543 | 2.46E-05 | --------- | 5.343 | 8 | 0.7203 |
| A Score | chr2 | 131,026,605 | chr2:131026605:C:T | t | c | -0.6503 | 0.1545 | 2.57E-05 | --------- | 5.192 | 8 | 0.7369 |
| A Score | chr2 | 131,028,537 | chr2:131028537:T:A | a | t | -0.5803 | 0.1487 | 9.50E-05 | --------- | 6.854 | 8 | 0.5525 |
| A Score | chr2 | 131,033,001 | chr2:131033001:T:C | t | c | 0.6113 | 0.1567 | 9.55E-05 | +++++++-+ | 7.533 | 8 | 0.4804 |
| A Score | chr2 | 132,023,323 | chr2:132023323:A:C | a | c | -1.1523 | 0.2938 | 8.76E-05 | ----??+-? | 12.523 | 5 | 0.02828 |
| A Score | chr2 | 149,572,905 | chr2:149572905:G:T | t | g | 0.837 | 0.1745 | 1.61E-06 | +-+++++-+ | 3.455 | 8 | 0.9027 |
| A Score | chr2 | 151,049,935 | chr2:151049935:GA:G | g | ga | 0.2295 | 0.0561 | 4.26E-05 | ++++-++-+ | 8.538 | 8 | 0.3827 |
| A Score | chr2 | 151,054,881 | chr2:151054881:C:T | t | c | 0.2037 | 0.052 | 8.83E-05 | +++--++-+ | 11.355 | 8 | 0.1824 |
| A Score | chr2 | 151,055,283 | chr2:151055283:C:T | t | c | 0.2318 | 0.056 | 3.44E-05 | ++++-++-+ | 8.862 | 8 | 0.3541 |
| A Score | chr2 | 151,058,641 | chr2:151058641:G:T | t | g | 0.204 | 0.0519 | 8.42E-05 | +++--++-+ | 11.623 | 8 | 0.1688 |
| A Score | chr2 | 151,064,285 | chr2:151064285:C:T | t | c | 0.2037 | 0.0519 | 8.59E-05 | +++--++-+ | 11.535 | 8 | 0.1732 |
| A Score | chr2 | 151,066,842 | chr2:151066842:T:G | t | g | 0.2029 | 0.0518 | 9.06E-05 | +++--++-+ | 10.642 | 8 | 0.2228 |
| A Score | chr2 | 151,068,773 | chr2:151068773:G:T | t | g | -0.2032 | 0.0519 | 8.96E-05 | ---++--+- | 10.581 | 8 | 0.2266 |
| A Score | chr2 | 151,069,958 | chr2:151069958:G:A | a | g | 0.2199 | 0.0559 | 8.33E-05 | ++++-+--+ | 7.64 | 8 | 0.4694 |
| A Score | chr2 | 202,088,541 | chr2:202088541:C:G | c | g | 0.3962 | 0.1013 | 9.16E-05 | ++++-++++ | 9.729 | 8 | 0.2845 |
| A Score | chr2 | 205,687,520 | chr2:205687520:G:A | a | g | -0.2641 | 0.0655 | 5.59E-05 | +---+---- | 8.311 | 8 | 0.4037 |
| A Score | chr2 | 208,668,013 | chr2:208668013:GC:G | g | gc | 0.919 | 0.2134 | 1.67E-05 | ++++?++++ | 6.06 | 7 | 0.5327 |
| A Score | chr2 | 208,746,279 | chr2:208746279:T:C | t | c | -0.8563 | 0.2151 | 6.85E-05 | ----?+--- | 7.259 | 7 | 0.4024 |
| A Score | chr2 | 208,795,449 | chr2:208795449:T:C | t | c | -0.8684 | 0.2159 | 5.77E-05 | ----?+--- | 7.67 | 7 | 0.3626 |
| A Score | chr2 | 208,833,781 | chr2:208833781:G:C | c | g | 0.8428 | 0.2081 | 5.12E-05 | +-+++-+++ | 9.516 | 8 | 0.3007 |
| A Score | chr2 | 217,731,865 | chr2:217731865:T:C | t | c | 11.8799 | 2.9956 | 7.32E-05 | +???-+??? | 5.987 | 2 | 0.0501 |
| A Score | chr2 | 220,800,302 | chr2:220800302:C:T | t | c | 0.5707 | 0.1409 | 5.14E-05 | +-+++++++ | 6.687 | 8 | 0.5708 |
| A Score | chr2 | 225,918,986 | chr2:225918986:A:C | a | c | -1.578 | 0.3703 | 2.04E-05 | ?-?-??-?? | 1.104 | 2 | 0.5759 |
| A Score | chr2 | 239,577,581 | chr2:239577581:C:T | t | c | 0.7991 | 0.1933 | 3.56E-05 | +++++++++ | 11.703 | 8 | 0.1649 |
| A Score | chr2 | 239,590,326 | chr2:239590326:C:T | t | c | 0.6618 | 0.1679 | 8.12E-05 | +-++++++- | 6.811 | 8 | 0.5572 |
| A Score | chr2 | 239,608,129 | chr2:239608129:G:A | a | g | 0.663 | 0.1661 | 6.54E-05 | +-++++++- | 6.172 | 8 | 0.628 |
| A Score | chr2 | 240,453,765 | chr2:240453765:T:C | t | c | -0.3256 | 0.083 | 8.68E-05 | --------- | 10.421 | 8 | 0.2367 |
| A Score | chr20 | 9,023,019 | chr20:9023019:A:G | a | g | -0.2897 | 0.0732 | 7.61E-05 | -+--+---- | 11.054 | 8 | 0.1987 |

| | | | | | | | | | | | | |
|---|---|---|---|---|---|---|---|---|---|---|---|---|
| A Score | chr20 | 32,388,229 | chr20:32388229:C:T | t | c | 0.3897 | 0.0979 | 6.86E-05 | +++++++++ | 6.01 | 8 | 0.6461 |
| A Score | chr20 | 40,544,088 | chr20:40544088:C:T | t | c | 0.2314 | 0.0543 | 2.02E-05 | --++-+--- | 22.105 | 8 | 0.004725 |
| A Score | chr20 | 53,289,563 | chr20:53289563:T:C | t | c | -0.2136 | 0.0546 | 9.26E-05 | +---+---- | 12.238 | 8 | 0.1409 |
| A Score | chr20 | 60,881,393 | chr20:60881393:C:A | a | c | -0.2372 | 0.0606 | 9.01E-05 | ----+---- | 7.736 | 8 | 0.4596 |
| A Score | chr21 | 17,083,590 | chr21:17083590:T:C | t | c | -1.2585 | 0.3115 | 5.34E-05 | ?--???-+- | 4.398 | 4 | 0.3549 |
| A Score | chr21 | 27,247,800 | chr21:27247800:A:G | a | g | -1.4743 | 0.3688 | 6.40E-05 | ?+?????-- | 1.886 | 2 | 0.3894 |
| A Score | chr21 | 44,795,325 | chr21:44795325:T:C | t | c | -1.3763 | 0.3358 | 4.15E-05 | ?---??-?? | 2.309 | 3 | 0.5109 |
| A Score | chr21 | 45,123,679 | chr21:45123679:C:T | t | c | -0.7888 | 0.2026 | 9.91E-05 | -?--+---- | 2.183 | 7 | 0.949 |
| A Score | chr21 | 45,141,195 | chr21:45141195:C:T | t | c | -0.7251 | 0.1675 | 1.50E-05 | -?------- | 1.619 | 7 | 0.9779 |
| A Score | chr22 | 17,850,701 | chr22:17850701:C:T | t | c | 0.5163 | 0.1165 | 9.30E-06 | +++++++++ | 11.624 | 8 | 0.1688 |
| A Score | chr22 | 49,272,438 | chr22:49272438:C:T | t | c | 0.5676 | 0.1363 | 3.14E-05 | +-+++++++ | 11.636 | 8 | 0.1682 |
| A Score | chr22 | 49,274,046 | chr22:49274046:C:T | t | c | 0.4839 | 0.1222 | 7.44E-05 | +++++++++ | 8.326 | 8 | 0.4023 |
| A Score | chr22 | 49,275,389 | chr22:49275389:A:G | a | g | -0.4844 | 0.1221 | 7.28E-05 | --------- | 8.331 | 8 | 0.4018 |
| A Score | chr22 | 49,275,671 | chr22:49275671:G:A | a | g | 0.4848 | 0.122 | 7.07E-05 | +++++++++ | 8.348 | 8 | 0.4002 |
| A Score | chr22 | 49,276,245 | chr22:49276245:T:G | t | g | -0.4844 | 0.1221 | 7.26E-05 | --------- | 8.332 | 8 | 0.4017 |
| A Score | chr3 | 2,192,346 | chr3:2192346:A:G | a | g | -0.2119 | 0.0537 | 7.95E-05 | -------+- | 4.259 | 8 | 0.833 |
| A Score | chr3 | 2,193,728 | chr3:2193728:A:G | a | g | -0.2143 | 0.0537 | 6.66E-05 | -------+- | 3.078 | 8 | 0.9293 |
| A Score | chr3 | 2,197,500 | chr3:2197500:C:T | t | c | 0.2124 | 0.0539 | 8.20E-05 | +++++++-+ | 4.025 | 8 | 0.8549 |
| A Score | chr3 | 10,979,316 | chr3:10979316:G:A | a | g | 0.3762 | 0.0954 | 8.03E-05 | ++++-++++ | 5.17 | 8 | 0.7393 |
| A Score | chr3 | 14,742,741 | chr3:14742741:T:C | t | c | -0.8358 | 0.1947 | 1.76E-05 | -+------- | 4.942 | 8 | 0.7637 |
| A Score | chr3 | 14,748,407 | chr3:14748407:C:G | c | g | -0.2885 | 0.0703 | 4.08E-05 | +-----++- | 5.927 | 8 | 0.6554 |
| A Score | chr3 | 30,639,653 | chr3:30639653:T:C | t | c | -1.7617 | 0.3883 | 5.71E-06 | ???-??-+? | 0.536 | 2 | 0.7648 |
| A Score | chr3 | 31,971,654 | chr3:31971654:C:A | a | c | -0.5419 | 0.1391 | 9.80E-05 | ----+-++- | 9.631 | 8 | 0.2919 |
| A Score | chr3 | 33,949,202 | chr3:33949202:C:A | a | c | 0.4171 | 0.1052 | 7.41E-05 | ++++++-++ | 6.188 | 8 | 0.6262 |
| A Score | chr3 | 38,783,660 | chr3:38783660:A:G | a | g | -0.9185 | 0.1945 | 2.34E-06 | +---+-+-- | 6.883 | 8 | 0.5493 |
| A Score | chr3 | 59,631,155 | chr3:59631155:G:T | t | g | -0.6152 | 0.1539 | 6.39E-05 | --------- | 2.423 | 8 | 0.9652 |
| A Score | chr3 | 62,414,654 | chr3:62414654:G:A | a | g | -0.2776 | 0.0705 | 8.27E-05 | -+----++- | 6.429 | 8 | 0.5993 |
| A Score | chr3 | 75,559,589 | chr3:75559589:T:C | t | c | -0.2557 | 0.0629 | 4.84E-05 | ------+-+ | 13.68 | 8 | 0.0905 |
| A Score | chr3 | 75,561,652 | chr3:75561652:C:T | t | c | 0.2442 | 0.0626 | 9.47E-05 | -+++++-+- | 11.886 | 8 | 0.1564 |
| A Score | chr3 | 75,594,035 | chr3:75594035:A:G | a | g | -0.2674 | 0.0613 | 1.29E-05 | ++----+-- | 9.248 | 8 | 0.3218 |
| A Score | chr3 | 75,594,526 | chr3:75594526:C:G | c | g | -0.248 | 0.0614 | 5.44E-05 | ++----+-- | 8.097 | 8 | 0.4241 |
| A Score | chr3 | 75,596,274 | chr3:75596274:G:A | a | g | 0.267 | 0.0612 | 1.29E-05 | --++++-++ | 8.876 | 8 | 0.3529 |
| A Score | chr3 | 75,597,199 | chr3:75597199:C:A | a | c | 0.2462 | 0.0614 | 6.05E-05 | --++++-++ | 7.668 | 8 | 0.4665 |
| A Score | chr3 | 75,597,359 | chr3:75597359:T:C | t | c | -0.2617 | 0.0608 | 1.69E-05 | ++----+-- | 8.268 | 8 | 0.4077 |
| A Score | chr3 | 75,598,793 | chr3:75598793:G:A | a | g | 0.2464 | 0.061 | 5.34E-05 | --++++-++ | 7.361 | 8 | 0.4983 |
| A Score | chr3 | 75,603,691 | chr3:75603691:C:T | t | c | 0.2561 | 0.0619 | 3.53E-05 | --++++-++ | 10.672 | 8 | 0.221 |
| A Score | chr3 | 75,606,127 | chr3:75606127:C:T | t | c | 0.2409 | 0.0609 | 7.74E-05 | --++++-++ | 8.04 | 8 | 0.4296 |
| A Score | chr3 | 75,608,958 | chr3:75608958:G:A | a | g | 0.2481 | 0.0605 | 4.14E-05 | +-++++-++ | 7.337 | 8 | 0.5008 |
| A Score | chr3 | 75,611,550 | chr3:75611550:A:G | a | g | -0.2454 | 0.0605 | 5.00E-05 | -+----+-- | 7.266 | 8 | 0.5083 |
| A Score | chr3 | 75,613,922 | chr3:75613922:A:G | a | g | -0.2443 | 0.0607 | 5.80E-05 | ++----+-- | 7.115 | 8 | 0.5243 |
| A Score | chr3 | 75,615,189 | chr3:75615189:G:A | a | g | 0.2454 | 0.0608 | 5.37E-05 | --++++-++ | 7.166 | 8 | 0.5188 |
| A Score | chr3 | 75,616,187 | chr3:75616187:ATTTAT:A | a | atttat | 0.2405 | 0.061 | 8.09E-05 | --++++-++ | 6.297 | 8 | 0.614 |
| A Score | chr3 | 75,632,681 | chr3:75632681:G:T | t | g | 0.2559 | 0.0597 | 1.81E-05 | +-++++-++ | 10.889 | 8 | 0.2081 |
| A Score | chr3 | 75,633,060 | chr3:75633060:G:A | a | g | 0.2718 | 0.0602 | 6.43E-06 | +-++++-++ | 11.297 | 8 | 0.1854 |
| A Score | chr3 | 75,633,727 | chr3:75633727:G:T | t | g | 0.2513 | 0.0596 | 2.48E-05 | +-++++-++ | 11.43 | 8 | 0.1785 |
| A Score | chr3 | 75,634,151 | chr3:75634151:G:T | t | g | 0.2516 | 0.0596 | 2.41E-05 | +-++++-++ | 11.387 | 8 | 0.1807 |

| | | | | | | | | | | | | |
|---|---|---|---|---|---|---|---|---|---|---|---|---|
| A Score | chr3 | 75,638,660 | chr3:75638660:G:T | t | g | 0.2529 | 0.0597 | 2.31E-05 | +-++++-++ | 11.408 | 8 | 0.1796 |
| A Score | chr3 | 75,639,583 | chr3:75639583:C:A | a | c | 0.253 | 0.0597 | 2.29E-05 | +-++++-++ | 11.396 | 8 | 0.1802 |
| A Score | chr3 | 75,639,945 | chr3:75639945:A:G | a | g | -0.2521 | 0.0597 | 2.45E-05 | -+----+-- | 11.367 | 8 | 0.1817 |
| A Score | chr3 | 75,644,766 | chr3:75644766:C:T | t | c | 0.261 | 0.0608 | 1.74E-05 | +-++++-++ | 12.128 | 8 | 0.1456 |
| A Score | chr3 | 75,645,663 | chr3:75645663:G:A | a | g | 0.2589 | 0.0608 | 2.03E-05 | +-++++-++ | 12.156 | 8 | 0.1444 |
| A Score | chr3 | 75,645,686 | chr3:75645686:T:C | t | c | -0.2584 | 0.0608 | 2.12E-05 | -+----+-- | 12.223 | 8 | 0.1415 |
| A Score | chr3 | 75,646,661 | chr3:75646661:C:T | t | c | 0.2595 | 0.0607 | 1.94E-05 | +-++++-++ | 11.806 | 8 | 0.1601 |
| A Score | chr3 | 75,647,124 | chr3:75647124:A:C | a | c | -0.2588 | 0.0608 | 2.05E-05 | -+----+-- | 12.266 | 8 | 0.1397 |
| A Score | chr3 | 75,651,002 | chr3:75651002:C:T | t | c | 0.258 | 0.0608 | 2.17E-05 | +-++++-++ | 11.439 | 8 | 0.178 |
| A Score | chr3 | 75,651,066 | chr3:75651066:A:G | a | g | -0.2546 | 0.0608 | 2.84E-05 | -+----+-- | 11.408 | 8 | 0.1796 |
| A Score | chr3 | 75,651,625 | chr3:75651625:G:T | t | g | 0.2572 | 0.0608 | 2.32E-05 | +-++++-++ | 11.531 | 8 | 0.1734 |
| A Score | chr3 | 75,651,893 | chr3:75651893:C:G | c | g | -0.2572 | 0.0608 | 2.32E-05 | -+----+-- | 11.533 | 8 | 0.1733 |
| A Score | chr3 | 75,652,937 | chr3:75652937:T:C | t | c | -0.2573 | 0.0608 | 2.32E-05 | -+----+-- | 11.551 | 8 | 0.1724 |
| A Score | chr3 | 75,654,770 | chr3:75654770:C:A | a | c | 0.2349 | 0.0602 | 9.60E-05 | +-++++-++ | 11.713 | 8 | 0.1645 |
| A Score | chr3 | 75,656,079 | chr3:75656079:A:T | a | t | -0.2545 | 0.061 | 3.04E-05 | ------+-- | 10.296 | 8 | 0.2449 |
| A Score | chr3 | 75,657,552 | chr3:75657552:C:T | t | c | 0.2432 | 0.0609 | 6.53E-05 | ++++++-++ | 9.981 | 8 | 0.2664 |
| A Score | chr3 | 104,191,920 | chr3:104191920:A:G | a | g | -0.2226 | 0.053 | 2.65E-05 | --------- | 4.15 | 8 | 0.8433 |
| A Score | chr3 | 149,021,012 | chr3:149021012:C:T | t | c | 0.7028 | 0.1778 | 7.69E-05 | --++++++- | 6.777 | 8 | 0.5609 |
| A Score | chr3 | 149,112,447 | chr3:149112447:G:A | a | g | 0.6951 | 0.1778 | 9.24E-05 | --++++++- | 6.332 | 8 | 0.6101 |
| A Score | chr3 | 190,741,279 | chr3:190741279:G:A | a | g | -0.2309 | 0.0592 | 9.67E-05 | --+------ | 15.051 | 8 | 0.05817 |
| A Score | chr3 | 190,744,075 | chr3:190744075:T:C | t | c | 0.2299 | 0.0579 | 7.09E-05 | ++++-++++ | 12.236 | 8 | 0.141 |
| A Score | chr3 | 190,746,658 | chr3:190746658:A:G | a | g | 0.2292 | 0.0578 | 7.38E-05 | ++++-++++ | 12.862 | 8 | 0.1167 |
| A Score | chr3 | 190,751,863 | chr3:190751863:A:G | a | g | 0.227 | 0.0577 | 8.34E-05 | ++++-++++ | 12.711 | 8 | 0.1222 |
| A Score | chr3 | 190,752,770 | chr3:190752770:T:G | t | g | 0.2254 | 0.0577 | 9.27E-05 | ++++-++++ | 12.535 | 8 | 0.1289 |
| A Score | chr3 | 190,753,433 | chr3:190753433:G:A | a | g | -0.2258 | 0.0577 | 9.06E-05 | ----+---- | 12.685 | 8 | 0.1232 |
| A Score | chr3 | 190,757,636 | chr3:190757636:G:A | a | g | -0.2266 | 0.0577 | 8.50E-05 | ----+---- | 12.7 | 8 | 0.1226 |
| A Score | chr4 | 4,168,918 | chr4:4168918:G:C | c | g | 0.3565 | 0.0902 | 7.68E-05 | +-+++++-+ | 5.622 | 8 | 0.6895 |
| A Score | chr4 | 27,328,987 | chr4:27328987:G:A | a | g | 0.4072 | 0.1046 | 9.97E-05 | --++-+-++ | 12.839 | 8 | 0.1175 |
| A Score | chr4 | 40,471,138 | chr4:40471138:T:C | t | c | -0.4899 | 0.1187 | 3.65E-05 | ++--+-++- | 12.836 | 8 | 0.1176 |
| A Score | chr4 | 43,235,809 | chr4:43235809:G:A | a | g | 0.9888 | 0.2277 | 1.41E-05 | ?+++?+++? | 7.897 | 5 | 0.162 |
| A Score | chr4 | 53,351,440 | chr4:53351440:T:C | t | c | -0.2948 | 0.0752 | 8.80E-05 | --------- | 10.055 | 8 | 0.2612 |
| A Score | chr4 | 53,354,006 | chr4:53354006:C:T | t | c | 0.2934 | 0.0751 | 9.41E-05 | +++++++++ | 10.103 | 8 | 0.2579 |
| A Score | chr4 | 53,354,303 | chr4:53354303:G:A | a | g | 0.2941 | 0.0751 | 9.04E-05 | +++++++++ | 10.104 | 8 | 0.2578 |
| A Score | chr4 | 53,359,686 | chr4:53359686:C:T | t | c | 0.2917 | 0.0749 | 9.96E-05 | +++++++++ | 10.135 | 8 | 0.2557 |
| A Score | chr4 | 53,372,091 | chr4:53372091:T:C | t | c | -0.2918 | 0.0749 | 9.85E-05 | --------- | 10.138 | 8 | 0.2555 |
| A Score | chr4 | 57,820,798 | chr4:57820798:G:A | a | g | 1.6393 | 0.3815 | 1.73E-05 | +??????++ | 0.61 | 2 | 0.737 |
| A Score | chr4 | 81,418,710 | chr4:81418710:A:G | a | g | -0.6626 | 0.1646 | 5.67E-05 | +------+- | 6.324 | 8 | 0.611 |
| A Score | chr4 | 84,524,852 | chr4:84524852:G:A | a | g | 1.0043 | 0.2544 | 7.90E-05 | ++++-+++- | 5.358 | 8 | 0.7187 |
| A Score | chr4 | 87,814,807 | chr4:87814807:G:C | c | g | 0.2319 | 0.0555 | 2.90E-05 | +++++++++ | 6.049 | 8 | 0.6418 |
| A Score | chr4 | 87,815,429 | chr4:87815429:T:G | t | g | -0.2273 | 0.0556 | 4.29E-05 | --------- | 6.57 | 8 | 0.5836 |
| A Score | chr4 | 87,817,020 | chr4:87817020:C:G | c | g | -0.2269 | 0.0555 | 4.28E-05 | --------- | 5.9 | 8 | 0.6584 |
| A Score | chr4 | 87,819,281 | chr4:87819281:C:T | t | c | 0.2293 | 0.0555 | 3.57E-05 | +++++++++ | 6.041 | 8 | 0.6427 |
| A Score | chr4 | 87,819,537 | chr4:87819537:C:G | c | g | -0.2314 | 0.0555 | 3.01E-05 | ---+----- | 6.339 | 8 | 0.6093 |
| A Score | chr4 | 87,820,846 | chr4:87820846:T:C | t | c | -0.2282 | 0.0553 | 3.63E-05 | --------- | 5.977 | 8 | 0.6498 |
| A Score | chr4 | 87,822,084 | chr4:87822084:TA:T | t | ta | 0.2268 | 0.0553 | 4.12E-05 | +++++++++ | 5.883 | 8 | 0.6603 |
| A Score | chr4 | 87,822,113 | chr4:87822113:G:A | a | g | 0.2223 | 0.0553 | 5.78E-05 | +++++++++ | 6.055 | 8 | 0.6411 |

| | | | | | | | | | | | | |
|---|---|---|---|---|---|---|---|---|---|---|---|---|
| A Score | chr4 | 87,822,389 | chr4:87822389:G:A | a | g | 0.2237 | 0.0553 | 5.18E-05 | +++++++++ | 6.004 | 8 | 0.6468 |
| A Score | chr4 | 87,823,650 | chr4:87823650:T:A | a | t | 0.2224 | 0.0551 | 5.50E-05 | +++++++++ | 6.043 | 8 | 0.6424 |
| A Score | chr4 | 87,823,973 | chr4:87823973:G:A | a | g | 0.2199 | 0.0551 | 6.48E-05 | +++++++++ | 6.052 | 8 | 0.6414 |
| A Score | chr4 | 87,824,695 | chr4:87824695:T:C | t | c | -0.2199 | 0.0551 | 6.63E-05 | --------- | 6.072 | 8 | 0.6391 |
| A Score | chr4 | 87,825,582 | chr4:87825582:A:G | a | g | -0.2209 | 0.055 | 5.97E-05 | --------- | 6.072 | 8 | 0.6392 |
| A Score | chr4 | 87,825,735 | chr4:87825735:G:A | a | g | 0.2223 | 0.0551 | 5.53E-05 | +++++++++ | 6.043 | 8 | 0.6424 |
| A Score | chr4 | 87,825,850 | chr4:87825850:C:A | a | c | 0.2223 | 0.0551 | 5.52E-05 | +++++++++ | 6.044 | 8 | 0.6423 |
| A Score | chr4 | 87,827,110 | chr4:87827110:CG:C | cg | c | -0.2202 | 0.0551 | 6.49E-05 | --------- | 6.079 | 8 | 0.6384 |
| A Score | chr4 | 87,831,235 | chr4:87831235:G:A | a | g | 0.2245 | 0.0551 | 4.65E-05 | +++++++++ | 5.624 | 8 | 0.6893 |
| A Score | chr4 | 87,831,726 | chr4:87831726:T:C | t | c | -0.2228 | 0.0551 | 5.33E-05 | --------- | 5.651 | 8 | 0.6862 |
| A Score | chr4 | 87,834,056 | chr4:87834056:G:A | a | g | 0.2216 | 0.0552 | 5.93E-05 | +++++++++ | 5.654 | 8 | 0.6859 |
| A Score | chr4 | 87,835,523 | chr4:87835523:C:T | t | c | 0.2267 | 0.0553 | 4.17E-05 | +++++++++ | 5.696 | 8 | 0.6812 |
| A Score | chr4 | 91,340,525 | chr4:91340525:G:T | t | g | 0.2275 | 0.0572 | 6.90E-05 | +++++++++ | 9.273 | 8 | 0.3198 |
| A Score | chr4 | 91,342,961 | chr4:91342961:T:C | t | c | -0.2295 | 0.0572 | 5.95E-05 | --------- | 9.08 | 8 | 0.3356 |
| A Score | chr4 | 91,344,469 | chr4:91344469:G:C | c | g | 0.2302 | 0.0573 | 5.91E-05 | +++++++++ | 8.809 | 8 | 0.3586 |
| A Score | chr4 | 91,346,391 | chr4:91346391:G:A | a | g | 0.232 | 0.0573 | 5.13E-05 | +++++++++ | 8.966 | 8 | 0.3452 |
| A Score | chr4 | 91,349,429 | chr4:91349429:GT:G | g | gt | 0.2321 | 0.0572 | 5.00E-05 | +++++++++ | 9.02 | 8 | 0.3406 |
| A Score | chr4 | 98,989,152 | chr4:98989152:C:T | t | c | 0.3406 | 0.083 | 4.08E-05 | +-+++++++ | 12.596 | 8 | 0.1265 |
| A Score | chr4 | 99,002,556 | chr4:99002556:ACT:A | a | act | 0.3446 | 0.083 | 3.29E-05 | +-+++++++ | 11.676 | 8 | 0.1662 |
| A Score | chr4 | 99,005,274 | chr4:99005274:A:G | a | g | -0.343 | 0.083 | 3.59E-05 | -+------- | 11.766 | 8 | 0.162 |
| A Score | chr4 | 99,005,937 | chr4:99005937:G:A | a | g | 0.3412 | 0.083 | 3.96E-05 | +-+++++++ | 11.855 | 8 | 0.1578 |
| A Score | chr4 | 99,008,559 | chr4:99008559:A:G | a | g | -0.3448 | 0.083 | 3.25E-05 | -+------- | 11.681 | 8 | 0.166 |
| A Score | chr4 | 99,009,377 | chr4:99009377:A:G | a | g | -0.3374 | 0.0829 | 4.73E-05 | -+------- | 9.443 | 8 | 0.3063 |
| A Score | chr4 | 99,011,541 | chr4:99011541:G:T | t | g | 0.3412 | 0.083 | 3.95E-05 | +-+++++++ | 11.98 | 8 | 0.1521 |
| A Score | chr4 | 99,014,731 | chr4:99014731:G:A | a | g | 0.343 | 0.083 | 3.59E-05 | +-+++++++ | 11.912 | 8 | 0.1552 |
| A Score | chr4 | 99,015,066 | chr4:99015066:G:A | a | g | 0.3445 | 0.083 | 3.35E-05 | +-+++++++ | 11.909 | 8 | 0.1553 |
| A Score | chr4 | 99,017,570 | chr4:99017570:G:A | a | g | 0.3434 | 0.083 | 3.50E-05 | +-+++++++ | 11.923 | 8 | 0.1547 |
| A Score | chr4 | 99,019,348 | chr4:99019348:G:A | a | g | 0.3429 | 0.083 | 3.62E-05 | +-+++++++ | 11.91 | 8 | 0.1553 |
| A Score | chr4 | 99,024,056 | chr4:99024056:G:A | a | g | 0.345 | 0.083 | 3.22E-05 | +-+++++++ | 11.968 | 8 | 0.1527 |
| A Score | chr4 | 99,032,127 | chr4:99032127:C:T | t | c | 0.345 | 0.083 | 3.22E-05 | +-+++++++ | 11.967 | 8 | 0.1527 |
| A Score | chr4 | 99,032,690 | chr4:99032690:G:A | a | g | 0.345 | 0.083 | 3.23E-05 | +-+++++++ | 11.969 | 8 | 0.1526 |
| A Score | chr4 | 99,042,256 | chr4:99042256:G:T | t | g | 0.3391 | 0.0831 | 4.50E-05 | +-+++++++ | 12.665 | 8 | 0.1239 |
| A Score | chr4 | 99,046,632 | chr4:99046632:T:G | t | g | -0.3426 | 0.083 | 3.67E-05 | -+------- | 12.176 | 8 | 0.1435 |
| A Score | chr4 | 99,047,660 | chr4:99047660:C:T | t | c | 0.3329 | 0.0831 | 6.22E-05 | +-+++++++ | 12.333 | 8 | 0.137 |
| A Score | chr4 | 99,049,642 | chr4:99049642:A:G | a | g | -0.3257 | 0.0827 | 8.19E-05 | -+------- | 15.066 | 8 | 0.05788 |
| A Score | chr4 | 99,057,546 | chr4:99057546:G:A | a | g | 0.326 | 0.0828 | 8.30E-05 | +-+++++++ | 15.227 | 8 | 0.05488 |
| A Score | chr4 | 99,470,280 | chr4:99470280:T:G | t | g | -0.4676 | 0.1148 | 4.63E-05 | -?--+---- | 4.555 | 7 | 0.714 |
| A Score | chr4 | 99,470,405 | chr4:99470405:C:T | t | c | 0.463 | 0.1149 | 5.57E-05 | +?++-++++ | 4.358 | 7 | 0.7378 |
| A Score | chr4 | 99,470,483 | chr4:99470483:C:A | a | c | 0.4634 | 0.1149 | 5.49E-05 | +?++-++++ | 4.339 | 7 | 0.74 |
| A Score | chr4 | 99,470,764 | chr4:99470764:G:A | a | g | 0.4632 | 0.1148 | 5.49E-05 | +?++-++++ | 4.347 | 7 | 0.7391 |
| A Score | chr4 | 99,470,903 | chr4:99470903:A:G | a | g | -0.4632 | 0.1148 | 5.50E-05 | -?--+---- | 4.347 | 7 | 0.739 |
| A Score | chr4 | 109,304,500 | chr4:109304500:A:ATCTTC | a | atcttc | 0.2792 | 0.0716 | 9.61E-05 | ++++++--+ | 11.638 | 8 | 0.1681 |
| A Score | chr4 | 109,315,167 | chr4:109315167:C:T | t | c | -0.3378 | 0.0844 | 6.29E-05 | ------++- | 12.065 | 8 | 0.1483 |
| A Score | chr4 | 109,329,152 | chr4:109329152:T:C | t | c | 0.2746 | 0.0702 | 9.06E-05 | ++++++--+ | 13.225 | 8 | 0.1043 |
| A Score | chr4 | 109,330,050 | chr4:109330050:A:G | a | g | 0.2742 | 0.0702 | 9.30E-05 | ++++++--+ | 13.333 | 8 | 0.1009 |
| A Score | chr4 | 109,332,944 | chr4:109332944:A:G | a | g | 0.3388 | 0.0844 | 5.97E-05 | ++++++--+ | 11.991 | 8 | 0.1516 |

| | | | | | | | | | | | | |
|---|---|---|---|---|---|---|---|---|---|---|---|---|
| A Score | chr4 | 109,333,984 | chr4:109333984:A:G | a | g | 0.274 | 0.07 | 9.20E-05 | ++++++--+ | 13.239 | 8 | 0.1039 |
| A Score | chr4 | 109,335,535 | chr4:109335535:T:G | t | g | 0.3393 | 0.0843 | 5.72E-05 | ++++++--+ | 11.737 | 8 | 0.1633 |
| A Score | chr4 | 109,341,392 | chr4:109341392:G:A | a | g | -0.3388 | 0.0844 | 5.95E-05 | ------++- | 11.992 | 8 | 0.1515 |
| A Score | chr4 | 109,342,801 | chr4:109342801:T:C | t | c | 0.2748 | 0.0702 | 8.97E-05 | ++++++--+ | 13.214 | 8 | 0.1047 |
| A Score | chr4 | 109,354,002 | chr4:109354002:G:A | a | g | -0.2748 | 0.07 | 8.61E-05 | ------++- | 12.432 | 8 | 0.133 |
| A Score | chr4 | 109,354,447 | chr4:109354447:C:T | t | c | -0.2764 | 0.0699 | 7.77E-05 | ------++- | 12.486 | 8 | 0.1308 |
| A Score | chr4 | 109,354,619 | chr4:109354619:G:A | a | g | -0.2763 | 0.0699 | 7.80E-05 | ------++- | 12.483 | 8 | 0.1309 |
| A Score | chr4 | 109,356,010 | chr4:109356010:T:C | t | c | 0.2754 | 0.07 | 8.23E-05 | ++++++--+ | 12.494 | 8 | 0.1305 |
| A Score | chr4 | 109,357,660 | chr4:109357660:G:A | a | g | -0.275 | 0.0697 | 7.97E-05 | ------++- | 12.614 | 8 | 0.1258 |
| A Score | chr4 | 109,358,130 | chr4:109358130:C:G | c | g | 0.2764 | 0.0699 | 7.79E-05 | ++++++--+ | 12.483 | 8 | 0.1309 |
| A Score | chr4 | 109,358,568 | chr4:109358568:C:T | t | c | -0.2798 | 0.07 | 6.47E-05 | ------++- | 13.037 | 8 | 0.1106 |
| A Score | chr4 | 109,358,621 | chr4:109358621:C:T | t | c | -0.2763 | 0.0699 | 7.79E-05 | ------++- | 12.483 | 8 | 0.1309 |
| A Score | chr4 | 109,359,934 | chr4:109359934:AG:A | a | ag | -0.2812 | 0.0701 | 6.01E-05 | ------++- | 13.019 | 8 | 0.1112 |
| A Score | chr4 | 109,360,247 | chr4:109360247:C:T | t | c | -0.2763 | 0.07 | 7.82E-05 | ------++- | 12.488 | 8 | 0.1307 |
| A Score | chr4 | 109,360,396 | chr4:109360396:G:A | a | g | -0.2763 | 0.07 | 7.84E-05 | ------++- | 12.485 | 8 | 0.1308 |
| A Score | chr4 | 109,360,417 | chr4:109360417:A:G | a | g | 0.2815 | 0.0701 | 5.87E-05 | ++++++--+ | 12.977 | 8 | 0.1126 |
| A Score | chr4 | 109,360,452 | chr4:109360452:C:G | c | g | 0.2763 | 0.07 | 7.83E-05 | ++++++--+ | 12.486 | 8 | 0.1308 |
| A Score | chr4 | 109,360,697 | chr4:109360697:G:A | a | g | -0.2763 | 0.07 | 7.83E-05 | ------++- | 12.482 | 8 | 0.1309 |
| A Score | chr4 | 109,360,796 | chr4:109360796:G:C | c | g | -0.2762 | 0.07 | 7.85E-05 | ------++- | 12.486 | 8 | 0.1308 |
| A Score | chr4 | 109,360,923 | chr4:109360923:C:T | t | c | -0.2761 | 0.0699 | 7.93E-05 | ------++- | 12.469 | 8 | 0.1315 |
| A Score | chr4 | 109,361,207 | chr4:109361207:T:G | t | g | 0.2763 | 0.07 | 7.84E-05 | ++++++--+ | 12.486 | 8 | 0.1308 |
| A Score | chr4 | 109,361,422 | chr4:109361422:C:T | t | c | -0.2801 | 0.0701 | 6.38E-05 | ------++- | 13.06 | 8 | 0.1098 |
| A Score | chr4 | 109,361,729 | chr4:109361729:G:C | c | g | -0.2762 | 0.07 | 7.91E-05 | ------++- | 12.486 | 8 | 0.1308 |
| A Score | chr4 | 109,361,778 | chr4:109361778:G:C | c | g | -0.28 | 0.07 | 6.38E-05 | ------++- | 13.045 | 8 | 0.1103 |
| A Score | chr4 | 109,365,610 | chr4:109365610:A:G | a | g | 0.2786 | 0.0713 | 9.37E-05 | ++++++--+ | 14.862 | 8 | 0.06189 |
| A Score | chr4 | 109,366,038 | chr4:109366038:C:T | t | c | -0.2785 | 0.0713 | 9.43E-05 | ------++- | 14.858 | 8 | 0.06196 |
| A Score | chr4 | 109,366,949 | chr4:109366949:C:T | t | c | -0.2846 | 0.0715 | 6.91E-05 | ------++- | 14.511 | 8 | 0.06939 |
| A Score | chr4 | 109,368,439 | chr4:109368439:C:T | t | c | -0.2845 | 0.0715 | 6.97E-05 | ------++- | 14.515 | 8 | 0.0693 |
| A Score | chr4 | 109,373,227 | chr4:109373227:T:A | a | t | -0.3435 | 0.0847 | 4.98E-05 | ------++- | 12.505 | 8 | 0.1301 |
| A Score | chr4 | 109,377,899 | chr4:109377899:A:C | a | c | 0.2835 | 0.0715 | 7.28E-05 | ++++++--+ | 14.904 | 8 | 0.06105 |
| A Score | chr4 | 109,382,211 | chr4:109382211:A:G | a | g | 0.2807 | 0.0713 | 8.29E-05 | ++++++--+ | 15.131 | 8 | 0.05665 |
| A Score | chr4 | 109,386,841 | chr4:109386841:G:T | t | g | -0.2807 | 0.0714 | 8.45E-05 | ------++- | 13.848 | 8 | 0.08582 |
| A Score | chr4 | 109,389,970 | chr4:109389970:T:C | t | c | 0.2814 | 0.0714 | 8.12E-05 | ++++++--+ | 14.036 | 8 | 0.08084 |
| A Score | chr4 | 112,789,337 | chr4:112789337:T:C | t | c | -0.6709 | 0.1601 | 2.79E-05 | +-----+-- | 7.198 | 8 | 0.5154 |
| A Score | chr4 | 114,404,480 | chr4:114404480:T:C | t | c | -1.125 | 0.243 | 3.67E-06 | ?---?---? | 4.463 | 5 | 0.4848 |
| A Score | chr4 | 114,454,138 | chr4:114454138:A:G | a | g | -0.209 | 0.0531 | 8.23E-05 | ----+---- | 4.263 | 8 | 0.8326 |
| A Score | chr4 | 114,495,265 | chr4:114495265:G:A | a | g | 0.7903 | 0.1925 | 4.04E-05 | +++++++++ | 2.177 | 8 | 0.9751 |
| A Score | chr4 | 114,577,462 | chr4:114577462:G:C | c | g | 0.9072 | 0.2271 | 6.50E-05 | ++++++++? | 3.061 | 7 | 0.8793 |
| A Score | chr4 | 114,595,032 | chr4:114595032:G:C | c | g | 1.0494 | 0.2493 | 2.55E-05 | -+++?+++? | 4.727 | 6 | 0.5793 |
| A Score | chr4 | 148,447,966 | chr4:148447966:A:AT | a | at | -0.6616 | 0.1661 | 6.79E-05 | -------++ | 6.859 | 8 | 0.552 |
| A Score | chr4 | 187,686,669 | chr4:187686669:AT:A | a | at | -0.4324 | 0.0975 | 9.21E-06 | --------- | 5.785 | 8 | 0.6713 |
| A Score | chr5 | 2,159,800 | chr5:2159800:C:T | t | c | 0.6585 | 0.169 | 9.71E-05 | --+++++-+ | 2.748 | 8 | 0.9491 |
| A Score | chr5 | 2,240,892 | chr5:2240892:C:T | t | c | 0.7296 | 0.1823 | 6.28E-05 | --++?++-+ | 6.857 | 7 | 0.444 |
| A Score | chr5 | 29,296,715 | chr5:29296715:T:C | t | c | -0.2538 | 0.0626 | 5.06E-05 | ----+---- | 6.272 | 8 | 0.6168 |
| A Score | chr5 | 29,310,828 | chr5:29310828:G:A | a | g | 0.2258 | 0.0579 | 9.65E-05 | +++--++++ | 8.094 | 8 | 0.4244 |
| A Score | chr5 | 29,312,648 | chr5:29312648:A:C | a | c | -0.2272 | 0.0579 | 8.70E-05 | ---++---- | 8.045 | 8 | 0.4291 |

| | | | | | | | | | | | | |
|---|---|---|---|---|---|---|---|---|---|---|---|---|
| A Score | chr5 | 29,314,947 | chr5:29314947:A:T | a | t | -0.2255 | 0.0579 | 9.74E-05 | ----+---- | 8.084 | 8 | 0.4253 |
| A Score | chr5 | 29,315,283 | chr5:29315283:T:C | t | c | -0.2257 | 0.0579 | 9.67E-05 | ----+---- | 8.097 | 8 | 0.4241 |
| A Score | chr5 | 29,316,432 | chr5:29316432:G:A | a | g | 0.2251 | 0.0579 | 9.96E-05 | +++--++++ | 8.277 | 8 | 0.4068 |
| A Score | chr5 | 29,324,352 | chr5:29324352:G:T | t | g | 0.2274 | 0.0579 | 8.59E-05 | ++++-++++ | 7.985 | 8 | 0.4349 |
| A Score | chr5 | 29,325,456 | chr5:29325456:T:A | a | t | 0.2252 | 0.0578 | 9.89E-05 | +++--++++ | 8.284 | 8 | 0.4063 |
| A Score | chr5 | 30,343,285 | chr5:30343285:G:A | a | g | -0.2608 | 0.0661 | 8.08E-05 | ------+-- | 18.799 | 8 | 0.01597 |
| A Score | chr5 | 30,364,702 | chr5:30364702:G:A | a | g | -0.258 | 0.0661 | 9.62E-05 | ------+-- | 20.36 | 8 | 0.009058 |
| A Score | chr5 | 30,365,543 | chr5:30365543:C:G | c | g | 0.2577 | 0.0661 | 9.78E-05 | ++++++-++ | 20.379 | 8 | 0.008994 |
| A Score | chr5 | 30,366,068 | chr5:30366068:G:A | a | g | -0.2581 | 0.0661 | 9.57E-05 | ------+-- | 20.37 | 8 | 0.009023 |
| A Score | chr5 | 31,311,350 | chr5:31311350:C:T | t | c | 0.8963 | 0.2219 | 5.37E-05 | +-++?+?-+ | 5.682 | 6 | 0.4598 |
| A Score | chr5 | 31,382,711 | chr5:31382711:C:T | t | c | 0.7637 | 0.1727 | 9.75E-06 | ++++----+ | 9.52 | 8 | 0.3004 |
| A Score | chr5 | 39,451,510 | chr5:39451510:G:A | a | g | 0.6422 | 0.1614 | 6.89E-05 | --+++++-- | 15.573 | 8 | 0.04892 |
| A Score | chr5 | 40,098,077 | chr5:40098077:T:C | t | c | -0.944 | 0.2351 | 5.94E-05 | ?+--?-+?- | 6.66 | 5 | 0.2472 |
| A Score | chr5 | 40,170,452 | chr5:40170452:C:T | t | c | 1.0365 | 0.238 | 1.33E-05 | ?-++?+-?+ | 1.183 | 5 | 0.9465 |
| A Score | chr5 | 44,021,715 | chr5:44021715:G:A | a | g | 0.5595 | 0.1402 | 6.59E-05 | -+++-++++ | 3.842 | 8 | 0.8711 |
| A Score | chr5 | 44,134,175 | chr5:44134175:T:C | t | c | -0.6035 | 0.1415 | 2.01E-05 | +-------- | 4.278 | 8 | 0.8312 |
| A Score | chr5 | 44,151,147 | chr5:44151147:T:A | a | t | 0.6054 | 0.1415 | 1.88E-05 | -++++++++ | 4.328 | 8 | 0.8263 |
| A Score | chr5 | 44,210,935 | chr5:44210935:G:A | a | g | 0.5971 | 0.1422 | 2.70E-05 | -++++++++ | 4.553 | 8 | 0.8041 |
| A Score | chr5 | 44,218,463 | chr5:44218463:CT:C | ct | c | -0.604 | 0.1419 | 2.09E-05 | +-------- | 4.633 | 8 | 0.796 |
| A Score | chr5 | 80,105,594 | chr5:80105594:T:C | t | c | -0.246 | 0.0628 | 8.95E-05 | -------+- | 5.516 | 8 | 0.7013 |
| A Score | chr5 | 83,237,865 | chr5:83237865:A:G | a | g | -0.5295 | 0.1351 | 8.89E-05 | -+------- | 6.507 | 8 | 0.5906 |
| A Score | chr5 | 107,823,640 | chr5:107823640:G:T | t | g | 0.8452 | 0.1952 | 1.50E-05 | --+-+++++ | 8.005 | 8 | 0.433 |
| A Score | chr5 | 117,439,914 | chr5:117439914:A:T | a | t | -0.3341 | 0.0814 | 4.02E-05 | ----+-+-- | 10.562 | 8 | 0.2278 |
| A Score | chr5 | 117,458,247 | chr5:117458247:T:C | t | c | -0.3412 | 0.0832 | 4.08E-05 | ----+-+-- | 9.266 | 8 | 0.3203 |
| A Score | chr5 | 117,458,610 | chr5:117458610:C:T | t | c | 0.3381 | 0.0837 | 5.37E-05 | ++++-+-++ | 9.098 | 8 | 0.3341 |
| A Score | chr5 | 117,459,015 | chr5:117459015:A:T | a | t | -0.3324 | 0.0837 | 7.11E-05 | ----+-+-- | 8.894 | 8 | 0.3513 |
| A Score | chr5 | 117,464,803 | chr5:117464803:A:G | a | g | -0.3338 | 0.0836 | 6.51E-05 | ----+-+-- | 9.417 | 8 | 0.3083 |
| A Score | chr5 | 117,467,746 | chr5:117467746:CT:C | ct | c | -0.3274 | 0.0837 | 9.19E-05 | ----+-+-- | 8.508 | 8 | 0.3855 |
| A Score | chr5 | 117,476,326 | chr5:117476326:C:T | t | c | 0.3367 | 0.0825 | 4.46E-05 | ++++-+-++ | 8.757 | 8 | 0.3632 |
| A Score | chr5 | 117,477,017 | chr5:117477017:T:C | t | c | -0.3359 | 0.0824 | 4.62E-05 | ----+-+-- | 8.79 | 8 | 0.3603 |
| A Score | chr5 | 117,477,043 | chr5:117477043:A:T | a | t | -0.3351 | 0.0824 | 4.81E-05 | ----+-+-- | 8.762 | 8 | 0.3628 |
| A Score | chr5 | 117,478,020 | chr5:117478020:A:G | a | g | -0.331 | 0.0825 | 5.97E-05 | ----+-+-- | 8.727 | 8 | 0.3659 |
| A Score | chr5 | 117,478,127 | chr5:117478127:A:G | a | g | -0.3334 | 0.0824 | 5.26E-05 | ----+-+-- | 8.742 | 8 | 0.3645 |
| A Score | chr5 | 117,478,418 | chr5:117478418:T:C | t | c | -0.3334 | 0.0824 | 5.25E-05 | ----+-+-- | 8.753 | 8 | 0.3636 |
| A Score | chr5 | 117,479,492 | chr5:117479492:T:A | a | t | 0.3331 | 0.0824 | 5.32E-05 | ++++-+-++ | 8.763 | 8 | 0.3627 |
| A Score | chr5 | 117,479,693 | chr5:117479693:T:G | t | g | -0.3333 | 0.0824 | 5.27E-05 | ----+-+-- | 8.756 | 8 | 0.3633 |
| A Score | chr5 | 117,480,515 | chr5:117480515:CCTT:C | cctt | c | -0.3339 | 0.0825 | 5.21E-05 | ----+-+-- | 8.665 | 8 | 0.3713 |
| A Score | chr5 | 117,480,679 | chr5:117480679:T:C | t | c | -0.3363 | 0.0825 | 4.58E-05 | ----+-+-- | 8.692 | 8 | 0.3689 |
| A Score | chr5 | 117,482,861 | chr5:117482861:G:A | a | g | 0.3291 | 0.0827 | 6.96E-05 | ++++-+-++ | 8.827 | 8 | 0.3571 |
| A Score | chr5 | 117,483,151 | chr5:117483151:G:A | a | g | 0.3279 | 0.0827 | 7.29E-05 | ++++-+-++ | 8.909 | 8 | 0.35 |
| A Score | chr5 | 141,646,014 | chr5:141646014:A:G | a | g | 0.2933 | 0.0744 | 8.06E-05 | ++++-++-+ | 8.122 | 8 | 0.4216 |
| A Score | chr5 | 141,646,527 | chr5:141646527:G:A | a | g | -0.328 | 0.0787 | 3.07E-05 | ----+---- | 3.552 | 8 | 0.8951 |
| A Score | chr5 | 141,675,050 | chr5:141675050:C:T | t | c | 0.2598 | 0.0645 | 5.64E-05 | +++++++++ | 2.086 | 8 | 0.9783 |
| A Score | chr5 | 141,676,702 | chr5:141676702:G:A | a | g | 0.2599 | 0.0643 | 5.37E-05 | +++++++++ | 2.082 | 8 | 0.9784 |
| A Score | chr5 | 141,677,745 | chr5:141677745:GT:G | g | gt | 0.2617 | 0.0642 | 4.62E-05 | +++++++++ | 2.083 | 8 | 0.9784 |
| A Score | chr5 | 141,678,711 | chr5:141678711:C:T | t | c | 0.2604 | 0.0643 | 5.09E-05 | +++++++++ | 1.991 | 8 | 0.9813 |

| | | | | | | | | | | | | |
|---|---|---|---|---|---|---|---|---|---|---|---|---|
| A Score | chr5 | 141,678,920 | chr5:141678920:G:A | a | g | 0.2584 | 0.0643 | 5.83E-05 | +++++++++ | 2.047 | 8 | 0.9795 |
| A Score | chr5 | 144,225,476 | chr5:144225476:A:C | a | c | -0.3587 | 0.0898 | 6.54E-05 | ----+---- | 5.963 | 8 | 0.6514 |
| A Score | chr5 | 144,308,643 | chr5:144308643:G:T | t | g | 0.4099 | 0.104 | 8.10E-05 | -+++?++++ | 3.875 | 7 | 0.7941 |
| A Score | chr5 | 144,354,180 | chr5:144354180:C:T | t | c | 0.4117 | 0.1051 | 8.93E-05 | -+++-++++ | 2.822 | 8 | 0.945 |
| A Score | chr5 | 162,435,356 | chr5:162435356:T:C | t | c | -0.9237 | 0.218 | 2.26E-05 | -+--?--+- | 10.748 | 7 | 0.15 |
| A Score | chr6 | 1,256,369 | chr6:1256369:C:CA | ca | c | 0.3846 | 0.097 | 7.34E-05 | +-+++++++ | 4.516 | 8 | 0.8078 |
| A Score | chr6 | 4,009,995 | chr6:4009995:T:C | t | c | -0.6704 | 0.1666 | 5.73E-05 | -------+- | 10.503 | 8 | 0.2314 |
| A Score | chr6 | 41,696,343 | chr6:41696343:C:T | t | c | 1.3592 | 0.2782 | 1.03E-06 | +??+?++++ | 5.147 | 5 | 0.3983 |
| A Score | chr6 | 67,224,559 | chr6:67224559:A:G | a | g | -1.3274 | 0.3148 | 2.48E-05 | +?--??-?? | 0.493 | 3 | 0.9204 |
| A Score | chr6 | 67,254,955 | chr6:67254955:C:A | a | c | 1.2848 | 0.3202 | 6.01E-05 | +?++??+?? | 0.54 | 3 | 0.9101 |
| A Score | chr6 | 70,239,404 | chr6:70239404:C:T | t | c | 0.2479 | 0.0624 | 7.12E-05 | ++++-++-+ | 9.556 | 8 | 0.2976 |
| A Score | chr6 | 70,348,892 | chr6:70348892:G:T | t | g | -0.2147 | 0.0524 | 4.24E-05 | ++-----+- | 13.22 | 8 | 0.1045 |
| A Score | chr6 | 70,354,489 | chr6:70354489:CT:C | ct | c | 0.821 | 0.2038 | 5.60E-05 | ++++-++-+ | 10.317 | 8 | 0.2434 |
| A Score | chr6 | 70,357,025 | chr6:70357025:G:C | c | g | -0.852 | 0.2077 | 4.09E-05 | ----+--+- | 10.806 | 8 | 0.2129 |
| A Score | chr6 | 70,357,254 | chr6:70357254:G:GTCTT | g | gtctt | -0.2165 | 0.0549 | 8.10E-05 | +---+--+- | 10.599 | 8 | 0.2255 |
| A Score | chr6 | 70,357,551 | chr6:70357551:TGAAAAA:T | t | tgaaaaa | -0.8513 | 0.2078 | 4.18E-05 | ----+--+- | 10.8 | 8 | 0.2133 |
| A Score | chr6 | 70,359,449 | chr6:70359449:C:T | t | c | 0.2191 | 0.055 | 6.72E-05 | -+++-++++ | 11.092 | 8 | 0.1966 |
| A Score | chr6 | 70,359,450 | chr6:70359450:A:G | a | g | -0.219 | 0.055 | 6.73E-05 | +---+---- | 11.095 | 8 | 0.1964 |
| A Score | chr6 | 70,359,451 | chr6:70359451:T:C | t | c | -0.219 | 0.055 | 6.73E-05 | +---+---- | 11.094 | 8 | 0.1964 |
| A Score | chr6 | 70,361,326 | chr6:70361326:A:G | a | g | -0.214 | 0.0548 | 9.34E-05 | +---+--+- | 11.418 | 8 | 0.1791 |
| A Score | chr6 | 70,361,886 | chr6:70361886:T:C | t | c | -0.2168 | 0.0547 | 7.41E-05 | +---+--+- | 11.455 | 8 | 0.1772 |
| A Score | chr6 | 70,362,835 | chr6:70362835:G:A | a | g | 0.2139 | 0.0547 | 9.36E-05 | -+++-++-+ | 10.943 | 8 | 0.2049 |
| A Score | chr6 | 70,365,764 | chr6:70365764:C:T | t | c | -0.2064 | 0.0523 | 7.98E-05 | +------+- | 13.082 | 8 | 0.1091 |
| A Score | chr6 | 70,366,970 | chr6:70366970:G:A | a | g | 0.2123 | 0.0545 | 9.93E-05 | -+++-++++ | 9.314 | 8 | 0.3165 |
| A Score | chr6 | 73,264,828 | chr6:73264828:G:A | a | g | 0.408 | 0.1048 | 9.85E-05 | ++++-++++ | 3.755 | 8 | 0.8785 |
| A Score | chr6 | 73,268,100 | chr6:73268100:G:A | a | g | 0.4077 | 0.1048 | 9.97E-05 | ++++-++++ | 3.746 | 8 | 0.8793 |
| A Score | chr6 | 73,279,062 | chr6:73279062:G:C | c | g | 0.4076 | 0.1048 | 9.97E-05 | ++++-++++ | 3.749 | 8 | 0.879 |
| A Score | chr6 | 73,279,133 | chr6:73279133:A:G | a | g | -0.4077 | 0.1048 | 9.96E-05 | ----+---- | 3.749 | 8 | 0.8791 |
| A Score | chr6 | 73,279,679 | chr6:73279679:G:C | c | g | 0.4083 | 0.1048 | 9.75E-05 | ++++-++++ | 3.752 | 8 | 0.8788 |
| A Score | chr6 | 73,280,590 | chr6:73280590:G:A | a | g | 0.4077 | 0.1048 | 9.95E-05 | ++++-++++ | 3.747 | 8 | 0.8792 |
| A Score | chr6 | 73,283,435 | chr6:73283435:T:G | t | g | -0.4105 | 0.1048 | 8.99E-05 | ----+---- | 3.645 | 8 | 0.8877 |
| A Score | chr6 | 73,283,830 | chr6:73283830:C:T | t | c | 0.4065 | 0.1043 | 9.75E-05 | ++++-++++ | 3.551 | 8 | 0.8952 |
| A Score | chr6 | 73,283,864 | chr6:73283864:C:T | t | c | 0.4062 | 0.1043 | 9.83E-05 | ++++-++++ | 3.544 | 8 | 0.8957 |
| A Score | chr6 | 73,285,629 | chr6:73285629:C:T | t | c | 0.4271 | 0.1073 | 6.85E-05 | ++++-++++ | 3.031 | 8 | 0.9324 |
| A Score | chr6 | 73,285,731 | chr6:73285731:C:T | t | c | 0.4159 | 0.104 | 6.40E-05 | ++++-++++ | 3.57 | 8 | 0.8937 |
| A Score | chr6 | 73,287,436 | chr6:73287436:A:G | a | g | -0.4077 | 0.1047 | 9.90E-05 | ----+---- | 3.858 | 8 | 0.8697 |
| A Score | chr6 | 73,288,594 | chr6:73288594:C:T | t | c | 0.4064 | 0.1043 | 9.79E-05 | ++++-++++ | 3.56 | 8 | 0.8945 |
| A Score | chr6 | 73,288,787 | chr6:73288787:A:G | a | g | -0.4064 | 0.1043 | 9.78E-05 | ----+---- | 3.563 | 8 | 0.8943 |
| A Score | chr6 | 73,296,705 | chr6:73296705:T:A | a | t | 0.4067 | 0.1043 | 9.65E-05 | ++++-++++ | 3.522 | 8 | 0.8975 |
| A Score | chr6 | 73,302,967 | chr6:73302967:C:T | t | c | 0.4106 | 0.1051 | 9.32E-05 | ++++-++++ | 3.943 | 8 | 0.8622 |
| A Score | chr6 | 73,311,751 | chr6:73311751:C:T | t | c | 0.425 | 0.1041 | 4.49E-05 | ++++-++++ | 4.021 | 8 | 0.8552 |
| A Score | chr6 | 73,313,794 | chr6:73313794:T:G | t | g | -0.4185 | 0.1045 | 6.22E-05 | ----+---- | 3.746 | 8 | 0.8793 |
| A Score | chr6 | 81,367,781 | chr6:81367781:A:G | a | g | -0.6216 | 0.1588 | 9.09E-05 | --------- | 6.26 | 8 | 0.6181 |
| A Score | chr6 | 88,550,699 | chr6:88550699:C:T | t | c | 0.3053 | 0.0775 | 8.15E-05 | --++-++-+ | 15.586 | 8 | 0.0487 |
| A Score | chr6 | 88,558,231 | chr6:88558231:G:A | a | g | 0.3122 | 0.0786 | 7.14E-05 | ++++-++-+ | 11.917 | 8 | 0.1549 |
| A Score | chr6 | 88,558,874 | chr6:88558874:T:C | t | c | -0.2983 | 0.0739 | 5.41E-05 | +---+--+- | 15.54 | 8 | 0.04946 |

| | | | | | | | | | | | | |
|---|---|---|---|---|---|---|---|---|---|---|---|---|
| A Score | chr6 | 88,558,996 | chr6:88558996:G:A | a | g | 0.2981 | 0.0738 | 5.41E-05 | -+++-++-+ | 15.504 | 8 | 0.05006 |
| A Score | chr6 | 96,048,937 | chr6:96048937:C:T | t | c | 0.7124 | 0.1782 | 6.37E-05 | -++++++-+ | 5.262 | 8 | 0.7292 |
| A Score | chr6 | 96,071,402 | chr6:96071402:A:T | a | t | -0.7157 | 0.1775 | 5.55E-05 | +------+- | 5.276 | 8 | 0.7278 |
| A Score | chr6 | 96,102,337 | chr6:96102337:A:C | a | c | -0.7396 | 0.1718 | 1.67E-05 | +-------- | 5.284 | 8 | 0.7268 |
| A Score | chr6 | 98,842,861 | chr6:98842861:C:T | t | c | 1.1427 | 0.2854 | 6.25E-05 | ??+????++ | 0.245 | 2 | 0.8848 |
| A Score | chr6 | 111,295,394 | chr6:111295394:T:C | t | c | -0.9649 | 0.23 | 2.73E-05 | ++----++- | 7.817 | 8 | 0.4516 |
| A Score | chr6 | 111,406,680 | chr6:111406680:G:A | a | g | 0.983 | 0.2272 | 1.52E-05 | --++++--+ | 9.042 | 8 | 0.3388 |
| A Score | chr6 | 144,276,371 | chr6:144276371:C:T | t | c | 0.6887 | 0.1712 | 5.77E-05 | ++++-++++ | 4.818 | 8 | 0.7768 |
| A Score | chr6 | 150,711,950 | chr6:150711950:T:C | t | c | 0.2119 | 0.0524 | 5.20E-05 | ++++-++-+ | 12.833 | 8 | 0.1177 |
| A Score | chr6 | 164,185,512 | chr6:164185512:G:A | a | g | 1.2556 | 0.2792 | 6.88E-06 | ?+++??+?? | 2.96 | 3 | 0.3979 |
| A Score | chr7 | 6,045,230 | chr7:6045230:C:G | c | g | -0.8262 | 0.1775 | 3.25E-06 | +-------- | 1.67 | 8 | 0.9895 |
| A Score | chr7 | 6,064,111 | chr7:6064111:G:A | a | g | 0.8509 | 0.1768 | 1.48E-06 | -++++++++ | 1.698 | 8 | 0.9889 |
| A Score | chr7 | 9,739,510 | chr7:9739510:G:A | a | g | -0.374 | 0.0932 | 5.98E-05 | -+--+-+-- | 14.972 | 8 | 0.0597 |
| A Score | chr7 | 23,393,184 | chr7:23393184:G:A | a | g | 1.0966 | 0.2766 | 7.37E-05 | ??++??-+? | 2.995 | 3 | 0.3924 |
| A Score | chr7 | 36,740,294 | chr7:36740294:A:T | a | t | 0.3056 | 0.0765 | 6.43E-05 | +++++++++ | 4.354 | 8 | 0.8238 |
| A Score | chr7 | 42,246,860 | chr7:42246860:A:T | a | t | -0.2094 | 0.0538 | 9.99E-05 | +------+- | 13.226 | 8 | 0.1043 |
| A Score | chr7 | 56,746,336 | chr7:56746336:C:T | t | c | 0.232 | 0.0579 | 6.16E-05 | +-+++-+++ | 10.547 | 8 | 0.2287 |
| A Score | chr7 | 56,749,629 | chr7:56749629:A:T | a | t | -0.2305 | 0.0577 | 6.53E-05 | -+---+--- | 10.336 | 8 | 0.2423 |
| A Score | chr7 | 56,749,997 | chr7:56749997:T:C | t | c | -0.2305 | 0.0577 | 6.53E-05 | -+---+--- | 10.321 | 8 | 0.2432 |
| A Score | chr7 | 56,751,124 | chr7:56751124:A:C | a | c | -0.227 | 0.0577 | 8.41E-05 | -+---+--- | 9.864 | 8 | 0.2747 |
| A Score | chr7 | 56,752,645 | chr7:56752645:C:G | c | g | -0.226 | 0.0577 | 9.04E-05 | -+---+--- | 9.995 | 8 | 0.2654 |
| A Score | chr7 | 56,753,328 | chr7:56753328:G:T | t | g | 0.226 | 0.0577 | 9.05E-05 | +-+++-+++ | 9.995 | 8 | 0.2654 |
| A Score | chr7 | 56,759,407 | chr7:56759407:T:C | t | c | -0.2307 | 0.0577 | 6.38E-05 | -+------- | 10.117 | 8 | 0.2569 |
| A Score | chr7 | 56,759,547 | chr7:56759547:C:T | t | c | 0.2326 | 0.0577 | 5.56E-05 | +-+++++++ | 9.962 | 8 | 0.2677 |
| A Score | chr7 | 56,764,271 | chr7:56764271:G:C | c | g | 0.2327 | 0.0578 | 5.59E-05 | +-+++++++ | 10.22 | 8 | 0.25 |
| A Score | chr7 | 56,773,194 | chr7:56773194:C:A | a | c | -0.2305 | 0.0579 | 6.78E-05 | -+-----+- | 11.513 | 8 | 0.1743 |
| A Score | chr7 | 92,904,524 | chr7:92904524:T:C | t | c | -0.4078 | 0.1001 | 4.66E-05 | -+------- | 5.053 | 8 | 0.7519 |
| A Score | chr7 | 92,950,386 | chr7:92950386:A:G | a | g | -0.3152 | 0.0806 | 9.22E-05 | ------+-- | 12.359 | 8 | 0.1359 |
| A Score | chr7 | 92,950,639 | chr7:92950639:C:T | t | c | 0.315 | 0.0806 | 9.34E-05 | ++++++-++ | 12.371 | 8 | 0.1354 |
| A Score | chr7 | 109,490,863 | chr7:109490863:A:G | a | g | -1.2009 | 0.2998 | 6.18E-05 | -?-???--? | 1.147 | 3 | 0.7657 |
| A Score | chr7 | 145,945,647 | chr7:145945647:T:A | a | t | -0.5157 | 0.1299 | 7.17E-05 | +-------- | 8.656 | 8 | 0.3721 |
| A Score | chr7 | 145,949,469 | chr7:145949469:G:A | a | g | -0.5059 | 0.1287 | 8.50E-05 | +-------- | 8.656 | 8 | 0.3721 |
| A Score | chr7 | 145,962,689 | chr7:145962689:G:C | c | g | -0.5174 | 0.1297 | 6.63E-05 | +-------- | 8.22 | 8 | 0.4123 |
| A Score | chr7 | 145,963,037 | chr7:145963037:G:A | a | g | -0.5216 | 0.1302 | 6.15E-05 | +-------- | 8.57 | 8 | 0.3799 |
| A Score | chr7 | 145,968,274 | chr7:145968274:C:A | a | c | -0.5388 | 0.1308 | 3.78E-05 | +-------- | 7.751 | 8 | 0.4582 |
| A Score | chr7 | 145,985,034 | chr7:145985034:G:T | t | g | -0.5253 | 0.1294 | 4.94E-05 | +-------- | 8.342 | 8 | 0.4008 |
| A Score | chr7 | 145,988,855 | chr7:145988855:TTC:T | t | ttc | -0.5299 | 0.1294 | 4.20E-05 | +-------- | 8.679 | 8 | 0.3701 |
| A Score | chr7 | 145,996,039 | chr7:145996039:G:A | a | g | -0.5374 | 0.1295 | 3.34E-05 | +-------- | 8.666 | 8 | 0.3712 |
| A Score | chr7 | 146,002,501 | chr7:146002501:C:T | t | c | -0.5373 | 0.1295 | 3.35E-05 | +-------- | 8.663 | 8 | 0.3715 |
| A Score | chr7 | 146,002,744 | chr7:146002744:C:CT | ct | c | -0.537 | 0.1295 | 3.40E-05 | +-------- | 8.676 | 8 | 0.3703 |
| A Score | chr7 | 146,004,623 | chr7:146004623:G:A | a | g | -0.5434 | 0.1289 | 2.48E-05 | +-------- | 9.789 | 8 | 0.2801 |
| A Score | chr7 | 146,008,105 | chr7:146008105:G:C | c | g | -0.5461 | 0.1295 | 2.49E-05 | +-------- | 9.786 | 8 | 0.2803 |
| A Score | chr7 | 146,011,013 | chr7:146011013:T:A | a | t | -0.5264 | 0.1298 | 4.98E-05 | +-------- | 8.53 | 8 | 0.3835 |
| A Score | chr7 | 146,012,389 | chr7:146012389:C:T | t | c | -0.5353 | 0.1298 | 3.73E-05 | +-------- | 8.925 | 8 | 0.3487 |
| A Score | chr7 | 146,013,452 | chr7:146013452:G:C | c | g | -0.5487 | 0.1294 | 2.23E-05 | +-------- | 10.122 | 8 | 0.2566 |
| A Score | chr7 | 146,014,094 | chr7:146014094:G:C | c | g | -0.5271 | 0.1298 | 4.88E-05 | +-------- | 11.941 | 8 | 0.1538 |

| | | | | | | | | | | | | |
|---|---|---|---|---|---|---|---|---|---|---|---|---|
| A Score | chr7 | 146,014,434 | chr7:146014434:T:G | t | g | 0.5084 | 0.1284 | 7.54E-05 | -++++++++ | 11.089 | 8 | 0.1967 |
| A Score | chr7 | 146,014,629 | chr7:146014629:C:T | t | c | -0.5396 | 0.1304 | 3.48E-05 | +-------- | 8.831 | 8 | 0.3568 |
| A Score | chr7 | 146,017,244 | chr7:146017244:G:C | c | g | -0.5487 | 0.1305 | 2.60E-05 | +-------- | 9.945 | 8 | 0.2689 |
| A Score | chr7 | 146,017,613 | chr7:146017613:G:A | a | g | -0.5283 | 0.13 | 4.81E-05 | +-------- | 8.749 | 8 | 0.3639 |
| A Score | chr7 | 146,017,792 | chr7:146017792:G:C | c | g | -0.5373 | 0.1301 | 3.60E-05 | +-------- | 9.841 | 8 | 0.2763 |
| A Score | chr7 | 146,017,972 | chr7:146017972:G:A | a | g | -0.5378 | 0.1301 | 3.54E-05 | +-------- | 9.88 | 8 | 0.2735 |
| A Score | chr7 | 146,018,079 | chr7:146018079:G:A | a | g | -0.5389 | 0.1301 | 3.45E-05 | +-------- | 9.803 | 8 | 0.2792 |
| A Score | chr7 | 146,018,101 | chr7:146018101:G:A | a | g | -0.5384 | 0.1301 | 3.51E-05 | +-------- | 9.936 | 8 | 0.2695 |
| A Score | chr7 | 146,018,122 | chr7:146018122:T:G | t | g | 0.5377 | 0.1301 | 3.57E-05 | -++++++++ | 9.881 | 8 | 0.2735 |
| A Score | chr7 | 146,018,688 | chr7:146018688:C:A | a | c | -0.5284 | 0.13 | 4.80E-05 | +-------- | 8.759 | 8 | 0.3631 |
| A Score | chr7 | 146,019,303 | chr7:146019303:A:C | a | c | 0.5378 | 0.13 | 3.54E-05 | -++++++++ | 9.881 | 8 | 0.2735 |
| A Score | chr7 | 146,019,439 | chr7:146019439:C:T | t | c | -0.5428 | 0.13 | 2.97E-05 | +-------- | 9.997 | 8 | 0.2653 |
| A Score | chr7 | 146,019,458 | chr7:146019458:C:A | a | c | -0.538 | 0.1301 | 3.52E-05 | +-------- | 9.881 | 8 | 0.2735 |
| A Score | chr7 | 146,019,852 | chr7:146019852:A:G | a | g | 0.5392 | 0.1302 | 3.44E-05 | -++++++++ | 9.993 | 8 | 0.2655 |
| A Score | chr7 | 146,020,622 | chr7:146020622:T:C | t | c | 0.5382 | 0.13 | 3.48E-05 | -++++++++ | 9.895 | 8 | 0.2725 |
| A Score | chr7 | 146,021,192 | chr7:146021192:A:T | a | t | 0.5258 | 0.13 | 5.27E-05 | -++++++++ | 8.985 | 8 | 0.3436 |
| A Score | chr7 | 146,021,242 | chr7:146021242:C:A | a | c | -0.5258 | 0.13 | 5.25E-05 | +-------- | 8.935 | 8 | 0.3478 |
| A Score | chr7 | 146,021,333 | chr7:146021333:G:A | a | g | -0.5268 | 0.13 | 5.08E-05 | +-------- | 8.958 | 8 | 0.3459 |
| A Score | chr7 | 146,021,691 | chr7:146021691:C:T | t | c | -0.5157 | 0.1296 | 6.89E-05 | +-------- | 8.72 | 8 | 0.3664 |
| A Score | chr7 | 155,596,117 | chr7:155596117:A:G | a | g | -0.6418 | 0.1623 | 7.63E-05 | +---+---- | 2.959 | 8 | 0.9369 |
| A Score | chr7 | 157,634,645 | chr7:157634645:G:A | a | g | 0.3398 | 0.0776 | 1.18E-05 | ++++-++-+ | 10.608 | 8 | 0.2249 |
| A Score | chr7 | 157,639,501 | chr7:157639501:C:T | t | c | 0.2457 | 0.0596 | 3.69E-05 | +++++++-+ | 9.833 | 8 | 0.277 |
| A Score | chr7 | 157,659,717 | chr7:157659717:C:A | a | c | 0.2391 | 0.0582 | 4.02E-05 | +++++++-+ | 9.62 | 8 | 0.2927 |
| A Score | chr8 | 5,783,136 | chr8:5783136:G:C | c | g | 0.343 | 0.0846 | 5.05E-05 | +-+++++-+ | 7.98 | 8 | 0.4354 |
| A Score | chr8 | 6,067,635 | chr8:6067635:G:A | a | g | 0.7603 | 0.1795 | 2.28E-05 | +++++++++ | 9.919 | 8 | 0.2707 |
| A Score | chr8 | 13,798,837 | chr8:13798837:C:G | c | g | -0.8323 | 0.2073 | 5.94E-05 | ++------- | 3.632 | 8 | 0.8887 |
| A Score | chr8 | 27,604,964 | chr8:27604964:A:G | a | g | 0.2206 | 0.0546 | 5.36E-05 | ++++-++++ | 7.054 | 8 | 0.5308 |
| A Score | chr8 | 27,607,002 | chr8:27607002:T:C | t | c | 0.2188 | 0.0541 | 5.29E-05 | +++++++++ | 7.332 | 8 | 0.5012 |
| A Score | chr8 | 27,607,412 | chr8:27607412:A:G | a | g | 0.212 | 0.0543 | 9.33E-05 | ++++-++++ | 6.469 | 8 | 0.5948 |
| A Score | chr8 | 27,608,664 | chr8:27608664:T:C | t | c | 0.2137 | 0.0547 | 9.26E-05 | ++++-++++ | 6.299 | 8 | 0.6138 |
| A Score | chr8 | 27,608,798 | chr8:27608798:T:C | t | c | 0.2206 | 0.0541 | 4.50E-05 | +++++++++ | 5.967 | 8 | 0.6509 |
| A Score | chr8 | 27,610,169 | chr8:27610169:C:T | t | c | -0.2292 | 0.0539 | 2.13E-05 | --------- | 8.875 | 8 | 0.3529 |
| A Score | chr8 | 27,610,304 | chr8:27610304:C:G | c | g | 0.2243 | 0.0546 | 4.00E-05 | +++++++++ | 7.642 | 8 | 0.4692 |
| A Score | chr8 | 27,931,789 | chr8:27931789:G:A | a | g | 0.8358 | 0.2143 | 9.62E-05 | ++++++++? | 11.2 | 7 | 0.1301 |
| A Score | chr8 | 27,936,912 | chr8:27936912:C:A | a | c | 0.8452 | 0.21 | 5.70E-05 | ++++++++? | 12.041 | 7 | 0.09923 |
| A Score | chr8 | 27,943,336 | chr8:27943336:C:T | t | c | 0.8493 | 0.2091 | 4.85E-05 | ++++++++? | 11.809 | 7 | 0.107 |
| A Score | chr8 | 27,948,548 | chr8:27948548:C:G | c | g | 0.8216 | 0.2061 | 6.73E-05 | ++++++++? | 12.249 | 7 | 0.09265 |
| A Score | chr8 | 27,949,643 | chr8:27949643:G:C | c | g | -0.8202 | 0.1966 | 3.02E-05 | --------? | 8.885 | 7 | 0.261 |
| A Score | chr8 | 27,950,976 | chr8:27950976:T:C | t | c | 0.8178 | 0.2015 | 4.93E-05 | ++++++++? | 11.579 | 7 | 0.1153 |
| A Score | chr8 | 27,969,969 | chr8:27969969:C:T | t | c | 0.8382 | 0.202 | 3.34E-05 | ++++++++? | 9.589 | 7 | 0.2131 |
| A Score | chr8 | 27,971,743 | chr8:27971743:T:C | t | c | -0.8382 | 0.2022 | 3.39E-05 | --------? | 9.79 | 7 | 0.2008 |
| A Score | chr8 | 27,973,938 | chr8:27973938:C:T | t | c | 0.81 | 0.2002 | 5.22E-05 | ++++-+++? | 9.628 | 7 | 0.2106 |
| A Score | chr8 | 39,809,022 | chr8:39809022:C:T | t | c | 0.6522 | 0.1646 | 7.39E-05 | +-++?++-+ | 5.683 | 7 | 0.5772 |
| A Score | chr8 | 62,979,755 | chr8:62979755:G:C | c | g | 1.5055 | 0.3617 | 3.16E-05 | ?++?+??+? | 2.081 | 3 | 0.5557 |
| A Score | chr8 | 63,136,903 | chr8:63136903:C:T | t | c | 1.2936 | 0.3278 | 7.95E-05 | ?++?+??++ | 6.039 | 4 | 0.1962 |
| A Score | chr8 | 63,153,040 | chr8:63153040:C:G | c | g | -1.3197 | 0.3224 | 4.25E-05 | ?--?-??-- | 6.031 | 4 | 0.1968 |

| | | | | | | | | | | | | |
|---|---|---|---|---|---|---|---|---|---|---|---|---|
| A Score | chr8 | 63,283,790 | chr8:63283790:A:G | a | g | -1.3971 | 0.3238 | 1.60E-05 | ?--?-??-- | 9.01 | 4 | 0.06084 |
| A Score | chr8 | 92,218,769 | chr8:92218769:C:A | a | c | 0.8079 | 0.1878 | 1.69E-05 | +-+++++++ | 10.371 | 8 | 0.2399 |
| A Score | chr8 | 97,421,113 | chr8:97421113:C:T | t | c | 0.9188 | 0.2244 | 4.23E-05 | +-++-++++ | 2.661 | 8 | 0.9538 |
| A Score | chr8 | 104,494,688 | chr8:104494688:T:C | t | c | -0.8816 | 0.2083 | 2.32E-05 | +-------- | 2.364 | 8 | 0.9678 |
| A Score | chr8 | 104,518,024 | chr8:104518024:A:T | a | t | -0.8808 | 0.2082 | 2.34E-05 | +-------- | 2.416 | 8 | 0.9655 |
| A Score | chr8 | 104,873,557 | chr8:104873557:C:G | c | g | -0.8146 | 0.2073 | 8.53E-05 | ?---?--+- | 9.665 | 6 | 0.1395 |
| A Score | chr8 | 104,909,332 | chr8:104909332:G:A | a | g | 0.7511 | 0.1873 | 6.08E-05 | ?+++++--+ | 10.158 | 7 | 0.1798 |
| A Score | chr8 | 105,209,318 | chr8:105209318:GA:G | g | ga | 0.5076 | 0.1283 | 7.56E-05 | -++++++++ | 6.399 | 8 | 0.6027 |
| A Score | chr8 | 114,673,963 | chr8:114673963:C:T | t | c | 0.9134 | 0.2139 | 1.95E-05 | ++++??+++ | 3.12 | 6 | 0.7937 |
| A Score | chr8 | 120,309,777 | chr8:120309777:A:G | a | g | -1.4306 | 0.3485 | 4.05E-05 | ?-?-??-?- | 2.227 | 3 | 0.5267 |
| A Score | chr8 | 123,596,186 | chr8:123596186:G:T | t | g | -0.2485 | 0.0619 | 5.97E-05 | -------++ | 7.823 | 8 | 0.451 |
| A Score | chr8 | 124,745,404 | chr8:124745404:A:G | a | g | -0.437 | 0.1001 | 1.27E-05 | -+----+-- | 6.067 | 8 | 0.6397 |
| A Score | chr8 | 124,755,740 | chr8:124755740:A:C | a | c | -0.6723 | 0.1434 | 2.75E-06 | -+------- | 5.84 | 8 | 0.6652 |
| A Score | chr8 | 135,559,415 | chr8:135559415:C:T | t | c | -0.2939 | 0.0747 | 8.41E-05 | ----+--+- | 6.548 | 8 | 0.5861 |
| A Score | chr8 | 135,563,789 | chr8:135563789:C:T | t | c | -0.2933 | 0.0745 | 8.25E-05 | ----+--+- | 6.987 | 8 | 0.538 |
| A Score | chr8 | 135,593,639 | chr8:135593639:G:A | a | g | -0.2941 | 0.0748 | 8.37E-05 | ----+--+- | 7.062 | 8 | 0.53 |
| A Score | chr8 | 135,605,456 | chr8:135605456:G:A | a | g | -0.2975 | 0.0748 | 6.93E-05 | ----+---- | 7.119 | 8 | 0.5238 |
| A Score | chr8 | 135,609,551 | chr8:135609551:T:A | a | t | -0.3181 | 0.0756 | 2.56E-05 | ----+---- | 6.835 | 8 | 0.5546 |
| A Score | chr8 | 135,626,589 | chr8:135626589:G:T | t | g | -0.3028 | 0.0749 | 5.24E-05 | ----+---- | 7.063 | 8 | 0.5298 |
| A Score | chr8 | 135,644,990 | chr8:135644990:C:T | t | c | -0.3054 | 0.0751 | 4.77E-05 | ----+---- | 6.97 | 8 | 0.5398 |
| A Score | chr8 | 135,648,887 | chr8:135648887:GA:G | g | ga | -0.2977 | 0.0751 | 7.32E-05 | ----+--+- | 6.686 | 8 | 0.5709 |
| A Score | chr8 | 135,651,227 | chr8:135651227:A:G | a | g | 0.2961 | 0.0749 | 7.72E-05 | ++++-++++ | 6.911 | 8 | 0.5463 |
| A Score | chr8 | 135,660,964 | chr8:135660964:G:C | c | g | 0.2753 | 0.0653 | 2.48E-05 | ++++-++++ | 5.008 | 8 | 0.7567 |
| A Score | chr8 | 135,661,604 | chr8:135661604:G:A | a | g | 0.2766 | 0.0652 | 2.22E-05 | ++++-++++ | 5.076 | 8 | 0.7494 |
| A Score | chr8 | 135,664,488 | chr8:135664488:G:A | a | g | -0.2757 | 0.0668 | 3.69E-05 | ----+---- | 6.972 | 8 | 0.5397 |
| A Score | chr8 | 135,665,648 | chr8:135665648:A:G | a | g | 0.2789 | 0.0673 | 3.38E-05 | ++++-++-+ | 7.384 | 8 | 0.4958 |
| A Score | chr8 | 135,666,718 | chr8:135666718:C:A | a | c | -0.2791 | 0.0673 | 3.39E-05 | ----+--+- | 7.205 | 8 | 0.5147 |
| A Score | chr8 | 140,307,779 | chr8:140307779:T:C | t | c | -0.2167 | 0.0551 | 8.41E-05 | ---++-++- | 21.853 | 8 | 0.005196 |
| A Score | chr8 | 140,308,161 | chr8:140308161:C:G | c | g | -0.2142 | 0.055 | 9.90E-05 | ---++-++- | 24.36 | 8 | 0.001994 |
| A Score | chr8 | 140,317,375 | chr8:140317375:T:C | t | c | -0.2201 | 0.0546 | 5.55E-05 | ---++-++- | 22.03 | 8 | 0.00486 |
| A Score | chr8 | 140,317,552 | chr8:140317552:A:T | a | t | -0.222 | 0.0551 | 5.66E-05 | ---++-++- | 21.708 | 8 | 0.005487 |
| A Score | chr8 | 140,330,261 | chr8:140330261:A:G | a | g | -0.225 | 0.0552 | 4.54E-05 | ---++-++- | 24.495 | 8 | 0.001892 |
| A Score | chr8 | 140,335,344 | chr8:140335344:G:A | a | g | 0.2243 | 0.0552 | 4.88E-05 | +++--+--+ | 22.97 | 8 | 0.003403 |
| A Score | chr8 | 140,337,560 | chr8:140337560:G:A | a | g | 0.2163 | 0.0553 | 9.27E-05 | +++--+--+ | 22.423 | 8 | 0.00419 |
| A Score | chr9 | 26,644,923 | chr9:26644923:G:A | a | g | 0.8768 | 0.2216 | 7.58E-05 | +-++?++?+ | 0.562 | 6 | 0.997 |
| A Score | chr9 | 74,399,959 | chr9:74399959:A:T | a | t | -0.9954 | 0.2388 | 3.08E-05 | -+---??-- | 3.547 | 6 | 0.7378 |
| A Score | chr9 | 75,074,878 | chr9:75074878:T:C | t | c | -0.8371 | 0.2025 | 3.58E-05 | ?---?---+ | 10.174 | 6 | 0.1175 |
| A Score | chr9 | 75,081,963 | chr9:75081963:C:T | t | c | 0.8367 | 0.2017 | 3.36E-05 | ?+++?+++- | 10.118 | 6 | 0.1198 |
| A Score | chr9 | 94,621,790 | chr9:94621790:C:T | t | c | 0.5482 | 0.1365 | 5.89E-05 | +-++-++++ | 9.256 | 8 | 0.3212 |
| A Score | chr9 | 105,116,147 | chr9:105116147:T:C | t | c | -0.2648 | 0.0671 | 8.01E-05 | --------- | 2.359 | 8 | 0.968 |
| A Score | chr9 | 135,112,415 | chr9:135112415:G:A | a | g | 0.433 | 0.1107 | 9.14E-05 | +-++-++++ | 2.25 | 8 | 0.9724 |
| AD Braak | chr1 | 6,384,349 | chr1:6384349:T:C | t | c | -0.5866 | 0.1448 | 5.09E-05 | ----???-?-?? | 13.801 | 8 | 0.08711 |
| AD Braak | chr1 | 7,188,154 | chr1:7188154:G:A | a | g | -0.1428 | 0.0365 | 9.17E-05 | -----------++ | 10.646 | 16 | 0.8308 |
| AD Braak | chr1 | 13,427,093 | chr1:13427093:C:T | t | c | -0.2073 | 0.0524 | 7.61E-05 | -+---+---++- | 7.318 | 16 | 0.9666 |
| AD Braak | chr1 | 30,684,235 | chr1:30684235:AGTTT:A | a | agttt | -0.3117 | 0.0762 | 4.34E-05 | ------+-+?-- | 7.752 | 15 | 0.9333 |
| AD Braak | chr1 | 32,024,252 | chr1:32024252:C:CA | ca | c | 0.4052 | 0.1026 | 7.83E-05 | +++++++-?? | 8.015 | 14 | 0.8885 |

| | | | | | | | | | | | | |
|---|---|---|---|---|---|---|---|---|---|---|---|---|
| AD Braak | chr1 | 37,161,356 | chr1:37161356:G:A | a | g | 0.9045 | 0.2248 | 5.74E-05 | ?+????-??? | 4.009 | 5 | 0.5482 |
| AD Braak | chr1 | 44,032,791 | chr1:44032791:G:C | c | g | 0.1915 | 0.0482 | 7.17E-05 | +++++++-+- | 13.359 | 16 | 0.6464 |
| AD Braak | chr1 | 54,811,673 | chr1:54811673:C:G | c | g | 0.2882 | 0.0726 | 7.24E-05 | ++-++++++- | 11.504 | 15 | 0.7161 |
| AD Braak | chr1 | 54,815,118 | chr1:54815118:G:A | a | g | -0.2723 | 0.0694 | 8.70E-05 | ----------?--+ | 8.114 | 15 | 0.9191 |
| AD Braak | chr1 | 54,892,224 | chr1:54892224:G:A | a | g | -0.2827 | 0.0687 | 3.87E-05 | ---+------?--- | 9.385 | 15 | 0.8565 |
| AD Braak | chr1 | 55,245,792 | chr1:55245792:A:G | a | g | 0.1886 | 0.0458 | 3.87E-05 | +-++++-++- | 20.162 | 16 | 0.213 |
| AD Braak | chr1 | 55,247,737 | chr1:55247737:A:C | a | c | 0.1825 | 0.0461 | 7.56E-05 | +-++++-++- | 20.641 | 16 | 0.1927 |
| AD Braak | chr1 | 55,247,998 | chr1:55247998:G:A | a | g | -0.1836 | 0.0461 | 6.91E-05 | -+----+--+--- | 20.576 | 16 | 0.1954 |
| AD Braak | chr1 | 55,248,274 | chr1:55248274:T:A | a | t | -0.1835 | 0.0461 | 6.97E-05 | -+----+--+--- | 20.559 | 16 | 0.1961 |
| AD Braak | chr1 | 59,275,243 | chr1:59275243:C:T | t | c | -0.1039 | 0.0257 | 5.18E-05 | ----------+--- | 5.795 | 16 | 0.9902 |
| AD Braak | chr1 | 65,797,571 | chr1:65797571:G:A | a | g | -0.8735 | 0.2209 | 7.69E-05 | ?-????-?--- | 2.738 | 6 | 0.841 |
| AD Braak | chr1 | 102,505,899 | chr1:102505899:G:T | t | g | 0.1643 | 0.0414 | 7.34E-05 | +++++++++ | 9.383 | 16 | 0.8968 |
| AD Braak | chr1 | 102,513,792 | chr1:102513792:T:C | t | c | -0.1624 | 0.0409 | 7.32E-05 | -----------+-+ | 9.439 | 16 | 0.8942 |
| AD Braak | chr1 | 102,537,886 | chr1:102537886:C:G | c | g | -0.1624 | 0.041 | 7.53E-05 | -----------+-+ | 9.412 | 16 | 0.8955 |
| AD Braak | chr1 | 102,546,410 | chr1:102546410:G:A | a | g | 0.1624 | 0.041 | 7.58E-05 | +++++++++ | 9.413 | 16 | 0.8954 |
| AD Braak | chr1 | 102,546,490 | chr1:102546490:G:T | t | g | 0.1624 | 0.041 | 7.49E-05 | +++++++++ | 9.41 | 16 | 0.8956 |
| AD Braak | chr1 | 102,552,971 | chr1:102552971:G:A | a | g | 0.1627 | 0.041 | 7.28E-05 | +++++++++ | 9.429 | 16 | 0.8947 |
| AD Braak | chr1 | 111,759,388 | chr1:111759388:G:GT | g | gt | -0.4571 | 0.112 | 4.45E-05 | ----?--------- | 6.787 | 14 | 0.9426 |
| AD Braak | chr1 | 147,838,192 | chr1:147838192:T:C | t | c | -0.2346 | 0.0602 | 9.73E-05 | -------+?-+- | 8.007 | 14 | 0.889 |
| AD Braak | chr1 | 147,838,341 | chr1:147838341:G:T | t | g | 0.2346 | 0.0602 | 9.79E-05 | +++++++-?- | 7.979 | 14 | 0.8904 |
| AD Braak | chr1 | 147,838,375 | chr1:147838375:G:GA | g | ga | -0.2361 | 0.0603 | 8.97E-05 | -------+?-+- | 7.995 | 14 | 0.8896 |
| AD Braak | chr1 | 147,839,430 | chr1:147839430:G:T | t | g | 0.2346 | 0.0603 | 9.97E-05 | +++++++-?- | 8.025 | 14 | 0.888 |
| AD Braak | chr1 | 147,839,592 | chr1:147839592:C:T | t | c | 0.2343 | 0.0602 | 9.93E-05 | +++++++-?- | 8.019 | 14 | 0.8883 |
| AD Braak | chr1 | 147,840,221 | chr1:147840221:C:T | t | c | 0.235 | 0.0603 | 9.64E-05 | +++++++-?- | 7.974 | 14 | 0.8907 |
| AD Braak | chr1 | 147,840,600 | chr1:147840600:GA:G | g | ga | 0.2385 | 0.0602 | 7.48E-05 | +++++++-?- | 8.363 | 14 | 0.8696 |
| AD Braak | chr1 | 147,840,825 | chr1:147840825:T:C | t | c | -0.2383 | 0.0602 | 7.62E-05 | -------+?-+- | 8.389 | 14 | 0.8681 |
| AD Braak | chr1 | 147,842,806 | chr1:147842806:T:G | t | g | -0.2139 | 0.0513 | 3.11E-05 | -------+?-+- | 9.951 | 14 | 0.7658 |
| AD Braak | chr1 | 147,844,318 | chr1:147844318:A:T | a | t | -0.2431 | 0.0575 | 2.31E-05 | -------+?---? | 7.863 | 14 | 0.8963 |
| AD Braak | chr1 | 147,844,695 | chr1:147844695:G:A | a | g | 0.2431 | 0.0574 | 2.32E-05 | +++++++-?- | 7.867 | 14 | 0.8961 |
| AD Braak | chr1 | 147,845,539 | chr1:147845539:C:T | t | c | 0.2456 | 0.0576 | 2.00E-05 | +++++++-?- | 7.862 | 14 | 0.8964 |
| AD Braak | chr1 | 148,158,667 | chr1:148158667:A:AG | a | ag | -0.2577 | 0.0662 | 9.88E-05 | -+------?---? | 9.63 | 14 | 0.7887 |
| AD Braak | chr1 | 148,295,055 | chr1:148295055:C:T | t | c | 0.2306 | 0.0516 | 7.82E-06 | +-+++++-?+ | 15.306 | 14 | 0.3576 |
| AD Braak | chr1 | 148,295,496 | chr1:148295496:C:T | t | c | 0.231 | 0.0514 | 6.97E-06 | +-+++++-?+ | 15.711 | 14 | 0.3313 |
| AD Braak | chr1 | 148,342,283 | chr1:148342283:C:T | t | c | 0.2128 | 0.0514 | 3.48E-05 | +++++++-?- | 15.688 | 14 | 0.3328 |
| AD Braak | chr1 | 148,344,090 | chr1:148344090:C:G | c | g | -0.2173 | 0.0519 | 2.79E-05 | -------+?---? | 16.502 | 14 | 0.2837 |
| AD Braak | chr1 | 148,344,146 | chr1:148344146:T:C | t | c | -0.2137 | 0.0515 | 3.32E-05 | -------+?---? | 15.632 | 14 | 0.3364 |
| AD Braak | chr1 | 148,347,597 | chr1:148347597:A:T | a | t | -0.2161 | 0.0515 | 2.70E-05 | -------+?---? | 15.802 | 14 | 0.3256 |
| AD Braak | chr1 | 148,347,726 | chr1:148347726:G:C | c | g | 0.2181 | 0.0516 | 2.39E-05 | +++++++-?- | 15.616 | 14 | 0.3374 |
| AD Braak | chr1 | 148,348,214 | chr1:148348214:A:G | a | g | -0.2135 | 0.0502 | 2.09E-05 | -------+?---? | 15.694 | 14 | 0.3324 |
| AD Braak | chr1 | 148,350,106 | chr1:148350106:A:G | a | g | -0.2169 | 0.0511 | 2.19E-05 | -------+?---? | 15.962 | 14 | 0.3157 |
| AD Braak | chr1 | 162,944,847 | chr1:162944847:C:T | t | c | 0.3134 | 0.0763 | 4.02E-05 | +-++-+-+++ | 17.49 | 16 | 0.3546 |
| AD Braak | chr1 | 165,680,478 | chr1:165680478:T:C | t | c | 0.2653 | 0.0661 | 6.00E-05 | +++++++++ | 14.181 | 16 | 0.5852 |
| AD Braak | chr1 | 217,400,281 | chr1:217400281:A:G | a | g | 0.1017 | 0.0252 | 5.29E-05 | +++++-+++- | 11.483 | 16 | 0.7787 |
| AD Braak | chr1 | 241,231,823 | chr1:241231823:A:C | a | c | 0.0977 | 0.025 | 8.98E-05 | ++++-+++-+ | 10.83 | 16 | 0.8199 |
| AD Braak | chr1 | 241,234,362 | chr1:241234362:G:A | a | g | -0.0996 | 0.0249 | 6.27E-05 | ----+---+---- | 10.825 | 16 | 0.8201 |
| AD Braak | chr1 | 248,911,119 | chr1:248911119:CT:C | ct | c | -1.3643 | 0.3464 | 8.20E-05 | -???--?-?-? | 4.451 | 5 | 0.4865 |

| | | | | | | | | | | | | |
|---|---|---|---|---|---|---|---|---|---|---|---|---|
| AD Braak | chr10 | 10,182,747 | chr10:10182747:G:A | a | g | 0.1332 | 0.0335 | 7.20E-05 | +++++-+++- | 12.661 | 16 | 0.6973 |
| AD Braak | chr10 | 10,188,006 | chr10:10188006:A:T | a | t | -0.1302 | 0.0335 | 9.99E-05 | -----+----+-- | 12.787 | 16 | 0.6882 |
| AD Braak | chr10 | 10,193,769 | chr10:10193769:T:C | t | c | -0.1359 | 0.0334 | 4.77E-05 | ----------+--- | 12.637 | 16 | 0.6991 |
| AD Braak | chr10 | 10,796,888 | chr10:10796888:A:T | a | t | -0.2305 | 0.0579 | 6.78E-05 | ------++----- | 21.386 | 16 | 0.1642 |
| AD Braak | chr10 | 11,676,140 | chr10:11676140:T:C | t | c | 0.1702 | 0.0436 | 9.48E-05 | +++++-+++- | 25.536 | 16 | 0.06092 |
| AD Braak | chr10 | 15,497,560 | chr10:15497560:C:T | t | c | 0.4018 | 0.1016 | 7.72E-05 | ++++++--+? | 10.329 | 15 | 0.7985 |
| AD Braak | chr10 | 17,784,969 | chr10:17784969:T:C | t | c | -0.1117 | 0.0284 | 8.30E-05 | ----+--+--?-- | 13.92 | 15 | 0.5316 |
| AD Braak | chr10 | 20,679,571 | chr10:20679571:C:T | t | c | 0.3922 | 0.0964 | 4.71E-05 | +-++-++-++ | 10.916 | 15 | 0.7585 |
| AD Braak | chr10 | 20,688,939 | chr10:20688939:G:A | a | g | 0.442 | 0.1012 | 1.25E-05 | +-++-++-++ | 7.908 | 14 | 0.8941 |
| AD Braak | chr10 | 20,713,515 | chr10:20713515:C:A | a | c | 0.412 | 0.1044 | 7.88E-05 | +-++-++++? | 6.852 | 13 | 0.9096 |
| AD Braak | chr10 | 29,709,493 | chr10:29709493:G:A | a | g | 0.3778 | 0.0966 | 9.22E-05 | +++-++-+++ | 17.281 | 15 | 0.3023 |
| AD Braak | chr10 | 72,610,333 | chr10:72610333:AT:A | a | at | 0.3931 | 0.089 | 9.95E-06 | ++++-++++- | 20.294 | 15 | 0.1609 |
| AD Braak | chr10 | 72,658,578 | chr10:72658578:C:T | t | c | 0.3782 | 0.0949 | 6.70E-05 | ++++-++++- | 14.708 | 15 | 0.4727 |
| AD Braak | chr10 | 72,833,818 | chr10:72833818:G:A | a | g | 0.3791 | 0.0947 | 6.30E-05 | ++++-++++- | 15.387 | 15 | 0.4239 |
| AD Braak | chr10 | 88,855,813 | chr10:88855813:G:C | c | g | 0.1154 | 0.0285 | 5.31E-05 | ++++-++-+- | 26.163 | 16 | 0.05177 |
| AD Braak | chr10 | 107,075,446 | chr10:107075446:A:T | a | t | -0.0946 | 0.0243 | 9.73E-05 | -+--+++---+- | 15.862 | 16 | 0.4626 |
| AD Braak | chr10 | 107,085,258 | chr10:107085258:C:G | c | g | -0.0949 | 0.0243 | 9.36E-05 | -+--+++--++ | 17.998 | 16 | 0.324 |
| AD Braak | chr10 | 107,086,132 | chr10:107086132:C:G | c | g | -0.0946 | 0.0243 | 9.91E-05 | -+--+++---+- | 17.46 | 16 | 0.3564 |
| AD Braak | chr10 | 119,365,091 | chr10:119365091:G:A | a | g | 0.2552 | 0.0638 | 6.39E-05 | ++++++-++- | 22.973 | 16 | 0.1144 |
| AD Braak | chr10 | 119,691,120 | chr10:119691120:G:C | c | g | 0.1152 | 0.0275 | 2.84E-05 | ++++-++++- | 18.642 | 16 | 0.2877 |
| AD Braak | chr10 | 119,691,613 | chr10:119691613:CGTG:C | cgtg | c | -0.1192 | 0.0275 | 1.44E-05 | ----+----+--- | 18.229 | 16 | 0.3107 |
| AD Braak | chr10 | 119,693,684 | chr10:119693684:C:G | c | g | -0.1175 | 0.0276 | 2.01E-05 | ----+----++-- | 18.92 | 16 | 0.2728 |
| AD Braak | chr10 | 130,146,988 | chr10:130146988:T:C | t | c | 0.0949 | 0.0242 | 8.99E-05 | ++++--+++- | 18.164 | 16 | 0.3144 |
| AD Braak | chr10 | 130,186,291 | chr10:130186291:G:A | a | g | 0.099 | 0.0251 | 8.28E-05 | +-++--+++-- | 33.23 | 16 | 0.006889 |
| AD Braak | chr10 | 132,049,587 | chr10:132049587:T:C | t | c | -0.1114 | 0.0282 | 7.82E-05 | --------+----+ | 17.085 | 16 | 0.3801 |
| AD Braak | chr11 | 1,848,180 | chr11:1848180:T:A | a | t | -0.119 | 0.0285 | 2.94E-05 | ----+-----?-+ | 20.296 | 15 | 0.1609 |
| AD Braak | chr11 | 24,747,359 | chr11:24747359:A:G | a | g | -0.1185 | 0.0304 | 9.72E-05 | -------+-++-- | 15.471 | 16 | 0.4904 |
| AD Braak | chr11 | 47,227,672 | chr11:47227672:G:A | a | g | -0.1044 | 0.0247 | 2.37E-05 | -+--+-------+ | 13.222 | 16 | 0.6565 |
| AD Braak | chr11 | 47,228,768 | chr11:47228768:C:T | t | c | -0.1043 | 0.0247 | 2.42E-05 | -+--+-------+ | 13.134 | 16 | 0.6629 |
| AD Braak | chr11 | 47,239,709 | chr11:47239709:G:A | a | g | -0.0998 | 0.0244 | 4.33E-05 | -+--+-------+ | 15.887 | 16 | 0.4609 |
| AD Braak | chr11 | 47,241,289 | chr11:47241289:G:A | a | g | -0.1001 | 0.0245 | 4.30E-05 | -+--+-------+ | 15.824 | 16 | 0.4653 |
| AD Braak | chr11 | 47,255,124 | chr11:47255124:A:G | a | g | 0.1031 | 0.0264 | 9.19E-05 | +-++-+++++ | 19.166 | 16 | 0.2602 |
| AD Braak | chr11 | 47,256,951 | chr11:47256951:C:T | t | c | -0.1047 | 0.0264 | 7.14E-05 | -+--+-------- | 18.902 | 16 | 0.2738 |
| AD Braak | chr11 | 47,272,470 | chr11:47272470:G:T | t | g | -0.0954 | 0.0245 | 9.76E-05 | -+--+-------+ | 15.484 | 16 | 0.4895 |
| AD Braak | chr11 | 47,281,182 | chr11:47281182:C:T | t | c | -0.0954 | 0.0245 | 9.72E-05 | -+--+-------+ | 15.568 | 16 | 0.4835 |
| AD Braak | chr11 | 47,296,606 | chr11:47296606:T:C | t | c | 0.0956 | 0.0244 | 9.02E-05 | +-++-+++++ | 15.896 | 16 | 0.4602 |
| AD Braak | chr11 | 47,314,769 | chr11:47314769:A:T | a | t | 0.1028 | 0.0264 | 9.73E-05 | +-++-+++++ | 19.733 | 16 | 0.2325 |
| AD Braak | chr11 | 47,325,389 | chr11:47325389:G:A | a | g | -0.0976 | 0.0248 | 8.48E-05 | -+--+-------+ | 13.077 | 16 | 0.6671 |
| AD Braak | chr11 | 47,339,533 | chr11:47339533:T:C | t | c | -0.1014 | 0.0249 | 4.52E-05 | -+--+-------+ | 11.203 | 16 | 0.7968 |
| AD Braak | chr11 | 47,340,287 | chr11:47340287:A:G | a | g | -0.1038 | 0.0249 | 2.95E-05 | -+--+-------+ | 11.304 | 16 | 0.7903 |
| AD Braak | chr11 | 47,341,664 | chr11:47341664:G:T | t | g | 0.104 | 0.0249 | 2.92E-05 | +-++-+++++ | 12.071 | 16 | 0.7391 |
| AD Braak | chr11 | 47,341,734 | chr11:47341734:G:A | a | g | -0.1052 | 0.0248 | 2.31E-05 | -+--+-------+ | 13.012 | 16 | 0.6718 |
| AD Braak | chr11 | 47,355,732 | chr11:47355732:A:G | a | g | 0.1185 | 0.0248 | 1.76E-06 | +-+--+++++ | 17.759 | 16 | 0.3382 |
| AD Braak | chr11 | 47,356,694 | chr11:47356694:T:C | t | c | -0.1168 | 0.0248 | 2.55E-06 | -+-++-------- | 17.925 | 16 | 0.3283 |
| AD Braak | chr11 | 47,356,845 | chr11:47356845:C:T | t | c | -0.1 | 0.0242 | 3.71E-05 | -+-++--+---- | 16.995 | 16 | 0.3859 |
| AD Braak | chr11 | 47,366,663 | chr11:47366663:G:A | a | g | -0.1085 | 0.0248 | 1.19E-05 | -+-++--+---- | 17.225 | 16 | 0.3712 |

| | | | | | | | | | | | | |
|---|---|---|---|---|---|---|---|---|---|---|---|---|
| AD Braak | chr11 | 47,368,087 | chr11:47368087:C:T | t | c | -0.1068 | 0.0247 | 1.56E-05 | -+-++--+---- | 15.939 | 16 | 0.4573 |
| AD Braak | chr11 | 47,370,397 | chr11:47370397:G:A | a | g | -0.109 | 0.0244 | 8.11E-06 | -+-++--+---- | 18.288 | 16 | 0.3073 |
| AD Braak | chr11 | 47,372,754 | chr11:47372754:C:T | t | c | -0.1088 | 0.0244 | 8.16E-06 | -+-++--+---- | 18.96 | 16 | 0.2708 |
| AD Braak | chr11 | 47,378,051 | chr11:47378051:G:A | a | g | -0.1067 | 0.0247 | 1.56E-05 | -+-++--+---- | 16.815 | 16 | 0.3976 |
| AD Braak | chr11 | 47,379,051 | chr11:47379051:G:A | a | g | -0.1067 | 0.0247 | 1.55E-05 | -+-++--+---- | 17.163 | 16 | 0.3751 |
| AD Braak | chr11 | 47,385,888 | chr11:47385888:C:A | a | c | -0.1042 | 0.0247 | 2.40E-05 | -+-++--+---- | 17.947 | 16 | 0.327 |
| AD Braak | chr11 | 47,386,310 | chr11:47386310:CT:C | ct | c | 0.1087 | 0.0244 | 8.29E-06 | +-+--++-++- | 19.147 | 16 | 0.2611 |
| AD Braak | chr11 | 47,387,041 | chr11:47387041:G:A | a | g | -0.105 | 0.0247 | 2.11E-05 | -+-++--+---- | 17.461 | 16 | 0.3564 |
| AD Braak | chr11 | 47,387,500 | chr11:47387500:T:A | a | t | -0.1045 | 0.0248 | 2.56E-05 | -+-+---+----- | 19.805 | 16 | 0.2291 |
| AD Braak | chr11 | 47,389,337 | chr11:47389337:G:T | t | g | -0.1087 | 0.0244 | 8.24E-06 | -+-++--+---- | 18.874 | 16 | 0.2753 |
| AD Braak | chr11 | 47,395,195 | chr11:47395195:C:T | t | c | -0.1104 | 0.0244 | 5.82E-06 | -+--+--+----- | 17.619 | 16 | 0.3467 |
| AD Braak | chr11 | 47,395,632 | chr11:47395632:A:C | a | c | 0.1095 | 0.0244 | 7.12E-06 | +-+--++-++- | 18.248 | 16 | 0.3096 |
| AD Braak | chr11 | 47,397,402 | chr11:47397402:C:G | c | g | 0.1062 | 0.0244 | 1.37E-05 | +-+--++-++- | 18.504 | 16 | 0.2952 |
| AD Braak | chr11 | 47,398,112 | chr11:47398112:G:T | t | g | -0.1064 | 0.0244 | 1.30E-05 | -+-++--+---- | 18.153 | 16 | 0.315 |
| AD Braak | chr11 | 47,401,789 | chr11:47401789:C:T | t | c | -0.1074 | 0.0244 | 1.07E-05 | -+-++--+---- | 17.685 | 16 | 0.3427 |
| AD Braak | chr11 | 47,403,380 | chr11:47403380:G:A | a | g | -0.1073 | 0.0244 | 1.10E-05 | -+-++--+---- | 17.816 | 16 | 0.3348 |
| AD Braak | chr11 | 47,406,116 | chr11:47406116:T:C | t | c | 0.1062 | 0.0244 | 1.36E-05 | +-+--++-++- | 17.425 | 16 | 0.3586 |
| AD Braak | chr11 | 47,406,188 | chr11:47406188:C:T | t | c | -0.1057 | 0.0244 | 1.47E-05 | -+-++--+---- | 17.686 | 16 | 0.3426 |
| AD Braak | chr11 | 47,408,353 | chr11:47408353:G:A | a | g | -0.106 | 0.0244 | 1.39E-05 | -+-++--+---- | 17.508 | 16 | 0.3535 |
| AD Braak | chr11 | 47,409,048 | chr11:47409048:G:A | a | g | -0.1059 | 0.0244 | 1.43E-05 | -+-++--+---- | 17.517 | 16 | 0.3529 |
| AD Braak | chr11 | 47,411,174 | chr11:47411174:T:G | t | g | 0.1056 | 0.0244 | 1.50E-05 | +-+--++-++- | 17.566 | 16 | 0.3499 |
| AD Braak | chr11 | 47,412,670 | chr11:47412670:C:G | c | g | 0.1065 | 0.0244 | 1.27E-05 | +-+--++-++- | 17.549 | 16 | 0.351 |
| AD Braak | chr11 | 47,413,435 | chr11:47413435:G:A | a | g | -0.0999 | 0.0251 | 6.74E-05 | -+--+-------- | 17.084 | 16 | 0.3802 |
| AD Braak | chr11 | 47,419,207 | chr11:47419207:G:A | a | g | -0.1079 | 0.0244 | 9.73E-06 | -+-++--+---- | 17.757 | 16 | 0.3383 |
| AD Braak | chr11 | 47,425,795 | chr11:47425795:C:T | t | c | -0.106 | 0.0244 | 1.38E-05 | -+-++--+---- | 18.001 | 16 | 0.3238 |
| AD Braak | chr11 | 47,427,521 | chr11:47427521:G:A | a | g | -0.1058 | 0.0243 | 1.36E-05 | -+-++--+---- | 17.889 | 16 | 0.3304 |
| AD Braak | chr11 | 47,430,474 | chr11:47430474:AC:A | a | ac | -0.102 | 0.0251 | 4.96E-05 | -+--+-------- | 17.981 | 16 | 0.325 |
| AD Braak | chr11 | 47,433,000 | chr11:47433000:A:T | a | t | 0.1069 | 0.0244 | 1.18E-05 | +-+--++-++- | 17.767 | 16 | 0.3377 |
| AD Braak | chr11 | 47,440,232 | chr11:47440232:A:G | a | g | 0.1069 | 0.0244 | 1.19E-05 | +-+--++-++- | 17.937 | 16 | 0.3276 |
| AD Braak | chr11 | 47,440,589 | chr11:47440589:A:G | a | g | 0.1069 | 0.0244 | 1.19E-05 | +-+--++-++- | 17.909 | 16 | 0.3292 |
| AD Braak | chr11 | 47,444,890 | chr11:47444890:G:A | a | g | -0.1007 | 0.0252 | 6.33E-05 | -+--+-------- | 17.85 | 16 | 0.3327 |
| AD Braak | chr11 | 47,445,620 | chr11:47445620:A:T | a | t | 0.1021 | 0.0252 | 5.24E-05 | +-+--+++++ | 17.812 | 16 | 0.335 |
| AD Braak | chr11 | 47,678,616 | chr11:47678616:G:T | t | g | -0.102 | 0.0261 | 9.34E-05 | -+-++------- | 19.099 | 16 | 0.2636 |
| AD Braak | chr11 | 67,732,707 | chr11:67732707:A:T | a | t | -0.1221 | 0.0297 | 4.01E-05 | --------++--- | 33.137 | 16 | 0.007087 |
| AD Braak | chr11 | 67,733,291 | chr11:67733291:T:C | t | c | -0.122 | 0.0297 | 4.05E-05 | --------++--+ | 33.516 | 16 | 0.006309 |
| AD Braak | chr11 | 81,341,051 | chr11:81341051:C:G | c | g | 0.1524 | 0.038 | 6.01E-05 | ++-+-++-++ | 21.218 | 16 | 0.1703 |
| AD Braak | chr11 | 85,941,208 | chr11:85941208:C:T | t | c | 0.1377 | 0.0338 | 4.67E-05 | +++++-+-++ | 21.093 | 16 | 0.175 |
| AD Braak | chr11 | 85,942,959 | chr11:85942959:C:CA | ca | c | -0.1196 | 0.0292 | 4.13E-05 | -------+-----+ | 22.461 | 16 | 0.1289 |
| AD Braak | chr11 | 85,944,062 | chr11:85944062:A:C | a | c | 0.126 | 0.0303 | 3.14E-05 | +++++++-+- | 22.254 | 16 | 0.1352 |
| AD Braak | chr11 | 85,948,804 | chr11:85948804:C:T | t | c | -0.126 | 0.0303 | 3.13E-05 | -------+-----+ | 23.08 | 16 | 0.1116 |
| AD Braak | chr11 | 85,953,861 | chr11:85953861:G:C | c | g | -0.1204 | 0.0302 | 6.56E-05 | -+-----+----- | 23.764 | 16 | 0.0948 |
| AD Braak | chr11 | 85,955,095 | chr11:85955095:T:C | t | c | 0.124 | 0.0303 | 4.20E-05 | +-+++++-++ | 23.262 | 16 | 0.1069 |
| AD Braak | chr11 | 85,955,397 | chr11:85955397:A:C | a | c | 0.1245 | 0.0303 | 3.88E-05 | +-+++++-++ | 23.314 | 16 | 0.1056 |
| AD Braak | chr11 | 85,964,490 | chr11:85964490:C:T | t | c | -0.1232 | 0.0304 | 4.92E-05 | -+-----+----- | 22.649 | 16 | 0.1234 |
| AD Braak | chr11 | 85,988,957 | chr11:85988957:C:A | a | c | -0.1197 | 0.0304 | 8.14E-05 | -+-----+----- | 21.227 | 16 | 0.17 |
| AD Braak | chr11 | 85,990,579 | chr11:85990579:T:C | t | c | 0.1232 | 0.0303 | 4.71E-05 | +-+++++-++ | 23.603 | 16 | 0.09853 |

| | | | | | | | | | | | | |
|---|---|---|---|---|---|---|---|---|---|---|---|---|
| AD Braak | chr11 | 85,990,698 | chr11:85990698:T:C | t | c | 0.1235 | 0.0303 | 4.59E-05 | +-+++++-++ | 22.46 | 16 | 0.1289 |
| AD Braak | chr11 | 85,996,185 | chr11:85996185:A:G | a | g | 0.1224 | 0.0302 | 4.94E-05 | +-+++++-++ | 23.568 | 16 | 0.09936 |
| AD Braak | chr11 | 85,998,305 | chr11:85998305:C:G | c | g | 0.123 | 0.0304 | 5.18E-05 | +-+++++-++ | 21.117 | 16 | 0.1741 |
| AD Braak | chr11 | 85,998,516 | chr11:85998516:G:A | a | g | -0.1195 | 0.0303 | 7.98E-05 | -+-----+----- | 21.687 | 16 | 0.1536 |
| AD Braak | chr11 | 85,999,126 | chr11:85999126:G:A | a | g | -0.1186 | 0.0303 | 8.95E-05 | -+-----+----- | 21.755 | 16 | 0.1513 |
| AD Braak | chr11 | 85,999,137 | chr11:85999137:G:A | a | g | -0.1223 | 0.0304 | 5.65E-05 | -+-----+----- | 21.107 | 16 | 0.1744 |
| AD Braak | chr11 | 86,002,522 | chr11:86002522:A:C | a | c | 0.1204 | 0.0303 | 7.07E-05 | +-+++++-++ | 24.015 | 16 | 0.08918 |
| AD Braak | chr11 | 86,032,407 | chr11:86032407:A:G | a | g | 0.1231 | 0.0304 | 5.27E-05 | +-+++++-++ | 23.895 | 16 | 0.09183 |
| AD Braak | chr11 | 86,044,583 | chr11:86044583:C:CAT | cat | c | -0.1237 | 0.0303 | 4.46E-05 | -+-----+----- | 22.916 | 16 | 0.116 |
| AD Braak | chr11 | 86,053,833 | chr11:86053833:T:G | t | g | 0.1234 | 0.0301 | 4.15E-05 | +-+++++-++ | 24.2 | 16 | 0.08522 |
| AD Braak | chr11 | 86,054,695 | chr11:86054695:T:C | t | c | 0.1261 | 0.0303 | 3.08E-05 | +-+++++-++ | 23.758 | 16 | 0.09493 |
| AD Braak | chr11 | 86,055,502 | chr11:86055502:A:ATGTC | a | atgtc | 0.1246 | 0.0302 | 3.79E-05 | +-+++++-++ | 23.664 | 16 | 0.0971 |
| AD Braak | chr11 | 86,055,540 | chr11:86055540:A:G | a | g | 0.1237 | 0.0302 | 4.14E-05 | +-+++++-++ | 24.464 | 16 | 0.07985 |
| AD Braak | chr11 | 86,060,023 | chr11:86060023:T:C | t | c | 0.1225 | 0.0302 | 4.94E-05 | +-+++++-++ | 23.944 | 16 | 0.09073 |
| AD Braak | chr11 | 86,060,128 | chr11:86060128:T:C | t | c | 0.1265 | 0.0303 | 2.94E-05 | +-+++++-++ | 23.517 | 16 | 0.1006 |
| AD Braak | chr11 | 86,067,882 | chr11:86067882:C:T | t | c | -0.1218 | 0.0301 | 5.28E-05 | -+-----+----- | 25.057 | 16 | 0.06884 |
| AD Braak | chr11 | 86,068,120 | chr11:86068120:C:G | c | g | 0.1256 | 0.0303 | 3.32E-05 | +-+++++-++ | 25.066 | 16 | 0.06866 |
| AD Braak | chr11 | 86,068,255 | chr11:86068255:T:G | t | g | 0.1248 | 0.0303 | 3.78E-05 | +-+++++-++ | 24.622 | 16 | 0.07678 |
| AD Braak | chr11 | 86,068,268 | chr11:86068268:A:G | a | g | 0.1251 | 0.0303 | 3.54E-05 | +-+++++-++ | 24.62 | 16 | 0.07681 |
| AD Braak | chr11 | 86,069,406 | chr11:86069406:T:C | t | c | 0.1241 | 0.0304 | 4.37E-05 | +-+++++-++ | 24.503 | 16 | 0.07908 |
| AD Braak | chr11 | 86,070,280 | chr11:86070280:C:T | t | c | -0.1197 | 0.0302 | 7.35E-05 | -+-----+----- | 24.186 | 16 | 0.08553 |
| AD Braak | chr11 | 86,070,555 | chr11:86070555:CA:C | ca | c | 0.119 | 0.0303 | 8.59E-05 | +-+++++-++ | 23.168 | 16 | 0.1093 |
| AD Braak | chr11 | 86,072,833 | chr11:86072833:A:G | a | g | 0.1276 | 0.0305 | 2.80E-05 | +-+++++-++ | 22.907 | 16 | 0.1162 |
| AD Braak | chr11 | 86,145,145 | chr11:86145145:C:G | c | g | 0.1036 | 0.0262 | 7.87E-05 | +++++++-+- | 17.281 | 16 | 0.3676 |
| AD Braak | chr11 | 86,147,496 | chr11:86147496:G:A | a | g | -0.1025 | 0.0262 | 9.11E-05 | -------+--+-- | 17.651 | 16 | 0.3447 |
| AD Braak | chr11 | 96,664,644 | chr11:96664644:A:AG | a | ag | -0.1687 | 0.0405 | 3.14E-05 | -+--+-------+ | 18.892 | 16 | 0.2743 |
| AD Braak | chr11 | 111,968,253 | chr11:111968253:T:C | t | c | 0.3906 | 0.0989 | 7.79E-05 | ++++-+++-+ | 16.073 | 16 | 0.4479 |
| AD Braak | chr11 | 114,345,216 | chr11:114345216:G:A | a | g | 0.4027 | 0.1024 | 8.38E-05 | ++++-++++? | 5.979 | 14 | 0.967 |
| AD Braak | chr12 | 1,535,561 | chr12:1535561:G:A | a | g | -0.2368 | 0.0589 | 5.76E-05 | ---------+--+ | 17.067 | 16 | 0.3813 |
| AD Braak | chr12 | 33,086,326 | chr12:33086326:G:A | a | g | 0.3462 | 0.0872 | 7.11E-05 | ++++-+--++ | 11.022 | 16 | 0.8081 |
| AD Braak | chr12 | 33,090,922 | chr12:33090922:A:G | a | g | -0.3378 | 0.0858 | 8.28E-05 | ----+-++--+- | 13.043 | 16 | 0.6696 |
| AD Braak | chr12 | 41,300,860 | chr12:41300860:T:TA | t | ta | -0.3745 | 0.0901 | 3.25E-05 | ------+---?-- | 22.356 | 15 | 0.09879 |
| AD Braak | chr12 | 72,419,173 | chr12:72419173:G:A | a | g | -0.2045 | 0.0516 | 7.29E-05 | ---------++-- | 14.668 | 16 | 0.549 |
| AD Braak | chr12 | 72,423,996 | chr12:72423996:C:T | t | c | -0.2071 | 0.0515 | 5.78E-05 | ---------++-- | 14.918 | 16 | 0.5307 |
| AD Braak | chr12 | 72,428,705 | chr12:72428705:G:A | a | g | -0.2 | 0.0513 | 9.64E-05 | ---------++-- | 14.669 | 16 | 0.549 |
| AD Braak | chr12 | 72,430,414 | chr12:72430414:C:T | t | c | -0.2054 | 0.0513 | 6.20E-05 | ---------++-- | 15.592 | 16 | 0.4818 |
| AD Braak | chr12 | 72,437,294 | chr12:72437294:G:T | t | g | -0.202 | 0.0514 | 8.55E-05 | ---------++-- | 14.769 | 16 | 0.5416 |
| AD Braak | chr12 | 72,437,397 | chr12:72437397:G:A | a | g | -0.2008 | 0.0514 | 9.20E-05 | ---------++-- | 14.768 | 16 | 0.5417 |
| AD Braak | chr12 | 72,454,926 | chr12:72454926:C:A | a | c | -0.2011 | 0.0512 | 8.46E-05 | ---------++-- | 14.801 | 16 | 0.5393 |
| AD Braak | chr12 | 72,455,251 | chr12:72455251:C:A | a | c | -0.2003 | 0.0512 | 9.26E-05 | ---------++-- | 15.052 | 16 | 0.5208 |
| AD Braak | chr12 | 72,460,122 | chr12:72460122:G:A | a | g | -0.2019 | 0.0513 | 8.40E-05 | ---------++-- | 14.848 | 16 | 0.5358 |
| AD Braak | chr12 | 72,464,190 | chr12:72464190:C:A | a | c | -0.2033 | 0.0514 | 7.66E-05 | ---------++-- | 15.046 | 16 | 0.5213 |
| AD Braak | chr12 | 72,470,927 | chr12:72470927:A:T | a | t | 0.2056 | 0.0518 | 7.21E-05 | +++++++++ | 16.009 | 16 | 0.4524 |
| AD Braak | chr12 | 72,471,004 | chr12:72471004:G:A | a | g | -0.2037 | 0.0517 | 8.05E-05 | ---------++-- | 16.318 | 16 | 0.431 |
| AD Braak | chr12 | 78,987,606 | chr12:78987606:G:A | a | g | -0.57 | 0.144 | 7.56E-05 | ----??-+-??- | 11.934 | 9 | 0.217 |
| AD Braak | chr12 | 91,495,461 | chr12:91495461:T:C | t | c | -0.1023 | 0.0246 | 3.26E-05 | ----------+--+ | 12.856 | 16 | 0.6833 |

| | | | | | | | | | | | | |
|---|---|---|---|---|---|---|---|---|---|---|---|---|
| AD Braak | chr12 | 96,079,327 | chr12:96079327:T:TC | t | tc | -0.131 | 0.0332 | 8.01E-05 | ----+-------+ | 11.529 | 16 | 0.7757 |
| AD Braak | chr12 | 119,784,403 | chr12:119784403:C:G | c | g | 0.2496 | 0.0629 | 7.28E-05 | -++++++++- | 12.105 | 16 | 0.7367 |
| AD Braak | chr12 | 132,513,044 | chr12:132513044:G:A | a | g | 0.2218 | 0.0531 | 2.92E-05 | ++++-+-++- | 21.362 | 16 | 0.165 |
| AD Braak | chr13 | 22,771,894 | chr13:22771894:T:C | t | c | -0.3497 | 0.0897 | 9.66E-05 | +------+--?-- | 15.566 | 15 | 0.4115 |
| AD Braak | chr13 | 30,365,688 | chr13:30365688:A:C | a | c | -0.4838 | 0.1175 | 3.84E-05 | ---+?--+----- | 20.521 | 12 | 0.05785 |
| AD Braak | chr13 | 40,264,456 | chr13:40264456:G:T | t | g | 0.1102 | 0.0273 | 5.57E-05 | +-+-++++++ | 12.151 | 16 | 0.7335 |
| AD Braak | chr13 | 47,722,423 | chr13:47722423:G:A | a | g | 0.208 | 0.0533 | 9.52E-05 | ++-++++-++ | 19.756 | 16 | 0.2314 |
| AD Braak | chr13 | 47,725,917 | chr13:47725917:G:A | a | g | 0.2109 | 0.0534 | 7.74E-05 | ++-++++-++ | 19.295 | 16 | 0.2537 |
| AD Braak | chr13 | 47,727,634 | chr13:47727634:T:C | t | c | -0.2119 | 0.0533 | 7.01E-05 | --+----+----- | 20.084 | 16 | 0.2164 |
| AD Braak | chr13 | 47,733,383 | chr13:47733383:A:C | a | c | -0.2148 | 0.054 | 7.04E-05 | --+---------- | 19.403 | 16 | 0.2483 |
| AD Braak | chr13 | 47,753,020 | chr13:47753020:T:C | t | c | -0.21 | 0.0535 | 8.79E-05 | --+----+----- | 20.023 | 16 | 0.2192 |
| AD Braak | chr13 | 47,772,776 | chr13:47772776:T:C | t | c | -0.2086 | 0.0531 | 8.63E-05 | --+-----+---- | 21.059 | 16 | 0.1763 |
| AD Braak | chr13 | 47,790,250 | chr13:47790250:T:C | t | c | -0.208 | 0.0531 | 9.00E-05 | --++----+---- | 19.967 | 16 | 0.2217 |
| AD Braak | chr13 | 47,792,146 | chr13:47792146:C:G | c | g | -0.2076 | 0.0531 | 9.28E-05 | --++----+---- | 20.019 | 16 | 0.2194 |
| AD Braak | chr13 | 47,800,100 | chr13:47800100:G:A | a | g | 0.206 | 0.0526 | 9.14E-05 | ++-++++++- | 18.008 | 16 | 0.3234 |
| AD Braak | chr13 | 47,802,440 | chr13:47802440:T:C | t | c | -0.2048 | 0.0524 | 9.45E-05 | --+---------- | 17.901 | 16 | 0.3297 |
| AD Braak | chr13 | 61,497,971 | chr13:61497971:C:A | a | c | -0.1308 | 0.0325 | 5.82E-05 | ---+--------- | 9.7 | 16 | 0.8818 |
| AD Braak | chr13 | 61,498,826 | chr13:61498826:T:A | a | t | -0.1316 | 0.0325 | 5.03E-05 | ---+--------- | 9.916 | 16 | 0.871 |
| AD Braak | chr13 | 61,499,898 | chr13:61499898:C:T | t | c | -0.1307 | 0.0325 | 5.73E-05 | ---+--------- | 9.877 | 16 | 0.873 |
| AD Braak | chr13 | 61,499,945 | chr13:61499945:C:T | t | c | -0.1305 | 0.0324 | 5.72E-05 | ---+--------- | 9.87 | 16 | 0.8733 |
| AD Braak | chr13 | 61,500,028 | chr13:61500028:T:A | a | t | -0.1332 | 0.0325 | 4.07E-05 | ---+--------- | 9.526 | 16 | 0.8902 |
| AD Braak | chr13 | 61,500,173 | chr13:61500173:C:A | a | c | -0.1302 | 0.0324 | 5.78E-05 | ---+--------- | 10.049 | 16 | 0.8641 |
| AD Braak | chr13 | 61,500,907 | chr13:61500907:T:C | t | c | 0.1326 | 0.0323 | 3.95E-05 | +++-+++++- | 9.822 | 16 | 0.8758 |
| AD Braak | chr13 | 61,501,159 | chr13:61501159:CT:C | ct | c | 0.1336 | 0.0323 | 3.46E-05 | +++-+++++- | 9.806 | 16 | 0.8766 |
| AD Braak | chr13 | 61,501,386 | chr13:61501386:T:C | t | c | 0.125 | 0.032 | 9.28E-05 | +++-+++++- | 9.116 | 16 | 0.9085 |
| AD Braak | chr13 | 61,501,820 | chr13:61501820:GT:G | g | gt | -0.1338 | 0.0323 | 3.41E-05 | ---+--------- | 9.771 | 16 | 0.8783 |
| AD Braak | chr13 | 61,502,012 | chr13:61502012:A:T | a | t | 0.133 | 0.0323 | 3.76E-05 | +++-+++++- | 9.983 | 16 | 0.8675 |
| AD Braak | chr13 | 61,502,427 | chr13:61502427:A:G | a | g | 0.1326 | 0.0323 | 3.93E-05 | +++-+++++- | 9.746 | 16 | 0.8795 |
| AD Braak | chr13 | 61,502,570 | chr13:61502570:T:C | t | c | 0.1303 | 0.0323 | 5.39E-05 | +++-+++++- | 9.871 | 16 | 0.8733 |
| AD Braak | chr13 | 61,502,657 | chr13:61502657:G:T | t | g | -0.131 | 0.0322 | 4.79E-05 | ---+--------- | 9.497 | 16 | 0.8915 |
| AD Braak | chr13 | 61,503,293 | chr13:61503293:T:A | a | t | -0.1293 | 0.0323 | 6.21E-05 | ---+--------- | 9.389 | 16 | 0.8965 |
| AD Braak | chr13 | 61,503,416 | chr13:61503416:C:T | t | c | -0.1312 | 0.0322 | 4.75E-05 | ---+--------- | 9.8 | 16 | 0.8769 |
| AD Braak | chr13 | 61,503,452 | chr13:61503452:G:C | c | g | -0.1309 | 0.0322 | 4.93E-05 | ---+--------- | 9.579 | 16 | 0.8877 |
| AD Braak | chr13 | 61,503,732 | chr13:61503732:C:T | t | c | -0.1325 | 0.0323 | 4.03E-05 | ---+--------- | 9.822 | 16 | 0.8757 |
| AD Braak | chr13 | 61,503,762 | chr13:61503762:A:T | a | t | 0.1308 | 0.0322 | 4.95E-05 | +++-+++++- | 9.579 | 16 | 0.8877 |
| AD Braak | chr13 | 61,503,773 | chr13:61503773:A:ATGG | a | atgg | 0.1325 | 0.0323 | 4.06E-05 | +++-+++++- | 9.689 | 16 | 0.8824 |
| AD Braak | chr13 | 61,504,156 | chr13:61504156:C:CAT | cat | c | -0.1313 | 0.0324 | 4.99E-05 | ---+--------- | 9.736 | 16 | 0.88 |
| AD Braak | chr13 | 61,504,307 | chr13:61504307:G:C | c | g | -0.1299 | 0.0322 | 5.61E-05 | ---+--------- | 9.635 | 16 | 0.885 |
| AD Braak | chr13 | 61,504,712 | chr13:61504712:ATT:A | a | att | -0.1294 | 0.0322 | 5.97E-05 | ---+--------- | 9.881 | 16 | 0.8728 |
| AD Braak | chr13 | 61,504,913 | chr13:61504913:A:G | a | g | 0.133 | 0.0324 | 4.00E-05 | +++-+++++- | 9.785 | 16 | 0.8776 |
| AD Braak | chr13 | 61,505,149 | chr13:61505149:T:C | t | c | 0.1297 | 0.0322 | 5.76E-05 | +++-+++++- | 9.632 | 16 | 0.8851 |
| AD Braak | chr13 | 61,505,469 | chr13:61505469:G:A | a | g | -0.1298 | 0.0322 | 5.67E-05 | ---+--------- | 9.627 | 16 | 0.8854 |
| AD Braak | chr13 | 61,505,505 | chr13:61505505:G:T | t | g | -0.1292 | 0.0322 | 6.19E-05 | ---+--------- | 9.649 | 16 | 0.8843 |
| AD Braak | chr13 | 61,505,725 | chr13:61505725:G:T | t | g | -0.1302 | 0.0322 | 5.37E-05 | ---+--------- | 9.605 | 16 | 0.8864 |
| AD Braak | chr13 | 61,505,873 | chr13:61505873:A:C | a | c | 0.1298 | 0.0322 | 5.64E-05 | +++-+++++- | 9.634 | 16 | 0.885 |
| AD Braak | chr13 | 61,505,935 | chr13:61505935:A:T | a | t | 0.1298 | 0.0322 | 5.66E-05 | +++-+++++- | 9.625 | 16 | 0.8854 |

| | | | | | | | | | | | | |
|---|---|---|---|---|---|---|---|---|---|---|---|---|
| AD Braak | chr13 | 61,506,060 | chr13:61506060:A:G | a | g | 0.1298 | 0.0322 | 5.64E-05 | +++-+++++- | 9.631 | 16 | 0.8852 |
| AD Braak | chr13 | 61,506,068 | chr13:61506068:C:G | c | g | 0.1298 | 0.0322 | 5.68E-05 | +++-+++++- | 9.628 | 16 | 0.8853 |
| AD Braak | chr13 | 61,506,228 | chr13:61506228:G:T | t | g | -0.1298 | 0.0322 | 5.67E-05 | ---+--------- | 9.629 | 16 | 0.8853 |
| AD Braak | chr13 | 61,506,675 | chr13:61506675:G:T | t | g | -0.1278 | 0.0322 | 7.05E-05 | ---+--------- | 9.561 | 16 | 0.8885 |
| AD Braak | chr13 | 61,506,781 | chr13:61506781:GA:G | g | ga | -0.1296 | 0.0322 | 5.81E-05 | ---+--------- | 9.625 | 16 | 0.8855 |
| AD Braak | chr13 | 61,506,786 | chr13:61506786:A:G | a | g | 0.1296 | 0.0322 | 5.83E-05 | +++-+++++- | 9.623 | 16 | 0.8855 |
| AD Braak | chr13 | 61,506,913 | chr13:61506913:G:A | a | g | -0.1286 | 0.0322 | 6.58E-05 | ---+--------- | 9.605 | 16 | 0.8864 |
| AD Braak | chr13 | 61,506,977 | chr13:61506977:G:T | t | g | -0.1295 | 0.0322 | 5.86E-05 | ---+--------- | 9.635 | 16 | 0.885 |
| AD Braak | chr13 | 61,506,980 | chr13:61506980:A:G | a | g | 0.1295 | 0.0322 | 5.88E-05 | +++-+++++- | 9.636 | 16 | 0.8849 |
| AD Braak | chr13 | 61,506,996 | chr13:61506996:A:T | a | t | 0.1296 | 0.0322 | 5.77E-05 | +++-+++++- | 9.617 | 16 | 0.8859 |
| AD Braak | chr13 | 61,507,213 | chr13:61507213:T:C | t | c | 0.129 | 0.0322 | 6.30E-05 | +++-+++++- | 9.638 | 16 | 0.8849 |
| AD Braak | chr13 | 61,507,423 | chr13:61507423:T:C | t | c | 0.1296 | 0.0322 | 5.78E-05 | +++-+++++- | 9.63 | 16 | 0.8852 |
| AD Braak | chr13 | 61,507,445 | chr13:61507445:A:G | a | g | 0.1296 | 0.0322 | 5.78E-05 | +++-+++++- | 9.63 | 16 | 0.8852 |
| AD Braak | chr13 | 61,507,461 | chr13:61507461:T:A | a | t | -0.1295 | 0.0322 | 5.72E-05 | ---+--------- | 9.375 | 16 | 0.8972 |
| AD Braak | chr13 | 61,507,546 | chr13:61507546:AAAC:A | a | aaac | -0.1297 | 0.0322 | 5.76E-05 | ---+--------- | 9.633 | 16 | 0.8851 |
| AD Braak | chr13 | 61,507,603 | chr13:61507603:A:C | a | c | 0.1296 | 0.0322 | 5.78E-05 | +++-+++++- | 9.636 | 16 | 0.8849 |
| AD Braak | chr13 | 61,507,712 | chr13:61507712:A:G | a | g | 0.1296 | 0.0322 | 5.77E-05 | +++-+++++- | 9.634 | 16 | 0.885 |
| AD Braak | chr13 | 61,507,897 | chr13:61507897:T:C | t | c | 0.1272 | 0.0323 | 8.07E-05 | +++-+++++- | 9.479 | 16 | 0.8924 |
| AD Braak | chr13 | 61,508,824 | chr13:61508824:A:G | a | g | 0.1295 | 0.0322 | 5.70E-05 | +++-+++++- | 9.708 | 16 | 0.8814 |
| AD Braak | chr13 | 61,509,381 | chr13:61509381:G:A | a | g | -0.1302 | 0.0322 | 5.25E-05 | ---+--------- | 9.837 | 16 | 0.875 |
| AD Braak | chr13 | 61,509,581 | chr13:61509581:T:A | a | t | -0.1303 | 0.0322 | 5.32E-05 | ---+--------- | 9.829 | 16 | 0.8754 |
| AD Braak | chr13 | 61,509,585 | chr13:61509585:G:A | a | g | -0.1303 | 0.0322 | 5.32E-05 | ---+--------- | 9.829 | 16 | 0.8754 |
| AD Braak | chr13 | 61,510,848 | chr13:61510848:A:C | a | c | 0.1284 | 0.0322 | 6.73E-05 | +++-+++++- | 9.888 | 16 | 0.8724 |
| AD Braak | chr13 | 61,511,450 | chr13:61511450:T:C | t | c | 0.1277 | 0.0322 | 7.49E-05 | +++-+++++- | 10.581 | 16 | 0.8346 |
| AD Braak | chr13 | 62,383,559 | chr13:62383559:C:T | t | c | -0.2332 | 0.057 | 4.32E-05 | ------------+ | 20.551 | 16 | 0.1964 |
| AD Braak | chr13 | 65,726,430 | chr13:65726430:G:A | a | g | 0.1235 | 0.0317 | 9.73E-05 | ++++++--++ | 20.711 | 16 | 0.1899 |
| AD Braak | chr13 | 65,728,567 | chr13:65728567:T:A | a | t | 0.1237 | 0.0317 | 9.68E-05 | ++++++--++ | 21.235 | 16 | 0.1696 |
| AD Braak | chr13 | 81,662,345 | chr13:81662345:T:C | t | c | -0.4588 | 0.1108 | 3.48E-05 | ----?+-----+- | 20.832 | 15 | 0.1423 |
| AD Braak | chr13 | 97,680,873 | chr13:97680873:A:G | a | g | -0.4363 | 0.1102 | 7.53E-05 | ----+--+--+-- | 20.859 | 13 | 0.07577 |
| AD Braak | chr13 | 101,616,552 | chr13:101616552:G:A | a | g | 0.6126 | 0.1574 | 9.92E-05 | +++???++? | 3.569 | 6 | 0.7348 |
| AD Braak | chr13 | 105,288,925 | chr13:105288925:G:A | a | g | -0.1591 | 0.0377 | 2.48E-05 | -+--+-------- | 17.612 | 16 | 0.3471 |
| AD Braak | chr13 | 105,289,593 | chr13:105289593:T:C | t | c | 0.1587 | 0.0377 | 2.57E-05 | +-++-+++++ | 17.328 | 16 | 0.3647 |
| AD Braak | chr13 | 105,293,992 | chr13:105293992:C:T | t | c | -0.1586 | 0.0377 | 2.64E-05 | -+--+-------- | 16.573 | 16 | 0.4137 |
| AD Braak | chr13 | 105,296,296 | chr13:105296296:T:C | t | c | 0.1585 | 0.0377 | 2.66E-05 | +-++-+++++ | 16.588 | 16 | 0.4127 |
| AD Braak | chr13 | 105,296,722 | chr13:105296722:C:T | t | c | -0.1584 | 0.0377 | 2.68E-05 | -+--+-------- | 16.499 | 16 | 0.4187 |
| AD Braak | chr13 | 105,296,811 | chr13:105296811:C:T | t | c | -0.1582 | 0.0377 | 2.75E-05 | -+--+-------- | 16.508 | 16 | 0.4181 |
| AD Braak | chr13 | 105,298,535 | chr13:105298535:G:T | t | g | -0.156 | 0.0377 | 3.51E-05 | -+--+-------- | 16.461 | 16 | 0.4212 |
| AD Braak | chr13 | 105,299,074 | chr13:105299074:C:G | c | g | 0.1558 | 0.0377 | 3.60E-05 | +-++-+++++ | 16.46 | 16 | 0.4214 |
| AD Braak | chr13 | 105,299,242 | chr13:105299242:A:G | a | g | 0.1576 | 0.0377 | 2.89E-05 | +-++-+++++ | 16.409 | 16 | 0.4248 |
| AD Braak | chr13 | 105,301,787 | chr13:105301787:C:T | t | c | -0.1566 | 0.0375 | 3.00E-05 | -+--+-------- | 16.267 | 16 | 0.4345 |
| AD Braak | chr13 | 105,308,422 | chr13:105308422:C:T | t | c | -0.1564 | 0.0377 | 3.40E-05 | -+--+-------- | 16.988 | 16 | 0.3864 |
| AD Braak | chr14 | 21,209,118 | chr14:21209118:TA:T | t | ta | 0.1552 | 0.0369 | 2.61E-05 | -+++-++++- | 20.101 | 16 | 0.2157 |
| AD Braak | chr14 | 21,210,257 | chr14:21210257:G:C | c | g | 0.1572 | 0.0368 | 1.98E-05 | -+++-++++- | 20.015 | 16 | 0.2195 |
| AD Braak | chr14 | 21,212,265 | chr14:21212265:G:A | a | g | 0.1539 | 0.0369 | 3.01E-05 | -+++-++++- | 19.732 | 16 | 0.2326 |
| AD Braak | chr14 | 21,213,539 | chr14:21213539:A:T | a | t | -0.1558 | 0.0369 | 2.40E-05 | +---+----+--- | 19.944 | 16 | 0.2227 |
| AD Braak | chr14 | 21,214,079 | chr14:21214079:T:G | t | g | -0.1543 | 0.0369 | 2.84E-05 | +---+----+--- | 20.311 | 16 | 0.2065 |

| | | | | | | | | | | | | |
|---|---|---|---|---|---|---|---|---|---|---|---|---|
| AD Braak | chr14 | 21,215,828 | chr14:21215828:G:A | a | g | 0.1534 | 0.0369 | 3.20E-05 | -+++-++++- | 20.139 | 16 | 0.214 |
| AD Braak | chr14 | 21,216,907 | chr14:21216907:T:C | t | c | -0.155 | 0.0369 | 2.63E-05 | +---+----+--- | 20.147 | 16 | 0.2137 |
| AD Braak | chr14 | 21,218,531 | chr14:21218531:C:G | c | g | -0.1517 | 0.0362 | 2.78E-05 | ----+----+--- | 19.471 | 16 | 0.245 |
| AD Braak | chr14 | 21,219,501 | chr14:21219501:A:T | a | t | -0.1535 | 0.0369 | 3.15E-05 | +---+----+--- | 20.165 | 16 | 0.2129 |
| AD Braak | chr14 | 21,221,385 | chr14:21221385:C:T | t | c | 0.1529 | 0.0369 | 3.34E-05 | -+++-++++- | 20.07 | 16 | 0.2171 |
| AD Braak | chr14 | 21,222,971 | chr14:21222971:C:T | t | c | 0.1518 | 0.0362 | 2.72E-05 | ++++-++++- | 19.477 | 16 | 0.2447 |
| AD Braak | chr14 | 21,223,379 | chr14:21223379:C:T | t | c | 0.154 | 0.0368 | 2.93E-05 | -+++-++++- | 20.214 | 16 | 0.2107 |
| AD Braak | chr14 | 21,226,566 | chr14:21226566:TA:T | t | ta | 0.1492 | 0.036 | 3.38E-05 | -+++-++++- | 19.844 | 16 | 0.2273 |
| AD Braak | chr14 | 21,230,700 | chr14:21230700:C:T | t | c | 0.1497 | 0.0367 | 4.52E-05 | -+++-++++- | 20.219 | 16 | 0.2105 |
| AD Braak | chr14 | 21,235,285 | chr14:21235285:G:A | a | g | 0.1516 | 0.0367 | 3.54E-05 | -+++-++++- | 20.729 | 16 | 0.1892 |
| AD Braak | chr14 | 21,235,430 | chr14:21235430:A:G | a | g | -0.1524 | 0.0367 | 3.25E-05 | +---+----+--- | 20.569 | 16 | 0.1957 |
| AD Braak | chr14 | 21,238,517 | chr14:21238517:C:T | t | c | 0.1477 | 0.036 | 4.07E-05 | -+++-++++- | 19.688 | 16 | 0.2346 |
| AD Braak | chr14 | 21,239,045 | chr14:21239045:TGAACATGG:T | t | tgaacatgg | 0.15 | 0.036 | 3.13E-05 | -+++-++++- | 20.283 | 16 | 0.2078 |
| AD Braak | chr14 | 21,239,369 | chr14:21239369:A:G | a | g | -0.1522 | 0.0367 | 3.32E-05 | +---+----+--- | 21.325 | 16 | 0.1664 |
| AD Braak | chr14 | 21,240,833 | chr14:21240833:A:T | a | t | -0.149 | 0.036 | 3.44E-05 | +---+----+--- | 19.501 | 16 | 0.2435 |
| AD Braak | chr14 | 21,240,843 | chr14:21240843:A:G | a | g | -0.1487 | 0.036 | 3.57E-05 | +---+----+--- | 19.505 | 16 | 0.2434 |
| AD Braak | chr14 | 21,241,953 | chr14:21241953:T:C | t | c | -0.1497 | 0.0359 | 3.08E-05 | +---+----+--- | 19.518 | 16 | 0.2427 |
| AD Braak | chr14 | 21,242,109 | chr14:21242109:C:T | t | c | 0.1487 | 0.036 | 3.57E-05 | -+++-++++- | 19.487 | 16 | 0.2442 |
| AD Braak | chr14 | 21,242,267 | chr14:21242267:G:A | a | g | 0.1523 | 0.0367 | 3.25E-05 | -+++-++++- | 20.509 | 16 | 0.1981 |
| AD Braak | chr14 | 21,242,611 | chr14:21242611:G:C | c | g | 0.1522 | 0.0367 | 3.29E-05 | -+++-++++- | 20.53 | 16 | 0.1973 |
| AD Braak | chr14 | 21,242,646 | chr14:21242646:G:A | a | g | 0.1498 | 0.036 | 3.15E-05 | -+++-++++- | 19.58 | 16 | 0.2398 |
| AD Braak | chr14 | 21,243,105 | chr14:21243105:A:G | a | g | -0.1487 | 0.036 | 3.58E-05 | +---+----+--- | 19.5 | 16 | 0.2436 |
| AD Braak | chr14 | 21,243,386 | chr14:21243386:T:C | t | c | -0.15 | 0.036 | 3.07E-05 | ----+----+--- | 19.526 | 16 | 0.2423 |
| AD Braak | chr14 | 21,243,967 | chr14:21243967:CCT:C | cct | c | -0.1516 | 0.0367 | 3.56E-05 | +---+----+--- | 20.834 | 16 | 0.185 |
| AD Braak | chr14 | 21,245,505 | chr14:21245505:T:A | a | t | 0.1478 | 0.0361 | 4.23E-05 | -+++-++++- | 20.01 | 16 | 0.2198 |
| AD Braak | chr14 | 21,245,507 | chr14:21245507:T:A | a | t | 0.1469 | 0.0361 | 4.73E-05 | -+++-++++- | 20.957 | 16 | 0.1802 |
| AD Braak | chr14 | 21,245,932 | chr14:21245932:A:G | a | g | -0.1497 | 0.0367 | 4.63E-05 | +---+----+--- | 20.36 | 16 | 0.2044 |
| AD Braak | chr14 | 21,247,143 | chr14:21247143:C:T | t | c | 0.151 | 0.0366 | 3.78E-05 | -+++-++++- | 20.672 | 16 | 0.1915 |
| AD Braak | chr14 | 21,248,318 | chr14:21248318:C:G | c | g | -0.1506 | 0.0366 | 3.90E-05 | +---+----+--- | 21.034 | 16 | 0.1772 |
| AD Braak | chr14 | 21,248,734 | chr14:21248734:T:C | t | c | -0.1464 | 0.0358 | 4.31E-05 | +---+----+--- | 20.114 | 16 | 0.2151 |
| AD Braak | chr14 | 21,248,926 | chr14:21248926:C:CAATG | caatg | c | 0.1456 | 0.036 | 5.35E-05 | -+++-++++- | 20.536 | 16 | 0.197 |
| AD Braak | chr14 | 21,250,621 | chr14:21250621:TTC:T | t | ttc | 0.155 | 0.0366 | 2.26E-05 | ++++-++++- | 19.55 | 16 | 0.2412 |
| AD Braak | chr14 | 21,250,759 | chr14:21250759:C:T | t | c | 0.1532 | 0.0367 | 2.92E-05 | ++++-++++- | 18.909 | 16 | 0.2734 |
| AD Braak | chr14 | 21,256,567 | chr14:21256567:G:A | a | g | 0.1514 | 0.0365 | 3.34E-05 | ++++-++++- | 19.916 | 16 | 0.224 |
| AD Braak | chr14 | 21,258,157 | chr14:21258157:T:C | t | c | -0.1486 | 0.0358 | 3.34E-05 | ----+----+--- | 18.841 | 16 | 0.277 |
| AD Braak | chr14 | 21,258,282 | chr14:21258282:C:G | c | g | -0.1504 | 0.0365 | 3.81E-05 | ----+----+--- | 19.86 | 16 | 0.2266 |
| AD Braak | chr14 | 21,258,322 | chr14:21258322:G:A | a | g | 0.1515 | 0.0365 | 3.31E-05 | ++++-++++- | 19.82 | 16 | 0.2284 |
| AD Braak | chr14 | 21,258,925 | chr14:21258925:C:T | t | c | 0.1502 | 0.0362 | 3.29E-05 | ++++-++++- | 19.902 | 16 | 0.2247 |
| AD Braak | chr14 | 21,260,417 | chr14:21260417:A:G | a | g | -0.1487 | 0.0358 | 3.33E-05 | ----+----+--- | 18.823 | 16 | 0.278 |
| AD Braak | chr14 | 21,260,579 | chr14:21260579:G:A | a | g | 0.1515 | 0.0365 | 3.31E-05 | ++++-++++- | 19.773 | 16 | 0.2306 |
| AD Braak | chr14 | 21,265,661 | chr14:21265661:T:G | t | g | -0.151 | 0.0366 | 3.75E-05 | ----+----+--- | 19.018 | 16 | 0.2677 |
| AD Braak | chr14 | 21,267,549 | chr14:21267549:GA:G | g | ga | 0.1512 | 0.0366 | 3.62E-05 | ++++-++++- | 19.47 | 16 | 0.2451 |
| AD Braak | chr14 | 21,270,103 | chr14:21270103:C:T | t | c | 0.1474 | 0.0366 | 5.67E-05 | -+++-++++- | 19.825 | 16 | 0.2282 |
| AD Braak | chr14 | 26,687,466 | chr14:26687466:G:A | a | g | 0.5061 | 0.1288 | 8.54E-05 | ++++++?++ | 10.05 | 11 | 0.5259 |
| AD Braak | chr14 | 34,332,257 | chr14:34332257:C:A | a | c | -0.0987 | 0.025 | 8.02E-05 | ----------+-- | 13.052 | 16 | 0.669 |
| AD Braak | chr14 | 34,334,702 | chr14:34334702:AT:A | a | at | -0.0999 | 0.0256 | 9.42E-05 | ----------+-- | 14.135 | 16 | 0.5887 |

| | | | | | | | | | | | | |
|---|---|---|---|---|---|---|---|---|---|---|---|---|
| AD Braak | chr14 | 34,337,631 | chr14:34337631:G:A | a | g | -0.1002 | 0.0254 | 7.94E-05 | -----------+-- | 13.756 | 16 | 0.6169 |
| AD Braak | chr14 | 34,340,486 | chr14:34340486:C:T | t | c | -0.1027 | 0.026 | 7.82E-05 | -----------+-- | 11.163 | 16 | 0.7993 |
| AD Braak | chr14 | 48,339,230 | chr14:48339230:A:G | a | g | -0.3123 | 0.0786 | 7.04E-05 | ----+--+-+--- | 22.604 | 16 | 0.1247 |
| AD Braak | chr14 | 48,339,233 | chr14:48339233:C:T | t | c | 0.3163 | 0.0785 | 5.57E-05 | ++++-++-+- | 22.936 | 16 | 0.1155 |
| AD Braak | chr14 | 52,135,935 | chr14:52135935:A:T | a | t | 0.611 | 0.1524 | 6.12E-05 | +?++??+??- | 5.948 | 6 | 0.429 |
| AD Braak | chr14 | 55,726,076 | chr14:55726076:G:A | a | g | 0.1002 | 0.0255 | 8.77E-05 | +++++++-+- | 14.439 | 16 | 0.5661 |
| AD Braak | chr14 | 55,726,594 | chr14:55726594:AT:A | a | at | 0.1009 | 0.0255 | 7.85E-05 | +++++++-+- | 14.424 | 16 | 0.5672 |
| AD Braak | chr14 | 66,030,966 | chr14:66030966:G:A | a | g | 0.7381 | 0.1861 | 7.29E-05 | ?+?+??+-?? | 3.586 | 8 | 0.8924 |
| AD Braak | chr14 | 91,527,312 | chr14:91527312:TGACA:T | t | tgaca | -0.3625 | 0.089 | 4.59E-05 | -------+--+-- | 10.362 | 16 | 0.8471 |
| AD Braak | chr14 | 91,550,694 | chr14:91550694:GTCTC:G | g | gtctc | -0.3005 | 0.0751 | 6.24E-05 | -------+--?+- | 11.546 | 15 | 0.713 |
| AD Braak | chr14 | 92,090,920 | chr14:92090920:C:T | t | c | 0.289 | 0.0742 | 9.75E-05 | +++++++++ | 11.057 | 16 | 0.8059 |
| AD Braak | chr14 | 92,460,608 | chr14:92460608:G:T | t | g | 0.1187 | 0.0291 | 4.45E-05 | +++++++-+- | 17.902 | 16 | 0.3297 |
| AD Braak | chr14 | 92,465,237 | chr14:92465237:C:A | a | c | 0.1096 | 0.0278 | 8.12E-05 | +-+++++++- | 12.73 | 16 | 0.6924 |
| AD Braak | chr14 | 92,465,654 | chr14:92465654:A:G | a | g | -0.1509 | 0.0386 | 9.44E-05 | -+--+--+----- | 16.806 | 16 | 0.3982 |
| AD Braak | chr14 | 92,465,679 | chr14:92465679:T:C | t | c | -0.1529 | 0.0385 | 7.30E-05 | -+--+--+----- | 15.511 | 16 | 0.4876 |
| AD Braak | chr14 | 92,466,484 | chr14:92466484:T:C | t | c | -0.1205 | 0.0292 | 3.69E-05 | -------+--+-- | 18.439 | 16 | 0.2988 |
| AD Braak | chr14 | 92,467,925 | chr14:92467925:A:G | a | g | 0.1128 | 0.0279 | 5.38E-05 | +-+++++++- | 11.852 | 16 | 0.7541 |
| AD Braak | chr14 | 92,467,971 | chr14:92467971:G:A | a | g | 0.1123 | 0.0281 | 6.36E-05 | +-+++++++- | 11.544 | 16 | 0.7748 |
| AD Braak | chr14 | 92,468,165 | chr14:92468165:A:G | a | g | -0.1516 | 0.0389 | 9.80E-05 | -+--+--+----- | 17.199 | 16 | 0.3729 |
| AD Braak | chr14 | 92,470,339 | chr14:92470339:C:G | c | g | -0.1301 | 0.0294 | 9.83E-06 | -------+-+--- | 12.358 | 16 | 0.719 |
| AD Braak | chr14 | 92,470,346 | chr14:92470346:G:A | a | g | 0.1303 | 0.0294 | 9.44E-06 | +++++++-+- | 12.338 | 16 | 0.7204 |
| AD Braak | chr14 | 92,470,595 | chr14:92470595:G:A | a | g | 0.119 | 0.0289 | 3.85E-05 | ++++-++-++ | 11.572 | 16 | 0.7728 |
| AD Braak | chr14 | 92,470,627 | chr14:92470627:T:C | t | c | -0.129 | 0.0294 | 1.14E-05 | -+-----+-+--- | 12.598 | 16 | 0.7019 |
| AD Braak | chr14 | 92,470,634 | chr14:92470634:G:A | a | g | 0.1298 | 0.0294 | 1.01E-05 | +++++++-+- | 12.178 | 16 | 0.7316 |
| AD Braak | chr14 | 92,470,675 | chr14:92470675:G:T | t | g | 0.1298 | 0.0294 | 1.04E-05 | +++++++-+- | 12.179 | 16 | 0.7316 |
| AD Braak | chr14 | 92,470,683 | chr14:92470683:C:T | t | c | 0.1287 | 0.0294 | 1.25E-05 | +++++++-+- | 12.541 | 16 | 0.706 |
| AD Braak | chr14 | 92,470,949 | chr14:92470949:C:T | t | c | 0.1311 | 0.0294 | 8.17E-06 | +++++++-+- | 12.292 | 16 | 0.7237 |
| AD Braak | chr14 | 92,472,038 | chr14:92472038:T:C | t | c | -0.1189 | 0.0293 | 4.85E-05 | -------+--+-- | 17.475 | 16 | 0.3555 |
| AD Braak | chr14 | 92,472,071 | chr14:92472071:T:C | t | c | -0.1281 | 0.0294 | 1.28E-05 | -+-------+--- | 12.256 | 16 | 0.7262 |
| AD Braak | chr14 | 100,816,739 | chr14:100816739:C:G | c | g | -0.2697 | 0.0688 | 8.88E-05 | ----------+-+ | 21.479 | 16 | 0.1608 |
| AD Braak | chr14 | 102,069,736 | chr14:102069736:T:C | t | c | -0.2476 | 0.0623 | 7.14E-05 | ----------+--+ | 15.117 | 16 | 0.5161 |
| AD Braak | chr14 | 102,069,835 | chr14:102069835:G:A | a | g | 0.2496 | 0.0625 | 6.58E-05 | +++++++++ | 14.918 | 16 | 0.5306 |
| AD Braak | chr14 | 102,069,940 | chr14:102069940:G:T | t | g | 0.2454 | 0.0625 | 8.68E-05 | +++++++++ | 14.25 | 16 | 0.5801 |
| AD Braak | chr14 | 102,072,984 | chr14:102072984:C:G | c | g | -0.2478 | 0.0625 | 7.37E-05 | ----------+--+ | 14.411 | 16 | 0.5681 |
| AD Braak | chr15 | 29,274,675 | chr15:29274675:C:A | a | c | 1.1748 | 0.2877 | 4.44E-05 | ????++???- | 0.631 | 5 | 0.9865 |
| AD Braak | chr15 | 29,279,895 | chr15:29279895:C:G | c | g | -1.1502 | 0.2937 | 9.00E-05 | ????--???-? | 3.148 | 5 | 0.6772 |
| AD Braak | chr15 | 45,298,519 | chr15:45298519:G:A | a | g | -0.4192 | 0.0975 | 1.70E-05 | ---+------++- | 11.953 | 16 | 0.7472 |
| AD Braak | chr15 | 45,337,309 | chr15:45337309:A:C | a | c | 0.3973 | 0.0973 | 4.42E-05 | +++-+++++- | 11.456 | 16 | 0.7805 |
| AD Braak | chr15 | 67,937,813 | chr15:67937813:T:TAGA | t | taga | 0.1007 | 0.0248 | 4.92E-05 | +-++-++-++ | 11.564 | 16 | 0.7734 |
| AD Braak | chr15 | 81,622,618 | chr15:81622618:T:TGGGGTCTA | t | tggggtcta | -0.4984 | 0.1243 | 6.09E-05 | ----+--??--?- | 9.557 | 12 | 0.6547 |
| AD Braak | chr15 | 81,625,644 | chr15:81625644:A:C | a | c | -0.5021 | 0.1237 | 4.93E-05 | ----+--??--?- | 9.529 | 12 | 0.6572 |
| AD Braak | chr15 | 81,638,110 | chr15:81638110:G:A | a | g | 0.5139 | 0.1302 | 7.92E-05 | ++++-+???+ | 16.004 | 11 | 0.141 |
| AD Braak | chr15 | 81,742,190 | chr15:81742190:G:A | a | g | 0.4954 | 0.1062 | 3.11E-06 | ++++-+++?- | 10.872 | 15 | 0.7616 |
| AD Braak | chr15 | 81,744,145 | chr15:81744145:A:G | a | g | -0.4011 | 0.0922 | 1.37E-05 | ----+---+---- | 14.511 | 16 | 0.5607 |
| AD Braak | chr15 | 81,744,307 | chr15:81744307:T:C | t | c | -0.4015 | 0.0924 | 1.39E-05 | ----+---+---- | 14.54 | 16 | 0.5585 |
| AD Braak | chr15 | 81,745,053 | chr15:81745053:T:C | t | c | -0.4025 | 0.0931 | 1.53E-05 | ----+---+---- | 14.539 | 16 | 0.5586 |

| | | | | | | | | | | | | |
|---|---|---|---|---|---|---|---|---|---|---|---|---|
| AD Braak | chr15 | 81,746,067 | chr15:81746067:G:A | a | g | 0.4092 | 0.0927 | 1.01E-05 | ++++-+++-+ | 13.844 | 16 | 0.6103 |
| AD Braak | chr15 | 81,746,108 | chr15:81746108:C:T | t | c | 0.4148 | 0.0925 | 7.25E-06 | ++++-+++-+ | 14.512 | 16 | 0.5606 |
| AD Braak | chr15 | 81,746,279 | chr15:81746279:C:T | t | c | 0.4311 | 0.0932 | 3.71E-06 | ++++-+++-+ | 15.715 | 16 | 0.473 |
| AD Braak | chr15 | 81,747,008 | chr15:81747008:G:T | t | g | 0.4189 | 0.0932 | 6.91E-06 | ++++-+++-+ | 14.916 | 16 | 0.5308 |
| AD Braak | chr15 | 81,806,434 | chr15:81806434:C:T | t | c | 0.3613 | 0.0904 | 6.48E-05 | ++++-+++-+ | 18.06 | 16 | 0.3204 |
| AD Braak | chr15 | 86,414,458 | chr15:86414458:C:T | t | c | 0.107 | 0.027 | 7.56E-05 | ++++-++++- | 9.639 | 16 | 0.8848 |
| AD Braak | chr15 | 89,780,133 | chr15:89780133:A:G | a | g | -0.101 | 0.0247 | 4.47E-05 | -----+-------- | 15.112 | 16 | 0.5165 |
| AD Braak | chr15 | 101,382,478 | chr15:101382478:G:A | a | g | 0.19 | 0.0468 | 4.90E-05 | +-++-+++++ | 9.91 | 16 | 0.8713 |
| AD Braak | chr16 | 2,353,294 | chr16:2353294:G:A | a | g | 0.4355 | 0.1104 | 8.05E-05 | ++++?+?++ | 6.231 | 11 | 0.8575 |
| AD Braak | chr16 | 54,069,282 | chr16:54069282:G:A | a | g | 0.4098 | 0.1017 | 5.64E-05 | +++-+++-+? | 17.044 | 14 | 0.2539 |
| AD Braak | chr16 | 58,643,903 | chr16:58643903:C:T | t | c | 0.2192 | 0.056 | 9.07E-05 | +++++++-+- | 15.209 | 16 | 0.5094 |
| AD Braak | chr16 | 58,644,323 | chr16:58644323:ATG:A | a | atg | 0.2194 | 0.0556 | 8.09E-05 | +++++++-+- | 15.512 | 16 | 0.4875 |
| AD Braak | chr16 | 58,649,955 | chr16:58649955:C:T | t | c | 0.2195 | 0.0558 | 8.42E-05 | +++++++-+- | 18.312 | 16 | 0.306 |
| AD Braak | chr16 | 58,651,109 | chr16:58651109:CA:C | ca | c | -0.2218 | 0.056 | 7.38E-05 | -------+-+--- | 18.694 | 16 | 0.2848 |
| AD Braak | chr16 | 58,651,291 | chr16:58651291:C:T | t | c | 0.2192 | 0.0558 | 8.68E-05 | +++++++-+- | 18.315 | 16 | 0.3058 |
| AD Braak | chr16 | 58,735,432 | chr16:58735432:A:T | a | t | -0.277 | 0.0616 | 6.93E-06 | -------+-+--- | 16.735 | 16 | 0.403 |
| AD Braak | chr16 | 58,735,608 | chr16:58735608:T:G | t | g | -0.2762 | 0.0616 | 7.33E-06 | -------+-+--- | 16.884 | 16 | 0.3932 |
| AD Braak | chr16 | 58,735,871 | chr16:58735871:A:G | a | g | -0.2757 | 0.0616 | 7.58E-06 | -------+-+--- | 16.636 | 16 | 0.4095 |
| AD Braak | chr16 | 58,737,095 | chr16:58737095:T:C | t | c | -0.2778 | 0.0615 | 6.28E-06 | -------+-+--- | 16.534 | 16 | 0.4164 |
| AD Braak | chr16 | 58,737,332 | chr16:58737332:A:G | a | g | -0.2779 | 0.0615 | 6.20E-06 | -------+-+--- | 16.538 | 16 | 0.4161 |
| AD Braak | chr16 | 58,739,815 | chr16:58739815:A:AT | a | at | -0.2761 | 0.0616 | 7.33E-06 | -------+-+--- | 14.763 | 16 | 0.5421 |
| AD Braak | chr16 | 58,740,387 | chr16:58740387:C:T | t | c | 0.2703 | 0.0614 | 1.08E-05 | +++++++-+- | 14.793 | 16 | 0.5398 |
| AD Braak | chr16 | 58,740,655 | chr16:58740655:C:T | t | c | 0.2754 | 0.0611 | 6.53E-06 | +++++++-+- | 15.354 | 16 | 0.4989 |
| AD Braak | chr16 | 58,741,707 | chr16:58741707:G:T | t | g | 0.2739 | 0.0614 | 8.26E-06 | +++++++-+- | 14.776 | 16 | 0.5411 |
| AD Braak | chr16 | 58,742,937 | chr16:58742937:G:A | a | g | 0.2727 | 0.0615 | 9.22E-06 | +++++++-+- | 15.098 | 16 | 0.5175 |
| AD Braak | chr16 | 58,743,040 | chr16:58743040:T:C | t | c | -0.274 | 0.0614 | 8.23E-06 | -------+-+--- | 14.781 | 16 | 0.5407 |
| AD Braak | chr16 | 58,743,783 | chr16:58743783:A:G | a | g | -0.2708 | 0.0614 | 1.04E-05 | -------+-+--- | 14.914 | 16 | 0.5309 |
| AD Braak | chr16 | 58,749,404 | chr16:58749404:GT:G | g | gt | 0.2617 | 0.0605 | 1.53E-05 | +++++++-+- | 15.377 | 16 | 0.4972 |
| AD Braak | chr16 | 58,749,710 | chr16:58749710:T:C | t | c | -0.262 | 0.0612 | 1.84E-05 | -------+-+--- | 14.669 | 16 | 0.549 |
| AD Braak | chr16 | 58,749,807 | chr16:58749807:C:T | t | c | 0.262 | 0.0612 | 1.83E-05 | +++++++-+- | 14.664 | 16 | 0.5493 |
| AD Braak | chr16 | 58,750,848 | chr16:58750848:T:C | t | c | -0.2619 | 0.0612 | 1.85E-05 | -------+-+--- | 14.662 | 16 | 0.5495 |
| AD Braak | chr16 | 58,751,106 | chr16:58751106:C:T | t | c | 0.262 | 0.0612 | 1.84E-05 | +++++++-+- | 14.665 | 16 | 0.5493 |
| AD Braak | chr16 | 58,751,232 | chr16:58751232:C:G | c | g | -0.2554 | 0.0609 | 2.73E-05 | -------+-+--+ | 15.281 | 16 | 0.5041 |
| AD Braak | chr16 | 58,752,020 | chr16:58752020:AAAAC:A | a | aaaac | 0.2514 | 0.0609 | 3.66E-05 | +++++++-+- | 15.67 | 16 | 0.4762 |
| AD Braak | chr16 | 58,753,032 | chr16:58753032:C:T | t | c | 0.2544 | 0.0616 | 3.59E-05 | +++++++-+- | 15.01 | 16 | 0.5239 |
| AD Braak | chr16 | 58,753,160 | chr16:58753160:C:T | t | c | 0.2555 | 0.0609 | 2.72E-05 | +++++++-+- | 15.296 | 16 | 0.5031 |
| AD Braak | chr16 | 58,753,313 | chr16:58753313:A:G | a | g | -0.2545 | 0.0616 | 3.56E-05 | -------+-+--+ | 15.046 | 16 | 0.5212 |
| AD Braak | chr16 | 58,753,940 | chr16:58753940:G:T | t | g | 0.2543 | 0.0616 | 3.61E-05 | +++++++-+- | 15.002 | 16 | 0.5245 |
| AD Braak | chr16 | 58,754,307 | chr16:58754307:G:A | a | g | 0.2544 | 0.0616 | 3.60E-05 | +++++++-+- | 14.999 | 16 | 0.5247 |
| AD Braak | chr16 | 58,754,918 | chr16:58754918:C:T | t | c | 0.2518 | 0.0616 | 4.36E-05 | +++++++-+- | 15.797 | 16 | 0.4672 |
| AD Braak | chr16 | 58,756,790 | chr16:58756790:TTG:T | t | ttg | 0.2515 | 0.0615 | 4.37E-05 | +++++++-+- | 15.699 | 16 | 0.4742 |
| AD Braak | chr16 | 58,758,314 | chr16:58758314:A:C | a | c | -0.2561 | 0.0609 | 2.58E-05 | -------+-+--+ | 15.773 | 16 | 0.4689 |
| AD Braak | chr16 | 58,759,598 | chr16:58759598:CTT:C | ctt | c | -0.2529 | 0.0616 | 3.97E-05 | -------+-+--+ | 15.591 | 16 | 0.4819 |
| AD Braak | chr16 | 58,759,759 | chr16:58759759:A:G | a | g | -0.2514 | 0.0615 | 4.40E-05 | -------+-+--+ | 15.693 | 16 | 0.4746 |
| AD Braak | chr16 | 58,761,821 | chr16:58761821:A:C | a | c | -0.2518 | 0.0614 | 4.14E-05 | -------+-+--+ | 15.962 | 16 | 0.4556 |
| AD Braak | chr16 | 58,761,929 | chr16:58761929:A:T | a | t | -0.2527 | 0.0615 | 3.95E-05 | -------+-+--+ | 15.59 | 16 | 0.4819 |

| | | | | | | | | | | | | |
|---|---|---|---|---|---|---|---|---|---|---|---|---|
| AD Braak | chr16 | 58,762,119 | chr16:58762119:G:T | t | g | 0.2526 | 0.0615 | 3.98E-05 | +++++++-+- | 15.593 | 16 | 0.4817 |
| AD Braak | chr16 | 58,763,169 | chr16:58763169:C:T | t | c | 0.254 | 0.0608 | 2.92E-05 | +++++++-+- | 15.568 | 16 | 0.4835 |
| AD Braak | chr16 | 58,764,250 | chr16:58764250:GCTT:G | g | gctt | 0.2526 | 0.0615 | 3.98E-05 | +++++++-+- | 15.626 | 16 | 0.4793 |
| AD Braak | chr16 | 58,765,078 | chr16:58765078:T:G | t | g | -0.2525 | 0.0615 | 4.00E-05 | -------+-+--+ | 15.585 | 16 | 0.4823 |
| AD Braak | chr16 | 58,765,348 | chr16:58765348:A:G | a | g | -0.2533 | 0.0614 | 3.74E-05 | -------+-+--+ | 16.558 | 16 | 0.4147 |
| AD Braak | chr16 | 58,766,359 | chr16:58766359:T:C | t | c | -0.2532 | 0.0614 | 3.75E-05 | -------+-+--+ | 16.559 | 16 | 0.4147 |
| AD Braak | chr16 | 58,766,576 | chr16:58766576:C:CT | ct | c | 0.2516 | 0.0615 | 4.34E-05 | +++++++-+- | 15.575 | 16 | 0.483 |
| AD Braak | chr16 | 58,766,823 | chr16:58766823:T:C | t | c | -0.257 | 0.0608 | 2.40E-05 | -------+-+--+ | 16.005 | 16 | 0.4526 |
| AD Braak | chr16 | 58,767,205 | chr16:58767205:T:G | t | g | -0.2531 | 0.0614 | 3.81E-05 | -------+-+--+ | 16.532 | 16 | 0.4165 |
| AD Braak | chr16 | 58,767,371 | chr16:58767371:G:A | a | g | 0.2531 | 0.0614 | 3.78E-05 | +++++++-+- | 16.597 | 16 | 0.4121 |
| AD Braak | chr16 | 58,767,431 | chr16:58767431:G:A | a | g | 0.2529 | 0.0615 | 3.88E-05 | +++++++-+- | 15.577 | 16 | 0.4828 |
| AD Braak | chr16 | 58,768,829 | chr16:58768829:C:T | t | c | 0.2524 | 0.0608 | 3.33E-05 | +++++++-+- | 15.793 | 16 | 0.4675 |
| AD Braak | chr16 | 58,769,651 | chr16:58769651:T:G | t | g | -0.2559 | 0.0615 | 3.16E-05 | -------+-+--+ | 16.188 | 16 | 0.4399 |
| AD Braak | chr16 | 58,769,875 | chr16:58769875:A:G | a | g | -0.2483 | 0.0615 | 5.41E-05 | -------+-+--+ | 15.934 | 16 | 0.4576 |
| AD Braak | chr16 | 58,769,890 | chr16:58769890:A:G | a | g | -0.2483 | 0.0615 | 5.43E-05 | -------+-+--+ | 15.939 | 16 | 0.4572 |
| AD Braak | chr16 | 58,770,000 | chr16:58770000:C:T | t | c | 0.2474 | 0.0615 | 5.80E-05 | +++++++-+- | 16.042 | 16 | 0.4501 |
| AD Braak | chr16 | 58,770,003 | chr16:58770003:G:A | a | g | 0.2484 | 0.0615 | 5.43E-05 | +++++++-+- | 15.939 | 16 | 0.4572 |
| AD Braak | chr16 | 58,770,077 | chr16:58770077:C:T | t | c | 0.2484 | 0.0615 | 5.41E-05 | +++++++-+- | 15.946 | 16 | 0.4568 |
| AD Braak | chr16 | 58,770,440 | chr16:58770440:TTTAA:T | t | tttaa | 0.2468 | 0.0615 | 5.98E-05 | +++++++-+- | 15.953 | 16 | 0.4562 |
| AD Braak | chr16 | 58,770,477 | chr16:58770477:T:C | t | c | -0.2551 | 0.0615 | 3.32E-05 | -------+-+--+ | 16.251 | 16 | 0.4356 |
| AD Braak | chr16 | 58,771,954 | chr16:58771954:C:T | t | c | 0.2554 | 0.0614 | 3.24E-05 | +++++++-+- | 16.335 | 16 | 0.4298 |
| AD Braak | chr16 | 58,773,323 | chr16:58773323:T:A | a | t | 0.2538 | 0.0615 | 3.68E-05 | +++++++-+- | 15.444 | 16 | 0.4924 |
| AD Braak | chr16 | 58,773,599 | chr16:58773599:A:G | a | g | -0.2551 | 0.0614 | 3.31E-05 | -------+-+--+ | 16.145 | 16 | 0.4429 |
| AD Braak | chr16 | 58,774,659 | chr16:58774659:A:G | a | g | -0.2535 | 0.0614 | 3.70E-05 | -------+-+--+ | 15.553 | 16 | 0.4846 |
| AD Braak | chr16 | 58,774,692 | chr16:58774692:C:T | t | c | 0.255 | 0.0614 | 3.31E-05 | +++++++-+- | 16.144 | 16 | 0.443 |
| AD Braak | chr16 | 58,774,792 | chr16:58774792:A:C | a | c | -0.255 | 0.0614 | 3.31E-05 | -------+-+--+ | 16.145 | 16 | 0.4429 |
| AD Braak | chr16 | 58,775,728 | chr16:58775728:C:T | t | c | 0.2508 | 0.0615 | 4.50E-05 | +++++++-+- | 14.597 | 16 | 0.5544 |
| AD Braak | chr16 | 58,776,599 | chr16:58776599:G:A | a | g | 0.2573 | 0.0614 | 2.80E-05 | +++++++-+- | 14.909 | 16 | 0.5313 |
| AD Braak | chr16 | 58,776,837 | chr16:58776837:T:C | t | c | -0.2571 | 0.0607 | 2.25E-05 | -------+-+--+ | 15.566 | 16 | 0.4837 |
| AD Braak | chr16 | 58,783,887 | chr16:58783887:ATTG:A | a | attg | 0.2589 | 0.0615 | 2.55E-05 | +++++++-+- | 15.053 | 16 | 0.5208 |
| AD Braak | chr16 | 58,783,984 | chr16:58783984:T:A | a | t | 0.2614 | 0.0614 | 2.08E-05 | +++++++-+- | 15.29 | 16 | 0.5035 |
| AD Braak | chr16 | 58,785,368 | chr16:58785368:G:A | a | g | 0.258 | 0.0614 | 2.66E-05 | +++++++-+- | 14.723 | 16 | 0.545 |
| AD Braak | chr16 | 58,787,631 | chr16:58787631:A:G | a | g | -0.2605 | 0.0613 | 2.16E-05 | -------+-+--+ | 14.715 | 16 | 0.5456 |
| AD Braak | chr16 | 58,787,724 | chr16:58787724:C:T | t | c | 0.2605 | 0.0613 | 2.16E-05 | +++++++-+- | 14.736 | 16 | 0.544 |
| AD Braak | chr16 | 58,789,915 | chr16:58789915:C:T | t | c | 0.2576 | 0.0617 | 2.95E-05 | +++++++-+- | 14.359 | 16 | 0.572 |
| AD Braak | chr16 | 58,792,825 | chr16:58792825:T:G | t | g | -0.2624 | 0.0613 | 1.88E-05 | -------+-+--+ | 14.443 | 16 | 0.5657 |
| AD Braak | chr16 | 58,795,177 | chr16:58795177:A:C | a | c | -0.2597 | 0.0614 | 2.34E-05 | -------+-+--+ | 14.502 | 16 | 0.5613 |
| AD Braak | chr16 | 58,796,259 | chr16:58796259:C:T | t | c | 0.2603 | 0.0614 | 2.24E-05 | +++++++-+- | 14.496 | 16 | 0.5618 |
| AD Braak | chr16 | 58,796,817 | chr16:58796817:A:G | a | g | -0.2602 | 0.0614 | 2.27E-05 | -------+-+--+ | 14.498 | 16 | 0.5617 |
| AD Braak | chr16 | 58,797,076 | chr16:58797076:T:G | t | g | -0.26 | 0.0614 | 2.28E-05 | -------+-+--+ | 14.49 | 16 | 0.5622 |
| AD Braak | chr16 | 58,800,096 | chr16:58800096:G:T | t | g | 0.2531 | 0.0608 | 3.19E-05 | +++++++-+- | 13.231 | 16 | 0.6558 |
| AD Braak | chr16 | 58,800,498 | chr16:58800498:A:C | a | c | -0.255 | 0.061 | 2.91E-05 | -------+-+--- | 12.618 | 16 | 0.7005 |
| AD Braak | chr16 | 58,801,258 | chr16:58801258:CA:C | ca | c | -0.2556 | 0.061 | 2.78E-05 | -------+-+--- | 12.631 | 16 | 0.6995 |
| AD Braak | chr16 | 58,802,131 | chr16:58802131:T:G | t | g | -0.2551 | 0.061 | 2.91E-05 | -------+-+--- | 12.569 | 16 | 0.704 |
| AD Braak | chr16 | 58,802,133 | chr16:58802133:T:C | t | c | -0.2551 | 0.061 | 2.91E-05 | -------+-+--- | 12.569 | 16 | 0.704 |
| AD Braak | chr16 | 58,802,424 | chr16:58802424:A:G | a | g | -0.2557 | 0.061 | 2.78E-05 | -------+-+--- | 12.562 | 16 | 0.7045 |

| | | | | | | | | | | | | |
|---|---|---|---|---|---|---|---|---|---|---|---|---|
| AD Braak | chr16 | 58,802,860 | chr16:58802860:G:A | a | g | 0.2549 | 0.061 | 2.93E-05 | +++++++-+- | 12.569 | 16 | 0.704 |
| AD Braak | chr16 | 58,802,936 | chr16:58802936:A:ATC | a | atc | -0.255 | 0.061 | 2.91E-05 | -------+-+--- | 12.568 | 16 | 0.704 |
| AD Braak | chr16 | 58,803,106 | chr16:58803106:G:C | c | g | 0.2578 | 0.061 | 2.40E-05 | +++++++-+- | 12.736 | 16 | 0.692 |
| AD Braak | chr16 | 58,803,310 | chr16:58803310:C:A | a | c | 0.2552 | 0.061 | 2.89E-05 | +++++++-+- | 12.576 | 16 | 0.7035 |
| AD Braak | chr16 | 58,803,451 | chr16:58803451:A:G | a | g | -0.2551 | 0.061 | 2.91E-05 | -------+-+--- | 12.568 | 16 | 0.704 |
| AD Braak | chr16 | 58,803,566 | chr16:58803566:AAAAGG:A | a | aaaagg | 0.255 | 0.061 | 2.92E-05 | +++++++-+- | 12.576 | 16 | 0.7034 |
| AD Braak | chr16 | 58,803,702 | chr16:58803702:T:C | t | c | -0.2551 | 0.061 | 2.91E-05 | -------+-+--- | 12.568 | 16 | 0.704 |
| AD Braak | chr16 | 58,803,775 | chr16:58803775:G:A | a | g | 0.2553 | 0.061 | 2.88E-05 | +++++++-+- | 12.586 | 16 | 0.7027 |
| AD Braak | chr16 | 58,803,863 | chr16:58803863:G:A | a | g | 0.2551 | 0.061 | 2.91E-05 | +++++++-+- | 12.568 | 16 | 0.704 |
| AD Braak | chr16 | 58,803,880 | chr16:58803880:A:G | a | g | -0.2551 | 0.061 | 2.91E-05 | -------+-+--- | 12.568 | 16 | 0.704 |
| AD Braak | chr16 | 58,804,009 | chr16:58804009:G:C | c | g | 0.2537 | 0.061 | 3.24E-05 | +++++++-+- | 12.842 | 16 | 0.6843 |
| AD Braak | chr16 | 58,804,312 | chr16:58804312:T:G | t | g | -0.2551 | 0.061 | 2.91E-05 | -------+-+--- | 12.568 | 16 | 0.704 |
| AD Braak | chr16 | 58,804,363 | chr16:58804363:G:A | a | g | 0.2566 | 0.061 | 2.64E-05 | +++++++-+- | 12.465 | 16 | 0.7114 |
| AD Braak | chr16 | 58,804,509 | chr16:58804509:T:C | t | c | -0.2517 | 0.0612 | 3.87E-05 | -------+-+--- | 12.238 | 16 | 0.7275 |
| AD Braak | chr16 | 58,804,607 | chr16:58804607:C:T | t | c | 0.2551 | 0.061 | 2.89E-05 | +++++++-+- | 12.573 | 16 | 0.7037 |
| AD Braak | chr16 | 58,804,652 | chr16:58804652:G:T | t | g | 0.2551 | 0.061 | 2.89E-05 | +++++++-+- | 12.573 | 16 | 0.7037 |
| AD Braak | chr16 | 58,805,631 | chr16:58805631:A:G | a | g | -0.2556 | 0.061 | 2.80E-05 | -------+-+--- | 12.582 | 16 | 0.703 |
| AD Braak | chr16 | 58,805,845 | chr16:58805845:TTCAG:T | t | ttcag | 0.2541 | 0.061 | 3.12E-05 | +++++++-+- | 12.838 | 16 | 0.6845 |
| AD Braak | chr16 | 58,805,876 | chr16:58805876:C:T | t | c | 0.2326 | 0.0544 | 1.91E-05 | +++++++-+- | 13.38 | 16 | 0.6448 |
| AD Braak | chr16 | 58,805,963 | chr16:58805963:G:A | a | g | 0.2545 | 0.061 | 3.02E-05 | +++++++-+- | 12.631 | 16 | 0.6996 |
| AD Braak | chr16 | 58,806,839 | chr16:58806839:A:AAAT | a | aaat | -0.2551 | 0.061 | 2.90E-05 | -------+-+--- | 12.583 | 16 | 0.703 |
| AD Braak | chr16 | 58,806,951 | chr16:58806951:G:T | t | g | 0.2554 | 0.061 | 2.79E-05 | +++++++-+- | 12.569 | 16 | 0.704 |
| AD Braak | chr16 | 58,806,980 | chr16:58806980:G:A | a | g | 0.2557 | 0.061 | 2.77E-05 | +++++++-+- | 12.592 | 16 | 0.7023 |
| AD Braak | chr16 | 58,807,007 | chr16:58807007:T:G | t | g | -0.255 | 0.061 | 2.91E-05 | -------+-+--- | 12.593 | 16 | 0.7023 |
| AD Braak | chr16 | 58,807,825 | chr16:58807825:C:T | t | c | 0.2548 | 0.061 | 2.99E-05 | +++++++-+- | 12.558 | 16 | 0.7048 |
| AD Braak | chr16 | 58,807,872 | chr16:58807872:AG:A | a | ag | 0.2551 | 0.061 | 2.91E-05 | +++++++-+- | 12.576 | 16 | 0.7035 |
| AD Braak | chr16 | 58,808,452 | chr16:58808452:CAT:C | cat | c | -0.2521 | 0.061 | 3.63E-05 | -------+-+--- | 12.501 | 16 | 0.7089 |
| AD Braak | chr16 | 58,809,350 | chr16:58809350:A:AAC | a | aac | -0.255 | 0.0611 | 2.97E-05 | -------+-+--- | 12.609 | 16 | 0.7011 |
| AD Braak | chr16 | 58,809,673 | chr16:58809673:A:AAAAG | a | aaaag | -0.2563 | 0.0613 | 2.88E-05 | -------+-+--- | 12.411 | 16 | 0.7152 |
| AD Braak | chr16 | 58,811,061 | chr16:58811061:G:A | a | g | 0.2543 | 0.0611 | 3.13E-05 | +++++++-+- | 12.623 | 16 | 0.7001 |
| AD Braak | chr16 | 58,811,318 | chr16:58811318:A:G | a | g | -0.2549 | 0.0611 | 3.06E-05 | -------+-+--- | 12.504 | 16 | 0.7086 |
| AD Braak | chr16 | 58,811,974 | chr16:58811974:C:T | t | c | 0.2547 | 0.0612 | 3.12E-05 | +++++++-+- | 12.404 | 16 | 0.7157 |
| AD Braak | chr16 | 58,812,209 | chr16:58812209:A:T | a | t | -0.2548 | 0.0612 | 3.16E-05 | -------+-+--- | 12.387 | 16 | 0.717 |
| AD Braak | chr16 | 58,821,160 | chr16:58821160:G:A | a | g | 0.2569 | 0.0614 | 2.86E-05 | +++++++-+- | 13.004 | 16 | 0.6724 |
| AD Braak | chr16 | 62,552,721 | chr16:62552721:T:C | t | c | 0.2742 | 0.0652 | 2.58E-05 | +-+++++++- | 14.177 | 15 | 0.5122 |
| AD Braak | chr16 | 63,632,713 | chr16:63632713:C:T | t | c | 0.3539 | 0.0838 | 2.40E-05 | +-++++-++- | 19.087 | 16 | 0.2642 |
| AD Braak | chr16 | 79,431,916 | chr16:79431916:G:A | a | g | -0.1146 | 0.0286 | 6.26E-05 | ---+---+---+- | 14.135 | 16 | 0.5886 |
| AD Braak | chr17 | 5,382,643 | chr17:5382643:TTGGGAGGC:T | t | ttgggaggc | 0.2588 | 0.0658 | 8.50E-05 | ++++--+-++ | 12.15 | 16 | 0.7336 |
| AD Braak | chr17 | 5,412,875 | chr17:5412875:C:T | t | c | 0.4587 | 0.1145 | 6.18E-05 | +?++-++-+- | 23.478 | 14 | 0.05292 |
| AD Braak | chr17 | 9,229,492 | chr17:9229492:C:T | t | c | -0.0993 | 0.0252 | 8.13E-05 | -+-++-----+- | 24.568 | 16 | 0.07782 |
| AD Braak | chr17 | 33,381,129 | chr17:33381129:C:T | t | c | 0.4384 | 0.1089 | 5.64E-05 | +-++--+++? | 17.26 | 12 | 0.1401 |
| AD Braak | chr17 | 37,008,846 | chr17:37008846:T:C | t | c | -0.6474 | 0.1658 | 9.46E-05 | ?-+-???+-? | 11.912 | 8 | 0.1552 |
| AD Braak | chr17 | 37,066,051 | chr17:37066051:G:A | a | g | 0.6469 | 0.1646 | 8.53E-05 | ?+-+???-+? | 9.325 | 8 | 0.3156 |
| AD Braak | chr17 | 45,372,711 | chr17:45372711:A:G | a | g | -0.4087 | 0.1035 | 7.84E-05 | ----+--+--+- | 15.071 | 14 | 0.3733 |
| AD Braak | chr17 | 51,053,155 | chr17:51053155:G:A | a | g | 0.2236 | 0.0564 | 7.42E-05 | +-+++++-++ | 11.766 | 16 | 0.7599 |
| AD Braak | chr17 | 55,870,044 | chr17:55870044:T:C | t | c | -0.2499 | 0.064 | 9.51E-05 | -----------++ | 6.62 | 16 | 0.9799 |

| | | | | | | | | | | | | |
|---|---|---|---|---|---|---|---|---|---|---|---|---|
| AD Braak | chr17 | 55,878,529 | chr17:55878529:C:T | t | c | 0.2496 | 0.0641 | 9.73E-05 | +++++++++ | 6.633 | 16 | 0.9797 |
| AD Braak | chr17 | 64,487,814 | chr17:64487814:C:T | t | c | 0.1884 | 0.0468 | 5.75E-05 | ++++++-++- | 11.601 | 16 | 0.771 |
| AD Braak | chr17 | 64,534,767 | chr17:64534767:TAAG:T | t | taag | 0.1832 | 0.0464 | 7.98E-05 | ++++++-++- | 11.459 | 16 | 0.7803 |
| AD Braak | chr17 | 74,123,278 | chr17:74123278:C:A | a | c | -0.3739 | 0.091 | 3.98E-05 | +-----+---+-- | 19.807 | 16 | 0.229 |
| AD Braak | chr17 | 79,232,926 | chr17:79232926:C:T | t | c | 0.1297 | 0.0284 | 4.78E-06 | ++++-++++- | 17.008 | 16 | 0.3851 |
| AD Braak | chr17 | 79,235,041 | chr17:79235041:T:C | t | c | -0.1238 | 0.0295 | 2.63E-05 | --+-+--+----- | 20.953 | 16 | 0.1803 |
| AD Braak | chr17 | 79,235,375 | chr17:79235375:T:C | t | c | -0.1209 | 0.0278 | 1.38E-05 | --+-+---+---- | 20.954 | 16 | 0.1803 |
| AD Braak | chr17 | 79,236,004 | chr17:79236004:G:T | t | g | 0.1158 | 0.0267 | 1.41E-05 | ++-+-+++++ | 19.521 | 16 | 0.2426 |
| AD Braak | chr17 | 79,240,007 | chr17:79240007:A:G | a | g | -0.1096 | 0.0261 | 2.70E-05 | --+-+-------- | 16.357 | 16 | 0.4284 |
| AD Braak | chr17 | 79,240,123 | chr17:79240123:C:T | t | c | 0.1096 | 0.0257 | 2.03E-05 | ++-+-+++++ | 17.379 | 16 | 0.3615 |
| AD Braak | chr18 | 30,887,272 | chr18:30887272:C:T | t | c | 0.1993 | 0.048 | 3.30E-05 | ++++-+--++ | 10.773 | 16 | 0.8233 |
| AD Braak | chr18 | 31,006,457 | chr18:31006457:G:A | a | g | 0.1112 | 0.0282 | 7.89E-05 | ++++-++++- | 16.101 | 16 | 0.4459 |
| AD Braak | chr18 | 31,006,654 | chr18:31006654:T:C | t | c | -0.1112 | 0.0282 | 7.86E-05 | ----+----+-+- | 16.049 | 16 | 0.4495 |
| AD Braak | chr18 | 31,007,311 | chr18:31007311:C:G | c | g | -0.1114 | 0.0281 | 7.46E-05 | ----+----+-+- | 15.732 | 16 | 0.4718 |
| AD Braak | chr18 | 31,007,593 | chr18:31007593:G:A | a | g | 0.11 | 0.0281 | 8.94E-05 | ++++-++++- | 15.834 | 16 | 0.4646 |
| AD Braak | chr18 | 31,008,571 | chr18:31008571:C:T | t | c | 0.1137 | 0.0281 | 5.23E-05 | ++++-++++- | 15.819 | 16 | 0.4657 |
| AD Braak | chr18 | 31,008,899 | chr18:31008899:T:G | t | g | -0.111 | 0.0281 | 7.79E-05 | ----+----+-+- | 15.708 | 16 | 0.4735 |
| AD Braak | chr18 | 31,009,715 | chr18:31009715:A:G | a | g | -0.111 | 0.0282 | 8.21E-05 | ----+----+++ | 16.132 | 16 | 0.4438 |
| AD Braak | chr18 | 31,010,362 | chr18:31010362:G:C | c | g | 0.1111 | 0.0281 | 7.81E-05 | ++++-++++- | 15.737 | 16 | 0.4715 |
| AD Braak | chr18 | 31,010,566 | chr18:31010566:A:G | a | g | -0.1109 | 0.0281 | 8.16E-05 | ----+----+++ | 15.749 | 16 | 0.4706 |
| AD Braak | chr18 | 60,648,170 | chr18:60648170:A:G | a | g | 0.2597 | 0.063 | 3.73E-05 | +-++++--++ | 12.352 | 16 | 0.7194 |
| AD Braak | chr18 | 60,648,558 | chr18:60648558:T:C | t | c | 0.2633 | 0.063 | 2.89E-05 | +-++++--++ | 12.443 | 16 | 0.713 |
| AD Braak | chr18 | 60,648,641 | chr18:60648641:G:A | a | g | -0.2673 | 0.063 | 2.21E-05 | -+----++----- | 12.381 | 16 | 0.7173 |
| AD Braak | chr18 | 60,648,676 | chr18:60648676:G:C | c | g | -0.2661 | 0.0629 | 2.36E-05 | -+----++----- | 12.402 | 16 | 0.7159 |
| AD Braak | chr18 | 60,649,202 | chr18:60649202:T:C | t | c | 0.2649 | 0.0629 | 2.56E-05 | +-++++--++ | 12.444 | 16 | 0.7129 |
| AD Braak | chr18 | 60,649,491 | chr18:60649491:GAT:G | g | gat | -0.2648 | 0.0629 | 2.59E-05 | -+----++----- | 12.447 | 16 | 0.7127 |
| AD Braak | chr18 | 60,649,652 | chr18:60649652:G:C | c | g | -0.2649 | 0.0629 | 2.58E-05 | -+----++----- | 12.449 | 16 | 0.7125 |
| AD Braak | chr18 | 60,649,782 | chr18:60649782:C:G | c | g | 0.265 | 0.0629 | 2.55E-05 | +-++++--++ | 12.444 | 16 | 0.7129 |
| AD Braak | chr18 | 60,649,796 | chr18:60649796:G:C | c | g | -0.2649 | 0.0629 | 2.56E-05 | -+----++----- | 12.443 | 16 | 0.7129 |
| AD Braak | chr18 | 60,649,797 | chr18:60649797:A:C | a | c | 0.2649 | 0.0629 | 2.56E-05 | +-++++--++ | 12.444 | 16 | 0.7129 |
| AD Braak | chr18 | 60,659,516 | chr18:60659516:G:A | a | g | -0.2641 | 0.0632 | 2.90E-05 | -+----++----- | 12.447 | 16 | 0.7126 |
| AD Braak | chr18 | 60,678,653 | chr18:60678653:T:C | t | c | 0.2618 | 0.0629 | 3.12E-05 | +-++++--++ | 12.144 | 16 | 0.734 |
| AD Braak | chr18 | 60,681,380 | chr18:60681380:T:C | t | c | 0.4578 | 0.1026 | 8.04E-06 | +-++++--++ | 15.871 | 16 | 0.462 |
| AD Braak | chr18 | 60,681,417 | chr18:60681417:G:A | a | g | -0.2512 | 0.0612 | 4.00E-05 | -+----++--+- | 13.217 | 16 | 0.6568 |
| AD Braak | chr18 | 60,682,978 | chr18:60682978:T:C | t | c | 0.2524 | 0.0612 | 3.68E-05 | +-++++--++ | 12.879 | 16 | 0.6816 |
| AD Braak | chr18 | 60,683,210 | chr18:60683210:A:C | a | c | 0.2509 | 0.061 | 3.90E-05 | +-++++--++ | 13.21 | 16 | 0.6574 |
| AD Braak | chr18 | 60,684,626 | chr18:60684626:T:G | t | g | 0.2482 | 0.061 | 4.71E-05 | +-++++--++ | 13.881 | 16 | 0.6075 |
| AD Braak | chr18 | 60,684,784 | chr18:60684784:T:C | t | c | 0.2615 | 0.0627 | 3.05E-05 | +-++++--++ | 12.471 | 16 | 0.711 |
| AD Braak | chr18 | 60,685,137 | chr18:60685137:C:G | c | g | 0.4383 | 0.1012 | 1.49E-05 | +-+++++-++ | 15.43 | 16 | 0.4934 |
| AD Braak | chr18 | 60,686,664 | chr18:60686664:A:C | a | c | 0.4385 | 0.1012 | 1.48E-05 | +-+++++-++ | 15.43 | 16 | 0.4934 |
| AD Braak | chr18 | 60,687,617 | chr18:60687617:T:A | a | t | -0.4388 | 0.1012 | 1.44E-05 | -+-----+----- | 15.393 | 16 | 0.4961 |
| AD Braak | chr18 | 60,694,457 | chr18:60694457:A:G | a | g | 0.3727 | 0.0932 | 6.35E-05 | +-+++++-++ | 17.517 | 16 | 0.3529 |
| AD Braak | chr18 | 60,696,063 | chr18:60696063:A:G | a | g | 0.4421 | 0.102 | 1.47E-05 | +-+++++-++ | 15.953 | 16 | 0.4563 |
| AD Braak | chr18 | 60,696,352 | chr18:60696352:G:A | a | g | -0.4421 | 0.102 | 1.47E-05 | -+-----+----- | 15.929 | 16 | 0.4579 |
| AD Braak | chr18 | 77,904,085 | chr18:77904085:A:T | a | t | 0.2517 | 0.0633 | 6.97E-05 | ++++-++++- | 5.78 | 15 | 0.9832 |
| AD Braak | chr18 | 77,904,188 | chr18:77904188:T:C | t | c | 0.2484 | 0.0637 | 9.54E-05 | ++++-++++- | 5.164 | 15 | 0.9906 |

| | | | | | | | | | | | | |
|---|---|---|---|---|---|---|---|---|---|---|---|---|
| AD Braak | chr18 | 77,904,264 | chr18:77904264:C:T | t | c | 0.2517 | 0.0637 | 7.85E-05 | ++++-++++- | 5.584 | 16 | 0.992 |
| AD Braak | chr18 | 77,904,903 | chr18:77904903:C:T | t | c | 0.2522 | 0.0635 | 7.11E-05 | ++++-++++- | 5.527 | 16 | 0.9924 |
| AD Braak | chr18 | 77,910,216 | chr18:77910216:G:A | a | g | 0.2559 | 0.0636 | 5.69E-05 | ++++-++++- | 5.522 | 16 | 0.9925 |
| AD Braak | chr18 | 77,912,320 | chr18:77912320:C:A | a | c | -0.2424 | 0.0621 | 9.37E-05 | -------------- | 5.856 | 16 | 0.9896 |
| AD Braak | chr18 | 77,914,049 | chr18:77914049:G:C | c | g | 0.1676 | 0.0415 | 5.34E-05 | ++++++-++- | 16.525 | 16 | 0.4169 |
| AD Braak | chr18 | 77,914,243 | chr18:77914243:A:C | a | c | -0.2664 | 0.063 | 2.33E-05 | ----------+--- | 7.262 | 16 | 0.9679 |
| AD Braak | chr18 | 77,915,819 | chr18:77915819:G:C | c | g | 0.2537 | 0.0625 | 4.93E-05 | +++++++++ | 6.789 | 16 | 0.9771 |
| AD Braak | chr18 | 77,918,139 | chr18:77918139:C:A | a | c | 0.261 | 0.0629 | 3.30E-05 | +++++++++ | 6.695 | 16 | 0.9787 |
| AD Braak | chr18 | 77,918,303 | chr18:77918303:C:T | t | c | 0.2657 | 0.0628 | 2.34E-05 | +++++++++ | 6.936 | 16 | 0.9745 |
| AD Braak | chr18 | 77,920,892 | chr18:77920892:C:T | t | c | 0.2687 | 0.0625 | 1.72E-05 | +++++++++ | 6.45 | 16 | 0.9825 |
| AD Braak | chr18 | 77,923,214 | chr18:77923214:A:G | a | g | -0.2605 | 0.0638 | 4.45E-05 | ----------+--- | 8.532 | 16 | 0.9314 |
| AD Braak | chr18 | 77,924,739 | chr18:77924739:T:G | t | g | -0.2649 | 0.0639 | 3.35E-05 | ----------+--- | 8.429 | 16 | 0.935 |
| AD Braak | chr18 | 77,926,081 | chr18:77926081:C:G | c | g | -0.2664 | 0.064 | 3.16E-05 | ----------+--- | 8.602 | 16 | 0.9289 |
| AD Braak | chr18 | 77,927,188 | chr18:77927188:T:C | t | c | -0.265 | 0.0643 | 3.78E-05 | ----------+--- | 8.697 | 16 | 0.9254 |
| AD Braak | chr18 | 77,927,874 | chr18:77927874:G:A | a | g | 0.2649 | 0.0639 | 3.39E-05 | +++++++++ | 8.366 | 16 | 0.9372 |
| AD Braak | chr18 | 77,928,391 | chr18:77928391:G:A | a | g | 0.2591 | 0.0641 | 5.28E-05 | +++++++++ | 8.494 | 16 | 0.9328 |
| AD Braak | chr18 | 77,928,625 | chr18:77928625:A:AC | a | ac | -0.2612 | 0.0624 | 2.81E-05 | ----------+--- | 10.426 | 16 | 0.8434 |
| AD Braak | chr18 | 77,928,646 | chr18:77928646:A:C | a | c | -0.2576 | 0.0638 | 5.41E-05 | ----------+--- | 8.044 | 16 | 0.9475 |
| AD Braak | chr19 | 651,852 | chr19:651852:C:T | t | c | 0.2646 | 0.0606 | 1.24E-05 | ++++-+++?- | 5.692 | 14 | 0.9737 |
| AD Braak | chr19 | 8,403,601 | chr19:8403601:C:T | t | c | 0.1342 | 0.0344 | 9.56E-05 | ++++-+++-- | 16.384 | 15 | 0.357 |
| AD Braak | chr19 | 15,585,748 | chr19:15585748:G:A | a | g | -0.1088 | 0.0275 | 7.67E-05 | ------++----? | 17.779 | 15 | 0.2745 |
| AD Braak | chr19 | 16,745,204 | chr19:16745204:T:C | t | c | 0.1146 | 0.0278 | 3.82E-05 | +-+++++-++ | 10.605 | 14 | 0.7167 |
| AD Braak | chr19 | 32,224,971 | chr19:32224971:C:T | t | c | 0.1575 | 0.039 | 5.49E-05 | +-+++++++- | 15.644 | 16 | 0.4781 |
| AD Braak | chr19 | 32,228,552 | chr19:32228552:T:C | t | c | -0.1613 | 0.0391 | 3.74E-05 | -+----------+ | 14.626 | 16 | 0.5522 |
| AD Braak | chr19 | 41,729,928 | chr19:41729928:A:G | a | g | -0.3122 | 0.0792 | 8.00E-05 | ------+----+- | 11.251 | 16 | 0.7938 |
| AD Braak | chr19 | 41,737,071 | chr19:41737071:C:A | a | c | 0.3141 | 0.0792 | 7.35E-05 | ++++++-++- | 11.196 | 16 | 0.7972 |
| AD Braak | chr19 | 41,739,968 | chr19:41739968:C:A | a | c | 0.3089 | 0.0787 | 8.62E-05 | ++++++-++- | 10.724 | 16 | 0.8262 |
| AD Braak | chr19 | 41,739,970 | chr19:41739970:A:G | a | g | -0.3089 | 0.0787 | 8.61E-05 | ------+----+- | 10.724 | 16 | 0.8262 |
| AD Braak | chr19 | 41,739,972 | chr19:41739972:C:T | t | c | 0.3088 | 0.0787 | 8.69E-05 | ++++++-++- | 10.723 | 16 | 0.8262 |
| AD Braak | chr19 | 41,742,866 | chr19:41742866:A:G | a | g | -0.3154 | 0.0792 | 6.88E-05 | ------+----+- | 11.328 | 16 | 0.7888 |
| AD Braak | chr19 | 41,755,328 | chr19:41755328:G:A | a | g | 0.3102 | 0.0795 | 9.45E-05 | ++++++-++- | 11.821 | 16 | 0.7562 |
| AD Braak | chr19 | 44,728,555 | chr19:44728555:G:A | a | g | 0.2156 | 0.0451 | 1.75E-06 | ++++++--++ | 15.026 | 16 | 0.5227 |
| AD Braak | chr19 | 44,728,895 | chr19:44728895:G:A | a | g | 0.3561 | 0.0677 | 1.46E-07 | +++++++-+- | 18.382 | 16 | 0.302 |
| AD Braak | chr19 | 44,730,062 | chr19:44730062:AAATTTATAG:A | a | aaatttatag | 0.2144 | 0.045 | 1.93E-06 | ++++++--++ | 13.561 | 16 | 0.6314 |
| AD Braak | chr19 | 44,730,118 | chr19:44730118:A:G | a | g | -0.216 | 0.0452 | 1.73E-06 | ------++--+-- | 12.831 | 16 | 0.685 |
| AD Braak | chr19 | 44,732,593 | chr19:44732593:AAG:A | a | aag | 0.2144 | 0.0451 | 1.99E-06 | ++++++--++ | 13.446 | 16 | 0.6399 |
| AD Braak | chr19 | 44,733,473 | chr19:44733473:C:T | t | c | -0.1302 | 0.0307 | 2.20E-05 | -------------+ | 10.933 | 16 | 0.8136 |
| AD Braak | chr19 | 44,734,556 | chr19:44734556:T:C | t | c | -0.1077 | 0.0263 | 4.14E-05 | -------+-----+ | 14.603 | 16 | 0.5539 |
| AD Braak | chr19 | 44,735,929 | chr19:44735929:A:G | a | g | -0.1095 | 0.0265 | 3.50E-05 | -------+-----+ | 14.05 | 16 | 0.595 |
| AD Braak | chr19 | 44,736,280 | chr19:44736280:C:A | a | c | 0.2156 | 0.045 | 1.64E-06 | ++++-+--++ | 12.579 | 16 | 0.7032 |
| AD Braak | chr19 | 44,737,327 | chr19:44737327:T:C | t | c | 0.1432 | 0.0357 | 6.14E-05 | ++++++--++ | 22.882 | 16 | 0.1169 |
| AD Braak | chr19 | 44,737,991 | chr19:44737991:C:CA | ca | c | 0.4615 | 0.109 | 2.30E-05 | +-+++++++- | 7.196 | 15 | 0.952 |
| AD Braak | chr19 | 44,738,093 | chr19:44738093:T:C | t | c | -0.3598 | 0.0873 | 3.78E-05 | -+--+-+++-+ | 16.493 | 16 | 0.4191 |
| AD Braak | chr19 | 44,738,134 | chr19:44738134:T:C | t | c | -0.3542 | 0.0869 | 4.59E-05 | -+--+-+++-+ | 16.574 | 16 | 0.4136 |
| AD Braak | chr19 | 44,738,215 | chr19:44738215:A:T | a | t | -0.3554 | 0.0873 | 4.68E-05 | -+--+-+++-+ | 16.61 | 16 | 0.4112 |
| AD Braak | chr19 | 44,738,381 | chr19:44738381:G:A | a | g | -0.1419 | 0.0308 | 4.18E-06 | -------------+ | 10.693 | 16 | 0.828 |

| | | | | | | | | | | | | |
|---|---|---|---|---|---|---|---|---|---|---|---|---|
| AD Braak | chr19 | 44,738,783 | chr19:44738783:A:G | a | g | -0.3576 | 0.0872 | 4.09E-05 | ----+-+++-+- | 16.454 | 16 | 0.4218 |
| AD Braak | chr19 | 44,738,977 | chr19:44738977:G:A | a | g | 0.3655 | 0.0869 | 2.63E-05 | ++++-+---+- | 16.272 | 16 | 0.4342 |
| AD Braak | chr19 | 44,739,193 | chr19:44739193:A:G | a | g | -0.3587 | 0.087 | 3.76E-05 | ----+-+++-+- | 16.422 | 16 | 0.4239 |
| AD Braak | chr19 | 44,739,258 | chr19:44739258:T:G | t | g | -0.3595 | 0.087 | 3.62E-05 | ----+-+++-+- | 16.378 | 16 | 0.4269 |
| AD Braak | chr19 | 44,739,483 | chr19:44739483:A:G | a | g | -0.1177 | 0.0263 | 7.73E-06 | -------+-----+ | 13.259 | 16 | 0.6537 |
| AD Braak | chr19 | 44,739,520 | chr19:44739520:C:T | t | c | 0.3537 | 0.0874 | 5.15E-05 | +-++-+---+- | 19.896 | 16 | 0.2249 |
| AD Braak | chr19 | 44,739,813 | chr19:44739813:C:T | t | c | 0.3549 | 0.0872 | 4.67E-05 | ++++-+---+- | 16.041 | 16 | 0.4501 |
| AD Braak | chr19 | 44,740,307 | chr19:44740307:G:A | a | g | 0.3568 | 0.091 | 8.88E-05 | ++++-+--++ | 11.451 | 15 | 0.72 |
| AD Braak | chr19 | 44,741,295 | chr19:44741295:C:T | t | c | 0.3597 | 0.0864 | 3.13E-05 | ++++-+---+? | 19.33 | 15 | 0.1992 |
| AD Braak | chr19 | 44,742,164 | chr19:44742164:G:A | a | g | 0.35 | 0.0874 | 6.23E-05 | ++++-+--++ | 20.044 | 16 | 0.2182 |
| AD Braak | chr19 | 44,743,791 | chr19:44743791:C:T | t | c | 0.1299 | 0.0259 | 5.11E-07 | +++++++-+- | 16.829 | 16 | 0.3967 |
| AD Braak | chr19 | 44,746,404 | chr19:44746404:C:T | t | c | -0.1415 | 0.0277 | 3.20E-07 | -------------+ | 14.812 | 16 | 0.5384 |
| AD Braak | chr19 | 44,747,899 | chr19:44747899:A:C | a | c | -0.1263 | 0.0258 | 9.71E-07 | -------+-----+ | 17.293 | 16 | 0.3669 |
| AD Braak | chr19 | 44,748,549 | chr19:44748549:G:T | t | g | 0.4512 | 0.0998 | 6.11E-06 | +++++++++ | 7.666 | 14 | 0.906 |
| AD Braak | chr19 | 44,750,234 | chr19:44750234:C:T | t | c | 0.1662 | 0.0388 | 1.84E-05 | ++++++--++ | 17.159 | 16 | 0.3754 |
| AD Braak | chr19 | 44,750,354 | chr19:44750354:ATTGGC:A | a | attggc | 0.1662 | 0.0389 | 1.91E-05 | ++++++--++ | 14.86 | 16 | 0.5349 |
| AD Braak | chr19 | 44,750,911 | chr19:44750911:C:A | a | c | -0.1518 | 0.035 | 1.49E-05 | -+----++---+ | 20.484 | 16 | 0.1992 |
| AD Braak | chr19 | 44,753,536 | chr19:44753536:C:G | c | g | -0.4056 | 0.0935 | 1.44E-05 | ----+-++----- | 13.884 | 16 | 0.6074 |
| AD Braak | chr19 | 44,773,333 | chr19:44773333:GTTA:G | g | gtta | 0.3683 | 0.0916 | 5.78E-05 | +-+++++++- | 11.217 | 16 | 0.7959 |
| AD Braak | chr19 | 44,781,370 | chr19:44781370:G:A | a | g | -0.45 | 0.1032 | 1.30E-05 | --------?-+-- | 20.331 | 15 | 0.1596 |
| AD Braak | chr19 | 44,792,629 | chr19:44792629:G:A | a | g | 0.4683 | 0.0758 | 6.49E-10 | +-+++++-++ | 9.113 | 16 | 0.9087 |
| AD Braak | chr19 | 44,793,107 | chr19:44793107:G:A | a | g | 0.5258 | 0.1098 | 1.69E-06 | +-+++++++- | 9.778 | 15 | 0.8335 |
| AD Braak | chr19 | 44,793,549 | chr19:44793549:C:T | t | c | 0.4623 | 0.0753 | 8.50E-10 | +-+++++-++ | 9.189 | 16 | 0.9054 |
| AD Braak | chr19 | 44,795,204 | chr19:44795204:G:T | t | g | 0.4467 | 0.0779 | 9.97E-09 | +-+++++-++ | 8.396 | 16 | 0.9362 |
| AD Braak | chr19 | 44,795,942 | chr19:44795942:G:T | t | g | 0.4665 | 0.0779 | 2.15E-09 | +-+++++-++ | 9.316 | 16 | 0.8998 |
| AD Braak | chr19 | 44,797,922 | chr19:44797922:G:GC | g | gc | -0.1736 | 0.0358 | 1.21E-06 | -------+-++-- | 9.072 | 16 | 0.9104 |
| AD Braak | chr19 | 44,798,169 | chr19:44798169:C:CCTG | cctg | c | 0.234 | 0.0434 | 6.96E-08 | +-+++++-++ | 17.587 | 16 | 0.3486 |
| AD Braak | chr19 | 44,798,197 | chr19:44798197:TTTTG:T | t | ttttg | 0.1739 | 0.0355 | 9.68E-07 | +++++++-+- | 12.528 | 16 | 0.7069 |
| AD Braak | chr19 | 44,798,667 | chr19:44798667:T:C | t | c | -0.174 | 0.0398 | 1.23E-05 | -------+--+-- | 7.523 | 16 | 0.9618 |
| AD Braak | chr19 | 44,798,685 | chr19:44798685:A:G | a | g | -0.176 | 0.0398 | 9.83E-06 | -------+--+-- | 7.641 | 16 | 0.9588 |
| AD Braak | chr19 | 44,799,020 | chr19:44799020:C:T | t | c | -0.5423 | 0.1263 | 1.75E-05 | -----??-??--- | 7.982 | 11 | 0.7149 |
| AD Braak | chr19 | 44,799,247 | chr19:44799247:G:A | a | g | 0.5825 | 0.104 | 2.15E-08 | ++++++-++- | 8.739 | 15 | 0.8908 |
| AD Braak | chr19 | 44,799,256 | chr19:44799256:A:G | a | g | -0.1783 | 0.0398 | 7.42E-06 | -------+--+-- | 7.684 | 16 | 0.9577 |
| AD Braak | chr19 | 44,799,521 | chr19:44799521:A:G | a | g | -0.4113 | 0.0741 | 2.89E-08 | -------+--+-- | 9.579 | 16 | 0.8877 |
| AD Braak | chr19 | 44,799,694 | chr19:44799694:A:G | a | g | -0.181 | 0.0403 | 7.25E-06 | -------+--+-- | 8.099 | 16 | 0.9459 |
| AD Braak | chr19 | 44,799,865 | chr19:44799865:AAAAG:A | a | aaaag | 0.476 | 0.078 | 1.04E-09 | +-+++++-++ | 11.46 | 16 | 0.7802 |
| AD Braak | chr19 | 44,800,054 | chr19:44800054:G:A | a | g | 0.4589 | 0.0776 | 3.39E-09 | +-+++++-++ | 10.067 | 16 | 0.8631 |
| AD Braak | chr19 | 44,802,273 | chr19:44802273:GA:G | g | ga | 0.4455 | 0.076 | 4.53E-09 | +-+++++-++ | 10.446 | 16 | 0.8423 |
| AD Braak | chr19 | 44,806,377 | chr19:44806377:G:A | a | g | 0.4819 | 0.0776 | 5.24E-10 | +++++++-+- | 11.709 | 16 | 0.7637 |
| AD Braak | chr19 | 44,806,778 | chr19:44806778:G:A | a | g | 0.4824 | 0.0775 | 4.91E-10 | +++++++-+- | 11.88 | 16 | 0.7522 |
| AD Braak | chr19 | 44,807,827 | chr19:44807827:G:A | a | g | -0.3003 | 0.0753 | 6.64E-05 | ----+-++---+ | 17.909 | 16 | 0.3292 |
| AD Braak | chr19 | 44,807,887 | chr19:44807887:C:A | a | c | 0.1692 | 0.0399 | 2.18E-05 | ++++-+++-+ | 18.701 | 16 | 0.2845 |
| AD Braak | chr19 | 44,808,042 | chr19:44808042:C:G | c | g | -0.4849 | 0.0778 | 4.67E-10 | -------+--+-- | 12.246 | 16 | 0.7269 |
| AD Braak | chr19 | 44,808,053 | chr19:44808053:T:G | t | g | -0.4848 | 0.0778 | 4.58E-10 | -------+--+-- | 12.241 | 16 | 0.7273 |
| AD Braak | chr19 | 44,810,458 | chr19:44810458:T:C | t | c | -0.1708 | 0.04 | 1.94E-05 | -------+-++-- | 9.1 | 16 | 0.9092 |
| AD Braak | chr19 | 44,812,188 | chr19:44812188:G:A | a | g | 0.468 | 0.0783 | 2.23E-09 | +-+++++-++ | 12.723 | 16 | 0.6929 |

| | | | | | | | | | | | | |
|---|---|---|---|---|---|---|---|---|---|---|---|---|
| AD Braak | chr19 | 44,812,964 | chr19:44812964:A:G | a | g | 0.4784 | 0.0896 | 9.21E-08 | +++++++++ | 10.064 | 15 | 0.8157 |
| AD Braak | chr19 | 44,813,331 | chr19:44813331:G:A | a | g | 0.4667 | 0.0784 | 2.61E-09 | +-+++++-++ | 11.123 | 16 | 0.8018 |
| AD Braak | chr19 | 44,815,094 | chr19:44815094:A:G | a | g | -0.4796 | 0.0789 | 1.21E-09 | -+-----+--+-- | 12.921 | 16 | 0.6785 |
| AD Braak | chr19 | 44,816,374 | chr19:44816374:G:A | a | g | 0.5137 | 0.0713 | 5.68E-13 | +++++++-+- | 17.884 | 16 | 0.3308 |
| AD Braak | chr19 | 44,820,881 | chr19:44820881:G:A | a | g | -0.2139 | 0.0265 | 6.20E-16 | ----+-++----- | 27.41 | 16 | 0.03715 |
| AD Braak | chr19 | 44,821,499 | chr19:44821499:C:G | c | g | 0.454 | 0.0729 | 4.71E-10 | ++++-++-++ | 15.398 | 16 | 0.4957 |
| AD Braak | chr19 | 44,822,481 | chr19:44822481:T:G | t | g | 0.1124 | 0.0277 | 4.96E-05 | ++-++++-++ | 17.113 | 16 | 0.3783 |
| AD Braak | chr19 | 44,822,960 | chr19:44822960:T:C | t | c | 0.1469 | 0.0254 | 7.08E-09 | ++++-+--++ | 17.447 | 16 | 0.3573 |
| AD Braak | chr19 | 44,823,407 | chr19:44823407:C:T | t | c | -0.1452 | 0.0254 | 1.03E-08 | ----+-++----- | 17.354 | 16 | 0.363 |
| AD Braak | chr19 | 44,824,052 | chr19:44824052:C:T | t | c | -0.1464 | 0.0254 | 7.88E-09 | ----+-++----- | 17.512 | 16 | 0.3532 |
| AD Braak | chr19 | 44,824,166 | chr19:44824166:G:T | t | g | -0.1458 | 0.0254 | 8.87E-09 | ----+-++----- | 16.815 | 16 | 0.3977 |
| AD Braak | chr19 | 44,824,202 | chr19:44824202:T:C | t | c | 0.1242 | 0.0273 | 5.31E-06 | ++++-++-++ | 16.393 | 16 | 0.4259 |
| AD Braak | chr19 | 44,825,110 | chr19:44825110:A:T | a | t | 0.1477 | 0.0253 | 5.55E-09 | ++++-+--++ | 18.114 | 16 | 0.3173 |
| AD Braak | chr19 | 44,825,122 | chr19:44825122:T:A | a | t | -0.1478 | 0.0254 | 5.62E-09 | ----+-++----- | 18.111 | 16 | 0.3174 |
| AD Braak | chr19 | 44,825,123 | chr19:44825123:C:A | a | c | -0.1486 | 0.0253 | 4.54E-09 | ----+-++----- | 18.16 | 16 | 0.3146 |
| AD Braak | chr19 | 44,825,957 | chr19:44825957:T:G | t | g | 0.1457 | 0.0251 | 6.14E-09 | ++++-+--++ | 18.054 | 16 | 0.3207 |
| AD Braak | chr19 | 44,826,087 | chr19:44826087:T:C | t | c | 0.1484 | 0.0254 | 5.15E-09 | ++++-+--++ | 18.201 | 16 | 0.3122 |
| AD Braak | chr19 | 44,827,852 | chr19:44827852:G:C | c | g | -0.5075 | 0.1144 | 9.21E-06 | -----??-??-- | 7.281 | 11 | 0.7759 |
| AD Braak | chr19 | 44,828,468 | chr19:44828468:C:T | t | c | -0.1625 | 0.0258 | 2.84E-10 | ----+-++----- | 22.881 | 16 | 0.117 |
| AD Braak | chr19 | 44,832,778 | chr19:44832778:T:C | t | c | 0.1451 | 0.0258 | 1.93E-08 | ++++-+--++ | 21.601 | 16 | 0.1565 |
| AD Braak | chr19 | 44,833,186 | chr19:44833186:G:A | a | g | -0.1451 | 0.0259 | 1.99E-08 | ----+-++----- | 22.235 | 16 | 0.1358 |
| AD Braak | chr19 | 44,833,992 | chr19:44833992:C:A | a | c | -0.1216 | 0.0256 | 1.98E-06 | ----+-++----- | 16.907 | 16 | 0.3916 |
| AD Braak | chr19 | 44,834,128 | chr19:44834128:G:C | c | g | -0.1453 | 0.0258 | 1.87E-08 | ----+-++----- | 22.146 | 16 | 0.1385 |
| AD Braak | chr19 | 44,834,247 | chr19:44834247:G:C | c | g | -0.1336 | 0.0256 | 1.87E-07 | ----+-++----- | 19.614 | 16 | 0.2381 |
| AD Braak | chr19 | 44,834,480 | chr19:44834480:C:T | t | c | -0.1294 | 0.0274 | 2.29E-06 | ------++--+-- | 16.533 | 16 | 0.4164 |
| AD Braak | chr19 | 44,834,606 | chr19:44834606:T:C | t | c | 0.1383 | 0.0272 | 3.81E-07 | ++++-+--++ | 19.125 | 16 | 0.2622 |
| AD Braak | chr19 | 44,834,661 | chr19:44834661:G:A | a | g | -0.4968 | 0.0566 | 1.74E-18 | ----+--+----- | 14.571 | 16 | 0.5563 |
| AD Braak | chr19 | 44,834,963 | chr19:44834963:A:G | a | g | 0.1296 | 0.0274 | 2.22E-06 | ++++++--++ | 16.68 | 16 | 0.4066 |
| AD Braak | chr19 | 44,834,993 | chr19:44834993:AT:A | a | at | -0.1446 | 0.0259 | 2.30E-08 | ----+-++----- | 21.562 | 16 | 0.1579 |
| AD Braak | chr19 | 44,835,236 | chr19:44835236:T:C | t | c | 0.1291 | 0.0274 | 2.41E-06 | ++++++--++ | 16.584 | 16 | 0.413 |
| AD Braak | chr19 | 44,835,266 | chr19:44835266:G:C | c | g | -0.1453 | 0.0258 | 1.86E-08 | ----+-++----- | 22.285 | 16 | 0.1342 |
| AD Braak | chr19 | 44,836,881 | chr19:44836881:G:A | a | g | -0.1423 | 0.0272 | 1.73E-07 | ----+-++--+- | 19.193 | 16 | 0.2588 |
| AD Braak | chr19 | 44,837,479 | chr19:44837479:G:A | a | g | -0.1451 | 0.0258 | 1.93E-08 | ----+-++----- | 22.144 | 16 | 0.1386 |
| AD Braak | chr19 | 44,838,502 | chr19:44838502:C:G | c | g | 0.1335 | 0.027 | 7.34E-07 | ++++-++-++ | 19.259 | 16 | 0.2555 |
| AD Braak | chr19 | 44,838,647 | chr19:44838647:G:A | a | g | -0.145 | 0.0259 | 2.03E-08 | ----+-++----- | 21.994 | 16 | 0.1434 |
| AD Braak | chr19 | 44,838,691 | chr19:44838691:A:G | a | g | 0.1294 | 0.0274 | 2.31E-06 | ++++++--++ | 15.937 | 16 | 0.4573 |
| AD Braak | chr19 | 44,838,750 | chr19:44838750:G:C | c | g | -0.1463 | 0.0259 | 1.65E-08 | ----+-++----- | 21.46 | 16 | 0.1615 |
| AD Braak | chr19 | 44,838,857 | chr19:44838857:A:G | a | g | 0.1287 | 0.0258 | 5.97E-07 | ++++-+--++ | 17.608 | 16 | 0.3473 |
| AD Braak | chr19 | 44,838,904 | chr19:44838904:G:A | a | g | -0.1409 | 0.0259 | 5.21E-08 | ----+-++----- | 20.575 | 16 | 0.1954 |
| AD Braak | chr19 | 44,838,984 | chr19:44838984:A:G | a | g | 0.133 | 0.0274 | 1.17E-06 | ++++-+--++ | 16.801 | 16 | 0.3986 |
| AD Braak | chr19 | 44,840,322 | chr19:44840322:G:A | a | g | 0.1539 | 0.0381 | 5.46E-05 | ++++--++++ | 21.934 | 16 | 0.1453 |
| AD Braak | chr19 | 44,841,465 | chr19:44841465:G:A | a | g | -0.4069 | 0.0916 | 8.87E-06 | -------------- | 6.471 | 16 | 0.9822 |
| AD Braak | chr19 | 44,842,026 | chr19:44842026:A:C | a | c | 0.4493 | 0.1123 | 6.34E-05 | ++++++??? | 8.702 | 11 | 0.6494 |
| AD Braak | chr19 | 44,842,366 | chr19:44842366:G:A | a | g | -0.1501 | 0.0261 | 9.40E-09 | ----+-++----- | 18.623 | 16 | 0.2887 |
| AD Braak | chr19 | 44,842,530 | chr19:44842530:A:T | a | t | -0.261 | 0.0567 | 4.11E-06 | -+----+-----+ | 9.968 | 16 | 0.8683 |
| AD Braak | chr19 | 44,843,409 | chr19:44843409:C:G | c | g | 0.5575 | 0.0996 | 2.21E-08 | +++++?++? | 26.29 | 14 | 0.02377 |

| | | | | | | | | | | | | |
|---|---|---|---|---|---|---|---|---|---|---|---|---|
| AD Braak | chr19 | 44,845,265 | chr19:44845265:G:T | t | g | -0.1494 | 0.0261 | 1.07E-08 | ----+-++----- | 18.457 | 16 | 0.2978 |
| AD Braak | chr19 | 44,848,259 | chr19:44848259:C:G | c | g | 0.4579 | 0.0529 | 4.91E-18 | ++++-++-++ | 17.06 | 16 | 0.3817 |
| AD Braak | chr19 | 44,849,230 | chr19:44849230:A:T | a | t | 0.1005 | 0.0248 | 4.94E-05 | ++++-+-+++ | 9.356 | 16 | 0.898 |
| AD Braak | chr19 | 44,850,981 | chr19:44850981:G:C | c | g | -0.1402 | 0.0339 | 3.59E-05 | ------++--+-- | 22.8 | 16 | 0.1192 |
| AD Braak | chr19 | 44,852,010 | chr19:44852010:T:A | a | t | -0.4912 | 0.1087 | 6.16E-06 | -------??-+-- | 8.226 | 13 | 0.8286 |
| AD Braak | chr19 | 44,852,031 | chr19:44852031:C:T | t | c | -0.4695 | 0.1156 | 4.90E-05 | -----?????+ | 7.809 | 10 | 0.6475 |
| AD Braak | chr19 | 44,852,464 | chr19:44852464:C:G | c | g | 0.1755 | 0.0269 | 6.86E-11 | ++++-+--++ | 16.489 | 16 | 0.4194 |
| AD Braak | chr19 | 44,852,884 | chr19:44852884:C:T | t | c | -0.141 | 0.0267 | 1.36E-07 | -----------+- | 12.348 | 16 | 0.7197 |
| AD Braak | chr19 | 44,853,207 | chr19:44853207:T:A | a | t | -0.1782 | 0.027 | 3.88E-11 | ----+-++----- | 16.505 | 16 | 0.4183 |
| AD Braak | chr19 | 44,853,746 | chr19:44853746:C:G | c | g | 0.1689 | 0.0266 | 2.04E-10 | ++++-+--++ | 15.974 | 16 | 0.4548 |
| AD Braak | chr19 | 44,854,034 | chr19:44854034:G:C | c | g | -0.1828 | 0.0269 | 1.07E-11 | ----+-++----- | 16.71 | 16 | 0.4046 |
| AD Braak | chr19 | 44,854,120 | chr19:44854120:T:C | t | c | 0.1823 | 0.0269 | 1.26E-11 | ++++-+--++ | 16.306 | 16 | 0.4318 |
| AD Braak | chr19 | 44,854,682 | chr19:44854682:A:G | a | g | 0.1665 | 0.0247 | 1.72E-11 | ++++++--++ | 13.816 | 16 | 0.6124 |
| AD Braak | chr19 | 44,854,769 | chr19:44854769:C:T | t | c | -0.4549 | 0.1062 | 1.82E-05 | -------??---- | 9.467 | 13 | 0.7368 |
| AD Braak | chr19 | 44,855,096 | chr19:44855096:T:C | t | c | 0.1679 | 0.0249 | 1.41E-11 | +++++++-+- | 13.762 | 16 | 0.6165 |
| AD Braak | chr19 | 44,856,329 | chr19:44856329:C:T | t | c | -0.1831 | 0.0269 | 1.04E-11 | ----+-++----- | 16.407 | 16 | 0.4249 |
| AD Braak | chr19 | 44,856,410 | chr19:44856410:A:G | a | g | 0.4652 | 0.1065 | 1.25E-05 | +++++++?? | 9.598 | 13 | 0.7264 |
| AD Braak | chr19 | 44,856,449 | chr19:44856449:G:A | a | g | -0.1852 | 0.0269 | 6.00E-12 | ----+-++----- | 15.457 | 16 | 0.4914 |
| AD Braak | chr19 | 44,856,688 | chr19:44856688:G:A | a | g | -0.52 | 0.1112 | 2.92E-06 | -----?-+??-- | 17.654 | 13 | 0.1711 |
| AD Braak | chr19 | 44,857,231 | chr19:44857231:T:G | t | g | 0.4347 | 0.1107 | 8.61E-05 | ++++++??? | 8.272 | 11 | 0.6888 |
| AD Braak | chr19 | 44,857,967 | chr19:44857967:G:A | a | g | 0.1146 | 0.0266 | 1.65E-05 | ++++-++-++ | 12.752 | 16 | 0.6908 |
| AD Braak | chr19 | 44,858,389 | chr19:44858389:A:G | a | g | -0.2673 | 0.0413 | 9.27E-11 | ------++----- | 30.197 | 16 | 0.01701 |
| AD Braak | chr19 | 44,860,135 | chr19:44860135:C:T | t | c | 0.1315 | 0.0337 | 9.33E-05 | ++++++++-- | 21.061 | 16 | 0.1762 |
| AD Braak | chr19 | 44,860,443 | chr19:44860443:A:G | a | g | -0.1395 | 0.0325 | 1.81E-05 | ---------+-+- | 20.437 | 16 | 0.2012 |
| AD Braak | chr19 | 44,860,563 | chr19:44860563:T:G | t | g | 0.5222 | 0.1151 | 5.72E-06 | +++++??+? | 8.331 | 11 | 0.6834 |
| AD Braak | chr19 | 44,862,190 | chr19:44862190:G:A | a | g | -0.4863 | 0.0703 | 4.66E-12 | ----+--+----- | 15.61 | 16 | 0.4805 |
| AD Braak | chr19 | 44,864,245 | chr19:44864245:G:C | c | g | 0.1315 | 0.0335 | 8.79E-05 | +++++++++ | 21.037 | 16 | 0.1771 |
| AD Braak | chr19 | 44,864,520 | chr19:44864520:C:T | t | c | 0.1751 | 0.0376 | 3.12E-06 | +++++++++ | 15.562 | 16 | 0.4839 |
| AD Braak | chr19 | 44,864,715 | chr19:44864715:G:C | c | g | 0.2166 | 0.0359 | 1.67E-09 | +++++++++ | 17.165 | 16 | 0.375 |
| AD Braak | chr19 | 44,864,753 | chr19:44864753:A:G | a | g | -0.1766 | 0.0341 | 2.18E-07 | -------------+ | 23.209 | 16 | 0.1083 |
| AD Braak | chr19 | 44,865,946 | chr19:44865946:G:A | a | g | 0.5805 | 0.0801 | 4.34E-13 | +++++++++ | 5.055 | 16 | 0.9955 |
| AD Braak | chr19 | 44,867,581 | chr19:44867581:G:A | a | g | 0.1357 | 0.033 | 3.81E-05 | +++++++++ | 18.746 | 16 | 0.282 |
| AD Braak | chr19 | 44,874,887 | chr19:44874887:C:T | t | c | -0.1996 | 0.0382 | 1.72E-07 | -------------+ | 13.716 | 16 | 0.6199 |
| AD Braak | chr19 | 44,876,259 | chr19:44876259:G:A | a | g | -0.2089 | 0.0383 | 4.78E-08 | -------------+ | 14.227 | 16 | 0.5818 |
| AD Braak | chr19 | 44,877,704 | chr19:44877704:C:A | a | c | -0.2089 | 0.0386 | 6.07E-08 | -------------+ | 14.814 | 16 | 0.5383 |
| AD Braak | chr19 | 44,877,713 | chr19:44877713:G:T | t | g | 0.1459 | 0.0248 | 4.20E-09 | +++++++++ | 21.408 | 16 | 0.1633 |
| AD Braak | chr19 | 44,878,777 | chr19:44878777:A:G | a | g | -0.2527 | 0.0245 | 7.67E-25 | -------------+ | 19.145 | 16 | 0.2612 |
| AD Braak | chr19 | 44,879,418 | chr19:44879418:G:A | a | g | 0.5699 | 0.068 | 5.43E-17 | +++++++-+- | 17.651 | 16 | 0.3447 |
| AD Braak | chr19 | 44,879,460 | chr19:44879460:G:A | a | g | 0.1415 | 0.0248 | 1.17E-08 | +++++++++ | 11.127 | 16 | 0.8016 |
| AD Braak | chr19 | 44,879,709 | chr19:44879709:C:T | t | c | 0.2058 | 0.0255 | 7.18E-16 | +++++++++ | 18.775 | 16 | 0.2805 |
| AD Braak | chr19 | 44,879,804 | chr19:44879804:G:T | t | g | 0.2043 | 0.0255 | 1.11E-15 | +++++++++ | 18.957 | 16 | 0.2709 |
| AD Braak | chr19 | 44,879,822 | chr19:44879822:T:C | t | c | 0.2131 | 0.0387 | 3.82E-08 | +++++++++ | 14.717 | 16 | 0.5455 |
| AD Braak | chr19 | 44,879,834 | chr19:44879834:A:AC | a | ac | 0.2172 | 0.0387 | 2.04E-08 | +++++++++ | 14.941 | 16 | 0.529 |
| AD Braak | chr19 | 44,879,858 | chr19:44879858:A:G | a | g | 0.2123 | 0.0386 | 3.92E-08 | +++++++++ | 15.37 | 16 | 0.4978 |
| AD Braak | chr19 | 44,879,882 | chr19:44879882:T:A | a | t | -0.2156 | 0.0386 | 2.42E-08 | -------------+ | 14.906 | 16 | 0.5315 |
| AD Braak | chr19 | 44,880,859 | chr19:44880859:G:A | a | g | 0.1312 | 0.0249 | 1.43E-07 | +++++++++ | 12.035 | 16 | 0.7416 |

| | | | | | | | | | | | | |
|---|---|---|---|---|---|---|---|---|---|---|---|---|
| AD Braak | chr19 | 44,881,148 | chr19:44881148:T:C | t | c | -0.2585 | 0.0387 | 2.30E-11 | ------+------ | 21.204 | 16 | 0.1708 |
| AD Braak | chr19 | 44,881,674 | chr19:44881674:G:A | a | g | -0.5082 | 0.0951 | 9.24E-08 | +-------?---- | 31.401 | 15 | 0.00776 |
| AD Braak | chr19 | 44,882,099 | chr19:44882099:C:A | a | c | -0.4878 | 0.0929 | 1.51E-07 | --------??--- | 14.368 | 14 | 0.4227 |
| AD Braak | chr19 | 44,882,502 | chr19:44882502:G:C | c | g | 0.2019 | 0.0259 | 6.71E-15 | +++++++-+- | 27.059 | 16 | 0.04083 |
| AD Braak | chr19 | 44,883,210 | chr19:44883210:G:GTAA | g | gtaa | 0.5812 | 0.0306 | 1.14E-80 | +++++++++ | 31.838 | 16 | 0.0105 |
| AD Braak | chr19 | 44,883,377 | chr19:44883377:C:T | t | c | -0.4605 | 0.0577 | 1.40E-15 | -------------- | 21.362 | 16 | 0.165 |
| AD Braak | chr19 | 44,883,777 | chr19:44883777:A:C | a | c | -0.3601 | 0.0651 | 3.16E-08 | ----------++- | 10.111 | 16 | 0.8608 |
| AD Braak | chr19 | 44,883,800 | chr19:44883800:A:G | a | g | -0.3602 | 0.0651 | 3.12E-08 | ----------++- | 10.122 | 16 | 0.8602 |
| AD Braak | chr19 | 44,884,202 | chr19:44884202:C:G | c | g | 0.5808 | 0.0305 | 7.43E-81 | +++++++++ | 30.785 | 16 | 0.01433 |
| AD Braak | chr19 | 44,884,339 | chr19:44884339:G:A | a | g | -0.5812 | 0.0305 | 4.86E-81 | -------------- | 30.453 | 16 | 0.01579 |
| AD Braak | chr19 | 44,884,873 | chr19:44884873:G:A | a | g | -0.5715 | 0.0303 | 2.51E-79 | -------------- | 28.935 | 16 | 0.02438 |
| AD Braak | chr19 | 44,885,243 | chr19:44885243:A:G | a | g | 0.5084 | 0.0272 | 8.69E-78 | +++++++++ | 34.943 | 16 | 0.004047 |
| AD Braak | chr19 | 44,885,967 | chr19:44885967:G:A | a | g | 0.2266 | 0.0486 | 3.21E-06 | ++++-++-++ | 18.884 | 16 | 0.2747 |
| AD Braak | chr19 | 44,886,339 | chr19:44886339:G:A | a | g | 0.5259 | 0.0773 | 9.95E-12 | +++++++++ | 7.687 | 16 | 0.9577 |
| AD Braak | chr19 | 44,887,076 | chr19:44887076:A:G | a | g | 0.5177 | 0.0272 | 5.87E-81 | +++++++++ | 35.292 | 16 | 0.003624 |
| AD Braak | chr19 | 44,888,997 | chr19:44888997:C:T | t | c | -0.6406 | 0.0288 | 1.63E-109 | -------------- | 40.915 | 16 | 0.000572 |
| AD Braak | chr19 | 44,889,415 | chr19:44889415:AT:A | a | at | 0.5533 | 0.0814 | 1.10E-11 | +++++++++ | 6.781 | 16 | 0.9773 |
| AD Braak | chr19 | 44,890,259 | chr19:44890259:C:T | t | c | -0.4673 | 0.0929 | 4.86E-07 | --------??--- | 12.291 | 14 | 0.5829 |
| AD Braak | chr19 | 44,890,954 | chr19:44890954:C:T | t | c | 0.2329 | 0.0467 | 6.17E-07 | +++++++-+- | 8.939 | 16 | 0.9159 |
| AD Braak | chr19 | 44,891,079 | chr19:44891079:T:C | t | c | 0.5926 | 0.0303 | 4.75E-85 | +++++++++ | 31.019 | 16 | 0.01338 |
| AD Braak | chr19 | 44,891,712 | chr19:44891712:T:G | t | g | 0.5238 | 0.0272 | 1.15E-82 | +++++++++ | 34.964 | 16 | 0.00402 |
| AD Braak | chr19 | 44,892,009 | chr19:44892009:G:A | a | g | -0.2928 | 0.0259 | 1.16E-29 | ------+------ | 34.659 | 16 | 0.004423 |
| AD Braak | chr19 | 44,892,073 | chr19:44892073:A:G | a | g | -0.1961 | 0.0258 | 2.90E-14 | -------+------ | 24.665 | 16 | 0.07595 |
| AD Braak | chr19 | 44,892,362 | chr19:44892362:A:G | a | g | 0.6 | 0.0302 | 1.35E-87 | +++++++++ | 30.881 | 16 | 0.01394 |
| AD Braak | chr19 | 44,892,457 | chr19:44892457:T:C | t | c | 0.5271 | 0.0271 | 4.80E-84 | +++++++++ | 35.705 | 16 | 0.003179 |
| AD Braak | chr19 | 44,892,587 | chr19:44892587:G:A | a | g | -0.522 | 0.0332 | 1.01E-55 | -------+------ | 21.273 | 16 | 0.1682 |
| AD Braak | chr19 | 44,892,652 | chr19:44892652:C:G | c | g | 0.6019 | 0.0304 | 2.85E-87 | +++++++++ | 30.756 | 16 | 0.01446 |
| AD Braak | chr19 | 44,892,887 | chr19:44892887:C:T | t | c | -0.6027 | 0.0304 | 1.80E-87 | -------------- | 30.936 | 16 | 0.01371 |
| AD Braak | chr19 | 44,892,962 | chr19:44892962:C:T | t | c | -0.5303 | 0.0271 | 5.10E-85 | -------------- | 34.399 | 16 | 0.004799 |
| AD Braak | chr19 | 44,893,408 | chr19:44893408:G:T | t | g | -0.5455 | 0.0274 | 5.72E-88 | -------------- | 33.946 | 16 | 0.005524 |
| AD Braak | chr19 | 44,893,642 | chr19:44893642:T:C | t | c | -0.1555 | 0.0245 | 2.38E-10 | -------+------ | 15.935 | 16 | 0.4575 |
| AD Braak | chr19 | 44,893,716 | chr19:44893716:G:A | a | g | -0.4596 | 0.0573 | 1.03E-15 | -------------- | 22.49 | 16 | 0.1281 |
| AD Braak | chr19 | 44,893,972 | chr19:44893972:G:A | a | g | 0.579 | 0.082 | 1.63E-12 | +++++++++ | 7.605 | 16 | 0.9598 |
| AD Braak | chr19 | 44,894,050 | chr19:44894050:C:T | t | c | -0.4628 | 0.0573 | 6.44E-16 | -------------- | 22.444 | 16 | 0.1294 |
| AD Braak | chr19 | 44,894,255 | chr19:44894255:A:C | a | c | -0.1712 | 0.0246 | 3.39E-12 | -------+------ | 17.627 | 16 | 0.3462 |
| AD Braak | chr19 | 44,894,695 | chr19:44894695:T:C | t | c | 0.4586 | 0.0571 | 9.83E-16 | +++++++++ | 21.304 | 16 | 0.1671 |
| AD Braak | chr19 | 44,895,007 | chr19:44895007:C:T | t | c | 0.1698 | 0.0246 | 5.05E-12 | +++++++-+- | 17.275 | 16 | 0.368 |
| AD Braak | chr19 | 44,895,376 | chr19:44895376:G:C | c | g | 0.2719 | 0.0355 | 1.94E-14 | ++++++-++- | 21.616 | 16 | 0.156 |
| AD Braak | chr19 | 44,895,459 | chr19:44895459:A:C | a | c | -0.1742 | 0.0246 | 1.50E-12 | -------+------ | 17.147 | 16 | 0.3762 |
| AD Braak | chr19 | 44,895,528 | chr19:44895528:C:T | t | c | -0.4669 | 0.0573 | 3.80E-16 | -------------- | 21.974 | 16 | 0.144 |
| AD Braak | chr19 | 44,895,560 | chr19:44895560:C:T | t | c | 0.23 | 0.0257 | 3.85E-19 | +++++++++ | 26.271 | 16 | 0.05033 |
| AD Braak | chr19 | 44,896,087 | chr19:44896087:G:T | t | g | -0.4669 | 0.0573 | 3.69E-16 | -------------- | 21.978 | 16 | 0.1439 |
| AD Braak | chr19 | 44,896,639 | chr19:44896639:G:A | a | g | -0.4671 | 0.0573 | 3.79E-16 | -------------- | 21.935 | 16 | 0.1453 |
| AD Braak | chr19 | 44,897,227 | chr19:44897227:T:TG | t | tg | 0.4663 | 0.0573 | 4.06E-16 | +++++++++ | 22.204 | 16 | 0.1367 |
| AD Braak | chr19 | 44,897,468 | chr19:44897468:C:T | t | c | -0.4669 | 0.0573 | 3.71E-16 | -------------- | 21.977 | 16 | 0.1439 |
| AD Braak | chr19 | 44,897,490 | chr19:44897490:T:A | a | t | 0.5847 | 0.0818 | 8.61E-13 | +++++++++ | 7.806 | 16 | 0.9544 |

| | | | | | | | | | | | | |
|---|---|---|---|---|---|---|---|---|---|---|---|---|
| AD Braak | chr19 | 44,898,409 | chr19:44898409:A:G | a | g | -0.2353 | 0.0255 | 2.56E-20 | -------------- | 24.617 | 16 | 0.07688 |
| AD Braak | chr19 | 44,898,611 | chr19:44898611:T:C | t | c | 0.3752 | 0.051 | 1.97E-13 | ++++++-++- | 18.686 | 16 | 0.2853 |
| AD Braak | chr19 | 44,899,005 | chr19:44899005:T:G | t | g | -0.5882 | 0.0817 | 6.16E-13 | ---------+--- | 7.52 | 16 | 0.9619 |
| AD Braak | chr19 | 44,899,220 | chr19:44899220:C:T | t | c | 0.2403 | 0.0257 | 9.42E-21 | +++++++++ | 24.294 | 16 | 0.08328 |
| AD Braak | chr19 | 44,899,959 | chr19:44899959:C:T | t | c | -0.4782 | 0.0557 | 9.16E-18 | ------------+ | 24.702 | 16 | 0.07526 |
| AD Braak | chr19 | 44,900,155 | chr19:44900155:C:T | t | c | 0.2821 | 0.025 | 1.64E-29 | +++++++++ | 22.418 | 16 | 0.1302 |
| AD Braak | chr19 | 44,900,601 | chr19:44900601:A:G | a | g | -0.2857 | 0.0252 | 9.95E-30 | -------------- | 22.593 | 16 | 0.1251 |
| AD Braak | chr19 | 44,901,174 | chr19:44901174:T:C | t | c | -0.2863 | 0.0253 | 8.57E-30 | -------------- | 22.851 | 16 | 0.1178 |
| AD Braak | chr19 | 44,901,434 | chr19:44901434:A:G | a | g | 0.2212 | 0.0288 | 1.66E-14 | ++++-++++- | 18.073 | 16 | 0.3196 |
| AD Braak | chr19 | 44,901,600 | chr19:44901600:T:C | t | c | 0.4802 | 0.0557 | 6.71E-18 | +++++++++ | 25.073 | 16 | 0.06854 |
| AD Braak | chr19 | 44,901,715 | chr19:44901715:T:C | t | c | -0.2863 | 0.0253 | 8.58E-30 | -------------- | 22.485 | 16 | 0.1282 |
| AD Braak | chr19 | 44,901,805 | chr19:44901805:A:G | a | g | -0.2848 | 0.0252 | 1.50E-29 | -------------- | 22.787 | 16 | 0.1195 |
| AD Braak | chr19 | 44,902,264 | chr19:44902264:G:C | c | g | 0.2383 | 0.0257 | 1.97E-20 | +++++++++ | 24.588 | 16 | 0.07743 |
| AD Braak | chr19 | 44,903,281 | chr19:44903281:C:CG | cg | c | 0.2843 | 0.0255 | 6.31E-29 | +++++++++ | 21.444 | 16 | 0.1621 |
| AD Braak | chr19 | 44,903,416 | chr19:44903416:G:A | a | g | -0.4618 | 0.0261 | 4.25E-70 | -------------- | 28.321 | 16 | 0.02894 |
| AD Braak | chr19 | 44,904,531 | chr19:44904531:G:A | a | g | 0.2896 | 0.0253 | 2.34E-30 | +++++++++ | 22.804 | 16 | 0.1191 |
| AD Braak | chr19 | 44,905,307 | chr19:44905307:A:T | a | t | -0.271 | 0.0359 | 4.36E-14 | ------+------- | 21.4 | 16 | 0.1636 |
| AD Braak | chr19 | 44,905,579 | chr19:44905579:T:G | t | g | -0.2527 | 0.0245 | 5.68E-25 | -------+------ | 18.673 | 16 | 0.286 |
| AD Braak | chr19 | 44,905,910 | chr19:44905910:C:G | c | g | 0.2307 | 0.0264 | 2.75E-18 | ++++-++++- | 21.743 | 16 | 0.1517 |
| AD Braak | chr19 | 44,906,745 | chr19:44906745:G:A | a | g | -0.7163 | 0.0316 | 5.06E-114 | -------------- | 36.071 | 16 | 0.002828 |
| AD Braak | chr19 | 44,907,187 | chr19:44907187:G:A | a | g | 0.2506 | 0.0259 | 4.06E-22 | +++++++++ | 24.971 | 16 | 0.07034 |
| AD Braak | chr19 | 44,908,684 | chr19:44908684:T:C | t | c | 0.7616 | 0.0295 | 8.06E-147 | +++++++++ | 45.18 | 16 | 0.00013 |
| AD Braak | chr19 | 44,908,822 | chr19:44908822:C:T | t | c | 0.6512 | 0.0535 | 4.94E-34 | +++++++-+- | 22.393 | 16 | 0.1309 |
| AD Braak | chr19 | 44,909,698 | chr19:44909698:A:C | a | c | 0.4812 | 0.0556 | 5.13E-18 | +++++++++ | 23.171 | 16 | 0.1092 |
| AD Braak | chr19 | 44,909,976 | chr19:44909976:G:T | t | g | 0.6525 | 0.0537 | 5.71E-34 | +++++++-+- | 22.253 | 16 | 0.1352 |
| AD Braak | chr19 | 44,910,109 | chr19:44910109:T:C | t | c | -0.281 | 0.0483 | 5.99E-09 | -------+------ | 10.025 | 16 | 0.8653 |
| AD Braak | chr19 | 44,910,319 | chr19:44910319:C:T | t | c | -0.4206 | 0.0284 | 1.34E-49 | -------------- | 15.316 | 16 | 0.5016 |
| AD Braak | chr19 | 44,911,194 | chr19:44911194:T:C | t | c | 0.2174 | 0.0264 | 1.75E-16 | ++++-+-++- | 22.067 | 16 | 0.141 |
| AD Braak | chr19 | 44,912,456 | chr19:44912456:G:A | a | g | -0.7197 | 0.0316 | 1.31E-114 | -------------- | 35.272 | 16 | 0.003647 |
| AD Braak | chr19 | 44,912,678 | chr19:44912678:G:T | t | g | -0.7179 | 0.0316 | 3.17E-114 | -------------- | 35.603 | 16 | 0.003284 |
| AD Braak | chr19 | 44,912,921 | chr19:44912921:G:T | t | g | -0.4983 | 0.0271 | 1.29E-75 | -------------- | 24.208 | 16 | 0.08506 |
| AD Braak | chr19 | 44,913,034 | chr19:44913034:C:T | t | c | 0.27 | 0.0263 | 1.08E-24 | +++++++++ | 29.045 | 16 | 0.02364 |
| AD Braak | chr19 | 44,913,221 | chr19:44913221:A:G | a | g | 0.2185 | 0.0267 | 2.54E-16 | ++++-+-++- | 22.839 | 16 | 0.1181 |
| AD Braak | chr19 | 44,913,484 | chr19:44913484:C:T | t | c | -0.4999 | 0.0271 | 4.37E-76 | -------------- | 23.871 | 16 | 0.09237 |
| AD Braak | chr19 | 44,915,229 | chr19:44915229:G:A | a | g | -0.5011 | 0.0683 | 2.21E-13 | ----+--+----- | 16.363 | 16 | 0.4279 |
| AD Braak | chr19 | 44,915,533 | chr19:44915533:T:C | t | c | 0.4956 | 0.0272 | 2.13E-74 | +++++++++ | 23.447 | 16 | 0.1023 |
| AD Braak | chr19 | 44,915,704 | chr19:44915704:T:C | t | c | 0.2171 | 0.0267 | 3.94E-16 | ++++-+-++- | 19.685 | 16 | 0.2347 |
| AD Braak | chr19 | 44,916,825 | chr19:44916825:A:C | a | c | 0.685 | 0.0313 | 3.72E-106 | +++++++-+- | 32.107 | 16 | 0.009684 |
| AD Braak | chr19 | 44,917,843 | chr19:44917843:G:A | a | g | 0.276 | 0.0265 | 2.37E-25 | +++++++++ | 23.372 | 16 | 0.1041 |
| AD Braak | chr19 | 44,917,947 | chr19:44917947:C:T | t | c | -0.4549 | 0.0553 | 1.88E-16 | ------------+ | 21.305 | 16 | 0.1671 |
| AD Braak | chr19 | 44,917,997 | chr19:44917997:G:A | a | g | -0.6143 | 0.0303 | 2.13E-91 | -------+------ | 30.957 | 16 | 0.01363 |
| AD Braak | chr19 | 44,918,393 | chr19:44918393:G:A | a | g | -0.4548 | 0.0552 | 1.84E-16 | ------------+ | 20.974 | 16 | 0.1795 |
| AD Braak | chr19 | 44,918,487 | chr19:44918487:G:T | t | g | 0.2517 | 0.0278 | 1.27E-19 | +++++++++ | 29.308 | 16 | 0.02194 |
| AD Braak | chr19 | 44,918,620 | chr19:44918620:A:G | a | g | 0.2689 | 0.0264 | 2.50E-24 | ++++-++-+- | 20.474 | 16 | 0.1996 |
| AD Braak | chr19 | 44,918,715 | chr19:44918715:AG:A | a | ag | 0.2529 | 0.0278 | 9.09E-20 | +++++++++ | 29.17 | 16 | 0.02282 |
| AD Braak | chr19 | 44,918,903 | chr19:44918903:C:G | c | g | 0.6589 | 0.0285 | 5.25E-118 | +++++++-+- | 34.65 | 16 | 0.004436 |

| | | | | | | | | | | | | |
|---|---|---|---|---|---|---|---|---|---|---|---|---|
| AD Braak | chr19 | 44,919,589 | chr19:44919589:G:A | a | g | -0.6566 | 0.0285 | 2.98E-117 | -------+------ | 34.253 | 16 | 0.005022 |
| AD Braak | chr19 | 44,919,689 | chr19:44919689:A:G | a | g | 0.6566 | 0.0283 | 7.27E-119 | +++++++-+- | 32.836 | 16 | 0.00777 |
| AD Braak | chr19 | 44,920,379 | chr19:44920379:G:A | a | g | 0.2442 | 0.0279 | 2.01E-18 | +++++++++ | 27.109 | 16 | 0.04029 |
| AD Braak | chr19 | 44,920,687 | chr19:44920687:C:T | t | c | 0.5495 | 0.1046 | 1.50E-07 | ++++-++-++ | 24.346 | 16 | 0.08222 |
| AD Braak | chr19 | 44,921,809 | chr19:44921809:G:A | a | g | -0.4449 | 0.0571 | 6.63E-15 | -------------- | 22.117 | 16 | 0.1395 |
| AD Braak | chr19 | 44,921,918 | chr19:44921918:T:G | t | g | 0.2687 | 0.0267 | 7.21E-24 | ++++-++-+- | 19.176 | 16 | 0.2596 |
| AD Braak | chr19 | 44,921,921 | chr19:44921921:G:A | a | g | 0.6113 | 0.0659 | 1.67E-20 | +++++++-+- | 20.718 | 16 | 0.1896 |
| AD Braak | chr19 | 44,922,203 | chr19:44922203:A:G | a | g | 0.269 | 0.0267 | 7.22E-24 | ++++-++-+- | 19.302 | 16 | 0.2533 |
| AD Braak | chr19 | 44,923,535 | chr19:44923535:G:A | a | g | 0.6034 | 0.0656 | 3.54E-20 | +++++++-+- | 19.491 | 16 | 0.244 |
| AD Braak | chr19 | 44,923,868 | chr19:44923868:T:A | a | t | -0.6227 | 0.0306 | 3.54E-92 | -------+------ | 31.938 | 16 | 0.01019 |
| AD Braak | chr19 | 44,924,096 | chr19:44924096:C:G | c | g | -0.2463 | 0.0273 | 1.63E-19 | -------------- | 35.386 | 16 | 0.003518 |
| AD Braak | chr19 | 44,924,977 | chr19:44924977:G:A | a | g | -0.6184 | 0.0306 | 4.71E-91 | -------+------ | 31.907 | 16 | 0.01028 |
| AD Braak | chr19 | 44,925,202 | chr19:44925202:C:T | t | c | 0.2542 | 0.0283 | 2.31E-19 | +++++++++ | 28.387 | 16 | 0.02841 |
| AD Braak | chr19 | 44,926,286 | chr19:44926286:C:T | t | c | 0.2064 | 0.0289 | 9.24E-13 | +++++++++ | 19.651 | 16 | 0.2363 |
| AD Braak | chr19 | 44,926,451 | chr19:44926451:G:C | c | g | -0.2239 | 0.0423 | 1.20E-07 | -------+-+--- | 18.59 | 16 | 0.2905 |
| AD Braak | chr19 | 44,928,401 | chr19:44928401:A:G | a | g | -0.138 | 0.0267 | 2.22E-07 | ----------+--- | 9.084 | 16 | 0.9099 |
| AD Braak | chr19 | 44,932,959 | chr19:44932959:A:G | a | g | -0.1265 | 0.0275 | 4.04E-06 | -------------- | 10.452 | 16 | 0.842 |
| AD Braak | chr19 | 44,932,987 | chr19:44932987:T:C | t | c | -0.1264 | 0.0274 | 4.08E-06 | -------------- | 10.483 | 16 | 0.8402 |
| AD Braak | chr19 | 44,932,990 | chr19:44932990:T:C | t | c | -0.1263 | 0.0274 | 4.15E-06 | -------------- | 10.496 | 16 | 0.8395 |
| AD Braak | chr19 | 44,935,297 | chr19:44935297:C:T | t | c | -0.2226 | 0.028 | 1.71E-15 | -------------- | 9.979 | 16 | 0.8677 |
| AD Braak | chr19 | 44,943,964 | chr19:44943964:G:A | a | g | 0.4624 | 0.0965 | 1.66E-06 | +++++++++ | 15.094 | 15 | 0.4447 |
| AD Braak | chr19 | 44,945,208 | chr19:44945208:T:G | t | g | 0.0996 | 0.0252 | 7.92E-05 | +-+-+++-++ | 24.967 | 16 | 0.07041 |
| AD Braak | chr19 | 45,013,455 | chr19:45013455:T:TAC | t | tac | 0.1729 | 0.0433 | 6.66E-05 | +++++++-+- | 15.335 | 16 | 0.5002 |
| AD Braak | chr19 | 45,017,701 | chr19:45017701:G:T | t | g | -0.3297 | 0.0785 | 2.69E-05 | ----+--+----- | 25.369 | 16 | 0.06358 |
| AD Braak | chr19 | 45,019,031 | chr19:45019031:G:A | a | g | -0.3624 | 0.0921 | 8.39E-05 | ---+---+-+--- | 25.632 | 16 | 0.05943 |
| AD Braak | chr19 | 45,019,803 | chr19:45019803:G:C | c | g | -0.373 | 0.0856 | 1.30E-05 | ----+-++--+- | 13.418 | 16 | 0.642 |
| AD Braak | chr19 | 45,020,861 | chr19:45020861:C:T | t | c | -0.3288 | 0.0779 | 2.46E-05 | ----+--+-+--- | 16.162 | 16 | 0.4417 |
| AD Braak | chr19 | 45,027,093 | chr19:45027093:C:T | t | c | 0.1123 | 0.0258 | 1.33E-05 | +-+++-++++ | 28.113 | 16 | 0.03065 |
| AD Braak | chr19 | 45,040,497 | chr19:45040497:A:G | a | g | 0.3169 | 0.0671 | 2.31E-06 | +++++++++ | 15.551 | 16 | 0.4847 |
| AD Braak | chr19 | 45,045,877 | chr19:45045877:G:A | a | g | -0.3987 | 0.0756 | 1.32E-07 | ---------++-- | 13.488 | 16 | 0.6368 |
| AD Braak | chr19 | 45,059,984 | chr19:45059984:C:T | t | c | -0.4932 | 0.1035 | 1.87E-06 | ---------?+-- | 10.244 | 15 | 0.8041 |
| AD Braak | chr19 | 45,078,695 | chr19:45078695:G:C | c | g | -0.3355 | 0.073 | 4.32E-06 | ---------+---- | 21.054 | 16 | 0.1764 |
| AD Braak | chr19 | 45,086,866 | chr19:45086866:G:A | a | g | -0.4236 | 0.099 | 1.90E-05 | ------+--?+-- | 10.8 | 15 | 0.7667 |
| AD Braak | chr19 | 45,087,826 | chr19:45087826:C:T | t | c | -0.3129 | 0.0702 | 8.30E-06 | -------------- | 13.749 | 16 | 0.6174 |
| AD Braak | chr19 | 45,088,980 | chr19:45088980:C:T | t | c | -0.1722 | 0.0435 | 7.48E-05 | ----+--+---+- | 18.616 | 16 | 0.2891 |
| AD Braak | chr19 | 45,102,867 | chr19:45102867:C:T | t | c | -0.2002 | 0.0397 | 4.51E-07 | ----+--------+ | 10.856 | 16 | 0.8183 |
| AD Braak | chr19 | 45,108,968 | chr19:45108968:C:T | t | c | -0.1954 | 0.0397 | 8.30E-07 | ----++------- | 11.584 | 16 | 0.7721 |
| AD Braak | chr19 | 45,111,343 | chr19:45111343:C:T | t | c | -0.1957 | 0.0396 | 7.93E-07 | ----++------- | 11.138 | 16 | 0.8009 |
| AD Braak | chr19 | 45,115,701 | chr19:45115701:G:A | a | g | -0.1976 | 0.0396 | 6.02E-07 | ----++------- | 10.981 | 16 | 0.8107 |
| AD Braak | chr19 | 45,118,499 | chr19:45118499:TG:T | t | tg | -0.1928 | 0.0397 | 1.22E-06 | ----++------- | 11.047 | 16 | 0.8066 |
| AD Braak | chr19 | 45,123,977 | chr19:45123977:C:T | t | c | -0.1876 | 0.0396 | 2.14E-06 | ----++------- | 11.065 | 16 | 0.8055 |
| AD Braak | chr19 | 45,128,558 | chr19:45128558:G:A | a | g | -0.1899 | 0.0394 | 1.46E-06 | ----++------- | 10.834 | 16 | 0.8196 |
| AD Braak | chr19 | 45,130,428 | chr19:45130428:G:A | a | g | -0.1956 | 0.0395 | 7.52E-07 | ----++------- | 11.192 | 16 | 0.7975 |
| AD Braak | chr19 | 45,132,943 | chr19:45132943:C:T | t | c | -0.1967 | 0.0387 | 3.67E-07 | ----++------- | 12.302 | 16 | 0.7229 |
| AD Braak | chr19 | 45,133,061 | chr19:45133061:C:A | a | c | -0.1941 | 0.0396 | 9.82E-07 | ----++------- | 11.69 | 16 | 0.765 |
| AD Braak | chr19 | 45,134,987 | chr19:45134987:C:T | t | c | -0.1949 | 0.0396 | 8.78E-07 | ----++------- | 11.763 | 16 | 0.7601 |

| | | | | | | | | | | | | |
|---|---|---|---|---|---|---|---|---|---|---|---|---|
| AD Braak | chr19 | 45,137,848 | chr19:45137848:C:A | a | c | -0.192 | 0.0392 | 9.81E-07 | ----++------- | 13.222 | 16 | 0.6565 |
| AD Braak | chr19 | 45,138,786 | chr19:45138786:G:A | a | g | -0.1955 | 0.0396 | 8.15E-07 | ----++------- | 11.735 | 16 | 0.762 |
| AD Braak | chr19 | 45,139,287 | chr19:45139287:C:G | c | g | 0.1965 | 0.0396 | 6.85E-07 | ++++--++++ | 11.936 | 16 | 0.7484 |
| AD Braak | chr19 | 45,141,096 | chr19:45141096:A:T | a | t | 0.1968 | 0.0396 | 6.59E-07 | ++++--++++ | 11.691 | 16 | 0.765 |
| AD Braak | chr19 | 45,146,841 | chr19:45146841:G:T | t | g | -0.1917 | 0.0395 | 1.21E-06 | ----++------- | 12.067 | 16 | 0.7393 |
| AD Braak | chr19 | 45,147,128 | chr19:45147128:C:T | t | c | -0.1907 | 0.0394 | 1.28E-06 | ----++------- | 12.246 | 16 | 0.7269 |
| AD Braak | chr19 | 45,147,979 | chr19:45147979:G:A | a | g | -0.1929 | 0.0395 | 1.06E-06 | ----++------- | 12.217 | 16 | 0.7289 |
| AD Braak | chr19 | 45,150,625 | chr19:45150625:TAGTC:T | t | tagtc | 0.4159 | 0.0968 | 1.73E-05 | +++++++-+- | 23.185 | 15 | 0.08029 |
| AD Braak | chr19 | 45,152,075 | chr19:45152075:T:C | t | c | 0.1897 | 0.0394 | 1.51E-06 | ++++--++++ | 13.369 | 16 | 0.6456 |
| AD Braak | chr19 | 45,204,659 | chr19:45204659:T:G | t | g | 0.1408 | 0.0325 | 1.45E-05 | +-+++++++- | 22.707 | 16 | 0.1218 |
| AD Braak | chr19 | 45,204,981 | chr19:45204981:A:G | a | g | 0.1404 | 0.0326 | 1.62E-05 | +-+++++++- | 22.774 | 16 | 0.1199 |
| AD Braak | chr19 | 45,205,150 | chr19:45205150:T:C | t | c | 0.1404 | 0.0326 | 1.61E-05 | +-+++++++- | 22.806 | 16 | 0.119 |
| AD Braak | chr19 | 45,205,166 | chr19:45205166:TTG:T | t | ttg | -0.1415 | 0.0326 | 1.42E-05 | -+----------- | 22.681 | 16 | 0.1225 |
| AD Braak | chr19 | 45,205,500 | chr19:45205500:A:G | a | g | 0.142 | 0.0323 | 1.11E-05 | +-+++++++- | 22.532 | 16 | 0.1268 |
| AD Braak | chr19 | 45,205,630 | chr19:45205630:T:C | t | c | 0.1427 | 0.0324 | 1.08E-05 | +-+++++++- | 22.393 | 16 | 0.131 |
| AD Braak | chr19 | 45,205,689 | chr19:45205689:G:A | a | g | -0.1426 | 0.0326 | 1.22E-05 | -+----------- | 22.666 | 16 | 0.1229 |
| AD Braak | chr19 | 45,205,778 | chr19:45205778:C:T | t | c | -0.1426 | 0.0326 | 1.22E-05 | -+----------- | 22.668 | 16 | 0.1229 |
| AD Braak | chr19 | 45,206,729 | chr19:45206729:A:G | a | g | 0.1424 | 0.0326 | 1.26E-05 | +-+++++++- | 22.773 | 16 | 0.1199 |
| AD Braak | chr19 | 45,207,018 | chr19:45207018:A:G | a | g | 0.1426 | 0.0326 | 1.22E-05 | +-+++++++- | 22.622 | 16 | 0.1242 |
| AD Braak | chr19 | 45,207,281 | chr19:45207281:C:T | t | c | -0.1434 | 0.0327 | 1.14E-05 | -+----------- | 22.525 | 16 | 0.127 |
| AD Braak | chr19 | 45,207,605 | chr19:45207605:C:T | t | c | -0.1429 | 0.0326 | 1.19E-05 | -+----------- | 22.502 | 16 | 0.1277 |
| AD Braak | chr19 | 45,208,340 | chr19:45208340:C:T | t | c | -0.1417 | 0.0326 | 1.34E-05 | -+----------- | 20.697 | 16 | 0.1904 |
| AD Braak | chr19 | 45,209,045 | chr19:45209045:GT:G | g | gt | -0.1411 | 0.0343 | 3.78E-05 | ----+-------- | 15.694 | 16 | 0.4745 |
| AD Braak | chr2 | 13,857,338 | chr2:13857338:G:A | a | g | -0.3934 | 0.1 | 8.34E-05 | ----+----++-- | 10.383 | 16 | 0.8459 |
| AD Braak | chr2 | 13,858,926 | chr2:13858926:A:G | a | g | 0.405 | 0.0998 | 4.95E-05 | ++++-++++- | 10.33 | 16 | 0.8489 |
| AD Braak | chr2 | 22,235,362 | chr2:22235362:T:A | a | t | -0.7479 | 0.1868 | 6.25E-05 | ?--????--?? | 2.274 | 5 | 0.8101 |
| AD Braak | chr2 | 38,420,849 | chr2:38420849:A:G | a | g | -0.7281 | 0.1812 | 5.84E-05 | --?????-??? | 4.348 | 5 | 0.5004 |
| AD Braak | chr2 | 40,240,280 | chr2:40240280:C:T | t | c | 0.2862 | 0.0705 | 4.92E-05 | ++++-++++- | 12.662 | 16 | 0.6973 |
| AD Braak | chr2 | 46,024,649 | chr2:46024649:T:C | t | c | -0.294 | 0.0705 | 3.09E-05 | ---+-+---+-- | 20.255 | 16 | 0.209 |
| AD Braak | chr2 | 46,028,227 | chr2:46028227:C:G | c | g | -0.2906 | 0.0703 | 3.55E-05 | ---+-+-+----- | 22.474 | 16 | 0.1285 |
| AD Braak | chr2 | 46,029,315 | chr2:46029315:C:G | c | g | -0.2955 | 0.0707 | 2.93E-05 | ---+-+------- | 21.529 | 16 | 0.1591 |
| AD Braak | chr2 | 51,759,174 | chr2:51759174:C:T | t | c | -0.103 | 0.0264 | 9.68E-05 | --------+-++- | 28.132 | 16 | 0.03049 |
| AD Braak | chr2 | 55,769,824 | chr2:55769824:T:A | a | t | -0.0983 | 0.0241 | 4.51E-05 | ----------+++ | 10.017 | 16 | 0.8657 |
| AD Braak | chr2 | 55,770,511 | chr2:55770511:A:G | a | g | 0.0967 | 0.0242 | 6.53E-05 | +++++++-+- | 13.76 | 16 | 0.6166 |
| AD Braak | chr2 | 55,775,269 | chr2:55775269:G:A | a | g | -0.0981 | 0.0242 | 5.06E-05 | -------+--+-+ | 13.768 | 16 | 0.616 |
| AD Braak | chr2 | 55,777,004 | chr2:55777004:A:G | a | g | 0.0966 | 0.0242 | 6.60E-05 | +++++++-+- | 13.492 | 16 | 0.6365 |
| AD Braak | chr2 | 56,432,689 | chr2:56432689:G:C | c | g | 0.4165 | 0.088 | 2.19E-06 | +++++++++ | 8.993 | 15 | 0.8779 |
| AD Braak | chr2 | 56,462,732 | chr2:56462732:A:T | a | t | -0.4235 | 0.0876 | 1.35E-06 | ----------+--- | 9.746 | 15 | 0.8354 |
| AD Braak | chr2 | 56,533,659 | chr2:56533659:A:G | a | g | -0.4194 | 0.0872 | 1.50E-06 | ----------+--- | 11.636 | 15 | 0.7063 |
| AD Braak | chr2 | 56,553,150 | chr2:56553150:C:T | t | c | 0.4239 | 0.0876 | 1.30E-06 | +++++++++ | 12.346 | 15 | 0.6527 |
| AD Braak | chr2 | 56,556,504 | chr2:56556504:A:G | a | g | -0.4276 | 0.0875 | 1.02E-06 | -+--------+-- | 12.304 | 15 | 0.6559 |
| AD Braak | chr2 | 56,574,563 | chr2:56574563:G:T | t | g | 0.4063 | 0.0817 | 6.63E-07 | +++++++-+- | 8.408 | 16 | 0.9358 |
| AD Braak | chr2 | 56,577,372 | chr2:56577372:G:C | c | g | 0.4219 | 0.0874 | 1.39E-06 | +-+++++++- | 12.561 | 15 | 0.6362 |
| AD Braak | chr2 | 56,579,781 | chr2:56579781:T:C | t | c | -0.4245 | 0.0872 | 1.14E-06 | -+--------+-- | 12.981 | 15 | 0.6038 |
| AD Braak | chr2 | 56,590,059 | chr2:56590059:G:A | a | g | 0.4203 | 0.0873 | 1.48E-06 | +-+++++++- | 12.571 | 15 | 0.6354 |
| AD Braak | chr2 | 56,614,631 | chr2:56614631:G:A | a | g | 0.4145 | 0.0871 | 1.92E-06 | +-+++++++- | 12.933 | 15 | 0.6075 |

| | | | | | | | | | | | | |
|---|---|---|---|---|---|---|---|---|---|---|---|---|
| AD Braak | chr2 | 56,631,220 | chr2:56631220:G:A | a | g | 0.4074 | 0.0867 | 2.64E-06 | +-++++++++ | 12.934 | 15 | 0.6074 |
| AD Braak | chr2 | 56,663,462 | chr2:56663462:T:C | t | c | -0.4092 | 0.0871 | 2.60E-06 | -+--------+-- | 12.909 | 15 | 0.6094 |
| AD Braak | chr2 | 56,732,590 | chr2:56732590:G:T | t | g | 0.4119 | 0.0874 | 2.45E-06 | +-++++++++ | 13.606 | 15 | 0.5556 |
| AD Braak | chr2 | 58,291,600 | chr2:58291600:C:T | t | c | 0.4034 | 0.1032 | 9.23E-05 | -+++-++++? | 26.235 | 13 | 0.01581 |
| AD Braak | chr2 | 75,439,080 | chr2:75439080:G:T | t | g | -0.1351 | 0.0341 | 7.53E-05 | -----+-----+- | 11.327 | 16 | 0.7889 |
| AD Braak | chr2 | 75,440,148 | chr2:75440148:G:T | t | g | -0.1355 | 0.0343 | 7.83E-05 | -----+-----+- | 11.495 | 16 | 0.778 |
| AD Braak | chr2 | 76,941,261 | chr2:76941261:A:G | a | g | -0.4288 | 0.1037 | 3.56E-05 | ----+----+--- | 16.465 | 16 | 0.421 |
| AD Braak | chr2 | 95,588,124 | chr2:95588124:A:G | a | g | -0.5853 | 0.1413 | 3.46E-05 | --------?-?-? | 10.124 | 12 | 0.6051 |
| AD Braak | chr2 | 103,962,075 | chr2:103962075:T:C | t | c | -0.1383 | 0.0326 | 2.21E-05 | -----+-+----+ | 17.234 | 16 | 0.3706 |
| AD Braak | chr2 | 103,964,045 | chr2:103964045:C:T | t | c | 0.1311 | 0.0326 | 5.77E-05 | +++++-+-++ | 17.229 | 16 | 0.3709 |
| AD Braak | chr2 | 103,970,893 | chr2:103970893:T:A | a | t | 0.1304 | 0.0327 | 6.58E-05 | +++++-+-++ | 17.025 | 16 | 0.384 |
| AD Braak | chr2 | 103,972,680 | chr2:103972680:G:A | a | g | 0.1297 | 0.0326 | 6.94E-05 | +++++-+-++ | 17.054 | 16 | 0.3821 |
| AD Braak | chr2 | 127,063,876 | chr2:127063876:C:T | t | c | -0.118 | 0.0299 | 7.86E-05 | -+--++---+-- | 9.821 | 16 | 0.8758 |
| AD Braak | chr2 | 127,064,584 | chr2:127064584:C:T | t | c | -0.1174 | 0.0299 | 8.64E-05 | -+--++---+-- | 9.986 | 16 | 0.8674 |
| AD Braak | chr2 | 127,066,110 | chr2:127066110:A:G | a | g | 0.1183 | 0.0299 | 7.53E-05 | +-++--+++-- | 9.529 | 16 | 0.8901 |
| AD Braak | chr2 | 127,068,683 | chr2:127068683:A:G | a | g | -0.1213 | 0.0303 | 6.38E-05 | -+----++-++- | 22.863 | 16 | 0.1174 |
| AD Braak | chr2 | 127,068,957 | chr2:127068957:A:G | a | g | 0.1361 | 0.0261 | 1.76E-07 | +++++-+++- | 17.508 | 16 | 0.3535 |
| AD Braak | chr2 | 127,069,257 | chr2:127069257:G:A | a | g | -0.1164 | 0.0299 | 9.84E-05 | -+--++---+-- | 10.555 | 16 | 0.8361 |
| AD Braak | chr2 | 127,081,898 | chr2:127081898:G:A | a | g | -0.1181 | 0.0257 | 4.46E-06 | -----------+- | 12.158 | 16 | 0.733 |
| AD Braak | chr2 | 127,082,205 | chr2:127082205:C:A | a | c | -0.1235 | 0.0255 | 1.28E-06 | -----------+- | 11.846 | 16 | 0.7545 |
| AD Braak | chr2 | 127,085,031 | chr2:127085031:A:G | a | g | 0.1012 | 0.0248 | 4.43E-05 | +++++-+++- | 8.907 | 16 | 0.9172 |
| AD Braak | chr2 | 127,086,585 | chr2:127086585:G:A | a | g | -0.0973 | 0.0245 | 7.39E-05 | ----++--++-- | 18.867 | 16 | 0.2756 |
| AD Braak | chr2 | 127,090,354 | chr2:127090354:G:A | a | g | -0.1278 | 0.0253 | 4.14E-07 | ----++---+--- | 17.466 | 16 | 0.3561 |
| AD Braak | chr2 | 127,104,557 | chr2:127104557:C:T | t | c | -0.1048 | 0.0245 | 1.83E-05 | ----++---+--- | 20.519 | 16 | 0.1978 |
| AD Braak | chr2 | 127,105,077 | chr2:127105077:T:G | t | g | 0.1139 | 0.0291 | 9.08E-05 | +++-+-+++- | 28.355 | 16 | 0.02867 |
| AD Braak | chr2 | 127,106,970 | chr2:127106970:G:A | a | g | -0.1222 | 0.0307 | 6.75E-05 | -------+-+--- | 8.423 | 16 | 0.9353 |
| AD Braak | chr2 | 127,107,524 | chr2:127107524:G:A | a | g | -0.1001 | 0.0256 | 9.31E-05 | ---------++-- | 20.54 | 16 | 0.1969 |
| AD Braak | chr2 | 127,107,528 | chr2:127107528:C:T | t | c | -0.1226 | 0.0307 | 6.39E-05 | -------+-+--- | 8.503 | 16 | 0.9325 |
| AD Braak | chr2 | 127,111,157 | chr2:127111157:C:T | t | c | 0.1219 | 0.0309 | 8.15E-05 | +-+++-+-+-- | 36.883 | 16 | 0.002178 |
| AD Braak | chr2 | 127,111,491 | chr2:127111491:G:A | a | g | 0.1206 | 0.031 | 9.88E-05 | +-+++-+-+-- | 36.753 | 16 | 0.002272 |
| AD Braak | chr2 | 127,111,790 | chr2:127111790:A:G | a | g | 0.1221 | 0.0307 | 6.96E-05 | +++++++-+- | 8.391 | 16 | 0.9364 |
| AD Braak | chr2 | 127,115,459 | chr2:127115459:T:C | t | c | 0.115 | 0.0292 | 8.32E-05 | +++-+-+++- | 32.24 | 16 | 0.009304 |
| AD Braak | chr2 | 127,116,759 | chr2:127116759:T:C | t | c | 0.123 | 0.0307 | 6.01E-05 | +++++++-+- | 8.448 | 16 | 0.9344 |
| AD Braak | chr2 | 127,117,813 | chr2:127117813:T:G | t | g | 0.1151 | 0.0293 | 8.60E-05 | +++-+-+++- | 31.685 | 16 | 0.01099 |
| AD Braak | chr2 | 127,118,084 | chr2:127118084:A:G | a | g | 0.1146 | 0.0293 | 9.28E-05 | +++-+-+++- | 31.789 | 16 | 0.01065 |
| AD Braak | chr2 | 127,120,512 | chr2:127120512:G:A | a | g | -0.124 | 0.0307 | 5.36E-05 | -------+-+--- | 8.714 | 16 | 0.9247 |
| AD Braak | chr2 | 127,121,263 | chr2:127121263:T:C | t | c | 0.1147 | 0.0292 | 8.71E-05 | +++-+-+++- | 32.035 | 16 | 0.009896 |
| AD Braak | chr2 | 127,123,357 | chr2:127123357:T:A | a | t | 0.122 | 0.0311 | 8.50E-05 | +++++-+-+- | 37.115 | 16 | 0.00202 |
| AD Braak | chr2 | 127,123,667 | chr2:127123667:G:A | a | g | 0.1224 | 0.0311 | 8.08E-05 | +++++-+-+- | 37.339 | 16 | 0.001878 |
| AD Braak | chr2 | 127,124,606 | chr2:127124606:T:C | t | c | 0.1159 | 0.0251 | 4.04E-06 | +++++++-+- | 13.261 | 16 | 0.6536 |
| AD Braak | chr2 | 127,124,766 | chr2:127124766:T:C | t | c | 0.1274 | 0.0307 | 3.31E-05 | +++++++-+- | 8.911 | 16 | 0.917 |
| AD Braak | chr2 | 127,125,710 | chr2:127125710:A:G | a | g | 0.1285 | 0.0307 | 2.81E-05 | +++++++-+- | 8.723 | 16 | 0.9244 |
| AD Braak | chr2 | 127,126,329 | chr2:127126329:A:C | a | c | 0.1286 | 0.0307 | 2.77E-05 | +++++++-+- | 8.732 | 16 | 0.9241 |
| AD Braak | chr2 | 127,126,547 | chr2:127126547:A:C | a | c | -0.1224 | 0.0311 | 8.23E-05 | -----+-+-++- | 37.127 | 16 | 0.002012 |
| AD Braak | chr2 | 127,127,671 | chr2:127127671:A:G | a | g | -0.1063 | 0.0272 | 9.39E-05 | ---+--------+ | 26.323 | 16 | 0.04965 |
| AD Braak | chr2 | 127,127,783 | chr2:127127783:A:T | a | t | 0.1273 | 0.0307 | 3.38E-05 | +++++++-+- | 8.634 | 16 | 0.9277 |

| | | | | | | | | | | | | |
|---|---|---|---|---|---|---|---|---|---|---|---|---|
| AD Braak | chr2 | 127,128,048 | chr2:127128048:T:A | a | t | -0.1271 | 0.0307 | 3.47E-05 | -------+-+--- | 8.461 | 16 | 0.9339 |
| AD Braak | chr2 | 127,128,075 | chr2:127128075:G:A | a | g | 0.1068 | 0.0272 | 8.74E-05 | +++-+++++- | 23.679 | 16 | 0.09675 |
| AD Braak | chr2 | 127,128,582 | chr2:127128582:C:T | t | c | -0.1074 | 0.0265 | 5.01E-05 | ---------++-- | 14.835 | 16 | 0.5367 |
| AD Braak | chr2 | 127,128,657 | chr2:127128657:C:T | t | c | -0.1071 | 0.0264 | 5.11E-05 | ---------++-- | 14.818 | 16 | 0.538 |
| AD Braak | chr2 | 127,128,795 | chr2:127128795:T:C | t | c | 0.1073 | 0.0265 | 5.10E-05 | +++++++++ | 14.592 | 16 | 0.5547 |
| AD Braak | chr2 | 127,128,840 | chr2:127128840:A:C | a | c | 0.1068 | 0.0265 | 5.55E-05 | +++++++++ | 14.685 | 16 | 0.5478 |
| AD Braak | chr2 | 127,128,970 | chr2:127128970:AAAG:A | a | aaag | -0.1064 | 0.0265 | 5.94E-05 | ---------++-- | 14.839 | 16 | 0.5365 |
| AD Braak | chr2 | 127,130,289 | chr2:127130289:T:C | t | c | 0.1265 | 0.0307 | 3.75E-05 | ++++--+-+-- | 8.846 | 16 | 0.9196 |
| AD Braak | chr2 | 127,130,545 | chr2:127130545:C:T | t | c | -0.1273 | 0.0307 | 3.31E-05 | ----++-+-+-- | 8.785 | 16 | 0.922 |
| AD Braak | chr2 | 127,131,181 | chr2:127131181:A:C | a | c | 0.1346 | 0.0253 | 1.05E-07 | ++++--+++- | 17.024 | 16 | 0.3841 |
| AD Braak | chr2 | 127,131,208 | chr2:127131208:C:G | c | g | 0.1262 | 0.0307 | 3.91E-05 | +++++-+-+- | 8.951 | 16 | 0.9154 |
| AD Braak | chr2 | 127,131,777 | chr2:127131777:GAA:G | g | gaa | -0.1257 | 0.0307 | 4.26E-05 | ----++-+-+-- | 8.721 | 16 | 0.9245 |
| AD Braak | chr2 | 127,132,061 | chr2:127132061:G:A | a | g | -0.134 | 0.0252 | 1.09E-07 | ----++---+-- | 16.773 | 16 | 0.4005 |
| AD Braak | chr2 | 127,132,677 | chr2:127132677:G:T | t | g | -0.1299 | 0.0309 | 2.60E-05 | ----++-+-+-- | 7.913 | 16 | 0.9514 |
| AD Braak | chr2 | 127,132,740 | chr2:127132740:C:A | a | c | -0.124 | 0.0307 | 5.39E-05 | ----++-+-+-- | 8.948 | 16 | 0.9155 |
| AD Braak | chr2 | 127,133,210 | chr2:127133210:G:C | c | g | -0.1223 | 0.0308 | 7.03E-05 | ----++-+-+-- | 10.14 | 16 | 0.8592 |
| AD Braak | chr2 | 127,133,302 | chr2:127133302:G:C | c | g | -0.1189 | 0.0305 | 9.98E-05 | ----++-+-+-- | 9.315 | 16 | 0.8999 |
| AD Braak | chr2 | 127,133,540 | chr2:127133540:T:C | t | c | 0.1277 | 0.0309 | 3.68E-05 | ++++--+-+-- | 9.848 | 16 | 0.8745 |
| AD Braak | chr2 | 127,133,631 | chr2:127133631:C:G | c | g | 0.1255 | 0.0308 | 4.69E-05 | ++++--+-+-- | 9.948 | 16 | 0.8693 |
| AD Braak | chr2 | 127,133,750 | chr2:127133750:G:A | a | g | -0.126 | 0.0309 | 4.51E-05 | ----++-+-+-- | 10.722 | 16 | 0.8263 |
| AD Braak | chr2 | 127,133,780 | chr2:127133780:G:A | a | g | -0.1269 | 0.0309 | 4.03E-05 | ----++-+-+-- | 10.669 | 16 | 0.8294 |
| AD Braak | chr2 | 127,133,851 | chr2:127133851:A:C | a | c | 0.1742 | 0.0247 | 1.71E-12 | +++++++-+- | 10.987 | 16 | 0.8103 |
| AD Braak | chr2 | 127,133,935 | chr2:127133935:G:A | a | g | -0.1274 | 0.0309 | 3.70E-05 | ----++-+-+-- | 10.628 | 16 | 0.8319 |
| AD Braak | chr2 | 127,134,047 | chr2:127134047:C:T | t | c | -0.1263 | 0.0309 | 4.30E-05 | ----++-+-+-- | 10.007 | 16 | 0.8663 |
| AD Braak | chr2 | 127,134,087 | chr2:127134087:T:C | t | c | 0.1264 | 0.0309 | 4.24E-05 | ++++--+-+-- | 10.023 | 16 | 0.8654 |
| AD Braak | chr2 | 127,134,219 | chr2:127134219:A:G | a | g | 0.1258 | 0.0309 | 4.62E-05 | ++++--+-+-- | 9.91 | 16 | 0.8713 |
| AD Braak | chr2 | 127,135,192 | chr2:127135192:G:T | t | g | -0.1471 | 0.0265 | 2.78E-08 | -------+------ | 14.358 | 16 | 0.5721 |
| AD Braak | chr2 | 127,135,234 | chr2:127135234:C:T | t | c | -0.1942 | 0.025 | 7.22E-15 | ----+--+----- | 11.573 | 16 | 0.7728 |
| AD Braak | chr2 | 127,135,380 | chr2:127135380:C:G | c | g | 0.1267 | 0.031 | 4.34E-05 | ++++--+-+-- | 9.16 | 16 | 0.9067 |
| AD Braak | chr2 | 127,135,449 | chr2:127135449:C:A | a | c | -0.128 | 0.031 | 3.64E-05 | ----++-+-+-- | 9.009 | 16 | 0.9131 |
| AD Braak | chr2 | 127,136,522 | chr2:127136522:G:A | a | g | -0.1501 | 0.0265 | 1.55E-08 | -------+------ | 14.866 | 16 | 0.5345 |
| AD Braak | chr2 | 127,136,908 | chr2:127136908:A:T | a | t | 0.1397 | 0.0264 | 1.21E-07 | ++++-+-++- | 12.077 | 16 | 0.7386 |
| AD Braak | chr2 | 127,137,039 | chr2:127137039:A:G | a | g | 0.1411 | 0.0262 | 7.15E-08 | ++++-+-++- | 12.062 | 16 | 0.7397 |
| AD Braak | chr2 | 144,456,071 | chr2:144456071:G:C | c | g | -0.1744 | 0.0448 | 9.97E-05 | -+----++--+- | 14.311 | 16 | 0.5755 |
| AD Braak | chr2 | 149,662,689 | chr2:149662689:A:G | a | g | -0.3852 | 0.097 | 7.19E-05 | +---------?-- | 15.488 | 15 | 0.4168 |
| AD Braak | chr2 | 149,678,048 | chr2:149678048:C:T | t | c | 0.4318 | 0.1055 | 4.25E-05 | +++++++++ | 15.331 | 15 | 0.4278 |
| AD Braak | chr2 | 149,718,386 | chr2:149718386:A:G | a | g | -0.4095 | 0.0996 | 3.94E-05 | ----------?--+ | 12.133 | 15 | 0.669 |
| AD Braak | chr2 | 149,721,361 | chr2:149721361:AT:A | a | at | 0.4014 | 0.0993 | 5.26E-05 | +++++++++ | 12.444 | 15 | 0.6452 |
| AD Braak | chr2 | 169,564,675 | chr2:169564675:A:AT | a | at | -0.4618 | 0.1181 | 9.19E-05 | -+--?---??-- | 7.906 | 12 | 0.7924 |
| AD Braak | chr2 | 169,577,669 | chr2:169577669:C:T | t | c | 0.4811 | 0.119 | 5.25E-05 | ++++?+++? | 6.399 | 12 | 0.8946 |
| AD Braak | chr2 | 216,227,750 | chr2:216227750:CGGGAGAGGGA | cgggagagg | c | 0.2913 | 0.0709 | 4.00E-05 | +++++++++ | 13.143 | 15 | 0.5912 |
| AD Braak | chr2 | 236,694,356 | chr2:236694356:G:A | a | g | 0.4617 | 0.1172 | 8.21E-05 | +++++++++ | 7.155 | 13 | 0.894 |
| AD Braak | chr20 | 723,135 | chr20:723135:T:C | t | c | -0.7381 | 0.1874 | 8.17E-05 | --??+-???-? | 2.298 | 6 | 0.8904 |
| AD Braak | chr20 | 6,437,048 | chr20:6437048:C:T | t | c | -0.0959 | 0.0246 | 9.69E-05 | ----+-----?-- | 14.668 | 15 | 0.4756 |
| AD Braak | chr20 | 6,437,155 | chr20:6437155:C:A | a | c | -0.0968 | 0.0246 | 8.35E-05 | ----+-----?-- | 14.455 | 15 | 0.4913 |
| AD Braak | chr20 | 6,437,541 | chr20:6437541:G:A | a | g | -0.0978 | 0.0246 | 7.11E-05 | ----+-----?-- | 15.022 | 15 | 0.4499 |

| | | | | | | | | | | | | |
|---|---|---|---|---|---|---|---|---|---|---|---|---|
| AD Braak | chr20 | 6,437,998 | chr20:6437998:C:T | t | c | -0.0978 | 0.0246 | 6.84E-05 | ----+--+--?-- | 14.063 | 15 | 0.5207 |
| AD Braak | chr20 | 6,438,129 | chr20:6438129:A:G | a | g | 0.0976 | 0.0246 | 7.09E-05 | ++++-++++- | 14.021 | 15 | 0.5239 |
| AD Braak | chr20 | 6,438,167 | chr20:6438167:T:G | t | g | 0.0972 | 0.0246 | 7.65E-05 | ++++-++++- | 13.952 | 15 | 0.5292 |
| AD Braak | chr20 | 6,438,653 | chr20:6438653:A:G | a | g | 0.0989 | 0.0246 | 5.65E-05 | ++++-++++- | 13.784 | 15 | 0.542 |
| AD Braak | chr20 | 6,438,673 | chr20:6438673:A:G | a | g | 0.0989 | 0.0246 | 5.67E-05 | ++++-++++- | 13.796 | 15 | 0.5411 |
| AD Braak | chr20 | 6,438,731 | chr20:6438731:A:G | a | g | 0.0991 | 0.0246 | 5.47E-05 | ++++-++++- | 13.784 | 15 | 0.542 |
| AD Braak | chr20 | 6,440,526 | chr20:6440526:T:G | t | g | 0.0959 | 0.0246 | 9.58E-05 | ++++-++++- | 13.563 | 15 | 0.5589 |
| AD Braak | chr20 | 6,481,955 | chr20:6481955:G:C | c | g | -0.097 | 0.0246 | 7.96E-05 | ----+-----?+- | 13.209 | 15 | 0.5862 |
| AD Braak | chr20 | 16,749,834 | chr20:16749834:C:T | t | c | -0.1331 | 0.026 | 3.03E-07 | ----+-----?+- | 15.288 | 15 | 0.4308 |
| AD Braak | chr20 | 16,750,455 | chr20:16750455:T:C | t | c | 0.1177 | 0.0265 | 8.95E-06 | +-+++++++- | 18.704 | 15 | 0.2275 |
| AD Braak | chr20 | 18,034,282 | chr20:18034282:C:T | t | c | 0.5018 | 0.1248 | 5.80E-05 | +++++++-? | 16.986 | 13 | 0.2 |
| AD Braak | chr20 | 18,382,533 | chr20:18382533:C:T | t | c | 0.2655 | 0.0639 | 3.29E-05 | +-+-++++++ | 23.978 | 15 | 0.06547 |
| AD Braak | chr20 | 19,073,410 | chr20:19073410:T:G | t | g | -0.5958 | 0.1293 | 4.07E-06 | -??----?-+?- | 3.763 | 10 | 0.9574 |
| AD Braak | chr20 | 32,450,354 | chr20:32450354:T:C | t | c | -0.1268 | 0.032 | 7.45E-05 | ----+++---++ | 19.782 | 16 | 0.2302 |
| AD Braak | chr20 | 35,857,220 | chr20:35857220:G:A | a | g | -0.1322 | 0.0339 | 9.53E-05 | ----+------+- | 5.337 | 16 | 0.9938 |
| AD Braak | chr20 | 35,859,787 | chr20:35859787:T:C | t | c | 0.1324 | 0.0339 | 9.39E-05 | ++++-++++- | 5.296 | 16 | 0.9941 |
| AD Braak | chr20 | 35,860,888 | chr20:35860888:G:A | a | g | -0.1322 | 0.0339 | 9.51E-05 | ----+------+- | 5.294 | 16 | 0.9941 |
| AD Braak | chr20 | 35,868,782 | chr20:35868782:A:G | a | g | 0.1328 | 0.0339 | 8.95E-05 | ++++-++++- | 5.307 | 16 | 0.994 |
| AD Braak | chr20 | 35,873,105 | chr20:35873105:A:AT | a | at | 0.1393 | 0.0344 | 5.21E-05 | ++++-++++- | 5.476 | 16 | 0.9928 |
| AD Braak | chr20 | 35,876,463 | chr20:35876463:C:T | t | c | -0.135 | 0.0343 | 8.25E-05 | ----+------+- | 4.711 | 16 | 0.997 |
| AD Braak | chr20 | 35,895,859 | chr20:35895859:A:G | a | g | 0.1356 | 0.0343 | 7.59E-05 | ++++-++++- | 4.729 | 16 | 0.9969 |
| AD Braak | chr20 | 35,924,684 | chr20:35924684:C:G | c | g | 0.1367 | 0.0342 | 6.42E-05 | ++++-++++- | 4.997 | 16 | 0.9958 |
| AD Braak | chr20 | 35,930,670 | chr20:35930670:C:T | t | c | -0.137 | 0.0342 | 6.22E-05 | ----+------+- | 4.942 | 16 | 0.996 |
| AD Braak | chr20 | 35,933,443 | chr20:35933443:C:T | t | c | -0.1373 | 0.0342 | 6.00E-05 | ----+------+- | 4.969 | 16 | 0.9959 |
| AD Braak | chr20 | 35,959,317 | chr20:35959317:ATATTAT:A | a | atattat | -0.1374 | 0.0335 | 4.18E-05 | ----+------+- | 8.619 | 16 | 0.9283 |
| AD Braak | chr20 | 42,947,985 | chr20:42947985:T:TTGAA | t | ttgaa | 1.0473 | 0.2479 | 2.39E-05 | +???++++? | 3.243 | 5 | 0.6625 |
| AD Braak | chr20 | 43,645,731 | chr20:43645731:G:A | a | g | 0.2948 | 0.074 | 6.73E-05 | +++++-+++- | 19.841 | 15 | 0.1781 |
| AD Braak | chr21 | 13,555,301 | chr21:13555301:C:T | t | c | -0.1271 | 0.0323 | 8.47E-05 | ----------?--- | 6.958 | 14 | 0.9363 |
| AD Braak | chr21 | 13,556,060 | chr21:13556060:A:G | a | g | 0.1277 | 0.0323 | 7.91E-05 | +++++++++ | 7.04 | 14 | 0.9331 |
| AD Braak | chr21 | 13,559,391 | chr21:13559391:A:C | a | c | 0.1246 | 0.032 | 9.69E-05 | +++++++++ | 7.246 | 14 | 0.9248 |
| AD Braak | chr21 | 13,559,714 | chr21:13559714:G:A | a | g | -0.1246 | 0.032 | 9.91E-05 | ----------?--- | 7.25 | 14 | 0.9246 |
| AD Braak | chr21 | 16,828,919 | chr21:16828919:C:T | t | c | 0.508 | 0.1261 | 5.62E-05 | +?++??+-? | 6.411 | 8 | 0.6013 |
| AD Braak | chr21 | 26,265,385 | chr21:26265385:G:A | a | g | -0.1102 | 0.0248 | 8.54E-06 | ----+--------- | 11.962 | 16 | 0.7466 |
| AD Braak | chr21 | 26,269,344 | chr21:26269344:G:A | a | g | -0.1021 | 0.0245 | 3.03E-05 | ----+--------- | 11.038 | 16 | 0.8071 |
| AD Braak | chr21 | 26,274,124 | chr21:26274124:A:G | a | g | 0.1025 | 0.0245 | 2.84E-05 | +++++++++ | 10.659 | 16 | 0.8301 |
| AD Braak | chr21 | 26,276,173 | chr21:26276173:G:A | a | g | -0.1006 | 0.0245 | 3.99E-05 | ----+--------- | 11.235 | 16 | 0.7948 |
| AD Braak | chr21 | 26,277,931 | chr21:26277931:A:C | a | c | 0.1009 | 0.0245 | 3.85E-05 | ++++-++++- | 10.35 | 16 | 0.8477 |
| AD Braak | chr21 | 27,043,838 | chr21:27043838:C:T | t | c | 0.2004 | 0.0494 | 4.98E-05 | +++++++++ | 8.323 | 16 | 0.9387 |
| AD Braak | chr21 | 42,390,674 | chr21:42390674:G:A | a | g | -0.2239 | 0.0531 | 2.49E-05 | -------+------ | 14.544 | 16 | 0.5583 |
| AD Braak | chr21 | 42,391,002 | chr21:42391002:A:C | a | c | 0.2297 | 0.0534 | 1.69E-05 | +++++++-+- | 14.225 | 16 | 0.582 |
| AD Braak | chr21 | 42,392,459 | chr21:42392459:C:G | c | g | 0.256 | 0.0542 | 2.35E-06 | +++++++-+- | 13.699 | 16 | 0.6212 |
| AD Braak | chr21 | 42,392,672 | chr21:42392672:C:T | t | c | -0.2571 | 0.0542 | 2.14E-06 | -------+----- | 13.789 | 16 | 0.6144 |
| AD Braak | chr21 | 42,393,421 | chr21:42393421:G:A | a | g | -0.2461 | 0.0534 | 4.02E-06 | -------+----- | 13.36 | 16 | 0.6463 |
| AD Braak | chr21 | 42,395,056 | chr21:42395056:A:G | a | g | 0.1813 | 0.0427 | 2.16E-05 | +++++++-+- | 16.536 | 16 | 0.4162 |
| AD Braak | chr21 | 42,395,720 | chr21:42395720:C:T | t | c | -0.1923 | 0.0429 | 7.35E-06 | -------+----- | 16.057 | 16 | 0.449 |
| AD Braak | chr21 | 42,399,684 | chr21:42399684:T:C | t | c | 0.2469 | 0.0543 | 5.41E-06 | +++++++-+- | 14.694 | 16 | 0.5471 |

| | | | | | | | | | | | | |
|---|---|---|---|---|---|---|---|---|---|---|---|---|
| AD Braak | chr21 | 42,399,757 | chr21:42399757:A:T | a | t | 0.2495 | 0.054 | 3.87E-06 | +++++++-+- | 15.152 | 16 | 0.5136 |
| AD Braak | chr21 | 42,402,065 | chr21:42402065:C:T | t | c | -0.2472 | 0.054 | 4.64E-06 | -------+----- | 14.74 | 16 | 0.5437 |
| AD Braak | chr21 | 42,404,027 | chr21:42404027:C:T | t | c | -0.2496 | 0.0539 | 3.65E-06 | -------+----- | 14.504 | 16 | 0.5612 |
| AD Braak | chr21 | 42,405,896 | chr21:42405896:G:A | a | g | -0.2593 | 0.0568 | 4.92E-06 | ----+--+----- | 13.293 | 16 | 0.6512 |
| AD Braak | chr21 | 42,406,075 | chr21:42406075:G:A | a | g | -0.2488 | 0.054 | 4.07E-06 | -------+----- | 14.521 | 16 | 0.5599 |
| AD Braak | chr21 | 42,410,896 | chr21:42410896:C:A | a | c | -0.2429 | 0.0541 | 7.23E-06 | -------+----- | 14.585 | 16 | 0.5552 |
| AD Braak | chr21 | 46,186,597 | chr21:46186597:CA:C | ca | c | 0.1163 | 0.0245 | 2.16E-06 | ++++-++++- | 12.862 | 16 | 0.6828 |
| AD Braak | chr21 | 46,186,837 | chr21:46186837:TTTTG:T | t | ttttg | -0.1131 | 0.0245 | 3.75E-06 | ----+--------- | 12.588 | 16 | 0.7026 |
| AD Braak | chr21 | 46,188,666 | chr21:46188666:G:A | a | g | -0.0999 | 0.0245 | 4.70E-05 | ----+--------- | 9.262 | 16 | 0.9022 |
| AD Braak | chr21 | 46,191,396 | chr21:46191396:A:G | a | g | 0.0989 | 0.0245 | 5.55E-05 | ++++-++++- | 9.33 | 16 | 0.8992 |
| AD Braak | chr21 | 46,196,166 | chr21:46196166:G:A | a | g | -0.0997 | 0.0246 | 5.03E-05 | ----+--------- | 9.266 | 16 | 0.902 |
| AD Braak | chr21 | 46,196,999 | chr21:46196999:G:A | a | g | -0.0996 | 0.0246 | 5.11E-05 | ----+--------- | 9.267 | 16 | 0.902 |
| AD Braak | chr21 | 46,199,125 | chr21:46199125:G:A | a | g | -0.0995 | 0.0246 | 5.14E-05 | ----+--------- | 9.152 | 16 | 0.907 |
| AD Braak | chr21 | 46,199,386 | chr21:46199386:G:A | a | g | -0.0995 | 0.0246 | 5.14E-05 | ----+--------- | 9.155 | 16 | 0.9069 |
| AD Braak | chr21 | 46,202,813 | chr21:46202813:A:G | a | g | -0.1003 | 0.0246 | 4.58E-05 | -------------- | 9.092 | 16 | 0.9096 |
| AD Braak | chr21 | 46,203,659 | chr21:46203659:G:A | a | g | 0.101 | 0.0245 | 3.91E-05 | +++++++++ | 10.129 | 16 | 0.8598 |
| AD Braak | chr21 | 46,240,131 | chr21:46240131:A:T | a | t | -0.1047 | 0.0265 | 7.70E-05 | ----+--------- | 12.332 | 16 | 0.7208 |
| AD Braak | chr21 | 46,241,321 | chr21:46241321:C:T | t | c | 0.0973 | 0.0245 | 7.17E-05 | +++++++++ | 4.995 | 16 | 0.9958 |
| AD Braak | chr21 | 46,241,673 | chr21:46241673:A:G | a | g | -0.0967 | 0.0245 | 8.05E-05 | -------------- | 4.929 | 16 | 0.9961 |
| AD Braak | chr21 | 46,244,361 | chr21:46244361:G:C | c | g | 0.0965 | 0.0245 | 8.25E-05 | +++++++++ | 4.991 | 16 | 0.9958 |
| AD Braak | chr21 | 46,272,884 | chr21:46272884:C:T | t | c | 0.1 | 0.0256 | 9.38E-05 | ++++-++++- | 15.47 | 16 | 0.4905 |
| AD Braak | chr22 | 17,573,019 | chr22:17573019:G:A | a | g | -0.4197 | 0.1055 | 7.00E-05 | -+--+-----+-- | 19.953 | 16 | 0.2224 |
| AD Braak | chr22 | 18,990,561 | chr22:18990561:G:T | t | g | -0.5232 | 0.1273 | 3.95E-05 | ----+--?-?-- | 10.855 | 14 | 0.6974 |
| AD Braak | chr22 | 21,062,931 | chr22:21062931:T:C | t | c | -0.1324 | 0.0312 | 2.25E-05 | ----+-------- | 10.038 | 16 | 0.8646 |
| AD Braak | chr22 | 22,754,136 | chr22:22754136:C:A | a | c | 0.4781 | 0.1145 | 3.00E-05 | ++++?+-?+- | 19.496 | 13 | 0.1085 |
| AD Braak | chr22 | 22,776,030 | chr22:22776030:T:C | t | c | -0.4499 | 0.1134 | 7.32E-05 | ---+?-++-?+ | 19.624 | 13 | 0.105 |
| AD Braak | chr22 | 23,198,877 | chr22:23198877:T:C | t | c | 0.5567 | 0.1376 | 5.24E-05 | +?++?++++ | 4.305 | 9 | 0.8902 |
| AD Braak | chr22 | 23,203,194 | chr22:23203194:A:T | a | t | 0.5535 | 0.1369 | 5.26E-05 | +?++?++++ | 4.263 | 9 | 0.8933 |
| AD Braak | chr22 | 23,616,668 | chr22:23616668:C:T | t | c | 0.1123 | 0.0274 | 4.02E-05 | +++++++++ | 8.006 | 16 | 0.9487 |
| AD Braak | chr22 | 32,050,818 | chr22:32050818:G:A | a | g | 0.1517 | 0.0379 | 6.18E-05 | +++++-+++- | 20.591 | 16 | 0.1948 |
| AD Braak | chr22 | 35,886,831 | chr22:35886831:C:T | t | c | 0.5002 | 0.1157 | 1.55E-05 | +++-++-+++ | 14.074 | 13 | 0.3686 |
| AD Braak | chr3 | 36,835,413 | chr3:36835413:C:T | t | c | -0.4802 | 0.1155 | 3.24E-05 | -+--+-+--+-- | 27.937 | 15 | 0.02196 |
| AD Braak | chr3 | 36,942,051 | chr3:36942051:T:C | t | c | 0.4764 | 0.1144 | 3.10E-05 | +-++-++++- | 23.337 | 15 | 0.07725 |
| AD Braak | chr3 | 37,025,301 | chr3:37025301:T:C | t | c | 0.4574 | 0.1129 | 5.12E-05 | +-++-++++- | 23.628 | 15 | 0.07169 |
| AD Braak | chr3 | 37,077,046 | chr3:37077046:A:G | a | g | 0.4635 | 0.1129 | 4.06E-05 | +-++-++++- | 20.775 | 15 | 0.1442 |
| AD Braak | chr3 | 37,133,606 | chr3:37133606:A:G | a | g | 0.4556 | 0.113 | 5.50E-05 | +-++-+-++-- | 21.514 | 15 | 0.1212 |
| AD Braak | chr3 | 55,027,563 | chr3:55027563:C:T | t | c | 0.1158 | 0.0297 | 9.42E-05 | +++++++-+- | 12.002 | 16 | 0.7439 |
| AD Braak | chr3 | 60,501,934 | chr3:60501934:G:A | a | g | 0.2223 | 0.0538 | 3.57E-05 | +++++++++ | 16.626 | 16 | 0.4102 |
| AD Braak | chr3 | 62,760,820 | chr3:62760820:C:T | t | c | 0.4582 | 0.11 | 3.13E-05 | ++++?++++ | 6.125 | 12 | 0.9096 |
| AD Braak | chr3 | 66,656,766 | chr3:66656766:G:A | a | g | 0.2258 | 0.0544 | 3.37E-05 | +-+++-++++ | 8.885 | 16 | 0.9181 |
| AD Braak | chr3 | 66,671,650 | chr3:66671650:A:G | a | g | -0.2069 | 0.0532 | 9.91E-05 | -+--++------ | 16.543 | 16 | 0.4157 |
| AD Braak | chr3 | 66,676,719 | chr3:66676719:C:T | t | c | 0.2722 | 0.0691 | 8.25E-05 | +-+++-++++ | 17.924 | 16 | 0.3284 |
| AD Braak | chr3 | 66,692,025 | chr3:66692025:T:C | t | c | -0.2905 | 0.0699 | 3.22E-05 | -+---+------- | 19.864 | 16 | 0.2264 |
| AD Braak | chr3 | 66,710,366 | chr3:66710366:G:T | t | g | 0.2935 | 0.0731 | 5.88E-05 | ++++--+-++ | 15.128 | 16 | 0.5153 |
| AD Braak | chr3 | 73,120,284 | chr3:73120284:A:G | a | g | -0.0952 | 0.0243 | 9.03E-05 | ----+--+--+-- | 14.101 | 16 | 0.5912 |
| AD Braak | chr3 | 76,132,270 | chr3:76132270:C:T | t | c | -0.3287 | 0.0781 | 2.60E-05 | ----------+-- | 19.342 | 16 | 0.2513 |

| | | | | | | | | | | | | |
|---|---|---|---|---|---|---|---|---|---|---|---|---|
| AD Braak | chr3 | 77,125,014 | chr3:77125014:A:G | a | g | 0.2481 | 0.0604 | 3.93E-05 | +-++-+++++ | 19.78 | 16 | 0.2303 |
| AD Braak | chr3 | 77,127,036 | chr3:77127036:G:A | a | g | -0.2564 | 0.0604 | 2.23E-05 | ----+------+- | 18.311 | 16 | 0.306 |
| AD Braak | chr3 | 77,127,309 | chr3:77127309:C:T | t | c | -0.2492 | 0.0592 | 2.57E-05 | ----+--------- | 15.337 | 16 | 0.5001 |
| AD Braak | chr3 | 77,128,524 | chr3:77128524:A:G | a | g | 0.2393 | 0.0586 | 4.44E-05 | ++++-++++- | 15.718 | 16 | 0.4728 |
| AD Braak | chr3 | 77,129,631 | chr3:77129631:G:T | t | g | -0.2527 | 0.0603 | 2.82E-05 | ----+------+- | 18.54 | 16 | 0.2932 |
| AD Braak | chr3 | 77,132,031 | chr3:77132031:G:A | a | g | -0.2468 | 0.0601 | 4.03E-05 | ----+------+- | 18.9 | 16 | 0.2739 |
| AD Braak | chr3 | 77,135,872 | chr3:77135872:C:T | t | c | -0.2411 | 0.0599 | 5.71E-05 | ----+------+- | 18.321 | 16 | 0.3055 |
| AD Braak | chr3 | 77,136,042 | chr3:77136042:G:A | a | g | -0.2452 | 0.06 | 4.31E-05 | ----+------+- | 18.694 | 16 | 0.2848 |
| AD Braak | chr3 | 77,139,205 | chr3:77139205:C:T | t | c | -0.2369 | 0.0587 | 5.51E-05 | ----+--------- | 15.737 | 16 | 0.4715 |
| AD Braak | chr3 | 77,139,424 | chr3:77139424:T:A | a | t | -0.2276 | 0.0537 | 2.23E-05 | -+--+-------- | 15.149 | 16 | 0.5138 |
| AD Braak | chr3 | 77,148,488 | chr3:77148488:G:A | a | g | -0.2472 | 0.0598 | 3.61E-05 | ----+------+- | 19.324 | 16 | 0.2522 |
| AD Braak | chr3 | 77,148,705 | chr3:77148705:A:G | a | g | 0.2386 | 0.0597 | 6.52E-05 | ++++-++++- | 18.658 | 16 | 0.2868 |
| AD Braak | chr3 | 77,154,006 | chr3:77154006:C:T | t | c | -0.2416 | 0.0536 | 6.49E-06 | -+--+-------- | 14.302 | 16 | 0.5762 |
| AD Braak | chr3 | 77,154,266 | chr3:77154266:G:A | a | g | -0.247 | 0.0586 | 2.47E-05 | -+--+-------- | 17.338 | 16 | 0.3641 |
| AD Braak | chr3 | 77,154,368 | chr3:77154368:C:T | t | c | -0.2495 | 0.0587 | 2.15E-05 | -+--+-------- | 17.405 | 16 | 0.3599 |
| AD Braak | chr3 | 77,155,643 | chr3:77155643:C:T | t | c | -0.2507 | 0.0601 | 3.04E-05 | ----+------+- | 17.881 | 16 | 0.3309 |
| AD Braak | chr3 | 77,157,743 | chr3:77157743:C:G | c | g | 0.2397 | 0.0587 | 4.47E-05 | +-++-+++++ | 15.866 | 16 | 0.4623 |
| AD Braak | chr3 | 77,158,033 | chr3:77158033:T:C | t | c | 0.2394 | 0.0587 | 4.58E-05 | +-++-+++++ | 15.914 | 16 | 0.459 |
| AD Braak | chr3 | 77,161,094 | chr3:77161094:C:T | t | c | -0.2467 | 0.0601 | 4.10E-05 | ----+------+- | 17.719 | 16 | 0.3406 |
| AD Braak | chr3 | 77,162,712 | chr3:77162712:T:A | a | t | -0.2484 | 0.0601 | 3.62E-05 | ----+------+- | 18.022 | 16 | 0.3226 |
| AD Braak | chr3 | 77,163,599 | chr3:77163599:G:A | a | g | -0.2538 | 0.0602 | 2.52E-05 | ----+------+- | 18.423 | 16 | 0.2997 |
| AD Braak | chr3 | 77,164,139 | chr3:77164139:G:A | a | g | -0.2527 | 0.0603 | 2.73E-05 | ----+------+- | 18.383 | 16 | 0.3019 |
| AD Braak | chr3 | 77,166,607 | chr3:77166607:G:A | a | g | -0.2585 | 0.0603 | 1.79E-05 | -+--+------+- | 17.356 | 16 | 0.3629 |
| AD Braak | chr3 | 77,167,895 | chr3:77167895:C:T | t | c | -0.2556 | 0.0603 | 2.23E-05 | -+--+------+- | 18.537 | 16 | 0.2934 |
| AD Braak | chr3 | 77,170,674 | chr3:77170674:G:A | a | g | -0.2577 | 0.0602 | 1.87E-05 | -+--+------+- | 17.971 | 16 | 0.3256 |
| AD Braak | chr3 | 77,172,414 | chr3:77172414:T:A | a | t | -0.2576 | 0.06 | 1.75E-05 | -+--+-------- | 18.006 | 16 | 0.3235 |
| AD Braak | chr3 | 77,178,917 | chr3:77178917:T:C | t | c | 0.2545 | 0.0603 | 2.44E-05 | +-++-+++++ | 18.348 | 16 | 0.3039 |
| AD Braak | chr3 | 77,181,914 | chr3:77181914:C:G | c | g | 0.2526 | 0.0602 | 2.75E-05 | +-++-+++++ | 18.184 | 16 | 0.3132 |
| AD Braak | chr3 | 77,183,732 | chr3:77183732:C:T | t | c | -0.2552 | 0.0602 | 2.25E-05 | -+--+-------- | 18.428 | 16 | 0.2995 |
| AD Braak | chr3 | 77,222,211 | chr3:77222211:A:G | a | g | 0.2561 | 0.0613 | 2.91E-05 | +-++-+++++ | 18.479 | 16 | 0.2966 |
| AD Braak | chr3 | 113,105,042 | chr3:113105042:A:G | a | g | -0.4785 | 0.1152 | 3.25E-05 | ----+--+???- | 11.996 | 12 | 0.446 |
| AD Braak | chr3 | 113,168,082 | chr3:113168082:C:T | t | c | 0.5869 | 0.1313 | 7.79E-06 | ++++??+-?? | 10.991 | 9 | 0.2763 |
| AD Braak | chr3 | 146,668,928 | chr3:146668928:T:C | t | c | -0.369 | 0.0913 | 5.33E-05 | ----------?--- | 13.567 | 15 | 0.5586 |
| AD Braak | chr3 | 148,120,176 | chr3:148120176:CAT:C | cat | c | 0.442 | 0.1068 | 3.51E-05 | +?++-++++? | 14.461 | 12 | 0.2722 |
| AD Braak | chr3 | 150,465,174 | chr3:150465174:T:A | a | t | 0.256 | 0.0655 | 9.24E-05 | -+++-++-++ | 13.559 | 16 | 0.6315 |
| AD Braak | chr3 | 150,465,175 | chr3:150465175:C:A | a | c | 0.256 | 0.0655 | 9.24E-05 | -+++-++-++ | 13.559 | 16 | 0.6315 |
| AD Braak | chr3 | 151,030,098 | chr3:151030098:A:C | a | c | -0.1804 | 0.0454 | 7.01E-05 | ---++-------+ | 13.749 | 16 | 0.6174 |
| AD Braak | chr3 | 151,366,669 | chr3:151366669:C:T | t | c | -0.135 | 0.0338 | 6.46E-05 | -----+-------- | 21.693 | 16 | 0.1534 |
| AD Braak | chr3 | 173,023,137 | chr3:173023137:A:ATG | a | atg | -0.1857 | 0.043 | 1.58E-05 | ------+---+-- | 10.665 | 16 | 0.8297 |
| AD Braak | chr3 | 177,454,275 | chr3:177454275:C:T | t | c | 0.3911 | 0.0934 | 2.84E-05 | +++++++-+- | 11.638 | 16 | 0.7685 |
| AD Braak | chr3 | 196,140,979 | chr3:196140979:C:T | t | c | -0.0975 | 0.0247 | 7.78E-05 | -----+-++---- | 22.927 | 16 | 0.1157 |
| AD Braak | chr3 | 196,143,290 | chr3:196143290:C:T | t | c | -0.1023 | 0.0245 | 2.91E-05 | ----++-+++-- | 21.143 | 16 | 0.1731 |
| AD Braak | chr4 | 13,756,738 | chr4:13756738:G:A | a | g | 0.1501 | 0.0385 | 9.73E-05 | +-+++-+++- | 22.912 | 16 | 0.1161 |
| AD Braak | chr4 | 13,758,405 | chr4:13758405:G:A | a | g | 0.1516 | 0.0386 | 8.47E-05 | +-+++-+++- | 22.644 | 16 | 0.1236 |
| AD Braak | chr4 | 13,759,232 | chr4:13759232:T:G | t | g | -0.1509 | 0.0386 | 9.10E-05 | -+---+---++- | 22.619 | 16 | 0.1243 |
| AD Braak | chr4 | 13,759,497 | chr4:13759497:T:A | a | t | 0.1505 | 0.0386 | 9.44E-05 | +-+++-+++- | 22.65 | 16 | 0.1234 |

| | | | | | | | | | | | | |
|---|---|---|---|---|---|---|---|---|---|---|---|---|
| AD Braak | chr4 | 13,761,983 | chr4:13761983:G:T | t | g | 0.1502 | 0.0385 | 9.66E-05 | +++++-+++- | 22.571 | 16 | 0.1257 |
| AD Braak | chr4 | 18,840,933 | chr4:18840933:A:G | a | g | 0.3496 | 0.0805 | 1.41E-05 | ++++++++-- | 11.143 | 15 | 0.7424 |
| AD Braak | chr4 | 18,863,832 | chr4:18863832:T:G | t | g | 0.3946 | 0.0874 | 6.37E-06 | ++++++++-- | 13.754 | 15 | 0.5443 |
| AD Braak | chr4 | 18,874,947 | chr4:18874947:C:T | t | c | -0.3966 | 0.0877 | 6.14E-06 | --------+-?-- | 13.806 | 15 | 0.5403 |
| AD Braak | chr4 | 26,401,783 | chr4:26401783:A:G | a | g | 0.3092 | 0.0731 | 2.32E-05 | +++++++++ | 10.822 | 15 | 0.7651 |
| AD Braak | chr4 | 45,342,130 | chr4:45342130:A:G | a | g | 0.1045 | 0.0268 | 9.41E-05 | ++++-++-++ | 11.658 | 16 | 0.7672 |
| AD Braak | chr4 | 79,686,295 | chr4:79686295:A:G | a | g | 0.1421 | 0.0362 | 8.86E-05 | ++++-++-++ | 15.652 | 16 | 0.4775 |
| AD Braak | chr4 | 79,690,813 | chr4:79690813:A:G | a | g | 0.1414 | 0.0362 | 9.24E-05 | ++++-++-++ | 14.999 | 16 | 0.5247 |
| AD Braak | chr4 | 79,692,994 | chr4:79692994:T:C | t | c | 0.1432 | 0.0359 | 6.66E-05 | ++++-++-++ | 14.54 | 16 | 0.5586 |
| AD Braak | chr4 | 79,693,711 | chr4:79693711:T:G | t | g | 0.1454 | 0.0359 | 5.14E-05 | ++++-++-++ | 14.156 | 16 | 0.5871 |
| AD Braak | chr4 | 79,694,325 | chr4:79694325:A:G | a | g | 0.1447 | 0.0359 | 5.57E-05 | ++++-++-++ | 14.325 | 16 | 0.5745 |
| AD Braak | chr4 | 79,695,186 | chr4:79695186:C:T | t | c | -0.1443 | 0.0359 | 5.81E-05 | ----+--+----- | 14.294 | 16 | 0.5769 |
| AD Braak | chr4 | 79,696,069 | chr4:79696069:C:G | c | g | 0.1454 | 0.0359 | 5.10E-05 | ++++-++-++ | 14.183 | 16 | 0.5851 |
| AD Braak | chr4 | 79,696,964 | chr4:79696964:G:T | t | g | -0.1453 | 0.0359 | 5.13E-05 | ----+--+----- | 14.184 | 16 | 0.585 |
| AD Braak | chr4 | 79,697,166 | chr4:79697166:T:A | a | t | -0.1453 | 0.0359 | 5.12E-05 | ----+--+----- | 14.181 | 16 | 0.5853 |
| AD Braak | chr4 | 79,698,158 | chr4:79698158:TTTTTC:T | t | tttttc | -0.1453 | 0.0359 | 5.17E-05 | ----+--+----- | 14.16 | 16 | 0.5868 |
| AD Braak | chr4 | 79,701,988 | chr4:79701988:G:A | a | g | -0.1448 | 0.0359 | 5.42E-05 | ----+--+----- | 14.287 | 16 | 0.5773 |
| AD Braak | chr4 | 79,702,087 | chr4:79702087:TA:T | t | ta | -0.1445 | 0.0361 | 6.17E-05 | ----+--+----- | 14.151 | 16 | 0.5874 |
| AD Braak | chr4 | 79,710,982 | chr4:79710982:C:T | t | c | -0.1453 | 0.0359 | 5.05E-05 | ----+--+----- | 14.391 | 16 | 0.5696 |
| AD Braak | chr4 | 79,711,577 | chr4:79711577:C:T | t | c | -0.1433 | 0.0359 | 6.39E-05 | ----+--+----- | 13.581 | 16 | 0.6299 |
| AD Braak | chr4 | 79,712,235 | chr4:79712235:C:A | a | c | -0.1453 | 0.0359 | 5.06E-05 | ----+--+----- | 14.39 | 16 | 0.5697 |
| AD Braak | chr4 | 79,712,741 | chr4:79712741:G:C | c | g | -0.1452 | 0.0359 | 5.21E-05 | ----+--+----- | 14.315 | 16 | 0.5753 |
| AD Braak | chr4 | 79,712,905 | chr4:79712905:G:A | a | g | -0.1453 | 0.0359 | 5.07E-05 | ----+--+----- | 14.4 | 16 | 0.5689 |
| AD Braak | chr4 | 79,714,869 | chr4:79714869:C:T | t | c | -0.1454 | 0.0359 | 5.03E-05 | ----+--+----- | 14.382 | 16 | 0.5703 |
| AD Braak | chr4 | 79,715,710 | chr4:79715710:G:A | a | g | -0.1449 | 0.0358 | 5.24E-05 | ----+--+----- | 14.476 | 16 | 0.5633 |
| AD Braak | chr4 | 79,717,485 | chr4:79717485:T:C | t | c | 0.146 | 0.0358 | 4.66E-05 | ++++-++-++ | 14.327 | 16 | 0.5743 |
| AD Braak | chr4 | 79,717,707 | chr4:79717707:C:G | c | g | 0.1452 | 0.0358 | 5.13E-05 | ++++-++-++ | 14.396 | 16 | 0.5693 |
| AD Braak | chr4 | 79,717,753 | chr4:79717753:G:T | t | g | -0.1452 | 0.0358 | 5.12E-05 | ----+--+----- | 14.406 | 16 | 0.5685 |
| AD Braak | chr4 | 79,718,120 | chr4:79718120:G:C | c | g | -0.144 | 0.0358 | 5.89E-05 | ----+--+----- | 14.335 | 16 | 0.5738 |
| AD Braak | chr4 | 79,718,846 | chr4:79718846:A:C | a | c | 0.1451 | 0.0358 | 5.14E-05 | ++++-++-++ | 14.403 | 16 | 0.5687 |
| AD Braak | chr4 | 79,718,868 | chr4:79718868:T:C | t | c | 0.1456 | 0.0358 | 4.89E-05 | ++++-++-++ | 14.404 | 16 | 0.5686 |
| AD Braak | chr4 | 79,720,193 | chr4:79720193:T:A | a | t | -0.1452 | 0.0358 | 5.13E-05 | ----+--+----- | 14.407 | 16 | 0.5684 |
| AD Braak | chr4 | 79,722,055 | chr4:79722055:A:G | a | g | 0.1436 | 0.0359 | 6.20E-05 | ++++-++-++ | 13.756 | 16 | 0.6169 |
| AD Braak | chr4 | 79,723,228 | chr4:79723228:C:T | t | c | -0.1435 | 0.0359 | 6.23E-05 | ----+--+----+ | 13.757 | 16 | 0.6168 |
| AD Braak | chr4 | 79,724,345 | chr4:79724345:T:C | t | c | 0.1438 | 0.0359 | 6.31E-05 | ++++-++-++ | 12.965 | 16 | 0.6753 |
| AD Braak | chr4 | 79,728,859 | chr4:79728859:C:T | t | c | -0.1457 | 0.0359 | 4.96E-05 | ----+--+----- | 13.316 | 16 | 0.6495 |
| AD Braak | chr4 | 79,731,272 | chr4:79731272:G:A | a | g | -0.1445 | 0.0359 | 5.69E-05 | ----+--+----+ | 13.471 | 16 | 0.638 |
| AD Braak | chr4 | 79,733,028 | chr4:79733028:A:C | a | c | 0.1436 | 0.0359 | 6.35E-05 | ++++-++-++ | 13.475 | 16 | 0.6378 |
| AD Braak | chr4 | 79,735,151 | chr4:79735151:A:G | a | g | 0.1449 | 0.0358 | 5.28E-05 | ++++-++-++ | 13.416 | 16 | 0.6421 |
| AD Braak | chr4 | 79,739,664 | chr4:79739664:ATATT:A | a | atatt | -0.14 | 0.0357 | 8.76E-05 | ----+--+---+- | 15.65 | 16 | 0.4777 |
| AD Braak | chr4 | 79,749,134 | chr4:79749134:C:G | c | g | 0.1396 | 0.0357 | 9.06E-05 | ++++-++-++ | 16.148 | 16 | 0.4427 |
| AD Braak | chr4 | 85,992,912 | chr4:85992912:C:T | t | c | 0.4071 | 0.0968 | 2.62E-05 | +++++++-+- | 23.503 | 15 | 0.07403 |
| AD Braak | chr4 | 86,061,793 | chr4:86061793:A:T | a | t | -0.5129 | 0.1053 | 1.12E-06 | ----?--?---+- | 26.786 | 13 | 0.01331 |
| AD Braak | chr4 | 103,078,063 | chr4:103078063:A:G | a | g | -0.3158 | 0.0806 | 8.99E-05 | ---++------+- | 9.765 | 16 | 0.8786 |
| AD Braak | chr4 | 104,792,508 | chr4:104792508:T:C | t | c | -0.6414 | 0.1484 | 1.54E-05 | -+----?+--?- | 11.269 | 13 | 0.5883 |
| AD Braak | chr4 | 114,822,051 | chr4:114822051:A:G | a | g | -0.3944 | 0.0977 | 5.42E-05 | -----+------- | 13.406 | 15 | 0.5709 |

| | | | | | | | | | | | | |
|---|---|---|---|---|---|---|---|---|---|---|---|---|
| AD Braak | chr4 | 114,823,691 | chr4:114823691:A:G | a | g | -0.4289 | 0.1004 | 1.95E-05 | -----+-------- | 13.104 | 15 | 0.5943 |
| AD Braak | chr4 | 114,897,581 | chr4:114897581:C:G | c | g | -0.4842 | 0.1052 | 4.13E-06 | -+---+------- | 20.996 | 15 | 0.137 |
| AD Braak | chr4 | 115,084,809 | chr4:115084809:G:T | t | g | 0.4891 | 0.0998 | 9.53E-07 | +++-+++++- | 14.773 | 14 | 0.3939 |
| AD Braak | chr4 | 115,170,537 | chr4:115170537:C:A | a | c | 0.3866 | 0.0892 | 1.46E-05 | +-+-+++-++ | 18.277 | 16 | 0.308 |
| AD Braak | chr4 | 115,333,555 | chr4:115333555:A:G | a | g | -0.4688 | 0.1017 | 4.04E-06 | ---+---+-+--- | 12.782 | 15 | 0.6192 |
| AD Braak | chr4 | 115,374,803 | chr4:115374803:C:G | c | g | -0.4757 | 0.1022 | 3.23E-06 | -------+-+--- | 12.386 | 15 | 0.6496 |
| AD Braak | chr4 | 115,420,709 | chr4:115420709:T:C | t | c | -0.4806 | 0.1026 | 2.81E-06 | -------+-+--- | 12.041 | 15 | 0.6759 |
| AD Braak | chr4 | 132,580,462 | chr4:132580462:A:C | a | c | 0.3931 | 0.0991 | 7.24E-05 | +-+--+++-+- | 17.697 | 16 | 0.3419 |
| AD Braak | chr4 | 159,259,641 | chr4:159259641:C:G | c | g | 0.115 | 0.0292 | 8.28E-05 | +++++-+++- | 17.563 | 16 | 0.3501 |
| AD Braak | chr4 | 159,304,037 | chr4:159304037:T:C | t | c | 0.1806 | 0.046 | 8.57E-05 | +++++++++ | 8.001 | 16 | 0.9488 |
| AD Braak | chr4 | 188,139,882 | chr4:188139882:C:T | t | c | 0.1239 | 0.0294 | 2.41E-05 | ++++++-++- | 17.382 | 16 | 0.3613 |
| AD Braak | chr4 | 188,140,015 | chr4:188140015:C:T | t | c | 0.1248 | 0.0294 | 2.13E-05 | ++++++-++- | 17.339 | 16 | 0.364 |
| AD Braak | chr4 | 188,140,450 | chr4:188140450:C:T | t | c | 0.1278 | 0.0293 | 1.34E-05 | ++++++-++- | 17.924 | 16 | 0.3283 |
| AD Braak | chr4 | 188,140,521 | chr4:188140521:A:AT | a | at | -0.1268 | 0.0294 | 1.57E-05 | ------+----+- | 17.916 | 16 | 0.3288 |
| AD Braak | chr4 | 188,140,701 | chr4:188140701:G:GA | g | ga | -0.1303 | 0.0297 | 1.18E-05 | ------+------- | 18.024 | 16 | 0.3225 |
| AD Braak | chr4 | 188,141,230 | chr4:188141230:C:T | t | c | 0.1196 | 0.03 | 6.58E-05 | +++++--+++ | 19.092 | 16 | 0.2639 |
| AD Braak | chr4 | 188,141,448 | chr4:188141448:G:A | a | g | 0.1192 | 0.03 | 6.92E-05 | +++++--+++ | 19.102 | 16 | 0.2634 |
| AD Braak | chr4 | 188,141,805 | chr4:188141805:A:T | a | t | -0.1192 | 0.03 | 6.92E-05 | -----++----+- | 19.033 | 16 | 0.267 |
| AD Braak | chr4 | 188,926,289 | chr4:188926289:T:C | t | c | -0.2729 | 0.0682 | 6.34E-05 | ----+--+---+- | 16.085 | 16 | 0.4471 |
| AD Braak | chr4 | 188,930,917 | chr4:188930917:C:A | a | c | 0.2748 | 0.0682 | 5.61E-05 | ++++-++-++ | 15.921 | 16 | 0.4585 |
| AD Braak | chr5 | 2,159,800 | chr5:2159800:C:T | t | c | 0.3774 | 0.082 | 4.14E-06 | +-++-++-++ | 15.702 | 16 | 0.4739 |
| AD Braak | chr5 | 6,674,037 | chr5:6674037:A:G | a | g | -0.5821 | 0.1431 | 4.76E-05 | ----???--?? | 3.925 | 5 | 0.5602 |
| AD Braak | chr5 | 68,300,260 | chr5:68300260:G:A | a | g | 0.0994 | 0.0244 | 4.76E-05 | +++++++-+- | 17.075 | 16 | 0.3808 |
| AD Braak | chr5 | 86,969,332 | chr5:86969332:G:T | t | g | 0.1133 | 0.029 | 9.54E-05 | +++++++++ | 4.226 | 16 | 0.9985 |
| AD Braak | chr5 | 86,986,612 | chr5:86986612:A:G | a | g | -0.1127 | 0.0289 | 9.75E-05 | -------------- | 4.336 | 16 | 0.9982 |
| AD Braak | chr5 | 86,998,852 | chr5:86998852:C:T | t | c | 0.1178 | 0.0287 | 3.93E-05 | +++++++++ | 5.032 | 16 | 0.9956 |
| AD Braak | chr5 | 87,002,832 | chr5:87002832:A:G | a | g | -0.1139 | 0.029 | 8.71E-05 | -------------- | 4.313 | 16 | 0.9983 |
| AD Braak | chr5 | 87,003,353 | chr5:87003353:C:T | t | c | 0.1148 | 0.029 | 7.68E-05 | +++++++++ | 4.119 | 16 | 0.9987 |
| AD Braak | chr5 | 87,006,556 | chr5:87006556:T:C | t | c | -0.1134 | 0.029 | 9.31E-05 | -------------- | 4.639 | 16 | 0.9973 |
| AD Braak | chr5 | 87,006,620 | chr5:87006620:C:T | t | c | 0.1132 | 0.029 | 9.48E-05 | +++++++++ | 4.7 | 16 | 0.9971 |
| AD Braak | chr5 | 87,013,081 | chr5:87013081:T:G | t | g | 0.1144 | 0.0292 | 8.81E-05 | +++++++++ | 5.544 | 16 | 0.9923 |
| AD Braak | chr5 | 87,013,307 | chr5:87013307:A:C | a | c | 0.1177 | 0.0288 | 4.31E-05 | +++++++++ | 5.583 | 16 | 0.992 |
| AD Braak | chr5 | 87,015,638 | chr5:87015638:A:G | a | g | 0.1084 | 0.0278 | 9.86E-05 | +++++++++ | 6.623 | 16 | 0.9799 |
| AD Braak | chr5 | 87,020,609 | chr5:87020609:C:T | t | c | -0.1086 | 0.0279 | 9.72E-05 | -------------- | 5.896 | 16 | 0.9892 |
| AD Braak | chr5 | 87,021,376 | chr5:87021376:T:C | t | c | 0.122 | 0.0288 | 2.23E-05 | +++++++++ | 5.623 | 16 | 0.9917 |
| AD Braak | chr5 | 87,023,211 | chr5:87023211:A:G | a | g | 0.1152 | 0.0291 | 7.70E-05 | +++++++++ | 5.247 | 16 | 0.9944 |
| AD Braak | chr5 | 87,023,522 | chr5:87023522:A:C | a | c | 0.1157 | 0.0291 | 7.10E-05 | +++++++++ | 5.408 | 16 | 0.9933 |
| AD Braak | chr5 | 87,024,119 | chr5:87024119:C:T | t | c | -0.1199 | 0.0287 | 2.96E-05 | -------------- | 5.791 | 16 | 0.9902 |
| AD Braak | chr5 | 87,057,174 | chr5:87057174:T:C | t | c | 0.1086 | 0.0279 | 9.93E-05 | +++++++++ | 6.254 | 16 | 0.9851 |
| AD Braak | chr5 | 87,072,777 | chr5:87072777:G:A | a | g | -0.1142 | 0.0285 | 5.99E-05 | -------------- | 4.502 | 16 | 0.9977 |
| AD Braak | chr5 | 111,902,082 | chr5:111902082:G:C | c | g | -0.2227 | 0.0529 | 2.59E-05 | -------+-+--- | 13.017 | 16 | 0.6715 |
| AD Braak | chr5 | 119,592,051 | chr5:119592051:C:T | t | c | -0.2277 | 0.0537 | 2.21E-05 | ------------+- | 11.275 | 16 | 0.7922 |
| AD Braak | chr5 | 119,602,685 | chr5:119602685:G:A | a | g | -0.2298 | 0.0537 | 1.91E-05 | ------------+- | 12.167 | 16 | 0.7324 |
| AD Braak | chr5 | 119,603,270 | chr5:119603270:ACT:A | a | act | -0.2308 | 0.0538 | 1.78E-05 | ------------+- | 12.273 | 16 | 0.725 |
| AD Braak | chr5 | 119,604,035 | chr5:119604035:A:G | a | g | 0.2293 | 0.0537 | 1.92E-05 | +++++++++ | 12.065 | 16 | 0.7395 |
| AD Braak | chr5 | 119,674,221 | chr5:119674221:T:C | t | c | 0.3667 | 0.0906 | 5.16E-05 | +++++++-+- | 11.385 | 16 | 0.7851 |

| | | | | | | | | | | | | |
|---|---|---|---|---|---|---|---|---|---|---|---|---|
| AD Braak | chr5 | 141,644,156 | chr5:141644156:T:C | t | c | -0.1138 | 0.0284 | 6.10E-05 | --------++--- | 15.014 | 16 | 0.5236 |
| AD Braak | chr5 | 141,645,323 | chr5:141645323:T:TAGC | t | tagc | -0.1051 | 0.026 | 5.23E-05 | -------++---- | 15.078 | 16 | 0.5189 |
| AD Braak | chr5 | 141,647,317 | chr5:141647317:T:C | t | c | -0.1143 | 0.0282 | 5.17E-05 | --------++--- | 17.21 | 16 | 0.3722 |
| AD Braak | chr5 | 141,647,833 | chr5:141647833:G:C | c | g | 0.1042 | 0.026 | 6.23E-05 | +++++++--+ | 15.244 | 16 | 0.5068 |
| AD Braak | chr5 | 176,374,082 | chr5:176374082:C:G | c | g | -0.1291 | 0.0307 | 2.58E-05 | ---+-----+--- | 18.857 | 16 | 0.2761 |
| AD Braak | chr5 | 178,302,673 | chr5:178302673:C:T | t | c | 0.6223 | 0.1589 | 8.94E-05 | ++++??+-?? | 3.912 | 6 | 0.6886 |
| AD Braak | chr6 | 1,478,638 | chr6:1478638:G:A | a | g | -0.1079 | 0.0266 | 5.04E-05 | ----+---+-+-- | 21.262 | 16 | 0.1687 |
| AD Braak | chr6 | 13,318,624 | chr6:13318624:A:C | a | c | 0.1699 | 0.0429 | 7.50E-05 | +++++++++ | 12.398 | 16 | 0.7161 |
| AD Braak | chr6 | 13,319,014 | chr6:13319014:AC:A | a | ac | -0.172 | 0.0427 | 5.53E-05 | ---------+---+ | 13.578 | 16 | 0.6301 |
| AD Braak | chr6 | 13,324,197 | chr6:13324197:G:A | a | g | -0.1659 | 0.0423 | 8.87E-05 | ---------+---+ | 11.566 | 16 | 0.7733 |
| AD Braak | chr6 | 13,324,658 | chr6:13324658:T:C | t | c | 0.1674 | 0.0422 | 7.16E-05 | +++++++++ | 12.523 | 16 | 0.7073 |
| AD Braak | chr6 | 13,325,851 | chr6:13325851:G:A | a | g | -0.1668 | 0.0422 | 7.61E-05 | ---------+---+ | 12.574 | 16 | 0.7036 |
| AD Braak | chr6 | 13,328,419 | chr6:13328419:G:C | c | g | -0.1706 | 0.0425 | 5.85E-05 | ---------+---+ | 10.304 | 16 | 0.8503 |
| AD Braak | chr6 | 13,328,485 | chr6:13328485:C:G | c | g | 0.1675 | 0.0423 | 7.62E-05 | +++++++++ | 9.782 | 16 | 0.8778 |
| AD Braak | chr6 | 32,621,638 | chr6:32621638:G:A | a | g | 0.1331 | 0.0333 | 6.49E-05 | +-+++++++- | 34.405 | 16 | 0.00479 |
| AD Braak | chr6 | 32,622,860 | chr6:32622860:T:A | a | t | 0.1326 | 0.0333 | 6.96E-05 | +-+++++++- | 34.733 | 16 | 0.004322 |
| AD Braak | chr6 | 32,622,864 | chr6:32622864:T:G | t | g | -0.1326 | 0.0333 | 6.98E-05 | -+---------+- | 34.713 | 16 | 0.00435 |
| AD Braak | chr6 | 32,634,360 | chr6:32634360:C:G | c | g | -0.1328 | 0.0334 | 6.98E-05 | -+---------+- | 34.982 | 16 | 0.003997 |
| AD Braak | chr6 | 32,636,021 | chr6:32636021:A:T | a | t | -0.1324 | 0.0334 | 7.28E-05 | -+---------+- | 34.338 | 16 | 0.004892 |
| AD Braak | chr6 | 32,636,679 | chr6:32636679:G:C | c | g | 0.1331 | 0.0334 | 6.75E-05 | +-+++++++- | 34.922 | 16 | 0.004073 |
| AD Braak | chr6 | 33,057,036 | chr6:33057036:TTC:T | t | ttc | 0.2476 | 0.0632 | 8.95E-05 | +++++++++ | 17.609 | 16 | 0.3473 |
| AD Braak | chr6 | 33,077,018 | chr6:33077018:C:T | t | c | 0.251 | 0.0628 | 6.37E-05 | +-+++++++- | 17.191 | 16 | 0.3733 |
| AD Braak | chr6 | 33,080,342 | chr6:33080342:C:T | t | c | 0.2479 | 0.0627 | 7.77E-05 | +-+++++++- | 16.76 | 16 | 0.4013 |
| AD Braak | chr6 | 33,108,332 | chr6:33108332:G:A | a | g | 0.2442 | 0.0626 | 9.67E-05 | +-+++++-++ | 17.628 | 16 | 0.3461 |
| AD Braak | chr6 | 39,164,822 | chr6:39164822:C:A | a | c | -0.4589 | 0.1062 | 1.56E-05 | -+--?----+?- | 8.661 | 13 | 0.798 |
| AD Braak | chr6 | 39,165,615 | chr6:39165615:G:A | a | g | -0.4585 | 0.1063 | 1.60E-05 | -+--?----+?- | 8.686 | 13 | 0.7962 |
| AD Braak | chr6 | 41,661,306 | chr6:41661306:C:T | t | c | 0.2459 | 0.0624 | 8.09E-05 | +++++++++ | 21.651 | 15 | 0.1173 |
| AD Braak | chr6 | 41,745,795 | chr6:41745795:C:T | t | c | 0.4503 | 0.1121 | 5.88E-05 | ++++-++++? | 16.344 | 15 | 0.3596 |
| AD Braak | chr6 | 43,990,869 | chr6:43990869:G:A | a | g | -0.2915 | 0.0736 | 7.48E-05 | ----+-+----+- | 15.522 | 16 | 0.4868 |
| AD Braak | chr6 | 47,462,031 | chr6:47462031:C:A | a | c | -0.11 | 0.027 | 4.74E-05 | -+---------+- | 13.881 | 16 | 0.6076 |
| AD Braak | chr6 | 47,463,548 | chr6:47463548:A:C | a | c | 0.1054 | 0.0269 | 8.94E-05 | +-+++++++- | 13.891 | 16 | 0.6069 |
| AD Braak | chr6 | 47,466,015 | chr6:47466015:A:C | a | c | 0.1101 | 0.027 | 4.56E-05 | +-+++++++- | 13.421 | 16 | 0.6418 |
| AD Braak | chr6 | 47,472,829 | chr6:47472829:C:A | a | c | -0.1114 | 0.027 | 3.75E-05 | -+---------+- | 13.213 | 16 | 0.6571 |
| AD Braak | chr6 | 47,474,135 | chr6:47474135:T:C | t | c | 0.1113 | 0.027 | 3.79E-05 | +-+++++++- | 13.49 | 16 | 0.6366 |
| AD Braak | chr6 | 47,474,418 | chr6:47474418:TA:T | t | ta | -0.1113 | 0.027 | 3.79E-05 | -+-------+-+- | 13.488 | 16 | 0.6368 |
| AD Braak | chr6 | 47,474,641 | chr6:47474641:C:T | t | c | -0.1113 | 0.027 | 3.82E-05 | -+-------+-+- | 13.479 | 16 | 0.6375 |
| AD Braak | chr6 | 47,476,070 | chr6:47476070:A:G | a | g | 0.11 | 0.027 | 4.65E-05 | +-+++++++- | 13.359 | 16 | 0.6464 |
| AD Braak | chr6 | 47,477,281 | chr6:47477281:C:T | t | c | -0.1107 | 0.027 | 4.11E-05 | -+-------+-+- | 13.425 | 16 | 0.6414 |
| AD Braak | chr6 | 47,479,305 | chr6:47479305:T:A | a | t | -0.1101 | 0.027 | 4.64E-05 | -+-------+-+- | 13.685 | 16 | 0.6221 |
| AD Braak | chr6 | 47,480,600 | chr6:47480600:A:G | a | g | 0.1102 | 0.027 | 4.51E-05 | +-+++++++- | 13.628 | 16 | 0.6264 |
| AD Braak | chr6 | 47,482,882 | chr6:47482882:G:A | a | g | -0.1108 | 0.027 | 4.18E-05 | -+-------+-+- | 13.568 | 16 | 0.6309 |
| AD Braak | chr6 | 47,484,147 | chr6:47484147:C:T | t | c | -0.1103 | 0.027 | 4.40E-05 | -+-------+-+- | 13.512 | 16 | 0.635 |
| AD Braak | chr6 | 47,484,534 | chr6:47484534:T:C | t | c | 0.11 | 0.027 | 4.58E-05 | +-+++++++- | 13.52 | 16 | 0.6344 |
| AD Braak | chr6 | 47,485,642 | chr6:47485642:G:C | c | g | -0.1101 | 0.027 | 4.65E-05 | -+-------+-+- | 13.512 | 16 | 0.635 |
| AD Braak | chr6 | 47,488,382 | chr6:47488382:A:G | a | g | 0.1074 | 0.0269 | 6.68E-05 | +-+++++++- | 14.401 | 16 | 0.5689 |
| AD Braak | chr6 | 47,494,177 | chr6:47494177:C:A | a | c | -0.1068 | 0.0269 | 7.26E-05 | -+-------+-+- | 14.304 | 16 | 0.5761 |

| | | | | | | | | | | | | |
|---|---|---|---|---|---|---|---|---|---|---|---|---|
| AD Braak | chr6 | 47,495,390 | chr6:47495390:G:T | t | g | -0.1069 | 0.0269 | 7.20E-05 | -+-------+-+- | 14.27 | 16 | 0.5786 |
| AD Braak | chr6 | 47,497,531 | chr6:47497531:T:C | t | c | 0.107 | 0.0269 | 7.08E-05 | +-++++++++- | 14.308 | 16 | 0.5758 |
| AD Braak | chr6 | 47,501,537 | chr6:47501537:A:G | a | g | 0.107 | 0.0269 | 7.02E-05 | +-++++++++- | 14.301 | 16 | 0.5763 |
| AD Braak | chr6 | 47,504,687 | chr6:47504687:G:GAGA | g | gaga | 0.1073 | 0.027 | 6.92E-05 | +-++++++++- | 14.359 | 16 | 0.572 |
| AD Braak | chr6 | 47,505,471 | chr6:47505471:C:T | t | c | -0.107 | 0.0269 | 7.06E-05 | -+-------+-+- | 14.344 | 16 | 0.5731 |
| AD Braak | chr6 | 47,507,226 | chr6:47507226:C:T | t | c | -0.1066 | 0.0269 | 7.39E-05 | -+-------+-+- | 14.361 | 16 | 0.5718 |
| AD Braak | chr6 | 47,507,603 | chr6:47507603:G:A | a | g | -0.1067 | 0.0269 | 7.33E-05 | -+-------+-+- | 14.412 | 16 | 0.5681 |
| AD Braak | chr6 | 47,511,400 | chr6:47511400:G:A | a | g | -0.1094 | 0.0271 | 5.32E-05 | -+-------+-+- | 14.631 | 16 | 0.5518 |
| AD Braak | chr6 | 47,511,882 | chr6:47511882:C:T | t | c | -0.1062 | 0.0269 | 7.81E-05 | -+-------+-+- | 14.229 | 16 | 0.5817 |
| AD Braak | chr6 | 47,512,940 | chr6:47512940:A:G | a | g | 0.1094 | 0.027 | 5.07E-05 | +-++++++++- | 13.29 | 16 | 0.6515 |
| AD Braak | chr6 | 47,513,239 | chr6:47513239:C:G | c | g | 0.1062 | 0.0269 | 7.81E-05 | +-++++++++- | 14.233 | 16 | 0.5814 |
| AD Braak | chr6 | 47,514,097 | chr6:47514097:C:G | c | g | 0.1093 | 0.027 | 5.05E-05 | +-++++++++- | 13.286 | 16 | 0.6518 |
| AD Braak | chr6 | 47,515,917 | chr6:47515917:C:G | c | g | 0.1094 | 0.027 | 5.04E-05 | +-++++++++- | 13.274 | 16 | 0.6526 |
| AD Braak | chr6 | 47,517,266 | chr6:47517266:T:C | t | c | 0.1058 | 0.0269 | 8.32E-05 | +-++++++++- | 14.244 | 16 | 0.5806 |
| AD Braak | chr6 | 47,517,390 | chr6:47517390:C:T | t | c | -0.1091 | 0.027 | 5.28E-05 | -+-------+-+- | 13.464 | 16 | 0.6386 |
| AD Braak | chr6 | 47,519,361 | chr6:47519361:TGTGATG:T | t | tgtgatg | -0.1058 | 0.0269 | 8.42E-05 | -+-------+-+- | 14.265 | 16 | 0.579 |
| AD Braak | chr6 | 47,520,026 | chr6:47520026:A:G | a | g | 0.1086 | 0.0269 | 5.57E-05 | +-++++++++- | 13.159 | 16 | 0.6611 |
| AD Braak | chr6 | 47,521,202 | chr6:47521202:G:A | a | g | -0.1061 | 0.0269 | 7.87E-05 | -+-------+-+- | 14.25 | 16 | 0.5801 |
| AD Braak | chr6 | 47,521,368 | chr6:47521368:A:G | a | g | 0.1093 | 0.027 | 5.09E-05 | +-++++++++- | 13.3 | 16 | 0.6507 |
| AD Braak | chr6 | 47,526,204 | chr6:47526204:G:A | a | g | -0.1061 | 0.0269 | 7.95E-05 | -+-------+-+- | 14.234 | 16 | 0.5813 |
| AD Braak | chr6 | 47,527,023 | chr6:47527023:A:G | a | g | 0.1061 | 0.0269 | 7.93E-05 | +-++++++++- | 14.232 | 16 | 0.5814 |
| AD Braak | chr6 | 47,534,288 | chr6:47534288:G:C | c | g | -0.1067 | 0.0269 | 7.23E-05 | -+-------+-+- | 14.25 | 16 | 0.5801 |
| AD Braak | chr6 | 47,535,761 | chr6:47535761:A:T | a | t | 0.1073 | 0.0269 | 6.64E-05 | +-++++++++- | 14.387 | 16 | 0.5699 |
| AD Braak | chr6 | 47,537,274 | chr6:47537274:C:T | t | c | -0.1063 | 0.0269 | 7.74E-05 | -+-------+-+- | 14.263 | 16 | 0.5791 |
| AD Braak | chr6 | 47,543,755 | chr6:47543755:A:G | a | g | 0.1092 | 0.0271 | 5.67E-05 | +-++++++++- | 14.772 | 16 | 0.5414 |
| AD Braak | chr6 | 47,546,510 | chr6:47546510:T:A | a | t | -0.106 | 0.0269 | 8.07E-05 | -+-------+-+- | 14.198 | 16 | 0.584 |
| AD Braak | chr6 | 47,547,022 | chr6:47547022:G:A | a | g | -0.106 | 0.0269 | 8.06E-05 | -+-------+-+- | 14.183 | 16 | 0.585 |
| AD Braak | chr6 | 47,547,894 | chr6:47547894:G:A | a | g | -0.108 | 0.0269 | 5.96E-05 | -+-------+-+- | 15.072 | 16 | 0.5194 |
| AD Braak | chr6 | 47,547,927 | chr6:47547927:T:C | t | c | 0.1061 | 0.0269 | 8.01E-05 | +-++++++++- | 14.209 | 16 | 0.5831 |
| AD Braak | chr6 | 47,548,075 | chr6:47548075:A:G | a | g | 0.1085 | 0.027 | 5.86E-05 | +-++++++++- | 13.405 | 16 | 0.643 |
| AD Braak | chr6 | 47,548,633 | chr6:47548633:A:G | a | g | 0.1066 | 0.0269 | 7.45E-05 | +-++++++++- | 14.205 | 16 | 0.5834 |
| AD Braak | chr6 | 47,561,028 | chr6:47561028:C:G | c | g | 0.1065 | 0.0269 | 7.58E-05 | +-++++++++- | 14.065 | 16 | 0.5939 |
| AD Braak | chr6 | 47,568,708 | chr6:47568708:C:T | t | c | -0.1066 | 0.0269 | 7.41E-05 | -+-------+-+- | 14.2 | 16 | 0.5838 |
| AD Braak | chr6 | 47,570,141 | chr6:47570141:CT:C | ct | c | 0.1067 | 0.0269 | 7.19E-05 | --++++++++- | 15.348 | 16 | 0.4993 |
| AD Braak | chr6 | 47,573,679 | chr6:47573679:C:G | c | g | 0.1068 | 0.0269 | 7.26E-05 | +-++++++++- | 14.281 | 16 | 0.5778 |
| AD Braak | chr6 | 47,575,128 | chr6:47575128:C:T | t | c | -0.1068 | 0.0269 | 7.19E-05 | -+-------+-+- | 14.241 | 16 | 0.5808 |
| AD Braak | chr6 | 47,581,759 | chr6:47581759:C:T | t | c | -0.1065 | 0.0269 | 7.50E-05 | -+-------+-+- | 14.191 | 16 | 0.5845 |
| AD Braak | chr6 | 47,583,708 | chr6:47583708:T:C | t | c | 0.1066 | 0.0269 | 7.44E-05 | +-++++++++- | 14.199 | 16 | 0.5839 |
| AD Braak | chr6 | 47,584,125 | chr6:47584125:T:C | t | c | 0.1067 | 0.0269 | 7.40E-05 | +-++++++++- | 14.205 | 16 | 0.5835 |
| AD Braak | chr6 | 47,584,202 | chr6:47584202:G:A | a | g | -0.1067 | 0.0269 | 7.40E-05 | -+-------+-+- | 14.205 | 16 | 0.5835 |
| AD Braak | chr6 | 47,584,444 | chr6:47584444:A:T | a | t | 0.1095 | 0.027 | 4.97E-05 | +-++++++++- | 13.587 | 16 | 0.6294 |
| AD Braak | chr6 | 47,585,081 | chr6:47585081:C:A | a | c | -0.1067 | 0.0269 | 7.39E-05 | -+-------+-+- | 14.202 | 16 | 0.5837 |
| AD Braak | chr6 | 47,585,666 | chr6:47585666:G:A | a | g | -0.109 | 0.0271 | 5.80E-05 | -+-------+-+- | 14.455 | 16 | 0.5649 |
| AD Braak | chr6 | 47,586,441 | chr6:47586441:G:A | a | g | -0.1091 | 0.0271 | 5.58E-05 | -+-------+-+- | 14.424 | 16 | 0.5672 |
| AD Braak | chr6 | 47,586,732 | chr6:47586732:T:G | t | g | 0.1091 | 0.0271 | 5.67E-05 | +-++++++++- | 14.427 | 16 | 0.5669 |
| AD Braak | chr6 | 47,588,894 | chr6:47588894:A:C | a | c | 0.1069 | 0.0269 | 7.07E-05 | +-++++++++- | 14.39 | 16 | 0.5697 |

| | | | | | | | | | | | | |
|---|---|---|---|---|---|---|---|---|---|---|---|---|
| AD Braak | chr6 | 47,588,898 | chr6:47588898:C:T | t | c | -0.1067 | 0.0269 | 7.36E-05 | -+-------+-+- | 14.259 | 16 | 0.5794 |
| AD Braak | chr6 | 47,588,944 | chr6:47588944:C:T | t | c | -0.1068 | 0.0269 | 7.27E-05 | -+-------+-+- | 14.256 | 16 | 0.5796 |
| AD Braak | chr6 | 47,591,715 | chr6:47591715:A:G | a | g | 0.1071 | 0.0269 | 6.98E-05 | +-++++++++- | 14.171 | 16 | 0.586 |
| AD Braak | chr6 | 47,595,179 | chr6:47595179:A:G | a | g | 0.1068 | 0.0269 | 7.28E-05 | +-++++++++- | 14.32 | 16 | 0.5749 |
| AD Braak | chr6 | 47,600,960 | chr6:47600960:A:G | a | g | 0.1077 | 0.0269 | 6.40E-05 | +-++++++++- | 14.313 | 16 | 0.5754 |
| AD Braak | chr6 | 47,607,596 | chr6:47607596:T:C | t | c | 0.1086 | 0.0269 | 5.55E-05 | +-++++++++- | 14.443 | 16 | 0.5657 |
| AD Braak | chr6 | 47,608,631 | chr6:47608631:T:C | t | c | 0.1082 | 0.0269 | 5.88E-05 | +-++++++++- | 14.562 | 16 | 0.5569 |
| AD Braak | chr6 | 47,612,921 | chr6:47612921:A:G | a | g | 0.1073 | 0.0269 | 6.76E-05 | +-++++++++- | 14.058 | 16 | 0.5944 |
| AD Braak | chr6 | 47,612,959 | chr6:47612959:G:A | a | g | -0.1079 | 0.0269 | 6.19E-05 | -+-------+-+- | 14.328 | 16 | 0.5743 |
| AD Braak | chr6 | 47,615,928 | chr6:47615928:C:T | t | c | -0.1111 | 0.0272 | 4.32E-05 | -+-------+-+- | 15.223 | 16 | 0.5084 |
| AD Braak | chr6 | 47,617,370 | chr6:47617370:A:T | a | t | 0.1072 | 0.0269 | 6.95E-05 | +-++++++++- | 14.277 | 16 | 0.5781 |
| AD Braak | chr6 | 47,617,879 | chr6:47617879:A:G | a | g | 0.1069 | 0.0269 | 7.06E-05 | +-++++++++- | 14.392 | 16 | 0.5695 |
| AD Braak | chr6 | 47,618,176 | chr6:47618176:T:C | t | c | 0.109 | 0.0269 | 5.27E-05 | +-++++++++- | 13.942 | 16 | 0.603 |
| AD Braak | chr6 | 47,619,313 | chr6:47619313:AACAT:A | a | aacat | -0.1068 | 0.0269 | 7.33E-05 | -+---------+- | 14.554 | 16 | 0.5575 |
| AD Braak | chr6 | 47,619,558 | chr6:47619558:ACTT:A | a | actt | -0.1065 | 0.0269 | 7.71E-05 | -+-------+-+- | 14.693 | 16 | 0.5473 |
| AD Braak | chr6 | 47,622,368 | chr6:47622368:C:T | t | c | -0.1073 | 0.027 | 6.89E-05 | -+-------+-+- | 13.946 | 16 | 0.6027 |
| AD Braak | chr6 | 47,622,740 | chr6:47622740:C:T | t | c | -0.1075 | 0.027 | 6.60E-05 | -+-------+-+- | 13.95 | 16 | 0.6025 |
| AD Braak | chr6 | 47,701,044 | chr6:47701044:A:C | a | c | 0.1366 | 0.0304 | 6.99E-06 | +++++++++ | 11.749 | 16 | 0.7611 |
| AD Braak | chr6 | 76,657,585 | chr6:76657585:A:G | a | g | -0.1581 | 0.0405 | 9.52E-05 | ----+----+-+- | 22.48 | 16 | 0.1284 |
| AD Braak | chr6 | 133,827,449 | chr6:133827449:TAAG:T | t | taag | 0.3769 | 0.0941 | 6.22E-05 | ++++++-++- | 9.29 | 16 | 0.901 |
| AD Braak | chr6 | 154,927,895 | chr6:154927895:C:T | t | c | 0.2638 | 0.0639 | 3.59E-05 | --++-++++-- | 20.823 | 16 | 0.1854 |
| AD Braak | chr6 | 154,929,907 | chr6:154929907:G:A | a | g | 0.2692 | 0.0678 | 7.23E-05 | --++-++++-- | 21.15 | 16 | 0.1728 |
| AD Braak | chr6 | 154,930,845 | chr6:154930845:G:A | a | g | 0.2599 | 0.0641 | 5.05E-05 | --++-++++-- | 20.915 | 16 | 0.1818 |
| AD Braak | chr6 | 154,930,953 | chr6:154930953:G:A | a | g | 0.2621 | 0.065 | 5.53E-05 | --++-++++-- | 20.154 | 16 | 0.2134 |
| AD Braak | chr6 | 155,440,484 | chr6:155440484:A:T | a | t | 0.1021 | 0.025 | 4.48E-05 | ++++-++-+- | 14.716 | 16 | 0.5455 |
| AD Braak | chr6 | 157,733,254 | chr6:157733254:T:C | t | c | 0.0971 | 0.0245 | 7.53E-05 | ++++---+++ | 18.337 | 16 | 0.3046 |
| AD Braak | chr6 | 157,733,357 | chr6:157733357:A:G | a | g | 0.0975 | 0.0245 | 7.01E-05 | ++++---+++ | 18.584 | 16 | 0.2908 |
| AD Braak | chr6 | 157,735,038 | chr6:157735038:C:G | c | g | 0.0982 | 0.0245 | 6.16E-05 | +-++---+++- | 17.855 | 16 | 0.3324 |
| AD Braak | chr6 | 157,735,316 | chr6:157735316:G:C | c | g | -0.0987 | 0.0245 | 5.61E-05 | -+--+++----- | 17.4 | 16 | 0.3602 |
| AD Braak | chr6 | 157,735,460 | chr6:157735460:A:G | a | g | 0.0975 | 0.0245 | 6.87E-05 | +-++---+++- | 17.84 | 16 | 0.3333 |
| AD Braak | chr6 | 157,736,720 | chr6:157736720:G:C | c | g | -0.0989 | 0.0245 | 5.28E-05 | -+--+++----- | 17.228 | 16 | 0.371 |
| AD Braak | chr6 | 157,738,311 | chr6:157738311:C:T | t | c | -0.0983 | 0.0245 | 6.00E-05 | -+--+++----- | 16.951 | 16 | 0.3888 |
| AD Braak | chr6 | 157,738,464 | chr6:157738464:CT:C | ct | c | 0.0977 | 0.0245 | 6.71E-05 | +-++---+++- | 16.853 | 16 | 0.3952 |
| AD Braak | chr6 | 157,738,573 | chr6:157738573:C:G | c | g | 0.0983 | 0.0245 | 5.93E-05 | +-++---+++- | 16.85 | 16 | 0.3954 |
| AD Braak | chr6 | 157,738,615 | chr6:157738615:C:T | t | c | -0.0996 | 0.0245 | 4.74E-05 | -+--+++----- | 17.444 | 16 | 0.3574 |
| AD Braak | chr6 | 157,738,739 | chr6:157738739:G:A | a | g | -0.0987 | 0.0245 | 5.56E-05 | -+--+++----- | 17.403 | 16 | 0.36 |
| AD Braak | chr6 | 157,738,870 | chr6:157738870:T:C | t | c | 0.098 | 0.0245 | 6.31E-05 | +-++---+++- | 16.891 | 16 | 0.3927 |
| AD Braak | chr6 | 157,739,148 | chr6:157739148:A:G | a | g | 0.0989 | 0.0245 | 5.35E-05 | +-++---+++- | 17.37 | 16 | 0.362 |
| AD Braak | chr6 | 157,739,520 | chr6:157739520:T:C | t | c | 0.0988 | 0.0245 | 5.46E-05 | +-++---+++- | 16.932 | 16 | 0.39 |
| AD Braak | chr6 | 157,739,636 | chr6:157739636:T:G | t | g | 0.0983 | 0.0245 | 5.92E-05 | +-++---+++- | 16.897 | 16 | 0.3923 |
| AD Braak | chr6 | 157,740,013 | chr6:157740013:G:A | a | g | -0.0983 | 0.0245 | 5.92E-05 | -+--+++----- | 16.9 | 16 | 0.3921 |
| AD Braak | chr6 | 157,740,093 | chr6:157740093:TATA:T | t | tata | -0.098 | 0.0245 | 6.25E-05 | -+--+++----- | 17.575 | 16 | 0.3493 |
| AD Braak | chr6 | 157,740,528 | chr6:157740528:G:A | a | g | -0.0987 | 0.0245 | 5.56E-05 | -+--+++----- | 17.412 | 16 | 0.3594 |
| AD Braak | chr6 | 157,740,942 | chr6:157740942:G:A | a | g | -0.0984 | 0.0245 | 5.88E-05 | -+--+++----- | 17.672 | 16 | 0.3435 |
| AD Braak | chr6 | 157,741,162 | chr6:157741162:C:T | t | c | -0.0985 | 0.0245 | 5.72E-05 | -+--+++----- | 17.851 | 16 | 0.3327 |
| AD Braak | chr6 | 157,741,653 | chr6:157741653:G:A | a | g | -0.0983 | 0.0245 | 5.98E-05 | -+--+++----- | 17.653 | 16 | 0.3446 |

| | | | | | | | | | | | | |
|---|---|---|---|---|---|---|---|---|---|---|---|---|
| AD Braak | chr6 | 157,742,175 | chr6:157742175:G:T | t | g | -0.0969 | 0.0243 | 6.73E-05 | -+--+++----- | 17.929 | 16 | 0.3281 |
| AD Braak | chr6 | 157,742,238 | chr6:157742238:T:G | t | g | 0.098 | 0.0245 | 6.28E-05 | +-++---+++- | 17.627 | 16 | 0.3462 |
| AD Braak | chr6 | 157,745,974 | chr6:157745974:C:T | t | c | -0.0981 | 0.0245 | 6.20E-05 | -+--+++----- | 18.35 | 16 | 0.3038 |
| AD Braak | chr6 | 157,748,798 | chr6:157748798:C:T | t | c | -0.0994 | 0.0245 | 4.93E-05 | -+--+++----- | 17.172 | 16 | 0.3746 |
| AD Braak | chr6 | 157,748,868 | chr6:157748868:A:C | a | c | 0.0987 | 0.0245 | 5.47E-05 | +-++---+++- | 17.77 | 16 | 0.3375 |
| AD Braak | chr6 | 157,752,055 | chr6:157752055:G:A | a | g | -0.0977 | 0.0245 | 6.77E-05 | -+--+++----- | 18.09 | 16 | 0.3187 |
| AD Braak | chr6 | 157,981,669 | chr6:157981669:G:T | t | g | 0.148 | 0.0365 | 5.05E-05 | +-++-+-+++ | 31.269 | 16 | 0.01243 |
| AD Braak | chr6 | 160,135,623 | chr6:160135623:C:T | t | c | 0.607 | 0.1525 | 6.88E-05 | +-+++???? | 9.917 | 7 | 0.1933 |
| AD Braak | chr6 | 165,430,823 | chr6:165430823:T:C | t | c | -0.1072 | 0.0272 | 8.33E-05 | ---------+--- | 11.777 | 16 | 0.7592 |
| AD Braak | chr6 | 170,041,509 | chr6:170041509:C:T | t | c | -0.0992 | 0.0253 | 8.97E-05 | +----------- | 13.18 | 16 | 0.6596 |
| AD Braak | chr7 | 9,555,355 | chr7:9555355:A:G | a | g | -0.119 | 0.0283 | 2.68E-05 | ------++----- | 12.717 | 16 | 0.6933 |
| AD Braak | chr7 | 12,211,934 | chr7:12211934:C:G | c | g | 0.0962 | 0.0245 | 8.32E-05 | +++++++-+- | 13.437 | 16 | 0.6406 |
| AD Braak | chr7 | 12,212,045 | chr7:12212045:T:A | a | t | -0.0965 | 0.0245 | 7.92E-05 | -------+------ | 13.563 | 16 | 0.6313 |
| AD Braak | chr7 | 12,212,164 | chr7:12212164:G:C | c | g | -0.0959 | 0.0244 | 8.72E-05 | -------+------ | 13.611 | 16 | 0.6276 |
| AD Braak | chr7 | 12,212,914 | chr7:12212914:C:G | c | g | 0.0961 | 0.0245 | 8.49E-05 | +++++++-+- | 12.036 | 16 | 0.7415 |
| AD Braak | chr7 | 12,213,033 | chr7:12213033:G:A | a | g | -0.0966 | 0.0245 | 7.85E-05 | -------+------ | 12.062 | 16 | 0.7397 |
| AD Braak | chr7 | 12,213,115 | chr7:12213115:T:G | t | g | 0.0957 | 0.0245 | 9.03E-05 | +++++++-+- | 12.158 | 16 | 0.733 |
| AD Braak | chr7 | 12,213,267 | chr7:12213267:A:G | a | g | 0.0966 | 0.0245 | 7.87E-05 | +++++++-+- | 12.048 | 16 | 0.7407 |
| AD Braak | chr7 | 12,213,276 | chr7:12213276:A:ACT | a | act | 0.097 | 0.0245 | 7.36E-05 | +++++++-+- | 12.125 | 16 | 0.7353 |
| AD Braak | chr7 | 12,213,424 | chr7:12213424:A:C | a | c | 0.0962 | 0.0245 | 8.43E-05 | +++++++-+- | 12.148 | 16 | 0.7337 |
| AD Braak | chr7 | 12,213,462 | chr7:12213462:A:G | a | g | 0.0973 | 0.0244 | 6.75E-05 | +++++++-+- | 11.804 | 16 | 0.7574 |
| AD Braak | chr7 | 12,213,736 | chr7:12213736:T:C | t | c | 0.0973 | 0.0244 | 6.76E-05 | +++++++-+- | 11.802 | 16 | 0.7575 |
| AD Braak | chr7 | 12,213,746 | chr7:12213746:T:C | t | c | 0.0973 | 0.0244 | 6.76E-05 | +++++++-+- | 11.803 | 16 | 0.7574 |
| AD Braak | chr7 | 12,213,917 | chr7:12213917:T:C | t | c | 0.0972 | 0.0244 | 6.85E-05 | +++++++-+- | 11.784 | 16 | 0.7587 |
| AD Braak | chr7 | 12,213,937 | chr7:12213937:C:T | t | c | -0.0973 | 0.0244 | 6.83E-05 | -------+------ | 11.742 | 16 | 0.7615 |
| AD Braak | chr7 | 12,214,254 | chr7:12214254:G:A | a | g | -0.0954 | 0.0244 | 9.39E-05 | -------+------ | 11.672 | 16 | 0.7662 |
| AD Braak | chr7 | 12,215,808 | chr7:12215808:C:T | t | c | -0.099 | 0.0244 | 4.98E-05 | -------+------ | 11.336 | 16 | 0.7883 |
| AD Braak | chr7 | 12,215,885 | chr7:12215885:G:A | a | g | -0.0983 | 0.0244 | 5.61E-05 | -------+------ | 11.424 | 16 | 0.7826 |
| AD Braak | chr7 | 12,216,038 | chr7:12216038:G:A | a | g | -0.0992 | 0.0244 | 4.87E-05 | -------+------ | 11.42 | 16 | 0.7828 |
| AD Braak | chr7 | 12,216,922 | chr7:12216922:T:C | t | c | 0.0982 | 0.0244 | 5.81E-05 | +++++++-+- | 11.341 | 16 | 0.7879 |
| AD Braak | chr7 | 12,217,901 | chr7:12217901:C:T | t | c | -0.0983 | 0.0244 | 5.69E-05 | -------+------ | 11.374 | 16 | 0.7858 |
| AD Braak | chr7 | 12,217,923 | chr7:12217923:A:G | a | g | 0.097 | 0.0244 | 7.15E-05 | +++++++-+- | 11.344 | 16 | 0.7878 |
| AD Braak | chr7 | 12,219,039 | chr7:12219039:T:TA | t | ta | 0.1 | 0.0244 | 4.24E-05 | +++++++-+- | 11.541 | 16 | 0.7749 |
| AD Braak | chr7 | 12,219,129 | chr7:12219129:G:C | c | g | -0.0991 | 0.0244 | 4.91E-05 | -------+------ | 11.184 | 16 | 0.798 |
| AD Braak | chr7 | 12,222,203 | chr7:12222203:A:G | a | g | 0.0974 | 0.0244 | 6.60E-05 | +++++++-+- | 10.729 | 16 | 0.8259 |
| AD Braak | chr7 | 12,222,616 | chr7:12222616:G:A | a | g | -0.0969 | 0.0244 | 7.17E-05 | -------+------ | 10.737 | 16 | 0.8254 |
| AD Braak | chr7 | 12,222,945 | chr7:12222945:C:T | t | c | -0.0966 | 0.0244 | 7.52E-05 | -------+------ | 10.785 | 16 | 0.8226 |
| AD Braak | chr7 | 12,223,091 | chr7:12223091:T:C | t | c | 0.0972 | 0.0244 | 6.82E-05 | +++++++-+- | 10.78 | 16 | 0.8229 |
| AD Braak | chr7 | 12,223,175 | chr7:12223175:G:A | a | g | -0.0971 | 0.0244 | 6.98E-05 | -------+------ | 10.772 | 16 | 0.8233 |
| AD Braak | chr7 | 12,223,342 | chr7:12223342:AT:A | a | at | -0.097 | 0.0244 | 7.16E-05 | -------+------ | 10.846 | 16 | 0.8189 |
| AD Braak | chr7 | 12,223,557 | chr7:12223557:TA:T | t | ta | -0.0972 | 0.0244 | 6.87E-05 | -------+------ | 10.787 | 16 | 0.8224 |
| AD Braak | chr7 | 12,223,752 | chr7:12223752:G:T | t | g | -0.0966 | 0.0244 | 7.67E-05 | -------+------ | 10.878 | 16 | 0.817 |
| AD Braak | chr7 | 12,223,811 | chr7:12223811:C:T | t | c | -0.0971 | 0.0244 | 6.97E-05 | -------+------ | 10.783 | 16 | 0.8227 |
| AD Braak | chr7 | 12,223,912 | chr7:12223912:T:C | t | c | 0.097 | 0.0244 | 7.09E-05 | +++++++-+- | 10.776 | 16 | 0.8231 |
| AD Braak | chr7 | 12,223,920 | chr7:12223920:T:C | t | c | 0.0972 | 0.0244 | 6.87E-05 | +++++++-+- | 10.792 | 16 | 0.8221 |
| AD Braak | chr7 | 12,223,961 | chr7:12223961:C:T | t | c | -0.0971 | 0.0244 | 6.97E-05 | -------+------ | 10.797 | 16 | 0.8218 |

| | | | | | | | | | | | | |
|---|---|---|---|---|---|---|---|---|---|---|---|---|
| AD Braak | chr7 | 12,224,173 | chr7:12224173:T:C | t | c | 0.0969 | 0.0244 | 7.22E-05 | +++++++-+- | 10.761 | 16 | 0.824 |
| AD Braak | chr7 | 12,224,174 | chr7:12224174:G:A | a | g | -0.0969 | 0.0244 | 7.22E-05 | -------+------ | 10.761 | 16 | 0.824 |
| AD Braak | chr7 | 12,224,635 | chr7:12224635:T:TA | t | ta | 0.0999 | 0.0245 | 4.53E-05 | +++++++-+- | 11.12 | 16 | 0.802 |
| AD Braak | chr7 | 12,224,671 | chr7:12224671:G:A | a | g | -0.0969 | 0.0244 | 7.16E-05 | -------+------ | 10.771 | 16 | 0.8234 |
| AD Braak | chr7 | 12,224,841 | chr7:12224841:T:C | t | c | 0.0972 | 0.0244 | 6.85E-05 | +++++++-+- | 10.787 | 16 | 0.8224 |
| AD Braak | chr7 | 12,225,040 | chr7:12225040:T:C | t | c | 0.0967 | 0.0244 | 7.39E-05 | +++++++-+- | 10.764 | 16 | 0.8238 |
| AD Braak | chr7 | 12,225,111 | chr7:12225111:C:T | t | c | -0.098 | 0.0244 | 5.86E-05 | -------+------ | 10.336 | 16 | 0.8485 |
| AD Braak | chr7 | 12,225,245 | chr7:12225245:G:A | a | g | -0.0951 | 0.0244 | 9.73E-05 | -------+------ | 10.778 | 16 | 0.823 |
| AD Braak | chr7 | 12,225,433 | chr7:12225433:A:G | a | g | 0.095 | 0.0244 | 9.82E-05 | +++++++-+- | 10.774 | 16 | 0.8232 |
| AD Braak | chr7 | 12,225,566 | chr7:12225566:T:C | t | c | 0.0966 | 0.0244 | 7.61E-05 | +++++++-+- | 10.771 | 16 | 0.8234 |
| AD Braak | chr7 | 12,226,222 | chr7:12226222:T:C | t | c | 0.097 | 0.0244 | 7.09E-05 | +++++++-+- | 11.087 | 16 | 0.8041 |
| AD Braak | chr7 | 12,226,326 | chr7:12226326:A:T | a | t | 0.097 | 0.0244 | 7.08E-05 | +++++++-+- | 11.09 | 16 | 0.8039 |
| AD Braak | chr7 | 12,226,362 | chr7:12226362:C:T | t | c | -0.0989 | 0.0244 | 4.84E-05 | -------+------ | 10.324 | 16 | 0.8492 |
| AD Braak | chr7 | 12,226,925 | chr7:12226925:TA:T | t | ta | -0.0971 | 0.0244 | 6.97E-05 | -------+------ | 10.812 | 16 | 0.8209 |
| AD Braak | chr7 | 12,227,188 | chr7:12227188:A:G | a | g | 0.0971 | 0.0244 | 6.94E-05 | +++++++-+- | 11.353 | 16 | 0.7872 |
| AD Braak | chr7 | 12,227,241 | chr7:12227241:G:T | t | g | -0.0983 | 0.0244 | 5.54E-05 | -------+------ | 10.235 | 16 | 0.8541 |
| AD Braak | chr7 | 12,227,595 | chr7:12227595:A:G | a | g | 0.0966 | 0.0244 | 7.50E-05 | +++++++-+- | 10.772 | 16 | 0.8234 |
| AD Braak | chr7 | 12,227,630 | chr7:12227630:C:T | t | c | -0.0968 | 0.0244 | 7.34E-05 | -------+------ | 10.668 | 16 | 0.8295 |
| AD Braak | chr7 | 12,227,869 | chr7:12227869:G:C | c | g | -0.0992 | 0.0244 | 4.78E-05 | -------+------ | 10.168 | 16 | 0.8577 |
| AD Braak | chr7 | 12,227,926 | chr7:12227926:G:A | a | g | -0.0983 | 0.0244 | 5.54E-05 | -------+------ | 10.221 | 16 | 0.8548 |
| AD Braak | chr7 | 12,227,933 | chr7:12227933:G:C | c | g | -0.0986 | 0.0244 | 5.26E-05 | -------+------ | 10.268 | 16 | 0.8522 |
| AD Braak | chr7 | 12,227,988 | chr7:12227988:T:C | t | c | 0.0983 | 0.0244 | 5.55E-05 | +++++++-+- | 10.232 | 16 | 0.8542 |
| AD Braak | chr7 | 12,228,108 | chr7:12228108:G:A | a | g | -0.0987 | 0.0244 | 5.24E-05 | -------+------ | 10.221 | 16 | 0.8548 |
| AD Braak | chr7 | 12,228,211 | chr7:12228211:A:G | a | g | 0.0985 | 0.0244 | 5.42E-05 | +++++++-+- | 10.44 | 16 | 0.8427 |
| AD Braak | chr7 | 12,228,456 | chr7:12228456:CTTAA:C | cttaa | c | 0.0985 | 0.0244 | 5.42E-05 | +++++++-+- | 10.228 | 16 | 0.8545 |
| AD Braak | chr7 | 12,228,491 | chr7:12228491:C:T | t | c | -0.0984 | 0.0244 | 5.54E-05 | -------+------ | 10.169 | 16 | 0.8576 |
| AD Braak | chr7 | 12,228,617 | chr7:12228617:C:A | a | c | -0.0982 | 0.0244 | 5.74E-05 | -------+------ | 10.264 | 16 | 0.8525 |
| AD Braak | chr7 | 12,228,842 | chr7:12228842:G:T | t | g | -0.0984 | 0.0244 | 5.43E-05 | -------+------ | 10.518 | 16 | 0.8382 |
| AD Braak | chr7 | 12,228,925 | chr7:12228925:G:A | a | g | -0.0987 | 0.0244 | 5.21E-05 | -------+------ | 10.522 | 16 | 0.838 |
| AD Braak | chr7 | 12,229,042 | chr7:12229042:A:C | a | c | 0.0957 | 0.0244 | 8.59E-05 | +++++++-+- | 10.996 | 16 | 0.8097 |
| AD Braak | chr7 | 12,229,091 | chr7:12229091:C:T | t | c | -0.0985 | 0.0244 | 5.38E-05 | -------+------ | 10.54 | 16 | 0.8369 |
| AD Braak | chr7 | 12,229,132 | chr7:12229132:A:C | a | c | 0.0968 | 0.0244 | 7.29E-05 | +++++++-+- | 11.108 | 16 | 0.8028 |
| AD Braak | chr7 | 12,229,166 | chr7:12229166:A:T | a | t | 0.0982 | 0.0244 | 5.64E-05 | +++++++-+- | 10.587 | 16 | 0.8342 |
| AD Braak | chr7 | 12,229,172 | chr7:12229172:C:T | t | c | -0.0985 | 0.0244 | 5.37E-05 | -------+------ | 10.539 | 16 | 0.837 |
| AD Braak | chr7 | 12,229,185 | chr7:12229185:G:C | c | g | -0.0981 | 0.0244 | 5.78E-05 | -------+------ | 10.395 | 16 | 0.8452 |
| AD Braak | chr7 | 12,229,791 | chr7:12229791:C:G | c | g | 0.0983 | 0.0244 | 5.55E-05 | +++++++-+- | 10.526 | 16 | 0.8378 |
| AD Braak | chr7 | 12,229,949 | chr7:12229949:G:C | c | g | -0.0985 | 0.0244 | 5.36E-05 | -------+------ | 10.539 | 16 | 0.837 |
| AD Braak | chr7 | 12,229,967 | chr7:12229967:C:A | a | c | -0.0968 | 0.0244 | 7.13E-05 | -------+------ | 10.587 | 16 | 0.8342 |
| AD Braak | chr7 | 12,230,053 | chr7:12230053:C:CA | ca | c | -0.1009 | 0.0245 | 3.82E-05 | -------+------ | 11.163 | 16 | 0.7993 |
| AD Braak | chr7 | 12,230,136 | chr7:12230136:G:T | t | g | -0.0987 | 0.0244 | 5.16E-05 | -------+------ | 10.544 | 16 | 0.8367 |
| AD Braak | chr7 | 12,230,178 | chr7:12230178:C:T | t | c | -0.0985 | 0.0244 | 5.38E-05 | -------+------ | 10.539 | 16 | 0.837 |
| AD Braak | chr7 | 12,230,191 | chr7:12230191:G:A | a | g | -0.0986 | 0.0244 | 5.24E-05 | -------+------ | 10.512 | 16 | 0.8386 |
| AD Braak | chr7 | 12,230,893 | chr7:12230893:G:A | a | g | -0.0985 | 0.0244 | 5.40E-05 | -------+------ | 10.531 | 16 | 0.8374 |
| AD Braak | chr7 | 12,230,939 | chr7:12230939:A:AT | a | at | 0.0974 | 0.0245 | 7.00E-05 | +++++++-+- | 10.434 | 16 | 0.843 |
| AD Braak | chr7 | 12,230,999 | chr7:12230999:G:A | a | g | -0.0987 | 0.0244 | 5.19E-05 | -------+------ | 10.643 | 16 | 0.831 |
| AD Braak | chr7 | 12,231,144 | chr7:12231144:T:A | a | t | -0.099 | 0.0244 | 4.95E-05 | -------+------ | 10.569 | 16 | 0.8353 |

| | | | | | | | | | | | | |
|---|---|---|---|---|---|---|---|---|---|---|---|---|
| AD Braak | chr7 | 12,231,189 | chr7:12231189:C:T | t | c | -0.099 | 0.0244 | 4.92E-05 | -------+------ | 10.557 | 16 | 0.836 |
| AD Braak | chr7 | 12,231,626 | chr7:12231626:T:C | t | c | 0.0965 | 0.0244 | 7.57E-05 | +++++++-+- | 11.159 | 16 | 0.7996 |
| AD Braak | chr7 | 12,232,371 | chr7:12232371:G:A | a | g | -0.0984 | 0.0244 | 5.46E-05 | -------+------ | 10.51 | 16 | 0.8386 |
| AD Braak | chr7 | 12,232,490 | chr7:12232490:C:G | c | g | 0.099 | 0.0244 | 4.93E-05 | +++++++-+- | 10.555 | 16 | 0.8361 |
| AD Braak | chr7 | 12,232,942 | chr7:12232942:A:T | a | t | 0.0984 | 0.0244 | 5.49E-05 | +++++++-+- | 10.546 | 16 | 0.8366 |
| AD Braak | chr7 | 12,233,787 | chr7:12233787:C:T | t | c | -0.0977 | 0.0244 | 6.22E-05 | -------+------ | 10.489 | 16 | 0.8398 |
| AD Braak | chr7 | 12,233,870 | chr7:12233870:G:A | a | g | -0.0979 | 0.0244 | 5.96E-05 | -------+------ | 10.326 | 16 | 0.8491 |
| AD Braak | chr7 | 12,234,787 | chr7:12234787:C:CT | ct | c | -0.0958 | 0.0246 | 9.60E-05 | -------+------ | 11.977 | 16 | 0.7456 |
| AD Braak | chr7 | 12,235,882 | chr7:12235882:T:C | t | c | 0.0969 | 0.0244 | 7.10E-05 | +++++++-+- | 10.982 | 16 | 0.8106 |
| AD Braak | chr7 | 12,235,998 | chr7:12235998:A:ATTTTC | a | attttc | 0.0978 | 0.0245 | 6.56E-05 | +++++++-+- | 9.879 | 16 | 0.8729 |
| AD Braak | chr7 | 12,236,049 | chr7:12236049:A:C | a | c | 0.0967 | 0.0244 | 7.34E-05 | +++++++-+- | 11.127 | 16 | 0.8016 |
| AD Braak | chr7 | 12,236,192 | chr7:12236192:A:G | a | g | 0.0962 | 0.0244 | 7.99E-05 | +++++++-+- | 11.012 | 16 | 0.8087 |
| AD Braak | chr7 | 12,236,385 | chr7:12236385:G:A | a | g | -0.0968 | 0.0244 | 7.17E-05 | -------+------ | 11.014 | 16 | 0.8086 |
| AD Braak | chr7 | 12,236,419 | chr7:12236419:G:C | c | g | -0.0971 | 0.0244 | 6.89E-05 | -------+------ | 11.03 | 16 | 0.8076 |
| AD Braak | chr7 | 12,237,259 | chr7:12237259:T:C | t | c | 0.098 | 0.0244 | 5.81E-05 | +++++++-+- | 10.555 | 16 | 0.8361 |
| AD Braak | chr7 | 12,238,147 | chr7:12238147:G:A | a | g | -0.0987 | 0.0244 | 5.15E-05 | -------+------ | 10.624 | 16 | 0.8321 |
| AD Braak | chr7 | 12,240,135 | chr7:12240135:A:C | a | c | 0.1 | 0.0244 | 4.05E-05 | +++++++-+- | 10.558 | 16 | 0.8359 |
| AD Braak | chr7 | 12,241,104 | chr7:12241104:T:A | a | t | -0.1003 | 0.0244 | 3.84E-05 | -------+------ | 10.648 | 16 | 0.8306 |
| AD Braak | chr7 | 12,242,825 | chr7:12242825:T:C | t | c | 0.0996 | 0.0244 | 4.53E-05 | +++++++-+- | 9.569 | 16 | 0.8881 |
| AD Braak | chr7 | 12,243,601 | chr7:12243601:A:G | a | g | 0.1007 | 0.0244 | 3.66E-05 | +++++++-+- | 10.212 | 16 | 0.8553 |
| AD Braak | chr7 | 12,243,650 | chr7:12243650:T:C | t | c | 0.1 | 0.0244 | 4.15E-05 | +++++++-+- | 10.167 | 16 | 0.8578 |
| AD Braak | chr7 | 12,243,703 | chr7:12243703:G:C | c | g | -0.1006 | 0.0244 | 3.74E-05 | -------+------ | 10.227 | 16 | 0.8545 |
| AD Braak | chr7 | 12,244,161 | chr7:12244161:A:G | a | g | 0.1006 | 0.0244 | 3.66E-05 | +++++++-+- | 10.124 | 16 | 0.8601 |
| AD Braak | chr7 | 12,244,247 | chr7:12244247:C:G | c | g | 0.0986 | 0.0244 | 5.34E-05 | +++++++-+- | 10.035 | 16 | 0.8648 |
| AD Braak | chr7 | 12,244,382 | chr7:12244382:A:G | a | g | 0.1004 | 0.0244 | 3.83E-05 | +++++++-+- | 10.174 | 16 | 0.8574 |
| AD Braak | chr7 | 12,244,752 | chr7:12244752:G:A | a | g | -0.1014 | 0.0245 | 3.58E-05 | -------+------ | 9.237 | 16 | 0.9033 |
| AD Braak | chr7 | 12,245,514 | chr7:12245514:C:T | t | c | -0.1008 | 0.0244 | 3.67E-05 | -------+------ | 10.026 | 16 | 0.8653 |
| AD Braak | chr7 | 52,736,121 | chr7:52736121:G:C | c | g | -0.2152 | 0.0516 | 3.06E-05 | -+-+--------- | 12.174 | 16 | 0.7319 |
| AD Braak | chr7 | 52,737,595 | chr7:52737595:C:T | t | c | -0.2152 | 0.0516 | 3.04E-05 | -+-+--------- | 12.182 | 16 | 0.7314 |
| AD Braak | chr7 | 52,739,191 | chr7:52739191:A:T | a | t | 0.2154 | 0.0516 | 3.00E-05 | +-+-++++++ | 12.182 | 16 | 0.7313 |
| AD Braak | chr7 | 52,739,826 | chr7:52739826:T:C | t | c | 0.2533 | 0.065 | 9.84E-05 | +-+-++++++ | 12.729 | 16 | 0.6925 |
| AD Braak | chr7 | 52,743,232 | chr7:52743232:C:T | t | c | -0.2106 | 0.0515 | 4.33E-05 | -+-+--------- | 12.406 | 16 | 0.7156 |
| AD Braak | chr7 | 52,743,331 | chr7:52743331:G:A | a | g | -0.2087 | 0.0514 | 4.98E-05 | -+-+--------- | 12.212 | 16 | 0.7292 |
| AD Braak | chr7 | 52,743,501 | chr7:52743501:G:A | a | g | -0.21 | 0.0515 | 4.54E-05 | -+-+--------- | 11.815 | 16 | 0.7566 |
| AD Braak | chr7 | 52,743,555 | chr7:52743555:G:A | a | g | -0.2119 | 0.0515 | 3.92E-05 | -+-+--------- | 12.013 | 16 | 0.7431 |
| AD Braak | chr7 | 52,747,681 | chr7:52747681:T:G | t | g | 0.2133 | 0.0515 | 3.47E-05 | +-+-++++++ | 12.125 | 16 | 0.7354 |
| AD Braak | chr7 | 52,749,053 | chr7:52749053:C:A | a | c | -0.214 | 0.0515 | 3.29E-05 | -+-+--------- | 12.298 | 16 | 0.7232 |
| AD Braak | chr7 | 52,754,441 | chr7:52754441:T:C | t | c | 0.2149 | 0.0514 | 2.94E-05 | +-+-++++++ | 12.87 | 16 | 0.6823 |
| AD Braak | chr7 | 52,756,342 | chr7:52756342:C:T | t | c | -0.2144 | 0.0514 | 3.05E-05 | -+-+--------- | 12.782 | 16 | 0.6886 |
| AD Braak | chr7 | 52,759,260 | chr7:52759260:G:T | t | g | -0.2145 | 0.0514 | 3.03E-05 | -+-+--------- | 12.954 | 16 | 0.6761 |
| AD Braak | chr7 | 52,760,645 | chr7:52760645:G:A | a | g | -0.2162 | 0.0514 | 2.65E-05 | -+-+--------- | 13.186 | 16 | 0.6591 |
| AD Braak | chr7 | 52,764,325 | chr7:52764325:T:G | t | g | 0.2127 | 0.0514 | 3.49E-05 | +-+-++++++ | 12.774 | 16 | 0.6892 |
| AD Braak | chr7 | 52,765,709 | chr7:52765709:G:A | a | g | -0.2147 | 0.0514 | 3.00E-05 | -+-+--------- | 12.951 | 16 | 0.6763 |
| AD Braak | chr7 | 52,770,515 | chr7:52770515:C:T | t | c | -0.2039 | 0.0507 | 5.75E-05 | -+-+--------+ | 12.872 | 16 | 0.6821 |
| AD Braak | chr7 | 52,773,527 | chr7:52773527:C:T | t | c | -0.2152 | 0.0517 | 3.14E-05 | -+-+--------- | 12.73 | 16 | 0.6924 |
| AD Braak | chr7 | 52,774,498 | chr7:52774498:C:T | t | c | -0.2161 | 0.0517 | 2.92E-05 | -+-+--------- | 12.323 | 16 | 0.7215 |

| | | | | | | | | | | | | |
|---|---|---|---|---|---|---|---|---|---|---|---|---|
| AD Braak | chr7 | 52,775,506 | chr7:52775506:C:T | t | c | -0.2617 | 0.0653 | 6.18E-05 | -+-+--------- | 13.285 | 16 | 0.6518 |
| AD Braak | chr7 | 52,778,583 | chr7:52778583:C:T | t | c | -0.2184 | 0.0517 | 2.37E-05 | -+-+--------- | 12.262 | 16 | 0.7258 |
| AD Braak | chr7 | 52,781,772 | chr7:52781772:A:G | a | g | 0.2674 | 0.0653 | 4.19E-05 | +-+-++++++ | 13.407 | 16 | 0.6428 |
| AD Braak | chr7 | 52,781,838 | chr7:52781838:C:A | a | c | -0.2674 | 0.0653 | 4.19E-05 | -+-+--------- | 13.407 | 16 | 0.6428 |
| AD Braak | chr7 | 52,785,147 | chr7:52785147:G:A | a | g | -0.2214 | 0.0516 | 1.79E-05 | -+-+--------- | 11.855 | 16 | 0.7539 |
| AD Braak | chr7 | 52,789,733 | chr7:52789733:C:T | t | c | -0.2732 | 0.0652 | 2.78E-05 | -+-+--------- | 13.222 | 16 | 0.6565 |
| AD Braak | chr7 | 52,791,061 | chr7:52791061:G:C | c | g | -0.2737 | 0.0652 | 2.72E-05 | -+-+--------- | 13.313 | 16 | 0.6497 |
| AD Braak | chr7 | 52,802,190 | chr7:52802190:G:A | a | g | -0.2189 | 0.0513 | 1.95E-05 | -+-+--------- | 12.888 | 16 | 0.6809 |
| AD Braak | chr7 | 52,802,873 | chr7:52802873:A:G | a | g | 0.2197 | 0.0513 | 1.84E-05 | +-+-++++++ | 12.951 | 16 | 0.6764 |
| AD Braak | chr7 | 52,805,457 | chr7:52805457:G:A | a | g | -0.2189 | 0.0513 | 1.99E-05 | -+-+--------- | 12.981 | 16 | 0.6741 |
| AD Braak | chr7 | 52,808,016 | chr7:52808016:T:C | t | c | 0.2622 | 0.0653 | 5.87E-05 | +-+-+++-++ | 16.187 | 16 | 0.44 |
| AD Braak | chr7 | 52,808,135 | chr7:52808135:G:A | a | g | -0.2622 | 0.0653 | 5.85E-05 | -+-+---+----- | 16.183 | 16 | 0.4402 |
| AD Braak | chr7 | 52,811,141 | chr7:52811141:C:T | t | c | -0.2632 | 0.0653 | 5.57E-05 | -+-+---+----- | 16.899 | 16 | 0.3921 |
| AD Braak | chr7 | 52,811,405 | chr7:52811405:T:A | a | t | -0.2632 | 0.0653 | 5.56E-05 | -+-+---+----- | 16.953 | 16 | 0.3887 |
| AD Braak | chr7 | 52,817,920 | chr7:52817920:CCAGA:C | ccaga | c | 0.2628 | 0.0653 | 5.73E-05 | +-+-+++-++ | 17.034 | 16 | 0.3834 |
| AD Braak | chr7 | 52,820,540 | chr7:52820540:G:A | a | g | -0.2627 | 0.0653 | 5.77E-05 | -+-+---+----- | 16.99 | 16 | 0.3862 |
| AD Braak | chr7 | 52,826,426 | chr7:52826426:G:T | t | g | -0.2624 | 0.0652 | 5.78E-05 | -+-+---+----- | 17.406 | 16 | 0.3598 |
| AD Braak | chr7 | 52,828,530 | chr7:52828530:G:T | t | g | -0.2644 | 0.0653 | 5.09E-05 | -+-+---+----- | 17.355 | 16 | 0.363 |
| AD Braak | chr7 | 52,832,579 | chr7:52832579:G:A | a | g | -0.2662 | 0.0654 | 4.66E-05 | -+-+---+----- | 17.453 | 16 | 0.3569 |
| AD Braak | chr7 | 52,833,066 | chr7:52833066:G:C | c | g | -0.2673 | 0.0655 | 4.45E-05 | -+-+---+----- | 17.561 | 16 | 0.3502 |
| AD Braak | chr7 | 52,836,465 | chr7:52836465:C:G | c | g | 0.2696 | 0.0654 | 3.76E-05 | +-+-+++-++ | 17.825 | 16 | 0.3342 |
| AD Braak | chr7 | 69,113,782 | chr7:69113782:TTATATC:T | t | ttatatc | 0.4934 | 0.1181 | 2.96E-05 | ++++++++-- | 14.922 | 12 | 0.2457 |
| AD Braak | chr7 | 75,662,953 | chr7:75662953:C:T | t | c | 0.4658 | 0.1026 | 5.61E-06 | ++++-+-+++ | 13.475 | 14 | 0.4895 |
| AD Braak | chr7 | 94,885,545 | chr7:94885545:T:C | t | c | -0.0962 | 0.0247 | 9.68E-05 | -+---------+-- | 15.728 | 16 | 0.4721 |
| AD Braak | chr8 | 5,769,614 | chr8:5769614:C:G | c | g | -0.1975 | 0.0485 | 4.69E-05 | -+-----+----+ | 27.352 | 16 | 0.03773 |
| AD Braak | chr8 | 5,783,136 | chr8:5783136:G:C | c | g | 0.1605 | 0.0397 | 5.20E-05 | +-++-++-++ | 24.02 | 16 | 0.08907 |
| AD Braak | chr8 | 5,837,977 | chr8:5837977:C:G | c | g | -0.2643 | 0.067 | 8.08E-05 | -+--------+-+ | 10.88 | 16 | 0.8168 |
| AD Braak | chr8 | 5,900,998 | chr8:5900998:G:A | a | g | 0.2682 | 0.0683 | 8.65E-05 | +++++++++ | 10.827 | 16 | 0.82 |
| AD Braak | chr8 | 22,420,411 | chr8:22420411:G:A | a | g | 0.779 | 0.1877 | 3.31E-05 | +??+??++? | 3.571 | 5 | 0.6127 |
| AD Braak | chr8 | 30,009,501 | chr8:30009501:C:G | c | g | 0.3255 | 0.0835 | 9.71E-05 | +++++++++ | 11.401 | 16 | 0.7841 |
| AD Braak | chr8 | 58,930,111 | chr8:58930111:A:C | a | c | 0.095 | 0.024 | 7.75E-05 | +++--++-++ | 20.525 | 16 | 0.1975 |
| AD Braak | chr8 | 78,328,082 | chr8:78328082:T:C | t | c | -0.4961 | 0.1265 | 8.75E-05 | ----+?-+-+?- | 6.952 | 13 | 0.9046 |
| AD Braak | chr8 | 84,813,327 | chr8:84813327:G:A | a | g | 0.4438 | 0.1106 | 6.01E-05 | ++++?++-+- | 10.299 | 13 | 0.6693 |
| AD Braak | chr8 | 103,126,664 | chr8:103126664:C:G | c | g | 0.0963 | 0.0243 | 7.69E-05 | ++++-++++- | 16.049 | 16 | 0.4496 |
| AD Braak | chr8 | 132,360,801 | chr8:132360801:A:G | a | g | -0.336 | 0.0775 | 1.44E-05 | +---------?+- | 15.793 | 15 | 0.396 |
| AD Braak | chr9 | 2,223,898 | chr9:2223898:A:G | a | g | -0.3464 | 0.087 | 6.85E-05 | ----+--+-?+- | 10.905 | 15 | 0.7593 |
| AD Braak | chr9 | 36,982,900 | chr9:36982900:G:GT | g | gt | -0.1403 | 0.0321 | 1.24E-05 | ----+-----+-- | 8.839 | 16 | 0.9199 |
| AD Braak | chr9 | 37,138,558 | chr9:37138558:A:G | a | g | -0.2262 | 0.0543 | 3.06E-05 | ----+------+- | 19.991 | 16 | 0.2206 |
| AD Braak | chr9 | 37,185,317 | chr9:37185317:TC:T | t | tc | 0.2618 | 0.0531 | 8.01E-07 | ++++-++++- | 23.469 | 16 | 0.1018 |
| AD Braak | chr9 | 37,236,286 | chr9:37236286:A:T | a | t | -0.2378 | 0.0536 | 9.21E-06 | ----+------+- | 21.768 | 16 | 0.1508 |
| AD Braak | chr9 | 37,302,158 | chr9:37302158:C:T | t | c | -0.0985 | 0.0245 | 5.66E-05 | ----+-++-++- | 16.036 | 16 | 0.4505 |
| AD Braak | chr9 | 37,302,649 | chr9:37302649:T:C | t | c | 0.0965 | 0.0244 | 7.80E-05 | ++++-+--++ | 18.51 | 16 | 0.2949 |
| AD Braak | chr9 | 37,343,410 | chr9:37343410:A:G | a | g | -0.2555 | 0.0527 | 1.23E-06 | ----+------+- | 23.454 | 16 | 0.1021 |
| AD Braak | chr9 | 73,183,571 | chr9:73183571:T:G | t | g | -0.4136 | 0.0967 | 1.90E-05 | ---+--+--+?+ | 35.797 | 15 | 0.001891 |
| AD Braak | chr9 | 73,191,010 | chr9:73191010:C:G | c | g | -0.4145 | 0.0967 | 1.80E-05 | ---+--+--+?+ | 35.691 | 15 | 0.001959 |
| AD Braak | chr9 | 73,269,613 | chr9:73269613:C:A | a | c | 0.379 | 0.0962 | 8.12E-05 | +++-++--+-? | 35.005 | 15 | 0.002455 |

| | | | | | | | | | | | | |
|---|---|---|---|---|---|---|---|---|---|---|---|---|
| AD Braak | chr9 | 73,390,917 | chr9:73390917:T:G | t | g | -0.3845 | 0.0967 | 7.03E-05 | ---+--++-+?- | 34.334 | 15 | 0.003056 |
| AD Braak | chr9 | 87,572,838 | chr9:87572838:G:T | t | g | 0.2508 | 0.0641 | 9.18E-05 | +++++++--+ | 17.728 | 16 | 0.3401 |
| AD Braak | chr9 | 90,608,147 | chr9:90608147:G:C | c | g | -0.1296 | 0.0324 | 6.19E-05 | -------+-++-- | 17.125 | 16 | 0.3776 |
| AD Braak | chr9 | 90,619,660 | chr9:90619660:A:G | a | g | 0.1281 | 0.0324 | 7.68E-05 | +++++++-+- | 17.386 | 16 | 0.361 |
| AD Braak | chr9 | 100,780,057 | chr9:100780057:C:T | t | c | 0.1365 | 0.0349 | 9.35E-05 | +++++++++ | 13.608 | 16 | 0.6279 |
| AD Braak | chr9 | 100,780,915 | chr9:100780915:T:C | t | c | -0.1415 | 0.035 | 5.39E-05 | ----------+--- | 13.091 | 16 | 0.6661 |
| AD Braak | chr9 | 100,780,990 | chr9:100780990:A:G | a | g | -0.1366 | 0.0347 | 8.07E-05 | ----------+--- | 14.536 | 16 | 0.5589 |
| AD Braak | chr9 | 100,781,410 | chr9:100781410:C:T | t | c | 0.1362 | 0.035 | 9.95E-05 | +++++++++ | 12.563 | 16 | 0.7044 |
| AD Braak | chr9 | 100,783,477 | chr9:100783477:C:A | a | c | 0.1366 | 0.0331 | 3.69E-05 | +++++++++ | 11.589 | 16 | 0.7717 |
| AD Braak | chr9 | 134,718,936 | chr9:134718936:G:A | a | g | -0.3748 | 0.0927 | 5.29E-05 | -----+----+-- | 13.571 | 16 | 0.6306 |
| ADNC Score | chr1 | 1,593,620 | chr1:1593620:G:C | c | g | -0.2702 | 0.0673 | 6.04E-05 | -?--+-?--- | 14.256 | 7 | 0.04682 |
| ADNC Score | chr1 | 1,594,077 | chr1:1594077:T:C | t | c | 0.2714 | 0.0673 | 5.59E-05 | +?++-+?++- | 14.002 | 7 | 0.05115 |
| ADNC Score | chr1 | 1,594,131 | chr1:1594131:T:C | t | c | 0.2611 | 0.0661 | 7.79E-05 | +?++-+?-++ | 14.681 | 7 | 0.04031 |
| ADNC Score | chr1 | 1,594,817 | chr1:1594817:A:C | a | c | 0.2741 | 0.0675 | 4.88E-05 | +?++-+?++- | 14.278 | 7 | 0.04646 |
| ADNC Score | chr1 | 1,594,820 | chr1:1594820:C:T | t | c | -0.2757 | 0.0674 | 4.26E-05 | -?--+-?--- | 14.451 | 7 | 0.04372 |
| ADNC Score | chr1 | 1,596,449 | chr1:1596449:G:A | a | g | -0.2776 | 0.0674 | 3.79E-05 | -?--++?--- | 14.91 | 7 | 0.03717 |
| ADNC Score | chr1 | 1,596,529 | chr1:1596529:A:G | a | g | 0.2724 | 0.0673 | 5.15E-05 | +?++-+?++- | 13.978 | 7 | 0.05157 |
| ADNC Score | chr1 | 1,599,234 | chr1:1599234:T:C | t | c | 0.2833 | 0.0663 | 1.90E-05 | +?++-+?++- | 14.936 | 7 | 0.03683 |
| ADNC Score | chr1 | 1,601,796 | chr1:1601796:A:C | a | c | -0.2439 | 0.06 | 4.80E-05 | -?--+-?--- | 14.47 | 7 | 0.04343 |
| ADNC Score | chr1 | 1,602,057 | chr1:1602057:C:T | t | c | -0.2803 | 0.066 | 2.16E-05 | -?--+-?--- | 14.651 | 7 | 0.04074 |
| ADNC Score | chr1 | 1,603,989 | chr1:1603989:T:C | t | c | -0.2406 | 0.0612 | 8.46E-05 | ??--+-?--- | 11.781 | 6 | 0.06704 |
| ADNC Score | chr1 | 1,604,202 | chr1:1604202:G:A | a | g | 0.2458 | 0.06 | 4.19E-05 | +?++-+?++- | 14.396 | 7 | 0.04458 |
| ADNC Score | chr1 | 1,608,244 | chr1:1608244:T:C | t | c | -0.2375 | 0.061 | 9.99E-05 | ??--+-?--- | 11.365 | 6 | 0.07772 |
| ADNC Score | chr1 | 1,609,909 | chr1:1609909:G:T | t | g | 0.2372 | 0.0609 | 9.90E-05 | ??++-+?+++ | 11.517 | 6 | 0.07366 |
| ADNC Score | chr1 | 1,611,248 | chr1:1611248:ATC:A | a | atc | -0.2766 | 0.0656 | 2.50E-05 | -?--+-?--- | 13.737 | 7 | 0.05607 |
| ADNC Score | chr1 | 1,613,322 | chr1:1613322:T:C | t | c | -0.2336 | 0.06 | 9.97E-05 | ??--+-?--- | 12.189 | 6 | 0.05788 |
| ADNC Score | chr1 | 1,613,421 | chr1:1613421:T:C | t | c | -0.2345 | 0.06 | 9.41E-05 | ??--+-?--- | 12.246 | 6 | 0.05669 |
| ADNC Score | chr1 | 1,616,547 | chr1:1616547:T:C | t | c | -0.2339 | 0.0598 | 9.26E-05 | ??--+-?--- | 12.053 | 6 | 0.0608 |
| ADNC Score | chr1 | 1,627,057 | chr1:1627057:C:G | c | g | 0.2586 | 0.0641 | 5.43E-05 | +?++--?-++ | 13.554 | 7 | 0.05971 |
| ADNC Score | chr1 | 1,653,319 | chr1:1653319:A:G | a | g | -0.259 | 0.066 | 8.74E-05 | ??--+-?+-- | 9.642 | 6 | 0.1406 |
| ADNC Score | chr1 | 1,661,169 | chr1:1661169:C:A | a | c | 0.2634 | 0.0669 | 8.18E-05 | +?++-+?-++ | 8.212 | 7 | 0.3142 |
| ADNC Score | chr1 | 1,671,724 | chr1:1671724:C:T | t | c | -0.2627 | 0.0654 | 5.85E-05 | ----++-+-- | 17.223 | 9 | 0.04533 |
| ADNC Score | chr1 | 1,672,155 | chr1:1672155:G:A | a | g | -0.294 | 0.0663 | 9.19E-06 | ----+--+-- | 13.518 | 9 | 0.1405 |
| ADNC Score | chr1 | 1,675,132 | chr1:1675132:A:G | a | g | -0.2647 | 0.0638 | 3.30E-05 | ----+----- | 10.125 | 9 | 0.3405 |
| ADNC Score | chr1 | 21,744,547 | chr1:21744547:G:A | a | g | -0.3267 | 0.0835 | 9.19E-05 | +--------? | 3.753 | 8 | 0.8787 |
| ADNC Score | chr1 | 21,745,130 | chr1:21745130:T:C | t | c | 0.3263 | 0.0836 | 9.51E-05 | -++++++++ | 3.725 | 8 | 0.8811 |
| ADNC Score | chr1 | 21,754,978 | chr1:21754978:G:A | a | g | -0.3278 | 0.0837 | 9.07E-05 | +--------? | 3.711 | 8 | 0.8822 |
| ADNC Score | chr1 | 21,759,137 | chr1:21759137:G:C | c | g | -0.3268 | 0.0838 | 9.65E-05 | +--------? | 3.68 | 8 | 0.8848 |
| ADNC Score | chr1 | 21,760,920 | chr1:21760920:C:T | t | c | -0.3266 | 0.0838 | 9.71E-05 | +--------? | 3.678 | 8 | 0.8849 |
| ADNC Score | chr1 | 21,762,668 | chr1:21762668:C:T | t | c | -0.3267 | 0.0838 | 9.71E-05 | +--------? | 3.679 | 8 | 0.8849 |
| ADNC Score | chr1 | 21,775,860 | chr1:21775860:T:C | t | c | 0.3278 | 0.0839 | 9.34E-05 | -++++++++ | 3.854 | 8 | 0.87 |
| ADNC Score | chr1 | 21,777,532 | chr1:21777532:G:C | c | g | -0.3297 | 0.084 | 8.74E-05 | +--------? | 3.678 | 8 | 0.8849 |
| ADNC Score | chr1 | 21,781,345 | chr1:21781345:A:G | a | g | 0.3295 | 0.0841 | 8.86E-05 | -++++++++ | 3.682 | 8 | 0.8846 |
| ADNC Score | chr1 | 21,785,485 | chr1:21785485:T:C | t | c | 0.3393 | 0.0833 | 4.65E-05 | -++++++++- | 3.921 | 9 | 0.9165 |
| ADNC Score | chr1 | 63,934,842 | chr1:63934842:C:T | t | c | 1.0641 | 0.2554 | 3.10E-05 | ++++??++? | 7.594 | 6 | 0.2694 |
| ADNC Score | chr1 | 79,271,598 | chr1:79271598:T:C | t | c | -0.2037 | 0.0489 | 3.13E-05 | ----+----+ | 7.325 | 9 | 0.6034 |

| | | | | | | | | | | | | |
|---|---|---|---|---|---|---|---|---|---|---|---|---|
| ADNC Score | chr1 | 79,275,797 | chr1:79275797:G:C | c | g | 0.1941 | 0.0492 | 8.07E-05 | ++++-++++- | 7.586 | 9 | 0.5763 |
| ADNC Score | chr1 | 79,276,059 | chr1:79276059:A:G | a | g | -0.2015 | 0.0487 | 3.52E-05 | ----+----+ | 7.306 | 9 | 0.6052 |
| ADNC Score | chr1 | 79,284,707 | chr1:79284707:A:G | a | g | -0.198 | 0.049 | 5.28E-05 | ----+----+ | 7.394 | 9 | 0.5961 |
| ADNC Score | chr1 | 93,003,712 | chr1:93003712:TAATA:T | t | taata | 0.1869 | 0.0474 | 7.95E-05 | +++++++++ | 5.814 | 9 | 0.7584 |
| ADNC Score | chr1 | 93,003,753 | chr1:93003753:G:A | a | g | 0.187 | 0.0473 | 7.72E-05 | +++++++++ | 5.682 | 9 | 0.7712 |
| ADNC Score | chr1 | 104,534,131 | chr1:104534131:T:A | a | t | 0.2331 | 0.0598 | 9.72E-05 | ++++-++++- | 10.544 | 9 | 0.3083 |
| ADNC Score | chr1 | 104,534,188 | chr1:104534188:C:T | t | c | 0.2326 | 0.0597 | 9.89E-05 | ++++-++++- | 11.596 | 9 | 0.237 |
| ADNC Score | chr1 | 111,324,087 | chr1:111324087:G:A | a | g | 0.6436 | 0.1603 | 5.96E-05 | ++++++-++- | 13.199 | 9 | 0.1538 |
| ADNC Score | chr1 | 111,609,639 | chr1:111609639:T:G | t | g | -0.4533 | 0.1145 | 7.50E-05 | ++-------+ | 4.012 | 9 | 0.9106 |
| ADNC Score | chr1 | 116,951,448 | chr1:116951448:C:T | t | c | 1.1259 | 0.281 | 6.17E-05 | ??++??+?? | 1.514 | 2 | 0.4691 |
| ADNC Score | chr1 | 117,000,501 | chr1:117000501:G:A | a | g | 1.0937 | 0.2789 | 8.79E-05 | +?++??+?? | 2.158 | 3 | 0.5403 |
| ADNC Score | chr1 | 177,880,828 | chr1:177880828:C:T | t | c | 0.2019 | 0.0505 | 6.29E-05 | +-+++++-++ | 10.013 | 9 | 0.3494 |
| ADNC Score | chr1 | 177,883,654 | chr1:177883654:C:A | a | c | 0.1996 | 0.0503 | 7.29E-05 | +-+++-+-++ | 9.759 | 9 | 0.3703 |
| ADNC Score | chr1 | 177,883,908 | chr1:177883908:G:T | t | g | 0.2001 | 0.0503 | 6.98E-05 | +-+++++-++ | 9.684 | 9 | 0.3767 |
| ADNC Score | chr1 | 177,884,085 | chr1:177884085:C:G | c | g | -0.2005 | 0.0503 | 6.63E-05 | -+-----+-- | 9.549 | 9 | 0.3882 |
| ADNC Score | chr1 | 177,885,264 | chr1:177885264:A:G | a | g | -0.2018 | 0.0503 | 6.01E-05 | -+---+-+-- | 9.886 | 9 | 0.3598 |
| ADNC Score | chr1 | 177,887,790 | chr1:177887790:C:T | t | c | 0.1992 | 0.0512 | 9.92E-05 | +-+++-+-++ | 11.737 | 9 | 0.2286 |
| ADNC Score | chr1 | 177,888,167 | chr1:177888167:T:C | t | c | -0.2027 | 0.0503 | 5.59E-05 | -+---+-+-- | 9.72 | 9 | 0.3736 |
| ADNC Score | chr1 | 177,890,507 | chr1:177890507:G:C | c | g | 0.2025 | 0.0503 | 5.63E-05 | +-+++-+-++ | 9.705 | 9 | 0.3749 |
| ADNC Score | chr1 | 177,891,196 | chr1:177891196:G:A | a | g | 0.2033 | 0.0503 | 5.29E-05 | +-+++-+-++ | 9.91 | 9 | 0.3578 |
| ADNC Score | chr1 | 177,897,657 | chr1:177897657:G:A | a | g | 0.2029 | 0.0503 | 5.46E-05 | +-+++-+-++ | 9.768 | 9 | 0.3696 |
| ADNC Score | chr1 | 177,898,473 | chr1:177898473:G:A | a | g | 0.2019 | 0.0502 | 5.73E-05 | +-+++-+-++ | 9.576 | 9 | 0.3859 |
| ADNC Score | chr1 | 177,900,615 | chr1:177900615:T:TGCATTC | t | tgcattc | -0.2038 | 0.0503 | 5.14E-05 | -+---+-+-- | 10.026 | 9 | 0.3484 |
| ADNC Score | chr1 | 177,901,122 | chr1:177901122:T:A | a | t | 0.2045 | 0.0503 | 4.73E-05 | +-+++-+-++ | 10.006 | 9 | 0.35 |
| ADNC Score | chr1 | 177,904,198 | chr1:177904198:A:G | a | g | -0.2027 | 0.0503 | 5.57E-05 | -+---+-+-- | 9.878 | 9 | 0.3605 |
| ADNC Score | chr1 | 177,917,982 | chr1:177917982:C:G | c | g | -0.1993 | 0.0502 | 7.23E-05 | -+---+-+-- | 9.685 | 9 | 0.3766 |
| ADNC Score | chr1 | 183,911,830 | chr1:183911830:C:T | t | c | 0.3228 | 0.0813 | 7.25E-05 | +++++++++ | 10.238 | 9 | 0.3315 |
| ADNC Score | chr1 | 183,911,955 | chr1:183911955:A:T | a | t | -0.362 | 0.0922 | 8.64E-05 | +--------- | 6.35 | 9 | 0.7045 |
| ADNC Score | chr1 | 196,081,932 | chr1:196081932:C:T | t | c | 1.0452 | 0.2606 | 6.05E-05 | +++++-+??? | 15.777 | 6 | 0.015 |
| ADNC Score | chr1 | 196,113,972 | chr1:196113972:T:C | t | c | -0.4596 | 0.1124 | 4.33E-05 | +----+-+-- | 12.061 | 9 | 0.2099 |
| ADNC Score | chr1 | 207,712,536 | chr1:207712536:C:T | t | c | 0.1934 | 0.0489 | 7.71E-05 | ++++-++++- | 10.275 | 9 | 0.3286 |
| ADNC Score | chr1 | 207,728,216 | chr1:207728216:G:A | a | g | 0.1899 | 0.0486 | 9.23E-05 | ++++-++++- | 10.628 | 9 | 0.302 |
| ADNC Score | chr1 | 207,731,491 | chr1:207731491:C:T | t | c | 0.1966 | 0.0486 | 5.28E-05 | ++++-++++- | 10.924 | 9 | 0.2809 |
| ADNC Score | chr1 | 207,731,664 | chr1:207731664:G:A | a | g | 0.197 | 0.0486 | 5.10E-05 | ++++-++++- | 10.879 | 9 | 0.2841 |
| ADNC Score | chr1 | 207,737,570 | chr1:207737570:G:C | c | g | 0.1969 | 0.0488 | 5.43E-05 | ++++-++++- | 10.136 | 9 | 0.3396 |
| ADNC Score | chr1 | 213,294,117 | chr1:213294117:G:C | c | g | -0.2008 | 0.0515 | 9.55E-05 | -+--+----- | 9.435 | 9 | 0.3981 |
| ADNC Score | chr1 | 222,533,805 | chr1:222533805:C:T | t | c | 0.6453 | 0.1522 | 2.23E-05 | +++++++++ | 4.975 | 9 | 0.8365 |
| ADNC Score | chr10 | 2,751,846 | chr10:2751846:G:T | t | g | 0.3527 | 0.0856 | 3.81E-05 | -++++++++- | 8.465 | 9 | 0.4881 |
| ADNC Score | chr10 | 2,762,193 | chr10:2762193:T:TTC | t | ttc | 0.3494 | 0.0875 | 6.46E-05 | -++++++++- | 10.615 | 9 | 0.303 |
| ADNC Score | chr10 | 2,762,297 | chr10:2762297:T:C | t | c | 0.3594 | 0.0874 | 3.96E-05 | -++++++++- | 10.365 | 9 | 0.3218 |
| ADNC Score | chr10 | 2,762,908 | chr10:2762908:T:C | t | c | -0.3704 | 0.0878 | 2.45E-05 | +--------+ | 10.735 | 9 | 0.2943 |
| ADNC Score | chr10 | 2,763,047 | chr10:2763047:T:G | t | g | 0.3608 | 0.0875 | 3.75E-05 | -++++++++- | 10.287 | 9 | 0.3278 |
| ADNC Score | chr10 | 2,773,632 | chr10:2773632:C:T | t | c | -0.3572 | 0.0848 | 2.51E-05 | +--------+ | 10.926 | 9 | 0.2808 |
| ADNC Score | chr10 | 5,536,252 | chr10:5536252:A:G | a | g | -0.6473 | 0.1609 | 5.74E-05 | -+----++-+ | 6.442 | 9 | 0.695 |
| ADNC Score | chr10 | 10,367,477 | chr10:10367477:C:T | t | c | 0.6685 | 0.1659 | 5.59E-05 | -++++++-++ | 5.886 | 9 | 0.7513 |
| ADNC Score | chr10 | 14,949,431 | chr10:14949431:A:C | a | c | -0.4195 | 0.1001 | 2.79E-05 | ---------+ | 4.14 | 9 | 0.902 |

| | | | | | | | | | | | | |
|---|---|---|---|---|---|---|---|---|---|---|---|---|
| ADNC Score | chr10 | 31,627,761 | chr10:31627761:T:C | t | c | -0.1983 | 0.0502 | 7.87E-05 | ++-------+ | 10.642 | 9 | 0.3011 |
| ADNC Score | chr10 | 33,319,678 | chr10:33319678:G:A | a | g | 0.3349 | 0.0858 | 9.50E-05 | -++++++-++ | 11.104 | 9 | 0.2687 |
| ADNC Score | chr10 | 52,203,746 | chr10:52203746:A:C | a | c | -0.6106 | 0.1534 | 6.87E-05 | ------++-+ | 6.718 | 9 | 0.6665 |
| ADNC Score | chr10 | 69,833,384 | chr10:69833384:C:T | t | c | -0.1972 | 0.0503 | 8.98E-05 | --------- | 5.34 | 9 | 0.8038 |
| ADNC Score | chr10 | 82,325,022 | chr10:82325022:G:A | a | g | 0.6533 | 0.1566 | 3.02E-05 | ++++?+--+? | 9.597 | 7 | 0.2126 |
| ADNC Score | chr10 | 82,343,370 | chr10:82343370:A:T | a | t | -0.6389 | 0.1573 | 4.89E-05 | ----?-++-? | 9.079 | 7 | 0.2471 |
| ADNC Score | chr10 | 82,347,092 | chr10:82347092:T:G | t | g | -0.6539 | 0.158 | 3.49E-05 | ----?-+--? | 8.788 | 7 | 0.2683 |
| ADNC Score | chr10 | 82,389,489 | chr10:82389489:C:T | t | c | 0.652 | 0.1662 | 8.75E-05 | ++++?+--+? | 10.759 | 7 | 0.1495 |
| ADNC Score | chr10 | 90,258,710 | chr10:90258710:T:C | t | c | -0.9108 | 0.2288 | 6.89E-05 | ?---?--?-? | 10.91 | 5 | 0.05318 |
| ADNC Score | chr10 | 97,909,571 | chr10:97909571:G:A | a | g | 0.431 | 0.108 | 6.61E-05 | +++++-+++- | 11.074 | 9 | 0.2706 |
| ADNC Score | chr10 | 97,913,426 | chr10:97913426:T:C | t | c | -0.4221 | 0.1074 | 8.46E-05 | -----+---? | 10.662 | 8 | 0.2216 |
| ADNC Score | chr10 | 103,490,365 | chr10:103490365:C:T | t | c | 0.6695 | 0.1708 | 8.82E-05 | ?++++++++ | 9.02 | 8 | 0.3406 |
| ADNC Score | chr10 | 103,572,177 | chr10:103572177:A:C | a | c | -0.4747 | 0.1135 | 2.91E-05 | ------+--- | 7.089 | 9 | 0.6279 |
| ADNC Score | chr10 | 114,809,785 | chr10:114809785:A:G | a | g | -0.7144 | 0.1824 | 8.97E-05 | -+----+--- | 6.791 | 9 | 0.6588 |
| ADNC Score | chr10 | 114,813,744 | chr10:114813744:C:T | t | c | 0.7222 | 0.1827 | 7.70E-05 | +-++++-+++ | 6.382 | 9 | 0.7012 |
| ADNC Score | chr10 | 114,814,636 | chr10:114814636:C:A | a | c | 0.722 | 0.1827 | 7.75E-05 | +-++++-+++ | 6.384 | 9 | 0.701 |
| ADNC Score | chr10 | 114,819,642 | chr10:114819642:A:G | a | g | -0.7231 | 0.1826 | 7.51E-05 | -+----+--- | 6.411 | 9 | 0.6982 |
| ADNC Score | chr10 | 114,822,730 | chr10:114822730:G:A | a | g | 0.7249 | 0.1822 | 6.94E-05 | +?++++-++- | 5.473 | 8 | 0.706 |
| ADNC Score | chr10 | 114,828,432 | chr10:114828432:A:G | a | g | -0.723 | 0.1786 | 5.18E-05 | -?----+--- | 4.989 | 8 | 0.7588 |
| ADNC Score | chr10 | 114,828,978 | chr10:114828978:A:G | a | g | -0.7385 | 0.1788 | 3.63E-05 | -?----+--- | 5.197 | 8 | 0.7364 |
| ADNC Score | chr10 | 114,838,454 | chr10:114838454:A:C | a | c | -0.7241 | 0.1786 | 5.01E-05 | -?----+--- | 5.003 | 8 | 0.7572 |
| ADNC Score | chr10 | 114,839,637 | chr10:114839637:G:C | c | g | 0.7364 | 0.1791 | 3.92E-05 | +?++++-++- | 5.243 | 8 | 0.7314 |
| ADNC Score | chr10 | 114,844,541 | chr10:114844541:A:G | a | g | -0.7363 | 0.1791 | 3.95E-05 | -?----+--- | 5.252 | 8 | 0.7303 |
| ADNC Score | chr10 | 114,846,911 | chr10:114846911:T:C | t | c | -0.7364 | 0.1791 | 3.94E-05 | -?----+--- | 5.255 | 8 | 0.7301 |
| ADNC Score | chr10 | 114,848,049 | chr10:114848049:T:C | t | c | -0.7265 | 0.1819 | 6.48E-05 | -?----+--- | 5.469 | 8 | 0.7065 |
| ADNC Score | chr10 | 114,849,677 | chr10:114849677:T:C | t | c | -0.7265 | 0.1819 | 6.48E-05 | -?----+--- | 5.469 | 8 | 0.7065 |
| ADNC Score | chr10 | 114,851,018 | chr10:114851018:A:G | a | g | -0.7256 | 0.1744 | 3.17E-05 | -?----+--- | 4.932 | 8 | 0.7649 |
| ADNC Score | chr10 | 114,853,023 | chr10:114853023:A:C | a | c | -0.7283 | 0.1818 | 6.18E-05 | -?----?--- | 5.241 | 7 | 0.6306 |
| ADNC Score | chr10 | 114,855,060 | chr10:114855060:C:T | t | c | 0.726 | 0.1785 | 4.78E-05 | +?++++?++ | 4.777 | 7 | 0.6872 |
| ADNC Score | chr10 | 114,859,281 | chr10:114859281:CTG:C | ctg | c | -0.7276 | 0.1818 | 6.30E-05 | -?----?--- | 5.256 | 7 | 0.6287 |
| ADNC Score | chr10 | 114,860,330 | chr10:114860330:G:A | a | g | 0.726 | 0.1786 | 4.78E-05 | +?++++?++ | 4.776 | 7 | 0.6873 |
| ADNC Score | chr10 | 114,861,209 | chr10:114861209:G:A | a | g | 0.7469 | 0.1826 | 4.29E-05 | +?++++?++ | 5.173 | 7 | 0.6388 |
| ADNC Score | chr10 | 114,867,057 | chr10:114867057:A:C | a | c | -0.7424 | 0.1793 | 3.45E-05 | -?----?--- | 4.976 | 7 | 0.6629 |
| ADNC Score | chr11 | 2,263,356 | chr11:2263356:G:A | a | g | 0.8992 | 0.2292 | 8.71E-05 | +?++-+?-+? | 2.842 | 6 | 0.8284 |
| ADNC Score | chr11 | 7,617,805 | chr11:7617805:C:G | c | g | -0.2278 | 0.0504 | 6.26E-06 | -+--+----- | 1.492 | 9 | 0.9972 |
| ADNC Score | chr11 | 7,688,219 | chr11:7688219:T:C | t | c | -0.2753 | 0.0661 | 3.11E-05 | +-------- | 6.001 | 9 | 0.7399 |
| ADNC Score | chr11 | 16,280,877 | chr11:16280877:A:G | a | g | 0.599 | 0.1514 | 7.58E-05 | +-+++++++ | 8.596 | 8 | 0.3775 |
| ADNC Score | chr11 | 57,028,853 | chr11:57028853:G:A | a | g | 0.2367 | 0.0575 | 3.82E-05 | +++++++++ | 4.907 | 9 | 0.8424 |
| ADNC Score | chr11 | 57,031,768 | chr11:57031768:G:A | a | g | 0.2399 | 0.0574 | 2.96E-05 | +++++++++ | 5.261 | 9 | 0.811 |
| ADNC Score | chr11 | 57,031,938 | chr11:57031938:C:A | a | c | 0.2173 | 0.0542 | 6.07E-05 | +++++++++ | 5.809 | 9 | 0.7589 |
| ADNC Score | chr11 | 57,032,438 | chr11:57032438:A:C | a | c | -0.2365 | 0.0575 | 3.85E-05 | --------- | 5.211 | 9 | 0.8155 |
| ADNC Score | chr11 | 57,037,282 | chr11:57037282:C:T | t | c | -0.215 | 0.0542 | 7.34E-05 | --------- | 5.92 | 9 | 0.7479 |
| ADNC Score | chr11 | 57,039,042 | chr11:57039042:G:A | a | g | -0.2144 | 0.0542 | 7.65E-05 | --------- | 5.829 | 9 | 0.7569 |
| ADNC Score | chr11 | 57,045,655 | chr11:57045655:G:C | c | g | 0.2331 | 0.0575 | 5.05E-05 | +++++++++ | 5.307 | 9 | 0.8067 |
| ADNC Score | chr11 | 57,049,822 | chr11:57049822:G:A | a | g | -0.2185 | 0.0543 | 5.70E-05 | --------- | 5.956 | 9 | 0.7443 |
| ADNC Score | chr11 | 57,051,191 | chr11:57051191:CAAGGG:C | caaggg | c | -0.2377 | 0.0576 | 3.62E-05 | --------- | 5.172 | 9 | 0.819 |

| | | | | | | | | | | | | |
|---|---|---|---|---|---|---|---|---|---|---|---|---|
| ADNC Score | chr11 | 57,052,523 | chr11:57052523:T:C | t | c | 0.2209 | 0.0543 | 4.71E-05 | +++++++++ | 6.229 | 9 | 0.7168 |
| ADNC Score | chr11 | 57,053,571 | chr11:57053571:T:C | t | c | 0.2202 | 0.0543 | 4.99E-05 | +++++++++ | 6.26 | 9 | 0.7136 |
| ADNC Score | chr11 | 57,055,956 | chr11:57055956:A:G | a | g | 0.2185 | 0.0543 | 5.73E-05 | +++++++++ | 6.007 | 9 | 0.7393 |
| ADNC Score | chr11 | 57,059,515 | chr11:57059515:C:CG | cg | c | -0.2167 | 0.0543 | 6.62E-05 | --------- | 6.059 | 9 | 0.734 |
| ADNC Score | chr11 | 57,063,651 | chr11:57063651:T:A | a | t | -0.2154 | 0.0543 | 7.19E-05 | --------- | 5.979 | 9 | 0.742 |
| ADNC Score | chr11 | 57,066,125 | chr11:57066125:G:C | c | g | -0.2154 | 0.0543 | 7.18E-05 | --------- | 5.961 | 9 | 0.7439 |
| ADNC Score | chr11 | 57,068,005 | chr11:57068005:G:A | a | g | -0.1966 | 0.0502 | 8.93E-05 | -+------- | 5.176 | 9 | 0.8187 |
| ADNC Score | chr11 | 57,073,914 | chr11:57073914:G:A | a | g | -0.2151 | 0.0542 | 7.28E-05 | --------- | 6.555 | 9 | 0.6834 |
| ADNC Score | chr11 | 57,074,144 | chr11:57074144:C:G | c | g | 0.2153 | 0.0543 | 7.36E-05 | +++++++++ | 6.714 | 9 | 0.6669 |
| ADNC Score | chr11 | 57,076,180 | chr11:57076180:C:T | t | c | -0.1984 | 0.0503 | 7.94E-05 | -+--+---- | 5.617 | 9 | 0.7776 |
| ADNC Score | chr11 | 57,079,923 | chr11:57079923:C:T | t | c | 0.231 | 0.0575 | 5.95E-05 | +++++++++ | 5.623 | 9 | 0.7769 |
| ADNC Score | chr11 | 57,084,623 | chr11:57084623:A:G | a | g | 0.217 | 0.0543 | 6.45E-05 | +++++++++ | 6.419 | 9 | 0.6974 |
| ADNC Score | chr11 | 57,094,878 | chr11:57094878:C:G | c | g | -0.2399 | 0.058 | 3.53E-05 | --------- | 5.873 | 9 | 0.7525 |
| ADNC Score | chr11 | 57,101,356 | chr11:57101356:T:C | t | c | -0.2334 | 0.0576 | 5.04E-05 | --------- | 5.56 | 9 | 0.783 |
| ADNC Score | chr11 | 57,102,828 | chr11:57102828:C:T | t | c | -0.2235 | 0.0544 | 3.94E-05 | --------- | 6.88 | 9 | 0.6497 |
| ADNC Score | chr11 | 81,986,583 | chr11:81986583:G:A | a | g | 0.6993 | 0.1793 | 9.66E-05 | ++++-+-++? | 9.558 | 8 | 0.2974 |
| ADNC Score | chr11 | 86,112,282 | chr11:86112282:C:T | t | c | -0.191 | 0.0476 | 6.03E-05 | --------+ | 9.324 | 9 | 0.408 |
| ADNC Score | chr11 | 86,130,466 | chr11:86130466:TGTGAAAA:T | t | tgtgaaaa | -0.1866 | 0.0476 | 8.93E-05 | ++------+ | 8.127 | 9 | 0.5214 |
| ADNC Score | chr11 | 86,142,209 | chr11:86142209:T:G | t | g | 0.1941 | 0.0473 | 4.06E-05 | +++++++++ | 8.559 | 9 | 0.479 |
| ADNC Score | chr11 | 86,143,134 | chr11:86143134:G:T | t | g | -0.1907 | 0.0471 | 5.15E-05 | ------+-+ | 8.61 | 9 | 0.474 |
| ADNC Score | chr11 | 86,147,845 | chr11:86147845:G:A | a | g | -0.1994 | 0.0478 | 3.04E-05 | ++------+ | 6.06 | 9 | 0.7339 |
| ADNC Score | chr11 | 106,429,272 | chr11:106429272:T:C | t | c | -0.2422 | 0.0621 | 9.72E-05 | -+----+-- | 6.267 | 9 | 0.713 |
| ADNC Score | chr11 | 107,752,294 | chr11:107752294:G:A | a | g | 0.2097 | 0.0485 | 1.54E-05 | ++++-+-+++ | 7.33 | 9 | 0.6028 |
| ADNC Score | chr11 | 107,792,064 | chr11:107792064:A:G | a | g | -0.1942 | 0.0473 | 4.05E-05 | -----+--- | 8.913 | 9 | 0.4453 |
| ADNC Score | chr11 | 107,799,748 | chr11:107799748:C:T | t | c | 0.1963 | 0.048 | 4.28E-05 | ++++++-++- | 7.23 | 9 | 0.6132 |
| ADNC Score | chr11 | 107,799,754 | chr11:107799754:A:G | a | g | -0.194 | 0.0477 | 4.73E-05 | -----+--- | 7.073 | 9 | 0.6295 |
| ADNC Score | chr11 | 107,800,447 | chr11:107800447:C:T | t | c | 0.1966 | 0.048 | 4.17E-05 | ++++++-++- | 7.23 | 9 | 0.6132 |
| ADNC Score | chr11 | 107,800,900 | chr11:107800900:C:T | t | c | 0.1977 | 0.048 | 3.78E-05 | ++++++-++- | 7.279 | 9 | 0.6081 |
| ADNC Score | chr11 | 107,802,574 | chr11:107802574:A:G | a | g | -0.1968 | 0.048 | 4.07E-05 | -----+--- | 7.273 | 9 | 0.6087 |
| ADNC Score | chr11 | 107,803,469 | chr11:107803469:C:T | t | c | 0.1986 | 0.048 | 3.49E-05 | ++++++-++- | 7.556 | 9 | 0.5794 |
| ADNC Score | chr11 | 107,806,673 | chr11:107806673:C:CA | ca | c | 0.196 | 0.048 | 4.52E-05 | ++++++-++- | 7.166 | 9 | 0.6199 |
| ADNC Score | chr11 | 107,808,194 | chr11:107808194:T:C | t | c | -0.197 | 0.048 | 4.10E-05 | -----+--- | 7.71 | 9 | 0.5636 |
| ADNC Score | chr11 | 107,810,140 | chr11:107810140:A:G | a | g | -0.1994 | 0.048 | 3.33E-05 | -----+--- | 7.824 | 9 | 0.552 |
| ADNC Score | chr11 | 107,812,471 | chr11:107812471:T:G | t | g | -0.2133 | 0.0485 | 1.10E-05 | -----+--- | 6.73 | 9 | 0.6653 |
| ADNC Score | chr11 | 107,879,727 | chr11:107879727:A:C | a | c | -0.2492 | 0.0604 | 3.64E-05 | +---+--+-- | 5.443 | 9 | 0.7941 |
| ADNC Score | chr11 | 129,325,931 | chr11:129325931:C:T | t | c | 0.686 | 0.1652 | 3.28E-05 | --++++++-? | 7.144 | 8 | 0.5212 |
| ADNC Score | chr11 | 129,375,888 | chr11:129375888:G:A | a | g | 0.5758 | 0.1395 | 3.67E-05 | --+++++++? | 5.301 | 8 | 0.725 |
| ADNC Score | chr11 | 129,466,743 | chr11:129466743:GGGGTTTC:G | g | ggggtttc | 0.8787 | 0.2153 | 4.47E-05 | -+++?+++?? | 2.028 | 6 | 0.9171 |
| ADNC Score | chr12 | 487,699 | chr12:487699:C:T | t | c | 0.4094 | 0.1049 | 9.53E-05 | +++++++++ | 10.439 | 9 | 0.3161 |
| ADNC Score | chr12 | 23,951,596 | chr12:23951596:T:A | a | t | -0.2577 | 0.0565 | 5.02E-06 | +---+----+ | 12.187 | 9 | 0.203 |
| ADNC Score | chr12 | 23,954,482 | chr12:23954482:G:A | a | g | -0.2619 | 0.0553 | 2.16E-06 | +-------+ | 7.594 | 9 | 0.5755 |
| ADNC Score | chr12 | 23,956,311 | chr12:23956311:T:C | t | c | 0.2309 | 0.054 | 1.88E-05 | -++++++++- | 9.854 | 9 | 0.3624 |
| ADNC Score | chr12 | 23,962,223 | chr12:23962223:C:A | a | c | 0.2331 | 0.0595 | 8.99E-05 | -++++++-+- | 14.33 | 9 | 0.1111 |
| ADNC Score | chr12 | 41,865,673 | chr12:41865673:T:C | t | c | -0.8385 | 0.1911 | 1.15E-05 | +?--+----? | 8.099 | 7 | 0.324 |
| ADNC Score | chr12 | 53,796,166 | chr12:53796166:A:AC | a | ac | -1.1119 | 0.2766 | 5.82E-05 | ?+--?---?? | 4.28 | 5 | 0.5098 |
| ADNC Score | chr12 | 53,797,895 | chr12:53797895:A:G | a | g | -1.1123 | 0.2765 | 5.77E-05 | ?+--?---?? | 4.283 | 5 | 0.5094 |

| | | | | | | | | | | | | |
|---|---|---|---|---|---|---|---|---|---|---|---|---|
| ADNC Score | chr12 | 54,856,226 | chr12:54856226:T:C | t | c | -0.7799 | 0.1959 | 6.88E-05 | +---+----? | 5.764 | 8 | 0.6737 |
| ADNC Score | chr12 | 56,285,425 | chr12:56285425:G:C | c | g | 1.3349 | 0.2882 | 3.63E-06 | ?+++??+-?- | 6.316 | 5 | 0.2767 |
| ADNC Score | chr12 | 56,343,467 | chr12:56343467:C:A | a | c | 1.3267 | 0.2885 | 4.26E-06 | ?+++??+-?- | 6.316 | 5 | 0.2767 |
| ADNC Score | chr12 | 59,157,582 | chr12:59157582:G:T | t | g | 1.5225 | 0.3707 | 4.00E-05 | ??+???+-?+ | 0.656 | 3 | 0.8836 |
| ADNC Score | chr12 | 67,748,119 | chr12:67748119:T:C | t | c | -0.719 | 0.1843 | 9.60E-05 | +--++----? | 21.333 | 8 | 0.006315 |
| ADNC Score | chr12 | 72,735,355 | chr12:72735355:T:C | t | c | -0.2522 | 0.0572 | 1.04E-05 | ----+--+-+ | 10.625 | 9 | 0.3023 |
| ADNC Score | chr12 | 104,106,107 | chr12:104106107:A:T | a | t | -0.6202 | 0.1587 | 9.32E-05 | -------+-- | 5.995 | 9 | 0.7404 |
| ADNC Score | chr12 | 104,108,261 | chr12:104108261:C:T | t | c | 0.6321 | 0.1574 | 5.94E-05 | +++++++-+- | 6.056 | 9 | 0.7343 |
| ADNC Score | chr12 | 104,195,232 | chr12:104195232:T:C | t | c | -0.6761 | 0.1584 | 1.97E-05 | -------+-- | 6.035 | 9 | 0.7364 |
| ADNC Score | chr12 | 123,656,970 | chr12:123656970:C:CT | ct | c | 0.9501 | 0.2429 | 9.17E-05 | +-++-+++?? | 9.084 | 7 | 0.2467 |
| ADNC Score | chr12 | 123,760,907 | chr12:123760907:A:C | a | c | -0.9568 | 0.2427 | 8.09E-05 | -+--+---?+ | 8.846 | 8 | 0.3554 |
| ADNC Score | chr12 | 126,018,531 | chr12:126018531:C:T | t | c | 0.3379 | 0.0862 | 8.88E-05 | ++++-++-++ | 10.812 | 9 | 0.2888 |
| ADNC Score | chr13 | 19,645,556 | chr13:19645556:G:C | c | g | 1.0019 | 0.2506 | 6.38E-05 | ?-++?+--+? | 6.913 | 6 | 0.329 |
| ADNC Score | chr13 | 19,654,162 | chr13:19654162:T:C | t | c | -1.0077 | 0.251 | 5.96E-05 | ?+--?-++-? | 6.907 | 6 | 0.3296 |
| ADNC Score | chr13 | 19,657,497 | chr13:19657497:G:C | c | g | 1.0013 | 0.2505 | 6.41E-05 | ?-++?+--+? | 6.91 | 6 | 0.3293 |
| ADNC Score | chr13 | 19,757,723 | chr13:19757723:G:A | a | g | 1.033 | 0.2542 | 4.81E-05 | ?-++?+--+? | 5.722 | 6 | 0.455 |
| ADNC Score | chr13 | 19,897,546 | chr13:19897546:C:T | t | c | 1.0587 | 0.253 | 2.87E-05 | ?-++?+--+? | 4.739 | 6 | 0.5777 |
| ADNC Score | chr13 | 35,679,539 | chr13:35679539:G:T | t | g | 1.1302 | 0.283 | 6.51E-05 | --+???+++? | 1.673 | 5 | 0.8923 |
| ADNC Score | chr13 | 40,587,842 | chr13:40587842:C:G | c | g | -0.247 | 0.0633 | 9.46E-05 | -+-------- | 4.455 | 9 | 0.879 |
| ADNC Score | chr13 | 40,631,152 | chr13:40631152:G:A | a | g | 0.2652 | 0.0635 | 2.98E-05 | +-+++++++- | 4.204 | 9 | 0.8975 |
| ADNC Score | chr13 | 40,655,925 | chr13:40655925:C:T | t | c | 0.262 | 0.0662 | 7.63E-05 | +-+++++++- | 3.442 | 9 | 0.9442 |
| ADNC Score | chr13 | 49,737,526 | chr13:49737526:CTGTTT:C | ctgttt | c | -1.0413 | 0.2589 | 5.79E-05 | ?---??-??? | 5.093 | 3 | 0.1651 |
| ADNC Score | chr13 | 54,523,454 | chr13:54523454:T:C | t | c | -0.724 | 0.1803 | 5.95E-05 | ++-------? | 5.269 | 8 | 0.7285 |
| ADNC Score | chr13 | 92,121,926 | chr13:92121926:G:A | a | g | 0.5519 | 0.1316 | 2.74E-05 | -+++++-+++ | 10.992 | 9 | 0.2762 |
| ADNC Score | chr13 | 94,653,707 | chr13:94653707:A:C | a | c | 0.2648 | 0.0677 | 9.14E-05 | ++++++--++ | 7.462 | 9 | 0.5892 |
| ADNC Score | chr13 | 94,654,060 | chr13:94654060:A:G | a | g | 0.2683 | 0.0677 | 7.49E-05 | ++++++--++ | 7.663 | 9 | 0.5685 |
| ADNC Score | chr13 | 107,796,381 | chr13:107796381:G:A | a | g | 0.1958 | 0.0497 | 8.07E-05 | +++++-+++- | 8.21 | 9 | 0.5131 |
| ADNC Score | chr13 | 107,797,776 | chr13:107797776:G:C | c | g | 0.1909 | 0.0488 | 9.10E-05 | +++++-+++- | 8.238 | 9 | 0.5103 |
| ADNC Score | chr13 | 109,643,517 | chr13:109643517:T:C | t | c | 0.2238 | 0.0562 | 6.89E-05 | +++++++-+- | 7.206 | 9 | 0.6157 |
| ADNC Score | chr14 | 23,275,591 | chr14:23275591:ACAT:A | a | acat | -0.2032 | 0.0504 | 5.44E-05 | +--------- | 10.323 | 9 | 0.325 |
| ADNC Score | chr14 | 36,354,877 | chr14:36354877:G:A | a | g | 0.9513 | 0.2348 | 5.10E-05 | +?++?+-++- | 4.553 | 7 | 0.7143 |
| ADNC Score | chr14 | 41,648,023 | chr14:41648023:C:T | t | c | 0.778 | 0.1992 | 9.42E-05 | ??++??-++? | 7.368 | 4 | 0.1177 |
| ADNC Score | chr14 | 83,326,615 | chr14:83326615:C:T | t | c | 0.6645 | 0.1641 | 5.13E-05 | ++++-++++- | 5.459 | 9 | 0.7926 |
| ADNC Score | chr14 | 96,295,934 | chr14:96295934:G:A | a | g | 0.2046 | 0.0523 | 9.23E-05 | ++++-+-++- | 8.968 | 9 | 0.4402 |
| ADNC Score | chr15 | 46,917,952 | chr15:46917952:G:A | a | g | 1.3154 | 0.3268 | 5.69E-05 | ++?++?+++ | 4.968 | 7 | 0.6638 |
| ADNC Score | chr15 | 49,508,592 | chr15:49508592:T:C | t | c | 0.1951 | 0.0501 | 9.92E-05 | ++++++-++- | 9.418 | 9 | 0.3996 |
| ADNC Score | chr15 | 49,837,386 | chr15:49837386:T:G | t | g | -0.5143 | 0.1295 | 7.11E-05 | ----+--?-- | 12.536 | 8 | 0.1289 |
| ADNC Score | chr15 | 49,842,266 | chr15:49842266:A:G | a | g | -0.5188 | 0.1299 | 6.52E-05 | ----+--?-- | 12.485 | 8 | 0.1308 |
| ADNC Score | chr15 | 49,854,505 | chr15:49854505:G:C | c | g | 0.5264 | 0.1306 | 5.60E-05 | ++++-++?+- | 12.501 | 8 | 0.1302 |
| ADNC Score | chr15 | 62,273,141 | chr15:62273141:G:A | a | g | 0.696 | 0.1746 | 6.73E-05 | ??+-+++-++ | 8.596 | 7 | 0.2829 |
| ADNC Score | chr15 | 62,276,155 | chr15:62276155:G:A | a | g | 0.7127 | 0.1766 | 5.45E-05 | ??+-+++-++ | 8.509 | 7 | 0.2898 |
| ADNC Score | chr15 | 72,408,716 | chr15:72408716:A:G | a | g | -0.5894 | 0.1491 | 7.75E-05 | ----?----+ | 7.52 | 8 | 0.4817 |
| ADNC Score | chr15 | 80,857,247 | chr15:80857247:G:A | a | g | 0.8834 | 0.2268 | 9.84E-05 | +++--+++?+ | 14.961 | 8 | 0.05991 |
| ADNC Score | chr15 | 99,096,865 | chr15:99096865:C:T | t | c | 1.2547 | 0.3218 | 9.65E-05 | +????+?++? | 0.966 | 3 | 0.8095 |
| ADNC Score | chr16 | 26,461,059 | chr16:26461059:G:C | c | g | 1.509 | 0.38 | 7.16E-05 | ?+????+?+? | 2.957 | 2 | 0.228 |
| ADNC Score | chr16 | 29,220,649 | chr16:29220649:C:A | a | c | 0.1864 | 0.0476 | 9.05E-05 | +-++++-+++ | 8.973 | 9 | 0.4398 |

| | | | | | | | | | | | | |
|---|---|---|---|---|---|---|---|---|---|---|---|---|
| ADNC Score | chr16 | 29,220,650 | chr16:29220650:T:G | t | g | -0.1884 | 0.0475 | 7.41E-05 | -+----+--- | 8.506 | 9 | 0.4841 |
| ADNC Score | chr16 | 46,553,821 | chr16:46553821:GTA:G | g | gta | 0.9333 | 0.2291 | 4.61E-05 | ?+++++--++ | 7.836 | 8 | 0.4497 |
| ADNC Score | chr16 | 46,654,343 | chr16:46654343:T:C | t | c | -0.8871 | 0.2189 | 5.09E-05 | -+----++-- | 8.74 | 9 | 0.4616 |
| ADNC Score | chr16 | 69,720,985 | chr16:69720985:A:G | a | g | 0.4426 | 0.1135 | 9.64E-05 | -++++++++ | 6.352 | 8 | 0.6079 |
| ADNC Score | chr16 | 69,742,777 | chr16:69742777:C:T | t | c | -0.4438 | 0.114 | 9.98E-05 | +--------? | 6.299 | 8 | 0.6138 |
| ADNC Score | chr16 | 71,330,858 | chr16:71330858:A:G | a | g | 0.2005 | 0.05 | 6.02E-05 | +++++++-+ | 4.896 | 9 | 0.8433 |
| ADNC Score | chr16 | 71,332,205 | chr16:71332205:A:G | a | g | 0.1977 | 0.0499 | 7.57E-05 | +++++++-+ | 4.824 | 9 | 0.8494 |
| ADNC Score | chr16 | 71,332,282 | chr16:71332282:A:T | a | t | 0.198 | 0.05 | 7.41E-05 | +++++++-+ | 4.852 | 9 | 0.847 |
| ADNC Score | chr16 | 71,334,614 | chr16:71334614:C:T | t | c | -0.1988 | 0.0499 | 6.85E-05 | -------+-- | 5.081 | 9 | 0.8272 |
| ADNC Score | chr16 | 71,335,057 | chr16:71335057:G:T | t | g | -0.2047 | 0.05 | 4.17E-05 | -------+-- | 4.804 | 9 | 0.8511 |
| ADNC Score | chr16 | 71,335,603 | chr16:71335603:AG:A | a | ag | -0.207 | 0.0501 | 3.66E-05 | -------+-- | 4.838 | 9 | 0.8482 |
| ADNC Score | chr16 | 71,336,115 | chr16:71336115:T:A | a | t | -0.2053 | 0.05 | 4.00E-05 | -------+-- | 4.854 | 9 | 0.8468 |
| ADNC Score | chr16 | 71,337,071 | chr16:71337071:G:A | a | g | -0.2059 | 0.05 | 3.84E-05 | -------+-- | 4.761 | 9 | 0.8546 |
| ADNC Score | chr16 | 71,343,408 | chr16:71343408:G:A | a | g | 0.2033 | 0.0488 | 3.04E-05 | +++++++-+ | 4.547 | 9 | 0.8719 |
| ADNC Score | chr16 | 71,344,010 | chr16:71344010:A:G | a | g | -0.2071 | 0.049 | 2.34E-05 | -------+-- | 4.506 | 9 | 0.8751 |
| ADNC Score | chr16 | 71,348,517 | chr16:71348517:T:G | t | g | -0.1971 | 0.0493 | 6.32E-05 | -+-----+-- | 7.253 | 9 | 0.6108 |
| ADNC Score | chr16 | 75,517,054 | chr16:75517054:G:A | a | g | 0.9625 | 0.2357 | 4.44E-05 | --++??-?++ | 13.781 | 6 | 0.03218 |
| ADNC Score | chr16 | 75,683,943 | chr16:75683943:C:T | t | c | 0.9112 | 0.2149 | 2.24E-05 | -?++---++? | 13.559 | 7 | 0.05961 |
| ADNC Score | chr16 | 75,837,636 | chr16:75837636:C:A | a | c | 0.8956 | 0.2232 | 6.02E-05 | -?++?--?+? | 7.611 | 5 | 0.179 |
| ADNC Score | chr16 | 81,943,852 | chr16:81943852:C:G | c | g | -0.1886 | 0.0477 | 7.64E-05 | ----+----- | 13.09 | 9 | 0.1586 |
| ADNC Score | chr17 | 44,351,170 | chr17:44351170:G:A | a | g | 0.4011 | 0.0912 | 1.09E-05 | ++++--+++- | 16.797 | 9 | 0.05198 |
| ADNC Score | chr17 | 44,502,360 | chr17:44502360:T:G | t | g | -0.3534 | 0.0887 | 6.75E-05 | +---++---+ | 15.525 | 9 | 0.07749 |
| ADNC Score | chr17 | 44,803,439 | chr17:44803439:C:T | t | c | 0.3268 | 0.084 | 9.98E-05 | -+++--+++- | 11.998 | 9 | 0.2134 |
| ADNC Score | chr17 | 44,810,282 | chr17:44810282:C:A | a | c | 0.3271 | 0.084 | 9.87E-05 | -+++--+++- | 12.009 | 9 | 0.2128 |
| ADNC Score | chr17 | 44,814,497 | chr17:44814497:C:T | t | c | 0.3272 | 0.084 | 9.82E-05 | -+++--+++- | 12.008 | 9 | 0.2128 |
| ADNC Score | chr17 | 44,816,071 | chr17:44816071:C:A | a | c | 0.33 | 0.0841 | 8.78E-05 | -+++--+++- | 12.096 | 9 | 0.2079 |
| ADNC Score | chr17 | 44,856,258 | chr17:44856258:G:A | a | g | 0.3272 | 0.0839 | 9.67E-05 | -+++--+++? | 12.114 | 8 | 0.1462 |
| ADNC Score | chr17 | 67,405,803 | chr17:67405803:C:T | t | c | -0.5781 | 0.1356 | 2.02E-05 | -------+-+ | 7.298 | 9 | 0.6062 |
| ADNC Score | chr17 | 72,278,079 | chr17:72278079:T:C | t | c | 0.2157 | 0.0548 | 8.18E-05 | ++++-++++ | 6.161 | 9 | 0.7237 |
| ADNC Score | chr17 | 72,291,383 | chr17:72291383:G:C | c | g | -0.2559 | 0.0531 | 1.45E-06 | ----+----- | 3.915 | 9 | 0.9169 |
| ADNC Score | chr17 | 72,293,175 | chr17:72293175:A:G | a | g | 0.2372 | 0.0525 | 6.27E-06 | ++++-++++ | 2.892 | 9 | 0.9684 |
| ADNC Score | chr17 | 72,294,754 | chr17:72294754:AT:A | a | at | -0.2383 | 0.0525 | 5.72E-06 | ----+----- | 2.886 | 9 | 0.9687 |
| ADNC Score | chr17 | 72,295,015 | chr17:72295015:C:A | a | c | -0.2379 | 0.0525 | 5.89E-06 | ----+----- | 2.888 | 9 | 0.9686 |
| ADNC Score | chr17 | 72,297,185 | chr17:72297185:G:A | a | g | -0.2403 | 0.0525 | 4.66E-06 | ----+--+-- | 3.33 | 9 | 0.9498 |
| ADNC Score | chr17 | 72,297,729 | chr17:72297729:G:A | a | g | -0.2399 | 0.0525 | 4.95E-06 | ----+--+-- | 3.253 | 9 | 0.9534 |
| ADNC Score | chr17 | 72,297,812 | chr17:72297812:G:A | a | g | -0.2421 | 0.0525 | 4.08E-06 | ----+--+-- | 3.208 | 9 | 0.9555 |
| ADNC Score | chr17 | 72,298,668 | chr17:72298668:CAA:C | caa | c | 0.2354 | 0.053 | 8.74E-06 | ++++-++-++ | 4.115 | 9 | 0.9037 |
| ADNC Score | chr17 | 72,299,461 | chr17:72299461:A:G | a | g | 0.2398 | 0.0524 | 4.78E-06 | ++++-++-++ | 3.082 | 9 | 0.961 |
| ADNC Score | chr17 | 72,299,891 | chr17:72299891:A:G | a | g | 0.2376 | 0.0523 | 5.50E-06 | ++++-++-++ | 3.863 | 9 | 0.9202 |
| ADNC Score | chr17 | 72,300,105 | chr17:72300105:A:G | a | g | 0.2598 | 0.0535 | 1.22E-06 | ++++-++++ | 4.54 | 9 | 0.8724 |
| ADNC Score | chr17 | 72,302,429 | chr17:72302429:T:C | t | c | 0.231 | 0.0524 | 1.06E-05 | ++++-++-++ | 3.852 | 9 | 0.9209 |
| ADNC Score | chr17 | 72,302,757 | chr17:72302757:G:C | c | g | -0.2325 | 0.0524 | 9.08E-06 | ----+--+-- | 3.878 | 9 | 0.9192 |
| ADNC Score | chr17 | 72,302,917 | chr17:72302917:AT:A | a | at | -0.2317 | 0.0524 | 9.70E-06 | ----+--+-- | 3.841 | 9 | 0.9216 |
| ADNC Score | chr17 | 72,304,311 | chr17:72304311:G:A | a | g | 0.225 | 0.0522 | 1.64E-05 | ++++-++++ | 8.429 | 9 | 0.4916 |
| ADNC Score | chr17 | 72,306,647 | chr17:72306647:A:T | a | t | -0.2111 | 0.0523 | 5.38E-05 | ----+----- | 5.511 | 9 | 0.7877 |
| ADNC Score | chr17 | 72,315,053 | chr17:72315053:G:T | t | g | 0.2298 | 0.0582 | 7.88E-05 | -+++-++-++ | 2.996 | 9 | 0.9645 |

| | | | | | | | | | | | | |
|---|---|---|---|---|---|---|---|---|---|---|---|---|
| ADNC Score | chr17 | 72,315,143 | chr17:72315143:G:A | a | g | 0.2296 | 0.0582 | 8.00E-05 | -+++-++-++ | 3.011 | 9 | 0.9639 |
| ADNC Score | chr17 | 72,315,214 | chr17:72315214:C:T | t | c | 0.2281 | 0.0582 | 8.93E-05 | -+++-++-++ | 3.07 | 9 | 0.9615 |
| ADNC Score | chr17 | 72,315,370 | chr17:72315370:C:T | t | c | 0.2253 | 0.0574 | 8.53E-05 | -+++-++-++ | 2.886 | 9 | 0.9686 |
| ADNC Score | chr17 | 72,315,979 | chr17:72315979:C:A | a | c | 0.2336 | 0.0581 | 5.81E-05 | -+++-++-++ | 3.281 | 9 | 0.9521 |
| ADNC Score | chr17 | 72,316,078 | chr17:72316078:A:G | a | g | -0.2347 | 0.0572 | 4.09E-05 | +------+-- | 5.364 | 9 | 0.8015 |
| ADNC Score | chr17 | 72,317,101 | chr17:72317101:G:A | a | g | 0.2277 | 0.0581 | 8.93E-05 | -+++-++-++ | 2.927 | 9 | 0.9671 |
| ADNC Score | chr17 | 72,317,158 | chr17:72317158:T:C | t | c | -0.2287 | 0.058 | 8.01E-05 | +---+--+-- | 2.942 | 9 | 0.9666 |
| ADNC Score | chr17 | 72,318,106 | chr17:72318106:A:G | a | g | -0.2295 | 0.058 | 7.55E-05 | +---+--+-- | 3.635 | 9 | 0.9338 |
| ADNC Score | chr17 | 72,318,596 | chr17:72318596:T:C | t | c | -0.2277 | 0.058 | 8.59E-05 | +---+--+-- | 3.549 | 9 | 0.9385 |
| ADNC Score | chr17 | 72,319,130 | chr17:72319130:A:C | a | c | -0.2264 | 0.058 | 9.40E-05 | +---+--+-- | 3.704 | 9 | 0.9298 |
| ADNC Score | chr17 | 72,319,151 | chr17:72319151:A:G | a | g | -0.2252 | 0.0578 | 9.85E-05 | +---+--+-- | 3.708 | 9 | 0.9296 |
| ADNC Score | chr17 | 72,319,749 | chr17:72319749:G:A | a | g | 0.2266 | 0.0579 | 8.97E-05 | -+++-++-++ | 4.277 | 9 | 0.8922 |
| ADNC Score | chr17 | 72,574,209 | chr17:72574209:G:A | a | g | 1.4479 | 0.3547 | 4.45E-05 | ?+????+?+ | 0.383 | 2 | 0.8257 |
| ADNC Score | chr17 | 78,133,130 | chr17:78133130:GA:G | g | ga | 0.3582 | 0.0815 | 1.10E-05 | --++-+--++ | 10.566 | 9 | 0.3066 |
| ADNC Score | chr17 | 78,133,258 | chr17:78133258:C:A | a | c | 0.3234 | 0.0811 | 6.70E-05 | +-++-+--++ | 7.83 | 9 | 0.5513 |
| ADNC Score | chr17 | 78,133,261 | chr17:78133261:T:C | t | c | -0.3278 | 0.0818 | 6.11E-05 | -+--+-++-- | 7.61 | 9 | 0.5738 |
| ADNC Score | chr17 | 78,459,501 | chr17:78459501:G:A | a | g | 0.6282 | 0.1459 | 1.67E-05 | ++++-++-+- | 15.682 | 9 | 0.07382 |
| ADNC Score | chr18 | 33,988,622 | chr18:33988622:T:C | t | c | 0.9358 | 0.2308 | 5.02E-05 | ?+++++++? | 6.362 | 7 | 0.4982 |
| ADNC Score | chr18 | 34,051,556 | chr18:34051556:AG:A | a | ag | -0.942 | 0.2295 | 4.04E-05 | ?-------?+ | 6.355 | 7 | 0.4989 |
| ADNC Score | chr18 | 50,923,320 | chr18:50923320:CAA:C | caa | c | -0.2817 | 0.0714 | 8.00E-05 | ---+-----+ | 8.91 | 9 | 0.4456 |
| ADNC Score | chr18 | 51,955,366 | chr18:51955366:G:A | a | g | 0.2098 | 0.0506 | 3.35E-05 | +++++++++ | 5.034 | 9 | 0.8313 |
| ADNC Score | chr18 | 51,958,632 | chr18:51958632:A:G | a | g | -0.216 | 0.0506 | 1.97E-05 | --------- | 5.816 | 9 | 0.7582 |
| ADNC Score | chr18 | 51,962,015 | chr18:51962015:C:T | t | c | 0.2055 | 0.0512 | 5.93E-05 | +++++++++ | 8.131 | 9 | 0.521 |
| ADNC Score | chr18 | 61,919,462 | chr18:61919462:G:C | c | g | 0.347 | 0.089 | 9.75E-05 | +-+++++-++ | 7.141 | 9 | 0.6224 |
| ADNC Score | chr18 | 73,490,746 | chr18:73490746:T:C | t | c | 0.4397 | 0.1091 | 5.61E-05 | +++-+++++- | 9.926 | 9 | 0.3565 |
| ADNC Score | chr19 | 4,665,213 | chr19:4665213:CTGTT:C | ctgtt | c | -0.2805 | 0.0714 | 8.54E-05 | ----+---+- | 13.008 | 9 | 0.1623 |
| ADNC Score | chr19 | 8,923,770 | chr19:8923770:T:C | t | c | 0.2975 | 0.0713 | 3.04E-05 | +-+++++++- | 12.87 | 9 | 0.1686 |
| ADNC Score | chr19 | 8,924,496 | chr19:8924496:C:T | t | c | -0.2994 | 0.0712 | 2.59E-05 | -+-------- | 12.93 | 9 | 0.1658 |
| ADNC Score | chr19 | 8,957,698 | chr19:8957698:A:G | a | g | -0.5546 | 0.1336 | 3.29E-05 | ++-------+ | 12.397 | 9 | 0.1919 |
| ADNC Score | chr19 | 8,960,886 | chr19:8960886:C:T | t | c | 0.5747 | 0.1343 | 1.87E-05 | --+++++++- | 13.05 | 9 | 0.1604 |
| ADNC Score | chr19 | 44,834,661 | chr19:44834661:G:A | a | g | -0.4991 | 0.1226 | 4.67E-05 | ---------+ | 3.696 | 9 | 0.9303 |
| ADNC Score | chr19 | 44,865,946 | chr19:44865946:G:A | a | g | 0.5731 | 0.147 | 9.73E-05 | ++++-++++- | 8.521 | 9 | 0.4826 |
| ADNC Score | chr19 | 44,878,777 | chr19:44878777:A:G | a | g | -0.2866 | 0.0481 | 2.47E-09 | --------- | 5.945 | 9 | 0.7454 |
| ADNC Score | chr19 | 44,879,418 | chr19:44879418:G:A | a | g | 0.5536 | 0.1316 | 2.61E-05 | ++++-++++- | 7.884 | 9 | 0.5458 |
| ADNC Score | chr19 | 44,879,709 | chr19:44879709:C:T | t | c | 0.2182 | 0.0493 | 9.54E-06 | +++++++++ | 7.022 | 9 | 0.6348 |
| ADNC Score | chr19 | 44,879,804 | chr19:44879804:G:T | t | g | 0.2157 | 0.0492 | 1.19E-05 | +++++++++ | 6.932 | 9 | 0.6442 |
| ADNC Score | chr19 | 44,881,148 | chr19:44881148:T:C | t | c | -0.3097 | 0.0727 | 2.02E-05 | +---+-+--- | 10.414 | 9 | 0.318 |
| ADNC Score | chr19 | 44,883,210 | chr19:44883210:G:GTAA | g | gtaa | 0.6785 | 0.0646 | 8.10E-26 | +++++++++ | 9.632 | 9 | 0.3811 |
| ADNC Score | chr19 | 44,883,377 | chr19:44883377:C:T | t | c | -0.5956 | 0.1147 | 2.06E-07 | --------- | 8.302 | 9 | 0.504 |
| ADNC Score | chr19 | 44,884,202 | chr19:44884202:C:G | c | g | 0.6805 | 0.0644 | 4.48E-26 | +++++++++ | 9.337 | 9 | 0.4068 |
| ADNC Score | chr19 | 44,884,339 | chr19:44884339:G:A | a | g | -0.6824 | 0.0644 | 2.96E-26 | --------- | 9.325 | 9 | 0.4078 |
| ADNC Score | chr19 | 44,884,873 | chr19:44884873:G:A | a | g | -0.6575 | 0.0638 | 6.10E-25 | ---------+ | 10.593 | 9 | 0.3046 |
| ADNC Score | chr19 | 44,885,243 | chr19:44885243:A:G | a | g | 0.5803 | 0.0549 | 4.49E-26 | +++++++++ | 7.577 | 9 | 0.5773 |
| ADNC Score | chr19 | 44,887,076 | chr19:44887076:A:G | a | g | 0.5977 | 0.0547 | 9.36E-28 | +++++++++ | 7.528 | 9 | 0.5823 |
| ADNC Score | chr19 | 44,888,997 | chr19:44888997:C:T | t | c | -0.7756 | 0.06 | 3.20E-38 | --------- | 7.727 | 9 | 0.5619 |
| ADNC Score | chr19 | 44,889,415 | chr19:44889415:AT:A | a | at | 0.6148 | 0.1488 | 3.61E-05 | ++++-++++- | 10.785 | 9 | 0.2907 |

| | | | | | | | | | | | | |
|---|---|---|---|---|---|---|---|---|---|---|---|---|
| ADNC Score | chr19 | 44,891,079 | chr19:44891079:T:C | t | c | 0.6955 | 0.0642 | 2.45E-27 | +++++++++ | 9.948 | 9 | 0.3547 |
| ADNC Score | chr19 | 44,891,712 | chr19:44891712:T:G | t | g | 0.6054 | 0.055 | 3.24E-28 | +++++++++ | 8.111 | 9 | 0.523 |
| ADNC Score | chr19 | 44,892,009 | chr19:44892009:G:A | a | g | -0.3265 | 0.0487 | 2.00E-11 | --------+ | 21.385 | 9 | 0.01105 |
| ADNC Score | chr19 | 44,892,362 | chr19:44892362:A:G | a | g | 0.7219 | 0.0644 | 3.75E-29 | +++++++++ | 7.685 | 9 | 0.5662 |
| ADNC Score | chr19 | 44,892,457 | chr19:44892457:T:C | t | c | 0.6096 | 0.0549 | 1.08E-28 | +++++++++ | 7.11 | 9 | 0.6257 |
| ADNC Score | chr19 | 44,892,587 | chr19:44892587:G:A | a | g | -0.5835 | 0.0706 | 1.43E-16 | --------- | 3.254 | 9 | 0.9534 |
| ADNC Score | chr19 | 44,892,652 | chr19:44892652:C:G | c | g | 0.7239 | 0.0646 | 3.61E-29 | +++++++++ | 8.201 | 9 | 0.514 |
| ADNC Score | chr19 | 44,892,887 | chr19:44892887:C:T | t | c | -0.7271 | 0.0646 | 2.24E-29 | --------- | 7.939 | 9 | 0.5403 |
| ADNC Score | chr19 | 44,892,962 | chr19:44892962:C:T | t | c | -0.604 | 0.0548 | 3.18E-28 | --------- | 6.439 | 9 | 0.6953 |
| ADNC Score | chr19 | 44,893,408 | chr19:44893408:G:T | t | g | -0.6528 | 0.0558 | 1.41E-31 | --------- | 5.519 | 9 | 0.787 |
| ADNC Score | chr19 | 44,893,716 | chr19:44893716:G:A | a | g | -0.6043 | 0.1148 | 1.39E-07 | --------- | 7.88 | 9 | 0.5463 |
| ADNC Score | chr19 | 44,893,972 | chr19:44893972:G:A | a | g | 0.6698 | 0.1498 | 7.79E-06 | ++++-++++ | 8.395 | 8 | 0.3958 |
| ADNC Score | chr19 | 44,894,050 | chr19:44894050:C:T | t | c | -0.6248 | 0.1151 | 5.66E-08 | --------- | 7.787 | 9 | 0.5557 |
| ADNC Score | chr19 | 44,894,695 | chr19:44894695:T:C | t | c | 0.605 | 0.1144 | 1.25E-07 | +++++++++ | 7.996 | 9 | 0.5346 |
| ADNC Score | chr19 | 44,895,376 | chr19:44895376:G:C | c | g | 0.3321 | 0.067 | 7.17E-07 | -+++-+-++- | 14.645 | 9 | 0.1012 |
| ADNC Score | chr19 | 44,895,528 | chr19:44895528:C:T | t | c | -0.6281 | 0.1154 | 5.28E-08 | --------- | 7.827 | 9 | 0.5516 |
| ADNC Score | chr19 | 44,895,560 | chr19:44895560:C:T | t | c | 0.2136 | 0.0488 | 1.23E-05 | --+++++++ | 9.749 | 9 | 0.3712 |
| ADNC Score | chr19 | 44,896,087 | chr19:44896087:G:T | t | g | -0.6264 | 0.1153 | 5.53E-08 | --------- | 7.823 | 9 | 0.5521 |
| ADNC Score | chr19 | 44,896,639 | chr19:44896639:G:A | a | g | -0.6275 | 0.1154 | 5.42E-08 | --------- | 7.823 | 9 | 0.5521 |
| ADNC Score | chr19 | 44,897,227 | chr19:44897227:T:TG | t | tg | 0.6267 | 0.1153 | 5.43E-08 | +++++++++ | 7.733 | 9 | 0.5613 |
| ADNC Score | chr19 | 44,897,468 | chr19:44897468:C:T | t | c | -0.6264 | 0.1153 | 5.53E-08 | --------- | 7.823 | 9 | 0.5521 |
| ADNC Score | chr19 | 44,897,490 | chr19:44897490:T:A | a | t | 0.6585 | 0.1494 | 1.05E-05 | ++++-++++ | 8.198 | 8 | 0.4144 |
| ADNC Score | chr19 | 44,898,409 | chr19:44898409:A:G | a | g | -0.2311 | 0.0484 | 1.82E-06 | -+-------- | 10.307 | 9 | 0.3262 |
| ADNC Score | chr19 | 44,898,611 | chr19:44898611:T:C | t | c | 0.5259 | 0.1038 | 4.06E-07 | +++++++++ | 5.984 | 9 | 0.7416 |
| ADNC Score | chr19 | 44,899,005 | chr19:44899005:T:G | t | g | -0.6566 | 0.1494 | 1.11E-05 | ----+----? | 8.242 | 8 | 0.4102 |
| ADNC Score | chr19 | 44,899,220 | chr19:44899220:C:T | t | c | 0.2279 | 0.0488 | 3.01E-06 | --+++++++ | 9.184 | 9 | 0.4204 |
| ADNC Score | chr19 | 44,899,959 | chr19:44899959:C:T | t | c | -0.6456 | 0.1127 | 1.01E-08 | --------- | 6.447 | 9 | 0.6944 |
| ADNC Score | chr19 | 44,900,155 | chr19:44900155:C:T | t | c | 0.2783 | 0.0475 | 4.84E-09 | +++++++++ | 13.649 | 9 | 0.1354 |
| ADNC Score | chr19 | 44,900,601 | chr19:44900601:A:G | a | g | -0.2797 | 0.0477 | 4.65E-09 | --------+ | 13.738 | 9 | 0.1319 |
| ADNC Score | chr19 | 44,901,174 | chr19:44901174:T:C | t | c | -0.2803 | 0.0479 | 4.70E-09 | --------+ | 13.74 | 9 | 0.1319 |
| ADNC Score | chr19 | 44,901,434 | chr19:44901434:A:G | a | g | 0.2589 | 0.0537 | 1.42E-06 | +++++++-+- | 13.272 | 9 | 0.1507 |
| ADNC Score | chr19 | 44,901,600 | chr19:44901600:T:C | t | c | 0.6597 | 0.1128 | 4.95E-09 | +++++++++ | 6.73 | 9 | 0.6652 |
| ADNC Score | chr19 | 44,901,715 | chr19:44901715:T:C | t | c | -0.2837 | 0.0479 | 3.17E-09 | --------+ | 13.526 | 9 | 0.1402 |
| ADNC Score | chr19 | 44,901,805 | chr19:44901805:A:G | a | g | -0.2831 | 0.0478 | 3.18E-09 | --------+ | 13.531 | 9 | 0.14 |
| ADNC Score | chr19 | 44,902,264 | chr19:44902264:G:C | c | g | 0.2255 | 0.0488 | 3.86E-06 | --+++++++ | 8.713 | 9 | 0.4642 |
| ADNC Score | chr19 | 44,903,281 | chr19:44903281:C:CG | cg | c | 0.2848 | 0.0483 | 3.82E-09 | +-+++++++- | 11.022 | 9 | 0.2742 |
| ADNC Score | chr19 | 44,903,416 | chr19:44903416:G:A | a | g | -0.5069 | 0.0508 | 1.82E-23 | --------- | 3.86 | 9 | 0.9204 |
| ADNC Score | chr19 | 44,904,531 | chr19:44904531:G:A | a | g | 0.292 | 0.0479 | 1.11E-09 | +++++++++ | 13.52 | 9 | 0.1404 |
| ADNC Score | chr19 | 44,905,307 | chr19:44905307:A:T | a | t | -0.3247 | 0.068 | 1.76E-06 | +-----+--+ | 11.285 | 9 | 0.2567 |
| ADNC Score | chr19 | 44,905,579 | chr19:44905579:T:G | t | g | -0.2342 | 0.047 | 6.28E-07 | -+-------+ | 12.752 | 9 | 0.1741 |
| ADNC Score | chr19 | 44,905,910 | chr19:44905910:C:G | c | g | 0.2809 | 0.0497 | 1.56E-08 | -++++++++- | 20.506 | 9 | 0.01504 |
| ADNC Score | chr19 | 44,906,745 | chr19:44906745:G:A | a | g | -0.8976 | 0.0685 | 2.92E-39 | --------+ | 6.927 | 9 | 0.6447 |
| ADNC Score | chr19 | 44,907,187 | chr19:44907187:G:A | a | g | 0.2487 | 0.0491 | 4.16E-07 | --+++++++- | 11.338 | 9 | 0.2532 |
| ADNC Score | chr19 | 44,908,684 | chr19:44908684:T:C | t | c | 0.9873 | 0.0629 | 1.92E-55 | +++++++++ | 7.513 | 9 | 0.5838 |
| ADNC Score | chr19 | 44,908,822 | chr19:44908822:C:T | t | c | 0.7067 | 0.1014 | 3.18E-12 | +-++-+++++ | 8.259 | 9 | 0.5082 |
| ADNC Score | chr19 | 44,909,698 | chr19:44909698:A:C | a | c | 0.6864 | 0.1133 | 1.36E-09 | +++++++++ | 5.822 | 9 | 0.7576 |

| | | | | | | | | | | | | |
|---|---|---|---|---|---|---|---|---|---|---|---|---|
| ADNC Score | chr19 | 44,909,976 | chr19:44909976:G:T | t | g | 0.6939 | 0.1016 | 8.59E-12 | +-++-+++++ | 8.035 | 9 | 0.5307 |
| ADNC Score | chr19 | 44,910,319 | chr19:44910319:C:T | t | c | -0.4907 | 0.0578 | 2.06E-17 | +--------+ | 11.911 | 9 | 0.2183 |
| ADNC Score | chr19 | 44,911,194 | chr19:44911194:T:C | t | c | 0.2159 | 0.0493 | 1.19E-05 | -++++++-++ | 17.74 | 9 | 0.03832 |
| ADNC Score | chr19 | 44,912,456 | chr19:44912456:G:A | a | g | -0.9142 | 0.0685 | 1.38E-40 | ---------+ | 6.383 | 9 | 0.7011 |
| ADNC Score | chr19 | 44,912,678 | chr19:44912678:G:T | t | g | -0.9145 | 0.0686 | 1.34E-40 | ---------+ | 6.42 | 9 | 0.6972 |
| ADNC Score | chr19 | 44,912,921 | chr19:44912921:G:T | t | g | -0.611 | 0.0546 | 5.10E-29 | +--------- | 9.368 | 9 | 0.404 |
| ADNC Score | chr19 | 44,913,034 | chr19:44913034:C:T | t | c | 0.3188 | 0.05 | 1.83E-10 | +++++++++ | 7.841 | 9 | 0.5502 |
| ADNC Score | chr19 | 44,913,221 | chr19:44913221:A:G | a | g | 0.2135 | 0.0498 | 1.79E-05 | -+++++--++ | 19.378 | 9 | 0.02216 |
| ADNC Score | chr19 | 44,913,484 | chr19:44913484:C:T | t | c | -0.6152 | 0.0546 | 2.01E-29 | +--------- | 9.272 | 9 | 0.4125 |
| ADNC Score | chr19 | 44,915,229 | chr19:44915229:G:A | a | g | -0.6555 | 0.1604 | 4.36E-05 | ?+--+----- | 4.86 | 8 | 0.7725 |
| ADNC Score | chr19 | 44,915,533 | chr19:44915533:T:C | t | c | 0.6167 | 0.055 | 3.18E-29 | -++++++++- | 9.387 | 9 | 0.4023 |
| ADNC Score | chr19 | 44,915,704 | chr19:44915704:T:C | t | c | 0.213 | 0.0498 | 1.87E-05 | -+++++--++ | 17.35 | 9 | 0.04351 |
| ADNC Score | chr19 | 44,916,825 | chr19:44916825:A:C | a | c | 0.8899 | 0.0673 | 6.52E-40 | -++++++++- | 11.558 | 9 | 0.2394 |
| ADNC Score | chr19 | 44,917,843 | chr19:44917843:G:A | a | g | 0.3507 | 0.0501 | 2.58E-12 | +++++++++ | 6.557 | 9 | 0.6831 |
| ADNC Score | chr19 | 44,917,947 | chr19:44917947:C:T | t | c | -0.6504 | 0.1128 | 8.04E-09 | ---------- | 4.707 | 9 | 0.8591 |
| ADNC Score | chr19 | 44,917,997 | chr19:44917997:G:A | a | g | -0.7853 | 0.0636 | 5.45E-35 | +--------+ | 9.924 | 9 | 0.3567 |
| ADNC Score | chr19 | 44,918,393 | chr19:44918393:G:A | a | g | -0.6532 | 0.1125 | 6.43E-09 | ---------- | 4.815 | 9 | 0.8501 |
| ADNC Score | chr19 | 44,918,487 | chr19:44918487:G:T | t | g | 0.3242 | 0.0522 | 5.21E-10 | +++++++++ | 7.896 | 9 | 0.5447 |
| ADNC Score | chr19 | 44,918,620 | chr19:44918620:A:G | a | g | 0.2637 | 0.0495 | 9.92E-08 | -+++++--++ | 18.276 | 9 | 0.0321 |
| ADNC Score | chr19 | 44,918,715 | chr19:44918715:AG:A | a | ag | 0.3232 | 0.0522 | 6.14E-10 | +++++++++ | 7.952 | 9 | 0.539 |
| ADNC Score | chr19 | 44,918,903 | chr19:44918903:C:G | c | g | 0.8684 | 0.0596 | 4.11E-48 | +++++++++ | 8.703 | 9 | 0.4651 |
| ADNC Score | chr19 | 44,919,589 | chr19:44919589:G:A | a | g | -0.8642 | 0.0594 | 5.37E-48 | ---------- | 8.578 | 9 | 0.4771 |
| ADNC Score | chr19 | 44,919,689 | chr19:44919689:A:G | a | g | 0.866 | 0.0591 | 1.49E-48 | +++++++++ | 8.688 | 9 | 0.4665 |
| ADNC Score | chr19 | 44,920,379 | chr19:44920379:G:A | a | g | 0.3273 | 0.0525 | 4.56E-10 | +++++++++ | 6.742 | 9 | 0.6639 |
| ADNC Score | chr19 | 44,921,809 | chr19:44921809:G:A | a | g | -0.6569 | 0.1174 | 2.21E-08 | ---------- | 4.254 | 9 | 0.8939 |
| ADNC Score | chr19 | 44,921,918 | chr19:44921918:T:G | t | g | 0.2613 | 0.0502 | 1.90E-07 | -+++++--++ | 16.81 | 9 | 0.05178 |
| ADNC Score | chr19 | 44,921,921 | chr19:44921921:G:A | a | g | 0.6492 | 0.1214 | 8.84E-08 | +-+++++-+? | 11.554 | 8 | 0.1722 |
| ADNC Score | chr19 | 44,922,203 | chr19:44922203:A:G | a | g | 0.2621 | 0.0503 | 1.88E-07 | -+++++--++ | 16.827 | 9 | 0.05149 |
| ADNC Score | chr19 | 44,923,535 | chr19:44923535:G:A | a | g | 0.6341 | 0.1199 | 1.23E-07 | +-+++++-+- | 11.547 | 9 | 0.2401 |
| ADNC Score | chr19 | 44,923,868 | chr19:44923868:T:A | a | t | -0.8131 | 0.0647 | 3.09E-36 | +--------+ | 10.42 | 9 | 0.3175 |
| ADNC Score | chr19 | 44,924,096 | chr19:44924096:C:G | c | g | -0.3427 | 0.0519 | 4.16E-11 | +--------+ | 6.424 | 9 | 0.6968 |
| ADNC Score | chr19 | 44,924,977 | chr19:44924977:G:A | a | g | -0.8088 | 0.0647 | 7.25E-36 | +--------- | 9.016 | 9 | 0.4358 |
| ADNC Score | chr19 | 44,925,202 | chr19:44925202:C:T | t | c | 0.3432 | 0.0536 | 1.48E-10 | ++++-++++- | 6.488 | 9 | 0.6903 |
| ADNC Score | chr19 | 44,926,286 | chr19:44926286:C:T | t | c | 0.2423 | 0.0553 | 1.20E-05 | +++++++++ | 5.256 | 9 | 0.8115 |
| ADNC Score | chr19 | 44,926,451 | chr19:44926451:G:C | c | g | -0.4113 | 0.0843 | 1.08E-06 | +--------+ | 8.348 | 9 | 0.4995 |
| ADNC Score | chr19 | 44,935,297 | chr19:44935297:C:T | t | c | -0.2666 | 0.0553 | 1.44E-06 | -------+-+ | 10.925 | 9 | 0.2809 |
| ADNC Score | chr19 | 45,204,659 | chr19:45204659:T:G | t | g | 0.263 | 0.0659 | 6.52E-05 | +++++++++ | 9.996 | 9 | 0.3508 |
| ADNC Score | chr19 | 45,204,981 | chr19:45204981:A:G | a | g | 0.2632 | 0.0659 | 6.49E-05 | +++++++++ | 9.958 | 9 | 0.3539 |
| ADNC Score | chr19 | 45,205,150 | chr19:45205150:T:C | t | c | 0.2633 | 0.0659 | 6.45E-05 | +++++++++ | 9.952 | 9 | 0.3544 |
| ADNC Score | chr19 | 45,205,166 | chr19:45205166:TTG:T | t | ttg | -0.2635 | 0.0659 | 6.40E-05 | ---------+ | 9.945 | 9 | 0.3549 |
| ADNC Score | chr19 | 45,205,500 | chr19:45205500:A:G | a | g | 0.2678 | 0.0656 | 4.51E-05 | +++++++++ | 9.007 | 9 | 0.4366 |
| ADNC Score | chr19 | 45,205,630 | chr19:45205630:T:C | t | c | 0.2689 | 0.066 | 4.64E-05 | +++++++++ | 9.002 | 9 | 0.4371 |
| ADNC Score | chr19 | 45,205,689 | chr19:45205689:G:A | a | g | -0.2701 | 0.0662 | 4.47E-05 | ---------+ | 9.553 | 9 | 0.3879 |
| ADNC Score | chr19 | 45,205,778 | chr19:45205778:C:T | t | c | -0.2701 | 0.0662 | 4.46E-05 | ---------+ | 9.552 | 9 | 0.388 |
| ADNC Score | chr19 | 45,206,729 | chr19:45206729:A:G | a | g | 0.27 | 0.0662 | 4.48E-05 | +++++++++ | 9.557 | 9 | 0.3876 |
| ADNC Score | chr19 | 45,207,018 | chr19:45207018:A:G | a | g | 0.2708 | 0.0661 | 4.24E-05 | +++++++++ | 9.531 | 9 | 0.3897 |

| | | | | | | | | | | | | |
|---|---|---|---|---|---|---|---|---|---|---|---|---|
| ADNC Score | chr19 | 45,207,281 | chr19:45207281:C:T | t | c | -0.2773 | 0.0664 | 2.96E-05 | ---------+ | 9.505 | 9 | 0.392 |
| ADNC Score | chr19 | 45,207,605 | chr19:45207605:C:T | t | c | -0.2688 | 0.0662 | 4.84E-05 | ---------+ | 9.644 | 9 | 0.3801 |
| ADNC Score | chr19 | 45,208,340 | chr19:45208340:C:T | t | c | -0.2705 | 0.0658 | 3.96E-05 | ---------- | 8.174 | 9 | 0.5167 |
| ADNC Score | chr2 | 319,464 | chr2:319464:C:T | t | c | 0.4495 | 0.1122 | 6.16E-05 | ++++--+++- | 10.73 | 9 | 0.2947 |
| ADNC Score | chr2 | 3,944,159 | chr2:3944159:C:T | t | c | 0.7711 | 0.1981 | 9.94E-05 | +++--++-+- | 10.618 | 9 | 0.3028 |
| ADNC Score | chr2 | 18,342,027 | chr2:18342027:TA:T | t | ta | 0.5313 | 0.133 | 6.47E-05 | +-+++++++- | 8.887 | 9 | 0.4478 |
| ADNC Score | chr2 | 24,787,549 | chr2:24787549:G:A | a | g | 0.2298 | 0.0577 | 6.79E-05 | -++++++-+- | 16.98 | 9 | 0.04904 |
| ADNC Score | chr2 | 24,798,168 | chr2:24798168:G:C | c | g | 0.221 | 0.0524 | 2.43E-05 | -++++++-+- | 19.555 | 9 | 0.02086 |
| ADNC Score | chr2 | 24,800,573 | chr2:24800573:G:A | a | g | 0.2396 | 0.0574 | 3.02E-05 | -++++++-+- | 18.317 | 9 | 0.03167 |
| ADNC Score | chr2 | 24,800,575 | chr2:24800575:T:C | t | c | -0.2391 | 0.0574 | 3.14E-05 | +------+-+ | 18.224 | 9 | 0.03266 |
| ADNC Score | chr2 | 24,804,442 | chr2:24804442:C:T | t | c | 0.2396 | 0.0573 | 2.91E-05 | -++++++-+- | 17.902 | 9 | 0.03632 |
| ADNC Score | chr2 | 24,811,460 | chr2:24811460:T:C | t | c | -0.2169 | 0.0523 | 3.43E-05 | +------+-+ | 19.238 | 9 | 0.02324 |
| ADNC Score | chr2 | 24,812,510 | chr2:24812510:C:G | c | g | -0.233 | 0.057 | 4.31E-05 | +------+-+ | 20.918 | 9 | 0.01302 |
| ADNC Score | chr2 | 24,813,989 | chr2:24813989:C:A | a | c | 0.2345 | 0.057 | 3.88E-05 | -++++++-+- | 18.721 | 9 | 0.02767 |
| ADNC Score | chr2 | 24,814,757 | chr2:24814757:G:T | t | g | 0.2344 | 0.0572 | 4.13E-05 | -++++++-+- | 18.608 | 9 | 0.02874 |
| ADNC Score | chr2 | 24,817,213 | chr2:24817213:C:T | t | c | 0.2331 | 0.0567 | 3.93E-05 | -++++++-+- | 20.762 | 9 | 0.01375 |
| ADNC Score | chr2 | 24,819,753 | chr2:24819753:A:G | a | g | -0.2367 | 0.0573 | 3.57E-05 | +------+-+ | 19.404 | 9 | 0.02197 |
| ADNC Score | chr2 | 24,832,761 | chr2:24832761:C:T | t | c | 0.2107 | 0.052 | 5.14E-05 | -++++++-+- | 19.563 | 9 | 0.02081 |
| ADNC Score | chr2 | 36,708,148 | chr2:36708148:G:A | a | g | -0.2786 | 0.0709 | 8.50E-05 | ---------- | 3.492 | 9 | 0.9416 |
| ADNC Score | chr2 | 78,825,371 | chr2:78825371:G:T | t | g | 0.9957 | 0.2367 | 2.60E-05 | +++++++++ | 4.319 | 8 | 0.8272 |
| ADNC Score | chr2 | 82,015,880 | chr2:82015880:G:T | t | g | 0.3392 | 0.0861 | 8.18E-05 | -+++++-+++ | 10.71 | 9 | 0.2961 |
| ADNC Score | chr2 | 82,077,015 | chr2:82077015:C:T | t | c | 0.2233 | 0.0573 | 9.66E-05 | ++++++-++- | 13.978 | 9 | 0.1231 |
| ADNC Score | chr2 | 82,088,055 | chr2:82088055:G:A | a | g | 0.2249 | 0.0569 | 7.75E-05 | ++++++-++- | 13.999 | 9 | 0.1224 |
| ADNC Score | chr2 | 82,117,165 | chr2:82117165:A:G | a | g | -0.2246 | 0.0572 | 8.57E-05 | ------+--+ | 14.175 | 9 | 0.1162 |
| ADNC Score | chr2 | 98,742,324 | chr2:98742324:G:A | a | g | 1.5087 | 0.3331 | 5.93E-06 | ?++?+????- | 1.365 | 3 | 0.7139 |
| ADNC Score | chr2 | 100,686,901 | chr2:100686901:T:A | a | t | 0.5273 | 0.1327 | 7.12E-05 | --++-+++++ | 7.846 | 9 | 0.5497 |
| ADNC Score | chr2 | 101,718,396 | chr2:101718396:T:C | t | c | -0.6004 | 0.142 | 2.35E-05 | +---?-++-- | 13.285 | 8 | 0.1024 |
| ADNC Score | chr2 | 102,016,636 | chr2:102016636:C:T | t | c | 0.7438 | 0.1853 | 5.94E-05 | -+++?+--++ | 13.13 | 8 | 0.1075 |
| ADNC Score | chr2 | 109,948,161 | chr2:109948161:G:A | a | g | 0.8344 | 0.2083 | 6.19E-05 | -+++++++?- | 4.905 | 8 | 0.7677 |
| ADNC Score | chr2 | 120,604,405 | chr2:120604405:G:A | a | g | 0.7828 | 0.1907 | 4.03E-05 | +++++++++ | 3.934 | 8 | 0.863 |
| ADNC Score | chr2 | 127,133,851 | chr2:127133851:A:C | a | c | 0.2263 | 0.0484 | 2.91E-06 | +++++++++ | 8.97 | 9 | 0.44 |
| ADNC Score | chr2 | 127,135,234 | chr2:127135234:C:T | t | c | -0.227 | 0.0487 | 3.15E-06 | ---------- | 9.08 | 9 | 0.43 |
| ADNC Score | chr2 | 127,137,039 | chr2:127137039:A:G | a | g | 0.2068 | 0.0521 | 7.13E-05 | -+++-+++++ | 9.718 | 9 | 0.3738 |
| ADNC Score | chr2 | 131,994,185 | chr2:131994185:A:G | a | g | 0.2181 | 0.0548 | 6.86E-05 | +-+++++++- | 7.517 | 9 | 0.5834 |
| ADNC Score | chr2 | 169,278,693 | chr2:169278693:G:A | a | g | -0.4223 | 0.1045 | 5.28E-05 | -+-------+ | 15.889 | 9 | 0.06923 |
| ADNC Score | chr2 | 181,119,900 | chr2:181119900:G:C | c | g | -0.201 | 0.0513 | 8.98E-05 | -------+-- | 4.01 | 9 | 0.9107 |
| ADNC Score | chr2 | 190,702,499 | chr2:190702499:T:C | t | c | 0.5243 | 0.1299 | 5.45E-05 | +++++++++ | 4.715 | 9 | 0.8584 |
| ADNC Score | chr2 | 190,702,500 | chr2:190702500:G:C | c | g | -0.526 | 0.1299 | 5.16E-05 | ---------+ | 4.651 | 9 | 0.8636 |
| ADNC Score | chr2 | 190,744,258 | chr2:190744258:T:A | a | t | -0.5152 | 0.1284 | 6.03E-05 | ---------+ | 3.162 | 9 | 0.9575 |
| ADNC Score | chr2 | 199,671,345 | chr2:199671345:C:T | t | c | -0.4895 | 0.1172 | 2.97E-05 | -+-------- | 6.31 | 9 | 0.7086 |
| ADNC Score | chr2 | 205,687,520 | chr2:205687520:G:A | a | g | -0.2265 | 0.0577 | 8.77E-05 | +---+----+ | 7.699 | 9 | 0.5648 |
| ADNC Score | chr2 | 205,717,138 | chr2:205717138:A:AGTGT | a | agtgt | 0.1949 | 0.0488 | 6.60E-05 | ++++-++++- | 5.69 | 9 | 0.7705 |
| ADNC Score | chr2 | 205,728,517 | chr2:205728517:T:C | t | c | -0.1848 | 0.0473 | 9.34E-05 | ---------+ | 6.171 | 9 | 0.7227 |
| ADNC Score | chr2 | 205,728,996 | chr2:205728996:A:G | a | g | 0.2039 | 0.0516 | 7.76E-05 | +++++++-+- | 8.303 | 9 | 0.5039 |
| ADNC Score | chr2 | 208,668,013 | chr2:208668013:GC:G | g | gc | 0.9713 | 0.2077 | 2.90E-06 | +-++?++++? | 5.365 | 7 | 0.6155 |
| ADNC Score | chr2 | 208,746,279 | chr2:208746279:T:C | t | c | -0.8696 | 0.209 | 3.18E-05 | -+--?+---? | 7.794 | 7 | 0.3511 |

| | | | | | | | | | | | | |
|---|---|---|---|---|---|---|---|---|---|---|---|---|
| ADNC Score | chr2 | 208,795,449 | chr2:208795449:T:C | t | c | -0.8925 | 0.2101 | 2.17E-05 | -+--?+---? | 8.472 | 7 | 0.2928 |
| ADNC Score | chr2 | 208,833,781 | chr2:208833781:G:C | c | g | 0.8912 | 0.2031 | 1.14E-05 | +-+++-+++- | 9.872 | 9 | 0.3609 |
| ADNC Score | chr2 | 228,367,334 | chr2:228367334:T:C | t | c | -0.2098 | 0.053 | 7.54E-05 | ---------+ | 5.632 | 9 | 0.7761 |
| ADNC Score | chr20 | 32,163,490 | chr20:32163490:T:TA | t | ta | -0.3993 | 0.0897 | 8.58E-06 | ----+----- | 1.588 | 9 | 0.9964 |
| ADNC Score | chr20 | 32,242,188 | chr20:32242188:C:T | t | c | 0.3895 | 0.0888 | 1.16E-05 | -+++-+++++ | 1.749 | 9 | 0.9948 |
| ADNC Score | chr20 | 32,276,738 | chr20:32276738:TTTTA:T | t | tttta | 0.2395 | 0.0602 | 6.93E-05 | ++++-++++- | 4.058 | 9 | 0.9076 |
| ADNC Score | chr20 | 32,316,974 | chr20:32316974:G:A | a | g | 0.234 | 0.06 | 9.65E-05 | ++++-++++- | 4.122 | 9 | 0.9032 |
| ADNC Score | chr20 | 32,331,380 | chr20:32331380:A:G | a | g | -0.2362 | 0.0599 | 7.94E-05 | ----+----- | 4.275 | 9 | 0.8924 |
| ADNC Score | chr20 | 32,335,231 | chr20:32335231:G:A | a | g | 0.2351 | 0.0599 | 8.74E-05 | ++++-++++- | 4.3 | 9 | 0.8906 |
| ADNC Score | chr20 | 32,364,378 | chr20:32364378:T:G | t | g | 0.2377 | 0.0605 | 8.44E-05 | ++++-++++- | 3.988 | 9 | 0.9122 |
| ADNC Score | chr20 | 32,372,684 | chr20:32372684:G:T | t | g | -0.2424 | 0.0604 | 5.98E-05 | ----+----- | 4.29 | 9 | 0.8913 |
| ADNC Score | chr20 | 32,379,841 | chr20:32379841:TA:T | t | ta | -0.2413 | 0.0612 | 8.09E-05 | ----+----- | 3.322 | 9 | 0.9502 |
| ADNC Score | chr20 | 32,388,229 | chr20:32388229:C:T | t | c | 0.4228 | 0.0904 | 2.92E-06 | -++++++++- | 2.666 | 9 | 0.9761 |
| ADNC Score | chr20 | 32,396,753 | chr20:32396753:A:G | a | g | 0.2393 | 0.0606 | 7.79E-05 | ++++-++++- | 4.146 | 9 | 0.9015 |
| ADNC Score | chr20 | 32,410,413 | chr20:32410413:C:T | t | c | -0.2402 | 0.0613 | 8.97E-05 | ----+----- | 3.035 | 9 | 0.9629 |
| ADNC Score | chr20 | 32,419,704 | chr20:32419704:A:G | a | g | 0.239 | 0.0614 | 9.86E-05 | ++++-++++- | 3.21 | 9 | 0.9554 |
| ADNC Score | chr20 | 32,421,237 | chr20:32421237:T:A | a | t | -0.2475 | 0.0615 | 5.63E-05 | ----+----- | 3.276 | 9 | 0.9523 |
| ADNC Score | chr20 | 32,431,221 | chr20:32431221:C:T | t | c | -0.2431 | 0.0612 | 7.16E-05 | ----+----- | 3.151 | 9 | 0.958 |
| ADNC Score | chr20 | 32,441,200 | chr20:32441200:G:C | c | g | 0.2448 | 0.0602 | 4.80E-05 | ++++-++++- | 4.321 | 9 | 0.8891 |
| ADNC Score | chr20 | 32,447,326 | chr20:32447326:A:G | a | g | -0.2513 | 0.0609 | 3.66E-05 | ----+----- | 3.821 | 9 | 0.9228 |
| ADNC Score | chr20 | 32,447,923 | chr20:32447923:A:AGTT | a | agtt | -0.2482 | 0.061 | 4.78E-05 | ----+----- | 3.615 | 9 | 0.9349 |
| ADNC Score | chr20 | 32,450,354 | chr20:32450354:T:C | t | c | -0.2562 | 0.0602 | 2.11E-05 | ----+----- | 4.661 | 9 | 0.8628 |
| ADNC Score | chr20 | 55,907,748 | chr20:55907748:A:T | a | t | 1.0438 | 0.2598 | 5.87E-05 | +-+++++++ | 8.879 | 8 | 0.3526 |
| ADNC Score | chr21 | 16,790,091 | chr21:16790091:G:A | a | g | -0.8021 | 0.1974 | 4.83E-05 | --------- | 3.703 | 9 | 0.9298 |
| ADNC Score | chr21 | 18,110,961 | chr21:18110961:T:C | t | c | -0.2601 | 0.0629 | 3.54E-05 | --------- | 2.833 | 9 | 0.9706 |
| ADNC Score | chr21 | 18,111,087 | chr21:18111087:A:G | a | g | -0.2563 | 0.0626 | 4.25E-05 | --------- | 2.918 | 9 | 0.9675 |
| ADNC Score | chr21 | 18,111,950 | chr21:18111950:C:T | t | c | 0.2503 | 0.0607 | 3.75E-05 | +++++++++ | 5.166 | 9 | 0.8196 |
| ADNC Score | chr21 | 18,112,772 | chr21:18112772:C:G | c | g | -0.2546 | 0.0626 | 4.77E-05 | --------- | 2.888 | 9 | 0.9686 |
| ADNC Score | chr21 | 18,115,098 | chr21:18115098:T:A | a | t | 0.261 | 0.063 | 3.42E-05 | +++++++++ | 3.406 | 9 | 0.946 |
| ADNC Score | chr21 | 44,795,325 | chr21:44795325:T:C | t | c | -1.4079 | 0.3351 | 2.65E-05 | ?---??-??? | 2.5 | 3 | 0.4752 |
| ADNC Score | chr22 | 17,850,701 | chr22:17850701:C:T | t | c | 0.4343 | 0.1082 | 5.94E-05 | +++++++++ | 16.003 | 9 | 0.06681 |
| ADNC Score | chr22 | 21,023,045 | chr22:21023045:C:T | t | c | 0.368 | 0.0922 | 6.58E-05 | +-++-+-+++ | 16.394 | 9 | 0.0591 |
| ADNC Score | chr22 | 21,025,775 | chr22:21025775:G:A | a | g | 0.3192 | 0.078 | 4.29E-05 | --++-+-+++ | 10.385 | 9 | 0.3203 |
| ADNC Score | chr22 | 21,028,193 | chr22:21028193:T:C | t | c | -0.3053 | 0.0779 | 8.82E-05 | ++--+-+--- | 9.605 | 9 | 0.3834 |
| ADNC Score | chr22 | 21,028,993 | chr22:21028993:T:C | t | c | -0.3056 | 0.0779 | 8.73E-05 | ++--+-+--- | 9.668 | 9 | 0.378 |
| ADNC Score | chr22 | 21,029,023 | chr22:21029023:G:A | a | g | 0.312 | 0.0782 | 6.57E-05 | --++-+-+++ | 10.237 | 9 | 0.3317 |
| ADNC Score | chr22 | 21,030,289 | chr22:21030289:C:T | t | c | 0.3009 | 0.0769 | 9.09E-05 | --++-+-+++ | 9.648 | 9 | 0.3797 |
| ADNC Score | chr22 | 21,032,188 | chr22:21032188:C:T | t | c | 0.3056 | 0.0781 | 9.14E-05 | --++-+-+++ | 9.674 | 9 | 0.3775 |
| ADNC Score | chr22 | 49,272,438 | chr22:49272438:C:T | t | c | 0.5626 | 0.1283 | 1.15E-05 | +-+++++++- | 10.222 | 9 | 0.3328 |
| ADNC Score | chr22 | 49,274,046 | chr22:49274046:C:T | t | c | 0.486 | 0.1136 | 1.87E-05 | +++++++++ | 7.402 | 9 | 0.5953 |
| ADNC Score | chr22 | 49,275,389 | chr22:49275389:A:G | a | g | -0.4847 | 0.1136 | 1.97E-05 | --------- | 7.483 | 9 | 0.587 |
| ADNC Score | chr22 | 49,275,671 | chr22:49275671:G:A | a | g | 0.486 | 0.1135 | 1.84E-05 | +++++++++ | 7.463 | 9 | 0.5891 |
| ADNC Score | chr22 | 49,276,245 | chr22:49276245:T:G | t | g | -0.4842 | 0.1136 | 2.01E-05 | --------- | 7.478 | 9 | 0.5875 |
| ADNC Score | chr3 | 14,742,741 | chr3:14742741:T:C | t | c | -0.7813 | 0.186 | 2.66E-05 | -+-------+ | 9.295 | 9 | 0.4105 |
| ADNC Score | chr3 | 33,962,116 | chr3:33962116:A:G | a | g | -0.6004 | 0.1397 | 1.73E-05 | ----?----- | 4.149 | 8 | 0.8434 |
| ADNC Score | chr3 | 38,783,660 | chr3:38783660:A:G | a | g | -0.8641 | 0.1832 | 2.38E-06 | +--------? | 5.805 | 8 | 0.669 |

| | | | | | | | | | | | | | |
|---|---|---|---|---|---|---|---|---|---|---|---|---|---|
| ADNC Score | chr3 | 62,071,217 | chr3:62071217:G:A | a | g | -0.2079 | 0.0498 | 2.94E-05 | -----+---+ | 15.713 |  | 9 | 0.07313 |
| ADNC Score | chr3 | 102,478,610 | chr3:102478610:C:T | t | c | -0.3037 | 0.0728 | 3.03E-05 | -+-----+-- | 7.018 |  | 9 | 0.6352 |
| ADNC Score | chr3 | 102,519,261 | chr3:102519261:G:A | a | g | -0.2771 | 0.0699 | 7.38E-05 | -+-----+-- | 10.809 |  | 9 | 0.289 |
| ADNC Score | chr3 | 102,545,468 | chr3:102545468:A:G | a | g | 0.2761 | 0.0706 | 9.23E-05 | +-+++++-++ | 11.224 | | 9 | 0.2607 |
| ADNC Score | chr3 | 102,548,552 | chr3:102548552:C:A | a | c | -0.2749 | 0.0701 | 8.92E-05 | -+-----+-- | 11.423 |  | 9 | 0.2478 |
| ADNC Score | chr3 | 102,549,634 | chr3:102549634:T:A | a | t | -0.2763 | 0.0706 | 9.17E-05 | -+-----+-- | 11.217 |  | 9 | 0.2612 |
| ADNC Score | chr3 | 104,191,920 | chr3:104191920:A:G | a | g | -0.2041 | 0.0479 | 2.07E-05 | --------- | 3.646 |  | 9 | 0.9331 |
| ADNC Score | chr3 | 104,192,219 | chr3:104192219:A:C | a | c | -0.2224 | 0.0533 | 3.06E-05 | ---------+ | 4.215 |  | 9 | 0.8967 |
| ADNC Score | chr3 | 129,082,494 | chr3:129082494:T:G | t | g | 0.2292 | 0.0587 | 9.43E-05 | +++++++-+- | 11.204 | | 9 | 0.262 |
| ADNC Score | chr3 | 129,243,779 | chr3:129243779:C:T | t | c | 0.4994 | 0.1162 | 1.73E-05 | +++++++++ | 3.092 | | 9 | 0.9605 |
| ADNC Score | chr3 | 144,999,676 | chr3:144999676:A:G | a | g | -0.7683 | 0.1726 | 8.54E-06 | -------+-? | 6.183 |  | 8 | 0.6267 |
| ADNC Score | chr3 | 193,576,295 | chr3:193576295:C:T | t | c | 0.3104 | 0.0791 | 8.66E-05 | -+++-+--+- | 8.767 | | 9 | 0.4591 |
| ADNC Score | chr3 | 193,596,628 | chr3:193596628:A:G | a | g | -0.248 | 0.062 | 6.28E-05 | ---++--+-+ | 14.8 | | 9 | 0.09658 |
| ADNC Score | chr3 | 193,598,916 | chr3:193598916:C:T | t | c | 0.2476 | 0.0619 | 6.35E-05 | +++--++-+- | 14.776 | | 9 | 0.09727 |
| ADNC Score | chr3 | 193,618,433 | chr3:193618433:G:A | a | g | 0.2505 | 0.062 | 5.25E-05 | ++++-++-+- | 14.488 | | 9 | 0.106 |
| ADNC Score | chr3 | 193,646,065 | chr3:193646065:A:G | a | g | -0.2478 | 0.0618 | 6.17E-05 | ----+-++-+ | 13.844 |  | 9 | 0.128 |
| ADNC Score | chr3 | 193,660,764 | chr3:193660764:G:A | a | g | 0.2525 | 0.062 | 4.60E-05 | ++++-+--+- | 14.657 | | 9 | 0.1008 |
| ADNC Score | chr3 | 193,668,673 | chr3:193668673:C:T | t | c | 0.2522 | 0.0619 | 4.68E-05 | ++++-+--+- | 14.642 | | 9 | 0.1013 |
| ADNC Score | chr3 | 193,671,674 | chr3:193671674:G:A | a | g | 0.2542 | 0.0621 | 4.22E-05 | ++++-+--+- | 15.327 | | 9 | 0.08235 |
| ADNC Score | chr3 | 193,676,922 | chr3:193676922:G:A | a | g | 0.2463 | 0.0628 | 8.72E-05 | ++++-+--+- | 14.247 | | 9 | 0.1138 |
| ADNC Score | chr4 | 6,411,041 | chr4:6411041:G:T | t | g | 0.2456 | 0.054 | 5.32E-06 | +++++++++ | 3.682 | | 9 | 0.9311 |
| ADNC Score | chr4 | 6,411,044 | chr4:6411044:T:C | t | c | -0.2586 | 0.0564 | 4.61E-06 | ---------+ | 5.18 |  | 9 | 0.8184 |
| ADNC Score | chr4 | 6,411,820 | chr4:6411820:A:G | a | g | 0.2237 | 0.0574 | 9.83E-05 | +++++++++ | 9.927 | | 9 | 0.3564 |
| ADNC Score | chr4 | 6,412,734 | chr4:6412734:G:A | a | g | -0.2417 | 0.055 | 1.10E-05 | ---------+ | 8.582 |  | 9 | 0.4767 |
| ADNC Score | chr4 | 6,412,914 | chr4:6412914:C:G | c | g | 0.2439 | 0.055 | 9.25E-06 | +++++++++ | 9 | | 9 | 0.4373 |
| ADNC Score | chr4 | 6,413,261 | chr4:6413261:GCACA:G | g | gcaca | 0.2395 | 0.061 | 8.60E-05 | +++++++++ | 7.287 | | 9 | 0.6073 |
| ADNC Score | chr4 | 6,414,301 | chr4:6414301:G:A | a | g | -0.2444 | 0.0546 | 7.53E-06 | ---------+ | 9.011 |  | 9 | 0.4362 |
| ADNC Score | chr4 | 6,415,835 | chr4:6415835:G:A | a | g | 0.2219 | 0.054 | 3.97E-05 | +++++++++ | 8.799 | | 9 | 0.456 |
| ADNC Score | chr4 | 6,418,277 | chr4:6418277:C:A | a | c | 0.2157 | 0.0541 | 6.76E-05 | +++++++++ | 8.107 | | 9 | 0.5234 |
| ADNC Score | chr4 | 20,416,685 | chr4:20416685:C:T | t | c | 0.3718 | 0.0948 | 8.84E-05 | +-++++++-+ | 12.849 | | 9 | 0.1696 |
| ADNC Score | chr4 | 40,705,804 | chr4:40705804:C:G | c | g | -0.2336 | 0.0517 | 6.30E-06 | +--------- | 4.835 |  | 9 | 0.8484 |
| ADNC Score | chr4 | 40,706,660 | chr4:40706660:C:A | a | c | 0.2385 | 0.0519 | 4.34E-06 | -++++++++- | 5.108 | | 9 | 0.8248 |
| ADNC Score | chr4 | 40,708,079 | chr4:40708079:T:C | t | c | -0.2393 | 0.0518 | 3.82E-06 | +--------- | 5.483 |  | 9 | 0.7903 |
| ADNC Score | chr4 | 40,710,327 | chr4:40710327:A:G | a | g | -0.2405 | 0.0517 | 3.26E-06 | ++-------- | 5.484 |  | 9 | 0.7903 |
| ADNC Score | chr4 | 40,710,577 | chr4:40710577:T:C | t | c | -0.2424 | 0.0518 | 2.91E-06 | +--------- | 5.427 |  | 9 | 0.7956 |
| ADNC Score | chr4 | 40,710,873 | chr4:40710873:G:A | a | g | 0.2417 | 0.0519 | 3.16E-06 | -++++++++- | 5.509 | | 9 | 0.7878 |
| ADNC Score | chr4 | 40,711,238 | chr4:40711238:A:T | a | t | -0.2387 | 0.0517 | 3.86E-06 | +--------- | 4.841 |  | 9 | 0.8479 |
| ADNC Score | chr4 | 40,711,817 | chr4:40711817:A:G | a | g | -0.2418 | 0.0517 | 2.95E-06 | +--------- | 5.505 |  | 9 | 0.7883 |
| ADNC Score | chr4 | 40,712,046 | chr4:40712046:A:G | a | g | -0.2429 | 0.0517 | 2.66E-06 | +--------- | 5.469 |  | 9 | 0.7917 |
| ADNC Score | chr4 | 40,712,101 | chr4:40712101:A:G | a | g | -0.237 | 0.0518 | 4.69E-06 | +--------- | 5.459 |  | 9 | 0.7926 |
| ADNC Score | chr4 | 40,713,166 | chr4:40713166:C:G | c | g | -0.2424 | 0.0518 | 2.94E-06 | +--------- | 5.428 |  | 9 | 0.7955 |
| ADNC Score | chr4 | 40,713,497 | chr4:40713497:C:T | t | c | 0.2384 | 0.0519 | 4.29E-06 | -++++++++- | 5.52 | | 9 | 0.7868 |
| ADNC Score | chr4 | 40,713,839 | chr4:40713839:G:A | a | g | 0.2384 | 0.0517 | 4.11E-06 | -++++++++- | 5.465 | | 9 | 0.792 |
| ADNC Score | chr4 | 40,713,843 | chr4:40713843:C:T | t | c | 0.2425 | 0.0517 | 2.75E-06 | -++++++++- | 5.521 | | 9 | 0.7867 |
| ADNC Score | chr4 | 40,714,722 | chr4:40714722:G:A | a | g | 0.2377 | 0.0517 | 4.24E-06 | -++++++++- | 5.339 | | 9 | 0.8038 |
| ADNC Score | chr4 | 40,715,412 | chr4:40715412:T:C | t | c | -0.2316 | 0.0523 | 9.62E-06 | ++-------- | 5.09 |  | 9 | 0.8264 |

| | | | | | | | | | | | | |
|---|---|---|---|---|---|---|---|---|---|---|---|---|
| ADNC Score | chr4 | 40,715,434 | chr4:40715434:CA:C | ca | c | -0.2256 | 0.0523 | 1.61E-05 | ++-------- | 5.047 | 9 | 0.8302 |
| ADNC Score | chr4 | 40,715,712 | chr4:40715712:T:C | t | c | -0.2302 | 0.0524 | 1.12E-05 | ++-------- | 4.871 | 9 | 0.8454 |
| ADNC Score | chr4 | 40,715,740 | chr4:40715740:T:C | t | c | -0.2281 | 0.0525 | 1.38E-05 | ++-------- | 5.064 | 9 | 0.8287 |
| ADNC Score | chr4 | 40,715,866 | chr4:40715866:C:T | t | c | 0.2308 | 0.0524 | 1.06E-05 | --++++++++ | 4.87 | 9 | 0.8455 |
| ADNC Score | chr4 | 40,716,011 | chr4:40716011:G:A | a | g | 0.213 | 0.0505 | 2.47E-05 | --++++++++ | 7.47 | 9 | 0.5883 |
| ADNC Score | chr4 | 40,716,839 | chr4:40716839:T:C | t | c | -0.2278 | 0.0526 | 1.48E-05 | ++-------- | 5.557 | 9 | 0.7833 |
| ADNC Score | chr4 | 54,714,836 | chr4:54714836:C:T | t | c | 0.7128 | 0.1783 | 6.39E-05 | --+++++++? | 7.944 | 8 | 0.4389 |
| ADNC Score | chr4 | 57,820,798 | chr4:57820798:G:A | a | g | 1.459 | 0.3737 | 9.44E-05 | +-?????++? | 3.234 | 3 | 0.3569 |
| ADNC Score | chr4 | 70,032,918 | chr4:70032918:T:A | a | t | 0.5541 | 0.1398 | 7.43E-05 | +++++++++ | 7.444 | 9 | 0.5909 |
| ADNC Score | chr4 | 70,033,486 | chr4:70033486:C:T | t | c | 0.5777 | 0.1385 | 3.01E-05 | +++++++++ | 8.211 | 9 | 0.5131 |
| ADNC Score | chr4 | 91,330,688 | chr4:91330688:C:T | t | c | 0.2028 | 0.0512 | 7.38E-05 | +++++++++ | 3.423 | 9 | 0.9452 |
| ADNC Score | chr4 | 91,340,525 | chr4:91340525:G:T | t | g | 0.2089 | 0.0513 | 4.66E-05 | +++++++++ | 3.668 | 9 | 0.9319 |
| ADNC Score | chr4 | 91,342,961 | chr4:91342961:T:C | t | c | -0.2121 | 0.0513 | 3.59E-05 | ---------+ | 3.345 | 9 | 0.949 |
| ADNC Score | chr4 | 91,344,469 | chr4:91344469:G:C | c | g | 0.2146 | 0.0515 | 3.05E-05 | +++++++++ | 3.459 | 9 | 0.9433 |
| ADNC Score | chr4 | 91,346,391 | chr4:91346391:G:A | a | g | 0.2169 | 0.0514 | 2.48E-05 | +++++++++ | 3.632 | 9 | 0.9339 |
| ADNC Score | chr4 | 91,349,429 | chr4:91349429:GT:G | g | gt | 0.221 | 0.0514 | 1.71E-05 | +++++++++ | 3.748 | 9 | 0.9272 |
| ADNC Score | chr4 | 91,349,559 | chr4:91349559:T:C | t | c | -0.2063 | 0.0516 | 6.34E-05 | ---------+ | 4.188 | 9 | 0.8986 |
| ADNC Score | chr4 | 109,304,500 | chr4:109304500:A:ATCTTC | a | atcttc | 0.2534 | 0.0641 | 7.65E-05 | ++++++-++- | 9.131 | 9 | 0.4252 |
| ADNC Score | chr4 | 109,305,702 | chr4:109305702:G:C | c | g | -0.2547 | 0.0629 | 5.16E-05 | ------+--+ | 7.926 | 9 | 0.5416 |
| ADNC Score | chr4 | 109,306,357 | chr4:109306357:C:T | t | c | -0.2571 | 0.063 | 4.46E-05 | ------+--+ | 7.715 | 9 | 0.5631 |
| ADNC Score | chr4 | 109,309,554 | chr4:109309554:A:T | a | t | 0.2511 | 0.0627 | 6.12E-05 | ++++++-++- | 8.102 | 9 | 0.5239 |
| ADNC Score | chr4 | 109,310,934 | chr4:109310934:G:T | t | g | -0.2549 | 0.0627 | 4.78E-05 | ------+--+ | 7.937 | 9 | 0.5406 |
| ADNC Score | chr4 | 109,311,775 | chr4:109311775:A:T | a | t | 0.2549 | 0.0627 | 4.78E-05 | ++++++-++- | 7.936 | 9 | 0.5406 |
| ADNC Score | chr4 | 109,312,269 | chr4:109312269:G:A | a | g | -0.2574 | 0.0627 | 4.02E-05 | ------+--+ | 7.457 | 9 | 0.5896 |
| ADNC Score | chr4 | 109,313,108 | chr4:109313108:G:A | a | g | -0.2546 | 0.0627 | 4.84E-05 | ------+--+ | 7.969 | 9 | 0.5373 |
| ADNC Score | chr4 | 109,314,951 | chr4:109314951:T:C | t | c | 0.2548 | 0.0627 | 4.79E-05 | ++++++-++- | 8.362 | 9 | 0.4982 |
| ADNC Score | chr4 | 109,315,167 | chr4:109315167:C:T | t | c | -0.2999 | 0.0754 | 7.03E-05 | ------++-- | 9.524 | 9 | 0.3904 |
| ADNC Score | chr4 | 109,316,413 | chr4:109316413:A:AT | a | at | 0.2861 | 0.0727 | 8.24E-05 | ++++++--++ | 9.012 | 9 | 0.4362 |
| ADNC Score | chr4 | 109,320,388 | chr4:109320388:C:T | t | c | -0.2875 | 0.0725 | 7.27E-05 | ------++-- | 9.035 | 9 | 0.434 |
| ADNC Score | chr4 | 109,320,677 | chr4:109320677:A:G | a | g | 0.2874 | 0.0725 | 7.31E-05 | ++++++--++ | 9.048 | 9 | 0.4329 |
| ADNC Score | chr4 | 109,321,458 | chr4:109321458:T:G | t | g | 0.2873 | 0.0725 | 7.36E-05 | ++++++--++ | 9.046 | 9 | 0.4331 |
| ADNC Score | chr4 | 109,322,078 | chr4:109322078:A:G | a | g | 0.2553 | 0.0627 | 4.59E-05 | ++++++-++- | 8.378 | 9 | 0.4966 |
| ADNC Score | chr4 | 109,322,809 | chr4:109322809:G:A | a | g | -0.2574 | 0.0627 | 4.00E-05 | ------+--+ | 8.305 | 9 | 0.5038 |
| ADNC Score | chr4 | 109,324,198 | chr4:109324198:T:C | t | c | 0.2876 | 0.0725 | 7.21E-05 | ++++++--++ | 9.091 | 9 | 0.4289 |
| ADNC Score | chr4 | 109,324,919 | chr4:109324919:C:T | t | c | -0.2876 | 0.0724 | 7.21E-05 | ------++-- | 9.091 | 9 | 0.4289 |
| ADNC Score | chr4 | 109,328,672 | chr4:109328672:T:G | t | g | 0.2537 | 0.0626 | 5.10E-05 | ++++++-++- | 8.615 | 9 | 0.4735 |
| ADNC Score | chr4 | 109,329,152 | chr4:109329152:T:C | t | c | 0.2715 | 0.063 | 1.65E-05 | ++++++-++- | 8.396 | 9 | 0.4948 |
| ADNC Score | chr4 | 109,330,050 | chr4:109330050:A:G | a | g | 0.2718 | 0.063 | 1.62E-05 | ++++++-++- | 8.364 | 9 | 0.498 |
| ADNC Score | chr4 | 109,330,199 | chr4:109330199:C:T | t | c | -0.2874 | 0.0725 | 7.32E-05 | ------++-- | 9.092 | 9 | 0.4288 |
| ADNC Score | chr4 | 109,330,934 | chr4:109330934:T:C | t | c | 0.2537 | 0.0626 | 5.11E-05 | ++++++-++- | 8.615 | 9 | 0.4736 |
| ADNC Score | chr4 | 109,330,953 | chr4:109330953:A:C | a | c | 0.2876 | 0.0725 | 7.22E-05 | ++++++--++ | 9.101 | 9 | 0.428 |
| ADNC Score | chr4 | 109,331,027 | chr4:109331027:A:T | a | t | 0.2538 | 0.0626 | 5.09E-05 | ++++++-++- | 8.703 | 9 | 0.4651 |
| ADNC Score | chr4 | 109,332,936 | chr4:109332936:C:T | t | c | -0.2534 | 0.0626 | 5.11E-05 | ------+--+ | 8.647 | 9 | 0.4705 |
| ADNC Score | chr4 | 109,332,944 | chr4:109332944:A:G | a | g | 0.3003 | 0.0754 | 6.83E-05 | ++++++--++ | 9.515 | 9 | 0.3911 |
| ADNC Score | chr4 | 109,333,685 | chr4:109333685:C:T | t | c | -0.2534 | 0.0626 | 5.17E-05 | ------+--+ | 8.651 | 9 | 0.4701 |
| ADNC Score | chr4 | 109,333,984 | chr4:109333984:A:G | a | g | 0.2714 | 0.063 | 1.63E-05 | ++++++-++- | 8.424 | 9 | 0.4921 |

| | | | | | | | | | | | | |
|---|---|---|---|---|---|---|---|---|---|---|---|---|
| ADNC Score | chr4 | 109,335,535 | chr4:109335535:T:G | t | g | 0.309 | 0.0755 | 4.28E-05 | ++++++--++ | 9.221 | 9 | 0.4171 |
| ADNC Score | chr4 | 109,338,225 | chr4:109338225:G:A | a | g | -0.2539 | 0.0626 | 5.06E-05 | ------+--+ | 8.618 | 9 | 0.4733 |
| ADNC Score | chr4 | 109,339,020 | chr4:109339020:C:T | t | c | -0.2537 | 0.0626 | 5.08E-05 | ------+--+ | 8.621 | 9 | 0.4729 |
| ADNC Score | chr4 | 109,339,463 | chr4:109339463:T:C | t | c | 0.2876 | 0.0725 | 7.24E-05 | ++++++--++ | 9.111 | 9 | 0.4271 |
| ADNC Score | chr4 | 109,340,132 | chr4:109340132:C:T | t | c | -0.2538 | 0.0626 | 5.06E-05 | ------+--+ | 8.619 | 9 | 0.4732 |
| ADNC Score | chr4 | 109,341,392 | chr4:109341392:G:A | a | g | -0.3004 | 0.0754 | 6.81E-05 | ------++-- | 9.518 | 9 | 0.3909 |
| ADNC Score | chr4 | 109,342,776 | chr4:109342776:A:C | a | c | 0.2668 | 0.063 | 2.26E-05 | ++++++-++- | 8.462 | 9 | 0.4883 |
| ADNC Score | chr4 | 109,342,801 | chr4:109342801:T:C | t | c | 0.2717 | 0.063 | 1.63E-05 | ++++++-++- | 8.392 | 9 | 0.4951 |
| ADNC Score | chr4 | 109,343,156 | chr4:109343156:A:C | a | c | 0.2538 | 0.0626 | 5.06E-05 | ++++++-++- | 8.619 | 9 | 0.4732 |
| ADNC Score | chr4 | 109,343,181 | chr4:109343181:A:G | a | g | 0.2538 | 0.0626 | 5.06E-05 | ++++++-++- | 8.618 | 9 | 0.4732 |
| ADNC Score | chr4 | 109,344,219 | chr4:109344219:G:A | a | g | -0.2475 | 0.0625 | 7.55E-05 | ------+--+ | 9.067 | 9 | 0.4311 |
| ADNC Score | chr4 | 109,345,035 | chr4:109345035:A:G | a | g | 0.2883 | 0.0724 | 6.83E-05 | ++++++--++ | 9.176 | 9 | 0.4212 |
| ADNC Score | chr4 | 109,345,724 | chr4:109345724:A:T | a | t | 0.2854 | 0.0725 | 8.22E-05 | ++++++--++ | 9.787 | 9 | 0.368 |
| ADNC Score | chr4 | 109,346,033 | chr4:109346033:G:A | a | g | -0.2708 | 0.063 | 1.72E-05 | ------+--+ | 8.341 | 9 | 0.5002 |
| ADNC Score | chr4 | 109,347,320 | chr4:109347320:G:C | c | g | -0.2688 | 0.0631 | 2.03E-05 | +-----+--+ | 8.732 | 9 | 0.4623 |
| ADNC Score | chr4 | 109,348,109 | chr4:109348109:T:C | t | c | 0.2514 | 0.0627 | 6.10E-05 | -+++++-++- | 9.013 | 9 | 0.4361 |
| ADNC Score | chr4 | 109,348,158 | chr4:109348158:G:A | a | g | -0.2639 | 0.0628 | 2.67E-05 | +-----+--+ | 8.426 | 9 | 0.4919 |
| ADNC Score | chr4 | 109,348,336 | chr4:109348336:G:A | a | g | -0.2512 | 0.0627 | 6.17E-05 | +-----+--+ | 8.93 | 9 | 0.4437 |
| ADNC Score | chr4 | 109,349,358 | chr4:109349358:T:C | t | c | 0.2637 | 0.0628 | 2.70E-05 | -+++++-++- | 8.398 | 9 | 0.4946 |
| ADNC Score | chr4 | 109,349,866 | chr4:109349866:C:G | c | g | 0.2573 | 0.0628 | 4.13E-05 | -+++++-++- | 8.375 | 9 | 0.4969 |
| ADNC Score | chr4 | 109,350,348 | chr4:109350348:G:A | a | g | -0.2639 | 0.0628 | 2.67E-05 | +-----+--+ | 8.408 | 9 | 0.4936 |
| ADNC Score | chr4 | 109,351,136 | chr4:109351136:G:A | a | g | -0.2573 | 0.0628 | 4.13E-05 | +-----+--+ | 8.374 | 9 | 0.497 |
| ADNC Score | chr4 | 109,351,278 | chr4:109351278:G:T | t | g | -0.2585 | 0.0628 | 3.82E-05 | +-----+--+ | 8.287 | 9 | 0.5055 |
| ADNC Score | chr4 | 109,351,913 | chr4:109351913:T:C | t | c | 0.2571 | 0.0628 | 4.20E-05 | -+++++-++- | 8.357 | 9 | 0.4986 |
| ADNC Score | chr4 | 109,351,990 | chr4:109351990:G:C | c | g | -0.2517 | 0.0627 | 6.02E-05 | +-----+--+ | 9.048 | 9 | 0.4329 |
| ADNC Score | chr4 | 109,352,303 | chr4:109352303:A:G | a | g | 0.2573 | 0.0628 | 4.13E-05 | -+++++-++- | 8.374 | 9 | 0.4969 |
| ADNC Score | chr4 | 109,353,067 | chr4:109353067:C:T | t | c | -0.2601 | 0.0628 | 3.44E-05 | +-----+--+ | 8.549 | 9 | 0.4799 |
| ADNC Score | chr4 | 109,353,364 | chr4:109353364:C:A | a | c | -0.2584 | 0.0663 | 9.81E-05 | -+----+--- | 10.091 | 9 | 0.3431 |
| ADNC Score | chr4 | 109,353,569 | chr4:109353569:A:AT | a | at | 0.2651 | 0.0632 | 2.74E-05 | -+++++-++- | 7.442 | 9 | 0.5912 |
| ADNC Score | chr4 | 109,353,677 | chr4:109353677:C:T | t | c | -0.2553 | 0.0628 | 4.77E-05 | +-----+--+ | 8.829 | 9 | 0.4532 |
| ADNC Score | chr4 | 109,354,002 | chr4:109354002:G:A | a | g | -0.2686 | 0.0628 | 1.90E-05 | +-----+--+ | 7.983 | 9 | 0.5358 |
| ADNC Score | chr4 | 109,354,447 | chr4:109354447:C:T | t | c | -0.268 | 0.0627 | 1.95E-05 | +-----+--+ | 7.826 | 9 | 0.5518 |
| ADNC Score | chr4 | 109,354,619 | chr4:109354619:G:A | a | g | -0.2679 | 0.0627 | 1.96E-05 | +-----+--+ | 7.829 | 9 | 0.5515 |
| ADNC Score | chr4 | 109,356,010 | chr4:109356010:T:C | t | c | 0.2674 | 0.0627 | 2.04E-05 | -+++++-++- | 7.834 | 9 | 0.551 |
| ADNC Score | chr4 | 109,356,052 | chr4:109356052:CCA:C | cca | c | 0.2551 | 0.0627 | 4.70E-05 | -+++++-+++ | 8.617 | 9 | 0.4733 |
| ADNC Score | chr4 | 109,356,238 | chr4:109356238:C:A | a | c | -0.2569 | 0.0626 | 4.12E-05 | +-----+--+ | 8.722 | 9 | 0.4634 |
| ADNC Score | chr4 | 109,357,327 | chr4:109357327:C:A | a | c | -0.2618 | 0.0626 | 2.91E-05 | +-----+--+ | 8.392 | 9 | 0.4952 |
| ADNC Score | chr4 | 109,357,660 | chr4:109357660:G:A | a | g | -0.2673 | 0.0625 | 1.87E-05 | +-----+--+ | 7.971 | 9 | 0.5371 |
| ADNC Score | chr4 | 109,357,959 | chr4:109357959:T:C | t | c | 0.2619 | 0.0627 | 2.95E-05 | -+++++-++- | 8.402 | 9 | 0.4942 |
| ADNC Score | chr4 | 109,358,130 | chr4:109358130:C:G | c | g | 0.2679 | 0.0627 | 1.96E-05 | -+++++-++- | 7.829 | 9 | 0.5515 |
| ADNC Score | chr4 | 109,358,568 | chr4:109358568:C:T | t | c | -0.2743 | 0.0628 | 1.27E-05 | +-----+--+ | 7.878 | 9 | 0.5465 |
| ADNC Score | chr4 | 109,358,621 | chr4:109358621:C:T | t | c | -0.2679 | 0.0627 | 1.96E-05 | +-----+--+ | 7.829 | 9 | 0.5515 |
| ADNC Score | chr4 | 109,359,934 | chr4:109359934:AG:A | a | ag | -0.2752 | 0.0628 | 1.20E-05 | +-----+--+ | 7.873 | 9 | 0.547 |
| ADNC Score | chr4 | 109,360,247 | chr4:109360247:C:T | t | c | -0.2678 | 0.0627 | 1.97E-05 | +-----+--+ | 7.832 | 9 | 0.5512 |
| ADNC Score | chr4 | 109,360,396 | chr4:109360396:G:A | a | g | -0.2678 | 0.0627 | 1.98E-05 | +-----+--+ | 7.829 | 9 | 0.5515 |
| ADNC Score | chr4 | 109,360,417 | chr4:109360417:A:G | a | g | 0.2736 | 0.0628 | 1.32E-05 | -+++++-++- | 7.921 | 9 | 0.5421 |

| | | | | | | | | | | | | |
|---|---|---|---|---|---|---|---|---|---|---|---|---|
| ADNC Score | chr4 | 109,360,452 | chr4:109360452:C:G | c | g | 0.2678 | 0.0627 | 1.97E-05 | -+++++-++- | 7.827 | 9 | 0.5516 |
| ADNC Score | chr4 | 109,360,697 | chr4:109360697:G:A | a | g | -0.2678 | 0.0627 | 1.98E-05 | +-----+--+ | 7.829 | 9 | 0.5515 |
| ADNC Score | chr4 | 109,360,796 | chr4:109360796:G:C | c | g | -0.2677 | 0.0627 | 1.98E-05 | +-----+--+ | 7.833 | 9 | 0.5511 |
| ADNC Score | chr4 | 109,360,923 | chr4:109360923:C:T | t | c | -0.2676 | 0.0627 | 2.00E-05 | +-----+--+ | 7.821 | 9 | 0.5523 |
| ADNC Score | chr4 | 109,361,207 | chr4:109361207:T:G | t | g | 0.2678 | 0.0627 | 1.98E-05 | -+++++-++- | 7.83 | 9 | 0.5513 |
| ADNC Score | chr4 | 109,361,422 | chr4:109361422:C:T | t | c | -0.2743 | 0.0628 | 1.27E-05 | +-----+--+ | 7.883 | 9 | 0.546 |
| ADNC Score | chr4 | 109,361,729 | chr4:109361729:G:C | c | g | -0.2677 | 0.0628 | 2.00E-05 | +-----+--+ | 7.83 | 9 | 0.5514 |
| ADNC Score | chr4 | 109,361,778 | chr4:109361778:G:C | c | g | -0.2743 | 0.0628 | 1.27E-05 | +-----+--+ | 7.88 | 9 | 0.5463 |
| ADNC Score | chr4 | 109,362,552 | chr4:109362552:G:A | a | g | -0.2636 | 0.0628 | 2.67E-05 | +-----+--+ | 8.43 | 9 | 0.4915 |
| ADNC Score | chr4 | 109,364,587 | chr4:109364587:G:C | c | g | -0.2537 | 0.0638 | 7.07E-05 | +-----+--+ | 9.501 | 9 | 0.3924 |
| ADNC Score | chr4 | 109,365,610 | chr4:109365610:A:G | a | g | 0.2603 | 0.0639 | 4.66E-05 | -+++++-++- | 9.616 | 9 | 0.3825 |
| ADNC Score | chr4 | 109,366,038 | chr4:109366038:C:T | t | c | -0.2603 | 0.0639 | 4.66E-05 | +-----+--+ | 9.609 | 9 | 0.383 |
| ADNC Score | chr4 | 109,366,949 | chr4:109366949:C:T | t | c | -0.2658 | 0.0641 | 3.36E-05 | +-----+--+ | 9.264 | 9 | 0.4133 |
| ADNC Score | chr4 | 109,368,439 | chr4:109368439:C:T | t | c | -0.2659 | 0.0641 | 3.35E-05 | +-----+--+ | 9.261 | 9 | 0.4136 |
| ADNC Score | chr4 | 109,370,273 | chr4:109370273:T:G | t | g | 0.2546 | 0.064 | 6.87E-05 | -+++++-++- | 9.617 | 9 | 0.3824 |
| ADNC Score | chr4 | 109,370,817 | chr4:109370817:C:A | a | c | -0.2545 | 0.064 | 6.93E-05 | +-----+--+ | 9.593 | 9 | 0.3844 |
| ADNC Score | chr4 | 109,370,982 | chr4:109370982:A:G | a | g | 0.2547 | 0.064 | 6.84E-05 | -+++++-++- | 9.617 | 9 | 0.3824 |
| ADNC Score | chr4 | 109,372,281 | chr4:109372281:A:G | a | g | 0.2547 | 0.064 | 6.86E-05 | -+++++-++- | 9.58 | 9 | 0.3855 |
| ADNC Score | chr4 | 109,373,227 | chr4:109373227:T:A | a | t | -0.3106 | 0.0758 | 4.13E-05 | ------++-- | 9.161 | 9 | 0.4226 |
| ADNC Score | chr4 | 109,377,899 | chr4:109377899:A:C | a | c | 0.265 | 0.0641 | 3.55E-05 | -+++++-++- | 9.641 | 9 | 0.3803 |
| ADNC Score | chr4 | 109,382,211 | chr4:109382211:A:G | a | g | 0.2631 | 0.0639 | 3.88E-05 | -+++++-++- | 9.824 | 9 | 0.3649 |
| ADNC Score | chr4 | 109,382,700 | chr4:109382700:T:C | t | c | 0.2502 | 0.0638 | 8.86E-05 | -+++++-++- | 10.018 | 9 | 0.3491 |
| ADNC Score | chr4 | 109,384,388 | chr4:109384388:G:A | a | g | -0.2504 | 0.0638 | 8.70E-05 | +-----+--+ | 10.049 | 9 | 0.3465 |
| ADNC Score | chr4 | 109,386,841 | chr4:109386841:G:T | t | g | -0.2581 | 0.064 | 5.54E-05 | +-----+--+ | 9.516 | 9 | 0.391 |
| ADNC Score | chr4 | 109,387,079 | chr4:109387079:A:G | a | g | 0.2541 | 0.0639 | 6.95E-05 | -+++++-++- | 9.337 | 9 | 0.4067 |
| ADNC Score | chr4 | 109,388,053 | chr4:109388053:A:G | a | g | 0.2539 | 0.0639 | 7.05E-05 | -+++++-++- | 9.359 | 9 | 0.4048 |
| ADNC Score | chr4 | 109,388,414 | chr4:109388414:T:A | a | t | -0.2539 | 0.0639 | 7.07E-05 | +-----+--+ | 9.36 | 9 | 0.4048 |
| ADNC Score | chr4 | 109,389,970 | chr4:109389970:T:C | t | c | 0.2582 | 0.0641 | 5.59E-05 | -+++++-++- | 9.525 | 9 | 0.3903 |
| ADNC Score | chr4 | 121,552,062 | chr4:121552062:A:G | a | g | -0.7698 | 0.1895 | 4.88E-05 | ++--+--?-? | 10.533 | 7 | 0.1603 |
| ADNC Score | chr4 | 174,313,278 | chr4:174313278:C:G | c | g | -0.2317 | 0.0591 | 8.87E-05 | -------+-- | 7.052 | 9 | 0.6317 |
| ADNC Score | chr4 | 174,318,221 | chr4:174318221:G:A | a | g | 0.2298 | 0.0591 | 9.96E-05 | +++++++-+- | 7.07 | 9 | 0.6298 |
| ADNC Score | chr4 | 174,320,434 | chr4:174320434:C:G | c | g | -0.2303 | 0.0591 | 9.74E-05 | -------+-- | 7.024 | 9 | 0.6346 |
| ADNC Score | chr4 | 177,351,876 | chr4:177351876:A:G | a | g | 0.3247 | 0.0822 | 7.82E-05 | -++++++++- | 10.009 | 9 | 0.3497 |
| ADNC Score | chr4 | 185,453,565 | chr4:185453565:C:T | t | c | -0.2623 | 0.0672 | 9.55E-05 | ++-------+ | 9.348 | 9 | 0.4058 |
| ADNC Score | chr4 | 187,686,669 | chr4:187686669:AT:A | a | at | -0.3576 | 0.0863 | 3.40E-05 | +---+----- | 4.081 | 9 | 0.906 |
| ADNC Score | chr5 | 25,892,157 | chr5:25892157:A:G | a | g | -0.221 | 0.0516 | 1.84E-05 | ----++---- | 15.085 | 9 | 0.08862 |
| ADNC Score | chr5 | 25,988,985 | chr5:25988985:C:T | t | c | -0.392 | 0.0966 | 4.97E-05 | -----+-+-- | 10.473 | 9 | 0.3136 |
| ADNC Score | chr5 | 40,170,452 | chr5:40170452:C:T | t | c | 0.9077 | 0.2312 | 8.65E-05 | ?-++?++?+? | 5.787 | 5 | 0.3275 |
| ADNC Score | chr5 | 51,936,547 | chr5:51936547:G:A | a | g | 0.6846 | 0.1698 | 5.55E-05 | --++?+-?+? | 15.903 | 6 | 0.01428 |
| ADNC Score | chr5 | 67,148,764 | chr5:67148764:G:A | a | g | -0.5829 | 0.1395 | 2.91E-05 | +---?--+-- | 8.575 | 8 | 0.3794 |
| ADNC Score | chr5 | 67,170,108 | chr5:67170108:AT:A | a | at | -0.5695 | 0.1385 | 3.92E-05 | +---?--+-- | 7.8 | 8 | 0.4533 |
| ADNC Score | chr5 | 76,153,810 | chr5:76153810:A:G | a | g | -0.198 | 0.0474 | 2.97E-05 | -+-------+ | 11.818 | 9 | 0.2238 |
| ADNC Score | chr5 | 76,153,885 | chr5:76153885:G:A | a | g | 0.1882 | 0.0475 | 7.42E-05 | +-++-+++++ | 8.838 | 9 | 0.4524 |
| ADNC Score | chr5 | 76,158,198 | chr5:76158198:A:T | a | t | 0.2001 | 0.0474 | 2.38E-05 | +-+++++++- | 11.362 | 9 | 0.2517 |
| ADNC Score | chr5 | 76,158,439 | chr5:76158439:T:C | t | c | 0.1916 | 0.0476 | 5.72E-05 | +-++-+++++ | 8.315 | 9 | 0.5027 |
| ADNC Score | chr5 | 76,159,825 | chr5:76159825:A:G | a | g | 0.1985 | 0.0474 | 2.80E-05 | +-+++++++- | 11.663 | 9 | 0.233 |

| | | | | | | | | | | | | |
|---|---|---|---|---|---|---|---|---|---|---|---|---|
| ADNC Score | chr5 | 76,160,114 | chr5:76160114:G:A | a | g | -0.1969 | 0.0474 | 3.25E-05 | -+-------+ | 11.748 | 9 | 0.2279 |
| ADNC Score | chr5 | 76,161,387 | chr5:76161387:T:C | t | c | 0.1984 | 0.0474 | 2.79E-05 | +-+++++++- | 11.739 | 9 | 0.2284 |
| ADNC Score | chr5 | 76,162,267 | chr5:76162267:G:C | c | g | -0.1927 | 0.0476 | 5.15E-05 | -+--+----- | 8.162 | 9 | 0.5179 |
| ADNC Score | chr5 | 76,162,576 | chr5:76162576:C:T | t | c | -0.1972 | 0.047 | 2.69E-05 | -+-------+ | 11.436 | 9 | 0.247 |
| ADNC Score | chr5 | 76,163,060 | chr5:76163060:C:T | t | c | -0.1919 | 0.0475 | 5.35E-05 | -+-------+ | 11.076 | 9 | 0.2705 |
| ADNC Score | chr5 | 76,166,269 | chr5:76166269:C:T | t | c | -0.1899 | 0.0476 | 6.57E-05 | -+-------+ | 11.129 | 9 | 0.267 |
| ADNC Score | chr5 | 76,168,605 | chr5:76168605:G:A | a | g | -0.1871 | 0.0477 | 8.75E-05 | -+-------+ | 12.958 | 9 | 0.1645 |
| ADNC Score | chr5 | 76,174,261 | chr5:76174261:T:C | t | c | 0.1904 | 0.0477 | 6.61E-05 | +-+++++++- | 9.676 | 9 | 0.3774 |
| ADNC Score | chr5 | 76,174,991 | chr5:76174991:T:G | t | g | 0.1896 | 0.0477 | 6.98E-05 | +-+++++++- | 9.766 | 9 | 0.3698 |
| ADNC Score | chr5 | 76,175,562 | chr5:76175562:C:G | c | g | 0.1929 | 0.0478 | 5.50E-05 | +-+++++++- | 9.41 | 9 | 0.4003 |
| ADNC Score | chr5 | 76,175,668 | chr5:76175668:A:G | a | g | 0.1929 | 0.0478 | 5.50E-05 | +-+++++++- | 9.41 | 9 | 0.4003 |
| ADNC Score | chr5 | 76,175,892 | chr5:76175892:C:T | t | c | -0.1925 | 0.0477 | 5.55E-05 | -+-------+ | 9.31 | 9 | 0.4091 |
| ADNC Score | chr5 | 76,176,379 | chr5:76176379:C:T | t | c | -0.1908 | 0.0479 | 6.68E-05 | -+--+----- | 7.601 | 9 | 0.5748 |
| ADNC Score | chr5 | 76,176,382 | chr5:76176382:G:A | a | g | -0.1921 | 0.0478 | 5.86E-05 | -+-------+ | 9.495 | 9 | 0.3929 |
| ADNC Score | chr5 | 76,177,091 | chr5:76177091:G:A | a | g | -0.1935 | 0.0478 | 5.16E-05 | -+-------+ | 9.567 | 9 | 0.3866 |
| ADNC Score | chr5 | 76,177,136 | chr5:76177136:A:G | a | g | 0.1915 | 0.0478 | 6.17E-05 | +-+++++++- | 9.588 | 9 | 0.3849 |
| ADNC Score | chr5 | 76,177,232 | chr5:76177232:G:A | a | g | -0.1913 | 0.0478 | 6.29E-05 | -+-------+ | 9.465 | 9 | 0.3955 |
| ADNC Score | chr5 | 76,177,492 | chr5:76177492:A:G | a | g | 0.1921 | 0.0478 | 5.86E-05 | +-+++++++- | 9.493 | 9 | 0.393 |
| ADNC Score | chr5 | 76,177,733 | chr5:76177733:C:A | a | c | -0.1921 | 0.0478 | 5.89E-05 | -+-------+ | 9.49 | 9 | 0.3934 |
| ADNC Score | chr5 | 76,178,384 | chr5:76178384:C:A | a | c | -0.1907 | 0.0479 | 6.83E-05 | -+--+----- | 7.711 | 9 | 0.5636 |
| ADNC Score | chr5 | 76,178,433 | chr5:76178433:G:A | a | g | -0.1921 | 0.0478 | 5.88E-05 | -+-------+ | 9.495 | 9 | 0.3929 |
| ADNC Score | chr5 | 76,178,625 | chr5:76178625:T:C | t | c | 0.1921 | 0.0478 | 5.85E-05 | +-+++++++- | 9.485 | 9 | 0.3938 |
| ADNC Score | chr5 | 76,178,955 | chr5:76178955:C:T | t | c | -0.1922 | 0.0478 | 5.75E-05 | -+-------+ | 9.397 | 9 | 0.4015 |
| ADNC Score | chr5 | 76,179,096 | chr5:76179096:C:T | t | c | -0.1921 | 0.0478 | 5.88E-05 | -+-------+ | 9.492 | 9 | 0.3931 |
| ADNC Score | chr5 | 76,179,177 | chr5:76179177:C:G | c | g | 0.1923 | 0.0478 | 5.76E-05 | +-+++++++- | 9.48 | 9 | 0.3942 |
| ADNC Score | chr5 | 76,179,197 | chr5:76179197:T:C | t | c | 0.1921 | 0.0478 | 5.88E-05 | +-+++++++- | 9.498 | 9 | 0.3927 |
| ADNC Score | chr5 | 76,180,281 | chr5:76180281:G:A | a | g | -0.1915 | 0.0478 | 6.17E-05 | -+-------+ | 9.486 | 9 | 0.3937 |
| ADNC Score | chr5 | 76,180,341 | chr5:76180341:T:C | t | c | 0.1915 | 0.0478 | 6.15E-05 | +-+++++++- | 9.485 | 9 | 0.3938 |
| ADNC Score | chr5 | 76,180,616 | chr5:76180616:A:G | a | g | 0.1915 | 0.0478 | 6.17E-05 | +-+++++++- | 9.486 | 9 | 0.3936 |
| ADNC Score | chr5 | 76,180,673 | chr5:76180673:C:T | t | c | -0.1916 | 0.0478 | 6.11E-05 | -+-------+ | 9.486 | 9 | 0.3937 |
| ADNC Score | chr5 | 76,180,902 | chr5:76180902:C:T | t | c | -0.1916 | 0.0478 | 6.11E-05 | -+-------+ | 9.478 | 9 | 0.3944 |
| ADNC Score | chr5 | 76,181,098 | chr5:76181098:T:TC | t | tc | 0.1933 | 0.0478 | 5.30E-05 | +-+++++++- | 9.445 | 9 | 0.3973 |
| ADNC Score | chr5 | 76,181,376 | chr5:76181376:T:C | t | c | 0.1931 | 0.0478 | 5.40E-05 | +-+++++++- | 9.58 | 9 | 0.3855 |
| ADNC Score | chr5 | 76,181,390 | chr5:76181390:T:C | t | c | 0.191 | 0.0478 | 6.50E-05 | +-+++++++- | 9.518 | 9 | 0.3909 |
| ADNC Score | chr5 | 80,105,594 | chr5:80105594:T:C | t | c | -0.2219 | 0.0569 | 9.60E-05 | -------+- | 12.477 | 9 | 0.1877 |
| ADNC Score | chr5 | 96,491,212 | chr5:96491212:A:G | a | g | 0.3787 | 0.0968 | 9.19E-05 | +-++-+++++ | 9.904 | 9 | 0.3583 |
| ADNC Score | chr5 | 96,491,795 | chr5:96491795:G:A | a | g | -0.382 | 0.0966 | 7.65E-05 | -+--+----- | 10.48 | 9 | 0.3131 |
| ADNC Score | chr5 | 96,492,472 | chr5:96492472:G:A | a | g | -0.3811 | 0.0968 | 8.24E-05 | -+--+----- | 10.564 | 9 | 0.3068 |
| ADNC Score | chr5 | 103,904,219 | chr5:103904219:A:T | a | t | -0.2105 | 0.0505 | 3.03E-05 | --------- | 13.132 | 9 | 0.1567 |
| ADNC Score | chr5 | 103,905,849 | chr5:103905849:C:T | t | c | 0.2108 | 0.0506 | 3.07E-05 | +++++++++ | 13.124 | 9 | 0.1571 |
| ADNC Score | chr5 | 103,907,015 | chr5:103907015:C:A | a | c | 0.206 | 0.0507 | 4.81E-05 | ++-++++++- | 13.634 | 9 | 0.136 |
| ADNC Score | chr5 | 103,911,133 | chr5:103911133:A:G | a | g | -0.2219 | 0.0507 | 1.18E-05 | --------- | 14.057 | 9 | 0.1203 |
| ADNC Score | chr5 | 124,165,293 | chr5:124165293:G:T | t | g | 0.8857 | 0.2225 | 6.86E-05 | +?++++-+?? | 9.942 | 6 | 0.1271 |
| ADNC Score | chr5 | 133,875,847 | chr5:133875847:T:C | t | c | -0.5124 | 0.1228 | 3.02E-05 | -----+---? | 12.228 | 8 | 0.1413 |
| ADNC Score | chr5 | 148,888,172 | chr5:148888172:T:G | t | g | 0.1963 | 0.0503 | 9.59E-05 | ++++-+-+++ | 20.052 | 9 | 0.01759 |
| ADNC Score | chr5 | 148,888,967 | chr5:148888967:T:C | t | c | 0.1968 | 0.0502 | 8.96E-05 | ++++-+-+++ | 19.551 | 9 | 0.02089 |

| | | | | | | | | | | | | |
|---|---|---|---|---|---|---|---|---|---|---|---|---|
| ADNC Score | chr5 | 162,435,356 | chr5:162435356:T:C | t | c | -0.8473 | 0.2041 | 3.31E-05 | ----?-++-+ | 15.667 | 8 | 0.0474 |
| ADNC Score | chr6 | 41,696,343 | chr6:41696343:C:T | t | c | 1.2432 | 0.2809 | 9.58E-06 | -??+?++++- | 6.219 | 6 | 0.3991 |
| ADNC Score | chr6 | 62,790,406 | chr6:62790406:C:A | a | c | 0.3607 | 0.0896 | 5.64E-05 | --+++++++- | 2.609 | 9 | 0.9778 |
| ADNC Score | chr6 | 62,802,119 | chr6:62802119:T:C | t | c | -0.3693 | 0.0895 | 3.66E-05 | ++-------+ | 2.73 | 9 | 0.974 |
| ADNC Score | chr6 | 62,816,813 | chr6:62816813:G:C | c | g | 0.3734 | 0.0896 | 3.07E-05 | -++++++++- | 2.704 | 9 | 0.9749 |
| ADNC Score | chr6 | 62,843,826 | chr6:62843826:A:T | a | t | -0.3607 | 0.0897 | 5.83E-05 | +--------+ | 2.871 | 9 | 0.9692 |
| ADNC Score | chr6 | 66,970,333 | chr6:66970333:T:A | a | t | 0.2619 | 0.0654 | 6.27E-05 | +-+++++++ | 10.914 | 8 | 0.2066 |
| ADNC Score | chr6 | 70,168,088 | chr6:70168088:G:A | a | g | 0.226 | 0.0565 | 6.39E-05 | ++++-++++- | 7.604 | 9 | 0.5745 |
| ADNC Score | chr6 | 70,169,525 | chr6:70169525:A:G | a | g | -0.2266 | 0.0565 | 6.14E-05 | ----+----- | 7.58 | 9 | 0.577 |
| ADNC Score | chr6 | 70,170,039 | chr6:70170039:A:G | a | g | -0.227 | 0.0565 | 5.95E-05 | ----+----- | 7.565 | 9 | 0.5785 |
| ADNC Score | chr6 | 70,178,050 | chr6:70178050:C:A | a | c | 0.2247 | 0.0564 | 6.88E-05 | ++++-++++- | 7.658 | 9 | 0.569 |
| ADNC Score | chr6 | 70,183,182 | chr6:70183182:C:T | t | c | 0.2246 | 0.0565 | 7.01E-05 | ++++-++++- | 7.619 | 9 | 0.5729 |
| ADNC Score | chr6 | 70,190,735 | chr6:70190735:G:A | a | g | 0.2261 | 0.0565 | 6.32E-05 | ++++-++++- | 7.524 | 9 | 0.5828 |
| ADNC Score | chr6 | 70,192,673 | chr6:70192673:G:A | a | g | 0.2259 | 0.0565 | 6.43E-05 | ++++-++++- | 7.522 | 9 | 0.583 |
| ADNC Score | chr6 | 70,225,443 | chr6:70225443:TTTTC:T | t | ttttc | 0.2327 | 0.0567 | 4.03E-05 | ++++-++++- | 6.743 | 9 | 0.6639 |
| ADNC Score | chr6 | 70,234,739 | chr6:70234739:G:C | c | g | 0.2298 | 0.0567 | 5.01E-05 | ++++-++++- | 6.584 | 9 | 0.6804 |
| ADNC Score | chr6 | 70,239,404 | chr6:70239404:C:T | t | c | 0.2398 | 0.0571 | 2.64E-05 | ++++-++++- | 6.362 | 9 | 0.7033 |
| ADNC Score | chr6 | 70,354,489 | chr6:70354489:CT:C | ct | c | 0.7788 | 0.1713 | 5.49E-06 | ++++-++-+- | 8.811 | 9 | 0.4549 |
| ADNC Score | chr6 | 70,357,025 | chr6:70357025:G:C | c | g | -0.7956 | 0.1734 | 4.50E-06 | ----+--+-+ | 9.411 | 9 | 0.4003 |
| ADNC Score | chr6 | 70,357,551 | chr6:70357551:TGAAAAA:T | t | tgaaaaa | -0.7998 | 0.1743 | 4.45E-06 | ----+--+-+ | 9.773 | 9 | 0.3692 |
| ADNC Score | chr6 | 96,048,937 | chr6:96048937:C:T | t | c | 0.6457 | 0.1616 | 6.45E-05 | -++++++++- | 4.947 | 9 | 0.8389 |
| ADNC Score | chr6 | 96,071,402 | chr6:96071402:A:T | a | t | -0.6384 | 0.162 | 8.08E-05 | +--------- | 4.844 | 9 | 0.8477 |
| ADNC Score | chr6 | 96,102,337 | chr6:96102337:A:C | a | c | -0.6609 | 0.1567 | 2.48E-05 | +--------- | 5.438 | 9 | 0.7946 |
| ADNC Score | chr6 | 137,167,429 | chr6:137167429:T:C | t | c | -0.8772 | 0.1979 | 9.34E-06 | -?--?----? | 1.383 | 6 | 0.9669 |
| ADNC Score | chr6 | 150,711,950 | chr6:150711950:T:C | t | c | 0.1832 | 0.0471 | 9.92E-05 | ++++-++++- | 6.649 | 9 | 0.6736 |
| ADNC Score | chr6 | 154,765,540 | chr6:154765540:G:A | a | g | 1.1258 | 0.2772 | 4.87E-05 | ++++??+?? | 0.21 | 4 | 0.9949 |
| ADNC Score | chr6 | 154,793,674 | chr6:154793674:G:T | t | g | 1.0906 | 0.2745 | 7.09E-05 | ++++??+?? | 0.154 | 4 | 0.9972 |
| ADNC Score | chr6 | 154,827,048 | chr6:154827048:T:C | t | c | -1.0888 | 0.2743 | 7.21E-05 | ----??-??? | 0.149 | 4 | 0.9974 |
| ADNC Score | chr6 | 164,185,512 | chr6:164185512:G:A | a | g | 1.1035 | 0.2785 | 7.42E-05 | ?+++??-??+ | 3.952 | 4 | 0.4126 |
| ADNC Score | chr6 | 166,710,006 | chr6:166710006:C:T | t | c | 0.5896 | 0.149 | 7.63E-05 | +-+++++-++ | 11.048 | 9 | 0.2724 |
| ADNC Score | chr6 | 166,726,817 | chr6:166726817:C:T | t | c | 0.7817 | 0.2009 | 9.96E-05 | -?+-+++-++ | 15.258 | 8 | 0.05433 |
| ADNC Score | chr6 | 167,568,583 | chr6:167568583:G:A | a | g | 0.7838 | 0.1963 | 6.51E-05 | --++-++-+? | 5.618 | 8 | 0.6899 |
| ADNC Score | chr6 | 167,588,023 | chr6:167588023:G:A | a | g | 0.7879 | 0.1974 | 6.58E-05 | --++-++-+? | 3.653 | 8 | 0.887 |
| ADNC Score | chr7 | 3,541,725 | chr7:3541725:G:A | a | g | -0.2998 | 0.0769 | 9.68E-05 | +--------+ | 7.859 | 9 | 0.5484 |
| ADNC Score | chr7 | 6,045,230 | chr7:6045230:C:G | c | g | -0.8242 | 0.1663 | 7.18E-07 | ---------- | 2.139 | 9 | 0.9891 |
| ADNC Score | chr7 | 6,064,111 | chr7:6064111:G:A | a | g | 0.8495 | 0.1663 | 3.24E-07 | +++++++++ | 2.721 | 9 | 0.9743 |
| ADNC Score | chr7 | 21,247,024 | chr7:21247024:A:G | a | g | 0.2069 | 0.05 | 3.50E-05 | ++++-++++- | 5.524 | 9 | 0.7864 |
| ADNC Score | chr7 | 21,283,617 | chr7:21283617:G:A | a | g | -0.1905 | 0.0488 | 9.42E-05 | -+-------+ | 3.323 | 9 | 0.9501 |
| ADNC Score | chr7 | 23,393,184 | chr7:23393184:G:A | a | g | 1.1443 | 0.2756 | 3.31E-05 | ??++??++? | 1.244 | 3 | 0.7425 |
| ADNC Score | chr7 | 23,926,416 | chr7:23926416:T:C | t | c | 0.1976 | 0.0503 | 8.58E-05 | +++++++++ | 9.703 | 9 | 0.375 |
| ADNC Score | chr7 | 23,926,424 | chr7:23926424:G:A | a | g | -0.2065 | 0.0506 | 4.48E-05 | ---------- | 9.864 | 9 | 0.3616 |
| ADNC Score | chr7 | 27,289,727 | chr7:27289727:G:C | c | g | 0.4456 | 0.1066 | 2.93E-05 | +-+++++++ | 3.328 | 8 | 0.9121 |
| ADNC Score | chr7 | 27,590,854 | chr7:27590854:C:T | t | c | -0.8459 | 0.2061 | 4.07E-05 | -+----+-?+ | 7.885 | 8 | 0.4448 |
| ADNC Score | chr7 | 42,229,672 | chr7:42229672:C:T | t | c | 0.8544 | 0.2099 | 4.70E-05 | ?+++?++?+ | 4.785 | 6 | 0.5717 |
| ADNC Score | chr7 | 50,470,354 | chr7:50470354:G:A | a | g | 0.7711 | 0.1966 | 8.77E-05 | ?+++?+?++ | 5.31 | 5 | 0.3792 |
| ADNC Score | chr7 | 50,968,202 | chr7:50968202:A:G | a | g | -0.1998 | 0.0475 | 2.63E-05 | ---------- | 7.36 | 9 | 0.5997 |

| | | | | | | | | | | | | |
|---|---|---|---|---|---|---|---|---|---|---|---|---|
| ADNC Score | chr7 | 97,384,119 | chr7:97384119:C:A | a | c | 0.3421 | 0.0842 | 4.89E-05 | +++++++++ | 6.706 | 9 | 0.6677 |
| ADNC Score | chr7 | 104,356,845 | chr7:104356845:C:T | t | c | -0.2208 | 0.0543 | 4.78E-05 | +-------- | 3.907 | 9 | 0.9174 |
| ADNC Score | chr7 | 104,358,266 | chr7:104358266:G:A | a | g | -0.215 | 0.0531 | 5.10E-05 | --------- | 3.119 | 9 | 0.9594 |
| ADNC Score | chr7 | 104,360,349 | chr7:104360349:C:G | c | g | 0.2224 | 0.0543 | 4.22E-05 | -++++++++ | 4.1 | 9 | 0.9047 |
| ADNC Score | chr7 | 130,971,087 | chr7:130971087:G:A | a | g | 0.3978 | 0.101 | 8.25E-05 | -+++++-+++ | 9.134 | 9 | 0.425 |
| ADNC Score | chr7 | 138,809,278 | chr7:138809278:C:G | c | g | -0.9375 | 0.239 | 8.74E-05 | ?----+?--- | 7.747 | 7 | 0.3555 |
| ADNC Score | chr7 | 138,818,355 | chr7:138818355:G:A | a | g | 0.7776 | 0.1975 | 8.21E-05 | ?-+++-++++ | 8.397 | 8 | 0.3957 |
| ADNC Score | chr8 | 5,783,136 | chr8:5783136:G:C | c | g | 0.3325 | 0.0781 | 2.08E-05 | +-+++++-+- | 6.506 | 9 | 0.6884 |
| ADNC Score | chr8 | 14,598,491 | chr8:14598491:C:A | a | c | -0.191 | 0.0485 | 8.37E-05 | ------+--+ | 13.357 | 9 | 0.1471 |
| ADNC Score | chr8 | 16,147,702 | chr8:16147702:G:C | c | g | 0.1892 | 0.0486 | 9.78E-05 | +++++++-+ | 3.791 | 9 | 0.9246 |
| ADNC Score | chr8 | 19,461,912 | chr8:19461912:G:A | a | g | -0.193 | 0.0475 | 4.80E-05 | +-------- | 5.75 | 9 | 0.7646 |
| ADNC Score | chr8 | 19,462,109 | chr8:19462109:G:A | a | g | -0.192 | 0.0474 | 5.18E-05 | +-------- | 5.659 | 9 | 0.7735 |
| ADNC Score | chr8 | 19,462,114 | chr8:19462114:G:A | a | g | -0.204 | 0.0503 | 5.07E-05 | ---------+ | 3.285 | 9 | 0.9519 |
| ADNC Score | chr8 | 19,462,685 | chr8:19462685:T:G | t | g | 0.2032 | 0.0503 | 5.27E-05 | +++++++++ | 3.29 | 9 | 0.9517 |
| ADNC Score | chr8 | 19,463,874 | chr8:19463874:A:G | a | g | 0.1898 | 0.0474 | 6.34E-05 | -++++++++ | 5.501 | 9 | 0.7887 |
| ADNC Score | chr8 | 19,465,551 | chr8:19465551:T:A | a | t | -0.2101 | 0.0535 | 8.47E-05 | --------- | 2.596 | 9 | 0.9782 |
| ADNC Score | chr8 | 19,469,976 | chr8:19469976:G:A | a | g | -0.1892 | 0.0481 | 8.49E-05 | -------+-+ | 3.95 | 9 | 0.9147 |
| ADNC Score | chr8 | 19,470,132 | chr8:19470132:G:A | a | g | -0.1884 | 0.0481 | 9.01E-05 | -------+-+ | 3.914 | 9 | 0.917 |
| ADNC Score | chr8 | 19,470,448 | chr8:19470448:G:A | a | g | -0.1925 | 0.0481 | 6.36E-05 | -------+-+ | 4.12 | 9 | 0.9033 |
| ADNC Score | chr8 | 19,470,708 | chr8:19470708:G:A | a | g | -0.1888 | 0.0482 | 8.80E-05 | -------+-+ | 4.082 | 9 | 0.9059 |
| ADNC Score | chr8 | 19,470,956 | chr8:19470956:C:A | a | c | -0.1886 | 0.0482 | 9.05E-05 | -------+-+ | 3.38 | 9 | 0.9473 |
| ADNC Score | chr8 | 19,471,053 | chr8:19471053:A:G | a | g | 0.1897 | 0.0482 | 8.28E-05 | +++++++-+ | 3.185 | 9 | 0.9565 |
| ADNC Score | chr8 | 19,471,792 | chr8:19471792:G:C | c | g | -0.1961 | 0.0482 | 4.75E-05 | -------+-+ | 3.793 | 9 | 0.9245 |
| ADNC Score | chr8 | 22,203,361 | chr8:22203361:G:A | a | g | 0.266 | 0.0676 | 8.30E-05 | ++++-+-+++ | 9.746 | 9 | 0.3714 |
| ADNC Score | chr8 | 27,917,960 | chr8:27917960:G:T | t | g | 1.0647 | 0.2372 | 7.16E-06 | +-++++++?- | 3.635 | 8 | 0.8884 |
| ADNC Score | chr8 | 27,920,192 | chr8:27920192:T:C | t | c | -0.913 | 0.2129 | 1.80E-05 | --------?- | 7.849 | 8 | 0.4483 |
| ADNC Score | chr8 | 27,921,779 | chr8:27921779:G:A | a | g | 0.9465 | 0.2158 | 1.15E-05 | ++++++++? | 8.969 | 8 | 0.3449 |
| ADNC Score | chr8 | 27,927,842 | chr8:27927842:T:C | t | c | -0.8479 | 0.2045 | 3.37E-05 | --------?- | 8.717 | 8 | 0.3667 |
| ADNC Score | chr8 | 27,928,867 | chr8:27928867:C:T | t | c | 0.8721 | 0.206 | 2.30E-05 | ++++++++? | 8.496 | 8 | 0.3866 |
| ADNC Score | chr8 | 27,929,726 | chr8:27929726:G:A | a | g | 0.9293 | 0.2084 | 8.20E-06 | ++++++++? | 7.275 | 8 | 0.5072 |
| ADNC Score | chr8 | 27,931,694 | chr8:27931694:C:T | t | c | 0.8954 | 0.205 | 1.26E-05 | ++++++++? | 10.516 | 8 | 0.2307 |
| ADNC Score | chr8 | 27,931,789 | chr8:27931789:G:A | a | g | 0.9254 | 0.2068 | 7.64E-06 | ++++++++? | 9.884 | 7 | 0.1952 |
| ADNC Score | chr8 | 27,933,461 | chr8:27933461:G:A | a | g | -0.7805 | 0.1965 | 7.12E-05 | --------?- | 10.577 | 8 | 0.2269 |
| ADNC Score | chr8 | 27,933,839 | chr8:27933839:A:G | a | g | -0.89 | 0.2032 | 1.18E-05 | --------?- | 10.668 | 8 | 0.2212 |
| ADNC Score | chr8 | 27,934,048 | chr8:27934048:G:A | a | g | 0.8718 | 0.2039 | 1.91E-05 | ++++++++? | 11.096 | 8 | 0.1963 |
| ADNC Score | chr8 | 27,936,912 | chr8:27936912:C:A | a | c | 0.9268 | 0.2036 | 5.34E-06 | ++++++++? | 10.409 | 7 | 0.1665 |
| ADNC Score | chr8 | 27,941,120 | chr8:27941120:G:A | a | g | -0.8505 | 0.199 | 1.92E-05 | --------?- | 10.392 | 8 | 0.2386 |
| ADNC Score | chr8 | 27,943,336 | chr8:27943336:C:T | t | c | 0.9334 | 0.2033 | 4.40E-06 | ++++++++? | 10.083 | 7 | 0.1839 |
| ADNC Score | chr8 | 27,948,548 | chr8:27948548:C:G | c | g | 0.8508 | 0.2013 | 2.38E-05 | ++++++++? | 10.696 | 8 | 0.2195 |
| ADNC Score | chr8 | 27,949,643 | chr8:27949643:G:C | c | g | -0.8265 | 0.1932 | 1.89E-05 | --------?- | 7.388 | 8 | 0.4954 |
| ADNC Score | chr8 | 27,950,976 | chr8:27950976:T:C | t | c | 0.8306 | 0.1974 | 2.58E-05 | ++++++++? | 9.839 | 8 | 0.2765 |
| ADNC Score | chr8 | 27,969,969 | chr8:27969969:C:T | t | c | 0.8597 | 0.1975 | 1.34E-05 | ++++++++? | 7.878 | 7 | 0.3434 |
| ADNC Score | chr8 | 27,971,743 | chr8:27971743:T:C | t | c | -0.8605 | 0.1976 | 1.33E-05 | --------?? | 8.037 | 7 | 0.3294 |
| ADNC Score | chr8 | 27,973,938 | chr8:27973938:C:T | t | c | 0.8279 | 0.196 | 2.39E-05 | ++++++++? | 7.967 | 7 | 0.3355 |
| ADNC Score | chr8 | 39,687,646 | chr8:39687646:G:A | a | g | 0.2163 | 0.0553 | 9.24E-05 | ++++-+-+++ | 14.445 | 9 | 0.1073 |
| ADNC Score | chr8 | 55,300,810 | chr8:55300810:A:G | a | g | -1.5118 | 0.3709 | 4.58E-05 | ?-?-??--?? | 0.63 | 3 | 0.8896 |

| | | | | | | | | | | | | |
|---|---|---|---|---|---|---|---|---|---|---|---|---|
| ADNC Score | chr8 | 55,301,719 | chr8:55301719:T:G | t | g | -1.5118 | 0.3709 | 4.58E-05 | ?-?-??--?? | 0.63 | 3 | 0.8896 |
| ADNC Score | chr8 | 62,979,755 | chr8:62979755:G:C | c | g | 1.5165 | 0.3551 | 1.95E-05 | ?++?+??+? | 0.811 | 3 | 0.8467 |
| ADNC Score | chr8 | 63,153,040 | chr8:63153040:C:G | c | g | -1.1687 | 0.2892 | 5.31E-05 | ?--?-??--? | 4.7 | 4 | 0.3195 |
| ADNC Score | chr8 | 63,283,790 | chr8:63283790:A:G | a | g | -1.2327 | 0.2859 | 1.62E-05 | ?--?-??--? | 7.337 | 4 | 0.1191 |
| ADNC Score | chr8 | 78,848,214 | chr8:78848214:ATC:A | a | atc | -0.4295 | 0.1093 | 8.48E-05 | ---------? | 3.449 | 8 | 0.9031 |
| ADNC Score | chr8 | 105,459,678 | chr8:105459678:G:A | a | g | -0.1906 | 0.0483 | 7.96E-05 | +---+----+ | 6.36 | 9 | 0.7034 |
| ADNC Score | chr8 | 105,462,743 | chr8:105462743:AT:A | a | at | -0.1895 | 0.0483 | 8.85E-05 | +---+----+ | 5.096 | 9 | 0.8258 |
| ADNC Score | chr8 | 105,502,887 | chr8:105502887:C:T | t | c | 0.1909 | 0.0489 | 9.48E-05 | +++++++++ | 5.086 | 9 | 0.8267 |
| ADNC Score | chr8 | 105,505,413 | chr8:105505413:C:T | t | c | 0.1897 | 0.0487 | 9.77E-05 | +++++++++ | 5.178 | 9 | 0.8185 |
| ADNC Score | chr8 | 105,506,982 | chr8:105506982:C:T | t | c | 0.1878 | 0.0481 | 9.58E-05 | +++++++++ | 4.248 | 9 | 0.8943 |
| ADNC Score | chr8 | 124,745,404 | chr8:124745404:A:G | a | g | -0.389 | 0.0937 | 3.32E-05 | -+-------+ | 5.248 | 9 | 0.8121 |
| ADNC Score | chr8 | 124,755,740 | chr8:124755740:A:C | a | c | -0.586 | 0.1355 | 1.53E-05 | -+-------+ | 5.776 | 9 | 0.7621 |
| ADNC Score | chr8 | 124,801,947 | chr8:124801947:G:A | a | g | 0.4093 | 0.1022 | 6.24E-05 | +++++++++ | 2.893 | 9 | 0.9684 |
| ADNC Score | chr8 | 137,231,719 | chr8:137231719:T:A | a | t | 0.7832 | 0.1871 | 2.83E-05 | -+++?++++- | 4.418 | 8 | 0.8176 |
| ADNC Score | chr8 | 137,282,843 | chr8:137282843:GCTT:G | g | gctt | 0.7674 | 0.1899 | 5.35E-05 | -+++?++++- | 6.459 | 8 | 0.596 |
| ADNC Score | chr8 | 142,984,728 | chr8:142984728:A:T | a | t | -0.8626 | 0.1937 | 8.43E-06 | +--------+ | 6.798 | 9 | 0.6581 |
| ADNC Score | chr9 | 1,710,381 | chr9:1710381:A:G | a | g | -0.5583 | 0.1305 | 1.89E-05 | ++----+--- | 5.126 | 9 | 0.8232 |
| ADNC Score | chr9 | 9,795,214 | chr9:9795214:A:G | a | g | -0.4424 | 0.1075 | 3.90E-05 | +-----+--+ | 10.24 | 9 | 0.3314 |
| ADNC Score | chr9 | 9,795,888 | chr9:9795888:G:C | c | g | 0.447 | 0.1095 | 4.45E-05 | -+++++-++- | 11.099 | 9 | 0.269 |
| ADNC Score | chr9 | 9,796,577 | chr9:9796577:A:C | a | c | -0.4298 | 0.1085 | 7.40E-05 | +-----+--+ | 11.229 | 9 | 0.2604 |
| ADNC Score | chr9 | 9,840,407 | chr9:9840407:T:C | t | c | -0.4109 | 0.1048 | 8.78E-05 | +-----+--- | 17.467 | 9 | 0.04189 |
| ADNC Score | chr9 | 14,647,363 | chr9:14647363:T:G | t | g | -0.2397 | 0.0611 | 8.80E-05 | ----+----- | 9.505 | 9 | 0.392 |
| ADNC Score | chr9 | 14,649,730 | chr9:14649730:G:A | a | g | 0.2453 | 0.0624 | 8.34E-05 | ++++-++++- | 9.497 | 9 | 0.3928 |
| ADNC Score | chr9 | 14,656,003 | chr9:14656003:G:GA | g | ga | -0.2553 | 0.0629 | 4.95E-05 | ----+----- | 10.529 | 9 | 0.3094 |
| ADNC Score | chr9 | 14,656,233 | chr9:14656233:G:C | c | g | 0.2492 | 0.0624 | 6.46E-05 | ++++-++++- | 8.805 | 9 | 0.4554 |
| ADNC Score | chr9 | 14,658,114 | chr9:14658114:A:C | a | c | -0.2558 | 0.0614 | 3.07E-05 | ----+----- | 10.436 | 9 | 0.3164 |
| ADNC Score | chr9 | 16,225,447 | chr9:16225447:A:G | a | g | -0.4365 | 0.099 | 1.04E-05 | +--------- | 6.714 | 9 | 0.6669 |
| ADNC Score | chr9 | 16,340,976 | chr9:16340976:C:T | t | c | 0.1953 | 0.0494 | 7.57E-05 | ++++-++-++ | 6.656 | 9 | 0.6729 |
| ADNC Score | chr9 | 23,479,947 | chr9:23479947:AT:A | a | at | -0.4919 | 0.1256 | 9.01E-05 | +-----+--- | 12.588 | 9 | 0.1821 |
| ADNC Score | chr9 | 34,973,810 | chr9:34973810:CT:C | ct | c | -0.6547 | 0.1617 | 5.16E-05 | +---?+---- | 6.818 | 8 | 0.5563 |
| ADNC Score | chr9 | 37,185,317 | chr9:37185317:TC:T | t | tc | 0.3992 | 0.1023 | 9.47E-05 | ++++-++++- | 6.191 | 9 | 0.7206 |
| ADNC Score | chr9 | 79,148,461 | chr9:79148461:G:A | a | g | -0.4739 | 0.1163 | 4.61E-05 | ----+----? | 10.766 | 8 | 0.2153 |
| ADNC Score | chr9 | 79,149,301 | chr9:79149301:A:G | a | g | 0.4993 | 0.1162 | 1.73E-05 | ++++-++++? | 10.325 | 8 | 0.243 |
| ADNC Score | chr9 | 79,153,447 | chr9:79153447:T:C | t | c | 0.4985 | 0.1152 | 1.52E-05 | ++++-++++? | 10.256 | 8 | 0.2475 |
| ADNC Score | chr9 | 83,296,421 | chr9:83296421:T:C | t | c | -0.7875 | 0.1896 | 3.28E-05 | ++--?--+-? | 4.998 | 7 | 0.6602 |
| ADNC Score | chr9 | 91,230,819 | chr9:91230819:G:C | c | g | 1.0587 | 0.2623 | 5.45E-05 | ?-?+??-++? | 7.667 | 4 | 0.1046 |
| ADNC Score | chr9 | 97,867,282 | chr9:97867282:T:C | t | c | -0.3435 | 0.0839 | 4.19E-05 | -------+-- | 8.461 | 9 | 0.4885 |
| ADNC Score | chr9 | 107,578,809 | chr9:107578809:C:T | t | c | 0.3865 | 0.0967 | 6.44E-05 | ++++-++-++ | 10.107 | 9 | 0.3419 |
| ADNC Score | chr9 | 107,580,631 | chr9:107580631:G:A | a | g | 0.3829 | 0.0968 | 7.60E-05 | ++++-++-++ | 10.107 | 9 | 0.3419 |
| ADNC Score | chr9 | 107,587,797 | chr9:107587797:C:T | t | c | 0.3795 | 0.096 | 7.75E-05 | ++++-++-++ | 10.025 | 9 | 0.3485 |
| ADNC Score | chr9 | 116,265,739 | chr9:116265739:A:G | a | g | 0.2525 | 0.0632 | 6.47E-05 | -++++++++- | 6.886 | 9 | 0.649 |
| ADNC Score | chr9 | 116,268,128 | chr9:116268128:C:G | c | g | 0.2539 | 0.0651 | 9.50E-05 | --++++++++ | 8.296 | 9 | 0.5046 |
| ADNC Score | chr9 | 116,268,378 | chr9:116268378:T:G | t | g | 0.2686 | 0.0642 | 2.85E-05 | -++++++++- | 7.977 | 9 | 0.5364 |
| ADNC Score | chr9 | 116,270,846 | chr9:116270846:GA:G | g | ga | -0.2562 | 0.0653 | 8.75E-05 | ++-------- | 9.333 | 9 | 0.4071 |
| ADNC Score | chr9 | 116,270,870 | chr9:116270870:T:A | a | t | -0.2609 | 0.0655 | 6.79E-05 | ++-------- | 8.949 | 9 | 0.442 |
| ADNC Score | chr9 | 116,271,055 | chr9:116271055:C:A | a | c | -0.2634 | 0.0655 | 5.78E-05 | ++-------- | 8.759 | 9 | 0.4598 |

| | | | | | | | | | | | | |
|---|---|---|---|---|---|---|---|---|---|---|---|---|
| ADNC Score | chr9 | 116,271,191 | chr9:116271191:G:A | a | g | -0.2539 | 0.0653 | 9.99E-05 | ++-------- | 9.285 | 9 | 0.4114 |
| ADNC Score | chr9 | 116,272,196 | chr9:116272196:T:C | t | c | 0.25 | 0.0631 | 7.54E-05 | --++++++++ | 8.104 | 9 | 0.5237 |
| ADNC Score | chr9 | 116,273,849 | chr9:116273849:T:C | t | c | 0.2593 | 0.0635 | 4.49E-05 | -++++++++- | 7.521 | 9 | 0.583 |
| ADNC Score | chr9 | 116,275,087 | chr9:116275087:C:T | t | c | -0.2643 | 0.0657 | 5.71E-05 | ++-------- | 8.565 | 9 | 0.4783 |
| ADNC Score | chr9 | 130,495,472 | chr9:130495472:T:C | t | c | 0.2187 | 0.0508 | 1.68E-05 | +-++++-+++ | 11.731 | 9 | 0.2289 |
| Any CBVD | chr1 | 4,348,260 | chr1:4348260:C:T | t | c | 0.4231 | 0.1044 | 5.05E-05 | +-+-+++-+ | 14.33 | 8 | 0.07355 |
| Any CBVD | chr1 | 22,218,298 | chr1:22218298:C:T | t | c | 2.0624 | 0.4827 | 1.93E-05 | ?+++????? | 1.666 | 2 | 0.4347 |
| Any CBVD | chr1 | 29,846,427 | chr1:29846427:C:T | t | c | 1.5742 | 0.3804 | 3.51E-05 | ??++??+?+ | 0.154 | 3 | 0.9846 |
| Any CBVD | chr1 | 42,715,273 | chr1:42715273:G:A | a | g | 0.9932 | 0.2373 | 2.84E-05 | ++++--+++ | 17.395 | 8 | 0.02625 |
| Any CBVD | chr1 | 43,963,230 | chr1:43963230:G:A | a | g | 0.532 | 0.1367 | 9.92E-05 | +-+++--++ | 14.087 | 8 | 0.07953 |
| Any CBVD | chr1 | 48,381,580 | chr1:48381580:C:T | t | c | 1.2089 | 0.2755 | 1.14E-05 | ++++?-+-+ | 10.044 | 7 | 0.1861 |
| Any CBVD | chr1 | 48,840,590 | chr1:48840590:T:C | t | c | -1.1755 | 0.302 | 9.92E-05 | -?--?-??- | 3.781 | 4 | 0.4364 |
| Any CBVD | chr1 | 48,854,995 | chr1:48854995:G:A | a | g | 0.7847 | 0.1966 | 6.57E-05 | ++++?++++ | 9.209 | 7 | 0.238 |
| Any CBVD | chr1 | 49,095,415 | chr1:49095415:C:T | t | c | 1.0355 | 0.2424 | 1.94E-05 | +-+++++++ | 15.662 | 8 | 0.04748 |
| Any CBVD | chr1 | 49,116,271 | chr1:49116271:C:T | t | c | 1.0026 | 0.244 | 3.96E-05 | +-++?++++ | 13.534 | 7 | 0.06012 |
| Any CBVD | chr1 | 49,213,515 | chr1:49213515:A:G | a | g | -1.0555 | 0.2405 | 1.14E-05 | -+------- | 13.258 | 8 | 0.1033 |
| Any CBVD | chr1 | 49,378,716 | chr1:49378716:G:A | a | g | 1.0207 | 0.2392 | 1.98E-05 | +-+++++++ | 11.923 | 8 | 0.1547 |
| Any CBVD | chr1 | 49,528,668 | chr1:49528668:G:T | t | g | 1.1147 | 0.2849 | 9.11E-05 | +?++?+?-+ | 7.293 | 5 | 0.1997 |
| Any CBVD | chr1 | 49,592,644 | chr1:49592644:A:G | a | g | -1.1094 | 0.2447 | 5.79E-06 | -?------- | 10.494 | 7 | 0.1623 |
| Any CBVD | chr1 | 49,721,560 | chr1:49721560:G:A | a | g | 1.1276 | 0.2466 | 4.81E-06 | +?+++++++ | 9.949 | 7 | 0.1915 |
| Any CBVD | chr1 | 49,749,006 | chr1:49749006:GT:G | g | gt | 1.0781 | 0.2439 | 9.88E-06 | +-+++++++ | 9.17 | 8 | 0.3281 |
| Any CBVD | chr1 | 49,768,855 | chr1:49768855:CT:C | ct | c | -1.0641 | 0.2444 | 1.34E-05 | -+------- | 9.701 | 8 | 0.2867 |
| Any CBVD | chr1 | 49,811,830 | chr1:49811830:G:A | a | g | 1.0695 | 0.2441 | 1.18E-05 | +-+++++++ | 9.126 | 8 | 0.3318 |
| Any CBVD | chr1 | 49,812,659 | chr1:49812659:C:T | t | c | 1.1285 | 0.2477 | 5.20E-06 | +?+++++++ | 9.683 | 7 | 0.2073 |
| Any CBVD | chr1 | 56,123,012 | chr1:56123012:T:C | t | c | -2.0631 | 0.5232 | 8.04E-05 | -???+-??? | 3.496 | 2 | 0.1741 |
| Any CBVD | chr1 | 56,131,866 | chr1:56131866:C:CT | ct | c | 1.9741 | 0.4966 | 7.04E-05 | +???-+??? | 2.275 | 2 | 0.3207 |
| Any CBVD | chr1 | 56,136,859 | chr1:56136859:C:T | t | c | 2.0624 | 0.5033 | 4.17E-05 | +???-+??? | 2.667 | 2 | 0.2635 |
| Any CBVD | chr1 | 56,138,196 | chr1:56138196:C:T | t | c | 2.0608 | 0.5031 | 4.21E-05 | +???-+??? | 2.665 | 2 | 0.2638 |
| Any CBVD | chr1 | 56,145,038 | chr1:56145038:T:G | t | g | -2.163 | 0.5312 | 4.66E-05 | -???+-??? | 4.007 | 2 | 0.1349 |
| Any CBVD | chr1 | 56,147,775 | chr1:56147775:G:A | a | g | 2.162 | 0.5314 | 4.73E-05 | +???-+??? | 4.004 | 2 | 0.135 |
| Any CBVD | chr1 | 56,171,157 | chr1:56171157:C:T | t | c | 1.0895 | 0.2729 | 6.55E-05 | +?+?-+?++ | 16.754 | 5 | 0.00499 |
| Any CBVD | chr1 | 56,172,038 | chr1:56172038:G:A | a | g | 1.0951 | 0.2731 | 6.07E-05 | +?+?-+?++ | 17.07 | 5 | 0.004368 |
| Any CBVD | chr1 | 56,172,147 | chr1:56172147:CTTG:C | cttg | c | -1.1696 | 0.2835 | 3.71E-05 | -?-?+-?-- | 18.347 | 5 | 0.002541 |
| Any CBVD | chr1 | 56,173,768 | chr1:56173768:T:A | a | t | 1.0923 | 0.2732 | 6.38E-05 | +?+?-+?++ | 16.928 | 5 | 0.004638 |
| Any CBVD | chr1 | 56,207,925 | chr1:56207925:C:G | c | g | -2.0582 | 0.5184 | 7.18E-05 | -???+-??? | 7.902 | 2 | 0.01924 |
| Any CBVD | chr1 | 58,881,480 | chr1:58881480:C:T | t | c | 0.3261 | 0.0808 | 5.46E-05 | +++++-+-+ | 16.758 | 8 | 0.03273 |
| Any CBVD | chr1 | 58,882,856 | chr1:58882856:G:A | a | g | 0.3353 | 0.0838 | 6.28E-05 | +++++++-+ | 16.477 | 8 | 0.03604 |
| Any CBVD | chr1 | 58,893,049 | chr1:58893049:G:C | c | g | 0.3499 | 0.0856 | 4.34E-05 | +++++++-+ | 19.97 | 8 | 0.01045 |
| Any CBVD | chr1 | 58,894,731 | chr1:58894731:G:A | a | g | 0.3384 | 0.0836 | 5.15E-05 | +++++++-+ | 17.385 | 8 | 0.02635 |
| Any CBVD | chr1 | 58,895,980 | chr1:58895980:C:G | c | g | -0.3321 | 0.0836 | 7.11E-05 | -------+- | 17.261 | 8 | 0.02751 |
| Any CBVD | chr1 | 59,245,647 | chr1:59245647:T:C | t | c | -1.0087 | 0.2349 | 1.76E-05 | ------+-? | 15.802 | 7 | 0.02698 |
| Any CBVD | chr1 | 62,454,885 | chr1:62454885:A:C | a | c | -1.1319 | 0.262 | 1.56E-05 | ----?+-?+ | 20.714 | 6 | 0.002064 |
| Any CBVD | chr1 | 62,506,878 | chr1:62506878:C:T | t | c | 1.2402 | 0.28 | 9.44E-06 | ++++??++- | 19.386 | 6 | 0.003558 |
| Any CBVD | chr1 | 62,555,600 | chr1:62555600:C:T | t | c | 1.2428 | 0.2794 | 8.66E-06 | ++++??++- | 19.102 | 6 | 0.003994 |
| Any CBVD | chr1 | 62,567,432 | chr1:62567432:C:T | t | c | 1.2424 | 0.2794 | 8.70E-06 | ++++??++- | 19.048 | 6 | 0.004083 |
| Any CBVD | chr1 | 62,623,810 | chr1:62623810:T:C | t | c | -1.2358 | 0.2789 | 9.39E-06 | ----??--+ | 18.905 | 6 | 0.004328 |

| | | | | | | | | | | | | |
|---|---|---|---|---|---|---|---|---|---|---|---|---|
| Any CBVD | chr1 | 62,654,501 | chr1:62654501:T:TA | t | ta | -1.1955 | 0.2819 | 2.22E-05 | ----??-?+ | 17.512 | 5 | 0.003625 |
| Any CBVD | chr1 | 69,874,575 | chr1:69874575:C:T | t | c | 0.9169 | 0.1959 | 2.88E-06 | ++++++-++ | 4.122 | 8 | 0.846 |
| Any CBVD | chr1 | 69,927,693 | chr1:69927693:C:A | a | c | 0.7968 | 0.185 | 1.66E-05 | ++++++-++ | 3.584 | 8 | 0.8926 |
| Any CBVD | chr1 | 79,387,706 | chr1:79387706:A:G | a | g | -1.449 | 0.3676 | 8.10E-05 | ?---?--+? | 6.08 | 5 | 0.2985 |
| Any CBVD | chr1 | 96,018,624 | chr1:96018624:TCTC:T | t | tctc | 0.3149 | 0.0808 | 9.78E-05 | ++-+-++++ | 16.253 | 8 | 0.0389 |
| Any CBVD | chr1 | 96,022,650 | chr1:96022650:G:A | a | g | 0.3185 | 0.0809 | 8.21E-05 | ++-+-++++ | 16.184 | 8 | 0.03982 |
| Any CBVD | chr1 | 96,022,813 | chr1:96022813:T:C | t | c | -0.3184 | 0.0809 | 8.24E-05 | --+-+---- | 16.179 | 8 | 0.03989 |
| Any CBVD | chr1 | 96,026,742 | chr1:96026742:A:G | a | g | -0.3232 | 0.0808 | 6.33E-05 | ----+---- | 15.785 | 8 | 0.04556 |
| Any CBVD | chr1 | 96,031,026 | chr1:96031026:A:G | a | g | -0.3175 | 0.0808 | 8.57E-05 | --+-+---- | 16.224 | 8 | 0.03928 |
| Any CBVD | chr1 | 96,039,280 | chr1:96039280:T:G | t | g | -0.3162 | 0.0797 | 7.35E-05 | --+++---- | 17.217 | 8 | 0.02793 |
| Any CBVD | chr1 | 96,040,044 | chr1:96040044:A:G | a | g | -0.3161 | 0.0797 | 7.35E-05 | --+++---- | 17.22 | 8 | 0.0279 |
| Any CBVD | chr1 | 96,040,592 | chr1:96040592:G:T | t | g | 0.3149 | 0.0797 | 7.81E-05 | ++---++++ | 17.172 | 8 | 0.02837 |
| Any CBVD | chr1 | 96,041,520 | chr1:96041520:T:C | t | c | -0.3284 | 0.0789 | 3.16E-05 | --+++---- | 18.234 | 8 | 0.01954 |
| Any CBVD | chr1 | 96,046,541 | chr1:96046541:T:C | t | c | -0.3316 | 0.0804 | 3.74E-05 | ----+---- | 15.358 | 8 | 0.05255 |
| Any CBVD | chr1 | 96,057,060 | chr1:96057060:G:C | c | g | 0.337 | 0.081 | 3.19E-05 | ++++-++++ | 16.798 | 8 | 0.03229 |
| Any CBVD | chr1 | 96,057,426 | chr1:96057426:C:T | t | c | 0.3316 | 0.08 | 3.43E-05 | ++++-++++ | 16.575 | 8 | 0.03485 |
| Any CBVD | chr1 | 96,116,229 | chr1:96116229:C:T | t | c | 0.3171 | 0.0753 | 2.54E-05 | ++++-++++ | 11.836 | 8 | 0.1587 |
| Any CBVD | chr1 | 96,116,862 | chr1:96116862:G:A | a | g | 0.3181 | 0.0754 | 2.43E-05 | ++++-++++ | 11.776 | 8 | 0.1615 |
| Any CBVD | chr1 | 96,119,011 | chr1:96119011:T:C | t | c | -0.3084 | 0.0753 | 4.24E-05 | ----+---- | 11.729 | 8 | 0.1637 |
| Any CBVD | chr1 | 96,122,024 | chr1:96122024:T:C | t | c | -0.3083 | 0.0754 | 4.30E-05 | ----+---- | 11.666 | 8 | 0.1668 |
| Any CBVD | chr1 | 96,128,058 | chr1:96128058:G:A | a | g | 0.3032 | 0.0756 | 6.03E-05 | ++++-++++ | 11.291 | 8 | 0.1858 |
| Any CBVD | chr1 | 98,361,937 | chr1:98361937:T:C | t | c | 0.2383 | 0.0565 | 2.43E-05 | ++++++-++ | 9.091 | 8 | 0.3347 |
| Any CBVD | chr1 | 98,377,414 | chr1:98377414:T:C | t | c | 0.2535 | 0.0619 | 4.19E-05 | +++++++++ | 4.214 | 8 | 0.8374 |
| Any CBVD | chr1 | 147,891,874 | chr1:147891874:A:G | a | g | -0.3039 | 0.0774 | 8.60E-05 | ------?-- | 3.397 | 7 | 0.846 |
| Any CBVD | chr1 | 163,878,354 | chr1:163878354:G:A | a | g | 1.3493 | 0.3359 | 5.89E-05 | +?+++?--? | 6.703 | 5 | 0.2437 |
| Any CBVD | chr1 | 163,878,355 | chr1:163878355:C:A | a | c | 1.3494 | 0.3359 | 5.88E-05 | +?+++?--? | 6.703 | 5 | 0.2437 |
| Any CBVD | chr1 | 178,555,324 | chr1:178555324:A:G | a | g | -0.7115 | 0.1729 | 3.88E-05 | -------+- | 3.713 | 8 | 0.882 |
| Any CBVD | chr1 | 196,078,702 | chr1:196078702:A:T | a | t | -0.6748 | 0.1524 | 9.57E-06 | --------- | 8.726 | 8 | 0.3659 |
| Any CBVD | chr1 | 196,081,346 | chr1:196081346:G:A | a | g | 0.6757 | 0.152 | 8.84E-06 | +++++++++ | 8.893 | 8 | 0.3514 |
| Any CBVD | chr1 | 196,149,336 | chr1:196149336:G:A | a | g | 0.8326 | 0.1907 | 1.27E-05 | +-+++++++ | 9.315 | 8 | 0.3164 |
| Any CBVD | chr1 | 196,166,295 | chr1:196166295:G:A | a | g | 0.7547 | 0.1568 | 1.48E-06 | +-+++++++ | 7.646 | 8 | 0.4688 |
| Any CBVD | chr1 | 196,197,372 | chr1:196197372:A:C | a | c | -0.8234 | 0.1875 | 1.13E-05 | -+------- | 8.469 | 8 | 0.389 |
| Any CBVD | chr1 | 196,972,144 | chr1:196972144:C:T | t | c | 0.5413 | 0.1391 | 9.95E-05 | +-++--+++ | 24.887 | 8 | 0.001624 |
| Any CBVD | chr1 | 200,932,288 | chr1:200932288:A:C | a | c | 0.4671 | 0.12 | 9.94E-05 | +-+++++++ | 7.614 | 8 | 0.4721 |
| Any CBVD | chr1 | 204,260,078 | chr1:204260078:T:G | t | g | -2.9349 | 0.6786 | 1.53E-05 | -???--??? | 4.118 | 2 | 0.1276 |
| Any CBVD | chr1 | 204,341,238 | chr1:204341238:G:A | a | g | 1.0815 | 0.2499 | 1.51E-05 | ++++-+--+ | 6.583 | 8 | 0.5822 |
| Any CBVD | chr1 | 216,059,034 | chr1:216059034:T:C | t | c | -1.3692 | 0.3442 | 6.96E-05 | -?--???+- | 11.223 | 4 | 0.02416 |
| Any CBVD | chr1 | 224,946,402 | chr1:224946402:T:G | t | g | -0.7243 | 0.1812 | 6.43E-05 | -+--+-+-- | 12.703 | 8 | 0.1225 |
| Any CBVD | chr1 | 231,929,385 | chr1:231929385:G:A | a | g | 1.3363 | 0.3281 | 4.65E-05 | ++-+?++?? | 6.771 | 5 | 0.2382 |
| Any CBVD | chr1 | 241,919,576 | chr1:241919576:A:G | a | g | -0.2495 | 0.0616 | 5.09E-05 | --------- | 4.294 | 8 | 0.8296 |
| Any CBVD | chr1 | 241,919,578 | chr1:241919578:C:A | a | c | 0.2499 | 0.062 | 5.55E-05 | +++++++++ | 4.367 | 8 | 0.8226 |
| Any CBVD | chr1 | 241,919,751 | chr1:241919751:C:T | t | c | 0.2556 | 0.0616 | 3.38E-05 | +++++++++ | 4.428 | 8 | 0.8166 |
| Any CBVD | chr1 | 244,000,236 | chr1:244000236:A:G | a | g | -0.3598 | 0.0923 | 9.63E-05 | ---+----- | 11.628 | 8 | 0.1686 |
| Any CBVD | chr1 | 244,005,653 | chr1:244005653:T:C | t | c | -0.364 | 0.0918 | 7.32E-05 | ---+----- | 10.887 | 8 | 0.2082 |
| Any CBVD | chr1 | 244,006,881 | chr1:244006881:A:C | a | c | -0.3634 | 0.0919 | 7.74E-05 | ---+----- | 10.813 | 8 | 0.2125 |
| Any CBVD | chr1 | 244,007,727 | chr1:244007727:T:C | t | c | -0.3614 | 0.092 | 8.52E-05 | ---+----- | 10.81 | 8 | 0.2127 |

| | | | | | | | | | | | | |
|---|---|---|---|---|---|---|---|---|---|---|---|---|
| Any CBVD | chr1 | 244,007,865 | chr1:244007865:G:A | a | g | 0.3631 | 0.0919 | 7.83E-05 | +++-+++++ | 10.862 | 8 | 0.2096 |
| Any CBVD | chr1 | 244,008,058 | chr1:244008058:G:A | a | g | 0.3619 | 0.092 | 8.36E-05 | +++-+++++ | 10.789 | 8 | 0.214 |
| Any CBVD | chr1 | 244,008,061 | chr1:244008061:A:G | a | g | -0.3619 | 0.092 | 8.37E-05 | ---+----- | 10.788 | 8 | 0.214 |
| Any CBVD | chr1 | 244,008,206 | chr1:244008206:A:G | a | g | -0.3615 | 0.0919 | 8.45E-05 | ---+----- | 10.806 | 8 | 0.213 |
| Any CBVD | chr1 | 244,008,282 | chr1:244008282:G:A | a | g | 0.3619 | 0.092 | 8.39E-05 | +++-+++++ | 10.785 | 8 | 0.2142 |
| Any CBVD | chr1 | 244,008,413 | chr1:244008413:G:A | a | g | 0.3604 | 0.092 | 8.89E-05 | +++-+++++ | 10.883 | 8 | 0.2084 |
| Any CBVD | chr1 | 244,008,463 | chr1:244008463:T:C | t | c | -0.3619 | 0.092 | 8.37E-05 | ---+----- | 10.788 | 8 | 0.214 |
| Any CBVD | chr1 | 244,008,828 | chr1:244008828:G:T | t | g | 0.3619 | 0.092 | 8.37E-05 | +++-+++++ | 10.789 | 8 | 0.214 |
| Any CBVD | chr1 | 244,009,211 | chr1:244009211:C:A | a | c | 0.362 | 0.092 | 8.35E-05 | +++-+++++ | 10.788 | 8 | 0.214 |
| Any CBVD | chr10 | 3,473,392 | chr10:3473392:C:T | t | c | 2.8613 | 0.7206 | 7.17E-05 | ?+??++??? | 6.101 | 2 | 0.04733 |
| Any CBVD | chr10 | 3,473,511 | chr10:3473511:G:A | a | g | 2.8078 | 0.7144 | 8.48E-05 | ?+??++??? | 6.456 | 2 | 0.03964 |
| Any CBVD | chr10 | 3,475,686 | chr10:3475686:C:T | t | c | 2.8612 | 0.7206 | 7.18E-05 | ?+??++??? | 6.102 | 2 | 0.04732 |
| Any CBVD | chr10 | 3,477,206 | chr10:3477206:G:T | t | g | 2.9335 | 0.7275 | 5.52E-05 | ?+??++??? | 6.609 | 2 | 0.03672 |
| Any CBVD | chr10 | 3,479,160 | chr10:3479160:G:T | t | g | 2.9335 | 0.7275 | 5.52E-05 | ?+??++??? | 6.609 | 2 | 0.03672 |
| Any CBVD | chr10 | 3,479,471 | chr10:3479471:G:A | a | g | 2.9289 | 0.7275 | 5.67E-05 | ?+??++??? | 6.595 | 2 | 0.03698 |
| Any CBVD | chr10 | 3,479,472 | chr10:3479472:C:A | a | c | 2.9335 | 0.7275 | 5.52E-05 | ?+??++??? | 6.609 | 2 | 0.03672 |
| Any CBVD | chr10 | 6,224,993 | chr10:6224993:G:T | t | g | 0.2593 | 0.0651 | 6.76E-05 | +-+++++++ | 7.647 | 8 | 0.4686 |
| Any CBVD | chr10 | 23,369,917 | chr10:23369917:G:A | a | g | 3.7862 | 0.9238 | 4.16E-05 | +???++??? | 1.822 | 2 | 0.4021 |
| Any CBVD | chr10 | 23,372,406 | chr10:23372406:G:A | a | g | 3.6695 | 0.9184 | 6.45E-05 | +???++??? | 2.064 | 2 | 0.3562 |
| Any CBVD | chr10 | 23,372,464 | chr10:23372464:C:T | t | c | 3.7825 | 0.9238 | 4.23E-05 | +???++??? | 1.836 | 2 | 0.3994 |
| Any CBVD | chr10 | 54,113,347 | chr10:54113347:A:G | a | g | -1.0228 | 0.2606 | 8.67E-05 | ----?--?- | 10.563 | 6 | 0.1029 |
| Any CBVD | chr10 | 55,067,809 | chr10:55067809:T:G | t | g | -0.8558 | 0.2145 | 6.64E-05 | -------+- | 2.897 | 8 | 0.9406 |
| Any CBVD | chr10 | 58,385,319 | chr10:58385319:A:G | a | g | 0.2443 | 0.0573 | 2.00E-05 | +++++++++ | 2.295 | 8 | 0.9706 |
| Any CBVD | chr10 | 58,387,510 | chr10:58387510:G:A | a | g | -0.2337 | 0.0569 | 4.05E-05 | -+------- | 3.369 | 8 | 0.9091 |
| Any CBVD | chr10 | 58,388,024 | chr10:58388024:G:A | a | g | -0.2332 | 0.0569 | 4.22E-05 | -+------- | 3.374 | 8 | 0.9088 |
| Any CBVD | chr10 | 58,388,932 | chr10:58388932:A:G | a | g | 0.2341 | 0.0567 | 3.70E-05 | +-+++++++ | 3.364 | 8 | 0.9095 |
| Any CBVD | chr10 | 58,391,266 | chr10:58391266:A:G | a | g | 0.2339 | 0.0564 | 3.41E-05 | +-+++++++ | 3.535 | 8 | 0.8965 |
| Any CBVD | chr10 | 58,396,824 | chr10:58396824:G:A | a | g | -0.2353 | 0.0568 | 3.41E-05 | -+------- | 3.535 | 8 | 0.8965 |
| Any CBVD | chr10 | 58,397,579 | chr10:58397579:C:T | t | c | -0.2348 | 0.0569 | 3.68E-05 | -+------- | 3.411 | 8 | 0.906 |
| Any CBVD | chr10 | 58,401,412 | chr10:58401412:G:A | a | g | -0.2303 | 0.0569 | 5.20E-05 | -+------- | 3.514 | 8 | 0.8981 |
| Any CBVD | chr10 | 58,401,880 | chr10:58401880:A:G | a | g | 0.235 | 0.0569 | 3.64E-05 | +-+++++++ | 3.4 | 8 | 0.9068 |
| Any CBVD | chr10 | 58,459,800 | chr10:58459800:G:A | a | g | 0.2389 | 0.0586 | 4.54E-05 | +-+++++++ | 5.387 | 8 | 0.7155 |
| Any CBVD | chr10 | 72,069,904 | chr10:72069904:C:T | t | c | 0.9563 | 0.2284 | 2.82E-05 | +++++++-+ | 16.651 | 8 | 0.03396 |
| Any CBVD | chr10 | 112,126,267 | chr10:112126267:G:A | a | g | 0.9584 | 0.242 | 7.48E-05 | +++++++++ | 8.557 | 8 | 0.381 |
| Any CBVD | chr10 | 124,562,674 | chr10:124562674:G:A | a | g | 1.4651 | 0.3663 | 6.35E-05 | ??+++?-?+ | 6.867 | 4 | 0.1431 |
| Any CBVD | chr10 | 124,589,191 | chr10:124589191:A:G | a | g | -1.5977 | 0.4063 | 8.42E-05 | ??-?-???- | 2.821 | 2 | 0.2441 |
| Any CBVD | chr10 | 129,672,079 | chr10:129672079:C:G | c | g | -1.0015 | 0.2424 | 3.61E-05 | -+-+--+-- | 24.512 | 8 | 0.00188 |
| Any CBVD | chr11 | 5,751,194 | chr11:5751194:TTA:T | t | tta | 0.2831 | 0.0695 | 4.67E-05 | +++-+++++ | 6.846 | 8 | 0.5534 |
| Any CBVD | chr11 | 6,052,819 | chr11:6052819:A:G | a | g | -0.4572 | 0.1067 | 1.82E-05 | --------- | 2.377 | 8 | 0.9672 |
| Any CBVD | chr11 | 23,650,092 | chr11:23650092:T:G | t | g | -1.4554 | 0.3722 | 9.23E-05 | ??--?-??- | 4.424 | 3 | 0.2192 |
| Any CBVD | chr11 | 24,957,220 | chr11:24957220:C:A | a | c | 1.1873 | 0.2985 | 6.96E-05 | ++++-+?-? | 5.77 | 6 | 0.4495 |
| Any CBVD | chr11 | 24,961,633 | chr11:24961633:G:A | a | g | 1.2495 | 0.3023 | 3.58E-05 | ++++-+?-? | 6.116 | 6 | 0.4103 |
| Any CBVD | chr11 | 24,963,451 | chr11:24963451:A:C | a | c | -1.1906 | 0.2635 | 6.26E-06 | ----+-?+- | 4.337 | 7 | 0.7402 |
| Any CBVD | chr11 | 24,970,741 | chr11:24970741:C:T | t | c | 1.198 | 0.2973 | 5.59E-05 | ++++-+?-? | 5.846 | 6 | 0.4406 |
| Any CBVD | chr11 | 24,973,833 | chr11:24973833:G:A | a | g | 1.2039 | 0.2625 | 4.53E-06 | ++++-+?-+ | 4.363 | 7 | 0.7372 |
| Any CBVD | chr11 | 24,973,869 | chr11:24973869:AGAGT:A | a | agagt | 1.1979 | 0.2973 | 5.59E-05 | ++++-+?-? | 5.845 | 6 | 0.4407 |

| | | | | | | | | | | | | |
|---|---|---|---|---|---|---|---|---|---|---|---|---|
| Any CBVD | chr11 | 24,985,813 | chr11:24985813:A:C | a | c | -1.2573 | 0.3006 | 2.88E-05 | ----+-?+? | 5.9 | 6 | 0.4345 |
| Any CBVD | chr11 | 24,991,724 | chr11:24991724:A:G | a | g | -1.2764 | 0.2658 | 1.56E-06 | ----+-?+- | 4.229 | 7 | 0.753 |
| Any CBVD | chr11 | 24,992,247 | chr11:24992247:T:C | t | c | -1.261 | 0.2649 | 1.93E-06 | ----+-?+- | 4.329 | 7 | 0.7412 |
| Any CBVD | chr11 | 25,002,543 | chr11:25002543:C:T | t | c | 1.2759 | 0.2934 | 1.37E-05 | ++++++?-? | 3.866 | 6 | 0.6948 |
| Any CBVD | chr11 | 25,004,162 | chr11:25004162:G:A | a | g | 1.2714 | 0.2929 | 1.43E-05 | ++++++?-? | 3.908 | 6 | 0.6891 |
| Any CBVD | chr11 | 25,008,061 | chr11:25008061:C:A | a | c | 1.2679 | 0.2929 | 1.50E-05 | ++++++?-? | 4.081 | 6 | 0.6658 |
| Any CBVD | chr11 | 25,011,309 | chr11:25011309:T:C | t | c | -1.21 | 0.2945 | 3.99E-05 | ----+-?+? | 4.691 | 6 | 0.584 |
| Any CBVD | chr11 | 25,016,972 | chr11:25016972:G:A | a | g | 1.4148 | 0.3088 | 4.62E-06 | ++++-+?-? | 7.447 | 6 | 0.2815 |
| Any CBVD | chr11 | 25,022,821 | chr11:25022821:C:A | a | c | 1.2668 | 0.2992 | 2.30E-05 | ++++-+?-? | 6.101 | 6 | 0.4119 |
| Any CBVD | chr11 | 25,025,003 | chr11:25025003:C:T | t | c | 1.2655 | 0.2991 | 2.33E-05 | ++++-+?-? | 6.079 | 6 | 0.4144 |
| Any CBVD | chr11 | 25,038,021 | chr11:25038021:T:C | t | c | 1.1467 | 0.277 | 3.48E-05 | ++++-+-+? | 8.377 | 7 | 0.3005 |
| Any CBVD | chr11 | 25,039,911 | chr11:25039911:T:G | t | g | -1.3359 | 0.3021 | 9.76E-06 | ----+-?+? | 6.44 | 6 | 0.3757 |
| Any CBVD | chr11 | 44,529,871 | chr11:44529871:A:G | a | g | -0.2614 | 0.0665 | 8.53E-05 | ------++- | 9.558 | 8 | 0.2974 |
| Any CBVD | chr11 | 47,820,165 | chr11:47820165:G:A | a | g | 0.5787 | 0.1459 | 7.32E-05 | +++++++++ | 4.252 | 8 | 0.8337 |
| Any CBVD | chr11 | 47,907,899 | chr11:47907899:C:T | t | c | 0.5754 | 0.1454 | 7.59E-05 | +++++++++ | 4.844 | 8 | 0.7741 |
| Any CBVD | chr11 | 48,543,119 | chr11:48543119:C:G | c | g | -1.6489 | 0.4191 | 8.35E-05 | --????-?? | 1.414 | 2 | 0.493 |
| Any CBVD | chr11 | 48,543,120 | chr11:48543120:T:A | a | t | 1.6526 | 0.4191 | 8.04E-05 | ++????+?? | 1.407 | 2 | 0.495 |
| Any CBVD | chr11 | 48,659,892 | chr11:48659892:G:C | c | g | 1.6535 | 0.4207 | 8.48E-05 | ++????+?? | 1.537 | 2 | 0.4638 |
| Any CBVD | chr11 | 65,025,091 | chr11:65025091:A:G | a | g | -0.403 | 0.1033 | 9.57E-05 | --------- | 10.68 | 8 | 0.2205 |
| Any CBVD | chr11 | 65,025,679 | chr11:65025679:ACCACT:A | a | accact | 0.4085 | 0.1035 | 7.89E-05 | +++++++++ | 10.592 | 8 | 0.2259 |
| Any CBVD | chr11 | 65,025,717 | chr11:65025717:C:T | t | c | 0.4059 | 0.1033 | 8.57E-05 | +++++++++ | 10.631 | 8 | 0.2235 |
| Any CBVD | chr11 | 65,026,393 | chr11:65026393:C:T | t | c | 0.4056 | 0.1033 | 8.67E-05 | +++++++++ | 10.636 | 8 | 0.2232 |
| Any CBVD | chr11 | 65,026,488 | chr11:65026488:A:G | a | g | -0.4075 | 0.1031 | 7.71E-05 | --------- | 10.529 | 8 | 0.2298 |
| Any CBVD | chr11 | 65,027,064 | chr11:65027064:C:G | c | g | -0.407 | 0.1032 | 7.98E-05 | --------- | 10.623 | 8 | 0.224 |
| Any CBVD | chr11 | 65,029,637 | chr11:65029637:T:C | t | c | -0.4366 | 0.1051 | 3.26E-05 | ----+---- | 13.428 | 8 | 0.09795 |
| Any CBVD | chr11 | 65,030,091 | chr11:65030091:G:T | t | g | 0.4386 | 0.1052 | 3.03E-05 | ++++-++++ | 13.376 | 8 | 0.09956 |
| Any CBVD | chr11 | 65,030,304 | chr11:65030304:G:A | a | g | 0.4435 | 0.1054 | 2.56E-05 | ++++-++++ | 13.504 | 8 | 0.09564 |
| Any CBVD | chr11 | 65,030,591 | chr11:65030591:T:C | t | c | -0.4385 | 0.1051 | 3.02E-05 | ----+---- | 13.099 | 8 | 0.1085 |
| Any CBVD | chr11 | 65,030,918 | chr11:65030918:A:T | a | t | -0.4386 | 0.1051 | 3.04E-05 | ----+---- | 13.366 | 8 | 0.09987 |
| Any CBVD | chr11 | 65,031,156 | chr11:65031156:A:G | a | g | -0.4386 | 0.1051 | 3.03E-05 | ----+---- | 13.367 | 8 | 0.09982 |
| Any CBVD | chr11 | 65,031,576 | chr11:65031576:T:G | t | g | -0.4389 | 0.1052 | 3.01E-05 | ----+---- | 13.371 | 8 | 0.09972 |
| Any CBVD | chr11 | 65,032,351 | chr11:65032351:G:A | a | g | 0.446 | 0.1051 | 2.19E-05 | ++++-++++ | 13.082 | 8 | 0.1091 |
| Any CBVD | chr11 | 65,032,369 | chr11:65032369:A:G | a | g | -0.4436 | 0.105 | 2.38E-05 | ----+---- | 13.14 | 8 | 0.1071 |
| Any CBVD | chr11 | 65,032,419 | chr11:65032419:T:C | t | c | -0.4419 | 0.105 | 2.55E-05 | ----+---- | 13.136 | 8 | 0.1073 |
| Any CBVD | chr11 | 65,035,734 | chr11:65035734:C:T | t | c | 0.4369 | 0.105 | 3.14E-05 | ++++-++++ | 14.619 | 8 | 0.067 |
| Any CBVD | chr11 | 65,036,165 | chr11:65036165:A:G | a | g | -0.4195 | 0.1047 | 6.19E-05 | ----+---- | 14.471 | 8 | 0.07029 |
| Any CBVD | chr11 | 65,036,320 | chr11:65036320:G:C | c | g | 0.4374 | 0.1047 | 2.94E-05 | ++++-++++ | 14.785 | 8 | 0.06347 |
| Any CBVD | chr11 | 65,036,766 | chr11:65036766:C:T | t | c | 0.419 | 0.1048 | 6.39E-05 | ++++-++++ | 14.549 | 8 | 0.06853 |
| Any CBVD | chr11 | 65,036,808 | chr11:65036808:G:T | t | g | 0.4188 | 0.1047 | 6.37E-05 | ++++-++++ | 14.56 | 8 | 0.06828 |
| Any CBVD | chr11 | 65,038,798 | chr11:65038798:C:T | t | c | 0.4181 | 0.1047 | 6.50E-05 | ++++-++++ | 14.411 | 8 | 0.07167 |
| Any CBVD | chr11 | 65,039,026 | chr11:65039026:C:T | t | c | 0.4192 | 0.1047 | 6.26E-05 | ++++-++++ | 14.499 | 8 | 0.06966 |
| Any CBVD | chr11 | 65,039,533 | chr11:65039533:T:A | a | t | 0.4273 | 0.1049 | 4.63E-05 | ++++-++++ | 14.35 | 8 | 0.07308 |
| Any CBVD | chr11 | 65,039,664 | chr11:65039664:G:A | a | g | 0.4273 | 0.1049 | 4.63E-05 | ++++-++++ | 14.353 | 8 | 0.07302 |
| Any CBVD | chr11 | 65,039,763 | chr11:65039763:C:T | t | c | 0.4261 | 0.1048 | 4.81E-05 | ++++-++++ | 14.31 | 8 | 0.07404 |
| Any CBVD | chr11 | 65,039,806 | chr11:65039806:A:C | a | c | -0.4276 | 0.1049 | 4.60E-05 | ----+---- | 14.346 | 8 | 0.07317 |
| Any CBVD | chr11 | 65,039,975 | chr11:65039975:A:C | a | c | 0.4294 | 0.1049 | 4.29E-05 | ++++-++++ | 14.363 | 8 | 0.07277 |

| | | | | | | | | | | | | |
|---|---|---|---|---|---|---|---|---|---|---|---|---|
| Any CBVD | chr11 | 65,040,740 | chr11:65040740:T:TC | t | tc | 0.4298 | 0.1046 | 3.99E-05 | ++++-++++ | 13.935 | 8 | 0.08348 |
| Any CBVD | chr11 | 65,041,175 | chr11:65041175:A:C | a | c | 0.4326 | 0.1051 | 3.84E-05 | ++++-++++ | 14.402 | 8 | 0.07187 |
| Any CBVD | chr11 | 65,041,661 | chr11:65041661:T:G | t | g | 0.4302 | 0.105 | 4.15E-05 | ++++-++++ | 14.267 | 8 | 0.07506 |
| Any CBVD | chr11 | 72,713,589 | chr11:72713589:T:C | t | c | 0.3095 | 0.0786 | 8.25E-05 | +++++++++ | 6.504 | 8 | 0.5909 |
| Any CBVD | chr11 | 72,718,534 | chr11:72718534:G:A | a | g | -0.3148 | 0.0797 | 7.75E-05 | --------- | 8.687 | 8 | 0.3694 |
| Any CBVD | chr11 | 72,718,751 | chr11:72718751:TGCA:T | t | tgca | -0.3186 | 0.0798 | 6.48E-05 | --------- | 8.483 | 8 | 0.3878 |
| Any CBVD | chr11 | 72,744,840 | chr11:72744840:C:T | t | c | -0.312 | 0.0801 | 9.81E-05 | --------- | 9.846 | 8 | 0.276 |
| Any CBVD | chr11 | 72,749,353 | chr11:72749353:A:C | a | c | 0.3099 | 0.0795 | 9.60E-05 | +++++++++ | 10.948 | 8 | 0.2047 |
| Any CBVD | chr11 | 72,749,649 | chr11:72749649:T:C | t | c | 0.3098 | 0.0795 | 9.66E-05 | +++++++++ | 10.941 | 8 | 0.2051 |
| Any CBVD | chr11 | 72,749,717 | chr11:72749717:A:T | a | t | 0.3092 | 0.0795 | 0.0001 | +++++++++ | 10.995 | 8 | 0.202 |
| Any CBVD | chr11 | 72,750,767 | chr11:72750767:A:AG | a | ag | 0.3098 | 0.0795 | 9.67E-05 | +++++++++ | 10.952 | 8 | 0.2044 |
| Any CBVD | chr11 | 72,762,959 | chr11:72762959:G:T | t | g | -0.3197 | 0.0764 | 2.85E-05 | --+------ | 12.355 | 8 | 0.136 |
| Any CBVD | chr11 | 72,763,526 | chr11:72763526:G:C | c | g | -0.3072 | 0.0757 | 4.97E-05 | --+------ | 10.29 | 8 | 0.2452 |
| Any CBVD | chr11 | 72,763,794 | chr11:72763794:A:G | a | g | 0.3096 | 0.0757 | 4.31E-05 | ++-++++++ | 11.428 | 8 | 0.1786 |
| Any CBVD | chr11 | 72,767,275 | chr11:72767275:G:A | a | g | -0.3052 | 0.076 | 5.90E-05 | --+------ | 10.682 | 8 | 0.2204 |
| Any CBVD | chr11 | 72,789,957 | chr11:72789957:C:T | t | c | -0.3133 | 0.0761 | 3.81E-05 | --------- | 10.454 | 8 | 0.2346 |
| Any CBVD | chr11 | 72,793,927 | chr11:72793927:G:C | c | g | -0.3142 | 0.076 | 3.54E-05 | --------- | 10.327 | 8 | 0.2429 |
| Any CBVD | chr11 | 72,795,835 | chr11:72795835:T:C | t | c | 0.3154 | 0.076 | 3.30E-05 | +++++++++ | 10.295 | 8 | 0.2449 |
| Any CBVD | chr11 | 72,800,476 | chr11:72800476:A:C | a | c | 0.3169 | 0.076 | 3.01E-05 | +++++++++ | 10.385 | 8 | 0.2391 |
| Any CBVD | chr11 | 72,803,458 | chr11:72803458:C:T | t | c | -0.3165 | 0.0759 | 3.07E-05 | --------- | 10.374 | 8 | 0.2398 |
| Any CBVD | chr11 | 72,811,097 | chr11:72811097:G:C | c | g | -0.3127 | 0.076 | 3.85E-05 | --------- | 9.886 | 8 | 0.2731 |
| Any CBVD | chr11 | 72,844,508 | chr11:72844508:G:A | a | g | -0.3318 | 0.0741 | 7.53E-06 | --------- | 8.122 | 8 | 0.4216 |
| Any CBVD | chr11 | 72,868,690 | chr11:72868690:AAAAC:A | a | aaaac | -0.3322 | 0.0742 | 7.63E-06 | --------- | 11.806 | 8 | 0.1601 |
| Any CBVD | chr11 | 72,877,132 | chr11:72877132:G:A | a | g | -0.3314 | 0.074 | 7.60E-06 | --------- | 7.998 | 8 | 0.4336 |
| Any CBVD | chr11 | 72,904,024 | chr11:72904024:C:G | c | g | 0.3243 | 0.0763 | 2.14E-05 | +++++++++ | 9.65 | 8 | 0.2904 |
| Any CBVD | chr11 | 72,908,815 | chr11:72908815:G:A | a | g | -0.3173 | 0.0762 | 3.17E-05 | --------- | 9.449 | 8 | 0.3058 |
| Any CBVD | chr11 | 72,918,901 | chr11:72918901:A:G | a | g | 0.3344 | 0.0739 | 5.95E-06 | +++++++++ | 8.419 | 8 | 0.3937 |
| Any CBVD | chr11 | 72,938,351 | chr11:72938351:A:T | a | t | 0.3289 | 0.0738 | 8.32E-06 | +++++++++ | 7.861 | 8 | 0.4472 |
| Any CBVD | chr11 | 72,958,732 | chr11:72958732:T:C | t | c | 0.3155 | 0.0757 | 3.11E-05 | +++++++++ | 9.059 | 8 | 0.3374 |
| Any CBVD | chr11 | 72,996,396 | chr11:72996396:A:C | a | c | 0.3334 | 0.0738 | 6.18E-06 | +++++++++ | 8.193 | 8 | 0.4148 |
| Any CBVD | chr11 | 73,010,226 | chr11:73010226:C:A | a | c | -0.3194 | 0.0763 | 2.85E-05 | --------- | 9.57 | 8 | 0.2965 |
| Any CBVD | chr11 | 73,016,795 | chr11:73016795:A:G | a | g | 0.3336 | 0.0736 | 5.83E-06 | +++++++++ | 8.306 | 8 | 0.4042 |
| Any CBVD | chr11 | 73,099,280 | chr11:73099280:CA:C | ca | c | 0.3125 | 0.0768 | 4.75E-05 | +++++++++ | 8.593 | 8 | 0.3778 |
| Any CBVD | chr11 | 73,140,418 | chr11:73140418:C:T | t | c | 0.2893 | 0.0734 | 8.11E-05 | +++++++++ | 9.042 | 8 | 0.3388 |
| Any CBVD | chr11 | 76,327,090 | chr11:76327090:C:T | t | c | 0.2249 | 0.0576 | 9.54E-05 | ++++-++++ | 7.813 | 8 | 0.452 |
| Any CBVD | chr11 | 76,327,126 | chr11:76327126:G:A | a | g | 0.2224 | 0.057 | 9.49E-05 | ++++-++++ | 8.52 | 8 | 0.3844 |
| Any CBVD | chr11 | 76,345,852 | chr11:76345852:A:G | a | g | -0.229 | 0.0568 | 5.47E-05 | ----+---- | 9.319 | 8 | 0.3161 |
| Any CBVD | chr11 | 76,346,843 | chr11:76346843:A:C | a | c | -0.2417 | 0.0576 | 2.71E-05 | ----+---- | 8.043 | 8 | 0.4293 |
| Any CBVD | chr11 | 76,347,036 | chr11:76347036:G:T | t | g | 0.2422 | 0.0576 | 2.62E-05 | ++++-++++ | 8.043 | 8 | 0.4292 |
| Any CBVD | chr11 | 76,348,185 | chr11:76348185:T:C | t | c | -0.2292 | 0.057 | 5.77E-05 | ----+---- | 8.596 | 8 | 0.3775 |
| Any CBVD | chr11 | 76,354,148 | chr11:76354148:T:C | t | c | -0.2421 | 0.0576 | 2.63E-05 | ----+---- | 7.754 | 8 | 0.4579 |
| Any CBVD | chr11 | 76,357,536 | chr11:76357536:C:T | t | c | 0.2431 | 0.0576 | 2.46E-05 | ++++-++++ | 7.979 | 8 | 0.4355 |
| Any CBVD | chr11 | 76,363,149 | chr11:76363149:A:T | a | t | -0.2274 | 0.0571 | 6.84E-05 | ----+---- | 9.062 | 8 | 0.3371 |
| Any CBVD | chr11 | 76,363,757 | chr11:76363757:T:A | a | t | 0.2442 | 0.0577 | 2.34E-05 | ++++-++++ | 8.574 | 8 | 0.3795 |
| Any CBVD | chr11 | 76,367,563 | chr11:76367563:T:C | t | c | -0.2302 | 0.0571 | 5.53E-05 | ----+---- | 10.632 | 8 | 0.2234 |
| Any CBVD | chr11 | 76,369,022 | chr11:76369022:A:G | a | g | -0.2454 | 0.0576 | 2.06E-05 | ----+---- | 9.862 | 8 | 0.2748 |

| | | | | | | | | | | | | |
|---|---|---|---|---|---|---|---|---|---|---|---|---|
| Any CBVD | chr11 | 76,372,527 | chr11:76372527:A:G | a | g | -0.2279 | 0.0572 | 6.68E-05 | ----+---- | 9.184 | 8 | 0.327 |
| Any CBVD | chr11 | 76,378,724 | chr11:76378724:G:A | a | g | 0.2477 | 0.0578 | 1.82E-05 | ++++-++++ | 8.496 | 8 | 0.3866 |
| Any CBVD | chr11 | 77,558,615 | chr11:77558615:C:T | t | c | 1.3537 | 0.2955 | 4.64E-06 | ++++-++?? | 12.599 | 6 | 0.04986 |
| Any CBVD | chr11 | 77,992,610 | chr11:77992610:C:A | a | c | 1.4179 | 0.3148 | 6.66E-06 | ++++?+??? | 5.616 | 4 | 0.2297 |
| Any CBVD | chr11 | 78,162,716 | chr11:78162716:T:G | t | g | -1.0629 | 0.2612 | 4.71E-05 | ----+-++? | 14.872 | 7 | 0.03767 |
| Any CBVD | chr11 | 79,594,268 | chr11:79594268:A:T | a | t | -0.4643 | 0.1155 | 5.80E-05 | ----+---- | 14.987 | 8 | 0.05941 |
| Any CBVD | chr11 | 80,739,563 | chr11:80739563:C:T | t | c | -0.4079 | 0.1029 | 7.34E-05 | ---+-++-- | 23.232 | 8 | 0.003079 |
| Any CBVD | chr11 | 80,759,434 | chr11:80759434:A:G | a | g | -0.9143 | 0.2169 | 2.50E-05 | -----+--- | 28.21 | 8 | 0.000436 |
| Any CBVD | chr11 | 80,761,051 | chr11:80761051:A:T | a | t | -0.9211 | 0.2153 | 1.89E-05 | -----+--- | 27.726 | 8 | 0.000529 |
| Any CBVD | chr11 | 80,766,905 | chr11:80766905:A:C | a | c | -0.9256 | 0.2155 | 1.75E-05 | -----+--- | 27.686 | 8 | 0.000538 |
| Any CBVD | chr11 | 80,767,708 | chr11:80767708:T:A | a | t | 0.9278 | 0.2156 | 1.69E-05 | +++++-+++ | 27.839 | 8 | 0.000506 |
| Any CBVD | chr11 | 80,768,883 | chr11:80768883:T:C | t | c | -0.93 | 0.2176 | 1.92E-05 | -----+--- | 26.449 | 8 | 0.00088 |
| Any CBVD | chr11 | 80,776,141 | chr11:80776141:T:C | t | c | -0.934 | 0.2182 | 1.86E-05 | -----+--- | 27.521 | 8 | 0.000574 |
| Any CBVD | chr11 | 80,789,164 | chr11:80789164:C:T | t | c | 0.9568 | 0.2367 | 5.28E-05 | +++++-+++ | 31.554 | 8 | 0.000112 |
| Any CBVD | chr11 | 80,789,712 | chr11:80789712:A:G | a | g | -0.924 | 0.2263 | 4.44E-05 | -----+--- | 31.915 | 8 | 9.65E-05 |
| Any CBVD | chr11 | 80,822,241 | chr11:80822241:G:A | a | g | 0.9333 | 0.237 | 8.21E-05 | +++++-+++ | 29.708 | 8 | 0.000238 |
| Any CBVD | chr11 | 101,855,396 | chr11:101855396:G:T | t | g | 0.6884 | 0.1669 | 3.70E-05 | ++++-+-++ | 6.456 | 8 | 0.5963 |
| Any CBVD | chr11 | 118,721,056 | chr11:118721056:G:A | a | g | -0.2586 | 0.0645 | 6.12E-05 | --------- | 10.452 | 8 | 0.2347 |
| Any CBVD | chr11 | 118,752,276 | chr11:118752276:T:C | t | c | 0.2588 | 0.0651 | 7.09E-05 | +++++++++ | 6.735 | 8 | 0.5655 |
| Any CBVD | chr11 | 119,779,194 | chr11:119779194:C:T | t | c | 0.4449 | 0.1118 | 6.93E-05 | +++-+++++ | 4.109 | 8 | 0.8471 |
| Any CBVD | chr11 | 126,057,635 | chr11:126057635:C:T | t | c | 2.6967 | 0.5995 | 6.84E-06 | ??++???-? | 7.967 | 2 | 0.01862 |
| Any CBVD | chr12 | 4,452,709 | chr12:4452709:C:A | a | c | 0.2772 | 0.0693 | 6.39E-05 | +++++++++ | 2.137 | 8 | 0.9765 |
| Any CBVD | chr12 | 8,246,061 | chr12:8246061:G:A | a | g | 0.8374 | 0.209 | 6.16E-05 | +++++-++? | 4.314 | 7 | 0.743 |
| Any CBVD | chr12 | 8,402,029 | chr12:8402029:GT:G | g | gt | 0.8067 | 0.188 | 1.78E-05 | ++++++--+ | 14.755 | 8 | 0.06408 |
| Any CBVD | chr12 | 19,976,590 | chr12:19976590:A:G | a | g | -1.8141 | 0.3759 | 1.40E-06 | ??--?--+- | 21.977 | 5 | 0.000529 |
| Any CBVD | chr12 | 23,716,730 | chr12:23716730:C:T | t | c | 2.0967 | 0.4416 | 2.05E-06 | +???++??? | 0.758 | 2 | 0.6845 |
| Any CBVD | chr12 | 23,731,636 | chr12:23731636:G:T | t | g | 2.1242 | 0.4643 | 4.76E-06 | +???++??? | 0.101 | 2 | 0.9509 |
| Any CBVD | chr12 | 23,750,976 | chr12:23750976:T:C | t | c | -2.1413 | 0.4655 | 4.22E-06 | -???--??? | 0.102 | 2 | 0.9501 |
| Any CBVD | chr12 | 24,043,425 | chr12:24043425:T:C | t | c | -0.4915 | 0.1241 | 7.47E-05 | -+-++---- | 13.818 | 8 | 0.08663 |
| Any CBVD | chr12 | 30,849,767 | chr12:30849767:G:A | a | g | -0.2512 | 0.0588 | 1.90E-05 | ---++-+-- | 9.561 | 8 | 0.2972 |
| Any CBVD | chr12 | 30,849,851 | chr12:30849851:G:A | a | g | -0.2535 | 0.0587 | 1.60E-05 | ---++-+-- | 8.55 | 8 | 0.3816 |
| Any CBVD | chr12 | 30,851,152 | chr12:30851152:G:A | a | g | -0.2414 | 0.0573 | 2.48E-05 | ----+-+-- | 7.612 | 8 | 0.4723 |
| Any CBVD | chr12 | 30,852,306 | chr12:30852306:T:C | t | c | 0.2398 | 0.0572 | 2.80E-05 | ++++-+-++ | 7.616 | 8 | 0.4718 |
| Any CBVD | chr12 | 30,855,451 | chr12:30855451:A:G | a | g | 0.2305 | 0.0572 | 5.49E-05 | ++++-+-++ | 8.469 | 8 | 0.389 |
| Any CBVD | chr12 | 30,856,623 | chr12:30856623:C:T | t | c | -0.2415 | 0.0585 | 3.62E-05 | ---++-+-- | 9.71 | 8 | 0.286 |
| Any CBVD | chr12 | 30,856,713 | chr12:30856713:T:C | t | c | 0.2332 | 0.0572 | 4.59E-05 | ++++-+-++ | 8.568 | 8 | 0.38 |
| Any CBVD | chr12 | 30,857,911 | chr12:30857911:C:T | t | c | -0.2378 | 0.0585 | 4.81E-05 | ---++-+-- | 9.586 | 8 | 0.2953 |
| Any CBVD | chr12 | 30,857,972 | chr12:30857972:C:T | t | c | -0.2414 | 0.0586 | 3.74E-05 | ---++-+-- | 9.671 | 8 | 0.2889 |
| Any CBVD | chr12 | 51,558,016 | chr12:51558016:C:T | t | c | 1.1554 | 0.2763 | 2.90E-05 | +-++??+-+ | 6.493 | 6 | 0.3703 |
| Any CBVD | chr12 | 51,943,308 | chr12:51943308:T:G | t | g | -1.4071 | 0.3555 | 7.57E-05 | ?-------? | 2.206 | 6 | 0.8998 |
| Any CBVD | chr12 | 58,243,903 | chr12:58243903:C:T | t | c | 0.5578 | 0.129 | 1.53E-05 | +++++++++ | 6.389 | 8 | 0.6038 |
| Any CBVD | chr12 | 58,300,065 | chr12:58300065:T:A | a | t | 0.4699 | 0.1201 | 9.16E-05 | +++++++++ | 6.506 | 8 | 0.5908 |
| Any CBVD | chr12 | 58,317,232 | chr12:58317232:A:G | a | g | -0.504 | 0.12 | 2.65E-05 | --------- | 6.094 | 8 | 0.6367 |
| Any CBVD | chr12 | 64,990,782 | chr12:64990782:G:A | a | g | 0.7215 | 0.183 | 8.08E-05 | +++--++-+ | 12.533 | 8 | 0.129 |
| Any CBVD | chr12 | 70,695,073 | chr12:70695073:A:C | a | c | -1.3776 | 0.3506 | 8.53E-05 | -+?????-- | 5.926 | 3 | 0.1153 |
| Any CBVD | chr12 | 75,705,761 | chr12:75705761:T:C | t | c | 0.346 | 0.0877 | 7.96E-05 | +-+++++-+ | 18.225 | 8 | 0.0196 |

| | | | | | | | | | | | | |
|---|---|---|---|---|---|---|---|---|---|---|---|---|
| Any CBVD | chr12 | 75,993,005 | chr12:75993005:T:C | t | c | -0.9396 | 0.2336 | 5.77E-05 | -------+- | 6.051 | 8 | 0.6415 |
| Any CBVD | chr12 | 75,996,040 | chr12:75996040:C:T | t | c | 0.9794 | 0.2388 | 4.12E-05 | +++++++-+ | 5.659 | 8 | 0.6854 |
| Any CBVD | chr12 | 76,000,602 | chr12:76000602:GT:G | g | gt | 1.0045 | 0.2402 | 2.90E-05 | +++++++-+ | 5.817 | 8 | 0.6677 |
| Any CBVD | chr12 | 80,348,365 | chr12:80348365:T:C | t | c | 0.3246 | 0.0784 | 3.47E-05 | ++++----+ | 19.253 | 8 | 0.01356 |
| Any CBVD | chr12 | 80,349,548 | chr12:80349548:G:A | a | g | -0.3234 | 0.0784 | 3.71E-05 | ----++++- | 19.354 | 8 | 0.01308 |
| Any CBVD | chr12 | 80,351,372 | chr12:80351372:G:T | t | g | -0.3217 | 0.0785 | 4.18E-05 | ----++++- | 18.642 | 8 | 0.0169 |
| Any CBVD | chr12 | 80,360,096 | chr12:80360096:G:T | t | g | -0.3118 | 0.0783 | 6.81E-05 | ----++++- | 15.945 | 8 | 0.04317 |
| Any CBVD | chr12 | 80,360,462 | chr12:80360462:C:T | t | c | -0.3092 | 0.0786 | 8.29E-05 | ----++++- | 18.407 | 8 | 0.01837 |
| Any CBVD | chr12 | 80,363,869 | chr12:80363869:G:A | a | g | -0.3095 | 0.0783 | 7.81E-05 | ----++++- | 18.277 | 8 | 0.01924 |
| Any CBVD | chr12 | 80,363,938 | chr12:80363938:A:T | a | t | 0.3089 | 0.0784 | 8.09E-05 | ++++----+ | 18.261 | 8 | 0.01936 |
| Any CBVD | chr12 | 80,364,923 | chr12:80364923:C:A | a | c | -0.3094 | 0.0784 | 7.92E-05 | ----++++- | 18.211 | 8 | 0.0197 |
| Any CBVD | chr12 | 80,367,650 | chr12:80367650:G:A | a | g | -0.3092 | 0.0781 | 7.57E-05 | ----++++- | 18.289 | 8 | 0.01916 |
| Any CBVD | chr12 | 80,368,466 | chr12:80368466:G:A | a | g | -0.3091 | 0.0784 | 8.04E-05 | ----++++- | 18.304 | 8 | 0.01906 |
| Any CBVD | chr12 | 80,368,974 | chr12:80368974:C:A | a | c | -0.305 | 0.0783 | 9.85E-05 | ----++++- | 18.12 | 8 | 0.02034 |
| Any CBVD | chr12 | 80,368,996 | chr12:80368996:A:C | a | c | 0.3099 | 0.0782 | 7.50E-05 | ++++----+ | 15.79 | 8 | 0.04548 |
| Any CBVD | chr12 | 80,369,544 | chr12:80369544:A:G | a | g | 0.3091 | 0.0784 | 8.02E-05 | ++++----+ | 18.261 | 8 | 0.01935 |
| Any CBVD | chr12 | 80,369,679 | chr12:80369679:C:T | t | c | -0.3111 | 0.0783 | 7.02E-05 | ----++++- | 15.848 | 8 | 0.04461 |
| Any CBVD | chr12 | 80,370,349 | chr12:80370349:A:C | a | c | 0.3086 | 0.0784 | 8.23E-05 | ++++----+ | 18.239 | 8 | 0.0195 |
| Any CBVD | chr12 | 80,370,424 | chr12:80370424:CTT:C | ctt | c | 0.3075 | 0.0784 | 8.77E-05 | ++++----+ | 18.187 | 8 | 0.01987 |
| Any CBVD | chr12 | 80,370,876 | chr12:80370876:G:A | a | g | -0.3088 | 0.0784 | 8.14E-05 | ----++++- | 18.228 | 8 | 0.01958 |
| Any CBVD | chr12 | 80,371,903 | chr12:80371903:TGAA:T | t | tgaa | -0.3104 | 0.0784 | 7.48E-05 | ----++++- | 18.135 | 8 | 0.02024 |
| Any CBVD | chr12 | 80,371,907 | chr12:80371907:G:A | a | g | -0.3086 | 0.0784 | 8.23E-05 | ----++++- | 18.217 | 8 | 0.01966 |
| Any CBVD | chr12 | 80,372,165 | chr12:80372165:G:A | a | g | -0.3087 | 0.0784 | 8.19E-05 | ----++++- | 18.214 | 8 | 0.01968 |
| Any CBVD | chr12 | 80,373,889 | chr12:80373889:G:T | t | g | -0.3084 | 0.0784 | 8.31E-05 | ----++++- | 18.206 | 8 | 0.01973 |
| Any CBVD | chr12 | 80,374,528 | chr12:80374528:G:A | a | g | -0.3089 | 0.0784 | 8.12E-05 | ----++++- | 18.353 | 8 | 0.01873 |
| Any CBVD | chr12 | 80,374,668 | chr12:80374668:C:G | c | g | 0.308 | 0.0784 | 8.49E-05 | ++++----+ | 18.304 | 8 | 0.01906 |
| Any CBVD | chr12 | 80,375,409 | chr12:80375409:G:A | a | g | -0.3077 | 0.0781 | 8.19E-05 | ----++++- | 18.276 | 8 | 0.01925 |
| Any CBVD | chr12 | 80,376,011 | chr12:80376011:A:G | a | g | 0.3079 | 0.0784 | 8.53E-05 | ++++----+ | 18.27 | 8 | 0.01929 |
| Any CBVD | chr12 | 80,376,690 | chr12:80376690:C:T | t | c | -0.3084 | 0.0784 | 8.34E-05 | ----++++- | 18.284 | 8 | 0.0192 |
| Any CBVD | chr12 | 80,376,984 | chr12:80376984:A:G | a | g | 0.3079 | 0.0784 | 8.59E-05 | ++++----+ | 18.267 | 8 | 0.01931 |
| Any CBVD | chr12 | 80,378,033 | chr12:80378033:G:A | a | g | -0.3082 | 0.0781 | 8.03E-05 | ----++++- | 18.256 | 8 | 0.01938 |
| Any CBVD | chr12 | 80,380,772 | chr12:80380772:C:T | t | c | -0.3094 | 0.0784 | 7.93E-05 | ----++++- | 18.289 | 8 | 0.01916 |
| Any CBVD | chr12 | 80,384,232 | chr12:80384232:T:C | t | c | 0.309 | 0.0784 | 8.14E-05 | ++++----+ | 18.333 | 8 | 0.01886 |
| Any CBVD | chr12 | 80,386,598 | chr12:80386598:A:G | a | g | 0.3092 | 0.0784 | 8.03E-05 | ++++----+ | 18.322 | 8 | 0.01894 |
| Any CBVD | chr12 | 80,386,988 | chr12:80386988:G:A | a | g | -0.3081 | 0.0784 | 8.50E-05 | ----++++- | 18.288 | 8 | 0.01917 |
| Any CBVD | chr12 | 80,387,850 | chr12:80387850:T:C | t | c | 0.3094 | 0.0784 | 7.93E-05 | ++++----+ | 18.282 | 8 | 0.01921 |
| Any CBVD | chr12 | 80,389,001 | chr12:80389001:T:G | t | g | 0.3125 | 0.0784 | 6.75E-05 | ++++----+ | 18.833 | 8 | 0.01578 |
| Any CBVD | chr12 | 80,390,074 | chr12:80390074:A:G | a | g | 0.3109 | 0.0783 | 7.13E-05 | ++++----+ | 15.853 | 8 | 0.04454 |
| Any CBVD | chr12 | 80,390,473 | chr12:80390473:C:T | t | c | -0.309 | 0.0784 | 8.10E-05 | ----++++- | 18.253 | 8 | 0.01941 |
| Any CBVD | chr12 | 80,390,483 | chr12:80390483:G:T | t | g | -0.309 | 0.0784 | 8.09E-05 | ----++++- | 18.252 | 8 | 0.01941 |
| Any CBVD | chr12 | 80,398,266 | chr12:80398266:G:A | a | g | -0.3078 | 0.0784 | 8.68E-05 | ----++++- | 18.224 | 8 | 0.01961 |
| Any CBVD | chr12 | 93,078,172 | chr12:93078172:C:G | c | g | 0.2239 | 0.0574 | 9.65E-05 | ++++--+++ | 10.327 | 8 | 0.2428 |
| Any CBVD | chr12 | 93,079,840 | chr12:93079840:G:A | a | g | -0.2249 | 0.057 | 8.05E-05 | ----++--- | 10.165 | 8 | 0.2536 |
| Any CBVD | chr12 | 93,082,967 | chr12:93082967:AT:A | a | at | -0.2252 | 0.0577 | 9.49E-05 | ----++--- | 10.549 | 8 | 0.2286 |
| Any CBVD | chr12 | 103,607,999 | chr12:103607999:A:C | a | c | -0.9065 | 0.2311 | 8.75E-05 | ----+--+- | 28.446 | 8 | 0.000396 |
| Any CBVD | chr12 | 103,610,759 | chr12:103610759:A:T | a | t | -0.9117 | 0.2317 | 8.31E-05 | ----+--+- | 28.41 | 8 | 0.000402 |

| | | | | | | | | | | | | |
|---|---|---|---|---|---|---|---|---|---|---|---|---|
| Any CBVD | chr12 | 103,783,306 | chr12:103783306:C:A | a | c | 0.3904 | 0.0846 | 3.97E-06 | +-+++++++ | 9.236 | 8 | 0.3228 |
| Any CBVD | chr12 | 116,396,322 | chr12:116396322:T:C | t | c | -0.4011 | 0.0994 | 5.44E-05 | -------+- | 6.716 | 8 | 0.5675 |
| Any CBVD | chr12 | 116,399,542 | chr12:116399542:C:A | a | c | 0.4446 | 0.1057 | 2.59E-05 | +++++++-+ | 5.835 | 8 | 0.6657 |
| Any CBVD | chr12 | 116,401,216 | chr12:116401216:C:T | t | c | 0.4498 | 0.1056 | 2.06E-05 | +++++++-+ | 5.766 | 8 | 0.6735 |
| Any CBVD | chr12 | 116,415,428 | chr12:116415428:G:A | a | g | 0.4483 | 0.1147 | 9.33E-05 | +++++++++ | 5.504 | 8 | 0.7026 |
| Any CBVD | chr12 | 116,415,594 | chr12:116415594:TTG:T | t | ttg | 0.4513 | 0.1148 | 8.51E-05 | +++++++++ | 5.355 | 8 | 0.719 |
| Any CBVD | chr12 | 116,416,695 | chr12:116416695:G:A | a | g | 0.4398 | 0.1118 | 8.39E-05 | +++++++-+ | 6.442 | 8 | 0.5978 |
| Any CBVD | chr12 | 117,137,625 | chr12:117137625:C:A | a | c | 2.2122 | 0.5558 | 6.88E-05 | +???++??? | 4.258 | 2 | 0.1189 |
| Any CBVD | chr12 | 120,602,486 | chr12:120602486:G:A | a | g | 1.075 | 0.2758 | 9.71E-05 | +-++-++-+ | 8.58 | 8 | 0.379 |
| Any CBVD | chr13 | 30,533,681 | chr13:30533681:T:C | t | c | 0.2938 | 0.0718 | 4.23E-05 | +-+++++-+ | 13.262 | 8 | 0.1031 |
| Any CBVD | chr13 | 37,716,139 | chr13:37716139:A:G | a | g | -0.2393 | 0.0577 | 3.41E-05 | ----+---- | 3.81 | 8 | 0.8739 |
| Any CBVD | chr13 | 44,782,601 | chr13:44782601:C:G | c | g | -0.6776 | 0.1577 | 1.73E-05 | --------- | 8.136 | 8 | 0.4203 |
| Any CBVD | chr13 | 44,783,520 | chr13:44783520:A:G | a | g | -0.6798 | 0.1576 | 1.61E-05 | --------- | 8.339 | 8 | 0.4011 |
| Any CBVD | chr13 | 49,017,798 | chr13:49017798:T:G | t | g | -0.3519 | 0.0892 | 8.02E-05 | --------- | 7.713 | 8 | 0.4619 |
| Any CBVD | chr13 | 49,175,988 | chr13:49175988:A:G | a | g | -0.3779 | 0.0941 | 5.92E-05 | -+------- | 5.757 | 8 | 0.6744 |
| Any CBVD | chr13 | 49,179,862 | chr13:49179862:C:G | c | g | -0.5543 | 0.1397 | 7.24E-05 | -+------- | 9.504 | 8 | 0.3016 |
| Any CBVD | chr13 | 49,265,939 | chr13:49265939:A:G | a | g | -0.7963 | 0.1914 | 3.18E-05 | -+--+---- | 8.554 | 8 | 0.3813 |
| Any CBVD | chr13 | 49,323,544 | chr13:49323544:C:G | c | g | -0.768 | 0.1968 | 9.54E-05 | -+--+---- | 8.24 | 8 | 0.4104 |
| Any CBVD | chr13 | 69,018,277 | chr13:69018277:G:A | a | g | 2.3997 | 0.4875 | 8.54E-07 | ??++??-?? | 10.618 | 2 | 0.004947 |
| Any CBVD | chr13 | 85,700,706 | chr13:85700706:T:C | t | c | -0.2336 | 0.0595 | 8.75E-05 | --------- | 2.981 | 8 | 0.9356 |
| Any CBVD | chr13 | 85,700,796 | chr13:85700796:G:A | a | g | 0.2333 | 0.0595 | 8.78E-05 | +++++++++ | 2.986 | 8 | 0.9352 |
| Any CBVD | chr13 | 102,901,050 | chr13:102901050:A:G | a | g | -1.0079 | 0.2521 | 6.38E-05 | ---+----? | 4.557 | 7 | 0.7138 |
| Any CBVD | chr13 | 111,347,883 | chr13:111347883:A:G | a | g | -1.2169 | 0.2912 | 2.93E-05 | ----?-?-? | 8.025 | 5 | 0.1549 |
| Any CBVD | chr14 | 29,993,846 | chr14:29993846:T:C | t | c | 0.313 | 0.0783 | 6.34E-05 | +++++++-+ | 7.524 | 8 | 0.4813 |
| Any CBVD | chr14 | 29,994,527 | chr14:29994527:G:T | t | g | -0.3191 | 0.0785 | 4.83E-05 | -------+- | 7.787 | 8 | 0.4546 |
| Any CBVD | chr14 | 29,998,298 | chr14:29998298:C:T | t | c | -0.3135 | 0.0782 | 6.08E-05 | -------+- | 7.368 | 8 | 0.4975 |
| Any CBVD | chr14 | 29,998,787 | chr14:29998787:A:T | a | t | 0.3161 | 0.0782 | 5.27E-05 | +++++++-+ | 7.668 | 8 | 0.4665 |
| Any CBVD | chr14 | 30,001,505 | chr14:30001505:G:T | t | g | -0.3088 | 0.0778 | 7.26E-05 | -------+- | 7.883 | 8 | 0.445 |
| Any CBVD | chr14 | 30,012,598 | chr14:30012598:T:C | t | c | 0.3184 | 0.0729 | 1.26E-05 | +++++++++ | 4.549 | 8 | 0.8045 |
| Any CBVD | chr14 | 30,019,493 | chr14:30019493:C:T | t | c | -0.3227 | 0.0727 | 9.11E-06 | --------- | 4.474 | 8 | 0.8121 |
| Any CBVD | chr14 | 30,031,147 | chr14:30031147:C:T | t | c | -0.3195 | 0.0726 | 1.09E-05 | --------- | 4.891 | 8 | 0.7691 |
| Any CBVD | chr14 | 30,037,683 | chr14:30037683:C:T | t | c | -0.3192 | 0.0729 | 1.18E-05 | --------- | 4.946 | 8 | 0.7633 |
| Any CBVD | chr14 | 30,042,868 | chr14:30042868:A:G | a | g | 0.3247 | 0.0729 | 8.51E-06 | +++++++++ | 4.696 | 8 | 0.7895 |
| Any CBVD | chr14 | 30,045,623 | chr14:30045623:A:G | a | g | 0.322 | 0.073 | 1.02E-05 | +++++++++ | 4.628 | 8 | 0.7965 |
| Any CBVD | chr14 | 30,046,165 | chr14:30046165:A:G | a | g | 0.3186 | 0.073 | 1.27E-05 | +++++++++ | 4.902 | 8 | 0.768 |
| Any CBVD | chr14 | 30,068,022 | chr14:30068022:CA:C | ca | c | -0.3169 | 0.081 | 9.08E-05 | -+-----+- | 13.532 | 8 | 0.09481 |
| Any CBVD | chr14 | 34,122,665 | chr14:34122665:G:A | a | g | -0.2283 | 0.0574 | 7.01E-05 | ---++--+- | 7.495 | 8 | 0.4843 |
| Any CBVD | chr14 | 34,686,678 | chr14:34686678:C:T | t | c | 1.4866 | 0.378 | 8.39E-05 | +???-??-+ | 8.166 | 3 | 0.0427 |
| Any CBVD | chr14 | 35,094,657 | chr14:35094657:TAC:T | t | tac | -0.2209 | 0.0563 | 8.73E-05 | ------+-- | 2.263 | 8 | 0.9718 |
| Any CBVD | chr14 | 35,107,252 | chr14:35107252:A:C | a | c | 0.221 | 0.0563 | 8.69E-05 | ++++++-++ | 2.333 | 8 | 0.9691 |
| Any CBVD | chr14 | 35,111,070 | chr14:35111070:A:G | a | g | 0.2194 | 0.0563 | 9.67E-05 | ++++++-++ | 2.237 | 8 | 0.9729 |
| Any CBVD | chr14 | 39,224,364 | chr14:39224364:G:A | a | g | 1.2356 | 0.2908 | 2.15E-05 | +?+?+??++ | 2.734 | 4 | 0.6033 |
| Any CBVD | chr14 | 50,749,097 | chr14:50749097:T:C | t | c | 0.2234 | 0.0572 | 9.41E-05 | ++++-+-++ | 11.139 | 8 | 0.1939 |
| Any CBVD | chr14 | 82,059,299 | chr14:82059299:A:G | a | g | 0.2261 | 0.058 | 9.72E-05 | +++++++++ | 3.263 | 8 | 0.9168 |
| Any CBVD | chr14 | 92,592,925 | chr14:92592925:C:T | t | c | 1.2565 | 0.3079 | 4.48E-05 | ++?+??+++ | 11.258 | 5 | 0.0465 |
| Any CBVD | chr14 | 94,577,881 | chr14:94577881:T:C | t | c | -0.24 | 0.0597 | 5.78E-05 | --------- | 3.651 | 8 | 0.8872 |

| | | | | | | | | | | | | |
|---|---|---|---|---|---|---|---|---|---|---|---|---|
| Any CBVD | chr14 | 94,577,934 | chr14:94577934:T:C | t | c | -0.2401 | 0.0597 | 5.76E-05 | --------- | 3.648 | 8 | 0.8874 |
| Any CBVD | chr14 | 94,577,985 | chr14:94577985:A:G | a | g | -0.2407 | 0.0597 | 5.49E-05 | --------- | 3.75 | 8 | 0.879 |
| Any CBVD | chr14 | 94,579,019 | chr14:94579019:A:G | a | g | -0.2374 | 0.0597 | 6.97E-05 | --------- | 4.021 | 8 | 0.8552 |
| Any CBVD | chr14 | 94,579,567 | chr14:94579567:T:C | t | c | -0.2401 | 0.0597 | 5.73E-05 | --------- | 3.802 | 8 | 0.8746 |
| Any CBVD | chr14 | 94,580,349 | chr14:94580349:C:T | t | c | 0.2401 | 0.0597 | 5.76E-05 | +++++++++ | 3.82 | 8 | 0.873 |
| Any CBVD | chr14 | 94,580,679 | chr14:94580679:G:A | a | g | 0.2399 | 0.0595 | 5.48E-05 | +++++++++ | 3.753 | 8 | 0.8787 |
| Any CBVD | chr14 | 94,581,060 | chr14:94581060:C:T | t | c | 0.2399 | 0.0597 | 5.83E-05 | +++++++++ | 3.804 | 8 | 0.8743 |
| Any CBVD | chr14 | 94,581,235 | chr14:94581235:T:C | t | c | -0.2401 | 0.0597 | 5.76E-05 | --------- | 3.806 | 8 | 0.8742 |
| Any CBVD | chr14 | 94,581,271 | chr14:94581271:G:C | c | g | 0.2402 | 0.0597 | 5.71E-05 | +++++++++ | 3.815 | 8 | 0.8734 |
| Any CBVD | chr14 | 94,581,473 | chr14:94581473:A:G | a | g | -0.2398 | 0.0596 | 5.83E-05 | --------- | 3.78 | 8 | 0.8764 |
| Any CBVD | chr14 | 94,581,500 | chr14:94581500:G:A | a | g | 0.2396 | 0.0597 | 5.93E-05 | +++++++++ | 3.78 | 8 | 0.8764 |
| Any CBVD | chr14 | 94,581,510 | chr14:94581510:A:C | a | c | -0.2401 | 0.0597 | 5.76E-05 | --------- | 3.808 | 8 | 0.874 |
| Any CBVD | chr14 | 94,582,125 | chr14:94582125:G:A | a | g | 0.2396 | 0.0597 | 5.98E-05 | +++++++++ | 3.836 | 8 | 0.8717 |
| Any CBVD | chr14 | 94,582,146 | chr14:94582146:A:G | a | g | -0.2394 | 0.0597 | 6.07E-05 | --------- | 3.845 | 8 | 0.8709 |
| Any CBVD | chr14 | 94,582,147 | chr14:94582147:G:A | a | g | 0.2394 | 0.0597 | 6.07E-05 | +++++++++ | 3.845 | 8 | 0.8709 |
| Any CBVD | chr14 | 94,582,202 | chr14:94582202:C:A | a | c | 0.2394 | 0.0597 | 6.07E-05 | +++++++++ | 3.849 | 8 | 0.8705 |
| Any CBVD | chr14 | 94,582,313 | chr14:94582313:A:T | a | t | -0.2392 | 0.0597 | 6.16E-05 | --------- | 3.853 | 8 | 0.8701 |
| Any CBVD | chr14 | 94,583,172 | chr14:94583172:G:T | t | g | 0.2398 | 0.0597 | 5.88E-05 | +++++++++ | 3.76 | 8 | 0.8781 |
| Any CBVD | chr14 | 94,583,330 | chr14:94583330:A:G | a | g | -0.2402 | 0.0597 | 5.72E-05 | --------- | 3.791 | 8 | 0.8755 |
| Any CBVD | chr14 | 94,584,159 | chr14:94584159:C:G | c | g | 0.2333 | 0.0564 | 3.49E-05 | +++++++++ | 1.88 | 8 | 0.9845 |
| Any CBVD | chr14 | 101,069,690 | chr14:101069690:C:A | a | c | 3.9928 | 0.9186 | 1.38E-05 | ?+??-+??? | 1.917 | 2 | 0.3835 |
| Any CBVD | chr15 | 22,870,354 | chr15:22870354:A:T | a | t | -1.4708 | 0.3487 | 2.46E-05 | ????--??- | 3.863 | 2 | 0.1449 |
| Any CBVD | chr15 | 27,369,773 | chr15:27369773:C:T | t | c | 0.9621 | 0.2021 | 1.93E-06 | ++++-++++ | 9.031 | 8 | 0.3397 |
| Any CBVD | chr15 | 35,846,141 | chr15:35846141:T:C | t | c | -0.2259 | 0.0565 | 6.40E-05 | ---+-+--- | 11.098 | 8 | 0.1962 |
| Any CBVD | chr15 | 35,846,823 | chr15:35846823:A:G | a | g | -0.2237 | 0.0561 | 6.61E-05 | ---+-+--- | 11.119 | 8 | 0.195 |
| Any CBVD | chr15 | 35,848,452 | chr15:35848452:G:A | a | g | 0.2275 | 0.0565 | 5.69E-05 | +++-+-+++ | 11.115 | 8 | 0.1953 |
| Any CBVD | chr15 | 35,857,196 | chr15:35857196:C:T | t | c | 0.2416 | 0.0572 | 2.40E-05 | ++++---++ | 11.657 | 8 | 0.1672 |
| Any CBVD | chr15 | 50,403,585 | chr15:50403585:A:G | a | g | -1.0948 | 0.2442 | 7.36E-06 | --++--+-- | 12.996 | 8 | 0.112 |
| Any CBVD | chr15 | 55,003,048 | chr15:55003048:T:C | t | c | -0.2246 | 0.0568 | 7.79E-05 | -------+- | 2.17 | 8 | 0.9753 |
| Any CBVD | chr15 | 55,005,520 | chr15:55005520:A:G | a | g | -0.2232 | 0.0568 | 8.52E-05 | -------+- | 2.029 | 8 | 0.9801 |
| Any CBVD | chr15 | 55,023,942 | chr15:55023942:T:C | t | c | 0.2262 | 0.0566 | 6.34E-05 | +++++++++ | 6.76 | 8 | 0.5627 |
| Any CBVD | chr15 | 55,024,897 | chr15:55024897:C:A | a | c | -0.2267 | 0.0567 | 6.41E-05 | --------- | 5.029 | 8 | 0.7545 |
| Any CBVD | chr15 | 55,025,321 | chr15:55025321:G:T | t | g | -0.2314 | 0.0566 | 4.37E-05 | --------- | 4.796 | 8 | 0.7792 |
| Any CBVD | chr15 | 58,671,486 | chr15:58671486:G:T | t | g | 1.2826 | 0.3054 | 2.67E-05 | +?+-+?++? | 8.187 | 5 | 0.1462 |
| Any CBVD | chr15 | 58,715,097 | chr15:58715097:T:C | t | c | -1.28 | 0.3053 | 2.75E-05 | -?-+-?--? | 7.708 | 5 | 0.1731 |
| Any CBVD | chr15 | 58,759,514 | chr15:58759514:C:T | t | c | 1.287 | 0.3058 | 2.57E-05 | +?+-+?++? | 7.852 | 5 | 0.1646 |
| Any CBVD | chr15 | 70,387,012 | chr15:70387012:G:A | a | g | 0.4349 | 0.1105 | 8.30E-05 | +++++++-+ | 11.189 | 8 | 0.1912 |
| Any CBVD | chr15 | 87,087,311 | chr15:87087311:GC:G | g | gc | 0.775 | 0.1952 | 7.17E-05 | ++++++-++ | 13.159 | 8 | 0.1065 |
| Any CBVD | chr16 | 286,888 | chr16:286888:C:CA | ca | c | 1.3408 | 0.3352 | 6.35E-05 | +-?++-??+ | 13.773 | 5 | 0.01712 |
| Any CBVD | chr16 | 6,400,239 | chr16:6400239:A:AT | a | at | -1.5665 | 0.3381 | 3.59E-06 | -?-+?-??? | 7.013 | 3 | 0.07148 |
| Any CBVD | chr16 | 6,410,626 | chr16:6410626:G:T | t | g | 1.0854 | 0.2717 | 6.49E-05 | +?+-?+??+ | 5.988 | 4 | 0.2001 |
| Any CBVD | chr16 | 6,412,500 | chr16:6412500:C:T | t | c | 1.2011 | 0.3054 | 8.41E-05 | +?+-?+??+ | 6.381 | 4 | 0.1724 |
| Any CBVD | chr16 | 22,712,625 | chr16:22712625:G:C | c | g | 0.2297 | 0.0587 | 9.10E-05 | ++++-++++ | 3.901 | 8 | 0.8659 |
| Any CBVD | chr16 | 24,626,795 | chr16:24626795:A:G | a | g | -0.3998 | 0.0911 | 1.14E-05 | ------+-- | 6.087 | 8 | 0.6375 |
| Any CBVD | chr16 | 50,727,755 | chr16:50727755:C:T | t | c | 0.7411 | 0.1893 | 9.00E-05 | +++--+-++ | 5.568 | 8 | 0.6955 |
| Any CBVD | chr16 | 51,391,686 | chr16:51391686:C:T | t | c | 0.4884 | 0.1167 | 2.85E-05 | ++++++-++ | 10.652 | 8 | 0.2222 |

| | | | | | | | | | | | | |
|---|---|---|---|---|---|---|---|---|---|---|---|---|
| Any CBVD | chr16 | 52,848,127 | chr16:52848127:C:T | t | c | 0.9221 | 0.219 | 2.54E-05 | ++++++-++ | 4.166 | 8 | 0.8418 |
| Any CBVD | chr16 | 57,329,084 | chr16:57329084:T:C | t | c | -1.2124 | 0.273 | 8.94E-06 | --+---+-- | 10.412 | 8 | 0.2373 |
| Any CBVD | chr16 | 73,404,471 | chr16:73404471:T:C | t | c | 0.4606 | 0.1151 | 6.30E-05 | ++++-++-+ | 6.513 | 8 | 0.59 |
| Any CBVD | chr16 | 78,747,054 | chr16:78747054:A:G | a | g | -1.47 | 0.3735 | 8.29E-05 | ---???--? | 1.593 | 4 | 0.81 |
| Any CBVD | chr16 | 83,760,390 | chr16:83760390:G:A | a | g | 0.2651 | 0.0627 | 2.35E-05 | +++++--++ | 14.213 | 8 | 0.07639 |
| Any CBVD | chr16 | 83,761,381 | chr16:83761381:G:A | a | g | 0.2407 | 0.0616 | 9.25E-05 | +++-+-+++ | 12.353 | 8 | 0.1361 |
| Any CBVD | chr16 | 83,761,386 | chr16:83761386:C:G | c | g | -0.2407 | 0.0616 | 9.25E-05 | ---+-+--- | 12.353 | 8 | 0.1361 |
| Any CBVD | chr16 | 83,761,454 | chr16:83761454:T:A | a | t | 0.2408 | 0.0616 | 9.22E-05 | +++-+--++ | 11.935 | 8 | 0.1541 |
| Any CBVD | chr16 | 83,761,878 | chr16:83761878:C:T | t | c | 0.2463 | 0.0617 | 6.62E-05 | +++++-+++ | 11.093 | 8 | 0.1965 |
| Any CBVD | chr16 | 83,763,969 | chr16:83763969:C:T | t | c | 0.2477 | 0.0603 | 3.96E-05 | +++++--++ | 11.606 | 8 | 0.1697 |
| Any CBVD | chr16 | 83,766,115 | chr16:83766115:C:T | t | c | 0.2461 | 0.0602 | 4.36E-05 | +++++-+++ | 8.047 | 8 | 0.4289 |
| Any CBVD | chr16 | 83,766,217 | chr16:83766217:T:A | a | t | 0.2408 | 0.0597 | 5.43E-05 | +++++-+++ | 6.64 | 8 | 0.576 |
| Any CBVD | chr16 | 83,768,112 | chr16:83768112:G:A | a | g | 0.2377 | 0.0597 | 6.89E-05 | +++++-+-+ | 6.277 | 8 | 0.6163 |
| Any CBVD | chr16 | 83,770,329 | chr16:83770329:A:C | a | c | -0.2791 | 0.0598 | 3.01E-06 | -----++-- | 15.51 | 8 | 0.04996 |
| Any CBVD | chr16 | 83,770,784 | chr16:83770784:T:A | a | t | 0.2578 | 0.0609 | 2.32E-05 | +++++--++ | 13.053 | 8 | 0.1101 |
| Any CBVD | chr16 | 83,771,536 | chr16:83771536:A:G | a | g | -0.2606 | 0.0609 | 1.87E-05 | -----++-- | 13.079 | 8 | 0.1092 |
| Any CBVD | chr16 | 83,772,244 | chr16:83772244:CT:C | ct | c | -0.2346 | 0.0586 | 6.31E-05 | -----+-+- | 4.639 | 8 | 0.7953 |
| Any CBVD | chr16 | 83,772,285 | chr16:83772285:G:A | a | g | 0.2738 | 0.0622 | 1.07E-05 | +++++--++ | 10.734 | 8 | 0.2172 |
| Any CBVD | chr16 | 83,772,348 | chr16:83772348:A:G | a | g | -0.273 | 0.0621 | 1.09E-05 | ---+-++-- | 11.127 | 8 | 0.1946 |
| Any CBVD | chr16 | 84,471,125 | chr16:84471125:G:C | c | g | 0.5449 | 0.1349 | 5.36E-05 | ++++-++++ | 13.667 | 8 | 0.09087 |
| Any CBVD | chr16 | 84,471,187 | chr16:84471187:A:G | a | g | -0.5634 | 0.1381 | 4.49E-05 | ----+---- | 13.231 | 8 | 0.1041 |
| Any CBVD | chr16 | 84,632,695 | chr16:84632695:C:G | c | g | -0.5814 | 0.146 | 6.84E-05 | -+--+---- | 9.144 | 8 | 0.3303 |
| Any CBVD | chr16 | 85,884,514 | chr16:85884514:G:T | t | g | 0.6613 | 0.1518 | 1.33E-05 | +-++-++-+ | 12.762 | 8 | 0.1203 |
| Any CBVD | chr16 | 85,884,798 | chr16:85884798:C:A | a | c | 0.6577 | 0.1517 | 1.46E-05 | +-++-++-+ | 12.684 | 8 | 0.1232 |
| Any CBVD | chr16 | 85,886,006 | chr16:85886006:G:A | a | g | 0.6575 | 0.1516 | 1.44E-05 | +-++-++-+ | 12.664 | 8 | 0.1239 |
| Any CBVD | chr16 | 85,887,073 | chr16:85887073:G:C | c | g | 0.6392 | 0.1538 | 3.24E-05 | +-++-++-+ | 12.024 | 8 | 0.1501 |
| Any CBVD | chr16 | 85,887,494 | chr16:85887494:AG:A | a | ag | 0.6747 | 0.1551 | 1.36E-05 | +-++-++-+ | 12.329 | 8 | 0.1371 |
| Any CBVD | chr16 | 85,888,421 | chr16:85888421:G:T | t | g | 0.684 | 0.1554 | 1.08E-05 | +-++-++-+ | 12.211 | 8 | 0.142 |
| Any CBVD | chr16 | 85,889,253 | chr16:85889253:A:G | a | g | -0.6578 | 0.1516 | 1.42E-05 | -+--+--+- | 12.654 | 8 | 0.1243 |
| Any CBVD | chr16 | 85,891,765 | chr16:85891765:C:G | c | g | -0.6747 | 0.1467 | 4.26E-06 | -+--+-++- | 13.079 | 8 | 0.1092 |
| Any CBVD | chr16 | 85,892,430 | chr16:85892430:T:C | t | c | -0.6368 | 0.1507 | 2.37E-05 | -+--+--+- | 12.506 | 8 | 0.13 |
| Any CBVD | chr16 | 85,892,795 | chr16:85892795:T:C | t | c | -0.6376 | 0.1506 | 2.31E-05 | -+--+--+- | 12.546 | 8 | 0.1285 |
| Any CBVD | chr16 | 85,893,113 | chr16:85893113:G:A | a | g | 0.636 | 0.1508 | 2.47E-05 | +-++-++-+ | 12.605 | 8 | 0.1262 |
| Any CBVD | chr16 | 85,893,138 | chr16:85893138:T:G | t | g | -0.636 | 0.1508 | 2.47E-05 | -+--+--+- | 12.601 | 8 | 0.1263 |
| Any CBVD | chr16 | 85,893,160 | chr16:85893160:T:G | t | g | -0.636 | 0.1508 | 2.46E-05 | -+--+--+- | 12.599 | 8 | 0.1264 |
| Any CBVD | chr16 | 85,899,975 | chr16:85899975:C:A | a | c | 0.684 | 0.1555 | 1.09E-05 | +-++-++-+ | 9.046 | 8 | 0.3384 |
| Any CBVD | chr16 | 85,903,756 | chr16:85903756:G:C | c | g | 0.6558 | 0.1517 | 1.55E-05 | +-++-++-+ | 10.52 | 8 | 0.2304 |
| Any CBVD | chr16 | 85,906,044 | chr16:85906044:T:A | a | t | 0.6546 | 0.1532 | 1.93E-05 | +-++-++-+ | 11.271 | 8 | 0.1868 |
| Any CBVD | chr16 | 85,906,060 | chr16:85906060:G:A | a | g | 0.6372 | 0.1533 | 3.24E-05 | +-++-++-+ | 10.968 | 8 | 0.2035 |
| Any CBVD | chr16 | 85,908,569 | chr16:85908569:G:C | c | g | 0.6304 | 0.15 | 2.63E-05 | +-++-++-+ | 14.287 | 8 | 0.07458 |
| Any CBVD | chr16 | 85,908,814 | chr16:85908814:T:A | a | t | 0.6636 | 0.1465 | 5.89E-06 | ++++-++-+ | 14.211 | 8 | 0.07643 |
| Any CBVD | chr16 | 85,909,562 | chr16:85909562:G:C | c | g | 0.5958 | 0.1522 | 9.01E-05 | ++++-++-+ | 9.132 | 8 | 0.3313 |
| Any CBVD | chr16 | 86,228,563 | chr16:86228563:T:C | t | c | -2.3701 | 0.588 | 5.57E-05 | ?---????? | 3.15 | 2 | 0.207 |
| Any CBVD | chr16 | 86,229,167 | chr16:86229167:T:C | t | c | -2.371 | 0.588 | 5.52E-05 | ?---????? | 3.16 | 2 | 0.206 |
| Any CBVD | chr16 | 86,230,483 | chr16:86230483:G:A | a | g | 2.2697 | 0.5796 | 9.01E-05 | ?+++????? | 3.409 | 2 | 0.1818 |
| Any CBVD | chr16 | 86,235,926 | chr16:86235926:A:C | a | c | -2.3615 | 0.5876 | 5.84E-05 | ?---????? | 3.228 | 2 | 0.1991 |

| | | | | | | | | | | | | |
|---|---|---|---|---|---|---|---|---|---|---|---|---|
| Any CBVD | chr16 | 86,238,132 | chr16:86238132:A:T | a | t | -2.3653 | 0.588 | 5.75E-05 | ?---????? | 3.224 | 2 | 0.1995 |
| Any CBVD | chr17 | 2,854,234 | chr17:2854234:T:C | t | c | -0.2364 | 0.0592 | 6.48E-05 | --------- | 0.913 | 8 | 0.9987 |
| Any CBVD | chr17 | 2,856,538 | chr17:2856538:C:T | t | c | 0.2343 | 0.059 | 7.07E-05 | +++++++++ | 1.661 | 8 | 0.9897 |
| Any CBVD | chr17 | 27,226,757 | chr17:27226757:A:G | a | g | -1.0964 | 0.2347 | 2.99E-06 | ---+----- | 21.442 | 8 | 0.006062 |
| Any CBVD | chr17 | 27,237,457 | chr17:27237457:G:A | a | g | 1.1043 | 0.2425 | 5.28E-06 | +++-+-+-+ | 23.656 | 8 | 0.002617 |
| Any CBVD | chr17 | 27,242,169 | chr17:27242169:G:A | a | g | 1.119 | 0.2416 | 3.64E-06 | +++-+-+-+ | 24.202 | 8 | 0.00212 |
| Any CBVD | chr17 | 35,209,546 | chr17:35209546:T:G | t | g | -0.7382 | 0.1645 | 7.22E-06 | -+--+---- | 16.689 | 8 | 0.03351 |
| Any CBVD | chr17 | 66,803,460 | chr17:66803460:C:T | t | c | 1.0137 | 0.2525 | 5.96E-05 | +++++--++ | 17.61 | 8 | 0.02435 |
| Any CBVD | chr17 | 68,103,580 | chr17:68103580:T:C | t | c | -0.3957 | 0.0952 | 3.25E-05 | -+--+--+- | 10.135 | 8 | 0.2556 |
| Any CBVD | chr17 | 68,103,584 | chr17:68103584:C:T | t | c | 0.3941 | 0.0953 | 3.57E-05 | +-++-++-+ | 10.101 | 8 | 0.258 |
| Any CBVD | chr17 | 68,103,650 | chr17:68103650:A:G | a | g | -0.3984 | 0.0922 | 1.56E-05 | ----+---- | 8.573 | 8 | 0.3796 |
| Any CBVD | chr17 | 68,103,876 | chr17:68103876:G:T | t | g | -0.232 | 0.0579 | 6.24E-05 | ----+-+-- | 12.771 | 8 | 0.12 |
| Any CBVD | chr17 | 68,110,503 | chr17:68110503:T:C | t | c | 0.231 | 0.058 | 6.71E-05 | ++++-+--+ | 14.498 | 8 | 0.06966 |
| Any CBVD | chr17 | 68,111,214 | chr17:68111214:T:C | t | c | 0.2301 | 0.0579 | 7.13E-05 | ++++-+--+ | 14.449 | 8 | 0.07079 |
| Any CBVD | chr17 | 68,111,294 | chr17:68111294:C:T | t | c | -0.2293 | 0.0579 | 7.57E-05 | ----+-++- | 14.395 | 8 | 0.07203 |
| Any CBVD | chr17 | 68,112,968 | chr17:68112968:A:C | a | c | 0.229 | 0.0579 | 7.75E-05 | ++++-+--+ | 14.416 | 8 | 0.07154 |
| Any CBVD | chr17 | 68,113,721 | chr17:68113721:A:T | a | t | 0.23 | 0.0579 | 7.12E-05 | ++++-+--+ | 13.716 | 8 | 0.08946 |
| Any CBVD | chr17 | 68,116,383 | chr17:68116383:T:C | t | c | 0.2317 | 0.0579 | 6.32E-05 | ++++-+--+ | 13.235 | 8 | 0.104 |
| Any CBVD | chr17 | 68,116,807 | chr17:68116807:G:A | a | g | -0.2253 | 0.0578 | 9.61E-05 | ----+-++- | 14.45 | 8 | 0.07077 |
| Any CBVD | chr17 | 68,116,851 | chr17:68116851:G:A | a | g | -0.2302 | 0.0579 | 6.92E-05 | ----+-++- | 13.839 | 8 | 0.08607 |
| Any CBVD | chr17 | 68,122,378 | chr17:68122378:A:G | a | g | -0.2267 | 0.0576 | 8.31E-05 | ----+-++- | 14.772 | 8 | 0.06374 |
| Any CBVD | chr17 | 68,124,068 | chr17:68124068:C:CTA | cta | c | 0.2308 | 0.0576 | 6.08E-05 | ++++-+--+ | 14.827 | 8 | 0.0626 |
| Any CBVD | chr17 | 68,124,415 | chr17:68124415:T:C | t | c | -0.227 | 0.0576 | 8.03E-05 | ----+-++- | 14.815 | 8 | 0.06284 |
| Any CBVD | chr17 | 68,125,159 | chr17:68125159:G:A | a | g | 0.229 | 0.0576 | 7.00E-05 | ++++-+--+ | 14.761 | 8 | 0.06396 |
| Any CBVD | chr17 | 68,127,291 | chr17:68127291:T:G | t | g | -0.2263 | 0.0575 | 8.38E-05 | ----+-++- | 14.863 | 8 | 0.06187 |
| Any CBVD | chr17 | 68,131,197 | chr17:68131197:A:T | a | t | -0.2278 | 0.0575 | 7.45E-05 | ----+-++- | 13.989 | 8 | 0.08204 |
| Any CBVD | chr17 | 68,131,217 | chr17:68131217:T:C | t | c | -0.2268 | 0.0574 | 7.84E-05 | ----+-++- | 13.944 | 8 | 0.08325 |
| Any CBVD | chr17 | 68,138,969 | chr17:68138969:G:A | a | g | 0.2244 | 0.0576 | 9.72E-05 | ++++-+--+ | 12.538 | 8 | 0.1287 |
| Any CBVD | chr17 | 68,140,179 | chr17:68140179:G:C | c | g | 0.225 | 0.0576 | 9.42E-05 | ++++-+--+ | 13.055 | 8 | 0.11 |
| Any CBVD | chr17 | 68,147,934 | chr17:68147934:C:A | a | c | 0.4746 | 0.1057 | 7.15E-06 | +++++++-+ | 1.142 | 8 | 0.9972 |
| Any CBVD | chr17 | 68,148,288 | chr17:68148288:C:T | t | c | 0.3981 | 0.0985 | 5.36E-05 | +++++++-+ | 2.222 | 8 | 0.9734 |
| Any CBVD | chr17 | 68,150,486 | chr17:68150486:C:G | c | g | -0.3969 | 0.0981 | 5.18E-05 | -------+- | 2.488 | 8 | 0.9623 |
| Any CBVD | chr17 | 68,151,065 | chr17:68151065:G:T | t | g | 0.5702 | 0.1407 | 5.06E-05 | +++++++-+ | 4.864 | 8 | 0.772 |
| Any CBVD | chr17 | 68,151,408 | chr17:68151408:G:C | c | g | 0.3385 | 0.0851 | 6.96E-05 | +++++++-+ | 2.671 | 8 | 0.9533 |
| Any CBVD | chr17 | 68,153,465 | chr17:68153465:G:A | a | g | 0.3957 | 0.098 | 5.39E-05 | +++++++-+ | 2.479 | 8 | 0.9627 |
| Any CBVD | chr17 | 68,153,545 | chr17:68153545:G:A | a | g | 0.3433 | 0.085 | 5.41E-05 | +++++++-+ | 2.548 | 8 | 0.9595 |
| Any CBVD | chr17 | 68,153,992 | chr17:68153992:T:A | a | t | 0.3607 | 0.0916 | 8.22E-05 | +++++++-+ | 1.948 | 8 | 0.9826 |
| Any CBVD | chr17 | 68,157,200 | chr17:68157200:C:T | t | c | 0.4116 | 0.0979 | 2.62E-05 | +++++++-+ | 2.801 | 8 | 0.9462 |
| Any CBVD | chr17 | 68,160,385 | chr17:68160385:T:C | t | c | -0.3416 | 0.0849 | 5.78E-05 | -------+- | 2.342 | 8 | 0.9687 |
| Any CBVD | chr17 | 68,160,405 | chr17:68160405:C:T | t | c | 0.3412 | 0.0849 | 5.90E-05 | +++++++-+ | 2.357 | 8 | 0.9681 |
| Any CBVD | chr17 | 81,439,956 | chr17:81439956:T:G | t | g | -0.2551 | 0.0644 | 7.46E-05 | -+-+----- | 7.242 | 8 | 0.5107 |
| Any CBVD | chr18 | 7,446,495 | chr18:7446495:G:C | c | g | -0.2952 | 0.0744 | 7.18E-05 | -+--+-++- | 8.755 | 8 | 0.3634 |
| Any CBVD | chr18 | 7,447,016 | chr18:7447016:T:C | t | c | 0.3004 | 0.0744 | 5.36E-05 | +-++-+--+ | 8.746 | 8 | 0.3641 |
| Any CBVD | chr18 | 7,447,700 | chr18:7447700:A:C | a | c | 0.2981 | 0.0744 | 6.10E-05 | +-++-+--+ | 9.148 | 8 | 0.33 |
| Any CBVD | chr18 | 14,815,044 | chr18:14815044:G:A | a | g | 1.1078 | 0.2835 | 9.30E-05 | +-+-++?+? | 9.833 | 6 | 0.1319 |
| Any CBVD | chr18 | 33,926,754 | chr18:33926754:C:T | t | c | 0.6032 | 0.1503 | 6.02E-05 | +-++-++++ | 8.076 | 8 | 0.4261 |

| | | | | | | | | | | | | |
|---|---|---|---|---|---|---|---|---|---|---|---|---|
| Any CBVD | chr18 | 45,633,164 | chr18:45633164:C:T | t | c | 1.1402 | 0.2751 | 3.40E-05 | +?+--?+++ | 12.204 | 6 | 0.05757 |
| Any CBVD | chr18 | 46,558,634 | chr18:46558634:A:T | a | t | -0.3486 | 0.0832 | 2.82E-05 | -----+--- | 10.033 | 8 | 0.2627 |
| Any CBVD | chr18 | 46,558,798 | chr18:46558798:T:G | t | g | -0.2891 | 0.0649 | 8.53E-06 | --------- | 7.249 | 8 | 0.51 |
| Any CBVD | chr18 | 46,559,930 | chr18:46559930:A:G | a | g | -0.3324 | 0.0788 | 2.45E-05 | -----+--- | 11.409 | 8 | 0.1796 |
| Any CBVD | chr18 | 46,561,833 | chr18:46561833:T:C | t | c | -0.3255 | 0.0801 | 4.87E-05 | -----+--- | 11.721 | 8 | 0.1641 |
| Any CBVD | chr18 | 46,564,093 | chr18:46564093:AGT:A | a | agt | 0.2913 | 0.0731 | 6.75E-05 | +++++-+-+ | 14.231 | 8 | 0.07593 |
| Any CBVD | chr18 | 46,564,117 | chr18:46564117:T:C | t | c | -0.3155 | 0.0804 | 8.68E-05 | -----+--- | 10.098 | 8 | 0.2582 |
| Any CBVD | chr18 | 46,565,340 | chr18:46565340:A:G | a | g | -0.3154 | 0.0802 | 8.36E-05 | -----+--- | 10.366 | 8 | 0.2403 |
| Any CBVD | chr18 | 46,567,796 | chr18:46567796:T:G | t | g | -0.3196 | 0.0801 | 6.56E-05 | -----+--- | 10.986 | 8 | 0.2025 |
| Any CBVD | chr18 | 46,569,511 | chr18:46569511:G:A | a | g | 0.3165 | 0.0799 | 7.42E-05 | +++++-+++ | 9.567 | 8 | 0.2968 |
| Any CBVD | chr18 | 47,758,270 | chr18:47758270:A:G | a | g | -0.712 | 0.1614 | 1.02E-05 | ----+---- | 6.051 | 8 | 0.6415 |
| Any CBVD | chr18 | 47,790,752 | chr18:47790752:C:T | t | c | 0.6674 | 0.1713 | 9.75E-05 | ++++-++++ | 5.7 | 8 | 0.6808 |
| Any CBVD | chr18 | 47,843,171 | chr18:47843171:A:G | a | g | -0.7405 | 0.1768 | 2.82E-05 | --------- | 6.311 | 8 | 0.6124 |
| Any CBVD | chr18 | 48,775,638 | chr18:48775638:G:T | t | g | 0.5552 | 0.1416 | 8.84E-05 | +++++++++ | 16.294 | 8 | 0.03836 |
| Any CBVD | chr18 | 48,776,429 | chr18:48776429:G:A | a | g | 0.559 | 0.1417 | 8.04E-05 | +++++++++ | 16.232 | 8 | 0.03918 |
| Any CBVD | chr19 | 5,548,968 | chr19:5548968:T:G | t | g | -0.6483 | 0.1595 | 4.80E-05 | --------- | 5.168 | 8 | 0.7394 |
| Any CBVD | chr19 | 7,040,211 | chr19:7040211:G:A | a | g | 1.2719 | 0.3072 | 3.48E-05 | +++++++-+ | 8.894 | 8 | 0.3513 |
| Any CBVD | chr19 | 28,919,507 | chr19:28919507:C:A | a | c | 0.4072 | 0.1019 | 6.41E-05 | +++-+++++ | 10.361 | 8 | 0.2406 |
| Any CBVD | chr19 | 28,976,108 | chr19:28976108:A:ATG | a | atg | -0.3559 | 0.0887 | 6.00E-05 | ---+----- | 5.8 | 8 | 0.6697 |
| Any CBVD | chr19 | 31,140,034 | chr19:31140034:G:A | a | g | 1.2264 | 0.3008 | 4.56E-05 | +?+++++-? | 16.806 | 6 | 0.01002 |
| Any CBVD | chr19 | 31,143,807 | chr19:31143807:A:G | a | g | -1.2398 | 0.2996 | 3.50E-05 | -?-----+? | 16.992 | 6 | 0.009314 |
| Any CBVD | chr19 | 41,592,189 | chr19:41592189:G:A | a | g | 0.2557 | 0.0627 | 4.59E-05 | +-+++++-+ | 26.411 | 8 | 0.000893 |
| Any CBVD | chr19 | 44,883,210 | chr19:44883210:G:GTAA | g | gtaa | 0.2763 | 0.0652 | 2.26E-05 | ++-++++++ | 18.635 | 8 | 0.01694 |
| Any CBVD | chr19 | 44,884,202 | chr19:44884202:C:G | c | g | 0.2778 | 0.0651 | 1.95E-05 | ++-++++++ | 18.859 | 8 | 0.01563 |
| Any CBVD | chr19 | 44,884,339 | chr19:44884339:G:A | a | g | -0.2779 | 0.065 | 1.92E-05 | --+------ | 18.876 | 8 | 0.01554 |
| Any CBVD | chr19 | 44,884,873 | chr19:44884873:G:A | a | g | -0.2637 | 0.0645 | 4.40E-05 | --+------ | 17.614 | 8 | 0.02431 |
| Any CBVD | chr19 | 44,885,243 | chr19:44885243:A:G | a | g | 0.2838 | 0.0589 | 1.47E-06 | ++-++++++ | 23.048 | 8 | 0.003303 |
| Any CBVD | chr19 | 44,887,076 | chr19:44887076:A:G | a | g | 0.2935 | 0.0587 | 5.75E-07 | +++++++++ | 22.2 | 8 | 0.004559 |
| Any CBVD | chr19 | 44,888,997 | chr19:44888997:C:T | t | c | -0.3747 | 0.0611 | 8.43E-10 | --------- | 24.428 | 8 | 0.001942 |
| Any CBVD | chr19 | 44,891,079 | chr19:44891079:T:C | t | c | 0.2718 | 0.0644 | 2.47E-05 | ++-++++++ | 16.083 | 8 | 0.04121 |
| Any CBVD | chr19 | 44,891,712 | chr19:44891712:T:G | t | g | 0.285 | 0.0588 | 1.26E-06 | +++++++++ | 20.54 | 8 | 0.008476 |
| Any CBVD | chr19 | 44,892,362 | chr19:44892362:A:G | a | g | 0.2856 | 0.0643 | 8.85E-06 | ++-++++++ | 18.323 | 8 | 0.01893 |
| Any CBVD | chr19 | 44,892,457 | chr19:44892457:T:C | t | c | 0.2925 | 0.0587 | 6.17E-07 | ++-++++++ | 22.049 | 8 | 0.004826 |
| Any CBVD | chr19 | 44,892,652 | chr19:44892652:C:G | c | g | 0.289 | 0.0647 | 7.83E-06 | ++-++++++ | 18.154 | 8 | 0.0201 |
| Any CBVD | chr19 | 44,892,887 | chr19:44892887:C:T | t | c | -0.2884 | 0.0647 | 8.22E-06 | --+------ | 18.221 | 8 | 0.01963 |
| Any CBVD | chr19 | 44,892,962 | chr19:44892962:C:T | t | c | -0.2912 | 0.0586 | 6.73E-07 | --+------ | 22.11 | 8 | 0.004716 |
| Any CBVD | chr19 | 44,893,408 | chr19:44893408:G:T | t | g | -0.3001 | 0.0592 | 3.92E-07 | --+------ | 26.409 | 8 | 0.000894 |
| Any CBVD | chr19 | 44,894,050 | chr19:44894050:C:T | t | c | -0.4747 | 0.1205 | 8.19E-05 | -+------- | 10.837 | 8 | 0.2111 |
| Any CBVD | chr19 | 44,894,695 | chr19:44894695:T:C | t | c | 0.4775 | 0.1201 | 7.01E-05 | +-+++++++ | 10.657 | 8 | 0.2219 |
| Any CBVD | chr19 | 44,895,528 | chr19:44895528:C:T | t | c | -0.4774 | 0.1207 | 7.60E-05 | -+------- | 10.719 | 8 | 0.2181 |
| Any CBVD | chr19 | 44,896,087 | chr19:44896087:G:T | t | g | -0.4779 | 0.1205 | 7.35E-05 | -+------- | 10.67 | 8 | 0.2211 |
| Any CBVD | chr19 | 44,896,639 | chr19:44896639:G:A | a | g | -0.4782 | 0.1207 | 7.40E-05 | -+------- | 10.709 | 8 | 0.2188 |
| Any CBVD | chr19 | 44,897,227 | chr19:44897227:T:TG | t | tg | 0.4847 | 0.1205 | 5.81E-05 | +-+++++++ | 10.864 | 8 | 0.2095 |
| Any CBVD | chr19 | 44,897,468 | chr19:44897468:C:T | t | c | -0.4779 | 0.1205 | 7.34E-05 | -+------- | 10.67 | 8 | 0.2211 |
| Any CBVD | chr19 | 44,898,611 | chr19:44898611:T:C | t | c | 0.5195 | 0.1105 | 2.59E-06 | +-+++++++ | 9.113 | 8 | 0.3328 |
| Any CBVD | chr19 | 44,899,959 | chr19:44899959:C:T | t | c | -0.4836 | 0.1173 | 3.77E-05 | -+------- | 9.315 | 8 | 0.3164 |

| | | | | | | | | | | | | |
|---|---|---|---|---|---|---|---|---|---|---|---|---|
| Any CBVD | chr19 | 44,900,155 | chr19:44900155:C:T | t | c | 0.2458 | 0.0579 | 2.21E-05 | +-+++++++ | 10.722 | 8 | 0.218 |
| Any CBVD | chr19 | 44,900,601 | chr19:44900601:A:G | a | g | -0.2475 | 0.0582 | 2.10E-05 | -+------- | 10.746 | 8 | 0.2165 |
| Any CBVD | chr19 | 44,901,174 | chr19:44901174:T:C | t | c | -0.2474 | 0.0583 | 2.19E-05 | -+------- | 10.865 | 8 | 0.2095 |
| Any CBVD | chr19 | 44,901,600 | chr19:44901600:T:C | t | c | 0.4875 | 0.1173 | 3.26E-05 | +-+++++++ | 9.394 | 8 | 0.3101 |
| Any CBVD | chr19 | 44,901,715 | chr19:44901715:T:C | t | c | -0.2466 | 0.0583 | 2.32E-05 | -+------- | 10.92 | 8 | 0.2063 |
| Any CBVD | chr19 | 44,901,805 | chr19:44901805:A:G | a | g | -0.2437 | 0.0582 | 2.80E-05 | -+------- | 10.701 | 8 | 0.2192 |
| Any CBVD | chr19 | 44,903,281 | chr19:44903281:C:CG | cg | c | 0.2364 | 0.0589 | 6.04E-05 | +-+++++++ | 10.569 | 8 | 0.2274 |
| Any CBVD | chr19 | 44,903,416 | chr19:44903416:G:A | a | g | -0.3678 | 0.0573 | 1.34E-10 | -+------- | 29.68 | 8 | 0.000241 |
| Any CBVD | chr19 | 44,904,531 | chr19:44904531:G:A | a | g | 0.2562 | 0.0583 | 1.11E-05 | +-+++++++ | 10.381 | 8 | 0.2393 |
| Any CBVD | chr19 | 44,906,745 | chr19:44906745:G:A | a | g | -0.3623 | 0.0659 | 3.83E-08 | --+------ | 24.217 | 8 | 0.002107 |
| Any CBVD | chr19 | 44,908,684 | chr19:44908684:T:C | t | c | 0.4542 | 0.0614 | 1.41E-13 | +++++++++ | 33.667 | 8 | 4.67E-05 |
| Any CBVD | chr19 | 44,909,698 | chr19:44909698:A:C | a | c | 0.4962 | 0.117 | 2.21E-05 | +-+++++++ | 9.034 | 8 | 0.3394 |
| Any CBVD | chr19 | 44,910,319 | chr19:44910319:C:T | t | c | -0.2517 | 0.0624 | 5.50E-05 | --+------ | 19.571 | 8 | 0.01209 |
| Any CBVD | chr19 | 44,912,456 | chr19:44912456:G:A | a | g | -0.351 | 0.0661 | 1.08E-07 | --------- | 20.819 | 8 | 0.007643 |
| Any CBVD | chr19 | 44,912,678 | chr19:44912678:G:T | t | g | -0.3467 | 0.066 | 1.53E-07 | --+------ | 21.459 | 8 | 0.006023 |
| Any CBVD | chr19 | 44,912,921 | chr19:44912921:G:T | t | g | -0.3412 | 0.059 | 7.22E-09 | --------- | 25.895 | 8 | 0.001095 |
| Any CBVD | chr19 | 44,913,034 | chr19:44913034:C:T | t | c | 0.2672 | 0.0609 | 1.16E-05 | +-+++++++ | 8.232 | 8 | 0.4111 |
| Any CBVD | chr19 | 44,913,484 | chr19:44913484:C:T | t | c | -0.346 | 0.0589 | 4.37E-09 | --------- | 25.584 | 8 | 0.001237 |
| Any CBVD | chr19 | 44,915,533 | chr19:44915533:T:C | t | c | 0.3544 | 0.0592 | 2.13E-09 | +++++++++ | 25.109 | 8 | 0.00149 |
| Any CBVD | chr19 | 44,916,825 | chr19:44916825:A:C | a | c | 0.3065 | 0.0663 | 3.74E-06 | ++-++++++ | 19.557 | 8 | 0.01215 |
| Any CBVD | chr19 | 44,917,843 | chr19:44917843:G:A | a | g | 0.2677 | 0.061 | 1.16E-05 | +-++-++++ | 11.419 | 8 | 0.1791 |
| Any CBVD | chr19 | 44,917,947 | chr19:44917947:C:T | t | c | -0.5057 | 0.1168 | 1.49E-05 | -+------- | 8.581 | 8 | 0.3789 |
| Any CBVD | chr19 | 44,918,393 | chr19:44918393:G:A | a | g | -0.4935 | 0.1166 | 2.33E-05 | -+------- | 9.21 | 8 | 0.3249 |
| Any CBVD | chr19 | 44,918,903 | chr19:44918903:C:G | c | g | 0.3656 | 0.0611 | 2.21E-09 | +++++++++ | 28.904 | 8 | 0.00033 |
| Any CBVD | chr19 | 44,919,589 | chr19:44919589:G:A | a | g | -0.3724 | 0.0609 | 9.84E-10 | --------- | 28.752 | 8 | 0.000351 |
| Any CBVD | chr19 | 44,919,689 | chr19:44919689:A:G | a | g | 0.3699 | 0.0607 | 1.09E-09 | +++++++++ | 29.121 | 8 | 0.000302 |
| Any CBVD | chr19 | 44,920,379 | chr19:44920379:G:A | a | g | 0.2561 | 0.0647 | 7.44E-05 | +-+++++++ | 13.852 | 8 | 0.08571 |
| Any CBVD | chr19 | 44,921,809 | chr19:44921809:G:A | a | g | -0.501 | 0.1207 | 3.32E-05 | -+------- | 9.876 | 8 | 0.2738 |
| Any CBVD | chr19 | 44,923,868 | chr19:44923868:T:A | a | t | -0.2613 | 0.0657 | 7.04E-05 | --+--+--- | 22.343 | 8 | 0.004319 |
| Any CBVD | chr19 | 44,924,977 | chr19:44924977:G:A | a | g | -0.26 | 0.0657 | 7.61E-05 | --+--+--- | 22.794 | 8 | 0.003639 |
| Any CBVD | chr19 | 44,932,959 | chr19:44932959:A:G | a | g | -0.2552 | 0.064 | 6.71E-05 | --------- | 14.167 | 8 | 0.07751 |
| Any CBVD | chr19 | 44,932,987 | chr19:44932987:T:C | t | c | -0.2529 | 0.064 | 7.80E-05 | --------- | 12.6 | 8 | 0.1264 |
| Any CBVD | chr19 | 44,932,990 | chr19:44932990:T:C | t | c | -0.2516 | 0.064 | 8.48E-05 | --------- | 12.624 | 8 | 0.1255 |
| Any CBVD | chr19 | 44,935,297 | chr19:44935297:C:T | t | c | -0.2658 | 0.0641 | 3.34E-05 | -+------- | 16.885 | 8 | 0.03133 |
| Any CBVD | chr19 | 52,420,510 | chr19:52420510:T:C | t | c | 0.2359 | 0.0602 | 9.00E-05 | +++++++-+ | 3.258 | 8 | 0.9171 |
| Any CBVD | chr19 | 54,394,495 | chr19:54394495:C:T | t | c | -0.3334 | 0.0749 | 8.59E-06 | --------- | 5.952 | 8 | 0.6526 |
| Any CBVD | chr19 | 54,400,808 | chr19:54400808:G:A | a | g | -0.3499 | 0.073 | 1.61E-06 | --------- | 3.427 | 8 | 0.9048 |
| Any CBVD | chr19 | 54,401,120 | chr19:54401120:G:A | a | g | -0.2792 | 0.07 | 6.62E-05 | -------+- | 5.215 | 8 | 0.7344 |
| Any CBVD | chr19 | 54,403,408 | chr19:54403408:G:T | t | g | -0.355 | 0.0723 | 9.24E-07 | --------- | 3.636 | 8 | 0.8884 |
| Any CBVD | chr19 | 54,403,663 | chr19:54403663:T:C | t | c | 0.2583 | 0.0662 | 9.62E-05 | +++++++++ | 2.699 | 8 | 0.9518 |
| Any CBVD | chr19 | 54,404,017 | chr19:54404017:C:T | t | c | -0.3527 | 0.0723 | 1.08E-06 | --------- | 3.545 | 8 | 0.8956 |
| Any CBVD | chr19 | 54,404,276 | chr19:54404276:T:C | t | c | 0.3394 | 0.0729 | 3.20E-06 | +++++++++ | 3.844 | 8 | 0.8709 |
| Any CBVD | chr19 | 54,407,306 | chr19:54407306:A:G | a | g | 0.3246 | 0.0702 | 3.74E-06 | +++++++++ | 2.069 | 8 | 0.9788 |
| Any CBVD | chr19 | 54,412,034 | chr19:54412034:C:T | t | c | -0.353 | 0.0731 | 1.36E-06 | --------- | 4.167 | 8 | 0.8417 |
| Any CBVD | chr2 | 10,159,720 | chr2:10159720:C:T | t | c | 0.6282 | 0.1585 | 7.42E-05 | ++++-++++ | 9.347 | 8 | 0.3139 |
| Any CBVD | chr2 | 11,887,219 | chr2:11887219:T:C | t | c | -1.5901 | 0.3977 | 6.37E-05 | ?--?????- | 2.815 | 2 | 0.2448 |

| | | | | | | | | | | | | |
|---|---|---|---|---|---|---|---|---|---|---|---|---|
| Any CBVD | chr2 | 15,027,822 | chr2:15027822:C:T | t | c | 0.9318 | 0.2325 | 6.11E-05 | +++++++++ | 15.017 | 8 | 0.05881 |
| Any CBVD | chr2 | 15,073,448 | chr2:15073448:C:G | c | g | -0.9682 | 0.2396 | 5.34E-05 | ----?---- | 13.661 | 7 | 0.05754 |
| Any CBVD | chr2 | 15,591,780 | chr2:15591780:T:C | t | c | -1.3706 | 0.3423 | 6.24E-05 | -+--?-??? | 5.996 | 4 | 0.1994 |
| Any CBVD | chr2 | 41,237,409 | chr2:41237409:C:T | t | c | 0.4021 | 0.1016 | 7.55E-05 | +++++++-+ | 4.088 | 8 | 0.8491 |
| Any CBVD | chr2 | 41,241,426 | chr2:41241426:G:A | a | g | 0.426 | 0.1053 | 5.25E-05 | +++++++++ | 7.449 | 8 | 0.489 |
| Any CBVD | chr2 | 46,703,374 | chr2:46703374:G:A | a | g | -0.2448 | 0.0623 | 8.57E-05 | -+--+--+- | 8.85 | 8 | 0.3551 |
| Any CBVD | chr2 | 46,710,202 | chr2:46710202:A:G | a | g | 0.2493 | 0.0624 | 6.47E-05 | +-++-++-+ | 8.778 | 8 | 0.3614 |
| Any CBVD | chr2 | 46,723,282 | chr2:46723282:G:T | t | g | -0.2427 | 0.062 | 9.15E-05 | -+--+--+- | 6.935 | 8 | 0.5436 |
| Any CBVD | chr2 | 46,728,046 | chr2:46728046:T:C | t | c | 0.2445 | 0.0624 | 8.90E-05 | +-++-++-+ | 8.696 | 8 | 0.3686 |
| Any CBVD | chr2 | 46,728,808 | chr2:46728808:T:C | t | c | 0.2432 | 0.0624 | 9.75E-05 | +-++-++-+ | 8.657 | 8 | 0.3721 |
| Any CBVD | chr2 | 46,765,424 | chr2:46765424:G:A | a | g | -0.2432 | 0.0622 | 9.27E-05 | -+--+--+- | 10.369 | 8 | 0.2401 |
| Any CBVD | chr2 | 50,987,893 | chr2:50987893:T:C | t | c | -0.9748 | 0.2339 | 3.07E-05 | --+------ | 13.524 | 8 | 0.09506 |
| Any CBVD | chr2 | 51,069,561 | chr2:51069561:C:G | c | g | -0.9065 | 0.23 | 8.10E-05 | --------- | 12.26 | 8 | 0.14 |
| Any CBVD | chr2 | 51,098,339 | chr2:51098339:G:A | a | g | 0.9965 | 0.247 | 5.46E-05 | ++++++-+? | 7.441 | 7 | 0.3845 |
| Any CBVD | chr2 | 57,584,545 | chr2:57584545:T:C | t | c | -1.3402 | 0.337 | 6.97E-05 | -??????+- | 0.89 | 2 | 0.641 |
| Any CBVD | chr2 | 57,584,547 | chr2:57584547:A:G | a | g | -1.3396 | 0.337 | 7.02E-05 | -??????+- | 0.889 | 2 | 0.6411 |
| Any CBVD | chr2 | 60,539,250 | chr2:60539250:C:T | t | c | 0.92 | 0.2276 | 5.31E-05 | ++++-+++- | 18.47 | 8 | 0.01797 |
| Any CBVD | chr2 | 60,541,647 | chr2:60541647:A:G | a | g | -0.6608 | 0.1556 | 2.18E-05 | ----+---- | 12.12 | 8 | 0.1459 |
| Any CBVD | chr2 | 65,842,094 | chr2:65842094:G:A | a | g | 0.589 | 0.1403 | 2.70E-05 | +++++++-+ | 5.306 | 8 | 0.7244 |
| Any CBVD | chr2 | 65,854,472 | chr2:65854472:A:G | a | g | -0.2634 | 0.0668 | 7.95E-05 | -+-----+- | 9.376 | 8 | 0.3116 |
| Any CBVD | chr2 | 65,863,794 | chr2:65863794:C:T | t | c | 0.2654 | 0.0672 | 7.89E-05 | +-++++--+ | 9.846 | 8 | 0.276 |
| Any CBVD | chr2 | 75,718,832 | chr2:75718832:C:A | a | c | 0.4773 | 0.1183 | 5.43E-05 | ++++++-++ | 5.678 | 8 | 0.6832 |
| Any CBVD | chr2 | 75,721,063 | chr2:75721063:T:C | t | c | -0.4672 | 0.119 | 8.69E-05 | ------+-- | 5.205 | 8 | 0.7354 |
| Any CBVD | chr2 | 88,568,327 | chr2:88568327:A:G | a | g | -1.9832 | 0.4849 | 4.32E-05 | ?---??--? | 5.171 | 4 | 0.2702 |
| Any CBVD | chr2 | 111,805,188 | chr2:111805188:A:G | a | g | -0.9265 | 0.2209 | 2.75E-05 | ------++- | 5.804 | 8 | 0.6691 |
| Any CBVD | chr2 | 122,368,293 | chr2:122368293:A:G | a | g | -0.2683 | 0.0671 | 6.38E-05 | -+----+-- | 12.42 | 8 | 0.1334 |
| Any CBVD | chr2 | 160,908,890 | chr2:160908890:T:A | a | t | 0.5868 | 0.1445 | 4.87E-05 | +++-++-++ | 11.196 | 8 | 0.1908 |
| Any CBVD | chr2 | 162,602,595 | chr2:162602595:C:G | c | g | -1.1729 | 0.2743 | 1.90E-05 | -+-+----? | 13.315 | 7 | 0.06481 |
| Any CBVD | chr2 | 162,602,894 | chr2:162602894:TGAG:T | t | tgag | 0.933 | 0.2255 | 3.51E-05 | +++++++++ | 10.47 | 8 | 0.2336 |
| Any CBVD | chr2 | 162,620,221 | chr2:162620221:TAGAG:T | t | tagag | 1.2217 | 0.273 | 7.66E-06 | +-+-++++? | 13.445 | 7 | 0.06198 |
| Any CBVD | chr2 | 162,624,379 | chr2:162624379:A:T | a | t | -0.8701 | 0.2228 | 9.38E-05 | --------- | 8.225 | 8 | 0.4118 |
| Any CBVD | chr2 | 162,638,611 | chr2:162638611:G:A | a | g | 0.9134 | 0.2269 | 5.71E-05 | +++++++++ | 8.752 | 8 | 0.3637 |
| Any CBVD | chr2 | 162,642,766 | chr2:162642766:A:G | a | g | -0.9092 | 0.2268 | 6.11E-05 | --------- | 8.79 | 8 | 0.3603 |
| Any CBVD | chr2 | 162,644,263 | chr2:162644263:TAAG:T | t | taag | 0.9111 | 0.2267 | 5.84E-05 | +++++++++ | 8.804 | 8 | 0.3591 |
| Any CBVD | chr2 | 162,644,469 | chr2:162644469:G:C | c | g | 0.9117 | 0.2267 | 5.79E-05 | +++++++++ | 8.788 | 8 | 0.3605 |
| Any CBVD | chr2 | 170,491,769 | chr2:170491769:G:T | t | g | 2.0331 | 0.5184 | 8.78E-05 | +???-+??? | 1.61 | 2 | 0.4471 |
| Any CBVD | chr2 | 170,532,853 | chr2:170532853:C:T | t | c | 0.9584 | 0.2027 | 2.27E-06 | +-+++++++ | 8.671 | 8 | 0.3708 |
| Any CBVD | chr2 | 170,560,456 | chr2:170560456:A:G | a | g | -0.7918 | 0.1842 | 1.72E-05 | -+-+----- | 13.292 | 8 | 0.1022 |
| Any CBVD | chr2 | 171,769,066 | chr2:171769066:C:T | t | c | 2.1031 | 0.4711 | 8.04E-06 | ?-+?????+ | 3.084 | 2 | 0.214 |
| Any CBVD | chr2 | 171,935,019 | chr2:171935019:G:A | a | g | 2.1392 | 0.4763 | 7.09E-06 | ?-+?????+ | 2.381 | 2 | 0.3041 |
| Any CBVD | chr2 | 175,748,333 | chr2:175748333:TA:T | t | ta | 1.7452 | 0.3945 | 9.69E-06 | ??++???++ | 3.383 | 3 | 0.3362 |
| Any CBVD | chr2 | 175,783,910 | chr2:175783910:A:T | a | t | -1.5979 | 0.3672 | 1.35E-05 | ??--??--- | 7.287 | 4 | 0.1215 |
| Any CBVD | chr2 | 202,458,477 | chr2:202458477:A:T | a | t | -2.0499 | 0.5156 | 7.00E-05 | -???+-??? | 1.457 | 2 | 0.4826 |
| Any CBVD | chr2 | 203,305,226 | chr2:203305226:G:T | t | g | -0.3499 | 0.0887 | 7.95E-05 | ----+---- | 5.617 | 8 | 0.69 |
| Any CBVD | chr2 | 206,476,773 | chr2:206476773:T:G | t | g | -0.9585 | 0.24 | 6.52E-05 | -+-+---+- | 10.235 | 8 | 0.2489 |
| Any CBVD | chr2 | 211,416,947 | chr2:211416947:G:A | a | g | 0.844 | 0.1964 | 1.72E-05 | +++++++++ | 8.538 | 8 | 0.3827 |

| | | | | | | | | | | | | |
|---|---|---|---|---|---|---|---|---|---|---|---|---|
| Any CBVD | chr2 | 211,420,614 | chr2:211420614:AAAAG:A | a | aaaag | 0.8331 | 0.1969 | 2.33E-05 | +++++++++ | 7.951 | 8 | 0.4383 |
| Any CBVD | chr2 | 211,421,030 | chr2:211421030:G:T | t | g | 0.8584 | 0.1978 | 1.43E-05 | +++++++++ | 9.11 | 8 | 0.3331 |
| Any CBVD | chr2 | 211,445,583 | chr2:211445583:C:G | c | g | -0.8979 | 0.2079 | 1.57E-05 | --------- | 8.075 | 8 | 0.4261 |
| Any CBVD | chr2 | 211,453,672 | chr2:211453672:T:C | t | c | -0.8945 | 0.2081 | 1.71E-05 | --------- | 8.077 | 8 | 0.4259 |
| Any CBVD | chr2 | 211,456,181 | chr2:211456181:G:A | a | g | 0.9025 | 0.2177 | 3.38E-05 | +++++++++ | 7.898 | 8 | 0.4435 |
| Any CBVD | chr2 | 211,456,370 | chr2:211456370:C:G | c | g | -0.9049 | 0.2178 | 3.25E-05 | --------- | 7.968 | 8 | 0.4366 |
| Any CBVD | chr2 | 211,458,675 | chr2:211458675:TTC:T | t | ttc | 0.875 | 0.2188 | 6.38E-05 | +++++++++ | 6.873 | 8 | 0.5504 |
| Any CBVD | chr2 | 211,465,137 | chr2:211465137:A:C | a | c | -0.8882 | 0.2087 | 2.08E-05 | --------- | 8.268 | 8 | 0.4077 |
| Any CBVD | chr2 | 211,465,178 | chr2:211465178:T:C | t | c | -0.893 | 0.2046 | 1.27E-05 | --------- | 8.24 | 8 | 0.4104 |
| Any CBVD | chr2 | 215,278,058 | chr2:215278058:A:C | a | c | -0.226 | 0.0569 | 7.18E-05 | ----+---- | 6.163 | 8 | 0.629 |
| Any CBVD | chr2 | 215,280,008 | chr2:215280008:A:T | a | t | -0.2245 | 0.0577 | 9.90E-05 | ----+---- | 7.702 | 8 | 0.4631 |
| Any CBVD | chr2 | 215,280,451 | chr2:215280451:T:C | t | c | -0.2367 | 0.0568 | 3.12E-05 | ----+---- | 6.53 | 8 | 0.5881 |
| Any CBVD | chr2 | 215,280,961 | chr2:215280961:A:T | a | t | 0.2374 | 0.0568 | 2.92E-05 | ++++-++++ | 6.712 | 8 | 0.568 |
| Any CBVD | chr2 | 215,281,148 | chr2:215281148:A:G | a | g | 0.2356 | 0.0568 | 3.31E-05 | ++++-++++ | 6.681 | 8 | 0.5714 |
| Any CBVD | chr2 | 215,283,070 | chr2:215283070:G:T | t | g | 0.248 | 0.0625 | 7.15E-05 | +++++++++ | 4.161 | 8 | 0.8424 |
| Any CBVD | chr2 | 215,284,231 | chr2:215284231:A:C | a | c | 0.2325 | 0.0567 | 4.17E-05 | ++++-++++ | 6.728 | 8 | 0.5662 |
| Any CBVD | chr2 | 215,284,370 | chr2:215284370:C:A | a | c | -0.2328 | 0.0567 | 4.01E-05 | ----+---- | 6.757 | 8 | 0.563 |
| Any CBVD | chr2 | 215,288,287 | chr2:215288287:G:A | a | g | 0.2491 | 0.0623 | 6.37E-05 | +++++++++ | 4.607 | 8 | 0.7987 |
| Any CBVD | chr2 | 215,290,493 | chr2:215290493:T:C | t | c | -0.2453 | 0.0625 | 8.72E-05 | --------- | 4.86 | 8 | 0.7724 |
| Any CBVD | chr2 | 235,094,660 | chr2:235094660:A:T | a | t | -0.679 | 0.1721 | 7.97E-05 | ------+-- | 11.132 | 8 | 0.1943 |
| Any CBVD | chr2 | 238,729,659 | chr2:238729659:T:C | t | c | -0.3128 | 0.0742 | 2.52E-05 | --------- | 3.55 | 8 | 0.8953 |
| Any CBVD | chr20 | 4,120,278 | chr20:4120278:A:C | a | c | -3.2417 | 0.7602 | 2.01E-05 | ?-??-??-? | 1.207 | 2 | 0.5468 |
| Any CBVD | chr20 | 8,588,467 | chr20:8588467:G:T | t | g | 0.9428 | 0.2414 | 9.37E-05 | +++++-+++ | 10.711 | 8 | 0.2186 |
| Any CBVD | chr20 | 8,588,808 | chr20:8588808:C:T | t | c | 0.9428 | 0.2414 | 9.38E-05 | +++++-+++ | 10.711 | 8 | 0.2186 |
| Any CBVD | chr20 | 8,591,829 | chr20:8591829:C:T | t | c | 0.9391 | 0.2413 | 9.95E-05 | +++++--++ | 10.66 | 8 | 0.2217 |
| Any CBVD | chr20 | 13,244,004 | chr20:13244004:A:G | a | g | 0.5301 | 0.1321 | 6.01E-05 | +++++-+++ | 7.168 | 8 | 0.5187 |
| Any CBVD | chr20 | 13,256,256 | chr20:13256256:C:T | t | c | -0.5397 | 0.1337 | 5.43E-05 | -----+-+- | 7.001 | 8 | 0.5366 |
| Any CBVD | chr20 | 17,237,647 | chr20:17237647:C:T | t | c | -0.2213 | 0.0569 | 9.91E-05 | -+------- | 8.341 | 8 | 0.4008 |
| Any CBVD | chr20 | 17,240,995 | chr20:17240995:C:T | t | c | -0.223 | 0.0568 | 8.62E-05 | -+------- | 7.897 | 8 | 0.4436 |
| Any CBVD | chr20 | 17,248,099 | chr20:17248099:A:G | a | g | 0.2208 | 0.0565 | 9.37E-05 | +-+++++++ | 8.459 | 8 | 0.3899 |
| Any CBVD | chr20 | 37,933,305 | chr20:37933305:A:G | a | g | -1.0737 | 0.2629 | 4.43E-05 | -+--+?-?- | 3.787 | 6 | 0.7054 |
| Any CBVD | chr20 | 41,878,120 | chr20:41878120:G:A | a | g | 0.6592 | 0.1604 | 3.98E-05 | +++++-+++ | 16.416 | 8 | 0.0368 |
| Any CBVD | chr21 | 19,395,033 | chr21:19395033:G:A | a | g | -0.6645 | 0.1631 | 4.62E-05 | ----+---- | 5.952 | 8 | 0.6526 |
| Any CBVD | chr21 | 19,398,235 | chr21:19398235:A:T | a | t | 0.6934 | 0.1623 | 1.93E-05 | ++++-++++ | 6.316 | 8 | 0.6119 |
| Any CBVD | chr21 | 19,399,807 | chr21:19399807:C:G | c | g | 0.6713 | 0.1632 | 3.92E-05 | ++++-++++ | 5.885 | 8 | 0.6601 |
| Any CBVD | chr21 | 19,403,275 | chr21:19403275:A:G | a | g | 0.6652 | 0.1632 | 4.56E-05 | ++++-++++ | 5.75 | 8 | 0.6752 |
| Any CBVD | chr21 | 20,812,426 | chr21:20812426:T:C | t | c | -1.627 | 0.4131 | 8.21E-05 | -???+-??? | 0.679 | 2 | 0.7121 |
| Any CBVD | chr21 | 21,463,392 | chr21:21463392:C:A | a | c | 0.7168 | 0.1829 | 8.93E-05 | +-+++-+++ | 16.661 | 8 | 0.03384 |
| Any CBVD | chr21 | 21,466,563 | chr21:21466563:G:A | a | g | 0.7176 | 0.183 | 8.80E-05 | +-+++-+++ | 16.665 | 8 | 0.03379 |
| Any CBVD | chr21 | 21,476,705 | chr21:21476705:G:A | a | g | 0.7465 | 0.1846 | 5.27E-05 | +-+++-+++ | 17.771 | 8 | 0.02301 |
| Any CBVD | chr21 | 21,488,212 | chr21:21488212:CAA:C | caa | c | -0.7089 | 0.1819 | 9.74E-05 | -+---+--- | 17.022 | 8 | 0.02988 |
| Any CBVD | chr21 | 21,489,457 | chr21:21489457:C:T | t | c | -0.7514 | 0.1754 | 1.83E-05 | -+------- | 18.723 | 8 | 0.01641 |
| Any CBVD | chr21 | 30,825,454 | chr21:30825454:T:C | t | c | -0.2408 | 0.0592 | 4.74E-05 | ----+---- | 6.523 | 8 | 0.5889 |
| Any CBVD | chr21 | 39,399,854 | chr21:39399854:T:C | t | c | -1.2657 | 0.3125 | 5.13E-05 | ?--+?--?- | 3.555 | 5 | 0.615 |
| Any CBVD | chr21 | 39,435,724 | chr21:39435724:C:T | t | c | 1.2417 | 0.3117 | 6.80E-05 | ?++-?++?+ | 3.433 | 5 | 0.6336 |
| Any CBVD | chr21 | 39,450,009 | chr21:39450009:T:C | t | c | -1.2782 | 0.3153 | 5.04E-05 | ?--+?--?- | 3.348 | 5 | 0.6465 |

| | | | | | | | | | | | | |
|---|---|---|---|---|---|---|---|---|---|---|---|---|
| Any CBVD | chr21 | 39,453,954 | chr21:39453954:ATTAG:A | a | attag | 1.2703 | 0.3161 | 5.86E-05 | ?++-?++?+ | 3.404 | 5 | 0.638 |
| Any CBVD | chr21 | 42,288,927 | chr21:42288927:A:T | a | t | -1.0654 | 0.2725 | 9.22E-05 | -+--?-?+- | 7.017 | 6 | 0.3193 |
| Any CBVD | chr22 | 17,170,692 | chr22:17170692:G:A | a | g | 1.0912 | 0.2733 | 6.55E-05 | +?++-++?+ | 8.067 | 6 | 0.2332 |
| Any CBVD | chr22 | 32,565,198 | chr22:32565198:C:G | c | g | -0.4976 | 0.1266 | 8.44E-05 | -+--+---- | 10.791 | 8 | 0.2138 |
| Any CBVD | chr22 | 36,398,522 | chr22:36398522:A:G | a | g | 0.3055 | 0.0771 | 7.40E-05 | ++++-++++ | 10.262 | 8 | 0.2471 |
| Any CBVD | chr22 | 36,426,502 | chr22:36426502:G:T | t | g | 0.5898 | 0.1478 | 6.62E-05 | ++++-+-++ | 8.084 | 8 | 0.4253 |
| Any CBVD | chr22 | 36,428,198 | chr22:36428198:G:A | a | g | 0.6024 | 0.1485 | 4.98E-05 | ++++-+-++ | 8.957 | 8 | 0.3459 |
| Any CBVD | chr22 | 36,428,312 | chr22:36428312:A:G | a | g | -0.5924 | 0.1477 | 6.07E-05 | ----+-+-- | 7.725 | 8 | 0.4608 |
| Any CBVD | chr22 | 36,439,383 | chr22:36439383:A:T | a | t | -0.5604 | 0.144 | 9.90E-05 | ----+-+-- | 10.54 | 8 | 0.2291 |
| Any CBVD | chr22 | 36,439,401 | chr22:36439401:G:A | a | g | 0.565 | 0.1443 | 9.02E-05 | ++++-+-++ | 10.772 | 8 | 0.215 |
| Any CBVD | chr22 | 36,442,432 | chr22:36442432:A:G | a | g | -0.5664 | 0.1431 | 7.57E-05 | ----+-+-- | 9.845 | 8 | 0.2761 |
| Any CBVD | chr22 | 36,443,585 | chr22:36443585:C:A | a | c | 0.5615 | 0.1442 | 9.90E-05 | ++++-+-++ | 10.379 | 8 | 0.2394 |
| Any CBVD | chr22 | 36,443,587 | chr22:36443587:A:C | a | c | -0.5612 | 0.144 | 9.72E-05 | ----+-+-- | 10.49 | 8 | 0.2323 |
| Any CBVD | chr22 | 36,443,641 | chr22:36443641:T:C | t | c | -0.5633 | 0.1447 | 9.87E-05 | ----+-+-- | 10.598 | 8 | 0.2255 |
| Any CBVD | chr22 | 36,443,694 | chr22:36443694:C:T | t | c | 0.5657 | 0.1442 | 8.76E-05 | ++++-+-++ | 10.494 | 8 | 0.2321 |
| Any CBVD | chr22 | 36,443,930 | chr22:36443930:G:A | a | g | 0.5599 | 0.1439 | 9.96E-05 | ++++-+-++ | 10.528 | 8 | 0.2299 |
| Any CBVD | chr22 | 36,445,431 | chr22:36445431:A:G | a | g | -0.5599 | 0.1439 | 9.97E-05 | ----+-+-- | 10.531 | 8 | 0.2297 |
| Any CBVD | chr22 | 38,913,223 | chr22:38913223:A:G | a | g | 0.4027 | 0.0963 | 2.88E-05 | +++-++--+ | 19.014 | 8 | 0.01479 |
| Any CBVD | chr22 | 38,918,321 | chr22:38918321:C:T | t | c | -0.3621 | 0.0926 | 9.26E-05 | ---+-++-+ | 19.34 | 8 | 0.01314 |
| Any CBVD | chr3 | 11,352,303 | chr3:11352303:A:C | a | c | -1.4147 | 0.3522 | 5.91E-05 | ---?+?-?? | 2.79 | 4 | 0.5936 |
| Any CBVD | chr3 | 21,003,444 | chr3:21003444:T:TTA | t | tta | -0.9725 | 0.2233 | 1.33E-05 | --------- | 14.003 | 8 | 0.0817 |
| Any CBVD | chr3 | 21,067,963 | chr3:21067963:G:A | a | g | 1.0335 | 0.2304 | 7.25E-06 | +++++-+++ | 16.184 | 8 | 0.03983 |
| Any CBVD | chr3 | 21,153,057 | chr3:21153057:T:A | a | t | 0.6989 | 0.1786 | 9.09E-05 | +++++++++ | 11.526 | 8 | 0.1736 |
| Any CBVD | chr3 | 21,234,820 | chr3:21234820:T:C | t | c | -0.7684 | 0.1945 | 7.80E-05 | --------- | 10.07 | 8 | 0.2601 |
| Any CBVD | chr3 | 53,526,177 | chr3:53526177:G:A | a | g | 1.9496 | 0.4304 | 5.90E-06 | +???-+??+ | 3.307 | 3 | 0.3467 |
| Any CBVD | chr3 | 53,526,434 | chr3:53526434:G:A | a | g | 1.9488 | 0.4298 | 5.78E-06 | +???-+??+ | 3.289 | 3 | 0.3492 |
| Any CBVD | chr3 | 53,526,916 | chr3:53526916:A:G | a | g | -1.7288 | 0.3926 | 1.07E-05 | -???+-??- | 2.958 | 3 | 0.3981 |
| Any CBVD | chr3 | 53,531,601 | chr3:53531601:C:G | c | g | -1.8315 | 0.4032 | 5.58E-06 | -???--??- | 1.321 | 3 | 0.7243 |
| Any CBVD | chr3 | 53,533,698 | chr3:53533698:T:G | t | g | -1.9581 | 0.4246 | 4.00E-06 | -???+-??- | 3.508 | 3 | 0.3198 |
| Any CBVD | chr3 | 53,535,281 | chr3:53535281:C:T | t | c | 1.992 | 0.4275 | 3.17E-06 | +???-+??+ | 3.611 | 3 | 0.3066 |
| Any CBVD | chr3 | 53,536,469 | chr3:53536469:A:C | a | c | -1.8804 | 0.4193 | 7.32E-06 | -???+-??- | 3.514 | 3 | 0.3189 |
| Any CBVD | chr3 | 53,538,614 | chr3:53538614:A:T | a | t | -1.8578 | 0.4181 | 8.86E-06 | -???+-??- | 3.873 | 3 | 0.2755 |
| Any CBVD | chr3 | 53,540,214 | chr3:53540214:G:T | t | g | 1.8191 | 0.4178 | 1.33E-05 | +???-+??+ | 3.646 | 3 | 0.3024 |
| Any CBVD | chr3 | 53,557,024 | chr3:53557024:G:A | a | g | 2.2622 | 0.4843 | 3.00E-06 | +????-??+ | 2.052 | 2 | 0.3584 |
| Any CBVD | chr3 | 53,559,191 | chr3:53559191:C:T | t | c | 2.3373 | 0.471 | 6.98E-07 | +???+-??+ | 2.454 | 3 | 0.4837 |
| Any CBVD | chr3 | 55,212,238 | chr3:55212238:A:G | a | g | -0.2928 | 0.0681 | 1.70E-05 | ------+-- | 6.128 | 8 | 0.6329 |
| Any CBVD | chr3 | 55,212,492 | chr3:55212492:G:A | a | g | 0.3087 | 0.0685 | 6.51E-06 | ++++++-++ | 6.232 | 8 | 0.6212 |
| Any CBVD | chr3 | 55,213,195 | chr3:55213195:G:A | a | g | 0.2977 | 0.0682 | 1.25E-05 | ++++++-++ | 6.129 | 8 | 0.6327 |
| Any CBVD | chr3 | 69,736,039 | chr3:69736039:C:T | t | c | 1.6771 | 0.4298 | 9.53E-05 | ?++???+?+ | 0.396 | 3 | 0.9411 |
| Any CBVD | chr3 | 103,967,768 | chr3:103967768:C:G | c | g | -1.1289 | 0.2649 | 2.03E-05 | -+-----+? | 7.294 | 7 | 0.399 |
| Any CBVD | chr3 | 159,693,813 | chr3:159693813:A:T | a | t | -0.3114 | 0.0797 | 9.43E-05 | ----++++- | 9.324 | 8 | 0.3157 |
| Any CBVD | chr3 | 176,545,866 | chr3:176545866:G:A | a | g | 1.9807 | 0.4019 | 8.27E-07 | +??+??-+? | 8.346 | 3 | 0.03937 |
| Any CBVD | chr3 | 176,551,963 | chr3:176551963:C:G | c | g | -1.9847 | 0.3906 | 3.77E-07 | -+?-??+-? | 11.092 | 4 | 0.02554 |
| Any CBVD | chr3 | 181,133,416 | chr3:181133416:T:C | t | c | -0.6404 | 0.1612 | 7.11E-05 | -+-++---- | 17.501 | 8 | 0.02529 |
| Any CBVD | chr3 | 181,198,671 | chr3:181198671:C:A | a | c | 0.6473 | 0.1626 | 6.83E-05 | +-+--++++ | 17.32 | 8 | 0.02694 |
| Any CBVD | chr3 | 181,268,811 | chr3:181268811:G:C | c | g | 0.6401 | 0.1631 | 8.69E-05 | +-+--++++ | 17.45 | 8 | 0.02575 |

| | | | | | | | | | | | | |
|---|---|---|---|---|---|---|---|---|---|---|---|---|
| Any CBVD | chr3 | 187,134,820 | chr3:187134820:C:T | t | c | 1.1342 | 0.2441 | 3.39E-06 | +-+-?+-++ | 10.838 | 7 | 0.1458 |
| Any CBVD | chr3 | 190,216,195 | chr3:190216195:A:G | a | g | 0.5881 | 0.146 | 5.64E-05 | ++++-++++ | 3.344 | 8 | 0.911 |
| Any CBVD | chr3 | 190,261,421 | chr3:190261421:G:T | t | g | -0.5972 | 0.1468 | 4.72E-05 | ----+---- | 3.24 | 8 | 0.9184 |
| Any CBVD | chr3 | 190,287,234 | chr3:190287234:C:T | t | c | -0.6104 | 0.1476 | 3.56E-05 | ----+---- | 3.098 | 8 | 0.9281 |
| Any CBVD | chr3 | 197,455,709 | chr3:197455709:G:GAC | g | gac | -0.3675 | 0.0929 | 7.66E-05 | ----+---- | 10.327 | 8 | 0.2428 |
| Any CBVD | chr4 | 7,310,903 | chr4:7310903:C:T | t | c | 1.4827 | 0.3486 | 2.11E-05 | +?+???++? | 6.15 | 3 | 0.1045 |
| Any CBVD | chr4 | 18,795,032 | chr4:18795032:G:A | a | g | 1.3219 | 0.3247 | 4.67E-05 | +?+++-++? | 4.633 | 6 | 0.5917 |
| Any CBVD | chr4 | 20,280,007 | chr4:20280007:T:C | t | c | -0.752 | 0.1885 | 6.62E-05 | -+--+---- | 6.384 | 8 | 0.6043 |
| Any CBVD | chr4 | 20,281,126 | chr4:20281126:C:T | t | c | 0.7822 | 0.1894 | 3.63E-05 | +-++-++++ | 6.624 | 8 | 0.5777 |
| Any CBVD | chr4 | 20,939,131 | chr4:20939131:C:T | t | c | 0.9276 | 0.2362 | 8.62E-05 | +?+-+++-+ | 13.109 | 7 | 0.0695 |
| Any CBVD | chr4 | 32,428,850 | chr4:32428850:C:A | a | c | 0.6322 | 0.1606 | 8.29E-05 | ++-+-++-+ | 16.167 | 8 | 0.04005 |
| Any CBVD | chr4 | 32,485,316 | chr4:32485316:T:C | t | c | -0.74 | 0.1658 | 8.03E-06 | -+--+--+- | 13.838 | 8 | 0.08608 |
| Any CBVD | chr4 | 32,494,556 | chr4:32494556:G:A | a | g | 0.6383 | 0.1622 | 8.34E-05 | +-++-++-+ | 11.884 | 8 | 0.1565 |
| Any CBVD | chr4 | 34,388,852 | chr4:34388852:A:G | a | g | -0.3856 | 0.0983 | 8.74E-05 | -----+-+- | 6.905 | 8 | 0.5469 |
| Any CBVD | chr4 | 34,389,473 | chr4:34389473:G:A | a | g | 0.3857 | 0.0983 | 8.70E-05 | +++++-+-+ | 6.9 | 8 | 0.5474 |
| Any CBVD | chr4 | 34,390,299 | chr4:34390299:C:A | a | c | 0.3883 | 0.0983 | 7.79E-05 | +++++-+-+ | 7.053 | 8 | 0.5309 |
| Any CBVD | chr4 | 34,390,458 | chr4:34390458:G:C | c | g | 0.3893 | 0.0984 | 7.62E-05 | +++++-+-+ | 6.968 | 8 | 0.54 |
| Any CBVD | chr4 | 34,390,529 | chr4:34390529:C:A | a | c | 0.386 | 0.0983 | 8.61E-05 | +++++-+-+ | 6.912 | 8 | 0.5462 |
| Any CBVD | chr4 | 34,391,814 | chr4:34391814:C:T | t | c | 0.3862 | 0.0983 | 8.52E-05 | +++++-+-+ | 6.862 | 8 | 0.5516 |
| Any CBVD | chr4 | 34,391,844 | chr4:34391844:A:C | a | c | -0.3857 | 0.0983 | 8.70E-05 | -----+-+- | 6.899 | 8 | 0.5475 |
| Any CBVD | chr4 | 34,392,215 | chr4:34392215:A:C | a | c | -0.3872 | 0.0983 | 8.23E-05 | -----+-+- | 6.836 | 8 | 0.5545 |
| Any CBVD | chr4 | 34,392,357 | chr4:34392357:T:G | t | g | -0.3859 | 0.0983 | 8.67E-05 | -----+-+- | 6.846 | 8 | 0.5534 |
| Any CBVD | chr4 | 34,393,137 | chr4:34393137:C:A | a | c | 0.3847 | 0.0983 | 9.13E-05 | +++++-+-+ | 6.881 | 8 | 0.5495 |
| Any CBVD | chr4 | 34,393,629 | chr4:34393629:C:G | c | g | -0.385 | 0.0983 | 8.98E-05 | -----+-+- | 6.899 | 8 | 0.5476 |
| Any CBVD | chr4 | 34,396,451 | chr4:34396451:C:A | a | c | 0.4164 | 0.1009 | 3.66E-05 | +++++++-+ | 6.341 | 8 | 0.6091 |
| Any CBVD | chr4 | 34,404,913 | chr4:34404913:C:T | t | c | 0.3887 | 0.0984 | 7.74E-05 | +++++-+-+ | 6.62 | 8 | 0.5782 |
| Any CBVD | chr4 | 34,406,738 | chr4:34406738:T:C | t | c | -0.3896 | 0.0984 | 7.49E-05 | -----+-+- | 6.706 | 8 | 0.5687 |
| Any CBVD | chr4 | 34,406,806 | chr4:34406806:G:A | a | g | 0.391 | 0.0985 | 7.19E-05 | +++++-+-+ | 6.687 | 8 | 0.5707 |
| Any CBVD | chr4 | 34,407,150 | chr4:34407150:T:C | t | c | -0.3867 | 0.0984 | 8.45E-05 | -----+-+- | 6.456 | 8 | 0.5962 |
| Any CBVD | chr4 | 34,408,066 | chr4:34408066:C:T | t | c | 0.392 | 0.0985 | 6.88E-05 | +++++-+-+ | 6.715 | 8 | 0.5677 |
| Any CBVD | chr4 | 34,413,971 | chr4:34413971:A:G | a | g | -0.3869 | 0.0983 | 8.32E-05 | -----+-+- | 6.852 | 8 | 0.5527 |
| Any CBVD | chr4 | 34,415,418 | chr4:34415418:A:T | a | t | -0.3886 | 0.0984 | 7.79E-05 | -----+-+- | 6.558 | 8 | 0.5849 |
| Any CBVD | chr4 | 34,415,451 | chr4:34415451:G:T | t | g | 0.3918 | 0.0985 | 6.94E-05 | +++++-+-+ | 6.664 | 8 | 0.5733 |
| Any CBVD | chr4 | 34,416,084 | chr4:34416084:T:G | t | g | -0.3898 | 0.0985 | 7.52E-05 | -----+-+- | 6.555 | 8 | 0.5853 |
| Any CBVD | chr4 | 34,418,208 | chr4:34418208:CAT:C | cat | c | -0.3952 | 0.0985 | 5.98E-05 | -----+-+- | 6.867 | 8 | 0.5511 |
| Any CBVD | chr4 | 34,418,327 | chr4:34418327:G:A | a | g | 0.3916 | 0.0985 | 7.02E-05 | +++++-+-+ | 6.605 | 8 | 0.5798 |
| Any CBVD | chr4 | 34,418,809 | chr4:34418809:A:T | a | t | -0.3876 | 0.0984 | 8.16E-05 | -----+-+- | 6.581 | 8 | 0.5824 |
| Any CBVD | chr4 | 38,151,666 | chr4:38151666:CG:C | cg | c | -0.9642 | 0.2266 | 2.09E-05 | --------? | 4.769 | 7 | 0.6881 |
| Any CBVD | chr4 | 38,157,904 | chr4:38157904:G:A | a | g | 0.3099 | 0.0738 | 2.67E-05 | +++++++++ | 5.914 | 8 | 0.6569 |
| Any CBVD | chr4 | 54,428,699 | chr4:54428699:G:T | t | g | 0.7578 | 0.1895 | 6.37E-05 | ++++++--+ | 12.41 | 8 | 0.1338 |
| Any CBVD | chr4 | 54,474,111 | chr4:54474111:C:T | t | c | 0.775 | 0.1897 | 4.40E-05 | ++++++--+ | 13.087 | 8 | 0.1089 |
| Any CBVD | chr4 | 56,581,548 | chr4:56581548:AG:A | a | ag | 1.2163 | 0.3117 | 9.55E-05 | -+++?+-+? | 29.31 | 6 | 5.31E-05 |
| Any CBVD | chr4 | 61,448,510 | chr4:61448510:ATACTT:A | a | atactt | 1.0864 | 0.2774 | 8.99E-05 | ++++????+ | 0.692 | 4 | 0.9523 |
| Any CBVD | chr4 | 64,980,390 | chr4:64980390:A:G | a | g | -0.7989 | 0.1964 | 4.77E-05 | ------+-- | 11.615 | 8 | 0.1692 |
| Any CBVD | chr4 | 96,837,140 | chr4:96837140:T:G | t | g | -0.6518 | 0.1493 | 1.27E-05 | ------+-- | 13.795 | 8 | 0.08727 |
| Any CBVD | chr4 | 96,837,656 | chr4:96837656:T:C | t | c | -1.7797 | 0.4569 | 9.83E-05 | ??--???+- | 5.595 | 3 | 0.1331 |

| | | | | | | | | | | | | |
|---|---|---|---|---|---|---|---|---|---|---|---|---|
| Any CBVD | chr4 | 135,910,869 | chr4:135910869:C:T | t | c | 0.989 | 0.2538 | 9.77E-05 | ++++-+-++ | 13.599 | 8 | 0.09285 |
| Any CBVD | chr4 | 148,828,957 | chr4:148828957:C:CA | ca | c | 1.1799 | 0.2659 | 9.13E-06 | +-++-++?? | 19.02 | 6 | 0.00413 |
| Any CBVD | chr4 | 148,835,486 | chr4:148835486:G:A | a | g | 1.1799 | 0.2659 | 9.12E-06 | +-++-++?? | 19.034 | 6 | 0.004107 |
| Any CBVD | chr4 | 157,056,241 | chr4:157056241:T:C | t | c | 0.3881 | 0.0954 | 4.75E-05 | ++++-++++ | 6.986 | 8 | 0.5381 |
| Any CBVD | chr4 | 157,063,463 | chr4:157063463:C:T | t | c | -0.383 | 0.0943 | 4.90E-05 | ----+---- | 6.19 | 8 | 0.626 |
| Any CBVD | chr4 | 157,066,577 | chr4:157066577:T:G | t | g | 0.3746 | 0.0943 | 7.07E-05 | ++++-++++ | 6.349 | 8 | 0.6082 |
| Any CBVD | chr4 | 161,461,831 | chr4:161461831:T:A | a | t | 0.7348 | 0.1882 | 9.41E-05 | +-++-++-+ | 14.46 | 8 | 0.07054 |
| Any CBVD | chr4 | 179,155,996 | chr4:179155996:C:G | c | g | 0.7011 | 0.1798 | 9.60E-05 | +-++++++- | 10.041 | 8 | 0.2622 |
| Any CBVD | chr4 | 180,618,582 | chr4:180618582:G:A | a | g | 1.3669 | 0.2695 | 3.93E-07 | +++-+++?+ | 15.89 | 7 | 0.02614 |
| Any CBVD | chr4 | 180,623,257 | chr4:180623257:G:T | t | g | 1.292 | 0.2765 | 2.97E-06 | +++-?++?+ | 15.282 | 6 | 0.01818 |
| Any CBVD | chr4 | 180,632,467 | chr4:180632467:G:A | a | g | 1.2647 | 0.2402 | 1.40E-07 | +++++++++ | 12.893 | 8 | 0.1156 |
| Any CBVD | chr4 | 180,633,067 | chr4:180633067:C:T | t | c | 1.3346 | 0.2355 | 1.45E-08 | +++++++++ | 12.011 | 8 | 0.1507 |
| Any CBVD | chr4 | 180,874,641 | chr4:180874641:ATTGT:A | a | attgt | 1.0695 | 0.2709 | 7.89E-05 | ++-+-++++ | 13.824 | 8 | 0.08648 |
| Any CBVD | chr5 | 4,625,086 | chr5:4625086:T:C | t | c | -0.9207 | 0.212 | 1.41E-05 | -------+- | 8.763 | 8 | 0.3627 |
| Any CBVD | chr5 | 24,825,239 | chr5:24825239:C:A | a | c | 3.8293 | 0.909 | 2.52E-05 | +???++??? | 0.785 | 2 | 0.6754 |
| Any CBVD | chr5 | 24,829,783 | chr5:24829783:G:A | a | g | 3.8484 | 0.912 | 2.45E-05 | +???++??? | 0.84 | 2 | 0.6571 |
| Any CBVD | chr5 | 24,833,091 | chr5:24833091:T:C | t | c | -3.8617 | 0.913 | 2.34E-05 | -???--??? | 0.87 | 2 | 0.6472 |
| Any CBVD | chr5 | 24,833,107 | chr5:24833107:T:A | a | t | 3.8617 | 0.913 | 2.34E-05 | +???++??? | 0.87 | 2 | 0.6472 |
| Any CBVD | chr5 | 24,833,108 | chr5:24833108:A:C | a | c | -3.8617 | 0.913 | 2.34E-05 | -???--??? | 0.87 | 2 | 0.6472 |
| Any CBVD | chr5 | 39,325,581 | chr5:39325581:T:G | t | g | -0.7269 | 0.1833 | 7.31E-05 | ------+-- | 8.026 | 8 | 0.4309 |
| Any CBVD | chr5 | 51,095,134 | chr5:51095134:A:AT | a | at | -0.9884 | 0.247 | 6.28E-05 | ----+---- | 15.541 | 8 | 0.04944 |
| Any CBVD | chr5 | 51,098,259 | chr5:51098259:A:T | a | t | -0.994 | 0.2461 | 5.39E-05 | ----+---- | 15.37 | 8 | 0.05234 |
| Any CBVD | chr5 | 62,713,299 | chr5:62713299:A:C | a | c | -1.0275 | 0.2584 | 6.98E-05 | --------? | 2.735 | 7 | 0.9084 |
| Any CBVD | chr5 | 62,755,742 | chr5:62755742:A:C | a | c | -1.0916 | 0.2606 | 2.80E-05 | --------? | 5.457 | 7 | 0.6044 |
| Any CBVD | chr5 | 62,774,091 | chr5:62774091:C:T | t | c | 1.0195 | 0.2583 | 7.89E-05 | ++++++++? | 4.593 | 7 | 0.7095 |
| Any CBVD | chr5 | 62,829,451 | chr5:62829451:G:A | a | g | 1.0189 | 0.2574 | 7.56E-05 | ++++++++? | 4.84 | 7 | 0.6794 |
| Any CBVD | chr5 | 62,837,939 | chr5:62837939:T:C | t | c | -1.0255 | 0.258 | 7.03E-05 | --------? | 4.834 | 7 | 0.6802 |
| Any CBVD | chr5 | 65,828,750 | chr5:65828750:T:G | t | g | 0.3029 | 0.0767 | 7.79E-05 | +++++++-+ | 5.197 | 8 | 0.7364 |
| Any CBVD | chr5 | 65,829,398 | chr5:65829398:A:C | a | c | 0.3108 | 0.079 | 8.44E-05 | +++++++++ | 4 | 8 | 0.8571 |
| Any CBVD | chr5 | 66,935,194 | chr5:66935194:C:T | t | c | 0.4618 | 0.1114 | 3.41E-05 | +++++--++ | 14.505 | 8 | 0.06952 |
| Any CBVD | chr5 | 66,979,972 | chr5:66979972:C:T | t | c | 0.4986 | 0.1207 | 3.62E-05 | +++++--++ | 9.153 | 8 | 0.3295 |
| Any CBVD | chr5 | 71,821,454 | chr5:71821454:T:TTTG | t | tttg | -0.5708 | 0.1325 | 1.65E-05 | ---++---- | 7.662 | 8 | 0.4672 |
| Any CBVD | chr5 | 71,832,272 | chr5:71832272:A:G | a | g | -0.5096 | 0.1308 | 9.71E-05 | ---++---- | 9.552 | 8 | 0.2979 |
| Any CBVD | chr5 | 71,832,284 | chr5:71832284:A:G | a | g | -0.525 | 0.1314 | 6.46E-05 | ---++---- | 7.351 | 8 | 0.4993 |
| Any CBVD | chr5 | 74,536,188 | chr5:74536188:G:A | a | g | 1.5326 | 0.3868 | 7.44E-05 | ++????+-? | 4.887 | 3 | 0.1803 |
| Any CBVD | chr5 | 81,753,721 | chr5:81753721:T:G | t | g | 0.4186 | 0.1043 | 6.03E-05 | +++++-+++ | 5.972 | 8 | 0.6504 |
| Any CBVD | chr5 | 83,346,579 | chr5:83346579:GGTAAA:G | g | ggtaaa | 1.0557 | 0.2496 | 2.35E-05 | ++++-+--+ | 10.666 | 8 | 0.2214 |
| Any CBVD | chr5 | 83,347,293 | chr5:83347293:C:T | t | c | 1.0925 | 0.2626 | 3.18E-05 | ++++-+--+ | 10.805 | 8 | 0.213 |
| Any CBVD | chr5 | 83,352,047 | chr5:83352047:C:A | a | c | 1.033 | 0.2483 | 3.18E-05 | ++++-+--+ | 10.146 | 8 | 0.2549 |
| Any CBVD | chr5 | 83,352,114 | chr5:83352114:G:T | t | g | 1.0409 | 0.2487 | 2.85E-05 | ++++-+--+ | 9.963 | 8 | 0.2676 |
| Any CBVD | chr5 | 83,355,622 | chr5:83355622:T:A | a | t | 1.0225 | 0.2512 | 4.70E-05 | ++++-+--+ | 10.246 | 8 | 0.2482 |
| Any CBVD | chr5 | 83,371,819 | chr5:83371819:G:A | a | g | 1.0602 | 0.2367 | 7.52E-06 | ++++-+-++ | 11.731 | 8 | 0.1636 |
| Any CBVD | chr5 | 88,747,493 | chr5:88747493:C:T | t | c | 1.3282 | 0.2749 | 1.35E-06 | +++++++-+ | 9.906 | 8 | 0.2717 |
| Any CBVD | chr5 | 96,489,113 | chr5:96489113:C:A | a | c | 1.686 | 0.3967 | 2.13E-05 | ?+++??+-+ | 7.898 | 5 | 0.1619 |
| Any CBVD | chr5 | 113,801,485 | chr5:113801485:T:A | a | t | 0.6044 | 0.1545 | 9.14E-05 | +++++-+-+ | 9.472 | 8 | 0.3041 |
| Any CBVD | chr5 | 113,849,228 | chr5:113849228:C:T | t | c | 0.7674 | 0.1576 | 1.12E-06 | +++++++-+ | 11.555 | 8 | 0.1722 |

| | | | | | | | | | | | | |
|---|---|---|---|---|---|---|---|---|---|---|---|---|
| Any CBVD | chr5 | 113,855,704 | chr5:113855704:A:G | a | g | -0.7069 | 0.1538 | 4.32E-06 | -----+-+- | 12.604 | 8 | 0.1262 |
| Any CBVD | chr5 | 113,897,765 | chr5:113897765:T:C | t | c | -0.6275 | 0.1528 | 4.01E-05 | -----+-+- | 10.297 | 8 | 0.2448 |
| Any CBVD | chr5 | 127,621,439 | chr5:127621439:C:T | t | c | 2.3446 | 0.5172 | 5.81E-06 | ??++???-? | 4.032 | 2 | 0.1332 |
| Any CBVD | chr5 | 127,625,977 | chr5:127625977:T:C | t | c | -2.3473 | 0.5176 | 5.77E-06 | ??--???+? | 4.057 | 2 | 0.1315 |
| Any CBVD | chr5 | 132,368,496 | chr5:132368496:G:A | a | g | 1.0465 | 0.2598 | 5.61E-05 | +-++-++-+ | 13.527 | 8 | 0.09494 |
| Any CBVD | chr5 | 133,057,205 | chr5:133057205:G:A | a | g | 1.7709 | 0.4212 | 2.61E-05 | ++?????+? | 6.354 | 2 | 0.04171 |
| Any CBVD | chr5 | 151,989,299 | chr5:151989299:G:A | a | g | 1.1749 | 0.2675 | 1.12E-05 | +-++-?-?+ | 10.397 | 6 | 0.1089 |
| Any CBVD | chr5 | 164,111,903 | chr5:164111903:CAGGA:C | cagga | c | -1.2763 | 0.3252 | 8.70E-05 | ----?-??? | 9.325 | 4 | 0.05347 |
| Any CBVD | chr5 | 166,923,781 | chr5:166923781:G:C | c | g | -0.5622 | 0.1437 | 9.17E-05 | -+-----+- | 7.421 | 8 | 0.492 |
| Any CBVD | chr5 | 170,431,029 | chr5:170431029:G:A | a | g | 0.3798 | 0.0956 | 7.13E-05 | +++--++++ | 8.514 | 8 | 0.385 |
| Any CBVD | chr5 | 170,431,719 | chr5:170431719:C:T | t | c | 0.3814 | 0.0956 | 6.66E-05 | +++--++++ | 8.145 | 8 | 0.4194 |
| Any CBVD | chr5 | 170,434,541 | chr5:170434541:G:A | a | g | 0.3793 | 0.0956 | 7.23E-05 | +++--++++ | 8.246 | 8 | 0.4098 |
| Any CBVD | chr5 | 170,435,589 | chr5:170435589:A:C | a | c | -0.3737 | 0.0956 | 9.24E-05 | ---++---- | 9.285 | 8 | 0.3188 |
| Any CBVD | chr5 | 170,439,390 | chr5:170439390:G:A | a | g | 0.3918 | 0.0957 | 4.24E-05 | +++--++++ | 7.487 | 8 | 0.4851 |
| Any CBVD | chr5 | 170,441,456 | chr5:170441456:C:G | c | g | -0.3993 | 0.0958 | 3.04E-05 | ---++---- | 7.137 | 8 | 0.5219 |
| Any CBVD | chr6 | 934,893 | chr6:934893:C:T | t | c | 0.2676 | 0.0687 | 9.88E-05 | +++++++++ | 7.766 | 8 | 0.4567 |
| Any CBVD | chr6 | 1,276,640 | chr6:1276640:A:G | a | g | -0.3889 | 0.0963 | 5.40E-05 | --------- | 1.947 | 8 | 0.9826 |
| Any CBVD | chr6 | 7,542,890 | chr6:7542890:A:C | a | c | -0.2824 | 0.0723 | 9.44E-05 | ----+---- | 2.957 | 8 | 0.937 |
| Any CBVD | chr6 | 9,488,976 | chr6:9488976:G:A | a | g | -0.2278 | 0.0569 | 6.33E-05 | --------- | 4.138 | 8 | 0.8445 |
| Any CBVD | chr6 | 10,540,335 | chr6:10540335:C:T | t | c | 2.2793 | 0.5754 | 7.45E-05 | ????++??+ | 0.129 | 2 | 0.9376 |
| Any CBVD | chr6 | 17,553,376 | chr6:17553376:TA:T | t | ta | -0.587 | 0.1391 | 2.45E-05 | ----+--+- | 5.634 | 8 | 0.6882 |
| Any CBVD | chr6 | 17,572,514 | chr6:17572514:G:A | a | g | 0.5798 | 0.1401 | 3.48E-05 | ++++-++-+ | 6.382 | 8 | 0.6045 |
| Any CBVD | chr6 | 17,572,547 | chr6:17572547:A:C | a | c | -0.5794 | 0.1401 | 3.54E-05 | ----+--+- | 6.379 | 8 | 0.6049 |
| Any CBVD | chr6 | 17,579,997 | chr6:17579997:G:T | t | g | 0.5902 | 0.1404 | 2.62E-05 | ++++-++-+ | 6.374 | 8 | 0.6054 |
| Any CBVD | chr6 | 21,951,833 | chr6:21951833:T:G | t | g | 0.29 | 0.0694 | 2.93E-05 | ++++-+-++ | 6.885 | 8 | 0.5491 |
| Any CBVD | chr6 | 21,952,879 | chr6:21952879:T:G | t | g | 0.2943 | 0.0695 | 2.31E-05 | ++++-+-++ | 7.376 | 8 | 0.4966 |
| Any CBVD | chr6 | 22,761,548 | chr6:22761548:CT:C | ct | c | -0.7263 | 0.1866 | 9.90E-05 | -+---+-+- | 19.796 | 8 | 0.01114 |
| Any CBVD | chr6 | 25,174,159 | chr6:25174159:CA:C | ca | c | 0.242 | 0.0617 | 8.84E-05 | +-++-+--+ | 23.865 | 8 | 0.002414 |
| Any CBVD | chr6 | 25,175,917 | chr6:25175917:T:C | t | c | 0.2367 | 0.0608 | 9.93E-05 | +-++-++-+ | 18.361 | 8 | 0.01868 |
| Any CBVD | chr6 | 27,466,315 | chr6:27466315:G:A | a | g | 0.8421 | 0.2102 | 6.18E-05 | ++++++--+ | 9.953 | 8 | 0.2683 |
| Any CBVD | chr6 | 29,564,041 | chr6:29564041:A:G | a | g | -0.491 | 0.1163 | 2.41E-05 | -+----+-- | 18.451 | 8 | 0.01809 |
| Any CBVD | chr6 | 29,566,814 | chr6:29566814:A:T | a | t | -0.4806 | 0.1158 | 3.31E-05 | -+------- | 17.665 | 8 | 0.02389 |
| Any CBVD | chr6 | 29,602,888 | chr6:29602888:C:T | t | c | 0.4881 | 0.1171 | 3.05E-05 | +-+++++++ | 14.053 | 8 | 0.08038 |
| Any CBVD | chr6 | 29,606,854 | chr6:29606854:C:T | t | c | 0.4634 | 0.112 | 3.49E-05 | +-+++++++ | 11.776 | 8 | 0.1615 |
| Any CBVD | chr6 | 29,633,783 | chr6:29633783:G:A | a | g | 0.5008 | 0.1197 | 2.86E-05 | +-+++++++ | 12.598 | 8 | 0.1265 |
| Any CBVD | chr6 | 29,634,290 | chr6:29634290:C:A | a | c | 0.4938 | 0.1195 | 3.60E-05 | +-+++++++ | 12.776 | 8 | 0.1198 |
| Any CBVD | chr6 | 30,487,330 | chr6:30487330:G:T | t | g | 0.401 | 0.1 | 6.05E-05 | ++++++-++ | 16.483 | 8 | 0.03597 |
| Any CBVD | chr6 | 30,690,608 | chr6:30690608:C:T | t | c | 0.8576 | 0.1849 | 3.50E-06 | ++++-++-+ | 7.978 | 8 | 0.4356 |
| Any CBVD | chr6 | 30,691,351 | chr6:30691351:A:G | a | g | -0.8617 | 0.1854 | 3.35E-06 | ----+--+- | 7.906 | 8 | 0.4427 |
| Any CBVD | chr6 | 30,691,695 | chr6:30691695:T:A | a | t | 0.8619 | 0.1854 | 3.35E-06 | ++++-++-+ | 7.908 | 8 | 0.4425 |
| Any CBVD | chr6 | 30,693,230 | chr6:30693230:T:C | t | c | -0.6855 | 0.1528 | 7.20E-06 | ------++- | 4.905 | 8 | 0.7677 |
| Any CBVD | chr6 | 30,693,286 | chr6:30693286:C:T | t | c | 0.8618 | 0.1854 | 3.36E-06 | ++++-++-+ | 7.905 | 8 | 0.4429 |
| Any CBVD | chr6 | 30,693,489 | chr6:30693489:T:C | t | c | -0.8621 | 0.1854 | 3.34E-06 | ----+--+- | 7.9 | 8 | 0.4433 |
| Any CBVD | chr6 | 30,695,789 | chr6:30695789:T:A | a | t | 0.8424 | 0.185 | 5.26E-06 | ++++-++-+ | 7.362 | 8 | 0.4981 |
| Any CBVD | chr6 | 30,695,943 | chr6:30695943:C:T | t | c | 0.8613 | 0.1853 | 3.34E-06 | ++++-++-+ | 7.909 | 8 | 0.4424 |
| Any CBVD | chr6 | 30,696,188 | chr6:30696188:C:T | t | c | 0.8617 | 0.1853 | 3.32E-06 | ++++-++-+ | 7.901 | 8 | 0.4432 |

| | | | | | | | | | | | | |
|---|---|---|---|---|---|---|---|---|---|---|---|---|
| Any CBVD | chr6 | 30,696,837 | chr6:30696837:C:T | t | c | 0.86 | 0.1851 | 3.38E-06 | ++++-++-+ | 7.882 | 8 | 0.4451 |
| Any CBVD | chr6 | 30,697,539 | chr6:30697539:C:T | t | c | 0.8626 | 0.1854 | 3.26E-06 | ++++-++-+ | 7.916 | 8 | 0.4417 |
| Any CBVD | chr6 | 30,698,250 | chr6:30698250:A:G | a | g | -0.8502 | 0.2177 | 9.39E-05 | -------+- | 5.919 | 8 | 0.6563 |
| Any CBVD | chr6 | 30,698,817 | chr6:30698817:A:G | a | g | -0.6957 | 0.1536 | 5.89E-06 | ------++- | 4.885 | 8 | 0.7697 |
| Any CBVD | chr6 | 30,699,574 | chr6:30699574:C:G | c | g | -0.8617 | 0.1855 | 3.38E-06 | ----+--+- | 7.9 | 8 | 0.4433 |
| Any CBVD | chr6 | 30,701,393 | chr6:30701393:C:G | c | g | -0.8619 | 0.1854 | 3.34E-06 | ----+--+- | 7.905 | 8 | 0.4428 |
| Any CBVD | chr6 | 30,708,269 | chr6:30708269:C:G | c | g | -0.8774 | 0.1874 | 2.84E-06 | ----+--+- | 8.695 | 8 | 0.3687 |
| Any CBVD | chr6 | 36,131,273 | chr6:36131273:C:T | t | c | 0.9493 | 0.2391 | 7.15E-05 | +++--++++ | 17.999 | 8 | 0.02123 |
| Any CBVD | chr6 | 69,378,816 | chr6:69378816:T:C | t | c | -0.7924 | 0.2029 | 9.39E-05 | -+----++- | 13.454 | 8 | 0.09716 |
| Any CBVD | chr6 | 76,773,476 | chr6:76773476:C:T | t | c | 0.8924 | 0.2055 | 1.40E-05 | +++-+++++ | 10.156 | 8 | 0.2542 |
| Any CBVD | chr6 | 77,415,132 | chr6:77415132:T:C | t | c | -0.9258 | 0.2367 | 9.19E-05 | ---+----- | 4.3 | 8 | 0.8291 |
| Any CBVD | chr6 | 82,118,197 | chr6:82118197:A:T | a | t | -0.9985 | 0.2485 | 5.88E-05 | -+--?---- | 4.725 | 7 | 0.6935 |
| Any CBVD | chr6 | 95,379,762 | chr6:95379762:C:T | t | c | 2.9998 | 0.7625 | 8.34E-05 | ?-??++??? | 10.081 | 2 | 0.00647 |
| Any CBVD | chr6 | 95,422,047 | chr6:95422047:G:A | a | g | 3.0074 | 0.7587 | 7.37E-05 | ?-??++??? | 10.114 | 2 | 0.006363 |
| Any CBVD | chr6 | 105,071,932 | chr6:105071932:A:C | a | c | -2.0578 | 0.4799 | 1.80E-05 | ??--??-+? | 6.271 | 3 | 0.09912 |
| Any CBVD | chr6 | 105,992,372 | chr6:105992372:AC:A | a | ac | 0.5564 | 0.1416 | 8.51E-05 | +-++-++++ | 8.293 | 8 | 0.4054 |
| Any CBVD | chr6 | 106,015,534 | chr6:106015534:A:C | a | c | -0.578 | 0.1412 | 4.27E-05 | -+--+-+-- | 8.597 | 8 | 0.3774 |
| Any CBVD | chr6 | 106,728,154 | chr6:106728154:G:C | c | g | 0.2426 | 0.0592 | 4.16E-05 | +++++++++ | 8.247 | 8 | 0.4097 |
| Any CBVD | chr6 | 133,870,824 | chr6:133870824:G:T | t | g | 3.3579 | 0.795 | 2.40E-05 | +???-+??? | 2.518 | 2 | 0.284 |
| Any CBVD | chr6 | 133,893,695 | chr6:133893695:A:G | a | g | -3.4336 | 0.799 | 1.73E-05 | -???+-??? | 2.658 | 2 | 0.2648 |
| Any CBVD | chr6 | 133,937,524 | chr6:133937524:G:A | a | g | 0.9523 | 0.2409 | 7.74E-05 | +++--++-+ | 13.343 | 8 | 0.1006 |
| Any CBVD | chr6 | 133,962,649 | chr6:133962649:A:G | a | g | -0.9727 | 0.24 | 5.07E-05 | ---++--+- | 9.653 | 8 | 0.2902 |
| Any CBVD | chr6 | 147,694,586 | chr6:147694586:GTGGGTGGAT:G | g | gtgggtggat | 0.6387 | 0.1423 | 7.20E-06 | ++++++-++ | 5.35 | 8 | 0.7196 |
| Any CBVD | chr6 | 154,326,624 | chr6:154326624:G:A | a | g | 1.2213 | 0.2964 | 3.78E-05 | +++-?++-? | 5.671 | 6 | 0.4611 |
| Any CBVD | chr6 | 159,931,487 | chr6:159931487:G:A | a | g | 1.0917 | 0.2782 | 8.72E-05 | +++--+?++ | 12.85 | 7 | 0.07584 |
| Any CBVD | chr6 | 166,226,968 | chr6:166226968:G:A | a | g | 0.2419 | 0.0564 | 1.82E-05 | ++++++-++ | 6.642 | 8 | 0.5757 |
| Any CBVD | chr6 | 169,231,899 | chr6:169231899:C:G | c | g | 0.2676 | 0.0664 | 5.57E-05 | +-+++++++ | 6.069 | 8 | 0.6395 |
| Any CBVD | chr7 | 124,277 | chr7:124277:G:T | t | g | 0.477 | 0.1188 | 5.92E-05 | +?+++++++ | 2.312 | 7 | 0.9406 |
| Any CBVD | chr7 | 124,587 | chr7:124587:T:C | t | c | 0.5501 | 0.1273 | 1.56E-05 | +++++++++ | 5.707 | 8 | 0.68 |
| Any CBVD | chr7 | 16,016,910 | chr7:16016910:TTA:T | t | tta | 0.3168 | 0.0774 | 4.23E-05 | ++++-++++ | 9.621 | 8 | 0.2926 |
| Any CBVD | chr7 | 17,420,495 | chr7:17420495:C:G | c | g | -0.3342 | 0.0848 | 8.10E-05 | -+-+----- | 10.754 | 8 | 0.216 |
| Any CBVD | chr7 | 19,617,173 | chr7:19617173:A:G | a | g | -1.6809 | 0.4258 | 7.90E-05 | ?+??---+- | 5.167 | 5 | 0.3958 |
| Any CBVD | chr7 | 25,430,368 | chr7:25430368:G:A | a | g | 0.5509 | 0.1393 | 7.64E-05 | +++++++++ | 5.008 | 8 | 0.7567 |
| Any CBVD | chr7 | 29,068,177 | chr7:29068177:T:G | t | g | -1.1136 | 0.261 | 1.99E-05 | -+----+-? | 11.58 | 7 | 0.1152 |
| Any CBVD | chr7 | 30,493,255 | chr7:30493255:A:T | a | t | -2.4584 | 0.5603 | 1.15E-05 | ?--???-?? | 4.839 | 2 | 0.08899 |
| Any CBVD | chr7 | 30,934,705 | chr7:30934705:T:A | a | t | 0.6824 | 0.1706 | 6.35E-05 | +-++-++++ | 9.094 | 8 | 0.3344 |
| Any CBVD | chr7 | 31,386,531 | chr7:31386531:TACTTC:T | t | tacttc | 0.4402 | 0.1119 | 8.34E-05 | +++++++++ | 4.186 | 8 | 0.8399 |
| Any CBVD | chr7 | 31,387,703 | chr7:31387703:C:T | t | c | 0.4433 | 0.1128 | 8.47E-05 | +++++++++ | 4.318 | 8 | 0.8273 |
| Any CBVD | chr7 | 31,388,890 | chr7:31388890:A:G | a | g | -0.4427 | 0.1128 | 8.65E-05 | --------- | 4.323 | 8 | 0.8269 |
| Any CBVD | chr7 | 44,564,314 | chr7:44564314:C:G | c | g | -1.4133 | 0.3616 | 9.31E-05 | -?--?-??? | 2.046 | 3 | 0.563 |
| Any CBVD | chr7 | 45,457,104 | chr7:45457104:C:T | t | c | 0.8001 | 0.2047 | 9.24E-05 | ++++++-++ | 7.292 | 8 | 0.5055 |
| Any CBVD | chr7 | 53,926,911 | chr7:53926911:G:C | c | g | 0.2324 | 0.0564 | 3.80E-05 | +--++++++ | 12.592 | 8 | 0.1267 |
| Any CBVD | chr7 | 65,182,345 | chr7:65182345:A:G | a | g | 0.5127 | 0.1204 | 2.06E-05 | +++++++++ | 2.101 | 8 | 0.9778 |
| Any CBVD | chr7 | 65,236,568 | chr7:65236568:TGATGCACTTCAT | t | tgatgcactt | -0.5138 | 0.1199 | 1.83E-05 | --------- | 1.95 | 8 | 0.9825 |
| Any CBVD | chr7 | 65,269,752 | chr7:65269752:G:A | a | g | -0.4783 | 0.1208 | 7.55E-05 | --------- | 1.852 | 8 | 0.9852 |
| Any CBVD | chr7 | 65,269,753 | chr7:65269753:C:A | a | c | -0.4783 | 0.1208 | 7.55E-05 | --------- | 1.852 | 8 | 0.9852 |

| | | | | | | | | | | | | |
|---|---|---|---|---|---|---|---|---|---|---|---|---|
| Any CBVD | chr7 | 71,442,478 | chr7:71442478:C:T | t | c | 1.0545 | 0.2512 | 2.69E-05 | +-++-++++ | 8.117 | 8 | 0.4221 |
| Any CBVD | chr7 | 84,931,780 | chr7:84931780:G:GA | g | ga | -1.2405 | 0.3102 | 6.38E-05 | ------?+? | 7.941 | 6 | 0.2425 |
| Any CBVD | chr7 | 84,933,918 | chr7:84933918:AAAGT:A | a | aaagt | 1.2398 | 0.3157 | 8.62E-05 | ++++++?-? | 9.144 | 6 | 0.1656 |
| Any CBVD | chr7 | 84,940,910 | chr7:84940910:C:T | t | c | 1.2271 | 0.3142 | 9.42E-05 | ++++++?-? | 9.43 | 6 | 0.1508 |
| Any CBVD | chr7 | 84,941,293 | chr7:84941293:A:G | a | g | -1.2268 | 0.3143 | 9.48E-05 | ------?+? | 9.423 | 6 | 0.1512 |
| Any CBVD | chr7 | 84,945,289 | chr7:84945289:T:A | a | t | 1.2271 | 0.3142 | 9.42E-05 | ++++++?-? | 9.43 | 6 | 0.1508 |
| Any CBVD | chr7 | 84,949,223 | chr7:84949223:G:A | a | g | 1.2266 | 0.3142 | 9.48E-05 | ++++++?-? | 9.432 | 6 | 0.1507 |
| Any CBVD | chr7 | 84,958,668 | chr7:84958668:C:G | c | g | -1.2266 | 0.3142 | 9.48E-05 | ------?+? | 9.432 | 6 | 0.1507 |
| Any CBVD | chr7 | 90,900,306 | chr7:90900306:T:A | a | t | 1.3277 | 0.34 | 9.44E-05 | ++++????? | 5.343 | 3 | 0.1483 |
| Any CBVD | chr7 | 99,486,115 | chr7:99486115:C:T | t | c | 0.9348 | 0.2365 | 7.70E-05 | +-+++++++ | 6.255 | 8 | 0.6187 |
| Any CBVD | chr7 | 103,604,002 | chr7:103604002:A:AGTGG | a | agtgg | -0.703 | 0.1649 | 2.02E-05 | -+------+ | 18.469 | 8 | 0.01797 |
| Any CBVD | chr7 | 103,613,663 | chr7:103613663:T:C | t | c | -0.6841 | 0.165 | 3.39E-05 | -+------+ | 18.195 | 8 | 0.01981 |
| Any CBVD | chr7 | 103,618,201 | chr7:103618201:A:G | a | g | -0.3274 | 0.0807 | 4.94E-05 | --------- | 8.932 | 8 | 0.3481 |
| Any CBVD | chr7 | 103,623,862 | chr7:103623862:T:C | t | c | -0.3413 | 0.081 | 2.49E-05 | --------- | 6.407 | 8 | 0.6018 |
| Any CBVD | chr7 | 103,625,809 | chr7:103625809:G:A | a | g | 0.339 | 0.0809 | 2.80E-05 | +-+++++++ | 6.594 | 8 | 0.581 |
| Any CBVD | chr7 | 103,634,386 | chr7:103634386:C:G | c | g | -0.685 | 0.1651 | 3.35E-05 | -+------+ | 17.008 | 8 | 0.03003 |
| Any CBVD | chr7 | 103,637,550 | chr7:103637550:T:C | t | c | -0.3127 | 0.0795 | 8.41E-05 | --------- | 5.67 | 8 | 0.6842 |
| Any CBVD | chr7 | 103,731,490 | chr7:103731490:A:AT | a | at | -0.7155 | 0.1733 | 3.66E-05 | -+------+ | 17.726 | 8 | 0.02338 |
| Any CBVD | chr7 | 103,754,988 | chr7:103754988:C:T | t | c | 0.6991 | 0.1662 | 2.60E-05 | +-+++-+++ | 16.279 | 8 | 0.03856 |
| Any CBVD | chr7 | 136,889,422 | chr7:136889422:A:C | a | c | -2.173 | 0.5101 | 2.04E-05 | ?-?-??-?- | 5.764 | 3 | 0.1237 |
| Any CBVD | chr7 | 155,389,971 | chr7:155389971:C:T | t | c | 1.4733 | 0.3545 | 3.24E-05 | +?++??+-? | 5.268 | 4 | 0.2609 |
| Any CBVD | chr8 | 1,507,095 | chr8:1507095:G:C | c | g | 0.7004 | 0.177 | 7.57E-05 | +-+++++++ | 13.963 | 8 | 0.08273 |
| Any CBVD | chr8 | 3,027,943 | chr8:3027943:G:A | a | g | 0.3033 | 0.071 | 1.93E-05 | +-++++-++ | 9.156 | 8 | 0.3293 |
| Any CBVD | chr8 | 3,045,338 | chr8:3045338:G:T | t | g | 1.0043 | 0.2543 | 7.85E-05 | +-++++?++ | 6.32 | 7 | 0.5029 |
| Any CBVD | chr8 | 18,409,062 | chr8:18409062:T:C | t | c | -0.3123 | 0.0795 | 8.50E-05 | --------- | 7.776 | 8 | 0.4557 |
| Any CBVD | chr8 | 18,409,264 | chr8:18409264:G:A | a | g | 0.3118 | 0.0795 | 8.72E-05 | +++++++++ | 7.879 | 8 | 0.4453 |
| Any CBVD | chr8 | 18,409,414 | chr8:18409414:G:A | a | g | 0.3117 | 0.0798 | 9.32E-05 | +++++++++ | 7.67 | 8 | 0.4664 |
| Any CBVD | chr8 | 18,409,828 | chr8:18409828:A:G | a | g | -0.3114 | 0.0794 | 8.85E-05 | --------- | 7.833 | 8 | 0.45 |
| Any CBVD | chr8 | 18,409,919 | chr8:18409919:A:C | a | c | -0.3103 | 0.0795 | 9.44E-05 | --------- | 7.693 | 8 | 0.464 |
| Any CBVD | chr8 | 18,409,951 | chr8:18409951:G:C | c | g | 0.3106 | 0.0794 | 9.13E-05 | +++++++++ | 7.652 | 8 | 0.4682 |
| Any CBVD | chr8 | 18,410,368 | chr8:18410368:G:A | a | g | 0.3112 | 0.0796 | 9.29E-05 | +++++++++ | 8.352 | 8 | 0.3999 |
| Any CBVD | chr8 | 18,411,833 | chr8:18411833:A:G | a | g | -0.3102 | 0.0796 | 9.77E-05 | --------- | 8.422 | 8 | 0.3934 |
| Any CBVD | chr8 | 18,412,071 | chr8:18412071:A:T | a | t | -0.3163 | 0.0796 | 7.12E-05 | --------- | 8.001 | 8 | 0.4333 |
| Any CBVD | chr8 | 18,415,508 | chr8:18415508:A:G | a | g | -0.3095 | 0.0795 | 9.92E-05 | --------- | 7.898 | 8 | 0.4434 |
| Any CBVD | chr8 | 18,587,214 | chr8:18587214:C:G | c | g | -0.996 | 0.2515 | 7.48E-05 | ---+----- | 13.124 | 8 | 0.1077 |
| Any CBVD | chr8 | 18,587,400 | chr8:18587400:T:C | t | c | -1.0521 | 0.2544 | 3.54E-05 | ---+----- | 13.692 | 8 | 0.09015 |
| Any CBVD | chr8 | 18,587,932 | chr8:18587932:G:A | a | g | 1.0554 | 0.2545 | 3.36E-05 | +++-+++++ | 13.692 | 8 | 0.09015 |
| Any CBVD | chr8 | 18,587,990 | chr8:18587990:C:T | t | c | 1.0573 | 0.2544 | 3.23E-05 | +++-+++++ | 13.559 | 8 | 0.09402 |
| Any CBVD | chr8 | 18,589,139 | chr8:18589139:T:C | t | c | -1.0353 | 0.2531 | 4.30E-05 | ---+----- | 13.74 | 8 | 0.08881 |
| Any CBVD | chr8 | 18,589,472 | chr8:18589472:A:G | a | g | -1.0471 | 0.2538 | 3.68E-05 | ---+----- | 13.596 | 8 | 0.09291 |
| Any CBVD | chr8 | 18,590,433 | chr8:18590433:G:A | a | g | 1.0559 | 0.2543 | 3.28E-05 | +++-+++++ | 13.716 | 8 | 0.08948 |
| Any CBVD | chr8 | 18,590,802 | chr8:18590802:G:C | c | g | 0.9994 | 0.251 | 6.85E-05 | +++-+++++ | 13.192 | 8 | 0.1054 |
| Any CBVD | chr8 | 18,590,948 | chr8:18590948:A:T | a | t | -1.0488 | 0.2539 | 3.62E-05 | ---+----- | 13.597 | 8 | 0.09288 |
| Any CBVD | chr8 | 18,591,032 | chr8:18591032:T:G | t | g | -1.0473 | 0.2538 | 3.67E-05 | ---+----- | 13.595 | 8 | 0.09295 |
| Any CBVD | chr8 | 18,591,344 | chr8:18591344:C:G | c | g | -1.0912 | 0.2526 | 1.57E-05 | ---+----- | 12.786 | 8 | 0.1194 |
| Any CBVD | chr8 | 18,592,128 | chr8:18592128:G:C | c | g | 1.0874 | 0.2524 | 1.65E-05 | +++-+++++ | 12.908 | 8 | 0.1151 |

| | | | | | | | | | | | | |
|---|---|---|---|---|---|---|---|---|---|---|---|---|
| Any CBVD | chr8 | 18,592,290 | chr8:18592290:T:C | t | c | -1.0517 | 0.2541 | 3.49E-05 | ---+----- | 13.494 | 8 | 0.09594 |
| Any CBVD | chr8 | 18,600,936 | chr8:18600936:G:C | c | g | 1.002 | 0.2456 | 4.51E-05 | +++-+++++ | 12.028 | 8 | 0.15 |
| Any CBVD | chr8 | 18,604,757 | chr8:18604757:C:T | t | c | 1.1367 | 0.2887 | 8.25E-05 | +++-++++? | 12.745 | 7 | 0.07856 |
| Any CBVD | chr8 | 36,441,298 | chr8:36441298:G:A | a | g | 0.6997 | 0.1636 | 1.89E-05 | ++++++-++ | 11.255 | 8 | 0.1876 |
| Any CBVD | chr8 | 36,474,206 | chr8:36474206:A:G | a | g | -0.6866 | 0.1623 | 2.34E-05 | ------+-- | 12.676 | 8 | 0.1235 |
| Any CBVD | chr8 | 38,616,874 | chr8:38616874:G:T | t | g | 0.4697 | 0.1175 | 6.38E-05 | +++--++++ | 8.494 | 8 | 0.3868 |
| Any CBVD | chr8 | 55,876,849 | chr8:55876849:C:T | t | c | 0.4095 | 0.0995 | 3.84E-05 | +-+++++++ | 6.024 | 8 | 0.6446 |
| Any CBVD | chr8 | 55,877,397 | chr8:55877397:C:T | t | c | 0.4071 | 0.0994 | 4.23E-05 | +-+++++++ | 5.594 | 8 | 0.6926 |
| Any CBVD | chr8 | 55,880,435 | chr8:55880435:C:G | c | g | -0.4432 | 0.1008 | 1.10E-05 | -+------- | 5.701 | 8 | 0.6807 |
| Any CBVD | chr8 | 67,807,116 | chr8:67807116:C:T | t | c | 2.2599 | 0.5471 | 3.62E-05 | ?++???++? | 2.475 | 3 | 0.4798 |
| Any CBVD | chr8 | 74,974,062 | chr8:74974062:C:T | t | c | 0.2497 | 0.0638 | 9.12E-05 | +++++++++ | 4.382 | 8 | 0.8211 |
| Any CBVD | chr8 | 78,977,011 | chr8:78977011:A:C | a | c | -1.1554 | 0.2802 | 3.74E-05 | -?----+?- | 4.063 | 6 | 0.6681 |
| Any CBVD | chr8 | 78,977,712 | chr8:78977712:T:C | t | c | -1.1554 | 0.2803 | 3.75E-05 | -?----+?- | 4.08 | 6 | 0.6659 |
| Any CBVD | chr8 | 78,979,018 | chr8:78979018:G:A | a | g | 1.1547 | 0.2804 | 3.81E-05 | +?++++-?+ | 4.059 | 6 | 0.6687 |
| Any CBVD | chr8 | 93,243,030 | chr8:93243030:T:A | a | t | 1.01 | 0.2388 | 2.35E-05 | +++++++-+ | 8.655 | 8 | 0.3722 |
| Any CBVD | chr8 | 93,351,590 | chr8:93351590:A:G | a | g | -0.6041 | 0.1513 | 6.51E-05 | ------+-- | 24.465 | 8 | 0.001914 |
| Any CBVD | chr8 | 97,263,739 | chr8:97263739:T:C | t | c | 0.2271 | 0.0565 | 5.81E-05 | +++++++-+ | 5.154 | 8 | 0.741 |
| Any CBVD | chr8 | 97,265,159 | chr8:97265159:T:G | t | g | -0.7096 | 0.1633 | 1.39E-05 | -------+- | 5.771 | 8 | 0.6729 |
| Any CBVD | chr8 | 97,291,811 | chr8:97291811:T:C | t | c | 0.2353 | 0.0567 | 3.29E-05 | +++++++-+ | 2.181 | 8 | 0.975 |
| Any CBVD | chr8 | 97,295,337 | chr8:97295337:C:T | t | c | -0.2189 | 0.0562 | 9.94E-05 | -------+- | 3.852 | 8 | 0.8702 |
| Any CBVD | chr8 | 97,310,644 | chr8:97310644:C:T | t | c | 0.2304 | 0.0586 | 8.50E-05 | +++++++++ | 5.586 | 8 | 0.6935 |
| Any CBVD | chr8 | 97,311,563 | chr8:97311563:GATAATTTTGTTC: | g | gataattttgt | 0.2316 | 0.0586 | 7.79E-05 | +++++++++ | 6.751 | 8 | 0.5637 |
| Any CBVD | chr8 | 97,719,461 | chr8:97719461:C:G | c | g | -3.8572 | 0.7958 | 1.25E-06 | ?-?-??-?? | 0.983 | 2 | 0.6116 |
| Any CBVD | chr8 | 109,791,323 | chr8:109791323:G:C | c | g | 0.8148 | 0.2071 | 8.33E-05 | ++++++-++ | 7.203 | 8 | 0.5149 |
| Any CBVD | chr8 | 124,124,334 | chr8:124124334:G:GC | g | gc | -0.6871 | 0.1695 | 5.05E-05 | +-------- | 20.425 | 8 | 0.008842 |
| Any CBVD | chr8 | 126,357,027 | chr8:126357027:G:C | c | g | 1.1074 | 0.272 | 4.66E-05 | +++++++-+ | 12.115 | 8 | 0.1461 |
| Any CBVD | chr8 | 128,266,935 | chr8:128266935:G:A | a | g | 0.9351 | 0.2314 | 5.31E-05 | ++++++++? | 5.494 | 7 | 0.5999 |
| Any CBVD | chr9 | 4,550,731 | chr9:4550731:C:G | c | g | -0.2603 | 0.0642 | 5.01E-05 | ---++--+- | 10.291 | 8 | 0.2452 |
| Any CBVD | chr9 | 18,879,778 | chr9:18879778:A:C | a | c | 0.631 | 0.1506 | 2.79E-05 | +-++-+--+ | 8.969 | 8 | 0.345 |
| Any CBVD | chr9 | 18,883,198 | chr9:18883198:A:C | a | c | 0.6187 | 0.1522 | 4.78E-05 | +-++-+--+ | 8.751 | 8 | 0.3638 |
| Any CBVD | chr9 | 18,887,910 | chr9:18887910:G:C | c | g | -0.6949 | 0.1679 | 3.51E-05 | -+--+-++- | 7.491 | 8 | 0.4847 |
| Any CBVD | chr9 | 27,186,859 | chr9:27186859:C:T | t | c | 0.8627 | 0.2148 | 5.89E-05 | +-++--+-+ | 20.04 | 8 | 0.01019 |
| Any CBVD | chr9 | 29,377,926 | chr9:29377926:C:T | t | c | 1.9541 | 0.4751 | 3.91E-05 | ??++++??? | 0.834 | 3 | 0.8412 |
| Any CBVD | chr9 | 31,051,799 | chr9:31051799:C:G | c | g | -0.9374 | 0.2305 | 4.78E-05 | --------- | 11.88 | 8 | 0.1566 |
| Any CBVD | chr9 | 72,051,237 | chr9:72051237:G:A | a | g | 1.0262 | 0.2295 | 7.73E-06 | +++++++-+ | 15.578 | 8 | 0.04883 |
| Any CBVD | chr9 | 87,032,468 | chr9:87032468:CTGTG:C | ctgtg | c | -0.4451 | 0.1138 | 9.14E-05 | --------- | 5.776 | 8 | 0.6724 |
| Any CBVD | chr9 | 103,175,897 | chr9:103175897:G:A | a | g | 2.1215 | 0.5443 | 9.70E-05 | ??+???+-? | 8.038 | 2 | 0.01797 |
| Any CBVD | chr9 | 117,233,686 | chr9:117233686:A:T | a | t | -0.4757 | 0.1119 | 2.13E-05 | --------- | 12.849 | 8 | 0.1171 |
| Any CBVD | chr9 | 120,181,968 | chr9:120181968:A:G | a | g | -0.731 | 0.1759 | 3.26E-05 | ------+-- | 7.882 | 8 | 0.4451 |
| Any CBVD | chr9 | 126,049,824 | chr9:126049824:C:T | t | c | 0.4054 | 0.0979 | 3.48E-05 | ++++-++++ | 10.231 | 8 | 0.2492 |
| Any CBVD | chr9 | 131,567,327 | chr9:131567327:C:T | t | c | 1.3942 | 0.3276 | 2.09E-05 | +???++??+ | 1.523 | 3 | 0.6771 |
| Any CBVD | chr9 | 133,305,960 | chr9:133305960:G:T | t | g | 0.5214 | 0.1301 | 6.16E-05 | +-++-++++ | 6.381 | 8 | 0.6047 |
| Any CBVD | chr9 | 133,306,241 | chr9:133306241:C:T | t | c | 0.5179 | 0.13 | 6.78E-05 | +-+--++++ | 6.254 | 8 | 0.6188 |
| Any CBVD | chr9 | 133,307,219 | chr9:133307219:C:T | t | c | 0.5164 | 0.1299 | 7.06E-05 | +-+--++++ | 6.234 | 8 | 0.621 |
| Any CBVD | chr9 | 133,309,392 | chr9:133309392:C:T | t | c | 0.5058 | 0.1294 | 9.28E-05 | +-+--++++ | 6.224 | 8 | 0.6222 |
| Any CBVD | chr9 | 133,317,867 | chr9:133317867:A:G | a | g | -0.5184 | 0.1319 | 8.52E-05 | -+-++---- | 6.217 | 8 | 0.6229 |

| | | | | | | | | | | | | |
|---|---|---|---|---|---|---|---|---|---|---|---|---|
| Any CBVD | chr9 | 136,298,330 | chr9:136298330:G:C | c | g | 0.59 | 0.144 | 4.17E-05 | +-+++++-+ | 17.661 | 8 | 0.02392 |
| AP (any/none) | chr1 | 4,677,606 | chr1:4677606:A:G | a | g | -0.5388 | 0.1343 | 6.01E-05 | -+----+------ | 17.069 | 12 | 0.147 |
| AP (any/none) | chr1 | 4,677,632 | chr1:4677632:C:T | t | c | 0.5266 | 0.134 | 8.53E-05 | +-++++-+++ | 17.237 | 12 | 0.1409 |
| AP (any/none) | chr1 | 4,677,756 | chr1:4677756:C:T | t | c | 0.5362 | 0.1344 | 6.58E-05 | +-++++-+++ | 16.998 | 12 | 0.1497 |
| AP (any/none) | chr1 | 4,677,763 | chr1:4677763:C:A | a | c | 0.5401 | 0.1345 | 5.93E-05 | +-++++-+++ | 17.053 | 12 | 0.1476 |
| AP (any/none) | chr1 | 4,677,780 | chr1:4677780:C:T | t | c | 0.5401 | 0.1345 | 5.93E-05 | +-++++-+++ | 17.053 | 12 | 0.1476 |
| AP (any/none) | chr1 | 4,678,334 | chr1:4678334:A:G | a | g | -0.5378 | 0.1344 | 6.32E-05 | -+----+------ | 17.085 | 12 | 0.1464 |
| AP (any/none) | chr1 | 4,678,360 | chr1:4678360:C:T | t | c | 0.5387 | 0.1345 | 6.19E-05 | +-++++-+++ | 17.105 | 12 | 0.1457 |
| AP (any/none) | chr1 | 4,678,383 | chr1:4678383:A:G | a | g | -0.535 | 0.1343 | 6.80E-05 | -+----+------ | 16.951 | 12 | 0.1515 |
| AP (any/none) | chr1 | 4,678,860 | chr1:4678860:T:C | t | c | -0.535 | 0.1343 | 6.79E-05 | -+----+------ | 16.95 | 12 | 0.1515 |
| AP (any/none) | chr1 | 4,679,028 | chr1:4679028:G:A | a | g | 0.5387 | 0.1345 | 6.18E-05 | +-++++-+++ | 17.104 | 12 | 0.1457 |
| AP (any/none) | chr1 | 4,679,432 | chr1:4679432:G:A | a | g | 0.5398 | 0.1345 | 6.03E-05 | +-++++-+++ | 17.137 | 12 | 0.1445 |
| AP (any/none) | chr1 | 4,680,757 | chr1:4680757:G:C | c | g | 0.528 | 0.1348 | 8.95E-05 | +-++++-+++ | 17.379 | 12 | 0.1359 |
| AP (any/none) | chr1 | 4,680,964 | chr1:4680964:G:A | a | g | 0.539 | 0.1346 | 6.20E-05 | +-++++-+++ | 17.117 | 12 | 0.1453 |
| AP (any/none) | chr1 | 4,681,077 | chr1:4681077:A:G | a | g | -0.5391 | 0.1346 | 6.16E-05 | -+----+------ | 17.114 | 12 | 0.1454 |
| AP (any/none) | chr1 | 4,681,090 | chr1:4681090:G:A | a | g | 0.5351 | 0.1344 | 6.83E-05 | +-++++-+++ | 16.943 | 12 | 0.1518 |
| AP (any/none) | chr1 | 4,682,605 | chr1:4682605:C:T | t | c | 0.5398 | 0.1347 | 6.18E-05 | +-++++-+++ | 17.069 | 12 | 0.147 |
| AP (any/none) | chr1 | 4,682,716 | chr1:4682716:T:C | t | c | -0.5441 | 0.1344 | 5.17E-05 | -+----+------ | 16.952 | 12 | 0.1514 |
| AP (any/none) | chr1 | 4,685,274 | chr1:4685274:A:G | a | g | -0.5405 | 0.1348 | 6.07E-05 | -+----+------ | 16.98 | 12 | 0.1504 |
| AP (any/none) | chr1 | 4,686,570 | chr1:4686570:C:T | t | c | 0.5296 | 0.135 | 8.80E-05 | +-++++-+++ | 17.261 | 12 | 0.14 |
| AP (any/none) | chr1 | 4,688,130 | chr1:4688130:C:A | a | c | 0.538 | 0.1353 | 6.97E-05 | +-++++-+++ | 16.27 | 12 | 0.1792 |
| AP (any/none) | chr1 | 4,688,680 | chr1:4688680:G:A | a | g | 0.5388 | 0.1353 | 6.87E-05 | +-++++-+++ | 16.278 | 12 | 0.1788 |
| AP (any/none) | chr1 | 5,725,334 | chr1:5725334:T:G | t | g | -0.491 | 0.1262 | 9.99E-05 | ----------+-- | 4.413 | 12 | 0.9748 |
| AP (any/none) | chr1 | 20,318,024 | chr1:20318024:T:C | t | c | -0.6558 | 0.1647 | 6.84E-05 | -------+----+ | 7.443 | 12 | 0.827 |
| AP (any/none) | chr1 | 30,212,182 | chr1:30212182:T:A | a | t | 0.4713 | 0.1123 | 2.70E-05 | -++++++-++ | 13.008 | 12 | 0.3685 |
| AP (any/none) | chr1 | 30,212,851 | chr1:30212851:G:A | a | g | 0.4701 | 0.1126 | 2.96E-05 | -++++++-++ | 12.534 | 12 | 0.4038 |
| AP (any/none) | chr1 | 30,212,953 | chr1:30212953:G:T | t | g | 0.4739 | 0.1125 | 2.53E-05 | -++++++-++ | 13.288 | 12 | 0.3485 |
| AP (any/none) | chr1 | 30,214,020 | chr1:30214020:C:T | t | c | 0.4435 | 0.1126 | 8.20E-05 | -++++++-++ | 12.614 | 12 | 0.3977 |
| AP (any/none) | chr1 | 30,225,138 | chr1:30225138:G:C | c | g | 0.4569 | 0.1128 | 5.09E-05 | -++++++-++ | 12.309 | 12 | 0.4212 |
| AP (any/none) | chr1 | 30,239,517 | chr1:30239517:C:G | c | g | -0.4494 | 0.1134 | 7.45E-05 | -------+--+-+ | 10.405 | 12 | 0.5805 |
| AP (any/none) | chr1 | 30,245,125 | chr1:30245125:G:A | a | g | 0.4452 | 0.1137 | 8.99E-05 | -++++++-++ | 10.763 | 12 | 0.5494 |
| AP (any/none) | chr1 | 30,245,674 | chr1:30245674:G:A | a | g | 0.4497 | 0.1139 | 7.93E-05 | -++++++-++ | 11.469 | 12 | 0.4892 |
| AP (any/none) | chr1 | 30,249,995 | chr1:30249995:A:T | a | t | -0.4434 | 0.1137 | 9.65E-05 | +------+--+-- | 10.936 | 12 | 0.5344 |
| AP (any/none) | chr1 | 30,252,118 | chr1:30252118:T:C | t | c | -0.3426 | 0.0845 | 4.98E-05 | -------+--+-+ | 16.843 | 12 | 0.1556 |
| AP (any/none) | chr1 | 30,252,771 | chr1:30252771:T:C | t | c | -0.4605 | 0.1142 | 5.55E-05 | +------+--+-- | 11.468 | 12 | 0.4893 |
| AP (any/none) | chr1 | 30,254,185 | chr1:30254185:A:G | a | g | -0.4503 | 0.1139 | 7.65E-05 | +------+--+-- | 10.781 | 12 | 0.5478 |
| AP (any/none) | chr1 | 30,256,125 | chr1:30256125:G:A | a | g | 0.4577 | 0.1155 | 7.44E-05 | -++++++-++ | 11.172 | 12 | 0.5142 |
| AP (any/none) | chr1 | 68,592,719 | chr1:68592719:T:A | a | t | -0.1749 | 0.0436 | 6.05E-05 | -+--+-----+-- | 11.535 | 12 | 0.4837 |
| AP (any/none) | chr1 | 101,393,497 | chr1:101393497:T:C | t | c | -0.3241 | 0.0823 | 8.23E-05 | -----------+- | 11.978 | 12 | 0.4475 |
| AP (any/none) | chr1 | 168,507,926 | chr1:168507926:G:C | c | g | 0.7706 | 0.1952 | 7.90E-05 | +-+++++--+ | 15.954 | 12 | 0.1934 |
| AP (any/none) | chr1 | 225,472,748 | chr1:225472748:G:A | a | g | 0.5841 | 0.1452 | 5.76E-05 | +++-++-+++ | 16.927 | 12 | 0.1524 |
| AP (any/none) | chr1 | 231,874,448 | chr1:231874448:C:T | t | c | 0.1862 | 0.0448 | 3.21E-05 | +++++++++ | 3.992 | 12 | 0.9836 |
| AP (any/none) | chr1 | 237,536,874 | chr1:237536874:A:G | a | g | 0.7245 | 0.1849 | 8.89E-05 | +-+++++++- | 7.266 | 12 | 0.8395 |
| AP (any/none) | chr1 | 239,308,883 | chr1:239308883:G:C | c | g | 0.1825 | 0.0464 | 8.36E-05 | -+++-++-++ | 13.308 | 12 | 0.3471 |
| AP (any/none) | chr1 | 239,332,589 | chr1:239332589:C:T | t | c | 0.1807 | 0.0459 | 8.31E-05 | ++++++--++ | 5.83 | 12 | 0.9244 |
| AP (any/none) | chr10 | 6,051,026 | chr10:6051026:C:T | t | c | 0.7459 | 0.1847 | 5.39E-05 | ++++?++-+- | 7.573 | 11 | 0.751 |

| | | | | | | | | | | | | |
|---|---|---|---|---|---|---|---|---|---|---|---|---|
| AP (any/none) | chr10 | 8,216,537 | chr10:8216537:C:T | t | c | 0.2303 | 0.0582 | 7.61E-05 | ++++++-++- | 14.423 | 12 | 0.2745 |
| AP (any/none) | chr10 | 8,217,865 | chr10:8217865:A:G | a | g | -0.2341 | 0.0582 | 5.82E-05 | ------+-----+ | 14.736 | 12 | 0.2562 |
| AP (any/none) | chr10 | 8,218,033 | chr10:8218033:C:T | t | c | 0.235 | 0.0582 | 5.47E-05 | ++++++-++- | 14.786 | 12 | 0.2534 |
| AP (any/none) | chr10 | 8,218,418 | chr10:8218418:A:C | a | c | -0.234 | 0.0582 | 5.82E-05 | ------+-----+ | 14.753 | 12 | 0.2552 |
| AP (any/none) | chr10 | 8,220,163 | chr10:8220163:GA:G | g | ga | 0.2303 | 0.0584 | 8.06E-05 | ++++++-++- | 14.272 | 12 | 0.2837 |
| AP (any/none) | chr10 | 8,221,639 | chr10:8221639:T:C | t | c | -0.2336 | 0.0582 | 6.07E-05 | +-----+-----+ | 15.533 | 12 | 0.2136 |
| AP (any/none) | chr10 | 8,221,868 | chr10:8221868:C:T | t | c | 0.2327 | 0.0581 | 6.22E-05 | -+++++-+++ | 15.453 | 12 | 0.2176 |
| AP (any/none) | chr10 | 9,079,325 | chr10:9079325:A:G | a | g | 0.4198 | 0.0995 | 2.43E-05 | +++++++++ | 11.174 | 11 | 0.4288 |
| AP (any/none) | chr10 | 15,085,167 | chr10:15085167:C:T | t | c | 0.2586 | 0.0639 | 5.24E-05 | +-+++++-+- | 18.169 | 12 | 0.1107 |
| AP (any/none) | chr10 | 15,085,820 | chr10:15085820:GTC:G | g | gtc | 0.2594 | 0.064 | 4.99E-05 | +-+++++-+- | 18.178 | 12 | 0.1104 |
| AP (any/none) | chr10 | 15,087,127 | chr10:15087127:C:T | t | c | 0.2604 | 0.0643 | 5.15E-05 | +-+++++-+- | 18.489 | 12 | 0.1016 |
| AP (any/none) | chr10 | 15,087,556 | chr10:15087556:C:T | t | c | 0.2614 | 0.064 | 4.41E-05 | +-+++++-+- | 18.371 | 12 | 0.1049 |
| AP (any/none) | chr10 | 15,087,799 | chr10:15087799:G:A | a | g | 0.2583 | 0.064 | 5.41E-05 | +-+++++-+- | 18.236 | 12 | 0.1087 |
| AP (any/none) | chr10 | 15,088,676 | chr10:15088676:C:T | t | c | 0.247 | 0.0604 | 4.34E-05 | +++++++-+- | 12.573 | 12 | 0.4008 |
| AP (any/none) | chr10 | 15,090,885 | chr10:15090885:C:T | t | c | 0.2414 | 0.0603 | 6.30E-05 | +++++++-+- | 12.416 | 12 | 0.4129 |
| AP (any/none) | chr10 | 15,091,029 | chr10:15091029:G:C | c | g | 0.2418 | 0.0603 | 6.09E-05 | +++++++-+- | 12.418 | 12 | 0.4127 |
| AP (any/none) | chr10 | 15,091,424 | chr10:15091424:A:G | a | g | -0.2389 | 0.0603 | 7.39E-05 | -------+-++-- | 12.537 | 12 | 0.4036 |
| AP (any/none) | chr10 | 15,095,348 | chr10:15095348:C:G | c | g | -0.2372 | 0.055 | 1.59E-05 | -+-------+--- | 12.294 | 12 | 0.4224 |
| AP (any/none) | chr10 | 15,095,907 | chr10:15095907:C:T | t | c | 0.2351 | 0.0601 | 9.24E-05 | +++++++-+- | 13.443 | 12 | 0.3377 |
| AP (any/none) | chr10 | 15,097,466 | chr10:15097466:G:A | a | g | 0.2414 | 0.0603 | 6.31E-05 | +++++++-+- | 12.702 | 12 | 0.3911 |
| AP (any/none) | chr10 | 15,099,290 | chr10:15099290:G:C | c | g | 0.246 | 0.0602 | 4.32E-05 | +++++++-+- | 13.158 | 12 | 0.3577 |
| AP (any/none) | chr10 | 15,099,846 | chr10:15099846:T:C | t | c | -0.2009 | 0.051 | 8.25E-05 | -+-+-----+--- | 11.756 | 12 | 0.4655 |
| AP (any/none) | chr10 | 15,100,294 | chr10:15100294:CCT:C | cct | c | -0.2453 | 0.0602 | 4.58E-05 | -------+-++-- | 13.268 | 12 | 0.3498 |
| AP (any/none) | chr10 | 15,100,451 | chr10:15100451:G:A | a | g | 0.2495 | 0.0603 | 3.48E-05 | +++++++++ | 12.9 | 12 | 0.3764 |
| AP (any/none) | chr10 | 15,101,824 | chr10:15101824:A:G | a | g | -0.2417 | 0.0602 | 5.89E-05 | -------+-++-- | 13.839 | 12 | 0.3111 |
| AP (any/none) | chr10 | 15,102,060 | chr10:15102060:C:T | t | c | 0.2427 | 0.0603 | 5.75E-05 | +++++++++ | 13.275 | 12 | 0.3494 |
| AP (any/none) | chr10 | 15,102,688 | chr10:15102688:G:C | c | g | 0.2395 | 0.0603 | 7.22E-05 | +++++++++ | 13.153 | 12 | 0.358 |
| AP (any/none) | chr10 | 15,102,708 | chr10:15102708:T:TA | t | ta | -0.2446 | 0.0604 | 5.08E-05 | ---------++-- | 13.48 | 12 | 0.3352 |
| AP (any/none) | chr10 | 15,103,362 | chr10:15103362:G:A | a | g | 0.2471 | 0.0595 | 3.26E-05 | +-+-+++-+-- | 13.493 | 12 | 0.3343 |
| AP (any/none) | chr10 | 15,105,749 | chr10:15105749:A:G | a | g | -0.2474 | 0.0602 | 4.02E-05 | ---------++-- | 14.012 | 12 | 0.2999 |
| AP (any/none) | chr10 | 15,126,244 | chr10:15126244:T:C | t | c | -0.2357 | 0.0586 | 5.67E-05 | -+-+-----++- | 14.233 | 12 | 0.2861 |
| AP (any/none) | chr10 | 15,144,041 | chr10:15144041:CA:C | ca | c | -0.2717 | 0.0659 | 3.78E-05 | ---+-----+--- | 9.893 | 12 | 0.6254 |
| AP (any/none) | chr10 | 65,556,265 | chr10:65556265:A:G | a | g | -0.1783 | 0.0443 | 5.67E-05 | -+--+-------- | 8.419 | 12 | 0.7516 |
| AP (any/none) | chr10 | 105,084,729 | chr10:105084729:T:G | t | g | -0.3666 | 0.0937 | 9.14E-05 | -+-+------?-- | 10.373 | 11 | 0.4972 |
| AP (any/none) | chr10 | 115,998,599 | chr10:115998599:A:T | a | t | -0.465 | 0.1118 | 3.21E-05 | +---+-----?+ | 12.531 | 11 | 0.3251 |
| AP (any/none) | chr10 | 116,000,805 | chr10:116000805:G:A | a | g | 0.4091 | 0.1019 | 5.97E-05 | ++++-++++- | 10.334 | 11 | 0.5006 |
| AP (any/none) | chr10 | 116,798,131 | chr10:116798131:GC:G | g | gc | 1.4042 | 0.3533 | 7.05E-05 | +???++?+? | 1.912 | 4 | 0.7519 |
| AP (any/none) | chr10 | 116,805,795 | chr10:116805795:G:C | c | g | 1.3897 | 0.3455 | 5.75E-05 | +???++?+? | 1.34 | 4 | 0.8545 |
| AP (any/none) | chr10 | 116,835,204 | chr10:116835204:C:T | t | c | 1.4104 | 0.3479 | 5.02E-05 | +???++?+? | 1.725 | 4 | 0.7862 |
| AP (any/none) | chr10 | 123,606,699 | chr10:123606699:C:T | t | c | -0.2433 | 0.0595 | 4.39E-05 | ---+------+-- | 9.044 | 12 | 0.6992 |
| AP (any/none) | chr10 | 123,608,890 | chr10:123608890:A:G | a | g | 0.2356 | 0.0594 | 7.33E-05 | +-+-++++++ | 8.921 | 12 | 0.7097 |
| AP (any/none) | chr10 | 123,609,090 | chr10:123609090:T:A | a | t | -0.2361 | 0.0602 | 8.89E-05 | -+-+------+-- | 8.78 | 12 | 0.7216 |
| AP (any/none) | chr10 | 123,615,641 | chr10:123615641:A:G | a | g | 0.2392 | 0.0602 | 7.14E-05 | +-+-++++++ | 8.408 | 12 | 0.7525 |
| AP (any/none) | chr10 | 123,616,551 | chr10:123616551:A:C | a | c | 0.2419 | 0.0603 | 5.97E-05 | +++-+++++- | 8.016 | 12 | 0.7839 |
| AP (any/none) | chr10 | 123,618,211 | chr10:123618211:AT:A | a | at | -0.2329 | 0.0595 | 9.10E-05 | ---+------+-- | 8.708 | 12 | 0.7277 |
| AP (any/none) | chr10 | 124,469,067 | chr10:124469067:C:T | t | c | -0.2176 | 0.0555 | 8.71E-05 | ---------+-- | 13.504 | 12 | 0.3335 |

| | | | | | | | | | | | | |
|---|---|---|---|---|---|---|---|---|---|---|---|---|
| AP (any/none) | chr10 | 124,469,789 | chr10:124469789:A:G | a | g | 0.2171 | 0.0549 | 7.66E-05 | +++++++++ | 13.923 | 12 | 0.3056 |
| AP (any/none) | chr10 | 124,470,382 | chr10:124470382:GC:G | g | gc | -0.2159 | 0.0553 | 9.32E-05 | ----------+-- | 13.657 | 12 | 0.3231 |
| AP (any/none) | chr10 | 124,471,753 | chr10:124471753:A:G | a | g | 0.2157 | 0.0551 | 8.98E-05 | +++++++++ | 14.015 | 12 | 0.2997 |
| AP (any/none) | chr10 | 124,471,954 | chr10:124471954:G:A | a | g | -0.2143 | 0.0549 | 9.51E-05 | ----------+-- | 14.014 | 12 | 0.2998 |
| AP (any/none) | chr10 | 124,472,715 | chr10:124472715:G:C | c | g | -0.2179 | 0.055 | 7.45E-05 | ----------+-- | 13.904 | 12 | 0.3069 |
| AP (any/none) | chr10 | 124,472,716 | chr10:124472716:T:C | t | c | 0.2156 | 0.0549 | 8.66E-05 | +++++++++ | 13.952 | 12 | 0.3038 |
| AP (any/none) | chr10 | 124,473,059 | chr10:124473059:G:A | a | g | -0.2151 | 0.055 | 9.16E-05 | ----------+-- | 14.052 | 12 | 0.2974 |
| AP (any/none) | chr10 | 124,473,270 | chr10:124473270:G:C | c | g | -0.2151 | 0.055 | 9.18E-05 | ----------+-- | 14.055 | 12 | 0.2972 |
| AP (any/none) | chr10 | 124,473,517 | chr10:124473517:T:A | a | t | -0.2152 | 0.055 | 9.09E-05 | ----------+-- | 14.058 | 12 | 0.297 |
| AP (any/none) | chr10 | 124,474,474 | chr10:124474474:A:G | a | g | 0.2146 | 0.055 | 9.54E-05 | +++++++++ | 14.086 | 12 | 0.2953 |
| AP (any/none) | chr10 | 124,474,774 | chr10:124474774:C:T | t | c | -0.2144 | 0.055 | 9.67E-05 | ----------+-- | 14.001 | 12 | 0.3006 |
| AP (any/none) | chr10 | 124,476,928 | chr10:124476928:C:T | t | c | -0.2148 | 0.055 | 9.33E-05 | ----------+-- | 13.78 | 12 | 0.315 |
| AP (any/none) | chr10 | 124,479,602 | chr10:124479602:C:T | t | c | -0.2184 | 0.0548 | 6.68E-05 | ----------++- | 14.218 | 12 | 0.287 |
| AP (any/none) | chr10 | 124,480,804 | chr10:124480804:G:A | a | g | -0.2155 | 0.0545 | 7.60E-05 | ----------++- | 14.527 | 12 | 0.2683 |
| AP (any/none) | chr10 | 124,481,076 | chr10:124481076:G:T | t | g | -0.218 | 0.0548 | 6.93E-05 | ----------++- | 14.246 | 12 | 0.2853 |
| AP (any/none) | chr10 | 124,481,633 | chr10:124481633:G:A | a | g | -0.2177 | 0.0548 | 7.10E-05 | ----------++- | 14.2 | 12 | 0.2881 |
| AP (any/none) | chr10 | 124,481,698 | chr10:124481698:G:T | t | g | -0.2176 | 0.0548 | 7.13E-05 | ----------++- | 14.222 | 12 | 0.2868 |
| AP (any/none) | chr10 | 124,482,537 | chr10:124482537:G:A | a | g | -0.2141 | 0.0549 | 9.65E-05 | ----------++- | 14.554 | 12 | 0.2667 |
| AP (any/none) | chr10 | 124,482,953 | chr10:124482953:G:T | t | g | -0.2154 | 0.0553 | 9.90E-05 | ----------++- | 13.667 | 12 | 0.3225 |
| AP (any/none) | chr10 | 124,482,997 | chr10:124482997:C:T | t | c | -0.2172 | 0.0553 | 8.58E-05 | ----------++- | 13.569 | 12 | 0.3291 |
| AP (any/none) | chr10 | 124,483,075 | chr10:124483075:T:A | a | t | -0.2224 | 0.0562 | 7.51E-05 | ----------+-- | 13.158 | 12 | 0.3577 |
| AP (any/none) | chr11 | 7,422,666 | chr11:7422666:A:G | a | g | -0.1938 | 0.0493 | 8.56E-05 | -+--------+-+ | 5.483 | 12 | 0.9399 |
| AP (any/none) | chr11 | 7,423,574 | chr11:7423574:C:A | a | c | 0.2019 | 0.0494 | 4.40E-05 | +-+++++++- | 5.479 | 12 | 0.94 |
| AP (any/none) | chr11 | 7,557,350 | chr11:7557350:T:G | t | g | -0.4268 | 0.1058 | 5.49E-05 | ----+--+++-- | 25.47 | 12 | 0.01275 |
| AP (any/none) | chr11 | 7,575,156 | chr11:7575156:C:A | a | c | 0.3196 | 0.0807 | 7.41E-05 | ++++++++-- | 25.444 | 12 | 0.01285 |
| AP (any/none) | chr11 | 12,096,394 | chr11:12096394:G:A | a | g | -0.207 | 0.0513 | 5.39E-05 | ----+-------+ | 12.291 | 12 | 0.4226 |
| AP (any/none) | chr11 | 41,416,931 | chr11:41416931:A:G | a | g | -0.635 | 0.1535 | 3.52E-05 | ++-------+--- | 11.45 | 12 | 0.4908 |
| AP (any/none) | chr11 | 67,281,771 | chr11:67281771:G:A | a | g | 1.2675 | 0.2969 | 1.96E-05 | +-++?+?+? | 6.315 | 6 | 0.3888 |
| AP (any/none) | chr11 | 85,959,342 | chr11:85959342:CAA:C | caa | c | 0.1863 | 0.0454 | 4.00E-05 | ++-++++++- | 13.248 | 12 | 0.3513 |
| AP (any/none) | chr11 | 85,959,902 | chr11:85959902:A:C | a | c | 0.1869 | 0.0454 | 3.78E-05 | ++-++++++- | 13.202 | 12 | 0.3546 |
| AP (any/none) | chr11 | 85,960,208 | chr11:85960208:G:GATCT | g | gatct | 0.1875 | 0.0454 | 3.59E-05 | ++-++++++- | 13.744 | 12 | 0.3174 |
| AP (any/none) | chr11 | 85,970,540 | chr11:85970540:A:G | a | g | 0.1802 | 0.0454 | 7.25E-05 | +++++++++ | 12.738 | 12 | 0.3884 |
| AP (any/none) | chr11 | 85,971,774 | chr11:85971774:A:G | a | g | 0.1852 | 0.0453 | 4.40E-05 | ++-++++++- | 13.409 | 12 | 0.34 |
| AP (any/none) | chr11 | 85,978,669 | chr11:85978669:A:T | a | t | 0.1841 | 0.0454 | 4.96E-05 | ++-++++++- | 13.445 | 12 | 0.3375 |
| AP (any/none) | chr11 | 85,980,767 | chr11:85980767:C:A | a | c | -0.1884 | 0.0453 | 3.22E-05 | --+---------- | 13.559 | 12 | 0.3297 |
| AP (any/none) | chr11 | 86,006,880 | chr11:86006880:A:C | a | c | 0.1883 | 0.0454 | 3.42E-05 | +++++++-+- | 12.195 | 12 | 0.4301 |
| AP (any/none) | chr11 | 86,007,188 | chr11:86007188:G:T | t | g | -0.189 | 0.0452 | 2.92E-05 | --+----+----- | 13.769 | 12 | 0.3157 |
| AP (any/none) | chr11 | 86,013,618 | chr11:86013618:T:C | t | c | 0.1891 | 0.0454 | 3.13E-05 | +++++++-+- | 11.976 | 12 | 0.4476 |
| AP (any/none) | chr11 | 86,022,772 | chr11:86022772:C:CAT | cat | c | -0.1896 | 0.0453 | 2.91E-05 | -------+----- | 10.935 | 12 | 0.5345 |
| AP (any/none) | chr11 | 86,022,967 | chr11:86022967:C:T | t | c | -0.1891 | 0.0453 | 3.02E-05 | -------+----- | 11.15 | 12 | 0.5161 |
| AP (any/none) | chr11 | 86,029,367 | chr11:86029367:A:G | a | g | 0.1869 | 0.0453 | 3.73E-05 | +++++++-+- | 10.914 | 12 | 0.5363 |
| AP (any/none) | chr11 | 86,030,088 | chr11:86030088:C:T | t | c | -0.1834 | 0.0454 | 5.41E-05 | -------+----- | 10.615 | 12 | 0.5622 |
| AP (any/none) | chr11 | 86,031,175 | chr11:86031175:CA:C | ca | c | 0.1856 | 0.0454 | 4.42E-05 | +++++++-+- | 10.533 | 12 | 0.5693 |
| AP (any/none) | chr11 | 86,034,995 | chr11:86034995:C:T | t | c | -0.1904 | 0.0454 | 2.71E-05 | -------+----- | 10.764 | 12 | 0.5492 |
| AP (any/none) | chr11 | 86,035,124 | chr11:86035124:CT:C | ct | c | 0.1938 | 0.0435 | 8.36E-06 | +++++++++ | 7.9 | 12 | 0.7929 |
| AP (any/none) | chr11 | 86,035,769 | chr11:86035769:T:C | t | c | 0.1859 | 0.0456 | 4.53E-05 | +++++++-+- | 10.924 | 12 | 0.5354 |

| | | | | | | | | | | | | |
|---|---|---|---|---|---|---|---|---|---|---|---|---|
| AP (any/none) | chr11 | 86,035,780 | chr11:86035780:TA:T | t | ta | -0.1888 | 0.0457 | 3.54E-05 | -------+----- | 10.822 | 12 | 0.5443 |
| AP (any/none) | chr11 | 86,037,344 | chr11:86037344:T:C | t | c | 0.1857 | 0.0455 | 4.51E-05 | +++++++-+- | 10.325 | 12 | 0.5875 |
| AP (any/none) | chr11 | 86,039,629 | chr11:86039629:A:T | a | t | 0.1857 | 0.0455 | 4.46E-05 | +++++++-+- | 10.218 | 12 | 0.5969 |
| AP (any/none) | chr11 | 86,046,547 | chr11:86046547:C:T | t | c | -0.1891 | 0.0453 | 3.01E-05 | -------+----- | 11.065 | 12 | 0.5234 |
| AP (any/none) | chr11 | 86,046,571 | chr11:86046571:C:G | c | g | 0.179 | 0.0455 | 8.23E-05 | +++++++-+- | 10.999 | 12 | 0.529 |
| AP (any/none) | chr11 | 86,046,575 | chr11:86046575:C:T | t | c | -0.1789 | 0.0455 | 8.27E-05 | -------+----- | 10.99 | 12 | 0.5298 |
| AP (any/none) | chr11 | 86,048,986 | chr11:86048986:T:C | t | c | 0.1872 | 0.0455 | 3.85E-05 | +++++++-+- | 11.228 | 12 | 0.5095 |
| AP (any/none) | chr11 | 86,051,202 | chr11:86051202:C:T | t | c | -0.1803 | 0.0455 | 7.36E-05 | -------+----- | 10.118 | 12 | 0.6056 |
| AP (any/none) | chr11 | 86,054,256 | chr11:86054256:G:A | a | g | -0.1814 | 0.0454 | 6.58E-05 | -------+----- | 10.195 | 12 | 0.5989 |
| AP (any/none) | chr11 | 86,063,377 | chr11:86063377:GTA:G | g | gta | -0.179 | 0.0455 | 8.19E-05 | ------------- | 10.242 | 12 | 0.5947 |
| AP (any/none) | chr11 | 86,065,502 | chr11:86065502:G:A | a | g | -0.1798 | 0.0455 | 7.85E-05 | ------------- | 10.803 | 12 | 0.5458 |
| AP (any/none) | chr11 | 86,076,782 | chr11:86076782:C:T | t | c | -0.1808 | 0.0458 | 7.98E-05 | ---------+--- | 9.804 | 12 | 0.6331 |
| AP (any/none) | chr11 | 86,077,309 | chr11:86077309:C:T | t | c | -0.1844 | 0.0458 | 5.58E-05 | ---------+--- | 10.21 | 12 | 0.5975 |
| AP (any/none) | chr11 | 86,083,500 | chr11:86083500:A:G | a | g | 0.1969 | 0.0462 | 2.07E-05 | +++++++-+- | 8.857 | 12 | 0.7151 |
| AP (any/none) | chr11 | 86,083,718 | chr11:86083718:C:T | t | c | -0.19 | 0.046 | 3.63E-05 | -------+-+--- | 10.568 | 12 | 0.5663 |
| AP (any/none) | chr11 | 86,083,719 | chr11:86083719:A:G | a | g | 0.1929 | 0.0465 | 3.30E-05 | +++++++-+- | 8.866 | 12 | 0.7144 |
| AP (any/none) | chr11 | 86,084,482 | chr11:86084482:T:C | t | c | 0.1923 | 0.0465 | 3.48E-05 | +++++++-+- | 8.79 | 12 | 0.7208 |
| AP (any/none) | chr11 | 86,084,908 | chr11:86084908:A:G | a | g | 0.1888 | 0.0466 | 5.04E-05 | +++++++-+- | 8.176 | 12 | 0.7712 |
| AP (any/none) | chr11 | 86,086,978 | chr11:86086978:T:G | t | g | 0.1956 | 0.0465 | 2.61E-05 | +++++++-+- | 8.422 | 12 | 0.7514 |
| AP (any/none) | chr11 | 101,532,593 | chr11:101532593:G:A | a | g | -0.5806 | 0.1333 | 1.33E-05 | ----+-------- | 13.734 | 12 | 0.318 |
| AP (any/none) | chr11 | 101,594,639 | chr11:101594639:T:C | t | c | 0.5855 | 0.1364 | 1.78E-05 | ++++-++++- | 15.591 | 12 | 0.2107 |
| AP (any/none) | chr11 | 101,671,405 | chr11:101671405:A:G | a | g | 0.6548 | 0.161 | 4.77E-05 | ++++-++++- | 13.355 | 12 | 0.3438 |
| AP (any/none) | chr11 | 101,689,617 | chr11:101689617:G:T | t | g | -0.5247 | 0.1288 | 4.66E-05 | ----+----+--- | 15.75 | 12 | 0.2029 |
| AP (any/none) | chr11 | 129,406,767 | chr11:129406767:C:T | t | c | 0.2888 | 0.0734 | 8.26E-05 | -+++-++-++ | 15.395 | 12 | 0.2206 |
| AP (any/none) | chr11 | 129,407,090 | chr11:129407090:C:CT | ct | c | 0.2889 | 0.0734 | 8.20E-05 | -+++-++-++ | 15.346 | 12 | 0.223 |
| AP (any/none) | chr11 | 129,407,322 | chr11:129407322:T:G | t | g | -0.2888 | 0.0733 | 8.09E-05 | +---+--+--+- | 15.012 | 12 | 0.2408 |
| AP (any/none) | chr11 | 129,407,383 | chr11:129407383:A:G | a | g | -0.289 | 0.0732 | 7.85E-05 | +---+--+--+- | 14.906 | 12 | 0.2466 |
| AP (any/none) | chr12 | 702,402 | chr12:702402:A:G | a | g | 0.2402 | 0.0584 | 3.91E-05 | +++-+++-++ | 16.002 | 12 | 0.1912 |
| AP (any/none) | chr12 | 713,498 | chr12:713498:T:G | t | g | 0.2583 | 0.0612 | 2.45E-05 | +++-+++++- | 15.55 | 12 | 0.2127 |
| AP (any/none) | chr12 | 720,413 | chr12:720413:T:A | a | t | -0.2415 | 0.0615 | 8.74E-05 | ---+--------+ | 14.863 | 12 | 0.249 |
| AP (any/none) | chr12 | 720,610 | chr12:720610:G:T | t | g | -0.2413 | 0.0614 | 8.55E-05 | ---+--------+ | 15.659 | 12 | 0.2073 |
| AP (any/none) | chr12 | 732,756 | chr12:732756:G:A | a | g | -0.2482 | 0.0622 | 6.66E-05 | ---+--------+ | 15.753 | 12 | 0.2028 |
| AP (any/none) | chr12 | 738,675 | chr12:738675:G:T | t | g | -0.2639 | 0.0655 | 5.52E-05 | ------------+ | 14.198 | 12 | 0.2883 |
| AP (any/none) | chr12 | 880,258 | chr12:880258:C:T | t | c | 0.6008 | 0.1537 | 9.26E-05 | +-+++++++- | 8.53 | 12 | 0.7425 |
| AP (any/none) | chr12 | 886,452 | chr12:886452:A:C | a | c | -0.6136 | 0.1543 | 6.96E-05 | -+---------+- | 8.249 | 12 | 0.7654 |
| AP (any/none) | chr12 | 24,957,105 | chr12:24957105:G:GT | g | gt | 0.2638 | 0.0664 | 7.05E-05 | +-++-+++++ | 15.813 | 12 | 0.1999 |
| AP (any/none) | chr12 | 24,960,889 | chr12:24960889:A:G | a | g | 0.246 | 0.0629 | 9.30E-05 | +-++-+++++ | 16.011 | 12 | 0.1907 |
| AP (any/none) | chr12 | 24,969,880 | chr12:24969880:T:C | t | c | 0.251 | 0.0643 | 9.47E-05 | +-++-+++++ | 14.2 | 12 | 0.2881 |
| AP (any/none) | chr12 | 24,970,927 | chr12:24970927:T:C | t | c | 0.295 | 0.0676 | 1.28E-05 | +-++-+++++ | 15.346 | 12 | 0.223 |
| AP (any/none) | chr12 | 24,998,193 | chr12:24998193:T:C | t | c | 0.2293 | 0.0549 | 2.93E-05 | ++++-++++- | 12.004 | 12 | 0.4454 |
| AP (any/none) | chr12 | 24,998,293 | chr12:24998293:A:G | a | g | 0.2286 | 0.0549 | 3.17E-05 | ++++-++++- | 11.999 | 12 | 0.4458 |
| AP (any/none) | chr12 | 24,998,310 | chr12:24998310:TTAA:T | t | ttaa | -0.2283 | 0.055 | 3.26E-05 | ----+-------+ | 12.021 | 12 | 0.444 |
| AP (any/none) | chr12 | 24,998,690 | chr12:24998690:A:T | a | t | 0.2245 | 0.0552 | 4.81E-05 | ++++-++++- | 12.209 | 12 | 0.429 |
| AP (any/none) | chr12 | 25,000,181 | chr12:25000181:C:G | c | g | 0.2255 | 0.0549 | 4.07E-05 | ++++-++++- | 11.856 | 12 | 0.4573 |
| AP (any/none) | chr12 | 25,001,800 | chr12:25001800:C:G | c | g | 0.23 | 0.0551 | 2.95E-05 | ++++-++++- | 11.885 | 12 | 0.4549 |
| AP (any/none) | chr12 | 64,854,968 | chr12:64854968:A:G | a | g | -0.3174 | 0.0803 | 7.69E-05 | -------+---+- | 15.982 | 12 | 0.1921 |

**Table S4: Association of Bellenguez et al AD risk loci with AD related dementia pathologies**

| Bellenguez *et al* | | | | | Lewy bodies (full) | | Lewy bodies (partial) | | Lewy bodies (binary) | | CAA (ordinal) | | CAA (binary) | | HS | | TDP-43 (ordinal) | | TDP-43 (binary) | |
|---|---|---|---|---|---|---|---|---|---|---|---|---|---|---|---|---|---|---|---|---|
| **Variant** | **RA** | **OA** | **Closest Gene** | **OR** | **OR** | **P** | **OR** | **P** | **OR** | **P** | **OR** | **P** | **OR** | **P** | **OR** | **P** | **OR** | **P** | **OR** | **P** |
| rs74685827 | G | T | SORL1 | 1.11 | 1.23 | 4.72E-02 | 1.22 | 6.98E-02 | 1.17 | 1.47E-01 | 1.19 | 9.93E-02 | 1.06 | 6.34E-01 | 1.40 | 4.23E-02 | 1.46 | 3.33E-02 | 1.30 | 1.19E-01 |
| rs61762319 | G | A | MME | 1.18 | 0.86 | 1.44E-01 | 0.86 | 1.46E-01 | 0.86 | 1.29E-01 | 1.08 | 3.96E-01 | 0.98 | 8.69E-01 | 1.07 | 6.39E-01 | 1.42 | 2.42E-02 | 1.37 | 3.97E-02 |
| rs11218343 | T | C | SORL1 | 1.22 | 1.12 | 1.45E-01 | 1.12 | 1.65E-01 | 1.09 | 2.86E-01 | 0.91 | 2.01E-01 | 0.82 | 2.02E-02 | 0.85 | 1.86E-01 | 0.88 | 2.86E-01 | 0.86 | 1.82E-01 |
| rs6489896 | C | T | TPCN1 | 1.08 | 0.99 | 8.14E-01 | 0.99 | 7.99E-01 | 0.98 | 7.28E-01 | 1.23 | 2.20E-04 | 1.16 | 1.63E-02 | 1.06 | 5.06E-01 | 1.17 | 9.98E-02 | 1.14 | 1.50E-01 |
| rs34173062 | A | G | SHARPIN | 1.14 | 0.97 | 5.70E-01 | 0.98 | 7.45E-01 | 0.98 | 7.99E-01 | 1.02 | 7.72E-01 | 1.03 | 6.58E-01 | 0.77 | 3.18E-03 | 0.91 | 3.03E-01 | 0.89 | 1.94E-01 |
| rs6014724 | A | G | CASS4 | 1.12 | 0.98 | 6.92E-01 | 0.96 | 4.07E-01 | 0.96 | 4.96E-01 | 0.97 | 5.45E-01 | 1.00 | 9.96E-01 | 1.20 | 2.22E-02 | 1.06 | 4.68E-01 | 1.10 | 2.28E-01 |
| rs10952097 | T | C | ICA1 | 1.07 | 0.98 | 6.81E-01 | 0.98 | 6.62E-01 | 1.00 | 9.57E-01 | 0.95 | 2.72E-01 | 0.96 | 4.17E-01 | 0.83 | 1.50E-02 | 0.86 | 5.51E-02 | 0.93 | 3.75E-01 |
| rs7225151 | A | G | SC1MP | 1.05 | 0.92 | 5.51E-02 | 0.92 | 6.51E-02 | 0.91 | 4.45E-02 | 0.98 | 6.43E-01 | 0.99 | 8.44E-01 | 0.94 | 3.90E-01 | 0.84 | 1.30E-02 | 0.87 | 3.72E-02 |
| rs10131280 | G | A | IGH cluster | 1.06 | 1.11 | 2.84E-02 | 1.11 | 3.43E-02 | 1.11 | 2.45E-02 | 0.97 | 4.84E-01 | 1.00 | 9.53E-01 | 1.07 | 3.63E-01 | 1.12 | 1.07E-01 | 1.06 | 3.84E-01 |
| rs12592898 | G | A | CTSH | 1.06 | 0.99 | 8.03E-01 | 0.99 | 8.37E-01 | 1.01 | 8.95E-01 | 1.08 | 5.93E-02 | 1.02 | 6.42E-01 | 1.03 | 7.12E-01 | 1.14 | 6.64E-02 | 1.15 | 3.89E-02 |
| rs72777026 | G | A | ADAM17 | 1.06 | 1.14 | 1.99E-03 | 1.14 | 1.71E-03 | 1.13 | 7.31E-03 | 0.97 | 5.33E-01 | 0.97 | 4.50E-01 | 1.06 | 3.39E-01 | 1.03 | 6.19E-01 | 1.00 | 9.73E-01 |
| rs6605556 | A | G | HLA-DOA1 | 1.07 | 0.96 | 3.11E-01 | 0.95 | 2.67E-01 | 0.96 | 3.00E-01 | 1.00 | 9.82E-01 | 1.01 | 7.55E-01 | 0.90 | 1.03E-01 | 0.84 | 8.78E-03 | 0.92 | 1.94E-01 |
| rs199515 | C | G | WNT3 | 1.07 | 1.01 | 7.56E-01 | 1.03 | 4.44E-01 | 1.04 | 3.63E-01 | 0.99 | 6.95E-01 | 1.03 | 4.35E-01 | 0.83 | 1.29E-03 | 0.90 | 6.40E-02 | 0.88 | 2.76E-02 |
| rs3848143 | G | A | SNX1 | 1.07 | 0.97 | 4.20E-01 | 0.97 | 3.67E-01 | 0.97 | 3.91E-01 | 1.00 | 9.12E-01 | 1.00 | 9.10E-01 | 1.08 | 1.64E-01 | 1.16 | 9.37E-03 | 1.07 | 1.94E-01 |
| rs6742 | C | T | SLC2A4RG | 1.06 | 1.04 | 3.52E-01 | 1.05 | 2.35E-01 | 1.04 | 3.65E-01 | 0.93 | 5.10E-02 | 0.91 | 2.52E-02 | 1.02 | 7.60E-01 | 0.90 | 7.21E-02 | 0.91 | 1.14E-01 |
| rs62374257 | C | T | COX7C | 1.06 | 0.97 | 3.59E-01 | 0.97 | 3.29E-01 | 0.97 | 3.29E-01 | 0.96 | 1.88E-01 | 0.93 | 4.12E-02 | 1.00 | 9.53E-01 | 1.13 | 3.01E-02 | 1.08 | 1.39E-01 |
| rs602602 | T | A | MINDY2 | 1.05 | 0.99 | 6.76E-01 | 0.98 | 6.41E-01 | 0.97 | 4.32E-01 | 1.07 | 3.17E-02 | 1.10 | 6.73E-03 | 1.12 | 2.31E-02 | 0.98 | 7.62E-01 | 1.01 | 8.71E-01 |
| rs5848 | T | C | GRN | 1.08 | 0.99 | 8.22E-01 | 1.00 | 9.03E-01 | 1.00 | 9.82E-01 | 0.97 | 3.06E-01 | 1.00 | 9.46E-01 | 0.72 | 1.29E-10 | 0.77 | 8.33E-07 | 0.79 | 4.16E-06 |
| rs871269 | C | T | TNIP1 | 1.05 | 1.02 | 5.81E-01 | 1.02 | 5.58E-01 | 1.02 | 5.46E-01 | 1.02 | 5.21E-01 | 1.03 | 3.32E-01 | 1.10 | 5.29E-02 | 1.10 | 6.57E-02 | 1.12 | 2.09E-02 |
| rs3851179 | C | T | EED | 1.10 | 1.05 | 1.29E-01 | 1.05 | 1.15E-01 | 1.05 | 1.14E-01 | 1.08 | 1.23E-02 | 1.14 | 1.20E-04 | 0.99 | 9.03E-01 | 0.95 | 3.27E-01 | 0.97 | 4.92E-01 |
| rs7157106 | A | G | IGH cluster | 1.04 | 0.99 | 7.71E-01 | 0.99 | 7.79E-01 | 0.98 | 7.43E-01 | 0.98 | 6.05E-01 | 0.91 | 4.41E-02 | 0.93 | 3.47E-01 | 0.85 | 1.41E-02 | 0.86 | 3.38E-02 |
| rs73223431 | T | C | PTK2B | 1.07 | 1.02 | 6.05E-01 | 1.01 | 7.46E-01 | 1.01 | 7.84E-01 | 1.03 | 2.39E-01 | 1.01 | 8.34E-01 | 1.13 | 8.24E-03 | 1.02 | 6.71E-01 | 1.04 | 3.63E-01 |
| rs450674 | T | C | MAF | 1.03 | 0.99 | 8.06E-01 | 0.99 | 8.39E-01 | 0.98 | 5.71E-01 | 0.98 | 4.67E-01 | 0.93 | 3.42E-02 | 1.01 | 7.89E-01 | 0.96 | 3.72E-01 | 1.00 | 9.25E-01 |
| rs4277405 | T | C | ACE | 1.06 | 0.94 | 5.73E-02 | 0.94 | 6.27E-02 | 0.95 | 8.88E-02 | 0.95 | 8.90E-02 | 0.92 | 1.66E-02 | 1.07 | 1.84E-01 | 0.97 | 5.92E-01 | 0.94 | 1.58E-01 |
| rs6733839 | T | C | BIN1 | 1.16 | 0.93 | 2.34E-02 | 0.93 | 1.21E-02 | 0.93 | 1.79E-02 | 0.99 | 6.48E-01 | 0.98 | 5.97E-01 | 1.04 | 4.33E-01 | 0.95 | 2.97E-01 | 0.94 | 2.09E-01 |
| rs11787077 | C | T | CLU | 1.09 | 1.01 | 8.30E-01 | 1.01 | 6.62E-01 | 1.01 | 7.02E-01 | 1.04 | 2.08E-01 | 1.07 | 4.26E-02 | 0.95 | 3.21E-01 | 0.96 | 3.52E-01 | 1.03 | 5.49E-01 |
| rs10437655 | A | G | SPI1 | 1.05 | 0.93 | 1.86E-02 | 0.93 | 2.03E-02 | 0.93 | 2.44E-02 | 0.97 | 3.49E-01 | 0.98 | 4.74E-01 | 1.05 | 3.08E-01 | 1.07 | 1.55E-01 | 1.07 | 1.46E-01 |
| rs13237518 | C | A | TMEM106B | 1.05 | 0.95 | 8.40E-02 | 0.94 | 4.66E-02 | 0.94 | 4.96E-02 | 0.98 | 5.34E-01 | 0.96 | 1.72E-01 | 1.43 | 8.59E-15 | 1.38 | 2.80E-11 | 1.34 | 1.59E-10 |
| rs6943429 | T | C | UMAD1 | 1.05 | 1.05 | 9.73E-02 | 1.05 | 8.24E-02 | 1.06 | 5.53E-02 | 1.00 | 8.88E-01 | 1.01 | 7.05E-01 | 0.97 | 4.82E-01 | 0.91 | 6.23E-02 | 0.91 | 4.87E-02 |
| rs7912495 | G | A | USP6NL | 1.07 | 1.06 | 6.70E-02 | 1.07 | 2.57E-02 | 1.06 | 5.20E-02 | 1.06 | 5.54E-02 | 1.06 | 9.16E-02 | 1.04 | 4.35E-01 | 1.05 | 3.11E-01 | 1.03 | 4.94E-01 |

Table 3.5 Summary statistics from Bellenguez et al. clinical AD candidate-variant analysis for ADRD pathologic phenotypes. Refer to Table 3.1 for additional information and sample size. Odds ratios (OR) refer to the risk allele (RA) from Bellenguez et al. GWAS. Highlighted and bolded ORs indicate significance (P < 0.05). Only variants showing significant association with at least one lesion is included in the table.

Abbreviations: RA, risk allele; OA, other allele; OR, odds ratio.

Variants lacking nominal association (p > 0.05) not shown

**Table S5: Association of Bellenguez et al AD risk loci with cerebrovascular and vascular brain injury pathologies**

| Variant | Chr | Pos | Gene | Min | Maj | MAF | OR | Arteriolosclerosis | | Arteriolosclerosis (any/none) | | Atherosclerosis | | Atherosclerosis (any/none) | | Any CBVD | | Hemorrhages | | Infarcts/Lacunes | | Microinfarcts | | WBVD | | WMR | | WMR (any/none) | |
|---|---|---|---|---|---|---|---|---|---|---|---|---|---|---|---|---|---|---|---|---|---|---|---|---|---|---|---|---|---|
| | | | | | | | | Beta | OR | Beta | OR | Beta | Pval | Beta | Pval | Beta | Pval | Beta | Pval | Beta | Pval | Beta | Pval | Beta | Pval | Beta | Pval | Beta | Pval |
| rs679515 | 1 | 207577223 | CR1 | T | C | 0.188 | 1.13 | 0.0406 | 0.3782 | 0.0832 | 0.1826 | -0.0082 | 0.8546 | 0.0463 | 0.46 | 0.0382 | 0.668 | 0.0208 | 0.8623 | 0.0337 | 0.5603 | -0.0519 | 0.3608 | -0.0035 | 0.9404 | -0.0003 | 0.9974 | 0.0636 | 0.3304 |
| rs72777026 | 2 | 9558882 | ADAM17 | G | A | 0.144 | 1.06 | 0.025 | 0.3954 | 0.0533 | 0.1793 | -0.0748 | 0.008845 | -0.0291 | 0.4639 | -0.0205 | 0.7174 | 0.0867 | 0.2557 | 0.0126 | 0.7356 | 0.0446 | 0.2223 | 0.0155 | 0.602 | -0.0483 | 0.3872 | 0.0173 | 0.6783 |
| rs17020490 | 2 | 37304796 | PRKD3 | C | T | 0.145 | 1.06 | -0.1202 | 0.05282 | -0.1207 | 0.1509 | -0.0288 | 0.6346 | -0.0103 | 0.9034 | -0.0048 | 0.9696 | -0.0056 | 0.9727 | -0.0239 | 0.7622 | -0.0692 | 0.3746 | -0.0599 | 0.3512 | -0.0071 | 0.9531 | 0.0195 | 0.826 |
| rs6733839 | 2 | 127135234 | BIN1 | T | C | 0.389 | 1.17 | -0.0167 | 0.6446 | 0.0087 | 0.8576 | -0.0108 | 0.7572 | -0.0196 | 0.6882 | -0.0979 | 0.1582 | -0.0644 | 0.4928 | -0.0589 | 0.1975 | 0.0043 | 0.9238 | -0.0274 | 0.4534 | 0.0051 | 0.9406 | -0.0131 | 0.7976 |
| rs139643391 | 2 | 202878716 | WDR12 | T | TC | 0.131 | 0.94 | 0.0231 | 0.5861 | 0.0573 | 0.316 | 0.0338 | 0.4088 | 0.0331 | 0.5661 | -0.0215 | 0.7971 | 0.0769 | 0.4861 | 0.0529 | 0.3172 | -0.0052 | 0.9206 | -0.0031 | 0.9421 | -0.0594 | 0.4612 | 0.0547 | 0.362 |
| rs10933431 | 2 | 233117202 | INPP5D | G | C | 0.234 | 0.93 | -0.0065 | 0.8951 | -0.0362 | 0.587 | -0.0011 | 0.9824 | 0.0173 | 0.795 | -0.1997 | 0.03485 | -0.2314 | 0.07613 | 0.0457 | 0.4681 | -0.0056 | 0.9278 | -0.0198 | 0.6939 | -0.0083 | 0.9281 | -0.0861 | 0.2204 |
| rs16824536 | 3 | 155069722 | MME | A | G | 0.054 | 0.92 | 0.1073 | 0.1587 | 0.0311 | 0.7652 | 0.1731 | 0.01885 | 0.0665 | 0.5266 | -0.1094 | 0.4688 | 0.1035 | 0.6019 | 0.0192 | 0.8438 | 0.094 | 0.3258 | -0.0445 | 0.5726 | -0.0791 | 0.5652 | -0.0558 | 0.6056 |
| rs61762319 | 3 | 155084189 | MME | G | A | 0.026 | 1.16 | -0.0063 | 0.9087 | 0.0316 | 0.6706 | 0.0854 | 0.1089 | 0.0161 | 0.8279 | -0.1629 | 0.1281 | 0.3781 | 0.008943 | 0.0637 | 0.3656 | -0.0175 | 0.801 | -0.0037 | 0.9483 | 0.0326 | 0.7555 | -0.0243 | 0.7564 |
| rs3822030 | 4 | 993555 | IDUA | G | T | 0.429 | 0.95 | -0.0153 | 0.7493 | 0.0129 | 0.84 | 0.0132 | 0.7754 | 0.0063 | 0.9224 | 0.0432 | 0.6336 | 0.0442 | 0.7221 | -0.0338 | 0.5784 | 0.0377 | 0.5276 | 0.0192 | 0.6901 | 0.0757 | 0.4217 | 0.046 | 0.5012 |
| rs6846529 | 4 | 11023507 | CLNK | C | T | 0.283 | 1.07 | 0.014 | 0.6422 | 0.0246 | 0.5447 | -0.0061 | 0.8352 | 0.0237 | 0.5621 | -0.0493 | 0.3985 | -0.0008 | 0.9921 | 0.0053 | 0.8897 | -0.0065 | 0.8618 | 0.0057 | 0.8535 | 0.004 | 0.9441 | -0.0238 | 0.5798 |
| rs2245466 | 4 | 40197226 | RHOH | G | C | 0.343 | 1.05 | -0.0094 | 0.789 | 0.0408 | 0.3876 | -0.028 | 0.4132 | 0.0049 | 0.9175 | -0.0244 | 0.7207 | 0.0801 | 0.3874 | 0.0053 | 0.9055 | 0.0556 | 0.2013 | 0.0222 | 0.5351 | 0.0043 | 0.949 | 0.0318 | 0.5293 |
| rs112403360 | 5 | 14724304 | ANKH | A | T | 0.073 | 1.09 | -0.0111 | 0.8037 | 0.0124 | 0.8365 | -0.0127 | 0.7703 | 0.0422 | 0.4794 | 0.0099 | 0.9035 | 0.0718 | 0.5357 | -0.0083 | 0.8839 | 0.0745 | 0.1822 | 0.0463 | 0.298 | -0.1406 | 0.09635 | -0.0305 | 0.6276 |
| rs62374257 | 5 | 86927378 | COX7C | C | T | 0.23 | 1.07 | -0.0043 | 0.8906 | -0.0142 | 0.734 | 0.0157 | 0.5995 | 0.0059 | 0.8882 | -0.0732 | 0.2164 | -0.0263 | 0.7428 | 0.0355 | 0.3652 | -0.0209 | 0.5863 | 0.0176 | 0.5746 | 0.0903 | 0.1295 | 0.0531 | 0.2282 |
| rs871269 | 5 | 151052827 | TNIP1 | T | C | 0.326 | 0.96 | -0.0018 | 0.9517 | -0.016 | 0.6959 | -0.001 | 0.9717 | -0.0059 | 0.8858 | 0.0541 | 0.3602 | -0.0618 | 0.4332 | -0.0292 | 0.447 | -0.0315 | 0.4031 | -0.0182 | 0.5563 | 0.0012 | 0.9827 | -0.0039 | 0.9286 |
| rs113706587 | 5 | 180201150 | RASGEF1C | A | G | 0.11 | 1.09 | 0.0166 | 0.6054 | 0.0739 | 0.08609 | -0.0604 | 0.04963 | -0.022 | 0.6075 | -0.0451 | 0.4505 | -0.0049 | 0.9518 | -0.0031 | 0.9397 | 0.0142 | 0.7209 | -0.0306 | 0.3398 | -0.0649 | 0.275 | -0.0774 | 0.08043 |
| rs6605556 | 6 | 32615322 | HLA-DQA1 | G | A | 0.161 | 0.91 | -0.033 | 0.2832 | -0.0979 | 0.01807 | -0.0075 | 0.8004 | -0.0055 | 0.8953 | 0.0232 | 0.6942 | 0.0197 | 0.8057 | -0.0303 | 0.4365 | 0.1153 | 0.002507 | -0.025 | 0.4227 | 0.0115 | 0.8431 | -0.0436 | 0.3177 |
| rs10947943 | 6 | 41036354 | UNC5CL | A | G | 0.142 | 0.94 | 0.0305 | 0.3304 | 0.0172 | 0.6826 | 0.0224 | 0.4605 | 0.0026 | 0.9516 | 0.0686 | 0.2616 | 0.11 | 0.1772 | -0.072 | 0.06983 | 0.0317 | 0.4156 | -0.0275 | 0.3897 | 0.0216 | 0.7187 | 0.0284 | 0.5249 |
| rs143332484 | 6 | 41161469 | TREM2 | T | C | 0.013 | 1.41 | 0.0517 | 0.2478 | 0.0132 | 0.8269 | 0.0606 | 0.1629 | 0.0588 | 0.3327 | 0.0279 | 0.7483 | -0.0518 | 0.6583 | 0.0151 | 0.7876 | 0.0221 | 0.6877 | 0.0323 | 0.4758 | 0.0609 | 0.4656 | 0.0146 | 0.8179 |
| rs60755019 | 6 | 41181270 | TREML2 | G | A | 0.004 | 1.55 | 0.0285 | 0.3302 | -0.0411 | 0.2944 | -0.0201 | 0.4797 | -0.0498 | 0.2067 | -0.1009 | 0.07437 | -0.0554 | 0.4664 | -0.0172 | 0.6438 | -0.0152 | 0.6774 | -0.0266 | 0.3718 | 0.0357 | 0.5233 | 0.0683 | 0.1021 |
| rs7767350 | 6 | 47517390 | CD2AP | T | C | 0.271 | 1.08 | 0.0417 | 0.2319 | 0.0396 | 0.3971 | 0.0501 | 0.1414 | 0.0815 | 0.08497 | 0.0333 | 0.6167 | 0.1616 | 0.07308 | 0.0274 | 0.5384 | 0.006 | 0.8912 | 0.0099 | 0.7783 | -0.0735 | 0.2648 | -0.0198 | 0.6875 |
| rs785129 | 6 | 114291731 | HS3ST5 | T | C | 0.35 | 1.04 | 0.0863 | 0.05253 | 0.1302 | 0.02922 | 0.0269 | 0.5313 | 0.0275 | 0.6447 | 0.0386 | 0.6422 | 0.0358 | 0.7546 | 0.0121 | 0.829 | 0.1119 | 0.04131 | 0.0732 | 0.09909 | 0.0605 | 0.4755 | 0.1038 | 0.09734 |
| rs6943429 | 7 | 7817263 | UMAD1 | T | C | 0.42 | 1.05 | -0.1097 | 0.4225 | 0.0068 | 0.9688 | -0.0527 | 0.6893 | -0.2244 | 0.2145 | -0.3729 | 0.1278 | -0.06 | 0.8597 | 0.0354 | 0.8455 | -0.2204 | 0.2061 | -0.1379 | 0.3149 | 0.0996 | 0.6874 | 0.1544 | 0.4018 |
| rs10952097 | 7 | 8204382 | ICA1 | T | C | 0.114 | 1.07 | -0.0247 | 0.4137 | 0.0143 | 0.7239 | 0.0013 | 0.9643 | -0.0095 | 0.8148 | 0.021 | 0.7152 | -0.0814 | 0.2969 | -0.046 | 0.2271 | -0.0238 | 0.5228 | -0.0389 | 0.2019 | -0.1326 | 0.02173 | -0.0548 | 0.1998 |
| rs13237518 | 7 | 12229967 | TMEM106 | A | C | 0.411 | 0.96 | 0.001 | 0.974 | -0.0052 | 0.8998 | -0.0316 | 0.2844 | -0.0746 | 0.07072 | -0.0253 | 0.6683 | 0.0083 | 0.9166 | -0.0068 | 0.8603 | -0.0294 | 0.4365 | 0.0063 | 0.8392 | 0.0105 | 0.8571 | 0.0215 | 0.6219 |
| rs1160871 | 7 | 28129126 | JAZF1 | G | GTCTT | 0.222 | 0.95 | 0.0086 | 0.899 | 0.0992 | 0.2801 | 0.046 | 0.4834 | 0.044 | 0.6331 | 0.1205 | 0.3745 | -0.2797 | 0.1177 | 0.0387 | 0.6441 | 0.0009 | 0.9914 | 0.1274 | 0.06536 | 0.1138 | 0.3923 | 0.1304 | 0.1785 |
| rs6966331 | 7 | 37844191 | EPDR1 | T | C | 0.349 | 0.96 | -0.1194 | 0.1832 | -0.2144 | 0.07704 | 0.002 | 0.9819 | -0.024 | 0.8441 | -0.1965 | 0.2563 | -0.1762 | 0.4552 | -0.1818 | 0.1188 | -0.1435 | 0.2111 | -0.1516 | 0.09832 | 0.0122 | 0.9392 | -0.0653 | 0.6085 |
| rs76928645 | 7 | 54873635 | SEC61G | T | C | 0.103 | 0.93 | -0.0316 | 0.4493 | -0.0265 | 0.6359 | -0.0113 | 0.7791 | -0.0714 | 0.2028 | 0.1218 | 0.1249 | -0.1843 | 0.08854 | -0.0015 | 0.977 | -0.0239 | 0.6458 | 0.0067 | 0.874 | 0.0609 | 0.4398 | 0.0759 | 0.1969 |
| rs7384878 | 7 | 100334426 | SPDYE3 | C | T | 0.31 | 0.92 | -0.0574 | 0.2653 | -0.0387 | 0.5754 | 0.0238 | 0.6339 | 0.0089 | 0.8978 | 0.1315 | 0.1926 | -0.0931 | 0.4944 | -0.048 | 0.461 | 0.0818 | 0.1986 | 0.0264 | 0.6162 | -0.0644 | 0.5309 | 0.0091 | 0.9029 |
| rs11771145 | 7 | 143413669 | EPHA1 | A | G | 0.348 | 0.95 | 0.0057 | 0.8936 | 0.0134 | 0.8163 | 0.0466 | 0.2632 | 0.0736 | 0.2065 | -0.0778 | 0.3418 | 0.0483 | 0.6598 | -0.0423 | 0.435 | -0.028 | 0.5972 | -0.057 | 0.1893 | -0.0999 | 0.2131 | -0.0197 | 0.7446 |
| rs1065712 | 8 | 11844613 | CTSB | C | G | 0.053 | 1.09 | -0.0036 | 0.9208 | -0.0327 | 0.5059 | -0.0354 | 0.3178 | -0.0611 | 0.2153 | -0.1024 | 0.1376 | 0 | 0.9998 | -0.0283 | 0.5417 | -0.05 | 0.273 | -0.0897 | 0.01508 | -0.1671 | 0.01888 | -0.0757 | 0.1494 |
| rs73223431 | 8 | 27362470 | PTK2B | T | C | 0.369 | 1.07 | 0.0413 | 0.1594 | 0.0812 | 0.03937 | 0.0269 | 0.3444 | 0.028 | 0.4799 | 0.0193 | 0.7359 | 0.0173 | 0.8207 | -0.0548 | 0.1396 | 0.0298 | 0.4122 | -0.0077 | 0.7967 | -0.0026 | 0.9622 | 0.0103 | 0.8047 |
| rs11787077 | 8 | 27607795 | CLU | T | C | 0.392 | 0.91 | 0.0303 | 0.5084 | -0.0192 | 0.7553 | 0.0125 | 0.7772 | -0.0479 | 0.4374 | -0.0872 | 0.3374 | -0.0567 | 0.6343 | -0.0974 | 0.09619 | 0.0175 | 0.7605 | -0.0377 | 0.4246 | 0.1544 | 0.08182 | 0.0547 | 0.4034 |
| rs34173062 | 8 | 144103704 | SHARPIN | A | G | 0.081 | 1.13 | -0.0125 | 0.6978 | -0.0061 | 0.8885 | 0.0009 | 0.9764 | -0.0217 | 0.6205 | 0.0782 | 0.2099 | -0.137 | 0.1018 | 0.0105 | 0.7995 | 0.018 | 0.6561 | 0.0243 | 0.4599 | -0.0281 | 0.6454 | 0.0413 | 0.3687 |
| rs1800978 | 9 | 104903697 | ABCA1 | G | C | 0.13 | 1.06 | 0.007 | 0.8118 | -0.0131 | 0.7412 | 0.0105 | 0.7134 | -0.0571 | 0.1532 | -0.1107 | 0.05495 | 0.0845 | 0.2727 | 0.0233 | 0.5342 | 0.0216 | 0.5551 | -0.0133 | 0.6587 | -0.0208 | 0.7129 | -0.0028 | 0.9469 |
| rs7912495 | 10 | 11676714 | USP6NL | G | A | 0.462 | 1.06 | 0.0346 | 0.2889 | 0.0514 | 0.2419 | 0.0028 | 0.9294 | -0.0422 | 0.3403 | 0.0223 | 0.7232 | 0.0412 | 0.6282 | -0.0279 | 0.4998 | 0.0371 | 0.3599 | -0.0233 | 0.482 | -0.0339 | 0.5911 | -0.0176 | 0.7082 |
| rs7068231 | 10 | 60025170 | ANK3 | T | G | 0.403 | 0.95 | -0.0773 | 0.1694 | -0.0136 | 0.858 | -0.0599 | 0.2731 | -0.1136 | 0.1375 | -0.2083 | 0.06012 | 0.0727 | 0.6114 | -0.1764 | 0.01531 | 0.0032 | 0.9645 | -0.0704 | 0.2237 | -0.0257 | 0.8046 | -0.0074 | 0.9237 |
| rs6586028 | 10 | 80494228 | TSPAN14 | C | T | 0.196 | 0.93 | 0.0056 | 0.8774 | -0.0016 | 0.9735 | -0.0202 | 0.5619 | -0.0204 | 0.6737 | -0.0073 | 0.917 | 0.0318 | 0.7337 | -0.0428 | 0.3506 | 0.0703 | 0.1176 | -0.0079 | 0.8301 | -0.0549 | 0.4232 | -0.0583 | 0.2526 |
| rs6584063 | 10 | 96266650 | BLNK | G | A | 0.043 | 0.89 | 0.0422 | 0.1648 | 0.0261 | 0.5214 | -0.0043 | 0.8847 | -0.0399 | 0.3266 | 0.0277 | 0.6292 | 0.0381 | 0.6277 | -0.0543 | 0.1565 | -0.0379 | 0.3135 | -0.0533 | 0.08103 | 0.0613 | 0.2854 | 0.0157 | 0.7149 |
| rs7908662 | 10 | 122413396 | PLEKHA1 | G | A | 0.467 | 0.96 | -0.0296 | 0.4065 | 0.0133 | 0.7809 | -0.0107 | 0.7561 | -0.0375 | 0.432 | 0.0003 | 0.9962 | 0.0004 | 0.9964 | 0.0337 | 0.4562 | -0.0224 | 0.6132 | 0.0122 | 0.7339 | -0.0642 | 0.3369 | 0.0183 | 0.7154 |
| rs10437655 | 11 | 47370397 | SPI1 | A | G | 0.399 | 1.06 | 0.0128 | 0.6737 | 0.0253 | 0.5375 | -0.0015 | 0.9599 | -0.0563 | 0.1725 | 0.0741 | 0.2085 | -0.0474 | 0.5491 | -0.0119 | 0.7584 | 0.0099 | 0.7944 | -0.0069 | 0.8236 | 0.0081 | 0.8878 | -0.0086 | 0.8417 |
| rs1582763 | 11 | 60254475 | MS4A4A | A | G | 0.371 | 0.91 | 0.0028 | 0.9278 | -0.0055 | 0.8929 | 0.018 | 0.5438 | 0.0506 | 0.2182 | 0.0731 | 0.2067 | 0.0418 | 0.5968 | 0.012 | 0.7561 | 0.0536 | 0.1574 | 0.0313 | 0.3105 | 0.0988 | 0.08898 | 0.0121 | 0.7793 |
| rs3851179 | 11 | 86157598 | EED | T | C | 0.358 | 0.9 | 0.0399 | 0.1903 | 0.0108 | 0.7916 | -0.0181 | 0.537 | -0.0357 | 0.3838 | 0.0561 | 0.3386 | 0.0073 | 0.9264 | -0.04 | 0.2988 | 0.061 | 0.1065 | 0.0039 | 0.9007 | 0.0708 | 0.2228 | 0.0318 | 0.4604 |
| rs74685827 | 11 | 121482368 | SORL1 | G | T | 0.019 | 1.19 | 0.0313 | 0.4975 | -0.0211 | 0.7331 | -0.0212 | 0.6378 | 0.0172 | 0.7835 | 0.1608 | 0.07208 | -0.1013 | 0.3999 | 0.0687 | 0.2452 | -0.0068 | 0.9065 | -0.0313 | 0.5088 | -0.1106 | 0.2038 | -0.0654 | 0.3202 |
| rs11218343 | 11 | 121564878 | SORL1 | C | T | 0.039 | 0.84 | 0.0338 | 0.5824 | -0.0021 | 0.9799 | -0.0306 | 0.6074 | -0.0123 | 0.8807 | 0.0617 | 0.5935 | 0.0012 | 0.9938 | 0.066 | 0.3987 | 0.0745 | 0.3295 | -0.0353 | 0.5686 | -0.1097 | 0.3324 | -0.1332 | 0.1211 |
| rs6489896 | 12 | 113281983 | TPCN1 | C | T | 0.076 | 1.08 | -0.0598 | 0.06367 | -0.0359 | 0.4048 | -0.046 | 0.1422 | -0.0618 | 0.154 | -0.0478 | 0.4387 | -0.1575 | 0.05887 | -0.0361 | 0.3795 | -0.1112 | 0.005748 | -0.0685 | 0.03667 | -0.1424 | 0.01787 | -0.0653 | 0.15 |
| rs17125924 | 14 | 52924962 | FERMT2 | G | A | 0.089 | 1.1 | 0.0099 | 0.7604 | 0.01 | 0.82 | -0.0144 | 0.6481 | 0.0136 | 0.7584 | 0.0539 | 0.386 | -0.0928 | 0.2651 | 0.039 | 0.3451 | 0.0179 | 0.6588 | 0.0134 | 0.6832 | 0.0925 | 0.1298 | 0.0423 | 0.3546 |
| rs7401792 | 14 | 92464917 | SLC24A4 | G | A | 0.371 | 1.04 | 0.0937 | 0.07243 | 0.0933 | 0.1853 | 0.1152 | 0.02407 | 0.146 | 0.04075 | 0.0793 | 0.433 | -0.0982 | 0.4645 | -0.0074 | 0.9115 | -0.0396 | 0.5433 | -0.0428 | 0.418 | 0.219 | 0.02352 | 0.0262 | 0.721 |
| rs12590654 | 14 | 92472511 | SLC24A4 | A | G | 0.328 | 0.93 | 0.0283 | 0.3852 | -0.0056 | 0.8985 | -0.0151 | 0.6298 | -0.0089 | 0.8395 | 0.0594 | 0.3422 | -0.0366 | 0.6658 | 0.003 | 0.9424 | -0.0214 | 0.5939 | 0.0445 | 0.1779 | 0.1528 | 0.01473 | 0.0373 | 0.42 |
| rs7157106 | 14 | 105761758 | IGH gene c | A | G | 0.36 | 1.05 | -0.3909 | 0.396 | -0.364 | 0.5084 | -0.079 | 0.8686 | -0.085 | 0.8852 | 0.0666 | 0.9328 | -0.447 | 0.7186 | -- | -- | -0.6194 | 0.3649 | -- | | -- | | -1.5454 | 0.04472 |
| rs10131280 | 14 | 106665591 | IGH gene c | A | G | 0.133 | 0.94 | -0.0451 | 0.7829 | 0.0115 | 0.9584 | -0.1199 | 0.4502 | -0.327 | 0.1379 | 0.0454 | 0.8912 | 0.0561 | 0.8914 | -0.4017 | 0.05683 | -0.1644 | 0.4276 | -0.2479 | 0.1539 | 0.1198 | 0.6981 | 0.1007 | 0.6684 |
| rs8025980 | 15 | 50701814 | SPPL2A | G | A | 0.345 | 0.96 | -0.0487 | 0.6141 | -0.1209 | 0.3549 | 0.1924 | 0.03951 | 0.1729 | 0.1787 | 0.1788 | 0.3188 | 0.0533 | 0.8265 | 0.0756 | 0.5466 | 0.2714 | 0.02557 | 0.18 | 0.06252 | -0.151 | 0.4056 | -0.0328 | 0.808 |
| rs602602 | 15 | 58764824 | MINDY2 | A | T | 0.28 | 0.94 | 0.0168 | 0.6293 | 0.0291 | 0.5336 | 0.0871 | 0.01011 | 0.1593 | 0.000671 | 0.1707 | 0.009367 | -0.0994 | 0.2729 | -0.0106 | 0.8113 | 0.0025 | 0.9539 | 0.0018 | 0.9595 | -0.0201 | 0.7584 | -0.0264 | 0.5954 |
| rs117618017 | 15 | 63277703 | APH1B | T | C | 0.144 | 1.11 | -0.1276 | 0.02856 | -0.1063 | 0.1784 | -0.0979 | 0.08313 | -0.0942 | 0.2306 | -0.273 | 0.01127 | 0.043 | 0.7757 | 0.0302 | 0.6851 | -0.256 | 0.000452 | -0.0704 | 0.2275 | -0.1162 | 0.3094 | 0.0003 | 0.9972 |
| rs3848143 | 15 | 64131307 | SNX1 | G | A | 0.22 | 1.05 | -0.0825 | 0.2874 | -0.0366 | 0.727 | 0.0151 | 0.8396 | -0.0966 | 0.3638 | 0.2279 | 0.1276 | 0.0904 | 0.6664 | -0.0909 | 0.353 | -0.0303 | 0.7538 | -0.0007 | 0.9933 | 0.1349 | 0.3633 | -0.0601 | 0.6009 |
| rs12592898 | 15 | 78936857 | CTSH | A | G | 0.133 | 0.94 | 0.009 | 0.8073 | -0.0619 | 0.2145 | -0.0782 | 0.03006 | -0.0988 | 0.04825 | -0.1554 | 0.03117 | 0.1225 | 0.2064 | -0.028 | 0.5532 | 0.0546 | 0.237 | -0.0196 | 0.6035 | -0.0732 | 0.3012 | -0.0146 | 0.7836 |
| rs1140239 | 16 | 30010081 | DOC2A | T | C | 0.379 | 0.94 | -0.0601 | 0.1385 | -0.0399 | 0.4673 | -0.0429 | 0.2735 | -0.056 | 0.3075 | -0.1671 | 0.03874 | -0.0408 | 0.7047 | -0.0385 | 0.4546 | -0.1164 | 0.02061 | -0.1339 | 0.001357 | -0.0765 | 0.3065 | -0.0232 | 0.6851 |
| rs889555 | 16 | 31111250 | BCKDK | T | C | 0.281 | 0.95 | 0.0743 | 0.01301 | 0.0347 | 0.3877 | 0.0157 | 0.59 | -0.0061 | 0.8803 | -0.0033 | 0.9533 | -0.0296 | 0.703 | 0.0147 | 0.7013 | 0.0687 | 0.06747 | 0.0436 | 0.1528 | 0.0778 | 0.171 | 0.0281 | 0.509 |
| rs4985556 | 16 | 70660097 | IL34 | A | C | 0.115 | 1.07 | -0.0239 | 0.5287 | -0.0505 | 0.3234 | 0.0035 | 0.9241 | 0.0018 | 0.9717 | -0.141 | 0.05432 | -0.0114 | 0.9079 | -0.0479 | 0.3191 | -0.076 | 0.1064 | -0.0642 | 0.09551 | 0.0056 | 0.9374 | -0.048 | 0.3699 |
| rs450674 | 16 | 79574511 | MAF | C | T | 0.373 | 0.96 | 0.0831 | 0.02649 | 0.1081 | 0.0309 | -0.0485 | 0.181 | 0.0154 | 0.7597 | 0.0943 | 0.1878 | 0.0476 | 0.6229 | -0.0327 | 0.4856 | 0.0137 | 0.7665 | -0.0029 | 0.9385 | -0.0734 | 0.3153 | -0.0053 | 0.9193 |
| rs12446759 | 16 | 81739398 | PLCG2 | G | A | 0.403 | 0.95 | 0.0372 | 0.2573 | 0.0635 | 0.1473 | -0.0179 | 0.5735 | 0.0356 | 0.4177 | 0.0205 | 0.7421 | 0.0206 | 0.8066 | 0.0597 | 0.1499 | 0.0206 | 0.6127 | 0.0411 | 0.2145 | 0.0937 | 0.1303 | 0.0648 | 0.1604 |
| rs16941239 | 16 | 86420604 | FOXF1 | A | T | 0.029 | 1.13 | -0.0164 | 0.5844 | 0.0423 | 0.2937 | -0.0757 | 0.009253 | -0.0498 | 0.2195 | -0.0158 | 0.7845 | -0.1326 | 0.0888 | 0.0334 | 0.3776 | -0.0548 | 0.1398 | -0.0019 | 0.9511 | 0.0174 | 0.7613 | 0.0211 | 0.6199 |
| rs56407236 | 16 | 90103687 | PRDM7 | A | G | 0.069 | 1.11 | -0.0058 | 0.851 | -0.0175 | 0.6741 | 0.0418 | 0.1652 | 0.0993 | 0.01793 | 0.0549 | 0.3571 | -0.1057 | 0.193 | -0.0053 | 0.8928 | -0.0058 | 0.8795 | -0.0235 | 0.4559 | 0.0362 | 0.5413 | -0.0404 | 0.3639 |
| rs35048651 | 17 | 1728046 | WDR81 | T | TGAG | 0.214 | 1.06 | -0.0078 | 0.7945 | -0.0348 | 0.3872 | -0.0281 | 0.3372 | -0.0349 | 0.3886 | -0.095 | 0.1013 | 0.0543 | 0.4874 | 0.0258 | 0.4984 | -0.0226 | 0.5449 | -0.0112 | 0.7141 | -0.0261 | 0.644 | -0.0682 | 0.1103 |
| rs7225151 | 17 | 5233752 | SCIMP | A | G | 0.124 | 1.08 | 0.0415 | 0.3201 | 0.0388 | 0.4881 | 0.0363 | 0.3735 | 0.0126 | 0.825 | -0.1166 | 0.1966 | -0.0934 | 0.4038 | -0.0612 | 0.2445 | 0.0284 | 0.5817 | 0.0119 | 0.7891 | 0.0677 | 0.4158 | -0.0254 | 0.6791 |
| rs2242595 | 17 | 18156140 | MYO15A | A | G | 0.112 | 0.94 | 0.0031 | 0.9454 | -0.0304 | 0.6137 | -0.0124 | 0.7758 | -0.0342 | 0.5719 | -0.0073 | 0.9322 | -0.0508 | 0.6622 | -0.0666 | 0.2418 | -0.0493 | 0.3758 | -0.0156 | 0.7317 | -0.0348 | 0.6753 | 0.0292 | 0.6448 |
| rs5848 | 17 | 44352876 | GRN | T | C | 0.289 | 1.07 | 0.0075 | 0.8592 | -0.0123 | 0.8283 | 0.0137 | 0.7392 | 0.017 | 0.7656 | -0.037 | 0.6494 | 0.0761 | 0.4904 | -0.0424 | 0.4282 | 0.0216 | 0.681 | -0.0065 | 0.8797 | -0.1603 | 0.0516 | -0.0167 | 0.7831 |
| rs199515 | 17 | 46779275 | WNT3 | G | C | 0.219 | 0.94 | 0.0459 | 0.1293 | 0.0417 | 0.3052 | 0.0603 | 0.0401 | 0.0975 | 0.01663 | 0.0025 | 0.966 | -0.0005 | 0.9953 | -0.047 | 0.2211 | 0.0081 | 0.8286 | -0.0036 | 0.9054 | -0.0762 | 0.1927 | -0.0112 | 0.796 |
| rs616338 | 17 | 49219935 | ABI3 | T | C | 0.012 | 1.32 | 0.0037 | 0.9096 | -0.0091 | 0.837 | 0.0002 | 0.9948 | 0.0163 | 0.7128 | 0.0334 | 0.5983 | 0.0768 | 0.3644 | -0.0575 | 0.1615 | 0.0247 | 0.5399 | -0.0385 | 0.2459 | -0.0322 | 0.5991 | 0.0002 | 0.9971 |
| rs2526377 | 17 | 58332680 | TSPOAP1 | G | A | 0.445 | 0.95 | -0.0231 | 0.4516 | -0.0229 | 0.5776 | 0.0459 | 0.1224 | 0.0767 | 0.06329 | 0.0974 | 0.09499 | 0.1524 | 0.05436 | 0.0287 | 0.4619 | 0.0266 | 0.4847 | 0.0433 | 0.1619 | -0.0162 | 0.7832 | 0.006 | 0.8904 |
| rs4277405 | 17 | 63471557 | ACE | C | T | 0.384 | 0.94 | 0.0422 | 0.6954 | 0.0098 | 0.9469 | -0.0306 | 0.7713 | 0.0345 | 0.8124 | -0.2169 | 0.2791 | 0.1035 | 0.7115 | 0.1331 | 0.3345 | 0.1229 | 0.3609 | 0.111 | 0.3069 | -0.0722 | 0.7289 | 0.0358 | 0.8117 |
| rs12151021 | 19 | 1050875 | ABCA7 | A | G | 0.336 | 1.1 | -0.0437 | 0.3675 | -0.0207 | 0.7501 | -0.0553 | 0.2438 | 0.0173 | 0.7925 | 0.1865 | 0.04388 | 0.0042 | 0.9742 | 0.068 | 0.2711 | -0.0993 | 0.09981 | -0.0048 | 0.922 | 0.1036 | 0.2646 | -0.0028 | 0.9682 |
| rs149080927 | 19 | 1854254 | KLF16 | G | GC | 0.48 | 1.05 | 0.0525 | 0.1078 | 0.0318 | 0.466 | -0.0595 | 0.06094 | -0.0092 | 0.8353 | -0.0497 | 0.4288 | 0.005 | 0.9528 | 0.0131 | 0.751 | 0.0579 | 0.1542 | -0.0021 | 0.9486 | 0.0096 | 0.8758 | -0.0396 | 0.396 |
| rs9304690 | 19 | 49950060 | SIGLEC11 | T | C | 0.24 | 1.05 | -0.0127 | 0.6811 | -0.0463 | 0.264 | -0.0147 | 0.6258 | -0.047 | 0.261 | -0.0862 | 0.1504 | 0.0905 | 0.2629 | 0.0701 | 0.07541 | 0.0728 | 0.05894 | 0.0668 | 0.03446 | -0.0172 | 0.7732 | -0.0064 | 0.8851 |
| rs587709 | 19 | 54267597 | LILRB2 | C | T | 0.325 | 1.05 | -0.0185 | 0.5257 | 0.0044 | 0.9099 | 0.0285 | 0.3108 | -0.008 | 0.8396 | -0.0421 | 0.4542 | 0.0822 | 0.2783 | -0.0058 | 0.8755 | 0.018 | 0.6195 | -0.0078 | 0.7907 | 0.0188 | 0.7282 | 0.0095 | 0.8195 |
| rs1358782 | 20 | 413334 | RBCK1 | A | G | 0.246 | 0.95 | -0.0837 | 0.005194 | -0.0744 | 0.0648 | 0.0136 | 0.641 | 0.0295 | 0.4639 | 0.0003 | 0.9961 | 0.0391 | 0.6096 | -0.0183 | 0.6304 | -0.0482 | 0.1959 | -0.0162 | 0.5927 | -0.1091 | 0.05113 | -0.019 | 0.6498 |

| | | | | | | | | | | | | | | | | | | | | | | | | | | | | | |
|---|---|---|---|---|---|---|---|---|---|---|---|---|---|---|---|---|---|---|---|---|---|---|---|---|---|---|---|---|---|
| rs6014724 | 20 | 56423488 | CASS4 | G | A | 0.09 | 0.89 | 0.0042 | 0.8894 | 0.0274 | 0.5011 | -0.0101 | 0.735 | 0.0252 | 0.541 | -0.0023 | 0.9692 | -0.0789 | 0.3243 | 0.044 | 0.2564 | -0.0639 | 0.09314 | -0.0346 | 0.2671 | -0.0505 | 0.386 | -0.0228 | 0.6016 |
| rs6742 | 20 | 63743088 | SLC2A4RG | T | C | 0.221 | 0.95 | 0.0354 | 0.2667 | 0.0667 | 0.1182 | 0.0011 | 0.9706 | -0.0065 | 0.8802 | 0.0281 | 0.6469 | -0.1453 | 0.07828 | -0.028 | 0.4874 | -0.0081 | 0.8371 | -0.0096 | 0.7662 | 0.0033 | 0.956 | 0.0523 | 0.2479 |
| rs2154481 | 21 | 26101558 | APP | C | T | 0.476 | 0.95 | 0.0087 | 0.7891 | 0.0273 | 0.5341 | 0.017 | 0.5899 | -0.0204 | 0.6442 | -0.1023 | 0.1029 | 0.0701 | 0.4086 | 0.0608 | 0.1388 | 0.0251 | 0.534 | 0.0587 | 0.07597 | 0.0195 | 0.7527 | 0.0222 | 0.6317 |
| rs2830489 | 21 | 26775872 | ADAMTS1 | T | C | 0.281 | 0.95 | 0.0308 | 0.3664 | 0.0479 | 0.291 | 0.036 | 0.275 | 0.0083 | 0.8559 | 0.0643 | 0.3253 | 0.1066 | 0.2314 | -0.0204 | 0.6361 | -0.0668 | 0.1151 | 0.0104 | 0.7642 | 0.0634 | 0.3364 | 0.0468 | 0.3366 |

**Table S6: Association of Nalls et al PD risk loci with lewy body pathology**

| SNP | CHR | POS_NEW | GENE | Effect allel | Referent | Beta | NFT Braak | | NFT Braak (3 cat) | | NFT Braak (any/none) | |
|---|---|---|---|---|---|---|---|---|---|---|---|---|
| | | | | | | | Beta | Pvalue | Beta | Pvalue | Beta | Pvalue |
| rs114138760 | 1 | 154925709 | PMVK | c | g | 0.2812 | -0.2847 | 0.1026 | -0.2466 | 0.1654 | -0.1579 | 0.3762 |
| rs35749011 | 1 | 155162560 | KRTCAP2 | a | g | 0.6068 | -0.6556 | 2.99E-07 | -0.594 | 7.49E-06 | -0.5944 | 1.48E-05 |
| rs35643925 | 1 | 156185069 | SEMA4A | a | g | 0.389 | -0.3111 | 0.007467 | -0.2137 | 0.07355 | -0.1999 | 0.1004 |
| rs6658353 | 1 | 161499264 | FCGR2A | c | g | 0.065 | 0.0413 | 0.1685 | 0.0497 | 0.1035 | 0.0443 | 0.1529 |
| rs11578699 | 1 | 171750629 | VAMP4 | t | c | -0.0704 | -0.0436 | 0.2416 | -0.0374 | 0.3243 | -0.0371 | 0.3368 |
| rs823118 | 1 | 205754444 | NUCKS1 | t | c | 0.1066 | -0.0328 | 0.2699 | -0.0318 | 0.293 | -0.0348 | 0.2579 |
| rs11557080 | 1 | 205768611 | RAB29 | a | g | 0.1315 | 0.048 | 0.2733 | 0.0524 | 0.2386 | 0.0492 | 0.2729 |
| rs4653767 | 1 | 226728377 | ITPKB | t | c | 0.0833 | -0.0411 | 0.2041 | -0.0401 | 0.2248 | -0.0422 | 0.2078 |
| rs10797576 | 1 | 232528865 | SIPA1L2 | t | c | 0.1114 | -0.0291 | 0.5029 | -0.0316 | 0.4747 | -0.026 | 0.5643 |
| rs76116224 | 2 | 17966582 | KCNS3 | a | t | 0.1104 | 0.1177 | 0.02444 | 0.1147 | 0.03193 | 0.1051 | 0.05558 |
| rs2042477 | 2 | 95335195 | KCNIP3 | a | t | -0.0657 | -0.0186 | 0.5996 | -0.0092 | 0.799 | -0.0088 | 0.8097 |
| rs11683001 | 2 | 101780501 | MAP4K4 | a | t | 0.0705 | 0.0206 | 0.5139 | 0.0275 | 0.3912 | 0.0259 | 0.427 |
| rs4954162 | 2 | 134681219 | TMEM163 | a | g | -0.0746 | 0.0819 | 0.04668 | 0.0784 | 0.06275 | 0.0895 | 0.03413 |
| rs57891859 | 2 | 134707046 | TMEM163 | a | g | 0.0807 | -0.0332 | 0.3113 | -0.0245 | 0.4646 | -0.022 | 0.5186 |
| rs1474055 | 2 | 168253884 | STK39 | t | c | 0.1796 | 0.0851 | 0.05916 | 0.0844 | 0.06607 | 0.063 | 0.1751 |
| rs73038319 | 3 | 18320267 | SATB1 | a | c | -0.1693 | 0.1679 | 0.02903 | 0.1876 | 0.0188 | 0.1685 | 0.03874 |
| rs6808178 | 3 | 28664199 | LINC00693 | t | c | 0.0658 | 0.0638 | 0.03632 | 0.0616 | 0.04727 | 0.0572 | 0.06929 |
| rs12497850 | 3 | 48711556 | IP6K2 | t | g | 0.0636 | -0.0305 | 0.3273 | -0.0421 | 0.1832 | -0.0453 | 0.1579 |
| rs55961674 | 3 | 122478045 | KPNA1 | t | c | 0.0861 | -0.0213 | 0.5936 | -0.0231 | 0.571 | -0.0215 | 0.6043 |
| rs11707416 | 3 | 151391177 | MED12L | a | t | -0.0627 | 0.0084 | 0.7859 | 0.0035 | 0.9108 | 0.0041 | 0.8992 |
| rs1450522 | 3 | 161359842 | SPTSSB | a | g | -0.0616 | 0.0129 | 0.6798 | 0.0142 | 0.6562 | 0.0173 | 0.5938 |
| rs10513789 | 3 | 183042285 | MCCC1 | t | g | 0.1485 | 0.0041 | 0.9133 | -0.0024 | 0.9496 | -0.0153 | 0.6912 |
| rs873786 | 4 | 931588 | GAK | t | c | -0.1731 | 0.0499 | 0.3045 | 0.0368 | 0.4572 | 0.0511 | 0.307 |
| rs34311866 | 4 | 958159 | TMEM175 | t | c | -0.2126 | 0.1279 | 0.000614 | 0.1204 | 0.001543 | 0.1163 | 0.002959 |
| rs4698412 | 4 | 15735725 | BST1 | a | g | 0.1035 | -0.0082 | 0.7821 | -0.0108 | 0.719 | -0.0102 | 0.74 |
| rs34025766 | 4 | 17967188 | LCORL | a | t | -0.0839 | -0.0153 | 0.6984 | -0.0261 | 0.5171 | -0.0145 | 0.7232 |
| rs6825004 | 4 | 76189212 | SCARB2 | c | g | 0.0622 | -0.0928 | 0.003801 | -0.0733 | 0.02505 | -0.0692 | 0.0364 |
| rs4101061 | 4 | 76226816 | FAM47E | a | g | -0.0912 | 0.055 | 0.08819 | 0.0568 | 0.08484 | 0.0602 | 0.07297 |
| rs6854006 | 4 | 76276901 | FAM47E-STBD1 | t | c | -0.0912 | 0.0266 | 0.3812 | 0.0266 | 0.3911 | 0.0231 | 0.4623 |
| rs356228 | 4 | 89685975 | SNCA | c | g | 0.1503 | 0.055 | 0.07331 | 0.0504 | 0.1068 | 0.0477 | 0.1329 |
| rs356182 | 4 | 89704960 | SNCA | a | g | -0.2774 | -0.0475 | 0.1391 | -0.0413 | 0.2046 | -0.0368 | 0.265 |
| rs5019538 | 4 | 89715479 | SNCA | a | g | -0.1565 | -0.0958 | 0.003919 | -0.0971 | 0.003865 | -0.0959 | 0.004742 |

| | | | | | | | | | | | | |
|---|---|---|---|---|---|---|---|---|---|---|---|---|
| rs356203 | 4 | 89744890 | SNCA | t | c | -0.2504 | -0.0299 | 0.3405 | -0.0276 | 0.3847 | -0.0209 | 0.5168 |
| rs13117519 | 4 | 113447909 | CAMK2D | t | c | 0.0875 | 0.0632 | 0.1155 | 0.0591 | 0.1488 | 0.0633 | 0.1261 |
| rs62333164 | 4 | 169662006 | CLCN3 | a | g | -0.0638 | 0.0487 | 0.1189 | 0.0525 | 0.09945 | 0.0618 | 0.05526 |
| rs1867598 | 5 | 60842132 | ELOVL7 | a | g | -0.1554 | 0.0525 | 0.2966 | 0.056 | 0.2775 | 0.0612 | 0.2437 |
| rs26431 | 5 | 103030090 | PAM | c | g | 0.0621 | -0.0186 | 0.5671 | -0.017 | 0.6062 | -0.0208 | 0.5354 |
| rs6875262 | 5 | 124774580 | ZNF608 | t | c | 0.1141 | 0.0793 | 0.144 | 0.0792 | 0.1524 | 0.0805 | 0.1466 |
| rs11950533 | 5 | 134863415 | C5orf24 | a | c | -0.0916 | -0.0094 | 0.848 | -0.0156 | 0.7557 | -0.0063 | 0.9006 |
| rs4140646 | 6 | 27771022 | LOC100131289 | a | g | 0.0833 | 0.0602 | 0.122 | 0.0653 | 0.09913 | 0.0773 | 0.05259 |
| rs9261484 | 6 | 30140906 | TRIM40 | t | c | -0.0635 | -0.0042 | 0.906 | 0.0071 | 0.8444 | 0.0085 | 0.8161 |
| rs9267659 | 6 | 31878457 | SLC44A4 | a | g | 0.079 | 0.0256 | 0.5134 | 0.032 | 0.4209 | 0.0177 | 0.6633 |
| rs112485576 | 6 | 32610995 | HLA-DRB5 | a | c | -0.1676 | 0.0337 | 0.4239 | 0.0415 | 0.3337 | 0.0376 | 0.3856 |
| rs12528068 | 6 | 71778059 | RIMS1 | t | c | 0.0657 | 0.0415 | 0.2074 | 0.0476 | 0.1555 | 0.0385 | 0.2575 |
| rs997368 | 6 | 111922088 | FYN | a | g | 0.0714 | -0.0506 | 0.1739 | -0.0637 | 0.0921 | -0.0598 | 0.1182 |
| rs75859381 | 6 | 132889222 | RPS12 | t | c | -0.2207 | 0.1044 | 0.2394 | 0.0953 | 0.2937 | 0.0097 | 0.9154 |
| rs199351 | 7 | 23260430 | GPNMB | a | c | 0.1016 | -0.0178 | 0.5493 | -0.0099 | 0.7443 | -0.0076 | 0.8054 |
| rs76949143 | 7 | 66544864 | GS1-124K5.11 | a | t | -0.1432 | -0.0985 | 0.1515 | -0.0928 | 0.186 | -0.0887 | 0.2197 |
| rs1293298 | 8 | 11854934 | CTSB | a | c | 0.093 | -0.0491 | 0.1479 | -0.0408 | 0.239 | -0.0356 | 0.3117 |
| rs620513 | 8 | 16840084 | FGF20 | t | g | -0.0856 | -0.0458 | 0.1722 | -0.0518 | 0.1298 | -0.0468 | 0.1797 |
| rs2280104 | 8 | 22668467 | BIN3 | t | c | 0.0556 | -0.0152 | 0.6208 | -0.0146 | 0.6421 | -0.0156 | 0.6262 |
| rs2086641 | 8 | 129889663 | FAM49B | t | c | -0.0605 | 0.0322 | 0.3302 | 0.0287 | 0.3957 | 0.0156 | 0.6497 |
| rs13294100 | 9 | 17579692 | SH3GL2 | t | g | -0.0859 | 0.0228 | 0.4637 | 0.0164 | 0.6043 | 0.0121 | 0.7057 |
| rs10756907 | 9 | 17727067 | SH3GL2 | a | g | -0.0926 | -0.0109 | 0.7573 | -0.0191 | 0.5948 | -0.0206 | 0.5709 |
| rs6476434 | 9 | 34046393 | UBAP2 | t | c | -0.0615 | 0.0251 | 0.4636 | 0.0161 | 0.6448 | 0.0132 | 0.7099 |
| rs896435 | 10 | 15515407 | ITGA8 | t | c | 0.0735 | -0.0256 | 0.4251 | -0.0249 | 0.4462 | -0.0206 | 0.536 |
| rs10748818 | 10 | 102255522 | GBF1 | a | g | -0.079 | -0.0212 | 0.6135 | -0.0163 | 0.7025 | -0.0137 | 0.7528 |
| rs72840788 | 10 | 119656173 | BAG3 | a | g | 0.0763 | -0.0092 | 0.8 | -0.0179 | 0.6294 | -0.0101 | 0.7892 |
| rs117896735 | 10 | 119776815 | INPP5F | a | g | 0.4354 | -0.3087 | 0.00846 | -0.2949 | 0.01496 | -0.1889 | 0.1251 |
| rs7938782 | 11 | 10537230 | RNF141 | a | g | 0.087 | 0.0345 | 0.4325 | 0.0345 | 0.44 | 0.025 | 0.5862 |
| rs12283611 | 11 | 83776234 | DLG2 | a | c | -0.0645 | 0.0197 | 0.5145 | 0.0125 | 0.6844 | 0.0055 | 0.8598 |
| rs3802920 | 11 | 133917106 | IGSF9B | t | g | 0.1073 | 0.0335 | 0.3629 | 0.0549 | 0.143 | 0.0654 | 0.08282 |
| rs76904798 | 12 | 40220632 | LRRK2 | t | c | 0.1439 | 0.0234 | 0.5874 | 0.0259 | 0.5574 | 0.0238 | 0.5917 |
| rs117073808 | 12 | 40528770 | LRRK2 | t | c | -0.3955 | 0.0828 | 0.4443 | 0.0497 | 0.6544 | 0.0043 | 0.9688 |
| rs141128804 | 12 | 40542837 | CNTN1 | t | c | 0.3468 | -0.1843 | 0.1963 | -0.1121 | 0.4462 | -0.1099 | 0.4615 |
| rs138017112 | 12 | 41901977 | GXYLT1 | c | g | 0.413 | 0.0596 | 0.6849 | 0.0763 | 0.6109 | 0.1198 | 0.422 |
| rs7134559 | 12 | 46025303 | SCAF11 | t | c | -0.0539 | -0.0001 | 0.9985 | -0.0029 | 0.9235 | -0.009 | 0.7727 |
| rs10847864 | 12 | 122842051 | HIP1R | t | g | 0.1478 | -0.0205 | 0.5325 | -0.0168 | 0.6149 | -0.0049 | 0.8862 |

| | | | | | | | | | | | | |
|---|---|---|---|---|---|---|---|---|---|---|---|---|
| rs11610045 | 12 | 132487182 | FBRSL1 | a | g | 0.0601 | 0.0254 | 0.3959 | 0.0353 | 0.2465 | 0.0281 | 0.3636 |
| rs9568188 | 13 | 49353596 | CAB39L | t | c | 0.0617 | -0.0439 | 0.1898 | -0.0374 | 0.2736 | -0.0495 | 0.1519 |
| rs4771268 | 13 | 97212767 | MBNL2 | t | c | 0.0675 | 0.0193 | 0.5835 | 0.0224 | 0.5325 | 0.0142 | 0.6966 |
| rs12147950 | 14 | 37520065 | MIPOL1 | t | c | -0.0529 | -0.0206 | 0.4916 | -0.0211 | 0.4893 | -0.0309 | 0.32 |
| rs11158026 | 14 | 54882151 | GCH1 | t | c | -0.0842 | -0.0093 | 0.7678 | -0.0083 | 0.7955 | -0.013 | 0.689 |
| rs3742785 | 14 | 74906331 | RPS6KL1 | a | c | 0.0707 | 0.0248 | 0.4876 | 0.0215 | 0.5547 | 0.0164 | 0.6582 |
| rs979812 | 14 | 87997920 | GALC | t | g | 0.061 | -0.0391 | 0.1838 | -0.0403 | 0.1797 | -0.0435 | 0.1547 |
| rs2251086 | 15 | 61705186 | VPS13C | t | c | -0.1186 | -0.0497 | 0.2326 | -0.0415 | 0.3269 | -0.0323 | 0.4532 |
| rs6497339 | 16 | 19266171 | SYT17 | a | t | 0.063 | -0.0121 | 0.6881 | -0.0126 | 0.6815 | -0.0092 | 0.7684 |
| rs2904880 | 16 | 28933075 | CD19 | c | g | -0.065 | -0.0052 | 0.8735 | -0.0021 | 0.95 | -0.0001 | 0.9971 |
| rs11150601 | 16 | 30966478 | SETD1A | a | g | 0.0907 | -0.0502 | 0.1024 | -0.0446 | 0.1559 | -0.0512 | 0.1081 |
| rs6500328 | 16 | 50702745 | NOD2 | a | g | 0.0586 | 0.0179 | 0.5493 | 0.0091 | 0.7645 | 0.0018 | 0.9531 |
| rs3104783 | 16 | 52602330 | CASC16 | a | c | 0.0668 | 0.0477 | 0.1127 | 0.0472 | 0.1219 | 0.0372 | 0.2307 |
| rs10221156 | 16 | 52935514 | CHD9 | a | g | -0.1156 | 0.0852 | 0.09052 | 0.0697 | 0.1722 | 0.0552 | 0.2885 |
| rs12600861 | 17 | 7452302 | CHRNB1 | a | c | -0.0565 | 0.0171 | 0.5833 | 0.0142 | 0.6555 | 0.0123 | 0.7029 |
| rs12951632 | 17 | 42588995 | RETREG3 | t | c | 0.0642 | -0.0123 | 0.7155 | -0.0096 | 0.778 | -0.0191 | 0.5825 |
| rs2269906 | 17 | 44216969 | UBTF | a | c | 0.0631 | -0.0191 | 0.5355 | -0.0181 | 0.5663 | -0.0168 | 0.5991 |
| rs850738 | 17 | 44357262 | FAM171A2 | a | g | -0.071 | 0.0436 | 0.1522 | 0.0366 | 0.2382 | 0.033 | 0.2955 |
| rs17686238 | 17 | 45339907 | MAP3K14 | t | g | -0.1896 | -0.0036 | 0.9415 | -0.0207 | 0.6775 | -0.0068 | 0.8917 |
| rs62053943 | 17 | 45666837 | CRHR1 | t | c | -0.27 | 0.0138 | 0.7505 | 0.0017 | 0.9698 | 0.0034 | 0.9394 |
| rs9912362 | 17 | 45706862 | CRHR1 | t | c | 0.077 | -0.0125 | 0.6829 | -0.0081 | 0.796 | -0.0017 | 0.9577 |
| rs117615688 | 17 | 45720942 | CRHR1 | a | g | -0.2324 | -0.0311 | 0.5997 | -0.0599 | 0.3247 | -0.0611 | 0.3275 |
| rs7221167 | 17 | 45855941 | MAPT-AS1 | t | c | 0.1443 | -0.0039 | 0.8982 | 0.0044 | 0.8868 | 0.0102 | 0.7466 |
| rs7225002 | 17 | 46111701 | KANSL1 | a | g | 0.1351 | 0.0105 | 0.7284 | 0.0226 | 0.4615 | 0.0215 | 0.4912 |
| rs199453 | 17 | 46723580 | NSF | t | c | -0.2045 | 0.0096 | 0.7859 | -0.0012 | 0.9723 | -0.0047 | 0.8972 |
| rs11658976 | 17 | 46789439 | WNT3 | a | g | -0.0624 | 0.0159 | 0.6074 | 0.0013 | 0.9661 | -0.0001 | 0.9977 |
| rs61169879 | 17 | 61840005 | BRIP1 | t | c | 0.082 | 0.0232 | 0.5649 | 0.0234 | 0.5695 | 0.0108 | 0.7965 |
| rs666463 | 17 | 78429399 | DNAH17 | a | t | 0.076 | -0.0267 | 0.5033 | -0.0187 | 0.6462 | -0.0233 | 0.5716 |
| rs1941685 | 18 | 33724354 | ASXL3 | t | g | 0.0531 | -0.0225 | 0.445 | -0.0256 | 0.3934 | -0.03 | 0.3245 |
| rs12456492 | 18 | 43093415 | RIT2 | a | g | -0.0983 | 0.0679 | 0.03117 | 0.0651 | 0.0425 | 0.0694 | 0.03428 |
| rs8087969 | 18 | 51157219 | MEX3C | t | g | -0.0578 | -0.031 | 0.3026 | -0.0201 | 0.5102 | -0.0315 | 0.3128 |
| rs55818311 | 19 | 2341049 | SPPL2B | t | c | -0.0696 | 0.0245 | 0.4505 | 0.0219 | 0.509 | 0.0212 | 0.5315 |
| rs2295545 | 20 | 3184040 | DDRGK1 | t | c | 0.0622 | -0.0042 | 0.8933 | -0.0113 | 0.7202 | -0.022 | 0.493 |
| rs77351827 | 20 | 6025395 | CRLS1 | t | c | 0.0802 | 0.0037 | 0.9374 | -0.0008 | 0.9864 | 0.0058 | 0.9029 |
| rs2248244 | 21 | 37480059 | DYRK1A | a | g | 0.0714 | 0.0083 | 0.804 | 0.0143 | 0.6754 | 0.0235 | 0.4968 |

**Table S7: Association of Nalls et al PD risk loci with AD hallmark pathologies**

| Nalls et al SNPs | | | | | | | A Score | | B Score | | CERAD (C Score) | | ADNC Score | |
|---|---|---|---|---|---|---|---|---|---|---|---|---|---|---|
| SNP | CHR | POS_NEW | GENE | Effect allel | Referent | Beta | Beta | Pval | Beta | Pval | Beta | Pval | Beta | Pval |
| rs1141387 | 1 | 1.55E+08 | PMVK | c | g | 0.2812 | 0.5722 | 0.04614 | -0.0688 | 0.6757 | 0.0622 | 0.6949 | 0.4056 | 0.1572 |
| rs3574901 | 1 | 1.55E+08 | KRTCAP2 | a | g | 0.6068 | 0.5187 | 0.04849 | 0.3225 | 0.00721 | 0.2756 | 0.0234 | 0.5746 | 0.02111 |
| rs3564392 | 1 | 1.56E+08 | SEMA4A | a | g | 0.389 | 0.1718 | 0.4731 | 0.2309 | 0.03107 | 0.3265 | 0.001731 | 0.293 | 0.1785 |
| rs6658353 | 1 | 1.61E+08 | FCGR2A | c | g | 0.065 | 0.0056 | 0.9155 | 0.0118 | 0.6753 | -0.0138 | 0.6238 | 0.0389 | 0.4179 |
| rs1157869 | 1 | 1.72E+08 | VAMP4 | t | c | -0.0704 | -0.063 | 0.336 | -0.0277 | 0.4294 | -0.0246 | 0.4813 | -0.0225 | 0.7024 |
| rs823118 | 1 | 2.06E+08 | NUCKS1 | t | c | 0.1066 | -0.0627 | 0.2345 | -0.043 | 0.1237 | -0.0329 | 0.237 | -0.0601 | 0.2072 |
| rs1155708 | 1 | 2.06E+08 | RAB29 | a | g | 0.1315 | 0.0402 | 0.5941 | 0.008 | 0.8438 | 0.0142 | 0.7254 | 0.0435 | 0.5243 |
| rs4653767 | 1 | 2.27E+08 | ITPKB | t | c | 0.0833 | -0.0164 | 0.7752 | -0.0527 | 0.08401 | -0.0196 | 0.5181 | -0.017 | 0.7432 |
| rs1079757 | 1 | 2.33E+08 | SIPA1L2 | t | c | 0.1114 | -0.0353 | 0.6469 | -0.0066 | 0.8729 | -0.0055 | 0.894 | 0.0193 | 0.7781 |
| rs7611622 | 2 | 17966582 | KCNS3 | a | t | 0.1104 | 0.0349 | 0.718 | 0.0539 | 0.2805 | 0.103 | 0.03768 | 0.0141 | 0.8718 |
| rs2042477 | 2 | 95335195 | KCNIP3 | a | t | -0.0657 | 0.0215 | 0.7293 | 0.0416 | 0.208 | 0.0775 | 0.01828 | 0.0003 | 0.9951 |
| rs1168300 | 2 | 1.02E+08 | MAP4K4 | a | t | 0.0705 | -0.048 | 0.3943 | 0.0291 | 0.3233 | 0.022 | 0.4541 | -0.0417 | 0.4102 |
| rs4954162 | 2 | 1.35E+08 | TMEM163 | a | g | -0.0746 | -0.1655 | 0.02303 | -0.0464 | 0.2287 | -0.0713 | 0.06399 | -0.0886 | 0.1662 |
| rs5789185 | 2 | 1.35E+08 | TMEM163 | a | g | 0.0807 | -0.0039 | 0.9463 | 0.0476 | 0.1253 | 0.0607 | 0.04937 | 0.015 | 0.774 |
| rs1474055 | 2 | 1.68E+08 | STK39 | t | c | 0.1796 | 0.1467 | 0.05684 | -0.0097 | 0.8184 | 0.0289 | 0.491 | 0.0531 | 0.4536 |
| rs7303831 | 3 | 18320267 | SATB1 | a | c | -0.1693 | -0.1777 | 0.1853 | 0.0376 | 0.6103 | -0.0058 | 0.9364 | -0.0623 | 0.6244 |
| rs6808178 | 3 | 28664199 | LINC00693 | t | c | 0.0658 | 0.0145 | 0.789 | -0.0026 | 0.9283 | 0.0007 | 0.9792 | 0.0605 | 0.2156 |
| rs1249785 | 3 | 48711556 | IP6K2 | t | g | 0.0636 | 0.0402 | 0.4558 | -0.0012 | 0.9671 | -0.0312 | 0.2812 | -0.0006 | 0.9903 |
| rs5596167 | 3 | 1.22E+08 | KPNA1 | t | c | 0.0861 | -0.006 | 0.9325 | 0.0053 | 0.8883 | 0.0199 | 0.5947 | -0.0035 | 0.9557 |
| rs1170741 | 3 | 1.51E+08 | MED12L | a | t | -0.0627 | -0.0172 | 0.754 | -0.0172 | 0.5528 | -0.0336 | 0.2456 | -0.0113 | 0.8199 |
| rs1450522 | 3 | 1.61E+08 | SPTSSB | a | g | -0.0616 | 0.0454 | 0.415 | 0.0355 | 0.2297 | -0.0065 | 0.8247 | 0.0607 | 0.229 |
| rs1051378 | 3 | 1.83E+08 | MCCC1 | t | g | 0.1485 | -0.0038 | 0.9543 | 0.0081 | 0.8168 | -0.0657 | 0.05812 | 0.0123 | 0.8375 |
| rs873786 | 4 | 931588 | GAK | t | c | -0.1731 | -0.1291 | 0.1304 | -0.1078 | 0.01942 | -0.037 | 0.4154 | -0.0596 | 0.4339 |
| rs3431186 | 4 | 958159 | TMEM175 | t | c | -0.2126 | -0.027 | 0.679 | -0.0983 | 0.005259 | -0.046 | 0.1894 | -0.0385 | 0.5177 |
| rs4698412 | 4 | 15735725 | BST1 | a | g | 0.1035 | -0.0525 | 0.3172 | -0.0073 | 0.7915 | 0.0045 | 0.8714 | -0.067 | 0.157 |
| rs3402576 | 4 | 17967188 | LCORL | a | t | -0.0839 | 0.1206 | 0.08306 | -0.0125 | 0.7379 | 0.0174 | 0.6397 | 0.0343 | 0.588 |
| rs6825004 | 4 | 76189212 | SCARB2 | c | g | 0.0622 | 0.0066 | 0.905 | 0.0771 | 0.0104 | 0.0264 | 0.3759 | 0.0597 | 0.237 |
| rs4101061 | 4 | 76226816 | FAM47E | a | g | -0.0912 | -0.0362 | 0.524 | -0.0365 | 0.2303 | -0.0289 | 0.3404 | -0.0511 | 0.3187 |
| rs6854006 | 4 | 76276901 | FAM47E-S1 | t | c | -0.0912 | 0.0328 | 0.5401 | -0.0062 | 0.827 | 0.0101 | 0.7236 | -0.0002 | 0.9974 |
| rs356228 | 4 | 89685975 | SNCA | c | g | 0.1503 | 0.052 | 0.3349 | -0.0162 | 0.5733 | -0.0185 | 0.5205 | 0.0191 | 0.6958 |
| rs356182 | 4 | 89704960 | SNCA | a | g | -0.2774 | 0.0302 | 0.5922 | -0.0063 | 0.8335 | -0.0085 | 0.7744 | 0.0257 | 0.6121 |
| rs5019538 | 4 | 89715479 | SNCA | a | g | -0.1565 | 0.0128 | 0.8238 | -0.0011 | 0.9705 | 0.0035 | 0.9095 | 0.0262 | 0.614 |
| rs356203 | 4 | 89744890 | SNCA | t | c | -0.2504 | 0.0184 | 0.7374 | -0.008 | 0.7853 | -0.0177 | 0.5438 | 0.0097 | 0.8458 |
| rs1311751 | 4 | 1.13E+08 | CAMK2D | t | c | 0.0875 | 0.0049 | 0.9445 | 0.0116 | 0.7578 | 0.0157 | 0.6752 | 0.0441 | 0.4796 |

| | | | | | | | | | | | | | | |
|---|---|---|---|---|---|---|---|---|---|---|---|---|---|---|
| rs62333164 | 4 | 1.7E+08 | CLCN3 | a | g | -0.0638 | -0.0077 | 0.8885 | 0.0063 | 0.831 | 0.0079 | 0.7874 | 0.0032 | 0.9488 |
| rs1867598 | 5 | 60842132 | ELOVL7 | a | g | -0.1554 | -0.1246 | 0.1585 | -0.0564 | 0.2393 | -0.0802 | 0.0921 | -0.0937 | 0.2403 |
| rs26431 | 5 | 1.03E+08 | PAM | c | g | 0.0621 | 0.0185 | 0.7508 | 0.0059 | 0.8469 | 0.0031 | 0.9197 | 0.0061 | 0.9073 |
| rs6875262 | 5 | 1.25E+08 | ZNF608 | t | c | 0.1141 | -0.0372 | 0.6993 | -0.0525 | 0.3052 | -0.0187 | 0.7147 | -0.012 | 0.8899 |
| rs11950533 | 5 | 1.35E+08 | C5orf24 | a | c | -0.0916 | 0.0775 | 0.3746 | 0.0399 | 0.3876 | 0.0261 | 0.5728 | -0.0109 | 0.892 |
| rs4140646 | 6 | 27771022 | LOC100131 | a | g | 0.0833 | -0.0637 | 0.3561 | 0.0033 | 0.9275 | -0.0135 | 0.7078 | -0.0378 | 0.5398 |
| rs9261484 | 6 | 30140906 | TRIM40 | t | c | -0.0635 | -0.0824 | 0.1949 | -0.0186 | 0.5778 | -0.0283 | 0.3958 | -0.0757 | 0.1872 |
| rs9267659 | 6 | 31878457 | SLC44A4 | a | g | 0.079 | 0.0328 | 0.6441 | -0.0357 | 0.3317 | -0.0487 | 0.1851 | -0.0059 | 0.9274 |
| rs11248557 | 6 | 32610995 | HLA-DRB5 | a | c | -0.1676 | -0.1474 | 0.04871 | 0.1518 | 9.86E-05 | 0.0395 | 0.3088 | -0.0037 | 0.9559 |
| rs12528068 | 6 | 71778059 | RIMS1 | t | c | 0.0657 | 0.0677 | 0.2401 | -0.013 | 0.6724 | 0.0202 | 0.5087 | 0.0275 | 0.598 |
| rs997368 | 6 | 1.12E+08 | FYN | a | g | 0.0714 | -0.0274 | 0.6752 | -0.0256 | 0.4611 | -0.0526 | 0.1283 | -0.0193 | 0.7411 |
| rs75859381 | 6 | 1.33E+08 | RPS12 | t | c | -0.2207 | 0.0221 | 0.8878 | -0.0843 | 0.3174 | -0.114 | 0.1728 | -0.0076 | 0.9573 |
| rs199351 | 7 | 23260430 | GPNMB | a | c | 0.1016 | 0.0209 | 0.687 | 0.0255 | 0.3616 | 0.053 | 0.05766 | 0.0309 | 0.5106 |
| rs76949143 | 7 | 66544864 | GS1-124K5 | a | t | -0.1432 | -0.108 | 0.395 | -0.0163 | 0.8041 | -0.042 | 0.5285 | -0.0611 | 0.5918 |
| rs1293298 | 8 | 11854934 | CTSB | a | c | 0.093 | -0.1247 | 0.03706 | 0.0302 | 0.3435 | 0.0154 | 0.6299 | -0.0571 | 0.2923 |
| rs620513 | 8 | 16840084 | FGF20 | t | g | -0.0856 | 0.0276 | 0.6439 | 0.0026 | 0.9352 | 0.0009 | 0.9775 | 0.0106 | 0.8446 |
| rs2280104 | 8 | 22668467 | BIN3 | t | c | 0.0556 | -0.0652 | 0.2348 | 0.0073 | 0.8003 | 0.0129 | 0.6547 | -0.0387 | 0.4314 |
| rs2086641 | 8 | 1.3E+08 | FAM49B | t | c | -0.0605 | -0.053 | 0.3632 | -0.0242 | 0.4389 | -0.0548 | 0.07851 | -0.072 | 0.1699 |
| rs13294100 | 9 | 17579692 | SH3GL2 | t | g | -0.0859 | -0.0297 | 0.5892 | -0.0485 | 0.0963 | 0.0325 | 0.2614 | -0.0393 | 0.4294 |
| rs10756907 | 9 | 17727067 | SH3GL2 | a | g | -0.0926 | -0.0041 | 0.948 | 0.0093 | 0.7776 | 0.0209 | 0.5275 | -0.0447 | 0.4257 |
| rs6476434 | 9 | 34046393 | UBAP2 | t | c | -0.0615 | 0.0849 | 0.1659 | -0.0119 | 0.7087 | 0.0004 | 0.9898 | -0.0209 | 0.7099 |
| rs896435 | 10 | 15515407 | ITGA8 | t | c | 0.0735 | 0.0303 | 0.5988 | -0.0232 | 0.4402 | 0.0379 | 0.2077 | 0.0335 | 0.5188 |
| rs10748818 | 10 | 1.02E+08 | GBF1 | a | g | -0.079 | -0.0072 | 0.9223 | -0.0613 | 0.1139 | -0.0422 | 0.2823 | -0.1009 | 0.1272 |
| rs72840788 | 10 | 1.2E+08 | BAG3 | a | g | 0.0763 | 0.0228 | 0.7197 | -0.0165 | 0.6297 | -0.0176 | 0.6046 | -0.019 | 0.743 |
| rs11789673 | 10 | 1.2E+08 | INPP5F | a | g | 0.4354 | -0.1325 | 0.5232 | -0.1439 | 0.2285 | -0.0873 | 0.4422 | -0.2517 | 0.185 |
| rs7938782 | 11 | 10537230 | RNF141 | a | g | 0.087 | -0.0478 | 0.5407 | -0.0409 | 0.328 | -0.0176 | 0.6696 | -0.0811 | 0.2429 |
| rs12283611 | 11 | 83776234 | DLG2 | a | c | -0.0645 | 0.0473 | 0.3766 | 0.0178 | 0.5291 | -0.0412 | 0.144 | 0.0002 | 0.9967 |
| rs3802920 | 11 | 1.34E+08 | IGSF9B | t | g | 0.1073 | 0.0053 | 0.9342 | -0.0196 | 0.5673 | -0.0327 | 0.3372 | 0.0519 | 0.3654 |
| rs76904798 | 12 | 40220632 | LRRK2 | t | c | 0.1439 | 0.0936 | 0.2101 | -0.0226 | 0.5751 | -0.0055 | 0.8905 | 0.0787 | 0.2515 |
| rs11707380 | 12 | 40528770 | LRRK2 | t | c | -0.3955 | -0.1718 | 0.3629 | -0.0737 | 0.4545 | -0.1095 | 0.2856 | -0.0154 | 0.928 |
| rs14112880 | 12 | 40542837 | CNTN1 | t | c | 0.3468 | -0.2945 | 0.323 | 0.0543 | 0.6823 | 0.0845 | 0.5452 | -0.3185 | 0.1975 |
| rs13801711 | 12 | 41901977 | GXYLT1 | c | g | 0.413 | 0.2362 | 0.2877 | 0.1004 | 0.4535 | 0.0467 | 0.7279 | 0.2611 | 0.213 |
| rs7134559 | 12 | 46025303 | SCAF11 | t | c | -0.0539 | -0.0706 | 0.1933 | -0.0093 | 0.7429 | 0.0167 | 0.5543 | -0.0822 | 0.09372 |
| rs10847864 | 12 | 1.23E+08 | HIP1R | t | g | 0.1478 | -0.0208 | 0.7189 | -0.0325 | 0.2939 | 0.0016 | 0.9589 | -0.0479 | 0.3619 |
| rs11610045 | 12 | 1.32E+08 | FBRSL1 | a | g | 0.0601 | 0.0329 | 0.5307 | 0.0857 | 0.002307 | 0.0546 | 0.05206 | 0.0796 | 0.09334 |
| rs9568188 | 13 | 49353596 | CAB39L | t | c | 0.0617 | -0.011 | 0.8524 | 0.0133 | 0.6732 | 0.043 | 0.1699 | -0.005 | 0.9264 |
| rs4771268 | 13 | 97212767 | MBNL2 | t | c | 0.0675 | 0.0347 | 0.5821 | -0.0173 | 0.6015 | -0.033 | 0.3186 | -0.0112 | 0.8444 |
| rs12147950 | 14 | 37520065 | MIPOL1 | t | c | -0.0529 | 0.0568 | 0.2901 | -0.0051 | 0.8577 | -0.0319 | 0.2609 | 0.0511 | 0.2921 |

| | | | | | | | | | | | | | | |
|---|---|---|---|---|---|---|---|---|---|---|---|---|---|---|
| rs1115802 | 14 | 54882151 | GCH1 | t | c | -0.0842 | 0.0052 | 0.9257 | 0.056 | 0.0573 | 0.0346 | 0.2382 | 0.0345 | 0.4932 |
| rs3742785 | 14 | 74906331 | RPS6KL1 | a | c | 0.0707 | 0.0414 | 0.5198 | 0.0389 | 0.2488 | 0.0838 | 0.01328 | 0.0167 | 0.7743 |
| rs979812 | 14 | 87997920 | GALC | t | g | 0.061 | -0.0711 | 0.1786 | 0.0651 | 0.01904 | 0.0266 | 0.3356 | -0.0094 | 0.8435 |
| rs2251086 | 15 | 61705186 | VPS13C | t | c | -0.1186 | -0.0154 | 0.8306 | 0.0026 | 0.9473 | 0.029 | 0.4495 | 0.0045 | 0.9454 |
| rs6497339 | 16 | 19266171 | SYT17 | a | t | 0.063 | -0.08 | 0.1386 | -0.0351 | 0.2163 | 0.0028 | 0.9209 | -0.0727 | 0.1368 |
| rs2904880 | 16 | 28933075 | CD19 | c | g | -0.065 | 0.127 | 0.0283 | -0.0193 | 0.5285 | -0.0324 | 0.2875 | 0.0709 | 0.178 |
| rs1115060 | 16 | 30966478 | SETD1A | a | g | 0.0907 | 0.0375 | 0.4984 | -0.0336 | 0.2464 | 0.0007 | 0.9814 | 0.004 | 0.9358 |
| rs6500328 | 16 | 50702745 | NOD2 | a | g | 0.0586 | 0.0984 | 0.06656 | 0.0629 | 0.02589 | 0.0926 | 0.00099 | 0.0983 | 0.04115 |
| rs3104783 | 16 | 52602330 | CASC16 | a | c | 0.0668 | 0.0048 | 0.9283 | 0.0199 | 0.4815 | -0.0143 | 0.6103 | 0.0157 | 0.7435 |
| rs1022115 | 16 | 52935514 | CHD9 | a | g | -0.1156 | 0.0922 | 0.2972 | 0.0043 | 0.9279 | 0.0199 | 0.6717 | 0.0813 | 0.3153 |
| rs1260086 | 17 | 7452302 | CHRNB1 | a | c | -0.0565 | -0.0926 | 0.09228 | 0.0039 | 0.8953 | -0.0303 | 0.2988 | -0.0399 | 0.4223 |
| rs1295163 | 17 | 42588995 | RETREG3 | t | c | 0.0642 | 0.1958 | 0.001277 | 0.0588 | 0.06482 | 0.1011 | 0.0014 | 0.1121 | 0.03838 |
| rs2269906 | 17 | 44216969 | UBTF | a | c | 0.0631 | 0.1448 | 0.008531 | -0.0083 | 0.7748 | 0.0204 | 0.4806 | 0.0957 | 0.05227 |
| rs850738 | 17 | 44357262 | FAM171A2 | a | g | -0.071 | -0.0902 | 0.094 | -0.0512 | 0.07455 | -0.0693 | 0.01561 | -0.1581 | 0.001202 |
| rs1768623 | 17 | 45339907 | MAP3K14 | t | g | -0.1896 | -0.1499 | 0.08866 | -0.0819 | 0.07819 | -0.0326 | 0.4799 | -0.1393 | 0.07444 |
| rs6205394 | 17 | 45666837 | CRHR1 | t | c | -0.27 | 0.033 | 0.6712 | -0.0445 | 0.2833 | -0.0796 | 0.05527 | 0.0206 | 0.7664 |
| rs9912362 | 17 | 45706862 | CRHR1 | t | c | 0.077 | 0.0232 | 0.6737 | 0.0121 | 0.6759 | -0.0042 | 0.8839 | -0.0122 | 0.8072 |
| rs1176156 | 17 | 45720942 | CRHR1 | a | g | -0.2324 | -0.1823 | 0.08694 | -0.0046 | 0.9361 | 0.1289 | 0.02346 | -0.1003 | 0.2745 |
| rs7221167 | 17 | 45855941 | MAPT-AS1 | t | c | 0.1443 | -0.0102 | 0.8479 | 0.0164 | 0.5659 | -0.0093 | 0.7431 | -0.0356 | 0.4599 |
| rs7225002 | 17 | 46111701 | KANSL1 | a | g | 0.1351 | 0.0144 | 0.7875 | 0.0187 | 0.5101 | 0.0188 | 0.5087 | -0.0197 | 0.6844 |
| rs199453 | 17 | 46723580 | NSF | t | c | -0.2045 | -0.0039 | 0.9514 | -0.04 | 0.2327 | -0.044 | 0.1881 | 0.0207 | 0.7156 |
| rs1165897 | 17 | 46789439 | WNT3 | a | g | -0.0624 | 0.0763 | 0.1573 | 0.0383 | 0.1893 | -0.0383 | 0.1868 | 0.0762 | 0.1197 |
| rs6116987 | 17 | 61840005 | BRIP1 | t | c | 0.082 | 0.0436 | 0.54 | -0.0571 | 0.1375 | -0.0284 | 0.458 | 0.0052 | 0.9354 |
| rs666463 | 17 | 78429399 | DNAH17 | a | t | 0.076 | 0.0076 | 0.9121 | -0.0229 | 0.5371 | 0.0271 | 0.4673 | -0.0057 | 0.9255 |
| rs1941685 | 18 | 33724354 | ASXL3 | t | g | 0.0531 | 0.0081 | 0.8765 | 0.0044 | 0.8731 | 0.0006 | 0.9825 | -0.0501 | 0.2885 |
| rs1245649 | 18 | 43093415 | RIT2 | a | g | -0.0983 | -0.0605 | 0.2764 | 0.0083 | 0.7794 | -0.0009 | 0.9761 | 0.0142 | 0.778 |
| rs8087969 | 18 | 51157219 | MEX3C | t | g | -0.0578 | 0.0524 | 0.333 | 0.0389 | 0.1716 | 0.0653 | 0.02117 | 0.0493 | 0.3085 |
| rs5581831 | 19 | 2341049 | SPPL2B | t | c | -0.0696 | 0.028 | 0.6251 | -0.0637 | 0.03882 | -0.0586 | 0.05877 | -0.0147 | 0.7756 |
| rs2295545 | 20 | 3184040 | DDRGK1 | t | c | 0.0622 | 0.0032 | 0.954 | 0.0496 | 0.08838 | 0.0165 | 0.5679 | 0.0225 | 0.6542 |
| rs7735182 | 20 | 6025395 | CRLS1 | t | c | 0.0802 | -0.0658 | 0.4188 | 0.0383 | 0.3762 | 0.061 | 0.1526 | 0.0152 | 0.8341 |
| rs2248244 | 21 | 37480059 | DYRK1A | a | g | 0.0714 | -0.0036 | 0.9522 | 0.0306 | 0.3309 | 0.0137 | 0.6634 | 0.0351 | 0.516 |

**Table S8: Association of Nalls et al PD risk loci with related dementia pathologies**

| Nalls et al SNPs | | | | | | | CAA | | CAA (any/none) | | HS | | TDP-43 (3 cat) | | TDP-43 (any/none) | |
|---|---|---|---|---|---|---|---|---|---|---|---|---|---|---|---|---|
| SNP | CHR | POS_NEW | GENE | Effect allel | Referent | Beta | Beta | Pval | Beta | Pval | Beta | Pval | Beta | Pval | Beta | Pval |
| rs1141387( | 1 | 154925709 | PMVK | c | g | 0.2812 | 0.1935 | 0.2185 | 0.1393 | 0.4657 | -0.3607 | 0.1849 | -0.0312 | 0.8928 | -0.1125 | 0.6486 |
| rs3574901 | 1 | 155162560 | KRTCAP2 | a | g | 0.6068 | 0.1095 | 0.3665 | 0.1612 | 0.2485 | 0.2216 | 0.3081 | -0.4763 | 0.03218 | -0.3217 | 0.1772 |
| rs3564392! | 1 | 156185069 | SEMA4A | a | g | 0.389 | 0.0379 | 0.722 | 0.0982 | 0.427 | 0.0489 | 0.7986 | -0.2014 | 0.2866 | -0.1178 | 0.5527 |
| rs6658353 | 1 | 161499264 | FCGR2A | c | g | 0.065 | 0.0073 | 0.799 | 0.0119 | 0.7122 | 0.0881 | 0.05491 | -0.0136 | 0.776 | 0.0073 | 0.874 |
| rs1157869! | 1 | 171750629 | VAMP4 | t | c | -0.0704 | 0.0184 | 0.606 | 0.0098 | 0.8077 | 0.004 | 0.9439 | 0.0031 | 0.9573 | 0.0049 | 0.9301 |
| rs823118 | 1 | 205754444 | NUCKS1 | t | c | 0.1066 | 0.0107 | 0.7055 | 0.0256 | 0.4271 | -0.0335 | 0.4641 | -0.0252 | 0.5903 | -0.0254 | 0.5738 |
| rs1155708( | 1 | 205768611 | RAB29 | a | g | 0.1315 | 0.0234 | 0.575 | 0.029 | 0.5343 | 0.1072 | 0.1053 | 0.133 | 0.0504 | 0.1098 | 0.09024 |
| rs4653767 | 1 | 226728377 | ITPKB | t | c | 0.0833 | 0.0022 | 0.9437 | 0.0054 | 0.8766 | 0.0151 | 0.7616 | -0.0692 | 0.1787 | -0.0692 | 0.1591 |
| rs1079757( | 1 | 232528865 | SIPA1L2 | t | c | 0.1114 | 0.0228 | 0.5898 | 0.0018 | 0.9692 | -0.1466 | 0.02897 | 0.1093 | 0.1285 | 0.0956 | 0.1537 |
| rs7611622 | 2 | 17966582 | KCNS3 | a | t | 0.1104 | 0.1108 | 0.02715 | 0.1281 | 0.02484 | 0.0402 | 0.625 | 0.0473 | 0.5691 | 0.1079 | 0.1877 |
| rs2042477 | 2 | 95335195 | KCNIP3 | a | t | -0.0657 | 0.0698 | 0.03751 | 0.0421 | 0.2675 | 0.0331 | 0.5456 | 0.0333 | 0.5555 | 0.0188 | 0.7328 |
| rs1168300 | 2 | 101780501 | MAP4K4 | a | t | 0.0705 | -0.0265 | 0.3801 | -0.0219 | 0.52 | 0.016 | 0.7415 | 0.0763 | 0.1269 | 0.0201 | 0.6751 |
| rs4954162 | 2 | 134681219 | TMEM163 | a | g | -0.0746 | -0.0054 | 0.8911 | -0.0211 | 0.63 | -0.0558 | 0.3648 | 0.1262 | 0.07117 | 0.0955 | 0.1297 |
| rs5789185! | 2 | 134707046 | TMEM163 | a | g | 0.0807 | 0.0462 | 0.1448 | 0.0589 | 0.09888 | 0.0812 | 0.1091 | -0.0445 | 0.3975 | -0.0335 | 0.5007 |
| rs1474055 | 2 | 168253884 | STK39 | t | c | 0.1796 | 0.0645 | 0.135 | 0.1084 | 0.02473 | 0.0352 | 0.6082 | 0.1745 | 0.01534 | 0.0884 | 0.1921 |
| rs7303831! | 3 | 18320267 | SATB1 | a | c | -0.1693 | 0.0955 | 0.192 | 0.0321 | 0.7021 | -0.053 | 0.6661 | -0.0865 | 0.4701 | -0.024 | 0.8434 |
| rs6808178 | 3 | 28664199 | LINC00693 | t | c | 0.0658 | 0.0081 | 0.7795 | -0.0097 | 0.7672 | -0.0022 | 0.9632 | 0.0046 | 0.9238 | -0.0009 | 0.9838 |
| rs1249785( | 3 | 48711556 | IP6K2 | t | g | 0.0636 | 0.0265 | 0.3652 | 0.0272 | 0.4147 | 0.0445 | 0.3515 | -0.0742 | 0.128 | -0.0339 | 0.4701 |
| rs5596167 | 3 | 122478045 | KPNA1 | t | c | 0.0861 | -0.0329 | 0.3951 | -0.0422 | 0.3296 | 0.1615 | 0.008322 | 0.0002 | 0.9976 | -0.0203 | 0.7393 |
| rs1170741( | 3 | 151391177 | MED12L | a | t | -0.0627 | 0.0313 | 0.2904 | -0.0089 | 0.7886 | -0.0009 | 0.9841 | -0.0168 | 0.7332 | -0.0044 | 0.9254 |
| rs1450522 | 3 | 161359842 | SPTSSB | a | g | -0.0616 | -0.0092 | 0.7581 | 0.0037 | 0.9114 | -0.0341 | 0.4817 | 0.1181 | 0.01781 | 0.068 | 0.1573 |
| rs1051378! | 3 | 183042285 | MCCC1 | t | g | 0.1485 | -0.0259 | 0.4632 | -0.067 | 0.09316 | -0.0021 | 0.9713 | 0.0698 | 0.2306 | 0.0355 | 0.5302 |
| rs873786 | 4 | 931588 | GAK | t | c | -0.1731 | -0.0298 | 0.5203 | 0.0374 | 0.4703 | -0.0097 | 0.8957 | 0.2237 | 0.004997 | 0.2471 | 0.000809 |
| rs3431186( | 4 | 958159 | TMEM175 | t | c | -0.2126 | -0.0662 | 0.06446 | -0.0612 | 0.1305 | 0.05 | 0.3944 | 0.0739 | 0.2156 | 0.043 | 0.4595 |
| rs4698412 | 4 | 15735725 | BST1 | a | g | 0.1035 | -0.0219 | 0.4398 | -0.0137 | 0.6693 | -0.0341 | 0.4562 | 0.044 | 0.3435 | 0.0626 | 0.1637 |
| rs3402576( | 4 | 17967188 | LCORL | a | t | -0.0839 | 0.0322 | 0.4043 | 0.0654 | 0.128 | -0.0145 | 0.8112 | -0.0754 | 0.2504 | 0.0015 | 0.9801 |
| rs6825004 | 4 | 76189212 | SCARB2 | c | g | 0.0622 | 0.0567 | 0.06281 | 0.065 | 0.05926 | 0.0324 | 0.5073 | -0.0037 | 0.9413 | -0.0287 | 0.5507 |
| rs4101061 | 4 | 76226816 | FAM47E | a | g | -0.0912 | 0.0443 | 0.1511 | 0.0448 | 0.1994 | -0.0212 | 0.6717 | -0.0157 | 0.7634 | -0.0257 | 0.6095 |
| rs6854006 | 4 | 76276901 | FAM47E-ST | t | c | -0.0912 | 0.0002 | 0.9956 | 0.0183 | 0.5787 | 0.0088 | 0.8502 | 0.0311 | 0.5213 | 0.0194 | 0.6739 |
| rs356228 | 4 | 89685975 | SNCA | c | g | 0.1503 | -0.0115 | 0.6929 | -0.0034 | 0.9171 | -0.0254 | 0.5886 | -0.073 | 0.1303 | -0.0528 | 0.2572 |
| rs356182 | 4 | 89704960 | SNCA | a | g | -0.2774 | 0.004 | 0.895 | 0.0032 | 0.9266 | 0.0184 | 0.7057 | 0.0405 | 0.4248 | 0.0185 | 0.7018 |
| rs5019538 | 4 | 89715479 | SNCA | a | g | -0.1565 | -0.0021 | 0.945 | 0.0125 | 0.7205 | 0.0446 | 0.3716 | 0.0844 | 0.09935 | 0.0502 | 0.3106 |
| rs356203 | 4 | 89744890 | SNCA | t | c | -0.2504 | 0.0106 | 0.7196 | 0.014 | 0.6752 | 0.0405 | 0.3958 | 0.0561 | 0.2573 | 0.0306 | 0.5219 |
| rs1311751! | 4 | 113447909 | CAMK2D | t | c | 0.0875 | -0.0138 | 0.7193 | -0.0314 | 0.4669 | 0.075 | 0.2193 | -0.0258 | 0.6861 | 0.0189 | 0.7553 |
| rs6233316 | 4 | 169662006 | CLCN3 | a | g | -0.0638 | -0.0109 | 0.714 | -0.0415 | 0.2173 | 0.0233 | 0.6278 | -0.0305 | 0.5361 | -0.0096 | 0.8395 |
| rs1867598 | 5 | 60842132 | ELOVL7 | a | g | -0.1554 | -0.0609 | 0.2272 | -0.1261 | 0.02355 | -0.1159 | 0.1366 | 0.0375 | 0.6533 | 0.0208 | 0.7919 |
| rs26431 | 5 | 103030090 | PAM | c | g | 0.0621 | -0.0167 | 0.5901 | 0.0225 | 0.522 | 0.0101 | 0.8407 | -0.0641 | 0.2175 | -0.0226 | 0.6489 |
| rs6875262 | 5 | 124774580 | ZNF608 | t | c | 0.1141 | -0.0227 | 0.6535 | -0.0062 | 0.9131 | 0.0218 | 0.7956 | 0.0142 | 0.8717 | 0.0105 | 0.9014 |
| rs1195053 | 5 | 134863415 | C5orf24 | a | c | -0.0916 | 0.0612 | 0.1885 | 0.0711 | 0.1754 | -0.0601 | 0.4317 | -0.1289 | 0.08923 | -0.0812 | 0.2716 |
| rs4140646 | 6 | 27771022 | LOC100131 | a | g | 0.0833 | -0.0005 | 0.9903 | -0.03 | 0.4739 | 0.0203 | 0.7327 | 0.0216 | 0.7275 | -0.0055 | 0.9263 |
| rs9261484 | 6 | 30140906 | TRIM40 | t | c | -0.0635 | -0.0328 | 0.3324 | -0.0258 | 0.5013 | 0.0342 | 0.5379 | 0.0124 | 0.825 | 0.0483 | 0.3761 |

| | | | | | | | | | | | | | | | | |
|---|---|---|---|---|---|---|---|---|---|---|---|---|---|---|---|---|
| rs9267659 | 6 | 31878457 | SLC44A4 | a | g | 0.079 | 0.0056 | 0.8805 | -0.0166 | 0.6938 | -0.1116 | 0.06671 | -0.0723 | 0.2317 | -0.0689 | 0.2512 |
| rs1124855 | 6 | 32610995 | HLA-DRB5 | a | c | -0.1676 | -0.0046 | 0.907 | -0.0219 | 0.6257 | 0.1221 | 0.06263 | 0.1668 | 0.0107 | 0.0881 | 0.1699 |
| rs1252806 | 6 | 71778059 | RIMS1 | t | c | 0.0657 | -0.0237 | 0.4506 | -0.0015 | 0.9666 | -0.0683 | 0.1753 | 0.0101 | 0.8468 | 0.0447 | 0.3721 |
| rs997368 | 6 | 111922088 | FYN | a | g | 0.0714 | -0.089 | 0.01305 | -0.0988 | 0.01363 | 0.0587 | 0.2999 | -0.0009 | 0.9885 | 0.0137 | 0.8074 |
| rs7585938 | 6 | 132889222 | RPS12 | t | c | -0.2207 | -0.0743 | 0.3915 | -0.1815 | 0.05943 | 0.0293 | 0.8325 | -0.198 | 0.173 | -0.1546 | 0.25 |
| rs199351 | 7 | 23260430 | GPNMB | a | c | 0.1016 | 0.0205 | 0.4724 | 0.037 | 0.2499 | -0.0648 | 0.1564 | -0.0382 | 0.4248 | -0.0652 | 0.1538 |
| rs7694914 | 7 | 66544864 | GS1-124K5. | a | t | -0.1432 | -0.0106 | 0.875 | 0.019 | 0.8005 | -0.021 | 0.8458 | -0.1784 | 0.1075 | -0.2619 | 0.01596 |
| rs1293298 | 8 | 11854934 | CTSB | a | c | 0.093 | 0.0068 | 0.8333 | 0.0048 | 0.8958 | -0.1007 | 0.05381 | -0.0886 | 0.09729 | -0.0364 | 0.4791 |
| rs620513 | 8 | 16840084 | FGF20 | t | g | -0.0856 | 0.0338 | 0.2956 | 0.0404 | 0.2673 | 0.0114 | 0.8273 | 0.0753 | 0.1595 | 0.0171 | 0.7392 |
| rs2280104 | 8 | 22668467 | BIN3 | t | c | 0.0556 | -0.0321 | 0.279 | 0.0031 | 0.9272 | -0.0273 | 0.5651 | -0.015 | 0.7589 | -0.0274 | 0.5594 |
| rs2086641 | 8 | 129889663 | FAM49B | t | c | -0.0605 | 0.017 | 0.596 | 0.0039 | 0.9126 | 0.0508 | 0.3182 | -0.1121 | 0.04224 | -0.0974 | 0.05694 |
| rs1329410 | 9 | 17579692 | SH3GL2 | t | g | -0.0859 | 0.0024 | 0.9347 | 0.0078 | 0.8147 | -0.0388 | 0.4156 | 0.0363 | 0.4618 | 0.009 | 0.8506 |
| rs1075690 | 9 | 17727067 | SH3GL2 | a | g | -0.0926 | -0.0229 | 0.4995 | -0.0188 | 0.6197 | 0.0805 | 0.1347 | 0.0339 | 0.5471 | -0.0169 | 0.7531 |
| rs6476434 | 9 | 34046393 | UBAP2 | t | c | -0.0615 | 0.0348 | 0.2729 | 0.0309 | 0.3964 | 0.0822 | 0.1266 | 0.0514 | 0.3241 | 0.0785 | 0.1371 |
| rs896435 | 10 | 15515407 | ITGA8 | t | c | 0.0735 | 0.0252 | 0.4121 | 0.0337 | 0.3307 | 0.0069 | 0.8892 | -0.0699 | 0.1736 | -0.0709 | 0.148 |
| rs1074881 | 10 | 102255522 | GBF1 | a | g | -0.079 | 0.0087 | 0.8291 | -0.0313 | 0.4905 | -0.0491 | 0.4487 | 0.01 | 0.8806 | -0.012 | 0.851 |
| rs7284078 | 10 | 119656173 | BAG3 | a | g | 0.0763 | 0.0164 | 0.6367 | 0.0231 | 0.5569 | 0.0538 | 0.3393 | -0.0019 | 0.9738 | -0.0288 | 0.6027 |
| rs1178967 | 10 | 119776815 | INPP5F | a | g | 0.4354 | -0.0325 | 0.7809 | 0.1307 | 0.3108 | -0.137 | 0.4531 | -0.0322 | 0.8706 | 0.0722 | 0.6995 |
| rs7938782 | 11 | 10537230 | RNF141 | a | g | 0.087 | -0.0079 | 0.8536 | -0.0039 | 0.9345 | -0.0328 | 0.6285 | -0.0283 | 0.6939 | 0.0049 | 0.9419 |
| rs1228361 | 11 | 83776234 | DLG2 | a | c | -0.0645 | -0.0105 | 0.7158 | 0.0068 | 0.8338 | 0.0415 | 0.3728 | 0.0058 | 0.9033 | 0.0395 | 0.3869 |
| rs3802920 | 11 | 133917106 | IGSF9B | t | g | 0.1073 | -0.0088 | 0.8001 | -0.022 | 0.5745 | -0.0938 | 0.09262 | -0.0056 | 0.9236 | 0.0009 | 0.9876 |
| rs7690479 | 12 | 40220632 | LRRK2 | t | c | 0.1439 | -0.0828 | 0.0423 | -0.0737 | 0.1121 | 0.0315 | 0.6344 | 0.0682 | 0.3125 | 0.0798 | 0.2154 |
| rs1170738 | 12 | 40528770 | LRRK2 | t | c | -0.3955 | -0.0263 | 0.7999 | 0.1218 | 0.3004 | -0.117 | 0.4841 | -0.2183 | 0.2518 | -0.1301 | 0.4454 |
| rs1411288 | 12 | 40542837 | CNTN1 | t | c | 0.3468 | -0.0689 | 0.6188 | -0.1696 | 0.2872 | -0.044 | 0.8461 | 0.3156 | 0.2378 | 0.2349 | 0.3171 |
| rs1380171 | 12 | 41901977 | GXYLT1 | c | g | 0.413 | 0.1297 | 0.358 | 0.2097 | 0.1695 | 0.2152 | 0.3307 | 0.022 | 0.9338 | 0.4253 | 0.06128 |
| rs7134559 | 12 | 46025303 | SCAF11 | t | c | -0.0539 | 0.0131 | 0.6482 | 0.0265 | 0.4157 | 0.0536 | 0.2495 | -0.0075 | 0.8758 | -0.0134 | 0.7717 |
| rs1084786 | 12 | 122842051 | HIP1R | t | g | 0.1478 | -0.026 | 0.4038 | -0.0388 | 0.2708 | 0.0587 | 0.247 | -0.0241 | 0.6527 | -0.028 | 0.5867 |
| rs1161004 | 12 | 132487182 | FBRSL1 | a | g | 0.0601 | 0.0397 | 0.1673 | 0.059 | 0.06753 | 0.0536 | 0.2455 | 0.0622 | 0.201 | 0.0354 | 0.4471 |
| rs9568188 | 13 | 49353596 | CAB39L | t | c | 0.0617 | -0.0225 | 0.4768 | 0.0073 | 0.8389 | -0.0096 | 0.854 | -0.0236 | 0.6576 | -0.0262 | 0.6138 |
| rs4771268 | 13 | 97212767 | MBNL2 | t | c | 0.0675 | 0.0023 | 0.9449 | 0.0091 | 0.81 | 0.0754 | 0.163 | -0.1244 | 0.02956 | -0.0701 | 0.1982 |
| rs1214795 | 14 | 37520065 | MIPOL1 | t | c | -0.0529 | 0.0261 | 0.3628 | 0.0274 | 0.3984 | 0.0335 | 0.4707 | -0.0329 | 0.4982 | -0.0393 | 0.3991 |
| rs1115802 | 14 | 54882151 | GCH1 | t | c | -0.0842 | 0.0384 | 0.1981 | 0.0482 | 0.1535 | 0.0476 | 0.3288 | -0.0237 | 0.6335 | -0.0056 | 0.9082 |
| rs3742785 | 14 | 74906331 | RPS6KL1 | a | c | 0.0707 | -0.0401 | 0.2411 | -0.056 | 0.1486 | 0.025 | 0.6544 | 0.0687 | 0.2309 | 0.0589 | 0.2924 |
| rs979812 | 14 | 87997920 | GALC | t | g | 0.061 | 0.0278 | 0.3248 | 0.0114 | 0.7215 | -0.0946 | 0.03827 | -0.024 | 0.6117 | -0.029 | 0.5225 |
| rs2251086 | 15 | 61705186 | VPS13C | t | c | -0.1186 | 0.0264 | 0.5073 | -0.0042 | 0.925 | 0.0623 | 0.3324 | -0.0127 | 0.8471 | -0.025 | 0.6946 |
| rs6497339 | 16 | 19266171 | SYT17 | a | t | 0.063 | 0.0105 | 0.7154 | -0.0247 | 0.4477 | 0.0008 | 0.9857 | 0.0195 | 0.6856 | -0.024 | 0.606 |
| rs2904880 | 16 | 28933075 | CD19 | c | g | -0.065 | 0.1013 | 0.001131 | 0.0949 | 0.006768 | -0.0381 | 0.4474 | -0.0022 | 0.9668 | -0.0223 | 0.6581 |
| rs1115060 | 16 | 30966478 | SETD1A | a | g | 0.0907 | 0.0099 | 0.7357 | 0.0257 | 0.4389 | -0.0459 | 0.3301 | -0.0999 | 0.04103 | -0.0498 | 0.2861 |
| rs6500328 | 16 | 50702745 | NOD2 | a | g | 0.0586 | 0.0421 | 0.1412 | 0.0879 | 0.006572 | 0.134 | 0.003671 | 0.0191 | 0.6923 | 0.0214 | 0.6412 |
| rs3104783 | 16 | 52602330 | CASC16 | a | c | 0.0668 | 0.0132 | 0.6462 | 0.0031 | 0.9243 | 0.0139 | 0.7638 | 0.0269 | 0.5789 | -0.0083 | 0.8577 |
| rs1022115 | 16 | 52935514 | CHD9 | a | g | -0.1156 | -0.0349 | 0.4664 | -0.0538 | 0.3207 | 0.0688 | 0.3744 | 0.0102 | 0.9026 | 0.0551 | 0.4806 |
| rs1260086 | 17 | 7452302 | CHRNB1 | a | c | -0.0565 | -0.0272 | 0.361 | -0.027 | 0.4213 | 0.0151 | 0.7535 | -0.0285 | 0.5631 | -0.0215 | 0.6493 |
| rs1295163 | 17 | 42588995 | RETREG3 | t | c | 0.0642 | 0.0282 | 0.3812 | 0.0152 | 0.6742 | -0.0267 | 0.6076 | 0.0377 | 0.4753 | 0.055 | 0.2782 |
| rs2269906 | 17 | 44216969 | UBTF | a | c | 0.0631 | -0.0171 | 0.5641 | -0.0202 | 0.546 | 0.0561 | 0.2374 | 0.0545 | 0.2675 | 0.0696 | 0.1398 |
| rs850738 | 17 | 44357262 | FAM171A2 | a | g | -0.071 | -0.0015 | 0.9581 | -0.0267 | 0.4174 | 0.226 | 1.67E-06 | 0.1894 | 7.86E-05 | 0.1807 | 0.000107 |

| | | | | | | | | | | | | | | | | |
|---|---|---|---|---|---|---|---|---|---|---|---|---|---|---|---|---|
| rs17686238 | 17 | 45339907 | MAP3K14 | t | g | -0.1896 | -0.015 | 0.7465 | -0.0756 | 0.1494 | 0.1459 | 0.04999 | 0.0767 | 0.326 | 0.0687 | 0.3565 |
| rs62053943 | 17 | 45666837 | CRHR1 | t | c | -0.27 | 0.0013 | 0.975 | -0.0271 | 0.5632 | 0.2325 | 0.000448 | 0.0675 | 0.3362 | 0.0554 | 0.3988 |
| rs9912362 | 17 | 45706862 | CRHR1 | t | c | 0.077 | 0.0156 | 0.5967 | 0.051 | 0.1253 | -0.1046 | 0.02755 | -0.0368 | 0.4513 | -0.0531 | 0.2616 |
| rs11761568 | 17 | 45720942 | CRHR1 | a | g | -0.2324 | 0.037 | 0.5291 | -0.0099 | 0.8777 | 0.138 | 0.1263 | 0.1667 | 0.1111 | 0.175 | 0.05816 |
| rs7221167 | 17 | 45855941 | MAPT-AS1 | t | c | 0.1443 | 0.0061 | 0.8315 | 0.042 | 0.1963 | -0.1597 | 0.000601 | -0.0137 | 0.7714 | 0.0134 | 0.769 |
| rs7225002 | 17 | 46111701 | KANSL1 | a | g | 0.1351 | 0.04 | 0.1682 | 0.0679 | 0.03758 | -0.216 | 3.55E-06 | -0.005 | 0.9167 | 0.0215 | 0.6427 |
| rs199453 | 17 | 46723580 | NSF | t | c | -0.2045 | 0.0124 | 0.7154 | -0.0123 | 0.7456 | 0.235 | 1.37E-05 | 0.081 | 0.1586 | 0.101 | 0.06134 |
| rs11658970 | 17 | 46789439 | WNT3 | a | g | -0.0624 | -0.0054 | 0.8566 | -0.0049 | 0.8832 | 0.0455 | 0.3406 | 0.0251 | 0.6087 | 0.0999 | 0.03512 |
| rs61169879 | 17 | 61840005 | BRIP1 | t | c | 0.082 | 0.0221 | 0.5668 | 0.0105 | 0.8103 | 0.0687 | 0.2706 | 0.0987 | 0.138 | 0.0566 | 0.3609 |
| rs666463 | 17 | 78429399 | DNAH17 | a | t | 0.076 | 0.0693 | 0.07041 | 0.0991 | 0.02156 | 0.0634 | 0.2992 | -0.0043 | 0.945 | 0.0084 | 0.8882 |
| rs1941685 | 18 | 33724354 | ASXL3 | t | g | 0.0531 | -0.0001 | 0.9965 | -0.0015 | 0.9626 | 0.0687 | 0.127 | -0.0129 | 0.7819 | -0.0395 | 0.3781 |
| rs12456492 | 18 | 43093415 | RIT2 | a | g | -0.0983 | 0.0006 | 0.9853 | -0.0219 | 0.5222 | 0.0923 | 0.05796 | 0.0452 | 0.3646 | 0.0205 | 0.6698 |
| rs8087969 | 18 | 51157219 | MEX3C | t | g | -0.0578 | 0.0062 | 0.8294 | 0.0135 | 0.6787 | -0.0189 | 0.6849 | 0.0609 | 0.2067 | 0.0237 | 0.607 |
| rs55818311 | 19 | 2341049 | SPPL2B | t | c | -0.0696 | -0.0547 | 0.08291 | -0.093 | 0.008573 | -0.0256 | 0.6117 | -0.0446 | 0.4049 | -0.0292 | 0.5631 |
| rs2295545 | 20 | 3184040 | DDRGK1 | t | c | 0.0622 | 0.0136 | 0.6406 | -0.0071 | 0.8311 | 0.0015 | 0.9741 | -0.0661 | 0.1809 | -0.0177 | 0.7086 |
| rs77351827 | 20 | 6025395 | CRLS1 | t | c | 0.0802 | 0.01 | 0.8205 | 0.0314 | 0.524 | -0.1024 | 0.1476 | -0.0061 | 0.9322 | -0.0277 | 0.6933 |
| rs2248244 | 21 | 37480059 | DYRK1A | a | g | 0.0714 | 0.0278 | 0.3861 | 0.0099 | 0.7836 | -0.0022 | 0.9665 | -0.0057 | 0.9152 | -0.0058 | 0.9106 |

**Table S9. Estimated SNP heritability across all AD/RD pathologies**

| Category | Phenotype | $h^2$ | SE | CI | P-value |
|---|---|---|---|---|---|
| ADNC | ADNC ABC Score | 0.554 | 0.165 | 0.231, 0.877 | $7.7 \times 10^{-4}$ |
| | A Score | 0.464 | 0.242 | -0.011, 0.939 | $5.5 \times 10^{-2}$ |
| | Amyloid plaques (any) | 0.424 | 0.114 | 0.202, 0.647 | $1.9 \times 10^{-4}$ |
| | C Score (CERAD) | 0.242 | 0.059 | 0.126, 0.358 | $4.4 \times 10^{-5}$ |
| | Thal Phase | 0.788 | 0.219 | 0.358, 1.218 | $3.3 \times 10^{-4}$ |
| | B Score | 0.253 | 0.068 | 0.12, 0.386 | $1.9 \times 10^{-4}$ |
| | Braak Stage for AD | 0.192 | 0.054 | 0.086, 0.298 | $4.0 \times 10^{-4}$ |
| CBVD | Arteriolosclerosis severity | 0.164 | 0.06 | 0.046, 0.281 | $6.5 \times 10^{-3}$ |
| | Arteriolosclerosis (any) | 0.163 | 0.075 | 0.017, 0.309 | $2.9 \times 10^{-2}$ |
| | Atherosclerosis severity | 0.113 | 0.053 | 0.01, 0.217 | $3.2 \times 10^{-2}$ |
| | Atherosclerosis (any) | 0.189 | 0.073 | 0.046, 0.332 | $9.5 \times 10^{-3}$ |
| | CAA severity | 0.103 | 0.051 | 0.003, 0.203 | $4.3 \times 10^{-2}$ |
| | CAA (any) | 0.256 | 0.058 | 0.143, 0.37 | $9.6 \times 10^{-6}$ |
| | Any CBVD | 0.202 | 0.127 | -0.047, 0.451 | $1.1 \times 10^{-1}$ |
| VBI | Hemorrhages | 0.251 | 0.203 | -0.147, 0.649 | $2.2 \times 10^{-1}$ |
| | Infarcts/lacunes | 0.131 | 0.065 | 0.005, 0.258 | $4.2 \times 10^{-2}$ |
| | Microinfarcts | 0.111 | 0.069 | -0.023, 0.246 | $1.1 \times 10^{-1}$ |
| | VBI (any) | 0.046 | 0.045 | -0.043, 0.134 | $3.1 \times 10^{-1}$ |
| | WMR severity | -0.1 | 0.278 | -0.644, 0.444 | $7.2 \times 10^{-1}$ |
| | WMR (any) | 0.212 | 0.103 | 0.01, 0.414 | $4.0 \times 10^{-2}$ |
| LBD | LBD (any/none) | 0.131 | 0.045 | 0.043, 0.219 | $3.6 \times 10^{-3}$ |
| | LBD (5 groups) | 0.15 | 0.055 | 0.042, 0.259 | $6.6 \times 10^{-3}$ |
| | LBD (3 groups) | 0.046 | 0.047 | -0.047, 0.139 | $3.3 \times 10^{-1}$ |
| TDP-43 | TDP-43 severity | 0.205 | 0.151 | -0.092, 0.502 | $1.8 \times 10^{-1}$ |
| | TDP-43 (any) | 0.41 | 0.128 | 0.159, 0.66 | $1.4 \times 10^{-3}$ |
| HS | HS | 0.123 | 0.083 | -0.04, 0.285 | $1.4 \times 10^{-1}$ |

Analyses used Model 2 throughout, excluding *APOE* region; liability scale; intercept constrained to 1. Shaded p-values indicate significance ($p < 0.05$).

Abbreviations: **AD**, Alzheimer's disease; **ADNC**, AD neuropathologic change; **CAA**, cerebral amyloid angiopathy; **CI**, confidence interval; **CERAD**, Consortium to Establish a Registry for AD neuritic plaque score; **CBVD**, cerebrovascular disease; **HS**, hippocampal sclerosis; **LBD**, Lewy body disease; **SE**, standard error; **TDP-43**, TAR DNA-binding protein 43; **VBI**, vascular brain injury; **WMR**, white matter rarefaction

**Table S10. Estimated SNP-based genetic correlation (shared heritability) across between neuropathological**

Genetic correlations only calculated when individual SNP heritabilities were significantly greater than zero (i.e

| Phenotyp1 | Phenotype2 | rg | rg (CI) | se | p |
|---|---|---|---|---|---|
| PD Braak | PD Braak (3 cat) | 1.04 | (0.91,1.16) | 0.07 | 0.000 |
| CAA | CAA (any/none) | 0.80 | (0.43,1.16) | 0.19 | 0.000 |
| AD Braak | B Score | 1.09 | (0.29,1.89) | 0.41 | 0.007 |
| CERAD (C Score) | AP (any/none) | 0.87 | (0.18,1.55) | 0.35 | 0.013 |
| AP (any/none) | CAA (any/none) | 0.98 | (0.05,1.92) | 0.48 | 0.040 |
| AP (any/none) | CAA | 1.06 | (0.03,2.08) | 0.52 | 0.044 |
| Atherosclerosis | Atherosclerosis (any/none) | 1.42 | (0.02,2.82) | 0.71 | 0.047 |
| CERAD (C Score) | CAA (any/none) | 1.68 | (-0.23,3.58) | 0.97 | 0.084 |
| CERAD (C Score) | CAA | 1.45 | (-0.39,3.28) | 0.94 | 0.122 |
| CERAD (C Score) | B Score | 1.70 | (-0.69,4.08) | 1.22 | 0.163 |
| AP (any/none) | PD Braak | 1.14 | (-0.78,3.07) | 0.98 | 0.245 |
| TDP-43 (3 cat) | CAA | -2.31 | (-6.24,1.62) | 2.01 | 0.250 |
| TDP-43 (any/none) | TDP-43 (3 cat) | 1.65 | (-1.29,4.58) | 1.50 | 0.271 |
| CERAD (C Score) | AD Braak | 2.11 | (-1.71,5.93) | 1.95 | 0.279 |
| AP (any/none) | PD Braak (3 cat) | 1.19 | (-0.98,3.36) | 1.11 | 0.284 |
| TDP-43 (3 cat) | CAA (any/none) | -1.55 | (-4.4,1.3) | 1.45 | 0.285 |
| B Score | CAA (any/none) | 1.14 | (-1,3.28) | 1.09 | 0.295 |
| AP (any/none) | AD Braak | 1.11 | (-1.03,3.24) | 1.09 | 0.310 |
| Arteriolosclerosis | Arteriolosclerosis (any/none) | 1.10 | (-1.04,3.25) | 1.09 | 0.314 |
| B Score | CAA | 1.16 | (-1.11,3.44) | 1.16 | 0.315 |
| AP (any/none) | B Score | 0.68 | (-0.65,2.01) | 0.68 | 0.317 |
| AP (any/none) | Atherosclerosis | -0.50 | (-1.5,0.49) | 0.51 | 0.321 |
| Atherosclerosis | CAA (any/none) | -0.50 | (-1.56,0.55) | 0.54 | 0.352 |
| CERAD (C Score) | PD Braak | 1.48 | (-1.87,4.83) | 1.71 | 0.387 |
| AD Braak | CAA (any/none) | 1.05 | (-1.43,3.53) | 1.27 | 0.408 |
| PD Braak | TDP-43 (3 cat) | -2.16 | (-7.31,2.98) | 2.62 | 0.410 |
| CERAD (C Score) | Atherosclerosis (any/none) | -1.18 | (-4.02,1.65) | 1.45 | 0.413 |
| CERAD (C Score) | PD Braak (3 cat) | 1.51 | (-2.1,5.11) | 1.84 | 0.413 |
| TDP-43 (3 cat) | Atherosclerosis (any/none) | -1.65 | (-5.67,2.36) | 2.05 | 0.420 |
| Atherosclerosis | CAA | -0.38 | (-1.33,0.57) | 0.49 | 0.436 |
| PD Braak (3 cat) | TDP-43 (3 cat) | -2.20 | (-7.82,3.42) | 2.87 | 0.443 |
| Atherosclerosis (any/none) | Arteriolosclerosis | 1.19 | (-1.95,4.33) | 1.60 | 0.456 |
| AP (any/none) | TDP-43 (3 cat) | -0.56 | (-2.02,0.91) | 0.75 | 0.459 |
| AD Braak | TDP-43 (3 cat) | -1.47 | (-5.4,2.47) | 2.01 | 0.465 |
| Atherosclerosis | Arteriolosclerosis | 0.49 | (-0.85,1.83) | 0.69 | 0.475 |
| B Score | PD Braak | 1.00 | (-1.91,3.91) | 1.49 | 0.500 |
| Atherosclerosis (any/none) | CAA (any/none) | -0.65 | (-2.54,1.25) | 0.97 | 0.505 |
| PD Braak (3 cat) | Atherosclerosis (any/none) | 0.70 | (-1.38,2.78) | 1.06 | 0.511 |
| TDP-43 (any/none) | CAA | -2.26 | (-9.23,4.72) | 3.56 | 0.526 |
| CERAD (C Score) | WMR (any/none) | 1.31 | (-2.75,5.38) | 2.07 | 0.527 |
| B Score | PD Braak (3 cat) | 0.94 | (-1.98,3.86) | 1.49 | 0.528 |
| B Score | TDP-43 (3 cat) | -1.04 | (-4.32,2.24) | 1.67 | 0.534 |
| B Score | Arteriolosclerosis | -1.08 | (-4.49,2.33) | 1.74 | 0.536 |

| | | | | | |
|---|---|---|---|---|---|
| TDP-43 (any/none) | CAA (any/none) | -1.69 | (-7.12,3.74) | 2.77 | 0.541 |
| B Score | Atherosclerosis | -0.82 | (-3.56,1.91) | 1.40 | 0.555 |
| PD Braak | Atherosclerosis (any/none) | 0.53 | (-1.26,2.33) | 0.91 | 0.559 |
| AD Braak | Arteriolosclerosis | -1.26 | (-5.55,3.02) | 2.18 | 0.563 |
| AD Braak | CAA | 0.56 | (-1.34,2.45) | 0.97 | 0.565 |
| B Score | TDP-43 (any/none) | -3.42 | (-15.07,8.24) | 5.95 | 0.566 |
| CERAD (C Score) | Atherosclerosis | -0.41 | (-1.8,0.99) | 0.71 | 0.568 |
| Atherosclerosis (any/none) | CAA | -0.47 | (-2.08,1.15) | 0.82 | 0.570 |
| TDP-43 (any/none) | Atherosclerosis (any/none) | -2.44 | (-11.06,6.18) | 4.40 | 0.579 |
| PD Braak | TDP-43 (any/none) | -2.50 | (-11.91,6.92) | 4.80 | 0.603 |
| AD Braak | TDP-43 (any/none) | -3.11 | (-15.11,8.88) | 6.12 | 0.611 |
| PD Braak (3 cat) | TDP-43 (any/none) | -3.01 | (-14.78,8.77) | 6.01 | 0.617 |
| WMR (any/none) | Atherosclerosis | -0.72 | (-3.54,2.1) | 1.44 | 0.617 |
| B Score | Atherosclerosis (any/none) | -0.79 | (-4.03,2.44) | 1.65 | 0.630 |
| TDP-43 (any/none) | Atherosclerosis | -0.68 | (-3.46,2.09) | 1.42 | 0.630 |
| WMR (any/none) | CAA (any/none) | -0.63 | (-3.35,2.09) | 1.39 | 0.652 |
| PD Braak | Arteriolosclerosis (any/none) | 2.23 | (-7.64,12.1) | 5.04 | 0.658 |
| AP (any/none) | TDP-43 (any/none) | -0.47 | (-2.54,1.61) | 1.06 | 0.659 |
| WMR (any/none) | Arteriolosclerosis | -2.14 | (-11.95,7.67) | 5.00 | 0.669 |
| PD Braak (3 cat) | WMR (any/none) | 0.75 | (-2.72,4.21) | 1.77 | 0.673 |
| PD Braak | WMR (any/none) | 0.69 | (-2.51,3.89) | 1.63 | 0.674 |
| TDP-43 (3 cat) | Atherosclerosis | -0.31 | (-1.78,1.15) | 0.75 | 0.674 |
| PD Braak (3 cat) | CAA | 0.27 | (-1.03,1.58) | 0.66 | 0.681 |
| WMR (any/none) | CAA | -0.60 | (-3.45,2.25) | 1.46 | 0.681 |
| AP (any/none) | Atherosclerosis (any/none) | -0.29 | (-1.66,1.09) | 0.70 | 0.682 |
| TDP-43 (any/none) | Arteriolosclerosis | -0.90 | (-5.24,3.45) | 2.22 | 0.687 |
| AD Braak | Atherosclerosis | -0.64 | (-3.74,2.47) | 1.58 | 0.688 |
| AP (any/none) | Arteriolosclerosis (any/none) | -0.42 | (-2.48,1.65) | 1.05 | 0.693 |
| PD Braak (3 cat) | Arteriolosclerosis (any/none) | 2.62 | (-10.73,15.96) | 6.81 | 0.701 |
| PD Braak (3 cat) | Arteriolosclerosis | 0.44 | (-1.83,2.72) | 1.16 | 0.702 |
| AP (any/none) | Arteriolosclerosis | -0.28 | (-1.75,1.18) | 0.75 | 0.705 |
| AD Braak | PD Braak | 0.35 | (-1.48,2.19) | 0.94 | 0.705 |
| AD Braak | PD Braak (3 cat) | 0.33 | (-1.54,2.19) | 0.95 | 0.732 |
| AD Braak | Atherosclerosis (any/none) | -0.46 | (-3.13,2.22) | 1.36 | 0.737 |
| AP (any/none) | WMR (any/none) | -0.32 | (-2.29,1.65) | 1.00 | 0.749 |
| Arteriolosclerosis | CAA (any/none) | 0.24 | (-1.22,1.69) | 0.74 | 0.750 |
| WMR (any/none) | TDP-43 (any/none) | 0.58 | (-3.25,4.42) | 1.96 | 0.765 |
| TDP-43 (3 cat) | Arteriolosclerosis (any/none) | 1.37 | (-8.25,11) | 4.91 | 0.780 |
| PD Braak | Arteriolosclerosis | 0.27 | (-1.66,2.21) | 0.99 | 0.784 |
| Atherosclerosis | Arteriolosclerosis (any/none) | -0.26 | (-2.19,1.68) | 0.99 | 0.795 |
| PD Braak | CAA (any/none) | -0.15 | (-1.49,1.18) | 0.68 | 0.822 |
| CERAD (C Score) | TDP-43 (any/none) | -0.34 | (-3.34,2.65) | 1.53 | 0.823 |
| PD Braak | Atherosclerosis | -0.12 | (-1.32,1.07) | 0.61 | 0.838 |
| PD Braak | CAA | 0.12 | (-1.1,1.34) | 0.62 | 0.844 |
| B Score | WMR (any/none) | -0.37 | (-4.17,3.43) | 1.94 | 0.848 |
| CERAD (C Score) | Arteriolosclerosis | 0.17 | (-1.66,2) | 0.93 | 0.855 |
| WMR (any/none) | Atherosclerosis (any/none) | 0.24 | (-2.37,2.85) | 1.33 | 0.856 |

| | | | | | |
|---|---|---|---|---|---|
| AD Braak | WMR (any/none) | -0.27 | (-3.65,3.11) | 1.73 | 0.875 |
| Arteriolosclerosis (any/none) | CAA (any/none) | 0.13 | (-1.7,1.95) | 0.93 | 0.892 |
| TDP-43 (3 cat) | Arteriolosclerosis | -0.13 | (-2.27,2) | 1.09 | 0.903 |
| CERAD (C Score) | Arteriolosclerosis (any/none) | 0.71 | (-11.09,12.51) | 6.02 | 0.906 |
| Atherosclerosis (any/none) | Arteriolosclerosis (any/none) | 0.14 | (-2.69,2.97) | 1.44 | 0.922 |
| PD Braak (3 cat) | CAA (any/none) | -0.07 | (-1.46,1.32) | 0.71 | 0.922 |
| CERAD (C Score) | TDP-43 (3 cat) | 0.09 | (-1.95,2.12) | 1.04 | 0.934 |
| Arteriolosclerosis | CAA | -0.07 | (-1.8,1.66) | 0.88 | 0.935 |
| AD Braak | Arteriolosclerosis (any/none) | -0.13 | (-4.14,3.89) | 2.05 | 0.951 |
| WMR (any/none) | TDP-43 (3 cat) | -0.05 | (-2.48,2.37) | 1.24 | 0.965 |
| B Score | Arteriolosclerosis (any/none) | -0.05 | (-3.23,3.14) | 1.63 | 0.977 |
| PD Braak (3 cat) | Atherosclerosis | 0.02 | (-1.24,1.27) | 0.64 | 0.979 |
| Arteriolosclerosis (any/none) | CAA | -0.02 | (-2.07,2.04) | 1.05 | 0.988 |

**l phenotypes**

e., $p < 0.05$ in Table S8). Some comparisons not included due to poor model convergence.

| h2_liab | h2_liab_se | h2_int | h2_int_se | gcov_int | gcov_int_se |
|---|---|---|---|---|---|
| 0.0484 | 0.06 | 1.00 | 0.01 | 0.99 | 0.01 |
| 0.1066 | 0.08 | 1.02 | 0.01 | 0.82 | 0.01 |
| 0.0485 | 0.08 | 1.02 | 0.01 | 0.90 | 0.01 |
| 0.2591 | 0.14 | 1.01 | 0.01 | 0.61 | 0.01 |
| 0.1042 | 0.08 | 1.02 | 0.01 | 0.24 | 0.00 |
| 0.0923 | 0.07 | 1.00 | 0.01 | 0.23 | 0.00 |
| 0.0595 | 0.09 | 1.01 | 0.01 | 0.72 | 0.01 |
| 0.1019 | 0.08 | 1.02 | 0.01 | 0.27 | 0.01 |
| 0.0893 | 0.07 | 1.00 | 0.01 | 0.24 | 0.01 |
| 0.0483 | 0.08 | 1.02 | 0.01 | 0.57 | 0.01 |
| 0.0632 | 0.07 | 1.01 | 0.01 | 0.07 | 0.00 |
| 0.0895 | 0.07 | 1.00 | 0.01 | 0.06 | 0.01 |
| 0.1535 | 0.20 | 1.00 | 0.01 | 0.86 | 0.01 |
| 0.0408 | 0.07 | 1.02 | 0.01 | 0.54 | 0.01 |
| 0.0485 | 0.06 | 1.00 | 0.01 | 0.08 | 0.01 |
| 0.1014 | 0.08 | 1.02 | 0.01 | 0.04 | 0.01 |
| 0.1010 | 0.08 | 1.02 | 0.01 | 0.26 | 0.00 |
| 0.0409 | 0.07 | 1.02 | 0.01 | 0.35 | 0.00 |
| 0.0468 | 0.11 | 1.01 | 0.01 | 0.75 | 0.01 |
| 0.0895 | 0.07 | 1.00 | 0.01 | 0.24 | 0.00 |
| 0.0494 | 0.08 | 1.02 | 0.01 | 0.38 | 0.00 |
| 0.1071 | 0.07 | 1.00 | 0.01 | 0.05 | 0.00 |
| 0.1073 | 0.08 | 1.02 | 0.01 | 0.05 | 0.00 |
| 0.0616 | 0.07 | 1.01 | 0.01 | 0.04 | 0.01 |
| 0.1012 | 0.08 | 1.02 | 0.01 | 0.26 | 0.00 |
| 0.1510 | 0.20 | 1.00 | 0.01 | 0.07 | 0.00 |
| 0.0648 | 0.09 | 1.01 | 0.01 | 0.05 | 0.00 |
| 0.0469 | 0.06 | 1.00 | 0.01 | 0.05 | 0.01 |
| 0.0549 | 0.09 | 1.01 | 0.01 | 0.04 | 0.00 |
| 0.0945 | 0.07 | 1.00 | 0.01 | 0.05 | 0.00 |
| 0.1512 | 0.20 | 1.00 | 0.01 | 0.06 | 0.00 |
| 0.0550 | 0.08 | 1.01 | 0.01 | 0.18 | 0.00 |
| 0.1554 | 0.20 | 1.00 | 0.01 | 0.05 | 0.00 |
| 0.1546 | 0.20 | 1.00 | 0.01 | 0.09 | 0.00 |
| 0.0560 | 0.08 | 1.01 | 0.01 | 0.22 | 0.00 |
| 0.0634 | 0.07 | 1.01 | 0.01 | 0.04 | 0.00 |
| 0.1070 | 0.08 | 1.02 | 0.01 | 0.09 | 0.00 |
| 0.0640 | 0.09 | 1.01 | 0.01 | 0.01 | 0.00 |
| 0.0982 | 0.07 | 1.00 | 0.01 | 0.04 | 0.01 |
| 0.0687 | 0.15 | 1.01 | 0.01 | -0.01 | 0.00 |
| 0.0482 | 0.06 | 1.00 | 0.01 | 0.06 | 0.00 |
| 0.1539 | 0.20 | 1.00 | 0.01 | 0.08 | 0.00 |
| 0.0580 | 0.08 | 1.01 | 0.01 | 0.04 | 0.00 |

| | | | | | |
|---|---|---|---|---|---|
| 0.1113 | 0.08 | 1.01 | 0.01 | 0.05 | 0.01 |
| 0.1002 | 0.07 | 1.00 | 0.01 | 0.05 | 0.00 |
| 0.0645 | 0.09 | 1.01 | 0.01 | 0.01 | 0.00 |
| 0.0566 | 0.08 | 1.01 | 0.01 | 0.04 | 0.00 |
| 0.0902 | 0.07 | 1.00 | 0.01 | 0.24 | 0.00 |
| 0.0652 | 0.16 | 1.02 | 0.01 | 0.10 | 0.00 |
| 0.1066 | 0.07 | 1.00 | 0.01 | 0.06 | 0.00 |
| 0.0943 | 0.07 | 1.00 | 0.01 | 0.07 | 0.00 |
| 0.0534 | 0.09 | 1.01 | 0.01 | 0.03 | 0.00 |
| 0.0638 | 0.16 | 1.02 | 0.01 | 0.07 | 0.00 |
| 0.0658 | 0.16 | 1.02 | 0.01 | 0.11 | 0.00 |
| 0.0615 | 0.16 | 1.02 | 0.01 | 0.07 | 0.00 |
| 0.1117 | 0.07 | 1.00 | 0.01 | 0.08 | 0.00 |
| 0.0606 | 0.09 | 1.01 | 0.01 | 0.05 | 0.00 |
| 0.0987 | 0.07 | 1.00 | 0.01 | 0.01 | 0.00 |
| 0.1025 | 0.08 | 1.02 | 0.01 | 0.05 | 0.00 |
| 0.0453 | 0.11 | 1.01 | 0.01 | 0.02 | 0.00 |
| 0.0672 | 0.16 | 1.02 | 0.01 | 0.04 | 0.00 |
| 0.0483 | 0.08 | 1.01 | 0.01 | 0.23 | 0.00 |
| 0.0604 | 0.15 | 1.01 | 0.01 | 0.05 | 0.00 |
| 0.0584 | 0.15 | 1.01 | 0.01 | 0.03 | 0.00 |
| 0.1007 | 0.07 | 1.00 | 0.01 | 0.02 | 0.00 |
| 0.0882 | 0.07 | 1.00 | 0.01 | 0.02 | 0.00 |
| 0.0875 | 0.07 | 1.00 | 0.01 | 0.05 | 0.00 |
| 0.0661 | 0.09 | 1.01 | 0.01 | 0.03 | 0.00 |
| 0.0537 | 0.08 | 1.01 | 0.01 | 0.05 | 0.01 |
| 0.1006 | 0.07 | 1.00 | 0.01 | 0.06 | 0.01 |
| 0.0460 | 0.11 | 1.01 | 0.01 | 0.02 | 0.00 |
| 0.0435 | 0.11 | 1.01 | 0.01 | 0.02 | 0.00 |
| 0.0544 | 0.08 | 1.01 | 0.01 | 0.03 | 0.00 |
| 0.0567 | 0.08 | 1.01 | 0.01 | 0.02 | 0.00 |
| 0.0641 | 0.07 | 1.01 | 0.01 | 0.03 | 0.00 |
| 0.0485 | 0.06 | 1.00 | 0.01 | 0.06 | 0.00 |
| 0.0610 | 0.09 | 1.01 | 0.01 | 0.05 | 0.00 |
| 0.0596 | 0.15 | 1.01 | 0.01 | 0.05 | 0.00 |
| 0.1050 | 0.08 | 1.02 | 0.01 | 0.07 | 0.00 |
| 0.0609 | 0.16 | 1.02 | 0.01 | 0.04 | 0.00 |
| 0.0412 | 0.11 | 1.01 | 0.01 | 0.02 | 0.00 |
| 0.0545 | 0.08 | 1.01 | 0.01 | 0.02 | 0.00 |
| 0.0469 | 0.11 | 1.01 | 0.01 | 0.15 | 0.00 |
| 0.1051 | 0.08 | 1.02 | 0.01 | 0.03 | 0.00 |
| 0.0673 | 0.16 | 1.02 | 0.01 | 0.08 | 0.00 |
| 0.1052 | 0.07 | 1.00 | 0.01 | 0.00 | 0.00 |
| 0.0889 | 0.07 | 1.00 | 0.01 | 0.02 | 0.00 |
| 0.0594 | 0.15 | 1.01 | 0.01 | 0.03 | 0.01 |
| 0.0557 | 0.08 | 1.01 | 0.01 | 0.03 | 0.00 |
| 0.0644 | 0.09 | 1.01 | 0.01 | 0.07 | 0.00 |

| | | | | | |
|---|---|---|---|---|---|
| 0.0579 | 0.15 | 1.01 | 0.01 | 0.04 | 0.00 |
| 0.1052 | 0.08 | 1.02 | 0.01 | 0.07 | 0.00 |
| 0.0544 | 0.08 | 1.01 | 0.01 | 0.05 | 0.00 |
| 0.0401 | 0.11 | 1.01 | 0.01 | 0.01 | 0.00 |
| 0.0477 | 0.11 | 1.01 | 0.01 | 0.17 | 0.00 |
| 0.1051 | 0.08 | 1.02 | 0.01 | 0.04 | 0.00 |
| 0.1574 | 0.20 | 1.00 | 0.01 | 0.07 | 0.00 |
| 0.0937 | 0.07 | 1.00 | 0.01 | 0.09 | 0.00 |
| 0.0415 | 0.11 | 1.01 | 0.01 | 0.00 | 0.00 |
| 0.1510 | 0.20 | 1.00 | 0.01 | 0.04 | 0.00 |
| 0.0419 | 0.11 | 1.01 | 0.01 | 0.01 | 0.00 |
| 0.1043 | 0.07 | 1.00 | 0.01 | 0.01 | 0.00 |
| 0.0952 | 0.07 | 1.00 | 0.01 | 0.07 | 0.00 |

**Table S11: Cell/Tissue-specific heritability enrichment p-values (GTEx, multi tissue)**

| Tissue/Cell Type | Tissue category | ADNC Score | AP (any/none) | CERAD (C Score) | Thal Phase | B Score | AD Braak | Arteriolo-sclerosis (any/none) | Arteriolo-sclerosis | Athero-sclerosis (any/none) | Athero-sclerosis | CAA (any/none) | CAA | Infarcts/lacunes | PD Braak (any/none) | PD Braak | TDP-43 (any/none) | WMR (any/none) |
|---|---|---|---|---|---|---|---|---|---|---|---|---|---|---|---|---|---|---|
| A10.165.114.830.500.750.Subcutaneous.Fat..Abdominal | Adipose | 0.4697 | 0.2904 | 0.3847 | 0.5423 | 0.6744 | 0.8709 | 0.4520 | 0.0374 | 0.6523 | 0.8824 | 0.8523 | 0.9239 | 0.4714 | 0.7783 | 0.9221 | 0.5495 | 0.7564 |
| A10.165.114.830.750.Subcutaneous.Fat | Adipose | 0.3465 | 0.3789 | 0.2860 | 0.5250 | 0.5072 | 0.8049 | 0.3441 | 0.0349 | 0.4196 | 0.6361 | 0.7052 | 0.8491 | 0.5705 | 0.7383 | 0.9227 | 0.4179 | 0.7652 |
| A11.329.114.Adipocytes | Adipose | 0.7394 | 0.4013 | 0.7952 | 0.7903 | 0.1862 | 0.1687 | 0.7010 | 0.0409 | 0.6943 | 0.8430 | 0.2168 | 0.2144 | 0.3879 | 0.6373 | 0.6978 | 0.6224 | 0.7416 |
| Adipose_Subcutaneous | Adipose | 0.2117 | 0.6014 | 0.1265 | 0.3056 | 0.5074 | 0.7184 | 0.3117 | 0.0190 | 0.9393 | 0.7856 | 0.8148 | 0.8261 | 0.3992 | 0.3251 | 0.3930 | 0.2380 | 0.5983 |
| Adipose_Visceral_(Omentum) | Adipose | 0.1905 | 0.4788 | 0.4954 | 0.6089 | 0.9079 | 0.9298 | 0.6663 | 0.1548 | 0.7111 | 0.9659 | 0.9456 | 0.9710 | 0.3087 | 0.0685 | 0.0781 | 0.1333 | 0.4092 |
| A02.835.583.443.800.800.Synovial.Fluid | Blood/Immune | 0.5386 | 0.2937 | 0.2913 | 0.5524 | 0.4105 | 0.2891 | 0.7815 | 0.7549 | 0.6812 | 0.3642 | 0.4696 | 0.6986 | 0.7992 | 0.8551 | 0.6671 | 0.3898 | 0.4007 |
| A10.549.400.Lymph.Nodes | Blood/Immune | 0.5206 | 0.7314 | 0.5829 | 0.8563 | 0.2841 | 0.3915 | 0.0658 | 0.5932 | 0.6465 | 0.1675 | 0.2432 | 0.6500 | 0.8937 | 0.9913 | 0.9820 | 0.8478 | 0.9929 |
| A10.549.Lymphoid.Tissue | Blood/Immune | 0.6393 | 0.7269 | 0.4672 | 0.7829 | 0.3890 | 0.4523 | 0.1320 | 0.4328 | 0.7476 | 0.1212 | 0.5262 | 0.5525 | 0.9093 | 0.9991 | 0.9982 | 0.7442 | 0.8469 |
| A11.118.637.555.567.562.440.Precursor.Cells..B.Lymphoid | Blood/Immune | 0.4566 | 0.4438 | 0.7175 | 0.4416 | 0.5611 | 0.5421 | 0.4623 | 0.5355 | 0.3803 | 0.4420 | 0.6055 | 0.4053 | 0.9644 | 0.8034 | 0.7600 | 0.3800 | 0.5545 |
| A11.118.637.555.567.562.B.Lymphocytes | Blood/Immune | 0.9380 | 0.7743 | 0.4272 | 0.8555 | 0.9247 | 0.9155 | 0.5243 | 0.5083 | 0.1372 | 0.1504 | 0.5736 | 0.2918 | 0.6130 | 0.8923 | 0.8093 | 0.3180 | 0.3400 |
| A11.118.637.555.567.569.200.700.T.Lymphocytes..Regulatory | Blood/Immune | 0.7097 | 0.3759 | 0.8332 | 0.7270 | 0.6737 | 0.6799 | 0.1220 | 0.1107 | 0.3324 | 0.1357 | 0.6638 | 0.7372 | 0.5996 | 0.9282 | 0.8475 | 0.7716 | 0.3600 |
| A11.118.637.555.567.569.T.Lymphocytes | Blood/Immune | 0.8934 | 0.7096 | 0.7779 | 0.9551 | 0.2261 | 0.1691 | 0.4963 | 0.4929 | 0.2398 | 0.2599 | 0.4520 | 0.5532 | 0.9578 | 0.9553 | 0.8888 | 0.2020 | 0.4271 |
| A11.118.637.Leukocytes | Blood/Immune | 0.0463 | 0.9525 | 0.1303 | 0.3108 | 0.1531 | 0.0297 | 0.3207 | 0.4330 | 0.6863 | 0.5645 | 0.6959 | 0.5592 | 0.6243 | 0.9316 | 0.9208 | 0.7848 | 0.3360 |
| A11.329.372.600.Macrophages..Alveolar | Blood/Immune | 0.1397 | 0.3922 | 0.0913 | 0.1958 | 0.2897 | 0.4264 | 0.3045 | 0.1178 | 0.4627 | 0.3401 | 0.2286 | 0.2653 | 0.3288 | 0.5889 | 0.5553 | 0.4766 | 0.3732 |
| A11.443.Erythroid.Cells | Blood/Immune | 0.2409 | 0.0675 | 0.1594 | 0.2904 | 0.4658 | 0.6916 | 0.4267 | 0.5293 | 0.7442 | 0.8042 | 0.6551 | 0.7845 | 0.8712 | 0.3482 | 0.2804 | 0.1403 | 0.2443 |
| A11.627.340.360.Granulocyte.Precursor.Cells | Blood/Immune | 0.1947 | 0.4284 | 0.5719 | 0.7741 | 0.5026 | 0.6357 | 0.9535 | 0.9837 | 0.0007 | 0.0044 | 0.8016 | 0.9671 | 0.3334 | 0.7862 | 0.6951 | 0.4240 | 0.2341 |
| A11.627.624.249.Monocyte.Macrophage.Precursor.Cells | Blood/Immune | 0.4367 | 0.1156 | 0.0432 | 0.5072 | 0.4376 | 0.4259 | 0.2006 | 0.4662 | 0.2330 | 0.3201 | 0.4852 | 0.8195 | 0.8974 | 0.4068 | 0.3127 | 0.7186 | 0.4827 |
| A11.627.635.Myeloid.Progenitor.Cells | Blood/Immune | 0.0258 | 0.0419 | 0.0046 | 0.2535 | 0.2350 | 0.3436 | 0.4246 | 0.5177 | 0.0106 | 0.1764 | 0.2067 | 0.6245 | 0.9316 | 0.4994 | 0.2282 | 0.5682 | 0.2392 |
| A11.872.378.590.635.Granulocyte.Macrophage.Progenitor.Cells | Blood/Immune | 0.1012 | 0.1701 | 0.0722 | 0.3245 | 0.4804 | 0.4191 | 0.4029 | 0.6656 | 0.0762 | 0.2364 | 0.5111 | 0.7494 | 0.3903 | 0.3856 | 0.2304 | 0.9535 | 0.4500 |
| A11.872.378.590.817.Megakaryocyte.Erythroid.Progenitor.Cells | Blood/Immune | 0.0881 | 0.0108 | 0.0122 | 0.0525 | 0.1521 | 0.5622 | 0.3000 | 0.7177 | 0.0272 | 0.2605 | 0.3734 | 0.5018 | 0.9922 | 0.9649 | 0.8433 | 0.1575 | 0.1888 |
| A11.872.378.Hematopoietic.Stem.Cells | Blood/Immune | 0.0691 | 0.1752 | 0.1691 | 0.3532 | 0.8227 | 0.8291 | 0.2393 | 0.7992 | 0.0164 | 0.3705 | 0.6200 | 0.7822 | 0.9484 | 0.6819 | 0.4390 | 0.6460 | 0.3124 |
| A15.145.229.188.Blood.Platelets | Blood/Immune | 0.0831 | 0.7310 | 0.3399 | 0.7974 | 0.7510 | 0.6518 | 0.9331 | 0.6652 | 0.8123 | 0.2434 | 0.5021 | 0.2444 | 0.1760 | 0.4075 | 0.5383 | 0.8549 | 0.5525 |
| A15.145.229.637.555.567.562.725.Plasma.Cells | Blood/Immune | 0.8289 | 0.8897 | 0.3585 | 0.8286 | 0.9441 | 0.8955 | 0.1018 | 0.1447 | 0.3152 | 0.1899 | 0.3779 | 0.2156 | 0.4181 | 0.8616 | 0.6954 | 0.2450 | 0.2036 |
| A15.145.229.637.555.567.569.200.CD4.Positive.T.Lymphocytes | Blood/Immune | 0.9161 | 0.5579 | 0.7612 | 0.8928 | 0.8145 | 0.7660 | 0.2675 | 0.2217 | 0.6119 | 0.4920 | 0.7337 | 0.5076 | 0.5605 | 0.9711 | 0.9364 | 0.4681 | 0.5007 |
| A15.145.229.637.555.Leukocytes..Mononuclear | Blood/Immune | 0.4958 | 0.6517 | 0.4490 | 0.7975 | 0.4043 | 0.3180 | 0.3666 | 0.5542 | 0.8762 | 0.8266 | 0.3605 | 0.5154 | 0.8740 | 0.9967 | 0.9960 | 0.1118 | 0.6597 |
| A15.145.229.Blood.Cells | Blood/Immune | 0.5918 | 0.6573 | 0.1689 | 0.8491 | 0.4646 | 0.1355 | 0.6539 | 0.8152 | 0.5623 | 0.5803 | 0.6224 | 0.7337 | 0.8522 | 0.9486 | 0.8836 | 0.7093 | 0.7209 |
| A15.145.300.Fetal.Blood | Blood/Immune | 0.0084 | 0.2774 | 0.2910 | 0.2392 | 0.3525 | 0.3786 | 0.6647 | 0.7040 | 0.2973 | 0.6289 | 0.5192 | 0.8709 | 0.3836 | 0.8908 | 0.8104 | 0.6338 | 0.0655 |
| A15.145.846.Serum | Blood/Immune | 0.4003 | 0.2898 | 0.1458 | 0.5370 | 0.5064 | 0.3875 | 0.7305 | 0.3086 | 0.0703 | 0.0479 | 0.6108 | 0.4580 | 0.1615 | 0.8493 | 0.7300 | 0.3772 | 0.1976 |
| A15.145.Blood | Blood/Immune | 0.6020 | 0.6533 | 0.2646 | 0.8526 | 0.4973 | 0.2827 | 0.7154 | 0.6634 | 0.6948 | 0.6019 | 0.6808 | 0.8098 | 0.8637 | 0.9866 | 0.9684 | 0.7543 | 0.1285 |
| A15.378.316.580.Monocytes | Blood/Immune | 0.0084 | 0.6670 | 0.0061 | 0.1223 | 0.0568 | 0.1056 | 0.2260 | 0.6397 | 0.2247 | 0.2691 | 0.4203 | 0.5052 | 0.0180 | 0.4913 | 0.3270 | 0.6970 | 0.1711 |
| A15.378.316.Bone.Marrow.Cells | Blood/Immune | 0.0819 | 0.2625 | 0.0351 | 0.4203 | 0.0502 | 0.0300 | 0.2445 | 0.8099 | 0.3222 | 0.2079 | 0.6181 | 0.5737 | 0.0230 | 0.4149 | 0.0951 | 0.9152 | 0.0680 |
| A15.382.490.315.583.Neutrophils | Blood/Immune | 0.1823 | 0.0039 | 0.0499 | 0.9210 | 0.2635 | 0.1246 | 0.5796 | 0.7089 | 0.3828 | 0.0671 | 0.3401 | 0.5579 | 0.1667 | 0.1667 | 0.0259 | 0.8940 | 0.0176 |
| A15.382.490.555.567.537.Killer.Cells..Natural | Blood/Immune | 0.4390 | 0.5243 | 0.7135 | 0.9078 | 0.7165 | 0.5207 | 0.8912 | 0.6084 | 0.1112 | 0.1052 | 0.9166 | 0.8843 | 0.9189 | 0.9892 | 0.9473 | 0.1660 | 0.0444 |
| A15.382.490.555.567.622.Lymphocytes..Null | Blood/Immune | 0.7926 | 0.3621 | 0.5979 | 0.7488 | 0.9394 | 0.9901 | 0.5333 | 0.0618 | 0.1236 | 0.1182 | 0.8991 | 0.5935 | 0.8536 | 0.9111 | 0.9725 | 0.6281 | 0.0153 |
| A15.382.490.555.567.Lymphocytes | Blood/Immune | 0.6122 | 0.4318 | 0.3832 | 0.9731 | 0.5099 | 0.4204 | 0.2412 | 0.4000 | 0.2976 | 0.6398 | 0.5680 | 0.2121 | 0.9350 | 0.9566 | 0.9448 | 0.2824 | 0.5333 |
| A15.382.520.604.700.Spleen | Blood/Immune | 0.7507 | 0.5765 | 0.2262 | 0.8140 | 0.1992 | 0.1315 | 0.4958 | 0.5544 | 0.8037 | 0.3240 | 0.6641 | 0.7671 | 0.5414 | 0.9259 | 0.9496 | 0.3124 | 0.8876 |
| A15.382.520.604.800.Palatine.Tonsil | Blood/Immune | 0.4500 | 0.4849 | 0.0686 | 0.2542 | 0.1378 | 0.2509 | 0.3761 | 0.5018 | 0.3779 | 0.0711 | 0.4575 | 0.5567 | 0.9998 | 0.6497 | 0.3482 | 0.8762 | 0.6234 |
| A15.382.680.Phagocytes | Blood/Immune | 0.0034 | 0.4397 | 0.0029 | 0.1880 | 0.1036 | 0.1845 | 0.1478 | 0.7404 | 0.6406 | 0.2505 | 0.1548 | 0.4367 | 0.0129 | 0.7124 | 0.4848 | 0.8340 | 0.1675 |
| A15.382.812.260.Dendritic.Cells | Blood/Immune | 0.1009 | 0.2685 | 0.0265 | 0.2654 | 0.0086 | 0.0709 | 0.8316 | 0.7424 | 0.3952 | 0.3343 | 0.5415 | 0.8151 | 0.0243 | 0.5289 | 0.6018 | 0.7651 | 0.2810 |
| A15.382.812.522.Macrophages | Blood/Immune | 0.0890 | 0.1800 | 0.0113 | 0.3429 | 0.0054 | 0.0263 | 0.3928 | 0.6434 | 0.3265 | 0.0469 | 0.6068 | 0.8072 | 0.0332 | 0.6197 | 0.6928 | 0.7145 | 0.1332 |
| A15.382.812.Mononuclear.Phagocyte.System | Blood/Immune | 0.0135 | 0.3925 | 0.0069 | 0.1190 | 0.0699 | 0.1011 | 0.2848 | 0.4558 | 0.3007 | 0.0980 | 0.3970 | 0.7255 | 0.0074 | 0.8681 | 0.8045 | 0.5895 | 0.0786 |
| A15.382.Immune.System | Blood/Immune | 0.4051 | 0.5882 | 0.2706 | 0.5711 | 0.5375 | 0.3273 | 0.6700 | 0.9512 | 0.6966 | 0.2503 | 0.1295 | 0.4081 | 0.5178 | 0.9859 | 0.9578 | 0.7074 | 0.9480 |
| Cells_EBV-transformed_lymphocytes | Blood/Immune | 0.1484 | 0.6444 | 0.0719 | 0.5931 | 0.0315 | 0.0717 | 0.5470 | 0.4996 | 0.3699 | 0.9597 | 0.6564 | 0.7832 | 0.4309 | 0.9883 | 0.9733 | 0.7534 | 0.1500 |
| Spleen | Blood/Immune | 0.0607 | 0.6258 | 0.4695 | 0.2902 | 0.1305 | 0.0398 | 0.7850 | 0.8706 | 0.2960 | 0.2887 | 0.1942 | 0.6339 | 0.2514 | 0.9961 | 0.9935 | 0.3940 | 0.9889 |
| Whole_Blood | Blood/Immune | 0.5354 | 0.1852 | 0.1239 | 0.9301 | 0.5904 | 0.2827 | 0.8105 | 0.3883 | 0.2225 | 0.0526 | 0.4202 | 0.4962 | 0.5603 | 0.4721 | 0.3555 | 0.8530 | 0.0516 |
| A07.231.114.Arteries | Cardiovascular | 0.4524 | 0.8873 | 0.7991 | 0.2170 | 0.9959 | 0.9899 | 0.3619 | 0.0413 | 0.9324 | 0.8449 | 0.7779 | 0.7895 | 0.1645 | 0.8687 | 0.7385 | 0.8269 | 0.9027 |
| A07.231.908.670.874.Umbilical.Veins | Cardiovascular | 0.6975 | 0.2813 | 0.1890 | 0.8047 | 0.5744 | 0.6187 | 0.7743 | 0.3267 | 0.0750 | 0.3400 | 0.7375 | 0.9430 | 0.4097 | 0.8929 | 0.7536 | 0.1213 | 0.2223 |
| A07.231.908.Veins | Cardiovascular | 0.8187 | 0.1657 | 0.2799 | 0.8220 | 0.5203 | 0.5308 | 0.9552 | 0.4820 | 0.1692 | 0.5125 | 0.7423 | 0.9172 | 0.3253 | 0.8420 | 0.7216 | 0.2370 | 0.3479 |
| A07.231.Blood.Vessels | Cardiovascular | 0.5344 | 0.0590 | 0.1011 | 0.5299 | 0.4914 | 0.4517 | 0.7917 | 0.0869 | 0.3939 | 0.5959 | 0.7299 | 0.8327 | 0.1238 | 0.9593 | 0.8959 | 0.1307 | 0.1036 |
| A07.541.358.100.Atrial.Appendage | Cardiovascular | 0.4515 | 0.5062 | 0.5713 | 0.9017 | 0.7042 | 0.5095 | 0.8845 | 0.2512 | 0.7048 | 0.5491 | 0.8753 | 0.3432 | 0.3795 | 0.4077 | 0.6381 | 0.0535 | 0.6425 |
| A07.541.358.Heart.Atria | Cardiovascular | 0.5991 | 0.4492 | 0.5013 | 0.9525 | 0.7212 | 0.4449 | 0.8207 | 0.3379 | 0.8069 | 0.6184 | 0.7170 | 0.1610 | 0.4695 | 0.5471 | 0.7789 | 0.0792 | 0.8312 |
| A07.541.510.110.Aortic.Valve | Cardiovascular | 0.4002 | 0.0842 | 0.1908 | 0.3730 | 0.9414 | 0.9243 | 0.3287 | 0.2299 | 0.6990 | 0.9333 | 0.8421 | 0.9047 | 0.1264 | 0.6014 | 0.7234 | 0.2823 | 0.4372 |
| A07.541.560.Heart.Ventricles | Cardiovascular | 0.7807 | 0.8345 | 0.6845 | 0.7995 | 0.3569 | 0.2777 | 0.9279 | 0.6971 | 0.5827 | 0.6984 | 0.7621 | 0.5918 | 0.8203 | 0.8913 | 0.9924 | 0.3478 | 0.9282 |
| A07.541.Heart | Cardiovascular | 0.6738 | 0.6299 | 0.5851 | 0.7695 | 0.5442 | 0.4896 | 0.9394 | 0.5853 | 0.5562 | 0.6891 | 0.6958 | 0.4977 | 0.7281 | 0.9475 | 0.9965 | 0.5355 | 0.9177 |
| Artery_Aorta | Cardiovascular | 0.3723 | 0.3842 | 0.4202 | 0.4961 | 0.3502 | 0.3124 | 0.5252 | 0.5539 | 0.3934 | 0.4306 | 0.3024 | 0.5803 | 0.4675 | 0.9757 | 0.9612 | 0.8910 | 0.7122 |
| Artery_Coronary | Cardiovascular | 0.1686 | 0.5338 | 0.4048 | 0.5689 | 0.7908 | 0.7261 | 0.8336 | 0.5539 | 0.4262 | 0.2850 | 0.0871 | 0.1264 | 0.8095 | 0.8244 | 0.8167 | 0.7404 | 0.7348 |
| Artery_Tibial | Cardiovascular | 0.0682 | 0.5671 | 0.5430 | 0.1109 | 0.4650 | 0.5574 | 0.6590 | 0.7021 | 0.3347 | 0.2726 | 0.0330 | 0.0147 | 0.1166 | 0.9878 | 0.9621 | 0.5736 | 0.4917 |
| Heart_Atrial_Appendage | Cardiovascular | 0.5740 | 0.2437 | 0.2598 | 0.7836 | 0.2380 | 0.3208 | 0.8523 | 0.7837 | 0.6702 | 0.1671 | 0.6332 | 0.5202 | 0.8425 | 0.8356 | 0.9388 | 0.5965 | 0.4944 |
| Heart_Left_Ventricle | Cardiovascular | 0.8262 | 0.4492 | 0.5311 | 0.9245 | 0.3279 | 0.4935 | 0.9954 | 0.9815 | 0.1704 | 0.4102 | 0.9701 | 0.7038 | 0.9417 | 0.7513 | 0.8676 | 0.3587 | 0.5602 |
| A08.186.211.132.810.428.200.Cerebellum | CNS | 0.1215 | 0.8109 | 0.6322 | 0.2902 | 0.0658 | 0.0433 | 0.1603 | 0.2430 | 0.5707 | 0.9721 | 0.8939 | 0.7715 | 0.3062 | 0.3279 | 0.4687 | 0.1876 | 0.3214 |
| A08.186.211.132.Brain.Stem | CNS | 0.2250 | 0.7942 | 0.5231 | 0.3057 | 0.0545 | 0.0206 | 0.2761 | 0.6239 | 0.8112 | 0.9846 | 0.8231 | 0.2450 | 0.3992 | 0.0665 | 0.1001 | 0.0469 | 0.7176 |
| A08.186.211.464.405.Hippocampus | CNS | 0.3004 | 0.4714 | 0.4466 | 0.5085 | 0.9252 | 0.5017 | 0.8092 | 0.8400 | 0.7558 | 0.9594 | 0.5183 | 0.2976 | 0.9543 | 0.0058 | 0.0047 | 0.7687 | 0.6516 |
| A08.186.211.464.710.225.Entorhinal.Cortex | CNS | 0.7284 | 0.1639 | 0.3230 | 0.8968 | 0.8301 | 0.5465 | 0.4356 | 0.6091 | 0.8121 | 0.9699 | 0.3613 | 0.0370 | 0.9214 | 0.0235 | 0.0117 | 0.7318 | 0.8997 |
| A08.186.211.464.Limbic.System | CNS | 0.4308 | 0.3349 | 0.3722 | 0.9003 | 0.7711 | 0.3834 | 0.5748 | 0.8334 | 0.8507 | 0.9796 | 0.4086 | 0.0373 | 0.9355 | 0.0097 | 0.0083 | 0.6581 | 0.8743 |
| A08.186.211.653.Mesencephalon | CNS | 0.3084 | 0.0100 | 0.0989 | 0.3850 | 0.8457 | 0.5395 | 0.9597 | 0.9910 | 0.5172 | 0.8188 | 0.1258 | 0.3728 | 0.7567 | 0.1032 | 0.1004 | 0.6407 | 0.9821 |
| A08.186.211.730.317.357.352.435.Hypothalamo.Hypophyseal.System | CNS | 0.9312 | 0.4162 | 0.3047 | 0.7675 | 0.9235 | 0.7317 | 0.4350 | 0.9047 | 0.8569 | 0.8724 | 0.9904 | 0.9987 | 0.9827 | 0.1592 | 0.4490 | 0.4053 | 0.3429 |
| A08.186.211.730.317.357.Hypothalamus | CNS | 0.8696 | 0.4543 | 0.3444 | 0.7909 | 0.9301 | 0.7912 | 0.4484 | 0.9188 | 0.8679 | 0.7617 | 0.9516 | 0.9967 | 0.9140 | 0.0424 | 0.2191 | 0.5294 | 0.9463 |

| | | | | | | | | | | | | | | | | | | |
|---|---|---|---|---|---|---|---|---|---|---|---|---|---|---|---|---|---|---|
| A08.186.211.730.317.Diencephalon | CNS | 0.8514 | 0.2924 | 0.2069 | 0.8011 | 0.6790 | 0.6344 | 0.5488 | 0.7604 | 0.2512 | 0.4866 | 0.2064 | 0.3330 | 0.7476 | 0.0297 | 0.0917 | 0.4877 | 0.9819 |
| A08.186.211.730.885.287.249.487.Corpus.Striatum | CNS | 0.1449 | 0.2553 | 0.8514 | 0.4226 | 0.8604 | 0.7906 | 0.9599 | 0.4308 | 0.1249 | 0.3781 | 0.8892 | 0.9499 | 0.5464 | 0.3992 | 0.2702 | 0.3919 | 0.7808 |
| A08.186.211.730.885.287.249.Basal.Ganglia | CNS | 0.0602 | 0.0094 | 0.2166 | 0.4749 | 0.2688 | 0.2198 | 0.8460 | 0.5026 | 0.3617 | 0.5550 | 0.6002 | 0.5820 | 0.9056 | 0.0495 | 0.0085 | 0.5232 | 0.7883 |
| A08.186.211.730.885.287.500.270.Frontal.Lobe | CNS | 0.6818 | 0.8187 | 0.8973 | 0.9329 | 0.9380 | 0.7132 | 0.7723 | 0.8584 | 0.4511 | 0.6655 | 0.2879 | 0.4012 | 0.9555 | 0.0330 | 0.0433 | 0.2907 | 0.7865 |
| A08.186.211.730.885.287.500.571.735.Visual.Cortex | CNS | 0.9433 | 0.5780 | 0.9585 | 0.8884 | 0.8830 | 0.8452 | 0.2651 | 0.6234 | 0.2584 | 0.7377 | 0.6942 | 0.2845 | 0.6937 | 0.1461 | 0.1209 | 0.3555 | 0.4777 |
| A08.186.211.730.885.287.500.670.Parietal.Lobe | CNS | 0.0484 | 0.2042 | 0.1971 | 0.3429 | 0.7764 | 0.2010 | 0.3060 | 0.6597 | 0.2493 | 0.7974 | 0.6331 | 0.1916 | 0.9901 | 0.0973 | 0.0812 | 0.4400 | 0.9405 |
| A08.186.211.730.885.287.500.Cerebral.Cortex | CNS | 0.5398 | 0.5116 | 0.3795 | 0.9459 | 0.7301 | 0.4306 | 0.7687 | 0.8626 | 0.6946 | 0.9361 | 0.4969 | 0.0598 | 0.9251 | 0.0159 | 0.0075 | 0.6926 | 0.7793 |
| A08.186.211.865.428.Metencephalon | CNS | 0.1260 | 0.8392 | 0.7799 | 0.2686 | 0.1107 | 0.0652 | 0.2302 | 0.3584 | 0.6102 | 0.9252 | 0.8962 | 0.7880 | 0.3650 | 0.2569 | 0.3478 | 0.1441 | 0.3117 |
| A08.186.211.Brain | CNS | 0.4411 | 0.3685 | 0.0691 | 0.7361 | 0.7875 | 0.4214 | 0.7078 | 0.8944 | 0.8620 | 0.8723 | 0.5073 | 0.2384 | 0.6516 | 0.0414 | 0.0125 | 0.5338 | 0.9906 |
| A09.371.729.Retina | CNS | 0.3501 | 0.9023 | 0.6030 | 0.5633 | 0.2696 | 0.2797 | 0.9925 | 0.9896 | 0.1298 | 0.2846 | 0.8190 | 0.8426 | 0.7829 | 0.1735 | 0.2524 | 0.1331 | 0.7021 |
| A11.872.653.Neural.Stem.Cells | CNS | 0.1894 | 0.0477 | 0.9546 | 0.1116 | 0.7364 | 0.6791 | 0.5327 | 0.9080 | 0.9970 | 0.8487 | 0.6548 | 0.2116 | 0.2760 | 0.2886 | 0.2117 | 0.0808 | 0.5829 |
| Brain_Amygdala | CNS | 0.7296 | 0.0503 | 0.7141 | 0.8526 | 0.8295 | 0.3641 | 0.6763 | 0.9774 | 0.2232 | 0.8763 | 0.5739 | 0.3795 | 0.6275 | 0.0020 | 0.0019 | 0.3524 | 0.2438 |
| Brain_Anterior_cingulate_cortex_(BA24) | CNS | 0.8923 | 0.0191 | 0.3977 | 0.9492 | 0.5940 | 0.1215 | 0.5750 | 0.9195 | 0.3216 | 0.8983 | 0.4750 | 0.2851 | 0.9056 | 0.0056 | 0.0011 | 0.5597 | 0.6960 |
| Brain_Caudate_(basal_ganglia) | CNS | 0.2002 | 0.0183 | 0.5040 | 0.8952 | 0.7599 | 0.3533 | 0.8729 | 0.8338 | 0.4875 | 0.8133 | 0.5212 | 0.2197 | 0.5806 | 0.0006 | 0.0002 | 0.5275 | 0.2247 |
| Brain_Cerebellar_Hemisphere | CNS | 0.1114 | 0.3538 | 0.3961 | 0.0854 | 0.3766 | 0.0983 | 0.4079 | 0.9403 | 0.1361 | 0.6724 | 0.7876 | 0.2956 | 0.9439 | 0.1465 | 0.1239 | 0.1215 | 0.6449 |
| Brain_Cerebellum | CNS | 0.6585 | 0.3227 | 0.8223 | 0.4328 | 0.6388 | 0.3168 | 0.4000 | 0.8764 | 0.2085 | 0.6459 | 0.8022 | 0.3827 | 0.8007 | 0.1531 | 0.0723 | 0.1639 | 0.8426 |
| Brain_Cortex | CNS | 0.8929 | 0.0117 | 0.3658 | 0.9232 | 0.5356 | 0.0577 | 0.6586 | 0.9622 | 0.2221 | 0.8235 | 0.3915 | 0.1258 | 0.9781 | 0.0622 | 0.0197 | 0.3648 | 0.1669 |
| Brain_Frontal_Cortex_(BA9) | CNS | 0.8306 | 0.0074 | 0.4695 | 0.8854 | 0.8892 | 0.4402 | 0.6182 | 0.9376 | 0.1613 | 0.8759 | 0.3718 | 0.3042 | 0.9400 | 0.0503 | 0.0175 | 0.6551 | 0.1793 |
| Brain_Hippocampus | CNS | 0.7487 | 0.1532 | 0.7253 | 0.8223 | 0.9127 | 0.2946 | 0.7212 | 0.9727 | 0.3922 | 0.9002 | 0.6308 | 0.3218 | 0.8344 | 0.0022 | 0.0023 | 0.6528 | 0.3755 |
| Brain_Hypothalamus | CNS | 0.7184 | 0.0093 | 0.7908 | 0.8711 | 0.8639 | 0.2252 | 0.6764 | 0.9319 | 0.3691 | 0.9238 | 0.6696 | 0.2543 | 0.5526 | 0.0039 | 0.0018 | 0.7553 | 0.1171 |
| Brain_Nucleus_accumbens_(basal_ganglia) | CNS | 0.5401 | 0.0260 | 0.6158 | 0.9315 | 0.7466 | 0.2569 | 0.7774 | 0.9428 | 0.4106 | 0.6816 | 0.5130 | 0.2161 | 0.5556 | 0.0201 | 0.0131 | 0.4947 | 0.2453 |
| Brain_Putamen_(basal_ganglia) | CNS | 0.2780 | 0.0224 | 0.7597 | 0.8119 | 0.7140 | 0.4113 | 0.7543 | 0.9035 | 0.4589 | 0.8826 | 0.6108 | 0.2712 | 0.7400 | 0.0049 | 0.0033 | 0.6964 | 0.2377 |
| Brain_Spinal_cord_(cervical_c-1) | CNS | 0.2443 | 0.1549 | 0.1469 | 0.6467 | 0.7616 | 0.3280 | 0.3580 | 0.8063 | 0.0601 | 0.7483 | 0.6924 | 0.5268 | 0.8364 | 0.0034 | 0.0011 | 0.3056 | 0.3263 |
| Brain_Substantia_nigra | CNS | 0.7520 | 0.0255 | 0.2922 | 0.9580 | 0.7274 | 0.2199 | 0.6441 | 0.9367 | 0.1406 | 0.9361 | 0.2284 | 0.3324 | 0.6777 | 0.0029 | 0.0011 | 0.1827 | 0.4900 |
| A03.556.124.369.Intestinal.Mucosa | Digestive | 0.6715 | 0.4674 | 0.9348 | 0.7571 | 0.7620 | 0.8375 | 0.3889 | 0.3388 | 0.5281 | 0.4933 | 0.0060 | 0.0493 | 0.7948 | 0.0479 | 0.0651 | 0.1914 | 0.1602 |
| A03.556.124.526.767.Rectum | Digestive | 0.0714 | 0.1714 | 0.4167 | 0.0811 | 0.9414 | 0.6496 | 0.0507 | 0.4453 | 0.3051 | 0.6835 | 0.0407 | 0.1490 | 0.9806 | 0.0823 | 0.0430 | 0.2368 | 0.4021 |
| A03.556.124.684.Intestine..Small | Digestive | 0.4872 | 0.3435 | 0.3159 | 0.2567 | 0.9389 | 0.4634 | 0.0553 | 0.5357 | 0.2039 | 0.3817 | 0.0529 | 0.1121 | 0.6049 | 0.6320 | 0.5367 | 0.5825 | 0.7982 |
| A03.556.124.Intestines | Digestive | 0.8015 | 0.4456 | 0.3955 | 0.7836 | 0.7891 | 0.5721 | 0.0269 | 0.2170 | 0.1572 | 0.5528 | 0.0568 | 0.0670 | 0.8841 | 0.0285 | 0.0259 | 0.2002 | 0.5602 |
| A03.556.249.124.Ileum | Digestive | 0.5578 | 0.3807 | 0.5722 | 0.5553 | 0.9983 | 0.9432 | 0.0170 | 0.3636 | 0.1686 | 0.1546 | 0.2470 | 0.4101 | 0.6093 | 0.2209 | 0.2703 | 0.2965 | 0.4967 |
| A03.556.249.249.209.Cecum | Digestive | 0.8273 | 0.1387 | 0.7868 | 0.7752 | 0.7976 | 0.4127 | 0.0889 | 0.7003 | 0.1914 | 0.6573 | 0.0152 | 0.0885 | 0.9151 | 0.2388 | 0.2513 | 0.4763 | 0.0160 |
| A03.556.249.249.356.668.Colon..Sigmoid | Digestive | 0.8029 | 0.3970 | 0.8843 | 0.8556 | 0.9439 | 0.8380 | 0.0016 | 0.2483 | 0.1549 | 0.3686 | 0.0494 | 0.0962 | 0.9436 | 0.2851 | 0.3446 | 0.5068 | 0.2674 |
| A03.556.249.249.356.Colon | Digestive | 0.7251 | 0.4686 | 0.4280 | 0.8246 | 0.9179 | 0.7047 | 0.0105 | 0.2679 | 0.1652 | 0.8494 | 0.0439 | 0.0682 | 0.8558 | 0.0274 | 0.0357 | 0.3771 | 0.4023 |
| A03.556.500.760.464.Parotid.Gland | Digestive | 0.7688 | 0.5796 | 0.6852 | 0.9624 | 0.5420 | 0.7180 | 0.5933 | 0.8208 | 0.7058 | 0.5466 | 0.8823 | 0.9716 | 0.2443 | 0.1386 | 0.1763 | 0.1802 | 0.7981 |
| A03.556.500.760.Salivary.Glands | Digestive | 0.7568 | 0.6797 | 0.7456 | 0.8914 | 0.5240 | 0.6910 | 0.5746 | 0.9501 | 0.7213 | 0.3214 | 0.8670 | 0.9873 | 0.4411 | 0.1388 | 0.1363 | 0.1643 | 0.9240 |
| A03.556.875.500.Esophagus | Digestive | 0.7726 | 0.1417 | 0.4315 | 0.8681 | 0.8364 | 0.5621 | 0.3421 | 0.8171 | 0.6006 | 0.2679 | 0.5420 | 0.5189 | 0.9912 | 0.0029 | 0.0050 | 0.2799 | 0.0814 |
| A03.556.875.875.Stomach | Digestive | 0.9158 | 0.0323 | 0.4103 | 0.7811 | 0.9214 | 0.6435 | 0.8849 | 0.9308 | 0.6303 | 0.5065 | 0.2185 | 0.4115 | 0.8044 | 0.6258 | 0.5594 | 0.1248 | 0.8391 |
| A03.556.875.Upper.Gastrointestinal.Tract | Digestive | 0.8980 | 0.0648 | 0.4261 | 0.7904 | 0.8643 | 0.5293 | 0.5153 | 0.6984 | 0.5596 | 0.2929 | 0.4640 | 0.5903 | 0.9645 | 0.5051 | 0.4642 | 0.1178 | 0.2705 |
| A03.556.Gastrointestinal.Tract | Digestive | 0.9580 | 0.2303 | 0.4027 | 0.8244 | 0.9322 | 0.7662 | 0.0289 | 0.1750 | 0.3684 | 0.7125 | 0.2071 | 0.1682 | 0.8923 | 0.0260 | 0.0435 | 0.3117 | 0.6781 |
| Colon_Sigmoid | Digestive | 0.3674 | 0.9567 | 0.3550 | 0.5869 | 0.6526 | 0.7267 | 0.3324 | 0.5007 | 0.5272 | 0.1381 | 0.7870 | 0.6876 | 0.3194 | 0.6533 | 0.6440 | 0.3327 | 0.1488 |
| Colon_Transverse | Digestive | 0.9542 | 0.9455 | 0.6752 | 0.9528 | 0.8357 | 0.5665 | 0.0685 | 0.6428 | 0.9372 | 0.6756 | 0.0360 | 0.1902 | 0.7606 | 0.7029 | 0.5134 | 0.1362 | 0.0476 |
| Esophagus_Gastroesophageal_Junction | Digestive | 0.0608 | 0.7766 | 0.9050 | 0.1352 | 0.9917 | 0.9748 | 0.6250 | 0.9217 | 0.4345 | 0.5222 | 0.3485 | 0.4083 | 0.6219 | 0.9971 | 0.9953 | 0.8655 | 0.3927 |
| Esophagus_Mucosa | Digestive | 0.8362 | 0.8930 | 0.5890 | 0.4872 | 0.7620 | 0.4336 | 0.0421 | 0.2228 | 0.9841 | 0.6749 | 0.8068 | 0.9090 | 0.2876 | 0.3870 | 0.1636 | 0.6469 | 0.3989 |
| Esophagus_Muscularis | Digestive | 0.0454 | 0.5577 | 0.7481 | 0.2040 | 0.9921 | 0.9447 | 0.8369 | 0.9126 | 0.1553 | 0.3135 | 0.4126 | 0.3868 | 0.5576 | 0.9978 | 0.9950 | 0.7853 | 0.1120 |
| Small_Intestine_Terminal_Ileum | Digestive | 0.8793 | 0.9172 | 0.2392 | 0.9189 | 0.7956 | 0.2838 | 0.0877 | 0.7689 | 0.7885 | 0.2593 | 0.1024 | 0.3228 | 0.6675 | 0.2726 | 0.1583 | 0.2538 | 0.6359 |
| Stomach | Digestive | 0.7707 | 0.4380 | 0.6792 | 0.7925 | 0.3874 | 0.2018 | 0.8517 | 0.9855 | 0.4967 | 0.1972 | 0.4470 | 0.9532 | 0.5454 | 0.2846 | 0.1282 | 0.4161 | 0.9528 |
| A03.620.Liver | Liver | 0.9881 | 0.9894 | 0.8508 | 0.6155 | 0.7352 | 0.4139 | 0.0868 | 0.3112 | 0.4878 | 0.2994 | 0.2770 | 0.5897 | 0.4566 | 0.8300 | 0.7386 | 0.6697 | 0.2840 |
| A11.436.348.Hepatocytes | Liver | 0.3164 | 0.7211 | 0.0882 | 0.2173 | 0.3582 | 0.2275 | 0.1984 | 0.6662 | 0.1357 | 0.1377 | 0.1097 | 0.0394 | 0.6524 | 0.3759 | 0.4749 | 0.9236 | 0.8514 |
| Liver | Liver | 0.9963 | 0.6945 | 0.7723 | 0.8078 | 0.8933 | 0.5801 | 0.9156 | 0.9310 | 0.4370 | 0.1104 | 0.2740 | 0.5265 | 0.7770 | 0.8270 | 0.5802 | 0.1443 | 0.5387 |
| A02.165.Cartilage | Musculoskeletal/Connective | 0.3701 | 0.5304 | 0.7909 | 0.4754 | 0.5786 | 0.5591 | 0.3626 | 0.4814 | 0.0360 | 0.1268 | 0.5852 | 0.7333 | 0.9685 | 0.7615 | 0.5224 | 0.5049 | 0.0364 |
| A02.633.567.850.Quadriceps.Muscle | Musculoskeletal/Connective | 0.0241 | 0.4233 | 0.3774 | 0.0804 | 0.2652 | 0.5045 | 0.9626 | 0.9962 | 0.4699 | 0.2774 | 0.6369 | 0.7500 | 0.8987 | 0.9195 | 0.9427 | 0.9693 | 0.7563 |
| A02.835.232.834.151.Cervical.Vertebrae | Musculoskeletal/Connective | 0.3293 | 0.5564 | 0.2115 | 0.7880 | 0.0319 | 0.3989 | 0.6263 | 0.3460 | 0.9898 | 0.7742 | 0.2626 | 0.1041 | 0.8540 | 0.7531 | 0.8283 | 0.9998 | 0.4302 |
| A02.835.583.443.800.Synovial.Membrane | Musculoskeletal/Connective | 0.3214 | 0.1611 | 0.6570 | 0.2275 | 0.3287 | 0.3005 | 0.2775 | 0.1502 | 0.5313 | 0.4977 | 0.7864 | 0.6799 | 0.1966 | 0.5594 | 0.6027 | 0.5907 | 0.8257 |
| A05.360.319.679.690.Myometrium | Musculoskeletal/Connective | 0.2964 | 0.4310 | 0.1140 | 0.2079 | 0.9575 | 0.9283 | 0.5672 | 0.2801 | 0.4055 | 0.3842 | 0.9827 | 0.7996 | 0.6769 | 0.2159 | 0.2238 | 0.1831 | 0.7123 |
| A10.165.450.300.425.Keloid | Musculoskeletal/Connective | 0.4485 | 0.0657 | 0.2793 | 0.1909 | 0.2890 | 0.1434 | 0.2171 | 0.2201 | 0.9272 | 0.7614 | 0.4635 | 0.8512 | 0.4204 | 0.6528 | 0.7302 | 0.3923 | 0.9167 |
| A10.165.450.300.Cicatrix | Musculoskeletal/Connective | 0.2720 | 0.1522 | 0.1336 | 0.1474 | 0.3821 | 0.3161 | 0.5197 | 0.5442 | 0.9163 | 0.7697 | 0.1750 | 0.7020 | 0.5534 | 0.7837 | 0.8228 | 0.2147 | 0.9632 |
| A10.690.467.Muscle..Smooth | Musculoskeletal/Connective | 0.2268 | 0.2705 | 0.1764 | 0.2748 | 0.0995 | 0.1662 | 0.5006 | 0.0966 | 0.1998 | 0.4769 | 0.1363 | 0.1890 | 0.4455 | 0.8580 | 0.7849 | 0.7257 | 0.5228 |
| A10.690.Muscles | Musculoskeletal/Connective | 0.0131 | 0.7572 | 0.6453 | 0.2821 | 0.2276 | 0.3764 | 0.8102 | 0.9320 | 0.6294 | 0.4496 | 0.6772 | 0.6829 | 0.8686 | 0.6734 | 0.7644 | 0.8879 | 0.5549 |
| A11.329.171.Chondrocytes | Musculoskeletal/Connective | 0.3232 | 0.0404 | 0.3347 | 0.3781 | 0.0847 | 0.1020 | 0.2159 | 0.0970 | 0.5894 | 0.8753 | 0.4246 | 0.5673 | 0.7575 | 0.9278 | 0.9583 | 0.4638 | 0.3760 |
| A11.329.228.Fibroblasts | Musculoskeletal/Connective | 0.7574 | 0.0734 | 0.2271 | 0.6853 | 0.3303 | 0.4995 | 0.1185 | 0.6520 | 0.2665 | 0.3977 | 0.5246 | 0.7284 | 0.1062 | 0.9175 | 0.9721 | 0.1420 | 0.9639 |
| A11.329.629.Osteoblasts | Musculoskeletal/Connective | 0.4491 | 0.1532 | 0.6139 | 0.1371 | 0.3284 | 0.6451 | 0.6244 | 0.1448 | 0.7163 | 0.7273 | 0.3499 | 0.1769 | 0.7201 | 0.8092 | 0.9446 | 0.4680 | 0.6579 |
| A11.329.830.Stromal.Cells | Musculoskeletal/Connective | 0.6685 | 0.2638 | 0.0125 | 0.7090 | 0.0555 | 0.0393 | 0.2086 | 0.2488 | 0.1469 | 0.2182 | 0.1751 | 0.3907 | 0.3694 | 0.9275 | 0.9651 | 0.7592 | 0.9413 |
| A11.329.Connective.Tissue.Cells | Musculoskeletal/Connective | 0.5018 | 0.3701 | 0.0063 | 0.6154 | 0.1234 | 0.0998 | 0.1169 | 0.0547 | 0.0829 | 0.4048 | 0.5913 | 0.7801 | 0.0146 | 0.6968 | 0.7804 | 0.5592 | 0.7478 |
| A11.620.520.Myocytes..Smooth.Muscle | Musculoskeletal/Connective | 0.4650 | 0.3685 | 0.4655 | 0.7865 | 0.0995 | 0.1370 | 0.3961 | 0.0893 | 0.7456 | 0.5382 | 0.8644 | 0.7811 | 0.3001 | 0.8346 | 0.9061 | 0.5133 | 0.4869 |
| Cells_Transformed_fibroblasts | Musculoskeletal/Connective | 0.7526 | 0.3424 | 0.0878 | 0.8077 | 0.1816 | 0.3881 | 0.0898 | 0.0826 | 0.7921 | 0.6230 | 0.3409 | 0.7583 | 0.4296 | 0.5381 | 0.5397 | 0.1677 | 0.5978 |
| Muscle_Skeletal | Musculoskeletal/Connective | 0.2883 | 0.9639 | 0.9991 | 0.5914 | 0.9241 | 0.8716 | 0.8753 | 0.9732 | 0.0926 | 0.1866 | 0.3857 | 0.8295 | 0.7556 | 0.7163 | 0.7744 | 0.8637 | 0.3356 |
| A04.411.Lung | Other | 0.4277 | 0.8834 | 0.5361 | 0.4008 | 0.9716 | 0.9918 | 0.1068 | 0.0810 | 0.4378 | 0.8651 | 0.7031 | 0.5556 | 0.5946 | 0.8094 | 0.9057 | 0.4233 | 0.3015 |
| A04.531.520.Nasal.Mucosa | Other | 0.6791 | 0.9221 | 0.6741 | 0.5995 | 0.9967 | 0.9703 | 0.2379 | 0.1216 | 0.5360 | 0.2008 | 0.8536 | 0.6947 | 0.8371 | 0.6935 | 0.6554 | 0.7072 | 0.2667 |
| A05.360.319.114.373.Fallopian.Tubes | Other | 0.5538 | 0.2021 | 0.2197 | 0.6133 | 0.1336 | 0.2780 | 0.2143 | 0.8400 | 0.4270 | 0.4288 | 0.3254 | 0.5675 | 0.8998 | 0.4806 | 0.4365 | 0.4727 | 0.5661 |
| A05.360.319.114.630.Ovary | Other | 0.3893 | 0.3075 | 0.2093 | 0.6760 | 0.3486 | 0.4069 | 0.7213 | 0.9013 | 0.6336 | 0.8243 | 0.4732 | 0.7785 | 0.9658 | 0.4195 | 0.4298 | 0.7749 | 0.6555 |
| A05.360.319.679.256.Cervix.Uteri | Other | 0.7984 | 0.2892 | 0.3421 | 0.8147 | 0.9936 | 0.9520 | 0.3297 | 0.5285 | 0.2676 | 0.1832 | 0.4303 | 0.8367 | 0.9071 | 0.8160 | 0.7741 | 0.8945 | 0.2806 |
| A05.360.319.679.490.Endometrium | Other | 0.8491 | 0.3926 | 0.4555 | 0.7169 | 0.6691 | 0.5682 | 0.4039 | 0.4245 | 0.3710 | 0.0757 | 0.5340 | 0.8078 | 0.4328 | 0.3553 | 0.3459 | 0.4870 | 0.9675 |
| A05.360.319.679.Uterus | Other | 0.7016 | 0.4282 | 0.3209 | 0.5826 | 0.8658 | 0.4207 | 0.2432 | 0.5898 | 0.8297 | 0.4030 | 0.1139 | 0.5593 | 0.2785 | 0.0590 | 0.0804 | 0.4747 | 0.8848 |
| A05.360.319.887.Vulva | Other | 0.9762 | 0.3082 | 0.2165 | 0.7321 | 0.6194 | 0.2870 | 0.1724 | 0.2707 | 0.4352 | 0.2074 | 0.2892 | 0.5566 | 0.8022 | 0.0125 | 0.0452 | 0.3860 | 0.7305 |

| | | | | | | | | | | | | | | | | | | |
|---|---|---|---|---|---|---|---|---|---|---|---|---|---|---|---|---|---|---|
| A05.360.319.Genitalia..Female | Other | 0.6628 | 0.1098 | 0.2302 | 0.7816 | 0.8002 | 0.5434 | 0.1896 | 0.2879 | 0.6641 | 0.5599 | 0.2725 | 0.7390 | 0.9137 | 0.3514 | 0.3585 | 0.6620 | 0.9417 |
| A05.360.444.492.362.Foreskin | Other | 0.2310 | 0.1822 | 0.0513 | 0.3063 | 0.0435 | 0.1487 | 0.2459 | 0.2764 | 0.5613 | 0.1524 | 0.3802 | 0.8426 | 0.6078 | 0.7654 | 0.7399 | 0.9233 | 0.5748 |
| A05.360.444.Genitalia..Male | Other | 0.2903 | 0.7395 | 0.0486 | 0.5539 | 0.7172 | 0.9097 | 0.7772 | 0.4955 | 0.7185 | 0.8114 | 0.8911 | 0.9861 | 0.9196 | 0.4498 | 0.4140 | 0.7094 | 0.9626 |
| A05.360.490.Germ.Cells | Other | 0.2345 | 0.9124 | 0.1654 | 0.1192 | 0.5236 | 0.6174 | 0.1626 | 0.3491 | 0.0288 | 0.0033 | 0.3211 | 0.3598 | 0.4859 | 0.8384 | 0.9429 | 0.1516 | 0.4077 |
| A05.360.Genitalia | Other | 0.6788 | 0.6830 | 0.5578 | 0.9619 | 0.7487 | 0.7951 | 0.7081 | 0.6261 | 0.8307 | 0.5575 | 0.3893 | 0.9037 | 0.9228 | 0.1939 | 0.2428 | 0.8979 | 0.9072 |
| A05.810.453.324.Kidney.Cortex | Other | 0.2073 | 0.0578 | 0.0104 | 0.0048 | 0.0992 | 0.1932 | 0.4931 | 0.2834 | 0.3677 | 0.1967 | 0.0023 | 0.0095 | 0.6723 | 0.9747 | 0.7783 | 0.5901 | 0.2485 |
| A05.810.453.Kidney | Other | 0.2852 | 0.8377 | 0.7949 | 0.3153 | 0.4100 | 0.5063 | 0.2737 | 0.1037 | 0.4931 | 0.5246 | 0.3098 | 0.2420 | 0.1215 | 0.8380 | 0.7318 | 0.4015 | 0.7932 |
| A05.810.890.Urinary.Bladder | Other | 0.8651 | 0.2511 | 0.6059 | 0.8642 | 0.9226 | 0.6025 | 0.5859 | 0.7812 | 0.9150 | 0.1941 | 0.4474 | 0.1708 | 0.2334 | 0.3097 | 0.2346 | 0.5075 | 0.6875 |
| A06.407.071.140.Adrenal.Cortex | Other | 0.3542 | 0.5101 | 0.6258 | 0.4386 | 0.8133 | 0.7591 | 0.1049 | 0.0066 | 0.7712 | 0.6129 | 0.9299 | 0.5819 | 0.7377 | 0.8109 | 0.8339 | 0.2853 | 0.9890 |
| A06.407.071.Adrenal.Glands | Other | 0.3107 | 0.2720 | 0.6737 | 0.4466 | 0.9316 | 0.8699 | 0.1456 | 0.0445 | 0.8938 | 0.7534 | 0.7956 | 0.7374 | 0.7206 | 0.7253 | 0.7480 | 0.2011 | 0.9960 |
| A06.407.312.782.Testis | Other | 0.2541 | 0.4796 | 0.4289 | 0.3189 | 0.4914 | 0.6724 | 0.4373 | 0.2712 | 0.3238 | 0.0313 | 0.9776 | 0.9853 | 0.4381 | 0.9284 | 0.9497 | 0.9913 | 0.2342 |
| A06.407.312.Gonads | Other | 0.4144 | 0.5879 | 0.3146 | 0.5518 | 0.3578 | 0.4767 | 0.5917 | 0.9060 | 0.5647 | 0.7603 | 0.4278 | 0.7157 | 0.9171 | 0.3937 | 0.4392 | 0.8175 | 0.4651 |
| A06.407.900.Thyroid.Gland | Other | 0.1564 | 0.5375 | 0.7590 | 0.8177 | 0.4747 | 0.5768 | 0.3345 | 0.1804 | 0.6624 | 0.6575 | 0.9674 | 0.8698 | 0.0773 | 0.1097 | 0.1835 | 0.5010 | 0.8350 |
| A06.407.Endocrine.Glands | Other | 0.1574 | 0.3820 | 0.3383 | 0.4804 | 0.2884 | 0.2545 | 0.1132 | 0.5608 | 0.3718 | 0.7072 | 0.4776 | 0.6523 | 0.9307 | 0.2279 | 0.1957 | 0.4293 | 0.7654 |
| A09.371.Eye | Other | 0.6992 | 0.5105 | 0.9728 | 0.6897 | 0.3657 | 0.2113 | 0.2007 | 0.6825 | 0.6067 | 0.8170 | 0.2295 | 0.3326 | 0.4001 | 0.6161 | 0.8123 | 0.4748 | 0.6178 |
| A10.272.497.Epidermis | Other | 0.9617 | 0.3967 | 0.1509 | 0.8485 | 0.4625 | 0.4067 | 0.2529 | 0.5153 | 0.2775 | 0.1946 | 0.9111 | 0.5243 | 0.5995 | 0.7460 | 0.8589 | 0.4469 | 0.3380 |
| A10.272.Epithelium | Other | 0.7157 | 0.9767 | 0.5726 | 0.7071 | 0.9200 | 0.8661 | 0.4859 | 0.1067 | 0.6764 | 0.2108 | 0.7952 | 0.6120 | 0.9491 | 0.6671 | 0.6554 | 0.7985 | 0.3113 |
| A10.336.707.Prostate | Other | 0.1231 | 0.6627 | 0.1308 | 0.3984 | 0.7258 | 0.9192 | 0.7535 | 0.5945 | 0.7212 | 0.8336 | 0.7163 | 0.8283 | 0.9459 | 0.4091 | 0.2326 | 0.7439 | 0.9765 |
| A10.615.284.473.Chorion | Other | 0.2648 | 0.8975 | 0.0688 | 0.3576 | 0.3495 | 0.2449 | 0.4217 | 0.2411 | 0.1446 | 0.0371 | 0.8096 | 0.6416 | 0.4707 | 0.9781 | 0.9800 | 0.4987 | 0.8917 |
| A10.615.550.599.Mouth.Mucosa | Other | 0.9127 | 0.9310 | 0.5976 | 0.6682 | 0.8649 | 0.2289 | 0.7859 | 0.7576 | 0.6303 | 0.6177 | 0.4819 | 0.5603 | 0.9893 | 0.0375 | 0.0668 | 0.9299 | 0.1258 |
| A10.615.550.Mucous.Membrane | Other | 0.9939 | 0.6920 | 0.3206 | 0.9607 | 0.9519 | 0.8522 | 0.0309 | 0.0938 | 0.8857 | 0.9814 | 0.3469 | 0.2470 | 0.7969 | 0.3365 | 0.5318 | 0.8023 | 0.1778 |
| A10.615.789.Serous.Membrane | Other | 0.2845 | 0.2251 | 0.2970 | 0.2252 | 0.7439 | 0.5946 | 0.5055 | 0.3542 | 0.5539 | 0.5647 | 0.6553 | 0.5168 | 0.4200 | 0.9782 | 0.9616 | 0.8112 | 0.7311 |
| A10.615.Membranes | Other | 0.9630 | 0.7583 | 0.3887 | 0.8784 | 0.9316 | 0.8266 | 0.0510 | 0.1960 | 0.8701 | 0.9571 | 0.4221 | 0.5081 | 0.6347 | 0.2233 | 0.4868 | 0.7178 | 0.2031 |
| A11.382.Endocrine.Cells | Other | 0.1671 | 0.5810 | 0.0942 | 0.0537 | 0.2111 | 0.1790 | 0.3715 | 0.8784 | 0.8341 | 0.5782 | 0.1716 | 0.0601 | 0.5248 | 0.9363 | 0.9426 | 0.8775 | 0.1284 |
| A11.436.275.Endothelial.Cells | Other | 0.8479 | 0.0444 | 0.1921 | 0.7721 | 0.0342 | 0.1041 | 0.7866 | 0.3603 | 0.1602 | 0.2424 | 0.5299 | 0.8803 | 0.3915 | 0.9131 | 0.7684 | 0.0059 | 0.1649 |
| A11.436.294.064.Glucagon.Secreting.Cells | Other | 0.0827 | 0.2751 | 0.1474 | 0.1688 | 0.2609 | 0.6472 | 0.0877 | 0.4343 | 0.0821 | 0.4028 | 0.8930 | 0.8424 | 0.9658 | 0.7568 | 0.7754 | 0.8537 | 0.1031 |
| A11.436.329.Granulosa.Cells | Other | 0.1408 | 0.9400 | 0.2574 | 0.0308 | 0.6342 | 0.3413 | 0.2241 | 0.4471 | 0.6123 | 0.4548 | 0.0740 | 0.1802 | 0.6655 | 0.9289 | 0.9649 | 0.7089 | 0.2280 |
| A11.436.397.Keratinocytes | Other | 0.7796 | 0.5884 | 0.0589 | 0.2136 | 0.4028 | 0.0871 | 0.2465 | 0.3308 | 0.7192 | 0.0330 | 0.3032 | 0.9084 | 0.8008 | 0.2555 | 0.3940 | 0.1499 | 0.4017 |
| A11.436.Epithelial.Cells | Other | 0.8373 | 0.9487 | 0.2953 | 0.7245 | 0.8934 | 0.9280 | 0.2930 | 0.1750 | 0.6231 | 0.3418 | 0.9161 | 0.8204 | 0.9971 | 0.4787 | 0.3907 | 0.2587 | 0.3038 |
| A11.497.497.600.Oocytes | Other | 0.7297 | 0.8436 | 0.2062 | 0.4634 | 0.6263 | 0.5176 | 0.2995 | 0.4171 | 0.0886 | 0.0048 | 0.2665 | 0.5137 | 0.5404 | 0.8669 | 0.8966 | 0.3218 | 0.4641 |
| A11.872.040.Adult.Stem.Cells | Other | 0.2081 | 0.2958 | 0.5361 | 0.6606 | 0.9539 | 0.9583 | 0.8375 | 0.4148 | 0.7748 | 0.4240 | 0.5926 | 0.6659 | 0.9834 | 0.8848 | 0.6668 | 0.0669 | 0.0813 |
| A11.872.190.260.Embryoid.Bodies | Other | 0.0844 | 0.3578 | 0.2940 | 0.8195 | 0.5972 | 0.5694 | 0.8034 | 0.5125 | 0.8908 | 0.9774 | 0.4042 | 0.8383 | 0.7030 | 0.3908 | 0.3876 | 0.8954 | 0.3736 |
| A11.872.190.Embryonic.Stem.Cells | Other | 0.1725 | 0.0951 | 0.0377 | 0.6922 | 0.6990 | 0.7600 | 0.5331 | 0.3706 | 0.6793 | 0.3581 | 0.8375 | 0.4542 | 0.9238 | 0.7358 | 0.5830 | 0.3776 | 0.7391 |
| A11.872.580.Mesenchymal.Stem.Cells | Other | 0.4803 | 0.8023 | 0.3561 | 0.6911 | 0.1335 | 0.4041 | 0.6983 | 0.2501 | 0.8181 | 0.6409 | 0.4260 | 0.5435 | 0.0368 | 0.9808 | 0.9983 | 0.6326 | 0.6552 |
| A11.872.700.500.Induced.Pluripotent.Stem.Cells | Other | 0.3920 | 0.2773 | 0.2986 | 0.7155 | 0.9600 | 0.9827 | 0.8413 | 0.4337 | 0.8799 | 0.4263 | 0.5761 | 0.7444 | 0.9995 | 0.8844 | 0.6704 | 0.3001 | 0.2955 |
| A11.872.Stem.Cells | Other | 0.0522 | 0.1290 | 0.0575 | 0.2916 | 0.1281 | 0.3984 | 0.3522 | 0.5969 | 0.4659 | 0.5810 | 0.1970 | 0.1549 | 0.8053 | 0.9823 | 0.9475 | 0.6969 | 0.0332 |
| A14.549.167.646.Periodontium | Other | 0.9389 | 0.6496 | 0.1803 | 0.6170 | 0.6841 | 0.0708 | 0.5037 | 0.1253 | 0.4575 | 0.0998 | 0.1781 | 0.6366 | 0.6108 | 0.0978 | 0.1968 | 0.4017 | 0.6134 |
| A14.549.167.Dentition | Other | 0.5685 | 0.4853 | 0.1645 | 0.5653 | 0.6444 | 0.1247 | 0.7396 | 0.4542 | 0.9304 | 0.6696 | 0.0861 | 0.7824 | 0.4184 | 0.3204 | 0.3770 | 0.6054 | 0.9080 |
| A14.549.885.Tongue | Other | 0.9867 | 0.2005 | 0.6556 | 0.9757 | 0.7510 | 0.7769 | 0.3034 | 0.2771 | 0.4544 | 0.0089 | 0.9798 | 0.9422 | 0.1121 | 0.6195 | 0.5780 | 0.5376 | 0.4443 |
| A14.549.Mouth | Other | 0.8702 | 0.8888 | 0.4163 | 0.5405 | 0.6765 | 0.0813 | 0.8226 | 0.7225 | 0.7634 | 0.4458 | 0.7897 | 0.6227 | 0.7074 | 0.0339 | 0.0686 | 0.8365 | 0.5485 |
| A14.724.557.Nasopharynx | Other | 0.5911 | 0.5671 | 0.6852 | 0.4027 | 0.5396 | 0.4949 | 0.2860 | 0.3870 | 0.9262 | 0.6856 | 0.2163 | 0.2041 | 0.2865 | 0.4524 | 0.6468 | 0.7233 | 0.9607 |
| A14.724.Pharynx | Other | 0.8166 | 0.6852 | 0.4968 | 0.4602 | 0.5111 | 0.5174 | 0.3800 | 0.5215 | 0.9628 | 0.5493 | 0.6314 | 0.6124 | 0.6498 | 0.4436 | 0.4158 | 0.7998 | 0.9963 |
| A17.815.Skin | Other | 0.8633 | 0.2357 | 0.0336 | 0.7714 | 0.0399 | 0.0128 | 0.4751 | 0.6484 | 0.9658 | 0.8467 | 0.6511 | 0.7125 | 0.9907 | 0.0969 | 0.2312 | 0.7860 | 0.5311 |
| Adrenal_Gland | Other | 0.2528 | 0.7553 | 0.6957 | 0.4828 | 0.4643 | 0.4368 | 0.0225 | 0.0228 | 0.7883 | 0.9815 | 0.6717 | 0.5814 | 0.6095 | 0.7806 | 0.8955 | 0.4209 | 0.6970 |
| Bladder | Other | 0.1414 | 0.6409 | 0.4069 | 0.6767 | 0.9052 | 0.5894 | 0.7955 | 0.9074 | 0.8624 | 0.3620 | 0.2931 | 0.6584 | 0.3923 | 0.7194 | 0.6565 | 0.9750 | 0.5297 |
| Breast_Mammary_Tissue | Other | 0.5840 | 0.7209 | 0.4407 | 0.7447 | 0.2658 | 0.6856 | 0.6246 | 0.3411 | 0.9798 | 0.9764 | 0.8915 | 0.9563 | 0.8500 | 0.0166 | 0.0294 | 0.0462 | 0.8948 |
| Cervix_Ectocervix | Other | 0.8783 | 0.6218 | 0.0368 | 0.7047 | 0.2172 | 0.1524 | 0.6271 | 0.1820 | 0.5082 | 0.0567 | 0.5313 | 0.9147 | 0.8077 | 0.2190 | 0.2346 | 0.0298 | 0.8602 |
| Cervix_Endocervix | Other | 0.2085 | 0.7510 | 0.2377 | 0.1983 | 0.1868 | 0.1653 | 0.7022 | 0.7536 | 0.4093 | 0.0141 | 0.0621 | 0.3502 | 0.4131 | 0.6184 | 0.2973 | 0.1466 | 0.7617 |
| Fallopian_Tube | Other | 0.5815 | 0.7998 | 0.7368 | 0.8137 | 0.3691 | 0.3752 | 0.8296 | 0.7218 | 0.7664 | 0.3420 | 0.7783 | 0.9184 | 0.5176 | 0.3091 | 0.3109 | 0.6517 | 0.8066 |
| Kidney_Cortex | Other | 0.1950 | 0.4720 | 0.0090 | 0.6737 | 0.0171 | 0.0205 | 0.3477 | 0.4070 | 0.7986 | 0.8521 | 0.2974 | 0.7247 | 0.9542 | 0.9489 | 0.7846 | 0.5071 | 0.9288 |
| Lung | Other | 0.3808 | 0.8853 | 0.1515 | 0.7593 | 0.0974 | 0.1729 | 0.6704 | 0.1199 | 0.3048 | 0.4507 | 0.6605 | 0.4696 | 0.3132 | 0.9728 | 0.9104 | 0.8145 | 0.1597 |
| Minor_Salivary_Gland | Other | 0.9789 | 0.6169 | 0.1378 | 0.9845 | 0.4989 | 0.3055 | 0.8116 | 0.5600 | 0.2760 | 0.6338 | 0.7288 | 0.8891 | 0.6131 | 0.3779 | 0.1790 | 0.0806 | 0.6826 |
| Nerve_Tibial | Other | 0.6528 | 0.7685 | 0.1774 | 0.4191 | 0.1006 | 0.2938 | 0.5664 | 0.8578 | 0.1912 | 0.3554 | 0.8478 | 0.7115 | 0.6454 | 0.8798 | 0.8502 | 0.3170 | 0.7206 |
| Ovary | Other | 0.3026 | 0.7021 | 0.8100 | 0.5885 | 0.6179 | 0.5889 | 0.7359 | 0.8620 | 0.3161 | 0.0600 | 0.4109 | 0.4588 | 0.9676 | 0.4151 | 0.2780 | 0.8393 | 0.5301 |
| Pituitary | Other | 0.3680 | 0.4670 | 0.0920 | 0.5037 | 0.5053 | 0.6496 | 0.0158 | 0.1240 | 0.4465 | 0.8424 | 0.9965 | 0.9538 | 0.3969 | 0.1798 | 0.0769 | 0.1645 | 0.9109 |
| Prostate | Other | 0.9822 | 0.7375 | 0.1363 | 0.9920 | 0.9360 | 0.8385 | 0.7175 | 0.6609 | 0.4873 | 0.2407 | 0.3673 | 0.8864 | 0.5386 | 0.6988 | 0.5313 | 0.2763 | 0.9090 |
| Skin_Not_Sun_Exposed_(Suprapubic) | Other | 0.8659 | 0.4402 | 0.0361 | 0.9815 | 0.7686 | 0.5606 | 0.4992 | 0.6414 | 0.4833 | 0.4117 | 0.9809 | 0.9516 | 0.9145 | 0.2134 | 0.2470 | 0.2291 | 0.5785 |
| Skin_Sun_Exposed_(Lower_leg) | Other | 0.8441 | 0.6468 | 0.0089 | 0.9737 | 0.7812 | 0.5318 | 0.6440 | 0.7619 | 0.4019 | 0.4322 | 0.9464 | 0.6606 | 0.9450 | 0.2754 | 0.3484 | 0.3108 | 0.7952 |
| Testis | Other | 0.1419 | 0.1166 | 0.3641 | 0.0276 | 0.3499 | 0.4639 | 0.0450 | 0.1330 | 0.5116 | 0.3160 | 0.9550 | 0.5843 | 0.9554 | 0.9707 | 0.9704 | 0.8518 | 0.9301 |
| Thyroid | Other | 0.9010 | 0.8823 | 0.4822 | 0.9452 | 0.4501 | 0.7842 | 0.0389 | 0.2105 | 0.3517 | 0.0861 | 0.9154 | 0.8052 | 0.5221 | 0.0610 | 0.0860 | 0.2204 | 0.4300 |
| Uterus | Other | 0.8981 | 0.4310 | 0.5051 | 0.8393 | 0.4523 | 0.2547 | 0.8509 | 0.6414 | 0.1070 | 0.0349 | 0.7128 | 0.8800 | 0.9289 | 0.4204 | 0.3717 | 0.2560 | 0.3541 |
| Vagina | Other | 0.9031 | 0.4843 | 0.2547 | 0.7383 | 0.7085 | 0.2035 | 0.7128 | 0.7497 | 0.7773 | 0.2637 | 0.4888 | 0.7914 | 0.9489 | 0.3211 | 0.4063 | 0.1894 | 0.8943 |
| A03.734.414.Islets.of.Langerhans | Pancreas | 0.0512 | 0.3043 | 0.1943 | 0.1497 | 0.4365 | 0.8438 | 0.2553 | 0.3557 | 0.1382 | 0.4746 | 0.7327 | 0.4700 | 0.7689 | 0.9147 | 0.9310 | 0.8442 | 0.2295 |
| A03.734.Pancreas | Pancreas | 0.6798 | 0.9969 | 0.9855 | 0.7171 | 0.9816 | 0.9925 | 0.0884 | 0.6089 | 0.1868 | 0.8325 | 0.4178 | 0.2334 | 0.8198 | 0.3412 | 0.2600 | 0.2836 | 0.7333 |
| Pancreas | Pancreas | 0.9601 | 0.9463 | 0.9086 | 0.9374 | 0.9120 | 0.7142 | 0.5273 | 0.6663 | 0.0518 | 0.2171 | 0.3859 | 0.8114 | 0.6732 | 0.5287 | 0.5754 | 0.7240 | 0.3687 |

**Table S12: Cell/Tissue-specific heritability enrichment coefficients and p-values (GTEx, multi tissue)**
This table contains the same pvalues as Table S10, but with coefficients and in "long" format

| Phenotype | Tissue/Cell Type | Category | Coefficient | Coefficient_std_error | Coefficient_P_value |
|---|---|---|---|---|---|
| AD Braak | A10.165.114.830.500.750.Subcutaneous.Fat..Abdominal | Adipose | -1.89E-08 | 1.67E-08 | 0.870925 |
| AD Braak | A10.165.114.830.750.Subcutaneous.Fat | Adipose | -1.47E-08 | 1.72E-08 | 0.804936 |
| AD Braak | A11.329.114.Adipocytes | Adipose | 1.49E-08 | 1.55E-08 | 0.168677 |
| AD Braak | Adipose_Subcutaneous | Adipose | -8.40E-09 | 1.45E-08 | 0.718389 |
| AD Braak | Adipose_Visceral_(Omentum) | Adipose | -2.26E-08 | 1.53E-08 | 0.929759 |
| AD Braak | A02.835.583.443.800.800.Synovial.Fluid | Blood/Immune | 1.31E-08 | 2.35E-08 | 0.289084 |
| AD Braak | A10.549.400.Lymph.Nodes | Blood/Immune | 5.27E-09 | 1.92E-08 | 0.391512 |
| AD Braak | A10.549.Lymphoid.Tissue | Blood/Immune | 2.29E-09 | 1.91E-08 | 0.452274 |
| AD Braak | A11.118.637.555.567.562.440.Precursor.Cells..B.Lymphoid | Blood/Immune | -2.12E-09 | 2.00E-08 | 0.542093 |
| AD Braak | A11.118.637.555.567.562.B.Lymphocytes | Blood/Immune | -2.80E-08 | 2.03E-08 | 0.915509 |
| AD Braak | A11.118.637.555.567.569.200.700.T.Lymphocytes..Regulatory | Blood/Immune | -8.28E-09 | 1.77E-08 | 0.679923 |
| AD Braak | A11.118.637.555.567.569.T.Lymphocytes | Blood/Immune | 1.78E-08 | 1.86E-08 | 0.169055 |
| AD Braak | A11.118.637.Leukocytes | Blood/Immune | 4.17E-08 | 2.21E-08 | 0.029708 |
| AD Braak | A11.329.372.600.Macrophages..Alveolar | Blood/Immune | 3.50E-09 | 1.89E-08 | 0.426406 |
| AD Braak | A11.443.Erythroid.Cells | Blood/Immune | -8.34E-09 | 1.67E-08 | 0.691552 |
| AD Braak | A11.627.340.360.Granulocyte.Precursor.Cells | Blood/Immune | -6.68E-09 | 1.92E-08 | 0.635694 |
| AD Braak | A11.627.624.249.Monocyte.Macrophage.Precursor.Cells | Blood/Immune | 3.21E-09 | 1.72E-08 | 0.425867 |
| AD Braak | A11.627.635.Myeloid.Progenitor.Cells | Blood/Immune | 7.82E-09 | 1.94E-08 | 0.343630 |
| AD Braak | A11.872.378.590.635.Granulocyte.Macrophage.Progenitor.Cells | Blood/Immune | 3.83E-09 | 1.87E-08 | 0.419090 |
| AD Braak | A11.872.378.590.817.Megakaryocyte.Erythroid.Progenitor.Cells | Blood/Immune | -2.84E-09 | 1.82E-08 | 0.562205 |
| AD Braak | A11.872.378.Hematopoietic.Stem.Cells | Blood/Immune | -1.79E-08 | 1.88E-08 | 0.829091 |
| AD Braak | A15.145.229.188.Blood.Platelets | Blood/Immune | -7.03E-09 | 1.80E-08 | 0.651848 |
| AD Braak | A15.145.229.637.555.567.562.725.Plasma.Cells | Blood/Immune | -2.30E-08 | 1.83E-08 | 0.895547 |
| AD Braak | A15.145.229.637.555.567.569.200.CD4.Positive.T.Lymphocytes | Blood/Immune | -1.35E-08 | 1.85E-08 | 0.766027 |
| AD Braak | A15.145.229.637.555.Leukocytes..Mononuclear | Blood/Immune | 9.79E-09 | 2.07E-08 | 0.318019 |
| AD Braak | A15.145.229.Blood.Cells | Blood/Immune | 2.40E-08 | 2.18E-08 | 0.135496 |
| AD Braak | A15.145.300.Fetal.Blood | Blood/Immune | 6.61E-09 | 2.14E-08 | 0.378601 |
| AD Braak | A15.145.846.Serum | Blood/Immune | 5.39E-09 | 1.89E-08 | 0.387460 |
| AD Braak | A15.145.Blood | Blood/Immune | 1.29E-08 | 2.25E-08 | 0.282657 |
| AD Braak | A15.378.316.580.Monocytes | Blood/Immune | 2.51E-08 | 2.00E-08 | 0.105552 |
| AD Braak | A15.378.316.Bone.Marrow.Cells | Blood/Immune | 4.02E-08 | 2.14E-08 | 0.029957 |
| AD Braak | A15.382.490.315.583.Neutrophils | Blood/Immune | 2.34E-08 | 2.03E-08 | 0.124591 |
| AD Braak | A15.382.490.555.567.537.Killer.Cells..Natural | Blood/Immune | -1.04E-09 | 2.01E-08 | 0.520683 |
| AD Braak | A15.382.490.555.567.622.Lymphocytes..Null | Blood/Immune | -4.43E-08 | 1.90E-08 | 0.990128 |
| AD Braak | A15.382.490.555.567.Lymphocytes | Blood/Immune | 3.94E-09 | 1.96E-08 | 0.420390 |
| AD Braak | A15.382.520.604.700.Spleen | Blood/Immune | 2.19E-08 | 1.95E-08 | 0.131540 |
| AD Braak | A15.382.520.604.800.Palatine.Tonsil | Blood/Immune | 1.25E-08 | 1.85E-08 | 0.250933 |
| AD Braak | A15.382.680.Phagocytes | Blood/Immune | 1.85E-08 | 2.06E-08 | 0.184517 |
| AD Braak | A15.382.812.260.Dendritic.Cells | Blood/Immune | 2.80E-08 | 1.91E-08 | 0.070928 |
| AD Braak | A15.382.812.522.Macrophages | Blood/Immune | 3.67E-08 | 1.89E-08 | 0.026309 |
| AD Braak | A15.382.812.Mononuclear.Phagocyte.System | Blood/Immune | 2.40E-08 | 1.88E-08 | 0.101063 |
| AD Braak | A15.382.Immune.System | Blood/Immune | 9.49E-09 | 2.12E-08 | 0.327307 |

| | | | | | |
|---|---|---|---|---|---|
| AD Braak | Cells_EBV-transformed_lymphocytes | Blood/Immune | 2.64E-08 | 1.80E-08 | 0.071682 |
| AD Braak | Spleen | Blood/Immune | 3.30E-08 | 1.88E-08 | 0.039752 |
| AD Braak | Whole_Blood | Blood/Immune | 1.20E-08 | 2.09E-08 | 0.282679 |
| AD Braak | A07.231.114.Arteries | Cardiovascular | -3.96E-08 | 1.71E-08 | 0.989873 |
| AD Braak | A07.231.908.670.874.Umbilical.Veins | Cardiovascular | -5.68E-09 | 1.88E-08 | 0.618655 |
| AD Braak | A07.231.908.Veins | Cardiovascular | -1.51E-09 | 1.95E-08 | 0.530752 |
| AD Braak | A07.231.Blood.Vessels | Cardiovascular | 2.25E-09 | 1.85E-08 | 0.451744 |
| AD Braak | A07.541.358.100.Atrial.Appendage | Cardiovascular | -3.72E-10 | 1.57E-08 | 0.509481 |
| AD Braak | A07.541.358.Heart.Atria | Cardiovascular | 2.18E-09 | 1.58E-08 | 0.444919 |
| AD Braak | A07.541.510.110.Aortic.Valve | Cardiovascular | -2.09E-08 | 1.46E-08 | 0.924344 |
| AD Braak | A07.541.560.Heart.Ventricles | Cardiovascular | 9.06E-09 | 1.54E-08 | 0.277693 |
| AD Braak | A07.541.Heart | Cardiovascular | 3.95E-10 | 1.52E-08 | 0.489627 |
| AD Braak | Artery_Aorta | Cardiovascular | 8.30E-09 | 1.70E-08 | 0.312368 |
| AD Braak | Artery_Coronary | Cardiovascular | -9.78E-09 | 1.63E-08 | 0.726088 |
| AD Braak | Artery_Tibial | Cardiovascular | -2.03E-09 | 1.41E-08 | 0.557362 |
| AD Braak | Heart_Atrial_Appendage | Cardiovascular | 6.94E-09 | 1.49E-08 | 0.320770 |
| AD Braak | Heart_Left_Ventricle | Cardiovascular | 2.40E-10 | 1.47E-08 | 0.493462 |
| AD Braak | A08.186.211.132.810.428.200.Cerebellum | CNS | 2.43E-08 | 1.41E-08 | 0.043267 |
| AD Braak | A08.186.211.132.Brain.Stem | CNS | 2.76E-08 | 1.35E-08 | 0.020571 |
| AD Braak | A08.186.211.464.405.Hippocampus | CNS | -6.10E-11 | 1.42E-08 | 0.501719 |
| AD Braak | A08.186.211.464.710.225.Entorhinal.Cortex | CNS | -1.73E-09 | 1.48E-08 | 0.546530 |
| AD Braak | A08.186.211.464.Limbic.System | CNS | 4.37E-09 | 1.47E-08 | 0.383376 |
| AD Braak | A08.186.211.653.Mesencephalon | CNS | -1.60E-09 | 1.61E-08 | 0.539511 |
| AD Braak | A08.186.211.730.317.357.352.435.Hypothalamo.Hypophyseal.System | CNS | -1.21E-08 | 1.96E-08 | 0.731702 |
| AD Braak | A08.186.211.730.317.357.Hypothalamus | CNS | -1.72E-08 | 2.12E-08 | 0.791170 |
| AD Braak | A08.186.211.730.317.Diencephalon | CNS | -7.16E-09 | 2.08E-08 | 0.634402 |
| AD Braak | A08.186.211.730.885.287.249.487.Corpus.Striatum | CNS | -1.24E-08 | 1.53E-08 | 0.790636 |
| AD Braak | A08.186.211.730.885.287.249.Basal.Ganglia | CNS | 1.24E-08 | 1.61E-08 | 0.219823 |
| AD Braak | A08.186.211.730.885.287.500.270.Frontal.Lobe | CNS | -8.88E-09 | 1.58E-08 | 0.713162 |
| AD Braak | A08.186.211.730.885.287.500.571.735.Visual.Cortex | CNS | -1.38E-08 | 1.36E-08 | 0.845163 |
| AD Braak | A08.186.211.730.885.287.500.670.Parietal.Lobe | CNS | 1.18E-08 | 1.41E-08 | 0.200956 |
| AD Braak | A08.186.211.730.885.287.500.Cerebral.Cortex | CNS | 2.56E-09 | 1.47E-08 | 0.430629 |
| AD Braak | A08.186.211.865.428.Metencephalon | CNS | 2.04E-08 | 1.35E-08 | 0.065219 |
| AD Braak | A08.186.211.Brain | CNS | 2.68E-09 | 1.35E-08 | 0.421406 |
| AD Braak | A09.371.729.Retina | CNS | 8.29E-09 | 1.42E-08 | 0.279716 |
| AD Braak | A11.872.653.Neural.Stem.Cells | CNS | -7.88E-09 | 1.69E-08 | 0.679051 |
| AD Braak | Brain_Amygdala | CNS | 4.49E-09 | 1.29E-08 | 0.364074 |
| AD Braak | Brain_Anterior_cingulate_cortex_(BA24) | CNS | 1.58E-08 | 1.36E-08 | 0.121517 |
| AD Braak | Brain_Caudate_(basal_ganglia) | CNS | 4.73E-09 | 1.26E-08 | 0.353251 |
| AD Braak | Brain_Cerebellar_Hemisphere | CNS | 1.89E-08 | 1.47E-08 | 0.098284 |
| AD Braak | Brain_Cerebellum | CNS | 6.88E-09 | 1.44E-08 | 0.316813 |
| AD Braak | Brain_Cortex | CNS | 2.01E-08 | 1.28E-08 | 0.057722 |
| AD Braak | Brain_Frontal_Cortex_(BA9) | CNS | 1.97E-09 | 1.31E-08 | 0.440153 |
| AD Braak | Brain_Hippocampus | CNS | 6.85E-09 | 1.27E-08 | 0.294604 |
| AD Braak | Brain_Hypothalamus | CNS | 9.14E-09 | 1.21E-08 | 0.225192 |
| AD Braak | Brain_Nucleus_accumbens_(basal_ganglia) | CNS | 8.26E-09 | 1.26E-08 | 0.256860 |

| | | | | | |
|---|---|---|---|---|---|
| AD Braak | Brain_Putamen_(basal_ganglia) | CNS | 2.83E-09 | 1.26E-08 | 0.411348 |
| AD Braak | Brain_Spinal_cord_(cervical_c-1) | CNS | 5.68E-09 | 1.28E-08 | 0.328034 |
| AD Braak | Brain_Substantia_nigra | CNS | 9.18E-09 | 1.19E-08 | 0.219930 |
| AD Braak | A03.556.124.369.Intestinal.Mucosa | Digestive | -1.85E-08 | 1.88E-08 | 0.837479 |
| AD Braak | A03.556.124.526.767.Rectum | Digestive | -7.70E-09 | 2.00E-08 | 0.649599 |
| AD Braak | A03.556.124.684.Intestine..Small | Digestive | 1.86E-09 | 2.03E-08 | 0.463361 |
| AD Braak | A03.556.124.Intestines | Digestive | -3.49E-09 | 1.92E-08 | 0.572113 |
| AD Braak | A03.556.249.124.Ileum | Digestive | -3.11E-08 | 1.96E-08 | 0.943178 |
| AD Braak | A03.556.249.249.209.Cecum | Digestive | 3.90E-09 | 1.77E-08 | 0.412676 |
| AD Braak | A03.556.249.249.356.668.Colon..Sigmoid | Digestive | -2.05E-08 | 2.08E-08 | 0.838035 |
| AD Braak | A03.556.249.249.356.Colon | Digestive | -1.08E-08 | 2.00E-08 | 0.704689 |
| AD Braak | A03.556.500.760.464.Parotid.Gland | Digestive | -1.08E-08 | 1.87E-08 | 0.717989 |
| AD Braak | A03.556.500.760.Salivary.Glands | Digestive | -8.91E-09 | 1.79E-08 | 0.691015 |
| AD Braak | A03.556.875.500.Esophagus | Digestive | -2.47E-09 | 1.58E-08 | 0.562106 |
| AD Braak | A03.556.875.875.Stomach | Digestive | -7.04E-09 | 1.92E-08 | 0.643474 |
| AD Braak | A03.556.875.Upper.Gastrointestinal.Tract | Digestive | -1.39E-09 | 1.89E-08 | 0.529295 |
| AD Braak | A03.556.Gastrointestinal.Tract | Digestive | -1.38E-08 | 1.89E-08 | 0.766239 |
| AD Braak | Colon_Sigmoid | Digestive | -9.28E-09 | 1.54E-08 | 0.726675 |
| AD Braak | Colon_Transverse | Digestive | -3.01E-09 | 1.80E-08 | 0.566471 |
| AD Braak | Esophagus_Gastroesophageal_Junction | Digestive | -3.00E-08 | 1.53E-08 | 0.974790 |
| AD Braak | Esophagus_Mucosa | Digestive | 2.83E-09 | 1.69E-08 | 0.433608 |
| AD Braak | Esophagus_Muscularis | Digestive | -2.39E-08 | 1.50E-08 | 0.944725 |
| AD Braak | Small_Intestine_Terminal_Ileum | Digestive | 1.04E-08 | 1.82E-08 | 0.283790 |
| AD Braak | Stomach | Digestive | 1.40E-08 | 1.68E-08 | 0.201792 |
| AD Braak | A03.620.Liver | Liver | 3.91E-09 | 1.79E-08 | 0.413869 |
| AD Braak | A11.436.348.Hepatocytes | Liver | 1.43E-08 | 1.92E-08 | 0.227461 |
| AD Braak | Liver | Liver | -3.39E-09 | 1.68E-08 | 0.580084 |
| AD Braak | A02.165.Cartilage | Musculoskeletal/Connective | -2.24E-09 | 1.51E-08 | 0.559109 |
| AD Braak | A02.633.567.850.Quadriceps.Muscle | Musculoskeletal/Connective | -2.04E-10 | 1.80E-08 | 0.504528 |
| AD Braak | A02.835.232.834.151.Cervical.Vertebrae | Musculoskeletal/Connective | 3.88E-09 | 1.51E-08 | 0.398887 |
| AD Braak | A02.835.583.443.800.Synovial.Membrane | Musculoskeletal/Connective | 8.38E-09 | 1.60E-08 | 0.300499 |
| AD Braak | A05.360.319.679.690.Myometrium | Musculoskeletal/Connective | -2.15E-08 | 1.47E-08 | 0.928309 |
| AD Braak | A10.165.450.300.425.Keloid | Musculoskeletal/Connective | 1.75E-08 | 1.64E-08 | 0.143400 |
| AD Braak | A10.165.450.300.Cicatrix | Musculoskeletal/Connective | 7.78E-09 | 1.63E-08 | 0.316149 |
| AD Braak | A10.690.467.Muscle..Smooth | Musculoskeletal/Connective | 1.76E-08 | 1.82E-08 | 0.166234 |
| AD Braak | A10.690.Muscles | Musculoskeletal/Connective | 5.58E-09 | 1.77E-08 | 0.376389 |
| AD Braak | A11.329.171.Chondrocytes | Musculoskeletal/Connective | 2.09E-08 | 1.64E-08 | 0.102002 |
| AD Braak | A11.329.228.Fibroblasts | Musculoskeletal/Connective | 2.22E-11 | 1.62E-08 | 0.499454 |
| AD Braak | A11.329.629.Osteoblasts | Musculoskeletal/Connective | -6.08E-09 | 1.63E-08 | 0.645078 |
| AD Braak | A11.329.830.Stromal.Cells | Musculoskeletal/Connective | 2.75E-08 | 1.57E-08 | 0.039263 |
| AD Braak | A11.329.Connective.Tissue.Cells | Musculoskeletal/Connective | 2.39E-08 | 1.86E-08 | 0.099838 |
| AD Braak | A11.620.520.Myocytes..Smooth.Muscle | Musculoskeletal/Connective | 2.01E-08 | 1.83E-08 | 0.136989 |
| AD Braak | Cells_Transformed_fibroblasts | Musculoskeletal/Connective | 4.39E-09 | 1.55E-08 | 0.388108 |
| AD Braak | Muscle_Skeletal | Musculoskeletal/Connective | -1.75E-08 | 1.54E-08 | 0.871579 |
| AD Braak | A04.411.Lung | Other | -4.25E-08 | 1.77E-08 | 0.991776 |
| AD Braak | A04.531.520.Nasal.Mucosa | Other | -2.93E-08 | 1.55E-08 | 0.970268 |

| | | | | | |
|---|---|---|---|---|---|
| AD Braak | A05.360.319.114.373.Fallopian.Tubes | Other | 1.06E-08 | 1.80E-08 | 0.278002 |
| AD Braak | A05.360.319.114.630.Ovary | Other | 3.91E-09 | 1.66E-08 | 0.406850 |
| AD Braak | A05.360.319.679.256.Cervix.Uteri | Other | -3.12E-08 | 1.88E-08 | 0.951951 |
| AD Braak | A05.360.319.679.490.Endometrium | Other | -2.76E-09 | 1.60E-08 | 0.568185 |
| AD Braak | A05.360.319.679.Uterus | Other | 3.17E-09 | 1.59E-08 | 0.420720 |
| AD Braak | A05.360.319.887.Vulva | Other | 9.47E-09 | 1.68E-08 | 0.286958 |
| AD Braak | A05.360.319.Genitalia..Female | Other | -1.84E-09 | 1.69E-08 | 0.543356 |
| AD Braak | A05.360.444.492.362.Foreskin | Other | 1.72E-08 | 1.66E-08 | 0.148682 |
| AD Braak | A05.360.444.Genitalia..Male | Other | -2.05E-08 | 1.53E-08 | 0.909691 |
| AD Braak | A05.360.490.Germ.Cells | Other | -4.51E-09 | 1.51E-08 | 0.617394 |
| AD Braak | A05.360.Genitalia | Other | -1.39E-08 | 1.68E-08 | 0.795124 |
| AD Braak | A05.810.453.324.Kidney.Cortex | Other | 1.36E-08 | 1.57E-08 | 0.193221 |
| AD Braak | A05.810.453.Kidney | Other | -2.54E-10 | 1.60E-08 | 0.506312 |
| AD Braak | A05.810.890.Urinary.Bladder | Other | -5.29E-09 | 2.04E-08 | 0.602499 |
| AD Braak | A06.407.071.140.Adrenal.Cortex | Other | -1.19E-08 | 1.69E-08 | 0.759126 |
| AD Braak | A06.407.071.Adrenal.Glands | Other | -1.91E-08 | 1.70E-08 | 0.869933 |
| AD Braak | A06.407.312.782.Testis | Other | -6.38E-09 | 1.43E-08 | 0.672399 |
| AD Braak | A06.407.312.Gonads | Other | 9.68E-10 | 1.66E-08 | 0.476729 |
| AD Braak | A06.407.900.Thyroid.Gland | Other | -3.04E-09 | 1.57E-08 | 0.576830 |
| AD Braak | A06.407.Endocrine.Glands | Other | 1.11E-08 | 1.69E-08 | 0.254520 |
| AD Braak | A09.371.Eye | Other | 1.52E-08 | 1.90E-08 | 0.211308 |
| AD Braak | A10.272.497.Epidermis | Other | 4.19E-09 | 1.77E-08 | 0.406682 |
| AD Braak | A10.272.Epithelium | Other | -1.72E-08 | 1.55E-08 | 0.866076 |
| AD Braak | A10.336.707.Prostate | Other | -2.15E-08 | 1.53E-08 | 0.919225 |
| AD Braak | A10.615.284.473.Chorion | Other | 1.18E-08 | 1.70E-08 | 0.244887 |
| AD Braak | A10.615.550.599.Mouth.Mucosa | Other | 1.39E-08 | 1.87E-08 | 0.228880 |
| AD Braak | A10.615.550.Mucous.Membrane | Other | -1.80E-08 | 1.73E-08 | 0.852195 |
| AD Braak | A10.615.789.Serous.Membrane | Other | -4.03E-09 | 1.68E-08 | 0.594638 |
| AD Braak | A10.615.Membranes | Other | -1.59E-08 | 1.69E-08 | 0.826637 |
| AD Braak | A11.382.Endocrine.Cells | Other | 1.64E-08 | 1.79E-08 | 0.179029 |
| AD Braak | A11.436.275.Endothelial.Cells | Other | 2.50E-08 | 1.99E-08 | 0.104135 |
| AD Braak | A11.436.294.064.Glucagon.Secreting.Cells | Other | -7.33E-09 | 1.94E-08 | 0.647187 |
| AD Braak | A11.436.329.Granulosa.Cells | Other | 7.03E-09 | 1.72E-08 | 0.341261 |
| AD Braak | A11.436.397.Keratinocytes | Other | 2.74E-08 | 2.02E-08 | 0.087143 |
| AD Braak | A11.436.Epithelial.Cells | Other | -2.46E-08 | 1.69E-08 | 0.927975 |
| AD Braak | A11.497.497.600.Oocytes | Other | -6.45E-10 | 1.46E-08 | 0.517572 |
| AD Braak | A11.872.040.Adult.Stem.Cells | Other | -2.81E-08 | 1.62E-08 | 0.958328 |
| AD Braak | A11.872.190.260.Embryoid.Bodies | Other | -2.80E-09 | 1.60E-08 | 0.569369 |
| AD Braak | A11.872.190.Embryonic.Stem.Cells | Other | -1.07E-08 | 1.51E-08 | 0.760047 |
| AD Braak | A11.872.580.Mesenchymal.Stem.Cells | Other | 3.81E-09 | 1.57E-08 | 0.404131 |
| AD Braak | A11.872.700.500.Induced.Pluripotent.Stem.Cells | Other | -3.31E-08 | 1.57E-08 | 0.982721 |
| AD Braak | A11.872.Stem.Cells | Other | 5.16E-09 | 2.00E-08 | 0.398411 |
| AD Braak | A14.549.167.646.Periodontium | Other | 3.04E-08 | 2.07E-08 | 0.070783 |
| AD Braak | A14.549.167.Dentition | Other | 2.02E-08 | 1.75E-08 | 0.124730 |
| AD Braak | A14.549.885.Tongue | Other | -1.49E-08 | 1.96E-08 | 0.776906 |
| AD Braak | A14.549.Mouth | Other | 2.46E-08 | 1.76E-08 | 0.081277 |

| | | | | | |
|---|---|---|---|---|---|
| AD Braak | A14.724.557.Nasopharynx | Other | 2.40E-10 | 1.87E-08 | 0.494885 |
| AD Braak | A14.724.Pharynx | Other | -8.30E-10 | 1.91E-08 | 0.517351 |
| AD Braak | A17.815.Skin | Other | 3.63E-08 | 1.63E-08 | 0.012776 |
| AD Braak | Adrenal_Gland | Other | 2.53E-09 | 1.59E-08 | 0.436845 |
| AD Braak | Bladder | Other | -3.18E-09 | 1.41E-08 | 0.589376 |
| AD Braak | Breast_Mammary_Tissue | Other | -7.90E-09 | 1.63E-08 | 0.685620 |
| AD Braak | Cervix_Ectocervix | Other | 1.67E-08 | 1.63E-08 | 0.152438 |
| AD Braak | Cervix_Endocervix | Other | 1.57E-08 | 1.61E-08 | 0.165256 |
| AD Braak | Fallopian_Tube | Other | 4.70E-09 | 1.48E-08 | 0.375241 |
| AD Braak | Kidney_Cortex | Other | 3.30E-08 | 1.61E-08 | 0.020543 |
| AD Braak | Lung | Other | 1.46E-08 | 1.55E-08 | 0.172899 |
| AD Braak | Minor_Salivary_Gland | Other | 8.15E-09 | 1.60E-08 | 0.305510 |
| AD Braak | Nerve_Tibial | Other | 8.39E-09 | 1.55E-08 | 0.293755 |
| AD Braak | Ovary | Other | -3.19E-09 | 1.42E-08 | 0.588872 |
| AD Braak | Pituitary | Other | -5.68E-09 | 1.48E-08 | 0.649642 |
| AD Braak | Prostate | Other | -1.66E-08 | 1.68E-08 | 0.838499 |
| AD Braak | Skin_Not_Sun_Exposed_(Suprapubic) | Other | -2.49E-09 | 1.63E-08 | 0.560615 |
| AD Braak | Skin_Sun_Exposed_(Lower_leg) | Other | -1.32E-09 | 1.65E-08 | 0.531782 |
| AD Braak | Testis | Other | 1.46E-09 | 1.61E-08 | 0.463885 |
| AD Braak | Thyroid | Other | -1.16E-08 | 1.48E-08 | 0.784241 |
| AD Braak | Uterus | Other | 1.02E-08 | 1.55E-08 | 0.254721 |
| AD Braak | Vagina | Other | 1.37E-08 | 1.66E-08 | 0.203474 |
| AD Braak | A03.734.414.Islets.of.Langerhans | Pancreas | -1.99E-08 | 1.97E-08 | 0.843783 |
| AD Braak | A03.734.Pancreas | Pancreas | -4.04E-08 | 1.66E-08 | 0.992510 |
| AD Braak | Pancreas | Pancreas | -9.34E-09 | 1.65E-08 | 0.714229 |
| ADNC Score | A10.165.114.830.500.750.Subcutaneous.Fat..Abdominal | Adipose | 4.45E-09 | 5.86E-08 | 0.469722 |
| ADNC Score | A10.165.114.830.750.Subcutaneous.Fat | Adipose | 2.30E-08 | 5.82E-08 | 0.346532 |
| ADNC Score | A11.329.114.Adipocytes | Adipose | -3.41E-08 | 5.32E-08 | 0.739399 |
| ADNC Score | Adipose_Subcutaneous | Adipose | 3.92E-08 | 4.90E-08 | 0.211690 |
| ADNC Score | Adipose_Visceral_(Omentum) | Adipose | 4.93E-08 | 5.63E-08 | 0.190490 |
| ADNC Score | A02.835.583.443.800.800.Synovial.Fluid | Blood/Immune | -6.14E-09 | 6.33E-08 | 0.538624 |
| ADNC Score | A10.549.400.Lymph.Nodes | Blood/Immune | -2.94E-09 | 5.69E-08 | 0.520610 |
| ADNC Score | A10.549.Lymphoid.Tissue | Blood/Immune | -2.01E-08 | 5.65E-08 | 0.639305 |
| ADNC Score | A11.118.637.555.567.562.440.Precursor.Cells..B.Lymphoid | Blood/Immune | 6.54E-09 | 5.99E-08 | 0.456570 |
| ADNC Score | A11.118.637.555.567.562.B.Lymphocytes | Blood/Immune | -8.53E-08 | 5.54E-08 | 0.938021 |
| ADNC Score | A11.118.637.555.567.569.200.700.T.Lymphocytes..Regulatory | Blood/Immune | -3.33E-08 | 6.02E-08 | 0.709738 |
| ADNC Score | A11.118.637.555.567.569.T.Lymphocytes | Blood/Immune | -7.23E-08 | 5.81E-08 | 0.893373 |
| ADNC Score | A11.118.637.Leukocytes | Blood/Immune | 1.03E-07 | 6.14E-08 | 0.046293 |
| ADNC Score | A11.329.372.600.Macrophages..Alveolar | Blood/Immune | 5.69E-08 | 5.26E-08 | 0.139723 |
| ADNC Score | A11.443.Erythroid.Cells | Blood/Immune | 3.75E-08 | 5.34E-08 | 0.240943 |
| ADNC Score | A11.627.340.360.Granulocyte.Precursor.Cells | Blood/Immune | 5.19E-08 | 6.03E-08 | 0.194673 |
| ADNC Score | A11.627.624.249.Monocyte.Macrophage.Precursor.Cells | Blood/Immune | 9.46E-09 | 5.93E-08 | 0.436662 |
| ADNC Score | A11.627.635.Myeloid.Progenitor.Cells | Blood/Immune | 1.19E-07 | 6.13E-08 | 0.025752 |
| ADNC Score | A11.872.378.590.635.Granulocyte.Macrophage.Progenitor.Cells | Blood/Immune | 7.46E-08 | 5.85E-08 | 0.101151 |
| ADNC Score | A11.872.378.590.817.Megakaryocyte.Erythroid.Progenitor.Cells | Blood/Immune | 7.75E-08 | 5.73E-08 | 0.088050 |
| ADNC Score | A11.872.378.Hematopoietic.Stem.Cells | Blood/Immune | 9.57E-08 | 6.46E-08 | 0.069072 |

| | | | | | |
|---|---|---|---|---|---|
| ADNC Score | A15.145.229.188.Blood.Platelets | Blood/Immune | 7.34E-08 | 5.30E-08 | 0.083099 |
| ADNC Score | A15.145.229.637.555.567.562.725.Plasma.Cells | Blood/Immune | -5.73E-08 | 6.03E-08 | 0.828895 |
| ADNC Score | A15.145.229.637.555.567.569.200.CD4.Positive.T.Lymphocytes | Blood/Immune | -8.01E-08 | 5.81E-08 | 0.916104 |
| ADNC Score | A15.145.229.637.555.Leukocytes..Mononuclear | Blood/Immune | 6.67E-10 | 6.33E-08 | 0.495800 |
| ADNC Score | A15.145.229.Blood.Cells | Blood/Immune | -1.51E-08 | 6.50E-08 | 0.591837 |
| ADNC Score | A15.145.300.Fetal.Blood | Blood/Immune | 1.60E-07 | 6.68E-08 | 0.008394 |
| ADNC Score | A15.145.846.Serum | Blood/Immune | 1.39E-08 | 5.48E-08 | 0.400278 |
| ADNC Score | A15.145.Blood | Blood/Immune | -1.66E-08 | 6.42E-08 | 0.602033 |
| ADNC Score | A15.378.316.580.Monocytes | Blood/Immune | 1.50E-07 | 6.26E-08 | 0.008431 |
| ADNC Score | A15.378.316.Bone.Marrow.Cells | Blood/Immune | 8.62E-08 | 6.19E-08 | 0.081859 |
| ADNC Score | A15.382.490.315.583.Neutrophils | Blood/Immune | 6.14E-08 | 6.77E-08 | 0.182279 |
| ADNC Score | A15.382.490.555.567.537.Killer.Cells..Natural | Blood/Immune | 9.44E-09 | 6.15E-08 | 0.439001 |
| ADNC Score | A15.382.490.555.567.622.Lymphocytes..Null | Blood/Immune | -5.09E-08 | 6.24E-08 | 0.792594 |
| ADNC Score | A15.382.490.555.567.Lymphocytes | Blood/Immune | -1.65E-08 | 5.80E-08 | 0.612240 |
| ADNC Score | A15.382.520.604.700.Spleen | Blood/Immune | -4.02E-08 | 5.94E-08 | 0.750652 |
| ADNC Score | A15.382.520.604.800.Palatine.Tonsil | Blood/Immune | 6.85E-09 | 5.45E-08 | 0.449953 |
| ADNC Score | A15.382.680.Phagocytes | Blood/Immune | 1.74E-07 | 6.44E-08 | 0.003388 |
| ADNC Score | A15.382.812.260.Dendritic.Cells | Blood/Immune | 7.53E-08 | 5.90E-08 | 0.100884 |
| ADNC Score | A15.382.812.522.Macrophages | Blood/Immune | 8.34E-08 | 6.19E-08 | 0.088996 |
| ADNC Score | A15.382.812.Mononuclear.Phagocyte.System | Blood/Immune | 1.31E-07 | 5.92E-08 | 0.013546 |
| ADNC Score | A15.382.Immune.System | Blood/Immune | 1.48E-08 | 6.17E-08 | 0.405052 |
| ADNC Score | Cells_EBV-transformed_lymphocytes | Blood/Immune | 5.84E-08 | 5.59E-08 | 0.148408 |
| ADNC Score | Spleen | Blood/Immune | 9.42E-08 | 6.08E-08 | 0.060673 |
| ADNC Score | Whole_Blood | Blood/Immune | -5.28E-09 | 5.95E-08 | 0.535358 |
| ADNC Score | A07.231.114.Arteries | Cardiovascular | 6.88E-09 | 5.75E-08 | 0.452379 |
| ADNC Score | A07.231.908.670.874.Umbilical.Veins | Cardiovascular | -2.84E-08 | 5.50E-08 | 0.697523 |
| ADNC Score | A07.231.908.Veins | Cardiovascular | -4.97E-08 | 5.46E-08 | 0.818652 |
| ADNC Score | A07.231.Blood.Vessels | Cardiovascular | -4.90E-09 | 5.67E-08 | 0.534415 |
| ADNC Score | A07.541.358.100.Atrial.Appendage | Cardiovascular | 6.27E-09 | 5.14E-08 | 0.451470 |
| ADNC Score | A07.541.358.Heart.Atria | Cardiovascular | -1.30E-08 | 5.18E-08 | 0.599112 |
| ADNC Score | A07.541.510.110.Aortic.Valve | Cardiovascular | 1.39E-08 | 5.49E-08 | 0.400237 |
| ADNC Score | A07.541.560.Heart.Ventricles | Cardiovascular | -4.26E-08 | 5.51E-08 | 0.780656 |
| ADNC Score | A07.541.Heart | Cardiovascular | -2.40E-08 | 5.33E-08 | 0.673805 |
| ADNC Score | Artery_Aorta | Cardiovascular | 1.67E-08 | 5.13E-08 | 0.372315 |
| ADNC Score | Artery_Coronary | Cardiovascular | 4.83E-08 | 5.03E-08 | 0.168574 |
| ADNC Score | Artery_Tibial | Cardiovascular | 6.09E-08 | 4.09E-08 | 0.068221 |
| ADNC Score | Heart_Atrial_Appendage | Cardiovascular | -8.99E-09 | 4.82E-08 | 0.574005 |
| ADNC Score | Heart_Left_Ventricle | Cardiovascular | -4.65E-08 | 4.95E-08 | 0.826250 |
| ADNC Score | A08.186.211.132.810.428.200.Cerebellum | CNS | 5.43E-08 | 4.66E-08 | 0.121513 |
| ADNC Score | A08.186.211.132.Brain.Stem | CNS | 3.46E-08 | 4.58E-08 | 0.225033 |
| ADNC Score | A08.186.211.464.405.Hippocampus | CNS | 2.37E-08 | 4.54E-08 | 0.300401 |
| ADNC Score | A08.186.211.464.710.225.Entorhinal.Cortex | CNS | -2.71E-08 | 4.45E-08 | 0.728430 |
| ADNC Score | A08.186.211.464.Limbic.System | CNS | 8.27E-09 | 4.74E-08 | 0.430777 |
| ADNC Score | A08.186.211.653.Mesencephalon | CNS | 2.58E-08 | 5.15E-08 | 0.308401 |
| ADNC Score | A08.186.211.730.317.357.352.435.Hypothalamo.Hypophyseal.System | CNS | -9.29E-08 | 6.25E-08 | 0.931184 |
| ADNC Score | A08.186.211.730.317.357.Hypothalamus | CNS | -7.22E-08 | 6.42E-08 | 0.869637 |

| | | | | | |
|---|---|---|---|---|---|
| ADNC Score | A08.186.211.730.317.Diencephalon | CNS | -6.23E-08 | 5.98E-08 | 0.851431 |
| ADNC Score | A08.186.211.730.885.287.249.487.Corpus.Striatum | CNS | 5.33E-08 | 5.03E-08 | 0.144916 |
| ADNC Score | A08.186.211.730.885.287.249.Basal.Ganglia | CNS | 8.00E-08 | 5.15E-08 | 0.060248 |
| ADNC Score | A08.186.211.730.885.287.500.270.Frontal.Lobe | CNS | -2.19E-08 | 4.63E-08 | 0.681823 |
| ADNC Score | A08.186.211.730.885.287.500.571.735.Visual.Cortex | CNS | -7.09E-08 | 4.48E-08 | 0.943342 |
| ADNC Score | A08.186.211.730.885.287.500.670.Parietal.Lobe | CNS | 8.29E-08 | 4.99E-08 | 0.048354 |
| ADNC Score | A08.186.211.730.885.287.500.Cerebral.Cortex | CNS | -4.62E-09 | 4.62E-08 | 0.539813 |
| ADNC Score | A08.186.211.865.428.Metencephalon | CNS | 5.50E-08 | 4.80E-08 | 0.125970 |
| ADNC Score | A08.186.211.Brain | CNS | 6.88E-09 | 4.64E-08 | 0.441080 |
| ADNC Score | A09.371.729.Retina | CNS | 1.91E-08 | 4.96E-08 | 0.350124 |
| ADNC Score | A11.872.653.Neural.Stem.Cells | CNS | 5.73E-08 | 6.51E-08 | 0.189367 |
| ADNC Score | Brain_Amygdala | CNS | -2.46E-08 | 4.02E-08 | 0.729586 |
| ADNC Score | Brain_Anterior_cingulate_cortex_(BA24) | CNS | -4.95E-08 | 4.00E-08 | 0.892303 |
| ADNC Score | Brain_Caudate_(basal_ganglia) | CNS | 3.54E-08 | 4.21E-08 | 0.200182 |
| ADNC Score | Brain_Cerebellar_Hemisphere | CNS | 5.55E-08 | 4.55E-08 | 0.111421 |
| ADNC Score | Brain_Cerebellum | CNS | -1.96E-08 | 4.79E-08 | 0.658477 |
| ADNC Score | Brain_Cortex | CNS | -5.08E-08 | 4.09E-08 | 0.892886 |
| ADNC Score | Brain_Frontal_Cortex_(BA9) | CNS | -3.77E-08 | 3.94E-08 | 0.830634 |
| ADNC Score | Brain_Hippocampus | CNS | -2.93E-08 | 4.37E-08 | 0.748656 |
| ADNC Score | Brain_Hypothalamus | CNS | -2.48E-08 | 4.30E-08 | 0.718351 |
| ADNC Score | Brain_Nucleus_accumbens_(basal_ganglia) | CNS | -4.30E-09 | 4.27E-08 | 0.540057 |
| ADNC Score | Brain_Putamen_(basal_ganglia) | CNS | 2.50E-08 | 4.25E-08 | 0.278007 |
| ADNC Score | Brain_Spinal_cord_(cervical_c-1) | CNS | 3.17E-08 | 4.58E-08 | 0.244315 |
| ADNC Score | Brain_Substantia_nigra | CNS | -2.96E-08 | 4.34E-08 | 0.751998 |
| ADNC Score | A03.556.124.369.Intestinal.Mucosa | Digestive | -2.74E-08 | 6.18E-08 | 0.671485 |
| ADNC Score | A03.556.124.526.767.Rectum | Digestive | 9.95E-08 | 6.79E-08 | 0.071414 |
| ADNC Score | A03.556.124.684.Intestine..Small | Digestive | 1.95E-09 | 6.09E-08 | 0.487199 |
| ADNC Score | A03.556.124.Intestines | Digestive | -5.38E-08 | 6.36E-08 | 0.801469 |
| ADNC Score | A03.556.249.124.Ileum | Digestive | -9.62E-09 | 6.62E-08 | 0.557798 |
| ADNC Score | A03.556.249.249.209.Cecum | Digestive | -5.44E-08 | 5.76E-08 | 0.827276 |
| ADNC Score | A03.556.249.249.356.668.Colon..Sigmoid | Digestive | -5.62E-08 | 6.60E-08 | 0.802904 |
| ADNC Score | A03.556.249.249.356.Colon | Digestive | -3.90E-08 | 6.52E-08 | 0.725087 |
| ADNC Score | A03.556.500.760.464.Parotid.Gland | Digestive | -5.31E-08 | 7.23E-08 | 0.768846 |
| ADNC Score | A03.556.500.760.Salivary.Glands | Digestive | -4.85E-08 | 6.96E-08 | 0.756805 |
| ADNC Score | A03.556.875.500.Esophagus | Digestive | -4.52E-08 | 6.05E-08 | 0.772625 |
| ADNC Score | A03.556.875.875.Stomach | Digestive | -8.37E-08 | 6.08E-08 | 0.915808 |
| ADNC Score | A03.556.875.Upper.Gastrointestinal.Tract | Digestive | -7.24E-08 | 5.70E-08 | 0.898050 |
| ADNC Score | A03.556.Gastrointestinal.Tract | Digestive | -1.08E-07 | 6.24E-08 | 0.958026 |
| ADNC Score | Colon_Sigmoid | Digestive | 1.76E-08 | 5.20E-08 | 0.367353 |
| ADNC Score | Colon_Transverse | Digestive | -9.84E-08 | 5.83E-08 | 0.954212 |
| ADNC Score | Esophagus_Gastroesophageal_Junction | Digestive | 8.50E-08 | 5.49E-08 | 0.060847 |
| ADNC Score | Esophagus_Mucosa | Digestive | -5.91E-08 | 6.04E-08 | 0.836205 |
| ADNC Score | Esophagus_Muscularis | Digestive | 9.24E-08 | 5.46E-08 | 0.045403 |
| ADNC Score | Small_Intestine_Terminal_Ileum | Digestive | -6.79E-08 | 5.80E-08 | 0.879307 |
| ADNC Score | Stomach | Digestive | -4.11E-08 | 5.54E-08 | 0.770663 |
| ADNC Score | A03.620.Liver | Liver | -1.36E-07 | 6.04E-08 | 0.988078 |

| | | | | | |
|---|---|---|---|---|---|
| ADNC Score | A11.436.348.Hepatocytes | Liver | 2.95E-08 | 6.17E-08 | 0.316434 |
| ADNC Score | Liver | Liver | -1.46E-07 | 5.45E-08 | 0.996262 |
| ADNC Score | A02.165.Cartilage | Musculoskeletal/Connective | 1.77E-08 | 5.33E-08 | 0.370142 |
| ADNC Score | A02.633.567.850.Quadriceps.Muscle | Musculoskeletal/Connective | 1.29E-07 | 6.51E-08 | 0.024067 |
| ADNC Score | A02.835.232.834.151.Cervical.Vertebrae | Musculoskeletal/Connective | 2.87E-08 | 6.49E-08 | 0.329316 |
| ADNC Score | A02.835.583.443.800.Synovial.Membrane | Musculoskeletal/Connective | 2.36E-08 | 5.08E-08 | 0.321365 |
| ADNC Score | A05.360.319.679.690.Myometrium | Musculoskeletal/Connective | 2.84E-08 | 5.31E-08 | 0.296405 |
| ADNC Score | A10.165.450.300.425.Keloid | Musculoskeletal/Connective | 7.09E-09 | 5.48E-08 | 0.448496 |
| ADNC Score | A10.165.450.300.Cicatrix | Musculoskeletal/Connective | 3.47E-08 | 5.73E-08 | 0.272013 |
| ADNC Score | A10.690.467.Muscle..Smooth | Musculoskeletal/Connective | 4.56E-08 | 6.08E-08 | 0.226790 |
| ADNC Score | A10.690.Muscles | Musculoskeletal/Connective | 1.47E-07 | 6.63E-08 | 0.013126 |
| ADNC Score | A11.329.171.Chondrocytes | Musculoskeletal/Connective | 3.00E-08 | 6.55E-08 | 0.323196 |
| ADNC Score | A11.329.228.Fibroblasts | Musculoskeletal/Connective | -4.09E-08 | 5.86E-08 | 0.757374 |
| ADNC Score | A11.329.629.Osteoblasts | Musculoskeletal/Connective | 6.57E-09 | 5.14E-08 | 0.449116 |
| ADNC Score | A11.329.830.Stromal.Cells | Musculoskeletal/Connective | -2.67E-08 | 6.13E-08 | 0.668498 |
| ADNC Score | A11.329.Connective.Tissue.Cells | Musculoskeletal/Connective | -2.43E-10 | 5.53E-08 | 0.501755 |
| ADNC Score | A11.620.520.Myocytes..Smooth.Muscle | Musculoskeletal/Connective | 5.28E-09 | 6.02E-08 | 0.465013 |
| ADNC Score | Cells_Transformed_fibroblasts | Musculoskeletal/Connective | -3.42E-08 | 5.00E-08 | 0.752615 |
| ADNC Score | Muscle_Skeletal | Musculoskeletal/Connective | 3.20E-08 | 5.74E-08 | 0.288251 |
| ADNC Score | A04.411.Lung | Other | 1.03E-08 | 5.63E-08 | 0.427717 |
| ADNC Score | A04.531.520.Nasal.Mucosa | Other | -2.30E-08 | 4.95E-08 | 0.679128 |
| ADNC Score | A05.360.319.114.373.Fallopian.Tubes | Other | -7.84E-09 | 5.79E-08 | 0.553849 |
| ADNC Score | A05.360.319.114.630.Ovary | Other | 1.61E-08 | 5.74E-08 | 0.389275 |
| ADNC Score | A05.360.319.679.256.Cervix.Uteri | Other | -5.16E-08 | 6.17E-08 | 0.798369 |
| ADNC Score | A05.360.319.679.490.Endometrium | Other | -5.55E-08 | 5.38E-08 | 0.849070 |
| ADNC Score | A05.360.319.679.Uterus | Other | -2.88E-08 | 5.44E-08 | 0.701612 |
| ADNC Score | A05.360.319.887.Vulva | Other | -1.23E-07 | 6.20E-08 | 0.976249 |
| ADNC Score | A05.360.319.Genitalia..Female | Other | -2.52E-08 | 6.00E-08 | 0.662837 |
| ADNC Score | A05.360.444.492.362.Foreskin | Other | 4.12E-08 | 5.60E-08 | 0.231004 |
| ADNC Score | A05.360.444.Genitalia..Male | Other | 3.05E-08 | 5.53E-08 | 0.290336 |
| ADNC Score | A05.360.490.Germ.Cells | Other | 3.70E-08 | 5.11E-08 | 0.234500 |
| ADNC Score | A05.360.Genitalia | Other | -2.59E-08 | 5.58E-08 | 0.678765 |
| ADNC Score | A05.810.453.324.Kidney.Cortex | Other | 3.86E-08 | 4.72E-08 | 0.207263 |
| ADNC Score | A05.810.453.Kidney | Other | 2.85E-08 | 5.02E-08 | 0.285183 |
| ADNC Score | A05.810.890.Urinary.Bladder | Other | -7.88E-08 | 7.14E-08 | 0.865054 |
| ADNC Score | A06.407.071.140.Adrenal.Cortex | Other | 2.40E-08 | 6.42E-08 | 0.354209 |
| ADNC Score | A06.407.071.Adrenal.Glands | Other | 3.24E-08 | 6.57E-08 | 0.310734 |
| ADNC Score | A06.407.312.782.Testis | Other | 3.44E-08 | 5.20E-08 | 0.254070 |
| ADNC Score | A06.407.312.Gonads | Other | 1.28E-08 | 5.93E-08 | 0.414426 |
| ADNC Score | A06.407.900.Thyroid.Gland | Other | 5.76E-08 | 5.71E-08 | 0.156363 |
| ADNC Score | A06.407.Endocrine.Glands | Other | 6.20E-08 | 6.16E-08 | 0.157368 |
| ADNC Score | A09.371.Eye | Other | -3.26E-08 | 6.25E-08 | 0.699233 |
| ADNC Score | A10.272.497.Epidermis | Other | -1.20E-07 | 6.76E-08 | 0.961685 |
| ADNC Score | A10.272.Epithelium | Other | -2.86E-08 | 5.03E-08 | 0.715669 |
| ADNC Score | A10.336.707.Prostate | Other | 6.58E-08 | 5.67E-08 | 0.123137 |
| ADNC Score | A10.615.284.473.Chorion | Other | 3.67E-08 | 5.84E-08 | 0.264833 |

| | | | | | |
|---|---|---|---|---|---|
| ADNC Score | A10.615.550.599.Mouth.Mucosa | Other | -8.34E-08 | 6.14E-08 | 0.912726 |
| ADNC Score | A10.615.550.Mucous.Membrane | Other | -1.46E-07 | 5.83E-08 | 0.993856 |
| ADNC Score | A10.615.789.Serous.Membrane | Other | 3.38E-08 | 5.93E-08 | 0.284550 |
| ADNC Score | A10.615.Membranes | Other | -1.02E-07 | 5.72E-08 | 0.963026 |
| ADNC Score | A11.382.Endocrine.Cells | Other | 5.58E-08 | 5.78E-08 | 0.167089 |
| ADNC Score | A11.436.275.Endothelial.Cells | Other | -5.57E-08 | 5.42E-08 | 0.847950 |
| ADNC Score | A11.436.294.064.Glucagon.Secreting.Cells | Other | 8.05E-08 | 5.80E-08 | 0.082664 |
| ADNC Score | A11.436.329.Granulosa.Cells | Other | 5.58E-08 | 5.18E-08 | 0.140801 |
| ADNC Score | A11.436.397.Keratinocytes | Other | -4.91E-08 | 6.38E-08 | 0.779574 |
| ADNC Score | A11.436.Epithelial.Cells | Other | -5.41E-08 | 5.50E-08 | 0.837285 |
| ADNC Score | A11.497.497.600.Oocytes | Other | -3.10E-08 | 5.07E-08 | 0.729663 |
| ADNC Score | A11.872.040.Adult.Stem.Cells | Other | 5.07E-08 | 6.24E-08 | 0.208131 |
| ADNC Score | A11.872.190.260.Embryoid.Bodies | Other | 7.90E-08 | 5.74E-08 | 0.084382 |
| ADNC Score | A11.872.190.Embryonic.Stem.Cells | Other | 5.69E-08 | 6.03E-08 | 0.172520 |
| ADNC Score | A11.872.580.Mesenchymal.Stem.Cells | Other | 2.83E-09 | 5.74E-08 | 0.480346 |
| ADNC Score | A11.872.700.500.Induced.Pluripotent.Stem.Cells | Other | 1.72E-08 | 6.29E-08 | 0.391983 |
| ADNC Score | A11.872.Stem.Cells | Other | 1.03E-07 | 6.36E-08 | 0.052186 |
| ADNC Score | A14.549.167.646.Periodontium | Other | -9.85E-08 | 6.37E-08 | 0.938880 |
| ADNC Score | A14.549.167.Dentition | Other | -1.10E-08 | 6.40E-08 | 0.568455 |
| ADNC Score | A14.549.885.Tongue | Other | -1.30E-07 | 5.87E-08 | 0.986657 |
| ADNC Score | A14.549.Mouth | Other | -6.61E-08 | 5.87E-08 | 0.870165 |
| ADNC Score | A14.724.557.Nasopharynx | Other | -1.30E-08 | 5.66E-08 | 0.591096 |
| ADNC Score | A14.724.Pharynx | Other | -5.32E-08 | 5.89E-08 | 0.816646 |
| ADNC Score | A17.815.Skin | Other | -6.16E-08 | 5.62E-08 | 0.863319 |
| ADNC Score | Adrenal_Gland | Other | 3.83E-08 | 5.75E-08 | 0.252824 |
| ADNC Score | Bladder | Other | 5.49E-08 | 5.12E-08 | 0.141378 |
| ADNC Score | Breast_Mammary_Tissue | Other | -1.02E-08 | 4.82E-08 | 0.584026 |
| ADNC Score | Cervix_Ectocervix | Other | -5.80E-08 | 4.98E-08 | 0.878300 |
| ADNC Score | Cervix_Endocervix | Other | 4.05E-08 | 4.99E-08 | 0.208498 |
| ADNC Score | Fallopian_Tube | Other | -1.15E-08 | 5.57E-08 | 0.581514 |
| ADNC Score | Kidney_Cortex | Other | 4.49E-08 | 5.22E-08 | 0.195046 |
| ADNC Score | Lung | Other | 1.71E-08 | 5.62E-08 | 0.380768 |
| ADNC Score | Minor_Salivary_Gland | Other | -9.88E-08 | 4.86E-08 | 0.978904 |
| ADNC Score | Nerve_Tibial | Other | -1.89E-08 | 4.80E-08 | 0.652807 |
| ADNC Score | Ovary | Other | 2.42E-08 | 4.67E-08 | 0.302631 |
| ADNC Score | Pituitary | Other | 1.80E-08 | 5.34E-08 | 0.367997 |
| ADNC Score | Prostate | Other | -1.12E-07 | 5.31E-08 | 0.982154 |
| ADNC Score | Skin_Not_Sun_Exposed_(Suprapubic) | Other | -6.51E-08 | 5.88E-08 | 0.865861 |
| ADNC Score | Skin_Sun_Exposed_(Lower_leg) | Other | -6.04E-08 | 5.97E-08 | 0.844141 |
| ADNC Score | Testis | Other | 5.25E-08 | 4.90E-08 | 0.141876 |
| ADNC Score | Thyroid | Other | -6.65E-08 | 5.17E-08 | 0.900999 |
| ADNC Score | Uterus | Other | -5.74E-08 | 4.51E-08 | 0.898123 |
| ADNC Score | Vagina | Other | -6.48E-08 | 4.99E-08 | 0.903084 |
| ADNC Score | A03.734.414.Islets.of.Langerhans | Pancreas | 9.87E-08 | 6.04E-08 | 0.051199 |
| ADNC Score | A03.734.Pancreas | Pancreas | -2.54E-08 | 5.43E-08 | 0.679776 |
| ADNC Score | Pancreas | Pancreas | -9.59E-08 | 5.48E-08 | 0.960104 |

| | | | | | |
|---|---|---|---|---|---|
| AP (any/none) | A10.165.114.830.500.750.Subcutaneous.Fat..Abdominal | Adipose | 1.03E-08 | 1.87E-08 | 0.290381 |
| AP (any/none) | A10.165.114.830.750.Subcutaneous.Fat | Adipose | 5.66E-09 | 1.83E-08 | 0.378881 |
| AP (any/none) | A11.329.114.Adipocytes | Adipose | 4.58E-09 | 1.83E-08 | 0.401268 |
| AP (any/none) | Adipose_Subcutaneous | Adipose | -4.31E-09 | 1.68E-08 | 0.601404 |
| AP (any/none) | Adipose_Visceral_(Omentum) | Adipose | 9.92E-10 | 1.87E-08 | 0.478835 |
| AP (any/none) | A02.835.583.443.800.800.Synovial.Fluid | Blood/Immune | 1.36E-08 | 2.50E-08 | 0.293728 |
| AP (any/none) | A10.549.400.Lymph.Nodes | Blood/Immune | -1.36E-08 | 2.20E-08 | 0.731389 |
| AP (any/none) | A10.549.Lymphoid.Tissue | Blood/Immune | -1.30E-08 | 2.15E-08 | 0.726912 |
| AP (any/none) | A11.118.637.555.567.562.440.Precursor.Cells..B.Lymphoid | Blood/Immune | 3.05E-09 | 2.16E-08 | 0.443805 |
| AP (any/none) | A11.118.637.555.567.562.B.Lymphocytes | Blood/Immune | -1.63E-08 | 2.16E-08 | 0.774270 |
| AP (any/none) | A11.118.637.555.567.569.200.700.T.Lymphocytes..Regulatory | Blood/Immune | 7.04E-09 | 2.23E-08 | 0.375942 |
| AP (any/none) | A11.118.637.555.567.569.T.Lymphocytes | Blood/Immune | -1.28E-08 | 2.31E-08 | 0.709584 |
| AP (any/none) | A11.118.637.Leukocytes | Blood/Immune | -3.63E-08 | 2.17E-08 | 0.952513 |
| AP (any/none) | A11.329.372.600.Macrophages..Alveolar | Blood/Immune | 4.93E-09 | 1.80E-08 | 0.392169 |
| AP (any/none) | A11.443.Erythroid.Cells | Blood/Immune | 2.83E-08 | 1.89E-08 | 0.067473 |
| AP (any/none) | A11.627.340.360.Granulocyte.Precursor.Cells | Blood/Immune | 4.39E-09 | 2.43E-08 | 0.428404 |
| AP (any/none) | A11.627.624.249.Monocyte.Macrophage.Precursor.Cells | Blood/Immune | 2.80E-08 | 2.34E-08 | 0.115587 |
| AP (any/none) | A11.627.635.Myeloid.Progenitor.Cells | Blood/Immune | 4.72E-08 | 2.73E-08 | 0.041941 |
| AP (any/none) | A11.872.378.590.635.Granulocyte.Macrophage.Progenitor.Cells | Blood/Immune | 2.40E-08 | 2.52E-08 | 0.170067 |
| AP (any/none) | A11.872.378.590.817.Megakaryocyte.Erythroid.Progenitor.Cells | Blood/Immune | 5.32E-08 | 2.32E-08 | 0.010814 |
| AP (any/none) | A11.872.378.Hematopoietic.Stem.Cells | Blood/Immune | 2.52E-08 | 2.70E-08 | 0.175175 |
| AP (any/none) | A15.145.229.188.Blood.Platelets | Blood/Immune | -1.27E-08 | 2.06E-08 | 0.731045 |
| AP (any/none) | A15.145.229.637.555.567.562.725.Plasma.Cells | Blood/Immune | -2.57E-08 | 2.10E-08 | 0.889723 |
| AP (any/none) | A15.145.229.637.555.567.569.200.CD4.Positive.T.Lymphocytes | Blood/Immune | -3.51E-09 | 2.41E-08 | 0.557872 |
| AP (any/none) | A15.145.229.637.555.Leukocytes..Mononuclear | Blood/Immune | -9.37E-09 | 2.40E-08 | 0.651691 |
| AP (any/none) | A15.145.229.Blood.Cells | Blood/Immune | -9.84E-09 | 2.43E-08 | 0.657263 |
| AP (any/none) | A15.145.300.Fetal.Blood | Blood/Immune | 1.47E-08 | 2.48E-08 | 0.277427 |
| AP (any/none) | A15.145.846.Serum | Blood/Immune | 1.19E-08 | 2.14E-08 | 0.289783 |
| AP (any/none) | A15.145.Blood | Blood/Immune | -1.02E-08 | 2.59E-08 | 0.653314 |
| AP (any/none) | A15.378.316.580.Monocytes | Blood/Immune | -1.01E-08 | 2.35E-08 | 0.666954 |
| AP (any/none) | A15.378.316.Bone.Marrow.Cells | Blood/Immune | 1.44E-08 | 2.26E-08 | 0.262467 |
| AP (any/none) | A15.382.490.315.583.Neutrophils | Blood/Immune | 5.83E-08 | 2.19E-08 | 0.003908 |
| AP (any/none) | A15.382.490.555.567.537.Killer.Cells..Natural | Blood/Immune | -1.33E-09 | 2.18E-08 | 0.524349 |
| AP (any/none) | A15.382.490.555.567.622.Lymphocytes..Null | Blood/Immune | 7.68E-09 | 2.18E-08 | 0.362062 |
| AP (any/none) | A15.382.490.555.567.Lymphocytes | Blood/Immune | 4.00E-09 | 2.33E-08 | 0.431830 |
| AP (any/none) | A15.382.520.604.700.Spleen | Blood/Immune | -4.11E-09 | 2.13E-08 | 0.576454 |
| AP (any/none) | A15.382.520.604.800.Palatine.Tonsil | Blood/Immune | 8.21E-10 | 2.17E-08 | 0.484896 |
| AP (any/none) | A15.382.680.Phagocytes | Blood/Immune | 3.41E-09 | 2.25E-08 | 0.439654 |
| AP (any/none) | A15.382.812.260.Dendritic.Cells | Blood/Immune | 1.49E-08 | 2.42E-08 | 0.268516 |
| AP (any/none) | A15.382.812.522.Macrophages | Blood/Immune | 2.09E-08 | 2.29E-08 | 0.179981 |
| AP (any/none) | A15.382.812.Mononuclear.Phagocyte.System | Blood/Immune | 6.57E-09 | 2.41E-08 | 0.392491 |
| AP (any/none) | A15.382.Immune.System | Blood/Immune | -5.32E-09 | 2.39E-08 | 0.588184 |
| AP (any/none) | Cells_EBV-transformed_lymphocytes | Blood/Immune | -7.99E-09 | 2.16E-08 | 0.644403 |
| AP (any/none) | Spleen | Blood/Immune | -8.36E-09 | 2.61E-08 | 0.625753 |
| AP (any/none) | Whole_Blood | Blood/Immune | 1.81E-08 | 2.02E-08 | 0.185243 |
| AP (any/none) | A07.231.114.Arteries | Cardiovascular | -2.45E-08 | 2.02E-08 | 0.887293 |

| | | | | | |
|---|---|---|---|---|---|
| AP (any/none) | A07.231.908.670.874.Umbilical.Veins | Cardiovascular | 1.27E-08 | 2.19E-08 | 0.281343 |
| AP (any/none) | A07.231.908.Veins | Cardiovascular | 2.23E-08 | 2.30E-08 | 0.165660 |
| AP (any/none) | A07.231.Blood.Vessels | Cardiovascular | 3.78E-08 | 2.42E-08 | 0.059022 |
| AP (any/none) | A07.541.358.100.Atrial.Appendage | Cardiovascular | -2.90E-10 | 1.85E-08 | 0.506249 |
| AP (any/none) | A07.541.358.Heart.Atria | Cardiovascular | 2.36E-09 | 1.85E-08 | 0.449172 |
| AP (any/none) | A07.541.510.110.Aortic.Valve | Cardiovascular | 2.46E-08 | 1.79E-08 | 0.084244 |
| AP (any/none) | A07.541.560.Heart.Ventricles | Cardiovascular | -1.83E-08 | 1.89E-08 | 0.834510 |
| AP (any/none) | A07.541.Heart | Cardiovascular | -6.13E-09 | 1.85E-08 | 0.629905 |
| AP (any/none) | Artery_Aorta | Cardiovascular | 5.19E-09 | 1.76E-08 | 0.384198 |
| AP (any/none) | Artery_Coronary | Cardiovascular | -1.54E-09 | 1.81E-08 | 0.533775 |
| AP (any/none) | Artery_Tibial | Cardiovascular | -2.90E-09 | 1.72E-08 | 0.567129 |
| AP (any/none) | Heart_Atrial_Appendage | Cardiovascular | 1.27E-08 | 1.83E-08 | 0.243707 |
| AP (any/none) | Heart_Left_Ventricle | Cardiovascular | 2.61E-09 | 2.04E-08 | 0.449162 |
| AP (any/none) | A08.186.211.132.810.428.200.Cerebellum | CNS | -1.47E-08 | 1.67E-08 | 0.810878 |
| AP (any/none) | A08.186.211.132.Brain.Stem | CNS | -1.31E-08 | 1.60E-08 | 0.794199 |
| AP (any/none) | A08.186.211.464.405.Hippocampus | CNS | 1.13E-09 | 1.57E-08 | 0.471366 |
| AP (any/none) | A08.186.211.464.710.225.Entorhinal.Cortex | CNS | 1.67E-08 | 1.70E-08 | 0.163873 |
| AP (any/none) | A08.186.211.464.Limbic.System | CNS | 6.81E-09 | 1.60E-08 | 0.334856 |
| AP (any/none) | A08.186.211.653.Mesencephalon | CNS | 3.94E-08 | 1.69E-08 | 0.010047 |
| AP (any/none) | A08.186.211.730.317.357.352.435.Hypothalamo.Hypophyseal.System | CNS | 4.60E-09 | 2.18E-08 | 0.416239 |
| AP (any/none) | A08.186.211.730.317.357.Hypothalamus | CNS | 2.65E-09 | 2.31E-08 | 0.454264 |
| AP (any/none) | A08.186.211.730.317.Diencephalon | CNS | 1.09E-08 | 2.00E-08 | 0.292418 |
| AP (any/none) | A08.186.211.730.885.287.249.487.Corpus.Striatum | CNS | 1.18E-08 | 1.79E-08 | 0.255285 |
| AP (any/none) | A08.186.211.730.885.287.249.Basal.Ganglia | CNS | 4.38E-08 | 1.86E-08 | 0.009413 |
| AP (any/none) | A08.186.211.730.885.287.500.270.Frontal.Lobe | CNS | -1.50E-08 | 1.65E-08 | 0.818736 |
| AP (any/none) | A08.186.211.730.885.287.500.571.735.Visual.Cortex | CNS | -3.25E-09 | 1.65E-08 | 0.577971 |
| AP (any/none) | A08.186.211.730.885.287.500.670.Parietal.Lobe | CNS | 1.46E-08 | 1.77E-08 | 0.204228 |
| AP (any/none) | A08.186.211.730.885.287.500.Cerebral.Cortex | CNS | -4.45E-10 | 1.52E-08 | 0.511635 |
| AP (any/none) | A08.186.211.865.428.Metencephalon | CNS | -1.64E-08 | 1.66E-08 | 0.839153 |
| AP (any/none) | A08.186.211.Brain | CNS | 5.59E-09 | 1.66E-08 | 0.368531 |
| AP (any/none) | A09.371.729.Retina | CNS | -2.21E-08 | 1.71E-08 | 0.902286 |
| AP (any/none) | A11.872.653.Neural.Stem.Cells | CNS | 3.55E-08 | 2.13E-08 | 0.047748 |
| AP (any/none) | Brain_Amygdala | CNS | 2.84E-08 | 1.73E-08 | 0.050343 |
| AP (any/none) | Brain_Anterior_cingulate_cortex_(BA24) | CNS | 3.59E-08 | 1.73E-08 | 0.019054 |
| AP (any/none) | Brain_Caudate_(basal_ganglia) | CNS | 3.52E-08 | 1.69E-08 | 0.018335 |
| AP (any/none) | Brain_Cerebellar_Hemisphere | CNS | 6.35E-09 | 1.69E-08 | 0.353801 |
| AP (any/none) | Brain_Cerebellum | CNS | 7.82E-09 | 1.70E-08 | 0.322653 |
| AP (any/none) | Brain_Cortex | CNS | 4.05E-08 | 1.78E-08 | 0.011656 |
| AP (any/none) | Brain_Frontal_Cortex_(BA9) | CNS | 4.19E-08 | 1.72E-08 | 0.007354 |
| AP (any/none) | Brain_Hippocampus | CNS | 1.65E-08 | 1.61E-08 | 0.153236 |
| AP (any/none) | Brain_Hypothalamus | CNS | 4.05E-08 | 1.72E-08 | 0.009322 |
| AP (any/none) | Brain_Nucleus_accumbens_(basal_ganglia) | CNS | 3.36E-08 | 1.73E-08 | 0.026030 |
| AP (any/none) | Brain_Putamen_(basal_ganglia) | CNS | 3.35E-08 | 1.67E-08 | 0.022447 |
| AP (any/none) | Brain_Spinal_cord_(cervical_c-1) | CNS | 1.63E-08 | 1.61E-08 | 0.154851 |
| AP (any/none) | Brain_Substantia_nigra | CNS | 3.26E-08 | 1.67E-08 | 0.025550 |
| AP (any/none) | A03.556.124.369.Intestinal.Mucosa | Digestive | 1.96E-09 | 2.39E-08 | 0.467365 |

| | | | | | |
|---|---|---|---|---|---|
| AP (any/none) | A03.556.124.526.767.Rectum | Digestive | 2.24E-08 | 2.37E-08 | 0.171437 |
| AP (any/none) | A03.556.124.684.Intestine..Small | Digestive | 9.45E-09 | 2.35E-08 | 0.343479 |
| AP (any/none) | A03.556.124.Intestines | Digestive | 3.05E-09 | 2.23E-08 | 0.445612 |
| AP (any/none) | A03.556.249.124.Ileum | Digestive | 7.80E-09 | 2.57E-08 | 0.380676 |
| AP (any/none) | A03.556.249.249.209.Cecum | Digestive | 2.15E-08 | 1.98E-08 | 0.138742 |
| AP (any/none) | A03.556.249.249.356.668.Colon..Sigmoid | Digestive | 5.66E-09 | 2.17E-08 | 0.397000 |
| AP (any/none) | A03.556.249.249.356.Colon | Digestive | 1.81E-09 | 2.30E-08 | 0.468641 |
| AP (any/none) | A03.556.500.760.464.Parotid.Gland | Digestive | -5.09E-09 | 2.53E-08 | 0.579627 |
| AP (any/none) | A03.556.500.760.Salivary.Glands | Digestive | -1.13E-08 | 2.42E-08 | 0.679663 |
| AP (any/none) | A03.556.875.500.Esophagus | Digestive | 2.37E-08 | 2.21E-08 | 0.141654 |
| AP (any/none) | A03.556.875.875.Stomach | Digestive | 4.59E-08 | 2.48E-08 | 0.032289 |
| AP (any/none) | A03.556.875.Upper.Gastrointestinal.Tract | Digestive | 3.67E-08 | 2.42E-08 | 0.064763 |
| AP (any/none) | A03.556.Gastrointestinal.Tract | Digestive | 1.57E-08 | 2.13E-08 | 0.230278 |
| AP (any/none) | Colon_Sigmoid | Digestive | -3.16E-08 | 1.84E-08 | 0.956718 |
| AP (any/none) | Colon_Transverse | Digestive | -3.12E-08 | 1.95E-08 | 0.945492 |
| AP (any/none) | Esophagus_Gastroesophageal_Junction | Digestive | -1.48E-08 | 1.95E-08 | 0.776578 |
| AP (any/none) | Esophagus_Mucosa | Digestive | -2.64E-08 | 2.13E-08 | 0.892976 |
| AP (any/none) | Esophagus_Muscularis | Digestive | -2.68E-09 | 1.85E-08 | 0.557717 |
| AP (any/none) | Small_Intestine_Terminal_Ileum | Digestive | -2.83E-08 | 2.04E-08 | 0.917235 |
| AP (any/none) | Stomach | Digestive | 3.23E-09 | 2.07E-08 | 0.437957 |
| AP (any/none) | A03.620.Liver | Liver | -4.73E-08 | 2.05E-08 | 0.989438 |
| AP (any/none) | A11.436.348.Hepatocytes | Liver | -1.26E-08 | 2.15E-08 | 0.721143 |
| AP (any/none) | Liver | Liver | -1.04E-08 | 2.05E-08 | 0.694454 |
| AP (any/none) | A02.165.Cartilage | Musculoskeletal/Connective | -1.34E-09 | 1.76E-08 | 0.530351 |
| AP (any/none) | A02.633.567.850.Quadriceps.Muscle | Musculoskeletal/Connective | 4.29E-09 | 2.22E-08 | 0.423280 |
| AP (any/none) | A02.835.232.834.151.Cervical.Vertebrae | Musculoskeletal/Connective | -3.01E-09 | 2.12E-08 | 0.556439 |
| AP (any/none) | A02.835.583.443.800.Synovial.Membrane | Musculoskeletal/Connective | 1.88E-08 | 1.89E-08 | 0.161053 |
| AP (any/none) | A05.360.319.679.690.Myometrium | Musculoskeletal/Connective | 3.00E-09 | 1.73E-08 | 0.431039 |
| AP (any/none) | A10.165.450.300.425.Keloid | Musculoskeletal/Connective | 3.01E-08 | 2.00E-08 | 0.065663 |
| AP (any/none) | A10.165.450.300.Cicatrix | Musculoskeletal/Connective | 2.09E-08 | 2.04E-08 | 0.152216 |
| AP (any/none) | A10.690.467.Muscle..Smooth | Musculoskeletal/Connective | 1.25E-08 | 2.04E-08 | 0.270548 |
| AP (any/none) | A10.690.Muscles | Musculoskeletal/Connective | -1.44E-08 | 2.07E-08 | 0.757160 |
| AP (any/none) | A11.329.171.Chondrocytes | Musculoskeletal/Connective | 3.36E-08 | 1.92E-08 | 0.040353 |
| AP (any/none) | A11.329.228.Fibroblasts | Musculoskeletal/Connective | 2.31E-08 | 1.59E-08 | 0.073354 |
| AP (any/none) | A11.329.629.Osteoblasts | Musculoskeletal/Connective | 1.92E-08 | 1.88E-08 | 0.153170 |
| AP (any/none) | A11.329.830.Stromal.Cells | Musculoskeletal/Connective | 1.14E-08 | 1.80E-08 | 0.263760 |
| AP (any/none) | A11.329.Connective.Tissue.Cells | Musculoskeletal/Connective | 6.85E-09 | 2.07E-08 | 0.370140 |
| AP (any/none) | A11.620.520.Myocytes..Smooth.Muscle | Musculoskeletal/Connective | 6.54E-09 | 1.95E-08 | 0.368531 |
| AP (any/none) | Cells_Transformed_fibroblasts | Musculoskeletal/Connective | 7.17E-09 | 1.77E-08 | 0.342409 |
| AP (any/none) | Muscle_Skeletal | Musculoskeletal/Connective | -3.67E-08 | 2.04E-08 | 0.963929 |
| AP (any/none) | A04.411.Lung | Other | -2.45E-08 | 2.05E-08 | 0.883387 |
| AP (any/none) | A04.531.520.Nasal.Mucosa | Other | -2.83E-08 | 2.00E-08 | 0.922077 |
| AP (any/none) | A05.360.319.114.373.Fallopian.Tubes | Other | 1.68E-08 | 2.01E-08 | 0.202115 |
| AP (any/none) | A05.360.319.114.630.Ovary | Other | 9.70E-09 | 1.93E-08 | 0.307483 |
| AP (any/none) | A05.360.319.679.256.Cervix.Uteri | Other | 1.24E-08 | 2.22E-08 | 0.289196 |
| AP (any/none) | A05.360.319.679.490.Endometrium | Other | 4.93E-09 | 1.81E-08 | 0.392590 |

| | | | | | |
|---|---|---|---|---|---|
| AP (any/none) | A05.360.319.679.Uterus | Other | 3.46E-09 | 1.92E-08 | 0.428230 |
| AP (any/none) | A05.360.319.887.Vulva | Other | 1.04E-08 | 2.08E-08 | 0.308155 |
| AP (any/none) | A05.360.319.Genitalia..Female | Other | 2.22E-08 | 1.81E-08 | 0.109829 |
| AP (any/none) | A05.360.444.492.362.Foreskin | Other | 1.57E-08 | 1.73E-08 | 0.182187 |
| AP (any/none) | A05.360.444.Genitalia..Male | Other | -1.14E-08 | 1.78E-08 | 0.739523 |
| AP (any/none) | A05.360.490.Germ.Cells | Other | -2.36E-08 | 1.74E-08 | 0.912384 |
| AP (any/none) | A05.360.Genitalia | Other | -8.73E-09 | 1.83E-08 | 0.683025 |
| AP (any/none) | A05.810.453.324.Kidney.Cortex | Other | 3.08E-08 | 1.95E-08 | 0.057770 |
| AP (any/none) | A05.810.453.Kidney | Other | -1.89E-08 | 1.92E-08 | 0.837692 |
| AP (any/none) | A05.810.890.Urinary.Bladder | Other | 1.56E-08 | 2.32E-08 | 0.251090 |
| AP (any/none) | A06.407.071.140.Adrenal.Cortex | Other | -5.10E-10 | 2.02E-08 | 0.510083 |
| AP (any/none) | A06.407.071.Adrenal.Glands | Other | 1.20E-08 | 1.98E-08 | 0.271966 |
| AP (any/none) | A06.407.312.782.Testis | Other | 8.63E-10 | 1.69E-08 | 0.479634 |
| AP (any/none) | A06.407.312.Gonads | Other | -4.15E-09 | 1.87E-08 | 0.587922 |
| AP (any/none) | A06.407.900.Thyroid.Gland | Other | -1.86E-09 | 1.98E-08 | 0.537486 |
| AP (any/none) | A06.407.Endocrine.Glands | Other | 5.62E-09 | 1.87E-08 | 0.382047 |
| AP (any/none) | A09.371.Eye | Other | -5.63E-10 | 2.14E-08 | 0.510471 |
| AP (any/none) | A10.272.497.Epidermis | Other | 6.24E-09 | 2.38E-08 | 0.396652 |
| AP (any/none) | A10.272.Epithelium | Other | -4.00E-08 | 2.01E-08 | 0.976659 |
| AP (any/none) | A10.336.707.Prostate | Other | -7.66E-09 | 1.83E-08 | 0.662718 |
| AP (any/none) | A10.615.284.473.Chorion | Other | -2.31E-08 | 1.82E-08 | 0.897536 |
| AP (any/none) | A10.615.550.599.Mouth.Mucosa | Other | -3.17E-08 | 2.13E-08 | 0.930963 |
| AP (any/none) | A10.615.550.Mucous.Membrane | Other | -1.09E-08 | 2.18E-08 | 0.691967 |
| AP (any/none) | A10.615.789.Serous.Membrane | Other | 1.76E-08 | 2.32E-08 | 0.225080 |
| AP (any/none) | A10.615.Membranes | Other | -1.40E-08 | 2.00E-08 | 0.758290 |
| AP (any/none) | A11.382.Endocrine.Cells | Other | -3.83E-09 | 1.87E-08 | 0.581015 |
| AP (any/none) | A11.436.275.Endothelial.Cells | Other | 4.07E-08 | 2.39E-08 | 0.044429 |
| AP (any/none) | A11.436.294.064.Glucagon.Secreting.Cells | Other | 1.25E-08 | 2.10E-08 | 0.275125 |
| AP (any/none) | A11.436.329.Granulosa.Cells | Other | -2.98E-08 | 1.91E-08 | 0.939975 |
| AP (any/none) | A11.436.397.Keratinocytes | Other | -4.85E-09 | 2.17E-08 | 0.588429 |
| AP (any/none) | A11.436.Epithelial.Cells | Other | -3.17E-08 | 1.94E-08 | 0.948721 |
| AP (any/none) | A11.497.497.600.Oocytes | Other | -1.73E-08 | 1.71E-08 | 0.843581 |
| AP (any/none) | A11.872.040.Adult.Stem.Cells | Other | 1.21E-08 | 2.25E-08 | 0.295847 |
| AP (any/none) | A11.872.190.260.Embryoid.Bodies | Other | 7.83E-09 | 2.15E-08 | 0.357806 |
| AP (any/none) | A11.872.190.Embryonic.Stem.Cells | Other | 2.74E-08 | 2.09E-08 | 0.095070 |
| AP (any/none) | A11.872.580.Mesenchymal.Stem.Cells | Other | -1.59E-08 | 1.87E-08 | 0.802293 |
| AP (any/none) | A11.872.700.500.Induced.Pluripotent.Stem.Cells | Other | 1.29E-08 | 2.19E-08 | 0.277305 |
| AP (any/none) | A11.872.Stem.Cells | Other | 2.55E-08 | 2.25E-08 | 0.128957 |
| AP (any/none) | A14.549.167.646.Periodontium | Other | -8.76E-09 | 2.28E-08 | 0.649629 |
| AP (any/none) | A14.549.167.Dentition | Other | 8.12E-10 | 2.20E-08 | 0.485283 |
| AP (any/none) | A14.549.885.Tongue | Other | 1.90E-08 | 2.26E-08 | 0.200468 |
| AP (any/none) | A14.549.Mouth | Other | -2.57E-08 | 2.11E-08 | 0.888839 |
| AP (any/none) | A14.724.557.Nasopharynx | Other | -3.51E-09 | 2.08E-08 | 0.567118 |
| AP (any/none) | A14.724.Pharynx | Other | -1.01E-08 | 2.10E-08 | 0.685150 |
| AP (any/none) | A17.815.Skin | Other | 1.47E-08 | 2.04E-08 | 0.235667 |
| AP (any/none) | Adrenal_Gland | Other | -1.39E-08 | 2.01E-08 | 0.755334 |

| | | | | | |
|---|---|---|---|---|---|
| AP (any/none) | Bladder | Other | -6.25E-09 | 1.73E-08 | 0.640947 |
| AP (any/none) | Breast_Mammary_Tissue | Other | -9.76E-09 | 1.67E-08 | 0.720897 |
| AP (any/none) | Cervix_Ectocervix | Other | -5.57E-09 | 1.80E-08 | 0.621792 |
| AP (any/none) | Cervix_Endocervix | Other | -1.24E-08 | 1.83E-08 | 0.751028 |
| AP (any/none) | Fallopian_Tube | Other | -1.62E-08 | 1.92E-08 | 0.799848 |
| AP (any/none) | Kidney_Cortex | Other | 1.32E-09 | 1.88E-08 | 0.471984 |
| AP (any/none) | Lung | Other | -2.26E-08 | 1.88E-08 | 0.885335 |
| AP (any/none) | Minor_Salivary_Gland | Other | -5.37E-09 | 1.81E-08 | 0.616887 |
| AP (any/none) | Nerve_Tibial | Other | -1.10E-08 | 1.50E-08 | 0.768549 |
| AP (any/none) | Ovary | Other | -9.34E-09 | 1.76E-08 | 0.702149 |
| AP (any/none) | Pituitary | Other | 1.45E-09 | 1.75E-08 | 0.466989 |
| AP (any/none) | Prostate | Other | -1.18E-08 | 1.85E-08 | 0.737476 |
| AP (any/none) | Skin_Not_Sun_Exposed_(Suprapubic) | Other | 3.23E-09 | 2.15E-08 | 0.440219 |
| AP (any/none) | Skin_Sun_Exposed_(Lower_leg) | Other | -7.38E-09 | 1.96E-08 | 0.646813 |
| AP (any/none) | Testis | Other | 2.53E-08 | 2.12E-08 | 0.116609 |
| AP (any/none) | Thyroid | Other | -2.01E-08 | 1.69E-08 | 0.882314 |
| AP (any/none) | Uterus | Other | 3.24E-09 | 1.86E-08 | 0.430972 |
| AP (any/none) | Vagina | Other | 7.15E-10 | 1.81E-08 | 0.484260 |
| AP (any/none) | A03.734.414.Islets.of.Langerhans | Pancreas | 1.07E-08 | 2.09E-08 | 0.304254 |
| AP (any/none) | A03.734.Pancreas | Pancreas | -5.14E-08 | 1.88E-08 | 0.996852 |
| AP (any/none) | Pancreas | Pancreas | -3.24E-08 | 2.01E-08 | 0.946322 |
| Arteriolosclerosis | A10.165.114.830.500.750.Subcutaneous.Fat..Abdominal | Adipose | 4.53E-08 | 2.54E-08 | 0.037356 |
| Arteriolosclerosis | A10.165.114.830.750.Subcutaneous.Fat | Adipose | 4.42E-08 | 2.44E-08 | 0.034895 |
| Arteriolosclerosis | A11.329.114.Adipocytes | Adipose | 4.39E-08 | 2.52E-08 | 0.040893 |
| Arteriolosclerosis | Adipose_Subcutaneous | Adipose | 4.75E-08 | 2.29E-08 | 0.019033 |
| Arteriolosclerosis | Adipose_Visceral_(Omentum) | Adipose | 2.25E-08 | 2.22E-08 | 0.154799 |
| Arteriolosclerosis | A02.835.583.443.800.800.Synovial.Fluid | Blood/Immune | -1.96E-08 | 2.84E-08 | 0.754946 |
| Arteriolosclerosis | A10.549.400.Lymph.Nodes | Blood/Immune | -5.71E-09 | 2.42E-08 | 0.593195 |
| Arteriolosclerosis | A10.549.Lymphoid.Tissue | Blood/Immune | 4.18E-09 | 2.47E-08 | 0.432826 |
| Arteriolosclerosis | A11.118.637.555.567.562.440.Precursor.Cells..B.Lymphoid | Blood/Immune | -2.59E-09 | 2.90E-08 | 0.535506 |
| Arteriolosclerosis | A11.118.637.555.567.562.B.Lymphocytes | Blood/Immune | -5.93E-10 | 2.85E-08 | 0.508300 |
| Arteriolosclerosis | A11.118.637.555.567.569.200.700.T.Lymphocytes..Regulatory | Blood/Immune | 3.28E-08 | 2.68E-08 | 0.110651 |
| Arteriolosclerosis | A11.118.637.555.567.569.T.Lymphocytes | Blood/Immune | 5.11E-10 | 2.88E-08 | 0.492913 |
| Arteriolosclerosis | A11.118.637.Leukocytes | Blood/Immune | 4.77E-09 | 2.83E-08 | 0.433041 |
| Arteriolosclerosis | A11.329.372.600.Macrophages..Alveolar | Blood/Immune | 2.94E-08 | 2.47E-08 | 0.117763 |
| Arteriolosclerosis | A11.443.Erythroid.Cells | Blood/Immune | -1.93E-09 | 2.63E-08 | 0.529299 |
| Arteriolosclerosis | A11.627.340.360.Granulocyte.Precursor.Cells | Blood/Immune | -6.08E-08 | 2.84E-08 | 0.983711 |
| Arteriolosclerosis | A11.627.624.249.Monocyte.Macrophage.Precursor.Cells | Blood/Immune | 2.37E-09 | 2.79E-08 | 0.466151 |
| Arteriolosclerosis | A11.627.635.Myeloid.Progenitor.Cells | Blood/Immune | -1.19E-09 | 2.68E-08 | 0.517656 |
| Arteriolosclerosis | A11.872.378.590.635.Granulocyte.Macrophage.Progenitor.Cells | Blood/Immune | -1.25E-08 | 2.92E-08 | 0.665566 |
| Arteriolosclerosis | A11.872.378.590.817.Megakaryocyte.Erythroid.Progenitor.Cells | Blood/Immune | -1.53E-08 | 2.65E-08 | 0.717681 |
| Arteriolosclerosis | A11.872.378.Hematopoietic.Stem.Cells | Blood/Immune | -2.27E-08 | 2.70E-08 | 0.799183 |
| Arteriolosclerosis | A15.145.229.188.Blood.Platelets | Blood/Immune | -1.15E-08 | 2.69E-08 | 0.665196 |
| Arteriolosclerosis | A15.145.229.637.555.567.562.725.Plasma.Cells | Blood/Immune | 3.47E-08 | 3.27E-08 | 0.144676 |
| Arteriolosclerosis | A15.145.229.637.555.567.569.200.CD4.Positive.T.Lymphocytes | Blood/Immune | 2.14E-08 | 2.79E-08 | 0.221733 |
| Arteriolosclerosis | A15.145.229.637.555.Leukocytes..Mononuclear | Blood/Immune | -3.77E-09 | 2.77E-08 | 0.554209 |

| | | | | | |
|---|---|---|---|---|---|
| Arteriolosclerosis | A15.145.229.Blood.Cells | Blood/Immune | -2.66E-08 | 2.97E-08 | 0.815217 |
| Arteriolosclerosis | A15.145.300.Fetal.Blood | Blood/Immune | -1.70E-08 | 3.17E-08 | 0.703997 |
| Arteriolosclerosis | A15.145.846.Serum | Blood/Immune | 1.37E-08 | 2.75E-08 | 0.308594 |
| Arteriolosclerosis | A15.145.Blood | Blood/Immune | -1.31E-08 | 3.12E-08 | 0.663421 |
| Arteriolosclerosis | A15.378.316.580.Monocytes | Blood/Immune | -9.12E-09 | 2.55E-08 | 0.639716 |
| Arteriolosclerosis | A15.378.316.Bone.Marrow.Cells | Blood/Immune | -2.65E-08 | 3.02E-08 | 0.809910 |
| Arteriolosclerosis | A15.382.490.315.583.Neutrophils | Blood/Immune | -1.61E-08 | 2.93E-08 | 0.708906 |
| Arteriolosclerosis | A15.382.490.555.567.537.Killer.Cells..Natural | Blood/Immune | -7.79E-09 | 2.83E-08 | 0.608368 |
| Arteriolosclerosis | A15.382.490.555.567.622.Lymphocytes..Null | Blood/Immune | 4.84E-08 | 3.14E-08 | 0.061772 |
| Arteriolosclerosis | A15.382.490.555.567.Lymphocytes | Blood/Immune | 7.71E-09 | 3.04E-08 | 0.399971 |
| Arteriolosclerosis | A15.382.520.604.700.Spleen | Blood/Immune | -3.60E-09 | 2.63E-08 | 0.554388 |
| Arteriolosclerosis | A15.382.520.604.800.Palatine.Tonsil | Blood/Immune | -1.12E-10 | 2.48E-08 | 0.501807 |
| Arteriolosclerosis | A15.382.680.Phagocytes | Blood/Immune | -1.84E-08 | 2.86E-08 | 0.740367 |
| Arteriolosclerosis | A15.382.812.260.Dendritic.Cells | Blood/Immune | -1.62E-08 | 2.49E-08 | 0.742358 |
| Arteriolosclerosis | A15.382.812.522.Macrophages | Blood/Immune | -8.86E-09 | 2.41E-08 | 0.643424 |
| Arteriolosclerosis | A15.382.812.Mononuclear.Phagocyte.System | Blood/Immune | 2.64E-09 | 2.38E-08 | 0.455764 |
| Arteriolosclerosis | A15.382.Immune.System | Blood/Immune | -4.69E-08 | 2.83E-08 | 0.951182 |
| Arteriolosclerosis | Cells_EBV-transformed_lymphocytes | Blood/Immune | 2.77E-11 | 2.48E-08 | 0.499555 |
| Arteriolosclerosis | Spleen | Blood/Immune | -3.02E-08 | 2.68E-08 | 0.870574 |
| Arteriolosclerosis | Whole_Blood | Blood/Immune | 7.79E-09 | 2.75E-08 | 0.388343 |
| Arteriolosclerosis | A07.231.114.Arteries | Cardiovascular | 4.54E-08 | 2.61E-08 | 0.041317 |
| Arteriolosclerosis | A07.231.908.670.874.Umbilical.Veins | Cardiovascular | 1.23E-08 | 2.75E-08 | 0.326717 |
| Arteriolosclerosis | A07.231.908.Veins | Cardiovascular | 1.28E-09 | 2.84E-08 | 0.482029 |
| Arteriolosclerosis | A07.231.Blood.Vessels | Cardiovascular | 3.98E-08 | 2.93E-08 | 0.086868 |
| Arteriolosclerosis | A07.541.358.100.Atrial.Appendage | Cardiovascular | 1.76E-08 | 2.62E-08 | 0.251204 |
| Arteriolosclerosis | A07.541.358.Heart.Atria | Cardiovascular | 1.07E-08 | 2.57E-08 | 0.337882 |
| Arteriolosclerosis | A07.541.510.110.Aortic.Valve | Cardiovascular | 1.90E-08 | 2.57E-08 | 0.229895 |
| Arteriolosclerosis | A07.541.560.Heart.Ventricles | Cardiovascular | -1.32E-08 | 2.55E-08 | 0.697075 |
| Arteriolosclerosis | A07.541.Heart | Cardiovascular | -5.56E-09 | 2.58E-08 | 0.585334 |
| Arteriolosclerosis | Artery_Aorta | Cardiovascular | -2.96E-09 | 2.19E-08 | 0.553855 |
| Arteriolosclerosis | Artery_Coronary | Cardiovascular | -3.12E-09 | 2.31E-08 | 0.553899 |
| Arteriolosclerosis | Artery_Tibial | Cardiovascular | -1.04E-08 | 1.96E-08 | 0.702091 |
| Arteriolosclerosis | Heart_Atrial_Appendage | Cardiovascular | -1.86E-08 | 2.37E-08 | 0.783682 |
| Arteriolosclerosis | Heart_Left_Ventricle | Cardiovascular | -5.16E-08 | 2.48E-08 | 0.981478 |
| Arteriolosclerosis | A08.186.211.132.810.428.200.Cerebellum | CNS | 1.60E-08 | 2.29E-08 | 0.242995 |
| Arteriolosclerosis | A08.186.211.132.Brain.Stem | CNS | -7.14E-09 | 2.26E-08 | 0.623876 |
| Arteriolosclerosis | A08.186.211.464.405.Hippocampus | CNS | -2.19E-08 | 2.20E-08 | 0.839950 |
| Arteriolosclerosis | A08.186.211.464.710.225.Entorhinal.Cortex | CNS | -5.81E-09 | 2.10E-08 | 0.609073 |
| Arteriolosclerosis | A08.186.211.464.Limbic.System | CNS | -2.04E-08 | 2.11E-08 | 0.833355 |
| Arteriolosclerosis | A08.186.211.653.Mesencephalon | CNS | -5.86E-08 | 2.48E-08 | 0.990978 |
| Arteriolosclerosis | A08.186.211.730.317.357.352.435.Hypothalamo.Hypophyseal.System | CNS | -3.36E-08 | 2.57E-08 | 0.904681 |
| Arteriolosclerosis | A08.186.211.730.317.357.Hypothalamus | CNS | -3.88E-08 | 2.78E-08 | 0.918751 |
| Arteriolosclerosis | A08.186.211.730.317.Diencephalon | CNS | -1.87E-08 | 2.64E-08 | 0.760381 |
| Arteriolosclerosis | A08.186.211.730.885.287.249.487.Corpus.Striatum | CNS | 3.99E-09 | 2.29E-08 | 0.430790 |
| Arteriolosclerosis | A08.186.211.730.885.287.249.Basal.Ganglia | CNS | -1.58E-10 | 2.39E-08 | 0.502641 |
| Arteriolosclerosis | A08.186.211.730.885.287.500.270.Frontal.Lobe | CNS | -2.59E-08 | 2.41E-08 | 0.858444 |

| | | | | | |
|---|---|---|---|---|---|
| Arteriolosclerosis | A08.186.211.730.885.287.500.571.735.Visual.Cortex | CNS | -6.80E-09 | 2.16E-08 | 0.623351 |
| Arteriolosclerosis | A08.186.211.730.885.287.500.670.Parietal.Lobe | CNS | -9.55E-09 | 2.32E-08 | 0.659691 |
| Arteriolosclerosis | A08.186.211.730.885.287.500.Cerebral.Cortex | CNS | -2.37E-08 | 2.17E-08 | 0.862626 |
| Arteriolosclerosis | A08.186.211.865.428.Metencephalon | CNS | 8.24E-09 | 2.27E-08 | 0.358441 |
| Arteriolosclerosis | A08.186.211.Brain | CNS | -2.68E-08 | 2.14E-08 | 0.894422 |
| Arteriolosclerosis | A09.371.729.Retina | CNS | -4.64E-08 | 2.01E-08 | 0.989563 |
| Arteriolosclerosis | A11.872.653.Neural.Stem.Cells | CNS | -3.37E-08 | 2.54E-08 | 0.907971 |
| Arteriolosclerosis | Brain_Amygdala | CNS | -4.18E-08 | 2.09E-08 | 0.977368 |
| Arteriolosclerosis | Brain_Anterior_cingulate_cortex_(BA24) | CNS | -2.84E-08 | 2.03E-08 | 0.919546 |
| Arteriolosclerosis | Brain_Caudate_(basal_ganglia) | CNS | -2.07E-08 | 2.13E-08 | 0.833789 |
| Arteriolosclerosis | Brain_Cerebellar_Hemisphere | CNS | -3.36E-08 | 2.16E-08 | 0.940327 |
| Arteriolosclerosis | Brain_Cerebellum | CNS | -2.52E-08 | 2.18E-08 | 0.876446 |
| Arteriolosclerosis | Brain_Cortex | CNS | -3.67E-08 | 2.06E-08 | 0.962211 |
| Arteriolosclerosis | Brain_Frontal_Cortex_(BA9) | CNS | -3.13E-08 | 2.04E-08 | 0.937582 |
| Arteriolosclerosis | Brain_Hippocampus | CNS | -4.08E-08 | 2.12E-08 | 0.972735 |
| Arteriolosclerosis | Brain_Hypothalamus | CNS | -3.25E-08 | 2.18E-08 | 0.931934 |
| Arteriolosclerosis | Brain_Nucleus_accumbens_(basal_ganglia) | CNS | -3.38E-08 | 2.14E-08 | 0.942807 |
| Arteriolosclerosis | Brain_Putamen_(basal_ganglia) | CNS | -2.91E-08 | 2.23E-08 | 0.903511 |
| Arteriolosclerosis | Brain_Spinal_cord_(cervical_c-1) | CNS | -1.87E-08 | 2.16E-08 | 0.806344 |
| Arteriolosclerosis | Brain_Substantia_nigra | CNS | -3.40E-08 | 2.22E-08 | 0.936675 |
| Arteriolosclerosis | A03.556.124.369.Intestinal.Mucosa | Digestive | 1.14E-08 | 2.74E-08 | 0.338798 |
| Arteriolosclerosis | A03.556.124.526.767.Rectum | Digestive | 3.74E-09 | 2.72E-08 | 0.445339 |
| Arteriolosclerosis | A03.556.124.684.Intestine..Small | Digestive | -2.68E-09 | 2.99E-08 | 0.535693 |
| Arteriolosclerosis | A03.556.124.Intestines | Digestive | 2.50E-08 | 3.19E-08 | 0.217040 |
| Arteriolosclerosis | A03.556.249.124.Ileum | Digestive | 1.01E-08 | 2.89E-08 | 0.363637 |
| Arteriolosclerosis | A03.556.249.249.209.Cecum | Digestive | -1.45E-08 | 2.77E-08 | 0.700275 |
| Arteriolosclerosis | A03.556.249.249.356.668.Colon..Sigmoid | Digestive | 2.07E-08 | 3.05E-08 | 0.248259 |
| Arteriolosclerosis | A03.556.249.249.356.Colon | Digestive | 1.88E-08 | 3.04E-08 | 0.267875 |
| Arteriolosclerosis | A03.556.500.760.464.Parotid.Gland | Digestive | -2.46E-08 | 2.68E-08 | 0.820759 |
| Arteriolosclerosis | A03.556.500.760.Salivary.Glands | Digestive | -4.43E-08 | 2.69E-08 | 0.950121 |
| Arteriolosclerosis | A03.556.875.500.Esophagus | Digestive | -2.44E-08 | 2.70E-08 | 0.817147 |
| Arteriolosclerosis | A03.556.875.875.Stomach | Digestive | -4.09E-08 | 2.76E-08 | 0.930844 |
| Arteriolosclerosis | A03.556.875.Upper.Gastrointestinal.Tract | Digestive | -1.45E-08 | 2.78E-08 | 0.698378 |
| Arteriolosclerosis | A03.556.Gastrointestinal.Tract | Digestive | 2.89E-08 | 3.09E-08 | 0.174985 |
| Arteriolosclerosis | Colon_Sigmoid | Digestive | -3.78E-11 | 2.29E-08 | 0.500659 |
| Arteriolosclerosis | Colon_Transverse | Digestive | -9.55E-09 | 2.61E-08 | 0.642842 |
| Arteriolosclerosis | Esophagus_Gastroesophageal_Junction | Digestive | -2.91E-08 | 2.06E-08 | 0.921651 |
| Arteriolosclerosis | Esophagus_Mucosa | Digestive | 2.00E-08 | 2.62E-08 | 0.222784 |
| Arteriolosclerosis | Esophagus_Muscularis | Digestive | -3.01E-08 | 2.22E-08 | 0.912561 |
| Arteriolosclerosis | Small_Intestine_Terminal_Ileum | Digestive | -1.94E-08 | 2.64E-08 | 0.768904 |
| Arteriolosclerosis | Stomach | Digestive | -5.46E-08 | 2.50E-08 | 0.985462 |
| Arteriolosclerosis | A03.620.Liver | Liver | 1.37E-08 | 2.79E-08 | 0.311208 |
| Arteriolosclerosis | A11.436.348.Hepatocytes | Liver | -1.32E-08 | 3.08E-08 | 0.666198 |
| Arteriolosclerosis | Liver | Liver | -3.57E-08 | 2.41E-08 | 0.931033 |
| Arteriolosclerosis | A02.165.Cartilage | Musculoskeletal/Connective | 1.12E-09 | 2.40E-08 | 0.481363 |
| Arteriolosclerosis | A02.633.567.850.Quadriceps.Muscle | Musculoskeletal/Connective | -6.82E-08 | 2.55E-08 | 0.996244 |

| | | | | | |
|---|---|---|---|---|---|
| Arteriolosclerosis | A02.835.232.834.151.Cervical.Vertebrae | Musculoskeletal/Connective | 1.02E-08 | 2.56E-08 | 0.346023 |
| Arteriolosclerosis | A02.835.583.443.800.Synovial.Membrane | Musculoskeletal/Connective | 2.39E-08 | 2.31E-08 | 0.150243 |
| Arteriolosclerosis | A05.360.319.679.690.Myometrium | Musculoskeletal/Connective | 1.40E-08 | 2.41E-08 | 0.280088 |
| Arteriolosclerosis | A10.165.450.300.425.Keloid | Musculoskeletal/Connective | 1.83E-08 | 2.38E-08 | 0.220066 |
| Arteriolosclerosis | A10.165.450.300.Cicatrix | Musculoskeletal/Connective | -2.79E-09 | 2.51E-08 | 0.544237 |
| Arteriolosclerosis | A10.690.467.Muscle..Smooth | Musculoskeletal/Connective | 3.94E-08 | 3.03E-08 | 0.096568 |
| Arteriolosclerosis | A10.690.Muscles | Musculoskeletal/Connective | -3.99E-08 | 2.68E-08 | 0.932019 |
| Arteriolosclerosis | A11.329.171.Chondrocytes | Musculoskeletal/Connective | 3.38E-08 | 2.60E-08 | 0.097036 |
| Arteriolosclerosis | A11.329.228.Fibroblasts | Musculoskeletal/Connective | -9.84E-09 | 2.52E-08 | 0.651991 |
| Arteriolosclerosis | A11.329.629.Osteoblasts | Musculoskeletal/Connective | 2.48E-08 | 2.34E-08 | 0.144791 |
| Arteriolosclerosis | A11.329.830.Stromal.Cells | Musculoskeletal/Connective | 1.81E-08 | 2.66E-08 | 0.248809 |
| Arteriolosclerosis | A11.329.Connective.Tissue.Cells | Musculoskeletal/Connective | 3.94E-08 | 2.46E-08 | 0.054662 |
| Arteriolosclerosis | A11.620.520.Myocytes..Smooth.Muscle | Musculoskeletal/Connective | 4.15E-08 | 3.09E-08 | 0.089331 |
| Arteriolosclerosis | Cells_Transformed_fibroblasts | Musculoskeletal/Connective | 3.29E-08 | 2.37E-08 | 0.082631 |
| Arteriolosclerosis | Muscle_Skeletal | Musculoskeletal/Connective | -4.71E-08 | 2.44E-08 | 0.973194 |
| Arteriolosclerosis | A04.411.Lung | Other | 3.94E-08 | 2.81E-08 | 0.080975 |
| Arteriolosclerosis | A04.531.520.Nasal.Mucosa | Other | 2.95E-08 | 2.53E-08 | 0.121611 |
| Arteriolosclerosis | A05.360.319.114.373.Fallopian.Tubes | Other | -2.45E-08 | 2.47E-08 | 0.839951 |
| Arteriolosclerosis | A05.360.319.114.630.Ovary | Other | -3.20E-08 | 2.49E-08 | 0.901324 |
| Arteriolosclerosis | A05.360.319.679.256.Cervix.Uteri | Other | -1.76E-09 | 2.46E-08 | 0.528541 |
| Arteriolosclerosis | A05.360.319.679.490.Endometrium | Other | 5.07E-09 | 2.66E-08 | 0.424489 |
| Arteriolosclerosis | A05.360.319.679.Uterus | Other | -5.71E-09 | 2.51E-08 | 0.589842 |
| Arteriolosclerosis | A05.360.319.887.Vulva | Other | 1.64E-08 | 2.69E-08 | 0.270659 |
| Arteriolosclerosis | A05.360.319.Genitalia..Female | Other | 1.45E-08 | 2.59E-08 | 0.287880 |
| Arteriolosclerosis | A05.360.444.492.362.Foreskin | Other | 1.44E-08 | 2.42E-08 | 0.276433 |
| Arteriolosclerosis | A05.360.444.Genitalia..Male | Other | 2.65E-10 | 2.34E-08 | 0.495483 |
| Arteriolosclerosis | A05.360.490.Germ.Cells | Other | 9.19E-09 | 2.37E-08 | 0.349148 |
| Arteriolosclerosis | A05.360.Genitalia | Other | -8.22E-09 | 2.56E-08 | 0.626092 |
| Arteriolosclerosis | A05.810.453.324.Kidney.Cortex | Other | 1.33E-08 | 2.32E-08 | 0.283404 |
| Arteriolosclerosis | A05.810.453.Kidney | Other | 3.12E-08 | 2.48E-08 | 0.103655 |
| Arteriolosclerosis | A05.810.890.Urinary.Bladder | Other | -2.11E-08 | 2.72E-08 | 0.781151 |
| Arteriolosclerosis | A06.407.071.140.Adrenal.Cortex | Other | 6.74E-08 | 2.72E-08 | 0.006566 |
| Arteriolosclerosis | A06.407.071.Adrenal.Glands | Other | 4.88E-08 | 2.87E-08 | 0.044533 |
| Arteriolosclerosis | A06.407.312.782.Testis | Other | 1.42E-08 | 2.33E-08 | 0.271243 |
| Arteriolosclerosis | A06.407.312.Gonads | Other | -3.17E-08 | 2.41E-08 | 0.905997 |
| Arteriolosclerosis | A06.407.900.Thyroid.Gland | Other | 2.18E-08 | 2.38E-08 | 0.180363 |
| Arteriolosclerosis | A06.407.Endocrine.Glands | Other | -3.91E-09 | 2.56E-08 | 0.560753 |
| Arteriolosclerosis | A09.371.Eye | Other | -1.30E-08 | 2.73E-08 | 0.682530 |
| Arteriolosclerosis | A10.272.497.Epidermis | Other | -9.72E-10 | 2.53E-08 | 0.515318 |
| Arteriolosclerosis | A10.272.Epithelium | Other | 3.15E-08 | 2.53E-08 | 0.106688 |
| Arteriolosclerosis | A10.336.707.Prostate | Other | -5.87E-09 | 2.45E-08 | 0.594484 |
| Arteriolosclerosis | A10.615.284.473.Chorion | Other | 1.77E-08 | 2.53E-08 | 0.241115 |
| Arteriolosclerosis | A10.615.550.599.Mouth.Mucosa | Other | -1.71E-08 | 2.45E-08 | 0.757636 |
| Arteriolosclerosis | A10.615.550.Mucous.Membrane | Other | 3.78E-08 | 2.87E-08 | 0.093813 |
| Arteriolosclerosis | A10.615.789.Serous.Membrane | Other | 9.83E-09 | 2.63E-08 | 0.354248 |
| Arteriolosclerosis | A10.615.Membranes | Other | 2.45E-08 | 2.86E-08 | 0.196030 |

| | | | | | |
|---|---|---|---|---|---|
| Arteriolosclerosis | A11.382.Endocrine.Cells | Other | -2.66E-08 | 2.28E-08 | 0.878407 |
| Arteriolosclerosis | A11.436.275.Endothelial.Cells | Other | 9.85E-09 | 2.75E-08 | 0.360324 |
| Arteriolosclerosis | A11.436.294.064.Glucagon.Secreting.Cells | Other | 4.40E-09 | 2.66E-08 | 0.434322 |
| Arteriolosclerosis | A11.436.329.Granulosa.Cells | Other | 3.13E-09 | 2.35E-08 | 0.447098 |
| Arteriolosclerosis | A11.436.397.Keratinocytes | Other | 1.16E-08 | 2.65E-08 | 0.330831 |
| Arteriolosclerosis | A11.436.Epithelial.Cells | Other | 2.75E-08 | 2.94E-08 | 0.174983 |
| Arteriolosclerosis | A11.497.497.600.Oocytes | Other | 4.98E-09 | 2.38E-08 | 0.417066 |
| Arteriolosclerosis | A11.872.040.Adult.Stem.Cells | Other | 5.99E-09 | 2.78E-08 | 0.414771 |
| Arteriolosclerosis | A11.872.190.260.Embryoid.Bodies | Other | -9.08E-10 | 2.90E-08 | 0.512480 |
| Arteriolosclerosis | A11.872.190.Embryonic.Stem.Cells | Other | 8.90E-09 | 2.70E-08 | 0.370637 |
| Arteriolosclerosis | A11.872.580.Mesenchymal.Stem.Cells | Other | 1.72E-08 | 2.56E-08 | 0.250057 |
| Arteriolosclerosis | A11.872.700.500.Induced.Pluripotent.Stem.Cells | Other | 4.85E-09 | 2.90E-08 | 0.433716 |
| Arteriolosclerosis | A11.872.Stem.Cells | Other | -7.00E-09 | 2.85E-08 | 0.596857 |
| Arteriolosclerosis | A14.549.167.646.Periodontium | Other | 3.29E-08 | 2.86E-08 | 0.125319 |
| Arteriolosclerosis | A14.549.167.Dentition | Other | 2.91E-09 | 2.52E-08 | 0.454164 |
| Arteriolosclerosis | A14.549.885.Tongue | Other | 1.72E-08 | 2.91E-08 | 0.277066 |
| Arteriolosclerosis | A14.549.Mouth | Other | -1.48E-08 | 2.52E-08 | 0.722455 |
| Arteriolosclerosis | A14.724.557.Nasopharynx | Other | 7.57E-09 | 2.64E-08 | 0.387003 |
| Arteriolosclerosis | A14.724.Pharynx | Other | -1.28E-09 | 2.37E-08 | 0.521457 |
| Arteriolosclerosis | A17.815.Skin | Other | -9.92E-09 | 2.60E-08 | 0.648370 |
| Arteriolosclerosis | Adrenal_Gland | Other | 5.27E-08 | 2.64E-08 | 0.022792 |
| Arteriolosclerosis | Bladder | Other | -3.07E-08 | 2.32E-08 | 0.907396 |
| Arteriolosclerosis | Breast_Mammary_Tissue | Other | 9.12E-09 | 2.23E-08 | 0.341132 |
| Arteriolosclerosis | Cervix_Ectocervix | Other | 2.04E-08 | 2.25E-08 | 0.181980 |
| Arteriolosclerosis | Cervix_Endocervix | Other | -1.53E-08 | 2.23E-08 | 0.753644 |
| Arteriolosclerosis | Fallopian_Tube | Other | -1.40E-08 | 2.38E-08 | 0.721785 |
| Arteriolosclerosis | Kidney_Cortex | Other | 5.67E-09 | 2.41E-08 | 0.406956 |
| Arteriolosclerosis | Lung | Other | 2.91E-08 | 2.48E-08 | 0.119878 |
| Arteriolosclerosis | Minor_Salivary_Gland | Other | -3.48E-09 | 2.31E-08 | 0.560018 |
| Arteriolosclerosis | Nerve_Tibial | Other | -2.40E-08 | 2.24E-08 | 0.857750 |
| Arteriolosclerosis | Ovary | Other | -2.55E-08 | 2.34E-08 | 0.862018 |
| Arteriolosclerosis | Pituitary | Other | 2.66E-08 | 2.30E-08 | 0.123952 |
| Arteriolosclerosis | Prostate | Other | -1.12E-08 | 2.70E-08 | 0.660902 |
| Arteriolosclerosis | Skin_Not_Sun_Exposed_(Suprapubic) | Other | -9.49E-09 | 2.62E-08 | 0.641382 |
| Arteriolosclerosis | Skin_Sun_Exposed_(Lower_leg) | Other | -1.90E-08 | 2.66E-08 | 0.761932 |
| Arteriolosclerosis | Testis | Other | 2.86E-08 | 2.57E-08 | 0.132954 |
| Arteriolosclerosis | Thyroid | Other | 1.86E-08 | 2.31E-08 | 0.210489 |
| Arteriolosclerosis | Uterus | Other | -8.16E-09 | 2.25E-08 | 0.641360 |
| Arteriolosclerosis | Vagina | Other | -1.62E-08 | 2.40E-08 | 0.749655 |
| Arteriolosclerosis | A03.734.414.Islets.of.Langerhans | Pancreas | 9.69E-09 | 2.62E-08 | 0.355674 |
| Arteriolosclerosis | A03.734.Pancreas | Pancreas | -7.16E-09 | 2.59E-08 | 0.608854 |
| Arteriolosclerosis | Pancreas | Pancreas | -1.03E-08 | 2.40E-08 | 0.666288 |
| Arteriolosclerosis (any/none) | A10.165.114.830.500.750.Subcutaneous.Fat..Abdominal | Adipose | 3.52E-09 | 2.92E-08 | 0.452018 |
| Arteriolosclerosis (any/none) | A10.165.114.830.750.Subcutaneous.Fat | Adipose | 1.10E-08 | 2.75E-08 | 0.344130 |
| Arteriolosclerosis (any/none) | A11.329.114.Adipocytes | Adipose | -1.38E-08 | 2.61E-08 | 0.700996 |
| Arteriolosclerosis (any/none) | Adipose_Subcutaneous | Adipose | 1.15E-08 | 2.33E-08 | 0.311663 |

| | | | | | |
|---|---|---|---|---|---|
| Arteriolosclerosis (any/none) | Adipose_Visceral_(Omentum) | Adipose | -9.82E-09 | 2.29E-08 | 0.666318 |
| Arteriolosclerosis (any/none) | A02.835.583.443.800.800.Synovial.Fluid | Blood/Immune | -2.32E-08 | 2.99E-08 | 0.781549 |
| Arteriolosclerosis (any/none) | A10.549.400.Lymph.Nodes | Blood/Immune | 4.58E-08 | 3.04E-08 | 0.065797 |
| Arteriolosclerosis (any/none) | A10.549.Lymphoid.Tissue | Blood/Immune | 3.38E-08 | 3.03E-08 | 0.132040 |
| Arteriolosclerosis (any/none) | A11.118.637.555.567.562.440.Precursor.Cells..B.Lymphoid | Blood/Immune | 2.71E-09 | 2.86E-08 | 0.462258 |
| Arteriolosclerosis (any/none) | A11.118.637.555.567.562.B.Lymphocytes | Blood/Immune | -1.87E-09 | 3.07E-08 | 0.524290 |
| Arteriolosclerosis (any/none) | A11.118.637.555.567.569.200.700.T.Lymphocytes..Regulatory | Blood/Immune | 3.33E-08 | 2.86E-08 | 0.121989 |
| Arteriolosclerosis (any/none) | A11.118.637.555.567.569.T.Lymphocytes | Blood/Immune | 2.75E-10 | 2.96E-08 | 0.496292 |
| Arteriolosclerosis (any/none) | A11.118.637.Leukocytes | Blood/Immune | 1.59E-08 | 3.41E-08 | 0.320706 |
| Arteriolosclerosis (any/none) | A11.329.372.600.Macrophages..Alveolar | Blood/Immune | 1.33E-08 | 2.60E-08 | 0.304464 |
| Arteriolosclerosis (any/none) | A11.443.Erythroid.Cells | Blood/Immune | 5.00E-09 | 2.71E-08 | 0.426684 |
| Arteriolosclerosis (any/none) | A11.627.340.360.Granulocyte.Precursor.Cells | Blood/Immune | -5.12E-08 | 3.05E-08 | 0.953469 |
| Arteriolosclerosis (any/none) | A11.627.624.249.Monocyte.Macrophage.Precursor.Cells | Blood/Immune | 2.48E-08 | 2.95E-08 | 0.200560 |
| Arteriolosclerosis (any/none) | A11.627.635.Myeloid.Progenitor.Cells | Blood/Immune | 6.19E-09 | 3.25E-08 | 0.424566 |
| Arteriolosclerosis (any/none) | A11.872.378.590.635.Granulocyte.Macrophage.Progenitor.Cells | Blood/Immune | 7.67E-09 | 3.12E-08 | 0.402944 |
| Arteriolosclerosis (any/none) | A11.872.378.590.817.Megakaryocyte.Erythroid.Progenitor.Cells | Blood/Immune | 1.55E-08 | 2.95E-08 | 0.299978 |
| Arteriolosclerosis (any/none) | A11.872.378.Hematopoietic.Stem.Cells | Blood/Immune | 2.38E-08 | 3.37E-08 | 0.239313 |
| Arteriolosclerosis (any/none) | A15.145.229.188.Blood.Platelets | Blood/Immune | -4.20E-08 | 2.80E-08 | 0.933067 |
| Arteriolosclerosis (any/none) | A15.145.229.637.555.567.562.725.Plasma.Cells | Blood/Immune | 4.17E-08 | 3.28E-08 | 0.101839 |
| Arteriolosclerosis (any/none) | A15.145.229.637.555.567.569.200.CD4.Positive.T.Lymphocytes | Blood/Immune | 1.89E-08 | 3.04E-08 | 0.267538 |
| Arteriolosclerosis (any/none) | A15.145.229.637.555.Leukocytes..Mononuclear | Blood/Immune | 1.06E-08 | 3.12E-08 | 0.366600 |
| Arteriolosclerosis (any/none) | A15.145.229.Blood.Cells | Blood/Immune | -1.24E-08 | 3.13E-08 | 0.653942 |
| Arteriolosclerosis (any/none) | A15.145.300.Fetal.Blood | Blood/Immune | -1.44E-08 | 3.40E-08 | 0.664689 |
| Arteriolosclerosis (any/none) | A15.145.846.Serum | Blood/Immune | -1.70E-08 | 2.77E-08 | 0.730450 |
| Arteriolosclerosis (any/none) | A15.145.Blood | Blood/Immune | -1.84E-08 | 3.24E-08 | 0.715413 |
| Arteriolosclerosis (any/none) | A15.378.316.580.Monocytes | Blood/Immune | 2.14E-08 | 2.85E-08 | 0.225992 |
| Arteriolosclerosis (any/none) | A15.378.316.Bone.Marrow.Cells | Blood/Immune | 2.23E-08 | 3.22E-08 | 0.244479 |
| Arteriolosclerosis (any/none) | A15.382.490.315.583.Neutrophils | Blood/Immune | -6.35E-09 | 3.16E-08 | 0.579569 |
| Arteriolosclerosis (any/none) | A15.382.490.555.567.537.Killer.Cells..Natural | Blood/Immune | -3.46E-08 | 2.80E-08 | 0.891200 |
| Arteriolosclerosis (any/none) | A15.382.490.555.567.622.Lymphocytes..Null | Blood/Immune | -2.67E-09 | 3.20E-08 | 0.533312 |
| Arteriolosclerosis (any/none) | A15.382.490.555.567.Lymphocytes | Blood/Immune | 2.30E-08 | 3.27E-08 | 0.241201 |
| Arteriolosclerosis (any/none) | A15.382.520.604.700.Spleen | Blood/Immune | 3.34E-10 | 3.18E-08 | 0.495806 |
| Arteriolosclerosis (any/none) | A15.382.520.604.800.Palatine.Tonsil | Blood/Immune | 9.50E-09 | 3.01E-08 | 0.376113 |
| Arteriolosclerosis (any/none) | A15.382.680.Phagocytes | Blood/Immune | 3.44E-08 | 3.29E-08 | 0.147834 |
| Arteriolosclerosis (any/none) | A15.382.812.260.Dendritic.Cells | Blood/Immune | -2.60E-08 | 2.71E-08 | 0.831554 |
| Arteriolosclerosis (any/none) | A15.382.812.522.Macrophages | Blood/Immune | 8.11E-09 | 2.98E-08 | 0.392772 |
| Arteriolosclerosis (any/none) | A15.382.812.Mononuclear.Phagocyte.System | Blood/Immune | 1.65E-08 | 2.89E-08 | 0.284783 |
| Arteriolosclerosis (any/none) | A15.382.Immune.System | Blood/Immune | -1.31E-08 | 2.99E-08 | 0.670036 |
| Arteriolosclerosis (any/none) | Cells_EBV-transformed_lymphocytes | Blood/Immune | -2.93E-09 | 2.48E-08 | 0.546986 |
| Arteriolosclerosis (any/none) | Spleen | Blood/Immune | -2.41E-08 | 3.06E-08 | 0.785032 |
| Arteriolosclerosis (any/none) | Whole_Blood | Blood/Immune | -2.56E-08 | 2.91E-08 | 0.810475 |
| Arteriolosclerosis (any/none) | A07.231.114.Arteries | Cardiovascular | 9.77E-09 | 2.76E-08 | 0.361863 |
| Arteriolosclerosis (any/none) | A07.231.908.670.874.Umbilical.Veins | Cardiovascular | -2.04E-08 | 2.71E-08 | 0.774324 |
| Arteriolosclerosis (any/none) | A07.231.908.Veins | Cardiovascular | -4.44E-08 | 2.61E-08 | 0.955171 |
| Arteriolosclerosis (any/none) | A07.231.Blood.Vessels | Cardiovascular | -2.17E-08 | 2.67E-08 | 0.791674 |
| Arteriolosclerosis (any/none) | A07.541.358.100.Atrial.Appendage | Cardiovascular | -2.93E-08 | 2.45E-08 | 0.884524 |

| | | | | | |
|---|---|---|---|---|---|
| Arteriolosclerosis (any/none) | A07.541.358.Heart.Atria | Cardiovascular | -2.27E-08 | 2.47E-08 | 0.820660 |
| Arteriolosclerosis (any/none) | A07.541.510.110.Aortic.Valve | Cardiovascular | 1.19E-08 | 2.69E-08 | 0.328653 |
| Arteriolosclerosis (any/none) | A07.541.560.Heart.Ventricles | Cardiovascular | -3.97E-08 | 2.72E-08 | 0.927907 |
| Arteriolosclerosis (any/none) | A07.541.Heart | Cardiovascular | -4.08E-08 | 2.64E-08 | 0.939364 |
| Arteriolosclerosis (any/none) | Artery_Aorta | Cardiovascular | -1.55E-09 | 2.45E-08 | 0.525218 |
| Arteriolosclerosis (any/none) | Artery_Coronary | Cardiovascular | -2.27E-08 | 2.35E-08 | 0.833619 |
| Arteriolosclerosis (any/none) | Artery_Tibial | Cardiovascular | -8.86E-09 | 2.16E-08 | 0.658994 |
| Arteriolosclerosis (any/none) | Heart_Atrial_Appendage | Cardiovascular | -2.35E-08 | 2.25E-08 | 0.852288 |
| Arteriolosclerosis (any/none) | Heart_Left_Ventricle | Cardiovascular | -6.10E-08 | 2.34E-08 | 0.995426 |
| Arteriolosclerosis (any/none) | A08.186.211.132.810.428.200.Cerebellum | CNS | 2.24E-08 | 2.26E-08 | 0.160260 |
| Arteriolosclerosis (any/none) | A08.186.211.132.Brain.Stem | CNS | 1.46E-08 | 2.46E-08 | 0.276138 |
| Arteriolosclerosis (any/none) | A08.186.211.464.405.Hippocampus | CNS | -2.09E-08 | 2.39E-08 | 0.809217 |
| Arteriolosclerosis (any/none) | A08.186.211.464.710.225.Entorhinal.Cortex | CNS | 3.96E-09 | 2.45E-08 | 0.435626 |
| Arteriolosclerosis (any/none) | A08.186.211.464.Limbic.System | CNS | -4.58E-09 | 2.43E-08 | 0.574772 |
| Arteriolosclerosis (any/none) | A08.186.211.653.Mesencephalon | CNS | -4.26E-08 | 2.44E-08 | 0.959741 |
| Arteriolosclerosis (any/none) | A08.186.211.730.317.357.352.435.Hypothalamo.Hypophyseal.System | CNS | 5.35E-09 | 3.26E-08 | 0.434952 |
| Arteriolosclerosis (any/none) | A08.186.211.730.317.357.Hypothalamus | CNS | 4.42E-09 | 3.41E-08 | 0.448386 |
| Arteriolosclerosis (any/none) | A08.186.211.730.317.Diencephalon | CNS | -3.60E-09 | 2.93E-08 | 0.548808 |
| Arteriolosclerosis (any/none) | A08.186.211.730.885.287.249.487.Corpus.Striatum | CNS | -4.01E-08 | 2.29E-08 | 0.959858 |
| Arteriolosclerosis (any/none) | A08.186.211.730.885.287.249.Basal.Ganglia | CNS | -2.42E-08 | 2.37E-08 | 0.845993 |
| Arteriolosclerosis (any/none) | A08.186.211.730.885.287.500.270.Frontal.Lobe | CNS | -1.86E-08 | 2.49E-08 | 0.772315 |
| Arteriolosclerosis (any/none) | A08.186.211.730.885.287.500.571.735.Visual.Cortex | CNS | 1.47E-08 | 2.34E-08 | 0.265106 |
| Arteriolosclerosis (any/none) | A08.186.211.730.885.287.500.670.Parietal.Lobe | CNS | 1.34E-08 | 2.64E-08 | 0.305996 |
| Arteriolosclerosis (any/none) | A08.186.211.730.885.287.500.Cerebral.Cortex | CNS | -1.86E-08 | 2.54E-08 | 0.768651 |
| Arteriolosclerosis (any/none) | A08.186.211.865.428.Metencephalon | CNS | 1.69E-08 | 2.29E-08 | 0.230228 |
| Arteriolosclerosis (any/none) | A08.186.211.Brain | CNS | -1.31E-08 | 2.39E-08 | 0.707800 |
| Arteriolosclerosis (any/none) | A09.371.729.Retina | CNS | -5.39E-08 | 2.22E-08 | 0.992475 |
| Arteriolosclerosis (any/none) | A11.872.653.Neural.Stem.Cells | CNS | -2.10E-09 | 2.56E-08 | 0.532748 |
| Arteriolosclerosis (any/none) | Brain_Amygdala | CNS | -1.09E-08 | 2.37E-08 | 0.676327 |
| Arteriolosclerosis (any/none) | Brain_Anterior_cingulate_cortex_(BA24) | CNS | -4.27E-09 | 2.25E-08 | 0.575008 |
| Arteriolosclerosis (any/none) | Brain_Caudate_(basal_ganglia) | CNS | -2.58E-08 | 2.26E-08 | 0.872863 |
| Arteriolosclerosis (any/none) | Brain_Cerebellar_Hemisphere | CNS | 5.08E-09 | 2.18E-08 | 0.407864 |
| Arteriolosclerosis (any/none) | Brain_Cerebellum | CNS | 5.57E-09 | 2.20E-08 | 0.399993 |
| Arteriolosclerosis (any/none) | Brain_Cortex | CNS | -9.46E-09 | 2.31E-08 | 0.658642 |
| Arteriolosclerosis (any/none) | Brain_Frontal_Cortex_(BA9) | CNS | -6.62E-09 | 2.20E-08 | 0.618199 |
| Arteriolosclerosis (any/none) | Brain_Hippocampus | CNS | -1.37E-08 | 2.34E-08 | 0.721187 |
| Arteriolosclerosis (any/none) | Brain_Hypothalamus | CNS | -1.10E-08 | 2.41E-08 | 0.676374 |
| Arteriolosclerosis (any/none) | Brain_Nucleus_accumbens_(basal_ganglia) | CNS | -1.71E-08 | 2.24E-08 | 0.777351 |
| Arteriolosclerosis (any/none) | Brain_Putamen_(basal_ganglia) | CNS | -1.65E-08 | 2.40E-08 | 0.754260 |
| Arteriolosclerosis (any/none) | Brain_Spinal_cord_(cervical_c-1) | CNS | 8.49E-09 | 2.33E-08 | 0.357974 |
| Arteriolosclerosis (any/none) | Brain_Substantia_nigra | CNS | -8.93E-09 | 2.42E-08 | 0.644062 |
| Arteriolosclerosis (any/none) | A03.556.124.369.Intestinal.Mucosa | Digestive | 8.19E-09 | 2.90E-08 | 0.388887 |
| Arteriolosclerosis (any/none) | A03.556.124.526.767.Rectum | Digestive | 5.28E-08 | 3.22E-08 | 0.050744 |
| Arteriolosclerosis (any/none) | A03.556.124.684.Intestine..Small | Digestive | 5.32E-08 | 3.34E-08 | 0.055333 |
| Arteriolosclerosis (any/none) | A03.556.124.Intestines | Digestive | 6.33E-08 | 3.28E-08 | 0.026877 |
| Arteriolosclerosis (any/none) | A03.556.249.124.Ileum | Digestive | 7.04E-08 | 3.32E-08 | 0.016980 |

| | | | | | |
|---|---|---|---|---|---|
| Arteriolosclerosis (any/none) | A03.556.249.249.209.Cecum | Digestive | 4.14E-08 | 3.08E-08 | 0.088921 |
| Arteriolosclerosis (any/none) | A03.556.249.249.356.668.Colon..Sigmoid | Digestive | 1.03E-07 | 3.50E-08 | 0.001561 |
| Arteriolosclerosis (any/none) | A03.556.249.249.356.Colon | Digestive | 7.36E-08 | 3.19E-08 | 0.010525 |
| Arteriolosclerosis (any/none) | A03.556.500.760.464.Parotid.Gland | Digestive | -7.34E-09 | 3.11E-08 | 0.593253 |
| Arteriolosclerosis (any/none) | A03.556.500.760.Salivary.Glands | Digestive | -5.69E-09 | 3.02E-08 | 0.574608 |
| Arteriolosclerosis (any/none) | A03.556.875.500.Esophagus | Digestive | 1.05E-08 | 2.59E-08 | 0.342115 |
| Arteriolosclerosis (any/none) | A03.556.875.875.Stomach | Digestive | -3.69E-08 | 3.08E-08 | 0.884855 |
| Arteriolosclerosis (any/none) | A03.556.875.Upper.Gastrointestinal.Tract | Digestive | -1.11E-09 | 2.90E-08 | 0.515255 |
| Arteriolosclerosis (any/none) | A03.556.Gastrointestinal.Tract | Digestive | 5.78E-08 | 3.05E-08 | 0.028899 |
| Arteriolosclerosis (any/none) | Colon_Sigmoid | Digestive | 1.05E-08 | 2.42E-08 | 0.332355 |
| Arteriolosclerosis (any/none) | Colon_Transverse | Digestive | 4.33E-08 | 2.91E-08 | 0.068543 |
| Arteriolosclerosis (any/none) | Esophagus_Gastroesophageal_Junction | Digestive | -7.34E-09 | 2.30E-08 | 0.625014 |
| Arteriolosclerosis (any/none) | Esophagus_Mucosa | Digestive | 4.44E-08 | 2.57E-08 | 0.042057 |
| Arteriolosclerosis (any/none) | Esophagus_Muscularis | Digestive | -2.16E-08 | 2.20E-08 | 0.836871 |
| Arteriolosclerosis (any/none) | Small_Intestine_Terminal_Ileum | Digestive | 3.98E-08 | 2.94E-08 | 0.087684 |
| Arteriolosclerosis (any/none) | Stomach | Digestive | -2.67E-08 | 2.56E-08 | 0.851741 |
| Arteriolosclerosis (any/none) | A03.620.Liver | Liver | 3.98E-08 | 2.92E-08 | 0.086799 |
| Arteriolosclerosis (any/none) | A11.436.348.Hepatocytes | Liver | 2.71E-08 | 3.19E-08 | 0.198432 |
| Arteriolosclerosis (any/none) | Liver | Liver | -3.55E-08 | 2.58E-08 | 0.915618 |
| Arteriolosclerosis (any/none) | A02.165.Cartilage | Musculoskeletal/Connective | 1.01E-08 | 2.87E-08 | 0.362649 |
| Arteriolosclerosis (any/none) | A02.633.567.850.Quadriceps.Muscle | Musculoskeletal/Connective | -4.94E-08 | 2.77E-08 | 0.962560 |
| Arteriolosclerosis (any/none) | A02.835.232.834.151.Cervical.Vertebrae | Musculoskeletal/Connective | -9.37E-09 | 2.91E-08 | 0.626335 |
| Arteriolosclerosis (any/none) | A02.835.583.443.800.Synovial.Membrane | Musculoskeletal/Connective | 1.55E-08 | 2.63E-08 | 0.277476 |
| Arteriolosclerosis (any/none) | A05.360.319.679.690.Myometrium | Musculoskeletal/Connective | -4.37E-09 | 2.58E-08 | 0.567233 |
| Arteriolosclerosis (any/none) | A10.165.450.300.425.Keloid | Musculoskeletal/Connective | 1.94E-08 | 2.48E-08 | 0.217112 |
| Arteriolosclerosis (any/none) | A10.165.450.300.Cicatrix | Musculoskeletal/Connective | -1.30E-09 | 2.63E-08 | 0.519655 |
| Arteriolosclerosis (any/none) | A10.690.467.Muscle..Smooth | Musculoskeletal/Connective | -4.04E-11 | 2.78E-08 | 0.500580 |
| Arteriolosclerosis (any/none) | A10.690.Muscles | Musculoskeletal/Connective | -2.51E-08 | 2.85E-08 | 0.810201 |
| Arteriolosclerosis (any/none) | A11.329.171.Chondrocytes | Musculoskeletal/Connective | 2.12E-08 | 2.70E-08 | 0.215878 |
| Arteriolosclerosis (any/none) | A11.329.228.Fibroblasts | Musculoskeletal/Connective | 3.06E-08 | 2.59E-08 | 0.118475 |
| Arteriolosclerosis (any/none) | A11.329.629.Osteoblasts | Musculoskeletal/Connective | -8.53E-09 | 2.69E-08 | 0.624394 |
| Arteriolosclerosis (any/none) | A11.329.830.Stromal.Cells | Musculoskeletal/Connective | 2.22E-08 | 2.73E-08 | 0.208598 |
| Arteriolosclerosis (any/none) | A11.329.Connective.Tissue.Cells | Musculoskeletal/Connective | 3.13E-08 | 2.63E-08 | 0.116863 |
| Arteriolosclerosis (any/none) | A11.620.520.Myocytes..Smooth.Muscle | Musculoskeletal/Connective | 7.16E-09 | 2.72E-08 | 0.396142 |
| Arteriolosclerosis (any/none) | Cells_Transformed_fibroblasts | Musculoskeletal/Connective | 3.39E-08 | 2.53E-08 | 0.089763 |
| Arteriolosclerosis (any/none) | Muscle_Skeletal | Musculoskeletal/Connective | -2.87E-08 | 2.49E-08 | 0.875267 |
| Arteriolosclerosis (any/none) | A04.411.Lung | Other | 3.90E-08 | 3.13E-08 | 0.106796 |
| Arteriolosclerosis (any/none) | A04.531.520.Nasal.Mucosa | Other | 2.05E-08 | 2.87E-08 | 0.237884 |
| Arteriolosclerosis (any/none) | A05.360.319.114.373.Fallopian.Tubes | Other | 2.25E-08 | 2.84E-08 | 0.214297 |
| Arteriolosclerosis (any/none) | A05.360.319.114.630.Ovary | Other | -1.63E-08 | 2.79E-08 | 0.721254 |
| Arteriolosclerosis (any/none) | A05.360.319.679.256.Cervix.Uteri | Other | 1.13E-08 | 2.57E-08 | 0.329699 |
| Arteriolosclerosis (any/none) | A05.360.319.679.490.Endometrium | Other | 7.01E-09 | 2.88E-08 | 0.403881 |
| Arteriolosclerosis (any/none) | A05.360.319.679.Uterus | Other | 1.95E-08 | 2.80E-08 | 0.243224 |
| Arteriolosclerosis (any/none) | A05.360.319.887.Vulva | Other | 2.78E-08 | 2.95E-08 | 0.172400 |
| Arteriolosclerosis (any/none) | A05.360.319.Genitalia..Female | Other | 2.39E-08 | 2.72E-08 | 0.189584 |
| Arteriolosclerosis (any/none) | A05.360.444.492.362.Foreskin | Other | 1.69E-08 | 2.45E-08 | 0.245865 |

| | | | | | |
|---|---|---|---|---|---|
| Arteriolosclerosis (any/none) | A05.360.444.Genitalia..Male | Other | -1.97E-08 | 2.58E-08 | 0.777200 |
| Arteriolosclerosis (any/none) | A05.360.490.Germ.Cells | Other | 2.42E-08 | 2.46E-08 | 0.162551 |
| Arteriolosclerosis (any/none) | A05.360.Genitalia | Other | -1.43E-08 | 2.61E-08 | 0.708109 |
| Arteriolosclerosis (any/none) | A05.810.453.324.Kidney.Cortex | Other | 4.71E-10 | 2.74E-08 | 0.493145 |
| Arteriolosclerosis (any/none) | A05.810.453.Kidney | Other | 1.50E-08 | 2.49E-08 | 0.273686 |
| Arteriolosclerosis (any/none) | A05.810.890.Urinary.Bladder | Other | -6.63E-09 | 3.05E-08 | 0.585920 |
| Arteriolosclerosis (any/none) | A06.407.071.140.Adrenal.Cortex | Other | 3.97E-08 | 3.16E-08 | 0.104904 |
| Arteriolosclerosis (any/none) | A06.407.071.Adrenal.Glands | Other | 3.38E-08 | 3.20E-08 | 0.145601 |
| Arteriolosclerosis (any/none) | A06.407.312.782.Testis | Other | 3.97E-09 | 2.52E-08 | 0.437327 |
| Arteriolosclerosis (any/none) | A06.407.312.Gonads | Other | -6.31E-09 | 2.72E-08 | 0.591686 |
| Arteriolosclerosis (any/none) | A06.407.900.Thyroid.Gland | Other | 1.00E-08 | 2.35E-08 | 0.334474 |
| Arteriolosclerosis (any/none) | A06.407.Endocrine.Glands | Other | 3.45E-08 | 2.85E-08 | 0.113216 |
| Arteriolosclerosis (any/none) | A09.371.Eye | Other | 2.38E-08 | 2.84E-08 | 0.200703 |
| Arteriolosclerosis (any/none) | A10.272.497.Epidermis | Other | 1.81E-08 | 2.72E-08 | 0.252927 |
| Arteriolosclerosis (any/none) | A10.272.Epithelium | Other | 1.03E-09 | 2.92E-08 | 0.485926 |
| Arteriolosclerosis (any/none) | A10.336.707.Prostate | Other | -1.78E-08 | 2.60E-08 | 0.753459 |
| Arteriolosclerosis (any/none) | A10.615.284.473.Chorion | Other | 5.40E-09 | 2.74E-08 | 0.421725 |
| Arteriolosclerosis (any/none) | A10.615.550.599.Mouth.Mucosa | Other | -2.09E-08 | 2.64E-08 | 0.785862 |
| Arteriolosclerosis (any/none) | A10.615.550.Mucous.Membrane | Other | 5.55E-08 | 2.97E-08 | 0.030919 |
| Arteriolosclerosis (any/none) | A10.615.789.Serous.Membrane | Other | -3.91E-10 | 2.84E-08 | 0.505488 |
| Arteriolosclerosis (any/none) | A10.615.Membranes | Other | 4.89E-08 | 2.99E-08 | 0.051046 |
| Arteriolosclerosis (any/none) | A11.382.Endocrine.Cells | Other | 8.07E-09 | 2.46E-08 | 0.371524 |
| Arteriolosclerosis (any/none) | A11.436.275.Endothelial.Cells | Other | -2.16E-08 | 2.71E-08 | 0.786638 |
| Arteriolosclerosis (any/none) | A11.436.294.064.Glucagon.Secreting.Cells | Other | 3.78E-08 | 2.79E-08 | 0.087669 |
| Arteriolosclerosis (any/none) | A11.436.329.Granulosa.Cells | Other | 2.05E-08 | 2.71E-08 | 0.224055 |
| Arteriolosclerosis (any/none) | A11.436.397.Keratinocytes | Other | 2.05E-08 | 2.99E-08 | 0.246489 |
| Arteriolosclerosis (any/none) | A11.436.Epithelial.Cells | Other | 1.53E-08 | 2.82E-08 | 0.293005 |
| Arteriolosclerosis (any/none) | A11.497.497.600.Oocytes | Other | 1.24E-08 | 2.36E-08 | 0.299543 |
| Arteriolosclerosis (any/none) | A11.872.040.Adult.Stem.Cells | Other | -2.86E-08 | 2.91E-08 | 0.837487 |
| Arteriolosclerosis (any/none) | A11.872.190.260.Embryoid.Bodies | Other | -2.14E-08 | 2.51E-08 | 0.803424 |
| Arteriolosclerosis (any/none) | A11.872.190.Embryonic.Stem.Cells | Other | -2.22E-09 | 2.67E-08 | 0.533144 |
| Arteriolosclerosis (any/none) | A11.872.580.Mesenchymal.Stem.Cells | Other | -1.32E-08 | 2.53E-08 | 0.698266 |
| Arteriolosclerosis (any/none) | A11.872.700.500.Induced.Pluripotent.Stem.Cells | Other | -2.79E-08 | 2.79E-08 | 0.841288 |
| Arteriolosclerosis (any/none) | A11.872.Stem.Cells | Other | 1.25E-08 | 3.29E-08 | 0.352245 |
| Arteriolosclerosis (any/none) | A14.549.167.646.Periodontium | Other | -2.68E-10 | 2.91E-08 | 0.503665 |
| Arteriolosclerosis (any/none) | A14.549.167.Dentition | Other | -1.74E-08 | 2.71E-08 | 0.739619 |
| Arteriolosclerosis (any/none) | A14.549.885.Tongue | Other | 1.42E-08 | 2.76E-08 | 0.303399 |
| Arteriolosclerosis (any/none) | A14.549.Mouth | Other | -2.36E-08 | 2.55E-08 | 0.822556 |
| Arteriolosclerosis (any/none) | A14.724.557.Nasopharynx | Other | 1.40E-08 | 2.47E-08 | 0.286033 |
| Arteriolosclerosis (any/none) | A14.724.Pharynx | Other | 8.10E-09 | 2.65E-08 | 0.379961 |
| Arteriolosclerosis (any/none) | A17.815.Skin | Other | 1.68E-09 | 2.69E-08 | 0.475112 |
| Arteriolosclerosis (any/none) | Adrenal_Gland | Other | 5.58E-08 | 2.78E-08 | 0.022488 |
| Arteriolosclerosis (any/none) | Bladder | Other | -1.96E-08 | 2.37E-08 | 0.795533 |
| Arteriolosclerosis (any/none) | Breast_Mammary_Tissue | Other | -8.20E-09 | 2.58E-08 | 0.624638 |
| Arteriolosclerosis (any/none) | Cervix_Ectocervix | Other | -7.91E-09 | 2.44E-08 | 0.627073 |
| Arteriolosclerosis (any/none) | Cervix_Endocervix | Other | -1.20E-08 | 2.26E-08 | 0.702231 |

| | | | | | |
|---|---|---|---|---|---|
| Arteriolosclerosis (any/none) | Fallopian_Tube | Other | -2.24E-08 | 2.35E-08 | 0.829605 |
| Arteriolosclerosis (any/none) | Kidney_Cortex | Other | 1.06E-08 | 2.71E-08 | 0.347683 |
| Arteriolosclerosis (any/none) | Lung | Other | -1.17E-08 | 2.65E-08 | 0.670438 |
| Arteriolosclerosis (any/none) | Minor_Salivary_Gland | Other | -2.14E-08 | 2.42E-08 | 0.811570 |
| Arteriolosclerosis (any/none) | Nerve_Tibial | Other | -4.11E-09 | 2.46E-08 | 0.566368 |
| Arteriolosclerosis (any/none) | Ovary | Other | -1.50E-08 | 2.38E-08 | 0.735890 |
| Arteriolosclerosis (any/none) | Pituitary | Other | 5.11E-08 | 2.38E-08 | 0.015751 |
| Arteriolosclerosis (any/none) | Prostate | Other | -1.69E-08 | 2.93E-08 | 0.717536 |
| Arteriolosclerosis (any/none) | Skin_Not_Sun_Exposed_(Suprapubic) | Other | 5.13E-11 | 2.54E-08 | 0.499193 |
| Arteriolosclerosis (any/none) | Skin_Sun_Exposed_(Lower_leg) | Other | -8.91E-09 | 2.41E-08 | 0.643994 |
| Arteriolosclerosis (any/none) | Testis | Other | 4.08E-08 | 2.41E-08 | 0.045042 |
| Arteriolosclerosis (any/none) | Thyroid | Other | 4.36E-08 | 2.47E-08 | 0.038859 |
| Arteriolosclerosis (any/none) | Uterus | Other | -2.65E-08 | 2.55E-08 | 0.850867 |
| Arteriolosclerosis (any/none) | Vagina | Other | -1.23E-08 | 2.19E-08 | 0.712805 |
| Arteriolosclerosis (any/none) | A03.734.414.Islets.of.Langerhans | Pancreas | 1.86E-08 | 2.83E-08 | 0.255284 |
| Arteriolosclerosis (any/none) | A03.734.Pancreas | Pancreas | 3.86E-08 | 2.86E-08 | 0.088428 |
| Arteriolosclerosis (any/none) | Pancreas | Pancreas | -1.62E-09 | 2.37E-08 | 0.527309 |
| Atherosclerosis | A10.165.114.830.500.750.Subcutaneous.Fat..Abdominal | Adipose | -2.25E-08 | 1.89E-08 | 0.882365 |
| Atherosclerosis | A10.165.114.830.750.Subcutaneous.Fat | Adipose | -6.74E-09 | 1.94E-08 | 0.636076 |
| Atherosclerosis | A11.329.114.Adipocytes | Adipose | -1.99E-08 | 1.98E-08 | 0.842981 |
| Atherosclerosis | Adipose_Subcutaneous | Adipose | -1.45E-08 | 1.83E-08 | 0.785553 |
| Atherosclerosis | Adipose_Visceral_(Omentum) | Adipose | -3.36E-08 | 1.84E-08 | 0.965887 |
| Atherosclerosis | A02.835.583.443.800.800.Synovial.Fluid | Blood/Immune | 8.86E-09 | 2.55E-08 | 0.364213 |
| Atherosclerosis | A10.549.400.Lymph.Nodes | Blood/Immune | 2.14E-08 | 2.22E-08 | 0.167516 |
| Atherosclerosis | A10.549.Lymphoid.Tissue | Blood/Immune | 2.64E-08 | 2.26E-08 | 0.121249 |
| Atherosclerosis | A11.118.637.555.567.562.440.Precursor.Cells..B.Lymphoid | Blood/Immune | 3.48E-09 | 2.39E-08 | 0.442035 |
| Atherosclerosis | A11.118.637.555.567.562.B.Lymphocytes | Blood/Immune | 2.48E-08 | 2.40E-08 | 0.150368 |
| Atherosclerosis | A11.118.637.555.567.569.200.700.T.Lymphocytes..Regulatory | Blood/Immune | 2.72E-08 | 2.48E-08 | 0.135702 |
| Atherosclerosis | A11.118.637.555.567.569.T.Lymphocytes | Blood/Immune | 1.55E-08 | 2.41E-08 | 0.259857 |
| Atherosclerosis | A11.118.637.Leukocytes | Blood/Immune | -4.50E-09 | 2.77E-08 | 0.564451 |
| Atherosclerosis | A11.329.372.600.Macrophages..Alveolar | Blood/Immune | 8.20E-09 | 1.99E-08 | 0.340092 |
| Atherosclerosis | A11.443.Erythroid.Cells | Blood/Immune | -1.93E-08 | 2.26E-08 | 0.804200 |
| Atherosclerosis | A11.627.340.360.Granulocyte.Precursor.Cells | Blood/Immune | 5.79E-08 | 2.21E-08 | 0.004391 |
| Atherosclerosis | A11.627.624.249.Monocyte.Macrophage.Precursor.Cells | Blood/Immune | 1.26E-08 | 2.70E-08 | 0.320084 |
| Atherosclerosis | A11.627.635.Myeloid.Progenitor.Cells | Blood/Immune | 2.19E-08 | 2.36E-08 | 0.176369 |
| Atherosclerosis | A11.872.378.590.635.Granulocyte.Macrophage.Progenitor.Cells | Blood/Immune | 2.03E-08 | 2.83E-08 | 0.236403 |
| Atherosclerosis | A11.872.378.590.817.Megakaryocyte.Erythroid.Progenitor.Cells | Blood/Immune | 1.53E-08 | 2.39E-08 | 0.260472 |
| Atherosclerosis | A11.872.378.Hematopoietic.Stem.Cells | Blood/Immune | 7.72E-09 | 2.34E-08 | 0.370527 |
| Atherosclerosis | A15.145.229.188.Blood.Platelets | Blood/Immune | 1.66E-08 | 2.39E-08 | 0.243423 |
| Atherosclerosis | A15.145.229.637.555.567.562.725.Plasma.Cells | Blood/Immune | 1.99E-08 | 2.26E-08 | 0.189920 |
| Atherosclerosis | A15.145.229.637.555.567.569.200.CD4.Positive.T.Lymphocytes | Blood/Immune | 5.24E-10 | 2.60E-08 | 0.491976 |
| Atherosclerosis | A15.145.229.637.555.Leukocytes..Mononuclear | Blood/Immune | -2.31E-08 | 2.46E-08 | 0.826626 |
| Atherosclerosis | A15.145.229.Blood.Cells | Blood/Immune | -4.97E-09 | 2.45E-08 | 0.580328 |
| Atherosclerosis | A15.145.300.Fetal.Blood | Blood/Immune | -9.55E-09 | 2.91E-08 | 0.628851 |
| Atherosclerosis | A15.145.846.Serum | Blood/Immune | 4.04E-08 | 2.43E-08 | 0.047910 |
| Atherosclerosis | A15.145.Blood | Blood/Immune | -6.41E-09 | 2.48E-08 | 0.601937 |

| | | | | | |
|---|---|---|---|---|---|
| Atherosclerosis | A15.378.316.580.Monocytes | Blood/Immune | 1.71E-08 | 2.78E-08 | 0.269102 |
| Atherosclerosis | A15.378.316.Bone.Marrow.Cells | Blood/Immune | 1.95E-08 | 2.40E-08 | 0.207900 |
| Atherosclerosis | A15.382.490.315.583.Neutrophils | Blood/Immune | 3.55E-08 | 2.37E-08 | 0.067082 |
| Atherosclerosis | A15.382.490.555.567.537.Killer.Cells..Natural | Blood/Immune | 3.01E-08 | 2.40E-08 | 0.105161 |
| Atherosclerosis | A15.382.490.555.567.622.Lymphocytes..Null | Blood/Immune | 2.75E-08 | 2.32E-08 | 0.118200 |
| Atherosclerosis | A15.382.490.555.567.Lymphocytes | Blood/Immune | -9.30E-09 | 2.60E-08 | 0.639779 |
| Atherosclerosis | A15.382.520.604.700.Spleen | Blood/Immune | 1.09E-08 | 2.38E-08 | 0.324003 |
| Atherosclerosis | A15.382.520.604.800.Palatine.Tonsil | Blood/Immune | 3.26E-08 | 2.22E-08 | 0.071064 |
| Atherosclerosis | A15.382.680.Phagocytes | Blood/Immune | 1.63E-08 | 2.42E-08 | 0.250477 |
| Atherosclerosis | A15.382.812.260.Dendritic.Cells | Blood/Immune | 1.10E-08 | 2.58E-08 | 0.334339 |
| Atherosclerosis | A15.382.812.522.Macrophages | Blood/Immune | 3.83E-08 | 2.29E-08 | 0.046851 |
| Atherosclerosis | A15.382.812.Mononuclear.Phagocyte.System | Blood/Immune | 3.31E-08 | 2.56E-08 | 0.098000 |
| Atherosclerosis | A15.382.Immune.System | Blood/Immune | 1.73E-08 | 2.56E-08 | 0.250296 |
| Atherosclerosis | Cells_EBV-transformed_lymphocytes | Blood/Immune | -3.75E-08 | 2.14E-08 | 0.959710 |
| Atherosclerosis | Spleen | Blood/Immune | 1.37E-08 | 2.47E-08 | 0.288716 |
| Atherosclerosis | Whole_Blood | Blood/Immune | 3.88E-08 | 2.40E-08 | 0.052629 |
| Atherosclerosis | A07.231.114.Arteries | Cardiovascular | -2.35E-08 | 2.32E-08 | 0.844874 |
| Atherosclerosis | A07.231.908.670.874.Umbilical.Veins | Cardiovascular | 9.61E-09 | 2.33E-08 | 0.339970 |
| Atherosclerosis | A07.231.908.Veins | Cardiovascular | -7.22E-10 | 2.30E-08 | 0.512509 |
| Atherosclerosis | A07.231.Blood.Vessels | Cardiovascular | -5.73E-09 | 2.36E-08 | 0.595944 |
| Atherosclerosis | A07.541.358.100.Atrial.Appendage | Cardiovascular | -2.66E-09 | 2.15E-08 | 0.549140 |
| Atherosclerosis | A07.541.358.Heart.Atria | Cardiovascular | -6.55E-09 | 2.17E-08 | 0.618371 |
| Atherosclerosis | A07.541.510.110.Aortic.Valve | Cardiovascular | -2.82E-08 | 1.88E-08 | 0.933327 |
| Atherosclerosis | A07.541.560.Heart.Ventricles | Cardiovascular | -1.08E-08 | 2.08E-08 | 0.698417 |
| Atherosclerosis | A07.541.Heart | Cardiovascular | -9.89E-09 | 2.01E-08 | 0.689066 |
| Atherosclerosis | Artery_Aorta | Cardiovascular | 3.16E-09 | 1.81E-08 | 0.430574 |
| Atherosclerosis | Artery_Coronary | Cardiovascular | 1.05E-08 | 1.84E-08 | 0.284951 |
| Atherosclerosis | Artery_Tibial | Cardiovascular | 1.15E-08 | 1.91E-08 | 0.272612 |
| Atherosclerosis | Heart_Atrial_Appendage | Cardiovascular | 1.77E-08 | 1.83E-08 | 0.167113 |
| Atherosclerosis | Heart_Left_Ventricle | Cardiovascular | 4.17E-09 | 1.84E-08 | 0.410189 |
| Atherosclerosis | A08.186.211.132.810.428.200.Cerebellum | CNS | -3.65E-08 | 1.91E-08 | 0.972068 |
| Atherosclerosis | A08.186.211.132.Brain.Stem | CNS | -3.94E-08 | 1.82E-08 | 0.984618 |
| Atherosclerosis | A08.186.211.464.405.Hippocampus | CNS | -3.55E-08 | 2.04E-08 | 0.959379 |
| Atherosclerosis | A08.186.211.464.710.225.Entorhinal.Cortex | CNS | -3.71E-08 | 1.97E-08 | 0.969870 |
| Atherosclerosis | A08.186.211.464.Limbic.System | CNS | -4.10E-08 | 2.01E-08 | 0.979568 |
| Atherosclerosis | A08.186.211.653.Mesencephalon | CNS | -2.01E-08 | 2.21E-08 | 0.818769 |
| Atherosclerosis | A08.186.211.730.317.357.352.435.Hypothalamo.Hypophyseal.System | CNS | -2.47E-08 | 2.18E-08 | 0.872371 |
| Atherosclerosis | A08.186.211.730.317.357.Hypothalamus | CNS | -1.59E-08 | 2.23E-08 | 0.761719 |
| Atherosclerosis | A08.186.211.730.317.Diencephalon | CNS | 7.46E-10 | 2.22E-08 | 0.486597 |
| Atherosclerosis | A08.186.211.730.885.287.249.487.Corpus.Striatum | CNS | 6.68E-09 | 2.15E-08 | 0.378052 |
| Atherosclerosis | A08.186.211.730.885.287.249.Basal.Ganglia | CNS | -3.06E-09 | 2.21E-08 | 0.555039 |
| Atherosclerosis | A08.186.211.730.885.287.500.270.Frontal.Lobe | CNS | -9.45E-09 | 2.21E-08 | 0.665514 |
| Atherosclerosis | A08.186.211.730.885.287.500.571.735.Visual.Cortex | CNS | -1.28E-08 | 2.01E-08 | 0.737680 |
| Atherosclerosis | A08.186.211.730.885.287.500.670.Parietal.Lobe | CNS | -1.71E-08 | 2.05E-08 | 0.797403 |
| Atherosclerosis | A08.186.211.730.885.287.500.Cerebral.Cortex | CNS | -3.12E-08 | 2.05E-08 | 0.936074 |
| Atherosclerosis | A08.186.211.865.428.Metencephalon | CNS | -2.73E-08 | 1.90E-08 | 0.925202 |

| | | | | | |
|---|---|---|---|---|---|
| Atherosclerosis | A08.186.211.Brain | CNS | -2.41E-08 | 2.12E-08 | 0.872340 |
| Atherosclerosis | A09.371.729.Retina | CNS | 1.17E-08 | 2.06E-08 | 0.284589 |
| Atherosclerosis | A11.872.653.Neural.Stem.Cells | CNS | -2.15E-08 | 2.09E-08 | 0.848657 |
| Atherosclerosis | Brain_Amygdala | CNS | -1.96E-08 | 1.69E-08 | 0.876318 |
| Atherosclerosis | Brain_Anterior_cingulate_cortex_(BA24) | CNS | -2.26E-08 | 1.78E-08 | 0.898310 |
| Atherosclerosis | Brain_Caudate_(basal_ganglia) | CNS | -1.59E-08 | 1.79E-08 | 0.813327 |
| Atherosclerosis | Brain_Cerebellar_Hemisphere | CNS | -8.33E-09 | 1.87E-08 | 0.672413 |
| Atherosclerosis | Brain_Cerebellum | CNS | -6.83E-09 | 1.83E-08 | 0.645921 |
| Atherosclerosis | Brain_Cortex | CNS | -1.62E-08 | 1.75E-08 | 0.823488 |
| Atherosclerosis | Brain_Frontal_Cortex_(BA9) | CNS | -1.96E-08 | 1.70E-08 | 0.875890 |
| Atherosclerosis | Brain_Hippocampus | CNS | -2.16E-08 | 1.68E-08 | 0.900235 |
| Atherosclerosis | Brain_Hypothalamus | CNS | -2.41E-08 | 1.68E-08 | 0.923763 |
| Atherosclerosis | Brain_Nucleus_accumbens_(basal_ganglia) | CNS | -8.27E-09 | 1.75E-08 | 0.681594 |
| Atherosclerosis | Brain_Putamen_(basal_ganglia) | CNS | -2.03E-08 | 1.71E-08 | 0.882567 |
| Atherosclerosis | Brain_Spinal_cord_(cervical_c-1) | CNS | -1.20E-08 | 1.80E-08 | 0.748257 |
| Atherosclerosis | Brain_Substantia_nigra | CNS | -2.62E-08 | 1.72E-08 | 0.936133 |
| Atherosclerosis | A03.556.124.369.Intestinal.Mucosa | Digestive | 4.04E-10 | 2.42E-08 | 0.493329 |
| Atherosclerosis | A03.556.124.526.767.Rectum | Digestive | -1.09E-08 | 2.28E-08 | 0.683481 |
| Atherosclerosis | A03.556.124.684.Intestine..Small | Digestive | 7.29E-09 | 2.42E-08 | 0.381692 |
| Atherosclerosis | A03.556.124.Intestines | Digestive | -3.04E-09 | 2.29E-08 | 0.552753 |
| Atherosclerosis | A03.556.249.124.Ileum | Digestive | 2.50E-08 | 2.46E-08 | 0.154639 |
| Atherosclerosis | A03.556.249.249.209.Cecum | Digestive | -9.22E-09 | 2.28E-08 | 0.657300 |
| Atherosclerosis | A03.556.249.249.356.668.Colon..Sigmoid | Digestive | 7.74E-09 | 2.30E-08 | 0.368569 |
| Atherosclerosis | A03.556.249.249.356.Colon | Digestive | -2.40E-08 | 2.32E-08 | 0.849367 |
| Atherosclerosis | A03.556.500.760.464.Parotid.Gland | Digestive | -3.11E-09 | 2.65E-08 | 0.546647 |
| Atherosclerosis | A03.556.500.760.Salivary.Glands | Digestive | 1.27E-08 | 2.74E-08 | 0.321378 |
| Atherosclerosis | A03.556.875.500.Esophagus | Digestive | 1.39E-08 | 2.25E-08 | 0.267903 |
| Atherosclerosis | A03.556.875.875.Stomach | Digestive | -4.32E-10 | 2.66E-08 | 0.506477 |
| Atherosclerosis | A03.556.875.Upper.Gastrointestinal.Tract | Digestive | 1.36E-08 | 2.50E-08 | 0.292873 |
| Atherosclerosis | A03.556.Gastrointestinal.Tract | Digestive | -1.23E-08 | 2.20E-08 | 0.712487 |
| Atherosclerosis | Colon_Sigmoid | Digestive | 2.32E-08 | 2.13E-08 | 0.138120 |
| Atherosclerosis | Colon_Transverse | Digestive | -1.11E-08 | 2.43E-08 | 0.675584 |
| Atherosclerosis | Esophagus_Gastroesophageal_Junction | Digestive | -1.18E-09 | 2.11E-08 | 0.522197 |
| Atherosclerosis | Esophagus_Mucosa | Digestive | -1.02E-08 | 2.25E-08 | 0.674902 |
| Atherosclerosis | Esophagus_Muscularis | Digestive | 1.04E-08 | 2.15E-08 | 0.313549 |
| Atherosclerosis | Small_Intestine_Terminal_Ileum | Digestive | 1.33E-08 | 2.07E-08 | 0.259255 |
| Atherosclerosis | Stomach | Digestive | 1.79E-08 | 2.10E-08 | 0.197223 |
| Atherosclerosis | A03.620.Liver | Liver | 1.26E-08 | 2.40E-08 | 0.299357 |
| Atherosclerosis | A11.436.348.Hepatocytes | Liver | 2.88E-08 | 2.64E-08 | 0.137740 |
| Atherosclerosis | Liver | Liver | 2.74E-08 | 2.24E-08 | 0.110356 |
| Atherosclerosis | A02.165.Cartilage | Musculoskeletal/Connective | 2.11E-08 | 1.85E-08 | 0.126768 |
| Atherosclerosis | A02.633.567.850.Quadriceps.Muscle | Musculoskeletal/Connective | 1.44E-08 | 2.43E-08 | 0.277448 |
| Atherosclerosis | A02.835.232.834.151.Cervical.Vertebrae | Musculoskeletal/Connective | -1.62E-08 | 2.15E-08 | 0.774161 |
| Atherosclerosis | A02.835.583.443.800.Synovial.Membrane | Musculoskeletal/Connective | 1.20E-10 | 2.11E-08 | 0.497730 |
| Atherosclerosis | A05.360.319.679.690.Myometrium | Musculoskeletal/Connective | 6.41E-09 | 2.18E-08 | 0.384190 |
| Atherosclerosis | A10.165.450.300.425.Keloid | Musculoskeletal/Connective | -1.53E-08 | 2.15E-08 | 0.761402 |

| | | | | | |
|---|---|---|---|---|---|
| Atherosclerosis | A10.165.450.300.Cicatrix | Musculoskeletal/Connective | -1.53E-08 | 2.08E-08 | 0.769725 |
| Atherosclerosis | A10.690.467.Muscle..Smooth | Musculoskeletal/Connective | 1.29E-09 | 2.23E-08 | 0.476918 |
| Atherosclerosis | A10.690.Muscles | Musculoskeletal/Connective | 3.27E-09 | 2.58E-08 | 0.449621 |
| Atherosclerosis | A11.329.171.Chondrocytes | Musculoskeletal/Connective | -2.31E-08 | 2.01E-08 | 0.875325 |
| Atherosclerosis | A11.329.228.Fibroblasts | Musculoskeletal/Connective | 5.68E-09 | 2.19E-08 | 0.397677 |
| Atherosclerosis | A11.329.629.Osteoblasts | Musculoskeletal/Connective | -1.28E-08 | 2.11E-08 | 0.727302 |
| Atherosclerosis | A11.329.830.Stromal.Cells | Musculoskeletal/Connective | 1.80E-08 | 2.31E-08 | 0.218164 |
| Atherosclerosis | A11.329.Connective.Tissue.Cells | Musculoskeletal/Connective | 5.45E-09 | 2.26E-08 | 0.404791 |
| Atherosclerosis | A11.620.520.Myocytes..Smooth.Muscle | Musculoskeletal/Connective | -2.26E-09 | 2.36E-08 | 0.538170 |
| Atherosclerosis | Cells_Transformed_fibroblasts | Musculoskeletal/Connective | -6.76E-09 | 2.15E-08 | 0.623045 |
| Atherosclerosis | Muscle_Skeletal | Musculoskeletal/Connective | 1.93E-08 | 2.17E-08 | 0.186612 |
| Atherosclerosis | A04.411.Lung | Other | -2.73E-08 | 2.47E-08 | 0.865110 |
| Atherosclerosis | A04.531.520.Nasal.Mucosa | Other | 1.97E-08 | 2.35E-08 | 0.200839 |
| Atherosclerosis | A05.360.319.114.373.Fallopian.Tubes | Other | 4.25E-09 | 2.37E-08 | 0.428808 |
| Atherosclerosis | A05.360.319.114.630.Ovary | Other | -2.20E-08 | 2.36E-08 | 0.824304 |
| Atherosclerosis | A05.360.319.679.256.Cervix.Uteri | Other | 2.16E-08 | 2.40E-08 | 0.183202 |
| Atherosclerosis | A05.360.319.679.490.Endometrium | Other | 3.06E-08 | 2.13E-08 | 0.075705 |
| Atherosclerosis | A05.360.319.679.Uterus | Other | 4.82E-09 | 1.96E-08 | 0.403016 |
| Atherosclerosis | A05.360.319.887.Vulva | Other | 1.94E-08 | 2.38E-08 | 0.207412 |
| Atherosclerosis | A05.360.319.Genitalia..Female | Other | -3.07E-09 | 2.04E-08 | 0.559871 |
| Atherosclerosis | A05.360.444.492.362.Foreskin | Other | 2.23E-08 | 2.17E-08 | 0.152354 |
| Atherosclerosis | A05.360.444.Genitalia..Male | Other | -1.61E-08 | 1.82E-08 | 0.811387 |
| Atherosclerosis | A05.360.490.Germ.Cells | Other | 6.03E-08 | 2.22E-08 | 0.003304 |
| Atherosclerosis | A05.360.Genitalia | Other | -2.92E-09 | 2.02E-08 | 0.557520 |
| Atherosclerosis | A05.810.453.324.Kidney.Cortex | Other | 1.85E-08 | 2.17E-08 | 0.196714 |
| Atherosclerosis | A05.810.453.Kidney | Other | -1.23E-09 | 1.99E-08 | 0.524613 |
| Atherosclerosis | A05.810.890.Urinary.Bladder | Other | 2.32E-08 | 2.69E-08 | 0.194114 |
| Atherosclerosis | A06.407.071.140.Adrenal.Cortex | Other | -5.93E-09 | 2.07E-08 | 0.612929 |
| Atherosclerosis | A06.407.071.Adrenal.Glands | Other | -1.37E-08 | 1.99E-08 | 0.753389 |
| Atherosclerosis | A06.407.312.782.Testis | Other | 3.53E-08 | 1.90E-08 | 0.031318 |
| Atherosclerosis | A06.407.312.Gonads | Other | -1.63E-08 | 2.30E-08 | 0.760349 |
| Atherosclerosis | A06.407.900.Thyroid.Gland | Other | -7.72E-09 | 1.90E-08 | 0.657542 |
| Atherosclerosis | A06.407.Endocrine.Glands | Other | -1.24E-08 | 2.27E-08 | 0.707214 |
| Atherosclerosis | A09.371.Eye | Other | -2.16E-08 | 2.39E-08 | 0.816985 |
| Atherosclerosis | A10.272.497.Epidermis | Other | 1.97E-08 | 2.29E-08 | 0.194600 |
| Atherosclerosis | A10.272.Epithelium | Other | 1.85E-08 | 2.31E-08 | 0.210786 |
| Atherosclerosis | A10.336.707.Prostate | Other | -1.72E-08 | 1.77E-08 | 0.833623 |
| Atherosclerosis | A10.615.284.473.Chorion | Other | 3.76E-08 | 2.11E-08 | 0.037103 |
| Atherosclerosis | A10.615.550.599.Mouth.Mucosa | Other | -6.61E-09 | 2.21E-08 | 0.617659 |
| Atherosclerosis | A10.615.550.Mucous.Membrane | Other | -4.68E-08 | 2.25E-08 | 0.981398 |
| Atherosclerosis | A10.615.789.Serous.Membrane | Other | -3.85E-09 | 2.36E-08 | 0.564677 |
| Atherosclerosis | A10.615.Membranes | Other | -3.85E-08 | 2.24E-08 | 0.957104 |
| Atherosclerosis | A11.382.Endocrine.Cells | Other | -4.49E-09 | 2.28E-08 | 0.578212 |
| Atherosclerosis | A11.436.275.Endothelial.Cells | Other | 1.54E-08 | 2.21E-08 | 0.242403 |
| Atherosclerosis | A11.436.294.064.Glucagon.Secreting.Cells | Other | 5.38E-09 | 2.19E-08 | 0.402820 |
| Atherosclerosis | A11.436.329.Granulosa.Cells | Other | 2.37E-09 | 2.09E-08 | 0.454802 |

| | | | | | |
|---|---|---|---|---|---|
| Atherosclerosis | A11.436.397.Keratinocytes | Other | 4.66E-08 | 2.53E-08 | 0.032969 |
| Atherosclerosis | A11.436.Epithelial.Cells | Other | 9.70E-09 | 2.38E-08 | 0.341796 |
| Atherosclerosis | A11.497.497.600.Oocytes | Other | 5.47E-08 | 2.11E-08 | 0.004813 |
| Atherosclerosis | A11.872.040.Adult.Stem.Cells | Other | 4.20E-09 | 2.19E-08 | 0.424001 |
| Atherosclerosis | A11.872.190.260.Embryoid.Bodies | Other | -4.32E-08 | 2.16E-08 | 0.977442 |
| Atherosclerosis | A11.872.190.Embryonic.Stem.Cells | Other | 8.40E-09 | 2.31E-08 | 0.358052 |
| Atherosclerosis | A11.872.580.Mesenchymal.Stem.Cells | Other | -7.64E-09 | 2.12E-08 | 0.640876 |
| Atherosclerosis | A11.872.700.500.Induced.Pluripotent.Stem.Cells | Other | 4.27E-09 | 2.30E-08 | 0.426273 |
| Atherosclerosis | A11.872.Stem.Cells | Other | -5.19E-09 | 2.54E-08 | 0.580970 |
| Atherosclerosis | A14.549.167.646.Periodontium | Other | 3.30E-08 | 2.57E-08 | 0.099793 |
| Atherosclerosis | A14.549.167.Dentition | Other | -1.08E-08 | 2.47E-08 | 0.669587 |
| Atherosclerosis | A14.549.885.Tongue | Other | 5.82E-08 | 2.46E-08 | 0.008895 |
| Atherosclerosis | A14.549.Mouth | Other | 3.17E-09 | 2.32E-08 | 0.445755 |
| Atherosclerosis | A14.724.557.Nasopharynx | Other | -9.83E-09 | 2.03E-08 | 0.685564 |
| Atherosclerosis | A14.724.Pharynx | Other | -2.55E-09 | 2.06E-08 | 0.549321 |
| Atherosclerosis | A17.815.Skin | Other | -2.15E-08 | 2.10E-08 | 0.846722 |
| Atherosclerosis | Adrenal_Gland | Other | -3.96E-08 | 1.90E-08 | 0.981453 |
| Atherosclerosis | Bladder | Other | 7.82E-09 | 2.22E-08 | 0.362037 |
| Atherosclerosis | Breast_Mammary_Tissue | Other | -3.94E-08 | 1.98E-08 | 0.976438 |
| Atherosclerosis | Cervix_Ectocervix | Other | 3.22E-08 | 2.03E-08 | 0.056695 |
| Atherosclerosis | Cervix_Endocervix | Other | 4.39E-08 | 2.00E-08 | 0.014063 |
| Atherosclerosis | Fallopian_Tube | Other | 8.37E-09 | 2.06E-08 | 0.341973 |
| Atherosclerosis | Kidney_Cortex | Other | -2.08E-08 | 1.99E-08 | 0.852067 |
| Atherosclerosis | Lung | Other | 2.76E-09 | 2.23E-08 | 0.450692 |
| Atherosclerosis | Minor_Salivary_Gland | Other | -6.96E-09 | 2.03E-08 | 0.633842 |
| Atherosclerosis | Nerve_Tibial | Other | 6.82E-09 | 1.84E-08 | 0.355415 |
| Atherosclerosis | Ovary | Other | 3.07E-08 | 1.98E-08 | 0.060026 |
| Atherosclerosis | Pituitary | Other | -1.97E-08 | 1.97E-08 | 0.842363 |
| Atherosclerosis | Prostate | Other | 1.59E-08 | 2.26E-08 | 0.240668 |
| Atherosclerosis | Skin_Not_Sun_Exposed_(Suprapubic) | Other | 4.65E-09 | 2.08E-08 | 0.411736 |
| Atherosclerosis | Skin_Sun_Exposed_(Lower_leg) | Other | 3.83E-09 | 2.25E-08 | 0.432223 |
| Atherosclerosis | Testis | Other | 1.08E-08 | 2.25E-08 | 0.315966 |
| Atherosclerosis | Thyroid | Other | 2.59E-08 | 1.90E-08 | 0.086093 |
| Atherosclerosis | Uterus | Other | 3.96E-08 | 2.19E-08 | 0.034884 |
| Atherosclerosis | Vagina | Other | 1.28E-08 | 2.02E-08 | 0.263653 |
| Atherosclerosis | A03.734.414.Islets.of.Langerhans | Pancreas | 1.36E-09 | 2.13E-08 | 0.474603 |
| Atherosclerosis | A03.734.Pancreas | Pancreas | -1.83E-08 | 1.90E-08 | 0.832547 |
| Atherosclerosis | Pancreas | Pancreas | 1.55E-08 | 1.99E-08 | 0.217055 |
| Atherosclerosis (any/none) | A10.165.114.830.500.750.Subcutaneous.Fat..Abdominal | Adipose | -8.10E-09 | 2.07E-08 | 0.652303 |
| Atherosclerosis (any/none) | A10.165.114.830.750.Subcutaneous.Fat | Adipose | 4.14E-09 | 2.04E-08 | 0.419626 |
| Atherosclerosis (any/none) | A11.329.114.Adipocytes | Adipose | -9.97E-09 | 1.96E-08 | 0.694316 |
| Atherosclerosis (any/none) | Adipose_Subcutaneous | Adipose | -2.95E-08 | 1.91E-08 | 0.939315 |
| Atherosclerosis (any/none) | Adipose_Visceral_(Omentum) | Adipose | -1.19E-08 | 2.14E-08 | 0.711115 |
| Atherosclerosis (any/none) | A02.835.583.443.800.800.Synovial.Fluid | Blood/Immune | -1.16E-08 | 2.47E-08 | 0.681208 |
| Atherosclerosis (any/none) | A10.549.400.Lymph.Nodes | Blood/Immune | -8.96E-09 | 2.38E-08 | 0.646488 |
| Atherosclerosis (any/none) | A10.549.Lymphoid.Tissue | Blood/Immune | -1.60E-08 | 2.39E-08 | 0.747574 |

| | | | | | |
|---|---|---|---|---|---|
| Atherosclerosis (any/none) | A11.118.637.555.567.562.440.Precursor.Cells..B.Lymphoid | Blood/Immune | 7.48E-09 | 2.45E-08 | 0.380296 |
| Atherosclerosis (any/none) | A11.118.637.555.567.562.B.Lymphocytes | Blood/Immune | 2.93E-08 | 2.68E-08 | 0.137241 |
| Atherosclerosis (any/none) | A11.118.637.555.567.569.200.700.T.Lymphocytes..Regulatory | Blood/Immune | 1.03E-08 | 2.38E-08 | 0.332370 |
| Atherosclerosis (any/none) | A11.118.637.555.567.569.T.Lymphocytes | Blood/Immune | 1.71E-08 | 2.41E-08 | 0.239759 |
| Atherosclerosis (any/none) | A11.118.637.Leukocytes | Blood/Immune | -1.26E-08 | 2.59E-08 | 0.686256 |
| Atherosclerosis (any/none) | A11.329.372.600.Macrophages..Alveolar | Blood/Immune | 1.96E-09 | 2.09E-08 | 0.462718 |
| Atherosclerosis (any/none) | A11.443.Erythroid.Cells | Blood/Immune | -1.35E-08 | 2.05E-08 | 0.744232 |
| Atherosclerosis (any/none) | A11.627.340.360.Granulocyte.Precursor.Cells | Blood/Immune | 6.70E-08 | 2.09E-08 | 0.000683 |
| Atherosclerosis (any/none) | A11.627.624.249.Monocyte.Macrophage.Precursor.Cells | Blood/Immune | 1.77E-08 | 2.43E-08 | 0.232959 |
| Atherosclerosis (any/none) | A11.627.635.Myeloid.Progenitor.Cells | Blood/Immune | 5.70E-08 | 2.47E-08 | 0.010600 |
| Atherosclerosis (any/none) | A11.872.378.590.635.Granulocyte.Macrophage.Progenitor.Cells | Blood/Immune | 3.47E-08 | 2.43E-08 | 0.076201 |
| Atherosclerosis (any/none) | A11.872.378.590.817.Megakaryocyte.Erythroid.Progenitor.Cells | Blood/Immune | 4.38E-08 | 2.28E-08 | 0.027189 |
| Atherosclerosis (any/none) | A11.872.378.Hematopoietic.Stem.Cells | Blood/Immune | 4.96E-08 | 2.33E-08 | 0.016440 |
| Atherosclerosis (any/none) | A15.145.229.188.Blood.Platelets | Blood/Immune | -1.92E-08 | 2.17E-08 | 0.812331 |
| Atherosclerosis (any/none) | A15.145.229.637.555.567.562.725.Plasma.Cells | Blood/Immune | 1.19E-08 | 2.47E-08 | 0.315226 |
| Atherosclerosis (any/none) | A15.145.229.637.555.567.569.200.CD4.Positive.T.Lymphocytes | Blood/Immune | -7.11E-09 | 2.50E-08 | 0.611928 |
| Atherosclerosis (any/none) | A15.145.229.637.555.Leukocytes..Mononuclear | Blood/Immune | -2.99E-08 | 2.59E-08 | 0.876219 |
| Atherosclerosis (any/none) | A15.145.229.Blood.Cells | Blood/Immune | -4.00E-09 | 2.55E-08 | 0.562284 |
| Atherosclerosis (any/none) | A15.145.300.Fetal.Blood | Blood/Immune | 1.46E-08 | 2.74E-08 | 0.297338 |
| Atherosclerosis (any/none) | A15.145.846.Serum | Blood/Immune | 3.62E-08 | 2.46E-08 | 0.070272 |
| Atherosclerosis (any/none) | A15.145.Blood | Blood/Immune | -1.22E-08 | 2.40E-08 | 0.694757 |
| Atherosclerosis (any/none) | A15.378.316.580.Monocytes | Blood/Immune | 2.05E-08 | 2.71E-08 | 0.224694 |
| Atherosclerosis (any/none) | A15.378.316.Bone.Marrow.Cells | Blood/Immune | 1.14E-08 | 2.47E-08 | 0.322202 |
| Atherosclerosis (any/none) | A15.382.490.315.583.Neutrophils | Blood/Immune | 6.84E-09 | 2.29E-08 | 0.382837 |
| Atherosclerosis (any/none) | A15.382.490.555.567.537.Killer.Cells..Natural | Blood/Immune | 2.86E-08 | 2.35E-08 | 0.111223 |
| Atherosclerosis (any/none) | A15.382.490.555.567.622.Lymphocytes..Null | Blood/Immune | 2.77E-08 | 2.39E-08 | 0.123550 |
| Atherosclerosis (any/none) | A15.382.490.555.567.Lymphocytes | Blood/Immune | 1.34E-08 | 2.53E-08 | 0.297583 |
| Atherosclerosis (any/none) | A15.382.520.604.700.Spleen | Blood/Immune | -2.28E-08 | 2.66E-08 | 0.803670 |
| Atherosclerosis (any/none) | A15.382.520.604.800.Palatine.Tonsil | Blood/Immune | 7.35E-09 | 2.36E-08 | 0.377948 |
| Atherosclerosis (any/none) | A15.382.680.Phagocytes | Blood/Immune | -8.47E-09 | 2.35E-08 | 0.640633 |
| Atherosclerosis (any/none) | A15.382.812.260.Dendritic.Cells | Blood/Immune | 6.92E-09 | 2.60E-08 | 0.395173 |
| Atherosclerosis (any/none) | A15.382.812.522.Macrophages | Blood/Immune | 1.02E-08 | 2.27E-08 | 0.326539 |
| Atherosclerosis (any/none) | A15.382.812.Mononuclear.Phagocyte.System | Blood/Immune | 1.36E-08 | 2.60E-08 | 0.300736 |
| Atherosclerosis (any/none) | A15.382.Immune.System | Blood/Immune | -1.32E-08 | 2.56E-08 | 0.696619 |
| Atherosclerosis (any/none) | Cells_EBV-transformed_lymphocytes | Blood/Immune | 7.24E-09 | 2.18E-08 | 0.369901 |
| Atherosclerosis (any/none) | Spleen | Blood/Immune | 1.37E-08 | 2.55E-08 | 0.295992 |
| Atherosclerosis (any/none) | Whole_Blood | Blood/Immune | 1.94E-08 | 2.54E-08 | 0.222509 |
| Atherosclerosis (any/none) | A07.231.114.Arteries | Cardiovascular | -3.22E-08 | 2.15E-08 | 0.932366 |
| Atherosclerosis (any/none) | A07.231.908.670.874.Umbilical.Veins | Cardiovascular | 3.61E-08 | 2.51E-08 | 0.074952 |
| Atherosclerosis (any/none) | A07.231.908.Veins | Cardiovascular | 2.34E-08 | 2.45E-08 | 0.169203 |
| Atherosclerosis (any/none) | A07.231.Blood.Vessels | Cardiovascular | 6.64E-09 | 2.47E-08 | 0.393894 |
| Atherosclerosis (any/none) | A07.541.358.100.Atrial.Appendage | Cardiovascular | -1.13E-08 | 2.10E-08 | 0.704780 |
| Atherosclerosis (any/none) | A07.541.358.Heart.Atria | Cardiovascular | -1.80E-08 | 2.07E-08 | 0.806929 |
| Atherosclerosis (any/none) | A07.541.510.110.Aortic.Valve | Cardiovascular | -9.89E-09 | 1.90E-08 | 0.698969 |
| Atherosclerosis (any/none) | A07.541.560.Heart.Ventricles | Cardiovascular | -4.79E-09 | 2.30E-08 | 0.582654 |
| Atherosclerosis (any/none) | A07.541.Heart | Cardiovascular | -3.16E-09 | 2.24E-08 | 0.556158 |

| | | | | | |
|---|---|---|---|---|---|
| Atherosclerosis (any/none) | Artery_Aorta | Cardiovascular | 4.94E-09 | 1.83E-08 | 0.393443 |
| Atherosclerosis (any/none) | Artery_Coronary | Cardiovascular | 3.31E-09 | 1.78E-08 | 0.426159 |
| Atherosclerosis (any/none) | Artery_Tibial | Cardiovascular | 7.70E-09 | 1.80E-08 | 0.334655 |
| Atherosclerosis (any/none) | Heart_Atrial_Appendage | Cardiovascular | -9.02E-09 | 2.05E-08 | 0.670191 |
| Atherosclerosis (any/none) | Heart_Left_Ventricle | Cardiovascular | 1.86E-08 | 1.95E-08 | 0.170435 |
| Atherosclerosis (any/none) | A08.186.211.132.810.428.200.Cerebellum | CNS | -3.50E-09 | 1.97E-08 | 0.570654 |
| Atherosclerosis (any/none) | A08.186.211.132.Brain.Stem | CNS | -1.79E-08 | 2.03E-08 | 0.811207 |
| Atherosclerosis (any/none) | A08.186.211.464.405.Hippocampus | CNS | -1.42E-08 | 2.05E-08 | 0.755778 |
| Atherosclerosis (any/none) | A08.186.211.464.710.225.Entorhinal.Cortex | CNS | -1.88E-08 | 2.13E-08 | 0.812101 |
| Atherosclerosis (any/none) | A08.186.211.464.Limbic.System | CNS | -2.26E-08 | 2.17E-08 | 0.850715 |
| Atherosclerosis (any/none) | A08.186.211.653.Mesencephalon | CNS | -9.65E-10 | 2.24E-08 | 0.517190 |
| Atherosclerosis (any/none) | A08.186.211.730.317.357.352.435.Hypothalamo.Hypophyseal.System | CNS | -2.60E-08 | 2.44E-08 | 0.856899 |
| Atherosclerosis (any/none) | A08.186.211.730.317.357.Hypothalamus | CNS | -2.88E-08 | 2.58E-08 | 0.867914 |
| Atherosclerosis (any/none) | A08.186.211.730.317.Diencephalon | CNS | 1.72E-08 | 2.56E-08 | 0.251185 |
| Atherosclerosis (any/none) | A08.186.211.730.885.287.249.487.Corpus.Striatum | CNS | 2.26E-08 | 1.96E-08 | 0.124879 |
| Atherosclerosis (any/none) | A08.186.211.730.885.287.249.Basal.Ganglia | CNS | 7.67E-09 | 2.17E-08 | 0.361657 |
| Atherosclerosis (any/none) | A08.186.211.730.885.287.500.270.Frontal.Lobe | CNS | 2.72E-09 | 2.21E-08 | 0.451068 |
| Atherosclerosis (any/none) | A08.186.211.730.885.287.500.571.735.Visual.Cortex | CNS | 1.44E-08 | 2.22E-08 | 0.258449 |
| Atherosclerosis (any/none) | A08.186.211.730.885.287.500.670.Parietal.Lobe | CNS | 1.55E-08 | 2.29E-08 | 0.249271 |
| Atherosclerosis (any/none) | A08.186.211.730.885.287.500.Cerebral.Cortex | CNS | -1.11E-08 | 2.18E-08 | 0.694552 |
| Atherosclerosis (any/none) | A08.186.211.865.428.Metencephalon | CNS | -5.47E-09 | 1.95E-08 | 0.610188 |
| Atherosclerosis (any/none) | A08.186.211.Brain | CNS | -2.41E-08 | 2.21E-08 | 0.862023 |
| Atherosclerosis (any/none) | A09.371.729.Retina | CNS | 2.41E-08 | 2.14E-08 | 0.129773 |
| Atherosclerosis (any/none) | A11.872.653.Neural.Stem.Cells | CNS | -5.79E-08 | 2.11E-08 | 0.996958 |
| Atherosclerosis (any/none) | Brain_Amygdala | CNS | 1.41E-08 | 1.85E-08 | 0.223231 |
| Atherosclerosis (any/none) | Brain_Anterior_cingulate_cortex_(BA24) | CNS | 8.37E-09 | 1.81E-08 | 0.321567 |
| Atherosclerosis (any/none) | Brain_Caudate_(basal_ganglia) | CNS | 6.19E-10 | 1.97E-08 | 0.487481 |
| Atherosclerosis (any/none) | Brain_Cerebellar_Hemisphere | CNS | 2.02E-08 | 1.84E-08 | 0.136137 |
| Atherosclerosis (any/none) | Brain_Cerebellum | CNS | 1.49E-08 | 1.84E-08 | 0.208513 |
| Atherosclerosis (any/none) | Brain_Cortex | CNS | 1.41E-08 | 1.84E-08 | 0.222057 |
| Atherosclerosis (any/none) | Brain_Frontal_Cortex_(BA9) | CNS | 1.72E-08 | 1.74E-08 | 0.161275 |
| Atherosclerosis (any/none) | Brain_Hippocampus | CNS | 4.96E-09 | 1.81E-08 | 0.392154 |
| Atherosclerosis (any/none) | Brain_Hypothalamus | CNS | 6.34E-09 | 1.90E-08 | 0.369137 |
| Atherosclerosis (any/none) | Brain_Nucleus_accumbens_(basal_ganglia) | CNS | 4.16E-09 | 1.84E-08 | 0.410551 |
| Atherosclerosis (any/none) | Brain_Putamen_(basal_ganglia) | CNS | 2.03E-09 | 1.96E-08 | 0.458905 |
| Atherosclerosis (any/none) | Brain_Spinal_cord_(cervical_c-1) | CNS | 2.97E-08 | 1.91E-08 | 0.060056 |
| Atherosclerosis (any/none) | Brain_Substantia_nigra | CNS | 2.02E-08 | 1.88E-08 | 0.140576 |
| Atherosclerosis (any/none) | A03.556.124.369.Intestinal.Mucosa | Digestive | -1.63E-09 | 2.31E-08 | 0.528129 |
| Atherosclerosis (any/none) | A03.556.124.526.767.Rectum | Digestive | 1.27E-08 | 2.49E-08 | 0.305103 |
| Atherosclerosis (any/none) | A03.556.124.684.Intestine..Small | Digestive | 1.96E-08 | 2.37E-08 | 0.203935 |
| Atherosclerosis (any/none) | A03.556.124.Intestines | Digestive | 2.45E-08 | 2.43E-08 | 0.157248 |
| Atherosclerosis (any/none) | A03.556.249.124.Ileum | Digestive | 2.22E-08 | 2.31E-08 | 0.168618 |
| Atherosclerosis (any/none) | A03.556.249.249.209.Cecum | Digestive | 2.14E-08 | 2.45E-08 | 0.191401 |
| Atherosclerosis (any/none) | A03.556.249.249.356.668.Colon..Sigmoid | Digestive | 2.39E-08 | 2.36E-08 | 0.154922 |
| Atherosclerosis (any/none) | A03.556.249.249.356.Colon | Digestive | 2.30E-08 | 2.37E-08 | 0.165239 |
| Atherosclerosis (any/none) | A03.556.500.760.464.Parotid.Gland | Digestive | -1.26E-08 | 2.33E-08 | 0.705759 |

| | | | | | |
|---|---|---|---|---|---|
| Atherosclerosis (any/none) | A03.556.500.760.Salivary.Glands | Digestive | -1.39E-08 | 2.37E-08 | 0.721256 |
| Atherosclerosis (any/none) | A03.556.875.500.Esophagus | Digestive | -5.82E-09 | 2.28E-08 | 0.600577 |
| Atherosclerosis (any/none) | A03.556.875.875.Stomach | Digestive | -9.09E-09 | 2.73E-08 | 0.630251 |
| Atherosclerosis (any/none) | A03.556.875.Upper.Gastrointestinal.Tract | Digestive | -3.98E-09 | 2.65E-08 | 0.559600 |
| Atherosclerosis (any/none) | A03.556.Gastrointestinal.Tract | Digestive | 8.36E-09 | 2.49E-08 | 0.368397 |
| Atherosclerosis (any/none) | Colon_Sigmoid | Digestive | -1.30E-09 | 1.90E-08 | 0.527224 |
| Atherosclerosis (any/none) | Colon_Transverse | Digestive | -3.47E-08 | 2.27E-08 | 0.937171 |
| Atherosclerosis (any/none) | Esophagus_Gastroesophageal_Junction | Digestive | 3.35E-09 | 2.03E-08 | 0.434544 |
| Atherosclerosis (any/none) | Esophagus_Mucosa | Digestive | -4.15E-08 | 1.93E-08 | 0.984087 |
| Atherosclerosis (any/none) | Esophagus_Muscularis | Digestive | 2.26E-08 | 2.23E-08 | 0.155323 |
| Atherosclerosis (any/none) | Small_Intestine_Terminal_Ileum | Digestive | -1.72E-08 | 2.15E-08 | 0.788500 |
| Atherosclerosis (any/none) | Stomach | Digestive | 1.62E-10 | 1.93E-08 | 0.496652 |
| Atherosclerosis (any/none) | A03.620.Liver | Liver | 7.72E-10 | 2.52E-08 | 0.487792 |
| Atherosclerosis (any/none) | A11.436.348.Hepatocytes | Liver | 2.95E-08 | 2.68E-08 | 0.135711 |
| Atherosclerosis (any/none) | Liver | Liver | 3.64E-09 | 2.29E-08 | 0.436988 |
| Atherosclerosis (any/none) | A02.165.Cartilage | Musculoskeletal/Connective | 3.95E-08 | 2.20E-08 | 0.036047 |
| Atherosclerosis (any/none) | A02.633.567.850.Quadriceps.Muscle | Musculoskeletal/Connective | 1.90E-09 | 2.51E-08 | 0.469941 |
| Atherosclerosis (any/none) | A02.835.232.834.151.Cervical.Vertebrae | Musculoskeletal/Connective | -5.17E-08 | 2.23E-08 | 0.989833 |
| Atherosclerosis (any/none) | A02.835.583.443.800.Synovial.Membrane | Musculoskeletal/Connective | -1.81E-09 | 2.30E-08 | 0.531334 |
| Atherosclerosis (any/none) | A05.360.319.679.690.Myometrium | Musculoskeletal/Connective | 5.25E-09 | 2.20E-08 | 0.405508 |
| Atherosclerosis (any/none) | A10.165.450.300.425.Keloid | Musculoskeletal/Connective | -3.05E-08 | 2.10E-08 | 0.927186 |
| Atherosclerosis (any/none) | A10.165.450.300.Cicatrix | Musculoskeletal/Connective | -2.87E-08 | 2.08E-08 | 0.916309 |
| Atherosclerosis (any/none) | A10.690.467.Muscle..Smooth | Musculoskeletal/Connective | 1.84E-08 | 2.19E-08 | 0.199838 |
| Atherosclerosis (any/none) | A10.690.Muscles | Musculoskeletal/Connective | -8.24E-09 | 2.50E-08 | 0.629427 |
| Atherosclerosis (any/none) | A11.329.171.Chondrocytes | Musculoskeletal/Connective | -4.78E-09 | 2.11E-08 | 0.589366 |
| Atherosclerosis (any/none) | A11.329.228.Fibroblasts | Musculoskeletal/Connective | 1.41E-08 | 2.26E-08 | 0.266482 |
| Atherosclerosis (any/none) | A11.329.629.Osteoblasts | Musculoskeletal/Connective | -1.24E-08 | 2.17E-08 | 0.716329 |
| Atherosclerosis (any/none) | A11.329.830.Stromal.Cells | Musculoskeletal/Connective | 2.25E-08 | 2.14E-08 | 0.146869 |
| Atherosclerosis (any/none) | A11.329.Connective.Tissue.Cells | Musculoskeletal/Connective | 3.20E-08 | 2.31E-08 | 0.082892 |
| Atherosclerosis (any/none) | A11.620.520.Myocytes..Smooth.Muscle | Musculoskeletal/Connective | -1.42E-08 | 2.15E-08 | 0.745564 |
| Atherosclerosis (any/none) | Cells_Transformed_fibroblasts | Musculoskeletal/Connective | -1.69E-08 | 2.07E-08 | 0.792054 |
| Atherosclerosis (any/none) | Muscle_Skeletal | Musculoskeletal/Connective | 2.87E-08 | 2.17E-08 | 0.092572 |
| Atherosclerosis (any/none) | A04.411.Lung | Other | 3.48E-09 | 2.22E-08 | 0.437839 |
| Atherosclerosis (any/none) | A04.531.520.Nasal.Mucosa | Other | -2.26E-09 | 2.50E-08 | 0.536043 |
| Atherosclerosis (any/none) | A05.360.319.114.373.Fallopian.Tubes | Other | 3.94E-09 | 2.14E-08 | 0.426990 |
| Atherosclerosis (any/none) | A05.360.319.114.630.Ovary | Other | -7.24E-09 | 2.12E-08 | 0.633627 |
| Atherosclerosis (any/none) | A05.360.319.679.256.Cervix.Uteri | Other | 1.46E-08 | 2.35E-08 | 0.267641 |
| Atherosclerosis (any/none) | A05.360.319.679.490.Endometrium | Other | 7.50E-09 | 2.28E-08 | 0.371034 |
| Atherosclerosis (any/none) | A05.360.319.679.Uterus | Other | -1.94E-08 | 2.04E-08 | 0.829722 |
| Atherosclerosis (any/none) | A05.360.319.887.Vulva | Other | 4.12E-09 | 2.53E-08 | 0.435209 |
| Atherosclerosis (any/none) | A05.360.319.Genitalia..Female | Other | -8.48E-09 | 2.00E-08 | 0.664113 |
| Atherosclerosis (any/none) | A05.360.444.492.362.Foreskin | Other | -3.45E-09 | 2.24E-08 | 0.561347 |
| Atherosclerosis (any/none) | A05.360.444.Genitalia..Male | Other | -1.10E-08 | 1.90E-08 | 0.718513 |
| Atherosclerosis (any/none) | A05.360.490.Germ.Cells | Other | 4.07E-08 | 2.15E-08 | 0.028827 |
| Atherosclerosis (any/none) | A05.360.Genitalia | Other | -2.08E-08 | 2.18E-08 | 0.830714 |
| Atherosclerosis (any/none) | A05.810.453.324.Kidney.Cortex | Other | 7.75E-09 | 2.29E-08 | 0.367713 |

| | | | | | |
|---|---|---|---|---|---|
| Atherosclerosis (any/none) | A05.810.453.Kidney | Other | 3.65E-10 | 2.11E-08 | 0.493117 |
| Atherosclerosis (any/none) | A05.810.890.Urinary.Bladder | Other | -3.16E-08 | 2.30E-08 | 0.914977 |
| Atherosclerosis (any/none) | A06.407.071.140.Adrenal.Cortex | Other | -1.61E-08 | 2.17E-08 | 0.771221 |
| Atherosclerosis (any/none) | A06.407.071.Adrenal.Glands | Other | -2.62E-08 | 2.10E-08 | 0.893761 |
| Atherosclerosis (any/none) | A06.407.312.782.Testis | Other | 9.52E-09 | 2.08E-08 | 0.323752 |
| Atherosclerosis (any/none) | A06.407.312.Gonads | Other | -3.27E-09 | 2.01E-08 | 0.564712 |
| Atherosclerosis (any/none) | A06.407.900.Thyroid.Gland | Other | -8.36E-09 | 2.00E-08 | 0.662370 |
| Atherosclerosis (any/none) | A06.407.Endocrine.Glands | Other | 7.08E-09 | 2.16E-08 | 0.371765 |
| Atherosclerosis (any/none) | A09.371.Eye | Other | -6.25E-09 | 2.31E-08 | 0.606661 |
| Atherosclerosis (any/none) | A10.272.497.Epidermis | Other | 1.48E-08 | 2.51E-08 | 0.277506 |
| Atherosclerosis (any/none) | A10.272.Epithelium | Other | -1.08E-08 | 2.36E-08 | 0.676427 |
| Atherosclerosis (any/none) | A10.336.707.Prostate | Other | -1.14E-08 | 1.94E-08 | 0.721206 |
| Atherosclerosis (any/none) | A10.615.284.473.Chorion | Other | 2.01E-08 | 1.90E-08 | 0.144589 |
| Atherosclerosis (any/none) | A10.615.550.599.Mouth.Mucosa | Other | -7.75E-09 | 2.33E-08 | 0.630341 |
| Atherosclerosis (any/none) | A10.615.550.Mucous.Membrane | Other | -2.74E-08 | 2.28E-08 | 0.885720 |
| Atherosclerosis (any/none) | A10.615.789.Serous.Membrane | Other | -3.24E-09 | 2.39E-08 | 0.553865 |
| Atherosclerosis (any/none) | A10.615.Membranes | Other | -2.57E-08 | 2.28E-08 | 0.870079 |
| Atherosclerosis (any/none) | A11.382.Endocrine.Cells | Other | -2.26E-08 | 2.32E-08 | 0.834112 |
| Atherosclerosis (any/none) | A11.436.275.Endothelial.Cells | Other | 2.41E-08 | 2.42E-08 | 0.160247 |
| Atherosclerosis (any/none) | A11.436.294.064.Glucagon.Secreting.Cells | Other | 3.02E-08 | 2.17E-08 | 0.082099 |
| Atherosclerosis (any/none) | A11.436.329.Granulosa.Cells | Other | -6.21E-09 | 2.18E-08 | 0.612308 |
| Atherosclerosis (any/none) | A11.436.397.Keratinocytes | Other | -1.45E-08 | 2.50E-08 | 0.719183 |
| Atherosclerosis (any/none) | A11.436.Epithelial.Cells | Other | -7.69E-09 | 2.45E-08 | 0.623139 |
| Atherosclerosis (any/none) | A11.497.497.600.Oocytes | Other | 2.90E-08 | 2.15E-08 | 0.088565 |
| Atherosclerosis (any/none) | A11.872.040.Adult.Stem.Cells | Other | -1.75E-08 | 2.32E-08 | 0.774788 |
| Atherosclerosis (any/none) | A11.872.190.260.Embryoid.Bodies | Other | -2.96E-08 | 2.41E-08 | 0.890794 |
| Atherosclerosis (any/none) | A11.872.190.Embryonic.Stem.Cells | Other | -1.05E-08 | 2.25E-08 | 0.679309 |
| Atherosclerosis (any/none) | A11.872.580.Mesenchymal.Stem.Cells | Other | -1.88E-08 | 2.07E-08 | 0.818107 |
| Atherosclerosis (any/none) | A11.872.700.500.Induced.Pluripotent.Stem.Cells | Other | -2.87E-08 | 2.44E-08 | 0.879934 |
| Atherosclerosis (any/none) | A11.872.Stem.Cells | Other | 2.08E-09 | 2.43E-08 | 0.465928 |
| Atherosclerosis (any/none) | A14.549.167.646.Periodontium | Other | 3.01E-09 | 2.82E-08 | 0.457483 |
| Atherosclerosis (any/none) | A14.549.167.Dentition | Other | -3.80E-08 | 2.57E-08 | 0.930449 |
| Atherosclerosis (any/none) | A14.549.885.Tongue | Other | 2.62E-09 | 2.29E-08 | 0.454384 |
| Atherosclerosis (any/none) | A14.549.Mouth | Other | -1.73E-08 | 2.41E-08 | 0.763439 |
| Atherosclerosis (any/none) | A14.724.557.Nasopharynx | Other | -3.13E-08 | 2.16E-08 | 0.926232 |
| Atherosclerosis (any/none) | A14.724.Pharynx | Other | -3.86E-08 | 2.16E-08 | 0.962793 |
| Atherosclerosis (any/none) | A17.815.Skin | Other | -4.10E-08 | 2.25E-08 | 0.965802 |
| Atherosclerosis (any/none) | Adrenal_Gland | Other | -1.63E-08 | 2.03E-08 | 0.788283 |
| Atherosclerosis (any/none) | Bladder | Other | -2.30E-08 | 2.11E-08 | 0.862375 |
| Atherosclerosis (any/none) | Breast_Mammary_Tissue | Other | -4.03E-08 | 1.97E-08 | 0.979780 |
| Atherosclerosis (any/none) | Cervix_Ectocervix | Other | -4.41E-10 | 2.15E-08 | 0.508188 |
| Atherosclerosis (any/none) | Cervix_Endocervix | Other | 5.27E-09 | 2.30E-08 | 0.409323 |
| Atherosclerosis (any/none) | Fallopian_Tube | Other | -1.59E-08 | 2.19E-08 | 0.766395 |
| Atherosclerosis (any/none) | Kidney_Cortex | Other | -1.63E-08 | 1.95E-08 | 0.798575 |
| Atherosclerosis (any/none) | Lung | Other | 1.17E-08 | 2.28E-08 | 0.304819 |
| Atherosclerosis (any/none) | Minor_Salivary_Gland | Other | 1.17E-08 | 1.96E-08 | 0.276007 |

| | | | | | |
|---|---|---|---|---|---|
| Atherosclerosis (any/none) | Nerve_Tibial | Other | 1.59E-08 | 1.82E-08 | 0.191172 |
| Atherosclerosis (any/none) | Ovary | Other | 1.02E-08 | 2.12E-08 | 0.316147 |
| Atherosclerosis (any/none) | Pituitary | Other | 2.87E-09 | 2.13E-08 | 0.446452 |
| Atherosclerosis (any/none) | Prostate | Other | 7.05E-10 | 2.22E-08 | 0.487311 |
| Atherosclerosis (any/none) | Skin_Not_Sun_Exposed_(Suprapubic) | Other | 8.79E-10 | 2.09E-08 | 0.483257 |
| Atherosclerosis (any/none) | Skin_Sun_Exposed_(Lower_leg) | Other | 5.47E-09 | 2.20E-08 | 0.401930 |
| Atherosclerosis (any/none) | Testis | Other | -5.93E-10 | 2.04E-08 | 0.511613 |
| Atherosclerosis (any/none) | Thyroid | Other | 8.18E-09 | 2.15E-08 | 0.351743 |
| Atherosclerosis (any/none) | Uterus | Other | 2.66E-08 | 2.14E-08 | 0.107031 |
| Atherosclerosis (any/none) | Vagina | Other | -1.52E-08 | 2.00E-08 | 0.777321 |
| Atherosclerosis (any/none) | A03.734.414.Islets.of.Langerhans | Pancreas | 2.40E-08 | 2.21E-08 | 0.138188 |
| Atherosclerosis (any/none) | A03.734.Pancreas | Pancreas | 1.77E-08 | 1.99E-08 | 0.186790 |
| Atherosclerosis (any/none) | Pancreas | Pancreas | 2.90E-08 | 1.78E-08 | 0.051778 |
| B Score | A10.165.114.830.500.750.Subcutaneous.Fat..Abdominal | Adipose | -8.23E-09 | 1.82E-08 | 0.674421 |
| B Score | A10.165.114.830.750.Subcutaneous.Fat | Adipose | -3.29E-10 | 1.83E-08 | 0.507163 |
| B Score | A11.329.114.Adipocytes | Adipose | 1.48E-08 | 1.65E-08 | 0.186174 |
| B Score | Adipose_Subcutaneous | Adipose | -2.78E-10 | 1.50E-08 | 0.507392 |
| B Score | Adipose_Visceral_(Omentum) | Adipose | -2.15E-08 | 1.62E-08 | 0.907930 |
| B Score | A02.835.583.443.800.800.Synovial.Fluid | Blood/Immune | 4.82E-09 | 2.13E-08 | 0.410517 |
| B Score | A10.549.400.Lymph.Nodes | Blood/Immune | 1.10E-08 | 1.93E-08 | 0.284127 |
| B Score | A10.549.Lymphoid.Tissue | Blood/Immune | 5.41E-09 | 1.92E-08 | 0.389012 |
| B Score | A11.118.637.555.567.562.440.Precursor.Cells..B.Lymphoid | Blood/Immune | -3.04E-09 | 1.98E-08 | 0.561069 |
| B Score | A11.118.637.555.567.562.B.Lymphocytes | Blood/Immune | -2.79E-08 | 1.94E-08 | 0.924741 |
| B Score | A11.118.637.555.567.569.200.700.T.Lymphocytes..Regulatory | Blood/Immune | -8.13E-09 | 1.81E-08 | 0.673745 |
| B Score | A11.118.637.555.567.569.T.Lymphocytes | Blood/Immune | 1.42E-08 | 1.89E-08 | 0.226087 |
| B Score | A11.118.637.Leukocytes | Blood/Immune | 2.06E-08 | 2.01E-08 | 0.153058 |
| B Score | A11.329.372.600.Macrophages..Alveolar | Blood/Immune | 1.05E-08 | 1.89E-08 | 0.289739 |
| B Score | A11.443.Erythroid.Cells | Blood/Immune | 1.50E-09 | 1.74E-08 | 0.465787 |
| B Score | A11.627.340.360.Granulocyte.Precursor.Cells | Blood/Immune | -1.33E-10 | 2.05E-08 | 0.502582 |
| B Score | A11.627.624.249.Monocyte.Macrophage.Precursor.Cells | Blood/Immune | 2.64E-09 | 1.68E-08 | 0.437583 |
| B Score | A11.627.635.Myeloid.Progenitor.Cells | Blood/Immune | 1.33E-08 | 1.84E-08 | 0.235017 |
| B Score | A11.872.378.590.635.Granulocyte.Macrophage.Progenitor.Cells | Blood/Immune | 9.45E-10 | 1.92E-08 | 0.480359 |
| B Score | A11.872.378.590.817.Megakaryocyte.Erythroid.Progenitor.Cells | Blood/Immune | 1.94E-08 | 1.88E-08 | 0.152102 |
| B Score | A11.872.378.Hematopoietic.Stem.Cells | Blood/Immune | -1.63E-08 | 1.76E-08 | 0.822684 |
| B Score | A15.145.229.188.Blood.Platelets | Blood/Immune | -1.14E-08 | 1.69E-08 | 0.750996 |
| B Score | A15.145.229.637.555.567.562.725.Plasma.Cells | Blood/Immune | -2.94E-08 | 1.85E-08 | 0.944148 |
| B Score | A15.145.229.637.555.567.569.200.CD4.Positive.T.Lymphocytes | Blood/Immune | -1.58E-08 | 1.77E-08 | 0.814467 |
| B Score | A15.145.229.637.555.Leukocytes..Mononuclear | Blood/Immune | 4.59E-09 | 1.89E-08 | 0.404333 |
| B Score | A15.145.229.Blood.Cells | Blood/Immune | 1.78E-09 | 2.01E-08 | 0.464594 |
| B Score | A15.145.300.Fetal.Blood | Blood/Immune | 7.82E-09 | 2.07E-08 | 0.352507 |
| B Score | A15.145.846.Serum | Blood/Immune | -2.90E-10 | 1.80E-08 | 0.506414 |
| B Score | A15.145.Blood | Blood/Immune | 1.39E-10 | 2.06E-08 | 0.497307 |
| B Score | A15.378.316.580.Monocytes | Blood/Immune | 3.46E-08 | 2.18E-08 | 0.056818 |
| B Score | A15.378.316.Bone.Marrow.Cells | Blood/Immune | 3.42E-08 | 2.08E-08 | 0.050206 |
| B Score | A15.382.490.315.583.Neutrophils | Blood/Immune | 1.16E-08 | 1.83E-08 | 0.263453 |
| B Score | A15.382.490.555.567.537.Killer.Cells..Natural | Blood/Immune | -1.18E-08 | 2.06E-08 | 0.716495 |

| | | | | | |
|---|---|---|---|---|---|
| B Score | A15.382.490.555.567.622.Lymphocytes..Null | Blood/Immune | -2.86E-08 | 1.85E-08 | 0.939441 |
| B Score | A15.382.490.555.567.Lymphocytes | Blood/Immune | -4.48E-10 | 1.81E-08 | 0.509855 |
| B Score | A15.382.520.604.700.Spleen | Blood/Immune | 1.49E-08 | 1.76E-08 | 0.199188 |
| B Score | A15.382.520.604.800.Palatine.Tonsil | Blood/Immune | 2.01E-08 | 1.84E-08 | 0.137839 |
| B Score | A15.382.680.Phagocytes | Blood/Immune | 2.62E-08 | 2.07E-08 | 0.103603 |
| B Score | A15.382.812.260.Dendritic.Cells | Blood/Immune | 4.72E-08 | 1.98E-08 | 0.008616 |
| B Score | A15.382.812.522.Macrophages | Blood/Immune | 5.03E-08 | 1.97E-08 | 0.005388 |
| B Score | A15.382.812.Mononuclear.Phagocyte.System | Blood/Immune | 2.95E-08 | 2.00E-08 | 0.069911 |
| B Score | A15.382.Immune.System | Blood/Immune | -1.76E-09 | 1.87E-08 | 0.537465 |
| B Score | Cells_EBV-transformed_lymphocytes | Blood/Immune | 3.14E-08 | 1.69E-08 | 0.031505 |
| B Score | Spleen | Blood/Immune | 2.11E-08 | 1.88E-08 | 0.130451 |
| B Score | Whole_Blood | Blood/Immune | -4.86E-09 | 2.12E-08 | 0.590447 |
| B Score | A07.231.114.Arteries | Cardiovascular | -4.64E-08 | 1.76E-08 | 0.995863 |
| B Score | A07.231.908.670.874.Umbilical.Veins | Cardiovascular | -3.64E-09 | 1.94E-08 | 0.574356 |
| B Score | A07.231.908.Veins | Cardiovascular | -1.00E-09 | 1.96E-08 | 0.520348 |
| B Score | A07.231.Blood.Vessels | Cardiovascular | 3.87E-10 | 1.79E-08 | 0.491403 |
| B Score | A07.541.358.100.Atrial.Appendage | Cardiovascular | -8.71E-09 | 1.62E-08 | 0.704169 |
| B Score | A07.541.358.Heart.Atria | Cardiovascular | -9.32E-09 | 1.59E-08 | 0.721218 |
| B Score | A07.541.510.110.Aortic.Valve | Cardiovascular | -2.37E-08 | 1.51E-08 | 0.941367 |
| B Score | A07.541.560.Heart.Ventricles | Cardiovascular | 5.95E-09 | 1.62E-08 | 0.356853 |
| B Score | A07.541.Heart | Cardiovascular | -1.74E-09 | 1.57E-08 | 0.544206 |
| B Score | Artery_Aorta | Cardiovascular | 6.28E-09 | 1.63E-08 | 0.350210 |
| B Score | Artery_Coronary | Cardiovascular | -1.34E-08 | 1.65E-08 | 0.790793 |
| B Score | Artery_Tibial | Cardiovascular | 1.23E-09 | 1.40E-08 | 0.465038 |
| B Score | Heart_Atrial_Appendage | Cardiovascular | 1.11E-08 | 1.56E-08 | 0.237982 |
| B Score | Heart_Left_Ventricle | Cardiovascular | 6.95E-09 | 1.56E-08 | 0.327901 |
| B Score | A08.186.211.132.810.428.200.Cerebellum | CNS | 2.19E-08 | 1.45E-08 | 0.065751 |
| B Score | A08.186.211.132.Brain.Stem | CNS | 2.25E-08 | 1.40E-08 | 0.054522 |
| B Score | A08.186.211.464.405.Hippocampus | CNS | -2.17E-08 | 1.50E-08 | 0.925189 |
| B Score | A08.186.211.464.710.225.Entorhinal.Cortex | CNS | -1.45E-08 | 1.52E-08 | 0.830127 |
| B Score | A08.186.211.464.Limbic.System | CNS | -1.13E-08 | 1.52E-08 | 0.771087 |
| B Score | A08.186.211.653.Mesencephalon | CNS | -1.57E-08 | 1.55E-08 | 0.845745 |
| B Score | A08.186.211.730.317.357.352.435.Hypothalamo.Hypophyseal.System | CNS | -2.67E-08 | 1.87E-08 | 0.923508 |
| B Score | A08.186.211.730.317.357.Hypothalamus | CNS | -2.94E-08 | 1.99E-08 | 0.930064 |
| B Score | A08.186.211.730.317.Diencephalon | CNS | -9.64E-09 | 2.07E-08 | 0.679041 |
| B Score | A08.186.211.730.885.287.249.487.Corpus.Striatum | CNS | -1.60E-08 | 1.47E-08 | 0.860416 |
| B Score | A08.186.211.730.885.287.249.Basal.Ganglia | CNS | 1.03E-08 | 1.67E-08 | 0.268811 |
| B Score | A08.186.211.730.885.287.500.270.Frontal.Lobe | CNS | -2.39E-08 | 1.55E-08 | 0.937966 |
| B Score | A08.186.211.730.885.287.500.571.735.Visual.Cortex | CNS | -1.69E-08 | 1.42E-08 | 0.883050 |
| B Score | A08.186.211.730.885.287.500.670.Parietal.Lobe | CNS | -1.11E-08 | 1.47E-08 | 0.776376 |
| B Score | A08.186.211.730.885.287.500.Cerebral.Cortex | CNS | -9.35E-09 | 1.52E-08 | 0.730113 |
| B Score | A08.186.211.865.428.Metencephalon | CNS | 1.69E-08 | 1.38E-08 | 0.110699 |
| B Score | A08.186.211.Brain | CNS | -1.13E-08 | 1.42E-08 | 0.787468 |
| B Score | A09.371.729.Retina | CNS | 9.09E-09 | 1.48E-08 | 0.269578 |
| B Score | A11.872.653.Neural.Stem.Cells | CNS | -1.00E-08 | 1.58E-08 | 0.736431 |
| B Score | Brain_Amygdala | CNS | -1.33E-08 | 1.39E-08 | 0.829462 |

| | | | | | |
|---|---|---|---|---|---|
| B Score | Brain_Anterior_cingulate_cortex_(BA24) | CNS | -3.36E-09 | 1.41E-08 | 0.594034 |
| B Score | Brain_Caudate_(basal_ganglia) | CNS | -9.98E-09 | 1.41E-08 | 0.759852 |
| B Score | Brain_Cerebellar_Hemisphere | CNS | 4.86E-09 | 1.55E-08 | 0.376646 |
| B Score | Brain_Cerebellum | CNS | -5.27E-09 | 1.48E-08 | 0.638844 |
| B Score | Brain_Cortex | CNS | -1.23E-09 | 1.37E-08 | 0.535637 |
| B Score | Brain_Frontal_Cortex_(BA9) | CNS | -1.69E-08 | 1.38E-08 | 0.889219 |
| B Score | Brain_Hippocampus | CNS | -1.77E-08 | 1.30E-08 | 0.912719 |
| B Score | Brain_Hypothalamus | CNS | -1.48E-08 | 1.35E-08 | 0.863934 |
| B Score | Brain_Nucleus_accumbens_(basal_ganglia) | CNS | -9.33E-09 | 1.40E-08 | 0.746643 |
| B Score | Brain_Putamen_(basal_ganglia) | CNS | -7.82E-09 | 1.38E-08 | 0.713970 |
| B Score | Brain_Spinal_cord_(cervical_c-1) | CNS | -9.24E-09 | 1.30E-08 | 0.761625 |
| B Score | Brain_Substantia_nigra | CNS | -7.64E-09 | 1.26E-08 | 0.727406 |
| B Score | A03.556.124.369.Intestinal.Mucosa | Digestive | -1.34E-08 | 1.88E-08 | 0.761958 |
| B Score | A03.556.124.526.767.Rectum | Digestive | -2.99E-08 | 1.91E-08 | 0.941382 |
| B Score | A03.556.124.684.Intestine..Small | Digestive | -2.98E-08 | 1.93E-08 | 0.938865 |
| B Score | A03.556.124.Intestines | Digestive | -1.55E-08 | 1.93E-08 | 0.789070 |
| B Score | A03.556.249.124.Ileum | Digestive | -5.86E-08 | 2.01E-08 | 0.998256 |
| B Score | A03.556.249.249.209.Cecum | Digestive | -1.47E-08 | 1.76E-08 | 0.797627 |
| B Score | A03.556.249.249.356.668.Colon..Sigmoid | Digestive | -3.25E-08 | 2.05E-08 | 0.943861 |
| B Score | A03.556.249.249.356.Colon | Digestive | -2.75E-08 | 1.98E-08 | 0.917904 |
| B Score | A03.556.500.760.464.Parotid.Gland | Digestive | -2.00E-09 | 1.89E-08 | 0.542022 |
| B Score | A03.556.500.760.Salivary.Glands | Digestive | -1.11E-09 | 1.85E-08 | 0.524010 |
| B Score | A03.556.875.500.Esophagus | Digestive | -1.61E-08 | 1.64E-08 | 0.836439 |
| B Score | A03.556.875.875.Stomach | Digestive | -2.73E-08 | 1.93E-08 | 0.921384 |
| B Score | A03.556.875.Upper.Gastrointestinal.Tract | Digestive | -2.07E-08 | 1.89E-08 | 0.864315 |
| B Score | A03.556.Gastrointestinal.Tract | Digestive | -2.80E-08 | 1.88E-08 | 0.932159 |
| B Score | Colon_Sigmoid | Digestive | -6.11E-09 | 1.56E-08 | 0.652615 |
| B Score | Colon_Transverse | Digestive | -1.73E-08 | 1.77E-08 | 0.835678 |
| B Score | Esophagus_Gastroesophageal_Junction | Digestive | -3.72E-08 | 1.55E-08 | 0.991716 |
| B Score | Esophagus_Mucosa | Digestive | -1.21E-08 | 1.70E-08 | 0.762029 |
| B Score | Esophagus_Muscularis | Digestive | -3.76E-08 | 1.56E-08 | 0.992113 |
| B Score | Small_Intestine_Terminal_Ileum | Digestive | -1.46E-08 | 1.76E-08 | 0.795563 |
| B Score | Stomach | Digestive | 5.39E-09 | 1.88E-08 | 0.387424 |
| B Score | A03.620.Liver | Liver | -1.09E-08 | 1.73E-08 | 0.735190 |
| B Score | A11.436.348.Hepatocytes | Liver | 7.18E-09 | 1.98E-08 | 0.358179 |
| B Score | Liver | Liver | -2.08E-08 | 1.67E-08 | 0.893335 |
| B Score | A02.165.Cartilage | Musculoskeletal/Connective | -2.95E-09 | 1.49E-08 | 0.578625 |
| B Score | A02.633.567.850.Quadriceps.Muscle | Musculoskeletal/Connective | 1.19E-08 | 1.90E-08 | 0.265204 |
| B Score | A02.835.232.834.151.Cervical.Vertebrae | Musculoskeletal/Connective | 2.91E-08 | 1.57E-08 | 0.031889 |
| B Score | A02.835.583.443.800.Synovial.Membrane | Musculoskeletal/Connective | 7.16E-09 | 1.61E-08 | 0.328664 |
| B Score | A05.360.319.679.690.Myometrium | Musculoskeletal/Connective | -2.41E-08 | 1.40E-08 | 0.957515 |
| B Score | A10.165.450.300.425.Keloid | Musculoskeletal/Connective | 8.56E-09 | 1.54E-08 | 0.288987 |
| B Score | A10.165.450.300.Cicatrix | Musculoskeletal/Connective | 4.53E-09 | 1.51E-08 | 0.382116 |
| B Score | A10.690.467.Muscle..Smooth | Musculoskeletal/Connective | 2.30E-08 | 1.79E-08 | 0.099488 |
| B Score | A10.690.Muscles | Musculoskeletal/Connective | 1.43E-08 | 1.91E-08 | 0.227629 |
| B Score | A11.329.171.Chondrocytes | Musculoskeletal/Connective | 2.18E-08 | 1.59E-08 | 0.084693 |

| | | | | | |
|---|---|---|---|---|---|
| B Score | A11.329.228.Fibroblasts | Musculoskeletal/Connective | 7.11E-09 | 1.62E-08 | 0.330277 |
| B Score | A11.329.629.Osteoblasts | Musculoskeletal/Connective | 7.33E-09 | 1.65E-08 | 0.328385 |
| B Score | A11.329.830.Stromal.Cells | Musculoskeletal/Connective | 2.62E-08 | 1.65E-08 | 0.055502 |
| B Score | A11.329.Connective.Tissue.Cells | Musculoskeletal/Connective | 2.13E-08 | 1.84E-08 | 0.123428 |
| B Score | A11.620.520.Myocytes..Smooth.Muscle | Musculoskeletal/Connective | 2.29E-08 | 1.79E-08 | 0.099538 |
| B Score | Cells_Transformed_fibroblasts | Musculoskeletal/Connective | 1.37E-08 | 1.50E-08 | 0.181560 |
| B Score | Muscle_Skeletal | Musculoskeletal/Connective | -2.26E-08 | 1.58E-08 | 0.924058 |
| B Score | A04.411.Lung | Other | -3.33E-08 | 1.75E-08 | 0.971594 |
| B Score | A04.531.520.Nasal.Mucosa | Other | -4.23E-08 | 1.56E-08 | 0.996657 |
| B Score | A05.360.319.114.373.Fallopian.Tubes | Other | 1.98E-08 | 1.79E-08 | 0.133569 |
| B Score | A05.360.319.114.630.Ovary | Other | 6.56E-09 | 1.69E-08 | 0.348635 |
| B Score | A05.360.319.679.256.Cervix.Uteri | Other | -4.62E-08 | 1.86E-08 | 0.993563 |
| B Score | A05.360.319.679.490.Endometrium | Other | -7.25E-09 | 1.66E-08 | 0.669064 |
| B Score | A05.360.319.679.Uterus | Other | -1.71E-08 | 1.55E-08 | 0.865798 |
| B Score | A05.360.319.887.Vulva | Other | -5.05E-09 | 1.66E-08 | 0.619408 |
| B Score | A05.360.319.Genitalia..Female | Other | -1.36E-08 | 1.61E-08 | 0.800219 |
| B Score | A05.360.444.492.362.Foreskin | Other | 2.57E-08 | 1.50E-08 | 0.043455 |
| B Score | A05.360.444.Genitalia..Male | Other | -9.16E-09 | 1.59E-08 | 0.717215 |
| B Score | A05.360.490.Germ.Cells | Other | -8.77E-10 | 1.48E-08 | 0.523560 |
| B Score | A05.360.Genitalia | Other | -1.10E-08 | 1.64E-08 | 0.748718 |
| B Score | A05.810.453.324.Kidney.Cortex | Other | 2.00E-08 | 1.55E-08 | 0.099210 |
| B Score | A05.810.453.Kidney | Other | 3.79E-09 | 1.67E-08 | 0.409987 |
| B Score | A05.810.890.Urinary.Bladder | Other | -2.98E-08 | 2.10E-08 | 0.922616 |
| B Score | A06.407.071.140.Adrenal.Cortex | Other | -1.64E-08 | 1.84E-08 | 0.813346 |
| B Score | A06.407.071.Adrenal.Glands | Other | -2.58E-08 | 1.73E-08 | 0.931634 |
| B Score | A06.407.312.782.Testis | Other | 2.97E-10 | 1.37E-08 | 0.491362 |
| B Score | A06.407.312.Gonads | Other | 5.88E-09 | 1.61E-08 | 0.357770 |
| B Score | A06.407.900.Thyroid.Gland | Other | 9.95E-10 | 1.57E-08 | 0.474699 |
| B Score | A06.407.Endocrine.Glands | Other | 9.20E-09 | 1.65E-08 | 0.288373 |
| B Score | A09.371.Eye | Other | 6.03E-09 | 1.76E-08 | 0.365685 |
| B Score | A10.272.497.Epidermis | Other | 1.65E-09 | 1.75E-08 | 0.462536 |
| B Score | A10.272.Epithelium | Other | -2.27E-08 | 1.61E-08 | 0.919983 |
| B Score | A10.336.707.Prostate | Other | -9.62E-09 | 1.60E-08 | 0.725761 |
| B Score | A10.615.284.473.Chorion | Other | 7.11E-09 | 1.84E-08 | 0.349496 |
| B Score | A10.615.550.599.Mouth.Mucosa | Other | -1.98E-08 | 1.79E-08 | 0.864933 |
| B Score | A10.615.550.Mucous.Membrane | Other | -2.88E-08 | 1.73E-08 | 0.951915 |
| B Score | A10.615.789.Serous.Membrane | Other | -1.11E-08 | 1.70E-08 | 0.743926 |
| B Score | A10.615.Membranes | Other | -2.50E-08 | 1.68E-08 | 0.931648 |
| B Score | A11.382.Endocrine.Cells | Other | 1.34E-08 | 1.67E-08 | 0.211141 |
| B Score | A11.436.275.Endothelial.Cells | Other | 3.71E-08 | 2.04E-08 | 0.034236 |
| B Score | A11.436.294.064.Glucagon.Secreting.Cells | Other | 1.18E-08 | 1.84E-08 | 0.260880 |
| B Score | A11.436.329.Granulosa.Cells | Other | -5.54E-09 | 1.62E-08 | 0.634249 |
| B Score | A11.436.397.Keratinocytes | Other | 4.80E-09 | 1.95E-08 | 0.402843 |
| B Score | A11.436.Epithelial.Cells | Other | -2.13E-08 | 1.71E-08 | 0.893435 |
| B Score | A11.497.497.600.Oocytes | Other | -4.63E-09 | 1.44E-08 | 0.626320 |
| B Score | A11.872.040.Adult.Stem.Cells | Other | -2.85E-08 | 1.69E-08 | 0.953851 |

| | | | | | |
|---|---|---|---|---|---|
| B Score | A11.872.190.260.Embryoid.Bodies | Other | -4.19E-09 | 1.70E-08 | 0.597221 |
| B Score | A11.872.190.Embryonic.Stem.Cells | Other | -8.68E-09 | 1.66E-08 | 0.698973 |
| B Score | A11.872.580.Mesenchymal.Stem.Cells | Other | 1.73E-08 | 1.56E-08 | 0.133523 |
| B Score | A11.872.700.500.Induced.Pluripotent.Stem.Cells | Other | -3.13E-08 | 1.78E-08 | 0.960027 |
| B Score | A11.872.Stem.Cells | Other | 2.23E-08 | 1.96E-08 | 0.128122 |
| B Score | A14.549.167.646.Periodontium | Other | -8.41E-09 | 1.75E-08 | 0.684101 |
| B Score | A14.549.167.Dentition | Other | -6.15E-09 | 1.66E-08 | 0.644431 |
| B Score | A14.549.885.Tongue | Other | -1.18E-08 | 1.75E-08 | 0.751012 |
| B Score | A14.549.Mouth | Other | -7.66E-09 | 1.67E-08 | 0.676489 |
| B Score | A14.724.557.Nasopharynx | Other | -1.76E-09 | 1.77E-08 | 0.539631 |
| B Score | A14.724.Pharynx | Other | -5.14E-10 | 1.86E-08 | 0.511059 |
| B Score | A17.815.Skin | Other | 2.80E-08 | 1.60E-08 | 0.039886 |
| B Score | Adrenal_Gland | Other | 1.34E-09 | 1.49E-08 | 0.464342 |
| B Score | Bladder | Other | -1.91E-08 | 1.46E-08 | 0.905152 |
| B Score | Breast_Mammary_Tissue | Other | 1.05E-08 | 1.67E-08 | 0.265758 |
| B Score | Cervix_Ectocervix | Other | 1.33E-08 | 1.70E-08 | 0.217220 |
| B Score | Cervix_Endocervix | Other | 1.37E-08 | 1.54E-08 | 0.186842 |
| B Score | Fallopian_Tube | Other | 5.26E-09 | 1.58E-08 | 0.369127 |
| B Score | Kidney_Cortex | Other | 3.85E-08 | 1.82E-08 | 0.017085 |
| B Score | Lung | Other | 2.23E-08 | 1.72E-08 | 0.097410 |
| B Score | Minor_Salivary_Gland | Other | 4.24E-11 | 1.61E-08 | 0.498950 |
| B Score | Nerve_Tibial | Other | 2.09E-08 | 1.63E-08 | 0.100633 |
| B Score | Ovary | Other | -4.18E-09 | 1.39E-08 | 0.617921 |
| B Score | Pituitary | Other | -2.14E-10 | 1.60E-08 | 0.505330 |
| B Score | Prostate | Other | -2.70E-08 | 1.78E-08 | 0.936001 |
| B Score | Skin_Not_Sun_Exposed_(Suprapubic) | Other | -1.20E-08 | 1.63E-08 | 0.768615 |
| B Score | Skin_Sun_Exposed_(Lower_leg) | Other | -1.33E-08 | 1.72E-08 | 0.781196 |
| B Score | Testis | Other | 6.02E-09 | 1.56E-08 | 0.349882 |
| B Score | Thyroid | Other | 1.76E-09 | 1.40E-08 | 0.450115 |
| B Score | Uterus | Other | 1.88E-09 | 1.57E-08 | 0.452283 |
| B Score | Vagina | Other | -9.33E-09 | 1.70E-08 | 0.708454 |
| B Score | A03.734.414.Islets.of.Langerhans | Pancreas | 3.07E-09 | 1.92E-08 | 0.436504 |
| B Score | A03.734.Pancreas | Pancreas | -3.34E-08 | 1.60E-08 | 0.981578 |
| B Score | Pancreas | Pancreas | -2.24E-08 | 1.65E-08 | 0.911985 |
| CAA | A10.165.114.830.500.750.Subcutaneous.Fat..Abdominal | Adipose | -2.85E-08 | 1.99E-08 | 0.923885 |
| CAA | A10.165.114.830.750.Subcutaneous.Fat | Adipose | -2.05E-08 | 1.99E-08 | 0.849105 |
| CAA | A11.329.114.Adipocytes | Adipose | 1.82E-08 | 2.30E-08 | 0.214436 |
| CAA | Adipose_Subcutaneous | Adipose | -2.00E-08 | 2.13E-08 | 0.826086 |
| CAA | Adipose_Visceral_(Omentum) | Adipose | -4.18E-08 | 2.21E-08 | 0.971002 |
| CAA | A02.835.583.443.800.800.Synovial.Fluid | Blood/Immune | -1.38E-08 | 2.65E-08 | 0.698615 |
| CAA | A10.549.400.Lymph.Nodes | Blood/Immune | -8.58E-09 | 2.23E-08 | 0.649960 |
| CAA | A10.549.Lymphoid.Tissue | Blood/Immune | -2.86E-09 | 2.17E-08 | 0.552525 |
| CAA | A11.118.637.555.567.562.440.Precursor.Cells..B.Lymphoid | Blood/Immune | 5.64E-09 | 2.35E-08 | 0.405290 |
| CAA | A11.118.637.555.567.562.B.Lymphocytes | Blood/Immune | 1.40E-08 | 2.55E-08 | 0.291789 |
| CAA | A11.118.637.555.567.569.200.700.T.Lymphocytes..Regulatory | Blood/Immune | -1.52E-08 | 2.40E-08 | 0.737238 |
| CAA | A11.118.637.555.567.569.T.Lymphocytes | Blood/Immune | -3.34E-09 | 2.50E-08 | 0.553158 |

| | | | | | |
|---|---|---|---|---|---|
| CAA | A11.118.637.Leukocytes | Blood/Immune | -3.84E-09 | 2.58E-08 | 0.559238 |
| CAA | A11.329.372.600.Macrophages..Alveolar | Blood/Immune | 1.35E-08 | 2.15E-08 | 0.265343 |
| CAA | A11.443.Erythroid.Cells | Blood/Immune | -1.62E-08 | 2.06E-08 | 0.784546 |
| CAA | A11.627.340.360.Granulocyte.Precursor.Cells | Blood/Immune | -4.63E-08 | 2.51E-08 | 0.967145 |
| CAA | A11.627.624.249.Monocyte.Macrophage.Precursor.Cells | Blood/Immune | -2.32E-08 | 2.54E-08 | 0.819484 |
| CAA | A11.627.635.Myeloid.Progenitor.Cells | Blood/Immune | -8.20E-09 | 2.58E-08 | 0.624522 |
| CAA | A11.872.378.590.635.Granulocyte.Macrophage.Progenitor.Cells | Blood/Immune | -1.73E-08 | 2.57E-08 | 0.749408 |
| CAA | A11.872.378.590.817.Megakaryocyte.Erythroid.Progenitor.Cells | Blood/Immune | -1.03E-10 | 2.31E-08 | 0.501790 |
| CAA | A11.872.378.Hematopoietic.Stem.Cells | Blood/Immune | -1.90E-08 | 2.43E-08 | 0.782201 |
| CAA | A15.145.229.188.Blood.Platelets | Blood/Immune | 1.44E-08 | 2.08E-08 | 0.244409 |
| CAA | A15.145.229.637.555.567.562.725.Plasma.Cells | Blood/Immune | 1.95E-08 | 2.48E-08 | 0.215593 |
| CAA | A15.145.229.637.555.567.569.200.CD4.Positive.T.Lymphocytes | Blood/Immune | -4.70E-10 | 2.47E-08 | 0.507586 |
| CAA | A15.145.229.637.555.Leukocytes..Mononuclear | Blood/Immune | -9.62E-10 | 2.49E-08 | 0.515411 |
| CAA | A15.145.229.Blood.Cells | Blood/Immune | -1.68E-08 | 2.68E-08 | 0.733696 |
| CAA | A15.145.300.Fetal.Blood | Blood/Immune | -3.22E-08 | 2.85E-08 | 0.870867 |
| CAA | A15.145.846.Serum | Blood/Immune | 2.46E-09 | 2.33E-08 | 0.458014 |
| CAA | A15.145.Blood | Blood/Immune | -2.36E-08 | 2.70E-08 | 0.809772 |
| CAA | A15.378.316.580.Monocytes | Blood/Immune | -3.15E-10 | 2.40E-08 | 0.505243 |
| CAA | A15.378.316.Bone.Marrow.Cells | Blood/Immune | -4.50E-09 | 2.42E-08 | 0.573681 |
| CAA | A15.382.490.315.583.Neutrophils | Blood/Immune | -3.45E-09 | 2.37E-08 | 0.557873 |
| CAA | A15.382.490.555.567.537.Killer.Cells..Natural | Blood/Immune | -2.68E-08 | 2.24E-08 | 0.884324 |
| CAA | A15.382.490.555.567.622.Lymphocytes..Null | Blood/Immune | -5.80E-09 | 2.45E-08 | 0.593534 |
| CAA | A15.382.490.555.567.Lymphocytes | Blood/Immune | 1.92E-08 | 2.40E-08 | 0.212079 |
| CAA | A15.382.520.604.700.Spleen | Blood/Immune | -1.71E-08 | 2.34E-08 | 0.767052 |
| CAA | A15.382.520.604.800.Palatine.Tonsil | Blood/Immune | -3.08E-09 | 2.16E-08 | 0.556740 |
| CAA | A15.382.680.Phagocytes | Blood/Immune | 3.95E-09 | 2.48E-08 | 0.436741 |
| CAA | A15.382.812.260.Dendritic.Cells | Blood/Immune | -1.95E-08 | 2.18E-08 | 0.815119 |
| CAA | A15.382.812.522.Macrophages | Blood/Immune | -1.95E-08 | 2.25E-08 | 0.807232 |
| CAA | A15.382.812.Mononuclear.Phagocyte.System | Blood/Immune | -1.42E-08 | 2.37E-08 | 0.725516 |
| CAA | A15.382.Immune.System | Blood/Immune | 5.87E-09 | 2.52E-08 | 0.408079 |
| CAA | Cells_EBV-transformed_lymphocytes | Blood/Immune | -1.63E-08 | 2.08E-08 | 0.783218 |
| CAA | Spleen | Blood/Immune | -9.13E-09 | 2.67E-08 | 0.633932 |
| CAA | Whole_Blood | Blood/Immune | 2.30E-10 | 2.41E-08 | 0.496196 |
| CAA | A07.231.114.Arteries | Cardiovascular | -1.81E-08 | 2.25E-08 | 0.789545 |
| CAA | A07.231.908.670.874.Umbilical.Veins | Cardiovascular | -3.84E-08 | 2.43E-08 | 0.943010 |
| CAA | A07.231.908.Veins | Cardiovascular | -3.40E-08 | 2.45E-08 | 0.917215 |
| CAA | A07.231.Blood.Vessels | Cardiovascular | -2.36E-08 | 2.45E-08 | 0.832722 |
| CAA | A07.541.358.100.Atrial.Appendage | Cardiovascular | 8.19E-09 | 2.03E-08 | 0.343234 |
| CAA | A07.541.358.Heart.Atria | Cardiovascular | 2.04E-08 | 2.06E-08 | 0.160958 |
| CAA | A07.541.510.110.Aortic.Valve | Cardiovascular | -2.71E-08 | 2.07E-08 | 0.904737 |
| CAA | A07.541.560.Heart.Ventricles | Cardiovascular | -5.10E-09 | 2.20E-08 | 0.591833 |
| CAA | A07.541.Heart | Cardiovascular | 1.20E-10 | 2.08E-08 | 0.497707 |
| CAA | Artery_Aorta | Cardiovascular | -3.82E-09 | 1.88E-08 | 0.580317 |
| CAA | Artery_Coronary | Cardiovascular | 2.20E-08 | 1.93E-08 | 0.126449 |
| CAA | Artery_Tibial | Cardiovascular | 3.98E-08 | 1.83E-08 | 0.014728 |
| CAA | Heart_Atrial_Appendage | Cardiovascular | -9.71E-10 | 1.91E-08 | 0.520230 |

| | | | | | |
|---|---|---|---|---|---|
| CAA | Heart_Left_Ventricle | Cardiovascular | -1.06E-08 | 1.98E-08 | 0.703809 |
| CAA | A08.186.211.132.810.428.200.Cerebellum | CNS | -1.28E-08 | 1.73E-08 | 0.771522 |
| CAA | A08.186.211.132.Brain.Stem | CNS | 1.20E-08 | 1.74E-08 | 0.244953 |
| CAA | A08.186.211.464.405.Hippocampus | CNS | 1.01E-08 | 1.91E-08 | 0.297554 |
| CAA | A08.186.211.464.710.225.Entorhinal.Cortex | CNS | 3.39E-08 | 1.90E-08 | 0.036985 |
| CAA | A08.186.211.464.Limbic.System | CNS | 3.45E-08 | 1.93E-08 | 0.037313 |
| CAA | A08.186.211.653.Mesencephalon | CNS | 7.07E-09 | 2.18E-08 | 0.372761 |
| CAA | A08.186.211.730.317.357.352.435.Hypothalamo.Hypophyseal.System | CNS | -7.35E-08 | 2.45E-08 | 0.998676 |
| CAA | A08.186.211.730.317.357.Hypothalamus | CNS | -6.64E-08 | 2.45E-08 | 0.996689 |
| CAA | A08.186.211.730.317.Diencephalon | CNS | 1.05E-08 | 2.43E-08 | 0.332988 |
| CAA | A08.186.211.730.885.287.249.487.Corpus.Striatum | CNS | -3.40E-08 | 2.07E-08 | 0.949916 |
| CAA | A08.186.211.730.885.287.249.Basal.Ganglia | CNS | -4.36E-09 | 2.11E-08 | 0.582007 |
| CAA | A08.186.211.730.885.287.500.270.Frontal.Lobe | CNS | 5.48E-09 | 2.19E-08 | 0.401180 |
| CAA | A08.186.211.730.885.287.500.571.735.Visual.Cortex | CNS | 1.05E-08 | 1.84E-08 | 0.284550 |
| CAA | A08.186.211.730.885.287.500.670.Parietal.Lobe | CNS | 1.81E-08 | 2.08E-08 | 0.191593 |
| CAA | A08.186.211.730.885.287.500.Cerebral.Cortex | CNS | 2.93E-08 | 1.88E-08 | 0.059784 |
| CAA | A08.186.211.865.428.Metencephalon | CNS | -1.37E-08 | 1.71E-08 | 0.788041 |
| CAA | A08.186.211.Brain | CNS | 1.44E-08 | 2.02E-08 | 0.238441 |
| CAA | A09.371.729.Retina | CNS | -1.93E-08 | 1.92E-08 | 0.842647 |
| CAA | A11.872.653.Neural.Stem.Cells | CNS | 1.93E-08 | 2.41E-08 | 0.211580 |
| CAA | Brain_Amygdala | CNS | 6.17E-09 | 2.01E-08 | 0.379480 |
| CAA | Brain_Anterior_cingulate_cortex_(BA24) | CNS | 1.08E-08 | 1.91E-08 | 0.285081 |
| CAA | Brain_Caudate_(basal_ganglia) | CNS | 1.51E-08 | 1.95E-08 | 0.219714 |
| CAA | Brain_Cerebellar_Hemisphere | CNS | 9.88E-09 | 1.84E-08 | 0.295639 |
| CAA | Brain_Cerebellum | CNS | 5.61E-09 | 1.88E-08 | 0.382740 |
| CAA | Brain_Cortex | CNS | 2.31E-08 | 2.01E-08 | 0.125773 |
| CAA | Brain_Frontal_Cortex_(BA9) | CNS | 9.52E-09 | 1.86E-08 | 0.304225 |
| CAA | Brain_Hippocampus | CNS | 9.07E-09 | 1.96E-08 | 0.321793 |
| CAA | Brain_Hypothalamus | CNS | 1.33E-08 | 2.01E-08 | 0.254251 |
| CAA | Brain_Nucleus_accumbens_(basal_ganglia) | CNS | 1.49E-08 | 1.90E-08 | 0.216064 |
| CAA | Brain_Putamen_(basal_ganglia) | CNS | 1.12E-08 | 1.84E-08 | 0.271188 |
| CAA | Brain_Spinal_cord_(cervical_c-1) | CNS | -1.37E-09 | 2.04E-08 | 0.526822 |
| CAA | Brain_Substantia_nigra | CNS | 8.27E-09 | 1.91E-08 | 0.332358 |
| CAA | A03.556.124.369.Intestinal.Mucosa | Digestive | 3.68E-08 | 2.23E-08 | 0.049309 |
| CAA | A03.556.124.526.767.Rectum | Digestive | 2.70E-08 | 2.60E-08 | 0.148964 |
| CAA | A03.556.124.684.Intestine..Small | Digestive | 3.33E-08 | 2.74E-08 | 0.112060 |
| CAA | A03.556.124.Intestines | Digestive | 3.80E-08 | 2.54E-08 | 0.066993 |
| CAA | A03.556.249.124.Ileum | Digestive | 6.31E-09 | 2.77E-08 | 0.410067 |
| CAA | A03.556.249.249.209.Cecum | Digestive | 3.15E-08 | 2.33E-08 | 0.088466 |
| CAA | A03.556.249.249.356.668.Colon..Sigmoid | Digestive | 3.23E-08 | 2.48E-08 | 0.096214 |
| CAA | A03.556.249.249.356.Colon | Digestive | 3.81E-08 | 2.56E-08 | 0.068162 |
| CAA | A03.556.500.760.464.Parotid.Gland | Digestive | -4.42E-08 | 2.32E-08 | 0.971618 |
| CAA | A03.556.500.760.Salivary.Glands | Digestive | -4.94E-08 | 2.21E-08 | 0.987292 |
| CAA | A03.556.875.500.Esophagus | Digestive | -1.02E-09 | 2.15E-08 | 0.518911 |
| CAA | A03.556.875.875.Stomach | Digestive | 5.37E-09 | 2.40E-08 | 0.411522 |
| CAA | A03.556.875.Upper.Gastrointestinal.Tract | Digestive | -5.45E-09 | 2.39E-08 | 0.590308 |

| | | | | | |
|---|---|---|---|---|---|
| CAA | A03.556.Gastrointestinal.Tract | Digestive | 2.49E-08 | 2.59E-08 | 0.168186 |
| CAA | Colon_Sigmoid | Digestive | -9.60E-09 | 1.96E-08 | 0.687614 |
| CAA | Colon_Transverse | Digestive | 2.22E-08 | 2.53E-08 | 0.190161 |
| CAA | Esophagus_Gastroesophageal_Junction | Digestive | 4.62E-09 | 1.99E-08 | 0.408280 |
| CAA | Esophagus_Mucosa | Digestive | -2.71E-08 | 2.03E-08 | 0.908988 |
| CAA | Esophagus_Muscularis | Digestive | 5.86E-09 | 2.04E-08 | 0.386824 |
| CAA | Small_Intestine_Terminal_Ileum | Digestive | 1.20E-08 | 2.62E-08 | 0.322819 |
| CAA | Stomach | Digestive | -3.34E-08 | 1.99E-08 | 0.953158 |
| CAA | A03.620.Liver | Liver | -5.12E-09 | 2.26E-08 | 0.589651 |
| CAA | A11.436.348.Hepatocytes | Liver | 4.66E-08 | 2.65E-08 | 0.039434 |
| CAA | Liver | Liver | -1.52E-09 | 2.29E-08 | 0.526480 |
| CAA | A02.165.Cartilage | Musculoskeletal/Connective | -1.29E-08 | 2.07E-08 | 0.733277 |
| CAA | A02.633.567.850.Quadriceps.Muscle | Musculoskeletal/Connective | -1.57E-08 | 2.33E-08 | 0.750011 |
| CAA | A02.835.232.834.151.Cervical.Vertebrae | Musculoskeletal/Connective | 2.92E-08 | 2.32E-08 | 0.104061 |
| CAA | A02.835.583.443.800.Synovial.Membrane | Musculoskeletal/Connective | -9.43E-09 | 2.02E-08 | 0.679914 |
| CAA | A05.360.319.679.690.Myometrium | Musculoskeletal/Connective | -1.74E-08 | 2.07E-08 | 0.799574 |
| CAA | A10.165.450.300.425.Keloid | Musculoskeletal/Connective | -1.95E-08 | 1.87E-08 | 0.851152 |
| CAA | A10.165.450.300.Cicatrix | Musculoskeletal/Connective | -9.83E-09 | 1.85E-08 | 0.702030 |
| CAA | A10.690.467.Muscle..Smooth | Musculoskeletal/Connective | 1.94E-08 | 2.20E-08 | 0.188952 |
| CAA | A10.690.Muscles | Musculoskeletal/Connective | -1.11E-08 | 2.33E-08 | 0.682893 |
| CAA | A11.329.171.Chondrocytes | Musculoskeletal/Connective | -4.12E-09 | 2.43E-08 | 0.567263 |
| CAA | A11.329.228.Fibroblasts | Musculoskeletal/Connective | -1.19E-08 | 1.96E-08 | 0.728380 |
| CAA | A11.329.629.Osteoblasts | Musculoskeletal/Connective | 2.16E-08 | 2.33E-08 | 0.176930 |
| CAA | A11.329.830.Stromal.Cells | Musculoskeletal/Connective | 6.02E-09 | 2.17E-08 | 0.390711 |
| CAA | A11.329.Connective.Tissue.Cells | Musculoskeletal/Connective | -1.63E-08 | 2.11E-08 | 0.780109 |
| CAA | A11.620.520.Myocytes..Smooth.Muscle | Musculoskeletal/Connective | -1.69E-08 | 2.18E-08 | 0.781123 |
| CAA | Cells_Transformed_fibroblasts | Musculoskeletal/Connective | -1.41E-08 | 2.01E-08 | 0.758349 |
| CAA | Muscle_Skeletal | Musculoskeletal/Connective | -1.89E-08 | 1.99E-08 | 0.829545 |
| CAA | A04.411.Lung | Other | -3.05E-09 | 2.18E-08 | 0.555644 |
| CAA | A04.531.520.Nasal.Mucosa | Other | -1.10E-08 | 2.16E-08 | 0.694662 |
| CAA | A05.360.319.114.373.Fallopian.Tubes | Other | -4.18E-09 | 2.46E-08 | 0.567548 |
| CAA | A05.360.319.114.630.Ovary | Other | -1.77E-08 | 2.31E-08 | 0.778495 |
| CAA | A05.360.319.679.256.Cervix.Uteri | Other | -2.14E-08 | 2.18E-08 | 0.836685 |
| CAA | A05.360.319.679.490.Endometrium | Other | -1.95E-08 | 2.24E-08 | 0.807754 |
| CAA | A05.360.319.679.Uterus | Other | -3.47E-09 | 2.32E-08 | 0.559313 |
| CAA | A05.360.319.887.Vulva | Other | -2.99E-09 | 2.10E-08 | 0.556633 |
| CAA | A05.360.319.Genitalia..Female | Other | -1.53E-08 | 2.39E-08 | 0.738964 |
| CAA | A05.360.444.492.362.Foreskin | Other | -2.15E-08 | 2.14E-08 | 0.842581 |
| CAA | A05.360.444.Genitalia..Male | Other | -4.22E-08 | 1.92E-08 | 0.986100 |
| CAA | A05.360.490.Germ.Cells | Other | 6.84E-09 | 1.91E-08 | 0.359848 |
| CAA | A05.360.Genitalia | Other | -2.82E-08 | 2.16E-08 | 0.903742 |
| CAA | A05.810.453.324.Kidney.Cortex | Other | 5.12E-08 | 2.18E-08 | 0.009471 |
| CAA | A05.810.453.Kidney | Other | 1.47E-08 | 2.09E-08 | 0.241979 |
| CAA | A05.810.890.Urinary.Bladder | Other | 2.34E-08 | 2.46E-08 | 0.170848 |
| CAA | A06.407.071.140.Adrenal.Cortex | Other | -4.37E-09 | 2.11E-08 | 0.581918 |
| CAA | A06.407.071.Adrenal.Glands | Other | -1.39E-08 | 2.19E-08 | 0.737450 |

| | | | | | |
|---|---|---|---|---|---|
| CAA | A06.407.312.782.Testis | Other | -4.20E-08 | 1.93E-08 | 0.985340 |
| CAA | A06.407.312.Gonads | Other | -1.33E-08 | 2.33E-08 | 0.715709 |
| CAA | A06.407.900.Thyroid.Gland | Other | -2.22E-08 | 1.97E-08 | 0.869846 |
| CAA | A06.407.Endocrine.Glands | Other | -9.00E-09 | 2.30E-08 | 0.652252 |
| CAA | A09.371.Eye | Other | 9.89E-09 | 2.29E-08 | 0.332585 |
| CAA | A10.272.497.Epidermis | Other | -1.48E-09 | 2.42E-08 | 0.524285 |
| CAA | A10.272.Epithelium | Other | -6.36E-09 | 2.23E-08 | 0.612015 |
| CAA | A10.336.707.Prostate | Other | -1.81E-08 | 1.91E-08 | 0.828279 |
| CAA | A10.615.284.473.Chorion | Other | -7.75E-09 | 2.13E-08 | 0.641628 |
| CAA | A10.615.550.599.Mouth.Mucosa | Other | -3.31E-09 | 2.18E-08 | 0.560287 |
| CAA | A10.615.550.Mucous.Membrane | Other | 1.62E-08 | 2.37E-08 | 0.246999 |
| CAA | A10.615.789.Serous.Membrane | Other | -9.96E-10 | 2.36E-08 | 0.516818 |
| CAA | A10.615.Membranes | Other | -4.62E-10 | 2.27E-08 | 0.508120 |
| CAA | A11.382.Endocrine.Cells | Other | 3.00E-08 | 1.93E-08 | 0.060058 |
| CAA | A11.436.275.Endothelial.Cells | Other | -2.86E-08 | 2.43E-08 | 0.880260 |
| CAA | A11.436.294.064.Glucagon.Secreting.Cells | Other | -2.27E-08 | 2.26E-08 | 0.842408 |
| CAA | A11.436.329.Granulosa.Cells | Other | 1.70E-08 | 1.86E-08 | 0.180247 |
| CAA | A11.436.397.Keratinocytes | Other | -2.91E-08 | 2.19E-08 | 0.908434 |
| CAA | A11.436.Epithelial.Cells | Other | -2.17E-08 | 2.36E-08 | 0.820364 |
| CAA | A11.497.497.600.Oocytes | Other | -6.76E-10 | 1.97E-08 | 0.513702 |
| CAA | A11.872.040.Adult.Stem.Cells | Other | -9.84E-09 | 2.30E-08 | 0.665859 |
| CAA | A11.872.190.260.Embryoid.Bodies | Other | -2.10E-08 | 2.13E-08 | 0.838308 |
| CAA | A11.872.190.Embryonic.Stem.Cells | Other | 2.56E-09 | 2.22E-08 | 0.454210 |
| CAA | A11.872.580.Mesenchymal.Stem.Cells | Other | -2.43E-09 | 2.22E-08 | 0.543480 |
| CAA | A11.872.700.500.Induced.Pluripotent.Stem.Cells | Other | -1.52E-08 | 2.31E-08 | 0.744352 |
| CAA | A11.872.Stem.Cells | Other | 2.46E-08 | 2.42E-08 | 0.154916 |
| CAA | A14.549.167.646.Periodontium | Other | -8.38E-09 | 2.40E-08 | 0.636621 |
| CAA | A14.549.167.Dentition | Other | -1.92E-08 | 2.46E-08 | 0.782418 |
| CAA | A14.549.885.Tongue | Other | -3.55E-08 | 2.26E-08 | 0.942240 |
| CAA | A14.549.Mouth | Other | -6.86E-09 | 2.19E-08 | 0.622748 |
| CAA | A14.724.557.Nasopharynx | Other | 1.90E-08 | 2.30E-08 | 0.204098 |
| CAA | A14.724.Pharynx | Other | -6.11E-09 | 2.14E-08 | 0.612422 |
| CAA | A17.815.Skin | Other | -1.21E-08 | 2.17E-08 | 0.712470 |
| CAA | Adrenal_Gland | Other | -4.41E-09 | 2.15E-08 | 0.581381 |
| CAA | Bladder | Other | -8.36E-09 | 2.05E-08 | 0.658418 |
| CAA | Breast_Mammary_Tissue | Other | -3.36E-08 | 1.97E-08 | 0.956287 |
| CAA | Cervix_Ectocervix | Other | -2.72E-08 | 1.98E-08 | 0.914748 |
| CAA | Cervix_Endocervix | Other | 8.19E-09 | 2.13E-08 | 0.350196 |
| CAA | Fallopian_Tube | Other | -2.72E-08 | 1.95E-08 | 0.918391 |
| CAA | Kidney_Cortex | Other | -1.24E-08 | 2.07E-08 | 0.724672 |
| CAA | Lung | Other | 1.61E-09 | 2.11E-08 | 0.469595 |
| CAA | Minor_Salivary_Gland | Other | -2.56E-08 | 2.10E-08 | 0.889135 |
| CAA | Nerve_Tibial | Other | -1.21E-08 | 2.17E-08 | 0.711498 |
| CAA | Ovary | Other | 2.11E-09 | 2.04E-08 | 0.458764 |
| CAA | Pituitary | Other | -3.49E-08 | 2.07E-08 | 0.953849 |
| CAA | Prostate | Other | -3.03E-08 | 2.51E-08 | 0.886385 |

| | | | | | |
|---|---|---|---|---|---|
| CAA | Skin_Not_Sun_Exposed_(Suprapubic) | Other | -3.56E-08 | 2.15E-08 | 0.951575 |
| CAA | Skin_Sun_Exposed_(Lower_leg) | Other | -9.11E-09 | 2.20E-08 | 0.660604 |
| CAA | Testis | Other | -3.94E-09 | 1.85E-08 | 0.584339 |
| CAA | Thyroid | Other | -1.71E-08 | 1.99E-08 | 0.805217 |
| CAA | Uterus | Other | -2.41E-08 | 2.05E-08 | 0.880038 |
| CAA | Vagina | Other | -1.57E-08 | 1.94E-08 | 0.791389 |
| CAA | A03.734.414.Islets.of.Langerhans | Pancreas | 1.76E-09 | 2.34E-08 | 0.470041 |
| CAA | A03.734.Pancreas | Pancreas | 1.65E-08 | 2.26E-08 | 0.233353 |
| CAA | Pancreas | Pancreas | -1.84E-08 | 2.08E-08 | 0.811424 |
| CAA (any/none) | A10.165.114.830.500.750.Subcutaneous.Fat..Abdominal | Adipose | -2.20E-08 | 2.10E-08 | 0.852296 |
| CAA (any/none) | A10.165.114.830.750.Subcutaneous.Fat | Adipose | -1.09E-08 | 2.02E-08 | 0.705153 |
| CAA (any/none) | A11.329.114.Adipocytes | Adipose | 1.85E-08 | 2.36E-08 | 0.216775 |
| CAA (any/none) | Adipose_Subcutaneous | Adipose | -1.78E-08 | 1.98E-08 | 0.814824 |
| CAA (any/none) | Adipose_Visceral_(Omentum) | Adipose | -3.49E-08 | 2.18E-08 | 0.945596 |
| CAA (any/none) | A02.835.583.443.800.800.Synovial.Fluid | Blood/Immune | 1.88E-09 | 2.46E-08 | 0.469562 |
| CAA (any/none) | A10.549.400.Lymph.Nodes | Blood/Immune | 1.56E-08 | 2.24E-08 | 0.243193 |
| CAA (any/none) | A10.549.Lymphoid.Tissue | Blood/Immune | -1.46E-09 | 2.22E-08 | 0.526222 |
| CAA (any/none) | A11.118.637.555.567.562.440.Precursor.Cells..B.Lymphoid | Blood/Immune | -6.20E-09 | 2.32E-08 | 0.605548 |
| CAA (any/none) | A11.118.637.555.567.562.B.Lymphocytes | Blood/Immune | -4.13E-09 | 2.23E-08 | 0.573611 |
| CAA (any/none) | A11.118.637.555.567.569.200.700.T.Lymphocytes..Regulatory | Blood/Immune | -1.05E-08 | 2.49E-08 | 0.663802 |
| CAA (any/none) | A11.118.637.555.567.569.T.Lymphocytes | Blood/Immune | 2.94E-09 | 2.44E-08 | 0.452039 |
| CAA (any/none) | A11.118.637.Leukocytes | Blood/Immune | -1.42E-08 | 2.76E-08 | 0.695903 |
| CAA (any/none) | A11.329.372.600.Macrophages..Alveolar | Blood/Immune | 1.55E-08 | 2.08E-08 | 0.228635 |
| CAA (any/none) | A11.443.Erythroid.Cells | Blood/Immune | -8.82E-09 | 2.21E-08 | 0.655076 |
| CAA (any/none) | A11.627.340.360.Granulocyte.Precursor.Cells | Blood/Immune | -2.14E-08 | 2.52E-08 | 0.801550 |
| CAA (any/none) | A11.627.624.249.Monocyte.Macrophage.Precursor.Cells | Blood/Immune | 9.45E-10 | 2.55E-08 | 0.485229 |
| CAA (any/none) | A11.627.635.Myeloid.Progenitor.Cells | Blood/Immune | 2.03E-08 | 2.48E-08 | 0.206698 |
| CAA (any/none) | A11.872.378.590.635.Granulocyte.Macrophage.Progenitor.Cells | Blood/Immune | -7.24E-10 | 2.60E-08 | 0.511105 |
| CAA (any/none) | A11.872.378.590.817.Megakaryocyte.Erythroid.Progenitor.Cells | Blood/Immune | 7.65E-09 | 2.37E-08 | 0.373421 |
| CAA (any/none) | A11.872.378.Hematopoietic.Stem.Cells | Blood/Immune | -7.56E-09 | 2.47E-08 | 0.620047 |
| CAA (any/none) | A15.145.229.188.Blood.Platelets | Blood/Immune | -1.18E-10 | 2.25E-08 | 0.502085 |
| CAA (any/none) | A15.145.229.637.555.567.562.725.Plasma.Cells | Blood/Immune | 7.00E-09 | 2.25E-08 | 0.377929 |
| CAA (any/none) | A15.145.229.637.555.567.569.200.CD4.Positive.T.Lymphocytes | Blood/Immune | -1.58E-08 | 2.53E-08 | 0.733733 |
| CAA (any/none) | A15.145.229.637.555.Leukocytes..Mononuclear | Blood/Immune | 9.14E-09 | 2.56E-08 | 0.360504 |
| CAA (any/none) | A15.145.229.Blood.Cells | Blood/Immune | -8.24E-09 | 2.64E-08 | 0.622382 |
| CAA (any/none) | A15.145.300.Fetal.Blood | Blood/Immune | -1.38E-09 | 2.86E-08 | 0.519186 |
| CAA (any/none) | A15.145.846.Serum | Blood/Immune | -6.63E-09 | 2.36E-08 | 0.610768 |
| CAA (any/none) | A15.145.Blood | Blood/Immune | -1.19E-08 | 2.54E-08 | 0.680808 |
| CAA (any/none) | A15.378.316.580.Monocytes | Blood/Immune | 4.83E-09 | 2.40E-08 | 0.420326 |
| CAA (any/none) | A15.378.316.Bone.Marrow.Cells | Blood/Immune | -7.40E-09 | 2.46E-08 | 0.618111 |
| CAA (any/none) | A15.382.490.315.583.Neutrophils | Blood/Immune | 1.03E-08 | 2.50E-08 | 0.340117 |
| CAA (any/none) | A15.382.490.555.567.537.Killer.Cells..Natural | Blood/Immune | -3.19E-08 | 2.31E-08 | 0.916565 |
| CAA (any/none) | A15.382.490.555.567.622.Lymphocytes..Null | Blood/Immune | -3.09E-08 | 2.42E-08 | 0.899111 |
| CAA (any/none) | A15.382.490.555.567.Lymphocytes | Blood/Immune | -4.31E-09 | 2.52E-08 | 0.567968 |
| CAA (any/none) | A15.382.520.604.700.Spleen | Blood/Immune | -1.01E-08 | 2.38E-08 | 0.664140 |
| CAA (any/none) | A15.382.520.604.800.Palatine.Tonsil | Blood/Immune | 2.34E-09 | 2.19E-08 | 0.457549 |

| | | | | | |
|---|---|---|---|---|---|
| CAA (any/none) | A15.382.680.Phagocytes | Blood/Immune | 2.50E-08 | 2.46E-08 | 0.154837 |
| CAA (any/none) | A15.382.812.260.Dendritic.Cells | Blood/Immune | -2.40E-09 | 2.30E-08 | 0.541469 |
| CAA (any/none) | A15.382.812.522.Macrophages | Blood/Immune | -6.91E-09 | 2.55E-08 | 0.606799 |
| CAA (any/none) | A15.382.812.Mononuclear.Phagocyte.System | Blood/Immune | 6.65E-09 | 2.55E-08 | 0.397044 |
| CAA (any/none) | A15.382.Immune.System | Blood/Immune | 2.67E-08 | 2.36E-08 | 0.129466 |
| CAA (any/none) | Cells_EBV-transformed_lymphocytes | Blood/Immune | -9.40E-09 | 2.34E-08 | 0.656366 |
| CAA (any/none) | Spleen | Blood/Immune | 2.30E-08 | 2.67E-08 | 0.194249 |
| CAA (any/none) | Whole_Blood | Blood/Immune | 5.29E-09 | 2.63E-08 | 0.420198 |
| CAA (any/none) | A07.231.114.Arteries | Cardiovascular | -1.71E-08 | 2.24E-08 | 0.777936 |
| CAA (any/none) | A07.231.908.670.874.Umbilical.Veins | Cardiovascular | -1.52E-08 | 2.39E-08 | 0.737513 |
| CAA (any/none) | A07.231.908.Veins | Cardiovascular | -1.59E-08 | 2.44E-08 | 0.742267 |
| CAA (any/none) | A07.231.Blood.Vessels | Cardiovascular | -1.46E-08 | 2.38E-08 | 0.729880 |
| CAA (any/none) | A07.541.358.100.Atrial.Appendage | Cardiovascular | -2.24E-08 | 1.95E-08 | 0.875325 |
| CAA (any/none) | A07.541.358.Heart.Atria | Cardiovascular | -1.14E-08 | 1.99E-08 | 0.717000 |
| CAA (any/none) | A07.541.510.110.Aortic.Valve | Cardiovascular | -1.98E-08 | 1.97E-08 | 0.842099 |
| CAA (any/none) | A07.541.560.Heart.Ventricles | Cardiovascular | -1.53E-08 | 2.15E-08 | 0.762137 |
| CAA (any/none) | A07.541.Heart | Cardiovascular | -1.04E-08 | 2.03E-08 | 0.695755 |
| CAA (any/none) | Artery_Aorta | Cardiovascular | 1.01E-08 | 1.95E-08 | 0.302381 |
| CAA (any/none) | Artery_Coronary | Cardiovascular | 2.66E-08 | 1.96E-08 | 0.087060 |
| CAA (any/none) | Artery_Tibial | Cardiovascular | 3.62E-08 | 1.97E-08 | 0.032990 |
| CAA (any/none) | Heart_Atrial_Appendage | Cardiovascular | -6.33E-09 | 1.86E-08 | 0.633184 |
| CAA (any/none) | Heart_Left_Ventricle | Cardiovascular | -3.47E-08 | 1.85E-08 | 0.970118 |
| CAA (any/none) | A08.186.211.132.810.428.200.Cerebellum | CNS | -2.27E-08 | 1.82E-08 | 0.893928 |
| CAA (any/none) | A08.186.211.132.Brain.Stem | CNS | -1.75E-08 | 1.89E-08 | 0.823101 |
| CAA (any/none) | A08.186.211.464.405.Hippocampus | CNS | -9.72E-10 | 2.12E-08 | 0.518289 |
| CAA (any/none) | A08.186.211.464.710.225.Entorhinal.Cortex | CNS | 7.09E-09 | 2.00E-08 | 0.361267 |
| CAA (any/none) | A08.186.211.464.Limbic.System | CNS | 4.94E-09 | 2.13E-08 | 0.408551 |
| CAA (any/none) | A08.186.211.653.Mesencephalon | CNS | 2.54E-08 | 2.21E-08 | 0.125835 |
| CAA (any/none) | A08.186.211.730.317.357.352.435.Hypothalamo.Hypophyseal.System | CNS | -4.94E-08 | 2.11E-08 | 0.990407 |
| CAA (any/none) | A08.186.211.730.317.357.Hypothalamus | CNS | -3.77E-08 | 2.27E-08 | 0.951565 |
| CAA (any/none) | A08.186.211.730.317.Diencephalon | CNS | 1.93E-08 | 2.35E-08 | 0.206418 |
| CAA (any/none) | A08.186.211.730.885.287.249.487.Corpus.Striatum | CNS | -2.39E-08 | 1.96E-08 | 0.889215 |
| CAA (any/none) | A08.186.211.730.885.287.249.Basal.Ganglia | CNS | -5.30E-09 | 2.09E-08 | 0.600167 |
| CAA (any/none) | A08.186.211.730.885.287.500.270.Frontal.Lobe | CNS | 1.22E-08 | 2.18E-08 | 0.287945 |
| CAA (any/none) | A08.186.211.730.885.287.500.571.735.Visual.Cortex | CNS | -9.84E-09 | 1.94E-08 | 0.694187 |
| CAA (any/none) | A08.186.211.730.885.287.500.670.Parietal.Lobe | CNS | -7.74E-09 | 2.28E-08 | 0.633096 |
| CAA (any/none) | A08.186.211.730.885.287.500.Cerebral.Cortex | CNS | 1.59E-10 | 2.07E-08 | 0.496936 |
| CAA (any/none) | A08.186.211.865.428.Metencephalon | CNS | -2.26E-08 | 1.79E-08 | 0.896190 |
| CAA (any/none) | A08.186.211.Brain | CNS | -3.93E-10 | 2.14E-08 | 0.507316 |
| CAA (any/none) | A09.371.729.Retina | CNS | -1.85E-08 | 2.03E-08 | 0.819004 |
| CAA (any/none) | A11.872.653.Neural.Stem.Cells | CNS | -9.96E-09 | 2.50E-08 | 0.654841 |
| CAA (any/none) | Brain_Amygdala | CNS | -3.60E-09 | 1.93E-08 | 0.573910 |
| CAA (any/none) | Brain_Anterior_cingulate_cortex_(BA24) | CNS | 1.18E-09 | 1.88E-08 | 0.474975 |
| CAA (any/none) | Brain_Caudate_(basal_ganglia) | CNS | -9.91E-10 | 1.86E-08 | 0.521228 |
| CAA (any/none) | Brain_Cerebellar_Hemisphere | CNS | -1.43E-08 | 1.80E-08 | 0.787587 |
| CAA (any/none) | Brain_Cerebellum | CNS | -1.52E-08 | 1.79E-08 | 0.802156 |

| | | | | | |
|---|---|---|---|---|---|
| CAA (any/none) | Brain_Cortex | CNS | 5.43E-09 | 1.97E-08 | 0.391536 |
| CAA (any/none) | Brain_Frontal_Cortex_(BA9) | CNS | 6.04E-09 | 1.84E-08 | 0.371762 |
| CAA (any/none) | Brain_Hippocampus | CNS | -6.08E-09 | 1.82E-08 | 0.630762 |
| CAA (any/none) | Brain_Hypothalamus | CNS | -8.61E-09 | 1.96E-08 | 0.669609 |
| CAA (any/none) | Brain_Nucleus_accumbens_(basal_ganglia) | CNS | -6.17E-10 | 1.89E-08 | 0.513007 |
| CAA (any/none) | Brain_Putamen_(basal_ganglia) | CNS | -5.09E-09 | 1.81E-08 | 0.610791 |
| CAA (any/none) | Brain_Spinal_cord_(cervical_c-1) | CNS | -9.36E-09 | 1.86E-08 | 0.692384 |
| CAA (any/none) | Brain_Substantia_nigra | CNS | 1.34E-08 | 1.80E-08 | 0.228385 |
| CAA (any/none) | A03.556.124.369.Intestinal.Mucosa | Digestive | 6.18E-08 | 2.46E-08 | 0.006045 |
| CAA (any/none) | A03.556.124.526.767.Rectum | Digestive | 4.52E-08 | 2.60E-08 | 0.040749 |
| CAA (any/none) | A03.556.124.684.Intestine..Small | Digestive | 4.40E-08 | 2.72E-08 | 0.052918 |
| CAA (any/none) | A03.556.124.Intestines | Digestive | 4.15E-08 | 2.62E-08 | 0.056776 |
| CAA (any/none) | A03.556.249.124.Ileum | Digestive | 1.84E-08 | 2.68E-08 | 0.246952 |
| CAA (any/none) | A03.556.249.249.209.Cecum | Digestive | 4.97E-08 | 2.29E-08 | 0.015174 |
| CAA (any/none) | A03.556.249.249.356.668.Colon..Sigmoid | Digestive | 4.03E-08 | 2.44E-08 | 0.049410 |
| CAA (any/none) | A03.556.249.249.356.Colon | Digestive | 4.32E-08 | 2.53E-08 | 0.043874 |
| CAA (any/none) | A03.556.500.760.464.Parotid.Gland | Digestive | -2.99E-08 | 2.52E-08 | 0.882316 |
| CAA (any/none) | A03.556.500.760.Salivary.Glands | Digestive | -2.59E-08 | 2.33E-08 | 0.867006 |
| CAA (any/none) | A03.556.875.500.Esophagus | Digestive | -2.45E-09 | 2.32E-08 | 0.541962 |
| CAA (any/none) | A03.556.875.875.Stomach | Digestive | 1.89E-08 | 2.43E-08 | 0.218550 |
| CAA (any/none) | A03.556.875.Upper.Gastrointestinal.Tract | Digestive | 2.17E-09 | 2.40E-08 | 0.463998 |
| CAA (any/none) | A03.556.Gastrointestinal.Tract | Digestive | 2.14E-08 | 2.63E-08 | 0.207067 |
| CAA (any/none) | Colon_Sigmoid | Digestive | -1.50E-08 | 1.89E-08 | 0.786959 |
| CAA (any/none) | Colon_Transverse | Digestive | 4.10E-08 | 2.28E-08 | 0.036010 |
| CAA (any/none) | Esophagus_Gastroesophageal_Junction | Digestive | 8.02E-09 | 2.06E-08 | 0.348516 |
| CAA (any/none) | Esophagus_Mucosa | Digestive | -1.89E-08 | 2.19E-08 | 0.806812 |
| CAA (any/none) | Esophagus_Muscularis | Digestive | 4.73E-09 | 2.14E-08 | 0.412576 |
| CAA (any/none) | Small_Intestine_Terminal_Ileum | Digestive | 3.19E-08 | 2.52E-08 | 0.102400 |
| CAA (any/none) | Stomach | Digestive | 2.98E-09 | 2.24E-08 | 0.447042 |
| CAA (any/none) | A03.620.Liver | Liver | 1.49E-08 | 2.52E-08 | 0.276969 |
| CAA (any/none) | A11.436.348.Hepatocytes | Liver | 3.21E-08 | 2.62E-08 | 0.109682 |
| CAA (any/none) | Liver | Liver | 1.48E-08 | 2.46E-08 | 0.274032 |
| CAA (any/none) | A02.165.Cartilage | Musculoskeletal/Connective | -4.40E-09 | 2.05E-08 | 0.585210 |
| CAA (any/none) | A02.633.567.850.Quadriceps.Muscle | Musculoskeletal/Connective | -8.09E-09 | 2.31E-08 | 0.636872 |
| CAA (any/none) | A02.835.232.834.151.Cervical.Vertebrae | Musculoskeletal/Connective | 1.52E-08 | 2.40E-08 | 0.262613 |
| CAA (any/none) | A02.835.583.443.800.Synovial.Membrane | Musculoskeletal/Connective | -1.73E-08 | 2.18E-08 | 0.786401 |
| CAA (any/none) | A05.360.319.679.690.Myometrium | Musculoskeletal/Connective | -4.04E-08 | 1.91E-08 | 0.982689 |
| CAA (any/none) | A10.165.450.300.425.Keloid | Musculoskeletal/Connective | 1.95E-09 | 2.13E-08 | 0.463491 |
| CAA (any/none) | A10.165.450.300.Cicatrix | Musculoskeletal/Connective | 1.94E-08 | 2.08E-08 | 0.175015 |
| CAA (any/none) | A10.690.467.Muscle..Smooth | Musculoskeletal/Connective | 2.29E-08 | 2.08E-08 | 0.136260 |
| CAA (any/none) | A10.690.Muscles | Musculoskeletal/Connective | -1.09E-08 | 2.38E-08 | 0.677170 |
| CAA (any/none) | A11.329.171.Chondrocytes | Musculoskeletal/Connective | 4.58E-09 | 2.41E-08 | 0.424590 |
| CAA (any/none) | A11.329.228.Fibroblasts | Musculoskeletal/Connective | -1.26E-09 | 2.04E-08 | 0.524615 |
| CAA (any/none) | A11.329.629.Osteoblasts | Musculoskeletal/Connective | 8.98E-09 | 2.33E-08 | 0.349854 |
| CAA (any/none) | A11.329.830.Stromal.Cells | Musculoskeletal/Connective | 2.03E-08 | 2.17E-08 | 0.175097 |
| CAA (any/none) | A11.329.Connective.Tissue.Cells | Musculoskeletal/Connective | -5.34E-09 | 2.31E-08 | 0.591279 |

| | | | | | |
|---|---|---|---|---|---|
| CAA (any/none) | A11.620.520.Myocytes..Smooth.Muscle | Musculoskeletal/Connective | -2.32E-08 | 2.11E-08 | 0.864360 |
| CAA (any/none) | Cells_Transformed_fibroblasts | Musculoskeletal/Connective | 8.22E-09 | 2.01E-08 | 0.340878 |
| CAA (any/none) | Muscle_Skeletal | Musculoskeletal/Connective | 5.84E-09 | 2.01E-08 | 0.385700 |
| CAA (any/none) | A04.411.Lung | Other | -1.14E-08 | 2.14E-08 | 0.703142 |
| CAA (any/none) | A04.531.520.Nasal.Mucosa | Other | -2.48E-08 | 2.36E-08 | 0.853623 |
| CAA (any/none) | A05.360.319.114.373.Fallopian.Tubes | Other | 1.12E-08 | 2.48E-08 | 0.325445 |
| CAA (any/none) | A05.360.319.114.630.Ovary | Other | 1.55E-09 | 2.31E-08 | 0.473199 |
| CAA (any/none) | A05.360.319.679.256.Cervix.Uteri | Other | 3.80E-09 | 2.16E-08 | 0.430292 |
| CAA (any/none) | A05.360.319.679.490.Endometrium | Other | -1.84E-09 | 2.16E-08 | 0.533975 |
| CAA (any/none) | A05.360.319.679.Uterus | Other | 2.75E-08 | 2.28E-08 | 0.113856 |
| CAA (any/none) | A05.360.319.887.Vulva | Other | 1.34E-08 | 2.42E-08 | 0.289151 |
| CAA (any/none) | A05.360.319.Genitalia..Female | Other | 1.44E-08 | 2.37E-08 | 0.272546 |
| CAA (any/none) | A05.360.444.492.362.Foreskin | Other | 6.71E-09 | 2.20E-08 | 0.380212 |
| CAA (any/none) | A05.360.444.Genitalia..Male | Other | -2.37E-08 | 1.92E-08 | 0.891072 |
| CAA (any/none) | A05.360.490.Germ.Cells | Other | 9.46E-09 | 2.04E-08 | 0.321150 |
| CAA (any/none) | A05.360.Genitalia | Other | 5.99E-09 | 2.13E-08 | 0.389337 |
| CAA (any/none) | A05.810.453.324.Kidney.Cortex | Other | 6.53E-08 | 2.30E-08 | 0.002292 |
| CAA (any/none) | A05.810.453.Kidney | Other | 1.02E-08 | 2.05E-08 | 0.309781 |
| CAA (any/none) | A05.810.890.Urinary.Bladder | Other | 3.02E-09 | 2.29E-08 | 0.447433 |
| CAA (any/none) | A06.407.071.140.Adrenal.Cortex | Other | -3.14E-08 | 2.13E-08 | 0.929902 |
| CAA (any/none) | A06.407.071.Adrenal.Glands | Other | -1.85E-08 | 2.24E-08 | 0.795635 |
| CAA (any/none) | A06.407.312.782.Testis | Other | -4.08E-08 | 2.03E-08 | 0.977602 |
| CAA (any/none) | A06.407.312.Gonads | Other | 4.29E-09 | 2.36E-08 | 0.427780 |
| CAA (any/none) | A06.407.900.Thyroid.Gland | Other | -3.78E-08 | 2.05E-08 | 0.967417 |
| CAA (any/none) | A06.407.Endocrine.Glands | Other | 1.31E-09 | 2.33E-08 | 0.477621 |
| CAA (any/none) | A09.371.Eye | Other | 1.77E-08 | 2.39E-08 | 0.229542 |
| CAA (any/none) | A10.272.497.Epidermis | Other | -3.11E-08 | 2.31E-08 | 0.911066 |
| CAA (any/none) | A10.272.Epithelium | Other | -1.98E-08 | 2.40E-08 | 0.795160 |
| CAA (any/none) | A10.336.707.Prostate | Other | -1.07E-08 | 1.87E-08 | 0.716323 |
| CAA (any/none) | A10.615.284.473.Chorion | Other | -1.68E-08 | 1.92E-08 | 0.809579 |
| CAA (any/none) | A10.615.550.599.Mouth.Mucosa | Other | 1.04E-09 | 2.29E-08 | 0.481948 |
| CAA (any/none) | A10.615.550.Mucous.Membrane | Other | 9.47E-09 | 2.40E-08 | 0.346896 |
| CAA (any/none) | A10.615.789.Serous.Membrane | Other | -9.97E-09 | 2.49E-08 | 0.655301 |
| CAA (any/none) | A10.615.Membranes | Other | 4.91E-09 | 2.50E-08 | 0.422105 |
| CAA (any/none) | A11.382.Endocrine.Cells | Other | 2.09E-08 | 2.20E-08 | 0.171596 |
| CAA (any/none) | A11.436.275.Endothelial.Cells | Other | -1.75E-09 | 2.33E-08 | 0.529855 |
| CAA (any/none) | A11.436.294.064.Glucagon.Secreting.Cells | Other | -2.97E-08 | 2.39E-08 | 0.892970 |
| CAA (any/none) | A11.436.329.Granulosa.Cells | Other | 2.94E-08 | 2.03E-08 | 0.074036 |
| CAA (any/none) | A11.436.397.Keratinocytes | Other | 1.24E-08 | 2.41E-08 | 0.303209 |
| CAA (any/none) | A11.436.Epithelial.Cells | Other | -3.03E-08 | 2.20E-08 | 0.916114 |
| CAA (any/none) | A11.497.497.600.Oocytes | Other | 1.23E-08 | 1.97E-08 | 0.266539 |
| CAA (any/none) | A11.872.040.Adult.Stem.Cells | Other | -5.17E-09 | 2.21E-08 | 0.592568 |
| CAA (any/none) | A11.872.190.260.Embryoid.Bodies | Other | 5.49E-09 | 2.26E-08 | 0.404152 |
| CAA (any/none) | A11.872.190.Embryonic.Stem.Cells | Other | -2.11E-08 | 2.14E-08 | 0.837466 |
| CAA (any/none) | A11.872.580.Mesenchymal.Stem.Cells | Other | 4.37E-09 | 2.34E-08 | 0.425967 |
| CAA (any/none) | A11.872.700.500.Induced.Pluripotent.Stem.Cells | Other | -4.09E-09 | 2.13E-08 | 0.576106 |

| | | | | | |
|---|---|---|---|---|---|
| CAA (any/none) | A11.872.Stem.Cells | Other | 2.18E-08 | 2.56E-08 | 0.197018 |
| CAA (any/none) | A14.549.167.646.Periodontium | Other | 2.27E-08 | 2.46E-08 | 0.178090 |
| CAA (any/none) | A14.549.167.Dentition | Other | 3.31E-08 | 2.42E-08 | 0.086100 |
| CAA (any/none) | A14.549.885.Tongue | Other | -4.15E-08 | 2.02E-08 | 0.979788 |
| CAA (any/none) | A14.549.Mouth | Other | -1.76E-08 | 2.19E-08 | 0.789701 |
| CAA (any/none) | A14.724.557.Nasopharynx | Other | 1.66E-08 | 2.12E-08 | 0.216253 |
| CAA (any/none) | A14.724.Pharynx | Other | -6.83E-09 | 2.04E-08 | 0.631362 |
| CAA (any/none) | A17.815.Skin | Other | -8.68E-09 | 2.24E-08 | 0.651109 |
| CAA (any/none) | Adrenal_Gland | Other | -8.72E-09 | 1.96E-08 | 0.671743 |
| CAA (any/none) | Bladder | Other | 1.03E-08 | 1.89E-08 | 0.293093 |
| CAA (any/none) | Breast_Mammary_Tissue | Other | -2.47E-08 | 2.00E-08 | 0.891485 |
| CAA (any/none) | Cervix_Ectocervix | Other | -1.72E-09 | 2.19E-08 | 0.531341 |
| CAA (any/none) | Cervix_Endocervix | Other | 3.29E-08 | 2.14E-08 | 0.062137 |
| CAA (any/none) | Fallopian_Tube | Other | -1.77E-08 | 2.31E-08 | 0.778288 |
| CAA (any/none) | Kidney_Cortex | Other | 1.21E-08 | 2.28E-08 | 0.297361 |
| CAA (any/none) | Lung | Other | -9.38E-09 | 2.27E-08 | 0.660519 |
| CAA (any/none) | Minor_Salivary_Gland | Other | -1.26E-08 | 2.07E-08 | 0.728808 |
| CAA (any/none) | Nerve_Tibial | Other | -2.17E-08 | 2.12E-08 | 0.847786 |
| CAA (any/none) | Ovary | Other | 4.23E-09 | 1.88E-08 | 0.410883 |
| CAA (any/none) | Pituitary | Other | -5.92E-08 | 2.19E-08 | 0.996500 |
| CAA (any/none) | Prostate | Other | 8.37E-09 | 2.47E-08 | 0.367252 |
| CAA (any/none) | Skin_Not_Sun_Exposed_(Suprapubic) | Other | -4.39E-08 | 2.12E-08 | 0.980914 |
| CAA (any/none) | Skin_Sun_Exposed_(Lower_leg) | Other | -3.37E-08 | 2.09E-08 | 0.946413 |
| CAA (any/none) | Testis | Other | -3.10E-08 | 1.83E-08 | 0.954964 |
| CAA (any/none) | Thyroid | Other | -2.89E-08 | 2.10E-08 | 0.915405 |
| CAA (any/none) | Uterus | Other | -1.15E-08 | 2.05E-08 | 0.712757 |
| CAA (any/none) | Vagina | Other | 5.71E-10 | 2.03E-08 | 0.488773 |
| CAA (any/none) | A03.734.414.Islets.of.Langerhans | Pancreas | -1.45E-08 | 2.33E-08 | 0.732673 |
| CAA (any/none) | A03.734.Pancreas | Pancreas | 5.31E-09 | 2.56E-08 | 0.417813 |
| CAA (any/none) | Pancreas | Pancreas | 6.09E-09 | 2.10E-08 | 0.385888 |
| CERAD (C Score) | A10.165.114.830.500.750.Subcutaneous.Fat..Abdominal | Adipose | 5.20E-09 | 1.77E-08 | 0.384690 |
| CERAD (C Score) | A10.165.114.830.750.Subcutaneous.Fat | Adipose | 1.02E-08 | 1.81E-08 | 0.285992 |
| CERAD (C Score) | A11.329.114.Adipocytes | Adipose | -1.46E-08 | 1.77E-08 | 0.795203 |
| CERAD (C Score) | Adipose_Subcutaneous | Adipose | 1.90E-08 | 1.66E-08 | 0.126500 |
| CERAD (C Score) | Adipose_Visceral_(Omentum) | Adipose | 1.82E-10 | 1.59E-08 | 0.495443 |
| CERAD (C Score) | A02.835.583.443.800.800.Synovial.Fluid | Blood/Immune | 1.30E-08 | 2.37E-08 | 0.291267 |
| CERAD (C Score) | A10.549.400.Lymph.Nodes | Blood/Immune | -4.07E-09 | 1.95E-08 | 0.582878 |
| CERAD (C Score) | A10.549.Lymphoid.Tissue | Blood/Immune | 1.71E-09 | 2.07E-08 | 0.467152 |
| CERAD (C Score) | A11.118.637.555.567.562.440.Precursor.Cells..B.Lymphoid | Blood/Immune | -1.20E-08 | 2.08E-08 | 0.717488 |
| CERAD (C Score) | A11.118.637.555.567.562.B.Lymphocytes | Blood/Immune | 3.93E-09 | 2.14E-08 | 0.427218 |
| CERAD (C Score) | A11.118.637.555.567.569.200.700.T.Lymphocytes..Regulatory | Blood/Immune | -1.86E-08 | 1.93E-08 | 0.833174 |
| CERAD (C Score) | A11.118.637.555.567.569.T.Lymphocytes | Blood/Immune | -1.33E-08 | 1.74E-08 | 0.777901 |
| CERAD (C Score) | A11.118.637.Leukocytes | Blood/Immune | 2.79E-08 | 2.48E-08 | 0.130324 |
| CERAD (C Score) | A11.329.372.600.Macrophages..Alveolar | Blood/Immune | 2.48E-08 | 1.86E-08 | 0.091323 |
| CERAD (C Score) | A11.443.Erythroid.Cells | Blood/Immune | 1.78E-08 | 1.78E-08 | 0.159386 |
| CERAD (C Score) | A11.627.340.360.Granulocyte.Precursor.Cells | Blood/Immune | -3.58E-09 | 1.98E-08 | 0.571877 |

| | | | | | |
|---|---|---|---|---|---|
| CERAD (C Score) | A11.627.624.249.Monocyte.Macrophage.Precursor.Cells | Blood/Immune | 3.09E-08 | 1.80E-08 | 0.043179 |
| CERAD (C Score) | A11.627.635.Myeloid.Progenitor.Cells | Blood/Immune | 5.04E-08 | 1.94E-08 | 0.004621 |
| CERAD (C Score) | A11.872.378.590.635.Granulocyte.Macrophage.Progenitor.Cells | Blood/Immune | 2.84E-08 | 1.94E-08 | 0.072160 |
| CERAD (C Score) | A11.872.378.590.817.Megakaryocyte.Erythroid.Progenitor.Cells | Blood/Immune | 4.60E-08 | 2.04E-08 | 0.012162 |
| CERAD (C Score) | A11.872.378.Hematopoietic.Stem.Cells | Blood/Immune | 1.94E-08 | 2.02E-08 | 0.169095 |
| CERAD (C Score) | A15.145.229.188.Blood.Platelets | Blood/Immune | 7.78E-09 | 1.88E-08 | 0.339918 |
| CERAD (C Score) | A15.145.229.637.555.567.562.725.Plasma.Cells | Blood/Immune | 7.33E-09 | 2.02E-08 | 0.358513 |
| CERAD (C Score) | A15.145.229.637.555.567.569.200.CD4.Positive.T.Lymphocytes | Blood/Immune | -1.31E-08 | 1.85E-08 | 0.761232 |
| CERAD (C Score) | A15.145.229.637.555.Leukocytes..Mononuclear | Blood/Immune | 2.89E-09 | 2.25E-08 | 0.448953 |
| CERAD (C Score) | A15.145.229.Blood.Cells | Blood/Immune | 2.35E-08 | 2.45E-08 | 0.168881 |
| CERAD (C Score) | A15.145.300.Fetal.Blood | Blood/Immune | 1.22E-08 | 2.22E-08 | 0.291006 |
| CERAD (C Score) | A15.145.846.Serum | Blood/Immune | 2.03E-08 | 1.93E-08 | 0.145805 |
| CERAD (C Score) | A15.145.Blood | Blood/Immune | 1.46E-08 | 2.32E-08 | 0.264556 |
| CERAD (C Score) | A15.378.316.580.Monocytes | Blood/Immune | 5.18E-08 | 2.07E-08 | 0.006097 |
| CERAD (C Score) | A15.378.316.Bone.Marrow.Cells | Blood/Immune | 4.10E-08 | 2.26E-08 | 0.035108 |
| CERAD (C Score) | A15.382.490.315.583.Neutrophils | Blood/Immune | 3.20E-08 | 1.95E-08 | 0.049925 |
| CERAD (C Score) | A15.382.490.555.567.537.Killer.Cells..Natural | Blood/Immune | -9.68E-09 | 1.72E-08 | 0.713537 |
| CERAD (C Score) | A15.382.490.555.567.622.Lymphocytes..Null | Blood/Immune | -4.91E-09 | 1.98E-08 | 0.597861 |
| CERAD (C Score) | A15.382.490.555.567.Lymphocytes | Blood/Immune | 6.63E-09 | 2.23E-08 | 0.383179 |
| CERAD (C Score) | A15.382.520.604.700.Spleen | Blood/Immune | 1.42E-08 | 1.89E-08 | 0.226224 |
| CERAD (C Score) | A15.382.520.604.800.Palatine.Tonsil | Blood/Immune | 2.82E-08 | 1.90E-08 | 0.068565 |
| CERAD (C Score) | A15.382.680.Phagocytes | Blood/Immune | 5.98E-08 | 2.17E-08 | 0.002918 |
| CERAD (C Score) | A15.382.812.260.Dendritic.Cells | Blood/Immune | 3.94E-08 | 2.04E-08 | 0.026529 |
| CERAD (C Score) | A15.382.812.522.Macrophages | Blood/Immune | 4.41E-08 | 1.93E-08 | 0.011266 |
| CERAD (C Score) | A15.382.812.Mononuclear.Phagocyte.System | Blood/Immune | 5.06E-08 | 2.06E-08 | 0.006884 |
| CERAD (C Score) | A15.382.Immune.System | Blood/Immune | 1.46E-08 | 2.39E-08 | 0.270614 |
| CERAD (C Score) | Cells_EBV-transformed_lymphocytes | Blood/Immune | 2.73E-08 | 1.87E-08 | 0.071883 |
| CERAD (C Score) | Spleen | Blood/Immune | 1.61E-09 | 2.11E-08 | 0.469526 |
| CERAD (C Score) | Whole_Blood | Blood/Immune | 2.24E-08 | 1.94E-08 | 0.123942 |
| CERAD (C Score) | A07.231.114.Arteries | Cardiovascular | -1.66E-08 | 1.98E-08 | 0.799139 |
| CERAD (C Score) | A07.231.908.670.874.Umbilical.Veins | Cardiovascular | 1.68E-08 | 1.90E-08 | 0.188989 |
| CERAD (C Score) | A07.231.908.Veins | Cardiovascular | 1.09E-08 | 1.87E-08 | 0.279902 |
| CERAD (C Score) | A07.231.Blood.Vessels | Cardiovascular | 2.56E-08 | 2.00E-08 | 0.101082 |
| CERAD (C Score) | A07.541.358.100.Atrial.Appendage | Cardiovascular | -2.70E-09 | 1.50E-08 | 0.571265 |
| CERAD (C Score) | A07.541.358.Heart.Atria | Cardiovascular | -4.89E-11 | 1.52E-08 | 0.501283 |
| CERAD (C Score) | A07.541.510.110.Aortic.Valve | Cardiovascular | 1.52E-08 | 1.74E-08 | 0.190819 |
| CERAD (C Score) | A07.541.560.Heart.Ventricles | Cardiovascular | -7.63E-09 | 1.59E-08 | 0.684549 |
| CERAD (C Score) | A07.541.Heart | Cardiovascular | -3.32E-09 | 1.54E-08 | 0.585053 |
| CERAD (C Score) | Artery_Aorta | Cardiovascular | 3.13E-09 | 1.55E-08 | 0.420240 |
| CERAD (C Score) | Artery_Coronary | Cardiovascular | 3.83E-09 | 1.59E-08 | 0.404772 |
| CERAD (C Score) | Artery_Tibial | Cardiovascular | -1.51E-09 | 1.40E-08 | 0.542963 |
| CERAD (C Score) | Heart_Atrial_Appendage | Cardiovascular | 9.43E-09 | 1.46E-08 | 0.259757 |
| CERAD (C Score) | Heart_Left_Ventricle | Cardiovascular | -1.07E-09 | 1.38E-08 | 0.531063 |
| CERAD (C Score) | A08.186.211.132.810.428.200.Cerebellum | CNS | -4.86E-09 | 1.44E-08 | 0.632227 |
| CERAD (C Score) | A08.186.211.132.Brain.Stem | CNS | -8.36E-10 | 1.44E-08 | 0.523118 |
| CERAD (C Score) | A08.186.211.464.405.Hippocampus | CNS | 2.02E-09 | 1.51E-08 | 0.446625 |

| | | | | | |
|---|---|---|---|---|---|
| CERAD (C Score) | A08.186.211.464.710.225.Entorhinal.Cortex | CNS | 7.14E-09 | 1.55E-08 | 0.322978 |
| CERAD (C Score) | A08.186.211.464.Limbic.System | CNS | 5.09E-09 | 1.56E-08 | 0.372174 |
| CERAD (C Score) | A08.186.211.653.Mesencephalon | CNS | 2.07E-08 | 1.61E-08 | 0.098905 |
| CERAD (C Score) | A08.186.211.730.317.357.352.435.Hypothalamo.Hypophyseal.System | CNS | 1.12E-08 | 2.20E-08 | 0.304695 |
| CERAD (C Score) | A08.186.211.730.317.357.Hypothalamus | CNS | 9.02E-09 | 2.25E-08 | 0.344363 |
| CERAD (C Score) | A08.186.211.730.317.Diencephalon | CNS | 1.93E-08 | 2.36E-08 | 0.206932 |
| CERAD (C Score) | A08.186.211.730.885.287.249.487.Corpus.Striatum | CNS | -1.62E-08 | 1.56E-08 | 0.851412 |
| CERAD (C Score) | A08.186.211.730.885.287.249.Basal.Ganglia | CNS | 1.33E-08 | 1.69E-08 | 0.216648 |
| CERAD (C Score) | A08.186.211.730.885.287.500.270.Frontal.Lobe | CNS | -2.01E-08 | 1.59E-08 | 0.897301 |
| CERAD (C Score) | A08.186.211.730.885.287.500.571.735.Visual.Cortex | CNS | -2.74E-08 | 1.58E-08 | 0.958468 |
| CERAD (C Score) | A08.186.211.730.885.287.500.670.Parietal.Lobe | CNS | 1.34E-08 | 1.57E-08 | 0.197111 |
| CERAD (C Score) | A08.186.211.730.885.287.500.Cerebral.Cortex | CNS | 4.69E-09 | 1.53E-08 | 0.379541 |
| CERAD (C Score) | A08.186.211.865.428.Metencephalon | CNS | -1.10E-08 | 1.42E-08 | 0.779892 |
| CERAD (C Score) | A08.186.211.Brain | CNS | 2.27E-08 | 1.53E-08 | 0.069149 |
| CERAD (C Score) | A09.371.729.Retina | CNS | -3.95E-09 | 1.51E-08 | 0.602972 |
| CERAD (C Score) | A11.872.653.Neural.Stem.Cells | CNS | -3.03E-08 | 1.79E-08 | 0.954576 |
| CERAD (C Score) | Brain_Amygdala | CNS | -8.52E-09 | 1.51E-08 | 0.714050 |
| CERAD (C Score) | Brain_Anterior_cingulate_cortex_(BA24) | CNS | 3.92E-09 | 1.51E-08 | 0.397730 |
| CERAD (C Score) | Brain_Caudate_(basal_ganglia) | CNS | -1.55E-10 | 1.53E-08 | 0.504046 |
| CERAD (C Score) | Brain_Cerebellar_Hemisphere | CNS | 3.87E-09 | 1.47E-08 | 0.396149 |
| CERAD (C Score) | Brain_Cerebellum | CNS | -1.37E-08 | 1.49E-08 | 0.822256 |
| CERAD (C Score) | Brain_Cortex | CNS | 5.11E-09 | 1.49E-08 | 0.365773 |
| CERAD (C Score) | Brain_Frontal_Cortex_(BA9) | CNS | 1.08E-09 | 1.41E-08 | 0.469487 |
| CERAD (C Score) | Brain_Hippocampus | CNS | -8.77E-09 | 1.46E-08 | 0.725323 |
| CERAD (C Score) | Brain_Hypothalamus | CNS | -1.22E-08 | 1.50E-08 | 0.790781 |
| CERAD (C Score) | Brain_Nucleus_accumbens_(basal_ganglia) | CNS | -4.46E-09 | 1.52E-08 | 0.615811 |
| CERAD (C Score) | Brain_Putamen_(basal_ganglia) | CNS | -1.03E-08 | 1.47E-08 | 0.759661 |
| CERAD (C Score) | Brain_Spinal_cord_(cervical_c-1) | CNS | 1.56E-08 | 1.48E-08 | 0.146906 |
| CERAD (C Score) | Brain_Substantia_nigra | CNS | 8.14E-09 | 1.49E-08 | 0.292173 |
| CERAD (C Score) | A03.556.124.369.Intestinal.Mucosa | Digestive | -2.79E-08 | 1.84E-08 | 0.934843 |
| CERAD (C Score) | A03.556.124.526.767.Rectum | Digestive | 4.05E-09 | 1.92E-08 | 0.416663 |
| CERAD (C Score) | A03.556.124.684.Intestine..Small | Digestive | 9.35E-09 | 1.95E-08 | 0.315889 |
| CERAD (C Score) | A03.556.124.Intestines | Digestive | 5.13E-09 | 1.94E-08 | 0.395512 |
| CERAD (C Score) | A03.556.249.124.Ileum | Digestive | -3.86E-09 | 2.12E-08 | 0.572173 |
| CERAD (C Score) | A03.556.249.249.209.Cecum | Digestive | -1.26E-08 | 1.58E-08 | 0.786777 |
| CERAD (C Score) | A03.556.249.249.356.668.Colon..Sigmoid | Digestive | -2.09E-08 | 1.75E-08 | 0.884346 |
| CERAD (C Score) | A03.556.249.249.356.Colon | Digestive | 3.26E-09 | 1.80E-08 | 0.427982 |
| CERAD (C Score) | A03.556.500.760.464.Parotid.Gland | Digestive | -1.01E-08 | 2.09E-08 | 0.685224 |
| CERAD (C Score) | A03.556.500.760.Salivary.Glands | Digestive | -1.31E-08 | 1.99E-08 | 0.745610 |
| CERAD (C Score) | A03.556.875.500.Esophagus | Digestive | 3.25E-09 | 1.88E-08 | 0.431454 |
| CERAD (C Score) | A03.556.875.875.Stomach | Digestive | 4.50E-09 | 1.99E-08 | 0.410272 |
| CERAD (C Score) | A03.556.875.Upper.Gastrointestinal.Tract | Digestive | 3.95E-09 | 2.12E-08 | 0.426056 |
| CERAD (C Score) | A03.556.Gastrointestinal.Tract | Digestive | 4.58E-09 | 1.86E-08 | 0.402682 |
| CERAD (C Score) | Colon_Sigmoid | Digestive | 5.89E-09 | 1.58E-08 | 0.354978 |
| CERAD (C Score) | Colon_Transverse | Digestive | -7.88E-09 | 1.73E-08 | 0.675163 |
| CERAD (C Score) | Esophagus_Gastroesophageal_Junction | Digestive | -2.17E-08 | 1.66E-08 | 0.905035 |

| | | | | | |
|---|---|---|---|---|---|
| CERAD (C Score) | Esophagus_Mucosa | Digestive | -4.22E-09 | 1.87E-08 | 0.589034 |
| CERAD (C Score) | Esophagus_Muscularis | Digestive | -1.10E-08 | 1.65E-08 | 0.748071 |
| CERAD (C Score) | Small_Intestine_Terminal_Ileum | Digestive | 1.30E-08 | 1.83E-08 | 0.239243 |
| CERAD (C Score) | Stomach | Digestive | -8.18E-09 | 1.76E-08 | 0.679173 |
| CERAD (C Score) | A03.620.Liver | Liver | -1.82E-08 | 1.75E-08 | 0.850846 |
| CERAD (C Score) | A11.436.348.Hepatocytes | Liver | 2.86E-08 | 2.12E-08 | 0.088243 |
| CERAD (C Score) | Liver | Liver | -1.31E-08 | 1.76E-08 | 0.772270 |
| CERAD (C Score) | A02.165.Cartilage | Musculoskeletal/Connective | -1.27E-08 | 1.57E-08 | 0.790906 |
| CERAD (C Score) | A02.633.567.850.Quadriceps.Muscle | Musculoskeletal/Connective | 5.63E-09 | 1.80E-08 | 0.377376 |
| CERAD (C Score) | A02.835.232.834.151.Cervical.Vertebrae | Musculoskeletal/Connective | 1.44E-08 | 1.80E-08 | 0.211472 |
| CERAD (C Score) | A02.835.583.443.800.Synovial.Membrane | Musculoskeletal/Connective | -7.15E-09 | 1.77E-08 | 0.656971 |
| CERAD (C Score) | A05.360.319.679.690.Myometrium | Musculoskeletal/Connective | 1.95E-08 | 1.62E-08 | 0.114016 |
| CERAD (C Score) | A10.165.450.300.425.Keloid | Musculoskeletal/Connective | 1.03E-08 | 1.77E-08 | 0.279336 |
| CERAD (C Score) | A10.165.450.300.Cicatrix | Musculoskeletal/Connective | 2.01E-08 | 1.81E-08 | 0.133581 |
| CERAD (C Score) | A10.690.467.Muscle..Smooth | Musculoskeletal/Connective | 2.04E-08 | 2.19E-08 | 0.176429 |
| CERAD (C Score) | A10.690.Muscles | Musculoskeletal/Connective | -6.78E-09 | 1.82E-08 | 0.645311 |
| CERAD (C Score) | A11.329.171.Chondrocytes | Musculoskeletal/Connective | 7.90E-09 | 1.85E-08 | 0.334656 |
| CERAD (C Score) | A11.329.228.Fibroblasts | Musculoskeletal/Connective | 1.30E-08 | 1.73E-08 | 0.227077 |
| CERAD (C Score) | A11.329.629.Osteoblasts | Musculoskeletal/Connective | -5.05E-09 | 1.74E-08 | 0.613934 |
| CERAD (C Score) | A11.329.830.Stromal.Cells | Musculoskeletal/Connective | 3.82E-08 | 1.70E-08 | 0.012468 |
| CERAD (C Score) | A11.329.Connective.Tissue.Cells | Musculoskeletal/Connective | 4.59E-08 | 1.84E-08 | 0.006275 |
| CERAD (C Score) | A11.620.520.Myocytes..Smooth.Muscle | Musculoskeletal/Connective | 1.83E-09 | 2.12E-08 | 0.465537 |
| CERAD (C Score) | Cells_Transformed_fibroblasts | Musculoskeletal/Connective | 2.30E-08 | 1.70E-08 | 0.087806 |
| CERAD (C Score) | Muscle_Skeletal | Musculoskeletal/Connective | -5.54E-08 | 1.77E-08 | 0.999131 |
| CERAD (C Score) | A04.411.Lung | Other | -1.66E-09 | 1.83E-08 | 0.536111 |
| CERAD (C Score) | A04.531.520.Nasal.Mucosa | Other | -8.35E-09 | 1.85E-08 | 0.674086 |
| CERAD (C Score) | A05.360.319.114.373.Fallopian.Tubes | Other | 1.39E-08 | 1.80E-08 | 0.219688 |
| CERAD (C Score) | A05.360.319.114.630.Ovary | Other | 1.41E-08 | 1.74E-08 | 0.209300 |
| CERAD (C Score) | A05.360.319.679.256.Cervix.Uteri | Other | 7.93E-09 | 1.95E-08 | 0.342137 |
| CERAD (C Score) | A05.360.319.679.490.Endometrium | Other | 1.87E-09 | 1.67E-08 | 0.455519 |
| CERAD (C Score) | A05.360.319.679.Uterus | Other | 8.46E-09 | 1.82E-08 | 0.320948 |
| CERAD (C Score) | A05.360.319.887.Vulva | Other | 1.50E-08 | 1.91E-08 | 0.216520 |
| CERAD (C Score) | A05.360.319.Genitalia..Female | Other | 1.29E-08 | 1.75E-08 | 0.230212 |
| CERAD (C Score) | A05.360.444.492.362.Foreskin | Other | 2.64E-08 | 1.62E-08 | 0.051271 |
| CERAD (C Score) | A05.360.444.Genitalia..Male | Other | 2.34E-08 | 1.41E-08 | 0.048562 |
| CERAD (C Score) | A05.360.490.Germ.Cells | Other | 1.67E-08 | 1.71E-08 | 0.165421 |
| CERAD (C Score) | A05.360.Genitalia | Other | -2.38E-09 | 1.64E-08 | 0.557756 |
| CERAD (C Score) | A05.810.453.324.Kidney.Cortex | Other | 4.31E-08 | 1.86E-08 | 0.010447 |
| CERAD (C Score) | A05.810.453.Kidney | Other | -1.43E-08 | 1.74E-08 | 0.794943 |
| CERAD (C Score) | A05.810.890.Urinary.Bladder | Other | -5.80E-09 | 2.16E-08 | 0.605891 |
| CERAD (C Score) | A06.407.071.140.Adrenal.Cortex | Other | -5.84E-09 | 1.82E-08 | 0.625792 |
| CERAD (C Score) | A06.407.071.Adrenal.Glands | Other | -7.95E-09 | 1.76E-08 | 0.673707 |
| CERAD (C Score) | A06.407.312.782.Testis | Other | 2.58E-09 | 1.44E-08 | 0.428910 |
| CERAD (C Score) | A06.407.312.Gonads | Other | 8.19E-09 | 1.70E-08 | 0.314593 |
| CERAD (C Score) | A06.407.900.Thyroid.Gland | Other | -1.12E-08 | 1.60E-08 | 0.759028 |
| CERAD (C Score) | A06.407.Endocrine.Glands | Other | 6.93E-09 | 1.66E-08 | 0.338336 |

| | | | | | |
|---|---|---|---|---|---|
| CERAD (C Score) | A09.371.Eye | Other | -3.54E-08 | 1.84E-08 | 0.972753 |
| CERAD (C Score) | A10.272.497.Epidermis | Other | 2.24E-08 | 2.17E-08 | 0.150933 |
| CERAD (C Score) | A10.272.Epithelium | Other | -3.51E-09 | 1.92E-08 | 0.572625 |
| CERAD (C Score) | A10.336.707.Prostate | Other | 1.78E-08 | 1.59E-08 | 0.130817 |
| CERAD (C Score) | A10.615.284.473.Chorion | Other | 2.83E-08 | 1.91E-08 | 0.068800 |
| CERAD (C Score) | A10.615.550.599.Mouth.Mucosa | Other | -4.70E-09 | 1.90E-08 | 0.597618 |
| CERAD (C Score) | A10.615.550.Mucous.Membrane | Other | 9.18E-09 | 1.97E-08 | 0.320578 |
| CERAD (C Score) | A10.615.789.Serous.Membrane | Other | 1.03E-08 | 1.94E-08 | 0.297048 |
| CERAD (C Score) | A10.615.Membranes | Other | 5.51E-09 | 1.95E-08 | 0.388719 |
| CERAD (C Score) | A11.382.Endocrine.Cells | Other | 2.15E-08 | 1.63E-08 | 0.094159 |
| CERAD (C Score) | A11.436.275.Endothelial.Cells | Other | 1.79E-08 | 2.06E-08 | 0.192147 |
| CERAD (C Score) | A11.436.294.064.Glucagon.Secreting.Cells | Other | 2.00E-08 | 1.91E-08 | 0.147375 |
| CERAD (C Score) | A11.436.329.Granulosa.Cells | Other | 1.10E-08 | 1.69E-08 | 0.257427 |
| CERAD (C Score) | A11.436.397.Keratinocytes | Other | 3.32E-08 | 2.12E-08 | 0.058871 |
| CERAD (C Score) | A11.436.Epithelial.Cells | Other | 9.68E-09 | 1.80E-08 | 0.295312 |
| CERAD (C Score) | A11.497.497.600.Oocytes | Other | 1.46E-08 | 1.78E-08 | 0.206190 |
| CERAD (C Score) | A11.872.040.Adult.Stem.Cells | Other | -1.74E-09 | 1.92E-08 | 0.536150 |
| CERAD (C Score) | A11.872.190.260.Embryoid.Bodies | Other | 9.77E-09 | 1.80E-08 | 0.294024 |
| CERAD (C Score) | A11.872.190.Embryonic.Stem.Cells | Other | 3.30E-08 | 1.86E-08 | 0.037723 |
| CERAD (C Score) | A11.872.580.Mesenchymal.Stem.Cells | Other | 6.54E-09 | 1.77E-08 | 0.356099 |
| CERAD (C Score) | A11.872.700.500.Induced.Pluripotent.Stem.Cells | Other | 1.02E-08 | 1.93E-08 | 0.298634 |
| CERAD (C Score) | A11.872.Stem.Cells | Other | 3.26E-08 | 2.07E-08 | 0.057521 |
| CERAD (C Score) | A14.549.167.646.Periodontium | Other | 1.90E-08 | 2.08E-08 | 0.180294 |
| CERAD (C Score) | A14.549.167.Dentition | Other | 1.87E-08 | 1.91E-08 | 0.164498 |
| CERAD (C Score) | A14.549.885.Tongue | Other | -7.56E-09 | 1.89E-08 | 0.655639 |
| CERAD (C Score) | A14.549.Mouth | Other | 3.94E-09 | 1.86E-08 | 0.416273 |
| CERAD (C Score) | A14.724.557.Nasopharynx | Other | -8.81E-09 | 1.83E-08 | 0.685177 |
| CERAD (C Score) | A14.724.Pharynx | Other | 1.51E-10 | 1.88E-08 | 0.496792 |
| CERAD (C Score) | A17.815.Skin | Other | 3.57E-08 | 1.95E-08 | 0.033571 |
| CERAD (C Score) | Adrenal_Gland | Other | -8.89E-09 | 1.74E-08 | 0.695691 |
| CERAD (C Score) | Bladder | Other | 3.34E-09 | 1.42E-08 | 0.406900 |
| CERAD (C Score) | Breast_Mammary_Tissue | Other | 2.47E-09 | 1.66E-08 | 0.440695 |
| CERAD (C Score) | Cervix_Ectocervix | Other | 2.88E-08 | 1.61E-08 | 0.036804 |
| CERAD (C Score) | Cervix_Endocervix | Other | 1.21E-08 | 1.70E-08 | 0.237667 |
| CERAD (C Score) | Fallopian_Tube | Other | -9.77E-09 | 1.54E-08 | 0.736763 |
| CERAD (C Score) | Kidney_Cortex | Other | 3.86E-08 | 1.63E-08 | 0.009014 |
| CERAD (C Score) | Lung | Other | 1.91E-08 | 1.85E-08 | 0.151550 |
| CERAD (C Score) | Minor_Salivary_Gland | Other | 1.84E-08 | 1.69E-08 | 0.137757 |
| CERAD (C Score) | Nerve_Tibial | Other | 1.43E-08 | 1.55E-08 | 0.177389 |
| CERAD (C Score) | Ovary | Other | -1.36E-08 | 1.55E-08 | 0.809959 |
| CERAD (C Score) | Pituitary | Other | 2.38E-08 | 1.80E-08 | 0.092030 |
| CERAD (C Score) | Prostate | Other | 1.97E-08 | 1.79E-08 | 0.136282 |
| CERAD (C Score) | Skin_Not_Sun_Exposed_(Suprapubic) | Other | 3.09E-08 | 1.72E-08 | 0.036106 |
| CERAD (C Score) | Skin_Sun_Exposed_(Lower_leg) | Other | 4.11E-08 | 1.74E-08 | 0.008915 |
| CERAD (C Score) | Testis | Other | 5.61E-09 | 1.61E-08 | 0.364059 |
| CERAD (C Score) | Thyroid | Other | 6.45E-10 | 1.45E-08 | 0.482237 |

| | | | | | |
|---|---|---|---|---|---|
| CERAD (C Score) | Uterus | Other | -2.09E-10 | 1.62E-08 | 0.505145 |
| CERAD (C Score) | Vagina | Other | 1.14E-08 | 1.73E-08 | 0.254657 |
| CERAD (C Score) | A03.734.414.Islets.of.Langerhans | Pancreas | 1.63E-08 | 1.89E-08 | 0.194290 |
| CERAD (C Score) | A03.734.Pancreas | Pancreas | -3.36E-08 | 1.54E-08 | 0.985509 |
| CERAD (C Score) | Pancreas | Pancreas | -2.21E-08 | 1.66E-08 | 0.908627 |
| Infarcts/lacunes | A10.165.114.830.500.750.Subcutaneous.Fat..Abdominal | Adipose | 1.58E-09 | 2.20E-08 | 0.471393 |
| Infarcts/lacunes | A10.165.114.830.750.Subcutaneous.Fat | Adipose | -3.77E-09 | 2.12E-08 | 0.570532 |
| Infarcts/lacunes | A11.329.114.Adipocytes | Adipose | 6.01E-09 | 2.11E-08 | 0.387895 |
| Infarcts/lacunes | Adipose_Subcutaneous | Adipose | 5.10E-09 | 2.00E-08 | 0.399239 |
| Infarcts/lacunes | Adipose_Visceral_(Omentum) | Adipose | 1.10E-08 | 2.20E-08 | 0.308740 |
| Infarcts/lacunes | A02.835.583.443.800.800.Synovial.Fluid | Blood/Immune | -2.28E-08 | 2.72E-08 | 0.799196 |
| Infarcts/lacunes | A10.549.400.Lymph.Nodes | Blood/Immune | -2.88E-08 | 2.31E-08 | 0.893722 |
| Infarcts/lacunes | A10.549.Lymphoid.Tissue | Blood/Immune | -2.98E-08 | 2.23E-08 | 0.909336 |
| Infarcts/lacunes | A11.118.637.555.567.562.440.Precursor.Cells..B.Lymphoid | Blood/Immune | -4.32E-08 | 2.39E-08 | 0.964360 |
| Infarcts/lacunes | A11.118.637.555.567.562.B.Lymphocytes | Blood/Immune | -7.05E-09 | 2.45E-08 | 0.612961 |
| Infarcts/lacunes | A11.118.637.555.567.569.200.700.T.Lymphocytes..Regulatory | Blood/Immune | -6.33E-09 | 2.51E-08 | 0.599561 |
| Infarcts/lacunes | A11.118.637.555.567.569.T.Lymphocytes | Blood/Immune | -4.33E-08 | 2.51E-08 | 0.957824 |
| Infarcts/lacunes | A11.118.637.Leukocytes | Blood/Immune | -8.00E-09 | 2.53E-08 | 0.624250 |
| Infarcts/lacunes | A11.329.372.600.Macrophages..Alveolar | Blood/Immune | 9.97E-09 | 2.25E-08 | 0.328834 |
| Infarcts/lacunes | A11.443.Erythroid.Cells | Blood/Immune | -2.63E-08 | 2.33E-08 | 0.871217 |
| Infarcts/lacunes | A11.627.340.360.Granulocyte.Precursor.Cells | Blood/Immune | 9.33E-09 | 2.17E-08 | 0.333404 |
| Infarcts/lacunes | A11.627.624.249.Monocyte.Macrophage.Precursor.Cells | Blood/Immune | -2.99E-08 | 2.36E-08 | 0.897374 |
| Infarcts/lacunes | A11.627.635.Myeloid.Progenitor.Cells | Blood/Immune | -3.24E-08 | 2.18E-08 | 0.931592 |
| Infarcts/lacunes | A11.872.378.590.635.Granulocyte.Macrophage.Progenitor.Cells | Blood/Immune | 7.02E-09 | 2.52E-08 | 0.390323 |
| Infarcts/lacunes | A11.872.378.590.817.Megakaryocyte.Erythroid.Progenitor.Cells | Blood/Immune | -6.00E-08 | 2.48E-08 | 0.992223 |
| Infarcts/lacunes | A11.872.378.Hematopoietic.Stem.Cells | Blood/Immune | -3.49E-08 | 2.14E-08 | 0.948404 |
| Infarcts/lacunes | A15.145.229.188.Blood.Platelets | Blood/Immune | 2.10E-08 | 2.25E-08 | 0.176049 |
| Infarcts/lacunes | A15.145.229.637.555.567.562.725.Plasma.Cells | Blood/Immune | 4.88E-09 | 2.36E-08 | 0.418052 |
| Infarcts/lacunes | A15.145.229.637.555.567.569.200.CD4.Positive.T.Lymphocytes | Blood/Immune | -3.71E-09 | 2.43E-08 | 0.560540 |
| Infarcts/lacunes | A15.145.229.637.555.Leukocytes..Mononuclear | Blood/Immune | -2.93E-08 | 2.56E-08 | 0.874029 |
| Infarcts/lacunes | A15.145.229.Blood.Cells | Blood/Immune | -2.79E-08 | 2.67E-08 | 0.852239 |
| Infarcts/lacunes | A15.145.300.Fetal.Blood | Blood/Immune | 8.06E-09 | 2.72E-08 | 0.383626 |
| Infarcts/lacunes | A15.145.846.Serum | Blood/Immune | 2.24E-08 | 2.26E-08 | 0.161461 |
| Infarcts/lacunes | A15.145.Blood | Blood/Immune | -2.74E-08 | 2.50E-08 | 0.863728 |
| Infarcts/lacunes | A15.378.316.580.Monocytes | Blood/Immune | 5.51E-08 | 2.62E-08 | 0.017959 |
| Infarcts/lacunes | A15.378.316.Bone.Marrow.Cells | Blood/Immune | 5.03E-08 | 2.52E-08 | 0.023044 |
| Infarcts/lacunes | A15.382.490.315.583.Neutrophils | Blood/Immune | 2.19E-08 | 2.26E-08 | 0.166725 |
| Infarcts/lacunes | A15.382.490.555.567.537.Killer.Cells..Natural | Blood/Immune | -3.34E-08 | 2.39E-08 | 0.918935 |
| Infarcts/lacunes | A15.382.490.555.567.622.Lymphocytes..Null | Blood/Immune | -2.61E-08 | 2.48E-08 | 0.853632 |
| Infarcts/lacunes | A15.382.490.555.567.Lymphocytes | Blood/Immune | -3.75E-08 | 2.48E-08 | 0.935044 |
| Infarcts/lacunes | A15.382.520.604.700.Spleen | Blood/Immune | -2.43E-09 | 2.34E-08 | 0.541441 |
| Infarcts/lacunes | A15.382.520.604.800.Palatine.Tonsil | Blood/Immune | -7.00E-08 | 1.94E-08 | 0.999842 |
| Infarcts/lacunes | A15.382.680.Phagocytes | Blood/Immune | 5.47E-08 | 2.45E-08 | 0.012899 |
| Infarcts/lacunes | A15.382.812.260.Dendritic.Cells | Blood/Immune | 5.40E-08 | 2.74E-08 | 0.024283 |
| Infarcts/lacunes | A15.382.812.522.Macrophages | Blood/Immune | 4.20E-08 | 2.29E-08 | 0.033162 |
| Infarcts/lacunes | A15.382.812.Mononuclear.Phagocyte.System | Blood/Immune | 6.36E-08 | 2.61E-08 | 0.007388 |

| | | | | | |
|---|---|---|---|---|---|
| Infarcts/lacunes | A15.382.Immune.System | Blood/Immune | -1.21E-09 | 2.71E-08 | 0.517763 |
| Infarcts/lacunes | Cells_EBV-transformed_lymphocytes | Blood/Immune | 3.82E-09 | 2.19E-08 | 0.430853 |
| Infarcts/lacunes | Spleen | Blood/Immune | 1.57E-08 | 2.34E-08 | 0.251408 |
| Infarcts/lacunes | Whole_Blood | Blood/Immune | -3.50E-09 | 2.31E-08 | 0.560294 |
| Infarcts/lacunes | A07.231.114.Arteries | Cardiovascular | 2.59E-08 | 2.65E-08 | 0.164524 |
| Infarcts/lacunes | A07.231.908.670.874.Umbilical.Veins | Cardiovascular | 6.07E-09 | 2.66E-08 | 0.409719 |
| Infarcts/lacunes | A07.231.908.Veins | Cardiovascular | 1.26E-08 | 2.79E-08 | 0.325338 |
| Infarcts/lacunes | A07.231.Blood.Vessels | Cardiovascular | 3.10E-08 | 2.68E-08 | 0.123806 |
| Infarcts/lacunes | A07.541.358.100.Atrial.Appendage | Cardiovascular | 6.14E-09 | 2.00E-08 | 0.379544 |
| Infarcts/lacunes | A07.541.358.Heart.Atria | Cardiovascular | 1.56E-09 | 2.04E-08 | 0.469459 |
| Infarcts/lacunes | A07.541.510.110.Aortic.Valve | Cardiovascular | 2.29E-08 | 2.00E-08 | 0.126439 |
| Infarcts/lacunes | A07.541.560.Heart.Ventricles | Cardiovascular | -2.00E-08 | 2.18E-08 | 0.820348 |
| Infarcts/lacunes | A07.541.Heart | Cardiovascular | -1.30E-08 | 2.15E-08 | 0.728065 |
| Infarcts/lacunes | Artery_Aorta | Cardiovascular | 1.55E-09 | 1.90E-08 | 0.467508 |
| Infarcts/lacunes | Artery_Coronary | Cardiovascular | -1.69E-08 | 1.92E-08 | 0.809472 |
| Infarcts/lacunes | Artery_Tibial | Cardiovascular | 2.41E-08 | 2.02E-08 | 0.116622 |
| Infarcts/lacunes | Heart_Atrial_Appendage | Cardiovascular | -1.75E-08 | 1.75E-08 | 0.842457 |
| Infarcts/lacunes | Heart_Left_Ventricle | Cardiovascular | -2.77E-08 | 1.77E-08 | 0.941667 |
| Infarcts/lacunes | A08.186.211.132.810.428.200.Cerebellum | CNS | 1.04E-08 | 2.04E-08 | 0.306182 |
| Infarcts/lacunes | A08.186.211.132.Brain.Stem | CNS | 5.11E-09 | 2.00E-08 | 0.399240 |
| Infarcts/lacunes | A08.186.211.464.405.Hippocampus | CNS | -3.23E-08 | 1.91E-08 | 0.954310 |
| Infarcts/lacunes | A08.186.211.464.710.225.Entorhinal.Cortex | CNS | -2.84E-08 | 2.01E-08 | 0.921411 |
| Infarcts/lacunes | A08.186.211.464.Limbic.System | CNS | -2.88E-08 | 1.90E-08 | 0.935549 |
| Infarcts/lacunes | A08.186.211.653.Mesencephalon | CNS | -1.48E-08 | 2.13E-08 | 0.756747 |
| Infarcts/lacunes | A08.186.211.730.317.357.352.435.Hypothalamo.Hypophyseal.System | CNS | -4.93E-08 | 2.33E-08 | 0.982692 |
| Infarcts/lacunes | A08.186.211.730.317.357.Hypothalamus | CNS | -3.16E-08 | 2.32E-08 | 0.913978 |
| Infarcts/lacunes | A08.186.211.730.317.Diencephalon | CNS | -1.47E-08 | 2.20E-08 | 0.747619 |
| Infarcts/lacunes | A08.186.211.730.885.287.249.487.Corpus.Striatum | CNS | -2.31E-09 | 1.98E-08 | 0.546355 |
| Infarcts/lacunes | A08.186.211.730.885.287.249.Basal.Ganglia | CNS | -2.71E-08 | 2.06E-08 | 0.905598 |
| Infarcts/lacunes | A08.186.211.730.885.287.500.270.Frontal.Lobe | CNS | -3.41E-08 | 2.01E-08 | 0.955519 |
| Infarcts/lacunes | A08.186.211.730.885.287.500.571.735.Visual.Cortex | CNS | -9.84E-09 | 1.94E-08 | 0.693712 |
| Infarcts/lacunes | A08.186.211.730.885.287.500.670.Parietal.Lobe | CNS | -4.43E-08 | 1.90E-08 | 0.990119 |
| Infarcts/lacunes | A08.186.211.730.885.287.500.Cerebral.Cortex | CNS | -2.79E-08 | 1.94E-08 | 0.925100 |
| Infarcts/lacunes | A08.186.211.865.428.Metencephalon | CNS | 6.86E-09 | 1.99E-08 | 0.365009 |
| Infarcts/lacunes | A08.186.211.Brain | CNS | -7.50E-09 | 1.92E-08 | 0.651610 |
| Infarcts/lacunes | A09.371.729.Retina | CNS | -1.55E-08 | 1.98E-08 | 0.782911 |
| Infarcts/lacunes | A11.872.653.Neural.Stem.Cells | CNS | 1.39E-08 | 2.34E-08 | 0.275979 |
| Infarcts/lacunes | Brain_Amygdala | CNS | -6.04E-09 | 1.86E-08 | 0.627456 |
| Infarcts/lacunes | Brain_Anterior_cingulate_cortex_(BA24) | CNS | -2.32E-08 | 1.76E-08 | 0.905610 |
| Infarcts/lacunes | Brain_Caudate_(basal_ganglia) | CNS | -3.73E-09 | 1.84E-08 | 0.580587 |
| Infarcts/lacunes | Brain_Cerebellar_Hemisphere | CNS | -3.11E-08 | 1.96E-08 | 0.943907 |
| Infarcts/lacunes | Brain_Cerebellum | CNS | -1.74E-08 | 2.06E-08 | 0.800651 |
| Infarcts/lacunes | Brain_Cortex | CNS | -3.66E-08 | 1.81E-08 | 0.978052 |
| Infarcts/lacunes | Brain_Frontal_Cortex_(BA9) | CNS | -2.65E-08 | 1.70E-08 | 0.939969 |
| Infarcts/lacunes | Brain_Hippocampus | CNS | -1.61E-08 | 1.66E-08 | 0.834388 |
| Infarcts/lacunes | Brain_Hypothalamus | CNS | -2.44E-09 | 1.84E-08 | 0.552588 |

| | | | | | |
|---|---|---|---|---|---|
| Infarcts/lacunes | Brain_Nucleus_accumbens_(basal_ganglia) | CNS | -2.67E-09 | 1.91E-08 | 0.555629 |
| Infarcts/lacunes | Brain_Putamen_(basal_ganglia) | CNS | -1.14E-08 | 1.77E-08 | 0.739983 |
| Infarcts/lacunes | Brain_Spinal_cord_(cervical_c-1) | CNS | -1.75E-08 | 1.79E-08 | 0.836430 |
| Infarcts/lacunes | Brain_Substantia_nigra | CNS | -8.08E-09 | 1.75E-08 | 0.677661 |
| Infarcts/lacunes | A03.556.124.369.Intestinal.Mucosa | Digestive | -1.93E-08 | 2.35E-08 | 0.794830 |
| Infarcts/lacunes | A03.556.124.526.767.Rectum | Digestive | -4.79E-08 | 2.32E-08 | 0.980605 |
| Infarcts/lacunes | A03.556.124.684.Intestine..Small | Digestive | -6.76E-09 | 2.54E-08 | 0.604877 |
| Infarcts/lacunes | A03.556.124.Intestines | Digestive | -2.91E-08 | 2.44E-08 | 0.884076 |
| Infarcts/lacunes | A03.556.249.124.Ileum | Digestive | -8.21E-09 | 2.96E-08 | 0.609346 |
| Infarcts/lacunes | A03.556.249.249.209.Cecum | Digestive | -2.98E-08 | 2.17E-08 | 0.915128 |
| Infarcts/lacunes | A03.556.249.249.356.668.Colon..Sigmoid | Digestive | -3.94E-08 | 2.48E-08 | 0.943552 |
| Infarcts/lacunes | A03.556.249.249.356.Colon | Digestive | -2.63E-08 | 2.48E-08 | 0.855782 |
| Infarcts/lacunes | A03.556.500.760.464.Parotid.Gland | Digestive | 1.71E-08 | 2.47E-08 | 0.244268 |
| Infarcts/lacunes | A03.556.500.760.Salivary.Glands | Digestive | 3.80E-09 | 2.56E-08 | 0.441074 |
| Infarcts/lacunes | A03.556.875.500.Esophagus | Digestive | -5.44E-08 | 2.29E-08 | 0.991171 |
| Infarcts/lacunes | A03.556.875.875.Stomach | Digestive | -1.95E-08 | 2.28E-08 | 0.804351 |
| Infarcts/lacunes | A03.556.875.Upper.Gastrointestinal.Tract | Digestive | -4.38E-08 | 2.42E-08 | 0.964469 |
| Infarcts/lacunes | A03.556.Gastrointestinal.Tract | Digestive | -3.07E-08 | 2.47E-08 | 0.892264 |
| Infarcts/lacunes | Colon_Sigmoid | Digestive | 9.74E-09 | 2.07E-08 | 0.319388 |
| Infarcts/lacunes | Colon_Transverse | Digestive | -1.73E-08 | 2.44E-08 | 0.760572 |
| Infarcts/lacunes | Esophagus_Gastroesophageal_Junction | Digestive | -6.09E-09 | 1.96E-08 | 0.621922 |
| Infarcts/lacunes | Esophagus_Mucosa | Digestive | 1.24E-08 | 2.21E-08 | 0.287637 |
| Infarcts/lacunes | Esophagus_Muscularis | Digestive | -2.72E-09 | 1.88E-08 | 0.557583 |
| Infarcts/lacunes | Small_Intestine_Terminal_Ileum | Digestive | -9.54E-09 | 2.20E-08 | 0.667481 |
| Infarcts/lacunes | Stomach | Digestive | -2.79E-09 | 2.44E-08 | 0.545429 |
| Infarcts/lacunes | A03.620.Liver | Liver | 2.68E-09 | 2.45E-08 | 0.456568 |
| Infarcts/lacunes | A11.436.348.Hepatocytes | Liver | -9.75E-09 | 2.49E-08 | 0.652414 |
| Infarcts/lacunes | Liver | Liver | -1.52E-08 | 1.99E-08 | 0.777020 |
| Infarcts/lacunes | A02.165.Cartilage | Musculoskeletal/Connective | -3.56E-08 | 1.92E-08 | 0.968534 |
| Infarcts/lacunes | A02.633.567.850.Quadriceps.Muscle | Musculoskeletal/Connective | -3.29E-08 | 2.58E-08 | 0.898687 |
| Infarcts/lacunes | A02.835.232.834.151.Cervical.Vertebrae | Musculoskeletal/Connective | -2.25E-08 | 2.14E-08 | 0.854023 |
| Infarcts/lacunes | A02.835.583.443.800.Synovial.Membrane | Musculoskeletal/Connective | 1.79E-08 | 2.09E-08 | 0.196598 |
| Infarcts/lacunes | A05.360.319.679.690.Myometrium | Musculoskeletal/Connective | -9.47E-09 | 2.06E-08 | 0.676919 |
| Infarcts/lacunes | A10.165.450.300.425.Keloid | Musculoskeletal/Connective | 4.14E-09 | 2.06E-08 | 0.420377 |
| Infarcts/lacunes | A10.165.450.300.Cicatrix | Musculoskeletal/Connective | -2.69E-09 | 2.00E-08 | 0.553437 |
| Infarcts/lacunes | A10.690.467.Muscle..Smooth | Musculoskeletal/Connective | 3.59E-09 | 2.62E-08 | 0.445529 |
| Infarcts/lacunes | A10.690.Muscles | Musculoskeletal/Connective | -2.76E-08 | 2.46E-08 | 0.868584 |
| Infarcts/lacunes | A11.329.171.Chondrocytes | Musculoskeletal/Connective | -1.39E-08 | 1.99E-08 | 0.757518 |
| Infarcts/lacunes | A11.329.228.Fibroblasts | Musculoskeletal/Connective | 2.83E-08 | 2.27E-08 | 0.106212 |
| Infarcts/lacunes | A11.329.629.Osteoblasts | Musculoskeletal/Connective | -1.14E-08 | 1.95E-08 | 0.720105 |
| Infarcts/lacunes | A11.329.830.Stromal.Cells | Musculoskeletal/Connective | 6.70E-09 | 2.01E-08 | 0.369394 |
| Infarcts/lacunes | A11.329.Connective.Tissue.Cells | Musculoskeletal/Connective | 4.75E-08 | 2.18E-08 | 0.014574 |
| Infarcts/lacunes | A11.620.520.Myocytes..Smooth.Muscle | Musculoskeletal/Connective | 1.29E-08 | 2.47E-08 | 0.300108 |
| Infarcts/lacunes | Cells_Transformed_fibroblasts | Musculoskeletal/Connective | 3.69E-09 | 2.08E-08 | 0.429602 |
| Infarcts/lacunes | Muscle_Skeletal | Musculoskeletal/Connective | -1.46E-08 | 2.11E-08 | 0.755609 |
| Infarcts/lacunes | A04.411.Lung | Other | -6.44E-09 | 2.69E-08 | 0.594589 |

| | | | | | |
|---|---|---|---|---|---|
| Infarcts/lacunes | A04.531.520.Nasal.Mucosa | Other | -2.27E-08 | 2.31E-08 | 0.837059 |
| Infarcts/lacunes | A05.360.319.114.373.Fallopian.Tubes | Other | -2.97E-08 | 2.32E-08 | 0.899840 |
| Infarcts/lacunes | A05.360.319.114.630.Ovary | Other | -3.93E-08 | 2.15E-08 | 0.965841 |
| Infarcts/lacunes | A05.360.319.679.256.Cervix.Uteri | Other | -3.03E-08 | 2.29E-08 | 0.907108 |
| Infarcts/lacunes | A05.360.319.679.490.Endometrium | Other | 3.70E-09 | 2.18E-08 | 0.432784 |
| Infarcts/lacunes | A05.360.319.679.Uterus | Other | 1.12E-08 | 1.91E-08 | 0.278505 |
| Infarcts/lacunes | A05.360.319.887.Vulva | Other | -2.04E-08 | 2.40E-08 | 0.802236 |
| Infarcts/lacunes | A05.360.319.Genitalia..Female | Other | -2.96E-08 | 2.17E-08 | 0.913654 |
| Infarcts/lacunes | A05.360.444.492.362.Foreskin | Other | -5.93E-09 | 2.17E-08 | 0.607798 |
| Infarcts/lacunes | A05.360.444.Genitalia..Male | Other | -3.10E-08 | 2.21E-08 | 0.919646 |
| Infarcts/lacunes | A05.360.490.Germ.Cells | Other | 8.12E-10 | 2.30E-08 | 0.485950 |
| Infarcts/lacunes | A05.360.Genitalia | Other | -3.08E-08 | 2.16E-08 | 0.922813 |
| Infarcts/lacunes | A05.810.453.324.Kidney.Cortex | Other | -1.04E-08 | 2.33E-08 | 0.672272 |
| Infarcts/lacunes | A05.810.453.Kidney | Other | 2.34E-08 | 2.00E-08 | 0.121524 |
| Infarcts/lacunes | A05.810.890.Urinary.Bladder | Other | 1.87E-08 | 2.57E-08 | 0.233384 |
| Infarcts/lacunes | A06.407.071.140.Adrenal.Cortex | Other | -1.32E-08 | 2.07E-08 | 0.737669 |
| Infarcts/lacunes | A06.407.071.Adrenal.Glands | Other | -1.27E-08 | 2.17E-08 | 0.720571 |
| Infarcts/lacunes | A06.407.312.782.Testis | Other | 2.98E-09 | 1.92E-08 | 0.438121 |
| Infarcts/lacunes | A06.407.312.Gonads | Other | -2.96E-08 | 2.13E-08 | 0.917147 |
| Infarcts/lacunes | A06.407.900.Thyroid.Gland | Other | 2.88E-08 | 2.02E-08 | 0.077281 |
| Infarcts/lacunes | A06.407.Endocrine.Glands | Other | -3.21E-08 | 2.17E-08 | 0.930707 |
| Infarcts/lacunes | A09.371.Eye | Other | 5.65E-09 | 2.23E-08 | 0.400105 |
| Infarcts/lacunes | A10.272.497.Epidermis | Other | -6.21E-09 | 2.46E-08 | 0.599511 |
| Infarcts/lacunes | A10.272.Epithelium | Other | -3.63E-08 | 2.22E-08 | 0.949145 |
| Infarcts/lacunes | A10.336.707.Prostate | Other | -3.34E-08 | 2.08E-08 | 0.945853 |
| Infarcts/lacunes | A10.615.284.473.Chorion | Other | 1.74E-09 | 2.37E-08 | 0.470680 |
| Infarcts/lacunes | A10.615.550.599.Mouth.Mucosa | Other | -5.10E-08 | 2.22E-08 | 0.989276 |
| Infarcts/lacunes | A10.615.550.Mucous.Membrane | Other | -2.04E-08 | 2.45E-08 | 0.796900 |
| Infarcts/lacunes | A10.615.789.Serous.Membrane | Other | 4.53E-09 | 2.25E-08 | 0.420038 |
| Infarcts/lacunes | A10.615.Membranes | Other | -8.20E-09 | 2.38E-08 | 0.634737 |
| Infarcts/lacunes | A11.382.Endocrine.Cells | Other | -1.32E-09 | 2.12E-08 | 0.524777 |
| Infarcts/lacunes | A11.436.275.Endothelial.Cells | Other | 6.99E-09 | 2.54E-08 | 0.391469 |
| Infarcts/lacunes | A11.436.294.064.Glucagon.Secreting.Cells | Other | -3.88E-08 | 2.13E-08 | 0.965828 |
| Infarcts/lacunes | A11.436.329.Granulosa.Cells | Other | -8.23E-09 | 1.92E-08 | 0.665467 |
| Infarcts/lacunes | A11.436.397.Keratinocytes | Other | -1.91E-08 | 2.26E-08 | 0.800800 |
| Infarcts/lacunes | A11.436.Epithelial.Cells | Other | -6.30E-08 | 2.29E-08 | 0.997070 |
| Infarcts/lacunes | A11.497.497.600.Oocytes | Other | -2.32E-09 | 2.29E-08 | 0.540438 |
| Infarcts/lacunes | A11.872.040.Adult.Stem.Cells | Other | -4.73E-08 | 2.22E-08 | 0.983437 |
| Infarcts/lacunes | A11.872.190.260.Embryoid.Bodies | Other | -1.23E-08 | 2.30E-08 | 0.703011 |
| Infarcts/lacunes | A11.872.190.Embryonic.Stem.Cells | Other | -2.93E-08 | 2.05E-08 | 0.923795 |
| Infarcts/lacunes | A11.872.580.Mesenchymal.Stem.Cells | Other | 4.09E-08 | 2.29E-08 | 0.036813 |
| Infarcts/lacunes | A11.872.700.500.Induced.Pluripotent.Stem.Cells | Other | -6.83E-08 | 2.07E-08 | 0.999519 |
| Infarcts/lacunes | A11.872.Stem.Cells | Other | -2.01E-08 | 2.33E-08 | 0.805254 |
| Infarcts/lacunes | A14.549.167.646.Periodontium | Other | -8.00E-09 | 2.84E-08 | 0.610820 |
| Infarcts/lacunes | A14.549.167.Dentition | Other | 5.20E-09 | 2.52E-08 | 0.418447 |
| Infarcts/lacunes | A14.549.885.Tongue | Other | 3.07E-08 | 2.53E-08 | 0.112140 |

| | | | | | |
|---|---|---|---|---|---|
| Infarcts/lacunes | A14.549.Mouth | Other | -1.30E-08 | 2.38E-08 | 0.707385 |
| Infarcts/lacunes | A14.724.557.Nasopharynx | Other | 1.28E-08 | 2.28E-08 | 0.286533 |
| Infarcts/lacunes | A14.724.Pharynx | Other | -8.95E-09 | 2.33E-08 | 0.649769 |
| Infarcts/lacunes | A17.815.Skin | Other | -4.97E-08 | 2.11E-08 | 0.990736 |
| Infarcts/lacunes | Adrenal_Gland | Other | -5.30E-09 | 1.91E-08 | 0.609532 |
| Infarcts/lacunes | Bladder | Other | 5.48E-09 | 2.01E-08 | 0.392327 |
| Infarcts/lacunes | Breast_Mammary_Tissue | Other | -2.01E-08 | 1.94E-08 | 0.849982 |
| Infarcts/lacunes | Cervix_Ectocervix | Other | -1.84E-08 | 2.11E-08 | 0.807698 |
| Infarcts/lacunes | Cervix_Endocervix | Other | 4.67E-09 | 2.13E-08 | 0.413126 |
| Infarcts/lacunes | Fallopian_Tube | Other | -9.83E-10 | 2.23E-08 | 0.517616 |
| Infarcts/lacunes | Kidney_Cortex | Other | -3.25E-08 | 1.93E-08 | 0.954201 |
| Infarcts/lacunes | Lung | Other | 1.12E-08 | 2.30E-08 | 0.313160 |
| Infarcts/lacunes | Minor_Salivary_Gland | Other | -5.96E-09 | 2.07E-08 | 0.613082 |
| Infarcts/lacunes | Nerve_Tibial | Other | -6.83E-09 | 1.83E-08 | 0.645420 |
| Infarcts/lacunes | Ovary | Other | -3.67E-08 | 1.99E-08 | 0.967563 |
| Infarcts/lacunes | Pituitary | Other | 5.16E-09 | 1.98E-08 | 0.396947 |
| Infarcts/lacunes | Prostate | Other | -2.15E-09 | 2.22E-08 | 0.538618 |
| Infarcts/lacunes | Skin_Not_Sun_Exposed_(Suprapubic) | Other | -3.09E-08 | 2.26E-08 | 0.914537 |
| Infarcts/lacunes | Skin_Sun_Exposed_(Lower_leg) | Other | -3.57E-08 | 2.23E-08 | 0.945049 |
| Infarcts/lacunes | Testis | Other | -3.38E-08 | 1.99E-08 | 0.955356 |
| Infarcts/lacunes | Thyroid | Other | -1.12E-09 | 2.02E-08 | 0.522080 |
| Infarcts/lacunes | Uterus | Other | -2.63E-08 | 1.79E-08 | 0.928857 |
| Infarcts/lacunes | Vagina | Other | -3.16E-08 | 1.94E-08 | 0.948882 |
| Infarcts/lacunes | A03.734.414.Islets.of.Langerhans | Pancreas | -1.66E-08 | 2.26E-08 | 0.768878 |
| Infarcts/lacunes | A03.734.Pancreas | Pancreas | -1.82E-08 | 1.99E-08 | 0.819816 |
| Infarcts/lacunes | Pancreas | Pancreas | -8.96E-09 | 2.00E-08 | 0.673247 |
| PD Braak | A10.165.114.830.500.750.Subcutaneous.Fat..Abdominal | Adipose | -2.32E-08 | 1.64E-08 | 0.922086 |
| PD Braak | A10.165.114.830.750.Subcutaneous.Fat | Adipose | -2.38E-08 | 1.67E-08 | 0.922674 |
| PD Braak | A11.329.114.Adipocytes | Adipose | -9.85E-09 | 1.90E-08 | 0.697832 |
| PD Braak | Adipose_Subcutaneous | Adipose | 4.68E-09 | 1.72E-08 | 0.393042 |
| PD Braak | Adipose_Visceral_(Omentum) | Adipose | 2.57E-08 | 1.81E-08 | 0.078138 |
| PD Braak | A02.835.583.443.800.800.Synovial.Fluid | Blood/Immune | -1.04E-08 | 2.42E-08 | 0.667136 |
| PD Braak | A10.549.400.Lymph.Nodes | Blood/Immune | -4.25E-08 | 2.03E-08 | 0.981960 |
| PD Braak | A10.549.Lymphoid.Tissue | Blood/Immune | -5.17E-08 | 1.78E-08 | 0.998155 |
| PD Braak | A11.118.637.555.567.562.440.Precursor.Cells..B.Lymphoid | Blood/Immune | -1.37E-08 | 1.93E-08 | 0.759961 |
| PD Braak | A11.118.637.555.567.562.B.Lymphocytes | Blood/Immune | -1.73E-08 | 1.98E-08 | 0.809284 |
| PD Braak | A11.118.637.555.567.569.200.700.T.Lymphocytes..Regulatory | Blood/Immune | -2.16E-08 | 2.10E-08 | 0.847543 |
| PD Braak | A11.118.637.555.567.569.T.Lymphocytes | Blood/Immune | -2.54E-08 | 2.08E-08 | 0.888819 |
| PD Braak | A11.118.637.Leukocytes | Blood/Immune | -3.40E-08 | 2.41E-08 | 0.920777 |
| PD Braak | A11.329.372.600.Macrophages..Alveolar | Blood/Immune | -2.84E-09 | 2.04E-08 | 0.555336 |
| PD Braak | A11.443.Erythroid.Cells | Blood/Immune | 1.21E-08 | 2.08E-08 | 0.280399 |
| PD Braak | A11.627.340.360.Granulocyte.Precursor.Cells | Blood/Immune | -1.17E-08 | 2.29E-08 | 0.695055 |
| PD Braak | A11.627.624.249.Monocyte.Macrophage.Precursor.Cells | Blood/Immune | 1.06E-08 | 2.17E-08 | 0.312692 |
| PD Braak | A11.627.635.Myeloid.Progenitor.Cells | Blood/Immune | 1.72E-08 | 2.31E-08 | 0.228223 |
| PD Braak | A11.872.378.590.635.Granulocyte.Macrophage.Progenitor.Cells | Blood/Immune | 1.74E-08 | 2.36E-08 | 0.230425 |
| PD Braak | A11.872.378.590.817.Megakaryocyte.Erythroid.Progenitor.Cells | Blood/Immune | -2.02E-08 | 2.00E-08 | 0.843284 |

| | | | | | |
|---|---|---|---|---|---|
| PD Braak | A11.872.378.Hematopoietic.Stem.Cells | Blood/Immune | 3.54E-09 | 2.31E-08 | 0.439000 |
| PD Braak | A15.145.229.188.Blood.Platelets | Blood/Immune | -2.17E-09 | 2.26E-08 | 0.538294 |
| PD Braak | A15.145.229.637.555.567.562.725.Plasma.Cells | Blood/Immune | -1.19E-08 | 2.33E-08 | 0.695412 |
| PD Braak | A15.145.229.637.555.567.569.200.CD4.Positive.T.Lymphocytes | Blood/Immune | -3.19E-08 | 2.09E-08 | 0.936385 |
| PD Braak | A15.145.229.637.555.Leukocytes..Mononuclear | Blood/Immune | -6.05E-08 | 2.28E-08 | 0.996035 |
| PD Braak | A15.145.229.Blood.Cells | Blood/Immune | -3.00E-08 | 2.52E-08 | 0.883593 |
| PD Braak | A15.145.300.Fetal.Blood | Blood/Immune | -2.46E-08 | 2.80E-08 | 0.810445 |
| PD Braak | A15.145.846.Serum | Blood/Immune | -1.36E-08 | 2.21E-08 | 0.729964 |
| PD Braak | A15.145.Blood | Blood/Immune | -4.41E-08 | 2.37E-08 | 0.968359 |
| PD Braak | A15.378.316.580.Monocytes | Blood/Immune | 1.06E-08 | 2.36E-08 | 0.327016 |
| PD Braak | A15.378.316.Bone.Marrow.Cells | Blood/Immune | 3.21E-08 | 2.45E-08 | 0.095113 |
| PD Braak | A15.382.490.315.583.Neutrophils | Blood/Immune | 4.55E-08 | 2.34E-08 | 0.025912 |
| PD Braak | A15.382.490.555.567.537.Killer.Cells..Natural | Blood/Immune | -3.03E-08 | 1.87E-08 | 0.947273 |
| PD Braak | A15.382.490.555.567.622.Lymphocytes..Null | Blood/Immune | -4.25E-08 | 2.22E-08 | 0.972546 |
| PD Braak | A15.382.490.555.567.Lymphocytes | Blood/Immune | -3.29E-08 | 2.06E-08 | 0.944816 |
| PD Braak | A15.382.520.604.700.Spleen | Blood/Immune | -3.10E-08 | 1.89E-08 | 0.949621 |
| PD Braak | A15.382.520.604.800.Palatine.Tonsil | Blood/Immune | 7.45E-09 | 1.91E-08 | 0.348177 |
| PD Braak | A15.382.680.Phagocytes | Blood/Immune | 8.79E-10 | 2.30E-08 | 0.484769 |
| PD Braak | A15.382.812.260.Dendritic.Cells | Blood/Immune | -5.55E-09 | 2.15E-08 | 0.601772 |
| PD Braak | A15.382.812.522.Macrophages | Blood/Immune | -1.12E-08 | 2.22E-08 | 0.692781 |
| PD Braak | A15.382.812.Mononuclear.Phagocyte.System | Blood/Immune | -1.86E-08 | 2.17E-08 | 0.804487 |
| PD Braak | A15.382.Immune.System | Blood/Immune | -3.73E-08 | 2.16E-08 | 0.957830 |
| PD Braak | Cells_EBV-transformed_lymphocytes | Blood/Immune | -4.15E-08 | 2.15E-08 | 0.973343 |
| PD Braak | Spleen | Blood/Immune | -5.10E-08 | 2.05E-08 | 0.993468 |
| PD Braak | Whole_Blood | Blood/Immune | 8.39E-09 | 2.26E-08 | 0.355510 |
| PD Braak | A07.231.114.Arteries | Cardiovascular | -1.36E-08 | 2.13E-08 | 0.738498 |
| PD Braak | A07.231.908.670.874.Umbilical.Veins | Cardiovascular | -1.65E-08 | 2.40E-08 | 0.753640 |
| PD Braak | A07.231.908.Veins | Cardiovascular | -1.43E-08 | 2.44E-08 | 0.721573 |
| PD Braak | A07.231.Blood.Vessels | Cardiovascular | -3.47E-08 | 2.76E-08 | 0.895909 |
| PD Braak | A07.541.358.100.Atrial.Appendage | Cardiovascular | -6.13E-09 | 1.73E-08 | 0.638112 |
| PD Braak | A07.541.358.Heart.Atria | Cardiovascular | -1.34E-08 | 1.75E-08 | 0.778906 |
| PD Braak | A07.541.510.110.Aortic.Valve | Cardiovascular | -1.19E-08 | 2.01E-08 | 0.723418 |
| PD Braak | A07.541.560.Heart.Ventricles | Cardiovascular | -4.25E-08 | 1.75E-08 | 0.992449 |
| PD Braak | A07.541.Heart | Cardiovascular | -4.71E-08 | 1.75E-08 | 0.996531 |
| PD Braak | Artery_Aorta | Cardiovascular | -2.89E-08 | 1.64E-08 | 0.961198 |
| PD Braak | Artery_Coronary | Cardiovascular | -1.57E-08 | 1.74E-08 | 0.816695 |
| PD Braak | Artery_Tibial | Cardiovascular | -2.78E-08 | 1.57E-08 | 0.962146 |
| PD Braak | Heart_Atrial_Appendage | Cardiovascular | -2.37E-08 | 1.53E-08 | 0.938770 |
| PD Braak | Heart_Left_Ventricle | Cardiovascular | -1.94E-08 | 1.74E-08 | 0.867593 |
| PD Braak | A08.186.211.132.810.428.200.Cerebellum | CNS | 1.58E-09 | 2.01E-08 | 0.468729 |
| PD Braak | A08.186.211.132.Brain.Stem | CNS | 2.55E-08 | 1.99E-08 | 0.100138 |
| PD Braak | A08.186.211.464.405.Hippocampus | CNS | 4.82E-08 | 1.85E-08 | 0.004666 |
| PD Braak | A08.186.211.464.710.225.Entorhinal.Cortex | CNS | 4.10E-08 | 1.81E-08 | 0.011653 |
| PD Braak | A08.186.211.464.Limbic.System | CNS | 4.46E-08 | 1.86E-08 | 0.008282 |
| PD Braak | A08.186.211.653.Mesencephalon | CNS | 2.60E-08 | 2.03E-08 | 0.100444 |
| PD Braak | A08.186.211.730.317.357.352.435.Hypothalamo.Hypophyseal.System | CNS | 2.73E-09 | 2.13E-08 | 0.449037 |

| | | | | | |
|---|---|---|---|---|---|
| PD Braak | A08.186.211.730.317.357.Hypothalamus | CNS | 1.62E-08 | 2.09E-08 | 0.219143 |
| PD Braak | A08.186.211.730.317.Diencephalon | CNS | 3.06E-08 | 2.30E-08 | 0.091675 |
| PD Braak | A08.186.211.730.885.287.249.487.Corpus.Striatum | CNS | 1.09E-08 | 1.77E-08 | 0.270204 |
| PD Braak | A08.186.211.730.885.287.249.Basal.Ganglia | CNS | 4.50E-08 | 1.88E-08 | 0.008491 |
| PD Braak | A08.186.211.730.885.287.500.270.Frontal.Lobe | CNS | 3.51E-08 | 2.05E-08 | 0.043297 |
| PD Braak | A08.186.211.730.885.287.500.571.735.Visual.Cortex | CNS | 2.11E-08 | 1.80E-08 | 0.120907 |
| PD Braak | A08.186.211.730.885.287.500.670.Parietal.Lobe | CNS | 2.90E-08 | 2.08E-08 | 0.081156 |
| PD Braak | A08.186.211.730.885.287.500.Cerebral.Cortex | CNS | 4.66E-08 | 1.92E-08 | 0.007498 |
| PD Braak | A08.186.211.865.428.Metencephalon | CNS | 7.80E-09 | 1.99E-08 | 0.347780 |
| PD Braak | A08.186.211.Brain | CNS | 4.03E-08 | 1.80E-08 | 0.012547 |
| PD Braak | A09.371.729.Retina | CNS | 1.24E-08 | 1.86E-08 | 0.252415 |
| PD Braak | A11.872.653.Neural.Stem.Cells | CNS | 1.61E-08 | 2.01E-08 | 0.211692 |
| PD Braak | Brain_Amygdala | CNS | 4.75E-08 | 1.64E-08 | 0.001936 |
| PD Braak | Brain_Anterior_cingulate_cortex_(BA24) | CNS | 5.03E-08 | 1.64E-08 | 0.001053 |
| PD Braak | Brain_Caudate_(basal_ganglia) | CNS | 5.69E-08 | 1.64E-08 | 0.000249 |
| PD Braak | Brain_Cerebellar_Hemisphere | CNS | 2.05E-08 | 1.78E-08 | 0.123913 |
| PD Braak | Brain_Cerebellum | CNS | 2.55E-08 | 1.75E-08 | 0.072266 |
| PD Braak | Brain_Cortex | CNS | 3.57E-08 | 1.73E-08 | 0.019740 |
| PD Braak | Brain_Frontal_Cortex_(BA9) | CNS | 3.35E-08 | 1.59E-08 | 0.017462 |
| PD Braak | Brain_Hippocampus | CNS | 4.48E-08 | 1.58E-08 | 0.002273 |
| PD Braak | Brain_Hypothalamus | CNS | 4.65E-08 | 1.59E-08 | 0.001752 |
| PD Braak | Brain_Nucleus_accumbens_(basal_ganglia) | CNS | 3.53E-08 | 1.59E-08 | 0.013077 |
| PD Braak | Brain_Putamen_(basal_ganglia) | CNS | 4.53E-08 | 1.67E-08 | 0.003258 |
| PD Braak | Brain_Spinal_cord_(cervical_c-1) | CNS | 4.95E-08 | 1.61E-08 | 0.001052 |
| PD Braak | Brain_Substantia_nigra | CNS | 4.76E-08 | 1.55E-08 | 0.001063 |
| PD Braak | A03.556.124.369.Intestinal.Mucosa | Digestive | 3.30E-08 | 2.18E-08 | 0.065063 |
| PD Braak | A03.556.124.526.767.Rectum | Digestive | 3.53E-08 | 2.06E-08 | 0.042954 |
| PD Braak | A03.556.124.684.Intestine..Small | Digestive | -1.78E-09 | 1.93E-08 | 0.536737 |
| PD Braak | A03.556.124.Intestines | Digestive | 3.74E-08 | 1.92E-08 | 0.025871 |
| PD Braak | A03.556.249.124.Ileum | Digestive | 1.27E-08 | 2.07E-08 | 0.270258 |
| PD Braak | A03.556.249.249.209.Cecum | Digestive | 1.34E-08 | 1.99E-08 | 0.251281 |
| PD Braak | A03.556.249.249.356.668.Colon..Sigmoid | Digestive | 8.46E-09 | 2.11E-08 | 0.344619 |
| PD Braak | A03.556.249.249.356.Colon | Digestive | 3.44E-08 | 1.91E-08 | 0.035671 |
| PD Braak | A03.556.500.760.464.Parotid.Gland | Digestive | 2.11E-08 | 2.27E-08 | 0.176284 |
| PD Braak | A03.556.500.760.Salivary.Glands | Digestive | 2.44E-08 | 2.22E-08 | 0.136251 |
| PD Braak | A03.556.875.500.Esophagus | Digestive | 5.22E-08 | 2.03E-08 | 0.005019 |
| PD Braak | A03.556.875.875.Stomach | Digestive | -2.93E-09 | 1.96E-08 | 0.559382 |
| PD Braak | A03.556.875.Upper.Gastrointestinal.Tract | Digestive | 1.71E-09 | 1.90E-08 | 0.464196 |
| PD Braak | A03.556.Gastrointestinal.Tract | Digestive | 3.52E-08 | 2.06E-08 | 0.043527 |
| PD Braak | Colon_Sigmoid | Digestive | -6.38E-09 | 1.73E-08 | 0.643957 |
| PD Braak | Colon_Transverse | Digestive | -6.71E-10 | 2.00E-08 | 0.513356 |
| PD Braak | Esophagus_Gastroesophageal_Junction | Digestive | -4.51E-08 | 1.74E-08 | 0.995307 |
| PD Braak | Esophagus_Mucosa | Digestive | 1.79E-08 | 1.82E-08 | 0.163563 |
| PD Braak | Esophagus_Muscularis | Digestive | -4.38E-08 | 1.70E-08 | 0.995001 |
| PD Braak | Small_Intestine_Terminal_Ileum | Digestive | 1.93E-08 | 1.93E-08 | 0.158252 |
| PD Braak | Stomach | Digestive | 2.18E-08 | 1.92E-08 | 0.128199 |

| | | | | | |
|---|---|---|---|---|---|
| PD Braak | A03.620.Liver | Liver | -1.40E-08 | 2.19E-08 | 0.738598 |
| PD Braak | A11.436.348.Hepatocytes | Liver | 1.42E-09 | 2.25E-08 | 0.474898 |
| PD Braak | Liver | Liver | -3.86E-09 | 1.91E-08 | 0.580225 |
| PD Braak | A02.165.Cartilage | Musculoskeletal/Connective | -1.09E-09 | 1.95E-08 | 0.522369 |
| PD Braak | A02.633.567.850.Quadriceps.Muscle | Musculoskeletal/Connective | -3.04E-08 | 1.93E-08 | 0.942679 |
| PD Braak | A02.835.232.834.151.Cervical.Vertebrae | Musculoskeletal/Connective | -1.85E-08 | 1.95E-08 | 0.828281 |
| PD Braak | A02.835.583.443.800.Synovial.Membrane | Musculoskeletal/Connective | -4.70E-09 | 1.81E-08 | 0.602661 |
| PD Braak | A05.360.319.679.690.Myometrium | Musculoskeletal/Connective | 1.49E-08 | 1.97E-08 | 0.223800 |
| PD Braak | A10.165.450.300.425.Keloid | Musculoskeletal/Connective | -1.08E-08 | 1.76E-08 | 0.730243 |
| PD Braak | A10.165.450.300.Cicatrix | Musculoskeletal/Connective | -1.76E-08 | 1.90E-08 | 0.822801 |
| PD Braak | A10.690.467.Muscle..Smooth | Musculoskeletal/Connective | -1.39E-08 | 1.76E-08 | 0.784943 |
| PD Braak | A10.690.Muscles | Musculoskeletal/Connective | -1.45E-08 | 2.01E-08 | 0.764406 |
| PD Braak | A11.329.171.Chondrocytes | Musculoskeletal/Connective | -3.24E-08 | 1.87E-08 | 0.958301 |
| PD Braak | A11.329.228.Fibroblasts | Musculoskeletal/Connective | -3.47E-08 | 1.82E-08 | 0.972075 |
| PD Braak | A11.329.629.Osteoblasts | Musculoskeletal/Connective | -3.08E-08 | 1.93E-08 | 0.944625 |
| PD Braak | A11.329.830.Stromal.Cells | Musculoskeletal/Connective | -3.20E-08 | 1.77E-08 | 0.965110 |
| PD Braak | A11.329.Connective.Tissue.Cells | Musculoskeletal/Connective | -1.51E-08 | 1.95E-08 | 0.780392 |
| PD Braak | A11.620.520.Myocytes..Smooth.Muscle | Musculoskeletal/Connective | -2.43E-08 | 1.84E-08 | 0.906091 |
| PD Braak | Cells_Transformed_fibroblasts | Musculoskeletal/Connective | -1.89E-09 | 1.89E-08 | 0.539724 |
| PD Braak | Muscle_Skeletal | Musculoskeletal/Connective | -1.39E-08 | 1.85E-08 | 0.774413 |
| PD Braak | A04.411.Lung | Other | -2.75E-08 | 2.09E-08 | 0.905659 |
| PD Braak | A04.531.520.Nasal.Mucosa | Other | -7.50E-09 | 1.88E-08 | 0.655385 |
| PD Braak | A05.360.319.114.373.Fallopian.Tubes | Other | 3.43E-09 | 2.15E-08 | 0.436495 |
| PD Braak | A05.360.319.114.630.Ovary | Other | 3.74E-09 | 2.11E-08 | 0.429786 |
| PD Braak | A05.360.319.679.256.Cervix.Uteri | Other | -1.40E-08 | 1.86E-08 | 0.774124 |
| PD Braak | A05.360.319.679.490.Endometrium | Other | 7.76E-09 | 1.96E-08 | 0.345904 |
| PD Braak | A05.360.319.679.Uterus | Other | 2.45E-08 | 1.75E-08 | 0.080378 |
| PD Braak | A05.360.319.887.Vulva | Other | 3.95E-08 | 2.33E-08 | 0.045201 |
| PD Braak | A05.360.319.Genitalia..Female | Other | 7.12E-09 | 1.96E-08 | 0.358462 |
| PD Braak | A05.360.444.492.362.Foreskin | Other | -1.07E-08 | 1.67E-08 | 0.739947 |
| PD Braak | A05.360.444.Genitalia..Male | Other | 3.99E-09 | 1.83E-08 | 0.413957 |
| PD Braak | A05.360.490.Germ.Cells | Other | -2.84E-08 | 1.80E-08 | 0.942900 |
| PD Braak | A05.360.Genitalia | Other | 1.43E-08 | 2.05E-08 | 0.242776 |
| PD Braak | A05.810.453.324.Kidney.Cortex | Other | -1.42E-08 | 1.86E-08 | 0.778313 |
| PD Braak | A05.810.453.Kidney | Other | -1.07E-08 | 1.73E-08 | 0.731817 |
| PD Braak | A05.810.890.Urinary.Bladder | Other | 1.42E-08 | 1.97E-08 | 0.234630 |
| PD Braak | A06.407.071.140.Adrenal.Cortex | Other | -1.92E-08 | 1.98E-08 | 0.833862 |
| PD Braak | A06.407.071.Adrenal.Glands | Other | -1.30E-08 | 1.94E-08 | 0.748029 |
| PD Braak | A06.407.312.782.Testis | Other | -2.75E-08 | 1.68E-08 | 0.949662 |
| PD Braak | A06.407.312.Gonads | Other | 3.19E-09 | 2.08E-08 | 0.439166 |
| PD Braak | A06.407.900.Thyroid.Gland | Other | 1.75E-08 | 1.94E-08 | 0.183496 |
| PD Braak | A06.407.Endocrine.Glands | Other | 1.87E-08 | 2.18E-08 | 0.195728 |
| PD Braak | A09.371.Eye | Other | -1.74E-08 | 1.96E-08 | 0.812272 |
| PD Braak | A10.272.497.Epidermis | Other | -2.22E-08 | 2.07E-08 | 0.858891 |
| PD Braak | A10.272.Epithelium | Other | -7.70E-09 | 1.93E-08 | 0.655435 |
| PD Braak | A10.336.707.Prostate | Other | 1.44E-08 | 1.98E-08 | 0.232585 |

| | | | | | |
|---|---|---|---|---|---|
| PD Braak | A10.615.284.473.Chorion | Other | -3.68E-08 | 1.79E-08 | 0.980043 |
| PD Braak | A10.615.550.599.Mouth.Mucosa | Other | 2.92E-08 | 1.94E-08 | 0.066843 |
| PD Braak | A10.615.550.Mucous.Membrane | Other | -1.63E-09 | 2.05E-08 | 0.531803 |
| PD Braak | A10.615.789.Serous.Membrane | Other | -3.93E-08 | 2.22E-08 | 0.961579 |
| PD Braak | A10.615.Membranes | Other | 6.49E-10 | 1.96E-08 | 0.486798 |
| PD Braak | A11.382.Endocrine.Cells | Other | -2.94E-08 | 1.86E-08 | 0.942559 |
| PD Braak | A11.436.275.Endothelial.Cells | Other | -1.92E-08 | 2.61E-08 | 0.768366 |
| PD Braak | A11.436.294.064.Glucagon.Secreting.Cells | Other | -1.52E-08 | 2.01E-08 | 0.775431 |
| PD Braak | A11.436.329.Granulosa.Cells | Other | -3.27E-08 | 1.81E-08 | 0.964892 |
| PD Braak | A11.436.397.Keratinocytes | Other | 5.14E-09 | 1.91E-08 | 0.393980 |
| PD Braak | A11.436.Epithelial.Cells | Other | 5.49E-09 | 1.98E-08 | 0.390741 |
| PD Braak | A11.497.497.600.Oocytes | Other | -2.26E-08 | 1.79E-08 | 0.896601 |
| PD Braak | A11.872.040.Adult.Stem.Cells | Other | -7.73E-09 | 1.79E-08 | 0.666774 |
| PD Braak | A11.872.190.260.Embryoid.Bodies | Other | 5.41E-09 | 1.89E-08 | 0.387561 |
| PD Braak | A11.872.190.Embryonic.Stem.Cells | Other | -3.87E-09 | 1.84E-08 | 0.583010 |
| PD Braak | A11.872.580.Mesenchymal.Stem.Cells | Other | -5.02E-08 | 1.71E-08 | 0.998317 |
| PD Braak | A11.872.700.500.Induced.Pluripotent.Stem.Cells | Other | -7.83E-09 | 1.78E-08 | 0.670418 |
| PD Braak | A11.872.Stem.Cells | Other | -3.42E-08 | 2.11E-08 | 0.947460 |
| PD Braak | A14.549.167.646.Periodontium | Other | 1.94E-08 | 2.28E-08 | 0.196831 |
| PD Braak | A14.549.167.Dentition | Other | 7.01E-09 | 2.24E-08 | 0.376969 |
| PD Braak | A14.549.885.Tongue | Other | -4.29E-09 | 2.18E-08 | 0.577969 |
| PD Braak | A14.549.Mouth | Other | 3.08E-08 | 2.07E-08 | 0.068551 |
| PD Braak | A14.724.557.Nasopharynx | Other | -7.41E-09 | 1.97E-08 | 0.646753 |
| PD Braak | A14.724.Pharynx | Other | 4.20E-09 | 1.97E-08 | 0.415787 |
| PD Braak | A17.815.Skin | Other | 1.54E-08 | 2.09E-08 | 0.231238 |
| PD Braak | Adrenal_Gland | Other | -2.14E-08 | 1.70E-08 | 0.895458 |
| PD Braak | Bladder | Other | -6.70E-09 | 1.66E-08 | 0.656468 |
| PD Braak | Breast_Mammary_Tissue | Other | 3.23E-08 | 1.71E-08 | 0.029394 |
| PD Braak | Cervix_Ectocervix | Other | 1.30E-08 | 1.80E-08 | 0.234623 |
| PD Braak | Cervix_Endocervix | Other | 9.48E-09 | 1.78E-08 | 0.297257 |
| PD Braak | Fallopian_Tube | Other | 9.42E-09 | 1.91E-08 | 0.310906 |
| PD Braak | Kidney_Cortex | Other | -1.41E-08 | 1.79E-08 | 0.784624 |
| PD Braak | Lung | Other | -2.53E-08 | 1.88E-08 | 0.910386 |
| PD Braak | Minor_Salivary_Gland | Other | 1.59E-08 | 1.73E-08 | 0.179033 |
| PD Braak | Nerve_Tibial | Other | -1.73E-08 | 1.67E-08 | 0.850237 |
| PD Braak | Ovary | Other | 1.05E-08 | 1.79E-08 | 0.278001 |
| PD Braak | Pituitary | Other | 2.35E-08 | 1.65E-08 | 0.076945 |
| PD Braak | Prostate | Other | -1.60E-09 | 2.04E-08 | 0.531300 |
| PD Braak | Skin_Not_Sun_Exposed_(Suprapubic) | Other | 1.27E-08 | 1.86E-08 | 0.246988 |
| PD Braak | Skin_Sun_Exposed_(Lower_leg) | Other | 7.14E-09 | 1.83E-08 | 0.348358 |
| PD Braak | Testis | Other | -3.04E-08 | 1.61E-08 | 0.970356 |
| PD Braak | Thyroid | Other | 2.60E-08 | 1.90E-08 | 0.086007 |
| PD Braak | Uterus | Other | 6.46E-09 | 1.97E-08 | 0.371745 |
| PD Braak | Vagina | Other | 4.38E-09 | 1.85E-08 | 0.406272 |
| PD Braak | A03.734.414.Islets.of.Langerhans | Pancreas | -3.01E-08 | 2.03E-08 | 0.931009 |
| PD Braak | A03.734.Pancreas | Pancreas | 1.21E-08 | 1.87E-08 | 0.259976 |

| | | | | | |
|---|---|---|---|---|---|
| PD Braak | Pancreas | Pancreas | -3.90E-09 | 2.05E-08 | 0.575436 |
| PD Braak (any/none) | A10.165.114.830.500.750.Subcutaneous.Fat..Abdominal | Adipose | -1.35E-08 | 1.77E-08 | 0.778274 |
| PD Braak (any/none) | A10.165.114.830.750.Subcutaneous.Fat | Adipose | -1.13E-08 | 1.77E-08 | 0.738341 |
| PD Braak (any/none) | A11.329.114.Adipocytes | Adipose | -6.56E-09 | 1.87E-08 | 0.637256 |
| PD Braak (any/none) | Adipose_Subcutaneous | Adipose | 7.77E-09 | 1.71E-08 | 0.325075 |
| PD Braak (any/none) | Adipose_Visceral_(Omentum) | Adipose | 2.71E-08 | 1.82E-08 | 0.068480 |
| PD Braak (any/none) | A02.835.583.443.800.800.Synovial.Fluid | Blood/Immune | -2.47E-08 | 2.33E-08 | 0.855070 |
| PD Braak (any/none) | A10.549.400.Lymph.Nodes | Blood/Immune | -4.78E-08 | 2.01E-08 | 0.991322 |
| PD Braak (any/none) | A10.549.Lymphoid.Tissue | Blood/Immune | -5.64E-08 | 1.80E-08 | 0.999123 |
| PD Braak (any/none) | A11.118.637.555.567.562.440.Precursor.Cells..B.Lymphoid | Blood/Immune | -1.55E-08 | 1.81E-08 | 0.803447 |
| PD Braak (any/none) | A11.118.637.555.567.562.B.Lymphocytes | Blood/Immune | -2.44E-08 | 1.97E-08 | 0.892287 |
| PD Braak (any/none) | A11.118.637.555.567.569.200.700.T.Lymphocytes..Regulatory | Blood/Immune | -2.98E-08 | 2.04E-08 | 0.928199 |
| PD Braak (any/none) | A11.118.637.555.567.569.T.Lymphocytes | Blood/Immune | -3.32E-08 | 1.96E-08 | 0.955309 |
| PD Braak (any/none) | A11.118.637.Leukocytes | Blood/Immune | -3.53E-08 | 2.37E-08 | 0.931606 |
| PD Braak (any/none) | A11.329.372.600.Macrophages..Alveolar | Blood/Immune | -4.55E-09 | 2.03E-08 | 0.588946 |
| PD Braak (any/none) | A11.443.Erythroid.Cells | Blood/Immune | 8.04E-09 | 2.06E-08 | 0.348207 |
| PD Braak (any/none) | A11.627.340.360.Granulocyte.Precursor.Cells | Blood/Immune | -1.68E-08 | 2.12E-08 | 0.786160 |
| PD Braak (any/none) | A11.627.624.249.Monocyte.Macrophage.Precursor.Cells | Blood/Immune | 4.92E-09 | 2.09E-08 | 0.406799 |
| PD Braak (any/none) | A11.627.635.Myeloid.Progenitor.Cells | Blood/Immune | 3.17E-11 | 2.16E-08 | 0.499415 |
| PD Braak (any/none) | A11.872.378.590.635.Granulocyte.Macrophage.Progenitor.Cells | Blood/Immune | 6.51E-09 | 2.24E-08 | 0.385649 |
| PD Braak (any/none) | A11.872.378.590.817.Megakaryocyte.Erythroid.Progenitor.Cells | Blood/Immune | -3.39E-08 | 1.87E-08 | 0.964945 |
| PD Braak (any/none) | A11.872.378.Hematopoietic.Stem.Cells | Blood/Immune | -1.02E-08 | 2.16E-08 | 0.681914 |
| PD Braak (any/none) | A15.145.229.188.Blood.Platelets | Blood/Immune | 5.52E-09 | 2.36E-08 | 0.407537 |
| PD Braak (any/none) | A15.145.229.637.555.567.562.725.Plasma.Cells | Blood/Immune | -2.47E-08 | 2.27E-08 | 0.861568 |
| PD Braak (any/none) | A15.145.229.637.555.567.569.200.CD4.Positive.T.Lymphocytes | Blood/Immune | -3.75E-08 | 1.98E-08 | 0.971062 |
| PD Braak (any/none) | A15.145.229.637.555.Leukocytes..Mononuclear | Blood/Immune | -6.40E-08 | 2.36E-08 | 0.996686 |
| PD Braak (any/none) | A15.145.229.Blood.Cells | Blood/Immune | -3.89E-08 | 2.38E-08 | 0.948564 |
| PD Braak (any/none) | A15.145.300.Fetal.Blood | Blood/Immune | -3.25E-08 | 2.64E-08 | 0.890839 |
| PD Braak (any/none) | A15.145.846.Serum | Blood/Immune | -2.25E-08 | 2.17E-08 | 0.849348 |
| PD Braak (any/none) | A15.145.Blood | Blood/Immune | -5.03E-08 | 2.27E-08 | 0.986628 |
| PD Braak (any/none) | A15.378.316.580.Monocytes | Blood/Immune | 5.01E-10 | 2.28E-08 | 0.491251 |
| PD Braak (any/none) | A15.378.316.Bone.Marrow.Cells | Blood/Immune | 5.00E-09 | 2.33E-08 | 0.414938 |
| PD Braak (any/none) | A15.382.490.315.583.Neutrophils | Blood/Immune | 2.13E-08 | 2.20E-08 | 0.166749 |
| PD Braak (any/none) | A15.382.490.555.567.537.Killer.Cells..Natural | Blood/Immune | -3.99E-08 | 1.74E-08 | 0.989242 |
| PD Braak (any/none) | A15.382.490.555.567.622.Lymphocytes..Null | Blood/Immune | -2.95E-08 | 2.19E-08 | 0.911086 |
| PD Braak (any/none) | A15.382.490.555.567.Lymphocytes | Blood/Immune | -3.60E-08 | 2.10E-08 | 0.956592 |
| PD Braak (any/none) | A15.382.520.604.700.Spleen | Blood/Immune | -2.72E-08 | 1.88E-08 | 0.925884 |
| PD Braak (any/none) | A15.382.520.604.800.Palatine.Tonsil | Blood/Immune | -6.99E-09 | 1.82E-08 | 0.649661 |
| PD Braak (any/none) | A15.382.680.Phagocytes | Blood/Immune | -1.22E-08 | 2.18E-08 | 0.712365 |
| PD Braak (any/none) | A15.382.812.260.Dendritic.Cells | Blood/Immune | -1.55E-09 | 2.14E-08 | 0.528946 |
| PD Braak (any/none) | A15.382.812.522.Macrophages | Blood/Immune | -6.63E-09 | 2.18E-08 | 0.619669 |
| PD Braak (any/none) | A15.382.812.Mononuclear.Phagocyte.System | Blood/Immune | -2.30E-08 | 2.06E-08 | 0.868089 |
| PD Braak (any/none) | A15.382.Immune.System | Blood/Immune | -4.74E-08 | 2.16E-08 | 0.985888 |
| PD Braak (any/none) | Cells_EBV-transformed_lymphocytes | Blood/Immune | -4.68E-08 | 2.07E-08 | 0.988298 |
| PD Braak (any/none) | Spleen | Blood/Immune | -5.67E-08 | 2.13E-08 | 0.996119 |
| PD Braak (any/none) | Whole_Blood | Blood/Immune | 1.63E-09 | 2.32E-08 | 0.472103 |

| | | | | | |
|---|---|---|---|---|---|
| PD Braak (any/none) | A07.231.114.Arteries | Cardiovascular | -2.42E-08 | 2.16E-08 | 0.868725 |
| PD Braak (any/none) | A07.231.908.670.874.Umbilical.Veins | Cardiovascular | -2.89E-08 | 2.32E-08 | 0.892880 |
| PD Braak (any/none) | A07.231.908.Veins | Cardiovascular | -2.34E-08 | 2.33E-08 | 0.842032 |
| PD Braak (any/none) | A07.231.Blood.Vessels | Cardiovascular | -4.58E-08 | 2.63E-08 | 0.959271 |
| PD Braak (any/none) | A07.541.358.100.Atrial.Appendage | Cardiovascular | 4.09E-09 | 1.75E-08 | 0.407684 |
| PD Braak (any/none) | A07.541.358.Heart.Atria | Cardiovascular | -2.10E-09 | 1.77E-08 | 0.547100 |
| PD Braak (any/none) | A07.541.510.110.Aortic.Valve | Cardiovascular | -4.87E-09 | 1.90E-08 | 0.601401 |
| PD Braak (any/none) | A07.541.560.Heart.Ventricles | Cardiovascular | -2.26E-08 | 1.83E-08 | 0.891336 |
| PD Braak (any/none) | A07.541.Heart | Cardiovascular | -2.97E-08 | 1.83E-08 | 0.947508 |
| PD Braak (any/none) | Artery_Aorta | Cardiovascular | -3.11E-08 | 1.58E-08 | 0.975714 |
| PD Braak (any/none) | Artery_Coronary | Cardiovascular | -1.56E-08 | 1.67E-08 | 0.824373 |
| PD Braak (any/none) | Artery_Tibial | Cardiovascular | -3.34E-08 | 1.48E-08 | 0.987815 |
| PD Braak (any/none) | Heart_Atrial_Appendage | Cardiovascular | -1.55E-08 | 1.58E-08 | 0.835606 |
| PD Braak (any/none) | Heart_Left_Ventricle | Cardiovascular | -1.17E-08 | 1.73E-08 | 0.751327 |
| PD Braak (any/none) | A08.186.211.132.810.428.200.Cerebellum | CNS | 8.79E-09 | 1.97E-08 | 0.327939 |
| PD Braak (any/none) | A08.186.211.132.Brain.Stem | CNS | 2.95E-08 | 1.96E-08 | 0.066505 |
| PD Braak (any/none) | A08.186.211.464.405.Hippocampus | CNS | 4.54E-08 | 1.80E-08 | 0.005844 |
| PD Braak (any/none) | A08.186.211.464.710.225.Entorhinal.Cortex | CNS | 3.56E-08 | 1.79E-08 | 0.023468 |
| PD Braak (any/none) | A08.186.211.464.Limbic.System | CNS | 4.18E-08 | 1.79E-08 | 0.009680 |
| PD Braak (any/none) | A08.186.211.653.Mesencephalon | CNS | 2.53E-08 | 2.00E-08 | 0.103215 |
| PD Braak (any/none) | A08.186.211.730.317.357.352.435.Hypothalamo.Hypophyseal.System | CNS | 2.19E-08 | 2.19E-08 | 0.159195 |
| PD Braak (any/none) | A08.186.211.730.317.357.Hypothalamus | CNS | 3.72E-08 | 2.16E-08 | 0.042412 |
| PD Braak (any/none) | A08.186.211.730.317.Diencephalon | CNS | 4.31E-08 | 2.28E-08 | 0.029704 |
| PD Braak (any/none) | A08.186.211.730.885.287.249.487.Corpus.Striatum | CNS | 4.48E-09 | 1.75E-08 | 0.399207 |
| PD Braak (any/none) | A08.186.211.730.885.287.249.Basal.Ganglia | CNS | 3.08E-08 | 1.87E-08 | 0.049530 |
| PD Braak (any/none) | A08.186.211.730.885.287.500.270.Frontal.Lobe | CNS | 3.64E-08 | 1.98E-08 | 0.033038 |
| PD Braak (any/none) | A08.186.211.730.885.287.500.571.735.Visual.Cortex | CNS | 1.92E-08 | 1.82E-08 | 0.146063 |
| PD Braak (any/none) | A08.186.211.730.885.287.500.670.Parietal.Lobe | CNS | 2.64E-08 | 2.04E-08 | 0.097279 |
| PD Braak (any/none) | A08.186.211.730.885.287.500.Cerebral.Cortex | CNS | 3.98E-08 | 1.85E-08 | 0.015855 |
| PD Braak (any/none) | A08.186.211.865.428.Metencephalon | CNS | 1.26E-08 | 1.92E-08 | 0.256939 |
| PD Braak (any/none) | A08.186.211.Brain | CNS | 2.94E-08 | 1.69E-08 | 0.041363 |
| PD Braak (any/none) | A09.371.729.Retina | CNS | 1.75E-08 | 1.86E-08 | 0.173489 |
| PD Braak (any/none) | A11.872.653.Neural.Stem.Cells | CNS | 1.16E-08 | 2.07E-08 | 0.288582 |
| PD Braak (any/none) | Brain_Amygdala | CNS | 4.65E-08 | 1.62E-08 | 0.002000 |
| PD Braak (any/none) | Brain_Anterior_cingulate_cortex_(BA24) | CNS | 4.13E-08 | 1.63E-08 | 0.005563 |
| PD Braak (any/none) | Brain_Caudate_(basal_ganglia) | CNS | 5.38E-08 | 1.67E-08 | 0.000638 |
| PD Braak (any/none) | Brain_Cerebellar_Hemisphere | CNS | 1.80E-08 | 1.72E-08 | 0.146462 |
| PD Braak (any/none) | Brain_Cerebellum | CNS | 1.78E-08 | 1.74E-08 | 0.153054 |
| PD Braak (any/none) | Brain_Cortex | CNS | 2.66E-08 | 1.73E-08 | 0.062239 |
| PD Braak (any/none) | Brain_Frontal_Cortex_(BA9) | CNS | 2.61E-08 | 1.59E-08 | 0.050297 |
| PD Braak (any/none) | Brain_Hippocampus | CNS | 4.36E-08 | 1.53E-08 | 0.002154 |
| PD Braak (any/none) | Brain_Hypothalamus | CNS | 4.27E-08 | 1.60E-08 | 0.003896 |
| PD Braak (any/none) | Brain_Nucleus_accumbens_(basal_ganglia) | CNS | 3.33E-08 | 1.62E-08 | 0.020130 |
| PD Braak (any/none) | Brain_Putamen_(basal_ganglia) | CNS | 4.29E-08 | 1.66E-08 | 0.004915 |
| PD Braak (any/none) | Brain_Spinal_cord_(cervical_c-1) | CNS | 4.34E-08 | 1.61E-08 | 0.003441 |
| PD Braak (any/none) | Brain_Substantia_nigra | CNS | 4.27E-08 | 1.55E-08 | 0.002894 |

| | | | | | |
|---|---|---|---|---|---|
| PD Braak (any/none) | A03.556.124.369.Intestinal.Mucosa | Digestive | 3.44E-08 | 2.07E-08 | 0.047946 |
| PD Braak (any/none) | A03.556.124.526.767.Rectum | Digestive | 2.85E-08 | 2.05E-08 | 0.082287 |
| PD Braak (any/none) | A03.556.124.684.Intestine..Small | Digestive | -6.46E-09 | 1.92E-08 | 0.631957 |
| PD Braak (any/none) | A03.556.124.Intestines | Digestive | 3.73E-08 | 1.96E-08 | 0.028470 |
| PD Braak (any/none) | A03.556.249.124.Ileum | Digestive | 1.62E-08 | 2.11E-08 | 0.220924 |
| PD Braak (any/none) | A03.556.249.249.209.Cecum | Digestive | 1.44E-08 | 2.02E-08 | 0.238780 |
| PD Braak (any/none) | A03.556.249.249.356.668.Colon..Sigmoid | Digestive | 1.20E-08 | 2.11E-08 | 0.285065 |
| PD Braak (any/none) | A03.556.249.249.356.Colon | Digestive | 3.75E-08 | 1.95E-08 | 0.027405 |
| PD Braak (any/none) | A03.556.500.760.464.Parotid.Gland | Digestive | 2.50E-08 | 2.30E-08 | 0.138618 |
| PD Braak (any/none) | A03.556.500.760.Salivary.Glands | Digestive | 2.48E-08 | 2.28E-08 | 0.138762 |
| PD Braak (any/none) | A03.556.875.500.Esophagus | Digestive | 5.77E-08 | 2.09E-08 | 0.002927 |
| PD Braak (any/none) | A03.556.875.875.Stomach | Digestive | -6.01E-09 | 1.87E-08 | 0.625779 |
| PD Braak (any/none) | A03.556.875.Upper.Gastrointestinal.Tract | Digestive | -2.45E-10 | 1.92E-08 | 0.505092 |
| PD Braak (any/none) | A03.556.Gastrointestinal.Tract | Digestive | 4.02E-08 | 2.07E-08 | 0.026046 |
| PD Braak (any/none) | Colon_Sigmoid | Digestive | -6.55E-09 | 1.66E-08 | 0.653334 |
| PD Braak (any/none) | Colon_Transverse | Digestive | -9.94E-09 | 1.87E-08 | 0.702858 |
| PD Braak (any/none) | Esophagus_Gastroesophageal_Junction | Digestive | -4.89E-08 | 1.78E-08 | 0.997076 |
| PD Braak (any/none) | Esophagus_Mucosa | Digestive | 5.32E-09 | 1.85E-08 | 0.386989 |
| PD Braak (any/none) | Esophagus_Muscularis | Digestive | -4.99E-08 | 1.75E-08 | 0.997824 |
| PD Braak (any/none) | Small_Intestine_Terminal_Ileum | Digestive | 1.09E-08 | 1.80E-08 | 0.272569 |
| PD Braak (any/none) | Stomach | Digestive | 1.04E-08 | 1.83E-08 | 0.284580 |
| PD Braak (any/none) | A03.620.Liver | Liver | -2.09E-08 | 2.19E-08 | 0.829974 |
| PD Braak (any/none) | A11.436.348.Hepatocytes | Liver | 7.15E-09 | 2.26E-08 | 0.375896 |
| PD Braak (any/none) | Liver | Liver | -1.84E-08 | 1.95E-08 | 0.826951 |
| PD Braak (any/none) | A02.165.Cartilage | Musculoskeletal/Connective | -1.34E-08 | 1.89E-08 | 0.761467 |
| PD Braak (any/none) | A02.633.567.850.Quadriceps.Muscle | Musculoskeletal/Connective | -2.62E-08 | 1.87E-08 | 0.919518 |
| PD Braak (any/none) | A02.835.232.834.151.Cervical.Vertebrae | Musculoskeletal/Connective | -1.31E-08 | 1.92E-08 | 0.753077 |
| PD Braak (any/none) | A02.835.583.443.800.Synovial.Membrane | Musculoskeletal/Connective | -2.78E-09 | 1.86E-08 | 0.559393 |
| PD Braak (any/none) | A05.360.319.679.690.Myometrium | Musculoskeletal/Connective | 1.49E-08 | 1.89E-08 | 0.215852 |
| PD Braak (any/none) | A10.165.450.300.425.Keloid | Musculoskeletal/Connective | -7.06E-09 | 1.80E-08 | 0.652849 |
| PD Braak (any/none) | A10.165.450.300.Cicatrix | Musculoskeletal/Connective | -1.46E-08 | 1.86E-08 | 0.783681 |
| PD Braak (any/none) | A10.690.467.Muscle..Smooth | Musculoskeletal/Connective | -1.92E-08 | 1.79E-08 | 0.858049 |
| PD Braak (any/none) | A10.690.Muscles | Musculoskeletal/Connective | -8.60E-09 | 1.91E-08 | 0.673432 |
| PD Braak (any/none) | A11.329.171.Chondrocytes | Musculoskeletal/Connective | -2.72E-08 | 1.87E-08 | 0.927817 |
| PD Braak (any/none) | A11.329.228.Fibroblasts | Musculoskeletal/Connective | -2.49E-08 | 1.80E-08 | 0.917475 |
| PD Braak (any/none) | A11.329.629.Osteoblasts | Musculoskeletal/Connective | -1.66E-08 | 1.89E-08 | 0.809196 |
| PD Braak (any/none) | A11.329.830.Stromal.Cells | Musculoskeletal/Connective | -2.43E-08 | 1.67E-08 | 0.927502 |
| PD Braak (any/none) | A11.329.Connective.Tissue.Cells | Musculoskeletal/Connective | -9.75E-09 | 1.89E-08 | 0.696759 |
| PD Braak (any/none) | A11.620.520.Myocytes..Smooth.Muscle | Musculoskeletal/Connective | -1.76E-08 | 1.81E-08 | 0.834630 |
| PD Braak (any/none) | Cells_Transformed_fibroblasts | Musculoskeletal/Connective | -1.73E-09 | 1.81E-08 | 0.538094 |
| PD Braak (any/none) | Muscle_Skeletal | Musculoskeletal/Connective | -1.01E-08 | 1.77E-08 | 0.716320 |
| PD Braak (any/none) | A04.411.Lung | Other | -1.81E-08 | 2.07E-08 | 0.809406 |
| PD Braak (any/none) | A04.531.520.Nasal.Mucosa | Other | -9.67E-09 | 1.91E-08 | 0.693492 |
| PD Braak (any/none) | A05.360.319.114.373.Fallopian.Tubes | Other | 1.02E-09 | 2.09E-08 | 0.480638 |
| PD Braak (any/none) | A05.360.319.114.630.Ovary | Other | 4.23E-09 | 2.08E-08 | 0.419503 |
| PD Braak (any/none) | A05.360.319.679.256.Cervix.Uteri | Other | -1.67E-08 | 1.85E-08 | 0.815957 |

| | | | | | |
|---|---|---|---|---|---|
| PD Braak (any/none) | A05.360.319.679.490.Endometrium | Other | 7.25E-09 | 1.95E-08 | 0.355266 |
| PD Braak (any/none) | A05.360.319.679.Uterus | Other | 2.76E-08 | 1.76E-08 | 0.058966 |
| PD Braak (any/none) | A05.360.319.887.Vulva | Other | 5.16E-08 | 2.30E-08 | 0.012452 |
| PD Braak (any/none) | A05.360.319.Genitalia..Female | Other | 7.46E-09 | 1.95E-08 | 0.351413 |
| PD Braak (any/none) | A05.360.444.492.362.Foreskin | Other | -1.17E-08 | 1.61E-08 | 0.765449 |
| PD Braak (any/none) | A05.360.444.Genitalia..Male | Other | 2.36E-09 | 1.87E-08 | 0.449759 |
| PD Braak (any/none) | A05.360.490.Germ.Cells | Other | -1.79E-08 | 1.81E-08 | 0.838380 |
| PD Braak (any/none) | A05.360.Genitalia | Other | 1.74E-08 | 2.01E-08 | 0.193910 |
| PD Braak (any/none) | A05.810.453.324.Kidney.Cortex | Other | -3.52E-08 | 1.80E-08 | 0.974741 |
| PD Braak (any/none) | A05.810.453.Kidney | Other | -1.70E-08 | 1.72E-08 | 0.838021 |
| PD Braak (any/none) | A05.810.890.Urinary.Bladder | Other | 1.02E-08 | 2.06E-08 | 0.309748 |
| PD Braak (any/none) | A06.407.071.140.Adrenal.Cortex | Other | -1.71E-08 | 1.94E-08 | 0.810868 |
| PD Braak (any/none) | A06.407.071.Adrenal.Glands | Other | -1.15E-08 | 1.92E-08 | 0.725314 |
| PD Braak (any/none) | A06.407.312.782.Testis | Other | -2.36E-08 | 1.61E-08 | 0.928401 |
| PD Braak (any/none) | A06.407.312.Gonads | Other | 5.59E-09 | 2.07E-08 | 0.393736 |
| PD Braak (any/none) | A06.407.900.Thyroid.Gland | Other | 2.33E-08 | 1.90E-08 | 0.109737 |
| PD Braak (any/none) | A06.407.Endocrine.Glands | Other | 1.63E-08 | 2.18E-08 | 0.227930 |
| PD Braak (any/none) | A09.371.Eye | Other | -5.71E-09 | 1.93E-08 | 0.616140 |
| PD Braak (any/none) | A10.272.497.Epidermis | Other | -1.44E-08 | 2.18E-08 | 0.745962 |
| PD Braak (any/none) | A10.272.Epithelium | Other | -8.66E-09 | 2.01E-08 | 0.667055 |
| PD Braak (any/none) | A10.336.707.Prostate | Other | 4.63E-09 | 2.01E-08 | 0.409060 |
| PD Braak (any/none) | A10.615.284.473.Chorion | Other | -3.63E-08 | 1.80E-08 | 0.978105 |
| PD Braak (any/none) | A10.615.550.599.Mouth.Mucosa | Other | 3.57E-08 | 2.01E-08 | 0.037495 |
| PD Braak (any/none) | A10.615.550.Mucous.Membrane | Other | 8.01E-09 | 1.90E-08 | 0.336514 |
| PD Braak (any/none) | A10.615.789.Serous.Membrane | Other | -4.40E-08 | 2.18E-08 | 0.978210 |
| PD Braak (any/none) | A10.615.Membranes | Other | 1.43E-08 | 1.87E-08 | 0.223313 |
| PD Braak (any/none) | A11.382.Endocrine.Cells | Other | -2.82E-08 | 1.85E-08 | 0.936273 |
| PD Braak (any/none) | A11.436.275.Endothelial.Cells | Other | -3.43E-08 | 2.52E-08 | 0.913058 |
| PD Braak (any/none) | A11.436.294.064.Glucagon.Secreting.Cells | Other | -1.41E-08 | 2.03E-08 | 0.756829 |
| PD Braak (any/none) | A11.436.329.Granulosa.Cells | Other | -2.51E-08 | 1.71E-08 | 0.928853 |
| PD Braak (any/none) | A11.436.397.Keratinocytes | Other | 1.29E-08 | 1.97E-08 | 0.255495 |
| PD Braak (any/none) | A11.436.Epithelial.Cells | Other | 1.06E-09 | 1.99E-08 | 0.478654 |
| PD Braak (any/none) | A11.497.497.600.Oocytes | Other | -1.93E-08 | 1.73E-08 | 0.866917 |
| PD Braak (any/none) | A11.872.040.Adult.Stem.Cells | Other | -2.12E-08 | 1.77E-08 | 0.884847 |
| PD Braak (any/none) | A11.872.190.260.Embryoid.Bodies | Other | 5.32E-09 | 1.92E-08 | 0.390757 |
| PD Braak (any/none) | A11.872.190.Embryonic.Stem.Cells | Other | -1.10E-08 | 1.74E-08 | 0.735780 |
| PD Braak (any/none) | A11.872.580.Mesenchymal.Stem.Cells | Other | -3.59E-08 | 1.73E-08 | 0.980819 |
| PD Braak (any/none) | A11.872.700.500.Induced.Pluripotent.Stem.Cells | Other | -2.10E-08 | 1.75E-08 | 0.884421 |
| PD Braak (any/none) | A11.872.Stem.Cells | Other | -4.24E-08 | 2.02E-08 | 0.982250 |
| PD Braak (any/none) | A14.549.167.646.Periodontium | Other | 3.05E-08 | 2.36E-08 | 0.097848 |
| PD Braak (any/none) | A14.549.167.Dentition | Other | 1.04E-08 | 2.22E-08 | 0.320446 |
| PD Braak (any/none) | A14.549.885.Tongue | Other | -6.61E-09 | 2.17E-08 | 0.619487 |
| PD Braak (any/none) | A14.549.Mouth | Other | 3.85E-08 | 2.11E-08 | 0.033945 |
| PD Braak (any/none) | A14.724.557.Nasopharynx | Other | 2.34E-09 | 1.96E-08 | 0.452410 |
| PD Braak (any/none) | A14.724.Pharynx | Other | 2.64E-09 | 1.86E-08 | 0.443590 |
| PD Braak (any/none) | A17.815.Skin | Other | 2.58E-08 | 1.99E-08 | 0.096865 |

| | | | | | |
|---|---|---|---|---|---|
| PD Braak (any/none) | Adrenal_Gland | Other | -1.33E-08 | 1.72E-08 | 0.780576 |
| PD Braak (any/none) | Bladder | Other | -9.36E-09 | 1.61E-08 | 0.719417 |
| PD Braak (any/none) | Breast_Mammary_Tissue | Other | 3.71E-08 | 1.74E-08 | 0.016558 |
| PD Braak (any/none) | Cervix_Ectocervix | Other | 1.35E-08 | 1.75E-08 | 0.218989 |
| PD Braak (any/none) | Cervix_Endocervix | Other | -5.41E-09 | 1.79E-08 | 0.618390 |
| PD Braak (any/none) | Fallopian_Tube | Other | 9.70E-09 | 1.95E-08 | 0.309134 |
| PD Braak (any/none) | Kidney_Cortex | Other | -2.99E-08 | 1.83E-08 | 0.948882 |
| PD Braak (any/none) | Lung | Other | -3.58E-08 | 1.86E-08 | 0.972817 |
| PD Braak (any/none) | Minor_Salivary_Gland | Other | 5.43E-09 | 1.75E-08 | 0.377945 |
| PD Braak (any/none) | Nerve_Tibial | Other | -1.95E-08 | 1.66E-08 | 0.879767 |
| PD Braak (any/none) | Ovary | Other | 3.81E-09 | 1.78E-08 | 0.415122 |
| PD Braak (any/none) | Pituitary | Other | 1.49E-08 | 1.62E-08 | 0.179753 |
| PD Braak (any/none) | Prostate | Other | -1.05E-08 | 2.01E-08 | 0.698841 |
| PD Braak (any/none) | Skin_Not_Sun_Exposed_(Suprapubic) | Other | 1.55E-08 | 1.95E-08 | 0.213409 |
| PD Braak (any/none) | Skin_Sun_Exposed_(Lower_leg) | Other | 1.15E-08 | 1.92E-08 | 0.275362 |
| PD Braak (any/none) | Testis | Other | -3.11E-08 | 1.64E-08 | 0.970749 |
| PD Braak (any/none) | Thyroid | Other | 2.92E-08 | 1.89E-08 | 0.060979 |
| PD Braak (any/none) | Uterus | Other | 3.81E-09 | 1.90E-08 | 0.420439 |
| PD Braak (any/none) | Vagina | Other | 8.32E-09 | 1.79E-08 | 0.321134 |
| PD Braak (any/none) | A03.734.414.Islets.of.Langerhans | Pancreas | -2.76E-08 | 2.01E-08 | 0.914658 |
| PD Braak (any/none) | A03.734.Pancreas | Pancreas | 7.68E-09 | 1.88E-08 | 0.341239 |
| PD Braak (any/none) | Pancreas | Pancreas | -1.43E-09 | 1.99E-08 | 0.528728 |
| TDP-43 (any/none) | A10.165.114.830.500.750.Subcutaneous.Fat..Abdominal | Adipose | -6.66E-09 | 5.35E-08 | 0.549499 |
| TDP-43 (any/none) | A10.165.114.830.750.Subcutaneous.Fat | Adipose | 1.12E-08 | 5.42E-08 | 0.417875 |
| TDP-43 (any/none) | A11.329.114.Adipocytes | Adipose | -1.59E-08 | 5.11E-08 | 0.622356 |
| TDP-43 (any/none) | Adipose_Subcutaneous | Adipose | 3.24E-08 | 4.54E-08 | 0.238010 |
| TDP-43 (any/none) | Adipose_Visceral_(Omentum) | Adipose | 4.98E-08 | 4.49E-08 | 0.133273 |
| TDP-43 (any/none) | A02.835.583.443.800.800.Synovial.Fluid | Blood/Immune | 1.71E-08 | 6.09E-08 | 0.389802 |
| TDP-43 (any/none) | A10.549.400.Lymph.Nodes | Blood/Immune | -5.62E-08 | 5.47E-08 | 0.847814 |
| TDP-43 (any/none) | A10.549.Lymphoid.Tissue | Blood/Immune | -3.64E-08 | 5.54E-08 | 0.744172 |
| TDP-43 (any/none) | A11.118.637.555.567.562.440.Precursor.Cells..B.Lymphoid | Blood/Immune | 1.81E-08 | 5.92E-08 | 0.379968 |
| TDP-43 (any/none) | A11.118.637.555.567.562.B.Lymphocytes | Blood/Immune | 2.72E-08 | 5.74E-08 | 0.318021 |
| TDP-43 (any/none) | A11.118.637.555.567.569.200.700.T.Lymphocytes..Regulatory | Blood/Immune | -4.16E-08 | 5.60E-08 | 0.771560 |
| TDP-43 (any/none) | A11.118.637.555.567.569.T.Lymphocytes | Blood/Immune | 4.32E-08 | 5.18E-08 | 0.201998 |
| TDP-43 (any/none) | A11.118.637.Leukocytes | Blood/Immune | -4.63E-08 | 5.87E-08 | 0.784841 |
| TDP-43 (any/none) | A11.329.372.600.Macrophages..Alveolar | Blood/Immune | 3.04E-09 | 5.18E-08 | 0.476577 |
| TDP-43 (any/none) | A11.443.Erythroid.Cells | Blood/Immune | 5.62E-08 | 5.21E-08 | 0.140338 |
| TDP-43 (any/none) | A11.627.340.360.Granulocyte.Precursor.Cells | Blood/Immune | 1.20E-08 | 6.25E-08 | 0.423951 |
| TDP-43 (any/none) | A11.627.624.249.Monocyte.Macrophage.Precursor.Cells | Blood/Immune | -3.11E-08 | 5.38E-08 | 0.718550 |
| TDP-43 (any/none) | A11.627.635.Myeloid.Progenitor.Cells | Blood/Immune | -9.76E-09 | 5.68E-08 | 0.568185 |
| TDP-43 (any/none) | A11.872.378.590.635.Granulocyte.Macrophage.Progenitor.Cells | Blood/Immune | -9.89E-08 | 5.89E-08 | 0.953461 |
| TDP-43 (any/none) | A11.872.378.590.817.Megakaryocyte.Erythroid.Progenitor.Cells | Blood/Immune | 5.36E-08 | 5.33E-08 | 0.157538 |
| TDP-43 (any/none) | A11.872.378.Hematopoietic.Stem.Cells | Blood/Immune | -2.10E-08 | 5.62E-08 | 0.646033 |
| TDP-43 (any/none) | A15.145.229.188.Blood.Platelets | Blood/Immune | -5.75E-08 | 5.44E-08 | 0.854925 |
| TDP-43 (any/none) | A15.145.229.637.555.567.562.725.Plasma.Cells | Blood/Immune | 4.04E-08 | 5.85E-08 | 0.244991 |
| TDP-43 (any/none) | A15.145.229.637.555.567.569.200.CD4.Positive.T.Lymphocytes | Blood/Immune | 4.58E-09 | 5.72E-08 | 0.468127 |

| | | | | | |
|---|---|---|---|---|---|
| TDP-43 (any/none) | A15.145.229.637.555.Leukocytes..Mononuclear | Blood/Immune | 7.71E-08 | 6.33E-08 | 0.111848 |
| TDP-43 (any/none) | A15.145.229.Blood.Cells | Blood/Immune | -3.39E-08 | 6.15E-08 | 0.709328 |
| TDP-43 (any/none) | A15.145.300.Fetal.Blood | Blood/Immune | -2.23E-08 | 6.54E-08 | 0.633805 |
| TDP-43 (any/none) | A15.145.846.Serum | Blood/Immune | 1.87E-08 | 5.97E-08 | 0.377235 |
| TDP-43 (any/none) | A15.145.Blood | Blood/Immune | -4.37E-08 | 6.34E-08 | 0.754346 |
| TDP-43 (any/none) | A15.378.316.580.Monocytes | Blood/Immune | -3.04E-08 | 5.90E-08 | 0.696965 |
| TDP-43 (any/none) | A15.378.316.Bone.Marrow.Cells | Blood/Immune | -7.83E-08 | 5.70E-08 | 0.915221 |
| TDP-43 (any/none) | A15.382.490.315.583.Neutrophils | Blood/Immune | -7.14E-08 | 5.72E-08 | 0.893988 |
| TDP-43 (any/none) | A15.382.490.555.567.537.Killer.Cells..Natural | Blood/Immune | 5.59E-08 | 5.76E-08 | 0.165972 |
| TDP-43 (any/none) | A15.382.490.555.567.622.Lymphocytes..Null | Blood/Immune | -1.98E-08 | 6.07E-08 | 0.628134 |
| TDP-43 (any/none) | A15.382.490.555.567.Lymphocytes | Blood/Immune | 3.53E-08 | 6.14E-08 | 0.282382 |
| TDP-43 (any/none) | A15.382.520.604.700.Spleen | Blood/Immune | 2.84E-08 | 5.81E-08 | 0.312440 |
| TDP-43 (any/none) | A15.382.520.604.800.Palatine.Tonsil | Blood/Immune | -6.28E-08 | 5.43E-08 | 0.876152 |
| TDP-43 (any/none) | A15.382.680.Phagocytes | Blood/Immune | -5.65E-08 | 5.83E-08 | 0.833985 |
| TDP-43 (any/none) | A15.382.812.260.Dendritic.Cells | Blood/Immune | -4.05E-08 | 5.61E-08 | 0.765052 |
| TDP-43 (any/none) | A15.382.812.522.Macrophages | Blood/Immune | -3.14E-08 | 5.55E-08 | 0.714521 |
| TDP-43 (any/none) | A15.382.812.Mononuclear.Phagocyte.System | Blood/Immune | -1.36E-08 | 6.00E-08 | 0.589480 |
| TDP-43 (any/none) | A15.382.Immune.System | Blood/Immune | -3.32E-08 | 6.09E-08 | 0.707400 |
| TDP-43 (any/none) | Cells_EBV-transformed_lymphocytes | Blood/Immune | -3.62E-08 | 5.28E-08 | 0.753425 |
| TDP-43 (any/none) | Spleen | Blood/Immune | 1.47E-08 | 5.47E-08 | 0.393953 |
| TDP-43 (any/none) | Whole_Blood | Blood/Immune | -5.84E-08 | 5.56E-08 | 0.853049 |
| TDP-43 (any/none) | A07.231.114.Arteries | Cardiovascular | -5.03E-08 | 5.34E-08 | 0.826857 |
| TDP-43 (any/none) | A07.231.908.670.874.Umbilical.Veins | Cardiovascular | 6.71E-08 | 5.74E-08 | 0.121278 |
| TDP-43 (any/none) | A07.231.908.Veins | Cardiovascular | 4.21E-08 | 5.89E-08 | 0.237040 |
| TDP-43 (any/none) | A07.231.Blood.Vessels | Cardiovascular | 7.21E-08 | 6.42E-08 | 0.130659 |
| TDP-43 (any/none) | A07.541.358.100.Atrial.Appendage | Cardiovascular | 7.91E-08 | 4.90E-08 | 0.053451 |
| TDP-43 (any/none) | A07.541.358.Heart.Atria | Cardiovascular | 7.10E-08 | 5.03E-08 | 0.079221 |
| TDP-43 (any/none) | A07.541.510.110.Aortic.Valve | Cardiovascular | 2.65E-08 | 4.61E-08 | 0.282307 |
| TDP-43 (any/none) | A07.541.560.Heart.Ventricles | Cardiovascular | 1.90E-08 | 4.87E-08 | 0.347810 |
| TDP-43 (any/none) | A07.541.Heart | Cardiovascular | -4.21E-09 | 4.72E-08 | 0.535545 |
| TDP-43 (any/none) | Artery_Aorta | Cardiovascular | -5.65E-08 | 4.59E-08 | 0.890970 |
| TDP-43 (any/none) | Artery_Coronary | Cardiovascular | -3.00E-08 | 4.65E-08 | 0.740433 |
| TDP-43 (any/none) | Artery_Tibial | Cardiovascular | -8.48E-09 | 4.57E-08 | 0.573608 |
| TDP-43 (any/none) | Heart_Atrial_Appendage | Cardiovascular | -1.15E-08 | 4.69E-08 | 0.596543 |
| TDP-43 (any/none) | Heart_Left_Ventricle | Cardiovascular | 1.63E-08 | 4.51E-08 | 0.358683 |
| TDP-43 (any/none) | A08.186.211.132.810.428.200.Cerebellum | CNS | 4.73E-08 | 5.33E-08 | 0.187592 |
| TDP-43 (any/none) | A08.186.211.132.Brain.Stem | CNS | 8.69E-08 | 5.19E-08 | 0.046945 |
| TDP-43 (any/none) | A08.186.211.464.405.Hippocampus | CNS | -3.19E-08 | 4.34E-08 | 0.768667 |
| TDP-43 (any/none) | A08.186.211.464.710.225.Entorhinal.Cortex | CNS | -2.79E-08 | 4.51E-08 | 0.731810 |
| TDP-43 (any/none) | A08.186.211.464.Limbic.System | CNS | -1.86E-08 | 4.57E-08 | 0.658119 |
| TDP-43 (any/none) | A08.186.211.653.Mesencephalon | CNS | -1.79E-08 | 4.97E-08 | 0.640735 |
| TDP-43 (any/none) | A08.186.211.730.317.357.352.435.Hypothalamo.Hypophyseal.System | CNS | 1.36E-08 | 5.68E-08 | 0.405310 |
| TDP-43 (any/none) | A08.186.211.730.317.357.Hypothalamus | CNS | -4.34E-09 | 5.90E-08 | 0.529351 |
| TDP-43 (any/none) | A08.186.211.730.317.Diencephalon | CNS | 1.55E-09 | 5.00E-08 | 0.487677 |
| TDP-43 (any/none) | A08.186.211.730.885.287.249.487.Corpus.Striatum | CNS | 1.18E-08 | 4.29E-08 | 0.391942 |
| TDP-43 (any/none) | A08.186.211.730.885.287.249.Basal.Ganglia | CNS | -2.59E-09 | 4.45E-08 | 0.523178 |

| | | | | | |
|---|---|---|---|---|---|
| TDP-43 (any/none) | A08.186.211.730.885.287.500.270.Frontal.Lobe | CNS | 2.72E-08 | 4.93E-08 | 0.290721 |
| TDP-43 (any/none) | A08.186.211.730.885.287.500.571.735.Visual.Cortex | CNS | 1.66E-08 | 4.47E-08 | 0.355453 |
| TDP-43 (any/none) | A08.186.211.730.885.287.500.670.Parietal.Lobe | CNS | 7.48E-09 | 4.96E-08 | 0.440048 |
| TDP-43 (any/none) | A08.186.211.730.885.287.500.Cerebral.Cortex | CNS | -2.29E-08 | 4.55E-08 | 0.692575 |
| TDP-43 (any/none) | A08.186.211.865.428.Metencephalon | CNS | 5.74E-08 | 5.40E-08 | 0.144066 |
| TDP-43 (any/none) | A08.186.211.Brain | CNS | -4.05E-09 | 4.78E-08 | 0.533762 |
| TDP-43 (any/none) | A09.371.729.Retina | CNS | 5.38E-08 | 4.84E-08 | 0.133109 |
| TDP-43 (any/none) | A11.872.653.Neural.Stem.Cells | CNS | 8.13E-08 | 5.81E-08 | 0.080797 |
| TDP-43 (any/none) | Brain_Amygdala | CNS | 1.72E-08 | 4.53E-08 | 0.352365 |
| TDP-43 (any/none) | Brain_Anterior_cingulate_cortex_(BA24) | CNS | -6.60E-09 | 4.39E-08 | 0.559731 |
| TDP-43 (any/none) | Brain_Caudate_(basal_ganglia) | CNS | -2.83E-09 | 4.10E-08 | 0.527504 |
| TDP-43 (any/none) | Brain_Cerebellar_Hemisphere | CNS | 5.39E-08 | 4.62E-08 | 0.121532 |
| TDP-43 (any/none) | Brain_Cerebellum | CNS | 4.59E-08 | 4.69E-08 | 0.163932 |
| TDP-43 (any/none) | Brain_Cortex | CNS | 1.57E-08 | 4.53E-08 | 0.364760 |
| TDP-43 (any/none) | Brain_Frontal_Cortex_(BA9) | CNS | -1.71E-08 | 4.28E-08 | 0.655105 |
| TDP-43 (any/none) | Brain_Hippocampus | CNS | -1.69E-08 | 4.30E-08 | 0.652820 |
| TDP-43 (any/none) | Brain_Hypothalamus | CNS | -2.97E-08 | 4.29E-08 | 0.755260 |
| TDP-43 (any/none) | Brain_Nucleus_accumbens_(basal_ganglia) | CNS | 5.83E-10 | 4.35E-08 | 0.494653 |
| TDP-43 (any/none) | Brain_Putamen_(basal_ganglia) | CNS | -2.17E-08 | 4.22E-08 | 0.696393 |
| TDP-43 (any/none) | Brain_Spinal_cord_(cervical_c-1) | CNS | 2.26E-08 | 4.44E-08 | 0.305615 |
| TDP-43 (any/none) | Brain_Substantia_nigra | CNS | 3.72E-08 | 4.11E-08 | 0.182736 |
| TDP-43 (any/none) | A03.556.124.369.Intestinal.Mucosa | Digestive | 4.66E-08 | 5.34E-08 | 0.191387 |
| TDP-43 (any/none) | A03.556.124.526.767.Rectum | Digestive | 3.79E-08 | 5.29E-08 | 0.236764 |
| TDP-43 (any/none) | A03.556.124.684.Intestine..Small | Digestive | -1.15E-08 | 5.51E-08 | 0.582521 |
| TDP-43 (any/none) | A03.556.124.Intestines | Digestive | 4.14E-08 | 4.92E-08 | 0.200185 |
| TDP-43 (any/none) | A03.556.249.124.Ileum | Digestive | 3.38E-08 | 6.33E-08 | 0.296534 |
| TDP-43 (any/none) | A03.556.249.249.209.Cecum | Digestive | 2.79E-09 | 4.70E-08 | 0.476302 |
| TDP-43 (any/none) | A03.556.249.249.356.668.Colon..Sigmoid | Digestive | -1.20E-09 | 7.09E-08 | 0.506773 |
| TDP-43 (any/none) | A03.556.249.249.356.Colon | Digestive | 1.62E-08 | 5.16E-08 | 0.377145 |
| TDP-43 (any/none) | A03.556.500.760.464.Parotid.Gland | Digestive | 5.12E-08 | 5.60E-08 | 0.180177 |
| TDP-43 (any/none) | A03.556.500.760.Salivary.Glands | Digestive | 5.28E-08 | 5.40E-08 | 0.164270 |
| TDP-43 (any/none) | A03.556.875.500.Esophagus | Digestive | 3.05E-08 | 5.22E-08 | 0.279860 |
| TDP-43 (any/none) | A03.556.875.875.Stomach | Digestive | 6.32E-08 | 5.49E-08 | 0.124844 |
| TDP-43 (any/none) | A03.556.875.Upper.Gastrointestinal.Tract | Digestive | 6.37E-08 | 5.37E-08 | 0.117787 |
| TDP-43 (any/none) | A03.556.Gastrointestinal.Tract | Digestive | 2.67E-08 | 5.45E-08 | 0.311709 |
| TDP-43 (any/none) | Colon_Sigmoid | Digestive | 2.19E-08 | 5.07E-08 | 0.332694 |
| TDP-43 (any/none) | Colon_Transverse | Digestive | 5.45E-08 | 4.97E-08 | 0.136217 |
| TDP-43 (any/none) | Esophagus_Gastroesophageal_Junction | Digestive | -5.72E-08 | 5.17E-08 | 0.865541 |
| TDP-43 (any/none) | Esophagus_Mucosa | Digestive | -1.78E-08 | 4.71E-08 | 0.646880 |
| TDP-43 (any/none) | Esophagus_Muscularis | Digestive | -4.11E-08 | 5.21E-08 | 0.785335 |
| TDP-43 (any/none) | Small_Intestine_Terminal_Ileum | Digestive | 3.64E-08 | 5.49E-08 | 0.253761 |
| TDP-43 (any/none) | Stomach | Digestive | 9.98E-09 | 4.71E-08 | 0.416090 |
| TDP-43 (any/none) | A03.620.Liver | Liver | -2.35E-08 | 5.36E-08 | 0.669693 |
| TDP-43 (any/none) | A11.436.348.Hepatocytes | Liver | -8.59E-08 | 6.01E-08 | 0.923582 |
| TDP-43 (any/none) | Liver | Liver | 5.49E-08 | 5.17E-08 | 0.144312 |
| TDP-43 (any/none) | A02.165.Cartilage | Musculoskeletal/Connective | -6.65E-10 | 5.40E-08 | 0.504911 |

| | | | | | |
|---|---|---|---|---|---|
| TDP-43 (any/none) | A02.633.567.850.Quadriceps.Muscle | Musculoskeletal/Connective | -9.07E-08 | 4.85E-08 | 0.969250 |
| TDP-43 (any/none) | A02.835.232.834.151.Cervical.Vertebrae | Musculoskeletal/Connective | -1.71E-07 | 4.83E-08 | 0.999800 |
| TDP-43 (any/none) | A02.835.583.443.800.Synovial.Membrane | Musculoskeletal/Connective | -1.23E-08 | 5.35E-08 | 0.590720 |
| TDP-43 (any/none) | A05.360.319.679.690.Myometrium | Musculoskeletal/Connective | 4.33E-08 | 4.79E-08 | 0.183109 |
| TDP-43 (any/none) | A10.165.450.300.425.Keloid | Musculoskeletal/Connective | 1.38E-08 | 5.05E-08 | 0.392342 |
| TDP-43 (any/none) | A10.165.450.300.Cicatrix | Musculoskeletal/Connective | 4.25E-08 | 5.39E-08 | 0.214747 |
| TDP-43 (any/none) | A10.690.467.Muscle..Smooth | Musculoskeletal/Connective | -3.03E-08 | 5.06E-08 | 0.725692 |
| TDP-43 (any/none) | A10.690.Muscles | Musculoskeletal/Connective | -5.68E-08 | 4.68E-08 | 0.887894 |
| TDP-43 (any/none) | A11.329.171.Chondrocytes | Musculoskeletal/Connective | 5.11E-09 | 5.62E-08 | 0.463810 |
| TDP-43 (any/none) | A11.329.228.Fibroblasts | Musculoskeletal/Connective | 5.67E-08 | 5.30E-08 | 0.142027 |
| TDP-43 (any/none) | A11.329.629.Osteoblasts | Musculoskeletal/Connective | 4.07E-09 | 5.06E-08 | 0.467983 |
| TDP-43 (any/none) | A11.329.830.Stromal.Cells | Musculoskeletal/Connective | -3.49E-08 | 4.95E-08 | 0.759223 |
| TDP-43 (any/none) | A11.329.Connective.Tissue.Cells | Musculoskeletal/Connective | -8.31E-09 | 5.58E-08 | 0.559238 |
| TDP-43 (any/none) | A11.620.520.Myocytes..Smooth.Muscle | Musculoskeletal/Connective | -1.69E-09 | 5.05E-08 | 0.513332 |
| TDP-43 (any/none) | Cells_Transformed_fibroblasts | Musculoskeletal/Connective | 4.68E-08 | 4.86E-08 | 0.167660 |
| TDP-43 (any/none) | Muscle_Skeletal | Musculoskeletal/Connective | -5.04E-08 | 4.59E-08 | 0.863655 |
| TDP-43 (any/none) | A04.411.Lung | Other | 1.06E-08 | 5.49E-08 | 0.423346 |
| TDP-43 (any/none) | A04.531.520.Nasal.Mucosa | Other | -3.09E-08 | 5.66E-08 | 0.707168 |
| TDP-43 (any/none) | A05.360.319.114.373.Fallopian.Tubes | Other | 3.52E-09 | 5.14E-08 | 0.472685 |
| TDP-43 (any/none) | A05.360.319.114.630.Ovary | Other | -3.65E-08 | 4.83E-08 | 0.774906 |
| TDP-43 (any/none) | A05.360.319.679.256.Cervix.Uteri | Other | -7.07E-08 | 5.65E-08 | 0.894536 |
| TDP-43 (any/none) | A05.360.319.679.490.Endometrium | Other | 1.84E-09 | 5.62E-08 | 0.486954 |
| TDP-43 (any/none) | A05.360.319.679.Uterus | Other | 3.59E-09 | 5.66E-08 | 0.474734 |
| TDP-43 (any/none) | A05.360.319.887.Vulva | Other | 1.56E-08 | 5.39E-08 | 0.386016 |
| TDP-43 (any/none) | A05.360.319.Genitalia..Female | Other | -2.34E-08 | 5.60E-08 | 0.661990 |
| TDP-43 (any/none) | A05.360.444.492.362.Foreskin | Other | -7.61E-08 | 5.33E-08 | 0.923321 |
| TDP-43 (any/none) | A05.360.444.Genitalia..Male | Other | -2.72E-08 | 4.94E-08 | 0.709396 |
| TDP-43 (any/none) | A05.360.490.Germ.Cells | Other | 4.85E-08 | 4.72E-08 | 0.151611 |
| TDP-43 (any/none) | A05.360.Genitalia | Other | -6.19E-08 | 4.87E-08 | 0.897877 |
| TDP-43 (any/none) | A05.810.453.324.Kidney.Cortex | Other | -1.09E-08 | 4.81E-08 | 0.590061 |
| TDP-43 (any/none) | A05.810.453.Kidney | Other | 1.26E-08 | 5.05E-08 | 0.401483 |
| TDP-43 (any/none) | A05.810.890.Urinary.Bladder | Other | -1.07E-09 | 5.70E-08 | 0.507477 |
| TDP-43 (any/none) | A06.407.071.140.Adrenal.Cortex | Other | 3.24E-08 | 5.70E-08 | 0.285265 |
| TDP-43 (any/none) | A06.407.071.Adrenal.Glands | Other | 5.20E-08 | 6.20E-08 | 0.201103 |
| TDP-43 (any/none) | A06.407.312.782.Testis | Other | -1.18E-07 | 4.95E-08 | 0.991272 |
| TDP-43 (any/none) | A06.407.312.Gonads | Other | -4.34E-08 | 4.79E-08 | 0.817505 |
| TDP-43 (any/none) | A06.407.900.Thyroid.Gland | Other | -1.31E-10 | 5.34E-08 | 0.500976 |
| TDP-43 (any/none) | A06.407.Endocrine.Glands | Other | 9.05E-09 | 5.08E-08 | 0.429333 |
| TDP-43 (any/none) | A09.371.Eye | Other | 3.18E-09 | 5.03E-08 | 0.474827 |
| TDP-43 (any/none) | A10.272.497.Epidermis | Other | 8.14E-09 | 6.10E-08 | 0.446942 |
| TDP-43 (any/none) | A10.272.Epithelium | Other | -4.68E-08 | 5.60E-08 | 0.798465 |
| TDP-43 (any/none) | A10.336.707.Prostate | Other | -3.31E-08 | 5.04E-08 | 0.743939 |
| TDP-43 (any/none) | A10.615.284.473.Chorion | Other | 1.51E-10 | 4.79E-08 | 0.498740 |
| TDP-43 (any/none) | A10.615.550.599.Mouth.Mucosa | Other | -8.40E-08 | 5.70E-08 | 0.929922 |
| TDP-43 (any/none) | A10.615.550.Mucous.Membrane | Other | -4.71E-08 | 5.54E-08 | 0.802257 |
| TDP-43 (any/none) | A10.615.789.Serous.Membrane | Other | -4.42E-08 | 5.00E-08 | 0.811230 |

| | | | | | |
|---|---|---|---|---|---|
| TDP-43 (any/none) | A10.615.Membranes | Other | -3.06E-08 | 5.31E-08 | 0.717821 |
| TDP-43 (any/none) | A11.382.Endocrine.Cells | Other | -5.76E-08 | 4.95E-08 | 0.877499 |
| TDP-43 (any/none) | A11.436.275.Endothelial.Cells | Other | 1.42E-07 | 5.64E-08 | 0.005909 |
| TDP-43 (any/none) | A11.436.294.064.Glucagon.Secreting.Cells | Other | -5.20E-08 | 4.94E-08 | 0.853717 |
| TDP-43 (any/none) | A11.436.329.Granulosa.Cells | Other | -2.58E-08 | 4.69E-08 | 0.708871 |
| TDP-43 (any/none) | A11.436.397.Keratinocytes | Other | 5.79E-08 | 5.58E-08 | 0.149941 |
| TDP-43 (any/none) | A11.436.Epithelial.Cells | Other | 3.38E-08 | 5.23E-08 | 0.258656 |
| TDP-43 (any/none) | A11.497.497.600.Oocytes | Other | 2.26E-08 | 4.88E-08 | 0.321846 |
| TDP-43 (any/none) | A11.872.040.Adult.Stem.Cells | Other | 7.40E-08 | 4.94E-08 | 0.066866 |
| TDP-43 (any/none) | A11.872.190.260.Embryoid.Bodies | Other | -6.98E-08 | 5.56E-08 | 0.895373 |
| TDP-43 (any/none) | A11.872.190.Embryonic.Stem.Cells | Other | 1.58E-08 | 5.06E-08 | 0.377626 |
| TDP-43 (any/none) | A11.872.580.Mesenchymal.Stem.Cells | Other | -1.93E-08 | 5.70E-08 | 0.632643 |
| TDP-43 (any/none) | A11.872.700.500.Induced.Pluripotent.Stem.Cells | Other | 2.92E-08 | 5.57E-08 | 0.300108 |
| TDP-43 (any/none) | A11.872.Stem.Cells | Other | -2.92E-08 | 5.67E-08 | 0.696945 |
| TDP-43 (any/none) | A14.549.167.646.Periodontium | Other | 1.47E-08 | 5.91E-08 | 0.401726 |
| TDP-43 (any/none) | A14.549.167.Dentition | Other | -1.45E-08 | 5.41E-08 | 0.605416 |
| TDP-43 (any/none) | A14.549.885.Tongue | Other | -6.00E-09 | 6.35E-08 | 0.537631 |
| TDP-43 (any/none) | A14.549.Mouth | Other | -5.48E-08 | 5.59E-08 | 0.836529 |
| TDP-43 (any/none) | A14.724.557.Nasopharynx | Other | -3.14E-08 | 5.31E-08 | 0.723304 |
| TDP-43 (any/none) | A14.724.Pharynx | Other | -4.22E-08 | 5.02E-08 | 0.799765 |
| TDP-43 (any/none) | A17.815.Skin | Other | -4.79E-08 | 6.05E-08 | 0.785955 |
| TDP-43 (any/none) | Adrenal_Gland | Other | 1.00E-08 | 5.03E-08 | 0.420868 |
| TDP-43 (any/none) | Bladder | Other | -8.43E-08 | 4.30E-08 | 0.975035 |
| TDP-43 (any/none) | Breast_Mammary_Tissue | Other | 7.73E-08 | 4.59E-08 | 0.046203 |
| TDP-43 (any/none) | Cervix_Ectocervix | Other | 1.03E-07 | 5.45E-08 | 0.029781 |
| TDP-43 (any/none) | Cervix_Endocervix | Other | 5.50E-08 | 5.23E-08 | 0.146569 |
| TDP-43 (any/none) | Fallopian_Tube | Other | -1.96E-08 | 5.03E-08 | 0.651746 |
| TDP-43 (any/none) | Kidney_Cortex | Other | -8.45E-10 | 4.73E-08 | 0.507124 |
| TDP-43 (any/none) | Lung | Other | -4.32E-08 | 4.83E-08 | 0.814540 |
| TDP-43 (any/none) | Minor_Salivary_Gland | Other | 7.21E-08 | 5.15E-08 | 0.080618 |
| TDP-43 (any/none) | Nerve_Tibial | Other | 2.00E-08 | 4.20E-08 | 0.317027 |
| TDP-43 (any/none) | Ovary | Other | -4.95E-08 | 4.99E-08 | 0.839254 |
| TDP-43 (any/none) | Pituitary | Other | 4.94E-08 | 5.07E-08 | 0.164549 |
| TDP-43 (any/none) | Prostate | Other | 3.06E-08 | 5.15E-08 | 0.276300 |
| TDP-43 (any/none) | Skin_Not_Sun_Exposed_(Suprapubic) | Other | 3.98E-08 | 5.36E-08 | 0.229115 |
| TDP-43 (any/none) | Skin_Sun_Exposed_(Lower_leg) | Other | 2.54E-08 | 5.15E-08 | 0.310805 |
| TDP-43 (any/none) | Testis | Other | -5.29E-08 | 5.07E-08 | 0.851816 |
| TDP-43 (any/none) | Thyroid | Other | 4.22E-08 | 5.48E-08 | 0.220440 |
| TDP-43 (any/none) | Uterus | Other | 3.47E-08 | 5.29E-08 | 0.255991 |
| TDP-43 (any/none) | Vagina | Other | 4.37E-08 | 4.96E-08 | 0.189444 |
| TDP-43 (any/none) | A03.734.414.Islets.of.Langerhans | Pancreas | -5.26E-08 | 5.20E-08 | 0.844168 |
| TDP-43 (any/none) | A03.734.Pancreas | Pancreas | 2.69E-08 | 4.70E-08 | 0.283620 |
| TDP-43 (any/none) | Pancreas | Pancreas | -2.80E-08 | 4.71E-08 | 0.724037 |
| Thal Phase | A10.165.114.830.500.750.Subcutaneous.Fat..Abdominal | Adipose | -8.28E-09 | 7.80E-08 | 0.542278 |
| Thal Phase | A10.165.114.830.750.Subcutaneous.Fat | Adipose | -4.72E-09 | 7.52E-08 | 0.525002 |
| Thal Phase | A11.329.114.Adipocytes | Adipose | -5.84E-08 | 7.23E-08 | 0.790284 |

| | | | | | |
|---|---|---|---|---|---|
| Thal Phase | Adipose_Subcutaneous | Adipose | 3.48E-08 | 6.85E-08 | 0.305603 |
| Thal Phase | Adipose_Visceral_(Omentum) | Adipose | -1.86E-08 | 6.71E-08 | 0.608907 |
| Thal Phase | A02.835.583.443.800.800.Synovial.Fluid | Blood/Immune | -1.18E-08 | 8.96E-08 | 0.552421 |
| Thal Phase | A10.549.400.Lymph.Nodes | Blood/Immune | -7.83E-08 | 7.36E-08 | 0.856333 |
| Thal Phase | A10.549.Lymphoid.Tissue | Blood/Immune | -5.95E-08 | 7.61E-08 | 0.782879 |
| Thal Phase | A11.118.637.555.567.562.440.Precursor.Cells..B.Lymphoid | Blood/Immune | 1.15E-08 | 7.86E-08 | 0.441640 |
| Thal Phase | A11.118.637.555.567.562.B.Lymphocytes | Blood/Immune | -8.26E-08 | 7.79E-08 | 0.855533 |
| Thal Phase | A11.118.637.555.567.569.200.700.T.Lymphocytes..Regulatory | Blood/Immune | -4.38E-08 | 7.25E-08 | 0.726988 |
| Thal Phase | A11.118.637.555.567.569.T.Lymphocytes | Blood/Immune | -1.28E-07 | 7.57E-08 | 0.955078 |
| Thal Phase | A11.118.637.Leukocytes | Blood/Immune | 4.02E-08 | 8.14E-08 | 0.310770 |
| Thal Phase | A11.329.372.600.Macrophages..Alveolar | Blood/Immune | 6.17E-08 | 7.20E-08 | 0.195835 |
| Thal Phase | A11.443.Erythroid.Cells | Blood/Immune | 4.21E-08 | 7.62E-08 | 0.290351 |
| Thal Phase | A11.627.340.360.Granulocyte.Precursor.Cells | Blood/Immune | -5.67E-08 | 7.53E-08 | 0.774083 |
| Thal Phase | A11.627.624.249.Monocyte.Macrophage.Precursor.Cells | Blood/Immune | -1.35E-09 | 7.46E-08 | 0.507224 |
| Thal Phase | A11.627.635.Myeloid.Progenitor.Cells | Blood/Immune | 5.04E-08 | 7.59E-08 | 0.253524 |
| Thal Phase | A11.872.378.590.635.Granulocyte.Macrophage.Progenitor.Cells | Blood/Immune | 3.51E-08 | 7.72E-08 | 0.324546 |
| Thal Phase | A11.872.378.590.817.Megakaryocyte.Erythroid.Progenitor.Cells | Blood/Immune | 1.28E-07 | 7.89E-08 | 0.052515 |
| Thal Phase | A11.872.378.Hematopoietic.Stem.Cells | Blood/Immune | 3.11E-08 | 8.25E-08 | 0.353157 |
| Thal Phase | A15.145.229.188.Blood.Platelets | Blood/Immune | -5.58E-08 | 6.70E-08 | 0.797350 |
| Thal Phase | A15.145.229.637.555.567.562.725.Plasma.Cells | Blood/Immune | -8.01E-08 | 8.45E-08 | 0.828576 |
| Thal Phase | A15.145.229.637.555.567.569.200.CD4.Positive.T.Lymphocytes | Blood/Immune | -8.81E-08 | 7.09E-08 | 0.892845 |
| Thal Phase | A15.145.229.637.555.Leukocytes..Mononuclear | Blood/Immune | -6.93E-08 | 8.33E-08 | 0.797521 |
| Thal Phase | A15.145.229.Blood.Cells | Blood/Immune | -8.84E-08 | 8.57E-08 | 0.849050 |
| Thal Phase | A15.145.300.Fetal.Blood | Blood/Immune | 6.50E-08 | 9.17E-08 | 0.239186 |
| Thal Phase | A15.145.846.Serum | Blood/Immune | -6.73E-09 | 7.26E-08 | 0.536963 |
| Thal Phase | A15.145.Blood | Blood/Immune | -9.59E-08 | 9.15E-08 | 0.852613 |
| Thal Phase | A15.378.316.580.Monocytes | Blood/Immune | 1.04E-07 | 8.95E-08 | 0.122305 |
| Thal Phase | A15.378.316.Bone.Marrow.Cells | Blood/Immune | 1.59E-08 | 7.93E-08 | 0.420308 |
| Thal Phase | A15.382.490.315.583.Neutrophils | Blood/Immune | -1.14E-07 | 8.09E-08 | 0.920983 |
| Thal Phase | A15.382.490.555.567.537.Killer.Cells..Natural | Blood/Immune | -1.08E-07 | 8.15E-08 | 0.907824 |
| Thal Phase | A15.382.490.555.567.622.Lymphocytes..Null | Blood/Immune | -6.24E-08 | 9.31E-08 | 0.748787 |
| Thal Phase | A15.382.490.555.567.Lymphocytes | Blood/Immune | -1.44E-07 | 7.48E-08 | 0.973097 |
| Thal Phase | A15.382.520.604.700.Spleen | Blood/Immune | -7.17E-08 | 8.03E-08 | 0.814022 |
| Thal Phase | A15.382.520.604.800.Palatine.Tonsil | Blood/Immune | 5.63E-08 | 8.51E-08 | 0.254155 |
| Thal Phase | A15.382.680.Phagocytes | Blood/Immune | 7.24E-08 | 8.18E-08 | 0.188047 |
| Thal Phase | A15.382.812.260.Dendritic.Cells | Blood/Immune | 5.22E-08 | 8.33E-08 | 0.265442 |
| Thal Phase | A15.382.812.522.Macrophages | Blood/Immune | 3.17E-08 | 7.84E-08 | 0.342879 |
| Thal Phase | A15.382.812.Mononuclear.Phagocyte.System | Blood/Immune | 9.84E-08 | 8.34E-08 | 0.118961 |
| Thal Phase | A15.382.Immune.System | Blood/Immune | -1.44E-08 | 8.06E-08 | 0.571056 |
| Thal Phase | Cells_EBV-transformed_lymphocytes | Blood/Immune | -1.71E-08 | 7.28E-08 | 0.593059 |
| Thal Phase | Spleen | Blood/Immune | 4.48E-08 | 8.11E-08 | 0.290214 |
| Thal Phase | Whole_Blood | Blood/Immune | -1.14E-07 | 7.73E-08 | 0.930075 |
| Thal Phase | A07.231.114.Arteries | Cardiovascular | 5.59E-08 | 7.15E-08 | 0.216955 |
| Thal Phase | A07.231.908.670.874.Umbilical.Veins | Cardiovascular | -6.40E-08 | 7.45E-08 | 0.804684 |
| Thal Phase | A07.231.908.Veins | Cardiovascular | -6.92E-08 | 7.50E-08 | 0.821999 |
| Thal Phase | A07.231.Blood.Vessels | Cardiovascular | -5.85E-09 | 7.80E-08 | 0.529913 |

| | | | | | |
|---|---|---|---|---|---|
| Thal Phase | A07.541.358.100.Atrial.Appendage | Cardiovascular | -7.69E-08 | 5.95E-08 | 0.901696 |
| Thal Phase | A07.541.358.Heart.Atria | Cardiovascular | -9.65E-08 | 5.78E-08 | 0.952491 |
| Thal Phase | A07.541.510.110.Aortic.Valve | Cardiovascular | 2.01E-08 | 6.19E-08 | 0.372968 |
| Thal Phase | A07.541.560.Heart.Ventricles | Cardiovascular | -5.59E-08 | 6.65E-08 | 0.799497 |
| Thal Phase | A07.541.Heart | Cardiovascular | -4.99E-08 | 6.77E-08 | 0.769533 |
| Thal Phase | Artery_Aorta | Cardiovascular | 5.87E-10 | 6.01E-08 | 0.496109 |
| Thal Phase | Artery_Coronary | Cardiovascular | -1.16E-08 | 6.68E-08 | 0.568925 |
| Thal Phase | Artery_Tibial | Cardiovascular | 6.87E-08 | 5.63E-08 | 0.110946 |
| Thal Phase | Heart_Atrial_Appendage | Cardiovascular | -4.43E-08 | 5.64E-08 | 0.783559 |
| Thal Phase | Heart_Left_Ventricle | Cardiovascular | -8.52E-08 | 5.93E-08 | 0.924506 |
| Thal Phase | A08.186.211.132.810.428.200.Cerebellum | CNS | 3.16E-08 | 5.73E-08 | 0.290234 |
| Thal Phase | A08.186.211.132.Brain.Stem | CNS | 2.85E-08 | 5.61E-08 | 0.305708 |
| Thal Phase | A08.186.211.464.405.Hippocampus | CNS | -1.21E-09 | 5.68E-08 | 0.508468 |
| Thal Phase | A08.186.211.464.710.225.Entorhinal.Cortex | CNS | -7.64E-08 | 6.05E-08 | 0.896764 |
| Thal Phase | A08.186.211.464.Limbic.System | CNS | -7.71E-08 | 6.01E-08 | 0.900260 |
| Thal Phase | A08.186.211.653.Mesencephalon | CNS | 1.94E-08 | 6.62E-08 | 0.384999 |
| Thal Phase | A08.186.211.730.317.357.352.435.Hypothalamo.Hypophyseal.System | CNS | -5.77E-08 | 7.90E-08 | 0.767504 |
| Thal Phase | A08.186.211.730.317.357.Hypothalamus | CNS | -6.36E-08 | 7.86E-08 | 0.790911 |
| Thal Phase | A08.186.211.730.317.Diencephalon | CNS | -5.87E-08 | 6.94E-08 | 0.801076 |
| Thal Phase | A08.186.211.730.885.287.249.487.Corpus.Striatum | CNS | 1.24E-08 | 6.34E-08 | 0.422623 |
| Thal Phase | A08.186.211.730.885.287.249.Basal.Ganglia | CNS | 4.32E-09 | 6.85E-08 | 0.474862 |
| Thal Phase | A08.186.211.730.885.287.500.270.Frontal.Lobe | CNS | -9.33E-08 | 6.23E-08 | 0.932869 |
| Thal Phase | A08.186.211.730.885.287.500.571.735.Visual.Cortex | CNS | -7.33E-08 | 6.02E-08 | 0.888354 |
| Thal Phase | A08.186.211.730.885.287.500.670.Parietal.Lobe | CNS | 2.76E-08 | 6.83E-08 | 0.342874 |
| Thal Phase | A08.186.211.730.885.287.500.Cerebral.Cortex | CNS | -9.15E-08 | 5.70E-08 | 0.945902 |
| Thal Phase | A08.186.211.865.428.Metencephalon | CNS | 3.58E-08 | 5.80E-08 | 0.268600 |
| Thal Phase | A08.186.211.Brain | CNS | -3.65E-08 | 5.77E-08 | 0.736110 |
| Thal Phase | A09.371.729.Retina | CNS | -1.03E-08 | 6.49E-08 | 0.563349 |
| Thal Phase | A11.872.653.Neural.Stem.Cells | CNS | 1.01E-07 | 8.30E-08 | 0.111625 |
| Thal Phase | Brain_Amygdala | CNS | -6.15E-08 | 5.87E-08 | 0.852587 |
| Thal Phase | Brain_Anterior_cingulate_cortex_(BA24) | CNS | -8.93E-08 | 5.46E-08 | 0.949220 |
| Thal Phase | Brain_Caudate_(basal_ganglia) | CNS | -7.13E-08 | 5.68E-08 | 0.895222 |
| Thal Phase | Brain_Cerebellar_Hemisphere | CNS | 7.49E-08 | 5.47E-08 | 0.085447 |
| Thal Phase | Brain_Cerebellum | CNS | 9.26E-09 | 5.47E-08 | 0.432753 |
| Thal Phase | Brain_Cortex | CNS | -7.64E-08 | 5.36E-08 | 0.923211 |
| Thal Phase | Brain_Frontal_Cortex_(BA9) | CNS | -6.26E-08 | 5.21E-08 | 0.885444 |
| Thal Phase | Brain_Hippocampus | CNS | -5.38E-08 | 5.82E-08 | 0.822278 |
| Thal Phase | Brain_Hypothalamus | CNS | -6.41E-08 | 5.67E-08 | 0.871105 |
| Thal Phase | Brain_Nucleus_accumbens_(basal_ganglia) | CNS | -8.49E-08 | 5.71E-08 | 0.931508 |
| Thal Phase | Brain_Putamen_(basal_ganglia) | CNS | -5.36E-08 | 6.06E-08 | 0.811877 |
| Thal Phase | Brain_Spinal_cord_(cervical_c-1) | CNS | -2.23E-08 | 5.92E-08 | 0.646717 |
| Thal Phase | Brain_Substantia_nigra | CNS | -1.01E-07 | 5.84E-08 | 0.958006 |
| Thal Phase | A03.556.124.369.Intestinal.Mucosa | Digestive | -5.22E-08 | 7.49E-08 | 0.757133 |
| Thal Phase | A03.556.124.526.767.Rectum | Digestive | 1.06E-07 | 7.60E-08 | 0.081149 |
| Thal Phase | A03.556.124.684.Intestine..Small | Digestive | 4.97E-08 | 7.60E-08 | 0.256715 |
| Thal Phase | A03.556.124.Intestines | Digestive | -5.69E-08 | 7.25E-08 | 0.783608 |

| | | | | | |
|---|---|---|---|---|---|
| Thal Phase | A03.556.249.124.Ileum | Digestive | -1.10E-08 | 7.92E-08 | 0.555343 |
| Thal Phase | A03.556.249.249.209.Cecum | Digestive | -5.47E-08 | 7.24E-08 | 0.775225 |
| Thal Phase | A03.556.249.249.356.668.Colon..Sigmoid | Digestive | -7.99E-08 | 7.54E-08 | 0.855557 |
| Thal Phase | A03.556.249.249.356.Colon | Digestive | -7.17E-08 | 7.69E-08 | 0.824603 |
| Thal Phase | A03.556.500.760.464.Parotid.Gland | Digestive | -1.38E-07 | 7.78E-08 | 0.962421 |
| Thal Phase | A03.556.500.760.Salivary.Glands | Digestive | -9.67E-08 | 7.83E-08 | 0.891379 |
| Thal Phase | A03.556.875.500.Esophagus | Digestive | -8.16E-08 | 7.31E-08 | 0.868123 |
| Thal Phase | A03.556.875.875.Stomach | Digestive | -6.03E-08 | 7.77E-08 | 0.781126 |
| Thal Phase | A03.556.875.Upper.Gastrointestinal.Tract | Digestive | -6.19E-08 | 7.66E-08 | 0.790411 |
| Thal Phase | A03.556.Gastrointestinal.Tract | Digestive | -7.12E-08 | 7.63E-08 | 0.824361 |
| Thal Phase | Colon_Sigmoid | Digestive | -1.43E-08 | 6.49E-08 | 0.586940 |
| Thal Phase | Colon_Transverse | Digestive | -1.12E-07 | 6.67E-08 | 0.952838 |
| Thal Phase | Esophagus_Gastroesophageal_Junction | Digestive | 7.47E-08 | 6.78E-08 | 0.135183 |
| Thal Phase | Esophagus_Mucosa | Digestive | 2.23E-09 | 6.96E-08 | 0.487249 |
| Thal Phase | Esophagus_Muscularis | Digestive | 5.72E-08 | 6.92E-08 | 0.203997 |
| Thal Phase | Small_Intestine_Terminal_Ileum | Digestive | -1.05E-07 | 7.49E-08 | 0.918897 |
| Thal Phase | Stomach | Digestive | -5.49E-08 | 6.73E-08 | 0.792521 |
| Thal Phase | A03.620.Liver | Liver | -2.43E-08 | 8.29E-08 | 0.615506 |
| Thal Phase | A11.436.348.Hepatocytes | Liver | 6.56E-08 | 8.40E-08 | 0.217285 |
| Thal Phase | Liver | Liver | -6.63E-08 | 7.63E-08 | 0.807823 |
| Thal Phase | A02.165.Cartilage | Musculoskeletal/Connective | 3.87E-09 | 6.28E-08 | 0.475432 |
| Thal Phase | A02.633.567.850.Quadriceps.Muscle | Musculoskeletal/Connective | 1.12E-07 | 7.99E-08 | 0.080390 |
| Thal Phase | A02.835.232.834.151.Cervical.Vertebrae | Musculoskeletal/Connective | -5.72E-08 | 7.15E-08 | 0.788041 |
| Thal Phase | A02.835.583.443.800.Synovial.Membrane | Musculoskeletal/Connective | 5.11E-08 | 6.83E-08 | 0.227477 |
| Thal Phase | A05.360.319.679.690.Myometrium | Musculoskeletal/Connective | 5.28E-08 | 6.49E-08 | 0.207910 |
| Thal Phase | A10.165.450.300.425.Keloid | Musculoskeletal/Connective | 5.90E-08 | 6.75E-08 | 0.190925 |
| Thal Phase | A10.165.450.300.Cicatrix | Musculoskeletal/Connective | 7.09E-08 | 6.76E-08 | 0.147358 |
| Thal Phase | A10.690.467.Muscle..Smooth | Musculoskeletal/Connective | 4.80E-08 | 8.02E-08 | 0.274808 |
| Thal Phase | A10.690.Muscles | Musculoskeletal/Connective | 4.53E-08 | 7.86E-08 | 0.282107 |
| Thal Phase | A11.329.171.Chondrocytes | Musculoskeletal/Connective | 2.31E-08 | 7.44E-08 | 0.378073 |
| Thal Phase | A11.329.228.Fibroblasts | Musculoskeletal/Connective | -3.18E-08 | 6.59E-08 | 0.685261 |
| Thal Phase | A11.329.629.Osteoblasts | Musculoskeletal/Connective | 7.87E-08 | 7.20E-08 | 0.137148 |
| Thal Phase | A11.329.830.Stromal.Cells | Musculoskeletal/Connective | -4.11E-08 | 7.47E-08 | 0.708973 |
| Thal Phase | A11.329.Connective.Tissue.Cells | Musculoskeletal/Connective | -2.14E-08 | 7.28E-08 | 0.615398 |
| Thal Phase | A11.620.520.Myocytes..Smooth.Muscle | Musculoskeletal/Connective | -6.08E-08 | 7.65E-08 | 0.786479 |
| Thal Phase | Cells_Transformed_fibroblasts | Musculoskeletal/Connective | -5.90E-08 | 6.79E-08 | 0.807667 |
| Thal Phase | Muscle_Skeletal | Musculoskeletal/Connective | -1.55E-08 | 6.69E-08 | 0.591409 |
| Thal Phase | A04.411.Lung | Other | 1.77E-08 | 7.03E-08 | 0.400813 |
| Thal Phase | A04.531.520.Nasal.Mucosa | Other | -1.67E-08 | 6.64E-08 | 0.599453 |
| Thal Phase | A05.360.319.114.373.Fallopian.Tubes | Other | -2.05E-08 | 7.12E-08 | 0.613302 |
| Thal Phase | A05.360.319.114.630.Ovary | Other | -3.29E-08 | 7.20E-08 | 0.675998 |
| Thal Phase | A05.360.319.679.256.Cervix.Uteri | Other | -6.46E-08 | 7.22E-08 | 0.814669 |
| Thal Phase | A05.360.319.679.490.Endometrium | Other | -3.69E-08 | 6.43E-08 | 0.716857 |
| Thal Phase | A05.360.319.679.Uterus | Other | -1.35E-08 | 6.46E-08 | 0.582608 |
| Thal Phase | A05.360.319.887.Vulva | Other | -4.90E-08 | 7.92E-08 | 0.732135 |
| Thal Phase | A05.360.319.Genitalia..Female | Other | -5.62E-08 | 7.22E-08 | 0.781635 |

| | | | | | |
|---|---|---|---|---|---|
| Thal Phase | A05.360.444.492.362.Foreskin | Other | 3.57E-08 | 7.05E-08 | 0.306293 |
| Thal Phase | A05.360.444.Genitalia..Male | Other | -9.38E-09 | 6.92E-08 | 0.553908 |
| Thal Phase | A05.360.490.Germ.Cells | Other | 7.44E-08 | 6.31E-08 | 0.119204 |
| Thal Phase | A05.360.Genitalia | Other | -1.20E-07 | 6.76E-08 | 0.961866 |
| Thal Phase | A05.810.453.324.Kidney.Cortex | Other | 1.65E-07 | 6.36E-08 | 0.004772 |
| Thal Phase | A05.810.453.Kidney | Other | 3.41E-08 | 7.10E-08 | 0.315253 |
| Thal Phase | A05.810.890.Urinary.Bladder | Other | -8.61E-08 | 7.83E-08 | 0.864202 |
| Thal Phase | A06.407.071.140.Adrenal.Cortex | Other | 1.14E-08 | 7.36E-08 | 0.438550 |
| Thal Phase | A06.407.071.Adrenal.Glands | Other | 1.03E-08 | 7.64E-08 | 0.446597 |
| Thal Phase | A06.407.312.782.Testis | Other | 2.99E-08 | 6.34E-08 | 0.318858 |
| Thal Phase | A06.407.312.Gonads | Other | -9.78E-09 | 7.51E-08 | 0.551832 |
| Thal Phase | A06.407.900.Thyroid.Gland | Other | -6.31E-08 | 6.96E-08 | 0.817705 |
| Thal Phase | A06.407.Endocrine.Glands | Other | 3.45E-09 | 7.01E-08 | 0.480363 |
| Thal Phase | A09.371.Eye | Other | -3.93E-08 | 7.94E-08 | 0.689745 |
| Thal Phase | A10.272.497.Epidermis | Other | -7.84E-08 | 7.61E-08 | 0.848542 |
| Thal Phase | A10.272.Epithelium | Other | -3.57E-08 | 6.55E-08 | 0.707127 |
| Thal Phase | A10.336.707.Prostate | Other | 1.73E-08 | 6.73E-08 | 0.398443 |
| Thal Phase | A10.615.284.473.Chorion | Other | 2.72E-08 | 7.46E-08 | 0.357559 |
| Thal Phase | A10.615.550.599.Mouth.Mucosa | Other | -3.50E-08 | 8.05E-08 | 0.668171 |
| Thal Phase | A10.615.550.Mucous.Membrane | Other | -1.34E-07 | 7.60E-08 | 0.960684 |
| Thal Phase | A10.615.789.Serous.Membrane | Other | 5.53E-08 | 7.32E-08 | 0.225234 |
| Thal Phase | A10.615.Membranes | Other | -8.69E-08 | 7.45E-08 | 0.878390 |
| Thal Phase | A11.382.Endocrine.Cells | Other | 1.08E-07 | 6.69E-08 | 0.053724 |
| Thal Phase | A11.436.275.Endothelial.Cells | Other | -5.38E-08 | 7.21E-08 | 0.772085 |
| Thal Phase | A11.436.294.064.Glucagon.Secreting.Cells | Other | 6.87E-08 | 7.17E-08 | 0.168786 |
| Thal Phase | A11.436.329.Granulosa.Cells | Other | 1.24E-07 | 6.61E-08 | 0.030808 |
| Thal Phase | A11.436.397.Keratinocytes | Other | 6.14E-08 | 7.74E-08 | 0.213574 |
| Thal Phase | A11.436.Epithelial.Cells | Other | -4.34E-08 | 7.27E-08 | 0.724544 |
| Thal Phase | A11.497.497.600.Oocytes | Other | 5.87E-09 | 6.40E-08 | 0.463423 |
| Thal Phase | A11.872.040.Adult.Stem.Cells | Other | -3.06E-08 | 7.39E-08 | 0.660564 |
| Thal Phase | A11.872.190.260.Embryoid.Bodies | Other | -6.28E-08 | 6.88E-08 | 0.819472 |
| Thal Phase | A11.872.190.Embryonic.Stem.Cells | Other | -3.68E-08 | 7.34E-08 | 0.692187 |
| Thal Phase | A11.872.580.Mesenchymal.Stem.Cells | Other | -3.52E-08 | 7.06E-08 | 0.691113 |
| Thal Phase | A11.872.700.500.Induced.Pluripotent.Stem.Cells | Other | -4.27E-08 | 7.50E-08 | 0.715452 |
| Thal Phase | A11.872.Stem.Cells | Other | 4.22E-08 | 7.69E-08 | 0.291610 |
| Thal Phase | A14.549.167.646.Periodontium | Other | -2.40E-08 | 8.07E-08 | 0.616994 |
| Thal Phase | A14.549.167.Dentition | Other | -1.34E-08 | 8.18E-08 | 0.565313 |
| Thal Phase | A14.549.885.Tongue | Other | -1.50E-07 | 7.61E-08 | 0.975690 |
| Thal Phase | A14.549.Mouth | Other | -7.76E-09 | 7.62E-08 | 0.540541 |
| Thal Phase | A14.724.557.Nasopharynx | Other | 1.65E-08 | 6.70E-08 | 0.402661 |
| Thal Phase | A14.724.Pharynx | Other | 6.97E-09 | 6.98E-08 | 0.460225 |
| Thal Phase | A17.815.Skin | Other | -5.12E-08 | 6.89E-08 | 0.771445 |
| Thal Phase | Adrenal_Gland | Other | 2.98E-09 | 6.90E-08 | 0.482798 |
| Thal Phase | Bladder | Other | -2.94E-08 | 6.42E-08 | 0.676730 |
| Thal Phase | Breast_Mammary_Tissue | Other | -4.18E-08 | 6.36E-08 | 0.744656 |
| Thal Phase | Cervix_Ectocervix | Other | -3.75E-08 | 6.96E-08 | 0.704692 |

| | | | | | |
|---|---|---|---|---|---|
| Thal Phase | Cervix_Endocervix | Other | 5.63E-08 | 6.64E-08 | 0.198261 |
| Thal Phase | Fallopian_Tube | Other | -5.78E-08 | 6.49E-08 | 0.813667 |
| Thal Phase | Kidney_Cortex | Other | -3.10E-08 | 6.89E-08 | 0.673730 |
| Thal Phase | Lung | Other | -5.24E-08 | 7.45E-08 | 0.759276 |
| Thal Phase | Minor_Salivary_Gland | Other | -1.38E-07 | 6.40E-08 | 0.984521 |
| Thal Phase | Nerve_Tibial | Other | 1.28E-08 | 6.26E-08 | 0.419107 |
| Thal Phase | Ovary | Other | -1.44E-08 | 6.46E-08 | 0.588489 |
| Thal Phase | Pituitary | Other | -6.22E-10 | 6.70E-08 | 0.503704 |
| Thal Phase | Prostate | Other | -1.67E-07 | 6.93E-08 | 0.992008 |
| Thal Phase | Skin_Not_Sun_Exposed_(Suprapubic) | Other | -1.40E-07 | 6.70E-08 | 0.981533 |
| Thal Phase | Skin_Sun_Exposed_(Lower_leg) | Other | -1.25E-07 | 6.45E-08 | 0.973653 |
| Thal Phase | Testis | Other | 1.26E-07 | 6.60E-08 | 0.027622 |
| Thal Phase | Thyroid | Other | -1.16E-07 | 7.22E-08 | 0.945155 |
| Thal Phase | Uterus | Other | -6.10E-08 | 6.15E-08 | 0.839306 |
| Thal Phase | Vagina | Other | -4.46E-08 | 6.98E-08 | 0.738297 |
| Thal Phase | A03.734.414.Islets.of.Langerhans | Pancreas | 7.91E-08 | 7.62E-08 | 0.149728 |
| Thal Phase | A03.734.Pancreas | Pancreas | -3.81E-08 | 6.63E-08 | 0.717090 |
| Thal Phase | Pancreas | Pancreas | -9.67E-08 | 6.31E-08 | 0.937363 |
| WMR (any/none) | A10.165.114.830.500.750.Subcutaneous.Fat..Abdominal | Adipose | -2.08E-08 | 2.99E-08 | 0.756371 |
| WMR (any/none) | A10.165.114.830.750.Subcutaneous.Fat | Adipose | -2.19E-08 | 3.03E-08 | 0.765194 |
| WMR (any/none) | A11.329.114.Adipocytes | Adipose | -1.99E-08 | 3.08E-08 | 0.741557 |
| WMR (any/none) | Adipose_Subcutaneous | Adipose | -7.39E-09 | 2.97E-08 | 0.598321 |
| WMR (any/none) | Adipose_Visceral_(Omentum) | Adipose | 7.19E-09 | 3.13E-08 | 0.409244 |
| WMR (any/none) | A02.835.583.443.800.800.Synovial.Fluid | Blood/Immune | 9.88E-09 | 3.93E-08 | 0.400670 |
| WMR (any/none) | A10.549.400.Lymph.Nodes | Blood/Immune | -8.06E-08 | 3.29E-08 | 0.992870 |
| WMR (any/none) | A10.549.Lymphoid.Tissue | Blood/Immune | -3.65E-08 | 3.57E-08 | 0.846872 |
| WMR (any/none) | A11.118.637.555.567.562.440.Precursor.Cells..B.Lymphoid | Blood/Immune | -5.12E-09 | 3.74E-08 | 0.554544 |
| WMR (any/none) | A11.118.637.555.567.562.B.Lymphocytes | Blood/Immune | 1.58E-08 | 3.82E-08 | 0.339980 |
| WMR (any/none) | A11.118.637.555.567.569.200.700.T.Lymphocytes..Regulatory | Blood/Immune | 1.26E-08 | 3.52E-08 | 0.360045 |
| WMR (any/none) | A11.118.637.555.567.569.T.Lymphocytes | Blood/Immune | 6.21E-09 | 3.38E-08 | 0.427137 |
| WMR (any/none) | A11.118.637.Leukocytes | Blood/Immune | 1.66E-08 | 3.92E-08 | 0.336041 |
| WMR (any/none) | A11.329.372.600.Macrophages..Alveolar | Blood/Immune | 1.18E-08 | 3.66E-08 | 0.373222 |
| WMR (any/none) | A11.443.Erythroid.Cells | Blood/Immune | 2.37E-08 | 3.42E-08 | 0.244313 |
| WMR (any/none) | A11.627.340.360.Granulocyte.Precursor.Cells | Blood/Immune | 2.66E-08 | 3.66E-08 | 0.234090 |
| WMR (any/none) | A11.627.624.249.Monocyte.Macrophage.Precursor.Cells | Blood/Immune | 1.52E-09 | 3.51E-08 | 0.482713 |
| WMR (any/none) | A11.627.635.Myeloid.Progenitor.Cells | Blood/Immune | 2.63E-08 | 3.71E-08 | 0.239212 |
| WMR (any/none) | A11.872.378.590.635.Granulocyte.Macrophage.Progenitor.Cells | Blood/Immune | 4.79E-09 | 3.81E-08 | 0.449962 |
| WMR (any/none) | A11.872.378.590.817.Megakaryocyte.Erythroid.Progenitor.Cells | Blood/Immune | 3.11E-08 | 3.53E-08 | 0.188808 |
| WMR (any/none) | A11.872.378.Hematopoietic.Stem.Cells | Blood/Immune | 1.74E-08 | 3.56E-08 | 0.312416 |
| WMR (any/none) | A15.145.229.188.Blood.Platelets | Blood/Immune | -4.14E-09 | 3.14E-08 | 0.552488 |
| WMR (any/none) | A15.145.229.637.555.567.562.725.Plasma.Cells | Blood/Immune | 2.85E-08 | 3.44E-08 | 0.203587 |
| WMR (any/none) | A15.145.229.637.555.567.569.200.CD4.Positive.T.Lymphocytes | Blood/Immune | -6.26E-11 | 3.68E-08 | 0.500678 |
| WMR (any/none) | A15.145.229.637.555.Leukocytes..Mononuclear | Blood/Immune | -1.62E-08 | 3.94E-08 | 0.659702 |
| WMR (any/none) | A15.145.229.Blood.Cells | Blood/Immune | -2.08E-08 | 3.55E-08 | 0.720922 |
| WMR (any/none) | A15.145.300.Fetal.Blood | Blood/Immune | 6.27E-08 | 4.15E-08 | 0.065473 |
| WMR (any/none) | A15.145.846.Serum | Blood/Immune | 2.91E-08 | 3.43E-08 | 0.197623 |

| | | | | | |
|---|---|---|---|---|---|
| WMR (any/none) | A15.145.Blood | Blood/Immune | 4.27E-08 | 3.77E-08 | 0.128519 |
| WMR (any/none) | A15.378.316.580.Monocytes | Blood/Immune | 3.83E-08 | 4.03E-08 | 0.171099 |
| WMR (any/none) | A15.378.316.Bone.Marrow.Cells | Blood/Immune | 5.90E-08 | 3.96E-08 | 0.068046 |
| WMR (any/none) | A15.382.490.315.583.Neutrophils | Blood/Immune | 7.49E-08 | 3.56E-08 | 0.017594 |
| WMR (any/none) | A15.382.490.555.567.537.Killer.Cells..Natural | Blood/Immune | 5.66E-08 | 3.33E-08 | 0.044398 |
| WMR (any/none) | A15.382.490.555.567.622.Lymphocytes..Null | Blood/Immune | 7.42E-08 | 3.43E-08 | 0.015259 |
| WMR (any/none) | A15.382.490.555.567.Lymphocytes | Blood/Immune | -3.40E-09 | 4.07E-08 | 0.533271 |
| WMR (any/none) | A15.382.520.604.700.Spleen | Blood/Immune | -4.54E-08 | 3.74E-08 | 0.887638 |
| WMR (any/none) | A15.382.520.604.800.Palatine.Tonsil | Blood/Immune | -1.02E-08 | 3.24E-08 | 0.623363 |
| WMR (any/none) | A15.382.680.Phagocytes | Blood/Immune | 3.80E-08 | 3.94E-08 | 0.167461 |
| WMR (any/none) | A15.382.812.260.Dendritic.Cells | Blood/Immune | 2.22E-08 | 3.83E-08 | 0.280970 |
| WMR (any/none) | A15.382.812.522.Macrophages | Blood/Immune | 4.14E-08 | 3.72E-08 | 0.133151 |
| WMR (any/none) | A15.382.812.Mononuclear.Phagocyte.System | Blood/Immune | 5.74E-08 | 4.06E-08 | 0.078643 |
| WMR (any/none) | A15.382.Immune.System | Blood/Immune | -5.69E-08 | 3.50E-08 | 0.947958 |
| WMR (any/none) | Cells_EBV-transformed_lymphocytes | Blood/Immune | 3.46E-08 | 3.34E-08 | 0.150013 |
| WMR (any/none) | Spleen | Blood/Immune | -7.84E-08 | 3.43E-08 | 0.988857 |
| WMR (any/none) | Whole_Blood | Blood/Immune | 6.59E-08 | 4.04E-08 | 0.051588 |
| WMR (any/none) | A07.231.114.Arteries | Cardiovascular | -4.09E-08 | 3.15E-08 | 0.902717 |
| WMR (any/none) | A07.231.908.670.874.Umbilical.Veins | Cardiovascular | 2.53E-08 | 3.31E-08 | 0.222311 |
| WMR (any/none) | A07.231.908.Veins | Cardiovascular | 1.33E-08 | 3.40E-08 | 0.347909 |
| WMR (any/none) | A07.231.Blood.Vessels | Cardiovascular | 4.59E-08 | 3.64E-08 | 0.103554 |
| WMR (any/none) | A07.541.358.100.Atrial.Appendage | Cardiovascular | -9.43E-09 | 2.58E-08 | 0.642531 |
| WMR (any/none) | A07.541.358.Heart.Atria | Cardiovascular | -2.43E-08 | 2.53E-08 | 0.831247 |
| WMR (any/none) | A07.541.510.110.Aortic.Valve | Cardiovascular | 4.59E-09 | 2.91E-08 | 0.437233 |
| WMR (any/none) | A07.541.560.Heart.Ventricles | Cardiovascular | -4.74E-08 | 3.24E-08 | 0.928223 |
| WMR (any/none) | A07.541.Heart | Cardiovascular | -4.19E-08 | 3.02E-08 | 0.917736 |
| WMR (any/none) | Artery_Aorta | Cardiovascular | -1.65E-08 | 2.95E-08 | 0.712166 |
| WMR (any/none) | Artery_Coronary | Cardiovascular | -1.79E-08 | 2.86E-08 | 0.734781 |
| WMR (any/none) | Artery_Tibial | Cardiovascular | 5.80E-10 | 2.77E-08 | 0.491657 |
| WMR (any/none) | Heart_Atrial_Appendage | Cardiovascular | 3.88E-10 | 2.77E-08 | 0.494410 |
| WMR (any/none) | Heart_Left_Ventricle | Cardiovascular | -4.07E-09 | 2.68E-08 | 0.560191 |
| WMR (any/none) | A08.186.211.132.810.428.200.Cerebellum | CNS | 1.27E-08 | 2.74E-08 | 0.321382 |
| WMR (any/none) | A08.186.211.132.Brain.Stem | CNS | -1.53E-08 | 2.65E-08 | 0.717608 |
| WMR (any/none) | A08.186.211.464.405.Hippocampus | CNS | -1.13E-08 | 2.90E-08 | 0.651552 |
| WMR (any/none) | A08.186.211.464.710.225.Entorhinal.Cortex | CNS | -3.73E-08 | 2.92E-08 | 0.899671 |
| WMR (any/none) | A08.186.211.464.Limbic.System | CNS | -3.49E-08 | 3.04E-08 | 0.874332 |
| WMR (any/none) | A08.186.211.653.Mesencephalon | CNS | -6.81E-08 | 3.25E-08 | 0.982074 |
| WMR (any/none) | A08.186.211.730.317.357.352.435.Hypothalamo.Hypophyseal.System | CNS | 1.69E-08 | 4.18E-08 | 0.342929 |
| WMR (any/none) | A08.186.211.730.317.357.Hypothalamus | CNS | -6.13E-08 | 3.81E-08 | 0.946341 |
| WMR (any/none) | A08.186.211.730.317.Diencephalon | CNS | -7.26E-08 | 3.47E-08 | 0.981853 |
| WMR (any/none) | A08.186.211.730.885.287.249.487.Corpus.Striatum | CNS | -2.23E-08 | 2.87E-08 | 0.780821 |
| WMR (any/none) | A08.186.211.730.885.287.249.Basal.Ganglia | CNS | -2.40E-08 | 3.00E-08 | 0.788342 |
| WMR (any/none) | A08.186.211.730.885.287.500.270.Frontal.Lobe | CNS | -2.50E-08 | 3.15E-08 | 0.786467 |
| WMR (any/none) | A08.186.211.730.885.287.500.571.735.Visual.Cortex | CNS | 1.68E-09 | 3.01E-08 | 0.477688 |
| WMR (any/none) | A08.186.211.730.885.287.500.670.Parietal.Lobe | CNS | -4.46E-08 | 2.86E-08 | 0.940502 |
| WMR (any/none) | A08.186.211.730.885.287.500.Cerebral.Cortex | CNS | -2.37E-08 | 3.08E-08 | 0.779345 |

| | | | | | |
|---|---|---|---|---|---|
| WMR (any/none) | A08.186.211.865.428.Metencephalon | CNS | 1.37E-08 | 2.79E-08 | 0.311728 |
| WMR (any/none) | A08.186.211.Brain | CNS | -6.76E-08 | 2.88E-08 | 0.990636 |
| WMR (any/none) | A09.371.729.Retina | CNS | -1.52E-08 | 2.86E-08 | 0.702149 |
| WMR (any/none) | A11.872.653.Neural.Stem.Cells | CNS | -7.44E-09 | 3.56E-08 | 0.582884 |
| WMR (any/none) | Brain_Amygdala | CNS | 1.79E-08 | 2.58E-08 | 0.243772 |
| WMR (any/none) | Brain_Anterior_cingulate_cortex_(BA24) | CNS | -1.28E-08 | 2.50E-08 | 0.695991 |
| WMR (any/none) | Brain_Caudate_(basal_ganglia) | CNS | 1.99E-08 | 2.63E-08 | 0.224727 |
| WMR (any/none) | Brain_Cerebellar_Hemisphere | CNS | -1.00E-08 | 2.69E-08 | 0.644913 |
| WMR (any/none) | Brain_Cerebellum | CNS | -2.76E-08 | 2.75E-08 | 0.842557 |
| WMR (any/none) | Brain_Cortex | CNS | 2.33E-08 | 2.42E-08 | 0.166915 |
| WMR (any/none) | Brain_Frontal_Cortex_(BA9) | CNS | 2.35E-08 | 2.56E-08 | 0.179298 |
| WMR (any/none) | Brain_Hippocampus | CNS | 8.22E-09 | 2.59E-08 | 0.375464 |
| WMR (any/none) | Brain_Hypothalamus | CNS | 3.01E-08 | 2.53E-08 | 0.117096 |
| WMR (any/none) | Brain_Nucleus_accumbens_(basal_ganglia) | CNS | 1.81E-08 | 2.63E-08 | 0.245265 |
| WMR (any/none) | Brain_Putamen_(basal_ganglia) | CNS | 1.83E-08 | 2.56E-08 | 0.237693 |
| WMR (any/none) | Brain_Spinal_cord_(cervical_c-1) | CNS | 1.21E-08 | 2.69E-08 | 0.326298 |
| WMR (any/none) | Brain_Substantia_nigra | CNS | 6.53E-10 | 2.59E-08 | 0.489950 |
| WMR (any/none) | A03.556.124.369.Intestinal.Mucosa | Digestive | 3.74E-08 | 3.76E-08 | 0.160174 |
| WMR (any/none) | A03.556.124.526.767.Rectum | Digestive | 8.45E-09 | 3.41E-08 | 0.402115 |
| WMR (any/none) | A03.556.124.684.Intestine..Small | Digestive | -2.93E-08 | 3.50E-08 | 0.798162 |
| WMR (any/none) | A03.556.124.Intestines | Digestive | -5.34E-09 | 3.53E-08 | 0.560155 |
| WMR (any/none) | A03.556.249.124.Ileum | Digestive | 3.28E-10 | 3.93E-08 | 0.496675 |
| WMR (any/none) | A03.556.249.249.209.Cecum | Digestive | 7.01E-08 | 3.27E-08 | 0.015989 |
| WMR (any/none) | A03.556.249.249.356.668.Colon..Sigmoid | Digestive | 2.21E-08 | 3.55E-08 | 0.267398 |
| WMR (any/none) | A03.556.249.249.356.Colon | Digestive | 8.25E-09 | 3.34E-08 | 0.402343 |
| WMR (any/none) | A03.556.500.760.464.Parotid.Gland | Digestive | -3.12E-08 | 3.73E-08 | 0.798137 |
| WMR (any/none) | A03.556.500.760.Salivary.Glands | Digestive | -5.42E-08 | 3.78E-08 | 0.923983 |
| WMR (any/none) | A03.556.875.500.Esophagus | Digestive | 4.47E-08 | 3.20E-08 | 0.081361 |
| WMR (any/none) | A03.556.875.875.Stomach | Digestive | -3.80E-08 | 3.84E-08 | 0.839149 |
| WMR (any/none) | A03.556.875.Upper.Gastrointestinal.Tract | Digestive | 2.27E-08 | 3.72E-08 | 0.270512 |
| WMR (any/none) | A03.556.Gastrointestinal.Tract | Digestive | -1.54E-08 | 3.33E-08 | 0.678094 |
| WMR (any/none) | Colon_Sigmoid | Digestive | 3.15E-08 | 3.03E-08 | 0.148848 |
| WMR (any/none) | Colon_Transverse | Digestive | 5.73E-08 | 3.43E-08 | 0.047561 |
| WMR (any/none) | Esophagus_Gastroesophageal_Junction | Digestive | 7.70E-09 | 2.83E-08 | 0.392654 |
| WMR (any/none) | Esophagus_Mucosa | Digestive | 8.59E-09 | 3.35E-08 | 0.398924 |
| WMR (any/none) | Esophagus_Muscularis | Digestive | 3.52E-08 | 2.89E-08 | 0.112046 |
| WMR (any/none) | Small_Intestine_Terminal_Ileum | Digestive | -1.27E-08 | 3.66E-08 | 0.635855 |
| WMR (any/none) | Stomach | Digestive | -5.46E-08 | 3.26E-08 | 0.952818 |
| WMR (any/none) | A03.620.Liver | Liver | 2.07E-08 | 3.62E-08 | 0.283987 |
| WMR (any/none) | A11.436.348.Hepatocytes | Liver | -3.99E-08 | 3.82E-08 | 0.851379 |
| WMR (any/none) | Liver | Liver | -3.50E-09 | 3.60E-08 | 0.538672 |
| WMR (any/none) | A02.165.Cartilage | Musculoskeletal/Connective | 6.15E-08 | 3.43E-08 | 0.036443 |
| WMR (any/none) | A02.633.567.850.Quadriceps.Muscle | Musculoskeletal/Connective | -2.29E-08 | 3.29E-08 | 0.756349 |
| WMR (any/none) | A02.835.232.834.151.Cervical.Vertebrae | Musculoskeletal/Connective | 5.41E-09 | 3.08E-08 | 0.430228 |
| WMR (any/none) | A02.835.583.443.800.Synovial.Membrane | Musculoskeletal/Connective | -3.01E-08 | 3.21E-08 | 0.825701 |
| WMR (any/none) | A05.360.319.679.690.Myometrium | Musculoskeletal/Connective | -1.72E-08 | 3.06E-08 | 0.712341 |

| | | | | | |
|---|---|---|---|---|---|
| WMR (any/none) | A10.165.450.300.425.Keloid | Musculoskeletal/Connective | -3.97E-08 | 2.87E-08 | 0.916747 |
| WMR (any/none) | A10.165.450.300.Cicatrix | Musculoskeletal/Connective | -5.09E-08 | 2.84E-08 | 0.963163 |
| WMR (any/none) | A10.690.467.Muscle..Smooth | Musculoskeletal/Connective | -1.88E-09 | 3.28E-08 | 0.522831 |
| WMR (any/none) | A10.690.Muscles | Musculoskeletal/Connective | -4.40E-09 | 3.19E-08 | 0.554908 |
| WMR (any/none) | A11.329.171.Chondrocytes | Musculoskeletal/Connective | 1.14E-08 | 3.61E-08 | 0.375978 |
| WMR (any/none) | A11.329.228.Fibroblasts | Musculoskeletal/Connective | -5.29E-08 | 2.94E-08 | 0.963898 |
| WMR (any/none) | A11.329.629.Osteoblasts | Musculoskeletal/Connective | -1.27E-08 | 3.12E-08 | 0.657896 |
| WMR (any/none) | A11.329.830.Stromal.Cells | Musculoskeletal/Connective | -4.86E-08 | 3.10E-08 | 0.941298 |
| WMR (any/none) | A11.329.Connective.Tissue.Cells | Musculoskeletal/Connective | -2.44E-08 | 3.65E-08 | 0.747829 |
| WMR (any/none) | A11.620.520.Myocytes..Smooth.Muscle | Musculoskeletal/Connective | 1.12E-09 | 3.39E-08 | 0.486874 |
| WMR (any/none) | Cells_Transformed_fibroblasts | Musculoskeletal/Connective | -7.24E-09 | 2.93E-08 | 0.597767 |
| WMR (any/none) | Muscle_Skeletal | Musculoskeletal/Connective | 1.32E-08 | 3.12E-08 | 0.335595 |
| WMR (any/none) | A04.411.Lung | Other | 1.77E-08 | 3.40E-08 | 0.301523 |
| WMR (any/none) | A04.531.520.Nasal.Mucosa | Other | 2.02E-08 | 3.25E-08 | 0.266673 |
| WMR (any/none) | A05.360.319.114.373.Fallopian.Tubes | Other | -4.89E-09 | 2.94E-08 | 0.566123 |
| WMR (any/none) | A05.360.319.114.630.Ovary | Other | -1.20E-08 | 3.00E-08 | 0.655525 |
| WMR (any/none) | A05.360.319.679.256.Cervix.Uteri | Other | 1.96E-08 | 3.37E-08 | 0.280562 |
| WMR (any/none) | A05.360.319.679.490.Endometrium | Other | -5.63E-08 | 3.05E-08 | 0.967523 |
| WMR (any/none) | A05.360.319.679.Uterus | Other | -3.59E-08 | 2.99E-08 | 0.884752 |
| WMR (any/none) | A05.360.319.887.Vulva | Other | -2.06E-08 | 3.36E-08 | 0.730514 |
| WMR (any/none) | A05.360.319.Genitalia..Female | Other | -4.80E-08 | 3.06E-08 | 0.941676 |
| WMR (any/none) | A05.360.444.492.362.Foreskin | Other | -5.70E-09 | 3.02E-08 | 0.574773 |
| WMR (any/none) | A05.360.444.Genitalia..Male | Other | -5.16E-08 | 2.90E-08 | 0.962576 |
| WMR (any/none) | A05.360.490.Germ.Cells | Other | 6.70E-09 | 2.87E-08 | 0.407719 |
| WMR (any/none) | A05.360.Genitalia | Other | -3.89E-08 | 2.94E-08 | 0.907204 |
| WMR (any/none) | A05.810.453.324.Kidney.Cortex | Other | 2.02E-08 | 2.98E-08 | 0.248454 |
| WMR (any/none) | A05.810.453.Kidney | Other | -2.50E-08 | 3.06E-08 | 0.793195 |
| WMR (any/none) | A05.810.890.Urinary.Bladder | Other | -1.74E-08 | 3.56E-08 | 0.687482 |
| WMR (any/none) | A06.407.071.140.Adrenal.Cortex | Other | -6.80E-08 | 2.97E-08 | 0.988978 |
| WMR (any/none) | A06.407.071.Adrenal.Glands | Other | -7.73E-08 | 2.92E-08 | 0.995970 |
| WMR (any/none) | A06.407.312.782.Testis | Other | 2.04E-08 | 2.81E-08 | 0.234239 |
| WMR (any/none) | A06.407.312.Gonads | Other | 2.65E-09 | 3.02E-08 | 0.465143 |
| WMR (any/none) | A06.407.900.Thyroid.Gland | Other | -2.99E-08 | 3.07E-08 | 0.835045 |
| WMR (any/none) | A06.407.Endocrine.Glands | Other | -2.11E-08 | 2.91E-08 | 0.765356 |
| WMR (any/none) | A09.371.Eye | Other | -1.10E-08 | 3.68E-08 | 0.617797 |
| WMR (any/none) | A10.272.497.Epidermis | Other | 1.44E-08 | 3.44E-08 | 0.338020 |
| WMR (any/none) | A10.272.Epithelium | Other | 1.63E-08 | 3.31E-08 | 0.311274 |
| WMR (any/none) | A10.336.707.Prostate | Other | -6.29E-08 | 3.17E-08 | 0.976501 |
| WMR (any/none) | A10.615.284.473.Chorion | Other | -3.74E-08 | 3.03E-08 | 0.891698 |
| WMR (any/none) | A10.615.550.599.Mouth.Mucosa | Other | 3.75E-08 | 3.27E-08 | 0.125834 |
| WMR (any/none) | A10.615.550.Mucous.Membrane | Other | 3.43E-08 | 3.71E-08 | 0.177820 |
| WMR (any/none) | A10.615.789.Serous.Membrane | Other | -1.97E-08 | 3.20E-08 | 0.731080 |
| WMR (any/none) | A10.615.Membranes | Other | 2.73E-08 | 3.28E-08 | 0.203136 |
| WMR (any/none) | A11.382.Endocrine.Cells | Other | 3.75E-08 | 3.31E-08 | 0.128434 |
| WMR (any/none) | A11.436.275.Endothelial.Cells | Other | 3.32E-08 | 3.40E-08 | 0.164874 |
| WMR (any/none) | A11.436.294.064.Glucagon.Secreting.Cells | Other | 4.42E-08 | 3.50E-08 | 0.103146 |

| | | | | | |
|---|---|---|---|---|---|
| WMR (any/none) | A11.436.329.Granulosa.Cells | Other | 2.25E-08 | 3.02E-08 | 0.228039 |
| WMR (any/none) | A11.436.397.Keratinocytes | Other | 8.92E-09 | 3.59E-08 | 0.401721 |
| WMR (any/none) | A11.436.Epithelial.Cells | Other | 1.71E-08 | 3.33E-08 | 0.303774 |
| WMR (any/none) | A11.497.497.600.Oocytes | Other | 2.53E-09 | 2.81E-08 | 0.464149 |
| WMR (any/none) | A11.872.040.Adult.Stem.Cells | Other | 4.48E-08 | 3.21E-08 | 0.081300 |
| WMR (any/none) | A11.872.190.260.Embryoid.Bodies | Other | 1.09E-08 | 3.39E-08 | 0.373628 |
| WMR (any/none) | A11.872.190.Embryonic.Stem.Cells | Other | -2.08E-08 | 3.25E-08 | 0.739146 |
| WMR (any/none) | A11.872.580.Mesenchymal.Stem.Cells | Other | -1.19E-08 | 2.98E-08 | 0.655199 |
| WMR (any/none) | A11.872.700.500.Induced.Pluripotent.Stem.Cells | Other | 1.81E-08 | 3.36E-08 | 0.295493 |
| WMR (any/none) | A11.872.Stem.Cells | Other | 6.56E-08 | 3.58E-08 | 0.033239 |
| WMR (any/none) | A14.549.167.646.Periodontium | Other | -1.10E-08 | 3.81E-08 | 0.613407 |
| WMR (any/none) | A14.549.167.Dentition | Other | -4.37E-08 | 3.29E-08 | 0.908003 |
| WMR (any/none) | A14.549.885.Tongue | Other | 4.97E-09 | 3.55E-08 | 0.444309 |
| WMR (any/none) | A14.549.Mouth | Other | -3.76E-09 | 3.09E-08 | 0.548510 |
| WMR (any/none) | A14.724.557.Nasopharynx | Other | -6.12E-08 | 3.48E-08 | 0.960731 |
| WMR (any/none) | A14.724.Pharynx | Other | -8.95E-08 | 3.34E-08 | 0.996344 |
| WMR (any/none) | A17.815.Skin | Other | -2.58E-09 | 3.30E-08 | 0.531145 |
| WMR (any/none) | Adrenal_Gland | Other | -1.69E-08 | 3.28E-08 | 0.697044 |
| WMR (any/none) | Bladder | Other | -2.32E-09 | 3.11E-08 | 0.529656 |
| WMR (any/none) | Breast_Mammary_Tissue | Other | -3.58E-08 | 2.86E-08 | 0.894834 |
| WMR (any/none) | Cervix_Ectocervix | Other | -3.01E-08 | 2.78E-08 | 0.860219 |
| WMR (any/none) | Cervix_Endocervix | Other | -2.11E-08 | 2.96E-08 | 0.761735 |
| WMR (any/none) | Fallopian_Tube | Other | -2.58E-08 | 2.98E-08 | 0.806587 |
| WMR (any/none) | Kidney_Cortex | Other | -5.25E-08 | 3.58E-08 | 0.928777 |
| WMR (any/none) | Lung | Other | 3.40E-08 | 3.42E-08 | 0.159733 |
| WMR (any/none) | Minor_Salivary_Gland | Other | -1.37E-08 | 2.88E-08 | 0.682567 |
| WMR (any/none) | Nerve_Tibial | Other | -1.55E-08 | 2.65E-08 | 0.720649 |
| WMR (any/none) | Ovary | Other | -2.39E-09 | 3.17E-08 | 0.530058 |
| WMR (any/none) | Pituitary | Other | -3.71E-08 | 2.76E-08 | 0.910928 |
| WMR (any/none) | Prostate | Other | -4.51E-08 | 3.38E-08 | 0.909003 |
| WMR (any/none) | Skin_Not_Sun_Exposed_(Suprapubic) | Other | -6.35E-09 | 3.21E-08 | 0.578516 |
| WMR (any/none) | Skin_Sun_Exposed_(Lower_leg) | Other | -2.45E-08 | 2.97E-08 | 0.795185 |
| WMR (any/none) | Testis | Other | -4.05E-08 | 2.75E-08 | 0.930055 |
| WMR (any/none) | Thyroid | Other | 5.75E-09 | 3.26E-08 | 0.430047 |
| WMR (any/none) | Uterus | Other | 1.18E-08 | 3.17E-08 | 0.354084 |
| WMR (any/none) | Vagina | Other | -3.71E-08 | 2.97E-08 | 0.894257 |
| WMR (any/none) | A03.734.414.Islets.of.Langerhans | Pancreas | 2.62E-08 | 3.53E-08 | 0.229463 |
| WMR (any/none) | A03.734.Pancreas | Pancreas | -2.05E-08 | 3.29E-08 | 0.733331 |
| WMR (any/none) | Pancreas | Pancreas | 1.04E-08 | 3.09E-08 | 0.368733 |

**Table S13: Cell/Tissue-specific heritability enrichment p-values (ImmGen)**

| Tissue/Cell Type | Tissue category | ADNC Score | AP (any/none) | CERAD (C Score) | Thal Phase | B Score | AD Braak | Arteriolo-sclerosis (any/none) | Arteriolo-sclerosis | Athero-sclerosis (any/none) | Athero-sclerosis | CAA (any/none) | CAA | Infarcts/ lacunes | PD Braak (any/none) | PD Braak | TDP-43 (any/none) | WMR (any/none) |
|---|---|---|---|---|---|---|---|---|---|---|---|---|---|---|---|---|---|---|
| B.Fo.LN | B | 0.8166 | 0.6011 | 0.0249 | 0.7811 | 0.0939 | 0.1427 | 0.1569 | 0.3389 | 0.4909 | 0.0394 | 0.7582 | 0.7230 | 0.5818 | 0.3981 | 0.1753 | 0.5249 | 0.5841 |
| B.Fo.MLN | B | 0.5072 | 0.4468 | 0.0895 | 0.4110 | 0.0145 | 0.0041 | 0.1471 | 0.5039 | 0.3088 | 0.0241 | 0.8734 | 0.8922 | 0.8738 | 0.3955 | 0.2300 | 0.6400 | 0.4123 |
| B.Fo.PC | B | 0.6562 | 0.2296 | 0.0785 | 0.4845 | 0.0097 | 0.0096 | 0.1551 | 0.1666 | 0.2537 | 0.0550 | 0.5524 | 0.7002 | 0.9151 | 0.4736 | 0.4102 | 0.6479 | 0.7073 |
| B.Fo.Sp | B | 0.5048 | 0.4557 | 0.3482 | 0.2912 | 0.1686 | 0.1056 | 0.2019 | 0.4454 | 0.4234 | 0.0702 | 0.9444 | 0.9480 | 0.8128 | 0.7751 | 0.6676 | 0.7066 | 0.6417 |
| B.FrE.BM | B | 0.8907 | 0.4453 | 0.0512 | 0.8584 | 0.0054 | 0.0112 | 0.2911 | 0.8258 | 0.3543 | 0.0468 | 0.9777 | 0.9902 | 0.8526 | 0.6940 | 0.5114 | 0.8951 | 0.7823 |
| B.FrE.FL | B | 0.9426 | 0.4583 | 0.3791 | 0.6879 | 0.1195 | 0.2845 | 0.1385 | 0.7731 | 0.6458 | 0.2632 | 0.3707 | 0.4013 | 0.9917 | 0.8787 | 0.7753 | 0.9902 | 0.5452 |
| B.FrF.BM | B | 0.6002 | 0.4500 | 0.2872 | 0.4611 | 0.0120 | 0.0087 | 0.3551 | 0.2736 | 0.2801 | 0.1522 | 0.9934 | 0.9954 | 0.7156 | 0.5540 | 0.4159 | 0.8480 | 0.6254 |
| B.GC.Sp | B | 0.9241 | 0.4475 | 0.5523 | 0.6748 | 0.1696 | 0.1933 | 0.0402 | 0.7871 | 0.1623 | 0.4214 | 0.4040 | 0.9241 | 0.3295 | 0.7884 | 0.6970 | 0.9782 | 0.4316 |
| B.Mem.Sp.v2 | B | 0.8604 | 0.6436 | 0.1373 | 0.8780 | 0.1016 | 0.0195 | 0.8761 | 0.4365 | 0.7849 | 0.6575 | 0.2909 | 0.7041 | 0.5920 | 0.2552 | 0.1775 | 0.8062 | 0.7326 |
| B.MZ.Sp | B | 0.8331 | 0.6716 | 0.1450 | 0.3504 | 0.0186 | 0.0059 | 0.0970 | 0.5007 | 0.4443 | 0.1717 | 0.8820 | 0.8798 | 0.8290 | 0.8030 | 0.5904 | 0.6474 | 0.9338 |
| B.T1.Sp | B | 0.4184 | 0.5760 | 0.0750 | 0.2729 | 0.0064 | 0.0039 | 0.0649 | 0.6211 | 0.0196 | 0.0060 | 0.8960 | 0.9415 | 0.9103 | 0.5461 | 0.5300 | 0.9260 | 0.2273 |
| B.T2.Sp | B | 0.6630 | 0.5383 | 0.2731 | 0.3147 | 0.0562 | 0.0244 | 0.0478 | 0.3913 | 0.1166 | 0.0465 | 0.8788 | 0.8367 | 0.8480 | 0.8746 | 0.7658 | 0.6764 | 0.5562 |
| B.T3.Sp | B | 0.8750 | 0.6998 | 0.5923 | 0.6147 | 0.0917 | 0.0345 | 0.0735 | 0.4784 | 0.1191 | 0.0141 | 0.9467 | 0.9099 | 0.9260 | 0.7624 | 0.6359 | 0.6523 | 0.7808 |
| B1a.PC | B | 0.4308 | 0.5391 | 0.1290 | 0.4007 | 0.0093 | 0.0118 | 0.4631 | 0.5774 | 0.2603 | 0.1041 | 0.6970 | 0.8409 | 0.7200 | 0.9056 | 0.7731 | 0.9738 | 0.9594 |
| B1a.Sp | B | 0.6921 | 0.9046 | 0.5238 | 0.3655 | 0.1859 | 0.1145 | 0.2342 | 0.5084 | 0.3425 | 0.2050 | 0.7717 | 0.7522 | 0.7207 | 0.9131 | 0.6845 | 0.7862 | 0.9142 |
| B1a.Sp.v2 | B | 0.8338 | 0.9358 | 0.5065 | 0.7725 | 0.8659 | 0.9581 | 0.4799 | 0.1506 | 0.8994 | 0.9733 | 0.8474 | 0.9878 | 0.7490 | 0.7334 | 0.7651 | 0.4646 | 0.1559 |
| B1b.PC | B | 0.3425 | 0.9063 | 0.3848 | 0.2962 | 0.0601 | 0.0410 | 0.2831 | 0.5681 | 0.3414 | 0.0784 | 0.9049 | 0.9255 | 0.4635 | 0.6742 | 0.4722 | 0.9792 | 0.9426 |
| CD19Control | B | 0.6508 | 0.7605 | 0.3635 | 0.3336 | 0.0716 | 0.0249 | 0.0716 | 0.2022 | 0.2382 | 0.0382 | 0.9106 | 0.9740 | 0.8304 | 0.7963 | 0.7614 | 0.7518 | 0.7581 |
| preB.FrC.BM | B | 0.7320 | 0.5309 | 0.4101 | 0.3441 | 0.2113 | 0.1033 | 0.1379 | 0.4324 | 0.2157 | 0.3047 | 0.0323 | 0.6114 | 0.7502 | 0.8877 | 0.9526 | 0.9924 | 0.1255 |
| preB.FrD.BM | B | 0.8583 | 0.7554 | 0.5768 | 0.5638 | 0.3117 | 0.3120 | 0.3728 | 0.6796 | 0.5262 | 0.0620 | 0.3610 | 0.7559 | 0.4014 | 0.7192 | 0.7135 | 0.6365 | 0.3005 |
| preB.FrD.FL | B | 0.4612 | 0.4323 | 0.2751 | 0.2937 | 0.3212 | 0.0986 | 0.0530 | 0.4208 | 0.1898 | 0.1747 | 0.0699 | 0.2881 | 0.7331 | 0.6986 | 0.6631 | 0.4269 | 0.0566 |
| proB.CLP.BM | B | 0.6799 | 0.3549 | 0.2508 | 0.3048 | 0.1723 | 0.1105 | 0.0325 | 0.0103 | 0.1493 | 0.4754 | 0.2009 | 0.4504 | 0.5841 | 0.6733 | 0.6775 | 0.8367 | 0.1698 |
| proB.CLP.FL | B | 0.3316 | 0.4315 | 0.5153 | 0.3784 | 0.8664 | 0.8525 | 0.0011 | 0.0190 | 0.0096 | 0.2827 | 0.2645 | 0.2326 | 0.9838 | 0.3619 | 0.3959 | 0.5371 | 0.0420 |
| proB.FrA.BM | B | 0.7078 | 0.0445 | 0.0037 | 0.5147 | 0.0367 | 0.0129 | 0.1765 | 0.0204 | 0.2426 | 0.1967 | 0.2207 | 0.7036 | 0.6066 | 0.7912 | 0.7681 | 0.8012 | 0.1666 |
| proB.FrA.FL | B | 0.6155 | 0.6889 | 0.3998 | 0.4153 | 0.6412 | 0.6813 | 0.0012 | 0.0147 | 0.0891 | 0.5423 | 0.3577 | 0.4547 | 0.8848 | 0.4397 | 0.4477 | 0.3363 | 0.0541 |
| proB.FrBC.BM | B | 0.5088 | 0.3772 | 0.1412 | 0.0565 | 0.2040 | 0.0758 | 0.0400 | 0.3218 | 0.2970 | 0.1116 | 0.0217 | 0.5885 | 0.7383 | 0.9751 | 0.9610 | 0.5863 | 0.5215 |
| proB.FrBC.FL | B | 0.3064 | 0.2601 | 0.3789 | 0.2696 | 0.6848 | 0.2950 | 0.0391 | 0.1231 | 0.1339 | 0.4436 | 0.2059 | 0.5480 | 0.5978 | 0.5928 | 0.5361 | 0.9302 | 0.0157 |
| BA.Bl | Myeloid | 0.5594 | 0.2197 | 0.5372 | 0.9847 | 0.5316 | 0.6913 | 0.3609 | 0.2197 | 0.6898 | 0.2247 | 0.8580 | 0.8450 | 0.8131 | 0.5133 | 0.6363 | 0.5984 | 0.1967 |
| BA.Sp | Myeloid | 0.6207 | 0.6428 | 0.7555 | 0.9750 | 0.6164 | 0.7883 | 0.4208 | 0.5475 | 0.3919 | 0.3912 | 0.4843 | 0.7814 | 0.3585 | 0.2615 | 0.2975 | 0.7303 | 0.2487 |
| DC.103+11b-.Lu | Myeloid | 0.7107 | 0.4123 | 0.1299 | 0.7658 | 0.1826 | 0.2225 | 0.1182 | 0.5183 | 0.5682 | 0.7443 | 0.8117 | 0.7785 | 0.3528 | 0.2511 | 0.4079 | 0.6581 | 0.5735 |
| DC.103+11b-.Lv | Myeloid | 0.2176 | 0.2236 | 0.2127 | 0.7837 | 0.3014 | 0.3742 | 0.2291 | 0.5932 | 0.6341 | 0.6908 | 0.8138 | 0.7673 | 0.7094 | 0.8149 | 0.8296 | 0.8052 | 0.6018 |
| DC.103-11b+24+.Lu | Myeloid | 0.3098 | 0.2077 | 0.1728 | 0.4318 | 0.2893 | 0.1371 | 0.3093 | 0.2239 | 0.5213 | 0.6755 | 0.7269 | 0.4989 | 0.1431 | 0.0825 | 0.1497 | 0.8820 | 0.2121 |
| DC.103-11b+F4_80lo.Kd | Myeloid | 0.0446 | 0.1326 | 0.0344 | 0.1112 | 0.1292 | 0.1105 | 0.1585 | 0.4380 | 0.8346 | 0.9596 | 0.5893 | 0.7201 | 0.2930 | 0.4843 | 0.3818 | 0.6390 | 0.4597 |
| DC.11b-.AT.v2 | Myeloid | 0.7565 | 0.1555 | 0.3871 | 0.5703 | 0.0619 | 0.0768 | 0.7985 | 0.7674 | 0.5891 | 0.6741 | 0.3477 | 0.3384 | 0.6577 | 0.2812 | 0.2212 | 0.6279 | 0.5094 |
| DC.11b+.AT.v2 | Myeloid | 0.7378 | 0.7116 | 0.1588 | 0.9927 | 0.1854 | 0.3379 | 0.2549 | 0.3689 | 0.7013 | 0.3484 | 0.8808 | 0.9124 | 0.0538 | 0.0073 | 0.0037 | 0.4580 | 0.4622 |
| DC.4+.MLN | Myeloid | 0.2806 | 0.0566 | 0.0389 | 0.5141 | 0.2600 | 0.4268 | 0.1098 | 0.2391 | 0.5728 | 0.7006 | 0.8376 | 0.8231 | 0.5629 | 0.1210 | 0.1954 | 0.9767 | 0.5212 |
| DC.4+.SLN | Myeloid | 0.0887 | 0.4024 | 0.6644 | 0.3673 | 0.4781 | 0.3770 | 0.1485 | 0.1404 | 0.6268 | 0.8541 | 0.5582 | 0.0409 | 0.1269 | 0.2105 | 0.1254 | 0.5670 | 0.2591 |
| DC.4+.Sp.ST | Myeloid | 0.2109 | 0.0787 | 0.0861 | 0.3232 | 0.1231 | 0.2252 | 0.2689 | 0.4206 | 0.6761 | 0.6692 | 0.8269 | 0.8339 | 0.6281 | 0.4144 | 0.3512 | 0.9470 | 0.5518 |
| DC.8-.Th | Myeloid | 0.3852 | 0.5397 | 0.7227 | 0.2391 | 0.6028 | 0.6831 | 0.5842 | 0.9563 | 0.6011 | 0.8189 | 0.7919 | 0.7575 | 0.3040 | 0.1190 | 0.2609 | 0.4117 | 0.5451 |
| DC.8+.MLN | Myeloid | 0.7114 | 0.4575 | 0.6088 | 0.7731 | 0.7566 | 0.6617 | 0.2155 | 0.8029 | 0.4575 | 0.4745 | 0.7694 | 0.9068 | 0.6032 | 0.3582 | 0.3284 | 0.2452 | 0.7835 |
| DC.8+.SLN | Myeloid | 0.4917 | 0.2036 | 0.3778 | 0.7759 | 0.4610 | 0.6409 | 0.1301 | 0.4697 | 0.2587 | 0.3916 | 0.5554 | 0.7927 | 0.7486 | 0.0580 | 0.0533 | 0.6130 | 0.6322 |
| DC.8+.Sp.ST | Myeloid | 0.8927 | 0.5496 | 0.5669 | 0.8900 | 0.7918 | 0.9134 | 0.5086 | 0.9446 | 0.4967 | 0.6527 | 0.8789 | 0.7975 | 0.3464 | 0.2634 | 0.2002 | 0.2217 | 0.9311 |
| DC.8+.Th | Myeloid | 0.4171 | 0.7216 | 0.8289 | 0.4735 | 0.6456 | 0.7162 | 0.2417 | 0.9493 | 0.4523 | 0.7975 | 0.7132 | 0.7306 | 0.2256 | 0.1198 | 0.2308 | 0.3790 | 0.5722 |
| DC.8-4-11b-.MLN | Myeloid | 0.8068 | 0.3694 | 0.3940 | 0.4107 | 0.7223 | 0.5712 | 0.5885 | 0.8980 | 0.2095 | 0.4501 | 0.8579 | 0.8936 | 0.6560 | 0.0358 | 0.0114 | 0.9703 | 0.1259 |
| DC.8-4-11b-.SLN | Myeloid | 0.6653 | 0.2304 | 0.4809 | 0.4247 | 0.7455 | 0.7237 | 0.4305 | 0.6161 | 0.6758 | 0.7149 | 0.4945 | 0.6418 | 0.3176 | 0.0237 | 0.0435 | 0.8600 | 0.7716 |
| DC.8-4-11b-.Sp | Myeloid | 0.5952 | 0.4940 | 0.2585 | 0.8928 | 0.3199 | 0.5391 | 0.3467 | 0.5134 | 0.7125 | 0.7196 | 0.7716 | 0.7307 | 0.4242 | 0.4093 | 0.3308 | 0.9117 | 0.5290 |
| DC.8-4-11b+.MLN | Myeloid | 0.8653 | 0.3294 | 0.4354 | 0.4512 | 0.5282 | 0.7611 | 0.7175 | 0.4916 | 0.8167 | 0.5664 | 0.7777 | 0.8543 | 0.5077 | 0.1162 | 0.0344 | 0.9653 | 0.5085 |
| DC.8-4-11b+.SLN | Myeloid | 0.5432 | 0.3806 | 0.5191 | 0.1847 | 0.8116 | 0.9011 | 0.3885 | 0.6950 | 0.7465 | 0.9009 | 0.7978 | 0.7027 | 0.5801 | 0.4128 | 0.2724 | 0.7811 | 0.0752 |
| DC.8-4-11b+.Sp | Myeloid | 0.1035 | 0.0860 | 0.0282 | 0.1941 | 0.2396 | 0.1808 | 0.5713 | 0.3035 | 0.4589 | 0.5150 | 0.8625 | 0.4300 | 0.2366 | 0.0391 | 0.0165 | 0.5929 | 0.4351 |
| DC.IIhilang+103+11blo.SLN | Myeloid | 0.5334 | 0.0598 | 0.6634 | 0.2264 | 0.6175 | 0.6957 | 0.2084 | 0.7310 | 0.7106 | 0.9386 | 0.7895 | 0.8047 | 0.9056 | 0.0948 | 0.1103 | 0.9673 | 0.1770 |
| DC.IIhilang+103-11b+.SLN | Myeloid | 0.2463 | 0.0270 | 0.2633 | 0.0916 | 0.4995 | 0.8526 | 0.2740 | 0.5791 | 0.5272 | 0.3798 | 0.6532 | 0.8291 | 0.5218 | 0.0738 | 0.0781 | 0.5722 | 0.4839 |
| DC.IIhilang-103-11b+.SLN | Myeloid | 0.5287 | 0.6026 | 0.5622 | 0.0604 | 0.5966 | 0.7319 | 0.0525 | 0.3634 | 0.6323 | 0.8519 | 0.7060 | 0.8952 | 0.7011 | 0.7543 | 0.6166 | 0.7357 | 0.0494 |
| DC.IIhilang-103-11blo.SLN | Myeloid | 0.6305 | 0.3502 | 0.6154 | 0.1313 | 0.7727 | 0.8275 | 0.2117 | 0.6651 | 0.4389 | 0.5380 | 0.8803 | 0.7659 | 0.7063 | 0.3077 | 0.1929 | 0.9972 | 0.2015 |
| DC.LC.Sk | Myeloid | 0.2650 | 0.4281 | 0.2579 | 0.1374 | 0.5943 | 0.5169 | 0.0832 | 0.0723 | 0.6204 | 0.4646 | 0.7511 | 0.5604 | 0.0600 | 0.0618 | 0.1126 | 0.8353 | 0.8368 |
| DC.pDC.8-.Sp | Myeloid | 0.9980 | 0.5675 | 0.5479 | 0.7267 | 0.9077 | 0.8267 | 0.3499 | 0.3140 | 0.8866 | 0.3865 | 0.4711 | 0.4256 | 0.2112 | 0.0044 | 0.0021 | 0.1151 | 0.3085 |
| DC.pDC.8+.MLN | Myeloid | 0.9963 | 0.8243 | 0.1763 | 0.7166 | 0.8219 | 0.7334 | 0.4307 | 0.5615 | 0.9068 | 0.1928 | 0.3846 | 0.3104 | 0.0797 | 0.0313 | 0.0368 | 0.0616 | 0.2567 |
| DC.pDC.8+.SLN | Myeloid | 0.9945 | 0.7839 | 0.2210 | 0.5864 | 0.8644 | 0.8945 | 0.2529 | 0.3972 | 0.7911 | 0.2634 | 0.4309 | 0.6168 | 0.1587 | 0.0098 | 0.0111 | 0.0567 | 0.1070 |
| DC.pDC.8+.Sp | Myeloid | 0.9880 | 0.7383 | 0.3986 | 0.4766 | 0.6909 | 0.6492 | 0.3787 | 0.3475 | 0.7985 | 0.3484 | 0.5228 | 0.6229 | 0.0800 | 0.0020 | 0.0011 | 0.1372 | 0.2251 |
| EO.AT.v2 | Myeloid | 0.4830 | 0.1921 | 0.5524 | 0.7994 | 0.3652 | 0.4453 | 0.7558 | 0.4282 | 0.6613 | 0.9546 | 0.7538 | 0.6829 | 0.7274 | 0.0103 | 0.0186 | 0.9891 | 0.7839 |

| | | | | | | | | | | | | | | | | | | |
|---|---|---|---|---|---|---|---|---|---|---|---|---|---|---|---|---|---|---|
| Eo.BL.v2 | Myeloid | 0.5086 | 0.5393 | 0.9509 | 0.9458 | 0.8820 | 0.9365 | 0.1385 | 0.0823 | 0.6327 | 0.4913 | 0.9295 | 0.8753 | 0.0892 | 0.1590 | 0.3893 | 0.8035 | 0.7438 |
| GN.Arth.BM | Myeloid | 0.6675 | 0.5383 | 0.0917 | 0.8475 | 0.6974 | 0.7321 | 0.5965 | 0.0929 | 0.5100 | 0.5050 | 0.9482 | 0.9599 | 0.3687 | 0.1241 | 0.2261 | 0.4860 | 0.0386 |
| GN.Arth.SynF | Myeloid | 0.1965 | 0.3567 | 0.0158 | 0.4378 | 0.5359 | 0.6020 | 0.3399 | 0.5108 | 0.7119 | 0.1269 | 0.6889 | 0.5775 | 0.5902 | 0.1314 | 0.1163 | 0.8424 | 0.2346 |
| GN.Bl | Myeloid | 0.2682 | 0.3248 | 0.0823 | 0.8927 | 0.8110 | 0.6804 | 0.6662 | 0.6352 | 0.6523 | 0.3222 | 0.8704 | 0.8484 | 0.4514 | 0.0523 | 0.0667 | 0.4533 | 0.3080 |
| GN.Bl.v2 | Myeloid | 0.5370 | 0.1405 | 0.0509 | 0.9875 | 0.2134 | 0.1425 | 0.1889 | 0.5900 | 0.8599 | 0.2211 | 0.9501 | 0.8403 | 0.4186 | 0.0058 | 0.0098 | 0.8497 | 0.2192 |
| GN.BM | Myeloid | 0.7661 | 0.2513 | 0.0314 | 0.9709 | 0.3618 | 0.6417 | 0.8428 | 0.5343 | 0.6419 | 0.7021 | 0.9320 | 0.9531 | 0.2172 | 0.2333 | 0.3682 | 0.4895 | 0.2131 |
| GN.Thio.PC | Myeloid | 0.0584 | 0.5365 | 0.1141 | 0.2921 | 0.5529 | 0.4420 | 0.6074 | 0.8375 | 0.1578 | 0.2574 | 0.7529 | 0.5428 | 0.5888 | 0.1830 | 0.1557 | 0.6456 | 0.3081 |
| GN.UrAc.PC | Myeloid | 0.2834 | 0.7648 | 0.1407 | 0.6704 | 0.6023 | 0.5407 | 0.2132 | 0.4461 | 0.3598 | 0.0868 | 0.7067 | 0.8239 | 0.3617 | 0.0006 | 0.0009 | 0.6768 | 0.6273 |
| MC.digest.PC | Myeloid | 0.4520 | 0.4514 | 0.6704 | 0.7120 | 0.7172 | 0.6671 | 0.2471 | 0.1636 | 0.5639 | 0.6479 | 0.5962 | 0.3082 | 0.1184 | 0.4615 | 0.5873 | 0.8755 | 0.0536 |
| MC.Es | Myeloid | 0.6689 | 0.1782 | 0.4385 | 0.6278 | 0.6607 | 0.7916 | 0.4915 | 0.0755 | 0.6764 | 0.3377 | 0.2245 | 0.2730 | 0.2271 | 0.1915 | 0.2758 | 0.5030 | 0.0705 |
| MC.PC | Myeloid | 0.5823 | 0.1924 | 0.6205 | 0.8006 | 0.9109 | 0.7727 | 0.1053 | 0.0969 | 0.4581 | 0.6539 | 0.4796 | 0.8159 | 0.1519 | 0.3287 | 0.3385 | 0.9054 | 0.3959 |
| MC.Sk | Myeloid | 0.5847 | 0.0320 | 0.2122 | 0.5757 | 0.9252 | 0.8383 | 0.7339 | 0.5541 | 0.6061 | 0.6875 | 0.4751 | 0.8197 | 0.4489 | 0.0894 | 0.1335 | 0.3102 | 0.1257 |
| MC.To | Myeloid | 0.6363 | 0.0603 | 0.3239 | 0.6766 | 0.2342 | 0.3559 | 0.4594 | 0.1661 | 0.6539 | 0.5307 | 0.4411 | 0.8325 | 0.1748 | 0.0660 | 0.1319 | 0.5638 | 0.0548 |
| MC.Tr | Myeloid | 0.9151 | 0.1025 | 0.5701 | 0.9617 | 0.9591 | 0.9493 | 0.6772 | 0.4404 | 0.2104 | 0.1423 | 0.7396 | 0.9356 | 0.1835 | 0.2488 | 0.4927 | 0.9862 | 0.0337 |
| MF.103-11b+.Lu | Myeloid | 0.4567 | 0.1520 | 0.1173 | 0.7018 | 0.7064 | 0.6920 | 0.0754 | 0.1649 | 0.4490 | 0.6863 | 0.7664 | 0.3736 | 0.5152 | 0.0630 | 0.0954 | 0.7362 | 0.3649 |
| MF.103-11b+.Salm3.SI | Myeloid | 0.2708 | 0.2697 | 0.0273 | 0.1267 | 0.5969 | 0.3220 | 0.0551 | 0.0788 | 0.5218 | 0.6829 | 0.3697 | 0.3821 | 0.0413 | 0.0575 | 0.0601 | 0.9832 | 0.8802 |
| MF.103-11b+.SI | Myeloid | 0.1160 | 0.1267 | 0.0047 | 0.0405 | 0.4305 | 0.4839 | 0.1815 | 0.2816 | 0.5532 | 0.7660 | 0.4517 | 0.4201 | 0.0118 | 0.0107 | 0.0059 | 0.9752 | 0.8103 |
| MF.11c-11b+.Lu | Myeloid | 0.1252 | 0.0485 | 0.0175 | 0.2641 | 0.1099 | 0.1767 | 0.3063 | 0.1578 | 0.8787 | 0.9908 | 0.4493 | 0.5808 | 0.0285 | 0.0565 | 0.0464 | 0.9634 | 0.3612 |
| MF.11cloSer.Salm3.SI | Myeloid | 0.4307 | 0.0832 | 0.0063 | 0.2192 | 0.3137 | 0.0504 | 0.0504 | 0.0829 | 0.7411 | 0.7966 | 0.1737 | 0.1761 | 0.0300 | 0.0403 | 0.0222 | 0.9822 | 0.7071 |
| MF.11cloSer.SI | Myeloid | 0.1967 | 0.1007 | 0.0308 | 0.1869 | 0.1477 | 0.0777 | 0.0601 | 0.0918 | 0.4621 | 0.7728 | 0.6116 | 0.7273 | 0.0545 | 0.0693 | 0.0395 | 0.9805 | 0.3433 |
| MF.169+11chi.SLN | Myeloid | 0.0992 | 0.0529 | 0.1050 | 0.4089 | 0.7203 | 0.4592 | 0.8029 | 0.6783 | 0.3475 | 0.7568 | 0.3029 | 0.0948 | 0.5341 | 0.0636 | 0.0742 | 0.9176 | 0.7995 |
| MF.480hi.LV.Naive | Myeloid | 0.4412 | 0.0439 | 0.1050 | 0.7560 | 0.0271 | 0.1128 | 0.1413 | 0.1706 | 0.6951 | 0.9224 | 0.5715 | 0.6406 | 0.2912 | 0.0277 | 0.0071 | 0.9055 | 0.6262 |
| MF.480int.LV.Naive | Myeloid | 0.6874 | 0.6522 | 0.5241 | 0.9586 | 0.1913 | 0.3947 | 0.3585 | 0.1622 | 0.8947 | 0.8650 | 0.7742 | 0.8034 | 0.1464 | 0.2709 | 0.1938 | 0.9057 | 0.8951 |
| MF.Alv.Lu | Myeloid | 0.0143 | 0.1669 | 0.0312 | 0.4301 | 0.0408 | 0.0831 | 0.2225 | 0.0269 | 0.2281 | 0.4895 | 0.6110 | 0.7409 | 0.0013 | 0.0455 | 0.0262 | 0.9872 | 0.6734 |
| MF.AT.v2 | Myeloid | 0.3690 | 0.0366 | 0.0460 | 0.3300 | 0.0157 | 0.0200 | 0.1603 | 0.0764 | 0.5714 | 0.6550 | 0.8564 | 0.4844 | 0.0240 | 0.0215 | 0.0192 | 0.9883 | 0.3171 |
| MF.BM | Myeloid | 0.8832 | 0.9918 | 0.5240 | 0.9726 | 0.8911 | 0.6984 | 0.9416 | 0.8183 | 0.5787 | 0.7240 | 0.7028 | 0.8157 | 0.5222 | 0.2289 | 0.2343 | 0.9549 | 0.9338 |
| MF.F480hi.ctrl.PC | Myeloid | 0.2354 | 0.2347 | 0.7600 | 0.8621 | 0.0430 | 0.0406 | 0.3981 | 0.1518 | 0.9261 | 0.6652 | 0.9035 | 0.7179 | 0.0942 | 0.0554 | 0.0569 | 0.7955 | 0.7055 |
| MF.F480hi.Gata6ko.PC | Myeloid | 0.1482 | 0.0376 | 0.1677 | 0.4596 | 0.0706 | 0.0732 | 0.5774 | 0.2007 | 0.8049 | 0.8555 | 0.1563 | 0.2911 | 0.2558 | 0.0672 | 0.0631 | 0.8096 | 0.5186 |
| MF.II+480lo.PC | Myeloid | 0.6007 | 0.4174 | 0.1157 | 0.7746 | 0.5711 | 0.2569 | 0.2009 | 0.0919 | 0.6452 | 0.7054 | 0.8238 | 0.6559 | 0.3974 | 0.1827 | 0.1453 | 0.8243 | 0.3225 |
| MF.II-480hi.PC | Myeloid | 0.4099 | 0.4342 | 0.4430 | 0.7753 | 0.2728 | 0.1615 | 0.3152 | 0.1108 | 0.6739 | 0.4189 | 0.6402 | 0.4662 | 0.7197 | 0.1587 | 0.1631 | 0.8805 | 0.5197 |
| MF.Lu | Myeloid | 0.0092 | 0.0605 | 0.0625 | 0.1586 | 0.0574 | 0.2091 | 0.0590 | 0.0061 | 0.6516 | 0.4434 | 0.7169 | 0.5834 | 0.0566 | 0.2214 | 0.1181 | 0.9455 | 0.8315 |
| MF.Medl.SLN | Myeloid | 0.0632 | 0.2946 | 0.0532 | 0.7030 | 0.3030 | 0.4553 | 0.3988 | 0.2601 | 0.1209 | 0.0190 | 0.7620 | 0.1542 | 0.1569 | 0.1110 | 0.0728 | 0.9738 | 0.5474 |
| MF.Microglia.CNS | Myeloid | 0.3701 | 0.1219 | 0.3017 | 0.5765 | 0.5472 | 0.5407 | 0.3766 | 0.0994 | 0.1158 | 0.1005 | 0.1232 | 0.1468 | 0.6658 | 0.1116 | 0.1582 | 0.7326 | 0.6979 |
| MF.PPAR-.Lu | Myeloid | 0.0516 | 0.0352 | 0.0600 | 0.2869 | 0.1223 | 0.0341 | 0.0058 | 0.0033 | 0.4087 | 0.3384 | 0.8210 | 0.9007 | 0.0393 | 0.0276 | 0.0321 | 0.9723 | 0.6664 |
| MF.RP.Sp | Myeloid | 0.3774 | 0.2478 | 0.3150 | 0.4511 | 0.1315 | 0.1948 | 0.0187 | 0.0048 | 0.4273 | 0.4707 | 0.8792 | 0.6325 | 0.2000 | 0.1616 | 0.1358 | 0.8549 | 0.7629 |
| MF.Sbcaps.SLN | Myeloid | 0.6758 | 0.8602 | 0.0460 | 0.8208 | 0.4748 | 0.4727 | 0.4125 | 0.2932 | 0.3773 | 0.1832 | 0.8388 | 0.3001 | 0.4175 | 0.3235 | 0.3771 | 0.6779 | 0.5981 |
| MF.Thio5.II+480int.PC | Myeloid | 0.3011 | 0.2517 | 0.0543 | 0.3575 | 0.1285 | 0.0367 | 0.0506 | 0.0199 | 0.7087 | 0.4909 | 0.6712 | 0.5287 | 0.2067 | 0.0865 | 0.0474 | 0.9005 | 0.9636 |
| MF.Thio5.II+480lo.PC | Myeloid | 0.3481 | 0.3314 | 0.1546 | 0.6736 | 0.4405 | 0.1468 | 0.2186 | 0.1587 | 0.7937 | 0.8051 | 0.6028 | 0.4877 | 0.3133 | 0.0639 | 0.0382 | 0.8013 | 0.6094 |
| MF.Thio5.II-480hi.PC | Myeloid | 0.1563 | 0.3474 | 0.1149 | 0.4683 | 0.1255 | 0.0534 | 0.1542 | 0.0140 | 0.5575 | 0.0969 | 0.5080 | 0.2111 | 0.7089 | 0.1554 | 0.1838 | 0.5079 | 0.8795 |
| MF.Thio5.II-480int.PC | Myeloid | 0.2565 | 0.2864 | 0.0230 | 0.4094 | 0.0232 | 0.0208 | 0.0699 | 0.0142 | 0.4155 | 0.0835 | 0.2873 | 0.2529 | 0.1128 | 0.0028 | 0.0032 | 0.6610 | 0.9037 |
| Mo.6+2-.BL | Myeloid | 0.4715 | 0.0048 | 0.1553 | 0.9833 | 0.1687 | 0.0528 | 0.9222 | 0.4851 | 0.5152 | 0.5700 | 0.4724 | 0.2620 | 0.3111 | 0.0603 | 0.0453 | 0.9660 | 0.1119 |
| Mo.6+2+.BL | Myeloid | 0.2130 | 0.0085 | 0.4383 | 0.7246 | 0.4019 | 0.1379 | 0.5317 | 0.2476 | 0.8486 | 0.7771 | 0.5519 | 0.2525 | 0.0516 | 0.0809 | 0.0520 | 0.9521 | 0.1204 |
| Mo.6+2+.MLN | Myeloid | 0.3665 | 0.0187 | 0.4182 | 0.7620 | 0.3071 | 0.1160 | 0.6511 | 0.0553 | 0.4030 | 0.6835 | 0.1430 | 0.0118 | 0.1414 | 0.0149 | 0.0141 | 0.4558 | 0.1565 |
| Mo.6+2+.SLN | Myeloid | 0.1703 | 0.0270 | 0.0266 | 0.6827 | 0.0827 | 0.0843 | 0.9408 | 0.6332 | 0.6612 | 0.8142 | 0.7255 | 0.5755 | 0.0780 | 0.0838 | 0.0552 | 0.8803 | 0.4967 |
| Mo.6C+II-.Bl | Myeloid | 0.0956 | 0.0113 | 0.0304 | 0.3574 | 0.3082 | 0.2761 | 0.5473 | 0.2296 | 0.4287 | 0.3506 | 0.8859 | 0.9163 | 0.2709 | 0.0313 | 0.0053 | 0.2506 | 0.2222 |
| Mo.6C+II-.BM | Myeloid | 0.5069 | 0.8640 | 0.5102 | 0.6413 | 0.0480 | 0.0037 | 0.6201 | 0.5499 | 0.6061 | 0.5738 | 0.7351 | 0.7850 | 0.0437 | 0.3655 | 0.4425 | 0.6237 | 0.8715 |
| Mo.6C+II-.LN | Myeloid | 0.7527 | 0.2258 | 0.1577 | 0.6490 | 0.6090 | 0.5062 | 0.9607 | 0.7207 | 0.5802 | 0.6847 | 0.5414 | 0.3235 | 0.1022 | 0.0886 | 0.0456 | 0.6853 | 0.2895 |
| Mo.6C+II+.Bl | Myeloid | 0.1660 | 0.0220 | 0.0015 | 0.5315 | 0.2591 | 0.2561 | 0.1433 | 0.0717 | 0.1790 | 0.1552 | 0.5324 | 0.3150 | 0.2457 | 0.0392 | 0.0146 | 0.6510 | 0.3330 |
| Mo.6C-II-.Bl | Myeloid | 0.8591 | 0.0285 | 0.0425 | 0.9996 | 0.5259 | 0.5662 | 0.2081 | 0.1360 | 0.1333 | 0.0880 | 0.7936 | 0.5363 | 0.2430 | 0.1094 | 0.0585 | 0.4953 | 0.6565 |
| Mo.6C-II-.BM | Myeloid | 0.5335 | 0.7297 | 0.5331 | 0.7052 | 0.2283 | 0.0454 | 0.3572 | 0.2675 | 0.1866 | 0.3814 | 0.5249 | 0.6007 | 0.2671 | 0.3129 | 0.2918 | 0.7225 | 0.7734 |
| Mo.6C-II+.Bl | Myeloid | 0.8671 | 0.1534 | 0.1420 | 0.9752 | 0.4839 | 0.3613 | 0.3063 | 0.4303 | 0.4654 | 0.2702 | 0.8834 | 0.2892 | 0.6106 | 0.1019 | 0.1376 | 0.9999 | 0.4250 |
| Mo.6C-IIint.Bl | Myeloid | 0.1509 | 0.0385 | 0.0250 | 0.7764 | 0.1639 | 0.1752 | 0.3665 | 0.2027 | 0.1851 | 0.1720 | 0.7542 | 0.5485 | 0.3329 | 0.0281 | 0.0131 | 0.1071 | 0.6077 |
| Mo.Lu | Myeloid | 0.3835 | 0.0173 | 0.1322 | 0.8189 | 0.1663 | 0.0923 | 0.6960 | 0.1988 | 0.8695 | 0.8440 | 0.4829 | 0.3005 | 0.0377 | 0.0077 | 0.0092 | 0.8593 | 0.1476 |
| SC.CDP.BM | Myeloid | 0.8731 | 0.7932 | 0.6514 | 0.9197 | 0.6850 | 0.3241 | 0.2499 | 0.3693 | 0.0856 | 0.4381 | 0.4910 | 0.8961 | 0.8026 | 0.6496 | 0.7294 | 0.6095 | 0.3829 |
| SC.CMP.BM.DR | Myeloid | 0.5223 | 0.9280 | 0.2383 | 0.7852 | 0.6681 | 0.3397 | 0.2278 | 0.2509 | 0.3297 | 0.3371 | 0.6767 | 0.8302 | 0.5173 | 0.2272 | 0.4065 | 0.4664 | 0.0628 |
| SC.GMP.BM | Myeloid | 0.6851 | 0.9803 | 0.7636 | 0.9482 | 0.3264 | 0.1277 | 0.5764 | 0.4072 | 0.5900 | 0.9193 | 0.3985 | 0.8035 | 0.8899 | 0.1885 | 0.1822 | 0.2815 | 0.8993 |
| SC.MDP.BM | Myeloid | 0.9009 | 0.7394 | 0.8309 | 0.7893 | 0.9368 | 0.8534 | 0.0406 | 0.2603 | 0.0592 | 0.2690 | 0.1258 | 0.6955 | 0.6074 | 0.7162 | 0.6480 | 0.5388 | 0.5430 |
| SC.MEP.BM | Myeloid | 0.2478 | 0.3455 | 0.6222 | 0.4615 | 0.9381 | 0.8258 | 0.1077 | 0.2272 | 0.1415 | 0.4932 | 0.1963 | 0.5361 | 0.8083 | 0.1075 | 0.1593 | 0.8940 | 0.1461 |
| ILC3.NKp46-.4-.SI | NK | 0.4150 | 0.4912 | 0.6377 | 0.6552 | 0.8297 | 0.9593 | 0.2596 | 0.3874 | 0.1291 | 0.4435 | 0.0796 | 0.3011 | 0.7973 | 0.7724 | 0.6150 | 0.7401 | 0.1001 |
| ILC3.NKp46-.4+.SI | NK | 0.1168 | 0.2459 | 0.7169 | 0.8425 | 0.4542 | 0.6631 | 0.0198 | 0.0749 | 0.2389 | 0.2015 | 0.4203 | 0.2982 | 0.9383 | 0.8102 | 0.8438 | 0.6551 | 0.1288 |
| ILC3.NKp46+.Rorgthi.SI | NK | 0.6554 | 0.8254 | 0.7076 | 0.8674 | 0.5812 | 0.9186 | 0.3170 | 0.7061 | 0.2542 | 0.7393 | 0.2079 | 0.3486 | 0.8865 | 0.8910 | 0.8810 | 0.0525 | 0.6273 |
| NK.49CI-.Sp | NK | 0.8023 | 0.6825 | 0.3911 | 0.9257 | 0.3625 | 0.4945 | 0.4699 | 0.0835 | 0.2102 | 0.0650 | 0.6245 | 0.2838 | 0.9123 | 0.9690 | 0.9539 | 0.5412 | 0.8590 |
| NK.49CI+.Sp | NK | 0.9004 | 0.6777 | 0.1689 | 0.9933 | 0.1559 | 0.2757 | 0.8991 | 0.7042 | 0.1772 | 0.1407 | 0.3524 | 0.2076 | 0.7609 | 0.4349 | 0.6671 | 0.7620 | 0.7230 |
| NK.49H-.Sp | NK | 0.9973 | 0.1843 | 0.0219 | 0.9832 | 0.4096 | 0.2679 | 0.8372 | 0.5667 | 0.0218 | 0.0075 | 0.1376 | 0.3906 | 0.8754 | 0.9718 | 0.9864 | 0.8868 | 0.6271 |

| | | | | | | | | | | | | | | | | | | |
|---|---|---|---|---|---|---|---|---|---|---|---|---|---|---|---|---|---|---|
| NK.49H+.Sp | NK | 0.9983 | 0.8596 | 0.1974 | 0.9739 | 0.3981 | 0.1518 | 0.7118 | 0.5831 | 0.4559 | 0.0668 | 0.3463 | 0.7351 | 0.7977 | 0.7024 | 0.8555 | 0.3776 | 0.6326 |
| NK.b2m-.Sp | NK | 0.9591 | 0.6869 | 0.2753 | 0.9915 | 0.3224 | 0.4917 | 0.5087 | 0.4262 | 0.0117 | 0.0068 | 0.7259 | 0.6156 | 0.9257 | 0.9640 | 0.9932 | 0.6972 | 0.8957 |
| NK.CD127-.SI | NK | 0.2852 | 0.6304 | 0.1366 | 0.8901 | 0.2934 | 0.7372 | 0.0715 | 0.0112 | 0.3779 | 0.9411 | 0.3969 | 0.2325 | 0.9739 | 0.2231 | 0.2217 | 0.0446 | 0.2410 |
| NK.CD127-.Sp | NK | 0.9986 | 0.9270 | 0.8128 | 0.9972 | 0.8685 | 0.9178 | 0.3536 | 0.0600 | 0.1180 | 0.4731 | 0.7913 | 0.9612 | 0.8123 | 0.4856 | 0.4867 | 0.3846 | 0.1394 |
| NK.CD49b+.Lv | NK | 0.9427 | 0.9904 | 0.7923 | 0.9222 | 0.9345 | 0.9636 | 0.4735 | 0.0515 | 0.5189 | 0.6578 | 0.5975 | 0.6643 | 0.7643 | 0.2772 | 0.3646 | 0.1689 | 0.0356 |
| NK.DAP10-.Sp | NK | 0.9991 | 0.8884 | 0.1775 | 0.9986 | 0.4531 | 0.4841 | 0.1476 | 0.0409 | 0.4775 | 0.5228 | 0.5881 | 0.2315 | 0.9841 | 0.8502 | 0.9265 | 0.4861 | 0.2572 |
| NK.DAP12-.Sp | NK | 0.9846 | 0.3176 | 0.1165 | 0.9939 | 0.2897 | 0.6703 | 0.7099 | 0.4781 | 0.4069 | 0.1213 | 0.4673 | 0.5474 | 0.7957 | 0.9867 | 0.9957 | 0.9488 | 0.9540 |
| NK.H+.MCMV1.Sp | NK | 0.6376 | 0.3549 | 0.1346 | 0.6159 | 0.4057 | 0.4298 | 0.7528 | 0.7319 | 0.2276 | 0.1117 | 0.3049 | 0.0494 | 0.6328 | 0.9860 | 0.9840 | 0.9250 | 0.0925 |
| NK.H+.MCMV7.Sp | NK | 0.8787 | 0.8133 | 0.1661 | 0.9745 | 0.1696 | 0.3891 | 0.8425 | 0.0386 | 0.0816 | 0.0024 | 0.3962 | 0.3207 | 0.2782 | 0.8697 | 0.9056 | 0.9133 | 0.8182 |
| NK.MCMV1.Sp | NK | 0.9334 | 0.3100 | 0.0869 | 0.9782 | 0.4631 | 0.3303 | 0.2284 | 0.1359 | 0.1994 | 0.4742 | 0.6338 | 0.3715 | 0.3908 | 0.9840 | 0.9942 | 0.9125 | 0.1151 |
| NK.MCMV7.Sp | NK | 0.8825 | 0.7363 | 0.2089 | 0.8838 | 0.5730 | 0.6918 | 0.3241 | 0.2539 | 0.5698 | 0.1319 | 0.2134 | 0.4933 | 0.7425 | 0.8319 | 0.8787 | 0.6860 | 0.6626 |
| NK.Sp | NK | 0.9045 | 0.6159 | 0.3799 | 0.9135 | 0.4300 | 0.4477 | 0.7133 | 0.6838 | 0.4321 | 0.1102 | 0.6155 | 0.1261 | 0.7491 | 0.7570 | 0.7007 | 0.7386 | 0.5227 |
| NKT.4-.Lv | NK | 0.4772 | 0.2760 | 0.3736 | 0.4718 | 0.7705 | 0.8152 | 0.2796 | 0.6050 | 0.5676 | 0.6329 | 0.1276 | 0.0217 | 0.8398 | 0.9875 | 0.9820 | 0.6950 | 0.5268 |
| NKT.4-.Sp | NK | 0.8646 | 0.1881 | 0.4708 | 0.3829 | 0.3645 | 0.4383 | 0.4170 | 0.3079 | 0.3927 | 0.6761 | 0.3941 | 0.2790 | 0.7103 | 0.9284 | 0.9123 | 0.1214 | 0.6603 |
| NKT.4+.Lv | NK | 0.4517 | 0.1225 | 0.3667 | 0.2976 | 0.1112 | 0.1523 | 0.1555 | 0.4831 | 0.1883 | 0.5293 | 0.1971 | 0.0427 | 0.9219 | 0.9993 | 0.9991 | 0.1761 | 0.3840 |
| NKT.4+.Sp | NK | 0.7490 | 0.1879 | 0.4391 | 0.7315 | 0.9863 | 0.9062 | 0.9371 | 0.8450 | 0.3912 | 0.5535 | 0.6551 | 0.6219 | 0.9948 | 0.6826 | 0.6367 | 0.9094 | 0.8066 |
| NKT.44+NK1.1-.Th | NK | 0.4151 | 0.1460 | 0.2788 | 0.3653 | 0.6161 | 0.6166 | 0.1639 | 0.2244 | 0.4276 | 0.7820 | 0.3085 | 0.1175 | 0.7712 | 0.8090 | 0.8253 | 0.4236 | 0.0162 |
| NKT.44+NK1.1+.Th | NK | 0.8459 | 0.0688 | 0.4441 | 0.8366 | 0.6040 | 0.4618 | 0.2557 | 0.2086 | 0.7961 | 0.4756 | 0.3338 | 0.1466 | 0.9642 | 0.9725 | 0.9202 | 0.6991 | 0.8065 |
| NKT.44-NK1.1-.Th | NK | 0.3641 | 0.1263 | 0.6876 | 0.5601 | 0.2665 | 0.6825 | 0.4527 | 0.1626 | 0.5870 | 0.5785 | 0.1478 | 0.0494 | 0.3651 | 0.8819 | 0.8849 | 0.2338 | 0.1962 |
| BEC.MLN | Other | 0.0250 | 0.5907 | 0.6299 | 0.3133 | 0.9050 | 0.8633 | 0.9148 | 0.5235 | 0.6049 | 0.4414 | 0.9107 | 0.9160 | 0.5935 | 0.4368 | 0.4445 | 0.3855 | 0.9577 |
| BEC.SLN | Other | 0.0223 | 0.6153 | 0.5315 | 0.2420 | 0.8795 | 0.8096 | 0.8989 | 0.5299 | 0.5192 | 0.5546 | 0.7077 | 0.9156 | 0.7162 | 0.5317 | 0.6412 | 0.4374 | 0.9826 |
| BEC.SLN.OT | Other | 0.0216 | 0.7713 | 0.4614 | 0.1418 | 0.9197 | 0.9219 | 0.7124 | 0.5171 | 0.6428 | 0.6729 | 0.7896 | 0.8656 | 0.3269 | 0.5922 | 0.6720 | 0.5078 | 0.9460 |
| DN.SLN.CFA.d6.v2 | Other | 0.6312 | 0.8863 | 0.4970 | 0.0773 | 0.8841 | 0.8738 | 0.8841 | 0.9483 | 0.8167 | 0.7051 | 0.3578 | 0.3608 | 0.3161 | 0.7792 | 0.7242 | 0.4406 | 0.9558 |
| DN.SLN.v2 | Other | 0.5840 | 0.8502 | 0.5244 | 0.0684 | 0.8695 | 0.7476 | 0.8272 | 0.9411 | 0.5808 | 0.4779 | 0.2877 | 0.3444 | 0.5291 | 0.5657 | 0.5603 | 0.4634 | 0.9278 |
| Ep.5wk.MEC.Sca1+.Th | Other | 0.3534 | 0.2106 | 0.3350 | 0.1704 | 0.9553 | 0.8746 | 0.5018 | 0.5288 | 0.2916 | 0.1501 | 0.4179 | 0.7979 | 0.6814 | 0.3587 | 0.2362 | 0.3873 | 0.9882 |
| Ep.5wk.MEChi.Th | Other | 0.6879 | 0.5445 | 0.8978 | 0.1034 | 0.9668 | 0.8561 | 0.4817 | 0.6424 | 0.9196 | 0.8768 | 0.0645 | 0.5445 | 0.4722 | 0.5864 | 0.4082 | 0.3918 | 0.8640 |
| Ep.5wk.MEClo.Th | Other | 0.8002 | 0.7518 | 0.8695 | 0.3394 | 0.9800 | 0.9392 | 0.4769 | 0.7794 | 0.8132 | 0.7904 | 0.5939 | 0.8410 | 0.7650 | 0.3082 | 0.1935 | 0.7355 | 0.9991 |
| Ep.8wk.CEC.Sca1+.Th | Other | 0.2205 | 0.3650 | 0.2702 | 0.0940 | 0.6519 | 0.4197 | 0.8985 | 0.8707 | 0.7907 | 0.3009 | 0.2874 | 0.6224 | 0.5763 | 0.6803 | 0.6716 | 0.0941 | 0.7738 |
| Ep.8wk.CEChi.Th | Other | 0.1341 | 0.1859 | 0.5177 | 0.2422 | 0.6359 | 0.6147 | 0.9705 | 0.9650 | 0.8952 | 0.8235 | 0.4184 | 0.6030 | 0.2402 | 0.2547 | 0.1796 | 0.6457 | 0.9925 |
| Ep.8wk.MEChi.Th | Other | 0.6507 | 0.4723 | 0.9151 | 0.2346 | 0.9667 | 0.8906 | 0.4993 | 0.7607 | 0.8622 | 0.8263 | 0.0625 | 0.5474 | 0.6012 | 0.1810 | 0.1753 | 0.3109 | 0.9169 |
| Ep.8wk.MEClo.Th | Other | 0.8156 | 0.6971 | 0.9427 | 0.4709 | 0.9042 | 0.7049 | 0.5908 | 0.7436 | 0.6886 | 0.7838 | 0.4183 | 0.7143 | 0.8265 | 0.2592 | 0.1503 | 0.4218 | 0.9996 |
| Ep.MEChi.Th | Other | 0.7587 | 0.4955 | 0.7259 | 0.1399 | 0.8461 | 0.7867 | 0.1497 | 0.6144 | 0.7469 | 0.8087 | 0.6530 | 0.9579 | 0.9126 | 0.2706 | 0.1027 | 0.4234 | 0.8783 |
| Fi.MTS15+.Th | Other | 0.1077 | 0.5550 | 0.0139 | 0.1148 | 0.5863 | 0.7289 | 0.7881 | 0.3345 | 0.5387 | 0.5671 | 0.2301 | 0.5216 | 0.4422 | 0.6678 | 0.6254 | 0.1684 | 0.9770 |
| Fi.Sk | Other | 0.0319 | 0.3854 | 0.1142 | 0.0263 | 0.2783 | 0.3754 | 0.9884 | 0.6813 | 0.6278 | 0.1473 | 0.1180 | 0.6391 | 0.5959 | 0.6653 | 0.6950 | 0.0853 | 0.8969 |
| FRC.Cad11.WT.v2 | Other | 0.3087 | 0.6606 | 0.0716 | 0.2233 | 0.9627 | 0.8689 | 0.9893 | 0.9411 | 0.3505 | 0.2746 | 0.2782 | 0.5138 | 0.5396 | 0.4921 | 0.3253 | 0.1928 | 0.9809 |
| FRC.MLN | Other | 0.1541 | 0.3847 | 0.1117 | 0.1240 | 0.8563 | 0.8185 | 0.9604 | 0.7779 | 0.6389 | 0.6167 | 0.2837 | 0.7455 | 0.9133 | 0.7132 | 0.7712 | 0.3203 | 0.9873 |
| FRC.SLN | Other | 0.1169 | 0.3131 | 0.0737 | 0.0920 | 0.8982 | 0.8000 | 0.9742 | 0.9155 | 0.7362 | 0.5890 | 0.2703 | 0.6359 | 0.8955 | 0.3440 | 0.4713 | 0.4814 | 0.9030 |
| FRC.SLN.CFA.d6.v2 | Other | 0.1281 | 0.6748 | 0.2648 | 0.0922 | 0.8565 | 0.5320 | 0.9882 | 0.9052 | 0.7773 | 0.4101 | 0.1516 | 0.3176 | 0.4510 | 0.2224 | 0.1896 | 0.3003 | 0.9858 |
| FRC.SLN.OT | Other | 0.1306 | 0.4093 | 0.1608 | 0.2072 | 0.8047 | 0.7703 | 0.9427 | 0.8830 | 0.6121 | 0.6296 | 0.1816 | 0.6935 | 0.8601 | 0.3796 | 0.4940 | 0.2952 | 0.9735 |
| FRC.SLN.v2 | Other | 0.1890 | 0.7458 | 0.2683 | 0.2543 | 0.9379 | 0.6877 | 0.9816 | 0.9441 | 0.4618 | 0.4157 | 0.1110 | 0.4500 | 0.5410 | 0.3867 | 0.3831 | 0.2279 | 0.9803 |
| ILC1.CD127+.SI | Other | 0.8099 | 0.0332 | 0.3236 | 0.9130 | 0.3033 | 0.6653 | 0.0477 | 0.1465 | 0.2416 | 0.3665 | 0.0501 | 0.3753 | 0.9879 | 0.8726 | 0.8380 | 0.2764 | 0.1956 |
| ILC1.CD127+.Sp | Other | 0.6619 | 0.5299 | 0.1430 | 0.8498 | 0.8013 | 0.9634 | 0.4863 | 0.0727 | 0.3368 | 0.6717 | 0.1078 | 0.1715 | 0.9969 | 0.4265 | 0.5094 | 0.0664 | 0.4264 |
| ILC1.CD49b-.Lv | Other | 0.9948 | 0.8127 | 0.8434 | 0.8865 | 0.8308 | 0.9753 | 0.4737 | 0.0473 | 0.2818 | 0.4542 | 0.8380 | 0.7106 | 0.7958 | 0.1225 | 0.1147 | 0.0753 | 0.0501 |
| ILC2.SI | Other | 0.5129 | 0.7064 | 0.9270 | 0.8659 | 0.9380 | 0.9628 | 0.2340 | 0.0291 | 0.6582 | 0.8845 | 0.2154 | 0.0984 | 0.8173 | 0.2047 | 0.2058 | 0.2077 | 0.0899 |
| LEC.MLN | Other | 0.0358 | 0.8813 | 0.6561 | 0.0391 | 0.7992 | 0.9477 | 0.7418 | 0.2500 | 0.8368 | 0.4838 | 0.1436 | 0.4892 | 0.3560 | 0.7401 | 0.6450 | 0.6420 | 0.9920 |
| LEC.SLN | Other | 0.0062 | 0.8219 | 0.7764 | 0.0073 | 0.6154 | 0.8293 | 0.6863 | 0.3868 | 0.7174 | 0.3925 | 0.0928 | 0.4985 | 0.4907 | 0.3952 | 0.4415 | 0.2235 | 0.9945 |
| LEC.SLN.CFA.d6.v2 | Other | 0.0175 | 0.9584 | 0.6165 | 0.0106 | 0.9534 | 0.9527 | 0.7727 | 0.6751 | 0.7226 | 0.6082 | 0.0857 | 0.3492 | 0.3050 | 0.1777 | 0.1139 | 0.2212 | 0.9979 |
| LEC.SLN.OT | Other | 0.0010 | 0.7508 | 0.5202 | 0.0145 | 0.8200 | 0.9679 | 0.6388 | 0.2948 | 0.6762 | 0.4623 | 0.1367 | 0.4441 | 0.5089 | 0.6975 | 0.6997 | 0.5141 | 0.9880 |
| LEC.SLN.v2 | Other | 0.0011 | 0.9287 | 0.8735 | 0.0026 | 0.7647 | 0.7438 | 0.5118 | 0.4020 | 0.3055 | 0.3580 | 0.0502 | 0.1374 | 0.5563 | 0.3565 | 0.1814 | 0.7622 | 0.9982 |
| MEChi.GFP-.Adult | Other | 0.8907 | 0.9200 | 0.7011 | 0.1908 | 0.9026 | 0.7951 | 0.6353 | 0.8672 | 0.7805 | 0.6791 | 0.6525 | 0.9742 | 0.6692 | 0.3036 | 0.2599 | 0.4032 | 0.8520 |
| MEChi.GFP+.Adult | Other | 0.7500 | 0.4689 | 0.8739 | 0.2874 | 0.9679 | 0.9528 | 0.3904 | 0.7412 | 0.8965 | 0.7738 | 0.3472 | 0.6777 | 0.5401 | 0.3129 | 0.2377 | 0.6572 | 0.6715 |
| MEChi.GFP+.Adult.KO | Other | 0.3452 | 0.7533 | 0.6805 | 0.1109 | 0.9510 | 0.9858 | 0.4732 | 0.7576 | 0.8871 | 0.8914 | 0.6461 | 0.7463 | 0.7695 | 0.3505 | 0.2448 | 0.3152 | 0.8901 |
| MLP.BM | Other | 0.6960 | 0.9636 | 0.4658 | 0.6588 | 0.7290 | 0.6400 | 0.0703 | 0.2624 | 0.2486 | 0.6242 | 0.0794 | 0.3088 | 0.5820 | 0.4734 | 0.5637 | 0.8109 | 0.6596 |
| MLP.FL | Other | 0.2974 | 0.8135 | 0.5293 | 0.5134 | 0.7879 | 0.5370 | 0.1541 | 0.2745 | 0.0107 | 0.5376 | 0.4217 | 0.1166 | 0.5555 | 0.1590 | 0.2985 | 0.9109 | 0.1745 |
| SC.LT34F.BM | Other | 0.8910 | 0.7439 | 0.8005 | 0.8406 | 0.9674 | 0.9832 | 0.5545 | 0.5534 | 0.4235 | 0.9371 | 0.7283 | 0.8040 | 0.2166 | 0.4322 | 0.6579 | 0.7740 | 0.7067 |
| SC.LTSL.FL | Other | 0.6814 | 0.2796 | 0.5867 | 0.4866 | 0.9480 | 0.9114 | 0.0845 | 0.1433 | 0.0492 | 0.8320 | 0.9614 | 0.8407 | 0.0048 | 0.8459 | 0.8731 | 0.9476 | 0.2147 |
| SC.MPP34F.BM | Other | 0.0641 | 0.8094 | 0.2466 | 0.2118 | 0.4563 | 0.3677 | 0.0785 | 0.3907 | 0.3262 | 0.7170 | 0.2371 | 0.2998 | 0.7599 | 0.6034 | 0.6830 | 0.2779 | 0.0810 |
| SC.ST34F.BM | Other | 0.2618 | 0.8728 | 0.1141 | 0.3260 | 0.8990 | 0.4438 | 0.0874 | 0.7689 | 0.4093 | 0.7424 | 0.5004 | 0.7768 | 0.6986 | 0.8083 | 0.9281 | 0.8586 | 0.3142 |
| SC.STSL.BM | Other | 0.8376 | 0.9214 | 0.6955 | 0.8958 | 0.7276 | 0.3468 | 0.1403 | 0.5345 | 0.8656 | 0.6403 | 0.8264 | 0.8160 | 0.3475 | 0.6176 | 0.6920 | 0.3571 | 0.5542 |
| SC.STSL.FL | Other | 0.7632 | 0.2509 | 0.5809 | 0.7073 | 0.9055 | 0.9115 | 0.0835 | 0.1721 | 0.0718 | 0.7702 | 0.9516 | 0.5790 | 0.2850 | 0.8790 | 0.9019 | 0.9608 | 0.0781 |
| St.31-38-44-.SLN | Other | 0.6342 | 0.8523 | 0.5447 | 0.2605 | 0.9465 | 0.8925 | 0.9562 | 0.9324 | 0.9131 | 0.7831 | 0.3587 | 0.3753 | 0.4129 | 0.2708 | 0.5027 | 0.5787 | 0.9520 |
| ABD.TR.14w.B6 | T | 0.7011 | 0.1128 | 0.3146 | 0.6315 | 0.8479 | 0.9314 | 0.2339 | 0.4369 | 0.3749 | 0.7989 | 0.2820 | 0.2304 | 0.6274 | 0.8664 | 0.9357 | 0.6791 | 0.9617 |
| CD4.1h.LN | T | 0.9484 | 0.8211 | 0.8818 | 0.6847 | 0.8293 | 0.8872 | 0.3741 | 0.2529 | 0.4115 | 0.7286 | 0.8191 | 0.4638 | 0.6992 | 0.1561 | 0.1112 | 0.4621 | 0.3986 |
| CD4.24h.LN | T | 0.5108 | 0.0926 | 0.3615 | 0.7743 | 0.2989 | 0.3938 | 0.9357 | 0.7524 | 0.7031 | 0.7297 | 0.3429 | 0.2468 | 0.9592 | 0.0528 | 0.0304 | 0.4808 | 0.4107 |

| | | | | | | | | | | | | | | | | | | |
|---|---|---|---|---|---|---|---|---|---|---|---|---|---|---|---|---|---|---|
| CD4.48h.LN | T | 0.7142 | 0.3055 | 0.0879 | 0.8659 | 0.2424 | 0.2421 | 0.9450 | 0.9948 | 0.0656 | 0.5280 | 0.4764 | 0.7066 | 0.2914 | 0.7604 | 0.6931 | 0.3979 | 0.4074 |
| CD4.5h.LN | T | 0.1586 | 0.6293 | 0.4733 | 0.0836 | 0.7478 | 0.5672 | 0.8361 | 0.8797 | 0.8850 | 0.5703 | 0.3098 | 0.1541 | 0.5019 | 0.2255 | 0.1819 | 0.2204 | 0.9188 |
| CD4.96h.LN | T | 0.3604 | 0.5747 | 0.9098 | 0.6522 | 0.4942 | 0.4553 | 0.7338 | 0.6885 | 0.3500 | 0.4544 | 0.4545 | 0.2973 | 0.3418 | 0.5399 | 0.7467 | 0.4158 | 0.6754 |
| CD4.CTR.LN | T | 0.3039 | 0.3325 | 0.1704 | 0.2438 | 0.6941 | 0.6562 | 0.9567 | 0.6713 | 0.0768 | 0.2969 | 0.6070 | 0.5092 | 0.6342 | 0.1082 | 0.1015 | 0.2262 | 0.8389 |
| CD4Control | T | 0.4013 | 0.4522 | 0.8330 | 0.4752 | 0.2395 | 0.2245 | 0.6129 | 0.8470 | 0.7348 | 0.5463 | 0.4280 | 0.4211 | 0.7507 | 0.9701 | 0.9841 | 0.4431 | 0.5418 |
| CD8.1h.LN | T | 0.2237 | 0.4849 | 0.6785 | 0.4580 | 0.8878 | 0.8161 | 0.5080 | 0.2372 | 0.8010 | 0.6118 | 0.5163 | 0.5324 | 0.5455 | 0.6240 | 0.4620 | 0.4190 | 0.9581 |
| CD8.24h.LN | T | 0.2379 | 0.2123 | 0.1285 | 0.8717 | 0.2255 | 0.3524 | 0.8885 | 0.7357 | 0.6126 | 0.3900 | 0.5401 | 0.3071 | 0.9356 | 0.1055 | 0.0451 | 0.4630 | 0.2796 |
| CD8.48h.LN | T | 0.7964 | 0.4273 | 0.0513 | 0.8626 | 0.1160 | 0.1337 | 0.7755 | 0.9557 | 0.1196 | 0.1273 | 0.4237 | 0.2169 | 0.0939 | 0.6984 | 0.6564 | 0.6707 | 0.7367 |
| CD8.5h.LN | T | 0.6631 | 0.5619 | 0.1575 | 0.5608 | 0.7130 | 0.6229 | 0.8714 | 0.8690 | 0.5766 | 0.3495 | 0.5718 | 0.5200 | 0.4611 | 0.1521 | 0.1059 | 0.4552 | 0.7949 |
| CD8.96h.LN | T | 0.7907 | 0.5881 | 0.9348 | 0.8911 | 0.7911 | 0.7112 | 0.9208 | 0.6980 | 0.3859 | 0.2879 | 0.4369 | 0.1921 | 0.6951 | 0.8063 | 0.7887 | 0.2181 | 0.7279 |
| CD8.CTR.LN | T | 0.5251 | 0.4769 | 0.1819 | 0.6631 | 0.6778 | 0.8107 | 0.8859 | 0.5686 | 0.3877 | 0.4957 | 0.5781 | 0.3424 | 0.8216 | 0.5037 | 0.4659 | 0.1119 | 0.9905 |
| LN.TR.14w.B6 | T | 0.6875 | 0.7525 | 0.4914 | 0.2117 | 0.2456 | 0.3799 | 0.5540 | 0.7318 | 0.9397 | 0.9419 | 0.5754 | 0.3555 | 0.8771 | 0.8600 | 0.9481 | 0.1508 | 0.1390 |
| preT.DN2.Th | T | 0.6882 | 0.5053 | 0.6047 | 0.6322 | 0.4765 | 0.6281 | 0.3159 | 0.6689 | 0.3190 | 0.2274 | 0.0354 | 0.2111 | 0.9043 | 0.7073 | 0.7728 | 0.7644 | 0.8852 |
| preT.DN2-3.Th | T | 0.9646 | 0.6511 | 0.5000 | 0.9097 | 0.7671 | 0.6958 | 0.9815 | 0.9321 | 0.1728 | 0.2109 | 0.2773 | 0.3977 | 0.7290 | 0.9169 | 0.9482 | 0.7984 | 0.8070 |
| preT.DN2A.Th | T | 0.5234 | 0.3119 | 0.5549 | 0.5527 | 0.5442 | 0.4684 | 0.0433 | 0.3590 | 0.3546 | 0.1481 | 0.0411 | 0.1815 | 0.8105 | 0.5879 | 0.5938 | 0.5867 | 0.7108 |
| preT.DN2B.Th | T | 0.3102 | 0.5075 | 0.3538 | 0.3700 | 0.6140 | 0.4368 | 0.0465 | 0.4510 | 0.5205 | 0.3856 | 0.2165 | 0.4371 | 0.5524 | 0.5711 | 0.5833 | 0.8414 | 0.8184 |
| preT.DN3-4.Th | T | 0.4694 | 0.3273 | 0.1986 | 0.7703 | 0.3194 | 0.5085 | 0.4525 | 0.7058 | 0.2710 | 0.1017 | 0.0335 | 0.1716 | 0.1610 | 0.1661 | 0.2259 | 0.5982 | 0.3030 |
| preT.DN3A.Th | T | 0.9585 | 0.9881 | 0.6949 | 0.9138 | 0.7364 | 0.5629 | 0.4743 | 0.9638 | 0.4190 | 0.5921 | 0.6937 | 0.5936 | 0.8482 | 0.7054 | 0.8096 | 0.4652 | 0.3766 |
| preT.DN3B.Th | T | 0.4065 | 0.4473 | 0.3438 | 0.8730 | 0.5475 | 0.7412 | 0.6277 | 0.6661 | 0.1339 | 0.1427 | 0.1186 | 0.3468 | 0.5691 | 0.5984 | 0.5958 | 0.6651 | 0.5718 |
| preT.ETP.Th | T | 0.4127 | 0.8607 | 0.6108 | 0.6562 | 0.4671 | 0.4157 | 0.0836 | 0.1923 | 0.2888 | 0.1620 | 0.0676 | 0.3175 | 0.8752 | 0.8327 | 0.8962 | 0.4669 | 0.8933 |
| preT.ETP-2A.Th | T | 0.8472 | 0.3154 | 0.8898 | 0.9490 | 0.7172 | 0.5891 | 0.1362 | 0.1297 | 0.3286 | 0.4449 | 0.2347 | 0.4600 | 0.7384 | 0.9649 | 0.9384 | 0.0160 | 0.9715 |
| T.4.LN.BDC | T | 0.6217 | 0.2328 | 0.7809 | 0.6818 | 0.6262 | 0.5986 | 0.6911 | 0.8265 | 0.7160 | 0.2018 | 0.2291 | 0.4171 | 0.7159 | 0.8281 | 0.8744 | 0.6096 | 0.6447 |
| T.4.Pa.BDC | T | 0.9944 | 0.2751 | 0.7977 | 0.8189 | 0.2650 | 0.3472 | 0.4810 | 0.6563 | 0.5898 | 0.3454 | 0.4022 | 0.2834 | 0.6839 | 0.8394 | 0.8821 | 0.3265 | 0.2378 |
| T.4.PLN.BDC | T | 0.9737 | 0.4754 | 0.8269 | 0.8329 | 0.6320 | 0.6955 | 0.7788 | 0.8102 | 0.7992 | 0.5050 | 0.8570 | 0.9318 | 0.7205 | 0.9689 | 0.9586 | 0.8912 | 0.7342 |
| T.4+8int.Th | T | 0.9864 | 0.6466 | 0.9829 | 0.7516 | 0.6952 | 0.8685 | 0.0929 | 0.4009 | 0.1942 | 0.1730 | 0.7812 | 0.2372 | 0.2933 | 0.9954 | 0.9986 | 0.7769 | 0.3985 |
| T.4Eff49d+11a+.Sp.d8.LCMV | T | 0.9154 | 0.0502 | 0.0129 | 0.8796 | 0.1377 | 0.1695 | 0.9863 | 0.9420 | 0.3864 | 0.5469 | 0.6366 | 0.2512 | 0.3578 | 0.9084 | 0.8718 | 0.1730 | 0.8124 |
| T.4FP3-.Sp | T | 0.7464 | 0.2314 | 0.9608 | 0.9281 | 0.9025 | 0.8557 | 0.5355 | 0.2449 | 0.6605 | 0.1478 | 0.6458 | 0.4101 | 0.3243 | 0.9620 | 0.9784 | 0.8192 | 0.6512 |
| T.4FP3+25+.Sp | T | 0.5849 | 0.3352 | 0.3314 | 0.6971 | 0.4609 | 0.5024 | 0.4725 | 0.4234 | 0.9571 | 0.9801 | 0.5712 | 0.5519 | 0.5085 | 0.8842 | 0.8689 | 0.8100 | 0.7076 |
| T.4int8+.Th | T | 0.8660 | 0.0634 | 0.2583 | 0.3889 | 0.2331 | 0.3579 | 0.6640 | 0.9039 | 0.2910 | 0.1679 | 0.3535 | 0.3125 | 0.6938 | 0.8924 | 0.9638 | 0.6441 | 0.3980 |
| T.4Mem.LN | T | 0.9503 | 0.3408 | 0.5924 | 0.8832 | 0.2758 | 0.3429 | 0.5662 | 0.5203 | 0.9464 | 0.9809 | 0.4293 | 0.8588 | 0.6856 | 0.6936 | 0.6905 | 0.7833 | 0.4939 |
| T.4Mem.Sp | T | 0.9029 | 0.0162 | 0.2084 | 0.7127 | 0.2060 | 0.1491 | 0.4701 | 0.9287 | 0.6790 | 0.7760 | 0.5315 | 0.6299 | 0.5326 | 0.9782 | 0.9560 | 0.0680 | 0.4956 |
| T.4Mem44h62l.LN | T | 0.9320 | 0.1424 | 0.3686 | 0.3894 | 0.3026 | 0.0855 | 0.6644 | 0.7779 | 0.9028 | 0.8314 | 0.2917 | 0.4422 | 0.9253 | 0.7023 | 0.6659 | 0.9089 | 0.7085 |
| T.4Mem44h62l.Sp | T | 0.9056 | 0.1010 | 0.4664 | 0.5070 | 0.7831 | 0.7383 | 0.8909 | 0.8307 | 0.5975 | 0.8928 | 0.6681 | 0.4458 | 0.8995 | 0.9589 | 0.9057 | 0.9156 | 0.2597 |
| T.4Mem49d+11a+.Sp.d30.LCMV | T | 0.8246 | 0.4205 | 0.3596 | 0.9082 | 0.4191 | 0.1809 | 0.9981 | 0.9699 | 0.7505 | 0.6170 | 0.5684 | 0.7721 | 0.3903 | 0.9584 | 0.9658 | 0.2082 | 0.6147 |
| T.4Nve.LN | T | 0.8299 | 0.5686 | 0.8848 | 0.7813 | 0.8788 | 0.4626 | 0.5036 | 0.5978 | 0.9310 | 0.6507 | 0.5531 | 0.8707 | 0.6188 | 0.7456 | 0.6851 | 0.9291 | 0.8810 |
| T.4Nve.MLN | T | 0.7854 | 0.2791 | 0.8026 | 0.6721 | 0.5854 | 0.5836 | 0.7445 | 0.5008 | 0.6237 | 0.0931 | 0.2292 | 0.2091 | 0.6415 | 0.5848 | 0.6119 | 0.8738 | 0.7387 |
| T.4Nve.PP | T | 0.3674 | 0.3392 | 0.7531 | 0.1384 | 0.2774 | 0.4335 | 0.1375 | 0.2564 | 0.8506 | 0.5371 | 0.8155 | 0.6924 | 0.8987 | 0.7528 | 0.8800 | 0.6145 | 0.1941 |
| T.4Nve.Sp | T | 0.8191 | 0.7196 | 0.9750 | 0.7639 | 0.9197 | 0.9494 | 0.1680 | 0.7792 | 0.7983 | 0.3665 | 0.6016 | 0.5565 | 0.9077 | 0.8813 | 0.9119 | 0.2139 | 0.1720 |
| T.4Nve44-49d-11a-.Sp | T | 0.8458 | 0.7421 | 0.8728 | 0.7050 | 0.5725 | 0.2929 | 0.9815 | 0.6859 | 0.4233 | 0.2222 | 0.3431 | 0.2007 | 0.3087 | 0.7707 | 0.8531 | 0.1594 | 0.6978 |
| T.4SP24-.Th | T | 0.9723 | 0.0656 | 0.3766 | 0.7239 | 0.4427 | 0.4295 | 0.9612 | 0.9128 | 0.8975 | 0.6833 | 0.5816 | 0.8360 | 0.6172 | 0.8225 | 0.7589 | 0.8112 | 0.9451 |
| T.4SP24int.Th | T | 0.9974 | 0.0926 | 0.3711 | 0.5299 | 0.2648 | 0.2064 | 0.7841 | 0.8834 | 0.6143 | 0.9271 | 0.5245 | 0.7104 | 0.6596 | 0.6255 | 0.6510 | 0.6341 | 0.8416 |
| T.4SP69+.Th | T | 0.9920 | 0.1653 | 0.4906 | 0.3672 | 0.6526 | 0.4580 | 0.4181 | 0.6575 | 0.6151 | 0.4652 | 0.5971 | 0.7666 | 0.6671 | 0.7957 | 0.7915 | 0.7925 | 0.4512 |
| T.8Eff.Sp.OT1.12hr.LisOva | T | 0.4557 | 0.8363 | 0.5468 | 0.1997 | 0.9868 | 0.9965 | 0.4865 | 0.8007 | 0.6847 | 0.3318 | 0.7248 | 0.2272 | 0.9451 | 0.1040 | 0.0669 | 0.4186 | 0.2037 |
| T.8Eff.Sp.OT1.24hr.LisOva | T | 0.5981 | 0.8384 | 0.8243 | 0.4381 | 0.9130 | 0.9383 | 0.4858 | 0.7611 | 0.7873 | 0.4301 | 0.5858 | 0.2101 | 0.8338 | 0.1084 | 0.0398 | 0.6190 | 0.1686 |
| T.8Eff.Sp.OT1.48hr.LisOva | T | 0.2558 | 0.3634 | 0.6569 | 0.7647 | 0.7986 | 0.9267 | 0.7179 | 0.5671 | 0.5440 | 0.2668 | 0.2652 | 0.2705 | 0.7977 | 0.1254 | 0.1381 | 0.5989 | 0.4873 |
| T.8Eff.Sp.OT1.d10.LisOva | T | 0.8823 | 0.3779 | 0.3889 | 0.7717 | 0.3964 | 0.4553 | 0.7678 | 0.2554 | 0.5910 | 0.1502 | 0.6377 | 0.4897 | 0.3661 | 0.8827 | 0.9766 | 0.2328 | 0.4983 |
| T.8Eff.Sp.OT1.d15.LisOva | T | 0.5689 | 0.3521 | 0.5632 | 0.7455 | 0.2173 | 0.1637 | 0.6078 | 0.3611 | 0.1905 | 0.0957 | 0.3552 | 0.2569 | 0.7859 | 0.4343 | 0.6615 | 0.3192 | 0.9620 |
| T.8Eff.Sp.OT1.d15.VSVOva | T | 0.4840 | 0.5142 | 0.4097 | 0.5797 | 0.3247 | 0.2970 | 0.5156 | 0.8763 | 0.3753 | 0.1233 | 0.5315 | 0.1441 | 0.4938 | 0.7328 | 0.8476 | 0.6683 | 0.1510 |
| T.8Eff.Sp.OT1.d5.VSVOva | T | 0.6052 | 0.2176 | 0.2635 | 0.7343 | 0.1126 | 0.2886 | 0.9592 | 0.7739 | 0.0316 | 0.0676 | 0.3030 | 0.4768 | 0.4090 | 0.5988 | 0.7217 | 0.9233 | 0.0830 |
| T.8Eff.Sp.OT1.d6.LisOva | T | 0.7306 | 0.2075 | 0.1556 | 0.9569 | 0.1378 | 0.1201 | 0.5090 | 0.0580 | 0.2853 | 0.3419 | 0.2099 | 0.3333 | 0.6772 | 0.8431 | 0.9294 | 0.8694 | 0.1664 |
| T.8Eff.Sp.OT1.d6.VSVOva | T | 0.3861 | 0.2194 | 0.2977 | 0.5836 | 0.3165 | 0.2828 | 0.5608 | 0.1895 | 0.0711 | 0.0865 | 0.3316 | 0.1279 | 0.4498 | 0.6261 | 0.6669 | 0.2957 | 0.2793 |
| T.8Eff.Sp.OT1.d8.LisOva | T | 0.8953 | 0.6510 | 0.5066 | 0.7632 | 0.2380 | 0.2811 | 0.5909 | 0.0383 | 0.4189 | 0.1664 | 0.7260 | 0.5818 | 0.3834 | 0.9877 | 0.9937 | 0.4090 | 0.7088 |
| T.8Eff.Sp.OT1.d8.VSVOva | T | 0.5750 | 0.7047 | 0.7219 | 0.6966 | 0.1554 | 0.2076 | 0.5504 | 0.3010 | 0.1400 | 0.0827 | 0.4254 | 0.2029 | 0.4802 | 0.9225 | 0.9616 | 0.2300 | 0.8848 |
| T.8Eff.Tbet-.Sp.OT1.d6LisOVA | T | 0.1893 | 0.7675 | 0.2094 | 0.6336 | 0.6393 | 0.3367 | 0.9885 | 0.9456 | 0.0886 | 0.7124 | 0.3737 | 0.3712 | 0.3916 | 0.5377 | 0.4954 | 0.2612 | 0.4702 |
| T.8Eff.Tbet+.Sp.OT1.d6LisOVA | T | 0.3238 | 0.5696 | 0.0459 | 0.4683 | 0.6149 | 0.3181 | 0.9767 | 0.9336 | 0.3157 | 0.4451 | 0.1177 | 0.2445 | 0.5294 | 0.2950 | 0.3312 | 0.3708 | 0.5533 |
| T.8EffKLRG1+CD127-.Sp.d8.LisOVA | T | 0.4867 | 0.6250 | 0.6768 | 0.4056 | 0.5361 | 0.4281 | 0.4546 | 0.2128 | 0.1982 | 0.4038 | 0.7932 | 0.6739 | 0.6681 | 0.0129 | 0.0482 | 0.2525 | 0.3781 |
| T.8Mem.LN | T | 0.1471 | 0.5961 | 0.5245 | 0.6910 | 0.8617 | 0.7296 | 0.6299 | 0.6377 | 0.3443 | 0.0698 | 0.2454 | 0.0971 | 0.9995 | 0.8074 | 0.9061 | 0.9411 | 0.5206 |
| T.8Mem.Sp | T | 0.5725 | 0.2306 | 0.8457 | 0.2427 | 0.3456 | 0.3950 | 0.5606 | 0.4825 | 0.5965 | 0.4277 | 0.2037 | 0.4010 | 0.9205 | 0.8595 | 0.9230 | 0.9040 | 0.5855 |
| T.8Mem.Sp.OT1.d100.LisOva | T | 0.8128 | 0.4751 | 0.8729 | 0.8309 | 0.2881 | 0.2055 | 0.4173 | 0.7155 | 0.5316 | 0.1747 | 0.5483 | 0.6166 | 0.5400 | 0.5403 | 0.7469 | 0.2421 | 0.6412 |
| T.8Mem.Sp.OT1.d106.VSVOva | T | 0.5378 | 0.3459 | 0.8189 | 0.7978 | 0.8179 | 0.3978 | 0.3280 | 0.3252 | 0.3776 | 0.0612 | 0.0822 | 0.2481 | 0.8108 | 0.6126 | 0.7012 | 0.2306 | 0.4393 |
| T.8Mem.Sp.OT1.d45.LisOva | T | 0.7323 | 0.2123 | 0.6258 | 0.7671 | 0.7319 | 0.5086 | 0.5139 | 0.2612 | 0.5639 | 0.3593 | 0.5499 | 0.7115 | 0.9197 | 0.8756 | 0.9750 | 0.5517 | 0.2707 |
| T.8Mem.Sp.OT1.d45.VSVOva | T | 0.6950 | 0.3216 | 0.5568 | 0.5663 | 0.1029 | 0.1293 | 0.3077 | 0.3620 | 0.0803 | 0.0688 | 0.3453 | 0.4956 | 0.9386 | 0.2763 | 0.4976 | 0.2139 | 0.8710 |
| T.8MemKLRG1-CD127+.Sp.d8.LisOVA | T | 0.5001 | 0.5905 | 0.6310 | 0.7027 | 0.7949 | 0.7289 | 0.1699 | 0.3135 | 0.2203 | 0.0482 | 0.9754 | 0.1347 | 0.6366 | 0.5182 | 0.7473 | 0.5025 | 0.4982 |
| T.8Nve.LN | T | 0.8755 | 0.5166 | 0.8637 | 0.9235 | 0.7458 | 0.6460 | 0.6547 | 0.7655 | 0.8733 | 0.8130 | 0.4399 | 0.6432 | 0.8183 | 0.7203 | 0.7339 | 0.5988 | 0.9733 |

| | | | | | | | | | | | | | | | | | | |
|---|---|---|---|---|---|---|---|---|---|---|---|---|---|---|---|---|---|---|
| T.8Nve.MLN | T | 0.6422 | 0.2381 | 0.4454 | 0.6942 | 0.3715 | 0.5064 | 0.3154 | 0.5931 | 0.6480 | 0.4018 | 0.3232 | 0.4989 | 0.7876 | 0.7434 | 0.8540 | 0.6838 | 0.9741 |
| T.8Nve.PP | T | 0.4970 | 0.4994 | 0.7614 | 0.6459 | 0.5246 | 0.5324 | 0.2839 | 0.7991 | 0.4580 | 0.1625 | 0.9209 | 0.9467 | 0.8392 | 0.2392 | 0.2872 | 0.5378 | 0.2239 |
| T.8Nve.Sp | T | 0.8105 | 0.1165 | 0.6710 | 0.7760 | 0.9013 | 0.7940 | 0.3411 | 0.1745 | 0.5687 | 0.6966 | 0.6430 | 0.3615 | 0.8328 | 0.5145 | 0.5861 | 0.9797 | 0.6091 |
| T.8Nve.Sp.OT1 | T | 0.8720 | 0.7014 | 0.7314 | 0.7917 | 0.8929 | 0.8069 | 0.0165 | 0.4790 | 0.7133 | 0.5447 | 0.6286 | 0.4987 | 0.8495 | 0.9187 | 0.9641 | 0.3201 | 0.3197 |
| T.8SP24-.Th | T | 0.8962 | 0.3744 | 0.7387 | 0.4759 | 0.5569 | 0.3763 | 0.6049 | 0.5660 | 0.8279 | 0.8561 | 0.6859 | 0.7393 | 0.5073 | 0.9047 | 0.8940 | 0.7950 | 0.9468 |
| T.8SP24int.Th | T | 0.9037 | 0.3430 | 0.6068 | 0.6330 | 0.6131 | 0.5335 | 0.4166 | 0.7734 | 0.4253 | 0.5305 | 0.6159 | 0.7691 | 0.5645 | 0.9543 | 0.9826 | 0.5312 | 0.5548 |
| T.8SP69+.Th | T | 0.9921 | 0.2990 | 0.7652 | 0.6098 | 0.4257 | 0.3099 | 0.8098 | 0.9579 | 0.4011 | 0.6504 | 0.3696 | 0.5967 | 0.8992 | 0.8807 | 0.8805 | 0.8984 | 0.8199 |
| T.DN4.Th | T | 0.7087 | 0.4391 | 0.3357 | 0.8271 | 0.6721 | 0.7891 | 0.3002 | 0.5656 | 0.2267 | 0.1210 | 0.1755 | 0.4825 | 0.1965 | 0.4569 | 0.5826 | 0.6199 | 0.3731 |
| T.DP.69-.e17.Th.v2 | T | 0.2820 | 0.4748 | 0.0308 | 0.5229 | 0.0795 | 0.2057 | 0.9826 | 0.9628 | 0.2027 | 0.2109 | 0.7942 | 0.8284 | 0.1431 | 0.5586 | 0.6652 | 0.8794 | 0.2335 |
| T.DP.Th | T | 0.9958 | 0.5701 | 0.9284 | 0.8181 | 0.7088 | 0.8187 | 0.1078 | 0.0605 | 0.0630 | 0.1396 | 0.6702 | 0.5347 | 0.2131 | 0.3288 | 0.2941 | 0.4744 | 0.8241 |
| T.DP.Th.v2 | T | 0.6944 | 0.1652 | 0.0166 | 0.4654 | 0.3132 | 0.2737 | 0.8420 | 0.4941 | 0.1394 | 0.1899 | 0.2569 | 0.5790 | 0.0224 | 0.5269 | 0.6598 | 0.3953 | 0.3171 |
| T.DP69+.Th | T | 0.9933 | 0.2494 | 0.8090 | 0.3066 | 0.7630 | 0.6321 | 0.4561 | 0.3885 | 0.3547 | 0.4322 | 0.3785 | 0.2104 | 0.2014 | 0.9221 | 0.9047 | 0.4303 | 0.9122 |
| T.DP69+.Th.v2 | T | 0.9647 | 0.5238 | 0.6304 | 0.7976 | 0.4781 | 0.5995 | 0.9877 | 0.8471 | 0.7708 | 0.1192 | 0.6743 | 0.4075 | 0.1529 | 0.9730 | 0.9597 | 0.3789 | 0.9494 |
| T.DPbl.Th | T | 0.8901 | 0.5474 | 0.7622 | 0.3784 | 0.6465 | 0.8520 | 0.2584 | 0.6401 | 0.2742 | 0.5517 | 0.0723 | 0.4871 | 0.0957 | 0.6247 | 0.6120 | 0.4210 | 0.2042 |
| T.DPsm.Th | T | 0.9618 | 0.8123 | 0.9711 | 0.6900 | 0.5949 | 0.5056 | 0.3727 | 0.6112 | 0.4725 | 0.3003 | 0.8956 | 0.9012 | 0.2277 | 0.8230 | 0.8156 | 0.0700 | 0.8743 |
| T.ISP.Th | T | 0.5848 | 0.5834 | 0.7950 | 0.3692 | 0.3419 | 0.5997 | 0.2308 | 0.4172 | 0.0309 | 0.1846 | 0.3091 | 0.5709 | 0.3589 | 0.3391 | 0.4104 | 0.8200 | 0.0679 |
| Tgd.Sp | T | 0.1277 | 0.0932 | 0.2142 | 0.2806 | 0.6778 | 0.9070 | 0.1711 | 0.1033 | 0.5750 | 0.0560 | 0.4013 | 0.1770 | 0.3813 | 0.4956 | 0.4704 | 0.7824 | 0.4791 |
| Tgd.Th | T | 0.8635 | 0.0961 | 0.7040 | 0.3597 | 0.8945 | 0.8424 | 0.0884 | 0.1337 | 0.2729 | 0.6917 | 0.2703 | 0.6304 | 0.3657 | 0.9260 | 0.8984 | 0.9265 | 0.4240 |
| Tgd.vg1+vd6+24ahi.Th | T | 0.8838 | 0.5343 | 0.6793 | 0.2887 | 0.9899 | 0.9448 | 0.0339 | 0.2893 | 0.0952 | 0.1521 | 0.6520 | 0.9015 | 0.1945 | 0.9989 | 0.9992 | 0.9820 | 0.6711 |
| Tgd.vg1+vd6+24alo.Th | T | 0.4418 | 0.6313 | 0.2288 | 0.4150 | 0.9292 | 0.7120 | 0.5940 | 0.9444 | 0.8944 | 0.8212 | 0.7481 | 0.5111 | 0.9380 | 0.9436 | 0.9193 | 0.2123 | 0.7164 |
| Tgd.vg1+vd6-24ahi.Th | T | 0.7322 | 0.1968 | 0.2420 | 0.0951 | 0.9679 | 0.7089 | 0.1260 | 0.5107 | 0.2427 | 0.3440 | 0.7878 | 0.8852 | 0.3904 | 0.9974 | 0.9906 | 0.6978 | 0.8269 |
| Tgd.vg1+vd6-24alo.Th | T | 0.5353 | 0.6932 | 0.4974 | 0.4520 | 0.9093 | 0.7439 | 0.0497 | 0.1227 | 0.2865 | 0.2128 | 0.6502 | 0.6889 | 0.4524 | 0.7627 | 0.7606 | 0.4289 | 0.9973 |
| Tgd.vg2-.act.Sp | T | 0.9705 | 0.1804 | 0.2780 | 0.8120 | 0.7961 | 0.4338 | 0.3866 | 0.0318 | 0.5953 | 0.3096 | 0.4178 | 0.0759 | 0.8611 | 0.9704 | 0.9932 | 0.7849 | 0.6022 |
| Tgd.vg2-.Sp | T | 0.9606 | 0.5908 | 0.9603 | 0.6673 | 0.5766 | 0.6773 | 0.5863 | 0.3408 | 0.2612 | 0.0843 | 0.2678 | 0.0752 | 0.1078 | 0.8868 | 0.9413 | 0.9153 | 0.9203 |
| Tgd.vg2-.Sp.TCRbko | T | 0.6610 | 0.5057 | 0.7603 | 0.3126 | 0.6910 | 0.6619 | 0.8149 | 0.5067 | 0.4249 | 0.1288 | 0.2285 | 0.2429 | 0.5800 | 0.9903 | 0.9951 | 0.6748 | 0.4384 |
| Tgd.vg2+.act.Sp | T | 0.6895 | 0.5741 | 0.5616 | 0.4974 | 0.1099 | 0.1059 | 0.3575 | 0.1034 | 0.4254 | 0.2025 | 0.1148 | 0.0276 | 0.4110 | 0.9404 | 0.9485 | 0.6215 | 0.8401 |
| Tgd.vg2+.Sp | T | 0.9574 | 0.2582 | 0.7090 | 0.6982 | 0.3900 | 0.2831 | 0.0709 | 0.1971 | 0.8583 | 0.7319 | 0.8377 | 0.6194 | 0.7079 | 0.9985 | 0.9998 | 0.9389 | 0.3933 |
| Tgd.vg2+.Sp.TCRbko | T | 0.6206 | 0.7409 | 0.9963 | 0.4752 | 0.8698 | 0.8569 | 0.4874 | 0.1624 | 0.2990 | 0.1409 | 0.2870 | 0.0425 | 0.4412 | 0.9884 | 0.9957 | 0.9567 | 0.3516 |
| Tgd.vg2+24ahi.e17.Th | T | 0.7085 | 0.0897 | 0.7590 | 0.6446 | 0.8965 | 0.9192 | 0.0109 | 0.1909 | 0.0688 | 0.5220 | 0.5126 | 0.7468 | 0.1467 | 0.7982 | 0.7898 | 0.6943 | 0.5888 |
| Tgd.vg2+24ahi.e17.Th.v2 | T | 0.2380 | 0.0888 | 0.0065 | 0.8665 | 0.3658 | 0.6006 | 0.5266 | 0.5829 | 0.1345 | 0.2245 | 0.5140 | 0.4840 | 0.5043 | 0.6956 | 0.6708 | 0.8568 | 0.4701 |
| Tgd.vg2+24ahi.Th | T | 0.9078 | 0.0681 | 0.9249 | 0.3850 | 0.9131 | 0.9534 | 0.2448 | 0.1451 | 0.2838 | 0.2899 | 0.6025 | 0.5227 | 0.4939 | 0.9982 | 0.9992 | 0.9045 | 0.1970 |
| Tgd.vg2+24alo.Th | T | 0.2252 | 0.6148 | 0.8869 | 0.2997 | 0.8739 | 0.9765 | 0.0264 | 0.1272 | 0.1292 | 0.1436 | 0.0442 | 0.0493 | 0.5921 | 0.7285 | 0.7495 | 0.3474 | 0.3818 |
| Tgd.vg3+24ahi.e17.Th | T | 0.8507 | 0.0208 | 0.2909 | 0.5935 | 0.9058 | 0.8928 | 0.1098 | 0.4867 | 0.2565 | 0.5941 | 0.2842 | 0.6880 | 0.6914 | 0.8025 | 0.8315 | 0.9973 | 0.3906 |
| Tgd.vg3+24alo.e17.Th | T | 0.4419 | 0.5384 | 0.3882 | 0.7883 | 0.7159 | 0.7480 | 0.2337 | 0.3933 | 0.2141 | 0.6323 | 0.8003 | 0.7634 | 0.2709 | 0.9366 | 0.9689 | 0.9305 | 0.1474 |
| Tgd.vg4+24ahi.e17.Th | T | 0.3813 | 0.0423 | 0.0085 | 0.9582 | 0.3776 | 0.5609 | 0.7239 | 0.5744 | 0.0935 | 0.2986 | 0.5158 | 0.5225 | 0.4510 | 0.5731 | 0.3517 | 0.7401 | 0.3264 |
| Tgd.vg4+24alo.e17.Th | T | 0.1656 | 0.1622 | 0.1428 | 0.8675 | 0.1261 | 0.2503 | 0.6393 | 0.4685 | 0.2277 | 0.3224 | 0.4234 | 0.2327 | 0.5706 | 0.2814 | 0.2192 | 0.5844 | 0.2086 |
| Tgd.vg5-.act.IEL | T | 0.8157 | 0.0248 | 0.2267 | 0.7985 | 0.0278 | 0.2782 | 0.0250 | 0.1930 | 0.1057 | 0.0933 | 0.4970 | 0.4677 | 0.1380 | 0.9980 | 0.9989 | 0.9721 | 0.4681 |
| Tgd.vg5-.IEL | T | 0.9688 | 0.4294 | 0.4403 | 0.9787 | 0.3228 | 0.7443 | 0.1898 | 0.0200 | 0.3631 | 0.2381 | 0.6216 | 0.2712 | 0.3987 | 0.9299 | 0.9696 | 0.8962 | 0.7952 |
| Tgd.vg5+.act.IEL | T | 0.8602 | 0.1235 | 0.1770 | 0.7160 | 0.1787 | 0.4701 | 0.1484 | 0.3460 | 0.3475 | 0.1383 | 0.6614 | 0.7081 | 0.2966 | 0.9921 | 0.9971 | 0.6613 | 0.9483 |
| Tgd.vg5+.IEL | T | 0.9761 | 0.4680 | 0.4055 | 0.9986 | 0.2032 | 0.7216 | 0.7132 | 0.7036 | 0.2173 | 0.3323 | 0.7222 | 0.7290 | 0.3371 | 0.9954 | 0.9971 | 0.9669 | 0.9449 |
| Tgd.vg5+24ahi.Th | T | 0.9088 | 0.6482 | 0.5344 | 0.1690 | 0.9458 | 0.8598 | 0.0327 | 0.2439 | 0.3598 | 0.7981 | 0.5561 | 0.7801 | 0.8783 | 0.8187 | 0.8521 | 0.9094 | 0.9432 |

**Table S14: Cell/Tissue-specific heritability enrichment coefficients and p-values (ImmGen)**

This table contains the same pvalues as Table S12, but with coefficients and in "long" format

| Phenotype | Tissue/Cell Type | Category | Coefficient | Coefficient_std_error | Coefficient_P_value |
|---|---|---|---|---|---|
| AD Braak | B.Fo.LN | B | 2.21E-08 | 2.07E-08 | 0.142707087 |
| ADNC Score | B.Fo.LN | B | -4.84E-08 | 5.36E-08 | 0.816620878 |
| AP (any/none) | B.Fo.LN | B | -5.07E-09 | 1.98E-08 | 0.601110218 |
| Arteriolosclerosis | B.Fo.LN | B | 1.10E-08 | 2.65E-08 | 0.338878203 |
| Arteriolosclerosis (any | B.Fo.LN | B | 2.78E-08 | 2.76E-08 | 0.156880327 |
| Atherosclerosis | B.Fo.LN | B | 4.49E-08 | 2.56E-08 | 0.039377166 |
| Atherosclerosis (any/r | B.Fo.LN | B | 5.83E-10 | 2.56E-08 | 0.490910488 |
| B Score | B.Fo.LN | B | 2.60E-08 | 1.97E-08 | 0.093929708 |
| CAA | B.Fo.LN | B | -1.36E-08 | 2.30E-08 | 0.723003428 |
| CAA (any/none) | B.Fo.LN | B | -1.65E-08 | 2.36E-08 | 0.75815797 |
| CERAD (C Score) | B.Fo.LN | B | 3.82E-08 | 1.95E-08 | 0.024884745 |
| Infarcts/lacunes | B.Fo.LN | B | -4.50E-09 | 2.18E-08 | 0.581808486 |
| PD Braak | B.Fo.LN | B | 1.73E-08 | 1.86E-08 | 0.175302523 |
| PD Braak (any/none) | B.Fo.LN | B | 4.46E-09 | 1.73E-08 | 0.39807582 |
| TDP-43 (any/none) | B.Fo.LN | B | -3.65E-09 | 5.86E-08 | 0.524871675 |
| Thal Phase | B.Fo.LN | B | -5.22E-08 | 6.73E-08 | 0.781105509 |
| WMR (any/none) | B.Fo.LN | B | -6.84E-09 | 3.22E-08 | 0.584097073 |
| #N/A | B.Fo.LN | B | 6.49E-08 | 4.27E-08 | 0.06456086 |
| AD Braak | B.Fo.MLN | B | 5.07E-08 | 1.92E-08 | 0.004127256 |
| ADNC Score | B.Fo.MLN | B | -1.04E-09 | 5.75E-08 | 0.507186272 |
| AP (any/none) | B.Fo.MLN | B | 2.83E-09 | 2.11E-08 | 0.446798244 |
| Arteriolosclerosis | B.Fo.MLN | B | -2.53E-10 | 2.59E-08 | 0.503888653 |
| Arteriolosclerosis (any | B.Fo.MLN | B | 3.10E-08 | 2.96E-08 | 0.147105954 |
| Atherosclerosis | B.Fo.MLN | B | 5.05E-08 | 2.56E-08 | 0.024096424 |
| Atherosclerosis (any/r | B.Fo.MLN | B | 1.26E-08 | 2.53E-08 | 0.308772173 |
| B Score | B.Fo.MLN | B | 4.14E-08 | 1.90E-08 | 0.014494482 |
| CAA | B.Fo.MLN | B | -3.03E-08 | 2.45E-08 | 0.892235655 |
| CAA (any/none) | B.Fo.MLN | B | -2.84E-08 | 2.48E-08 | 0.873431213 |
| CERAD (C Score) | B.Fo.MLN | B | 2.74E-08 | 2.04E-08 | 0.089487537 |
| Infarcts/lacunes | B.Fo.MLN | B | -2.62E-08 | 2.29E-08 | 0.873756458 |

| | | | | | |
|---|---|---|---|---|---|
| PD Braak | B.Fo.MLN | B | 1.46E-08 | 1.98E-08 | 0.230003375 |
| PD Braak (any/none) | B.Fo.MLN | B | 4.99E-09 | 1.88E-08 | 0.39545623 |
| TDP-43 (any/none) | B.Fo.MLN | B | -2.09E-08 | 5.83E-08 | 0.639952811 |
| Thal Phase | B.Fo.MLN | B | 1.62E-08 | 7.22E-08 | 0.410994453 |
| WMR (any/none) | B.Fo.MLN | B | 7.71E-09 | 3.48E-08 | 0.412285452 |
| #N/A | B.Fo.MLN | B | 5.20E-08 | 4.01E-08 | 0.097257756 |
| AD Braak | B.Fo.PC | B | 4.79E-08 | 2.04E-08 | 0.009557157 |
| ADNC Score | B.Fo.PC | B | -2.34E-08 | 5.83E-08 | 0.656188926 |
| AP (any/none) | B.Fo.PC | B | 1.55E-08 | 2.09E-08 | 0.229626328 |
| Arteriolosclerosis | B.Fo.PC | B | 2.58E-08 | 2.67E-08 | 0.166591291 |
| Arteriolosclerosis (any | B.Fo.PC | B | 3.12E-08 | 3.07E-08 | 0.155098287 |
| Atherosclerosis | B.Fo.PC | B | 3.87E-08 | 2.42E-08 | 0.054989169 |
| Atherosclerosis (any/r | B.Fo.PC | B | 1.60E-08 | 2.42E-08 | 0.253665965 |
| B Score | B.Fo.PC | B | 4.42E-08 | 1.89E-08 | 0.00966447 |
| CAA | B.Fo.PC | B | -1.22E-08 | 2.33E-08 | 0.700217327 |
| CAA (any/none) | B.Fo.PC | B | -3.32E-09 | 2.52E-08 | 0.552378169 |
| CERAD (C Score) | B.Fo.PC | B | 3.07E-08 | 2.17E-08 | 0.078479471 |
| Infarcts/lacunes | B.Fo.PC | B | -2.82E-08 | 2.06E-08 | 0.915114227 |
| PD Braak | B.Fo.PC | B | 4.67E-09 | 2.06E-08 | 0.410186078 |
| PD Braak (any/none) | B.Fo.PC | B | 1.28E-09 | 1.93E-08 | 0.473625873 |
| TDP-43 (any/none) | B.Fo.PC | B | -2.04E-08 | 5.38E-08 | 0.647859073 |
| Thal Phase | B.Fo.PC | B | 2.82E-09 | 7.24E-08 | 0.484450064 |
| WMR (any/none) | B.Fo.PC | B | -1.96E-08 | 3.59E-08 | 0.707322717 |
| #N/A | B.Fo.PC | B | 6.90E-08 | 3.93E-08 | 0.039721344 |
| AD Braak | B.Fo.Sp | B | 2.30E-08 | 1.84E-08 | 0.105610997 |
| ADNC Score | B.Fo.Sp | B | -6.39E-10 | 5.31E-08 | 0.504797714 |
| AP (any/none) | B.Fo.Sp | B | 2.22E-09 | 2.00E-08 | 0.455713956 |
| Arteriolosclerosis | B.Fo.Sp | B | 3.51E-09 | 2.55E-08 | 0.445353034 |
| Arteriolosclerosis (any | B.Fo.Sp | B | 2.37E-08 | 2.83E-08 | 0.201855997 |
| Atherosclerosis | B.Fo.Sp | B | 3.73E-08 | 2.53E-08 | 0.07018478 |
| Atherosclerosis (any/r | B.Fo.Sp | B | 4.43E-09 | 2.29E-08 | 0.423430913 |
| B Score | B.Fo.Sp | B | 1.74E-08 | 1.81E-08 | 0.168576411 |
| CAA | B.Fo.Sp | B | -3.89E-08 | 2.39E-08 | 0.948045799 |
| CAA (any/none) | B.Fo.Sp | B | -3.76E-08 | 2.36E-08 | 0.944366478 |

| | | | | | |
|---|---|---|---|---|---|
| CERAD (C Score) | B.Fo.Sp | B | 7.52E-09 | 1.93E-08 | 0.348179473 |
| Infarcts/lacunes | B.Fo.Sp | B | -2.08E-08 | 2.34E-08 | 0.812819434 |
| PD Braak | B.Fo.Sp | B | -8.74E-09 | 2.02E-08 | 0.667622256 |
| PD Braak (any/none) | B.Fo.Sp | B | -1.47E-08 | 1.95E-08 | 0.775118251 |
| TDP-43 (any/none) | B.Fo.Sp | B | -2.94E-08 | 5.41E-08 | 0.706609966 |
| Thal Phase | B.Fo.Sp | B | 3.74E-08 | 6.81E-08 | 0.291215011 |
| WMR (any/none) | B.Fo.Sp | B | -1.21E-08 | 3.32E-08 | 0.641706585 |
| #N/A | B.Fo.Sp | B | 1.44E-08 | 3.74E-08 | 0.350186284 |
| AD Braak | B.MZ.Sp | B | 5.01E-08 | 1.99E-08 | 0.005925845 |
| ADNC Score | B.MZ.Sp | B | -6.18E-08 | 6.40E-08 | 0.833091289 |
| AP (any/none) | B.MZ.Sp | B | -9.81E-09 | 2.21E-08 | 0.671598139 |
| Arteriolosclerosis | B.MZ.Sp | B | -4.45E-11 | 2.64E-08 | 0.500672018 |
| Arteriolosclerosis (any | B.MZ.Sp | B | 3.95E-08 | 3.04E-08 | 0.097020837 |
| Atherosclerosis | B.MZ.Sp | B | 2.62E-08 | 2.77E-08 | 0.171717421 |
| Atherosclerosis (any/r | B.MZ.Sp | B | 3.64E-09 | 2.60E-08 | 0.444321143 |
| B Score | B.MZ.Sp | B | 4.09E-08 | 1.96E-08 | 0.018568846 |
| CAA | B.MZ.Sp | B | -2.94E-08 | 2.50E-08 | 0.879847143 |
| CAA (any/none) | B.MZ.Sp | B | -2.83E-08 | 2.39E-08 | 0.882001961 |
| CERAD (C Score) | B.MZ.Sp | B | 2.28E-08 | 2.16E-08 | 0.145048459 |
| Infarcts/lacunes | B.MZ.Sp | B | -2.19E-08 | 2.30E-08 | 0.828962111 |
| PD Braak | B.MZ.Sp | B | -5.06E-09 | 2.21E-08 | 0.590446585 |
| PD Braak (any/none) | B.MZ.Sp | B | -1.82E-08 | 2.13E-08 | 0.8029661 |
| TDP-43 (any/none) | B.MZ.Sp | B | -2.05E-08 | 5.42E-08 | 0.647444664 |
| Thal Phase | B.MZ.Sp | B | 3.05E-08 | 7.94E-08 | 0.350401045 |
| WMR (any/none) | B.MZ.Sp | B | -5.27E-08 | 3.50E-08 | 0.933754666 |
| #N/A | B.MZ.Sp | B | 1.26E-08 | 4.39E-08 | 0.387384475 |
| AD Braak | B.T1.Sp | B | 5.07E-08 | 1.91E-08 | 0.003921763 |
| ADNC Score | B.T1.Sp | B | 1.13E-08 | 5.47E-08 | 0.418432537 |
| AP (any/none) | B.T1.Sp | B | -3.63E-09 | 1.89E-08 | 0.5759981 |
| Arteriolosclerosis | B.T1.Sp | B | -7.99E-09 | 2.59E-08 | 0.621140602 |
| Arteriolosclerosis (any | B.T1.Sp | B | 4.30E-08 | 2.84E-08 | 0.064941332 |
| Atherosclerosis | B.T1.Sp | B | 5.82E-08 | 2.32E-08 | 0.006034467 |
| Atherosclerosis (any/r | B.T1.Sp | B | 4.67E-08 | 2.26E-08 | 0.019614457 |
| B Score | B.T1.Sp | B | 4.59E-08 | 1.84E-08 | 0.00643667 |

| | | | | | |
|---|---|---|---|---|---|
| CAA | B.T1.Sp | B | -3.69E-08 | 2.35E-08 | 0.941544237 |
| CAA (any/none) | B.T1.Sp | B | -2.90E-08 | 2.30E-08 | 0.896043684 |
| CERAD (C Score) | B.T1.Sp | B | 2.95E-08 | 2.05E-08 | 0.074980484 |
| Infarcts/lacunes | B.T1.Sp | B | -3.05E-08 | 2.27E-08 | 0.910310518 |
| PD Braak | B.T1.Sp | B | -1.52E-09 | 2.02E-08 | 0.530025774 |
| PD Braak (any/none) | B.T1.Sp | B | -2.25E-09 | 1.94E-08 | 0.546053327 |
| TDP-43 (any/none) | B.T1.Sp | B | -7.38E-08 | 5.10E-08 | 0.925960075 |
| Thal Phase | B.T1.Sp | B | 4.43E-08 | 7.33E-08 | 0.272906501 |
| WMR (any/none) | B.T1.Sp | B | 2.51E-08 | 3.36E-08 | 0.22734944 |
| #N/A | B.T1.Sp | B | 7.61E-08 | 3.86E-08 | 0.024133239 |
| AD Braak | B.T2.Sp | B | 3.79E-08 | 1.92E-08 | 0.024422591 |
| ADNC Score | B.T2.Sp | B | -2.44E-08 | 5.80E-08 | 0.663027383 |
| AP (any/none) | B.T2.Sp | B | -1.93E-09 | 2.01E-08 | 0.538302318 |
| Arteriolosclerosis | B.T2.Sp | B | 7.33E-09 | 2.65E-08 | 0.391288249 |
| Arteriolosclerosis (any | B.T2.Sp | B | 4.93E-08 | 2.96E-08 | 0.047824995 |
| Atherosclerosis | B.T2.Sp | B | 4.09E-08 | 2.44E-08 | 0.04646843 |
| Atherosclerosis (any/r | B.T2.Sp | B | 2.64E-08 | 2.22E-08 | 0.116567881 |
| B Score | B.T2.Sp | B | 3.04E-08 | 1.92E-08 | 0.056177544 |
| CAA | B.T2.Sp | B | -2.38E-08 | 2.43E-08 | 0.836670885 |
| CAA (any/none) | B.T2.Sp | B | -2.82E-08 | 2.42E-08 | 0.878768749 |
| CERAD (C Score) | B.T2.Sp | B | 1.22E-08 | 2.03E-08 | 0.273097611 |
| Infarcts/lacunes | B.T2.Sp | B | -2.50E-08 | 2.43E-08 | 0.847988396 |
| PD Braak | B.T2.Sp | B | -1.45E-08 | 1.99E-08 | 0.765790225 |
| PD Braak (any/none) | B.T2.Sp | B | -2.11E-08 | 1.84E-08 | 0.874591015 |
| TDP-43 (any/none) | B.T2.Sp | B | -2.43E-08 | 5.32E-08 | 0.676367802 |
| Thal Phase | B.T2.Sp | B | 3.72E-08 | 7.70E-08 | 0.314690385 |
| WMR (any/none) | B.T2.Sp | B | -4.86E-09 | 3.44E-08 | 0.55619373 |
| #N/A | B.T2.Sp | B | 2.44E-08 | 4.02E-08 | 0.272353316 |
| AD Braak | B.T3.Sp | B | 3.40E-08 | 1.87E-08 | 0.034504636 |
| ADNC Score | B.T3.Sp | B | -6.11E-08 | 5.31E-08 | 0.874983036 |
| AP (any/none) | B.T3.Sp | B | -1.06E-08 | 2.03E-08 | 0.699780557 |
| Arteriolosclerosis | B.T3.Sp | B | 1.42E-09 | 2.63E-08 | 0.478448694 |
| Arteriolosclerosis (any | B.T3.Sp | B | 4.20E-08 | 2.89E-08 | 0.073478107 |
| Atherosclerosis | B.T3.Sp | B | 5.80E-08 | 2.64E-08 | 0.014074651 |

| | | | | | |
|---|---|---|---|---|---|
| Atherosclerosis (any/r | B.T3.Sp | B | 2.88E-08 | 2.44E-08 | 0.119088627 |
| B Score | B.T3.Sp | B | 2.44E-08 | 1.83E-08 | 0.091740333 |
| CAA | B.T3.Sp | B | -3.40E-08 | 2.54E-08 | 0.90993255 |
| CAA (any/none) | B.T3.Sp | B | -3.89E-08 | 2.41E-08 | 0.946716017 |
| CERAD (C Score) | B.T3.Sp | B | -4.56E-09 | 1.95E-08 | 0.592308689 |
| Infarcts/lacunes | B.T3.Sp | B | -3.32E-08 | 2.30E-08 | 0.926000114 |
| PD Braak | B.T3.Sp | B | -6.68E-09 | 1.92E-08 | 0.635893711 |
| PD Braak (any/none) | B.T3.Sp | B | -1.31E-08 | 1.83E-08 | 0.762402552 |
| TDP-43 (any/none) | B.T3.Sp | B | -2.01E-08 | 5.15E-08 | 0.652269148 |
| Thal Phase | B.T3.Sp | B | -2.11E-08 | 7.24E-08 | 0.614658473 |
| WMR (any/none) | B.T3.Sp | B | -2.43E-08 | 3.13E-08 | 0.780845303 |
| #N/A | B.T3.Sp | B | 3.80E-08 | 3.98E-08 | 0.169813009 |
| AD Braak | B1a.PC | B | 4.26E-08 | 1.88E-08 | 0.011754977 |
| ADNC Score | B1a.PC | B | 1.06E-08 | 6.10E-08 | 0.430781535 |
| AP (any/none) | B1a.PC | B | -2.14E-09 | 2.18E-08 | 0.539148192 |
| Arteriolosclerosis | B1a.PC | B | -5.01E-09 | 2.57E-08 | 0.577351534 |
| Arteriolosclerosis (any | B1a.PC | B | 2.70E-09 | 2.92E-08 | 0.463140126 |
| Atherosclerosis | B1a.PC | B | 2.85E-08 | 2.26E-08 | 0.104108951 |
| Atherosclerosis (any/r | B1a.PC | B | 1.67E-08 | 2.59E-08 | 0.260316871 |
| B Score | B1a.PC | B | 4.49E-08 | 1.91E-08 | 0.009304094 |
| CAA | B1a.PC | B | -2.27E-08 | 2.27E-08 | 0.840886366 |
| CAA (any/none) | B1a.PC | B | -1.18E-08 | 2.28E-08 | 0.697017495 |
| CERAD (C Score) | B1a.PC | B | 2.43E-08 | 2.15E-08 | 0.129026066 |
| Infarcts/lacunes | B1a.PC | B | -1.31E-08 | 2.25E-08 | 0.719986502 |
| PD Braak | B1a.PC | B | -1.45E-08 | 1.94E-08 | 0.773122553 |
| PD Braak (any/none) | B1a.PC | B | -2.60E-08 | 1.98E-08 | 0.905605343 |
| TDP-43 (any/none) | B1a.PC | B | -1.01E-07 | 5.22E-08 | 0.973811337 |
| Thal Phase | B1a.PC | B | 2.00E-08 | 7.93E-08 | 0.400659816 |
| WMR (any/none) | B1a.PC | B | -6.00E-08 | 3.44E-08 | 0.959391845 |
| #N/A | B1a.PC | B | 2.97E-08 | 4.43E-08 | 0.251513644 |
| AD Braak | B1a.Sp | B | 2.31E-08 | 1.92E-08 | 0.114469947 |
| ADNC Score | B1a.Sp | B | -2.92E-08 | 5.82E-08 | 0.692079883 |
| AP (any/none) | B1a.Sp | B | -2.89E-08 | 2.21E-08 | 0.904594922 |
| Arteriolosclerosis | B1a.Sp | B | -5.72E-10 | 2.71E-08 | 0.508403776 |

| | | | | | |
|---|---|---|---|---|---|
| Arteriolosclerosis (any | B1a.Sp | B | 2.32E-08 | 3.20E-08 | 0.234235483 |
| Atherosclerosis | B1a.Sp | B | 1.95E-08 | 2.36E-08 | 0.204982271 |
| Atherosclerosis (any/r | B1a.Sp | B | 1.02E-08 | 2.52E-08 | 0.342538582 |
| B Score | B1a.Sp | B | 1.80E-08 | 2.02E-08 | 0.18592986 |
| CAA | B1a.Sp | B | -1.67E-08 | 2.46E-08 | 0.752185462 |
| CAA (any/none) | B1a.Sp | B | -1.68E-08 | 2.26E-08 | 0.771691312 |
| CERAD (C Score) | B1a.Sp | B | -1.32E-09 | 2.22E-08 | 0.523763543 |
| Infarcts/lacunes | B1a.Sp | B | -1.31E-08 | 2.24E-08 | 0.720664639 |
| PD Braak | B1a.Sp | B | -1.03E-08 | 2.14E-08 | 0.684527214 |
| PD Braak (any/none) | B1a.Sp | B | -2.88E-08 | 2.12E-08 | 0.913140353 |
| TDP-43 (any/none) | B1a.Sp | B | -4.52E-08 | 5.70E-08 | 0.786239649 |
| Thal Phase | B1a.Sp | B | 2.47E-08 | 7.18E-08 | 0.365531736 |
| WMR (any/none) | B1a.Sp | B | -4.92E-08 | 3.60E-08 | 0.914221857 |
| #N/A | B1a.Sp | B | 4.24E-09 | 4.83E-08 | 0.465008407 |
| AD Braak | B1a.Sp.v2 | B | -3.04E-08 | 1.76E-08 | 0.958147785 |
| ADNC Score | B1a.Sp.v2 | B | -5.48E-08 | 5.65E-08 | 0.8338192 |
| AP (any/none) | B1a.Sp.v2 | B | -2.92E-08 | 1.92E-08 | 0.93582939 |
| Arteriolosclerosis | B1a.Sp.v2 | B | 2.68E-08 | 2.59E-08 | 0.150610678 |
| Arteriolosclerosis (any | B1a.Sp.v2 | B | 1.29E-09 | 2.56E-08 | 0.479899048 |
| Atherosclerosis | B1a.Sp.v2 | B | -3.82E-08 | 1.98E-08 | 0.973291268 |
| Atherosclerosis (any/r | B1a.Sp.v2 | B | -2.71E-08 | 2.12E-08 | 0.899407056 |
| B Score | B1a.Sp.v2 | B | -1.97E-08 | 1.78E-08 | 0.865872829 |
| CAA | B1a.Sp.v2 | B | -4.85E-08 | 2.16E-08 | 0.987826025 |
| CAA (any/none) | B1a.Sp.v2 | B | -2.11E-08 | 2.06E-08 | 0.847382241 |
| CERAD (C Score) | B1a.Sp.v2 | B | -3.04E-10 | 1.87E-08 | 0.50649945 |
| Infarcts/lacunes | B1a.Sp.v2 | B | -1.50E-08 | 2.23E-08 | 0.749022345 |
| PD Braak | B1a.Sp.v2 | B | -1.67E-08 | 2.31E-08 | 0.765055482 |
| PD Braak (any/none) | B1a.Sp.v2 | B | -1.40E-08 | 2.25E-08 | 0.73336354 |
| TDP-43 (any/none) | B1a.Sp.v2 | B | 4.79E-09 | 5.39E-08 | 0.464597464 |
| Thal Phase | B1a.Sp.v2 | B | -5.06E-08 | 6.77E-08 | 0.772537412 |
| WMR (any/none) | B1a.Sp.v2 | B | 3.27E-08 | 3.23E-08 | 0.155942401 |
| #N/A | B1a.Sp.v2 | B | -5.12E-08 | 4.12E-08 | 0.893286686 |
| AD Braak | B1b.PC | B | 3.56E-08 | 2.05E-08 | 0.040994299 |
| ADNC Score | B1b.PC | B | 2.59E-08 | 6.38E-08 | 0.342488687 |

| | | | | | |
|---|---|---|---|---|---|
| AP (any/none) | B1b.PC | B | -2.84E-08 | 2.16E-08 | 0.906271472 |
| Arteriolosclerosis | B1b.PC | B | -4.56E-09 | 2.66E-08 | 0.568081843 |
| Arteriolosclerosis (any | B1b.PC | B | 1.70E-08 | 2.96E-08 | 0.283103467 |
| Atherosclerosis | B1b.PC | B | 3.64E-08 | 2.57E-08 | 0.078449556 |
| Atherosclerosis (any/r | B1b.PC | B | 1.06E-08 | 2.58E-08 | 0.341409766 |
| B Score | B1b.PC | B | 3.11E-08 | 2.00E-08 | 0.060110724 |
| CAA | B1b.PC | B | -3.46E-08 | 2.40E-08 | 0.925509877 |
| CAA (any/none) | B1b.PC | B | -2.81E-08 | 2.15E-08 | 0.904913245 |
| CERAD (C Score) | B1b.PC | B | 6.10E-09 | 2.08E-08 | 0.384843071 |
| Infarcts/lacunes | B1b.PC | B | 2.21E-09 | 2.41E-08 | 0.463538649 |
| PD Braak | B1b.PC | B | 1.49E-09 | 2.13E-08 | 0.472238175 |
| PD Braak (any/none) | B1b.PC | B | -9.25E-09 | 2.05E-08 | 0.674197086 |
| TDP-43 (any/none) | B1b.PC | B | -1.13E-07 | 5.54E-08 | 0.979212716 |
| Thal Phase | B1b.PC | B | 4.38E-08 | 8.18E-08 | 0.296194082 |
| WMR (any/none) | B1b.PC | B | -5.34E-08 | 3.39E-08 | 0.942597549 |
| #N/A | B1b.PC | B | 1.70E-09 | 4.34E-08 | 0.484370161 |
| AD Braak | BA.Bl | B | -8.84E-09 | 1.77E-08 | 0.691277042 |
| ADNC Score | BA.Bl | B | -9.52E-09 | 6.37E-08 | 0.55938057 |
| AP (any/none) | BA.Bl | B | 1.60E-08 | 2.08E-08 | 0.219690554 |
| Arteriolosclerosis | BA.Bl | B | 1.92E-08 | 2.49E-08 | 0.219707957 |
| Arteriolosclerosis (any | BA.Bl | B | 9.67E-09 | 2.71E-08 | 0.360860111 |
| Atherosclerosis | BA.Bl | B | 1.72E-08 | 2.27E-08 | 0.224653001 |
| Atherosclerosis (any/r | BA.Bl | B | -1.09E-08 | 2.20E-08 | 0.689752209 |
| B Score | BA.Bl | B | -1.32E-09 | 1.67E-08 | 0.531571789 |
| CAA | BA.Bl | B | -2.42E-08 | 2.39E-08 | 0.844986724 |
| CAA (any/none) | BA.Bl | B | -2.27E-08 | 2.12E-08 | 0.858042423 |
| CERAD (C Score) | BA.Bl | B | -1.72E-09 | 1.84E-08 | 0.537160641 |
| Infarcts/lacunes | BA.Bl | B | -2.08E-08 | 2.34E-08 | 0.813141739 |
| PD Braak | BA.Bl | B | -7.21E-09 | 2.07E-08 | 0.636293512 |
| PD Braak (any/none) | BA.Bl | B | -6.29E-10 | 1.88E-08 | 0.513335423 |
| TDP-43 (any/none) | BA.Bl | B | -1.40E-08 | 5.61E-08 | 0.598405785 |
| Thal Phase | BA.Bl | B | -1.58E-07 | 7.29E-08 | 0.984651232 |
| WMR (any/none) | BA.Bl | B | 2.70E-08 | 3.17E-08 | 0.196672151 |
| #N/A | BA.Bl | B | 1.86E-08 | 4.24E-08 | 0.330790075 |

| | | | | | |
|---|---|---|---|---|---|
| AD Braak | BA.Sp | B | -1.45E-08 | 1.81E-08 | 0.788318777 |
| ADNC Score | BA.Sp | B | -1.71E-08 | 5.55E-08 | 0.620749522 |
| AP (any/none) | BA.Sp | B | -7.57E-09 | 2.07E-08 | 0.642766074 |
| Arteriolosclerosis | BA.Sp | B | -3.23E-09 | 2.71E-08 | 0.547452252 |
| Arteriolosclerosis (any | BA.Sp | B | 5.93E-09 | 2.97E-08 | 0.420824624 |
| Atherosclerosis | BA.Sp | B | 6.34E-09 | 2.30E-08 | 0.391195522 |
| Atherosclerosis (any/r | BA.Sp | B | 6.23E-09 | 2.27E-08 | 0.391914039 |
| B Score | BA.Sp | B | -5.24E-09 | 1.77E-08 | 0.616362166 |
| CAA | BA.Sp | B | -1.81E-08 | 2.33E-08 | 0.781416726 |
| CAA (any/none) | BA.Sp | B | 8.33E-10 | 2.12E-08 | 0.484293157 |
| CERAD (C Score) | BA.Sp | B | -1.34E-08 | 1.94E-08 | 0.7554989 |
| Infarcts/lacunes | BA.Sp | B | 8.61E-09 | 2.38E-08 | 0.358531827 |
| PD Braak | BA.Sp | B | 1.31E-08 | 2.46E-08 | 0.297506709 |
| PD Braak (any/none) | BA.Sp | B | 1.48E-08 | 2.32E-08 | 0.261476014 |
| TDP-43 (any/none) | BA.Sp | B | -3.16E-08 | 5.15E-08 | 0.730307123 |
| Thal Phase | BA.Sp | B | -1.39E-07 | 7.11E-08 | 0.975045076 |
| WMR (any/none) | BA.Sp | B | 2.13E-08 | 3.14E-08 | 0.248697612 |
| #N/A | BA.Sp | B | 2.98E-08 | 4.31E-08 | 0.244323756 |
| AD Braak | BEC.MLN | B | -1.79E-08 | 1.63E-08 | 0.863327181 |
| ADNC Score | BEC.MLN | B | 1.23E-07 | 6.27E-08 | 0.024963792 |
| AP (any/none) | BEC.MLN | B | -5.08E-09 | 2.22E-08 | 0.590708089 |
| Arteriolosclerosis | BEC.MLN | B | -1.60E-09 | 2.72E-08 | 0.523495627 |
| Arteriolosclerosis (any | BEC.MLN | B | -4.14E-08 | 3.02E-08 | 0.914829029 |
| Atherosclerosis | BEC.MLN | B | 3.52E-09 | 2.39E-08 | 0.441437795 |
| Atherosclerosis (any/r | BEC.MLN | B | -6.15E-09 | 2.31E-08 | 0.604936403 |
| B Score | BEC.MLN | B | -2.27E-08 | 1.73E-08 | 0.904997027 |
| CAA | BEC.MLN | B | -3.19E-08 | 2.31E-08 | 0.916003889 |
| CAA (any/none) | BEC.MLN | B | -3.09E-08 | 2.30E-08 | 0.910696 |
| CERAD (C Score) | BEC.MLN | B | -5.81E-09 | 1.75E-08 | 0.629865207 |
| Infarcts/lacunes | BEC.MLN | B | -5.59E-09 | 2.36E-08 | 0.593511803 |
| PD Braak | BEC.MLN | B | 2.76E-09 | 1.98E-08 | 0.444467871 |
| PD Braak (any/none) | BEC.MLN | B | 3.15E-09 | 1.98E-08 | 0.436792928 |
| TDP-43 (any/none) | BEC.MLN | B | 1.75E-08 | 6.01E-08 | 0.385547195 |
| Thal Phase | BEC.MLN | B | 3.67E-08 | 7.54E-08 | 0.31328639 |

| | | | | | |
|---|---|---|---|---|---|
| WMR (any/none) | BEC.MLN | B | -5.80E-08 | 3.36E-08 | 0.957704923 |
| #N/A | BEC.MLN | B | -3.17E-08 | 4.05E-08 | 0.783571505 |
| AD Braak | BEC.SLN | B | -1.38E-08 | 1.58E-08 | 0.809630226 |
| ADNC Score | BEC.SLN | B | 1.16E-07 | 5.77E-08 | 0.022252031 |
| AP (any/none) | BEC.SLN | B | -6.51E-09 | 2.22E-08 | 0.615273675 |
| Arteriolosclerosis | BEC.SLN | B | -1.98E-09 | 2.64E-08 | 0.529877158 |
| Arteriolosclerosis (any | BEC.SLN | B | -3.63E-08 | 2.84E-08 | 0.898853953 |
| Atherosclerosis | BEC.SLN | B | -3.28E-09 | 2.39E-08 | 0.554574933 |
| Atherosclerosis (any/r | BEC.SLN | B | -1.10E-09 | 2.28E-08 | 0.519222852 |
| B Score | BEC.SLN | B | -2.01E-08 | 1.71E-08 | 0.879483042 |
| CAA | BEC.SLN | B | -3.05E-08 | 2.22E-08 | 0.915557011 |
| CAA (any/none) | BEC.SLN | B | -1.23E-08 | 2.25E-08 | 0.707711686 |
| CERAD (C Score) | BEC.SLN | B | -1.37E-09 | 1.74E-08 | 0.531459941 |
| Infarcts/lacunes | BEC.SLN | B | -1.29E-08 | 2.26E-08 | 0.716208912 |
| PD Braak | BEC.SLN | B | -7.34E-09 | 2.03E-08 | 0.641227564 |
| PD Braak (any/none) | BEC.SLN | B | -1.58E-09 | 1.98E-08 | 0.53167218 |
| TDP-43 (any/none) | BEC.SLN | B | 9.19E-09 | 5.83E-08 | 0.437379074 |
| Thal Phase | BEC.SLN | B | 5.14E-08 | 7.34E-08 | 0.242030706 |
| WMR (any/none) | BEC.SLN | B | -7.08E-08 | 3.36E-08 | 0.982604042 |
| #N/A | BEC.SLN | B | -2.51E-08 | 4.10E-08 | 0.729762118 |
| AD Braak | BEC.SLN.OT | B | -2.35E-08 | 1.66E-08 | 0.921920754 |
| ADNC Score | BEC.SLN.OT | B | 1.23E-07 | 6.08E-08 | 0.021630094 |
| AP (any/none) | BEC.SLN.OT | B | -1.63E-08 | 2.19E-08 | 0.771325731 |
| Arteriolosclerosis | BEC.SLN.OT | B | -1.13E-09 | 2.64E-08 | 0.517068266 |
| Arteriolosclerosis (any | BEC.SLN.OT | B | -1.64E-08 | 2.93E-08 | 0.712407575 |
| Atherosclerosis | BEC.SLN.OT | B | -1.02E-08 | 2.28E-08 | 0.672891475 |
| Atherosclerosis (any/r | BEC.SLN.OT | B | -8.31E-09 | 2.27E-08 | 0.642782095 |
| B Score | BEC.SLN.OT | B | -2.46E-08 | 1.76E-08 | 0.919713375 |
| CAA | BEC.SLN.OT | B | -2.46E-08 | 2.23E-08 | 0.865587888 |
| CAA (any/none) | BEC.SLN.OT | B | -1.81E-08 | 2.25E-08 | 0.789649845 |
| CERAD (C Score) | BEC.SLN.OT | B | 1.67E-09 | 1.73E-08 | 0.46136433 |
| Infarcts/lacunes | BEC.SLN.OT | B | 1.05E-08 | 2.33E-08 | 0.326887119 |
| PD Braak | BEC.SLN.OT | B | -8.81E-09 | 1.98E-08 | 0.671983992 |
| PD Braak (any/none) | BEC.SLN.OT | B | -4.63E-09 | 1.98E-08 | 0.592239365 |

| | | | | | |
|---|---|---|---|---|---|
| TDP-43 (any/none) | BEC.SLN.OT | B | -1.17E-09 | 5.99E-08 | 0.507821209 |
| Thal Phase | BEC.SLN.OT | B | 7.97E-08 | 7.44E-08 | 0.141780861 |
| WMR (any/none) | BEC.SLN.OT | B | -5.46E-08 | 3.40E-08 | 0.945957225 |
| #N/A | BEC.SLN.OT | B | -2.00E-08 | 4.09E-08 | 0.687793743 |
| AD Braak | CD19Control | B | 3.89E-08 | 1.98E-08 | 0.024867868 |
| ADNC Score | CD19Control | B | -2.21E-08 | 5.70E-08 | 0.650820557 |
| AP (any/none) | CD19Control | B | -1.41E-08 | 1.99E-08 | 0.760456491 |
| Arteriolosclerosis | CD19Control | B | 2.15E-08 | 2.57E-08 | 0.202210527 |
| Arteriolosclerosis (any | CD19Control | B | 4.40E-08 | 3.01E-08 | 0.071596781 |
| Atherosclerosis | CD19Control | B | 4.36E-08 | 2.46E-08 | 0.038221001 |
| Atherosclerosis (any/r | CD19Control | B | 1.61E-08 | 2.27E-08 | 0.238157257 |
| B Score | CD19Control | B | 2.72E-08 | 1.85E-08 | 0.071605513 |
| CAA | CD19Control | B | -4.51E-08 | 2.32E-08 | 0.97404027 |
| CAA (any/none) | CD19Control | B | -3.16E-08 | 2.35E-08 | 0.910639271 |
| CERAD (C Score) | CD19Control | B | 7.36E-09 | 2.11E-08 | 0.36350756 |
| Infarcts/lacunes | CD19Control | B | -2.23E-08 | 2.33E-08 | 0.830361139 |
| PD Braak | CD19Control | B | -1.48E-08 | 2.08E-08 | 0.761423072 |
| PD Braak (any/none) | CD19Control | B | -1.61E-08 | 1.94E-08 | 0.796256292 |
| TDP-43 (any/none) | CD19Control | B | -3.62E-08 | 5.32E-08 | 0.751758286 |
| Thal Phase | CD19Control | B | 3.02E-08 | 7.02E-08 | 0.333606202 |
| WMR (any/none) | CD19Control | B | -2.25E-08 | 3.21E-08 | 0.758050452 |
| #N/A | CD19Control | B | 5.29E-08 | 3.52E-08 | 0.066520554 |
| AD Braak | preB.FrC.BM | B | 2.41E-08 | 1.91E-08 | 0.103344303 |
| ADNC Score | preB.FrC.BM | B | -3.69E-08 | 5.97E-08 | 0.731953485 |
| AP (any/none) | preB.FrC.BM | B | -1.74E-09 | 2.25E-08 | 0.530871371 |
| Arteriolosclerosis | preB.FrC.BM | B | 4.80E-09 | 2.82E-08 | 0.432363764 |
| Arteriolosclerosis (any | preB.FrC.BM | B | 3.17E-08 | 2.91E-08 | 0.137890097 |
| Atherosclerosis | preB.FrC.BM | B | 1.27E-08 | 2.48E-08 | 0.304685526 |
| Atherosclerosis (any/r | preB.FrC.BM | B | 1.81E-08 | 2.30E-08 | 0.215661294 |
| B Score | preB.FrC.BM | B | 1.60E-08 | 1.99E-08 | 0.211339167 |
| CAA | preB.FrC.BM | B | -6.83E-09 | 2.41E-08 | 0.61138736 |
| CAA (any/none) | preB.FrC.BM | B | 4.28E-08 | 2.31E-08 | 0.032293942 |
| CERAD (C Score) | preB.FrC.BM | B | 4.90E-09 | 2.15E-08 | 0.410082858 |
| Infarcts/lacunes | preB.FrC.BM | B | -1.68E-08 | 2.49E-08 | 0.750243172 |

| | | | | | |
|---|---|---|---|---|---|
| PD Braak | preB.FrC.BM | B | -3.49E-08 | 2.09E-08 | 0.952567839 |
| PD Braak (any/none) | preB.FrC.BM | B | -2.59E-08 | 2.13E-08 | 0.887666351 |
| TDP-43 (any/none) | preB.FrC.BM | B | -1.32E-07 | 5.44E-08 | 0.992442948 |
| Thal Phase | preB.FrC.BM | B | 3.16E-08 | 7.88E-08 | 0.344117842 |
| WMR (any/none) | preB.FrC.BM | B | 3.92E-08 | 3.42E-08 | 0.125540771 |
| #N/A | preB.FrC.BM | B | -9.90E-08 | 4.34E-08 | 0.988714456 |
| AD Braak | preB.FrD.BM | B | 1.02E-08 | 2.09E-08 | 0.311988322 |
| ADNC Score | preB.FrD.BM | B | -6.44E-08 | 6.00E-08 | 0.858337847 |
| AP (any/none) | preB.FrD.BM | B | -1.48E-08 | 2.14E-08 | 0.755426696 |
| Arteriolosclerosis | preB.FrD.BM | B | -1.27E-08 | 2.72E-08 | 0.67964487 |
| Arteriolosclerosis (any | preB.FrD.BM | B | 8.73E-09 | 2.69E-08 | 0.372788421 |
| Atherosclerosis | preB.FrD.BM | B | 3.44E-08 | 2.24E-08 | 0.061964313 |
| Atherosclerosis (any/r | preB.FrD.BM | B | -1.54E-09 | 2.33E-08 | 0.52623234 |
| B Score | preB.FrD.BM | B | 9.93E-09 | 2.02E-08 | 0.311727578 |
| CAA | preB.FrD.BM | B | -1.55E-08 | 2.24E-08 | 0.755902597 |
| CAA (any/none) | preB.FrD.BM | B | 7.58E-09 | 2.13E-08 | 0.361003167 |
| CERAD (C Score) | preB.FrD.BM | B | -4.08E-09 | 2.11E-08 | 0.576824656 |
| Infarcts/lacunes | preB.FrD.BM | B | 5.37E-09 | 2.15E-08 | 0.401417856 |
| PD Braak | preB.FrD.BM | B | -1.11E-08 | 1.96E-08 | 0.713540622 |
| PD Braak (any/none) | preB.FrD.BM | B | -1.13E-08 | 1.94E-08 | 0.719249095 |
| TDP-43 (any/none) | preB.FrD.BM | B | -2.01E-08 | 5.76E-08 | 0.636481809 |
| Thal Phase | preB.FrD.BM | B | -1.14E-08 | 7.08E-08 | 0.563842548 |
| WMR (any/none) | preB.FrD.BM | B | 1.66E-08 | 3.17E-08 | 0.30047569 |
| #N/A | preB.FrD.BM | B | 4.67E-09 | 4.25E-08 | 0.456230593 |
| AD Braak | preB.FrD.FL | B | 2.37E-08 | 1.84E-08 | 0.098640757 |
| ADNC Score | preB.FrD.FL | B | 5.47E-09 | 5.62E-08 | 0.461202811 |
| AP (any/none) | preB.FrD.FL | B | 3.58E-09 | 2.10E-08 | 0.432256388 |
| Arteriolosclerosis | preB.FrD.FL | B | 5.38E-09 | 2.69E-08 | 0.420832383 |
| Arteriolosclerosis (any | preB.FrD.FL | B | 4.37E-08 | 2.70E-08 | 0.053036409 |
| Atherosclerosis | preB.FrD.FL | B | 2.30E-08 | 2.46E-08 | 0.174670955 |
| Atherosclerosis (any/r | preB.FrD.FL | B | 2.03E-08 | 2.31E-08 | 0.18977488 |
| B Score | preB.FrD.FL | B | 8.31E-09 | 1.79E-08 | 0.321238898 |
| CAA | preB.FrD.FL | B | 1.36E-08 | 2.44E-08 | 0.28806307 |
| CAA (any/none) | preB.FrD.FL | B | 3.17E-08 | 2.15E-08 | 0.069927131 |

| | | | | | |
|---|---|---|---|---|---|
| CERAD (C Score) | preB.FrD.FL | B | 1.25E-08 | 2.10E-08 | 0.275095932 |
| Infarcts/lacunes | preB.FrD.FL | B | -1.44E-08 | 2.32E-08 | 0.733131178 |
| PD Braak | preB.FrD.FL | B | -9.46E-09 | 2.25E-08 | 0.663096681 |
| PD Braak (any/none) | preB.FrD.FL | B | -1.18E-08 | 2.27E-08 | 0.698605144 |
| TDP-43 (any/none) | preB.FrD.FL | B | 1.07E-08 | 5.82E-08 | 0.426858223 |
| Thal Phase | preB.FrD.FL | B | 3.61E-08 | 6.66E-08 | 0.29367998 |
| WMR (any/none) | preB.FrD.FL | B | 5.29E-08 | 3.34E-08 | 0.056605985 |
| #N/A | preB.FrD.FL | B | 1.10E-08 | 4.16E-08 | 0.395900707 |
| AD Braak | proB.CLP.BM | B | 2.16E-08 | 1.77E-08 | 0.110471317 |
| ADNC Score | proB.CLP.BM | B | -2.62E-08 | 5.61E-08 | 0.679898679 |
| AP (any/none) | proB.CLP.BM | B | 7.33E-09 | 1.97E-08 | 0.354865384 |
| Arteriolosclerosis | proB.CLP.BM | B | 7.23E-08 | 3.12E-08 | 0.010287278 |
| Arteriolosclerosis (any | proB.CLP.BM | B | 5.55E-08 | 3.01E-08 | 0.032459733 |
| Atherosclerosis | proB.CLP.BM | B | 1.41E-09 | 2.28E-08 | 0.475385881 |
| Atherosclerosis (any/r | proB.CLP.BM | B | 2.39E-08 | 2.30E-08 | 0.149305309 |
| B Score | proB.CLP.BM | B | 1.76E-08 | 1.86E-08 | 0.172309937 |
| CAA | proB.CLP.BM | B | 3.36E-09 | 2.70E-08 | 0.450372199 |
| CAA (any/none) | proB.CLP.BM | B | 2.04E-08 | 2.44E-08 | 0.200860803 |
| CERAD (C Score) | proB.CLP.BM | B | 1.43E-08 | 2.12E-08 | 0.250765199 |
| Infarcts/lacunes | proB.CLP.BM | B | -4.86E-09 | 2.29E-08 | 0.584060699 |
| PD Braak | proB.CLP.BM | B | -1.13E-08 | 2.44E-08 | 0.677502141 |
| PD Braak (any/none) | proB.CLP.BM | B | -1.03E-08 | 2.30E-08 | 0.6732675 |
| TDP-43 (any/none) | proB.CLP.BM | B | -5.35E-08 | 5.45E-08 | 0.836726593 |
| Thal Phase | proB.CLP.BM | B | 3.82E-08 | 7.48E-08 | 0.304831127 |
| WMR (any/none) | proB.CLP.BM | B | 3.20E-08 | 3.35E-08 | 0.169788587 |
| #N/A | proB.CLP.BM | B | -6.66E-08 | 3.94E-08 | 0.954583028 |
| AD Braak | proB.CLP.FL | B | -2.07E-08 | 1.98E-08 | 0.85248909 |
| ADNC Score | proB.CLP.FL | B | 2.55E-08 | 5.86E-08 | 0.331586652 |
| AP (any/none) | proB.CLP.FL | B | 3.76E-09 | 2.18E-08 | 0.431548693 |
| Arteriolosclerosis | proB.CLP.FL | B | 6.31E-08 | 3.04E-08 | 0.018968229 |
| Arteriolosclerosis (any | proB.CLP.FL | B | 1.03E-07 | 3.35E-08 | 0.001052035 |
| Atherosclerosis | proB.CLP.FL | B | 1.47E-08 | 2.56E-08 | 0.282692673 |
| Atherosclerosis (any/r | proB.CLP.FL | B | 6.25E-08 | 2.67E-08 | 0.009580496 |
| B Score | proB.CLP.FL | B | -2.27E-08 | 2.05E-08 | 0.866410409 |

| | | | | | |
|---|---|---|---|---|---|
| CAA | proB.CLP.FL | B | 1.81E-08 | 2.48E-08 | 0.232572114 |
| CAA (any/none) | proB.CLP.FL | B | 1.58E-08 | 2.51E-08 | 0.264490885 |
| CERAD (C Score) | proB.CLP.FL | B | -8.53E-10 | 2.23E-08 | 0.515293178 |
| Infarcts/lacunes | proB.CLP.FL | B | -5.17E-08 | 2.42E-08 | 0.983766362 |
| PD Braak | proB.CLP.FL | B | 7.00E-09 | 2.65E-08 | 0.395865649 |
| PD Braak (any/none) | proB.CLP.FL | B | 9.41E-09 | 2.66E-08 | 0.361862025 |
| TDP-43 (any/none) | proB.CLP.FL | B | -5.79E-09 | 6.22E-08 | 0.53711813 |
| Thal Phase | proB.CLP.FL | B | 2.35E-08 | 7.57E-08 | 0.378355207 |
| WMR (any/none) | proB.CLP.FL | B | 6.62E-08 | 3.83E-08 | 0.042037113 |
| #N/A | proB.CLP.FL | B | -1.14E-07 | 4.33E-08 | 0.995796645 |
| AD Braak | proB.FrA.BM | B | 4.14E-08 | 1.86E-08 | 0.012944185 |
| ADNC Score | proB.FrA.BM | B | -3.39E-08 | 6.20E-08 | 0.707842893 |
| AP (any/none) | proB.FrA.BM | B | 3.60E-08 | 2.12E-08 | 0.044518886 |
| Arteriolosclerosis | proB.FrA.BM | B | 6.49E-08 | 3.17E-08 | 0.02041376 |
| Arteriolosclerosis (any | proB.FrA.BM | B | 2.78E-08 | 2.99E-08 | 0.176513954 |
| Atherosclerosis | proB.FrA.BM | B | 1.84E-08 | 2.15E-08 | 0.196743817 |
| Atherosclerosis (any/r | proB.FrA.BM | B | 1.47E-08 | 2.11E-08 | 0.242600566 |
| B Score | proB.FrA.BM | B | 3.47E-08 | 1.94E-08 | 0.036672307 |
| CAA | proB.FrA.BM | B | -1.28E-08 | 2.39E-08 | 0.703630134 |
| CAA (any/none) | proB.FrA.BM | B | 1.73E-08 | 2.24E-08 | 0.220693799 |
| CERAD (C Score) | proB.FrA.BM | B | 5.97E-08 | 2.23E-08 | 0.003672683 |
| Infarcts/lacunes | proB.FrA.BM | B | -6.45E-09 | 2.39E-08 | 0.606587516 |
| PD Braak | proB.FrA.BM | B | -1.67E-08 | 2.28E-08 | 0.768080442 |
| PD Braak (any/none) | proB.FrA.BM | B | -1.72E-08 | 2.12E-08 | 0.79121567 |
| TDP-43 (any/none) | proB.FrA.BM | B | -4.63E-08 | 5.47E-08 | 0.801244415 |
| Thal Phase | proB.FrA.BM | B | -2.77E-09 | 7.54E-08 | 0.51465837 |
| WMR (any/none) | proB.FrA.BM | B | 3.11E-08 | 3.21E-08 | 0.166570355 |
| #N/A | proB.FrA.BM | B | -2.49E-08 | 4.12E-08 | 0.727299372 |
| AD Braak | proB.FrA.FL | B | -9.37E-09 | 1.99E-08 | 0.681283279 |
| ADNC Score | proB.FrA.FL | B | -1.69E-08 | 5.76E-08 | 0.61550939 |
| AP (any/none) | proB.FrA.FL | B | -1.01E-08 | 2.05E-08 | 0.688900625 |
| Arteriolosclerosis | proB.FrA.FL | B | 7.20E-08 | 3.31E-08 | 0.014727189 |
| Arteriolosclerosis (any | proB.FrA.FL | B | 9.73E-08 | 3.22E-08 | 0.001243356 |
| Atherosclerosis | proB.FrA.FL | B | -2.60E-09 | 2.44E-08 | 0.542313521 |

| | | | | | |
|---|---|---|---|---|---|
| Atherosclerosis (any/r | proB.FrA.FL | B | 3.47E-08 | 2.58E-08 | 0.089085907 |
| B Score | proB.FrA.FL | B | -7.18E-09 | 1.99E-08 | 0.641198517 |
| CAA | proB.FrA.FL | B | 2.78E-09 | 2.45E-08 | 0.454706712 |
| CAA (any/none) | proB.FrA.FL | B | 8.63E-09 | 2.37E-08 | 0.357699849 |
| CERAD (C Score) | proB.FrA.FL | B | 5.71E-09 | 2.25E-08 | 0.399764728 |
| Infarcts/lacunes | proB.FrA.FL | B | -2.97E-08 | 2.48E-08 | 0.884822662 |
| PD Braak | proB.FrA.FL | B | 3.35E-09 | 2.55E-08 | 0.447680789 |
| PD Braak (any/none) | proB.FrA.FL | B | 3.75E-09 | 2.47E-08 | 0.439732103 |
| TDP-43 (any/none) | proB.FrA.FL | B | 2.61E-08 | 6.18E-08 | 0.33627435 |
| Thal Phase | proB.FrA.FL | B | 1.74E-08 | 8.12E-08 | 0.415333519 |
| WMR (any/none) | proB.FrA.FL | B | 6.17E-08 | 3.84E-08 | 0.054085385 |
| #N/A | proB.FrA.FL | B | -6.01E-08 | 3.96E-08 | 0.935256399 |
| AD Braak | proB.FrBC.BM | B | 2.96E-08 | 2.07E-08 | 0.075781228 |
| ADNC Score | proB.FrBC.BM | B | -1.31E-09 | 5.95E-08 | 0.508750408 |
| AP (any/none) | proB.FrBC.BM | B | 7.03E-09 | 2.25E-08 | 0.37723672 |
| Arteriolosclerosis | proB.FrBC.BM | B | 1.32E-08 | 2.84E-08 | 0.321765194 |
| Arteriolosclerosis (any | proB.FrBC.BM | B | 5.27E-08 | 3.01E-08 | 0.039966493 |
| Atherosclerosis | proB.FrBC.BM | B | 2.79E-08 | 2.29E-08 | 0.111590385 |
| Atherosclerosis (any/r | proB.FrBC.BM | B | 1.17E-08 | 2.19E-08 | 0.29701479 |
| B Score | proB.FrBC.BM | B | 1.68E-08 | 2.03E-08 | 0.20399034 |
| CAA | proB.FrBC.BM | B | -5.85E-09 | 2.62E-08 | 0.588507787 |
| CAA (any/none) | proB.FrBC.BM | B | 4.70E-08 | 2.32E-08 | 0.02167711 |
| CERAD (C Score) | proB.FrBC.BM | B | 2.16E-08 | 2.01E-08 | 0.141197577 |
| Infarcts/lacunes | proB.FrBC.BM | B | -1.46E-08 | 2.29E-08 | 0.738259877 |
| PD Braak | proB.FrBC.BM | B | -3.97E-08 | 2.25E-08 | 0.960956421 |
| PD Braak (any/none) | proB.FrBC.BM | B | -4.43E-08 | 2.26E-08 | 0.975107519 |
| TDP-43 (any/none) | proB.FrBC.BM | B | -1.28E-08 | 5.85E-08 | 0.586307308 |
| Thal Phase | proB.FrBC.BM | B | 1.19E-07 | 7.48E-08 | 0.056497039 |
| WMR (any/none) | proB.FrBC.BM | B | -1.83E-09 | 3.40E-08 | 0.521474834 |
| #N/A | proB.FrBC.BM | B | -3.46E-08 | 4.22E-08 | 0.793742553 |
| AD Braak | proB.FrBC.FL | B | 1.17E-08 | 2.17E-08 | 0.294973753 |
| ADNC Score | proB.FrBC.FL | B | 3.49E-08 | 6.89E-08 | 0.306352316 |
| AP (any/none) | proB.FrBC.FL | B | 1.53E-08 | 2.39E-08 | 0.260109836 |
| Arteriolosclerosis | proB.FrBC.FL | B | 3.71E-08 | 3.19E-08 | 0.123067543 |

| | | | | | |
|---|---|---|---|---|---|
| Arteriolosclerosis (any | proB.FrBC.FL | B | 5.72E-08 | 3.25E-08 | 0.039148262 |
| Atherosclerosis | proB.FrBC.FL | B | 3.69E-09 | 2.60E-08 | 0.443563055 |
| Atherosclerosis (any/r | proB.FrBC.FL | B | 2.82E-08 | 2.55E-08 | 0.133921965 |
| B Score | proB.FrBC.FL | B | -1.01E-08 | 2.10E-08 | 0.684835646 |
| CAA | proB.FrBC.FL | B | -3.34E-09 | 2.77E-08 | 0.547964023 |
| CAA (any/none) | proB.FrBC.FL | B | 2.06E-08 | 2.50E-08 | 0.205881436 |
| CERAD (C Score) | proB.FrBC.FL | B | 6.92E-09 | 2.24E-08 | 0.378877339 |
| Infarcts/lacunes | proB.FrBC.FL | B | -6.51E-09 | 2.63E-08 | 0.597824237 |
| PD Braak | proB.FrBC.FL | B | -2.77E-09 | 3.07E-08 | 0.536051494 |
| PD Braak (any/none) | proB.FrBC.FL | B | -6.99E-09 | 2.98E-08 | 0.59279503 |
| TDP-43 (any/none) | proB.FrBC.FL | B | -8.20E-08 | 5.55E-08 | 0.930236551 |
| Thal Phase | proB.FrBC.FL | B | 5.35E-08 | 8.71E-08 | 0.269592647 |
| WMR (any/none) | proB.FrBC.FL | B | 8.65E-08 | 4.02E-08 | 0.015659201 |
| #N/A | proB.FrBC.FL | B | -8.56E-08 | 4.54E-08 | 0.970185369 |
| AD Braak | ILC1.CD127+.SI | Innate_lymphocyte | -8.06E-09 | 1.89E-08 | 0.665277791 |
| ADNC Score | ILC1.CD127+.SI | Innate_lymphocyte | -4.92E-08 | 5.61E-08 | 0.809896195 |
| AP (any/none) | ILC1.CD127+.SI | Innate_lymphocyte | 3.96E-08 | 2.15E-08 | 0.033154705 |
| Arteriolosclerosis | ILC1.CD127+.SI | Innate_lymphocyte | 3.06E-08 | 2.91E-08 | 0.146544976 |
| Arteriolosclerosis (any | ILC1.CD127+.SI | Innate_lymphocyte | 4.87E-08 | 2.92E-08 | 0.04767915 |
| Atherosclerosis | ILC1.CD127+.SI | Innate_lymphocyte | 7.34E-09 | 2.15E-08 | 0.36651894 |
| Atherosclerosis (any/r | ILC1.CD127+.SI | Innate_lymphocyte | 1.66E-08 | 2.37E-08 | 0.241556279 |
| B Score | ILC1.CD127+.SI | Innate_lymphocyte | 9.41E-09 | 1.83E-08 | 0.303349788 |
| CAA | ILC1.CD127+.SI | Innate_lymphocyte | 6.83E-09 | 2.15E-08 | 0.375271444 |
| CAA (any/none) | ILC1.CD127+.SI | Innate_lymphocyte | 3.59E-08 | 2.18E-08 | 0.050136503 |
| CERAD (C Score) | ILC1.CD127+.SI | Innate_lymphocyte | 8.73E-09 | 1.91E-08 | 0.323590321 |
| Infarcts/lacunes | ILC1.CD127+.SI | Innate_lymphocyte | -5.20E-08 | 2.31E-08 | 0.987896787 |
| PD Braak | ILC1.CD127+.SI | Innate_lymphocyte | -2.15E-08 | 2.18E-08 | 0.838008456 |
| PD Braak (any/none) | ILC1.CD127+.SI | Innate_lymphocyte | -2.52E-08 | 2.21E-08 | 0.872601702 |
| TDP-43 (any/none) | ILC1.CD127+.SI | Innate_lymphocyte | 3.21E-08 | 5.42E-08 | 0.276435173 |
| Thal Phase | ILC1.CD127+.SI | Innate_lymphocyte | -9.87E-08 | 7.26E-08 | 0.912979972 |
| WMR (any/none) | ILC1.CD127+.SI | Innate_lymphocyte | 3.02E-08 | 3.52E-08 | 0.195573968 |
| #N/A | ILC1.CD127+.SI | Innate_lymphocyte | -2.59E-11 | 4.19E-08 | 0.500246435 |
| AD Braak | ILC1.CD127+.Sp | Innate_lymphocyte | -3.49E-08 | 1.95E-08 | 0.963358044 |
| ADNC Score | ILC1.CD127+.Sp | Innate_lymphocyte | -2.17E-08 | 5.20E-08 | 0.661898296 |

| | | | | | |
|---|---|---|---|---|---|
| AP (any/none) | ILC1.CD127+.Sp | Innate_lymphocyte | -1.58E-09 | 2.10E-08 | 0.529889302 |
| Arteriolosclerosis | ILC1.CD127+.Sp | Innate_lymphocyte | 4.16E-08 | 2.86E-08 | 0.07269753 |
| Arteriolosclerosis (any | ILC1.CD127+.Sp | Innate_lymphocyte | 9.06E-10 | 2.63E-08 | 0.486252463 |
| Atherosclerosis | ILC1.CD127+.Sp | Innate_lymphocyte | -9.20E-09 | 2.07E-08 | 0.671697391 |
| Atherosclerosis (any/r | ILC1.CD127+.Sp | Innate_lymphocyte | 1.04E-08 | 2.47E-08 | 0.336764027 |
| B Score | ILC1.CD127+.Sp | Innate_lymphocyte | -1.59E-08 | 1.88E-08 | 0.801275478 |
| CAA | ILC1.CD127+.Sp | Innate_lymphocyte | 1.93E-08 | 2.04E-08 | 0.17152171 |
| CAA (any/none) | ILC1.CD127+.Sp | Innate_lymphocyte | 2.86E-08 | 2.31E-08 | 0.107844739 |
| CERAD (C Score) | ILC1.CD127+.Sp | Innate_lymphocyte | 2.08E-08 | 1.95E-08 | 0.143013999 |
| Infarcts/lacunes | ILC1.CD127+.Sp | Innate_lymphocyte | -5.41E-08 | 1.98E-08 | 0.996879007 |
| PD Braak | ILC1.CD127+.Sp | Innate_lymphocyte | -5.36E-10 | 2.28E-08 | 0.509384831 |
| PD Braak (any/none) | ILC1.CD127+.Sp | Innate_lymphocyte | 4.21E-09 | 2.27E-08 | 0.426460795 |
| TDP-43 (any/none) | ILC1.CD127+.Sp | Innate_lymphocyte | 8.07E-08 | 5.37E-08 | 0.066399597 |
| Thal Phase | ILC1.CD127+.Sp | Innate_lymphocyte | -7.01E-08 | 6.77E-08 | 0.849821007 |
| WMR (any/none) | ILC1.CD127+.Sp | Innate_lymphocyte | 6.64E-09 | 3.58E-08 | 0.426389111 |
| #N/A | ILC1.CD127+.Sp | Innate_lymphocyte | -5.31E-09 | 4.49E-08 | 0.547052512 |
| AD Braak | ILC1.CD49b-.Lv | Innate_lymphocyte | -3.52E-08 | 1.79E-08 | 0.975348574 |
| ADNC Score | ILC1.CD49b-.Lv | Innate_lymphocyte | -1.30E-07 | 5.05E-08 | 0.994819959 |
| AP (any/none) | ILC1.CD49b-.Lv | Innate_lymphocyte | -1.73E-08 | 1.95E-08 | 0.812725459 |
| Arteriolosclerosis | ILC1.CD49b-.Lv | Innate_lymphocyte | 4.58E-08 | 2.74E-08 | 0.047288752 |
| Arteriolosclerosis (any | ILC1.CD49b-.Lv | Innate_lymphocyte | 1.77E-09 | 2.68E-08 | 0.473714877 |
| Atherosclerosis | ILC1.CD49b-.Lv | Innate_lymphocyte | 2.17E-09 | 1.88E-08 | 0.454181165 |
| Atherosclerosis (any/r | ILC1.CD49b-.Lv | Innate_lymphocyte | 1.21E-08 | 2.09E-08 | 0.281828577 |
| B Score | ILC1.CD49b-.Lv | Innate_lymphocyte | -1.62E-08 | 1.69E-08 | 0.830753177 |
| CAA | ILC1.CD49b-.Lv | Innate_lymphocyte | -1.09E-08 | 1.96E-08 | 0.71064236 |
| CAA (any/none) | ILC1.CD49b-.Lv | Innate_lymphocyte | -2.18E-08 | 2.21E-08 | 0.838038514 |
| CERAD (C Score) | ILC1.CD49b-.Lv | Innate_lymphocyte | -1.79E-08 | 1.77E-08 | 0.843412224 |
| Infarcts/lacunes | ILC1.CD49b-.Lv | Innate_lymphocyte | -1.81E-08 | 2.18E-08 | 0.795815103 |
| PD Braak | ILC1.CD49b-.Lv | Innate_lymphocyte | 2.58E-08 | 2.14E-08 | 0.114654077 |
| PD Braak (any/none) | ILC1.CD49b-.Lv | Innate_lymphocyte | 2.43E-08 | 2.09E-08 | 0.122547601 |
| TDP-43 (any/none) | ILC1.CD49b-.Lv | Innate_lymphocyte | 7.80E-08 | 5.42E-08 | 0.075256664 |
| Thal Phase | ILC1.CD49b-.Lv | Innate_lymphocyte | -7.60E-08 | 6.29E-08 | 0.886532585 |
| WMR (any/none) | ILC1.CD49b-.Lv | Innate_lymphocyte | 5.68E-08 | 3.46E-08 | 0.050142389 |
| #N/A | ILC1.CD49b-.Lv | Innate_lymphocyte | -2.68E-08 | 3.82E-08 | 0.758594035 |

| | | | | | |
|---|---|---|---|---|---|
| AD Braak | ILC2.SI | Innate_lymphocyte | -3.10E-08 | 1.74E-08 | 0.9628156 |
| ADNC Score | ILC2.SI | Innate_lymphocyte | -1.76E-09 | 5.44E-08 | 0.512907996 |
| AP (any/none) | ILC2.SI | Innate_lymphocyte | -1.08E-08 | 1.99E-08 | 0.706365935 |
| Arteriolosclerosis | ILC2.SI | Innate_lymphocyte | 4.86E-08 | 2.57E-08 | 0.029054595 |
| Arteriolosclerosis (any | ILC2.SI | Innate_lymphocyte | 1.70E-08 | 2.34E-08 | 0.234043842 |
| Atherosclerosis | ILC2.SI | Innate_lymphocyte | -2.47E-08 | 2.06E-08 | 0.884520269 |
| Atherosclerosis (any/r | ILC2.SI | Innate_lymphocyte | -9.22E-09 | 2.26E-08 | 0.65823619 |
| B Score | ILC2.SI | Innate_lymphocyte | -2.26E-08 | 1.47E-08 | 0.93801781 |
| CAA | ILC2.SI | Innate_lymphocyte | 2.62E-08 | 2.03E-08 | 0.09837769 |
| CAA (any/none) | ILC2.SI | Innate_lymphocyte | 1.66E-08 | 2.10E-08 | 0.215437269 |
| CERAD (C Score) | ILC2.SI | Innate_lymphocyte | -2.45E-08 | 1.68E-08 | 0.926963388 |
| Infarcts/lacunes | ILC2.SI | Innate_lymphocyte | -1.94E-08 | 2.14E-08 | 0.817256886 |
| PD Braak | ILC2.SI | Innate_lymphocyte | 1.76E-08 | 2.14E-08 | 0.205821115 |
| PD Braak (any/none) | ILC2.SI | Innate_lymphocyte | 1.75E-08 | 2.12E-08 | 0.204693973 |
| TDP-43 (any/none) | ILC2.SI | Innate_lymphocyte | 4.31E-08 | 5.29E-08 | 0.207739411 |
| Thal Phase | ILC2.SI | Innate_lymphocyte | -8.05E-08 | 7.27E-08 | 0.865898597 |
| WMR (any/none) | ILC2.SI | Innate_lymphocyte | 4.38E-08 | 3.27E-08 | 0.089945656 |
| #N/A | ILC2.SI | Innate_lymphocyte | -4.68E-08 | 3.72E-08 | 0.89562168 |
| AD Braak | ILC3.NKp46-.4-.SI | Innate_lymphocyte | -2.65E-08 | 1.52E-08 | 0.959304386 |
| ADNC Score | ILC3.NKp46-.4-.SI | Innate_lymphocyte | 1.19E-08 | 5.55E-08 | 0.415039657 |
| AP (any/none) | ILC3.NKp46-.4-.SI | Innate_lymphocyte | 4.19E-10 | 1.89E-08 | 0.491161215 |
| Arteriolosclerosis | ILC3.NKp46-.4-.SI | Innate_lymphocyte | 8.19E-09 | 2.86E-08 | 0.387368738 |
| Arteriolosclerosis (any | ILC3.NKp46-.4-.SI | Innate_lymphocyte | 1.62E-08 | 2.52E-08 | 0.259587273 |
| Atherosclerosis | ILC3.NKp46-.4-.SI | Innate_lymphocyte | 2.97E-09 | 2.09E-08 | 0.443492138 |
| Atherosclerosis (any/r | ILC3.NKp46-.4-.SI | Innate_lymphocyte | 2.44E-08 | 2.16E-08 | 0.129139201 |
| B Score | ILC3.NKp46-.4-.SI | Innate_lymphocyte | -1.44E-08 | 1.51E-08 | 0.829749556 |
| CAA | ILC3.NKp46-.4-.SI | Innate_lymphocyte | 1.15E-08 | 2.21E-08 | 0.301075007 |
| CAA (any/none) | ILC3.NKp46-.4-.SI | Innate_lymphocyte | 3.06E-08 | 2.18E-08 | 0.079594641 |
| CERAD (C Score) | ILC3.NKp46-.4-.SI | Innate_lymphocyte | -5.99E-09 | 1.70E-08 | 0.637661789 |
| Infarcts/lacunes | ILC3.NKp46-.4-.SI | Innate_lymphocyte | -1.81E-08 | 2.18E-08 | 0.797312026 |
| PD Braak | ILC3.NKp46-.4-.SI | Innate_lymphocyte | -5.57E-09 | 1.91E-08 | 0.61501003 |
| PD Braak (any/none) | ILC3.NKp46-.4-.SI | Innate_lymphocyte | -1.45E-08 | 1.95E-08 | 0.77243867 |
| TDP-43 (any/none) | ILC3.NKp46-.4-.SI | Innate_lymphocyte | -3.28E-08 | 5.10E-08 | 0.740089336 |
| Thal Phase | ILC3.NKp46-.4-.SI | Innate_lymphocyte | -2.61E-08 | 6.54E-08 | 0.655213547 |

| | | | | | |
|---|---|---|---|---|---|
| WMR (any/none) | ILC3.NKp46-.4-.SI | Innate_lymphocyte | 4.04E-08 | 3.15E-08 | 0.100094993 |
| #N/A | ILC3.NKp46-.4-.SI | Innate_lymphocyte | 7.51E-09 | 3.96E-08 | 0.424771533 |
| AD Braak | ILC3.NKp46-.4+.SI | Innate_lymphocyte | -6.49E-09 | 1.54E-08 | 0.663064359 |
| ADNC Score | ILC3.NKp46-.4+.SI | Innate_lymphocyte | 6.16E-08 | 5.17E-08 | 0.116800968 |
| AP (any/none) | ILC3.NKp46-.4+.SI | Innate_lymphocyte | 1.25E-08 | 1.81E-08 | 0.245925125 |
| Arteriolosclerosis | ILC3.NKp46-.4+.SI | Innate_lymphocyte | 3.65E-08 | 2.53E-08 | 0.07489711 |
| Arteriolosclerosis (any | ILC3.NKp46-.4+.SI | Innate_lymphocyte | 5.05E-08 | 2.45E-08 | 0.019835271 |
| Atherosclerosis | ILC3.NKp46-.4+.SI | Innate_lymphocyte | 1.77E-08 | 2.12E-08 | 0.201493321 |
| Atherosclerosis (any/r | ILC3.NKp46-.4+.SI | Innate_lymphocyte | 1.48E-08 | 2.08E-08 | 0.238874081 |
| B Score | ILC3.NKp46-.4+.SI | Innate_lymphocyte | 1.97E-09 | 1.71E-08 | 0.454152593 |
| CAA | ILC3.NKp46-.4+.SI | Innate_lymphocyte | 1.23E-08 | 2.33E-08 | 0.298210773 |
| CAA (any/none) | ILC3.NKp46-.4+.SI | Innate_lymphocyte | 4.37E-09 | 2.17E-08 | 0.420304064 |
| CERAD (C Score) | ILC3.NKp46-.4+.SI | Innate_lymphocyte | -9.32E-09 | 1.63E-08 | 0.71691556 |
| Infarcts/lacunes | ILC3.NKp46-.4+.SI | Innate_lymphocyte | -3.34E-08 | 2.17E-08 | 0.938286306 |
| PD Braak | ILC3.NKp46-.4+.SI | Innate_lymphocyte | -1.89E-08 | 1.87E-08 | 0.843818888 |
| PD Braak (any/none) | ILC3.NKp46-.4+.SI | Innate_lymphocyte | -1.65E-08 | 1.87E-08 | 0.810165395 |
| TDP-43 (any/none) | ILC3.NKp46-.4+.SI | Innate_lymphocyte | -1.87E-08 | 4.68E-08 | 0.655126992 |
| Thal Phase | ILC3.NKp46-.4+.SI | Innate_lymphocyte | -6.56E-08 | 6.53E-08 | 0.842543714 |
| WMR (any/none) | ILC3.NKp46-.4+.SI | Innate_lymphocyte | 3.61E-08 | 3.19E-08 | 0.128842846 |
| #N/A | ILC3.NKp46-.4+.SI | Innate_lymphocyte | 6.80E-09 | 3.57E-08 | 0.424403 |
| AD Braak | ILC3.NKp46+.Rorgthi.SI | Innate_lymphocyte | -2.44E-08 | 1.75E-08 | 0.918644686 |
| ADNC Score | ILC3.NKp46+.Rorgthi.SI | Innate_lymphocyte | -2.13E-08 | 5.33E-08 | 0.655361988 |
| AP (any/none) | ILC3.NKp46+.Rorgthi.SI | Innate_lymphocyte | -1.71E-08 | 1.83E-08 | 0.825368034 |
| Arteriolosclerosis | ILC3.NKp46+.Rorgthi.SI | Innate_lymphocyte | -1.47E-08 | 2.72E-08 | 0.706058449 |
| Arteriolosclerosis (any | ILC3.NKp46+.Rorgthi.SI | Innate_lymphocyte | 1.32E-08 | 2.78E-08 | 0.317016266 |
| Atherosclerosis | ILC3.NKp46+.Rorgthi.SI | Innate_lymphocyte | -1.37E-08 | 2.14E-08 | 0.739291077 |
| Atherosclerosis (any/r | ILC3.NKp46+.Rorgthi.SI | Innate_lymphocyte | 1.41E-08 | 2.12E-08 | 0.25419431 |
| B Score | ILC3.NKp46+.Rorgthi.SI | Innate_lymphocyte | -3.43E-09 | 1.67E-08 | 0.581156303 |
| CAA | ILC3.NKp46+.Rorgthi.SI | Innate_lymphocyte | 8.35E-09 | 2.15E-08 | 0.348585816 |
| CAA (any/none) | ILC3.NKp46+.Rorgthi.SI | Innate_lymphocyte | 1.79E-08 | 2.19E-08 | 0.207893327 |
| CERAD (C Score) | ILC3.NKp46+.Rorgthi.SI | Innate_lymphocyte | -9.74E-09 | 1.78E-08 | 0.707579939 |
| Infarcts/lacunes | ILC3.NKp46+.Rorgthi.SI | Innate_lymphocyte | -2.68E-08 | 2.22E-08 | 0.886535352 |
| PD Braak | ILC3.NKp46+.Rorgthi.SI | Innate_lymphocyte | -2.56E-08 | 2.17E-08 | 0.881008298 |
| PD Braak (any/none) | ILC3.NKp46+.Rorgthi.SI | Innate_lymphocyte | -2.62E-08 | 2.12E-08 | 0.891021503 |

| | | | | | |
|---|---|---|---|---|---|
| TDP-43 (any/none) | ILC3.NKp46+.Rorgthi.SI | Innate_lymphocyte | 8.89E-08 | 5.48E-08 | 0.052510195 |
| Thal Phase | ILC3.NKp46+.Rorgthi.SI | Innate_lymphocyte | -7.89E-08 | 7.08E-08 | 0.867389795 |
| WMR (any/none) | ILC3.NKp46+.Rorgthi.SI | Innate_lymphocyte | -1.05E-08 | 3.24E-08 | 0.627257804 |
| #N/A | ILC3.NKp46+.Rorgthi.SI | Innate_lymphocyte | 1.49E-08 | 4.10E-08 | 0.357808752 |
| AD Braak | NK.49CI-.Sp | Innate_lymphocyte | 2.75E-10 | 2.01E-08 | 0.494542465 |
| ADNC Score | NK.49CI-.Sp | Innate_lymphocyte | -3.88E-08 | 4.56E-08 | 0.802322689 |
| AP (any/none) | NK.49CI-.Sp | Innate_lymphocyte | -8.66E-09 | 1.83E-08 | 0.682501813 |
| Arteriolosclerosis | NK.49CI-.Sp | Innate_lymphocyte | 3.52E-08 | 2.55E-08 | 0.083546673 |
| Arteriolosclerosis (any | NK.49CI-.Sp | Innate_lymphocyte | 2.17E-09 | 2.88E-08 | 0.46987896 |
| Atherosclerosis | NK.49CI-.Sp | Innate_lymphocyte | 3.45E-08 | 2.28E-08 | 0.064950572 |
| Atherosclerosis (any/r | NK.49CI-.Sp | Innate_lymphocyte | 2.00E-08 | 2.48E-08 | 0.210163386 |
| B Score | NK.49CI-.Sp | Innate_lymphocyte | 6.76E-09 | 1.92E-08 | 0.362509534 |
| CAA | NK.49CI-.Sp | Innate_lymphocyte | 1.22E-08 | 2.13E-08 | 0.283834525 |
| CAA (any/none) | NK.49CI-.Sp | Innate_lymphocyte | -6.89E-09 | 2.17E-08 | 0.624476557 |
| CERAD (C Score) | NK.49CI-.Sp | Innate_lymphocyte | 4.74E-09 | 1.72E-08 | 0.391104243 |
| Infarcts/lacunes | NK.49CI-.Sp | Innate_lymphocyte | -3.04E-08 | 2.24E-08 | 0.912316074 |
| PD Braak | NK.49CI-.Sp | Innate_lymphocyte | -2.95E-08 | 1.75E-08 | 0.953935075 |
| PD Braak (any/none) | NK.49CI-.Sp | Innate_lymphocyte | -2.95E-08 | 1.58E-08 | 0.968993315 |
| TDP-43 (any/none) | NK.49CI-.Sp | Innate_lymphocyte | -5.30E-09 | 5.13E-08 | 0.541163691 |
| Thal Phase | NK.49CI-.Sp | Innate_lymphocyte | -9.76E-08 | 6.76E-08 | 0.925682133 |
| WMR (any/none) | NK.49CI-.Sp | Innate_lymphocyte | -3.02E-08 | 2.81E-08 | 0.859024793 |
| #N/A | NK.49CI-.Sp | Innate_lymphocyte | 9.11E-09 | 4.12E-08 | 0.412516347 |
| AD Braak | NK.49CI+.Sp | Innate_lymphocyte | 1.16E-08 | 1.94E-08 | 0.27566032 |
| ADNC Score | NK.49CI+.Sp | Innate_lymphocyte | -6.87E-08 | 5.35E-08 | 0.900384036 |
| AP (any/none) | NK.49CI+.Sp | Innate_lymphocyte | -9.53E-09 | 2.07E-08 | 0.677667396 |
| Arteriolosclerosis | NK.49CI+.Sp | Innate_lymphocyte | -1.38E-08 | 2.56E-08 | 0.704175744 |
| Arteriolosclerosis (any | NK.49CI+.Sp | Innate_lymphocyte | -3.82E-08 | 2.99E-08 | 0.899079015 |
| Atherosclerosis | NK.49CI+.Sp | Innate_lymphocyte | 2.53E-08 | 2.35E-08 | 0.140749586 |
| Atherosclerosis (any/r | NK.49CI+.Sp | Innate_lymphocyte | 2.23E-08 | 2.40E-08 | 0.177221418 |
| B Score | NK.49CI+.Sp | Innate_lymphocyte | 1.91E-08 | 1.89E-08 | 0.155924438 |
| CAA | NK.49CI+.Sp | Innate_lymphocyte | 1.89E-08 | 2.32E-08 | 0.207628512 |
| CAA (any/none) | NK.49CI+.Sp | Innate_lymphocyte | 8.97E-09 | 2.37E-08 | 0.352363268 |
| CERAD (C Score) | NK.49CI+.Sp | Innate_lymphocyte | 1.68E-08 | 1.75E-08 | 0.168900701 |
| Infarcts/lacunes | NK.49CI+.Sp | Innate_lymphocyte | -1.69E-08 | 2.39E-08 | 0.760896082 |

| | | | | | |
|---|---|---|---|---|---|
| PD Braak | NK.49CI+.Sp | Innate_lymphocyte | -9.25E-09 | 2.14E-08 | 0.667077566 |
| PD Braak (any/none) | NK.49CI+.Sp | Innate_lymphocyte | 3.34E-09 | 2.04E-08 | 0.434920927 |
| TDP-43 (any/none) | NK.49CI+.Sp | Innate_lymphocyte | -3.99E-08 | 5.59E-08 | 0.76201549 |
| Thal Phase | NK.49CI+.Sp | Innate_lymphocyte | -1.70E-07 | 6.87E-08 | 0.993252872 |
| WMR (any/none) | NK.49CI+.Sp | Innate_lymphocyte | -2.02E-08 | 3.41E-08 | 0.72298219 |
| #N/A | NK.49CI+.Sp | Innate_lymphocyte | 3.25E-08 | 4.71E-08 | 0.245056486 |
| AD Braak | NK.49H-.Sp | Innate_lymphocyte | 1.24E-08 | 2.00E-08 | 0.26793422 |
| ADNC Score | NK.49H-.Sp | Innate_lymphocyte | -1.41E-07 | 5.07E-08 | 0.997349606 |
| AP (any/none) | NK.49H-.Sp | Innate_lymphocyte | 1.75E-08 | 1.94E-08 | 0.184348236 |
| Arteriolosclerosis | NK.49H-.Sp | Innate_lymphocyte | -4.44E-09 | 2.64E-08 | 0.566749306 |
| Arteriolosclerosis (any | NK.49H-.Sp | Innate_lymphocyte | -2.71E-08 | 2.76E-08 | 0.837213192 |
| Atherosclerosis | NK.49H-.Sp | Innate_lymphocyte | 6.11E-08 | 2.51E-08 | 0.007530589 |
| Atherosclerosis (any/r | NK.49H-.Sp | Innate_lymphocyte | 5.13E-08 | 2.54E-08 | 0.02178106 |
| B Score | NK.49H-.Sp | Innate_lymphocyte | 4.33E-09 | 1.89E-08 | 0.409639338 |
| CAA | NK.49H-.Sp | Innate_lymphocyte | 7.23E-09 | 2.60E-08 | 0.390617716 |
| CAA (any/none) | NK.49H-.Sp | Innate_lymphocyte | 2.60E-08 | 2.39E-08 | 0.137568132 |
| CERAD (C Score) | NK.49H-.Sp | Innate_lymphocyte | 3.64E-08 | 1.81E-08 | 0.021881524 |
| Infarcts/lacunes | NK.49H-.Sp | Innate_lymphocyte | -2.76E-08 | 2.40E-08 | 0.87542503 |
| PD Braak | NK.49H-.Sp | Innate_lymphocyte | -4.42E-08 | 2.00E-08 | 0.986419599 |
| PD Braak (any/none) | NK.49H-.Sp | Innate_lymphocyte | -3.64E-08 | 1.91E-08 | 0.971792042 |
| TDP-43 (any/none) | NK.49H-.Sp | Innate_lymphocyte | -7.41E-08 | 6.12E-08 | 0.886808299 |
| Thal Phase | NK.49H-.Sp | Innate_lymphocyte | -1.41E-07 | 6.62E-08 | 0.983178296 |
| WMR (any/none) | NK.49H-.Sp | Innate_lymphocyte | -1.06E-08 | 3.28E-08 | 0.627066044 |
| #N/A | NK.49H-.Sp | Innate_lymphocyte | -1.85E-08 | 4.49E-08 | 0.660143514 |
| AD Braak | NK.49H+.Sp | Innate_lymphocyte | 2.06E-08 | 2.01E-08 | 0.151766527 |
| ADNC Score | NK.49H+.Sp | Innate_lymphocyte | -1.56E-07 | 5.35E-08 | 0.998260364 |
| AP (any/none) | NK.49H+.Sp | Innate_lymphocyte | -2.21E-08 | 2.05E-08 | 0.85959798 |
| Arteriolosclerosis | NK.49H+.Sp | Innate_lymphocyte | -5.71E-09 | 2.72E-08 | 0.58311859 |
| Arteriolosclerosis (any | NK.49H+.Sp | Innate_lymphocyte | -1.68E-08 | 3.00E-08 | 0.711824842 |
| Atherosclerosis | NK.49H+.Sp | Innate_lymphocyte | 3.85E-08 | 2.57E-08 | 0.066778013 |
| Atherosclerosis (any/r | NK.49H+.Sp | Innate_lymphocyte | 2.68E-09 | 2.43E-08 | 0.455936263 |
| B Score | NK.49H+.Sp | Innate_lymphocyte | 5.29E-09 | 2.05E-08 | 0.398059901 |
| CAA | NK.49H+.Sp | Innate_lymphocyte | -1.58E-08 | 2.51E-08 | 0.735061143 |
| CAA (any/none) | NK.49H+.Sp | Innate_lymphocyte | 9.77E-09 | 2.47E-08 | 0.346326822 |

| | | | | | |
|---|---|---|---|---|---|
| CERAD (C Score) | NK.49H+.Sp | Innate_lymphocyte | 1.53E-08 | 1.80E-08 | 0.197391024 |
| Infarcts/lacunes | NK.49H+.Sp | Innate_lymphocyte | -2.04E-08 | 2.44E-08 | 0.797733528 |
| PD Braak | NK.49H+.Sp | Innate_lymphocyte | -2.14E-08 | 2.02E-08 | 0.855522936 |
| PD Braak (any/none) | NK.49H+.Sp | Innate_lymphocyte | -1.03E-08 | 1.93E-08 | 0.702400542 |
| TDP-43 (any/none) | NK.49H+.Sp | Innate_lymphocyte | 2.10E-08 | 6.75E-08 | 0.377554565 |
| Thal Phase | NK.49H+.Sp | Innate_lymphocyte | -1.36E-07 | 7.00E-08 | 0.973925639 |
| WMR (any/none) | NK.49H+.Sp | Innate_lymphocyte | -1.18E-08 | 3.48E-08 | 0.632632473 |
| #N/A | NK.49H+.Sp | Innate_lymphocyte | -2.04E-08 | 4.47E-08 | 0.675864644 |
| AD Braak | NK.b2m-.Sp | Innate_lymphocyte | 4.33E-10 | 2.09E-08 | 0.491716306 |
| ADNC Score | NK.b2m-.Sp | Innate_lymphocyte | -8.94E-08 | 5.14E-08 | 0.959120469 |
| AP (any/none) | NK.b2m-.Sp | Innate_lymphocyte | -9.68E-09 | 1.99E-08 | 0.686885201 |
| Arteriolosclerosis | NK.b2m-.Sp | Innate_lymphocyte | 4.95E-09 | 2.66E-08 | 0.426212073 |
| Arteriolosclerosis (any | NK.b2m-.Sp | Innate_lymphocyte | -6.52E-10 | 2.99E-08 | 0.508697712 |
| Atherosclerosis | NK.b2m-.Sp | Innate_lymphocyte | 5.88E-08 | 2.39E-08 | 0.006834551 |
| Atherosclerosis (any/r | NK.b2m-.Sp | Innate_lymphocyte | 5.54E-08 | 2.44E-08 | 0.011688677 |
| B Score | NK.b2m-.Sp | Innate_lymphocyte | 8.93E-09 | 1.94E-08 | 0.322404483 |
| CAA | NK.b2m-.Sp | Innate_lymphocyte | -7.21E-09 | 2.45E-08 | 0.615597003 |
| CAA (any/none) | NK.b2m-.Sp | Innate_lymphocyte | -1.44E-08 | 2.40E-08 | 0.725858933 |
| CERAD (C Score) | NK.b2m-.Sp | Innate_lymphocyte | 1.10E-08 | 1.84E-08 | 0.275349991 |
| Infarcts/lacunes | NK.b2m-.Sp | Innate_lymphocyte | -3.62E-08 | 2.50E-08 | 0.92565227 |
| PD Braak | NK.b2m-.Sp | Innate_lymphocyte | -4.38E-08 | 1.78E-08 | 0.993168929 |
| PD Braak (any/none) | NK.b2m-.Sp | Innate_lymphocyte | -2.90E-08 | 1.61E-08 | 0.964014416 |
| TDP-43 (any/none) | NK.b2m-.Sp | Innate_lymphocyte | -2.99E-08 | 5.80E-08 | 0.697190882 |
| Thal Phase | NK.b2m-.Sp | Innate_lymphocyte | -1.62E-07 | 6.77E-08 | 0.991514867 |
| WMR (any/none) | NK.b2m-.Sp | Innate_lymphocyte | -3.93E-08 | 3.12E-08 | 0.895692096 |
| #N/A | NK.b2m-.Sp | Innate_lymphocyte | -1.76E-08 | 4.61E-08 | 0.649050647 |
| AD Braak | NK.CD127-.SI | Innate_lymphocyte | -1.27E-08 | 2.00E-08 | 0.737249331 |
| ADNC Score | NK.CD127-.SI | Innate_lymphocyte | 3.35E-08 | 5.90E-08 | 0.285214474 |
| AP (any/none) | NK.CD127-.SI | Innate_lymphocyte | -6.87E-09 | 2.06E-08 | 0.630378256 |
| Arteriolosclerosis | NK.CD127-.SI | Innate_lymphocyte | 5.94E-08 | 2.60E-08 | 0.011173805 |
| Arteriolosclerosis (any | NK.CD127-.SI | Innate_lymphocyte | 4.39E-08 | 3.00E-08 | 0.071511395 |
| Atherosclerosis | NK.CD127-.SI | Innate_lymphocyte | -3.21E-08 | 2.05E-08 | 0.941092227 |
| Atherosclerosis (any/r | NK.CD127-.SI | Innate_lymphocyte | 6.91E-09 | 2.22E-08 | 0.377919988 |
| B Score | NK.CD127-.SI | Innate_lymphocyte | 1.04E-08 | 1.91E-08 | 0.293443133 |

| | | | | | |
|---|---|---|---|---|---|
| CAA | NK.CD127-.SI | Innate_lymphocyte | 1.62E-08 | 2.22E-08 | 0.232458247 |
| CAA (any/none) | NK.CD127-.SI | Innate_lymphocyte | 5.65E-09 | 2.16E-08 | 0.396895659 |
| CERAD (C Score) | NK.CD127-.SI | Innate_lymphocyte | 2.04E-08 | 1.87E-08 | 0.136550433 |
| Infarcts/lacunes | NK.CD127-.SI | Innate_lymphocyte | -4.27E-08 | 2.20E-08 | 0.973941227 |
| PD Braak | NK.CD127-.SI | Innate_lymphocyte | 1.77E-08 | 2.31E-08 | 0.221674241 |
| PD Braak (any/none) | NK.CD127-.SI | Innate_lymphocyte | 1.70E-08 | 2.23E-08 | 0.223081374 |
| TDP-43 (any/none) | NK.CD127-.SI | Innate_lymphocyte | 9.85E-08 | 5.80E-08 | 0.044627739 |
| Thal Phase | NK.CD127-.SI | Innate_lymphocyte | -9.15E-08 | 7.45E-08 | 0.890105435 |
| WMR (any/none) | NK.CD127-.SI | Innate_lymphocyte | 2.53E-08 | 3.60E-08 | 0.241023272 |
| #N/A | NK.CD127-.SI | Innate_lymphocyte | 1.02E-08 | 4.27E-08 | 0.405750403 |
| AD Braak | NK.CD127-.Sp | Innate_lymphocyte | -2.38E-08 | 1.71E-08 | 0.917773216 |
| ADNC Score | NK.CD127-.Sp | Innate_lymphocyte | -1.49E-07 | 4.99E-08 | 0.998587617 |
| AP (any/none) | NK.CD127-.Sp | Innate_lymphocyte | -2.89E-08 | 1.99E-08 | 0.926957149 |
| Arteriolosclerosis | NK.CD127-.Sp | Innate_lymphocyte | 4.10E-08 | 2.64E-08 | 0.060045131 |
| Arteriolosclerosis (any | NK.CD127-.Sp | Innate_lymphocyte | 1.02E-08 | 2.71E-08 | 0.353643179 |
| Atherosclerosis | NK.CD127-.Sp | Innate_lymphocyte | 1.41E-09 | 2.09E-08 | 0.473057061 |
| Atherosclerosis (any/r | NK.CD127-.Sp | Innate_lymphocyte | 2.55E-08 | 2.15E-08 | 0.117957616 |
| B Score | NK.CD127-.Sp | Innate_lymphocyte | -1.97E-08 | 1.76E-08 | 0.868462452 |
| CAA | NK.CD127-.Sp | Innate_lymphocyte | -3.32E-08 | 1.88E-08 | 0.961199453 |
| CAA (any/none) | NK.CD127-.Sp | Innate_lymphocyte | -1.68E-08 | 2.07E-08 | 0.791261123 |
| CERAD (C Score) | NK.CD127-.Sp | Innate_lymphocyte | -1.49E-08 | 1.68E-08 | 0.812772796 |
| Infarcts/lacunes | NK.CD127-.Sp | Innate_lymphocyte | -2.10E-08 | 2.37E-08 | 0.812287753 |
| PD Braak | NK.CD127-.Sp | Innate_lymphocyte | 7.44E-10 | 2.24E-08 | 0.486729724 |
| PD Braak (any/none) | NK.CD127-.Sp | Innate_lymphocyte | 8.02E-10 | 2.21E-08 | 0.485554244 |
| TDP-43 (any/none) | NK.CD127-.Sp | Innate_lymphocyte | 1.54E-08 | 5.26E-08 | 0.384621567 |
| Thal Phase | NK.CD127-.Sp | Innate_lymphocyte | -1.66E-07 | 6.00E-08 | 0.997224477 |
| WMR (any/none) | NK.CD127-.Sp | Innate_lymphocyte | 3.70E-08 | 3.42E-08 | 0.139434864 |
| #N/A | NK.CD127-.Sp | Innate_lymphocyte | -5.79E-08 | 4.04E-08 | 0.923873339 |
| AD Braak | NK.CD49b+.Lv | Innate_lymphocyte | -3.33E-08 | 1.86E-08 | 0.963564404 |
| ADNC Score | NK.CD49b+.Lv | Innate_lymphocyte | -8.63E-08 | 5.47E-08 | 0.942670977 |
| AP (any/none) | NK.CD49b+.Lv | Innate_lymphocyte | -4.59E-08 | 1.96E-08 | 0.990422717 |
| Arteriolosclerosis | NK.CD49b+.Lv | Innate_lymphocyte | 4.54E-08 | 2.78E-08 | 0.051456455 |
| Arteriolosclerosis (any | NK.CD49b+.Lv | Innate_lymphocyte | 1.97E-09 | 2.97E-08 | 0.473487864 |
| Atherosclerosis | NK.CD49b+.Lv | Innate_lymphocyte | -8.75E-09 | 2.15E-08 | 0.657804043 |

| | | | | | |
|---|---|---|---|---|---|
| Atherosclerosis (any/r | NK.CD49b+.Lv | Innate_lymphocyte | -9.71E-10 | 2.05E-08 | 0.518895647 |
| B Score | NK.CD49b+.Lv | Innate_lymphocyte | -2.72E-08 | 1.80E-08 | 0.934450874 |
| CAA | NK.CD49b+.Lv | Innate_lymphocyte | -7.71E-09 | 1.82E-08 | 0.664254463 |
| CAA (any/none) | NK.CD49b+.Lv | Innate_lymphocyte | -5.14E-09 | 2.08E-08 | 0.597502244 |
| CERAD (C Score) | NK.CD49b+.Lv | Innate_lymphocyte | -1.56E-08 | 1.92E-08 | 0.79234495 |
| Infarcts/lacunes | NK.CD49b+.Lv | Innate_lymphocyte | -1.81E-08 | 2.51E-08 | 0.76430506 |
| PD Braak | NK.CD49b+.Lv | Innate_lymphocyte | 7.98E-09 | 2.30E-08 | 0.364556126 |
| PD Braak (any/none) | NK.CD49b+.Lv | Innate_lymphocyte | 1.30E-08 | 2.19E-08 | 0.277168638 |
| TDP-43 (any/none) | NK.CD49b+.Lv | Innate_lymphocyte | 5.21E-08 | 5.43E-08 | 0.16887724 |
| Thal Phase | NK.CD49b+.Lv | Innate_lymphocyte | -9.57E-08 | 6.74E-08 | 0.92216208 |
| WMR (any/none) | NK.CD49b+.Lv | Innate_lymphocyte | 6.53E-08 | 3.62E-08 | 0.035587236 |
| #N/A | NK.CD49b+.Lv | Innate_lymphocyte | -6.56E-08 | 4.07E-08 | 0.946328702 |
| AD Braak | NK.DAP10-.Sp | Innate_lymphocyte | 8.39E-10 | 2.11E-08 | 0.484121515 |
| ADNC Score | NK.DAP10-.Sp | Innate_lymphocyte | -1.63E-07 | 5.21E-08 | 0.99910224 |
| AP (any/none) | NK.DAP10-.Sp | Innate_lymphocyte | -2.32E-08 | 1.91E-08 | 0.888359393 |
| Arteriolosclerosis | NK.DAP10-.Sp | Innate_lymphocyte | 5.15E-08 | 2.96E-08 | 0.040927 |
| Arteriolosclerosis (any | NK.DAP10-.Sp | Innate_lymphocyte | 3.24E-08 | 3.10E-08 | 0.147575527 |
| Atherosclerosis | NK.DAP10-.Sp | Innate_lymphocyte | -1.45E-09 | 2.54E-08 | 0.522799035 |
| Atherosclerosis (any/r | NK.DAP10-.Sp | Innate_lymphocyte | 1.37E-09 | 2.42E-08 | 0.477469403 |
| B Score | NK.DAP10-.Sp | Innate_lymphocyte | 2.19E-09 | 1.86E-08 | 0.453136281 |
| CAA | NK.DAP10-.Sp | Innate_lymphocyte | 1.84E-08 | 2.51E-08 | 0.231455711 |
| CAA (any/none) | NK.DAP10-.Sp | Innate_lymphocyte | -5.15E-09 | 2.31E-08 | 0.58809548 |
| CERAD (C Score) | NK.DAP10-.Sp | Innate_lymphocyte | 1.83E-08 | 1.98E-08 | 0.177519719 |
| Infarcts/lacunes | NK.DAP10-.Sp | Innate_lymphocyte | -5.06E-08 | 2.36E-08 | 0.984120597 |
| PD Braak | NK.DAP10-.Sp | Innate_lymphocyte | -2.91E-08 | 2.01E-08 | 0.92646302 |
| PD Braak (any/none) | NK.DAP10-.Sp | Innate_lymphocyte | -2.01E-08 | 1.94E-08 | 0.850238071 |
| TDP-43 (any/none) | NK.DAP10-.Sp | Innate_lymphocyte | 2.12E-09 | 6.09E-08 | 0.486140825 |
| Thal Phase | NK.DAP10-.Sp | Innate_lymphocyte | -2.06E-07 | 6.86E-08 | 0.998643698 |
| WMR (any/none) | NK.DAP10-.Sp | Innate_lymphocyte | 2.27E-08 | 3.48E-08 | 0.25722194 |
| #N/A | NK.DAP10-.Sp | Innate_lymphocyte | 1.46E-08 | 4.64E-08 | 0.376016038 |
| AD Braak | NK.DAP12-.Sp | Innate_lymphocyte | -8.76E-09 | 1.99E-08 | 0.670270178 |
| ADNC Score | NK.DAP12-.Sp | Innate_lymphocyte | -1.16E-07 | 5.36E-08 | 0.984606911 |
| AP (any/none) | NK.DAP12-.Sp | Innate_lymphocyte | 9.32E-09 | 1.97E-08 | 0.317630323 |
| Arteriolosclerosis | NK.DAP12-.Sp | Innate_lymphocyte | 1.42E-09 | 2.58E-08 | 0.478101582 |

| | | | | | |
|---|---|---|---|---|---|
| Arteriolosclerosis (any | NK.DAP12-.Sp | Innate_lymphocyte | -1.42E-08 | 2.57E-08 | 0.709946563 |
| Atherosclerosis | NK.DAP12-.Sp | Innate_lymphocyte | 2.80E-08 | 2.40E-08 | 0.121302928 |
| Atherosclerosis (any/r | NK.DAP12-.Sp | Innate_lymphocyte | 5.56E-09 | 2.36E-08 | 0.406945813 |
| B Score | NK.DAP12-.Sp | Innate_lymphocyte | 1.02E-08 | 1.84E-08 | 0.289674027 |
| CAA | NK.DAP12-.Sp | Innate_lymphocyte | -2.61E-09 | 2.19E-08 | 0.547422603 |
| CAA (any/none) | NK.DAP12-.Sp | Innate_lymphocyte | 1.87E-09 | 2.28E-08 | 0.467333615 |
| CERAD (C Score) | NK.DAP12-.Sp | Innate_lymphocyte | 2.09E-08 | 1.75E-08 | 0.116535622 |
| Infarcts/lacunes | NK.DAP12-.Sp | Innate_lymphocyte | -1.90E-08 | 2.30E-08 | 0.795716165 |
| PD Braak | NK.DAP12-.Sp | Innate_lymphocyte | -4.78E-08 | 1.82E-08 | 0.995718035 |
| PD Braak (any/none) | NK.DAP12-.Sp | Innate_lymphocyte | -4.07E-08 | 1.83E-08 | 0.986733234 |
| TDP-43 (any/none) | NK.DAP12-.Sp | Innate_lymphocyte | -9.22E-08 | 5.64E-08 | 0.948831005 |
| Thal Phase | NK.DAP12-.Sp | Innate_lymphocyte | -1.74E-07 | 6.94E-08 | 0.993948031 |
| WMR (any/none) | NK.DAP12-.Sp | Innate_lymphocyte | -5.58E-08 | 3.31E-08 | 0.953962922 |
| #N/A | NK.DAP12-.Sp | Innate_lymphocyte | -2.50E-08 | 4.31E-08 | 0.718781625 |
| AD Braak | NK.H+.MCMV1.Sp | Innate_lymphocyte | 3.55E-09 | 2.01E-08 | 0.429793283 |
| ADNC Score | NK.H+.MCMV1.Sp | Innate_lymphocyte | -2.45E-08 | 6.96E-08 | 0.637623275 |
| AP (any/none) | NK.H+.MCMV1.Sp | Innate_lymphocyte | 9.09E-09 | 2.44E-08 | 0.354944561 |
| Arteriolosclerosis | NK.H+.MCMV1.Sp | Innate_lymphocyte | -1.81E-08 | 2.92E-08 | 0.73190097 |
| Arteriolosclerosis (any | NK.H+.MCMV1.Sp | Innate_lymphocyte | -2.14E-08 | 3.14E-08 | 0.75284498 |
| Atherosclerosis | NK.H+.MCMV1.Sp | Innate_lymphocyte | 3.10E-08 | 2.55E-08 | 0.111663504 |
| Atherosclerosis (any/r | NK.H+.MCMV1.Sp | Innate_lymphocyte | 1.90E-08 | 2.54E-08 | 0.227577283 |
| B Score | NK.H+.MCMV1.Sp | Innate_lymphocyte | 4.83E-09 | 2.02E-08 | 0.405719911 |
| CAA | NK.H+.MCMV1.Sp | Innate_lymphocyte | 4.98E-08 | 3.01E-08 | 0.049405563 |
| CAA (any/none) | NK.H+.MCMV1.Sp | Innate_lymphocyte | 1.44E-08 | 2.82E-08 | 0.304946373 |
| CERAD (C Score) | NK.H+.MCMV1.Sp | Innate_lymphocyte | 2.71E-08 | 2.45E-08 | 0.134612753 |
| Infarcts/lacunes | NK.H+.MCMV1.Sp | Innate_lymphocyte | -8.58E-09 | 2.53E-08 | 0.632781209 |
| PD Braak | NK.H+.MCMV1.Sp | Innate_lymphocyte | -4.54E-08 | 2.12E-08 | 0.983995804 |
| PD Braak (any/none) | NK.H+.MCMV1.Sp | Innate_lymphocyte | -4.47E-08 | 2.03E-08 | 0.986047821 |
| TDP-43 (any/none) | NK.H+.MCMV1.Sp | Innate_lymphocyte | -7.97E-08 | 5.54E-08 | 0.925036581 |
| Thal Phase | NK.H+.MCMV1.Sp | Innate_lymphocyte | -2.20E-08 | 7.46E-08 | 0.615926603 |
| WMR (any/none) | NK.H+.MCMV1.Sp | Innate_lymphocyte | 4.78E-08 | 3.61E-08 | 0.092541487 |
| #N/A | NK.H+.MCMV1.Sp | Innate_lymphocyte | -4.16E-08 | 4.46E-08 | 0.824533621 |
| AD Braak | NK.H+.MCMV7.Sp | Innate_lymphocyte | 5.61E-09 | 1.99E-08 | 0.389133814 |
| ADNC Score | NK.H+.MCMV7.Sp | Innate_lymphocyte | -7.56E-08 | 6.47E-08 | 0.87872481 |

| | | | | | |
|---|---|---|---|---|---|
| AP (any/none) | NK.H+.MCMV7.Sp | Innate_lymphocyte | -1.76E-08 | 1.98E-08 | 0.81330858 |
| Arteriolosclerosis | NK.H+.MCMV7.Sp | Innate_lymphocyte | 4.33E-08 | 2.45E-08 | 0.03864718 |
| Arteriolosclerosis (any | NK.H+.MCMV7.Sp | Innate_lymphocyte | -2.87E-08 | 2.86E-08 | 0.842496958 |
| Atherosclerosis | NK.H+.MCMV7.Sp | Innate_lymphocyte | 6.63E-08 | 2.35E-08 | 0.002392706 |
| Atherosclerosis (any/r | NK.H+.MCMV7.Sp | Innate_lymphocyte | 3.20E-08 | 2.30E-08 | 0.081641322 |
| B Score | NK.H+.MCMV7.Sp | Innate_lymphocyte | 1.82E-08 | 1.90E-08 | 0.169629736 |
| CAA | NK.H+.MCMV7.Sp | Innate_lymphocyte | 1.17E-08 | 2.52E-08 | 0.320697551 |
| CAA (any/none) | NK.H+.MCMV7.Sp | Innate_lymphocyte | 6.33E-09 | 2.41E-08 | 0.396213706 |
| CERAD (C Score) | NK.H+.MCMV7.Sp | Innate_lymphocyte | 1.81E-08 | 1.87E-08 | 0.166097795 |
| Infarcts/lacunes | NK.H+.MCMV7.Sp | Innate_lymphocyte | 1.35E-08 | 2.30E-08 | 0.278187782 |
| PD Braak | NK.H+.MCMV7.Sp | Innate_lymphocyte | -2.47E-08 | 1.88E-08 | 0.90555793 |
| PD Braak (any/none) | NK.H+.MCMV7.Sp | Innate_lymphocyte | -1.97E-08 | 1.75E-08 | 0.869746127 |
| TDP-43 (any/none) | NK.H+.MCMV7.Sp | Innate_lymphocyte | -7.16E-08 | 5.26E-08 | 0.913314475 |
| Thal Phase | NK.H+.MCMV7.Sp | Innate_lymphocyte | -1.45E-07 | 7.41E-08 | 0.974458709 |
| WMR (any/none) | NK.H+.MCMV7.Sp | Innate_lymphocyte | -3.00E-08 | 3.30E-08 | 0.818220334 |
| #N/A | NK.H+.MCMV7.Sp | Innate_lymphocyte | -6.60E-08 | 4.42E-08 | 0.932292644 |
| AD Braak | NK.MCMV1.Sp | Innate_lymphocyte | 8.24E-09 | 1.88E-08 | 0.330325279 |
| ADNC Score | NK.MCMV1.Sp | Innate_lymphocyte | -8.68E-08 | 5.78E-08 | 0.933430988 |
| AP (any/none) | NK.MCMV1.Sp | Innate_lymphocyte | 1.08E-08 | 2.18E-08 | 0.309952145 |
| Arteriolosclerosis | NK.MCMV1.Sp | Innate_lymphocyte | 3.14E-08 | 2.86E-08 | 0.135865297 |
| Arteriolosclerosis (any | NK.MCMV1.Sp | Innate_lymphocyte | 2.39E-08 | 3.21E-08 | 0.228370172 |
| Atherosclerosis | NK.MCMV1.Sp | Innate_lymphocyte | 1.70E-09 | 2.63E-08 | 0.474232532 |
| Atherosclerosis (any/r | NK.MCMV1.Sp | Innate_lymphocyte | 2.32E-08 | 2.75E-08 | 0.199374902 |
| B Score | NK.MCMV1.Sp | Innate_lymphocyte | 1.69E-09 | 1.83E-08 | 0.463130713 |
| CAA | NK.MCMV1.Sp | Innate_lymphocyte | 8.33E-09 | 2.54E-08 | 0.371516926 |
| CAA (any/none) | NK.MCMV1.Sp | Innate_lymphocyte | -8.50E-09 | 2.49E-08 | 0.633782457 |
| CERAD (C Score) | NK.MCMV1.Sp | Innate_lymphocyte | 2.96E-08 | 2.18E-08 | 0.086946021 |
| Infarcts/lacunes | NK.MCMV1.Sp | Innate_lymphocyte | 6.62E-09 | 2.39E-08 | 0.390758309 |
| PD Braak | NK.MCMV1.Sp | Innate_lymphocyte | -4.85E-08 | 1.92E-08 | 0.994182428 |
| PD Braak (any/none) | NK.MCMV1.Sp | Innate_lymphocyte | -3.89E-08 | 1.82E-08 | 0.983968564 |
| TDP-43 (any/none) | NK.MCMV1.Sp | Innate_lymphocyte | -7.12E-08 | 5.25E-08 | 0.912488363 |
| Thal Phase | NK.MCMV1.Sp | Innate_lymphocyte | -1.58E-07 | 7.83E-08 | 0.978183166 |
| WMR (any/none) | NK.MCMV1.Sp | Innate_lymphocyte | 4.04E-08 | 3.37E-08 | 0.115120034 |
| #N/A | NK.MCMV1.Sp | Innate_lymphocyte | -3.62E-09 | 4.43E-08 | 0.532524527 |

| | | | | | |
|---|---|---|---|---|---|
| AD Braak | NK.MCMV7.Sp | Innate_lymphocyte | -9.19E-09 | 1.83E-08 | 0.691786802 |
| ADNC Score | NK.MCMV7.Sp | Innate_lymphocyte | -6.74E-08 | 5.68E-08 | 0.882512565 |
| AP (any/none) | NK.MCMV7.Sp | Innate_lymphocyte | -1.29E-08 | 2.04E-08 | 0.736259366 |
| Arteriolosclerosis | NK.MCMV7.Sp | Innate_lymphocyte | 1.75E-08 | 2.64E-08 | 0.253855359 |
| Arteriolosclerosis (any | NK.MCMV7.Sp | Innate_lymphocyte | 1.39E-08 | 3.04E-08 | 0.324111051 |
| Atherosclerosis | NK.MCMV7.Sp | Innate_lymphocyte | 2.67E-08 | 2.39E-08 | 0.13191528 |
| Atherosclerosis (any/r | NK.MCMV7.Sp | Innate_lymphocyte | -4.57E-09 | 2.60E-08 | 0.569777424 |
| B Score | NK.MCMV7.Sp | Innate_lymphocyte | -3.19E-09 | 1.73E-08 | 0.573033048 |
| CAA | NK.MCMV7.Sp | Innate_lymphocyte | 3.99E-10 | 2.37E-08 | 0.493283687 |
| CAA (any/none) | NK.MCMV7.Sp | Innate_lymphocyte | 1.79E-08 | 2.25E-08 | 0.213404849 |
| CERAD (C Score) | NK.MCMV7.Sp | Innate_lymphocyte | 1.54E-08 | 1.90E-08 | 0.208874947 |
| Infarcts/lacunes | NK.MCMV7.Sp | Innate_lymphocyte | -1.57E-08 | 2.41E-08 | 0.742511012 |
| PD Braak | NK.MCMV7.Sp | Innate_lymphocyte | -2.23E-08 | 1.91E-08 | 0.878733612 |
| PD Braak (any/none) | NK.MCMV7.Sp | Innate_lymphocyte | -1.71E-08 | 1.77E-08 | 0.831895967 |
| TDP-43 (any/none) | NK.MCMV7.Sp | Innate_lymphocyte | -2.67E-08 | 5.51E-08 | 0.685979444 |
| Thal Phase | NK.MCMV7.Sp | Innate_lymphocyte | -8.67E-08 | 7.26E-08 | 0.883782342 |
| WMR (any/none) | NK.MCMV7.Sp | Innate_lymphocyte | -1.36E-08 | 3.24E-08 | 0.662569509 |
| #N/A | NK.MCMV7.Sp | Innate_lymphocyte | 3.13E-08 | 4.20E-08 | 0.228328032 |
| AD Braak | NK.Sp | Innate_lymphocyte | 2.54E-09 | 1.94E-08 | 0.44773341 |
| ADNC Score | NK.Sp | Innate_lymphocyte | -7.86E-08 | 6.01E-08 | 0.904495183 |
| AP (any/none) | NK.Sp | Innate_lymphocyte | -6.16E-09 | 2.09E-08 | 0.615871095 |
| Arteriolosclerosis | NK.Sp | Innate_lymphocyte | -1.30E-08 | 2.71E-08 | 0.683795665 |
| Arteriolosclerosis (any | NK.Sp | Innate_lymphocyte | -1.60E-08 | 2.85E-08 | 0.713321435 |
| Atherosclerosis | NK.Sp | Innate_lymphocyte | 2.91E-08 | 2.37E-08 | 0.110189235 |
| Atherosclerosis (any/r | NK.Sp | Innate_lymphocyte | 4.65E-09 | 2.72E-08 | 0.432100194 |
| B Score | NK.Sp | Innate_lymphocyte | 3.22E-09 | 1.83E-08 | 0.430022769 |
| CAA | NK.Sp | Innate_lymphocyte | 2.81E-08 | 2.46E-08 | 0.126094127 |
| CAA (any/none) | NK.Sp | Innate_lymphocyte | -6.95E-09 | 2.37E-08 | 0.615499511 |
| CERAD (C Score) | NK.Sp | Innate_lymphocyte | 5.61E-09 | 1.83E-08 | 0.379864255 |
| Infarcts/lacunes | NK.Sp | Innate_lymphocyte | -1.68E-08 | 2.50E-08 | 0.749120371 |
| PD Braak | NK.Sp | Innate_lymphocyte | -1.08E-08 | 2.06E-08 | 0.700677027 |
| PD Braak (any/none) | NK.Sp | Innate_lymphocyte | -1.43E-08 | 2.05E-08 | 0.757030599 |
| TDP-43 (any/none) | NK.Sp | Innate_lymphocyte | -3.41E-08 | 5.34E-08 | 0.738579937 |
| Thal Phase | NK.Sp | Innate_lymphocyte | -1.04E-07 | 7.65E-08 | 0.913482781 |

| | | | | | |
|---|---|---|---|---|---|
| WMR (any/none) | NK.Sp | Innate_lymphocyte | -1.92E-09 | 3.37E-08 | 0.522723604 |
| #N/A | NK.Sp | Innate_lymphocyte | 5.86E-08 | 4.85E-08 | 0.113698227 |
| AD Braak | B.FrE.BM | Myeloid | 4.70E-08 | 2.06E-08 | 0.011168878 |
| ADNC Score | B.FrE.BM | Myeloid | -6.66E-08 | 5.41E-08 | 0.890708417 |
| AP (any/none) | B.FrE.BM | Myeloid | 3.08E-09 | 2.24E-08 | 0.445331765 |
| Arteriolosclerosis | B.FrE.BM | Myeloid | -2.51E-08 | 2.67E-08 | 0.825776662 |
| Arteriolosclerosis (any | B.FrE.BM | Myeloid | 1.75E-08 | 3.18E-08 | 0.291131181 |
| Atherosclerosis | B.FrE.BM | Myeloid | 3.88E-08 | 2.31E-08 | 0.046828977 |
| Atherosclerosis (any/r | B.FrE.BM | Myeloid | 8.33E-09 | 2.23E-08 | 0.354340817 |
| B Score | B.FrE.BM | Myeloid | 5.27E-08 | 2.06E-08 | 0.005360351 |
| CAA | B.FrE.BM | Myeloid | -4.81E-08 | 2.06E-08 | 0.990198448 |
| CAA (any/none) | B.FrE.BM | Myeloid | -4.04E-08 | 2.01E-08 | 0.977658791 |
| CERAD (C Score) | B.FrE.BM | Myeloid | 3.64E-08 | 2.23E-08 | 0.051221401 |
| Infarcts/lacunes | B.FrE.BM | Myeloid | -2.39E-08 | 2.28E-08 | 0.852581229 |
| PD Braak | B.FrE.BM | Myeloid | -6.10E-10 | 2.13E-08 | 0.511417384 |
| PD Braak (any/none) | B.FrE.BM | Myeloid | -1.04E-08 | 2.06E-08 | 0.694003307 |
| TDP-43 (any/none) | B.FrE.BM | Myeloid | -7.03E-08 | 5.61E-08 | 0.895106795 |
| Thal Phase | B.FrE.BM | Myeloid | -8.38E-08 | 7.80E-08 | 0.858406535 |
| WMR (any/none) | B.FrE.BM | Myeloid | -2.64E-08 | 3.39E-08 | 0.782277587 |
| #N/A | B.FrE.BM | Myeloid | 5.55E-08 | 4.48E-08 | 0.107748403 |
| AD Braak | B.FrE.FL | Myeloid | 1.09E-08 | 1.91E-08 | 0.284547599 |
| ADNC Score | B.FrE.FL | Myeloid | -8.67E-08 | 5.49E-08 | 0.94264577 |
| AP (any/none) | B.FrE.FL | Myeloid | 2.29E-09 | 2.19E-08 | 0.458328308 |
| Arteriolosclerosis | B.FrE.FL | Myeloid | -1.97E-08 | 2.63E-08 | 0.773125109 |
| Arteriolosclerosis (any | B.FrE.FL | Myeloid | 3.45E-08 | 3.18E-08 | 0.138530693 |
| Atherosclerosis | B.FrE.FL | Myeloid | 1.60E-08 | 2.52E-08 | 0.263196801 |
| Atherosclerosis (any/r | B.FrE.FL | Myeloid | -9.54E-09 | 2.55E-08 | 0.645767772 |
| B Score | B.FrE.FL | Myeloid | 2.27E-08 | 1.92E-08 | 0.119453862 |
| CAA | B.FrE.FL | Myeloid | 6.17E-09 | 2.47E-08 | 0.401299737 |
| CAA (any/none) | B.FrE.FL | Myeloid | 7.38E-09 | 2.24E-08 | 0.370717103 |
| CERAD (C Score) | B.FrE.FL | Myeloid | 6.71E-09 | 2.18E-08 | 0.379087313 |
| Infarcts/lacunes | B.FrE.FL | Myeloid | -5.21E-08 | 2.18E-08 | 0.991658025 |
| PD Braak | B.FrE.FL | Myeloid | -1.60E-08 | 2.12E-08 | 0.775295607 |
| PD Braak (any/none) | B.FrE.FL | Myeloid | -2.40E-08 | 2.05E-08 | 0.878690443 |

| | | | | | |
|---|---|---|---|---|---|
| TDP-43 (any/none) | B.FrE.FL | Myeloid | -1.23E-07 | 5.29E-08 | 0.990213437 |
| Thal Phase | B.FrE.FL | Myeloid | -3.58E-08 | 7.30E-08 | 0.68793271 |
| WMR (any/none) | B.FrE.FL | Myeloid | -4.07E-09 | 3.58E-08 | 0.545238685 |
| #N/A | B.FrE.FL | Myeloid | 1.30E-08 | 4.21E-08 | 0.378388345 |
| AD Braak | DC.103+11b-.Lu | Myeloid | 1.32E-08 | 1.73E-08 | 0.222495925 |
| ADNC Score | DC.103+11b-.Lu | Myeloid | -3.42E-08 | 6.16E-08 | 0.710668767 |
| AP (any/none) | DC.103+11b-.Lu | Myeloid | 4.82E-09 | 2.18E-08 | 0.412324153 |
| Arteriolosclerosis | DC.103+11b-.Lu | Myeloid | -1.30E-09 | 2.82E-08 | 0.518323308 |
| Arteriolosclerosis (any | DC.103+11b-.Lu | Myeloid | 3.33E-08 | 2.82E-08 | 0.118220676 |
| Atherosclerosis | DC.103+11b-.Lu | Myeloid | -1.55E-08 | 2.36E-08 | 0.744303567 |
| Atherosclerosis (any/r | DC.103+11b-.Lu | Myeloid | -3.64E-09 | 2.12E-08 | 0.568198234 |
| B Score | DC.103+11b-.Lu | Myeloid | 1.67E-08 | 1.85E-08 | 0.182565771 |
| CAA | DC.103+11b-.Lu | Myeloid | -1.83E-08 | 2.39E-08 | 0.778476928 |
| CAA (any/none) | DC.103+11b-.Lu | Myeloid | -2.17E-08 | 2.46E-08 | 0.811749749 |
| CERAD (C Score) | DC.103+11b-.Lu | Myeloid | 2.13E-08 | 1.89E-08 | 0.129948651 |
| Infarcts/lacunes | DC.103+11b-.Lu | Myeloid | 9.41E-09 | 2.49E-08 | 0.352812832 |
| PD Braak | DC.103+11b-.Lu | Myeloid | 4.72E-09 | 2.03E-08 | 0.407916119 |
| PD Braak (any/none) | DC.103+11b-.Lu | Myeloid | 1.29E-08 | 1.92E-08 | 0.251135621 |
| TDP-43 (any/none) | DC.103+11b-.Lu | Myeloid | -2.09E-08 | 5.13E-08 | 0.658128413 |
| Thal Phase | DC.103+11b-.Lu | Myeloid | -5.51E-08 | 7.60E-08 | 0.765820988 |
| WMR (any/none) | DC.103+11b-.Lu | Myeloid | -6.17E-09 | 3.33E-08 | 0.5735453 |
| #N/A | DC.103+11b-.Lu | Myeloid | -3.15E-08 | 4.24E-08 | 0.771561249 |
| AD Braak | DC.103+11b-.Lv | Myeloid | 5.31E-09 | 1.65E-08 | 0.374204532 |
| ADNC Score | DC.103+11b-.Lv | Myeloid | 5.09E-08 | 6.52E-08 | 0.217550983 |
| AP (any/none) | DC.103+11b-.Lv | Myeloid | 1.61E-08 | 2.12E-08 | 0.223598261 |
| Arteriolosclerosis | DC.103+11b-.Lv | Myeloid | -6.71E-09 | 2.85E-08 | 0.593165354 |
| Arteriolosclerosis (any | DC.103+11b-.Lv | Myeloid | 2.04E-08 | 2.75E-08 | 0.229116228 |
| Atherosclerosis | DC.103+11b-.Lv | Myeloid | -1.10E-08 | 2.20E-08 | 0.690811405 |
| Atherosclerosis (any/r | DC.103+11b-.Lv | Myeloid | -7.87E-09 | 2.30E-08 | 0.6340888 |
| B Score | DC.103+11b-.Lv | Myeloid | 8.99E-09 | 1.73E-08 | 0.301360641 |
| CAA | DC.103+11b-.Lv | Myeloid | -1.57E-08 | 2.15E-08 | 0.767276423 |
| CAA (any/none) | DC.103+11b-.Lv | Myeloid | -1.84E-08 | 2.06E-08 | 0.81378663 |
| CERAD (C Score) | DC.103+11b-.Lv | Myeloid | 1.51E-08 | 1.90E-08 | 0.212742703 |
| Infarcts/lacunes | DC.103+11b-.Lv | Myeloid | -1.30E-08 | 2.36E-08 | 0.709375821 |

| | | | | | |
|---|---|---|---|---|---|
| PD Braak | DC.103+11b-.Lv | Myeloid | -1.83E-08 | 1.92E-08 | 0.82963451 |
| PD Braak (any/none) | DC.103+11b-.Lv | Myeloid | -1.63E-08 | 1.82E-08 | 0.814940855 |
| TDP-43 (any/none) | DC.103+11b-.Lv | Myeloid | -4.36E-08 | 5.07E-08 | 0.805210668 |
| Thal Phase | DC.103+11b-.Lv | Myeloid | -5.72E-08 | 7.28E-08 | 0.783740319 |
| WMR (any/none) | DC.103+11b-.Lv | Myeloid | -8.88E-09 | 3.44E-08 | 0.601818416 |
| #N/A | DC.103+11b-.Lv | Myeloid | -3.60E-08 | 3.94E-08 | 0.82002164 |
| AD Braak | DC.103-11b+24+.Lu | Myeloid | 1.85E-08 | 1.69E-08 | 0.137071501 |
| ADNC Score | DC.103-11b+24+.Lu | Myeloid | 2.89E-08 | 5.82E-08 | 0.30978762 |
| AP (any/none) | DC.103-11b+24+.Lu | Myeloid | 1.80E-08 | 2.21E-08 | 0.207734838 |
| Arteriolosclerosis | DC.103-11b+24+.Lu | Myeloid | 2.15E-08 | 2.84E-08 | 0.223852778 |
| Arteriolosclerosis (any | DC.103-11b+24+.Lu | Myeloid | 1.35E-08 | 2.72E-08 | 0.309348852 |
| Atherosclerosis | DC.103-11b+24+.Lu | Myeloid | -1.07E-08 | 2.34E-08 | 0.675531204 |
| Atherosclerosis (any/r | DC.103-11b+24+.Lu | Myeloid | -1.19E-09 | 2.22E-08 | 0.521311401 |
| B Score | DC.103-11b+24+.Lu | Myeloid | 9.91E-09 | 1.78E-08 | 0.289327728 |
| CAA | DC.103-11b+24+.Lu | Myeloid | 5.56E-11 | 2.10E-08 | 0.498942478 |
| CAA (any/none) | DC.103-11b+24+.Lu | Myeloid | -1.25E-08 | 2.06E-08 | 0.726937675 |
| CERAD (C Score) | DC.103-11b+24+.Lu | Myeloid | 1.79E-08 | 1.90E-08 | 0.172837752 |
| Infarcts/lacunes | DC.103-11b+24+.Lu | Myeloid | 2.61E-08 | 2.45E-08 | 0.143140063 |
| PD Braak | DC.103-11b+24+.Lu | Myeloid | 2.10E-08 | 2.03E-08 | 0.149727402 |
| PD Braak (any/none) | DC.103-11b+24+.Lu | Myeloid | 2.72E-08 | 1.96E-08 | 0.082535931 |
| TDP-43 (any/none) | DC.103-11b+24+.Lu | Myeloid | -5.66E-08 | 4.78E-08 | 0.882009024 |
| Thal Phase | DC.103-11b+24+.Lu | Myeloid | 1.32E-08 | 7.69E-08 | 0.43182731 |
| WMR (any/none) | DC.103-11b+24+.Lu | Myeloid | 2.94E-08 | 3.68E-08 | 0.212061431 |
| #N/A | DC.103-11b+24+.Lu | Myeloid | 1.24E-08 | 3.89E-08 | 0.37547033 |
| AD Braak | DC.103-11b+F4_80lo.Kd | Myeloid | 2.30E-08 | 1.88E-08 | 0.110470599 |
| ADNC Score | DC.103-11b+F4_80lo.Kd | Myeloid | 1.13E-07 | 6.62E-08 | 0.044623208 |
| AP (any/none) | DC.103-11b+F4_80lo.Kd | Myeloid | 2.63E-08 | 2.36E-08 | 0.132561617 |
| Arteriolosclerosis | DC.103-11b+F4_80lo.Kd | Myeloid | 4.57E-09 | 2.93E-08 | 0.438047553 |
| Arteriolosclerosis (any | DC.103-11b+F4_80lo.Kd | Myeloid | 2.80E-08 | 2.80E-08 | 0.158537455 |
| Atherosclerosis | DC.103-11b+F4_80lo.Kd | Myeloid | -4.22E-08 | 2.42E-08 | 0.959558019 |
| Atherosclerosis (any/r | DC.103-11b+F4_80lo.Kd | Myeloid | -2.15E-08 | 2.21E-08 | 0.834643801 |
| B Score | DC.103-11b+F4_80lo.Kd | Myeloid | 2.34E-08 | 2.07E-08 | 0.129211929 |
| CAA | DC.103-11b+F4_80lo.Kd | Myeloid | -1.33E-08 | 2.27E-08 | 0.720122867 |
| CAA (any/none) | DC.103-11b+F4_80lo.Kd | Myeloid | -5.07E-09 | 2.24E-08 | 0.589323785 |

| | | | | | |
|---|---|---|---|---|---|
| CERAD (C Score) | DC.103-11b+F4_80lo.Kd | Myeloid | 3.83E-08 | 2.10E-08 | 0.034367997 |
| Infarcts/lacunes | DC.103-11b+F4_80lo.Kd | Myeloid | 1.28E-08 | 2.35E-08 | 0.293004427 |
| PD Braak | DC.103-11b+F4_80lo.Kd | Myeloid | 6.54E-09 | 2.18E-08 | 0.381840574 |
| PD Braak (any/none) | DC.103-11b+F4_80lo.Kd | Myeloid | 8.29E-10 | 2.10E-08 | 0.484281611 |
| TDP-43 (any/none) | DC.103-11b+F4_80lo.Kd | Myeloid | -2.02E-08 | 5.68E-08 | 0.638998986 |
| Thal Phase | DC.103-11b+F4_80lo.Kd | Myeloid | 9.89E-08 | 8.10E-08 | 0.11122268 |
| WMR (any/none) | DC.103-11b+F4_80lo.Kd | Myeloid | 3.89E-09 | 3.84E-08 | 0.459702602 |
| #N/A | DC.103-11b+F4_80lo.Kd | Myeloid | -3.50E-08 | 4.26E-08 | 0.794560711 |
| AD Braak | DC.11b-.AT.v2 | Myeloid | 2.50E-08 | 1.75E-08 | 0.076773747 |
| ADNC Score | DC.11b-.AT.v2 | Myeloid | -3.79E-08 | 5.45E-08 | 0.756472618 |
| AP (any/none) | DC.11b-.AT.v2 | Myeloid | 1.97E-08 | 1.95E-08 | 0.155517058 |
| Arteriolosclerosis | DC.11b-.AT.v2 | Myeloid | -2.07E-08 | 2.84E-08 | 0.767375072 |
| Arteriolosclerosis (any | DC.11b-.AT.v2 | Myeloid | -2.31E-08 | 2.76E-08 | 0.798525614 |
| Atherosclerosis | DC.11b-.AT.v2 | Myeloid | -9.42E-09 | 2.09E-08 | 0.674106534 |
| Atherosclerosis (any/r | DC.11b-.AT.v2 | Myeloid | -4.81E-09 | 2.14E-08 | 0.589118571 |
| B Score | DC.11b-.AT.v2 | Myeloid | 2.83E-08 | 1.84E-08 | 0.061901812 |
| CAA | DC.11b-.AT.v2 | Myeloid | 9.75E-09 | 2.34E-08 | 0.338412493 |
| CAA (any/none) | DC.11b-.AT.v2 | Myeloid | 8.52E-09 | 2.18E-08 | 0.347696787 |
| CERAD (C Score) | DC.11b-.AT.v2 | Myeloid | 4.97E-09 | 1.73E-08 | 0.387081174 |
| Infarcts/lacunes | DC.11b-.AT.v2 | Myeloid | -8.92E-09 | 2.20E-08 | 0.657724191 |
| PD Braak | DC.11b-.AT.v2 | Myeloid | 1.56E-08 | 2.04E-08 | 0.221230788 |
| PD Braak (any/none) | DC.11b-.AT.v2 | Myeloid | 1.14E-08 | 1.97E-08 | 0.281212749 |
| TDP-43 (any/none) | DC.11b-.AT.v2 | Myeloid | -1.66E-08 | 5.08E-08 | 0.627907406 |
| Thal Phase | DC.11b-.AT.v2 | Myeloid | -1.25E-08 | 7.07E-08 | 0.570343927 |
| WMR (any/none) | DC.11b-.AT.v2 | Myeloid | -7.70E-10 | 3.28E-08 | 0.509366182 |
| #N/A | DC.11b-.AT.v2 | Myeloid | -1.58E-08 | 3.91E-08 | 0.657410592 |
| AD Braak | DC.11b+.AT.v2 | Myeloid | 7.32E-09 | 1.75E-08 | 0.337936353 |
| ADNC Score | DC.11b+.AT.v2 | Myeloid | -3.26E-08 | 5.12E-08 | 0.737809103 |
| AP (any/none) | DC.11b+.AT.v2 | Myeloid | -1.14E-08 | 2.03E-08 | 0.711585427 |
| Arteriolosclerosis | DC.11b+.AT.v2 | Myeloid | 9.58E-09 | 2.86E-08 | 0.368934936 |
| Arteriolosclerosis (any | DC.11b+.AT.v2 | Myeloid | 1.78E-08 | 2.70E-08 | 0.254900264 |
| Atherosclerosis | DC.11b+.AT.v2 | Myeloid | 8.28E-09 | 2.13E-08 | 0.348401733 |
| Atherosclerosis (any/r | DC.11b+.AT.v2 | Myeloid | -1.16E-08 | 2.19E-08 | 0.701305438 |
| B Score | DC.11b+.AT.v2 | Myeloid | 1.60E-08 | 1.79E-08 | 0.185442181 |

| | | | | | |
|---|---|---|---|---|---|
| CAA | DC.11b+.AT.v2 | Myeloid | -3.07E-08 | 2.26E-08 | 0.912413829 |
| CAA (any/none) | DC.11b+.AT.v2 | Myeloid | -2.48E-08 | 2.10E-08 | 0.880832295 |
| CERAD (C Score) | DC.11b+.AT.v2 | Myeloid | 1.70E-08 | 1.70E-08 | 0.158846009 |
| Infarcts/lacunes | DC.11b+.AT.v2 | Myeloid | 3.70E-08 | 2.30E-08 | 0.053836015 |
| PD Braak | DC.11b+.AT.v2 | Myeloid | 5.46E-08 | 2.04E-08 | 0.003695885 |
| PD Braak (any/none) | DC.11b+.AT.v2 | Myeloid | 4.87E-08 | 1.99E-08 | 0.007330106 |
| TDP-43 (any/none) | DC.11b+.AT.v2 | Myeloid | 6.30E-09 | 5.98E-08 | 0.458033675 |
| Thal Phase | DC.11b+.AT.v2 | Myeloid | -1.55E-07 | 6.36E-08 | 0.992661821 |
| WMR (any/none) | DC.11b+.AT.v2 | Myeloid | 3.21E-09 | 3.38E-08 | 0.462181057 |
| #N/A | DC.11b+.AT.v2 | Myeloid | 4.12E-08 | 4.53E-08 | 0.181410951 |
| AD Braak | DC.4+.MLN | Myeloid | 3.19E-09 | 1.73E-08 | 0.426784873 |
| ADNC Score | DC.4+.MLN | Myeloid | 3.79E-08 | 6.52E-08 | 0.280632514 |
| AP (any/none) | DC.4+.MLN | Myeloid | 3.36E-08 | 2.12E-08 | 0.05660147 |
| Arteriolosclerosis | DC.4+.MLN | Myeloid | 1.93E-08 | 2.72E-08 | 0.239133452 |
| Arteriolosclerosis (any | DC.4+.MLN | Myeloid | 3.63E-08 | 2.96E-08 | 0.109784149 |
| Atherosclerosis | DC.4+.MLN | Myeloid | -1.18E-08 | 2.25E-08 | 0.700627507 |
| Atherosclerosis (any/r | DC.4+.MLN | Myeloid | -4.57E-09 | 2.49E-08 | 0.572824698 |
| B Score | DC.4+.MLN | Myeloid | 1.08E-08 | 1.68E-08 | 0.260047178 |
| CAA | DC.4+.MLN | Myeloid | -2.06E-08 | 2.22E-08 | 0.823122988 |
| CAA (any/none) | DC.4+.MLN | Myeloid | -2.07E-08 | 2.10E-08 | 0.83755706 |
| CERAD (C Score) | DC.4+.MLN | Myeloid | 3.32E-08 | 1.88E-08 | 0.038899507 |
| Infarcts/lacunes | DC.4+.MLN | Myeloid | -3.66E-09 | 2.31E-08 | 0.562913354 |
| PD Braak | DC.4+.MLN | Myeloid | 1.79E-08 | 2.09E-08 | 0.195377742 |
| PD Braak (any/none) | DC.4+.MLN | Myeloid | 2.36E-08 | 2.01E-08 | 0.120953868 |
| TDP-43 (any/none) | DC.4+.MLN | Myeloid | -1.11E-07 | 5.60E-08 | 0.976661842 |
| Thal Phase | DC.4+.MLN | Myeloid | -2.75E-09 | 7.79E-08 | 0.51410357 |
| WMR (any/none) | DC.4+.MLN | Myeloid | -1.98E-09 | 3.73E-08 | 0.521169958 |
| #N/A | DC.4+.MLN | Myeloid | -4.05E-11 | 3.90E-08 | 0.500414468 |
| AD Braak | DC.4+.SLN | Myeloid | 5.70E-09 | 1.82E-08 | 0.377018151 |
| ADNC Score | DC.4+.SLN | Myeloid | 8.73E-08 | 6.47E-08 | 0.088728622 |
| AP (any/none) | DC.4+.SLN | Myeloid | 5.13E-09 | 2.07E-08 | 0.402362122 |
| Arteriolosclerosis | DC.4+.SLN | Myeloid | 2.80E-08 | 2.60E-08 | 0.140418777 |
| Arteriolosclerosis (any | DC.4+.SLN | Myeloid | 2.88E-08 | 2.76E-08 | 0.148499071 |
| Atherosclerosis | DC.4+.SLN | Myeloid | -2.45E-08 | 2.32E-08 | 0.854111346 |

| | | | | | |
|---|---|---|---|---|---|
| Atherosclerosis (any/r | DC.4+.SLN | Myeloid | -7.88E-09 | 2.44E-08 | 0.626821181 |
| B Score | DC.4+.SLN | Myeloid | 9.56E-10 | 1.74E-08 | 0.478112939 |
| CAA | DC.4+.SLN | Myeloid | 3.88E-08 | 2.23E-08 | 0.040923627 |
| CAA (any/none) | DC.4+.SLN | Myeloid | -2.97E-09 | 2.03E-08 | 0.558168625 |
| CERAD (C Score) | DC.4+.SLN | Myeloid | -7.73E-09 | 1.82E-08 | 0.664428029 |
| Infarcts/lacunes | DC.4+.SLN | Myeloid | 2.79E-08 | 2.44E-08 | 0.126892857 |
| PD Braak | DC.4+.SLN | Myeloid | 2.09E-08 | 1.82E-08 | 0.125427217 |
| PD Braak (any/none) | DC.4+.SLN | Myeloid | 1.47E-08 | 1.83E-08 | 0.210537153 |
| TDP-43 (any/none) | DC.4+.SLN | Myeloid | -9.00E-09 | 5.34E-08 | 0.566978173 |
| Thal Phase | DC.4+.SLN | Myeloid | 2.64E-08 | 7.79E-08 | 0.367326957 |
| WMR (any/none) | DC.4+.SLN | Myeloid | 2.63E-08 | 4.07E-08 | 0.25905146 |
| #N/A | DC.4+.SLN | Myeloid | 1.39E-08 | 4.36E-08 | 0.374908037 |
| AD Braak | DC.4+.Sp.ST | Myeloid | 1.34E-08 | 1.78E-08 | 0.225152192 |
| ADNC Score | DC.4+.Sp.ST | Myeloid | 5.20E-08 | 6.47E-08 | 0.210865107 |
| AP (any/none) | DC.4+.Sp.ST | Myeloid | 3.28E-08 | 2.32E-08 | 0.078688194 |
| Arteriolosclerosis | DC.4+.Sp.ST | Myeloid | 5.64E-09 | 2.82E-08 | 0.420555845 |
| Arteriolosclerosis (any | DC.4+.Sp.ST | Myeloid | 1.64E-08 | 2.67E-08 | 0.268949186 |
| Atherosclerosis | DC.4+.Sp.ST | Myeloid | -9.60E-09 | 2.19E-08 | 0.669168647 |
| Atherosclerosis (any/r | DC.4+.Sp.ST | Myeloid | -1.10E-08 | 2.41E-08 | 0.67605403 |
| B Score | DC.4+.Sp.ST | Myeloid | 2.00E-08 | 1.73E-08 | 0.123148555 |
| CAA | DC.4+.Sp.ST | Myeloid | -2.38E-08 | 2.46E-08 | 0.833902628 |
| CAA (any/none) | DC.4+.Sp.ST | Myeloid | -2.07E-08 | 2.20E-08 | 0.826889833 |
| CERAD (C Score) | DC.4+.Sp.ST | Myeloid | 2.87E-08 | 2.10E-08 | 0.086132029 |
| Infarcts/lacunes | DC.4+.Sp.ST | Myeloid | -7.74E-09 | 2.37E-08 | 0.628080622 |
| PD Braak | DC.4+.Sp.ST | Myeloid | 7.30E-09 | 1.91E-08 | 0.351197668 |
| PD Braak (any/none) | DC.4+.Sp.ST | Myeloid | 3.98E-09 | 1.84E-08 | 0.414360402 |
| TDP-43 (any/none) | DC.4+.Sp.ST | Myeloid | -8.86E-08 | 5.48E-08 | 0.947047897 |
| Thal Phase | DC.4+.Sp.ST | Myeloid | 3.50E-08 | 7.63E-08 | 0.323181468 |
| WMR (any/none) | DC.4+.Sp.ST | Myeloid | -4.54E-09 | 3.48E-08 | 0.551789167 |
| #N/A | DC.4+.Sp.ST | Myeloid | 2.02E-08 | 4.24E-08 | 0.31684151 |
| AD Braak | DC.8-.Th | Myeloid | -7.89E-09 | 1.66E-08 | 0.683092196 |
| ADNC Score | DC.8-.Th | Myeloid | 1.96E-08 | 6.73E-08 | 0.385175697 |
| AP (any/none) | DC.8-.Th | Myeloid | -2.23E-09 | 2.23E-08 | 0.53969037 |
| Arteriolosclerosis | DC.8-.Th | Myeloid | -5.08E-08 | 2.97E-08 | 0.956331039 |

| | | | | | |
|---|---|---|---|---|---|
| Arteriolosclerosis (any | DC.8-.Th | Myeloid | -6.50E-09 | 3.06E-08 | 0.584207297 |
| Atherosclerosis | DC.8-.Th | Myeloid | -2.44E-08 | 2.68E-08 | 0.818869034 |
| Atherosclerosis (any/r | DC.8-.Th | Myeloid | -5.81E-09 | 2.27E-08 | 0.601058858 |
| B Score | DC.8-.Th | Myeloid | -4.51E-09 | 1.73E-08 | 0.602778454 |
| CAA | DC.8-.Th | Myeloid | -1.66E-08 | 2.38E-08 | 0.757466654 |
| CAA (any/none) | DC.8-.Th | Myeloid | -1.95E-08 | 2.39E-08 | 0.791867134 |
| CERAD (C Score) | DC.8-.Th | Myeloid | -1.11E-08 | 1.88E-08 | 0.72265309 |
| Infarcts/lacunes | DC.8-.Th | Myeloid | 1.22E-08 | 2.38E-08 | 0.304037635 |
| PD Braak | DC.8-.Th | Myeloid | 1.30E-08 | 2.03E-08 | 0.260877343 |
| PD Braak (any/none) | DC.8-.Th | Myeloid | 2.33E-08 | 1.97E-08 | 0.119002261 |
| TDP-43 (any/none) | DC.8-.Th | Myeloid | 1.23E-08 | 5.52E-08 | 0.411667161 |
| Thal Phase | DC.8-.Th | Myeloid | 6.02E-08 | 8.49E-08 | 0.23907046 |
| WMR (any/none) | DC.8-.Th | Myeloid | -4.55E-09 | 4.01E-08 | 0.545141812 |
| #N/A | DC.8-.Th | Myeloid | -3.31E-08 | 4.31E-08 | 0.778991207 |
| AD Braak | DC.8+.MLN | Myeloid | -6.77E-09 | 1.62E-08 | 0.661687817 |
| ADNC Score | DC.8+.MLN | Myeloid | -3.46E-08 | 6.22E-08 | 0.711358688 |
| AP (any/none) | DC.8+.MLN | Myeloid | 2.28E-09 | 2.14E-08 | 0.457532865 |
| Arteriolosclerosis | DC.8+.MLN | Myeloid | -2.63E-08 | 3.09E-08 | 0.802931849 |
| Arteriolosclerosis (any | DC.8+.MLN | Myeloid | 2.36E-08 | 3.00E-08 | 0.215514595 |
| Atherosclerosis | DC.8+.MLN | Myeloid | 1.45E-09 | 2.27E-08 | 0.474510232 |
| Atherosclerosis (any/r | DC.8+.MLN | Myeloid | 2.63E-09 | 2.47E-08 | 0.45752018 |
| B Score | DC.8+.MLN | Myeloid | -1.12E-08 | 1.61E-08 | 0.756627287 |
| CAA | DC.8+.MLN | Myeloid | -3.06E-08 | 2.31E-08 | 0.906768461 |
| CAA (any/none) | DC.8+.MLN | Myeloid | -1.67E-08 | 2.26E-08 | 0.76937149 |
| CERAD (C Score) | DC.8+.MLN | Myeloid | -5.19E-09 | 1.88E-08 | 0.608796338 |
| Infarcts/lacunes | DC.8+.MLN | Myeloid | -6.55E-09 | 2.50E-08 | 0.603235894 |
| PD Braak | DC.8+.MLN | Myeloid | 9.07E-09 | 2.04E-08 | 0.328396577 |
| PD Braak (any/none) | DC.8+.MLN | Myeloid | 6.84E-09 | 1.88E-08 | 0.358220328 |
| TDP-43 (any/none) | DC.8+.MLN | Myeloid | 3.73E-08 | 5.40E-08 | 0.245185018 |
| Thal Phase | DC.8+.MLN | Myeloid | -5.31E-08 | 7.09E-08 | 0.773130155 |
| WMR (any/none) | DC.8+.MLN | Myeloid | -2.71E-08 | 3.46E-08 | 0.783472589 |
| #N/A | DC.8+.MLN | Myeloid | -7.34E-08 | 4.29E-08 | 0.956279959 |
| AD Braak | DC.8+.SLN | Myeloid | -5.64E-09 | 1.56E-08 | 0.640905397 |
| ADNC Score | DC.8+.SLN | Myeloid | 1.30E-09 | 6.27E-08 | 0.491708823 |

| | | | | | |
|---|---|---|---|---|---|
| AP (any/none) | DC.8+.SLN | Myeloid | 1.83E-08 | 2.21E-08 | 0.203595672 |
| Arteriolosclerosis | DC.8+.SLN | Myeloid | 2.20E-09 | 2.90E-08 | 0.46967266 |
| Arteriolosclerosis (any | DC.8+.SLN | Myeloid | 3.19E-08 | 2.84E-08 | 0.130093016 |
| Atherosclerosis | DC.8+.SLN | Myeloid | 6.47E-09 | 2.35E-08 | 0.39158367 |
| Atherosclerosis (any/r | DC.8+.SLN | Myeloid | 1.68E-08 | 2.59E-08 | 0.258748154 |
| B Score | DC.8+.SLN | Myeloid | 1.49E-09 | 1.52E-08 | 0.461013342 |
| CAA | DC.8+.SLN | Myeloid | -1.86E-08 | 2.28E-08 | 0.792696956 |
| CAA (any/none) | DC.8+.SLN | Myeloid | -2.95E-09 | 2.11E-08 | 0.555415416 |
| CERAD (C Score) | DC.8+.SLN | Myeloid | 5.74E-09 | 1.84E-08 | 0.377760165 |
| Infarcts/lacunes | DC.8+.SLN | Myeloid | -1.67E-08 | 2.49E-08 | 0.748646297 |
| PD Braak | DC.8+.SLN | Myeloid | 3.00E-08 | 1.86E-08 | 0.053312777 |
| PD Braak (any/none) | DC.8+.SLN | Myeloid | 2.79E-08 | 1.78E-08 | 0.057993189 |
| TDP-43 (any/none) | DC.8+.SLN | Myeloid | -1.57E-08 | 5.47E-08 | 0.612952332 |
| Thal Phase | DC.8+.SLN | Myeloid | -6.13E-08 | 8.08E-08 | 0.775922966 |
| WMR (any/none) | DC.8+.SLN | Myeloid | -1.19E-08 | 3.52E-08 | 0.63219018 |
| #N/A | DC.8+.SLN | Myeloid | -2.32E-08 | 4.67E-08 | 0.690637079 |
| AD Braak | DC.8+.Sp.ST | Myeloid | -2.32E-08 | 1.70E-08 | 0.913416623 |
| ADNC Score | DC.8+.Sp.ST | Myeloid | -7.11E-08 | 5.73E-08 | 0.892735352 |
| AP (any/none) | DC.8+.Sp.ST | Myeloid | -2.61E-09 | 2.09E-08 | 0.549571861 |
| Arteriolosclerosis | DC.8+.Sp.ST | Myeloid | -4.22E-08 | 2.64E-08 | 0.94457447 |
| Arteriolosclerosis (any | DC.8+.Sp.ST | Myeloid | -5.94E-10 | 2.75E-08 | 0.508626343 |
| Atherosclerosis | DC.8+.Sp.ST | Myeloid | -8.69E-09 | 2.21E-08 | 0.652716548 |
| Atherosclerosis (any/r | DC.8+.Sp.ST | Myeloid | 2.06E-10 | 2.47E-08 | 0.496671462 |
| B Score | DC.8+.Sp.ST | Myeloid | -1.39E-08 | 1.71E-08 | 0.79180764 |
| CAA | DC.8+.Sp.ST | Myeloid | -1.88E-08 | 2.25E-08 | 0.797541552 |
| CAA (any/none) | DC.8+.Sp.ST | Myeloid | -2.48E-08 | 2.12E-08 | 0.878885885 |
| CERAD (C Score) | DC.8+.Sp.ST | Myeloid | -3.05E-09 | 1.81E-08 | 0.566923456 |
| Infarcts/lacunes | DC.8+.Sp.ST | Myeloid | 9.30E-09 | 2.36E-08 | 0.346447435 |
| PD Braak | DC.8+.Sp.ST | Myeloid | 1.66E-08 | 1.97E-08 | 0.200249435 |
| PD Braak (any/none) | DC.8+.Sp.ST | Myeloid | 1.20E-08 | 1.89E-08 | 0.263382536 |
| TDP-43 (any/none) | DC.8+.Sp.ST | Myeloid | 4.28E-08 | 5.59E-08 | 0.221736647 |
| Thal Phase | DC.8+.Sp.ST | Myeloid | -8.89E-08 | 7.25E-08 | 0.890019332 |
| WMR (any/none) | DC.8+.Sp.ST | Myeloid | -5.32E-08 | 3.58E-08 | 0.931141032 |
| #N/A | DC.8+.Sp.ST | Myeloid | -3.50E-08 | 4.38E-08 | 0.788055276 |

| | | | | | |
|---|---|---|---|---|---|
| AD Braak | DC.8+.Th | Myeloid | -9.82E-09 | 1.72E-08 | 0.71622635 |
| ADNC Score | DC.8+.Th | Myeloid | 1.34E-08 | 6.39E-08 | 0.417112994 |
| AP (any/none) | DC.8+.Th | Myeloid | -1.31E-08 | 2.22E-08 | 0.721599404 |
| Arteriolosclerosis | DC.8+.Th | Myeloid | -4.90E-08 | 2.99E-08 | 0.949261277 |
| Arteriolosclerosis (any | DC.8+.Th | Myeloid | 2.15E-08 | 3.07E-08 | 0.241666621 |
| Atherosclerosis | DC.8+.Th | Myeloid | -2.05E-08 | 2.46E-08 | 0.797518176 |
| Atherosclerosis (any/r | DC.8+.Th | Myeloid | 2.79E-09 | 2.33E-08 | 0.452295254 |
| B Score | DC.8+.Th | Myeloid | -6.46E-09 | 1.73E-08 | 0.645647756 |
| CAA | DC.8+.Th | Myeloid | -1.48E-08 | 2.41E-08 | 0.730613732 |
| CAA (any/none) | DC.8+.Th | Myeloid | -1.31E-08 | 2.32E-08 | 0.713193928 |
| CERAD (C Score) | DC.8+.Th | Myeloid | -1.82E-08 | 1.92E-08 | 0.828855803 |
| Infarcts/lacunes | DC.8+.Th | Myeloid | 1.76E-08 | 2.34E-08 | 0.225572445 |
| PD Braak | DC.8+.Th | Myeloid | 1.46E-08 | 1.99E-08 | 0.230778839 |
| PD Braak (any/none) | DC.8+.Th | Myeloid | 2.19E-08 | 1.86E-08 | 0.119810089 |
| TDP-43 (any/none) | DC.8+.Th | Myeloid | 1.67E-08 | 5.41E-08 | 0.378972281 |
| Thal Phase | DC.8+.Th | Myeloid | 5.21E-09 | 7.82E-08 | 0.473450804 |
| WMR (any/none) | DC.8+.Th | Myeloid | -7.09E-09 | 3.89E-08 | 0.572217336 |
| #N/A | DC.8+.Th | Myeloid | -6.66E-08 | 4.28E-08 | 0.940070721 |
| AD Braak | DC.8-4-11b-.MLN | Myeloid | -3.01E-09 | 1.68E-08 | 0.571246602 |
| ADNC Score | DC.8-4-11b-.MLN | Myeloid | -5.12E-08 | 5.91E-08 | 0.80682291 |
| AP (any/none) | DC.8-4-11b-.MLN | Myeloid | 7.64E-09 | 2.29E-08 | 0.369403602 |
| Arteriolosclerosis | DC.8-4-11b-.MLN | Myeloid | -3.32E-08 | 2.61E-08 | 0.898009788 |
| Arteriolosclerosis (any | DC.8-4-11b-.MLN | Myeloid | -5.99E-09 | 2.68E-08 | 0.588505225 |
| Atherosclerosis | DC.8-4-11b-.MLN | Myeloid | 2.85E-09 | 2.27E-08 | 0.45012819 |
| Atherosclerosis (any/r | DC.8-4-11b-.MLN | Myeloid | 1.88E-08 | 2.32E-08 | 0.20949146 |
| B Score | DC.8-4-11b-.MLN | Myeloid | -1.00E-08 | 1.70E-08 | 0.722309036 |
| CAA | DC.8-4-11b-.MLN | Myeloid | -2.90E-08 | 2.32E-08 | 0.893600858 |
| CAA (any/none) | DC.8-4-11b-.MLN | Myeloid | -2.58E-08 | 2.41E-08 | 0.857917151 |
| CERAD (C Score) | DC.8-4-11b-.MLN | Myeloid | 4.92E-09 | 1.83E-08 | 0.393987859 |
| Infarcts/lacunes | DC.8-4-11b-.MLN | Myeloid | -9.11E-09 | 2.27E-08 | 0.655987047 |
| PD Braak | DC.8-4-11b-.MLN | Myeloid | 4.14E-08 | 1.82E-08 | 0.01138153 |
| PD Braak (any/none) | DC.8-4-11b-.MLN | Myeloid | 3.19E-08 | 1.77E-08 | 0.035753847 |
| TDP-43 (any/none) | DC.8-4-11b-.MLN | Myeloid | -9.27E-08 | 4.92E-08 | 0.970280053 |
| Thal Phase | DC.8-4-11b-.MLN | Myeloid | 1.71E-08 | 7.56E-08 | 0.41072274 |

| | | | | | |
|---|---|---|---|---|---|
| WMR (any/none) | DC.8-4-11b-.MLN | Myeloid | 4.30E-08 | 3.75E-08 | 0.125912851 |
| #N/A | DC.8-4-11b-.MLN | Myeloid | 2.53E-08 | 4.31E-08 | 0.27883177 |
| AD Braak | DC.8-4-11b-.SLN | Myeloid | -1.02E-08 | 1.72E-08 | 0.723725945 |
| ADNC Score | DC.8-4-11b-.SLN | Myeloid | -2.68E-08 | 6.28E-08 | 0.665308922 |
| AP (any/none) | DC.8-4-11b-.SLN | Myeloid | 1.72E-08 | 2.33E-08 | 0.23038356 |
| Arteriolosclerosis | DC.8-4-11b-.SLN | Myeloid | -8.14E-09 | 2.76E-08 | 0.616103181 |
| Arteriolosclerosis (any | DC.8-4-11b-.SLN | Myeloid | 5.28E-09 | 3.01E-08 | 0.430496049 |
| Atherosclerosis | DC.8-4-11b-.SLN | Myeloid | -1.28E-08 | 2.26E-08 | 0.714875753 |
| Atherosclerosis (any/r | DC.8-4-11b-.SLN | Myeloid | -1.15E-08 | 2.53E-08 | 0.675830636 |
| B Score | DC.8-4-11b-.SLN | Myeloid | -1.19E-08 | 1.80E-08 | 0.745536555 |
| CAA | DC.8-4-11b-.SLN | Myeloid | -8.40E-09 | 2.31E-08 | 0.6417589 |
| CAA (any/none) | DC.8-4-11b-.SLN | Myeloid | 3.29E-10 | 2.39E-08 | 0.494514308 |
| CERAD (C Score) | DC.8-4-11b-.SLN | Myeloid | 9.42E-10 | 1.97E-08 | 0.480923739 |
| Infarcts/lacunes | DC.8-4-11b-.SLN | Myeloid | 1.07E-08 | 2.25E-08 | 0.317621089 |
| PD Braak | DC.8-4-11b-.SLN | Myeloid | 2.90E-08 | 1.69E-08 | 0.043515748 |
| PD Braak (any/none) | DC.8-4-11b-.SLN | Myeloid | 3.29E-08 | 1.66E-08 | 0.023706752 |
| TDP-43 (any/none) | DC.8-4-11b-.SLN | Myeloid | -5.38E-08 | 4.98E-08 | 0.859996404 |
| Thal Phase | DC.8-4-11b-.SLN | Myeloid | 1.44E-08 | 7.59E-08 | 0.424721823 |
| WMR (any/none) | DC.8-4-11b-.SLN | Myeloid | -2.98E-08 | 4.00E-08 | 0.771616797 |
| #N/A | DC.8-4-11b-.SLN | Myeloid | -5.21E-08 | 4.48E-08 | 0.87744516 |
| AD Braak | DC.8-4-11b-.Sp | Myeloid | -1.70E-09 | 1.74E-08 | 0.539052154 |
| ADNC Score | DC.8-4-11b-.Sp | Myeloid | -1.33E-08 | 5.54E-08 | 0.59522013 |
| AP (any/none) | DC.8-4-11b-.Sp | Myeloid | 3.12E-10 | 2.08E-08 | 0.494016167 |
| Arteriolosclerosis | DC.8-4-11b-.Sp | Myeloid | -1.01E-09 | 3.03E-08 | 0.513355988 |
| Arteriolosclerosis (any | DC.8-4-11b-.Sp | Myeloid | 1.11E-08 | 2.81E-08 | 0.346702885 |
| Atherosclerosis | DC.8-4-11b-.Sp | Myeloid | -1.36E-08 | 2.35E-08 | 0.719623521 |
| Atherosclerosis (any/r | DC.8-4-11b-.Sp | Myeloid | -1.36E-08 | 2.43E-08 | 0.712535935 |
| B Score | DC.8-4-11b-.Sp | Myeloid | 7.85E-09 | 1.68E-08 | 0.319883821 |
| CAA | DC.8-4-11b-.Sp | Myeloid | -1.40E-08 | 2.28E-08 | 0.730666398 |
| CAA (any/none) | DC.8-4-11b-.Sp | Myeloid | -1.57E-08 | 2.11E-08 | 0.771551721 |
| CERAD (C Score) | DC.8-4-11b-.Sp | Myeloid | 1.28E-08 | 1.98E-08 | 0.25852247 |
| Infarcts/lacunes | DC.8-4-11b-.Sp | Myeloid | 4.47E-09 | 2.34E-08 | 0.424182971 |
| PD Braak | DC.8-4-11b-.Sp | Myeloid | 8.42E-09 | 1.92E-08 | 0.330849695 |
| PD Braak (any/none) | DC.8-4-11b-.Sp | Myeloid | 4.16E-09 | 1.81E-08 | 0.409342807 |

| | | | | | |
|---|---|---|---|---|---|
| TDP-43 (any/none) | DC.8-4-11b-.Sp | Myeloid | -7.31E-08 | 5.41E-08 | 0.911652167 |
| Thal Phase | DC.8-4-11b-.Sp | Myeloid | -8.14E-08 | 6.56E-08 | 0.892770644 |
| WMR (any/none) | DC.8-4-11b-.Sp | Myeloid | -2.69E-09 | 3.69E-08 | 0.529042941 |
| #N/A | DC.8-4-11b-.Sp | Myeloid | 3.46E-09 | 4.02E-08 | 0.465664022 |
| AD Braak | DC.8-4-11b+.MLN | Myeloid | -1.24E-08 | 1.75E-08 | 0.761086777 |
| ADNC Score | DC.8-4-11b+.MLN | Myeloid | -6.83E-08 | 6.19E-08 | 0.865312768 |
| AP (any/none) | DC.8-4-11b+.MLN | Myeloid | 9.63E-09 | 2.18E-08 | 0.329373882 |
| Arteriolosclerosis | DC.8-4-11b+.MLN | Myeloid | 5.53E-10 | 2.61E-08 | 0.491558255 |
| Arteriolosclerosis (any | DC.8-4-11b+.MLN | Myeloid | -1.65E-08 | 2.87E-08 | 0.717484352 |
| Atherosclerosis | DC.8-4-11b+.MLN | Myeloid | -3.72E-09 | 2.23E-08 | 0.566367628 |
| Atherosclerosis (any/r | DC.8-4-11b+.MLN | Myeloid | -1.97E-08 | 2.18E-08 | 0.816742285 |
| B Score | DC.8-4-11b+.MLN | Myeloid | -1.21E-09 | 1.70E-08 | 0.528212403 |
| CAA | DC.8-4-11b+.MLN | Myeloid | -2.51E-08 | 2.38E-08 | 0.854272347 |
| CAA (any/none) | DC.8-4-11b+.MLN | Myeloid | -1.80E-08 | 2.36E-08 | 0.777711793 |
| CERAD (C Score) | DC.8-4-11b+.MLN | Myeloid | 3.07E-09 | 1.89E-08 | 0.435423714 |
| Infarcts/lacunes | DC.8-4-11b+.MLN | Myeloid | -4.70E-10 | 2.43E-08 | 0.507715964 |
| PD Braak | DC.8-4-11b+.MLN | Myeloid | 3.40E-08 | 1.87E-08 | 0.034380931 |
| PD Braak (any/none) | DC.8-4-11b+.MLN | Myeloid | 2.10E-08 | 1.76E-08 | 0.116227798 |
| TDP-43 (any/none) | DC.8-4-11b+.MLN | Myeloid | -8.60E-08 | 4.74E-08 | 0.965279918 |
| Thal Phase | DC.8-4-11b+.MLN | Myeloid | 9.77E-09 | 7.96E-08 | 0.451178037 |
| WMR (any/none) | DC.8-4-11b+.MLN | Myeloid | -8.08E-10 | 3.81E-08 | 0.508465655 |
| #N/A | DC.8-4-11b+.MLN | Myeloid | 2.39E-08 | 4.67E-08 | 0.304258794 |
| AD Braak | DC.8-4-11b+.SLN | Myeloid | -2.19E-08 | 1.70E-08 | 0.901113922 |
| ADNC Score | DC.8-4-11b+.SLN | Myeloid | -6.96E-09 | 6.42E-08 | 0.543159621 |
| AP (any/none) | DC.8-4-11b+.SLN | Myeloid | 6.73E-09 | 2.22E-08 | 0.380646088 |
| Arteriolosclerosis | DC.8-4-11b+.SLN | Myeloid | -1.34E-08 | 2.63E-08 | 0.695040979 |
| Arteriolosclerosis (any | DC.8-4-11b+.SLN | Myeloid | 8.32E-09 | 2.94E-08 | 0.38845228 |
| Atherosclerosis | DC.8-4-11b+.SLN | Myeloid | -2.88E-08 | 2.23E-08 | 0.900887119 |
| Atherosclerosis (any/r | DC.8-4-11b+.SLN | Myeloid | -1.54E-08 | 2.32E-08 | 0.746478862 |
| B Score | DC.8-4-11b+.SLN | Myeloid | -1.56E-08 | 1.76E-08 | 0.811564946 |
| CAA | DC.8-4-11b+.SLN | Myeloid | -1.22E-08 | 2.30E-08 | 0.702735697 |
| CAA (any/none) | DC.8-4-11b+.SLN | Myeloid | -1.82E-08 | 2.18E-08 | 0.797767729 |
| CERAD (C Score) | DC.8-4-11b+.SLN | Myeloid | -9.44E-10 | 1.97E-08 | 0.519143131 |
| Infarcts/lacunes | DC.8-4-11b+.SLN | Myeloid | -4.97E-09 | 2.46E-08 | 0.580090341 |

| | | | | | |
|---|---|---|---|---|---|
| PD Braak | DC.8-4-11b+.SLN | Myeloid | 1.10E-08 | 1.82E-08 | 0.272423275 |
| PD Braak (any/none) | DC.8-4-11b+.SLN | Myeloid | 3.85E-09 | 1.75E-08 | 0.412788245 |
| TDP-43 (any/none) | DC.8-4-11b+.SLN | Myeloid | -4.31E-08 | 5.56E-08 | 0.781086872 |
| Thal Phase | DC.8-4-11b+.SLN | Myeloid | 7.36E-08 | 8.20E-08 | 0.184666567 |
| WMR (any/none) | DC.8-4-11b+.SLN | Myeloid | 5.54E-08 | 3.85E-08 | 0.075192262 |
| #N/A | DC.8-4-11b+.SLN | Myeloid | 2.37E-09 | 4.67E-08 | 0.479775723 |
| AD Braak | DC.8-4-11b+.Sp | Myeloid | 1.76E-08 | 1.93E-08 | 0.180838922 |
| ADNC Score | DC.8-4-11b+.Sp | Myeloid | 7.50E-08 | 5.95E-08 | 0.103458178 |
| AP (any/none) | DC.8-4-11b+.Sp | Myeloid | 3.12E-08 | 2.28E-08 | 0.085982804 |
| Arteriolosclerosis | DC.8-4-11b+.Sp | Myeloid | 1.45E-08 | 2.83E-08 | 0.303504122 |
| Arteriolosclerosis (any | DC.8-4-11b+.Sp | Myeloid | -4.98E-09 | 2.77E-08 | 0.571295162 |
| Atherosclerosis | DC.8-4-11b+.Sp | Myeloid | -9.16E-10 | 2.43E-08 | 0.515004227 |
| Atherosclerosis (any/r | DC.8-4-11b+.Sp | Myeloid | 2.71E-09 | 2.63E-08 | 0.45894148 |
| B Score | DC.8-4-11b+.Sp | Myeloid | 1.27E-08 | 1.79E-08 | 0.239630753 |
| CAA | DC.8-4-11b+.Sp | Myeloid | 4.11E-09 | 2.33E-08 | 0.42998074 |
| CAA (any/none) | DC.8-4-11b+.Sp | Myeloid | -2.20E-08 | 2.01E-08 | 0.862543298 |
| CERAD (C Score) | DC.8-4-11b+.Sp | Myeloid | 3.61E-08 | 1.89E-08 | 0.028244154 |
| Infarcts/lacunes | DC.8-4-11b+.Sp | Myeloid | 1.70E-08 | 2.36E-08 | 0.236565683 |
| PD Braak | DC.8-4-11b+.Sp | Myeloid | 4.35E-08 | 2.04E-08 | 0.016546174 |
| PD Braak (any/none) | DC.8-4-11b+.Sp | Myeloid | 3.35E-08 | 1.90E-08 | 0.039072739 |
| TDP-43 (any/none) | DC.8-4-11b+.Sp | Myeloid | -1.27E-08 | 5.41E-08 | 0.592941212 |
| Thal Phase | DC.8-4-11b+.Sp | Myeloid | 7.06E-08 | 8.18E-08 | 0.194098471 |
| WMR (any/none) | DC.8-4-11b+.Sp | Myeloid | 5.93E-09 | 3.63E-08 | 0.435058307 |
| #N/A | DC.8-4-11b+.Sp | Myeloid | -3.74E-08 | 3.92E-08 | 0.830296906 |
| AD Braak | DC.IIhilang+103+11blo.SLN | Myeloid | -9.01E-09 | 1.76E-08 | 0.695736786 |
| ADNC Score | DC.IIhilang+103+11blo.SLN | Myeloid | -5.21E-09 | 6.21E-08 | 0.533424847 |
| AP (any/none) | DC.IIhilang+103+11blo.SLN | Myeloid | 3.33E-08 | 2.14E-08 | 0.059807397 |
| Arteriolosclerosis | DC.IIhilang+103+11blo.SLN | Myeloid | -1.79E-08 | 2.91E-08 | 0.731021686 |
| Arteriolosclerosis (any | DC.IIhilang+103+11blo.SLN | Myeloid | 2.48E-08 | 3.06E-08 | 0.208380835 |
| Atherosclerosis | DC.IIhilang+103+11blo.SLN | Myeloid | -3.56E-08 | 2.30E-08 | 0.938648119 |
| Atherosclerosis (any/r | DC.IIhilang+103+11blo.SLN | Myeloid | -1.37E-08 | 2.47E-08 | 0.710597793 |
| B Score | DC.IIhilang+103+11blo.SLN | Myeloid | -5.18E-09 | 1.73E-08 | 0.617497859 |
| CAA | DC.IIhilang+103+11blo.SLN | Myeloid | -1.91E-08 | 2.23E-08 | 0.804731104 |
| CAA (any/none) | DC.IIhilang+103+11blo.SLN | Myeloid | -1.87E-08 | 2.32E-08 | 0.789525194 |

| | | | | | |
|---|---|---|---|---|---|
| CERAD (C Score) | DC.IIhilang+103+11blo.SLN | Myeloid | -8.37E-09 | 1.99E-08 | 0.663364002 |
| Infarcts/lacunes | DC.IIhilang+103+11blo.SLN | Myeloid | -3.14E-08 | 2.39E-08 | 0.905613885 |
| PD Braak | DC.IIhilang+103+11blo.SLN | Myeloid | 2.42E-08 | 1.97E-08 | 0.110303665 |
| PD Braak (any/none) | DC.IIhilang+103+11blo.SLN | Myeloid | 2.50E-08 | 1.91E-08 | 0.094805273 |
| TDP-43 (any/none) | DC.IIhilang+103+11blo.SLN | Myeloid | -9.63E-08 | 5.23E-08 | 0.967294359 |
| Thal Phase | DC.IIhilang+103+11blo.SLN | Myeloid | 6.05E-08 | 8.06E-08 | 0.226393517 |
| WMR (any/none) | DC.IIhilang+103+11blo.SLN | Myeloid | 3.59E-08 | 3.88E-08 | 0.176960687 |
| #N/A | DC.IIhilang+103+11blo.SLN | Myeloid | -1.46E-08 | 4.52E-08 | 0.626577653 |
| AD Braak | DC.IIhilang+103-11b+.SLN | Myeloid | -1.73E-08 | 1.65E-08 | 0.852623118 |
| ADNC Score | DC.IIhilang+103-11b+.SLN | Myeloid | 4.05E-08 | 5.90E-08 | 0.246255741 |
| AP (any/none) | DC.IIhilang+103-11b+.SLN | Myeloid | 4.01E-08 | 2.08E-08 | 0.027001841 |
| Arteriolosclerosis | DC.IIhilang+103-11b+.SLN | Myeloid | -4.94E-09 | 2.47E-08 | 0.579103912 |
| Arteriolosclerosis (any | DC.IIhilang+103-11b+.SLN | Myeloid | 1.68E-08 | 2.80E-08 | 0.273986026 |
| Atherosclerosis | DC.IIhilang+103-11b+.SLN | Myeloid | 6.72E-09 | 2.19E-08 | 0.379781662 |
| Atherosclerosis (any/r | DC.IIhilang+103-11b+.SLN | Myeloid | -1.50E-09 | 2.19E-08 | 0.527189661 |
| B Score | DC.IIhilang+103-11b+.SLN | Myeloid | 2.32E-11 | 1.73E-08 | 0.49946278 |
| CAA | DC.IIhilang+103-11b+.SLN | Myeloid | -2.04E-08 | 2.14E-08 | 0.829126615 |
| CAA (any/none) | DC.IIhilang+103-11b+.SLN | Myeloid | -9.46E-09 | 2.40E-08 | 0.6532088 |
| CERAD (C Score) | DC.IIhilang+103-11b+.SLN | Myeloid | 1.26E-08 | 1.99E-08 | 0.263331341 |
| Infarcts/lacunes | DC.IIhilang+103-11b+.SLN | Myeloid | -1.26E-09 | 2.31E-08 | 0.52176036 |
| PD Braak | DC.IIhilang+103-11b+.SLN | Myeloid | 2.93E-08 | 2.06E-08 | 0.078065517 |
| PD Braak (any/none) | DC.IIhilang+103-11b+.SLN | Myeloid | 2.94E-08 | 2.03E-08 | 0.073802987 |
| TDP-43 (any/none) | DC.IIhilang+103-11b+.SLN | Myeloid | -9.60E-09 | 5.28E-08 | 0.572161746 |
| Thal Phase | DC.IIhilang+103-11b+.SLN | Myeloid | 1.06E-07 | 7.97E-08 | 0.091635424 |
| WMR (any/none) | DC.IIhilang+103-11b+.SLN | Myeloid | 1.45E-09 | 3.59E-08 | 0.483930814 |
| #N/A | DC.IIhilang+103-11b+.SLN | Myeloid | 7.66E-09 | 4.23E-08 | 0.42807015 |
| AD Braak | DC.IIhilang-103-11b+.SLN | Myeloid | -1.03E-08 | 1.67E-08 | 0.731947695 |
| ADNC Score | DC.IIhilang-103-11b+.SLN | Myeloid | -4.10E-09 | 5.70E-08 | 0.528716166 |
| AP (any/none) | DC.IIhilang-103-11b+.SLN | Myeloid | -5.66E-09 | 2.18E-08 | 0.602556124 |
| Arteriolosclerosis | DC.IIhilang-103-11b+.SLN | Myeloid | 9.31E-09 | 2.67E-08 | 0.363384578 |
| Arteriolosclerosis (any | DC.IIhilang-103-11b+.SLN | Myeloid | 5.20E-08 | 3.21E-08 | 0.052546573 |
| Atherosclerosis | DC.IIhilang-103-11b+.SLN | Myeloid | -2.36E-08 | 2.26E-08 | 0.851908821 |
| Atherosclerosis (any/r | DC.IIhilang-103-11b+.SLN | Myeloid | -7.98E-09 | 2.36E-08 | 0.632309043 |
| B Score | DC.IIhilang-103-11b+.SLN | Myeloid | -4.27E-09 | 1.75E-08 | 0.596619519 |

| | | | | | |
|---|---|---|---|---|---|
| CAA | DC.IIhilang-103-11b+.SLN | Myeloid | -2.54E-08 | 2.02E-08 | 0.89516794 |
| CAA (any/none) | DC.IIhilang-103-11b+.SLN | Myeloid | -1.15E-08 | 2.13E-08 | 0.705983658 |
| CERAD (C Score) | DC.IIhilang-103-11b+.SLN | Myeloid | -3.00E-09 | 1.92E-08 | 0.562192507 |
| Infarcts/lacunes | DC.IIhilang-103-11b+.SLN | Myeloid | -1.30E-08 | 2.46E-08 | 0.701092269 |
| PD Braak | DC.IIhilang-103-11b+.SLN | Myeloid | -5.19E-09 | 1.75E-08 | 0.616576334 |
| PD Braak (any/none) | DC.IIhilang-103-11b+.SLN | Myeloid | -1.12E-08 | 1.63E-08 | 0.754308008 |
| TDP-43 (any/none) | DC.IIhilang-103-11b+.SLN | Myeloid | -3.25E-08 | 5.16E-08 | 0.735665765 |
| Thal Phase | DC.IIhilang-103-11b+.SLN | Myeloid | 1.29E-07 | 8.29E-08 | 0.060445043 |
| WMR (any/none) | DC.IIhilang-103-11b+.SLN | Myeloid | 6.31E-08 | 3.82E-08 | 0.049418986 |
| #N/A | DC.IIhilang-103-11b+.SLN | Myeloid | -1.98E-08 | 4.42E-08 | 0.672975863 |
| AD Braak | DC.IIhilang-103-11blo.SLN | Myeloid | -1.62E-08 | 1.71E-08 | 0.827507933 |
| ADNC Score | DC.IIhilang-103-11blo.SLN | Myeloid | -2.07E-08 | 6.20E-08 | 0.63051407 |
| AP (any/none) | DC.IIhilang-103-11blo.SLN | Myeloid | 8.36E-09 | 2.17E-08 | 0.350168014 |
| Arteriolosclerosis | DC.IIhilang-103-11blo.SLN | Myeloid | -1.13E-08 | 2.65E-08 | 0.665095637 |
| Arteriolosclerosis (any | DC.IIhilang-103-11blo.SLN | Myeloid | 2.28E-08 | 2.84E-08 | 0.211719814 |
| Atherosclerosis | DC.IIhilang-103-11blo.SLN | Myeloid | -2.09E-09 | 2.19E-08 | 0.538006237 |
| Atherosclerosis (any/r | DC.IIhilang-103-11blo.SLN | Myeloid | 3.76E-09 | 2.45E-08 | 0.438934338 |
| B Score | DC.IIhilang-103-11blo.SLN | Myeloid | -1.27E-08 | 1.70E-08 | 0.772731527 |
| CAA | DC.IIhilang-103-11blo.SLN | Myeloid | -1.69E-08 | 2.33E-08 | 0.765932398 |
| CAA (any/none) | DC.IIhilang-103-11blo.SLN | Myeloid | -2.76E-08 | 2.35E-08 | 0.88029863 |
| CERAD (C Score) | DC.IIhilang-103-11blo.SLN | Myeloid | -5.90E-09 | 2.01E-08 | 0.61540824 |
| Infarcts/lacunes | DC.IIhilang-103-11blo.SLN | Myeloid | -1.33E-08 | 2.46E-08 | 0.706282316 |
| PD Braak | DC.IIhilang-103-11blo.SLN | Myeloid | 1.58E-08 | 1.82E-08 | 0.192879467 |
| PD Braak (any/none) | DC.IIhilang-103-11blo.SLN | Myeloid | 8.74E-09 | 1.74E-08 | 0.307708079 |
| TDP-43 (any/none) | DC.IIhilang-103-11blo.SLN | Myeloid | -1.35E-07 | 4.87E-08 | 0.997161693 |
| Thal Phase | DC.IIhilang-103-11blo.SLN | Myeloid | 9.94E-08 | 8.87E-08 | 0.131274707 |
| WMR (any/none) | DC.IIhilang-103-11blo.SLN | Myeloid | 3.28E-08 | 3.92E-08 | 0.201451112 |
| #N/A | DC.IIhilang-103-11blo.SLN | Myeloid | 1.69E-08 | 4.40E-08 | 0.349988526 |
| AD Braak | DC.LC.Sk | Myeloid | -8.28E-10 | 1.95E-08 | 0.516892646 |
| ADNC Score | DC.LC.Sk | Myeloid | 3.67E-08 | 5.85E-08 | 0.264968047 |
| AP (any/none) | DC.LC.Sk | Myeloid | 4.26E-09 | 2.35E-08 | 0.42806793 |
| Arteriolosclerosis | DC.LC.Sk | Myeloid | 4.72E-08 | 3.24E-08 | 0.07229137 |
| Arteriolosclerosis (any | DC.LC.Sk | Myeloid | 4.78E-08 | 3.46E-08 | 0.083234146 |
| Atherosclerosis | DC.LC.Sk | Myeloid | 2.17E-09 | 2.44E-08 | 0.464602269 |

| | | | | | |
|---|---|---|---|---|---|
| Atherosclerosis (any/r | DC.LC.Sk | Myeloid | -7.66E-09 | 2.50E-08 | 0.620382757 |
| B Score | DC.LC.Sk | Myeloid | -4.82E-09 | 2.02E-08 | 0.594256084 |
| CAA | DC.LC.Sk | Myeloid | -3.77E-09 | 2.48E-08 | 0.560403136 |
| CAA (any/none) | DC.LC.Sk | Myeloid | -1.63E-08 | 2.40E-08 | 0.751084887 |
| CERAD (C Score) | DC.LC.Sk | Myeloid | 1.33E-08 | 2.04E-08 | 0.257905606 |
| Infarcts/lacunes | DC.LC.Sk | Myeloid | 3.75E-08 | 2.41E-08 | 0.059995494 |
| PD Braak | DC.LC.Sk | Myeloid | 2.88E-08 | 2.38E-08 | 0.112622375 |
| PD Braak (any/none) | DC.LC.Sk | Myeloid | 3.75E-08 | 2.44E-08 | 0.061774336 |
| TDP-43 (any/none) | DC.LC.Sk | Myeloid | -5.70E-08 | 5.84E-08 | 0.835252008 |
| Thal Phase | DC.LC.Sk | Myeloid | 8.59E-08 | 7.87E-08 | 0.137369967 |
| WMR (any/none) | DC.LC.Sk | Myeloid | -3.77E-08 | 3.84E-08 | 0.836765305 |
| #N/A | DC.LC.Sk | Myeloid | -4.65E-09 | 4.37E-08 | 0.542328387 |
| AD Braak | DC.pDC.8-.Sp | Myeloid | -1.75E-08 | 1.86E-08 | 0.826690795 |
| ADNC Score | DC.pDC.8-.Sp | Myeloid | -1.55E-07 | 5.38E-08 | 0.997964299 |
| AP (any/none) | DC.pDC.8-.Sp | Myeloid | -3.66E-09 | 2.15E-08 | 0.567495351 |
| Arteriolosclerosis | DC.pDC.8-.Sp | Myeloid | 1.41E-08 | 2.92E-08 | 0.313955805 |
| Arteriolosclerosis (any | DC.pDC.8-.Sp | Myeloid | 1.14E-08 | 2.96E-08 | 0.349862451 |
| Atherosclerosis | DC.pDC.8-.Sp | Myeloid | 6.51E-09 | 2.26E-08 | 0.386509652 |
| Atherosclerosis (any/r | DC.pDC.8-.Sp | Myeloid | -2.72E-08 | 2.25E-08 | 0.886595201 |
| B Score | DC.pDC.8-.Sp | Myeloid | -2.37E-08 | 1.79E-08 | 0.907741901 |
| CAA | DC.pDC.8-.Sp | Myeloid | 4.84E-09 | 2.58E-08 | 0.425604315 |
| CAA (any/none) | DC.pDC.8-.Sp | Myeloid | 1.72E-09 | 2.37E-08 | 0.47107684 |
| CERAD (C Score) | DC.pDC.8-.Sp | Myeloid | -2.43E-09 | 2.02E-08 | 0.547924971 |
| Infarcts/lacunes | DC.pDC.8-.Sp | Myeloid | 1.74E-08 | 2.17E-08 | 0.211207046 |
| PD Braak | DC.pDC.8-.Sp | Myeloid | 5.73E-08 | 2.00E-08 | 0.002081653 |
| PD Braak (any/none) | DC.pDC.8-.Sp | Myeloid | 5.31E-08 | 2.03E-08 | 0.004437301 |
| TDP-43 (any/none) | DC.pDC.8-.Sp | Myeloid | 7.97E-08 | 6.64E-08 | 0.115074975 |
| Thal Phase | DC.pDC.8-.Sp | Myeloid | -4.33E-08 | 7.18E-08 | 0.726704551 |
| WMR (any/none) | DC.pDC.8-.Sp | Myeloid | 1.84E-08 | 3.69E-08 | 0.308451129 |
| #N/A | DC.pDC.8-.Sp | Myeloid | 3.18E-08 | 4.56E-08 | 0.242497186 |
| AD Braak | DC.pDC.8+.MLN | Myeloid | -1.22E-08 | 1.96E-08 | 0.733367945 |
| ADNC Score | DC.pDC.8+.MLN | Myeloid | -1.48E-07 | 5.52E-08 | 0.996324044 |
| AP (any/none) | DC.pDC.8+.MLN | Myeloid | -1.98E-08 | 2.12E-08 | 0.824266017 |
| Arteriolosclerosis | DC.pDC.8+.MLN | Myeloid | -4.51E-09 | 2.92E-08 | 0.561473846 |

| | | | | | |
|---|---|---|---|---|---|
| Arteriolosclerosis (any | DC.pDC.8+.MLN | Myeloid | 5.47E-09 | 3.13E-08 | 0.430694493 |
| Atherosclerosis | DC.pDC.8+.MLN | Myeloid | 2.02E-08 | 2.33E-08 | 0.192776692 |
| Atherosclerosis (any/r | DC.pDC.8+.MLN | Myeloid | -3.13E-08 | 2.37E-08 | 0.906829728 |
| B Score | DC.pDC.8+.MLN | Myeloid | -1.74E-08 | 1.89E-08 | 0.821874862 |
| CAA | DC.pDC.8+.MLN | Myeloid | 1.29E-08 | 2.62E-08 | 0.310401116 |
| CAA (any/none) | DC.pDC.8+.MLN | Myeloid | 6.91E-09 | 2.35E-08 | 0.384625923 |
| CERAD (C Score) | DC.pDC.8+.MLN | Myeloid | 1.88E-08 | 2.03E-08 | 0.176315828 |
| Infarcts/lacunes | DC.pDC.8+.MLN | Myeloid | 3.10E-08 | 2.21E-08 | 0.079695343 |
| PD Braak | DC.pDC.8+.MLN | Myeloid | 3.77E-08 | 2.11E-08 | 0.036762381 |
| PD Braak (any/none) | DC.pDC.8+.MLN | Myeloid | 3.77E-08 | 2.02E-08 | 0.031312133 |
| TDP-43 (any/none) | DC.pDC.8+.MLN | Myeloid | 8.99E-08 | 5.83E-08 | 0.061634839 |
| Thal Phase | DC.pDC.8+.MLN | Myeloid | -4.16E-08 | 7.27E-08 | 0.71660015 |
| WMR (any/none) | DC.pDC.8+.MLN | Myeloid | 2.47E-08 | 3.78E-08 | 0.256675078 |
| #N/A | DC.pDC.8+.MLN | Myeloid | 1.92E-09 | 4.70E-08 | 0.483733716 |
| AD Braak | DC.pDC.8+.SLN | Myeloid | -2.52E-08 | 2.02E-08 | 0.894521617 |
| ADNC Score | DC.pDC.8+.SLN | Myeloid | -1.40E-07 | 5.51E-08 | 0.99449451 |
| AP (any/none) | DC.pDC.8+.SLN | Myeloid | -1.66E-08 | 2.11E-08 | 0.783857182 |
| Arteriolosclerosis | DC.pDC.8+.SLN | Myeloid | 7.28E-09 | 2.79E-08 | 0.397155799 |
| Arteriolosclerosis (any | DC.pDC.8+.SLN | Myeloid | 2.08E-08 | 3.13E-08 | 0.252924645 |
| Atherosclerosis | DC.pDC.8+.SLN | Myeloid | 1.57E-08 | 2.48E-08 | 0.263395582 |
| Atherosclerosis (any/r | DC.pDC.8+.SLN | Myeloid | -1.95E-08 | 2.41E-08 | 0.791086178 |
| B Score | DC.pDC.8+.SLN | Myeloid | -2.11E-08 | 1.92E-08 | 0.864434301 |
| CAA | DC.pDC.8+.SLN | Myeloid | -7.73E-09 | 2.60E-08 | 0.616780158 |
| CAA (any/none) | DC.pDC.8+.SLN | Myeloid | 4.08E-09 | 2.34E-08 | 0.430932949 |
| CERAD (C Score) | DC.pDC.8+.SLN | Myeloid | 1.51E-08 | 1.96E-08 | 0.220980268 |
| Infarcts/lacunes | DC.pDC.8+.SLN | Myeloid | 2.11E-08 | 2.11E-08 | 0.158668698 |
| PD Braak | DC.pDC.8+.SLN | Myeloid | 4.66E-08 | 2.04E-08 | 0.011106626 |
| PD Braak (any/none) | DC.pDC.8+.SLN | Myeloid | 4.71E-08 | 2.02E-08 | 0.009785589 |
| TDP-43 (any/none) | DC.pDC.8+.SLN | Myeloid | 9.15E-08 | 5.78E-08 | 0.056659223 |
| Thal Phase | DC.pDC.8+.SLN | Myeloid | -1.54E-08 | 7.04E-08 | 0.58638955 |
| WMR (any/none) | DC.pDC.8+.SLN | Myeloid | 4.58E-08 | 3.69E-08 | 0.107041101 |
| #N/A | DC.pDC.8+.SLN | Myeloid | -1.37E-08 | 4.56E-08 | 0.618376755 |
| AD Braak | DC.pDC.8+.Sp | Myeloid | -7.17E-09 | 1.87E-08 | 0.649202292 |
| ADNC Score | DC.pDC.8+.Sp | Myeloid | -1.22E-07 | 5.41E-08 | 0.98799533 |

| | | | | | |
|---|---|---|---|---|---|
| AP (any/none) | DC.pDC.8+.Sp | Myeloid | -1.38E-08 | 2.17E-08 | 0.738325191 |
| Arteriolosclerosis | DC.pDC.8+.Sp | Myeloid | 1.12E-08 | 2.86E-08 | 0.347489778 |
| Arteriolosclerosis (any | DC.pDC.8+.Sp | Myeloid | 9.05E-09 | 2.93E-08 | 0.378657977 |
| Atherosclerosis | DC.pDC.8+.Sp | Myeloid | 8.75E-09 | 2.24E-08 | 0.348356363 |
| Atherosclerosis (any/r | DC.pDC.8+.Sp | Myeloid | -1.87E-08 | 2.24E-08 | 0.798549862 |
| B Score | DC.pDC.8+.Sp | Myeloid | -9.06E-09 | 1.82E-08 | 0.690913567 |
| CAA | DC.pDC.8+.Sp | Myeloid | -8.13E-09 | 2.60E-08 | 0.622872053 |
| CAA (any/none) | DC.pDC.8+.Sp | Myeloid | -1.32E-09 | 2.32E-08 | 0.522781337 |
| CERAD (C Score) | DC.pDC.8+.Sp | Myeloid | 5.17E-09 | 2.01E-08 | 0.398628915 |
| Infarcts/lacunes | DC.pDC.8+.Sp | Myeloid | 3.18E-08 | 2.26E-08 | 0.080020525 |
| PD Braak | DC.pDC.8+.Sp | Myeloid | 5.88E-08 | 1.92E-08 | 0.001088987 |
| PD Braak (any/none) | DC.pDC.8+.Sp | Myeloid | 5.57E-08 | 1.93E-08 | 0.001968217 |
| TDP-43 (any/none) | DC.pDC.8+.Sp | Myeloid | 7.05E-08 | 6.45E-08 | 0.137225026 |
| Thal Phase | DC.pDC.8+.Sp | Myeloid | 4.20E-09 | 7.14E-08 | 0.476586073 |
| WMR (any/none) | DC.pDC.8+.Sp | Myeloid | 2.71E-08 | 3.59E-08 | 0.225052416 |
| #N/A | DC.pDC.8+.Sp | Myeloid | 4.98E-08 | 4.93E-08 | 0.156219264 |
| AD Braak | DN.SLN.CFA.d6.v2 | Myeloid | -1.83E-08 | 1.60E-08 | 0.873848619 |
| ADNC Score | DN.SLN.CFA.d6.v2 | Myeloid | -1.74E-08 | 5.19E-08 | 0.631195793 |
| AP (any/none) | DN.SLN.CFA.d6.v2 | Myeloid | -2.12E-08 | 1.76E-08 | 0.886339943 |
| Arteriolosclerosis | DN.SLN.CFA.d6.v2 | Myeloid | -3.87E-08 | 2.37E-08 | 0.948328112 |
| Arteriolosclerosis (any | DN.SLN.CFA.d6.v2 | Myeloid | -3.10E-08 | 2.59E-08 | 0.884136704 |
| Atherosclerosis | DN.SLN.CFA.d6.v2 | Myeloid | -1.15E-08 | 2.13E-08 | 0.705067097 |
| Atherosclerosis (any/r | DN.SLN.CFA.d6.v2 | Myeloid | -1.83E-08 | 2.02E-08 | 0.816735701 |
| B Score | DN.SLN.CFA.d6.v2 | Myeloid | -1.94E-08 | 1.62E-08 | 0.884073858 |
| CAA | DN.SLN.CFA.d6.v2 | Myeloid | 7.33E-09 | 2.06E-08 | 0.360825932 |
| CAA (any/none) | DN.SLN.CFA.d6.v2 | Myeloid | 7.31E-09 | 2.01E-08 | 0.357750291 |
| CERAD (C Score) | DN.SLN.CFA.d6.v2 | Myeloid | 1.19E-10 | 1.55E-08 | 0.49695587 |
| Infarcts/lacunes | DN.SLN.CFA.d6.v2 | Myeloid | 1.07E-08 | 2.23E-08 | 0.316067793 |
| PD Braak | DN.SLN.CFA.d6.v2 | Myeloid | -1.13E-08 | 1.90E-08 | 0.724207302 |
| PD Braak (any/none) | DN.SLN.CFA.d6.v2 | Myeloid | -1.40E-08 | 1.82E-08 | 0.779167732 |
| TDP-43 (any/none) | DN.SLN.CFA.d6.v2 | Myeloid | 6.75E-09 | 4.52E-08 | 0.44061309 |
| Thal Phase | DN.SLN.CFA.d6.v2 | Myeloid | 9.00E-08 | 6.32E-08 | 0.077324799 |
| WMR (any/none) | DN.SLN.CFA.d6.v2 | Myeloid | -4.98E-08 | 2.92E-08 | 0.955799649 |
| #N/A | DN.SLN.CFA.d6.v2 | Myeloid | -5.68E-08 | 3.43E-08 | 0.951107019 |

| | | | | | |
|---|---|---|---|---|---|
| AD Braak | DN.SLN.v2 | Myeloid | -1.09E-08 | 1.63E-08 | 0.747552558 |
| ADNC Score | DN.SLN.v2 | Myeloid | -1.09E-08 | 5.15E-08 | 0.583958441 |
| AP (any/none) | DN.SLN.v2 | Myeloid | -1.80E-08 | 1.73E-08 | 0.850231702 |
| Arteriolosclerosis | DN.SLN.v2 | Myeloid | -3.82E-08 | 2.44E-08 | 0.941148744 |
| Arteriolosclerosis (any | DN.SLN.v2 | Myeloid | -2.43E-08 | 2.58E-08 | 0.827169771 |
| Atherosclerosis | DN.SLN.v2 | Myeloid | 1.19E-09 | 2.15E-08 | 0.477905247 |
| Atherosclerosis (any/r | DN.SLN.v2 | Myeloid | -4.43E-09 | 2.17E-08 | 0.580781183 |
| B Score | DN.SLN.v2 | Myeloid | -1.91E-08 | 1.70E-08 | 0.86951116 |
| CAA | DN.SLN.v2 | Myeloid | 8.74E-09 | 2.18E-08 | 0.344435039 |
| CAA (any/none) | DN.SLN.v2 | Myeloid | 1.16E-08 | 2.08E-08 | 0.287706116 |
| CERAD (C Score) | DN.SLN.v2 | Myeloid | -9.49E-10 | 1.55E-08 | 0.524388553 |
| Infarcts/lacunes | DN.SLN.v2 | Myeloid | -1.60E-09 | 2.19E-08 | 0.529138021 |
| PD Braak | DN.SLN.v2 | Myeloid | -2.96E-09 | 1.95E-08 | 0.560301178 |
| PD Braak (any/none) | DN.SLN.v2 | Myeloid | -3.12E-09 | 1.89E-08 | 0.565650869 |
| TDP-43 (any/none) | DN.SLN.v2 | Myeloid | 4.13E-09 | 4.51E-08 | 0.463442291 |
| Thal Phase | DN.SLN.v2 | Myeloid | 9.79E-08 | 6.58E-08 | 0.068352711 |
| WMR (any/none) | DN.SLN.v2 | Myeloid | -4.19E-08 | 2.87E-08 | 0.927841446 |
| #N/A | DN.SLN.v2 | Myeloid | -6.28E-08 | 3.64E-08 | 0.957929703 |
| AD Braak | EO.AT.v2 | Myeloid | 3.04E-09 | 2.21E-08 | 0.445311273 |
| ADNC Score | EO.AT.v2 | Myeloid | 2.85E-09 | 6.70E-08 | 0.483004441 |
| AP (any/none) | EO.AT.v2 | Myeloid | 1.90E-08 | 2.18E-08 | 0.19213422 |
| Arteriolosclerosis | EO.AT.v2 | Myeloid | 4.34E-09 | 2.40E-08 | 0.428163037 |
| Arteriolosclerosis (any | EO.AT.v2 | Myeloid | -1.88E-08 | 2.71E-08 | 0.755753712 |
| Atherosclerosis | EO.AT.v2 | Myeloid | -3.65E-08 | 2.16E-08 | 0.95456851 |
| Atherosclerosis (any/r | EO.AT.v2 | Myeloid | -9.46E-09 | 2.27E-08 | 0.661318327 |
| B Score | EO.AT.v2 | Myeloid | 7.14E-09 | 2.07E-08 | 0.365184747 |
| CAA | EO.AT.v2 | Myeloid | -1.07E-08 | 2.25E-08 | 0.682870522 |
| CAA (any/none) | EO.AT.v2 | Myeloid | -1.45E-08 | 2.12E-08 | 0.753806834 |
| CERAD (C Score) | EO.AT.v2 | Myeloid | -2.80E-09 | 2.12E-08 | 0.552408573 |
| Infarcts/lacunes | EO.AT.v2 | Myeloid | -1.33E-08 | 2.21E-08 | 0.727446036 |
| PD Braak | EO.AT.v2 | Myeloid | 4.46E-08 | 2.14E-08 | 0.018570725 |
| PD Braak (any/none) | EO.AT.v2 | Myeloid | 4.96E-08 | 2.14E-08 | 0.010297182 |
| TDP-43 (any/none) | EO.AT.v2 | Myeloid | -1.16E-07 | 5.04E-08 | 0.989066722 |
| Thal Phase | EO.AT.v2 | Myeloid | -6.26E-08 | 7.45E-08 | 0.799406702 |

| | | | | | |
|---|---|---|---|---|---|
| WMR (any/none) | EO.AT.v2 | Myeloid | -2.66E-08 | 3.39E-08 | 0.783929087 |
| #N/A | EO.AT.v2 | Myeloid | -2.94E-08 | 4.40E-08 | 0.748489951 |
| AD Braak | Eo.BL.v2 | Myeloid | -2.66E-08 | 1.74E-08 | 0.936483515 |
| ADNC Score | Eo.BL.v2 | Myeloid | -1.05E-09 | 4.90E-08 | 0.508565525 |
| AP (any/none) | Eo.BL.v2 | Myeloid | -1.87E-09 | 1.90E-08 | 0.539302531 |
| Arteriolosclerosis | Eo.BL.v2 | Myeloid | 3.33E-08 | 2.39E-08 | 0.082306417 |
| Arteriolosclerosis (any | Eo.BL.v2 | Myeloid | 2.77E-08 | 2.55E-08 | 0.138469306 |
| Atherosclerosis | Eo.BL.v2 | Myeloid | 4.56E-10 | 2.08E-08 | 0.491254678 |
| Atherosclerosis (any/r | Eo.BL.v2 | Myeloid | -7.17E-09 | 2.12E-08 | 0.632706378 |
| B Score | Eo.BL.v2 | Myeloid | -1.91E-08 | 1.61E-08 | 0.881968749 |
| CAA | Eo.BL.v2 | Myeloid | -2.57E-08 | 2.23E-08 | 0.875333555 |
| CAA (any/none) | Eo.BL.v2 | Myeloid | -3.03E-08 | 2.06E-08 | 0.92952785 |
| CERAD (C Score) | Eo.BL.v2 | Myeloid | -2.58E-08 | 1.56E-08 | 0.950866215 |
| Infarcts/lacunes | Eo.BL.v2 | Myeloid | 2.81E-08 | 2.08E-08 | 0.089166966 |
| PD Braak | Eo.BL.v2 | Myeloid | 5.69E-09 | 2.03E-08 | 0.389322322 |
| PD Braak (any/none) | Eo.BL.v2 | Myeloid | 1.92E-08 | 1.92E-08 | 0.159032049 |
| TDP-43 (any/none) | Eo.BL.v2 | Myeloid | -4.63E-08 | 5.42E-08 | 0.803503561 |
| Thal Phase | Eo.BL.v2 | Myeloid | -9.64E-08 | 6.00E-08 | 0.945848354 |
| WMR (any/none) | Eo.BL.v2 | Myeloid | -2.11E-08 | 3.22E-08 | 0.743828866 |
| #N/A | Eo.BL.v2 | Myeloid | 2.73E-09 | 3.87E-08 | 0.4718817 |
| AD Braak | Ep.5wk.MEC.Sca1+.Th | Myeloid | -1.73E-08 | 1.51E-08 | 0.874644735 |
| ADNC Score | Ep.5wk.MEC.Sca1+.Th | Myeloid | 1.95E-08 | 5.18E-08 | 0.353356516 |
| AP (any/none) | Ep.5wk.MEC.Sca1+.Th | Myeloid | 1.52E-08 | 1.89E-08 | 0.210582678 |
| Arteriolosclerosis | Ep.5wk.MEC.Sca1+.Th | Myeloid | -1.87E-09 | 2.60E-08 | 0.52878195 |
| Arteriolosclerosis (any | Ep.5wk.MEC.Sca1+.Th | Myeloid | -1.30E-10 | 2.89E-08 | 0.501791636 |
| Atherosclerosis | Ep.5wk.MEC.Sca1+.Th | Myeloid | 2.19E-08 | 2.12E-08 | 0.15014189 |
| Atherosclerosis (any/r | Ep.5wk.MEC.Sca1+.Th | Myeloid | 1.21E-08 | 2.20E-08 | 0.291564044 |
| B Score | Ep.5wk.MEC.Sca1+.Th | Myeloid | -2.64E-08 | 1.55E-08 | 0.95531388 |
| CAA | Ep.5wk.MEC.Sca1+.Th | Myeloid | -1.82E-08 | 2.18E-08 | 0.797908867 |
| CAA (any/none) | Ep.5wk.MEC.Sca1+.Th | Myeloid | 4.28E-09 | 2.07E-08 | 0.417903665 |
| CERAD (C Score) | Ep.5wk.MEC.Sca1+.Th | Myeloid | 7.09E-09 | 1.66E-08 | 0.334982309 |
| Infarcts/lacunes | Ep.5wk.MEC.Sca1+.Th | Myeloid | -1.04E-08 | 2.21E-08 | 0.681387706 |
| PD Braak | Ep.5wk.MEC.Sca1+.Th | Myeloid | 1.31E-08 | 1.83E-08 | 0.236225814 |
| PD Braak (any/none) | Ep.5wk.MEC.Sca1+.Th | Myeloid | 6.51E-09 | 1.80E-08 | 0.358708364 |

| | | | | | |
|---|---|---|---|---|---|
| TDP-43 (any/none) | Ep.5wk.MEC.Sca1+.Th | Myeloid | 1.41E-08 | 4.91E-08 | 0.38727619 |
| Thal Phase | Ep.5wk.MEC.Sca1+.Th | Myeloid | 6.29E-08 | 6.60E-08 | 0.170404134 |
| WMR (any/none) | Ep.5wk.MEC.Sca1+.Th | Myeloid | -6.49E-08 | 2.87E-08 | 0.988184658 |
| #N/A | Ep.5wk.MEC.Sca1+.Th | Myeloid | -6.28E-08 | 3.34E-08 | 0.970027003 |
| AD Braak | Ep.5wk.MEChi.Th | Myeloid | -1.65E-08 | 1.56E-08 | 0.856083092 |
| ADNC Score | Ep.5wk.MEChi.Th | Myeloid | -2.48E-08 | 5.07E-08 | 0.687905699 |
| AP (any/none) | Ep.5wk.MEChi.Th | Myeloid | -2.07E-09 | 1.85E-08 | 0.544452897 |
| Arteriolosclerosis | Ep.5wk.MEChi.Th | Myeloid | -8.42E-09 | 2.31E-08 | 0.642369151 |
| Arteriolosclerosis (any | Ep.5wk.MEChi.Th | Myeloid | 1.17E-09 | 2.55E-08 | 0.481723317 |
| Atherosclerosis | Ep.5wk.MEChi.Th | Myeloid | -2.20E-08 | 1.90E-08 | 0.87681562 |
| Atherosclerosis (any/r | Ep.5wk.MEChi.Th | Myeloid | -2.77E-08 | 1.98E-08 | 0.919587465 |
| B Score | Ep.5wk.MEChi.Th | Myeloid | -2.84E-08 | 1.54E-08 | 0.966825253 |
| CAA | Ep.5wk.MEChi.Th | Myeloid | -2.37E-09 | 2.12E-08 | 0.544498962 |
| CAA (any/none) | Ep.5wk.MEChi.Th | Myeloid | 3.00E-08 | 1.98E-08 | 0.064468609 |
| CERAD (C Score) | Ep.5wk.MEChi.Th | Myeloid | -1.89E-08 | 1.49E-08 | 0.897773335 |
| Infarcts/lacunes | Ep.5wk.MEChi.Th | Myeloid | 1.45E-09 | 2.08E-08 | 0.472177692 |
| PD Braak | Ep.5wk.MEChi.Th | Myeloid | 4.00E-09 | 1.72E-08 | 0.408236611 |
| PD Braak (any/none) | Ep.5wk.MEChi.Th | Myeloid | -3.80E-09 | 1.74E-08 | 0.586415989 |
| TDP-43 (any/none) | Ep.5wk.MEChi.Th | Myeloid | 1.46E-08 | 5.30E-08 | 0.391809231 |
| Thal Phase | Ep.5wk.MEChi.Th | Myeloid | 8.97E-08 | 7.11E-08 | 0.103437878 |
| WMR (any/none) | Ep.5wk.MEChi.Th | Myeloid | -3.18E-08 | 2.90E-08 | 0.863991787 |
| #N/A | Ep.5wk.MEChi.Th | Myeloid | -2.67E-08 | 3.75E-08 | 0.761708089 |
| AD Braak | Ep.5wk.MEClo.Th | Myeloid | -2.30E-08 | 1.49E-08 | 0.939156596 |
| ADNC Score | Ep.5wk.MEClo.Th | Myeloid | -4.00E-08 | 4.74E-08 | 0.800152362 |
| AP (any/none) | Ep.5wk.MEClo.Th | Myeloid | -1.23E-08 | 1.80E-08 | 0.751815354 |
| Arteriolosclerosis | Ep.5wk.MEClo.Th | Myeloid | -1.78E-08 | 2.31E-08 | 0.779425228 |
| Arteriolosclerosis (any | Ep.5wk.MEClo.Th | Myeloid | 1.56E-09 | 2.69E-08 | 0.476864971 |
| Atherosclerosis | Ep.5wk.MEClo.Th | Myeloid | -1.59E-08 | 1.97E-08 | 0.790440924 |
| Atherosclerosis (any/r | Ep.5wk.MEClo.Th | Myeloid | -1.68E-08 | 1.89E-08 | 0.813173746 |
| B Score | Ep.5wk.MEClo.Th | Myeloid | -3.16E-08 | 1.54E-08 | 0.980037955 |
| CAA | Ep.5wk.MEClo.Th | Myeloid | -1.97E-08 | 1.98E-08 | 0.840974415 |
| CAA (any/none) | Ep.5wk.MEClo.Th | Myeloid | -4.71E-09 | 1.98E-08 | 0.593940682 |
| CERAD (C Score) | Ep.5wk.MEClo.Th | Myeloid | -1.63E-08 | 1.45E-08 | 0.869542514 |
| Infarcts/lacunes | Ep.5wk.MEClo.Th | Myeloid | -1.50E-08 | 2.07E-08 | 0.764983679 |

| | | | | | |
|---|---|---|---|---|---|
| PD Braak | Ep.5wk.MEClo.Th | Myeloid | 1.49E-08 | 1.73E-08 | 0.193549682 |
| PD Braak (any/none) | Ep.5wk.MEClo.Th | Myeloid | 8.72E-09 | 1.74E-08 | 0.308153683 |
| TDP-43 (any/none) | Ep.5wk.MEClo.Th | Myeloid | -2.69E-08 | 4.27E-08 | 0.735533242 |
| Thal Phase | Ep.5wk.MEClo.Th | Myeloid | 2.78E-08 | 6.71E-08 | 0.339437498 |
| WMR (any/none) | Ep.5wk.MEClo.Th | Myeloid | -9.02E-08 | 2.90E-08 | 0.999055846 |
| #N/A | Ep.5wk.MEClo.Th | Myeloid | -3.48E-08 | 3.23E-08 | 0.859233605 |
| AD Braak | Ep.8wk.CEC.Sca1+.Th | Myeloid | 3.04E-09 | 1.50E-08 | 0.419655023 |
| ADNC Score | Ep.8wk.CEC.Sca1+.Th | Myeloid | 4.21E-08 | 5.47E-08 | 0.220463992 |
| AP (any/none) | Ep.8wk.CEC.Sca1+.Th | Myeloid | 6.43E-09 | 1.86E-08 | 0.365042019 |
| Arteriolosclerosis | Ep.8wk.CEC.Sca1+.Th | Myeloid | -2.74E-08 | 2.42E-08 | 0.870738024 |
| Arteriolosclerosis (any | Ep.8wk.CEC.Sca1+.Th | Myeloid | -3.60E-08 | 2.83E-08 | 0.898459677 |
| Atherosclerosis | Ep.8wk.CEC.Sca1+.Th | Myeloid | 1.15E-08 | 2.20E-08 | 0.300877022 |
| Atherosclerosis (any/r | Ep.8wk.CEC.Sca1+.Th | Myeloid | -1.70E-08 | 2.10E-08 | 0.790676982 |
| B Score | Ep.8wk.CEC.Sca1+.Th | Myeloid | -6.33E-09 | 1.62E-08 | 0.651937719 |
| CAA | Ep.8wk.CEC.Sca1+.Th | Myeloid | -6.43E-09 | 2.06E-08 | 0.622361009 |
| CAA (any/none) | Ep.8wk.CEC.Sca1+.Th | Myeloid | 1.20E-08 | 2.13E-08 | 0.287418267 |
| CERAD (C Score) | Ep.8wk.CEC.Sca1+.Th | Myeloid | 1.02E-08 | 1.67E-08 | 0.27020462 |
| Infarcts/lacunes | Ep.8wk.CEC.Sca1+.Th | Myeloid | -3.80E-09 | 1.97E-08 | 0.576333974 |
| PD Braak | Ep.8wk.CEC.Sca1+.Th | Myeloid | -8.18E-09 | 1.84E-08 | 0.671558618 |
| PD Braak (any/none) | Ep.8wk.CEC.Sca1+.Th | Myeloid | -8.28E-09 | 1.77E-08 | 0.680324305 |
| TDP-43 (any/none) | Ep.8wk.CEC.Sca1+.Th | Myeloid | 6.20E-08 | 4.71E-08 | 0.094057184 |
| Thal Phase | Ep.8wk.CEC.Sca1+.Th | Myeloid | 8.69E-08 | 6.60E-08 | 0.093979636 |
| WMR (any/none) | Ep.8wk.CEC.Sca1+.Th | Myeloid | -2.21E-08 | 2.94E-08 | 0.77381601 |
| #N/A | Ep.8wk.CEC.Sca1+.Th | Myeloid | -6.96E-08 | 3.18E-08 | 0.985752186 |
| AD Braak | Ep.8wk.CEChi.Th | Myeloid | -4.58E-09 | 1.57E-08 | 0.614749487 |
| ADNC Score | Ep.8wk.CEChi.Th | Myeloid | 6.31E-08 | 5.70E-08 | 0.134080771 |
| AP (any/none) | Ep.8wk.CEChi.Th | Myeloid | 1.85E-08 | 2.07E-08 | 0.185920845 |
| Arteriolosclerosis | Ep.8wk.CEChi.Th | Myeloid | -4.08E-08 | 2.25E-08 | 0.96502629 |
| Arteriolosclerosis (any | Ep.8wk.CEChi.Th | Myeloid | -5.12E-08 | 2.71E-08 | 0.970469005 |
| Atherosclerosis | Ep.8wk.CEChi.Th | Myeloid | -2.07E-08 | 2.22E-08 | 0.823522188 |
| Atherosclerosis (any/r | Ep.8wk.CEChi.Th | Myeloid | -2.88E-08 | 2.29E-08 | 0.895221704 |
| B Score | Ep.8wk.CEChi.Th | Myeloid | -5.79E-09 | 1.67E-08 | 0.63587463 |
| CAA | Ep.8wk.CEChi.Th | Myeloid | -5.50E-09 | 2.10E-08 | 0.603034695 |
| CAA (any/none) | Ep.8wk.CEChi.Th | Myeloid | 4.76E-09 | 2.31E-08 | 0.418437255 |

| | | | | | |
|---|---|---|---|---|---|
| CERAD (C Score) | Ep.8wk.CEChi.Th | Myeloid | -7.05E-10 | 1.59E-08 | 0.517673581 |
| Infarcts/lacunes | Ep.8wk.CEChi.Th | Myeloid | 1.46E-08 | 2.07E-08 | 0.240210398 |
| PD Braak | Ep.8wk.CEChi.Th | Myeloid | 1.66E-08 | 1.81E-08 | 0.17963475 |
| PD Braak (any/none) | Ep.8wk.CEChi.Th | Myeloid | 1.16E-08 | 1.76E-08 | 0.254681988 |
| TDP-43 (any/none) | Ep.8wk.CEChi.Th | Myeloid | -1.68E-08 | 4.48E-08 | 0.645739781 |
| Thal Phase | Ep.8wk.CEChi.Th | Myeloid | 4.62E-08 | 6.61E-08 | 0.242170905 |
| WMR (any/none) | Ep.8wk.CEChi.Th | Myeloid | -7.58E-08 | 3.12E-08 | 0.992530204 |
| #N/A | Ep.8wk.CEChi.Th | Myeloid | -8.44E-08 | 3.78E-08 | 0.987312254 |
| AD Braak | Ep.8wk.MEChi.Th | Myeloid | -2.03E-08 | 1.65E-08 | 0.89060498 |
| ADNC Score | Ep.8wk.MEChi.Th | Myeloid | -1.96E-08 | 5.07E-08 | 0.650695039 |
| AP (any/none) | Ep.8wk.MEChi.Th | Myeloid | 1.22E-09 | 1.76E-08 | 0.472289887 |
| Arteriolosclerosis | Ep.8wk.MEChi.Th | Myeloid | -1.56E-08 | 2.20E-08 | 0.760658102 |
| Arteriolosclerosis (any | Ep.8wk.MEChi.Th | Myeloid | 4.49E-11 | 2.55E-08 | 0.499295474 |
| Atherosclerosis | Ep.8wk.MEChi.Th | Myeloid | -1.87E-08 | 1.99E-08 | 0.826340079 |
| Atherosclerosis (any/r | Ep.8wk.MEChi.Th | Myeloid | -2.16E-08 | 1.98E-08 | 0.862205289 |
| B Score | Ep.8wk.MEChi.Th | Myeloid | -2.95E-08 | 1.61E-08 | 0.966733668 |
| CAA | Ep.8wk.MEChi.Th | Myeloid | -2.55E-09 | 2.14E-08 | 0.547370503 |
| CAA (any/none) | Ep.8wk.MEChi.Th | Myeloid | 3.00E-08 | 1.96E-08 | 0.062519687 |
| CERAD (C Score) | Ep.8wk.MEChi.Th | Myeloid | -2.06E-08 | 1.50E-08 | 0.915107786 |
| Infarcts/lacunes | Ep.8wk.MEChi.Th | Myeloid | -5.18E-09 | 2.02E-08 | 0.601160022 |
| PD Braak | Ep.8wk.MEChi.Th | Myeloid | 1.68E-08 | 1.80E-08 | 0.175279403 |
| PD Braak (any/none) | Ep.8wk.MEChi.Th | Myeloid | 1.69E-08 | 1.85E-08 | 0.180956255 |
| TDP-43 (any/none) | Ep.8wk.MEChi.Th | Myeloid | 2.66E-08 | 5.40E-08 | 0.310884078 |
| Thal Phase | Ep.8wk.MEChi.Th | Myeloid | 5.15E-08 | 7.11E-08 | 0.234580608 |
| WMR (any/none) | Ep.8wk.MEChi.Th | Myeloid | -3.81E-08 | 2.75E-08 | 0.916912522 |
| #N/A | Ep.8wk.MEChi.Th | Myeloid | 6.89E-09 | 3.67E-08 | 0.425624964 |
| AD Braak | Ep.8wk.MEClo.Th | Myeloid | -7.74E-09 | 1.44E-08 | 0.704895689 |
| ADNC Score | Ep.8wk.MEClo.Th | Myeloid | -4.35E-08 | 4.84E-08 | 0.815601661 |
| AP (any/none) | Ep.8wk.MEClo.Th | Myeloid | -9.79E-09 | 1.90E-08 | 0.697132535 |
| Arteriolosclerosis | Ep.8wk.MEClo.Th | Myeloid | -1.67E-08 | 2.55E-08 | 0.743563182 |
| Arteriolosclerosis (any | Ep.8wk.MEClo.Th | Myeloid | -6.55E-09 | 2.85E-08 | 0.590803775 |
| Atherosclerosis | Ep.8wk.MEClo.Th | Myeloid | -1.61E-08 | 2.05E-08 | 0.783809686 |
| Atherosclerosis (any/r | Ep.8wk.MEClo.Th | Myeloid | -9.95E-09 | 2.02E-08 | 0.688573016 |
| B Score | Ep.8wk.MEClo.Th | Myeloid | -2.03E-08 | 1.56E-08 | 0.904247414 |

| | | | | | |
|---|---|---|---|---|---|
| CAA | Ep.8wk.MEClo.Th | Myeloid | -1.18E-08 | 2.08E-08 | 0.714261617 |
| CAA (any/none) | Ep.8wk.MEClo.Th | Myeloid | 4.21E-09 | 2.04E-08 | 0.418327915 |
| CERAD (C Score) | Ep.8wk.MEClo.Th | Myeloid | -2.32E-08 | 1.47E-08 | 0.942692307 |
| Infarcts/lacunes | Ep.8wk.MEClo.Th | Myeloid | -2.08E-08 | 2.21E-08 | 0.82653263 |
| PD Braak | Ep.8wk.MEClo.Th | Myeloid | 1.82E-08 | 1.76E-08 | 0.15028788 |
| PD Braak (any/none) | Ep.8wk.MEClo.Th | Myeloid | 1.14E-08 | 1.77E-08 | 0.259194579 |
| TDP-43 (any/none) | Ep.8wk.MEClo.Th | Myeloid | 9.00E-09 | 4.56E-08 | 0.421785419 |
| Thal Phase | Ep.8wk.MEClo.Th | Myeloid | 4.73E-09 | 6.47E-08 | 0.470870379 |
| WMR (any/none) | Ep.8wk.MEClo.Th | Myeloid | -9.51E-08 | 2.85E-08 | 0.999574909 |
| #N/A | Ep.8wk.MEClo.Th | Myeloid | -3.33E-08 | 3.55E-08 | 0.825875786 |
| AD Braak | Ep.MEChi.Th | Myeloid | -1.24E-08 | 1.56E-08 | 0.786683023 |
| ADNC Score | Ep.MEChi.Th | Myeloid | -3.74E-08 | 5.33E-08 | 0.758717625 |
| AP (any/none) | Ep.MEChi.Th | Myeloid | 2.08E-10 | 1.84E-08 | 0.495490198 |
| Arteriolosclerosis | Ep.MEChi.Th | Myeloid | -6.57E-09 | 2.26E-08 | 0.614368978 |
| Arteriolosclerosis (any | Ep.MEChi.Th | Myeloid | 2.82E-08 | 2.71E-08 | 0.149744072 |
| Atherosclerosis | Ep.MEChi.Th | Myeloid | -1.68E-08 | 1.92E-08 | 0.808724819 |
| Atherosclerosis (any/r | Ep.MEChi.Th | Myeloid | -1.36E-08 | 2.05E-08 | 0.746895914 |
| B Score | Ep.MEChi.Th | Myeloid | -1.56E-08 | 1.53E-08 | 0.846132421 |
| CAA | Ep.MEChi.Th | Myeloid | -3.53E-08 | 2.04E-08 | 0.957922274 |
| CAA (any/none) | Ep.MEChi.Th | Myeloid | -8.15E-09 | 2.07E-08 | 0.652959372 |
| CERAD (C Score) | Ep.MEChi.Th | Myeloid | -9.48E-09 | 1.58E-08 | 0.72591865 |
| Infarcts/lacunes | Ep.MEChi.Th | Myeloid | -2.98E-08 | 2.20E-08 | 0.912631846 |
| PD Braak | Ep.MEChi.Th | Myeloid | 2.21E-08 | 1.74E-08 | 0.1026931 |
| PD Braak (any/none) | Ep.MEChi.Th | Myeloid | 1.03E-08 | 1.69E-08 | 0.2705759 |
| TDP-43 (any/none) | Ep.MEChi.Th | Myeloid | 1.00E-08 | 5.20E-08 | 0.423436883 |
| Thal Phase | Ep.MEChi.Th | Myeloid | 6.88E-08 | 6.36E-08 | 0.139881282 |
| WMR (any/none) | Ep.MEChi.Th | Myeloid | -3.56E-08 | 3.05E-08 | 0.878304398 |
| #N/A | Ep.MEChi.Th | Myeloid | 1.36E-08 | 4.04E-08 | 0.368299078 |
| AD Braak | Fi.MTS15+.Th | Myeloid | -9.20E-09 | 1.51E-08 | 0.728859929 |
| ADNC Score | Fi.MTS15+.Th | Myeloid | 6.11E-08 | 4.93E-08 | 0.107670288 |
| AP (any/none) | Fi.MTS15+.Th | Myeloid | -2.41E-09 | 1.74E-08 | 0.554961249 |
| Arteriolosclerosis | Fi.MTS15+.Th | Myeloid | 1.10E-08 | 2.58E-08 | 0.334538861 |
| Arteriolosclerosis (any | Fi.MTS15+.Th | Myeloid | -2.17E-08 | 2.72E-08 | 0.788116354 |
| Atherosclerosis | Fi.MTS15+.Th | Myeloid | -3.49E-09 | 2.07E-08 | 0.567064822 |

| | | | | | |
|---|---|---|---|---|---|
| Atherosclerosis (any/r | Fi.MTS15+.Th | Myeloid | -1.93E-09 | 1.98E-08 | 0.538692615 |
| B Score | Fi.MTS15+.Th | Myeloid | -3.48E-09 | 1.59E-08 | 0.586285707 |
| CAA | Fi.MTS15+.Th | Myeloid | -1.08E-09 | 1.99E-08 | 0.521591099 |
| CAA (any/none) | Fi.MTS15+.Th | Myeloid | 1.47E-08 | 1.99E-08 | 0.230061004 |
| CERAD (C Score) | Fi.MTS15+.Th | Myeloid | 3.57E-08 | 1.62E-08 | 0.013929269 |
| Infarcts/lacunes | Fi.MTS15+.Th | Myeloid | 3.04E-09 | 2.09E-08 | 0.442209648 |
| PD Braak | Fi.MTS15+.Th | Myeloid | -5.66E-09 | 1.77E-08 | 0.625358147 |
| PD Braak (any/none) | Fi.MTS15+.Th | Myeloid | -7.42E-09 | 1.71E-08 | 0.667834187 |
| TDP-43 (any/none) | Fi.MTS15+.Th | Myeloid | 5.18E-08 | 5.39E-08 | 0.168402777 |
| Thal Phase | Fi.MTS15+.Th | Myeloid | 8.55E-08 | 7.12E-08 | 0.114779012 |
| WMR (any/none) | Fi.MTS15+.Th | Myeloid | -6.01E-08 | 3.01E-08 | 0.977007338 |
| #N/A | Fi.MTS15+.Th | Myeloid | -3.63E-08 | 3.74E-08 | 0.834040509 |
| AD Braak | Fi.Sk | Myeloid | 5.49E-09 | 1.73E-08 | 0.375390022 |
| ADNC Score | Fi.Sk | Myeloid | 1.04E-07 | 5.62E-08 | 0.031930771 |
| AP (any/none) | Fi.Sk | Myeloid | 5.46E-09 | 1.87E-08 | 0.385362648 |
| Arteriolosclerosis | Fi.Sk | Myeloid | -1.24E-08 | 2.64E-08 | 0.681307484 |
| Arteriolosclerosis (any | Fi.Sk | Myeloid | -5.99E-08 | 2.64E-08 | 0.98837133 |
| Atherosclerosis | Fi.Sk | Myeloid | 2.20E-08 | 2.10E-08 | 0.147338427 |
| Atherosclerosis (any/r | Fi.Sk | Myeloid | -6.70E-09 | 2.06E-08 | 0.627835222 |
| B Score | Fi.Sk | Myeloid | 1.02E-08 | 1.73E-08 | 0.278308899 |
| CAA | Fi.Sk | Myeloid | -7.64E-09 | 2.15E-08 | 0.63910513 |
| CAA (any/none) | Fi.Sk | Myeloid | 2.67E-08 | 2.25E-08 | 0.117981673 |
| CERAD (C Score) | Fi.Sk | Myeloid | 2.08E-08 | 1.73E-08 | 0.114185403 |
| Infarcts/lacunes | Fi.Sk | Myeloid | -5.44E-09 | 2.24E-08 | 0.595874721 |
| PD Braak | Fi.Sk | Myeloid | -9.99E-09 | 1.96E-08 | 0.695038092 |
| PD Braak (any/none) | Fi.Sk | Myeloid | -8.34E-09 | 1.95E-08 | 0.665267095 |
| TDP-43 (any/none) | Fi.Sk | Myeloid | 8.12E-08 | 5.92E-08 | 0.085325321 |
| Thal Phase | Fi.Sk | Myeloid | 1.32E-07 | 6.82E-08 | 0.026294928 |
| WMR (any/none) | Fi.Sk | Myeloid | -4.08E-08 | 3.23E-08 | 0.896907512 |
| #N/A | Fi.Sk | Myeloid | -2.49E-08 | 4.02E-08 | 0.732126283 |
| AD Braak | FRC.Cad11.WT.v2 | Myeloid | -1.76E-08 | 1.57E-08 | 0.86890618 |
| ADNC Score | FRC.Cad11.WT.v2 | Myeloid | 2.77E-08 | 5.54E-08 | 0.308745484 |
| AP (any/none) | FRC.Cad11.WT.v2 | Myeloid | -7.54E-09 | 1.82E-08 | 0.660582097 |
| Arteriolosclerosis | FRC.Cad11.WT.v2 | Myeloid | -3.75E-08 | 2.40E-08 | 0.941090849 |

| | | | | | |
|---|---|---|---|---|---|
| Arteriolosclerosis (any | FRC.Cad11.WT.v2 | Myeloid | -6.23E-08 | 2.71E-08 | 0.989307863 |
| Atherosclerosis | FRC.Cad11.WT.v2 | Myeloid | 1.36E-08 | 2.26E-08 | 0.274629363 |
| Atherosclerosis (any/r | FRC.Cad11.WT.v2 | Myeloid | 7.89E-09 | 2.06E-08 | 0.350506015 |
| B Score | FRC.Cad11.WT.v2 | Myeloid | -2.95E-08 | 1.65E-08 | 0.962732421 |
| CAA | FRC.Cad11.WT.v2 | Myeloid | -7.66E-10 | 2.22E-08 | 0.513752558 |
| CAA (any/none) | FRC.Cad11.WT.v2 | Myeloid | 1.41E-08 | 2.40E-08 | 0.278228338 |
| CERAD (C Score) | FRC.Cad11.WT.v2 | Myeloid | 2.46E-08 | 1.68E-08 | 0.07155907 |
| Infarcts/lacunes | FRC.Cad11.WT.v2 | Myeloid | -2.34E-09 | 2.35E-08 | 0.539572609 |
| PD Braak | FRC.Cad11.WT.v2 | Myeloid | 8.41E-09 | 1.86E-08 | 0.32526564 |
| PD Braak (any/none) | FRC.Cad11.WT.v2 | Myeloid | 3.58E-10 | 1.81E-08 | 0.492111722 |
| TDP-43 (any/none) | FRC.Cad11.WT.v2 | Myeloid | 4.23E-08 | 4.87E-08 | 0.1927576 |
| Thal Phase | FRC.Cad11.WT.v2 | Myeloid | 4.83E-08 | 6.34E-08 | 0.223258109 |
| WMR (any/none) | FRC.Cad11.WT.v2 | Myeloid | -6.21E-08 | 3.00E-08 | 0.9809368 |
| #N/A | FRC.Cad11.WT.v2 | Myeloid | -9.80E-08 | 3.55E-08 | 0.997079176 |
| AD Braak | FRC.MLN | Myeloid | -1.42E-08 | 1.56E-08 | 0.818539872 |
| ADNC Score | FRC.MLN | Myeloid | 5.61E-08 | 5.51E-08 | 0.154096723 |
| AP (any/none) | FRC.MLN | Myeloid | 5.95E-09 | 2.03E-08 | 0.38472077 |
| Arteriolosclerosis | FRC.MLN | Myeloid | -1.87E-08 | 2.44E-08 | 0.777869712 |
| Arteriolosclerosis (any | FRC.MLN | Myeloid | -5.07E-08 | 2.89E-08 | 0.960427865 |
| Atherosclerosis | FRC.MLN | Myeloid | -7.21E-09 | 2.43E-08 | 0.616726928 |
| Atherosclerosis (any/r | FRC.MLN | Myeloid | -7.59E-09 | 2.13E-08 | 0.638946568 |
| B Score | FRC.MLN | Myeloid | -1.74E-08 | 1.64E-08 | 0.856256113 |
| CAA | FRC.MLN | Myeloid | -1.46E-08 | 2.21E-08 | 0.745513826 |
| CAA (any/none) | FRC.MLN | Myeloid | 1.36E-08 | 2.38E-08 | 0.283717884 |
| CERAD (C Score) | FRC.MLN | Myeloid | 2.15E-08 | 1.76E-08 | 0.111653097 |
| Infarcts/lacunes | FRC.MLN | Myeloid | -3.01E-08 | 2.21E-08 | 0.913298984 |
| PD Braak | FRC.MLN | Myeloid | -1.44E-08 | 1.94E-08 | 0.771215057 |
| PD Braak (any/none) | FRC.MLN | Myeloid | -1.09E-08 | 1.93E-08 | 0.71324615 |
| TDP-43 (any/none) | FRC.MLN | Myeloid | 2.47E-08 | 5.29E-08 | 0.320340039 |
| Thal Phase | FRC.MLN | Myeloid | 7.81E-08 | 6.76E-08 | 0.124002256 |
| WMR (any/none) | FRC.MLN | Myeloid | -7.13E-08 | 3.19E-08 | 0.987327352 |
| #N/A | FRC.MLN | Myeloid | -6.94E-08 | 3.99E-08 | 0.958796113 |
| AD Braak | FRC.SLN | Myeloid | -1.42E-08 | 1.69E-08 | 0.800038586 |
| ADNC Score | FRC.SLN | Myeloid | 6.37E-08 | 5.35E-08 | 0.116852267 |

| | | | | | |
|---|---|---|---|---|---|
| AP (any/none) | FRC.SLN | Myeloid | 9.00E-09 | 1.85E-08 | 0.31306022 |
| Arteriolosclerosis | FRC.SLN | Myeloid | -3.33E-08 | 2.42E-08 | 0.91552299 |
| Arteriolosclerosis (any | FRC.SLN | Myeloid | -5.31E-08 | 2.72E-08 | 0.974243016 |
| Atherosclerosis | FRC.SLN | Myeloid | -5.40E-09 | 2.40E-08 | 0.589015928 |
| Atherosclerosis (any/r | FRC.SLN | Myeloid | -1.31E-08 | 2.08E-08 | 0.736233972 |
| B Score | FRC.SLN | Myeloid | -2.15E-08 | 1.69E-08 | 0.898224201 |
| CAA | FRC.SLN | Myeloid | -7.86E-09 | 2.26E-08 | 0.63594776 |
| CAA (any/none) | FRC.SLN | Myeloid | 1.46E-08 | 2.39E-08 | 0.270272842 |
| CERAD (C Score) | FRC.SLN | Myeloid | 2.50E-08 | 1.73E-08 | 0.07374232 |
| Infarcts/lacunes | FRC.SLN | Myeloid | -2.97E-08 | 2.37E-08 | 0.895463591 |
| PD Braak | FRC.SLN | Myeloid | 1.42E-09 | 1.97E-08 | 0.471253965 |
| PD Braak (any/none) | FRC.SLN | Myeloid | 7.65E-09 | 1.90E-08 | 0.344024165 |
| TDP-43 (any/none) | FRC.SLN | Myeloid | 2.44E-09 | 5.22E-08 | 0.481360802 |
| Thal Phase | FRC.SLN | Myeloid | 8.72E-08 | 6.56E-08 | 0.092043536 |
| WMR (any/none) | FRC.SLN | Myeloid | -4.10E-08 | 3.16E-08 | 0.903036685 |
| #N/A | FRC.SLN | Myeloid | -4.91E-08 | 3.81E-08 | 0.901002903 |
| AD Braak | FRC.SLN.CFA.d6.v2 | Myeloid | -1.34E-09 | 1.68E-08 | 0.531960147 |
| ADNC Score | FRC.SLN.CFA.d6.v2 | Myeloid | 6.41E-08 | 5.64E-08 | 0.128073889 |
| AP (any/none) | FRC.SLN.CFA.d6.v2 | Myeloid | -8.71E-09 | 1.92E-08 | 0.674762176 |
| Arteriolosclerosis | FRC.SLN.CFA.d6.v2 | Myeloid | -3.21E-08 | 2.45E-08 | 0.905190218 |
| Arteriolosclerosis (any | FRC.SLN.CFA.d6.v2 | Myeloid | -6.03E-08 | 2.66E-08 | 0.988233161 |
| Atherosclerosis | FRC.SLN.CFA.d6.v2 | Myeloid | 5.04E-09 | 2.21E-08 | 0.41008457 |
| Atherosclerosis (any/r | FRC.SLN.CFA.d6.v2 | Myeloid | -1.56E-08 | 2.05E-08 | 0.777251392 |
| B Score | FRC.SLN.CFA.d6.v2 | Myeloid | -1.84E-08 | 1.73E-08 | 0.856459346 |
| CAA | FRC.SLN.CFA.d6.v2 | Myeloid | 1.07E-08 | 2.26E-08 | 0.317585869 |
| CAA (any/none) | FRC.SLN.CFA.d6.v2 | Myeloid | 2.52E-08 | 2.45E-08 | 0.151570703 |
| CERAD (C Score) | FRC.SLN.CFA.d6.v2 | Myeloid | 1.07E-08 | 1.70E-08 | 0.264843769 |
| Infarcts/lacunes | FRC.SLN.CFA.d6.v2 | Myeloid | 2.91E-09 | 2.37E-08 | 0.450970657 |
| PD Braak | FRC.SLN.CFA.d6.v2 | Myeloid | 1.77E-08 | 2.01E-08 | 0.189557131 |
| PD Braak (any/none) | FRC.SLN.CFA.d6.v2 | Myeloid | 1.51E-08 | 1.97E-08 | 0.222442013 |
| TDP-43 (any/none) | FRC.SLN.CFA.d6.v2 | Myeloid | 3.18E-08 | 6.08E-08 | 0.300279498 |
| Thal Phase | FRC.SLN.CFA.d6.v2 | Myeloid | 8.48E-08 | 6.39E-08 | 0.092218006 |
| WMR (any/none) | FRC.SLN.CFA.d6.v2 | Myeloid | -6.77E-08 | 3.09E-08 | 0.985763163 |
| #N/A | FRC.SLN.CFA.d6.v2 | Myeloid | -8.34E-08 | 3.64E-08 | 0.989030828 |

| | | | | | |
|---|---|---|---|---|---|
| AD Braak | FRC.SLN.OT | Myeloid | -1.26E-08 | 1.70E-08 | 0.7702525 |
| ADNC Score | FRC.SLN.OT | Myeloid | 5.67E-08 | 5.05E-08 | 0.130583573 |
| AP (any/none) | FRC.SLN.OT | Myeloid | 4.54E-09 | 1.98E-08 | 0.409335633 |
| Arteriolosclerosis | FRC.SLN.OT | Myeloid | -2.96E-08 | 2.48E-08 | 0.882997866 |
| Arteriolosclerosis (any | FRC.SLN.OT | Myeloid | -4.31E-08 | 2.73E-08 | 0.94271523 |
| Atherosclerosis | FRC.SLN.OT | Myeloid | -7.83E-09 | 2.37E-08 | 0.629569503 |
| Atherosclerosis (any/r | FRC.SLN.OT | Myeloid | -6.20E-09 | 2.18E-08 | 0.612081695 |
| B Score | FRC.SLN.OT | Myeloid | -1.47E-08 | 1.72E-08 | 0.80466221 |
| CAA | FRC.SLN.OT | Myeloid | -1.10E-08 | 2.17E-08 | 0.693524555 |
| CAA (any/none) | FRC.SLN.OT | Myeloid | 2.07E-08 | 2.28E-08 | 0.181630665 |
| CERAD (C Score) | FRC.SLN.OT | Myeloid | 1.71E-08 | 1.72E-08 | 0.160814124 |
| Infarcts/lacunes | FRC.SLN.OT | Myeloid | -2.51E-08 | 2.32E-08 | 0.860136539 |
| PD Braak | FRC.SLN.OT | Myeloid | 2.88E-10 | 1.93E-08 | 0.494039014 |
| PD Braak (any/none) | FRC.SLN.OT | Myeloid | 5.84E-09 | 1.91E-08 | 0.379643879 |
| TDP-43 (any/none) | FRC.SLN.OT | Myeloid | 2.65E-08 | 4.93E-08 | 0.295158675 |
| Thal Phase | FRC.SLN.OT | Myeloid | 5.48E-08 | 6.71E-08 | 0.207186571 |
| WMR (any/none) | FRC.SLN.OT | Myeloid | -6.41E-08 | 3.31E-08 | 0.973489151 |
| #N/A | FRC.SLN.OT | Myeloid | -7.86E-08 | 3.78E-08 | 0.981270736 |
| AD Braak | FRC.SLN.v2 | Myeloid | -7.77E-09 | 1.59E-08 | 0.687726155 |
| ADNC Score | FRC.SLN.v2 | Myeloid | 4.80E-08 | 5.45E-08 | 0.189019595 |
| AP (any/none) | FRC.SLN.v2 | Myeloid | -1.26E-08 | 1.90E-08 | 0.745844359 |
| Arteriolosclerosis | FRC.SLN.v2 | Myeloid | -3.77E-08 | 2.37E-08 | 0.944070107 |
| Arteriolosclerosis (any | FRC.SLN.v2 | Myeloid | -5.76E-08 | 2.76E-08 | 0.981586355 |
| Atherosclerosis | FRC.SLN.v2 | Myeloid | 4.87E-09 | 2.29E-08 | 0.415736878 |
| Atherosclerosis (any/r | FRC.SLN.v2 | Myeloid | 1.99E-09 | 2.08E-08 | 0.461783341 |
| B Score | FRC.SLN.v2 | Myeloid | -2.55E-08 | 1.66E-08 | 0.937903533 |
| CAA | FRC.SLN.v2 | Myeloid | 2.90E-09 | 2.31E-08 | 0.450000793 |
| CAA (any/none) | FRC.SLN.v2 | Myeloid | 2.97E-08 | 2.43E-08 | 0.111022821 |
| CERAD (C Score) | FRC.SLN.v2 | Myeloid | 1.06E-08 | 1.72E-08 | 0.268306857 |
| Infarcts/lacunes | FRC.SLN.v2 | Myeloid | -2.43E-09 | 2.36E-08 | 0.541030075 |
| PD Braak | FRC.SLN.v2 | Myeloid | 5.84E-09 | 1.97E-08 | 0.383108153 |
| PD Braak (any/none) | FRC.SLN.v2 | Myeloid | 5.53E-09 | 1.92E-08 | 0.386697528 |
| TDP-43 (any/none) | FRC.SLN.v2 | Myeloid | 3.72E-08 | 4.99E-08 | 0.22790515 |
| Thal Phase | FRC.SLN.v2 | Myeloid | 4.29E-08 | 6.49E-08 | 0.2543301 |

| | | | | | |
|---|---|---|---|---|---|
| WMR (any/none) | FRC.SLN.v2 | Myeloid | -6.29E-08 | 3.05E-08 | 0.980319827 |
| #N/A | FRC.SLN.v2 | Myeloid | -7.16E-08 | 3.52E-08 | 0.97888215 |
| AD Braak | GN.Arth.BM | Myeloid | -1.16E-08 | 1.87E-08 | 0.732074493 |
| ADNC Score | GN.Arth.BM | Myeloid | -2.55E-08 | 5.89E-08 | 0.667544673 |
| AP (any/none) | GN.Arth.BM | Myeloid | -2.14E-09 | 2.23E-08 | 0.53829474 |
| Arteriolosclerosis | GN.Arth.BM | Myeloid | 3.93E-08 | 2.97E-08 | 0.092907072 |
| Arteriolosclerosis (any | GN.Arth.BM | Myeloid | -8.44E-09 | 3.45E-08 | 0.596501549 |
| Atherosclerosis | GN.Arth.BM | Myeloid | -3.17E-10 | 2.51E-08 | 0.505035477 |
| Atherosclerosis (any/r | GN.Arth.BM | Myeloid | -6.19E-10 | 2.47E-08 | 0.509999583 |
| B Score | GN.Arth.BM | Myeloid | -9.82E-09 | 1.90E-08 | 0.697435942 |
| CAA | GN.Arth.BM | Myeloid | -4.30E-08 | 2.46E-08 | 0.959864797 |
| CAA (any/none) | GN.Arth.BM | Myeloid | -3.74E-08 | 2.30E-08 | 0.948188012 |
| CERAD (C Score) | GN.Arth.BM | Myeloid | 2.62E-08 | 1.97E-08 | 0.091732215 |
| Infarcts/lacunes | GN.Arth.BM | Myeloid | 8.84E-09 | 2.64E-08 | 0.368720389 |
| PD Braak | GN.Arth.BM | Myeloid | 1.89E-08 | 2.51E-08 | 0.226083967 |
| PD Braak (any/none) | GN.Arth.BM | Myeloid | 2.68E-08 | 2.32E-08 | 0.124113764 |
| TDP-43 (any/none) | GN.Arth.BM | Myeloid | 2.26E-09 | 6.43E-08 | 0.485989817 |
| Thal Phase | GN.Arth.BM | Myeloid | -8.99E-08 | 8.77E-08 | 0.847488566 |
| WMR (any/none) | GN.Arth.BM | Myeloid | 7.13E-08 | 4.03E-08 | 0.038635365 |
| #N/A | GN.Arth.BM | Myeloid | 2.72E-08 | 5.06E-08 | 0.295786849 |
| AD Braak | GN.Arth.SynF | Myeloid | -4.49E-09 | 1.74E-08 | 0.601959941 |
| ADNC Score | GN.Arth.SynF | Myeloid | 5.24E-08 | 6.14E-08 | 0.19646704 |
| AP (any/none) | GN.Arth.SynF | Myeloid | 8.27E-09 | 2.25E-08 | 0.35665115 |
| Arteriolosclerosis | GN.Arth.SynF | Myeloid | -7.08E-10 | 2.62E-08 | 0.510783552 |
| Arteriolosclerosis (any | GN.Arth.SynF | Myeloid | 1.26E-08 | 3.04E-08 | 0.339900603 |
| Atherosclerosis | GN.Arth.SynF | Myeloid | 2.49E-08 | 2.18E-08 | 0.126942537 |
| Atherosclerosis (any/r | GN.Arth.SynF | Myeloid | -1.22E-08 | 2.18E-08 | 0.711927895 |
| B Score | GN.Arth.SynF | Myeloid | -1.58E-09 | 1.76E-08 | 0.535898359 |
| CAA | GN.Arth.SynF | Myeloid | -5.09E-09 | 2.60E-08 | 0.57747583 |
| CAA (any/none) | GN.Arth.SynF | Myeloid | -1.13E-08 | 2.30E-08 | 0.688920016 |
| CERAD (C Score) | GN.Arth.SynF | Myeloid | 4.51E-08 | 2.10E-08 | 0.015752509 |
| Infarcts/lacunes | GN.Arth.SynF | Myeloid | -5.23E-09 | 2.29E-08 | 0.590164608 |
| PD Braak | GN.Arth.SynF | Myeloid | 2.71E-08 | 2.27E-08 | 0.116257404 |
| PD Braak (any/none) | GN.Arth.SynF | Myeloid | 2.42E-08 | 2.16E-08 | 0.131430206 |

| | | | | | |
|---|---|---|---|---|---|
| TDP-43 (any/none) | GN.Arth.SynF | Myeloid | -5.90E-08 | 5.88E-08 | 0.842393868 |
| Thal Phase | GN.Arth.SynF | Myeloid | 1.28E-08 | 8.20E-08 | 0.437762684 |
| WMR (any/none) | GN.Arth.SynF | Myeloid | 2.47E-08 | 3.42E-08 | 0.234575107 |
| #N/A | GN.Arth.SynF | Myeloid | 5.89E-08 | 4.76E-08 | 0.108113405 |
| AD Braak | GN.Bl | Myeloid | -8.54E-09 | 1.82E-08 | 0.680357714 |
| ADNC Score | GN.Bl | Myeloid | 4.16E-08 | 6.73E-08 | 0.26819807 |
| AP (any/none) | GN.Bl | Myeloid | 1.08E-08 | 2.38E-08 | 0.32483373 |
| Arteriolosclerosis | GN.Bl | Myeloid | -9.42E-09 | 2.72E-08 | 0.635158589 |
| Arteriolosclerosis (any | GN.Bl | Myeloid | -1.38E-08 | 3.21E-08 | 0.66624129 |
| Atherosclerosis | GN.Bl | Myeloid | 1.03E-08 | 2.23E-08 | 0.322231659 |
| Atherosclerosis (any/r | GN.Bl | Myeloid | -1.01E-08 | 2.57E-08 | 0.65231775 |
| B Score | GN.Bl | Myeloid | -1.69E-08 | 1.92E-08 | 0.810972636 |
| CAA | GN.Bl | Myeloid | -2.49E-08 | 2.42E-08 | 0.848418568 |
| CAA (any/none) | GN.Bl | Myeloid | -2.69E-08 | 2.39E-08 | 0.870403358 |
| CERAD (C Score) | GN.Bl | Myeloid | 2.98E-08 | 2.14E-08 | 0.082297592 |
| Infarcts/lacunes | GN.Bl | Myeloid | 3.14E-09 | 2.58E-08 | 0.451415584 |
| PD Braak | GN.Bl | Myeloid | 3.38E-08 | 2.25E-08 | 0.066708326 |
| PD Braak (any/none) | GN.Bl | Myeloid | 3.40E-08 | 2.10E-08 | 0.052250486 |
| TDP-43 (any/none) | GN.Bl | Myeloid | 6.99E-09 | 5.96E-08 | 0.453335706 |
| Thal Phase | GN.Bl | Myeloid | -1.06E-07 | 8.57E-08 | 0.892743501 |
| WMR (any/none) | GN.Bl | Myeloid | 1.85E-08 | 3.70E-08 | 0.308036281 |
| #N/A | GN.Bl | Myeloid | 5.27E-09 | 4.98E-08 | 0.457837296 |
| AD Braak | GN.Bl.v2 | Myeloid | 2.10E-08 | 1.96E-08 | 0.142511412 |
| ADNC Score | GN.Bl.v2 | Myeloid | -5.44E-09 | 5.85E-08 | 0.536994769 |
| AP (any/none) | GN.Bl.v2 | Myeloid | 2.69E-08 | 2.49E-08 | 0.140533371 |
| Arteriolosclerosis | GN.Bl.v2 | Myeloid | -6.00E-09 | 2.64E-08 | 0.590009419 |
| Arteriolosclerosis (any | GN.Bl.v2 | Myeloid | 2.60E-08 | 2.95E-08 | 0.188917184 |
| Atherosclerosis | GN.Bl.v2 | Myeloid | 1.70E-08 | 2.22E-08 | 0.22110342 |
| Atherosclerosis (any/r | GN.Bl.v2 | Myeloid | -2.56E-08 | 2.37E-08 | 0.859919879 |
| B Score | GN.Bl.v2 | Myeloid | 1.57E-08 | 1.97E-08 | 0.213416555 |
| CAA | GN.Bl.v2 | Myeloid | -2.33E-08 | 2.34E-08 | 0.840329013 |
| CAA (any/none) | GN.Bl.v2 | Myeloid | -3.74E-08 | 2.27E-08 | 0.950120811 |
| CERAD (C Score) | GN.Bl.v2 | Myeloid | 3.29E-08 | 2.01E-08 | 0.05089804 |
| Infarcts/lacunes | GN.Bl.v2 | Myeloid | 5.17E-09 | 2.52E-08 | 0.418598106 |

| | | | | | |
|---|---|---|---|---|---|
| PD Braak | GN.Bl.v2 | Myeloid | 5.06E-08 | 2.17E-08 | 0.009806704 |
| PD Braak (any/none) | GN.Bl.v2 | Myeloid | 5.20E-08 | 2.06E-08 | 0.005817216 |
| TDP-43 (any/none) | GN.Bl.v2 | Myeloid | -6.20E-08 | 5.99E-08 | 0.849712433 |
| Thal Phase | GN.Bl.v2 | Myeloid | -1.72E-07 | 7.66E-08 | 0.987505232 |
| WMR (any/none) | GN.Bl.v2 | Myeloid | 2.71E-08 | 3.50E-08 | 0.219198269 |
| #N/A | GN.Bl.v2 | Myeloid | 4.66E-08 | 4.44E-08 | 0.147074057 |
| AD Braak | GN.BM | Myeloid | -6.75E-09 | 1.86E-08 | 0.641727456 |
| ADNC Score | GN.BM | Myeloid | -4.35E-08 | 5.99E-08 | 0.76607048 |
| AP (any/none) | GN.BM | Myeloid | 1.59E-08 | 2.37E-08 | 0.251327638 |
| Arteriolosclerosis | GN.BM | Myeloid | -2.44E-09 | 2.84E-08 | 0.534306258 |
| Arteriolosclerosis (any | GN.BM | Myeloid | -3.39E-08 | 3.37E-08 | 0.842828792 |
| Atherosclerosis | GN.BM | Myeloid | -1.22E-08 | 2.30E-08 | 0.70214958 |
| Atherosclerosis (any/r | GN.BM | Myeloid | -9.72E-09 | 2.67E-08 | 0.641880921 |
| B Score | GN.BM | Myeloid | 6.88E-09 | 1.95E-08 | 0.361834529 |
| CAA | GN.BM | Myeloid | -4.29E-08 | 2.56E-08 | 0.95311392 |
| CAA (any/none) | GN.BM | Myeloid | -3.65E-08 | 2.45E-08 | 0.932000768 |
| CERAD (C Score) | GN.BM | Myeloid | 4.12E-08 | 2.21E-08 | 0.031364852 |
| Infarcts/lacunes | GN.BM | Myeloid | 1.95E-08 | 2.49E-08 | 0.217191887 |
| PD Braak | GN.BM | Myeloid | 8.00E-09 | 2.38E-08 | 0.368218644 |
| PD Braak (any/none) | GN.BM | Myeloid | 1.61E-08 | 2.22E-08 | 0.233311563 |
| TDP-43 (any/none) | GN.BM | Myeloid | 1.64E-09 | 6.22E-08 | 0.489504557 |
| Thal Phase | GN.BM | Myeloid | -1.56E-07 | 8.24E-08 | 0.970948331 |
| WMR (any/none) | GN.BM | Myeloid | 3.09E-08 | 3.89E-08 | 0.213144402 |
| #N/A | GN.BM | Myeloid | 4.53E-08 | 4.95E-08 | 0.179803482 |
| AD Braak | GN.Thio.PC | Myeloid | 2.62E-09 | 1.80E-08 | 0.441992285 |
| ADNC Score | GN.Thio.PC | Myeloid | 1.04E-07 | 6.62E-08 | 0.058433289 |
| AP (any/none) | GN.Thio.PC | Myeloid | -2.22E-09 | 2.43E-08 | 0.536471751 |
| Arteriolosclerosis | GN.Thio.PC | Myeloid | -2.58E-08 | 2.62E-08 | 0.837463712 |
| Arteriolosclerosis (any | GN.Thio.PC | Myeloid | -8.44E-09 | 3.09E-08 | 0.607431676 |
| Atherosclerosis | GN.Thio.PC | Myeloid | 1.60E-08 | 2.46E-08 | 0.257399645 |
| Atherosclerosis (any/r | GN.Thio.PC | Myeloid | 2.60E-08 | 2.59E-08 | 0.157795329 |
| B Score | GN.Thio.PC | Myeloid | -2.49E-09 | 1.87E-08 | 0.552922141 |
| CAA | GN.Thio.PC | Myeloid | -2.77E-09 | 2.58E-08 | 0.542840179 |
| CAA (any/none) | GN.Thio.PC | Myeloid | -1.60E-08 | 2.35E-08 | 0.752910556 |

| | | | | | |
|---|---|---|---|---|---|
| CERAD (C Score) | GN.Thio.PC | Myeloid | 2.64E-08 | 2.19E-08 | 0.114102819 |
| Infarcts/lacunes | GN.Thio.PC | Myeloid | -5.53E-09 | 2.46E-08 | 0.58879207 |
| PD Braak | GN.Thio.PC | Myeloid | 2.28E-08 | 2.25E-08 | 0.155670095 |
| PD Braak (any/none) | GN.Thio.PC | Myeloid | 1.96E-08 | 2.16E-08 | 0.182989956 |
| TDP-43 (any/none) | GN.Thio.PC | Myeloid | -2.23E-08 | 5.97E-08 | 0.645593933 |
| Thal Phase | GN.Thio.PC | Myeloid | 4.95E-08 | 9.04E-08 | 0.292133028 |
| WMR (any/none) | GN.Thio.PC | Myeloid | 1.89E-08 | 3.76E-08 | 0.308147886 |
| #N/A | GN.Thio.PC | Myeloid | 7.50E-08 | 5.03E-08 | 0.067933229 |
| AD Braak | GN.UrAc.PC | Myeloid | -1.91E-09 | 1.87E-08 | 0.54071892 |
| ADNC Score | GN.UrAc.PC | Myeloid | 3.66E-08 | 6.39E-08 | 0.283426949 |
| AP (any/none) | GN.UrAc.PC | Myeloid | -1.86E-08 | 2.57E-08 | 0.764801612 |
| Arteriolosclerosis | GN.UrAc.PC | Myeloid | 3.81E-09 | 2.81E-08 | 0.446129301 |
| Arteriolosclerosis (any | GN.UrAc.PC | Myeloid | 2.65E-08 | 3.33E-08 | 0.213179454 |
| Atherosclerosis | GN.UrAc.PC | Myeloid | 3.57E-08 | 2.62E-08 | 0.086825871 |
| Atherosclerosis (any/r | GN.UrAc.PC | Myeloid | 8.86E-09 | 2.47E-08 | 0.359765179 |
| B Score | GN.UrAc.PC | Myeloid | -4.80E-09 | 1.85E-08 | 0.602265118 |
| CAA | GN.UrAc.PC | Myeloid | -2.54E-08 | 2.74E-08 | 0.823856455 |
| CAA (any/none) | GN.UrAc.PC | Myeloid | -1.35E-08 | 2.48E-08 | 0.706718535 |
| CERAD (C Score) | GN.UrAc.PC | Myeloid | 2.38E-08 | 2.21E-08 | 0.140694746 |
| Infarcts/lacunes | GN.UrAc.PC | Myeloid | 9.03E-09 | 2.55E-08 | 0.361743887 |
| PD Braak | GN.UrAc.PC | Myeloid | 7.53E-08 | 2.41E-08 | 8.90E-04 |
| PD Braak (any/none) | GN.UrAc.PC | Myeloid | 7.34E-08 | 2.27E-08 | 5.99E-04 |
| TDP-43 (any/none) | GN.UrAc.PC | Myeloid | -2.62E-08 | 5.72E-08 | 0.676755298 |
| Thal Phase | GN.UrAc.PC | Myeloid | -3.87E-08 | 8.77E-08 | 0.670366815 |
| WMR (any/none) | GN.UrAc.PC | Myeloid | -1.11E-08 | 3.42E-08 | 0.627296848 |
| #N/A | GN.UrAc.PC | Myeloid | 3.47E-08 | 4.78E-08 | 0.233681882 |
| AD Braak | MC.digest.PC | Myeloid | -6.81E-09 | 1.58E-08 | 0.667096291 |
| ADNC Score | MC.digest.PC | Myeloid | 6.92E-09 | 5.73E-08 | 0.451973086 |
| AP (any/none) | MC.digest.PC | Myeloid | 2.54E-09 | 2.08E-08 | 0.451404892 |
| Arteriolosclerosis | MC.digest.PC | Myeloid | 2.53E-08 | 2.58E-08 | 0.163588091 |
| Arteriolosclerosis (any | MC.digest.PC | Myeloid | 1.72E-08 | 2.51E-08 | 0.247129962 |
| Atherosclerosis | MC.digest.PC | Myeloid | -8.21E-09 | 2.16E-08 | 0.647935974 |
| Atherosclerosis (any/r | MC.digest.PC | Myeloid | -3.56E-09 | 2.21E-08 | 0.563865054 |
| B Score | MC.digest.PC | Myeloid | -9.37E-09 | 1.63E-08 | 0.717223643 |

| | | | | | |
|---|---|---|---|---|---|
| CAA | MC.digest.PC | Myeloid | 1.05E-08 | 2.09E-08 | 0.3081798 |
| CAA (any/none) | MC.digest.PC | Myeloid | -5.02E-09 | 2.06E-08 | 0.596213008 |
| CERAD (C Score) | MC.digest.PC | Myeloid | -7.79E-09 | 1.77E-08 | 0.670395693 |
| Infarcts/lacunes | MC.digest.PC | Myeloid | 2.86E-08 | 2.41E-08 | 0.118423203 |
| PD Braak | MC.digest.PC | Myeloid | -4.21E-09 | 1.91E-08 | 0.587328759 |
| PD Braak (any/none) | MC.digest.PC | Myeloid | 1.84E-09 | 1.91E-08 | 0.461482232 |
| TDP-43 (any/none) | MC.digest.PC | Myeloid | -6.06E-08 | 5.26E-08 | 0.875531105 |
| Thal Phase | MC.digest.PC | Myeloid | -3.89E-08 | 6.95E-08 | 0.711983711 |
| WMR (any/none) | MC.digest.PC | Myeloid | 4.81E-08 | 2.98E-08 | 0.053609261 |
| #N/A | MC.digest.PC | Myeloid | -1.28E-08 | 4.12E-08 | 0.621742735 |
| AD Braak | MC.Es | Myeloid | -1.35E-08 | 1.67E-08 | 0.791602957 |
| ADNC Score | MC.Es | Myeloid | -2.40E-08 | 5.49E-08 | 0.668921867 |
| AP (any/none) | MC.Es | Myeloid | 2.07E-08 | 2.25E-08 | 0.178230722 |
| Arteriolosclerosis | MC.Es | Myeloid | 4.08E-08 | 2.84E-08 | 0.07545556 |
| Arteriolosclerosis (any | MC.Es | Myeloid | 5.30E-10 | 2.48E-08 | 0.49147418 |
| Atherosclerosis | MC.Es | Myeloid | 9.85E-09 | 2.35E-08 | 0.337671905 |
| Atherosclerosis (any/r | MC.Es | Myeloid | -1.03E-08 | 2.26E-08 | 0.676432568 |
| B Score | MC.Es | Myeloid | -6.98E-09 | 1.68E-08 | 0.660692175 |
| CAA | MC.Es | Myeloid | 1.36E-08 | 2.25E-08 | 0.272979108 |
| CAA (any/none) | MC.Es | Myeloid | 1.74E-08 | 2.30E-08 | 0.224466167 |
| CERAD (C Score) | MC.Es | Myeloid | 2.70E-09 | 1.75E-08 | 0.438482023 |
| Infarcts/lacunes | MC.Es | Myeloid | 1.75E-08 | 2.33E-08 | 0.227105193 |
| PD Braak | MC.Es | Myeloid | 1.30E-08 | 2.19E-08 | 0.275750358 |
| PD Braak (any/none) | MC.Es | Myeloid | 1.85E-08 | 2.12E-08 | 0.19151308 |
| TDP-43 (any/none) | MC.Es | Myeloid | -3.64E-10 | 4.77E-08 | 0.503039162 |
| Thal Phase | MC.Es | Myeloid | -2.18E-08 | 6.70E-08 | 0.627829422 |
| WMR (any/none) | MC.Es | Myeloid | 4.95E-08 | 3.36E-08 | 0.070481649 |
| #N/A | MC.Es | Myeloid | -7.16E-08 | 4.25E-08 | 0.953873203 |
| AD Braak | MC.PC | Myeloid | -1.26E-08 | 1.68E-08 | 0.772715361 |
| ADNC Score | MC.PC | Myeloid | -1.08E-08 | 5.18E-08 | 0.58231855 |
| AP (any/none) | MC.PC | Myeloid | 1.71E-08 | 1.96E-08 | 0.192390487 |
| Arteriolosclerosis | MC.PC | Myeloid | 3.49E-08 | 2.69E-08 | 0.096919218 |
| Arteriolosclerosis (any | MC.PC | Myeloid | 3.25E-08 | 2.60E-08 | 0.105339373 |
| Atherosclerosis | MC.PC | Myeloid | -8.09E-09 | 2.04E-08 | 0.653857447 |

| | | | | | |
|---|---|---|---|---|---|
| Atherosclerosis (any/r | MC.PC | Myeloid | 2.20E-09 | 2.09E-08 | 0.458059642 |
| B Score | MC.PC | Myeloid | -2.18E-08 | 1.62E-08 | 0.910927395 |
| CAA | MC.PC | Myeloid | -1.86E-08 | 2.06E-08 | 0.815871721 |
| CAA (any/none) | MC.PC | Myeloid | 1.09E-09 | 2.14E-08 | 0.479582862 |
| CERAD (C Score) | MC.PC | Myeloid | -5.15E-09 | 1.68E-08 | 0.620454306 |
| Infarcts/lacunes | MC.PC | Myeloid | 2.54E-08 | 2.47E-08 | 0.151892922 |
| PD Braak | MC.PC | Myeloid | 8.01E-09 | 1.92E-08 | 0.338530159 |
| PD Braak (any/none) | MC.PC | Myeloid | 8.89E-09 | 2.00E-08 | 0.328725684 |
| TDP-43 (any/none) | MC.PC | Myeloid | -7.28E-08 | 5.54E-08 | 0.90541844 |
| Thal Phase | MC.PC | Myeloid | -6.15E-08 | 7.29E-08 | 0.800618034 |
| WMR (any/none) | MC.PC | Myeloid | 8.16E-09 | 3.09E-08 | 0.395885312 |
| #N/A | MC.PC | Myeloid | -4.32E-08 | 4.08E-08 | 0.854869795 |
| AD Braak | MC.Sk | Myeloid | -1.61E-08 | 1.63E-08 | 0.838270133 |
| ADNC Score | MC.Sk | Myeloid | -1.07E-08 | 5.01E-08 | 0.584727257 |
| AP (any/none) | MC.Sk | Myeloid | 4.14E-08 | 2.24E-08 | 0.032030942 |
| Arteriolosclerosis | MC.Sk | Myeloid | -3.72E-09 | 2.73E-08 | 0.554075462 |
| Arteriolosclerosis (any | MC.Sk | Myeloid | -1.52E-08 | 2.43E-08 | 0.733887229 |
| Atherosclerosis | MC.Sk | Myeloid | -1.17E-08 | 2.39E-08 | 0.687451308 |
| Atherosclerosis (any/r | MC.Sk | Myeloid | -6.23E-09 | 2.31E-08 | 0.606116584 |
| B Score | MC.Sk | Myeloid | -2.21E-08 | 1.53E-08 | 0.925167237 |
| CAA | MC.Sk | Myeloid | -1.82E-08 | 1.99E-08 | 0.819667022 |
| CAA (any/none) | MC.Sk | Myeloid | 1.10E-09 | 1.77E-08 | 0.475146259 |
| CERAD (C Score) | MC.Sk | Myeloid | 1.43E-08 | 1.78E-08 | 0.212237519 |
| Infarcts/lacunes | MC.Sk | Myeloid | 2.91E-09 | 2.27E-08 | 0.448888836 |
| PD Braak | MC.Sk | Myeloid | 2.17E-08 | 1.95E-08 | 0.133453998 |
| PD Braak (any/none) | MC.Sk | Myeloid | 2.62E-08 | 1.95E-08 | 0.089351359 |
| TDP-43 (any/none) | MC.Sk | Myeloid | 2.61E-08 | 5.28E-08 | 0.310179761 |
| Thal Phase | MC.Sk | Myeloid | -1.35E-08 | 7.08E-08 | 0.575731005 |
| WMR (any/none) | MC.Sk | Myeloid | 3.90E-08 | 3.40E-08 | 0.12571858 |
| #N/A | MC.Sk | Myeloid | -5.98E-08 | 4.40E-08 | 0.913240201 |
| AD Braak | MC.To | Myeloid | 6.43E-09 | 1.74E-08 | 0.355888547 |
| ADNC Score | MC.To | Myeloid | -1.83E-08 | 5.25E-08 | 0.636302511 |
| AP (any/none) | MC.To | Myeloid | 3.36E-08 | 2.17E-08 | 0.060278749 |
| Arteriolosclerosis | MC.To | Myeloid | 2.59E-08 | 2.67E-08 | 0.166128373 |

| | | | | | |
|---|---|---|---|---|---|
| Arteriolosclerosis (any | MC.To | Myeloid | 2.87E-09 | 2.81E-08 | 0.459351184 |
| Atherosclerosis | MC.To | Myeloid | -1.78E-09 | 2.32E-08 | 0.530670228 |
| Atherosclerosis (any/r | MC.To | Myeloid | -8.85E-09 | 2.24E-08 | 0.653873441 |
| B Score | MC.To | Myeloid | 1.26E-08 | 1.74E-08 | 0.234163113 |
| CAA | MC.To | Myeloid | -1.90E-08 | 1.97E-08 | 0.832516486 |
| CAA (any/none) | MC.To | Myeloid | 3.03E-09 | 2.04E-08 | 0.441133982 |
| CERAD (C Score) | MC.To | Myeloid | 8.10E-09 | 1.77E-08 | 0.323924258 |
| Infarcts/lacunes | MC.To | Myeloid | 2.20E-08 | 2.35E-08 | 0.174753092 |
| PD Braak | MC.To | Myeloid | 2.27E-08 | 2.03E-08 | 0.131880686 |
| PD Braak (any/none) | MC.To | Myeloid | 3.02E-08 | 2.01E-08 | 0.06597502 |
| TDP-43 (any/none) | MC.To | Myeloid | -7.56E-09 | 4.71E-08 | 0.563794084 |
| Thal Phase | MC.To | Myeloid | -3.18E-08 | 6.94E-08 | 0.676560526 |
| WMR (any/none) | MC.To | Myeloid | 5.32E-08 | 3.32E-08 | 0.054821232 |
| #N/A | MC.To | Myeloid | -4.95E-08 | 4.22E-08 | 0.87954768 |
| AD Braak | MC.Tr | Myeloid | -2.66E-08 | 1.62E-08 | 0.949259423 |
| ADNC Score | MC.Tr | Myeloid | -7.64E-08 | 5.57E-08 | 0.915050204 |
| AP (any/none) | MC.Tr | Myeloid | 2.79E-08 | 2.20E-08 | 0.102537446 |
| Arteriolosclerosis | MC.Tr | Myeloid | 4.11E-09 | 2.74E-08 | 0.440382623 |
| Arteriolosclerosis (any | MC.Tr | Myeloid | -1.20E-08 | 2.61E-08 | 0.677217055 |
| Atherosclerosis | MC.Tr | Myeloid | 2.42E-08 | 2.26E-08 | 0.142344671 |
| Atherosclerosis (any/r | MC.Tr | Myeloid | 1.79E-08 | 2.22E-08 | 0.210351489 |
| B Score | MC.Tr | Myeloid | -2.74E-08 | 1.58E-08 | 0.959068815 |
| CAA | MC.Tr | Myeloid | -3.42E-08 | 2.25E-08 | 0.93562253 |
| CAA (any/none) | MC.Tr | Myeloid | -1.34E-08 | 2.08E-08 | 0.739633866 |
| CERAD (C Score) | MC.Tr | Myeloid | -2.96E-09 | 1.68E-08 | 0.570068643 |
| Infarcts/lacunes | MC.Tr | Myeloid | 2.10E-08 | 2.33E-08 | 0.183513414 |
| PD Braak | MC.Tr | Myeloid | 3.52E-10 | 1.93E-08 | 0.49271184 |
| PD Braak (any/none) | MC.Tr | Myeloid | 1.34E-08 | 1.97E-08 | 0.248831305 |
| TDP-43 (any/none) | MC.Tr | Myeloid | -1.00E-07 | 4.56E-08 | 0.986244688 |
| Thal Phase | MC.Tr | Myeloid | -1.26E-07 | 7.10E-08 | 0.961681489 |
| WMR (any/none) | MC.Tr | Myeloid | 5.98E-08 | 3.27E-08 | 0.033652855 |
| #N/A | MC.Tr | Myeloid | -9.37E-08 | 4.15E-08 | 0.988004163 |
| AD Braak | MF.103-11b+.Lu | Myeloid | -9.62E-09 | 1.92E-08 | 0.691973467 |
| ADNC Score | MF.103-11b+.Lu | Myeloid | 6.73E-09 | 6.19E-08 | 0.456712898 |

| | | | | | |
|---|---|---|---|---|---|
| AP (any/none) | MF.103-11b+.Lu | Myeloid | 2.39E-08 | 2.33E-08 | 0.151963494 |
| Arteriolosclerosis | MF.103-11b+.Lu | Myeloid | 2.65E-08 | 2.72E-08 | 0.164859418 |
| Arteriolosclerosis (any | MF.103-11b+.Lu | Myeloid | 4.49E-08 | 3.13E-08 | 0.075432725 |
| Atherosclerosis | MF.103-11b+.Lu | Myeloid | -1.06E-08 | 2.18E-08 | 0.686291132 |
| Atherosclerosis (any/r | MF.103-11b+.Lu | Myeloid | 3.04E-09 | 2.37E-08 | 0.448978664 |
| B Score | MF.103-11b+.Lu | Myeloid | -1.00E-08 | 1.85E-08 | 0.706446457 |
| CAA | MF.103-11b+.Lu | Myeloid | 7.34E-09 | 2.28E-08 | 0.373618863 |
| CAA (any/none) | MF.103-11b+.Lu | Myeloid | -1.58E-08 | 2.18E-08 | 0.766359043 |
| CERAD (C Score) | MF.103-11b+.Lu | Myeloid | 2.15E-08 | 1.81E-08 | 0.117316699 |
| Infarcts/lacunes | MF.103-11b+.Lu | Myeloid | -9.59E-10 | 2.52E-08 | 0.515186407 |
| PD Braak | MF.103-11b+.Lu | Myeloid | 3.00E-08 | 2.29E-08 | 0.095415283 |
| PD Braak (any/none) | MF.103-11b+.Lu | Myeloid | 3.34E-08 | 2.18E-08 | 0.062957141 |
| TDP-43 (any/none) | MF.103-11b+.Lu | Myeloid | -3.41E-08 | 5.40E-08 | 0.73618841 |
| Thal Phase | MF.103-11b+.Lu | Myeloid | -4.12E-08 | 7.78E-08 | 0.701814783 |
| WMR (any/none) | MF.103-11b+.Lu | Myeloid | 1.33E-08 | 3.86E-08 | 0.364926441 |
| #N/A | MF.103-11b+.Lu | Myeloid | -1.53E-08 | 3.92E-08 | 0.651509957 |
| AD Braak | MF.103-11b+.Salm3.Sl | Myeloid | 8.25E-09 | 1.79E-08 | 0.321964486 |
| ADNC Score | MF.103-11b+.Salm3.Sl | Myeloid | 4.24E-08 | 6.95E-08 | 0.270770297 |
| AP (any/none) | MF.103-11b+.Salm3.Sl | Myeloid | 1.52E-08 | 2.47E-08 | 0.269669752 |
| Arteriolosclerosis | MF.103-11b+.Salm3.Sl | Myeloid | 4.17E-08 | 2.95E-08 | 0.078776877 |
| Arteriolosclerosis (any | MF.103-11b+.Salm3.Sl | Myeloid | 5.03E-08 | 3.15E-08 | 0.055148561 |
| Atherosclerosis | MF.103-11b+.Salm3.Sl | Myeloid | -1.11E-08 | 2.32E-08 | 0.682949041 |
| Atherosclerosis (any/r | MF.103-11b+.Salm3.Sl | Myeloid | -1.23E-09 | 2.26E-08 | 0.521760805 |
| B Score | MF.103-11b+.Salm3.Sl | Myeloid | -4.27E-09 | 1.74E-08 | 0.596938689 |
| CAA | MF.103-11b+.Salm3.Sl | Myeloid | 6.96E-09 | 2.32E-08 | 0.38213595 |
| CAA (any/none) | MF.103-11b+.Salm3.Sl | Myeloid | 8.20E-09 | 2.47E-08 | 0.36972941 |
| CERAD (C Score) | MF.103-11b+.Salm3.Sl | Myeloid | 3.89E-08 | 2.02E-08 | 0.027253227 |
| Infarcts/lacunes | MF.103-11b+.Salm3.Sl | Myeloid | 4.48E-08 | 2.58E-08 | 0.041306705 |
| PD Braak | MF.103-11b+.Salm3.Sl | Myeloid | 3.50E-08 | 2.25E-08 | 0.060138336 |
| PD Braak (any/none) | MF.103-11b+.Salm3.Sl | Myeloid | 3.38E-08 | 2.14E-08 | 0.057463947 |
| TDP-43 (any/none) | MF.103-11b+.Salm3.Sl | Myeloid | -1.13E-07 | 5.31E-08 | 0.98319032 |
| Thal Phase | MF.103-11b+.Salm3.Sl | Myeloid | 8.71E-08 | 7.62E-08 | 0.126652371 |
| WMR (any/none) | MF.103-11b+.Salm3.Sl | Myeloid | -4.60E-08 | 3.91E-08 | 0.880179674 |
| #N/A | MF.103-11b+.Salm3.Sl | Myeloid | -2.94E-08 | 4.00E-08 | 0.769269073 |

| | | | | | |
|---|---|---|---|---|---|
| AD Braak | MF.103-11b+.SI | Myeloid | 7.32E-10 | 1.81E-08 | 0.48389571 |
| ADNC Score | MF.103-11b+.SI | Myeloid | 7.42E-08 | 6.21E-08 | 0.115969024 |
| AP (any/none) | MF.103-11b+.SI | Myeloid | 2.68E-08 | 2.34E-08 | 0.126693334 |
| Arteriolosclerosis | MF.103-11b+.SI | Myeloid | 1.67E-08 | 2.89E-08 | 0.281566846 |
| Arteriolosclerosis (any | MF.103-11b+.SI | Myeloid | 2.86E-08 | 3.14E-08 | 0.181461179 |
| Atherosclerosis | MF.103-11b+.SI | Myeloid | -1.70E-08 | 2.34E-08 | 0.765955594 |
| Atherosclerosis (any/r | MF.103-11b+.SI | Myeloid | -3.00E-09 | 2.24E-08 | 0.553220326 |
| B Score | MF.103-11b+.SI | Myeloid | 3.21E-09 | 1.83E-08 | 0.43046219 |
| CAA | MF.103-11b+.SI | Myeloid | 4.61E-09 | 2.29E-08 | 0.420064283 |
| CAA (any/none) | MF.103-11b+.SI | Myeloid | 2.86E-09 | 2.36E-08 | 0.45168035 |
| CERAD (C Score) | MF.103-11b+.SI | Myeloid | 4.96E-08 | 1.91E-08 | 0.004717661 |
| Infarcts/lacunes | MF.103-11b+.SI | Myeloid | 5.69E-08 | 2.52E-08 | 0.011830058 |
| PD Braak | MF.103-11b+.SI | Myeloid | 5.86E-08 | 2.33E-08 | 0.005864343 |
| PD Braak (any/none) | MF.103-11b+.SI | Myeloid | 4.85E-08 | 2.11E-08 | 0.010748471 |
| TDP-43 (any/none) | MF.103-11b+.SI | Myeloid | -1.02E-07 | 5.18E-08 | 0.975155397 |
| Thal Phase | MF.103-11b+.SI | Myeloid | 1.22E-07 | 6.98E-08 | 0.040541836 |
| WMR (any/none) | MF.103-11b+.SI | Myeloid | -3.57E-08 | 4.06E-08 | 0.810347745 |
| #N/A | MF.103-11b+.SI | Myeloid | 1.65E-09 | 4.08E-08 | 0.483838766 |
| AD Braak | MF.11c-11b+.Lu | Myeloid | 1.59E-08 | 1.71E-08 | 0.176658503 |
| ADNC Score | MF.11c-11b+.Lu | Myeloid | 6.74E-08 | 5.86E-08 | 0.125162873 |
| AP (any/none) | MF.11c-11b+.Lu | Myeloid | 3.84E-08 | 2.31E-08 | 0.048531297 |
| Arteriolosclerosis | MF.11c-11b+.Lu | Myeloid | 2.89E-08 | 2.88E-08 | 0.157828294 |
| Arteriolosclerosis (any | MF.11c-11b+.Lu | Myeloid | 1.61E-08 | 3.19E-08 | 0.306331463 |
| Atherosclerosis | MF.11c-11b+.Lu | Myeloid | -5.29E-08 | 2.24E-08 | 0.990793891 |
| Atherosclerosis (any/r | MF.11c-11b+.Lu | Myeloid | -2.34E-08 | 2.00E-08 | 0.878690998 |
| B Score | MF.11c-11b+.Lu | Myeloid | 2.20E-08 | 1.79E-08 | 0.109903039 |
| CAA | MF.11c-11b+.Lu | Myeloid | -4.75E-09 | 2.33E-08 | 0.58076207 |
| CAA (any/none) | MF.11c-11b+.Lu | Myeloid | 2.60E-09 | 2.04E-08 | 0.449326959 |
| CERAD (C Score) | MF.11c-11b+.Lu | Myeloid | 4.10E-08 | 1.95E-08 | 0.017546193 |
| Infarcts/lacunes | MF.11c-11b+.Lu | Myeloid | 4.31E-08 | 2.26E-08 | 0.028497207 |
| PD Braak | MF.11c-11b+.Lu | Myeloid | 3.56E-08 | 2.12E-08 | 0.046447121 |
| PD Braak (any/none) | MF.11c-11b+.Lu | Myeloid | 3.24E-08 | 2.05E-08 | 0.05650312 |
| TDP-43 (any/none) | MF.11c-11b+.Lu | Myeloid | -9.44E-08 | 5.27E-08 | 0.963406669 |
| Thal Phase | MF.11c-11b+.Lu | Myeloid | 4.32E-08 | 6.85E-08 | 0.26409302 |

| | | | | | |
|---|---|---|---|---|---|
| WMR (any/none) | MF.11c-11b+.Lu | Myeloid | 1.37E-08 | 3.87E-08 | 0.361228271 |
| #N/A | MF.11c-11b+.Lu | Myeloid | 1.28E-08 | 3.98E-08 | 0.373827996 |
| AD Braak | MF.11cloSer.Salm3.SI | Myeloid | 3.09E-08 | 1.89E-08 | 0.05040051 |
| ADNC Score | MF.11cloSer.Salm3.SI | Myeloid | 1.17E-08 | 6.71E-08 | 0.430687315 |
| AP (any/none) | MF.11cloSer.Salm3.SI | Myeloid | 3.42E-08 | 2.47E-08 | 0.083183016 |
| Arteriolosclerosis | MF.11cloSer.Salm3.SI | Myeloid | 4.27E-08 | 3.08E-08 | 0.082868815 |
| Arteriolosclerosis (any | MF.11cloSer.Salm3.SI | Myeloid | 5.23E-08 | 3.18E-08 | 0.050379769 |
| Atherosclerosis | MF.11cloSer.Salm3.SI | Myeloid | -1.76E-08 | 2.13E-08 | 0.796645695 |
| Atherosclerosis (any/r | MF.11cloSer.Salm3.SI | Myeloid | -1.37E-08 | 2.11E-08 | 0.741091484 |
| B Score | MF.11cloSer.Salm3.SI | Myeloid | 8.50E-09 | 1.75E-08 | 0.313708248 |
| CAA | MF.11cloSer.Salm3.SI | Myeloid | 2.22E-08 | 2.39E-08 | 0.176075672 |
| CAA (any/none) | MF.11cloSer.Salm3.SI | Myeloid | 2.30E-08 | 2.45E-08 | 0.173694449 |
| CERAD (C Score) | MF.11cloSer.Salm3.SI | Myeloid | 4.82E-08 | 1.93E-08 | 0.006339813 |
| Infarcts/lacunes | MF.11cloSer.Salm3.SI | Myeloid | 4.84E-08 | 2.57E-08 | 0.029976041 |
| PD Braak | MF.11cloSer.Salm3.SI | Myeloid | 4.62E-08 | 2.30E-08 | 0.022157926 |
| PD Braak (any/none) | MF.11cloSer.Salm3.SI | Myeloid | 3.75E-08 | 2.15E-08 | 0.0403303 |
| TDP-43 (any/none) | MF.11cloSer.Salm3.SI | Myeloid | -1.10E-07 | 5.23E-08 | 0.982198274 |
| Thal Phase | MF.11cloSer.Salm3.SI | Myeloid | 6.30E-08 | 8.12E-08 | 0.219181642 |
| WMR (any/none) | MF.11cloSer.Salm3.SI | Myeloid | -2.16E-08 | 3.96E-08 | 0.707137921 |
| #N/A | MF.11cloSer.Salm3.SI | Myeloid | -1.02E-08 | 3.92E-08 | 0.602303429 |
| AD Braak | MF.11cloSer.SI | Myeloid | 2.51E-08 | 1.76E-08 | 0.077717174 |
| ADNC Score | MF.11cloSer.SI | Myeloid | 5.14E-08 | 6.02E-08 | 0.196724692 |
| AP (any/none) | MF.11cloSer.SI | Myeloid | 2.95E-08 | 2.31E-08 | 0.100670191 |
| Arteriolosclerosis | MF.11cloSer.SI | Myeloid | 3.82E-08 | 2.87E-08 | 0.091824408 |
| Arteriolosclerosis (any | MF.11cloSer.SI | Myeloid | 4.73E-08 | 3.05E-08 | 0.060109528 |
| Atherosclerosis | MF.11cloSer.SI | Myeloid | -1.78E-08 | 2.38E-08 | 0.772810961 |
| Atherosclerosis (any/r | MF.11cloSer.SI | Myeloid | 2.06E-09 | 2.16E-08 | 0.462105393 |
| B Score | MF.11cloSer.SI | Myeloid | 1.84E-08 | 1.76E-08 | 0.147718887 |
| CAA | MF.11cloSer.SI | Myeloid | -1.33E-08 | 2.20E-08 | 0.727286568 |
| CAA (any/none) | MF.11cloSer.SI | Myeloid | -6.41E-09 | 2.26E-08 | 0.611579226 |
| CERAD (C Score) | MF.11cloSer.SI | Myeloid | 3.49E-08 | 1.87E-08 | 0.030781471 |
| Infarcts/lacunes | MF.11cloSer.SI | Myeloid | 4.13E-08 | 2.58E-08 | 0.054483045 |
| PD Braak | MF.11cloSer.SI | Myeloid | 3.80E-08 | 2.16E-08 | 0.039511272 |
| PD Braak (any/none) | MF.11cloSer.SI | Myeloid | 2.99E-08 | 2.02E-08 | 0.069268647 |

| | | | | | |
|---|---|---|---|---|---|
| TDP-43 (any/none) | MF.11cloSer.SI | Myeloid | -1.09E-07 | 5.29E-08 | 0.980467065 |
| Thal Phase | MF.11cloSer.SI | Myeloid | 6.28E-08 | 7.06E-08 | 0.186947479 |
| WMR (any/none) | MF.11cloSer.SI | Myeloid | 1.71E-08 | 4.25E-08 | 0.343289373 |
| #N/A | MF.11cloSer.SI | Myeloid | -3.24E-08 | 3.88E-08 | 0.798073483 |
| AD Braak | MF.169+11chi.SLN | Myeloid | 1.62E-09 | 1.58E-08 | 0.459182208 |
| ADNC Score | MF.169+11chi.SLN | Myeloid | 7.69E-08 | 5.98E-08 | 0.099156415 |
| AP (any/none) | MF.169+11chi.SLN | Myeloid | 3.55E-08 | 2.20E-08 | 0.052914337 |
| Arteriolosclerosis | MF.169+11chi.SLN | Myeloid | -1.09E-08 | 2.35E-08 | 0.678325705 |
| Arteriolosclerosis (any | MF.169+11chi.SLN | Myeloid | -2.29E-08 | 2.69E-08 | 0.80286789 |
| Atherosclerosis | MF.169+11chi.SLN | Myeloid | -1.58E-08 | 2.26E-08 | 0.756821191 |
| Atherosclerosis (any/r | MF.169+11chi.SLN | Myeloid | 9.37E-09 | 2.39E-08 | 0.347464242 |
| B Score | MF.169+11chi.SLN | Myeloid | -8.65E-09 | 1.48E-08 | 0.720285831 |
| CAA | MF.169+11chi.SLN | Myeloid | 3.23E-08 | 2.46E-08 | 0.094839072 |
| CAA (any/none) | MF.169+11chi.SLN | Myeloid | 1.12E-08 | 2.17E-08 | 0.302923432 |
| CERAD (C Score) | MF.169+11chi.SLN | Myeloid | 2.31E-08 | 1.85E-08 | 0.104974892 |
| Infarcts/lacunes | MF.169+11chi.SLN | Myeloid | -1.74E-09 | 2.04E-08 | 0.534115786 |
| PD Braak | MF.169+11chi.SLN | Myeloid | 3.12E-08 | 2.16E-08 | 0.074219119 |
| PD Braak (any/none) | MF.169+11chi.SLN | Myeloid | 3.22E-08 | 2.11E-08 | 0.063649158 |
| TDP-43 (any/none) | MF.169+11chi.SLN | Myeloid | -7.05E-08 | 5.08E-08 | 0.917631126 |
| Thal Phase | MF.169+11chi.SLN | Myeloid | 1.76E-08 | 7.62E-08 | 0.40889394 |
| WMR (any/none) | MF.169+11chi.SLN | Myeloid | -3.03E-08 | 3.61E-08 | 0.799500866 |
| #N/A | MF.169+11chi.SLN | Myeloid | -6.02E-08 | 4.00E-08 | 0.933907804 |
| AD Braak | MF.480hi.LV.Naive | Myeloid | 2.15E-08 | 1.78E-08 | 0.112822987 |
| ADNC Score | MF.480hi.LV.Naive | Myeloid | 8.52E-09 | 5.76E-08 | 0.441210681 |
| AP (any/none) | MF.480hi.LV.Naive | Myeloid | 3.74E-08 | 2.19E-08 | 0.043948367 |
| Arteriolosclerosis | MF.480hi.LV.Naive | Myeloid | 2.69E-08 | 2.83E-08 | 0.170588065 |
| Arteriolosclerosis (any | MF.480hi.LV.Naive | Myeloid | 3.07E-08 | 2.85E-08 | 0.141284313 |
| Atherosclerosis | MF.480hi.LV.Naive | Myeloid | -3.29E-08 | 2.32E-08 | 0.922422319 |
| Atherosclerosis (any/r | MF.480hi.LV.Naive | Myeloid | -1.09E-08 | 2.14E-08 | 0.695089471 |
| B Score | MF.480hi.LV.Naive | Myeloid | 3.32E-08 | 1.73E-08 | 0.027109906 |
| CAA | MF.480hi.LV.Naive | Myeloid | -7.86E-09 | 2.18E-08 | 0.640613143 |
| CAA (any/none) | MF.480hi.LV.Naive | Myeloid | -3.68E-09 | 2.04E-08 | 0.571528878 |
| CERAD (C Score) | MF.480hi.LV.Naive | Myeloid | 2.35E-08 | 1.88E-08 | 0.104976585 |
| Infarcts/lacunes | MF.480hi.LV.Naive | Myeloid | 1.30E-08 | 2.36E-08 | 0.291240857 |

| | | | | | |
|---|---|---|---|---|---|
| PD Braak | MF.480hi.LV.Naive | Myeloid | 5.11E-08 | 2.08E-08 | 0.007112461 |
| PD Braak (any/none) | MF.480hi.LV.Naive | Myeloid | 3.93E-08 | 2.05E-08 | 0.027737166 |
| TDP-43 (any/none) | MF.480hi.LV.Naive | Myeloid | -6.86E-08 | 5.22E-08 | 0.905517576 |
| Thal Phase | MF.480hi.LV.Naive | Myeloid | -4.78E-08 | 6.90E-08 | 0.755963227 |
| WMR (any/none) | MF.480hi.LV.Naive | Myeloid | -1.10E-08 | 3.42E-08 | 0.626246658 |
| #N/A | MF.480hi.LV.Naive | Myeloid | 2.43E-08 | 4.37E-08 | 0.289245338 |
| AD Braak | MF.480int.LV.Naive | Myeloid | 4.27E-09 | 1.60E-08 | 0.39474256 |
| ADNC Score | MF.480int.LV.Naive | Myeloid | -2.82E-08 | 5.76E-08 | 0.687429839 |
| AP (any/none) | MF.480int.LV.Naive | Myeloid | -8.10E-09 | 2.07E-08 | 0.652174507 |
| Arteriolosclerosis | MF.480int.LV.Naive | Myeloid | 2.85E-08 | 2.89E-08 | 0.162197802 |
| Arteriolosclerosis (any | MF.480int.LV.Naive | Myeloid | 1.00E-08 | 2.76E-08 | 0.35853827 |
| Atherosclerosis | MF.480int.LV.Naive | Myeloid | -2.25E-08 | 2.04E-08 | 0.865011691 |
| Atherosclerosis (any/r | MF.480int.LV.Naive | Myeloid | -2.58E-08 | 2.06E-08 | 0.894677426 |
| B Score | MF.480int.LV.Naive | Myeloid | 1.43E-08 | 1.63E-08 | 0.19128235 |
| CAA | MF.480int.LV.Naive | Myeloid | -1.89E-08 | 2.22E-08 | 0.803442921 |
| CAA (any/none) | MF.480int.LV.Naive | Myeloid | -1.48E-08 | 1.97E-08 | 0.774170462 |
| CERAD (C Score) | MF.480int.LV.Naive | Myeloid | -9.86E-10 | 1.63E-08 | 0.524125238 |
| Infarcts/lacunes | MF.480int.LV.Naive | Myeloid | 2.48E-08 | 2.36E-08 | 0.146425159 |
| PD Braak | MF.480int.LV.Naive | Myeloid | 1.71E-08 | 1.98E-08 | 0.193805534 |
| PD Braak (any/none) | MF.480int.LV.Naive | Myeloid | 1.19E-08 | 1.95E-08 | 0.270853537 |
| TDP-43 (any/none) | MF.480int.LV.Naive | Myeloid | -6.99E-08 | 5.32E-08 | 0.905749556 |
| Thal Phase | MF.480int.LV.Naive | Myeloid | -1.23E-07 | 7.09E-08 | 0.958617223 |
| WMR (any/none) | MF.480int.LV.Naive | Myeloid | -4.03E-08 | 3.22E-08 | 0.895107106 |
| #N/A | MF.480int.LV.Naive | Myeloid | -1.79E-08 | 4.12E-08 | 0.668118439 |
| AD Braak | MF.Alv.Lu | Myeloid | 2.57E-08 | 1.85E-08 | 0.083112955 |
| ADNC Score | MF.Alv.Lu | Myeloid | 1.34E-07 | 6.10E-08 | 0.014306078 |
| AP (any/none) | MF.Alv.Lu | Myeloid | 2.34E-08 | 2.42E-08 | 0.166851262 |
| Arteriolosclerosis | MF.Alv.Lu | Myeloid | 5.45E-08 | 2.82E-08 | 0.026944887 |
| Arteriolosclerosis (any | MF.Alv.Lu | Myeloid | 2.17E-08 | 2.84E-08 | 0.222494579 |
| Atherosclerosis | MF.Alv.Lu | Myeloid | 5.95E-10 | 2.25E-08 | 0.489464533 |
| Atherosclerosis (any/r | MF.Alv.Lu | Myeloid | 1.70E-08 | 2.28E-08 | 0.228137827 |
| B Score | MF.Alv.Lu | Myeloid | 3.31E-08 | 1.90E-08 | 0.040838566 |
| CAA | MF.Alv.Lu | Myeloid | -1.49E-08 | 2.31E-08 | 0.740872242 |
| CAA (any/none) | MF.Alv.Lu | Myeloid | -6.06E-09 | 2.15E-08 | 0.611013605 |

| | | | | | |
|---|---|---|---|---|---|
| CERAD (C Score) | MF.Alv.Lu | Myeloid | 3.61E-08 | 1.93E-08 | 0.031153106 |
| Infarcts/lacunes | MF.Alv.Lu | Myeloid | 6.99E-08 | 2.32E-08 | 0.001296993 |
| PD Braak | MF.Alv.Lu | Myeloid | 4.22E-08 | 2.17E-08 | 0.026159282 |
| PD Braak (any/none) | MF.Alv.Lu | Myeloid | 3.59E-08 | 2.12E-08 | 0.045539888 |
| TDP-43 (any/none) | MF.Alv.Lu | Myeloid | -1.06E-07 | 4.74E-08 | 0.987183037 |
| Thal Phase | MF.Alv.Lu | Myeloid | 1.37E-08 | 7.80E-08 | 0.430114017 |
| WMR (any/none) | MF.Alv.Lu | Myeloid | -1.45E-08 | 3.23E-08 | 0.673428702 |
| #N/A | MF.Alv.Lu | Myeloid | 5.63E-08 | 4.13E-08 | 0.086273512 |
| AD Braak | MF.AT.v2 | Myeloid | 3.20E-08 | 1.56E-08 | 0.019965103 |
| ADNC Score | MF.AT.v2 | Myeloid | 1.79E-08 | 5.34E-08 | 0.369009114 |
| AP (any/none) | MF.AT.v2 | Myeloid | 4.01E-08 | 2.24E-08 | 0.036629355 |
| Arteriolosclerosis | MF.AT.v2 | Myeloid | 4.34E-08 | 3.04E-08 | 0.07641268 |
| Arteriolosclerosis (any | MF.AT.v2 | Myeloid | 3.04E-08 | 3.06E-08 | 0.160275104 |
| Atherosclerosis | MF.AT.v2 | Myeloid | -8.67E-09 | 2.17E-08 | 0.655006398 |
| Atherosclerosis (any/r | MF.AT.v2 | Myeloid | -3.85E-09 | 2.14E-08 | 0.5714198 |
| B Score | MF.AT.v2 | Myeloid | 3.37E-08 | 1.57E-08 | 0.015688837 |
| CAA | MF.AT.v2 | Myeloid | 8.77E-10 | 2.24E-08 | 0.484386575 |
| CAA (any/none) | MF.AT.v2 | Myeloid | -2.26E-08 | 2.12E-08 | 0.856409059 |
| CERAD (C Score) | MF.AT.v2 | Myeloid | 2.96E-08 | 1.76E-08 | 0.046006021 |
| Infarcts/lacunes | MF.AT.v2 | Myeloid | 4.43E-08 | 2.24E-08 | 0.023968756 |
| PD Braak | MF.AT.v2 | Myeloid | 4.69E-08 | 2.26E-08 | 0.019245734 |
| PD Braak (any/none) | MF.AT.v2 | Myeloid | 4.27E-08 | 2.11E-08 | 0.021507652 |
| TDP-43 (any/none) | MF.AT.v2 | Myeloid | -1.20E-07 | 5.29E-08 | 0.988335719 |
| Thal Phase | MF.AT.v2 | Myeloid | 3.09E-08 | 7.03E-08 | 0.329998002 |
| WMR (any/none) | MF.AT.v2 | Myeloid | 1.83E-08 | 3.85E-08 | 0.31711935 |
| #N/A | MF.AT.v2 | Myeloid | -1.95E-08 | 4.48E-08 | 0.668354077 |
| AD Braak | MF.BM | Myeloid | -1.15E-08 | 2.22E-08 | 0.698384635 |
| ADNC Score | MF.BM | Myeloid | -8.87E-08 | 7.44E-08 | 0.883203676 |
| AP (any/none) | MF.BM | Myeloid | -5.65E-08 | 2.35E-08 | 0.991821237 |
| Arteriolosclerosis | MF.BM | Myeloid | -2.95E-08 | 3.25E-08 | 0.81834042 |
| Arteriolosclerosis (any | MF.BM | Myeloid | -5.37E-08 | 3.42E-08 | 0.941642401 |
| Atherosclerosis | MF.BM | Myeloid | -1.82E-08 | 3.05E-08 | 0.723974477 |
| Atherosclerosis (any/r | MF.BM | Myeloid | -5.79E-09 | 2.92E-08 | 0.578685629 |
| B Score | MF.BM | Myeloid | -2.74E-08 | 2.22E-08 | 0.891074698 |

| | | | | | |
|---|---|---|---|---|---|
| CAA | MF.BM | Myeloid | -2.63E-08 | 2.92E-08 | 0.815658014 |
| CAA (any/none) | MF.BM | Myeloid | -1.55E-08 | 2.91E-08 | 0.702755754 |
| CERAD (C Score) | MF.BM | Myeloid | -1.34E-09 | 2.23E-08 | 0.524017338 |
| Infarcts/lacunes | MF.BM | Myeloid | -1.61E-09 | 2.89E-08 | 0.522205003 |
| PD Braak | MF.BM | Myeloid | 2.30E-08 | 3.17E-08 | 0.23428911 |
| PD Braak (any/none) | MF.BM | Myeloid | 2.30E-08 | 3.10E-08 | 0.228916911 |
| TDP-43 (any/none) | MF.BM | Myeloid | -1.05E-07 | 6.21E-08 | 0.954881309 |
| Thal Phase | MF.BM | Myeloid | -1.81E-07 | 9.45E-08 | 0.972633257 |
| WMR (any/none) | MF.BM | Myeloid | -6.74E-08 | 4.48E-08 | 0.933842363 |
| #N/A | MF.BM | Myeloid | -9.78E-08 | 4.87E-08 | 0.977763041 |
| AD Braak | MF.F480hi.ctrl.PC | Myeloid | 2.67E-08 | 1.53E-08 | 0.040621288 |
| ADNC Score | MF.F480hi.ctrl.PC | Myeloid | 4.04E-08 | 5.60E-08 | 0.235367442 |
| AP (any/none) | MF.F480hi.ctrl.PC | Myeloid | 1.49E-08 | 2.06E-08 | 0.234743532 |
| Arteriolosclerosis | MF.F480hi.ctrl.PC | Myeloid | 2.67E-08 | 2.60E-08 | 0.151814545 |
| Arteriolosclerosis (any | MF.F480hi.ctrl.PC | Myeloid | 7.09E-09 | 2.75E-08 | 0.398103532 |
| Atherosclerosis | MF.F480hi.ctrl.PC | Myeloid | -8.51E-09 | 1.99E-08 | 0.665160562 |
| Atherosclerosis (any/r | MF.F480hi.ctrl.PC | Myeloid | -3.00E-08 | 2.08E-08 | 0.926123233 |
| B Score | MF.F480hi.ctrl.PC | Myeloid | 2.69E-08 | 1.57E-08 | 0.042991436 |
| CAA | MF.F480hi.ctrl.PC | Myeloid | -1.31E-08 | 2.27E-08 | 0.717934014 |
| CAA (any/none) | MF.F480hi.ctrl.PC | Myeloid | -2.71E-08 | 2.08E-08 | 0.903488319 |
| CERAD (C Score) | MF.F480hi.ctrl.PC | Myeloid | -1.29E-08 | 1.82E-08 | 0.760022276 |
| Infarcts/lacunes | MF.F480hi.ctrl.PC | Myeloid | 2.85E-08 | 2.17E-08 | 0.094158507 |
| PD Braak | MF.F480hi.ctrl.PC | Myeloid | 3.23E-08 | 2.04E-08 | 0.056859706 |
| PD Braak (any/none) | MF.F480hi.ctrl.PC | Myeloid | 3.31E-08 | 2.07E-08 | 0.055405561 |
| TDP-43 (any/none) | MF.F480hi.ctrl.PC | Myeloid | -3.97E-08 | 4.81E-08 | 0.795524015 |
| Thal Phase | MF.F480hi.ctrl.PC | Myeloid | -7.53E-08 | 6.91E-08 | 0.862120872 |
| WMR (any/none) | MF.F480hi.ctrl.PC | Myeloid | -1.79E-08 | 3.32E-08 | 0.705467184 |
| #N/A | MF.F480hi.ctrl.PC | Myeloid | 5.88E-08 | 4.54E-08 | 0.097705814 |
| AD Braak | MF.F480hi.Gata6ko.PC | Myeloid | 2.31E-08 | 1.59E-08 | 0.073212531 |
| ADNC Score | MF.F480hi.Gata6ko.PC | Myeloid | 6.51E-08 | 6.23E-08 | 0.148163379 |
| AP (any/none) | MF.F480hi.Gata6ko.PC | Myeloid | 3.93E-08 | 2.21E-08 | 0.037617237 |
| Arteriolosclerosis | MF.F480hi.Gata6ko.PC | Myeloid | 2.27E-08 | 2.71E-08 | 0.2007093 |
| Arteriolosclerosis (any | MF.F480hi.Gata6ko.PC | Myeloid | -5.46E-09 | 2.80E-08 | 0.577364902 |
| Atherosclerosis | MF.F480hi.Gata6ko.PC | Myeloid | -2.29E-08 | 2.16E-08 | 0.855500599 |

| | | | | | |
|---|---|---|---|---|---|
| Atherosclerosis (any/r | MF.F480hi.Gata6ko.PC | Myeloid | -1.84E-08 | 2.14E-08 | 0.804934038 |
| B Score | MF.F480hi.Gata6ko.PC | Myeloid | 2.32E-08 | 1.58E-08 | 0.070593008 |
| CAA | MF.F480hi.Gata6ko.PC | Myeloid | 1.26E-08 | 2.29E-08 | 0.291098942 |
| CAA (any/none) | MF.F480hi.Gata6ko.PC | Myeloid | 2.06E-08 | 2.04E-08 | 0.156285842 |
| CERAD (C Score) | MF.F480hi.Gata6ko.PC | Myeloid | 1.94E-08 | 2.02E-08 | 0.167668592 |
| Infarcts/lacunes | MF.F480hi.Gata6ko.PC | Myeloid | 1.50E-08 | 2.29E-08 | 0.255822956 |
| PD Braak | MF.F480hi.Gata6ko.PC | Myeloid | 3.09E-08 | 2.02E-08 | 0.063062914 |
| PD Braak (any/none) | MF.F480hi.Gata6ko.PC | Myeloid | 3.11E-08 | 2.08E-08 | 0.067205281 |
| TDP-43 (any/none) | MF.F480hi.Gata6ko.PC | Myeloid | -4.45E-08 | 5.07E-08 | 0.809605081 |
| Thal Phase | MF.F480hi.Gata6ko.PC | Myeloid | 7.57E-09 | 7.45E-08 | 0.459553422 |
| WMR (any/none) | MF.F480hi.Gata6ko.PC | Myeloid | -1.51E-09 | 3.24E-08 | 0.518610171 |
| #N/A | MF.F480hi.Gata6ko.PC | Myeloid | 7.85E-08 | 4.33E-08 | 0.034885521 |
| AD Braak | MF.II+480lo.PC | Myeloid | 1.12E-08 | 1.71E-08 | 0.256907973 |
| ADNC Score | MF.II+480lo.PC | Myeloid | -1.59E-08 | 6.24E-08 | 0.600685226 |
| AP (any/none) | MF.II+480lo.PC | Myeloid | 4.84E-09 | 2.32E-08 | 0.417448029 |
| Arteriolosclerosis | MF.II+480lo.PC | Myeloid | 4.27E-08 | 3.21E-08 | 0.091870135 |
| Arteriolosclerosis (any | MF.II+480lo.PC | Myeloid | 2.81E-08 | 3.36E-08 | 0.20092064 |
| Atherosclerosis | MF.II+480lo.PC | Myeloid | -1.37E-08 | 2.53E-08 | 0.7053508 |
| Atherosclerosis (any/r | MF.II+480lo.PC | Myeloid | -9.37E-09 | 2.52E-08 | 0.645170498 |
| B Score | MF.II+480lo.PC | Myeloid | -3.04E-09 | 1.70E-08 | 0.571124152 |
| CAA | MF.II+480lo.PC | Myeloid | -9.52E-09 | 2.37E-08 | 0.655906558 |
| CAA (any/none) | MF.II+480lo.PC | Myeloid | -2.15E-08 | 2.31E-08 | 0.823836717 |
| CERAD (C Score) | MF.II+480lo.PC | Myeloid | 2.53E-08 | 2.11E-08 | 0.115708009 |
| Infarcts/lacunes | MF.II+480lo.PC | Myeloid | 6.22E-09 | 2.39E-08 | 0.397395504 |
| PD Braak | MF.II+480lo.PC | Myeloid | 2.42E-08 | 2.29E-08 | 0.145348449 |
| PD Braak (any/none) | MF.II+480lo.PC | Myeloid | 2.09E-08 | 2.31E-08 | 0.182683618 |
| TDP-43 (any/none) | MF.II+480lo.PC | Myeloid | -5.30E-08 | 5.69E-08 | 0.824332848 |
| Thal Phase | MF.II+480lo.PC | Myeloid | -5.98E-08 | 7.94E-08 | 0.774569689 |
| WMR (any/none) | MF.II+480lo.PC | Myeloid | 1.65E-08 | 3.59E-08 | 0.322458783 |
| #N/A | MF.II+480lo.PC | Myeloid | -1.90E-09 | 4.39E-08 | 0.517258527 |
| AD Braak | MF.II-480hi.PC | Myeloid | 1.70E-08 | 1.72E-08 | 0.161499783 |
| ADNC Score | MF.II-480hi.PC | Myeloid | 1.39E-08 | 6.08E-08 | 0.409896471 |
| AP (any/none) | MF.II-480hi.PC | Myeloid | 3.53E-09 | 2.13E-08 | 0.434220012 |
| Arteriolosclerosis | MF.II-480hi.PC | Myeloid | 3.60E-08 | 2.95E-08 | 0.110757733 |

| | | | | | |
|---|---|---|---|---|---|
| Arteriolosclerosis (any | MF.II-480hi.PC | Myeloid | 1.46E-08 | 3.04E-08 | 0.315208444 |
| Atherosclerosis | MF.II-480hi.PC | Myeloid | 4.77E-09 | 2.33E-08 | 0.418910888 |
| Atherosclerosis (any/r | MF.II-480hi.PC | Myeloid | -9.37E-09 | 2.08E-08 | 0.673873797 |
| B Score | MF.II-480hi.PC | Myeloid | 1.00E-08 | 1.66E-08 | 0.272827092 |
| CAA | MF.II-480hi.PC | Myeloid | 1.83E-09 | 2.15E-08 | 0.466201959 |
| CAA (any/none) | MF.II-480hi.PC | Myeloid | -7.62E-09 | 2.12E-08 | 0.640158299 |
| CERAD (C Score) | MF.II-480hi.PC | Myeloid | 2.80E-09 | 1.95E-08 | 0.443019648 |
| Infarcts/lacunes | MF.II-480hi.PC | Myeloid | -1.34E-08 | 2.30E-08 | 0.719673627 |
| PD Braak | MF.II-480hi.PC | Myeloid | 2.15E-08 | 2.19E-08 | 0.163116634 |
| PD Braak (any/none) | MF.II-480hi.PC | Myeloid | 2.26E-08 | 2.26E-08 | 0.158650893 |
| TDP-43 (any/none) | MF.II-480hi.PC | Myeloid | -6.29E-08 | 5.34E-08 | 0.880509915 |
| Thal Phase | MF.II-480hi.PC | Myeloid | -5.43E-08 | 7.18E-08 | 0.775306924 |
| WMR (any/none) | MF.II-480hi.PC | Myeloid | -1.72E-09 | 3.48E-08 | 0.519711553 |
| #N/A | MF.II-480hi.PC | Myeloid | 3.04E-08 | 3.87E-08 | 0.216324309 |
| AD Braak | MF.Lu | Myeloid | 1.48E-08 | 1.83E-08 | 0.209084021 |
| ADNC Score | MF.Lu | Myeloid | 1.45E-07 | 6.17E-08 | 0.009243238 |
| AP (any/none) | MF.Lu | Myeloid | 3.83E-08 | 2.47E-08 | 0.060458307 |
| Arteriolosclerosis | MF.Lu | Myeloid | 7.21E-08 | 2.87E-08 | 0.006080699 |
| Arteriolosclerosis (any | MF.Lu | Myeloid | 4.60E-08 | 2.94E-08 | 0.059002478 |
| Atherosclerosis | MF.Lu | Myeloid | 3.27E-09 | 2.30E-08 | 0.443402531 |
| Atherosclerosis (any/r | MF.Lu | Myeloid | -8.23E-09 | 2.11E-08 | 0.651581876 |
| B Score | MF.Lu | Myeloid | 3.11E-08 | 1.97E-08 | 0.057402308 |
| CAA | MF.Lu | Myeloid | -5.07E-09 | 2.41E-08 | 0.58336089 |
| CAA (any/none) | MF.Lu | Myeloid | -1.32E-08 | 2.30E-08 | 0.716935962 |
| CERAD (C Score) | MF.Lu | Myeloid | 2.90E-08 | 1.89E-08 | 0.062518063 |
| Infarcts/lacunes | MF.Lu | Myeloid | 3.66E-08 | 2.31E-08 | 0.056648641 |
| PD Braak | MF.Lu | Myeloid | 2.27E-08 | 1.92E-08 | 0.118111415 |
| PD Braak (any/none) | MF.Lu | Myeloid | 1.43E-08 | 1.86E-08 | 0.221405836 |
| TDP-43 (any/none) | MF.Lu | Myeloid | -7.94E-08 | 4.95E-08 | 0.9454973 |
| Thal Phase | MF.Lu | Myeloid | 7.45E-08 | 7.44E-08 | 0.158591257 |
| WMR (any/none) | MF.Lu | Myeloid | -2.92E-08 | 3.04E-08 | 0.831486307 |
| #N/A | MF.Lu | Myeloid | 4.55E-08 | 4.38E-08 | 0.149451653 |
| AD Braak | MF.Medl.SLN | Myeloid | 1.83E-09 | 1.63E-08 | 0.455254299 |
| ADNC Score | MF.Medl.SLN | Myeloid | 7.65E-08 | 5.01E-08 | 0.063215031 |

| | | | | | |
|---|---|---|---|---|---|
| AP (any/none) | MF.Medl.SLN | Myeloid | 9.33E-09 | 1.73E-08 | 0.294594999 |
| Arteriolosclerosis | MF.Medl.SLN | Myeloid | 1.62E-08 | 2.52E-08 | 0.260068107 |
| Arteriolosclerosis (any | MF.Medl.SLN | Myeloid | 6.26E-09 | 2.44E-08 | 0.398834136 |
| Atherosclerosis | MF.Medl.SLN | Myeloid | 4.48E-08 | 2.16E-08 | 0.018976144 |
| Atherosclerosis (any/r | MF.Medl.SLN | Myeloid | 2.48E-08 | 2.12E-08 | 0.120897174 |
| B Score | MF.Medl.SLN | Myeloid | 8.86E-09 | 1.72E-08 | 0.302968428 |
| CAA | MF.Medl.SLN | Myeloid | 2.43E-08 | 2.39E-08 | 0.154228025 |
| CAA (any/none) | MF.Medl.SLN | Myeloid | -1.51E-08 | 2.12E-08 | 0.761954437 |
| CERAD (C Score) | MF.Medl.SLN | Myeloid | 2.89E-08 | 1.79E-08 | 0.053157168 |
| Infarcts/lacunes | MF.Medl.SLN | Myeloid | 2.06E-08 | 2.04E-08 | 0.156873077 |
| PD Braak | MF.Medl.SLN | Myeloid | 2.56E-08 | 1.76E-08 | 0.072753376 |
| PD Braak (any/none) | MF.Medl.SLN | Myeloid | 2.15E-08 | 1.76E-08 | 0.110977116 |
| TDP-43 (any/none) | MF.Medl.SLN | Myeloid | -9.90E-08 | 5.10E-08 | 0.973832981 |
| Thal Phase | MF.Medl.SLN | Myeloid | -3.80E-08 | 7.13E-08 | 0.703015455 |
| WMR (any/none) | MF.Medl.SLN | Myeloid | -3.86E-09 | 3.24E-08 | 0.547441821 |
| #N/A | MF.Medl.SLN | Myeloid | -3.89E-08 | 4.06E-08 | 0.831197235 |
| AD Braak | MF.Microglia.CNS | Myeloid | -2.02E-09 | 1.97E-08 | 0.540714947 |
| ADNC Score | MF.Microglia.CNS | Myeloid | 1.96E-08 | 5.90E-08 | 0.370078579 |
| AP (any/none) | MF.Microglia.CNS | Myeloid | 2.78E-08 | 2.39E-08 | 0.121939322 |
| Arteriolosclerosis | MF.Microglia.CNS | Myeloid | 3.66E-08 | 2.85E-08 | 0.099420458 |
| Arteriolosclerosis (any | MF.Microglia.CNS | Myeloid | 1.02E-08 | 3.23E-08 | 0.376614537 |
| Atherosclerosis | MF.Microglia.CNS | Myeloid | 3.35E-08 | 2.62E-08 | 0.100456912 |
| Atherosclerosis (any/r | MF.Microglia.CNS | Myeloid | 3.22E-08 | 2.69E-08 | 0.115773491 |
| B Score | MF.Microglia.CNS | Myeloid | -2.63E-09 | 2.22E-08 | 0.54723368 |
| CAA | MF.Microglia.CNS | Myeloid | 2.77E-08 | 2.64E-08 | 0.146811022 |
| CAA (any/none) | MF.Microglia.CNS | Myeloid | 2.88E-08 | 2.48E-08 | 0.123176449 |
| CERAD (C Score) | MF.Microglia.CNS | Myeloid | 1.09E-08 | 2.09E-08 | 0.301693872 |
| Infarcts/lacunes | MF.Microglia.CNS | Myeloid | -1.13E-08 | 2.63E-08 | 0.665813905 |
| PD Braak | MF.Microglia.CNS | Myeloid | 2.42E-08 | 2.42E-08 | 0.15820584 |
| PD Braak (any/none) | MF.Microglia.CNS | Myeloid | 2.88E-08 | 2.36E-08 | 0.111562975 |
| TDP-43 (any/none) | MF.Microglia.CNS | Myeloid | -3.27E-08 | 5.28E-08 | 0.732579072 |
| Thal Phase | MF.Microglia.CNS | Myeloid | -1.55E-08 | 8.05E-08 | 0.576507231 |
| WMR (any/none) | MF.Microglia.CNS | Myeloid | -1.96E-08 | 3.78E-08 | 0.69785536 |
| #N/A | MF.Microglia.CNS | Myeloid | 2.22E-08 | 4.30E-08 | 0.303181482 |

| | | | | | |
|---|---|---|---|---|---|
| AD Braak | MF.PPAR-.Lu | Myeloid | 3.06E-08 | 1.68E-08 | 0.034143381 |
| ADNC Score | MF.PPAR-.Lu | Myeloid | 9.16E-08 | 5.62E-08 | 0.051643943 |
| AP (any/none) | MF.PPAR-.Lu | Myeloid | 4.51E-08 | 2.49E-08 | 0.035233457 |
| Arteriolosclerosis | MF.PPAR-.Lu | Myeloid | 7.87E-08 | 2.90E-08 | 0.003347351 |
| Arteriolosclerosis (any | MF.PPAR-.Lu | Myeloid | 7.55E-08 | 2.99E-08 | 0.005775919 |
| Atherosclerosis | MF.PPAR-.Lu | Myeloid | 9.23E-09 | 2.21E-08 | 0.33839318 |
| Atherosclerosis (any/r | MF.PPAR-.Lu | Myeloid | 5.10E-09 | 2.21E-08 | 0.40874073 |
| B Score | MF.PPAR-.Lu | Myeloid | 2.06E-08 | 1.77E-08 | 0.122310493 |
| CAA | MF.PPAR-.Lu | Myeloid | -2.79E-08 | 2.17E-08 | 0.900726173 |
| CAA (any/none) | MF.PPAR-.Lu | Myeloid | -1.95E-08 | 2.12E-08 | 0.821030423 |
| CERAD (C Score) | MF.PPAR-.Lu | Myeloid | 2.96E-08 | 1.91E-08 | 0.059969201 |
| Infarcts/lacunes | MF.PPAR-.Lu | Myeloid | 4.21E-08 | 2.39E-08 | 0.039266264 |
| PD Braak | MF.PPAR-.Lu | Myeloid | 3.67E-08 | 1.98E-08 | 0.032119813 |
| PD Braak (any/none) | MF.PPAR-.Lu | Myeloid | 3.80E-08 | 1.98E-08 | 0.027637837 |
| TDP-43 (any/none) | MF.PPAR-.Lu | Myeloid | -9.83E-08 | 5.13E-08 | 0.972338349 |
| Thal Phase | MF.PPAR-.Lu | Myeloid | 4.13E-08 | 7.35E-08 | 0.286860884 |
| WMR (any/none) | MF.PPAR-.Lu | Myeloid | -1.36E-08 | 3.16E-08 | 0.666380261 |
| #N/A | MF.PPAR-.Lu | Myeloid | 3.68E-08 | 4.39E-08 | 0.201418053 |
| AD Braak | MF.RP.Sp | Myeloid | 1.43E-08 | 1.66E-08 | 0.194767627 |
| ADNC Score | MF.RP.Sp | Myeloid | 1.94E-08 | 6.20E-08 | 0.377439953 |
| AP (any/none) | MF.RP.Sp | Myeloid | 1.50E-08 | 2.21E-08 | 0.247845231 |
| Arteriolosclerosis | MF.RP.Sp | Myeloid | 7.32E-08 | 2.83E-08 | 0.004844608 |
| Arteriolosclerosis (any | MF.RP.Sp | Myeloid | 6.04E-08 | 2.90E-08 | 0.018678098 |
| Atherosclerosis | MF.RP.Sp | Myeloid | 1.77E-09 | 2.41E-08 | 0.47065188 |
| Atherosclerosis (any/r | MF.RP.Sp | Myeloid | 4.10E-09 | 2.24E-08 | 0.427286851 |
| B Score | MF.RP.Sp | Myeloid | 1.85E-08 | 1.66E-08 | 0.131531407 |
| CAA | MF.RP.Sp | Myeloid | -7.88E-09 | 2.33E-08 | 0.632537006 |
| CAA (any/none) | MF.RP.Sp | Myeloid | -2.42E-08 | 2.07E-08 | 0.879195591 |
| CERAD (C Score) | MF.RP.Sp | Myeloid | 9.44E-09 | 1.96E-08 | 0.315014575 |
| Infarcts/lacunes | MF.RP.Sp | Myeloid | 1.77E-08 | 2.11E-08 | 0.20000656 |
| PD Braak | MF.RP.Sp | Myeloid | 2.05E-08 | 1.86E-08 | 0.135776357 |
| PD Braak (any/none) | MF.RP.Sp | Myeloid | 1.87E-08 | 1.89E-08 | 0.161578995 |
| TDP-43 (any/none) | MF.RP.Sp | Myeloid | -5.07E-08 | 4.80E-08 | 0.854934823 |
| Thal Phase | MF.RP.Sp | Myeloid | 8.78E-09 | 7.15E-08 | 0.451086277 |

| | | | | | |
|---|---|---|---|---|---|
| WMR (any/none) | MF.RP.Sp | Myeloid | -2.38E-08 | 3.33E-08 | 0.76289281 |
| #N/A | MF.RP.Sp | Myeloid | -2.71E-08 | 3.83E-08 | 0.760286502 |
| AD Braak | MF.Sbcaps.SLN | Myeloid | 1.27E-09 | 1.85E-08 | 0.47269443 |
| ADNC Score | MF.Sbcaps.SLN | Myeloid | -2.68E-08 | 5.88E-08 | 0.675800975 |
| AP (any/none) | MF.Sbcaps.SLN | Myeloid | -1.97E-08 | 1.82E-08 | 0.860198607 |
| Arteriolosclerosis | MF.Sbcaps.SLN | Myeloid | 1.34E-08 | 2.47E-08 | 0.293182644 |
| Arteriolosclerosis (any | MF.Sbcaps.SLN | Myeloid | 6.01E-09 | 2.72E-08 | 0.412484521 |
| Atherosclerosis | MF.Sbcaps.SLN | Myeloid | 1.95E-08 | 2.16E-08 | 0.183216104 |
| Atherosclerosis (any/r | MF.Sbcaps.SLN | Myeloid | 6.72E-09 | 2.15E-08 | 0.377332276 |
| B Score | MF.Sbcaps.SLN | Myeloid | 1.17E-09 | 1.85E-08 | 0.474804571 |
| CAA | MF.Sbcaps.SLN | Myeloid | 1.20E-08 | 2.30E-08 | 0.300145674 |
| CAA (any/none) | MF.Sbcaps.SLN | Myeloid | -2.09E-08 | 2.11E-08 | 0.838778666 |
| CERAD (C Score) | MF.Sbcaps.SLN | Myeloid | 2.94E-08 | 1.74E-08 | 0.045958078 |
| Infarcts/lacunes | MF.Sbcaps.SLN | Myeloid | 4.77E-09 | 2.29E-08 | 0.417513049 |
| PD Braak | MF.Sbcaps.SLN | Myeloid | 6.39E-09 | 2.04E-08 | 0.377088329 |
| PD Braak (any/none) | MF.Sbcaps.SLN | Myeloid | 9.21E-09 | 2.01E-08 | 0.323473549 |
| TDP-43 (any/none) | MF.Sbcaps.SLN | Myeloid | -2.30E-08 | 4.99E-08 | 0.677910336 |
| Thal Phase | MF.Sbcaps.SLN | Myeloid | -6.72E-08 | 7.32E-08 | 0.820801015 |
| WMR (any/none) | MF.Sbcaps.SLN | Myeloid | -8.11E-09 | 3.26E-08 | 0.598116881 |
| #N/A | MF.Sbcaps.SLN | Myeloid | -1.79E-08 | 4.40E-08 | 0.657890021 |
| AD Braak | MF.Thio5.II+480int.PC | Myeloid | 2.94E-08 | 1.64E-08 | 0.036732955 |
| ADNC Score | MF.Thio5.II+480int.PC | Myeloid | 3.22E-08 | 6.17E-08 | 0.301097027 |
| AP (any/none) | MF.Thio5.II+480int.PC | Myeloid | 1.59E-08 | 2.38E-08 | 0.251705692 |
| Arteriolosclerosis | MF.Thio5.II+480int.PC | Myeloid | 6.22E-08 | 3.02E-08 | 0.019866526 |
| Arteriolosclerosis (any | MF.Thio5.II+480int.PC | Myeloid | 5.43E-08 | 3.31E-08 | 0.050626568 |
| Atherosclerosis | MF.Thio5.II+480int.PC | Myeloid | 5.47E-10 | 2.39E-08 | 0.49087516 |
| Atherosclerosis (any/r | MF.Thio5.II+480int.PC | Myeloid | -1.27E-08 | 2.31E-08 | 0.708660615 |
| B Score | MF.Thio5.II+480int.PC | Myeloid | 1.83E-08 | 1.62E-08 | 0.128534127 |
| CAA | MF.Thio5.II+480int.PC | Myeloid | -1.65E-09 | 2.29E-08 | 0.528692696 |
| CAA (any/none) | MF.Thio5.II+480int.PC | Myeloid | -9.69E-09 | 2.19E-08 | 0.671184042 |
| CERAD (C Score) | MF.Thio5.II+480int.PC | Myeloid | 3.37E-08 | 2.10E-08 | 0.05425067 |
| Infarcts/lacunes | MF.Thio5.II+480int.PC | Myeloid | 1.83E-08 | 2.24E-08 | 0.20672621 |
| PD Braak | MF.Thio5.II+480int.PC | Myeloid | 3.82E-08 | 2.28E-08 | 0.047367369 |
| PD Braak (any/none) | MF.Thio5.II+480int.PC | Myeloid | 3.11E-08 | 2.28E-08 | 0.086529451 |

| | | | | | |
|---|---|---|---|---|---|
| TDP-43 (any/none) | MF.Thio5.II+480int.PC | Myeloid | -6.61E-08 | 5.15E-08 | 0.900495736 |
| Thal Phase | MF.Thio5.II+480int.PC | Myeloid | 2.70E-08 | 7.40E-08 | 0.357536321 |
| WMR (any/none) | MF.Thio5.II+480int.PC | Myeloid | -6.52E-08 | 3.63E-08 | 0.963563264 |
| #N/A | MF.Thio5.II+480int.PC | Myeloid | 3.65E-08 | 4.28E-08 | 0.1967921 |
| AD Braak | MF.Thio5.II+480lo.PC | Myeloid | 1.83E-08 | 1.74E-08 | 0.146790863 |
| ADNC Score | MF.Thio5.II+480lo.PC | Myeloid | 2.55E-08 | 6.54E-08 | 0.34812536 |
| AP (any/none) | MF.Thio5.II+480lo.PC | Myeloid | 1.02E-08 | 2.34E-08 | 0.331350276 |
| Arteriolosclerosis | MF.Thio5.II+480lo.PC | Myeloid | 2.93E-08 | 2.93E-08 | 0.158659475 |
| Arteriolosclerosis (any | MF.Thio5.II+480lo.PC | Myeloid | 2.44E-08 | 3.14E-08 | 0.218624903 |
| Atherosclerosis | MF.Thio5.II+480lo.PC | Myeloid | -2.20E-08 | 2.56E-08 | 0.805099558 |
| Atherosclerosis (any/r | MF.Thio5.II+480lo.PC | Myeloid | -2.14E-08 | 2.61E-08 | 0.793749362 |
| B Score | MF.Thio5.II+480lo.PC | Myeloid | 2.53E-09 | 1.69E-08 | 0.440533827 |
| CAA | MF.Thio5.II+480lo.PC | Myeloid | 7.48E-10 | 2.42E-08 | 0.487688633 |
| CAA (any/none) | MF.Thio5.II+480lo.PC | Myeloid | -6.07E-09 | 2.33E-08 | 0.602839769 |
| CERAD (C Score) | MF.Thio5.II+480lo.PC | Myeloid | 2.20E-08 | 2.17E-08 | 0.15457959 |
| Infarcts/lacunes | MF.Thio5.II+480lo.PC | Myeloid | 1.14E-08 | 2.34E-08 | 0.31333738 |
| PD Braak | MF.Thio5.II+480lo.PC | Myeloid | 4.02E-08 | 2.27E-08 | 0.038159009 |
| PD Braak (any/none) | MF.Thio5.II+480lo.PC | Myeloid | 3.41E-08 | 2.24E-08 | 0.063906816 |
| TDP-43 (any/none) | MF.Thio5.II+480lo.PC | Myeloid | -4.64E-08 | 5.48E-08 | 0.801262922 |
| Thal Phase | MF.Thio5.II+480lo.PC | Myeloid | -3.49E-08 | 7.76E-08 | 0.673586465 |
| WMR (any/none) | MF.Thio5.II+480lo.PC | Myeloid | -1.07E-08 | 3.86E-08 | 0.609372376 |
| #N/A | MF.Thio5.II+480lo.PC | Myeloid | 2.42E-08 | 4.48E-08 | 0.294896298 |
| AD Braak | MF.Thio5.II-480hi.PC | Myeloid | 2.87E-08 | 1.78E-08 | 0.053356666 |
| ADNC Score | MF.Thio5.II-480hi.PC | Myeloid | 6.18E-08 | 6.12E-08 | 0.156346767 |
| AP (any/none) | MF.Thio5.II-480hi.PC | Myeloid | 8.45E-09 | 2.15E-08 | 0.347432675 |
| Arteriolosclerosis | MF.Thio5.II-480hi.PC | Myeloid | 6.24E-08 | 2.84E-08 | 0.013992132 |
| Arteriolosclerosis (any | MF.Thio5.II-480hi.PC | Myeloid | 3.21E-08 | 3.15E-08 | 0.154244019 |
| Atherosclerosis | MF.Thio5.II-480hi.PC | Myeloid | 2.98E-08 | 2.29E-08 | 0.096871635 |
| Atherosclerosis (any/r | MF.Thio5.II-480hi.PC | Myeloid | -3.14E-09 | 2.17E-08 | 0.557542714 |
| B Score | MF.Thio5.II-480hi.PC | Myeloid | 2.01E-08 | 1.75E-08 | 0.125460771 |
| CAA | MF.Thio5.II-480hi.PC | Myeloid | 1.82E-08 | 2.26E-08 | 0.211060863 |
| CAA (any/none) | MF.Thio5.II-480hi.PC | Myeloid | -4.44E-10 | 2.22E-08 | 0.507972057 |
| CERAD (C Score) | MF.Thio5.II-480hi.PC | Myeloid | 2.34E-08 | 1.94E-08 | 0.114876311 |
| Infarcts/lacunes | MF.Thio5.II-480hi.PC | Myeloid | -1.19E-08 | 2.17E-08 | 0.708940845 |

| | | | | | |
|---|---|---|---|---|---|
| PD Braak | MF.Thio5.II-480hi.PC | Myeloid | 1.81E-08 | 2.01E-08 | 0.183787936 |
| PD Braak (any/none) | MF.Thio5.II-480hi.PC | Myeloid | 2.11E-08 | 2.09E-08 | 0.155357332 |
| TDP-43 (any/none) | MF.Thio5.II-480hi.PC | Myeloid | -1.34E-09 | 6.77E-08 | 0.507892057 |
| Thal Phase | MF.Thio5.II-480hi.PC | Myeloid | 6.09E-09 | 7.67E-08 | 0.46834249 |
| WMR (any/none) | MF.Thio5.II-480hi.PC | Myeloid | -4.18E-08 | 3.57E-08 | 0.879453063 |
| #N/A | MF.Thio5.II-480hi.PC | Myeloid | -2.53E-08 | 4.11E-08 | 0.731237363 |
| AD Braak | MF.Thio5.II-480int.PC | Myeloid | 3.73E-08 | 1.83E-08 | 0.0208208 |
| ADNC Score | MF.Thio5.II-480int.PC | Myeloid | 3.94E-08 | 6.03E-08 | 0.256460881 |
| AP (any/none) | MF.Thio5.II-480int.PC | Myeloid | 1.33E-08 | 2.35E-08 | 0.286387137 |
| Arteriolosclerosis | MF.Thio5.II-480int.PC | Myeloid | 6.05E-08 | 2.76E-08 | 0.014223482 |
| Arteriolosclerosis (any | MF.Thio5.II-480int.PC | Myeloid | 4.59E-08 | 3.11E-08 | 0.069912926 |
| Atherosclerosis | MF.Thio5.II-480int.PC | Myeloid | 3.38E-08 | 2.45E-08 | 0.083522885 |
| Atherosclerosis (any/r | MF.Thio5.II-480int.PC | Myeloid | 5.11E-09 | 2.39E-08 | 0.41545216 |
| B Score | MF.Thio5.II-480int.PC | Myeloid | 3.88E-08 | 1.95E-08 | 0.02315148 |
| CAA | MF.Thio5.II-480int.PC | Myeloid | 1.60E-08 | 2.41E-08 | 0.252895837 |
| CAA (any/none) | MF.Thio5.II-480int.PC | Myeloid | 1.32E-08 | 2.34E-08 | 0.287280611 |
| CERAD (C Score) | MF.Thio5.II-480int.PC | Myeloid | 4.21E-08 | 2.11E-08 | 0.023008041 |
| Infarcts/lacunes | MF.Thio5.II-480int.PC | Myeloid | 2.62E-08 | 2.17E-08 | 0.112834653 |
| PD Braak | MF.Thio5.II-480int.PC | Myeloid | 6.00E-08 | 2.20E-08 | 0.003208436 |
| PD Braak (any/none) | MF.Thio5.II-480int.PC | Myeloid | 6.00E-08 | 2.16E-08 | 0.002761101 |
| TDP-43 (any/none) | MF.Thio5.II-480int.PC | Myeloid | -2.51E-08 | 6.04E-08 | 0.660994446 |
| Thal Phase | MF.Thio5.II-480int.PC | Myeloid | 1.82E-08 | 7.93E-08 | 0.40940035 |
| WMR (any/none) | MF.Thio5.II-480int.PC | Myeloid | -5.03E-08 | 3.86E-08 | 0.903743403 |
| #N/A | MF.Thio5.II-480int.PC | Myeloid | 3.49E-08 | 4.39E-08 | 0.213125037 |
| AD Braak | Mo.6+2-.BL | Myeloid | 3.13E-08 | 1.94E-08 | 0.052778259 |
| ADNC Score | Mo.6+2-.BL | Myeloid | 4.08E-09 | 5.70E-08 | 0.47147452 |
| AP (any/none) | Mo.6+2-.BL | Myeloid | 5.48E-08 | 2.12E-08 | 0.004818998 |
| Arteriolosclerosis | Mo.6+2-.BL | Myeloid | 1.03E-09 | 2.75E-08 | 0.485072478 |
| Arteriolosclerosis (any | Mo.6+2-.BL | Myeloid | -3.68E-08 | 2.59E-08 | 0.92222022 |
| Atherosclerosis | Mo.6+2-.BL | Myeloid | -3.89E-09 | 2.21E-08 | 0.569954701 |
| Atherosclerosis (any/r | Mo.6+2-.BL | Myeloid | -7.87E-10 | 2.07E-08 | 0.515163055 |
| B Score | Mo.6+2-.BL | Myeloid | 1.84E-08 | 1.92E-08 | 0.168661665 |
| CAA | Mo.6+2-.BL | Myeloid | 1.46E-08 | 2.30E-08 | 0.261978448 |
| CAA (any/none) | Mo.6+2-.BL | Myeloid | 1.41E-09 | 2.04E-08 | 0.472415417 |

| | | | | | |
|---|---|---|---|---|---|
| CERAD (C Score) | Mo.6+2-.BL | Myeloid | 2.02E-08 | 1.99E-08 | 0.155280118 |
| Infarcts/lacunes | Mo.6+2-.BL | Myeloid | 1.14E-08 | 2.31E-08 | 0.311133064 |
| PD Braak | Mo.6+2-.BL | Myeloid | 3.68E-08 | 2.17E-08 | 0.045347379 |
| PD Braak (any/none) | Mo.6+2-.BL | Myeloid | 3.20E-08 | 2.06E-08 | 0.060269674 |
| TDP-43 (any/none) | Mo.6+2-.BL | Myeloid | -9.55E-08 | 5.23E-08 | 0.96600408 |
| Thal Phase | Mo.6+2-.BL | Myeloid | -1.48E-07 | 6.96E-08 | 0.983258364 |
| WMR (any/none) | Mo.6+2-.BL | Myeloid | 4.19E-08 | 3.45E-08 | 0.111939995 |
| #N/A | Mo.6+2-.BL | Myeloid | 1.86E-08 | 4.01E-08 | 0.321535633 |
| AD Braak | Mo.6+2+.BL | Myeloid | 2.10E-08 | 1.92E-08 | 0.137909964 |
| ADNC Score | Mo.6+2+.BL | Myeloid | 4.78E-08 | 6.00E-08 | 0.212961186 |
| AP (any/none) | Mo.6+2+.BL | Myeloid | 5.30E-08 | 2.22E-08 | 0.008483278 |
| Arteriolosclerosis | Mo.6+2+.BL | Myeloid | 1.98E-08 | 2.90E-08 | 0.247555939 |
| Arteriolosclerosis (any | Mo.6+2+.BL | Myeloid | -2.39E-09 | 3.01E-08 | 0.53168796 |
| Atherosclerosis | Mo.6+2+.BL | Myeloid | -1.75E-08 | 2.30E-08 | 0.777083917 |
| Atherosclerosis (any/r | Mo.6+2+.BL | Myeloid | -2.26E-08 | 2.19E-08 | 0.848625358 |
| B Score | Mo.6+2+.BL | Myeloid | 4.70E-09 | 1.89E-08 | 0.401894432 |
| CAA | Mo.6+2+.BL | Myeloid | 1.52E-08 | 2.28E-08 | 0.252450859 |
| CAA (any/none) | Mo.6+2+.BL | Myeloid | -2.72E-09 | 2.08E-08 | 0.551918332 |
| CERAD (C Score) | Mo.6+2+.BL | Myeloid | 3.30E-09 | 2.12E-08 | 0.438288915 |
| Infarcts/lacunes | Mo.6+2+.BL | Myeloid | 3.90E-08 | 2.39E-08 | 0.051642276 |
| PD Braak | Mo.6+2+.BL | Myeloid | 3.71E-08 | 2.28E-08 | 0.052012146 |
| PD Braak (any/none) | Mo.6+2+.BL | Myeloid | 3.07E-08 | 2.20E-08 | 0.080886255 |
| TDP-43 (any/none) | Mo.6+2+.BL | Myeloid | -9.32E-08 | 5.60E-08 | 0.9520645 |
| Thal Phase | Mo.6+2+.BL | Myeloid | -4.52E-08 | 7.58E-08 | 0.72459558 |
| WMR (any/none) | Mo.6+2+.BL | Myeloid | 4.05E-08 | 3.46E-08 | 0.120404229 |
| #N/A | Mo.6+2+.BL | Myeloid | 3.33E-08 | 4.52E-08 | 0.230611627 |
| AD Braak | Mo.6+2+.MLN | Myeloid | 2.15E-08 | 1.80E-08 | 0.116018532 |
| ADNC Score | Mo.6+2+.MLN | Myeloid | 1.86E-08 | 5.45E-08 | 0.366482653 |
| AP (any/none) | Mo.6+2+.MLN | Myeloid | 4.53E-08 | 2.18E-08 | 0.018695199 |
| Arteriolosclerosis | Mo.6+2+.MLN | Myeloid | 4.18E-08 | 2.62E-08 | 0.055303885 |
| Arteriolosclerosis (any | Mo.6+2+.MLN | Myeloid | -9.23E-09 | 2.38E-08 | 0.65107148 |
| Atherosclerosis | Mo.6+2+.MLN | Myeloid | -1.03E-08 | 2.16E-08 | 0.683513956 |
| Atherosclerosis (any/r | Mo.6+2+.MLN | Myeloid | 5.10E-09 | 2.08E-08 | 0.40300871 |
| B Score | Mo.6+2+.MLN | Myeloid | 9.20E-09 | 1.83E-08 | 0.307114281 |

| | | | | | |
|---|---|---|---|---|---|
| CAA | Mo.6+2+.MLN | Myeloid | 4.75E-08 | 2.10E-08 | 0.011777929 |
| CAA (any/none) | Mo.6+2+.MLN | Myeloid | 2.16E-08 | 2.03E-08 | 0.14304894 |
| CERAD (C Score) | Mo.6+2+.MLN | Myeloid | 3.99E-09 | 1.93E-08 | 0.418204996 |
| Infarcts/lacunes | Mo.6+2+.MLN | Myeloid | 2.63E-08 | 2.45E-08 | 0.141390015 |
| PD Braak | Mo.6+2+.MLN | Myeloid | 5.33E-08 | 2.43E-08 | 0.014084625 |
| PD Braak (any/none) | Mo.6+2+.MLN | Myeloid | 4.86E-08 | 2.24E-08 | 0.014858624 |
| TDP-43 (any/none) | Mo.6+2+.MLN | Myeloid | 5.96E-09 | 5.36E-08 | 0.455750127 |
| Thal Phase | Mo.6+2+.MLN | Myeloid | -4.84E-08 | 6.79E-08 | 0.761981693 |
| WMR (any/none) | Mo.6+2+.MLN | Myeloid | 3.39E-08 | 3.36E-08 | 0.156450084 |
| #N/A | Mo.6+2+.MLN | Myeloid | -1.34E-08 | 4.02E-08 | 0.630988247 |
| AD Braak | Mo.6+2+.SLN | Myeloid | 2.56E-08 | 1.86E-08 | 0.084342887 |
| ADNC Score | Mo.6+2+.SLN | Myeloid | 5.60E-08 | 5.88E-08 | 0.170295075 |
| AP (any/none) | Mo.6+2+.SLN | Myeloid | 4.33E-08 | 2.25E-08 | 0.026970553 |
| Arteriolosclerosis | Mo.6+2+.SLN | Myeloid | -8.87E-09 | 2.61E-08 | 0.633180174 |
| Arteriolosclerosis (any | Mo.6+2+.SLN | Myeloid | -3.90E-08 | 2.50E-08 | 0.940823653 |
| Atherosclerosis | Mo.6+2+.SLN | Myeloid | -1.91E-08 | 2.14E-08 | 0.81423136 |
| Atherosclerosis (any/r | Mo.6+2+.SLN | Myeloid | -9.39E-09 | 2.26E-08 | 0.661150141 |
| B Score | Mo.6+2+.SLN | Myeloid | 2.63E-08 | 1.90E-08 | 0.082688731 |
| CAA | Mo.6+2+.SLN | Myeloid | -4.25E-09 | 2.23E-08 | 0.575515064 |
| CAA (any/none) | Mo.6+2+.SLN | Myeloid | -1.18E-08 | 1.97E-08 | 0.725539112 |
| CERAD (C Score) | Mo.6+2+.SLN | Myeloid | 4.15E-08 | 2.15E-08 | 0.026645708 |
| Infarcts/lacunes | Mo.6+2+.SLN | Myeloid | 3.40E-08 | 2.40E-08 | 0.077983621 |
| PD Braak | Mo.6+2+.SLN | Myeloid | 3.72E-08 | 2.33E-08 | 0.055226761 |
| PD Braak (any/none) | Mo.6+2+.SLN | Myeloid | 2.97E-08 | 2.15E-08 | 0.083751974 |
| TDP-43 (any/none) | Mo.6+2+.SLN | Myeloid | -5.80E-08 | 4.93E-08 | 0.880297457 |
| Thal Phase | Mo.6+2+.SLN | Myeloid | -3.52E-08 | 7.41E-08 | 0.682747468 |
| WMR (any/none) | Mo.6+2+.SLN | Myeloid | 2.81E-10 | 3.35E-08 | 0.496651572 |
| #N/A | Mo.6+2+.SLN | Myeloid | 2.44E-08 | 4.07E-08 | 0.274221877 |
| AD Braak | Mo.6C+II-.Bl | Myeloid | 1.00E-08 | 1.69E-08 | 0.276075707 |
| ADNC Score | Mo.6C+II-.Bl | Myeloid | 8.31E-08 | 6.36E-08 | 0.095614119 |
| AP (any/none) | Mo.6C+II-.Bl | Myeloid | 5.10E-08 | 2.24E-08 | 0.011335022 |
| Arteriolosclerosis | Mo.6C+II-.Bl | Myeloid | 2.19E-08 | 2.96E-08 | 0.229637846 |
| Arteriolosclerosis (any | Mo.6C+II-.Bl | Myeloid | -3.50E-09 | 2.94E-08 | 0.547311242 |
| Atherosclerosis | Mo.6C+II-.Bl | Myeloid | 8.86E-09 | 2.31E-08 | 0.350612217 |

| | | | | | |
|---|---|---|---|---|---|
| Atherosclerosis (any/r | Mo.6C+II-.Bl | Myeloid | 3.77E-09 | 2.10E-08 | 0.428723479 |
| B Score | Mo.6C+II-.Bl | Myeloid | 8.24E-09 | 1.65E-08 | 0.30822612 |
| CAA | Mo.6C+II-.Bl | Myeloid | -3.10E-08 | 2.24E-08 | 0.916272275 |
| CAA (any/none) | Mo.6C+II-.Bl | Myeloid | -2.61E-08 | 2.16E-08 | 0.88587956 |
| CERAD (C Score) | Mo.6C+II-.Bl | Myeloid | 3.96E-08 | 2.11E-08 | 0.030358132 |
| Infarcts/lacunes | Mo.6C+II-.Bl | Myeloid | 1.47E-08 | 2.41E-08 | 0.270930102 |
| PD Braak | Mo.6C+II-.Bl | Myeloid | 5.57E-08 | 2.18E-08 | 0.005284275 |
| PD Braak (any/none) | Mo.6C+II-.Bl | Myeloid | 3.95E-08 | 2.12E-08 | 0.031326372 |
| TDP-43 (any/none) | Mo.6C+II-.Bl | Myeloid | 3.74E-08 | 5.57E-08 | 0.25063589 |
| Thal Phase | Mo.6C+II-.Bl | Myeloid | 2.91E-08 | 7.97E-08 | 0.357400323 |
| WMR (any/none) | Mo.6C+II-.Bl | Myeloid | 2.85E-08 | 3.72E-08 | 0.222249326 |
| #N/A | Mo.6C+II-.Bl | Myeloid | -3.86E-08 | 4.24E-08 | 0.818752523 |
| AD Braak | Mo.6C+II-.BM | Myeloid | 5.66E-08 | 2.12E-08 | 0.003738518 |
| ADNC Score | Mo.6C+II-.BM | Myeloid | -1.19E-09 | 6.88E-08 | 0.506876505 |
| AP (any/none) | Mo.6C+II-.BM | Myeloid | -2.90E-08 | 2.64E-08 | 0.863950163 |
| Arteriolosclerosis | Mo.6C+II-.BM | Myeloid | -3.70E-09 | 2.95E-08 | 0.549898442 |
| Arteriolosclerosis (any | Mo.6C+II-.BM | Myeloid | -1.04E-08 | 3.40E-08 | 0.62008818 |
| Atherosclerosis | Mo.6C+II-.BM | Myeloid | -4.86E-09 | 2.61E-08 | 0.573758884 |
| Atherosclerosis (any/r | Mo.6C+II-.BM | Myeloid | -7.21E-09 | 2.68E-08 | 0.606052818 |
| B Score | Mo.6C+II-.BM | Myeloid | 3.52E-08 | 2.12E-08 | 0.048035573 |
| CAA | Mo.6C+II-.BM | Myeloid | -2.30E-08 | 2.91E-08 | 0.785003961 |
| CAA (any/none) | Mo.6C+II-.BM | Myeloid | -1.57E-08 | 2.50E-08 | 0.735142585 |
| CERAD (C Score) | Mo.6C+II-.BM | Myeloid | -5.94E-10 | 2.33E-08 | 0.510177506 |
| Infarcts/lacunes | Mo.6C+II-.BM | Myeloid | 4.90E-08 | 2.86E-08 | 0.043693622 |
| PD Braak | Mo.6C+II-.BM | Myeloid | 3.73E-09 | 2.57E-08 | 0.44246169 |
| PD Braak (any/none) | Mo.6C+II-.BM | Myeloid | 8.50E-09 | 2.47E-08 | 0.365529278 |
| TDP-43 (any/none) | Mo.6C+II-.BM | Myeloid | -2.27E-08 | 7.21E-08 | 0.623701041 |
| Thal Phase | Mo.6C+II-.BM | Myeloid | -3.32E-08 | 9.17E-08 | 0.64132806 |
| WMR (any/none) | Mo.6C+II-.BM | Myeloid | -5.09E-08 | 4.49E-08 | 0.87149378 |
| #N/A | Mo.6C+II-.BM | Myeloid | -9.01E-08 | 5.26E-08 | 0.956759551 |
| AD Braak | Mo.6C+II-.LN | Myeloid | -2.76E-10 | 1.78E-08 | 0.506164303 |
| ADNC Score | Mo.6C+II-.LN | Myeloid | -4.28E-08 | 6.27E-08 | 0.752707916 |
| AP (any/none) | Mo.6C+II-.LN | Myeloid | 2.05E-08 | 2.72E-08 | 0.225839924 |
| Arteriolosclerosis | Mo.6C+II-.LN | Myeloid | -1.74E-08 | 2.97E-08 | 0.720688563 |

| | | | | | |
|---|---|---|---|---|---|
| Arteriolosclerosis (any | Mo.6C+II-.LN | Myeloid | -5.21E-08 | 2.96E-08 | 0.960687612 |
| Atherosclerosis | Mo.6C+II-.LN | Myeloid | -1.21E-08 | 2.52E-08 | 0.684689134 |
| Atherosclerosis (any/r | Mo.6C+II-.LN | Myeloid | -5.09E-09 | 2.52E-08 | 0.580212172 |
| B Score | Mo.6C+II-.LN | Myeloid | -4.94E-09 | 1.79E-08 | 0.609012012 |
| CAA | Mo.6C+II-.LN | Myeloid | 1.14E-08 | 2.50E-08 | 0.32346584 |
| CAA (any/none) | Mo.6C+II-.LN | Myeloid | -2.59E-09 | 2.50E-08 | 0.541391844 |
| CERAD (C Score) | Mo.6C+II-.LN | Myeloid | 1.98E-08 | 1.97E-08 | 0.157690956 |
| Infarcts/lacunes | Mo.6C+II-.LN | Myeloid | 3.69E-08 | 2.90E-08 | 0.102179698 |
| PD Braak | Mo.6C+II-.LN | Myeloid | 3.94E-08 | 2.33E-08 | 0.045601159 |
| PD Braak (any/none) | Mo.6C+II-.LN | Myeloid | 3.01E-08 | 2.23E-08 | 0.088610883 |
| TDP-43 (any/none) | Mo.6C+II-.LN | Myeloid | -2.85E-08 | 5.91E-08 | 0.685256783 |
| Thal Phase | Mo.6C+II-.LN | Myeloid | -2.95E-08 | 7.71E-08 | 0.648962592 |
| WMR (any/none) | Mo.6C+II-.LN | Myeloid | 2.49E-08 | 4.48E-08 | 0.289549284 |
| #N/A | Mo.6C+II-.LN | Myeloid | 1.08E-08 | 4.79E-08 | 0.410732887 |
| AD Braak | Mo.6C+II+.Bl | Myeloid | 1.13E-08 | 1.73E-08 | 0.25610343 |
| ADNC Score | Mo.6C+II+.Bl | Myeloid | 5.95E-08 | 6.13E-08 | 0.165981296 |
| AP (any/none) | Mo.6C+II+.Bl | Myeloid | 4.83E-08 | 2.40E-08 | 0.022007354 |
| Arteriolosclerosis | Mo.6C+II+.Bl | Myeloid | 4.57E-08 | 3.12E-08 | 0.071681144 |
| Arteriolosclerosis (any | Mo.6C+II+.Bl | Myeloid | 3.18E-08 | 2.98E-08 | 0.143260074 |
| Atherosclerosis | Mo.6C+II+.Bl | Myeloid | 2.47E-08 | 2.43E-08 | 0.155227443 |
| Atherosclerosis (any/r | Mo.6C+II+.Bl | Myeloid | 2.17E-08 | 2.36E-08 | 0.178969056 |
| B Score | Mo.6C+II+.Bl | Myeloid | 1.15E-08 | 1.78E-08 | 0.259120611 |
| CAA | Mo.6C+II+.Bl | Myeloid | 1.12E-08 | 2.33E-08 | 0.315006755 |
| CAA (any/none) | Mo.6C+II+.Bl | Myeloid | -1.82E-09 | 2.24E-08 | 0.532400369 |
| CERAD (C Score) | Mo.6C+II+.Bl | Myeloid | 6.30E-08 | 2.12E-08 | 0.001474363 |
| Infarcts/lacunes | Mo.6C+II+.Bl | Myeloid | 1.66E-08 | 2.42E-08 | 0.245737036 |
| PD Braak | Mo.6C+II+.Bl | Myeloid | 5.13E-08 | 2.35E-08 | 0.014605562 |
| PD Braak (any/none) | Mo.6C+II+.Bl | Myeloid | 4.07E-08 | 2.31E-08 | 0.039245748 |
| TDP-43 (any/none) | Mo.6C+II+.Bl | Myeloid | -2.04E-08 | 5.26E-08 | 0.650963668 |
| Thal Phase | Mo.6C+II+.Bl | Myeloid | -6.07E-09 | 7.69E-08 | 0.531476152 |
| WMR (any/none) | Mo.6C+II+.Bl | Myeloid | 1.67E-08 | 3.88E-08 | 0.332987853 |
| #N/A | Mo.6C+II+.Bl | Myeloid | -2.51E-09 | 4.50E-08 | 0.522255431 |
| AD Braak | Mo.6C-II-.Bl | Myeloid | -3.28E-09 | 1.97E-08 | 0.566185849 |
| ADNC Score | Mo.6C-II-.Bl | Myeloid | -6.62E-08 | 6.15E-08 | 0.859076147 |

| | | | | | |
|---|---|---|---|---|---|
| AP (any/none) | Mo.6C-II-.Bl | Myeloid | 4.73E-08 | 2.49E-08 | 0.0284704 |
| Arteriolosclerosis | Mo.6C-II-.Bl | Myeloid | 3.26E-08 | 2.97E-08 | 0.136038352 |
| Arteriolosclerosis (any | Mo.6C-II-.Bl | Myeloid | 2.30E-08 | 2.83E-08 | 0.208085055 |
| Atherosclerosis | Mo.6C-II-.Bl | Myeloid | 3.32E-08 | 2.45E-08 | 0.08797593 |
| Atherosclerosis (any/r | Mo.6C-II-.Bl | Myeloid | 2.74E-08 | 2.47E-08 | 0.133309872 |
| B Score | Mo.6C-II-.Bl | Myeloid | -1.19E-09 | 1.84E-08 | 0.525876055 |
| CAA | Mo.6C-II-.Bl | Myeloid | -2.19E-09 | 2.41E-08 | 0.536274362 |
| CAA (any/none) | Mo.6C-II-.Bl | Myeloid | -1.76E-08 | 2.15E-08 | 0.793609923 |
| CERAD (C Score) | Mo.6C-II-.Bl | Myeloid | 3.55E-08 | 2.06E-08 | 0.042481278 |
| Infarcts/lacunes | Mo.6C-II-.Bl | Myeloid | 1.87E-08 | 2.69E-08 | 0.243027182 |
| PD Braak | Mo.6C-II-.Bl | Myeloid | 4.01E-08 | 2.56E-08 | 0.058459463 |
| PD Braak (any/none) | Mo.6C-II-.Bl | Myeloid | 2.89E-08 | 2.35E-08 | 0.109370439 |
| TDP-43 (any/none) | Mo.6C-II-.Bl | Myeloid | 6.49E-10 | 5.56E-08 | 0.495341556 |
| Thal Phase | Mo.6C-II-.Bl | Myeloid | -2.56E-07 | 7.70E-08 | 0.999558925 |
| WMR (any/none) | Mo.6C-II-.Bl | Myeloid | -1.58E-08 | 3.93E-08 | 0.656505232 |
| #N/A | Mo.6C-II-.Bl | Myeloid | 3.78E-08 | 4.67E-08 | 0.208948198 |
| AD Braak | Mo.6C-II-.BM | Myeloid | 4.00E-08 | 2.36E-08 | 0.045374996 |
| ADNC Score | Mo.6C-II-.BM | Myeloid | -5.28E-09 | 6.27E-08 | 0.533512944 |
| AP (any/none) | Mo.6C-II-.BM | Myeloid | -1.49E-08 | 2.44E-08 | 0.729678589 |
| Arteriolosclerosis | Mo.6C-II-.BM | Myeloid | 2.05E-08 | 3.31E-08 | 0.267489806 |
| Arteriolosclerosis (any | Mo.6C-II-.BM | Myeloid | 1.22E-08 | 3.34E-08 | 0.35720523 |
| Atherosclerosis | Mo.6C-II-.BM | Myeloid | 8.62E-09 | 2.86E-08 | 0.381414886 |
| Atherosclerosis (any/r | Mo.6C-II-.BM | Myeloid | 2.63E-08 | 2.95E-08 | 0.186575982 |
| B Score | Mo.6C-II-.BM | Myeloid | 1.71E-08 | 2.30E-08 | 0.228339491 |
| CAA | Mo.6C-II-.BM | Myeloid | -7.28E-09 | 2.85E-08 | 0.600739826 |
| CAA (any/none) | Mo.6C-II-.BM | Myeloid | -1.78E-09 | 2.85E-08 | 0.524896038 |
| CERAD (C Score) | Mo.6C-II-.BM | Myeloid | -1.91E-09 | 2.30E-08 | 0.53309006 |
| Infarcts/lacunes | Mo.6C-II-.BM | Myeloid | 1.77E-08 | 2.85E-08 | 0.267059896 |
| PD Braak | Mo.6C-II-.BM | Myeloid | 1.46E-08 | 2.66E-08 | 0.291767989 |
| PD Braak (any/none) | Mo.6C-II-.BM | Myeloid | 1.24E-08 | 2.55E-08 | 0.312898014 |
| TDP-43 (any/none) | Mo.6C-II-.BM | Myeloid | -4.05E-08 | 6.87E-08 | 0.722474472 |
| Thal Phase | Mo.6C-II-.BM | Myeloid | -4.66E-08 | 8.63E-08 | 0.705156839 |
| WMR (any/none) | Mo.6C-II-.BM | Myeloid | -3.35E-08 | 4.47E-08 | 0.773358371 |
| #N/A | Mo.6C-II-.BM | Myeloid | -5.39E-09 | 5.01E-08 | 0.542837106 |

| | | | | | |
|---|---|---|---|---|---|
| AD Braak | Mo.6C-II+.Bl | Myeloid | 6.59E-09 | 1.86E-08 | 0.361295114 |
| ADNC Score | Mo.6C-II+.Bl | Myeloid | -6.28E-08 | 5.64E-08 | 0.867083151 |
| AP (any/none) | Mo.6C-II+.Bl | Myeloid | 2.29E-08 | 2.24E-08 | 0.15336767 |
| Arteriolosclerosis | Mo.6C-II+.Bl | Myeloid | 4.54E-09 | 2.58E-08 | 0.430277428 |
| Arteriolosclerosis (any | Mo.6C-II+.Bl | Myeloid | 1.42E-08 | 2.80E-08 | 0.306261284 |
| Atherosclerosis | Mo.6C-II+.Bl | Myeloid | 1.50E-08 | 2.45E-08 | 0.27015741 |
| Atherosclerosis (any/r | Mo.6C-II+.Bl | Myeloid | 2.03E-09 | 2.34E-08 | 0.465410909 |
| B Score | Mo.6C-II+.Bl | Myeloid | 7.12E-10 | 1.77E-08 | 0.483925392 |
| CAA | Mo.6C-II+.Bl | Myeloid | 1.26E-08 | 2.27E-08 | 0.289240159 |
| CAA (any/none) | Mo.6C-II+.Bl | Myeloid | -2.47E-08 | 2.08E-08 | 0.883366369 |
| CERAD (C Score) | Mo.6C-II+.Bl | Myeloid | 2.07E-08 | 1.93E-08 | 0.142022665 |
| Infarcts/lacunes | Mo.6C-II+.Bl | Myeloid | -6.65E-09 | 2.37E-08 | 0.610624347 |
| PD Braak | Mo.6C-II+.Bl | Myeloid | 2.45E-08 | 2.24E-08 | 0.137596459 |
| PD Braak (any/none) | Mo.6C-II+.Bl | Myeloid | 2.83E-08 | 2.23E-08 | 0.101862789 |
| TDP-43 (any/none) | Mo.6C-II+.Bl | Myeloid | -1.87E-07 | 5.01E-08 | 0.999902491 |
| Thal Phase | Mo.6C-II+.Bl | Myeloid | -1.30E-07 | 6.61E-08 | 0.975153666 |
| WMR (any/none) | Mo.6C-II+.Bl | Myeloid | 6.92E-09 | 3.66E-08 | 0.424955986 |
| #N/A | Mo.6C-II+.Bl | Myeloid | 1.44E-08 | 4.36E-08 | 0.370913112 |
| AD Braak | Mo.6C-IIint.Bl | Myeloid | 1.87E-08 | 2.00E-08 | 0.175185789 |
| ADNC Score | Mo.6C-IIint.Bl | Myeloid | 6.49E-08 | 6.29E-08 | 0.150908609 |
| AP (any/none) | Mo.6C-IIint.Bl | Myeloid | 4.54E-08 | 2.57E-08 | 0.038470404 |
| Arteriolosclerosis | Mo.6C-IIint.Bl | Myeloid | 2.42E-08 | 2.91E-08 | 0.20270935 |
| Arteriolosclerosis (any | Mo.6C-IIint.Bl | Myeloid | 9.67E-09 | 2.84E-08 | 0.3664909 |
| Atherosclerosis | Mo.6C-IIint.Bl | Myeloid | 2.27E-08 | 2.40E-08 | 0.172000855 |
| Atherosclerosis (any/r | Mo.6C-IIint.Bl | Myeloid | 2.17E-08 | 2.42E-08 | 0.185071194 |
| B Score | Mo.6C-IIint.Bl | Myeloid | 1.98E-08 | 2.02E-08 | 0.163905367 |
| CAA | Mo.6C-IIint.Bl | Myeloid | -2.90E-09 | 2.38E-08 | 0.548491803 |
| CAA (any/none) | Mo.6C-IIint.Bl | Myeloid | -1.44E-08 | 2.10E-08 | 0.754236754 |
| CERAD (C Score) | Mo.6C-IIint.Bl | Myeloid | 4.12E-08 | 2.10E-08 | 0.024965267 |
| Infarcts/lacunes | Mo.6C-IIint.Bl | Myeloid | 1.14E-08 | 2.63E-08 | 0.332887697 |
| PD Braak | Mo.6C-IIint.Bl | Myeloid | 5.64E-08 | 2.54E-08 | 0.013090036 |
| PD Braak (any/none) | Mo.6C-IIint.Bl | Myeloid | 4.63E-08 | 2.42E-08 | 0.028132328 |
| TDP-43 (any/none) | Mo.6C-IIint.Bl | Myeloid | 6.96E-08 | 5.61E-08 | 0.107122092 |
| Thal Phase | Mo.6C-IIint.Bl | Myeloid | -5.97E-08 | 7.85E-08 | 0.776415021 |

| | | | | | |
|---|---|---|---|---|---|
| WMR (any/none) | Mo.6C-IIint.Bl | Myeloid | -1.08E-08 | 3.96E-08 | 0.607665455 |
| #N/A | Mo.6C-IIint.Bl | Myeloid | 1.79E-08 | 4.72E-08 | 0.351819058 |
| AD Braak | Mo.Lu | Myeloid | 2.60E-08 | 1.96E-08 | 0.092315073 |
| ADNC Score | Mo.Lu | Myeloid | 1.80E-08 | 6.07E-08 | 0.383464442 |
| AP (any/none) | Mo.Lu | Myeloid | 4.49E-08 | 2.12E-08 | 0.017295147 |
| Arteriolosclerosis | Mo.Lu | Myeloid | 2.49E-08 | 2.95E-08 | 0.19876063 |
| Arteriolosclerosis (any | Mo.Lu | Myeloid | -1.56E-08 | 3.03E-08 | 0.696001697 |
| Atherosclerosis | Mo.Lu | Myeloid | -2.34E-08 | 2.31E-08 | 0.843950254 |
| Atherosclerosis (any/r | Mo.Lu | Myeloid | -2.49E-08 | 2.22E-08 | 0.869507669 |
| B Score | Mo.Lu | Myeloid | 2.02E-08 | 2.09E-08 | 0.166277738 |
| CAA | Mo.Lu | Myeloid | 1.22E-08 | 2.33E-08 | 0.300513371 |
| CAA (any/none) | Mo.Lu | Myeloid | 9.43E-10 | 2.20E-08 | 0.482872325 |
| CERAD (C Score) | Mo.Lu | Myeloid | 2.40E-08 | 2.15E-08 | 0.132237241 |
| Infarcts/lacunes | Mo.Lu | Myeloid | 4.60E-08 | 2.59E-08 | 0.037713675 |
| PD Braak | Mo.Lu | Myeloid | 5.53E-08 | 2.35E-08 | 0.00924618 |
| PD Braak (any/none) | Mo.Lu | Myeloid | 5.32E-08 | 2.20E-08 | 0.007719132 |
| TDP-43 (any/none) | Mo.Lu | Myeloid | -5.73E-08 | 5.32E-08 | 0.859251383 |
| Thal Phase | Mo.Lu | Myeloid | -6.81E-08 | 7.47E-08 | 0.818942329 |
| WMR (any/none) | Mo.Lu | Myeloid | 3.83E-08 | 3.66E-08 | 0.147627035 |
| #N/A | Mo.Lu | Myeloid | -1.28E-08 | 4.31E-08 | 0.617185418 |
| AD Braak | MLP.BM | Stem | -6.83E-09 | 1.91E-08 | 0.640007032 |
| ADNC Score | MLP.BM | Stem | -2.98E-08 | 5.81E-08 | 0.695981701 |
| AP (any/none) | MLP.BM | Stem | -3.57E-08 | 1.99E-08 | 0.963639815 |
| Arteriolosclerosis | MLP.BM | Stem | 1.82E-08 | 2.87E-08 | 0.262413867 |
| Arteriolosclerosis (any | MLP.BM | Stem | 4.71E-08 | 3.20E-08 | 0.070330608 |
| Atherosclerosis | MLP.BM | Stem | -7.47E-09 | 2.36E-08 | 0.624152276 |
| Atherosclerosis (any/r | MLP.BM | Stem | 1.57E-08 | 2.32E-08 | 0.24855032 |
| B Score | MLP.BM | Stem | -1.17E-08 | 1.91E-08 | 0.728969824 |
| CAA | MLP.BM | Stem | 1.18E-08 | 2.37E-08 | 0.308784751 |
| CAA (any/none) | MLP.BM | Stem | 3.25E-08 | 2.30E-08 | 0.079422968 |
| CERAD (C Score) | MLP.BM | Stem | 1.80E-09 | 2.09E-08 | 0.465809484 |
| Infarcts/lacunes | MLP.BM | Stem | -4.95E-09 | 2.39E-08 | 0.581974244 |
| PD Braak | MLP.BM | Stem | -3.91E-09 | 2.44E-08 | 0.56371691 |
| PD Braak (any/none) | MLP.BM | Stem | 1.58E-09 | 2.38E-08 | 0.473421505 |

| | | | | | |
|---|---|---|---|---|---|
| TDP-43 (any/none) | MLP.BM | Stem | -4.85E-08 | 5.51E-08 | 0.810949395 |
| Thal Phase | MLP.BM | Stem | -3.07E-08 | 7.51E-08 | 0.658796186 |
| WMR (any/none) | MLP.BM | Stem | -1.46E-08 | 3.55E-08 | 0.659649839 |
| #N/A | MLP.BM | Stem | -4.46E-08 | 4.39E-08 | 0.845070204 |
| AD Braak | MLP.FL | Stem | -2.05E-09 | 2.21E-08 | 0.53696432 |
| ADNC Score | MLP.FL | Stem | 3.11E-08 | 5.85E-08 | 0.297444918 |
| AP (any/none) | MLP.FL | Stem | -1.95E-08 | 2.18E-08 | 0.813478406 |
| Arteriolosclerosis | MLP.FL | Stem | 2.11E-08 | 3.53E-08 | 0.274541022 |
| Arteriolosclerosis (any | MLP.FL | Stem | 3.42E-08 | 3.35E-08 | 0.154096747 |
| Atherosclerosis | MLP.FL | Stem | -2.45E-09 | 2.59E-08 | 0.537591034 |
| Atherosclerosis (any/r | MLP.FL | Stem | 5.60E-08 | 2.43E-08 | 0.010660032 |
| B Score | MLP.FL | Stem | -1.77E-08 | 2.21E-08 | 0.787921104 |
| CAA | MLP.FL | Stem | 3.26E-08 | 2.74E-08 | 0.116604528 |
| CAA (any/none) | MLP.FL | Stem | 5.31E-09 | 2.69E-08 | 0.421700625 |
| CERAD (C Score) | MLP.FL | Stem | -1.52E-09 | 2.07E-08 | 0.529344325 |
| Infarcts/lacunes | MLP.FL | Stem | -3.76E-09 | 2.69E-08 | 0.555544969 |
| PD Braak | MLP.FL | Stem | 1.40E-08 | 2.66E-08 | 0.298538287 |
| PD Braak (any/none) | MLP.FL | Stem | 2.62E-08 | 2.62E-08 | 0.158980961 |
| TDP-43 (any/none) | MLP.FL | Stem | -8.66E-08 | 6.43E-08 | 0.910887815 |
| Thal Phase | MLP.FL | Stem | -2.75E-09 | 8.19E-08 | 0.513378896 |
| WMR (any/none) | MLP.FL | Stem | 3.81E-08 | 4.07E-08 | 0.174488598 |
| #N/A | MLP.FL | Stem | -9.96E-08 | 4.64E-08 | 0.984159356 |
| AD Braak | SC.CDP.BM | Stem | 8.68E-09 | 1.90E-08 | 0.324088227 |
| ADNC Score | SC.CDP.BM | Stem | -7.95E-08 | 6.97E-08 | 0.873095795 |
| AP (any/none) | SC.CDP.BM | Stem | -1.89E-08 | 2.31E-08 | 0.793175627 |
| Arteriolosclerosis | SC.CDP.BM | Stem | 1.17E-08 | 3.52E-08 | 0.369292216 |
| Arteriolosclerosis (any | SC.CDP.BM | Stem | 2.42E-08 | 3.58E-08 | 0.249929483 |
| Atherosclerosis | SC.CDP.BM | Stem | 4.24E-09 | 2.73E-08 | 0.438148448 |
| Atherosclerosis (any/r | SC.CDP.BM | Stem | 3.67E-08 | 2.68E-08 | 0.085590158 |
| B Score | SC.CDP.BM | Stem | -8.77E-09 | 1.82E-08 | 0.685008084 |
| CAA | SC.CDP.BM | Stem | -3.32E-08 | 2.64E-08 | 0.896136785 |
| CAA (any/none) | SC.CDP.BM | Stem | 5.90E-10 | 2.62E-08 | 0.491018239 |
| CERAD (C Score) | SC.CDP.BM | Stem | -8.83E-09 | 2.27E-08 | 0.651438454 |
| Infarcts/lacunes | SC.CDP.BM | Stem | -1.98E-08 | 2.32E-08 | 0.802643915 |

| | | | | | |
|---|---|---|---|---|---|
| PD Braak | SC.CDP.BM | Stem | -1.42E-08 | 2.32E-08 | 0.729444728 |
| PD Braak (any/none) | SC.CDP.BM | Stem | -8.63E-09 | 2.25E-08 | 0.649605101 |
| TDP-43 (any/none) | SC.CDP.BM | Stem | -1.66E-08 | 5.96E-08 | 0.60951352 |
| Thal Phase | SC.CDP.BM | Stem | -1.20E-07 | 8.54E-08 | 0.919654349 |
| WMR (any/none) | SC.CDP.BM | Stem | 1.19E-08 | 4.01E-08 | 0.382882351 |
| #N/A | SC.CDP.BM | Stem | -1.03E-07 | 4.76E-08 | 0.984510484 |
| AD Braak | SC.CMP.BM.DR | Stem | 7.67E-09 | 1.86E-08 | 0.339715979 |
| ADNC Score | SC.CMP.BM.DR | Stem | -3.47E-09 | 6.19E-08 | 0.522337954 |
| AP (any/none) | SC.CMP.BM.DR | Stem | -2.87E-08 | 1.97E-08 | 0.928007741 |
| Arteriolosclerosis | SC.CMP.BM.DR | Stem | 2.10E-08 | 3.12E-08 | 0.250902247 |
| Arteriolosclerosis (any | SC.CMP.BM.DR | Stem | 2.29E-08 | 3.07E-08 | 0.227765668 |
| Atherosclerosis | SC.CMP.BM.DR | Stem | 1.04E-08 | 2.47E-08 | 0.337141957 |
| Atherosclerosis (any/r | SC.CMP.BM.DR | Stem | 9.88E-09 | 2.24E-08 | 0.329683376 |
| B Score | SC.CMP.BM.DR | Stem | -8.24E-09 | 1.89E-08 | 0.668141297 |
| CAA | SC.CMP.BM.DR | Stem | -2.39E-08 | 2.50E-08 | 0.830153693 |
| CAA (any/none) | SC.CMP.BM.DR | Stem | -1.13E-08 | 2.47E-08 | 0.67665537 |
| CERAD (C Score) | SC.CMP.BM.DR | Stem | 1.51E-08 | 2.12E-08 | 0.238346381 |
| Infarcts/lacunes | SC.CMP.BM.DR | Stem | -1.05E-09 | 2.43E-08 | 0.517337492 |
| PD Braak | SC.CMP.BM.DR | Stem | 5.90E-09 | 2.50E-08 | 0.406540999 |
| PD Braak (any/none) | SC.CMP.BM.DR | Stem | 1.86E-08 | 2.49E-08 | 0.227248434 |
| TDP-43 (any/none) | SC.CMP.BM.DR | Stem | 4.97E-09 | 5.88E-08 | 0.466358314 |
| Thal Phase | SC.CMP.BM.DR | Stem | -6.17E-08 | 7.81E-08 | 0.785184473 |
| WMR (any/none) | SC.CMP.BM.DR | Stem | 5.72E-08 | 3.74E-08 | 0.062794136 |
| #N/A | SC.CMP.BM.DR | Stem | 3.01E-08 | 4.61E-08 | 0.256543592 |
| AD Braak | SC.GMP.BM | Stem | 2.91E-08 | 2.56E-08 | 0.127704962 |
| ADNC Score | SC.GMP.BM | Stem | -3.17E-08 | 6.58E-08 | 0.685136947 |
| AP (any/none) | SC.GMP.BM | Stem | -4.64E-08 | 2.25E-08 | 0.980294567 |
| Arteriolosclerosis | SC.GMP.BM | Stem | 8.13E-09 | 3.46E-08 | 0.407198178 |
| Arteriolosclerosis (any | SC.GMP.BM | Stem | -7.30E-09 | 3.79E-08 | 0.576395948 |
| Atherosclerosis | SC.GMP.BM | Stem | -3.88E-08 | 2.77E-08 | 0.919268975 |
| Atherosclerosis (any/r | SC.GMP.BM | Stem | -6.52E-09 | 2.87E-08 | 0.58996891 |
| B Score | SC.GMP.BM | Stem | 1.15E-08 | 2.57E-08 | 0.326413263 |
| CAA | SC.GMP.BM | Stem | -2.92E-08 | 3.42E-08 | 0.803477239 |
| CAA (any/none) | SC.GMP.BM | Stem | 8.58E-09 | 3.34E-08 | 0.398528612 |

| | | | | | |
|---|---|---|---|---|---|
| CERAD (C Score) | SC.GMP.BM | Stem | -1.60E-08 | 2.23E-08 | 0.763607191 |
| Infarcts/lacunes | SC.GMP.BM | Stem | -3.47E-08 | 2.83E-08 | 0.889913482 |
| PD Braak | SC.GMP.BM | Stem | 2.49E-08 | 2.75E-08 | 0.182151675 |
| PD Braak (any/none) | SC.GMP.BM | Stem | 2.55E-08 | 2.88E-08 | 0.188451426 |
| TDP-43 (any/none) | SC.GMP.BM | Stem | 4.40E-08 | 7.61E-08 | 0.281522684 |
| Thal Phase | SC.GMP.BM | Stem | -1.39E-07 | 8.52E-08 | 0.948231998 |
| WMR (any/none) | SC.GMP.BM | Stem | -5.59E-08 | 4.38E-08 | 0.899291894 |
| #N/A | SC.GMP.BM | Stem | -1.30E-08 | 5.37E-08 | 0.59542211 |
| AD Braak | SC.LT34F.BM | Stem | -3.36E-08 | 1.58E-08 | 0.983248624 |
| ADNC Score | SC.LT34F.BM | Stem | -6.84E-08 | 5.55E-08 | 0.891035185 |
| AP (any/none) | SC.LT34F.BM | Stem | -1.28E-08 | 1.96E-08 | 0.743936279 |
| Arteriolosclerosis | SC.LT34F.BM | Stem | -3.58E-09 | 2.67E-08 | 0.553371094 |
| Arteriolosclerosis (any | SC.LT34F.BM | Stem | -3.62E-09 | 2.64E-08 | 0.554466063 |
| Atherosclerosis | SC.LT34F.BM | Stem | -3.06E-08 | 2.00E-08 | 0.937098476 |
| Atherosclerosis (any/r | SC.LT34F.BM | Stem | 4.10E-09 | 2.13E-08 | 0.423523517 |
| B Score | SC.LT34F.BM | Stem | -3.09E-08 | 1.68E-08 | 0.967368552 |
| CAA | SC.LT34F.BM | Stem | -1.83E-08 | 2.14E-08 | 0.804013768 |
| CAA (any/none) | SC.LT34F.BM | Stem | -1.31E-08 | 2.15E-08 | 0.728271045 |
| CERAD (C Score) | SC.LT34F.BM | Stem | -1.40E-08 | 1.66E-08 | 0.800531018 |
| Infarcts/lacunes | SC.LT34F.BM | Stem | 1.84E-08 | 2.34E-08 | 0.216648102 |
| PD Braak | SC.LT34F.BM | Stem | -8.63E-09 | 2.12E-08 | 0.657946109 |
| PD Braak (any/none) | SC.LT34F.BM | Stem | 3.60E-09 | 2.11E-08 | 0.432220817 |
| TDP-43 (any/none) | SC.LT34F.BM | Stem | -4.11E-08 | 5.46E-08 | 0.774046671 |
| Thal Phase | SC.LT34F.BM | Stem | -6.96E-08 | 6.98E-08 | 0.840593413 |
| WMR (any/none) | SC.LT34F.BM | Stem | -1.78E-08 | 3.28E-08 | 0.706716203 |
| #N/A | SC.LT34F.BM | Stem | -6.66E-08 | 3.71E-08 | 0.963731427 |
| AD Braak | SC.LTSL.FL | Stem | -2.34E-08 | 1.74E-08 | 0.911407665 |
| ADNC Score | SC.LTSL.FL | Stem | -2.49E-08 | 5.27E-08 | 0.681350646 |
| AP (any/none) | SC.LTSL.FL | Stem | 1.26E-08 | 2.16E-08 | 0.279557231 |
| Arteriolosclerosis | SC.LTSL.FL | Stem | 2.94E-08 | 2.75E-08 | 0.14331841 |
| Arteriolosclerosis (any | SC.LTSL.FL | Stem | 3.93E-08 | 2.86E-08 | 0.084504047 |
| Atherosclerosis | SC.LTSL.FL | Stem | -1.98E-08 | 2.06E-08 | 0.832029055 |
| Atherosclerosis (any/r | SC.LTSL.FL | Stem | 3.68E-08 | 2.23E-08 | 0.049216862 |
| B Score | SC.LTSL.FL | Stem | -2.67E-08 | 1.64E-08 | 0.947983025 |

| | | | | | |
|---|---|---|---|---|---|
| CAA | SC.LTSL.FL | Stem | -2.38E-08 | 2.38E-08 | 0.84072495 |
| CAA (any/none) | SC.LTSL.FL | Stem | -3.97E-08 | 2.25E-08 | 0.961440894 |
| CERAD (C Score) | SC.LTSL.FL | Stem | -4.03E-09 | 1.84E-08 | 0.586720922 |
| Infarcts/lacunes | SC.LTSL.FL | Stem | 5.91E-08 | 2.29E-08 | 0.00484478 |
| PD Braak | SC.LTSL.FL | Stem | -2.73E-08 | 2.39E-08 | 0.873055763 |
| PD Braak (any/none) | SC.LTSL.FL | Stem | -2.40E-08 | 2.35E-08 | 0.845921939 |
| TDP-43 (any/none) | SC.LTSL.FL | Stem | -8.53E-08 | 5.26E-08 | 0.94757598 |
| Thal Phase | SC.LTSL.FL | Stem | 2.38E-09 | 7.08E-08 | 0.486606537 |
| WMR (any/none) | SC.LTSL.FL | Stem | 2.53E-08 | 3.20E-08 | 0.214664858 |
| #N/A | SC.LTSL.FL | Stem | -7.36E-08 | 4.18E-08 | 0.960764377 |
| AD Braak | SC.MDP.BM | Stem | -2.08E-08 | 1.98E-08 | 0.853378051 |
| ADNC Score | SC.MDP.BM | Stem | -9.56E-08 | 7.43E-08 | 0.900935168 |
| AP (any/none) | SC.MDP.BM | Stem | -1.36E-08 | 2.13E-08 | 0.739378614 |
| Arteriolosclerosis | SC.MDP.BM | Stem | 2.37E-08 | 3.69E-08 | 0.260258075 |
| Arteriolosclerosis (any | SC.MDP.BM | Stem | 5.82E-08 | 3.33E-08 | 0.040554244 |
| Atherosclerosis | SC.MDP.BM | Stem | 1.54E-08 | 2.51E-08 | 0.268968646 |
| Atherosclerosis (any/r | SC.MDP.BM | Stem | 3.91E-08 | 2.50E-08 | 0.059203839 |
| B Score | SC.MDP.BM | Stem | -2.92E-08 | 1.91E-08 | 0.936808278 |
| CAA | SC.MDP.BM | Stem | -1.38E-08 | 2.70E-08 | 0.695514131 |
| CAA (any/none) | SC.MDP.BM | Stem | 3.10E-08 | 2.70E-08 | 0.12577225 |
| CERAD (C Score) | SC.MDP.BM | Stem | -2.07E-08 | 2.17E-08 | 0.830895464 |
| Infarcts/lacunes | SC.MDP.BM | Stem | -7.60E-09 | 2.79E-08 | 0.607390788 |
| PD Braak | SC.MDP.BM | Stem | -9.13E-09 | 2.40E-08 | 0.648031042 |
| PD Braak (any/none) | SC.MDP.BM | Stem | -1.30E-08 | 2.28E-08 | 0.716175066 |
| TDP-43 (any/none) | SC.MDP.BM | Stem | -5.95E-09 | 6.10E-08 | 0.538845261 |
| Thal Phase | SC.MDP.BM | Stem | -7.13E-08 | 8.86E-08 | 0.789317386 |
| WMR (any/none) | SC.MDP.BM | Stem | -4.19E-09 | 3.88E-08 | 0.543006142 |
| #N/A | SC.MDP.BM | Stem | -7.28E-08 | 4.77E-08 | 0.936700301 |
| AD Braak | SC.MEP.BM | Stem | -1.96E-08 | 2.09E-08 | 0.825753946 |
| ADNC Score | SC.MEP.BM | Stem | 4.68E-08 | 6.87E-08 | 0.247847034 |
| AP (any/none) | SC.MEP.BM | Stem | 1.01E-08 | 2.54E-08 | 0.345499864 |
| Arteriolosclerosis | SC.MEP.BM | Stem | 2.53E-08 | 3.38E-08 | 0.227169786 |
| Arteriolosclerosis (any | SC.MEP.BM | Stem | 4.36E-08 | 3.52E-08 | 0.107675456 |
| Atherosclerosis | SC.MEP.BM | Stem | 4.45E-10 | 2.61E-08 | 0.4932036 |

| | | | | | |
|---|---|---|---|---|---|
| Atherosclerosis (any/r | SC.MEP.BM | Stem | 2.65E-08 | 2.47E-08 | 0.141514435 |
| B Score | SC.MEP.BM | Stem | -3.14E-08 | 2.04E-08 | 0.938134038 |
| CAA | SC.MEP.BM | Stem | -2.57E-09 | 2.83E-08 | 0.536117694 |
| CAA (any/none) | SC.MEP.BM | Stem | 2.36E-08 | 2.76E-08 | 0.19629265 |
| CERAD (C Score) | SC.MEP.BM | Stem | -7.94E-09 | 2.55E-08 | 0.62220496 |
| Infarcts/lacunes | SC.MEP.BM | Stem | -2.39E-08 | 2.74E-08 | 0.808257116 |
| PD Braak | SC.MEP.BM | Stem | 2.66E-08 | 2.67E-08 | 0.159253793 |
| PD Braak (any/none) | SC.MEP.BM | Stem | 3.35E-08 | 2.70E-08 | 0.107506502 |
| TDP-43 (any/none) | SC.MEP.BM | Stem | -7.37E-08 | 5.90E-08 | 0.894002177 |
| Thal Phase | SC.MEP.BM | Stem | 7.97E-09 | 8.26E-08 | 0.461546084 |
| WMR (any/none) | SC.MEP.BM | Stem | 3.87E-08 | 3.67E-08 | 0.14609992 |
| #N/A | SC.MEP.BM | Stem | -4.58E-08 | 4.70E-08 | 0.83528824 |
| AD Braak | SC.MPP34F.BM | Stem | 6.13E-09 | 1.81E-08 | 0.36766799 |
| ADNC Score | SC.MPP34F.BM | Stem | 1.04E-07 | 6.80E-08 | 0.064106843 |
| AP (any/none) | SC.MPP34F.BM | Stem | -1.77E-08 | 2.02E-08 | 0.809426169 |
| Arteriolosclerosis | SC.MPP34F.BM | Stem | 7.78E-09 | 2.81E-08 | 0.39074702 |
| Arteriolosclerosis (any | SC.MPP34F.BM | Stem | 4.27E-08 | 3.02E-08 | 0.078470442 |
| Atherosclerosis | SC.MPP34F.BM | Stem | -1.29E-08 | 2.24E-08 | 0.716984109 |
| Atherosclerosis (any/r | SC.MPP34F.BM | Stem | 9.56E-09 | 2.12E-08 | 0.326213396 |
| B Score | SC.MPP34F.BM | Stem | 1.95E-09 | 1.77E-08 | 0.456271238 |
| CAA | SC.MPP34F.BM | Stem | 1.24E-08 | 2.36E-08 | 0.299754644 |
| CAA (any/none) | SC.MPP34F.BM | Stem | 1.58E-08 | 2.20E-08 | 0.237072006 |
| CERAD (C Score) | SC.MPP34F.BM | Stem | 1.38E-08 | 2.01E-08 | 0.246610221 |
| Infarcts/lacunes | SC.MPP34F.BM | Stem | -1.65E-08 | 2.34E-08 | 0.7599001 |
| PD Braak | SC.MPP34F.BM | Stem | -1.21E-08 | 2.55E-08 | 0.683044039 |
| PD Braak (any/none) | SC.MPP34F.BM | Stem | -6.55E-09 | 2.50E-08 | 0.60344286 |
| TDP-43 (any/none) | SC.MPP34F.BM | Stem | 3.40E-08 | 5.76E-08 | 0.277897828 |
| Thal Phase | SC.MPP34F.BM | Stem | 6.26E-08 | 7.83E-08 | 0.211842928 |
| WMR (any/none) | SC.MPP34F.BM | Stem | 4.95E-08 | 3.54E-08 | 0.081033114 |
| #N/A | SC.MPP34F.BM | Stem | 2.63E-08 | 4.10E-08 | 0.260438403 |
| AD Braak | SC.ST34F.BM | Stem | 2.49E-09 | 1.76E-08 | 0.443754162 |
| ADNC Score | SC.ST34F.BM | Stem | 3.98E-08 | 6.24E-08 | 0.261785903 |
| AP (any/none) | SC.ST34F.BM | Stem | -2.40E-08 | 2.10E-08 | 0.87278725 |
| Arteriolosclerosis | SC.ST34F.BM | Stem | -2.08E-08 | 2.83E-08 | 0.768914368 |

| | | | | | |
|---|---|---|---|---|---|
| Arteriolosclerosis (any | SC.ST34F.BM | Stem | 4.01E-08 | 2.95E-08 | 0.087355546 |
| Atherosclerosis | SC.ST34F.BM | Stem | -1.51E-08 | 2.32E-08 | 0.742402897 |
| Atherosclerosis (any/r | SC.ST34F.BM | Stem | 4.74E-09 | 2.07E-08 | 0.409341557 |
| B Score | SC.ST34F.BM | Stem | -2.16E-08 | 1.69E-08 | 0.898952154 |
| CAA | SC.ST34F.BM | Stem | -1.77E-08 | 2.32E-08 | 0.776832397 |
| CAA (any/none) | SC.ST34F.BM | Stem | -2.42E-11 | 2.17E-08 | 0.500444437 |
| CERAD (C Score) | SC.ST34F.BM | Stem | 2.13E-08 | 1.77E-08 | 0.114089319 |
| Infarcts/lacunes | SC.ST34F.BM | Stem | -1.22E-08 | 2.35E-08 | 0.698566494 |
| PD Braak | SC.ST34F.BM | Stem | -3.50E-08 | 2.40E-08 | 0.928122203 |
| PD Braak (any/none) | SC.ST34F.BM | Stem | -2.14E-08 | 2.46E-08 | 0.80832171 |
| TDP-43 (any/none) | SC.ST34F.BM | Stem | -5.83E-08 | 5.43E-08 | 0.858586711 |
| Thal Phase | SC.ST34F.BM | Stem | 3.53E-08 | 7.82E-08 | 0.325951023 |
| WMR (any/none) | SC.ST34F.BM | Stem | 1.61E-08 | 3.32E-08 | 0.314183818 |
| #N/A | SC.ST34F.BM | Stem | 1.13E-08 | 3.99E-08 | 0.38809551 |
| AD Braak | SC.STSL.BM | Stem | 7.02E-09 | 1.78E-08 | 0.346804508 |
| ADNC Score | SC.STSL.BM | Stem | -5.56E-08 | 5.65E-08 | 0.837629079 |
| AP (any/none) | SC.STSL.BM | Stem | -2.73E-08 | 1.93E-08 | 0.921409057 |
| Arteriolosclerosis | SC.STSL.BM | Stem | -2.24E-09 | 2.59E-08 | 0.534498286 |
| Arteriolosclerosis (any | SC.STSL.BM | Stem | 2.69E-08 | 2.49E-08 | 0.140334157 |
| Atherosclerosis | SC.STSL.BM | Stem | -8.44E-09 | 2.35E-08 | 0.640297972 |
| Atherosclerosis (any/r | SC.STSL.BM | Stem | -2.56E-08 | 2.31E-08 | 0.865611859 |
| B Score | SC.STSL.BM | Stem | -9.59E-09 | 1.58E-08 | 0.727642675 |
| CAA | SC.STSL.BM | Stem | -1.98E-08 | 2.19E-08 | 0.815963913 |
| CAA (any/none) | SC.STSL.BM | Stem | -2.15E-08 | 2.29E-08 | 0.826361724 |
| CERAD (C Score) | SC.STSL.BM | Stem | -9.14E-09 | 1.79E-08 | 0.695495061 |
| Infarcts/lacunes | SC.STSL.BM | Stem | 1.01E-08 | 2.57E-08 | 0.347491387 |
| PD Braak | SC.STSL.BM | Stem | -1.20E-08 | 2.39E-08 | 0.691980605 |
| PD Braak (any/none) | SC.STSL.BM | Stem | -7.02E-09 | 2.35E-08 | 0.617557134 |
| TDP-43 (any/none) | SC.STSL.BM | Stem | 2.01E-08 | 5.48E-08 | 0.357084232 |
| Thal Phase | SC.STSL.BM | Stem | -9.43E-08 | 7.50E-08 | 0.895824122 |
| WMR (any/none) | SC.STSL.BM | Stem | -4.89E-09 | 3.59E-08 | 0.554171766 |
| #N/A | SC.STSL.BM | Stem | -2.16E-08 | 3.89E-08 | 0.711085387 |
| AD Braak | SC.STSL.FL | Stem | -2.26E-08 | 1.68E-08 | 0.911517241 |
| ADNC Score | SC.STSL.FL | Stem | -4.17E-08 | 5.82E-08 | 0.763230352 |

| | | | | | |
|---|---|---|---|---|---|
| AP (any/none) | SC.STSL.FL | Stem | 1.41E-08 | 2.10E-08 | 0.250919909 |
| Arteriolosclerosis | SC.STSL.FL | Stem | 2.77E-08 | 2.93E-08 | 0.172068279 |
| Arteriolosclerosis (any | SC.STSL.FL | Stem | 4.09E-08 | 2.96E-08 | 0.083492924 |
| Atherosclerosis | SC.STSL.FL | Stem | -1.62E-08 | 2.19E-08 | 0.770182526 |
| Atherosclerosis (any/r | SC.STSL.FL | Stem | 3.51E-08 | 2.40E-08 | 0.071769832 |
| B Score | SC.STSL.FL | Stem | -2.23E-08 | 1.70E-08 | 0.905459462 |
| CAA | SC.STSL.FL | Stem | -4.91E-09 | 2.46E-08 | 0.579029687 |
| CAA (any/none) | SC.STSL.FL | Stem | -3.64E-08 | 2.19E-08 | 0.951579676 |
| CERAD (C Score) | SC.STSL.FL | Stem | -4.08E-09 | 2.00E-08 | 0.580888515 |
| Infarcts/lacunes | SC.STSL.FL | Stem | 1.17E-08 | 2.07E-08 | 0.284997777 |
| PD Braak | SC.STSL.FL | Stem | -2.89E-08 | 2.23E-08 | 0.901922942 |
| PD Braak (any/none) | SC.STSL.FL | Stem | -2.61E-08 | 2.23E-08 | 0.878971983 |
| TDP-43 (any/none) | SC.STSL.FL | Stem | -9.92E-08 | 5.64E-08 | 0.960840708 |
| Thal Phase | SC.STSL.FL | Stem | -4.14E-08 | 7.59E-08 | 0.707318502 |
| WMR (any/none) | SC.STSL.FL | Stem | 4.92E-08 | 3.47E-08 | 0.078061803 |
| #N/A | SC.STSL.FL | Stem | -6.41E-08 | 4.16E-08 | 0.938011024 |
| AD Braak | B.FrF.BM | Stromal | 4.69E-08 | 1.97E-08 | 0.008671294 |
| ADNC Score | B.FrF.BM | Stromal | -1.50E-08 | 5.91E-08 | 0.600167669 |
| AP (any/none) | B.FrF.BM | Stromal | 2.41E-09 | 1.92E-08 | 0.449994664 |
| Arteriolosclerosis | B.FrF.BM | Stromal | 1.67E-08 | 2.78E-08 | 0.273631616 |
| Arteriolosclerosis (any | B.FrF.BM | Stromal | 1.12E-08 | 3.01E-08 | 0.355137579 |
| Atherosclerosis | B.FrF.BM | Stromal | 2.56E-08 | 2.49E-08 | 0.152174093 |
| Atherosclerosis (any/r | B.FrF.BM | Stromal | 1.38E-08 | 2.37E-08 | 0.280136722 |
| B Score | B.FrF.BM | Stromal | 4.22E-08 | 1.87E-08 | 0.011991461 |
| CAA | B.FrF.BM | Stromal | -5.96E-08 | 2.29E-08 | 0.995387106 |
| CAA (any/none) | B.FrF.BM | Stromal | -5.48E-08 | 2.21E-08 | 0.993363121 |
| CERAD (C Score) | B.FrF.BM | Stromal | 1.19E-08 | 2.12E-08 | 0.287208031 |
| Infarcts/lacunes | B.FrF.BM | Stromal | -1.34E-08 | 2.35E-08 | 0.71555822 |
| PD Braak | B.FrF.BM | Stromal | 4.35E-09 | 2.05E-08 | 0.415896563 |
| PD Braak (any/none) | B.FrF.BM | Stromal | -2.57E-09 | 1.89E-08 | 0.553966187 |
| TDP-43 (any/none) | B.FrF.BM | Stromal | -5.63E-08 | 5.48E-08 | 0.848007998 |
| Thal Phase | B.FrF.BM | Stromal | 7.65E-09 | 7.83E-08 | 0.461088849 |
| WMR (any/none) | B.FrF.BM | Stromal | -1.07E-08 | 3.33E-08 | 0.625449123 |
| #N/A | B.FrF.BM | Stromal | 4.71E-08 | 3.87E-08 | 0.111778747 |

| | | | | | |
|---|---|---|---|---|---|
| AD Braak | B.GC.Sp | Stromal | 1.71E-08 | 1.97E-08 | 0.193265867 |
| ADNC Score | B.GC.Sp | Stromal | -8.80E-08 | 6.14E-08 | 0.924092761 |
| AP (any/none) | B.GC.Sp | Stromal | 2.92E-09 | 2.22E-08 | 0.447522121 |
| Arteriolosclerosis | B.GC.Sp | Stromal | -2.22E-08 | 2.78E-08 | 0.787051387 |
| Arteriolosclerosis (any | B.GC.Sp | Stromal | 5.31E-08 | 3.04E-08 | 0.04022738 |
| Atherosclerosis | B.GC.Sp | Stromal | 4.47E-09 | 2.26E-08 | 0.421435874 |
| Atherosclerosis (any/r | B.GC.Sp | Stromal | 2.35E-08 | 2.39E-08 | 0.16226171 |
| B Score | B.GC.Sp | Stromal | 2.06E-08 | 2.16E-08 | 0.16962698 |
| CAA | B.GC.Sp | Stromal | -3.38E-08 | 2.36E-08 | 0.924137195 |
| CAA (any/none) | B.GC.Sp | Stromal | 5.68E-09 | 2.34E-08 | 0.403996041 |
| CERAD (C Score) | B.GC.Sp | Stromal | -3.06E-09 | 2.33E-08 | 0.552305658 |
| Infarcts/lacunes | B.GC.Sp | Stromal | 9.37E-09 | 2.12E-08 | 0.329548367 |
| PD Braak | B.GC.Sp | Stromal | -1.12E-08 | 2.18E-08 | 0.696959609 |
| PD Braak (any/none) | B.GC.Sp | Stromal | -1.75E-08 | 2.19E-08 | 0.788395115 |
| TDP-43 (any/none) | B.GC.Sp | Stromal | -1.08E-07 | 5.37E-08 | 0.978248171 |
| Thal Phase | B.GC.Sp | Stromal | -3.40E-08 | 7.51E-08 | 0.674760843 |
| WMR (any/none) | B.GC.Sp | Stromal | 6.06E-09 | 3.52E-08 | 0.431571231 |
| #N/A | B.GC.Sp | Stromal | -1.28E-07 | 4.18E-08 | 0.998877245 |
| AD Braak | B.Mem.Sp.v2 | Stromal | 3.81E-08 | 1.85E-08 | 0.019474844 |
| ADNC Score | B.Mem.Sp.v2 | Stromal | -6.25E-08 | 5.78E-08 | 0.860367722 |
| AP (any/none) | B.Mem.Sp.v2 | Stromal | -7.20E-09 | 1.96E-08 | 0.643557852 |
| Arteriolosclerosis | B.Mem.Sp.v2 | Stromal | 3.93E-09 | 2.46E-08 | 0.436463851 |
| Arteriolosclerosis (any | B.Mem.Sp.v2 | Stromal | -3.09E-08 | 2.68E-08 | 0.876095012 |
| Atherosclerosis | B.Mem.Sp.v2 | Stromal | -8.38E-09 | 2.07E-08 | 0.65751066 |
| Atherosclerosis (any/r | B.Mem.Sp.v2 | Stromal | -1.77E-08 | 2.24E-08 | 0.784929836 |
| B Score | B.Mem.Sp.v2 | Stromal | 2.53E-08 | 1.99E-08 | 0.101567663 |
| CAA | B.Mem.Sp.v2 | Stromal | -1.25E-08 | 2.32E-08 | 0.704137007 |
| CAA (any/none) | B.Mem.Sp.v2 | Stromal | 1.16E-08 | 2.11E-08 | 0.290922858 |
| CERAD (C Score) | B.Mem.Sp.v2 | Stromal | 2.08E-08 | 1.91E-08 | 0.137292025 |
| Infarcts/lacunes | B.Mem.Sp.v2 | Stromal | -4.45E-09 | 1.91E-08 | 0.5919918 |
| PD Braak | B.Mem.Sp.v2 | Stromal | 1.78E-08 | 1.93E-08 | 0.177463254 |
| PD Braak (any/none) | B.Mem.Sp.v2 | Stromal | 1.24E-08 | 1.89E-08 | 0.255166092 |
| TDP-43 (any/none) | B.Mem.Sp.v2 | Stromal | -4.32E-08 | 5.00E-08 | 0.80622324 |
| Thal Phase | B.Mem.Sp.v2 | Stromal | -8.56E-08 | 7.35E-08 | 0.877967379 |

| | | | | | |
|---|---|---|---|---|---|
| WMR (any/none) | B.Mem.Sp.v2 | Stromal | -1.94E-08 | 3.13E-08 | 0.732573577 |
| #N/A | B.Mem.Sp.v2 | Stromal | 3.91E-08 | 3.94E-08 | 0.160201668 |
| AD Braak | LEC.MLN | Stromal | -2.58E-08 | 1.59E-08 | 0.947683319 |
| ADNC Score | LEC.MLN | Stromal | 1.00E-07 | 5.57E-08 | 0.035827501 |
| AP (any/none) | LEC.MLN | Stromal | -2.35E-08 | 1.99E-08 | 0.881297371 |
| Arteriolosclerosis | LEC.MLN | Stromal | 1.63E-08 | 2.42E-08 | 0.250038887 |
| Arteriolosclerosis (any | LEC.MLN | Stromal | -1.62E-08 | 2.49E-08 | 0.74182836 |
| Atherosclerosis | LEC.MLN | Stromal | 9.22E-10 | 2.28E-08 | 0.483839244 |
| Atherosclerosis (any/r | LEC.MLN | Stromal | -2.18E-08 | 2.22E-08 | 0.836844688 |
| B Score | LEC.MLN | Stromal | -1.41E-08 | 1.68E-08 | 0.799219775 |
| CAA | LEC.MLN | Stromal | 5.54E-10 | 2.04E-08 | 0.489168595 |
| CAA (any/none) | LEC.MLN | Stromal | 2.36E-08 | 2.21E-08 | 0.143574475 |
| CERAD (C Score) | LEC.MLN | Stromal | -7.42E-09 | 1.85E-08 | 0.656076406 |
| Infarcts/lacunes | LEC.MLN | Stromal | 8.62E-09 | 2.33E-08 | 0.356009294 |
| PD Braak | LEC.MLN | Stromal | -7.79E-09 | 2.09E-08 | 0.645045165 |
| PD Braak (any/none) | LEC.MLN | Stromal | -1.33E-08 | 2.07E-08 | 0.740076921 |
| TDP-43 (any/none) | LEC.MLN | Stromal | -1.79E-08 | 4.93E-08 | 0.642007644 |
| Thal Phase | LEC.MLN | Stromal | 1.23E-07 | 6.98E-08 | 0.039144452 |
| WMR (any/none) | LEC.MLN | Stromal | -6.94E-08 | 2.88E-08 | 0.99201026 |
| #N/A | LEC.MLN | Stromal | -5.19E-08 | 3.79E-08 | 0.914802182 |
| AD Braak | LEC.SLN | Stromal | -1.58E-08 | 1.66E-08 | 0.829287968 |
| ADNC Score | LEC.SLN | Stromal | 1.44E-07 | 5.77E-08 | 0.006174228 |
| AP (any/none) | LEC.SLN | Stromal | -1.87E-08 | 2.03E-08 | 0.82191325 |
| Arteriolosclerosis | LEC.SLN | Stromal | 7.30E-09 | 2.54E-08 | 0.386813129 |
| Arteriolosclerosis (any | LEC.SLN | Stromal | -1.33E-08 | 2.74E-08 | 0.68627965 |
| Atherosclerosis | LEC.SLN | Stromal | 5.92E-09 | 2.17E-08 | 0.392493309 |
| Atherosclerosis (any/r | LEC.SLN | Stromal | -1.31E-08 | 2.28E-08 | 0.717392986 |
| B Score | LEC.SLN | Stromal | -5.11E-09 | 1.74E-08 | 0.615404473 |
| CAA | LEC.SLN | Stromal | 8.26E-11 | 2.25E-08 | 0.49853863 |
| CAA (any/none) | LEC.SLN | Stromal | 3.10E-08 | 2.34E-08 | 0.092792252 |
| CERAD (C Score) | LEC.SLN | Stromal | -1.38E-08 | 1.82E-08 | 0.776386404 |
| Infarcts/lacunes | LEC.SLN | Stromal | 5.53E-10 | 2.37E-08 | 0.490677768 |
| PD Braak | LEC.SLN | Stromal | 2.97E-09 | 2.02E-08 | 0.441493363 |
| PD Braak (any/none) | LEC.SLN | Stromal | 5.38E-09 | 2.03E-08 | 0.395223433 |

| | | | | | |
|---|---|---|---|---|---|
| TDP-43 (any/none) | LEC.SLN | Stromal | 5.01E-08 | 6.59E-08 | 0.223524241 |
| Thal Phase | LEC.SLN | Stromal | 1.77E-07 | 7.23E-08 | 0.0073479 |
| WMR (any/none) | LEC.SLN | Stromal | -7.69E-08 | 3.02E-08 | 0.994524857 |
| #N/A | LEC.SLN | Stromal | -5.30E-08 | 3.63E-08 | 0.928189173 |
| AD Braak | LEC.SLN.CFA.d6.v2 | Stromal | -2.48E-08 | 1.48E-08 | 0.952694974 |
| ADNC Score | LEC.SLN.CFA.d6.v2 | Stromal | 1.10E-07 | 5.23E-08 | 0.017529006 |
| AP (any/none) | LEC.SLN.CFA.d6.v2 | Stromal | -3.54E-08 | 2.05E-08 | 0.958405861 |
| Arteriolosclerosis | LEC.SLN.CFA.d6.v2 | Stromal | -1.13E-08 | 2.49E-08 | 0.675142749 |
| Arteriolosclerosis (any | LEC.SLN.CFA.d6.v2 | Stromal | -2.09E-08 | 2.79E-08 | 0.772667138 |
| Atherosclerosis | LEC.SLN.CFA.d6.v2 | Stromal | -5.61E-09 | 2.04E-08 | 0.608232753 |
| Atherosclerosis (any/r | LEC.SLN.CFA.d6.v2 | Stromal | -1.29E-08 | 2.18E-08 | 0.722574411 |
| B Score | LEC.SLN.CFA.d6.v2 | Stromal | -2.79E-08 | 1.66E-08 | 0.953379814 |
| CAA | LEC.SLN.CFA.d6.v2 | Stromal | 7.94E-09 | 2.05E-08 | 0.349168055 |
| CAA (any/none) | LEC.SLN.CFA.d6.v2 | Stromal | 3.05E-08 | 2.23E-08 | 0.085707926 |
| CERAD (C Score) | LEC.SLN.CFA.d6.v2 | Stromal | -5.10E-09 | 1.72E-08 | 0.616499697 |
| Infarcts/lacunes | LEC.SLN.CFA.d6.v2 | Stromal | 1.15E-08 | 2.26E-08 | 0.304955216 |
| PD Braak | LEC.SLN.CFA.d6.v2 | Stromal | 2.33E-08 | 1.93E-08 | 0.113873062 |
| PD Braak (any/none) | LEC.SLN.CFA.d6.v2 | Stromal | 1.83E-08 | 1.98E-08 | 0.177679113 |
| TDP-43 (any/none) | LEC.SLN.CFA.d6.v2 | Stromal | 3.89E-08 | 5.06E-08 | 0.221159337 |
| Thal Phase | LEC.SLN.CFA.d6.v2 | Stromal | 1.52E-07 | 6.60E-08 | 0.010552935 |
| WMR (any/none) | LEC.SLN.CFA.d6.v2 | Stromal | -8.45E-08 | 2.96E-08 | 0.997879889 |
| #N/A | LEC.SLN.CFA.d6.v2 | Stromal | -7.88E-08 | 3.57E-08 | 0.986382501 |
| AD Braak | LEC.SLN.OT | Stromal | -3.13E-08 | 1.69E-08 | 0.967875636 |
| ADNC Score | LEC.SLN.OT | Stromal | 1.69E-07 | 5.47E-08 | 0.001037858 |
| AP (any/none) | LEC.SLN.OT | Stromal | -1.40E-08 | 2.07E-08 | 0.750752095 |
| Arteriolosclerosis | LEC.SLN.OT | Stromal | 1.31E-08 | 2.43E-08 | 0.294776042 |
| Arteriolosclerosis (any | LEC.SLN.OT | Stromal | -9.59E-09 | 2.70E-08 | 0.638840091 |
| Atherosclerosis | LEC.SLN.OT | Stromal | 2.13E-09 | 2.25E-08 | 0.462302011 |
| Atherosclerosis (any/r | LEC.SLN.OT | Stromal | -1.05E-08 | 2.30E-08 | 0.676168528 |
| B Score | LEC.SLN.OT | Stromal | -1.63E-08 | 1.78E-08 | 0.819970956 |
| CAA | LEC.SLN.OT | Stromal | 3.21E-09 | 2.28E-08 | 0.444086231 |
| CAA (any/none) | LEC.SLN.OT | Stromal | 2.55E-08 | 2.33E-08 | 0.136740363 |
| CERAD (C Score) | LEC.SLN.OT | Stromal | -9.39E-10 | 1.85E-08 | 0.520213347 |
| Infarcts/lacunes | LEC.SLN.OT | Stromal | -5.54E-10 | 2.47E-08 | 0.508945886 |

| | | | | | |
|---|---|---|---|---|---|
| PD Braak | LEC.SLN.OT | Stromal | -1.12E-08 | 2.13E-08 | 0.699689781 |
| PD Braak (any/none) | LEC.SLN.OT | Stromal | -1.09E-08 | 2.11E-08 | 0.697451379 |
| TDP-43 (any/none) | LEC.SLN.OT | Stromal | -2.26E-09 | 6.39E-08 | 0.514126574 |
| Thal Phase | LEC.SLN.OT | Stromal | 1.63E-07 | 7.45E-08 | 0.01451374 |
| WMR (any/none) | LEC.SLN.OT | Stromal | -6.75E-08 | 2.99E-08 | 0.987992911 |
| #N/A | LEC.SLN.OT | Stromal | -6.17E-08 | 3.84E-08 | 0.945939976 |
| AD Braak | LEC.SLN.v2 | Stromal | -1.04E-08 | 1.59E-08 | 0.743839389 |
| ADNC Score | LEC.SLN.v2 | Stromal | 1.71E-07 | 5.59E-08 | 0.001128834 |
| AP (any/none) | LEC.SLN.v2 | Stromal | -2.99E-08 | 2.04E-08 | 0.928657341 |
| Arteriolosclerosis | LEC.SLN.v2 | Stromal | 6.26E-09 | 2.52E-08 | 0.401997328 |
| Arteriolosclerosis (any | LEC.SLN.v2 | Stromal | -8.60E-10 | 2.91E-08 | 0.511803844 |
| Atherosclerosis | LEC.SLN.v2 | Stromal | 8.07E-09 | 2.22E-08 | 0.357993291 |
| Atherosclerosis (any/r | LEC.SLN.v2 | Stromal | 1.22E-08 | 2.39E-08 | 0.30547207 |
| B Score | LEC.SLN.v2 | Stromal | -1.29E-08 | 1.78E-08 | 0.764732124 |
| CAA | LEC.SLN.v2 | Stromal | 2.40E-08 | 2.20E-08 | 0.137385684 |
| CAA (any/none) | LEC.SLN.v2 | Stromal | 3.67E-08 | 2.23E-08 | 0.050246411 |
| CERAD (C Score) | LEC.SLN.v2 | Stromal | -2.04E-08 | 1.79E-08 | 0.873452351 |
| Infarcts/lacunes | LEC.SLN.v2 | Stromal | -3.22E-09 | 2.27E-08 | 0.556318623 |
| PD Braak | LEC.SLN.v2 | Stromal | 1.89E-08 | 2.08E-08 | 0.18141141 |
| PD Braak (any/none) | LEC.SLN.v2 | Stromal | 7.79E-09 | 2.12E-08 | 0.356519973 |
| TDP-43 (any/none) | LEC.SLN.v2 | Stromal | -3.53E-08 | 4.94E-08 | 0.762241987 |
| Thal Phase | LEC.SLN.v2 | Stromal | 1.95E-07 | 6.98E-08 | 0.002614549 |
| WMR (any/none) | LEC.SLN.v2 | Stromal | -8.93E-08 | 3.08E-08 | 0.99815002 |
| #N/A | LEC.SLN.v2 | Stromal | -4.27E-08 | 3.69E-08 | 0.876047363 |
| AD Braak | MEChi.GFP-.Adult | Stromal | -1.27E-08 | 1.54E-08 | 0.795121928 |
| ADNC Score | MEChi.GFP-.Adult | Stromal | -6.25E-08 | 5.09E-08 | 0.89065642 |
| AP (any/none) | MEChi.GFP-.Adult | Stromal | -2.51E-08 | 1.78E-08 | 0.91996839 |
| Arteriolosclerosis | MEChi.GFP-.Adult | Stromal | -2.61E-08 | 2.34E-08 | 0.867205258 |
| Arteriolosclerosis (any | MEChi.GFP-.Adult | Stromal | -8.52E-09 | 2.46E-08 | 0.635330799 |
| Atherosclerosis | MEChi.GFP-.Adult | Stromal | -9.29E-09 | 2.00E-08 | 0.679105102 |
| Atherosclerosis (any/r | MEChi.GFP-.Adult | Stromal | -1.57E-08 | 2.03E-08 | 0.780450826 |
| B Score | MEChi.GFP-.Adult | Stromal | -2.14E-08 | 1.65E-08 | 0.90260518 |
| CAA | MEChi.GFP-.Adult | Stromal | -4.04E-08 | 2.07E-08 | 0.974218707 |
| CAA (any/none) | MEChi.GFP-.Adult | Stromal | -7.80E-09 | 1.99E-08 | 0.652451002 |

| | | | | | |
|---|---|---|---|---|---|
| CERAD (C Score) | MEChi.GFP-.Adult | Stromal | -8.41E-09 | 1.59E-08 | 0.701120648 |
| Infarcts/lacunes | MEChi.GFP-.Adult | Stromal | -9.49E-09 | 2.17E-08 | 0.669191923 |
| PD Braak | MEChi.GFP-.Adult | Stromal | 1.15E-08 | 1.79E-08 | 0.259873543 |
| PD Braak (any/none) | MEChi.GFP-.Adult | Stromal | 9.21E-09 | 1.79E-08 | 0.303613881 |
| TDP-43 (any/none) | MEChi.GFP-.Adult | Stromal | 1.26E-08 | 5.13E-08 | 0.403234749 |
| Thal Phase | MEChi.GFP-.Adult | Stromal | 5.56E-08 | 6.35E-08 | 0.190843941 |
| WMR (any/none) | MEChi.GFP-.Adult | Stromal | -3.09E-08 | 2.96E-08 | 0.851960518 |
| #N/A | MEChi.GFP-.Adult | Stromal | -1.81E-08 | 3.84E-08 | 0.681281657 |
| AD Braak | MEChi.GFP+.Adult | Stromal | -2.85E-08 | 1.70E-08 | 0.952789378 |
| ADNC Score | MEChi.GFP+.Adult | Stromal | -3.35E-08 | 4.97E-08 | 0.749977303 |
| AP (any/none) | MEChi.GFP+.Adult | Stromal | 1.52E-09 | 1.95E-08 | 0.468917816 |
| Arteriolosclerosis | MEChi.GFP+.Adult | Stromal | -1.48E-08 | 2.28E-08 | 0.741211039 |
| Arteriolosclerosis (any | MEChi.GFP+.Adult | Stromal | 7.27E-09 | 2.61E-08 | 0.390373958 |
| Atherosclerosis | MEChi.GFP+.Adult | Stromal | -1.40E-08 | 1.86E-08 | 0.773785409 |
| Atherosclerosis (any/r | MEChi.GFP+.Adult | Stromal | -2.53E-08 | 2.01E-08 | 0.896457125 |
| B Score | MEChi.GFP+.Adult | Stromal | -3.04E-08 | 1.64E-08 | 0.967943683 |
| CAA | MEChi.GFP+.Adult | Stromal | -9.51E-09 | 2.06E-08 | 0.677735981 |
| CAA (any/none) | MEChi.GFP+.Adult | Stromal | 7.92E-09 | 2.02E-08 | 0.347245664 |
| CERAD (C Score) | MEChi.GFP+.Adult | Stromal | -1.88E-08 | 1.64E-08 | 0.873922967 |
| Infarcts/lacunes | MEChi.GFP+.Adult | Stromal | -2.07E-09 | 2.06E-08 | 0.540074533 |
| PD Braak | MEChi.GFP+.Adult | Stromal | 1.42E-08 | 1.99E-08 | 0.23773363 |
| PD Braak (any/none) | MEChi.GFP+.Adult | Stromal | 9.15E-09 | 1.88E-08 | 0.312922525 |
| TDP-43 (any/none) | MEChi.GFP+.Adult | Stromal | -2.19E-08 | 5.40E-08 | 0.657202456 |
| Thal Phase | MEChi.GFP+.Adult | Stromal | 4.02E-08 | 7.16E-08 | 0.287421905 |
| WMR (any/none) | MEChi.GFP+.Adult | Stromal | -1.33E-08 | 3.01E-08 | 0.671513996 |
| #N/A | MEChi.GFP+.Adult | Stromal | -1.70E-08 | 3.65E-08 | 0.679046778 |
| AD Braak | MEChi.GFP+.Adult.KO | Stromal | -3.36E-08 | 1.53E-08 | 0.985814746 |
| ADNC Score | MEChi.GFP+.Adult.KO | Stromal | 2.16E-08 | 5.42E-08 | 0.345190244 |
| AP (any/none) | MEChi.GFP+.Adult.KO | Stromal | -1.28E-08 | 1.88E-08 | 0.753296999 |
| Arteriolosclerosis | MEChi.GFP+.Adult.KO | Stromal | -1.63E-08 | 2.33E-08 | 0.757557073 |
| Arteriolosclerosis (any | MEChi.GFP+.Adult.KO | Stromal | 1.75E-09 | 2.60E-08 | 0.473241479 |
| Atherosclerosis | MEChi.GFP+.Adult.KO | Stromal | -2.42E-08 | 1.96E-08 | 0.891352637 |
| Atherosclerosis (any/r | MEChi.GFP+.Adult.KO | Stromal | -2.45E-08 | 2.03E-08 | 0.88714835 |
| B Score | MEChi.GFP+.Adult.KO | Stromal | -2.51E-08 | 1.51E-08 | 0.951008255 |

| | | | | | |
|---|---|---|---|---|---|
| CAA | MEChi.GFP+.Adult.KO | Stromal | -1.33E-08 | 2.01E-08 | 0.746317918 |
| CAA (any/none) | MEChi.GFP+.Adult.KO | Stromal | -7.62E-09 | 2.03E-08 | 0.646084661 |
| CERAD (C Score) | MEChi.GFP+.Adult.KO | Stromal | -7.06E-09 | 1.51E-08 | 0.680524189 |
| Infarcts/lacunes | MEChi.GFP+.Adult.KO | Stromal | -1.53E-08 | 2.08E-08 | 0.769460879 |
| PD Braak | MEChi.GFP+.Adult.KO | Stromal | 1.18E-08 | 1.71E-08 | 0.244818581 |
| PD Braak (any/none) | MEChi.GFP+.Adult.KO | Stromal | 6.56E-09 | 1.71E-08 | 0.350465321 |
| TDP-43 (any/none) | MEChi.GFP+.Adult.KO | Stromal | 2.36E-08 | 4.92E-08 | 0.315237913 |
| Thal Phase | MEChi.GFP+.Adult.KO | Stromal | 7.93E-08 | 6.49E-08 | 0.110854144 |
| WMR (any/none) | MEChi.GFP+.Adult.KO | Stromal | -3.68E-08 | 3.00E-08 | 0.890132999 |
| #N/A | MEChi.GFP+.Adult.KO | Stromal | -1.86E-08 | 3.57E-08 | 0.698955157 |
| AD Braak | St.31-38-44-.SLN | Stromal | -2.11E-08 | 1.70E-08 | 0.892527274 |
| ADNC Score | St.31-38-44-.SLN | Stromal | -1.92E-08 | 5.61E-08 | 0.634186612 |
| AP (any/none) | St.31-38-44-.SLN | Stromal | -1.76E-08 | 1.68E-08 | 0.852331964 |
| Arteriolosclerosis | St.31-38-44-.SLN | Stromal | -3.95E-08 | 2.65E-08 | 0.932404317 |
| Arteriolosclerosis (any | St.31-38-44-.SLN | Stromal | -4.74E-08 | 2.78E-08 | 0.956229273 |
| Atherosclerosis | St.31-38-44-.SLN | Stromal | -1.73E-08 | 2.21E-08 | 0.783083916 |
| Atherosclerosis (any/r | St.31-38-44-.SLN | Stromal | -2.65E-08 | 1.95E-08 | 0.91306562 |
| B Score | St.31-38-44-.SLN | Stromal | -2.84E-08 | 1.76E-08 | 0.946481265 |
| CAA | St.31-38-44-.SLN | Stromal | 6.69E-09 | 2.11E-08 | 0.375330162 |
| CAA (any/none) | St.31-38-44-.SLN | Stromal | 7.74E-09 | 2.14E-08 | 0.358685531 |
| CERAD (C Score) | St.31-38-44-.SLN | Stromal | -1.84E-09 | 1.64E-08 | 0.544717801 |
| Infarcts/lacunes | St.31-38-44-.SLN | Stromal | 5.19E-09 | 2.36E-08 | 0.412929222 |
| PD Braak | St.31-38-44-.SLN | Stromal | -1.34E-10 | 1.96E-08 | 0.502724375 |
| PD Braak (any/none) | St.31-38-44-.SLN | Stromal | 1.22E-08 | 1.99E-08 | 0.270794646 |
| TDP-43 (any/none) | St.31-38-44-.SLN | Stromal | -9.98E-09 | 5.03E-08 | 0.578713538 |
| Thal Phase | St.31-38-44-.SLN | Stromal | 4.61E-08 | 7.18E-08 | 0.260518976 |
| WMR (any/none) | St.31-38-44-.SLN | Stromal | -5.21E-08 | 3.13E-08 | 0.952006192 |
| #N/A | St.31-38-44-.SLN | Stromal | -6.53E-08 | 3.70E-08 | 0.961005659 |
| AD Braak | ABD.TR.14w.B6 | Tab | -2.62E-08 | 1.76E-08 | 0.931385469 |
| ADNC Score | ABD.TR.14w.B6 | Tab | -3.19E-08 | 6.05E-08 | 0.701075428 |
| AP (any/none) | ABD.TR.14w.B6 | Tab | 2.75E-08 | 2.27E-08 | 0.11277025 |
| Arteriolosclerosis | ABD.TR.14w.B6 | Tab | 4.43E-09 | 2.79E-08 | 0.436904969 |
| Arteriolosclerosis (any | ABD.TR.14w.B6 | Tab | 2.15E-08 | 2.97E-08 | 0.23386279 |
| Atherosclerosis | ABD.TR.14w.B6 | Tab | -1.68E-08 | 2.01E-08 | 0.798877958 |

| | | | | | |
|---|---|---|---|---|---|
| Atherosclerosis (any/r | ABD.TR.14w.B6 | Tab | 7.76E-09 | 2.43E-08 | 0.374871106 |
| B Score | ABD.TR.14w.B6 | Tab | -1.90E-08 | 1.85E-08 | 0.847939634 |
| CAA | ABD.TR.14w.B6 | Tab | 1.74E-08 | 2.37E-08 | 0.230384463 |
| CAA (any/none) | ABD.TR.14w.B6 | Tab | 1.35E-08 | 2.34E-08 | 0.282017207 |
| CERAD (C Score) | ABD.TR.14w.B6 | Tab | 9.53E-09 | 1.97E-08 | 0.314630409 |
| Infarcts/lacunes | ABD.TR.14w.B6 | Tab | -6.74E-09 | 2.07E-08 | 0.627385016 |
| PD Braak | ABD.TR.14w.B6 | Tab | -3.11E-08 | 2.05E-08 | 0.93566408 |
| PD Braak (any/none) | ABD.TR.14w.B6 | Tab | -2.31E-08 | 2.08E-08 | 0.866413185 |
| TDP-43 (any/none) | ABD.TR.14w.B6 | Tab | -2.59E-08 | 5.56E-08 | 0.679084745 |
| Thal Phase | ABD.TR.14w.B6 | Tab | -2.39E-08 | 7.12E-08 | 0.631525392 |
| WMR (any/none) | ABD.TR.14w.B6 | Tab | -5.77E-08 | 3.26E-08 | 0.961725624 |
| #N/A | ABD.TR.14w.B6 | Tab | 2.11E-08 | 4.55E-08 | 0.321357349 |
| AD Braak | CD4.1h.LN | Tab | -2.18E-08 | 1.80E-08 | 0.887206971 |
| ADNC Score | CD4.1h.LN | Tab | -8.62E-08 | 5.29E-08 | 0.948408397 |
| AP (any/none) | CD4.1h.LN | Tab | -2.09E-08 | 2.27E-08 | 0.8211196 |
| Arteriolosclerosis | CD4.1h.LN | Tab | 1.62E-08 | 2.44E-08 | 0.252914972 |
| Arteriolosclerosis (any | CD4.1h.LN | Tab | 8.55E-09 | 2.66E-08 | 0.374096727 |
| Atherosclerosis | CD4.1h.LN | Tab | -1.35E-08 | 2.21E-08 | 0.728619768 |
| Atherosclerosis (any/r | CD4.1h.LN | Tab | 5.15E-09 | 2.30E-08 | 0.411474505 |
| B Score | CD4.1h.LN | Tab | -1.69E-08 | 1.78E-08 | 0.829320385 |
| CAA | CD4.1h.LN | Tab | 1.97E-09 | 2.16E-08 | 0.463817509 |
| CAA (any/none) | CD4.1h.LN | Tab | -1.99E-08 | 2.18E-08 | 0.819093109 |
| CERAD (C Score) | CD4.1h.LN | Tab | -2.04E-08 | 1.72E-08 | 0.881771061 |
| Infarcts/lacunes | CD4.1h.LN | Tab | -1.14E-08 | 2.18E-08 | 0.699198596 |
| PD Braak | CD4.1h.LN | Tab | 2.68E-08 | 2.20E-08 | 0.111223168 |
| PD Braak (any/none) | CD4.1h.LN | Tab | 2.16E-08 | 2.14E-08 | 0.156101518 |
| TDP-43 (any/none) | CD4.1h.LN | Tab | 5.42E-09 | 5.69E-08 | 0.462066609 |
| Thal Phase | CD4.1h.LN | Tab | -3.42E-08 | 7.12E-08 | 0.684738027 |
| WMR (any/none) | CD4.1h.LN | Tab | 8.52E-09 | 3.31E-08 | 0.398575191 |
| #N/A | CD4.1h.LN | Tab | -5.94E-08 | 4.24E-08 | 0.919174891 |
| AD Braak | CD4.24h.LN | Tab | 5.78E-09 | 2.15E-08 | 0.393803825 |
| ADNC Score | CD4.24h.LN | Tab | -1.89E-09 | 6.95E-08 | 0.510822002 |
| AP (any/none) | CD4.24h.LN | Tab | 3.23E-08 | 2.44E-08 | 0.092604015 |
| Arteriolosclerosis | CD4.24h.LN | Tab | -2.38E-08 | 3.49E-08 | 0.752394108 |

| | | | | | |
|---|---|---|---|---|---|
| Arteriolosclerosis (any | CD4.24h.LN | Tab | -5.60E-08 | 3.69E-08 | 0.935703603 |
| Atherosclerosis | CD4.24h.LN | Tab | -1.86E-08 | 3.04E-08 | 0.72972338 |
| Atherosclerosis (any/r | CD4.24h.LN | Tab | -1.43E-08 | 2.68E-08 | 0.703110614 |
| B Score | CD4.24h.LN | Tab | 1.21E-08 | 2.28E-08 | 0.298900355 |
| CAA | CD4.24h.LN | Tab | 1.86E-08 | 2.72E-08 | 0.246845722 |
| CAA (any/none) | CD4.24h.LN | Tab | 1.16E-08 | 2.87E-08 | 0.342876114 |
| CERAD (C Score) | CD4.24h.LN | Tab | 8.23E-09 | 2.32E-08 | 0.361456762 |
| Infarcts/lacunes | CD4.24h.LN | Tab | -4.61E-08 | 2.64E-08 | 0.959182316 |
| PD Braak | CD4.24h.LN | Tab | 5.05E-08 | 2.69E-08 | 0.030359815 |
| PD Braak (any/none) | CD4.24h.LN | Tab | 4.19E-08 | 2.59E-08 | 0.052776245 |
| TDP-43 (any/none) | CD4.24h.LN | Tab | 3.05E-09 | 6.33E-08 | 0.480770873 |
| Thal Phase | CD4.24h.LN | Tab | -7.03E-08 | 9.33E-08 | 0.77432842 |
| WMR (any/none) | CD4.24h.LN | Tab | 1.01E-08 | 4.48E-08 | 0.410675064 |
| #N/A | CD4.24h.LN | Tab | -3.55E-08 | 5.16E-08 | 0.753930667 |
| AD Braak | CD4.48h.LN | Tab | 1.69E-08 | 2.42E-08 | 0.242107214 |
| ADNC Score | CD4.48h.LN | Tab | -3.98E-08 | 7.03E-08 | 0.71421859 |
| AP (any/none) | CD4.48h.LN | Tab | 1.23E-08 | 2.41E-08 | 0.305472788 |
| Arteriolosclerosis | CD4.48h.LN | Tab | -7.56E-08 | 2.95E-08 | 0.994841186 |
| Arteriolosclerosis (any | CD4.48h.LN | Tab | -4.92E-08 | 3.08E-08 | 0.944996182 |
| Atherosclerosis | CD4.48h.LN | Tab | -1.93E-09 | 2.74E-08 | 0.52804404 |
| Atherosclerosis (any/r | CD4.48h.LN | Tab | 3.92E-08 | 2.60E-08 | 0.065584372 |
| B Score | CD4.48h.LN | Tab | 1.56E-08 | 2.24E-08 | 0.242352678 |
| CAA | CD4.48h.LN | Tab | -1.40E-08 | 2.58E-08 | 0.706640562 |
| CAA (any/none) | CD4.48h.LN | Tab | 1.72E-09 | 2.90E-08 | 0.476402721 |
| CERAD (C Score) | CD4.48h.LN | Tab | 3.28E-08 | 2.42E-08 | 0.087861018 |
| Infarcts/lacunes | CD4.48h.LN | Tab | 1.37E-08 | 2.50E-08 | 0.291356671 |
| PD Braak | CD4.48h.LN | Tab | -1.14E-08 | 2.25E-08 | 0.693079773 |
| PD Braak (any/none) | CD4.48h.LN | Tab | -1.58E-08 | 2.23E-08 | 0.760427217 |
| TDP-43 (any/none) | CD4.48h.LN | Tab | 1.37E-08 | 5.30E-08 | 0.397876035 |
| Thal Phase | CD4.48h.LN | Tab | -9.62E-08 | 8.69E-08 | 0.865918634 |
| WMR (any/none) | CD4.48h.LN | Tab | 9.97E-09 | 4.26E-08 | 0.407384709 |
| #N/A | CD4.48h.LN | Tab | -4.47E-08 | 5.10E-08 | 0.809714288 |
| AD Braak | CD4.5h.LN | Tab | -3.56E-09 | 2.11E-08 | 0.567183986 |
| ADNC Score | CD4.5h.LN | Tab | 6.67E-08 | 6.67E-08 | 0.15859687 |

| | | | | | |
|---|---|---|---|---|---|
| AP (any/none) | CD4.5h.LN | Tab | -8.35E-09 | 2.53E-08 | 0.629262912 |
| Arteriolosclerosis | CD4.5h.LN | Tab | -3.72E-08 | 3.17E-08 | 0.879727424 |
| Arteriolosclerosis (any | CD4.5h.LN | Tab | -3.27E-08 | 3.34E-08 | 0.836099904 |
| Atherosclerosis | CD4.5h.LN | Tab | -5.08E-09 | 2.87E-08 | 0.570305275 |
| Atherosclerosis (any/r | CD4.5h.LN | Tab | -3.40E-08 | 2.84E-08 | 0.885039597 |
| B Score | CD4.5h.LN | Tab | -1.43E-08 | 2.15E-08 | 0.747800935 |
| CAA | CD4.5h.LN | Tab | 2.78E-08 | 2.73E-08 | 0.154091437 |
| CAA (any/none) | CD4.5h.LN | Tab | 1.28E-08 | 2.58E-08 | 0.30975348 |
| CERAD (C Score) | CD4.5h.LN | Tab | 1.46E-09 | 2.18E-08 | 0.473302849 |
| Infarcts/lacunes | CD4.5h.LN | Tab | -1.30E-10 | 2.73E-08 | 0.501898132 |
| PD Braak | CD4.5h.LN | Tab | 2.22E-08 | 2.45E-08 | 0.181860351 |
| PD Braak (any/none) | CD4.5h.LN | Tab | 1.74E-08 | 2.30E-08 | 0.225491977 |
| TDP-43 (any/none) | CD4.5h.LN | Tab | 4.86E-08 | 6.31E-08 | 0.220413862 |
| Thal Phase | CD4.5h.LN | Tab | 1.32E-07 | 9.52E-08 | 0.083639951 |
| WMR (any/none) | CD4.5h.LN | Tab | -5.95E-08 | 4.26E-08 | 0.918849871 |
| #N/A | CD4.5h.LN | Tab | -4.73E-08 | 4.83E-08 | 0.836056987 |
| AD Braak | CD4.96h.LN | Tab | 2.25E-09 | 2.01E-08 | 0.455297747 |
| ADNC Score | CD4.96h.LN | Tab | 2.36E-08 | 6.61E-08 | 0.360416061 |
| AP (any/none) | CD4.96h.LN | Tab | -4.07E-09 | 2.16E-08 | 0.574743424 |
| Arteriolosclerosis | CD4.96h.LN | Tab | -1.27E-08 | 2.59E-08 | 0.688514945 |
| Arteriolosclerosis (any | CD4.96h.LN | Tab | -1.80E-08 | 2.88E-08 | 0.733763605 |
| Atherosclerosis | CD4.96h.LN | Tab | 2.64E-09 | 2.30E-08 | 0.454384757 |
| Atherosclerosis (any/r | CD4.96h.LN | Tab | 9.46E-09 | 2.45E-08 | 0.350046469 |
| B Score | CD4.96h.LN | Tab | 3.09E-10 | 2.11E-08 | 0.494152751 |
| CAA | CD4.96h.LN | Tab | 1.18E-08 | 2.21E-08 | 0.29733539 |
| CAA (any/none) | CD4.96h.LN | Tab | 2.51E-09 | 2.20E-08 | 0.4545047 |
| CERAD (C Score) | CD4.96h.LN | Tab | -2.75E-08 | 2.06E-08 | 0.909774656 |
| Infarcts/lacunes | CD4.96h.LN | Tab | 8.98E-09 | 2.20E-08 | 0.341808783 |
| PD Braak | CD4.96h.LN | Tab | -1.40E-08 | 2.11E-08 | 0.746686049 |
| PD Braak (any/none) | CD4.96h.LN | Tab | -2.20E-09 | 2.20E-08 | 0.539851022 |
| TDP-43 (any/none) | CD4.96h.LN | Tab | 1.20E-08 | 5.63E-08 | 0.415767062 |
| Thal Phase | CD4.96h.LN | Tab | -3.09E-08 | 7.90E-08 | 0.65216297 |
| WMR (any/none) | CD4.96h.LN | Tab | -1.65E-08 | 3.64E-08 | 0.675432156 |
| #N/A | CD4.96h.LN | Tab | 2.57E-08 | 4.49E-08 | 0.28327698 |

| | | | | | |
|---|---|---|---|---|---|
| AD Braak | CD4.CTR.LN | Tab | -7.84E-09 | 1.95E-08 | 0.656198454 |
| ADNC Score | CD4.CTR.LN | Tab | 2.99E-08 | 5.83E-08 | 0.303935942 |
| AP (any/none) | CD4.CTR.LN | Tab | 9.15E-09 | 2.11E-08 | 0.332528399 |
| Arteriolosclerosis | CD4.CTR.LN | Tab | -1.09E-08 | 2.46E-08 | 0.671316505 |
| Arteriolosclerosis (any | CD4.CTR.LN | Tab | -4.61E-08 | 2.69E-08 | 0.956715366 |
| Atherosclerosis | CD4.CTR.LN | Tab | 1.33E-08 | 2.50E-08 | 0.296892475 |
| Atherosclerosis (any/r | CD4.CTR.LN | Tab | 3.27E-08 | 2.29E-08 | 0.076801685 |
| B Score | CD4.CTR.LN | Tab | -9.85E-09 | 1.94E-08 | 0.694081765 |
| CAA | CD4.CTR.LN | Tab | -5.33E-10 | 2.31E-08 | 0.509213529 |
| CAA (any/none) | CD4.CTR.LN | Tab | -6.22E-09 | 2.29E-08 | 0.607020654 |
| CERAD (C Score) | CD4.CTR.LN | Tab | 1.77E-08 | 1.86E-08 | 0.170365137 |
| Infarcts/lacunes | CD4.CTR.LN | Tab | -8.43E-09 | 2.46E-08 | 0.634241304 |
| PD Braak | CD4.CTR.LN | Tab | 2.61E-08 | 2.05E-08 | 0.101483495 |
| PD Braak (any/none) | CD4.CTR.LN | Tab | 2.52E-08 | 2.04E-08 | 0.108229955 |
| TDP-43 (any/none) | CD4.CTR.LN | Tab | 4.13E-08 | 5.50E-08 | 0.226186751 |
| Thal Phase | CD4.CTR.LN | Tab | 5.01E-08 | 7.21E-08 | 0.243794979 |
| WMR (any/none) | CD4.CTR.LN | Tab | -3.35E-08 | 3.38E-08 | 0.838926198 |
| #N/A | CD4.CTR.LN | Tab | -1.09E-09 | 4.25E-08 | 0.510215291 |
| AD Braak | CD4Control | Tab | 1.52E-08 | 2.01E-08 | 0.224473762 |
| ADNC Score | CD4Control | Tab | 1.45E-08 | 5.79E-08 | 0.401312165 |
| AP (any/none) | CD4Control | Tab | 2.55E-09 | 2.12E-08 | 0.452171936 |
| Arteriolosclerosis | CD4Control | Tab | -2.95E-08 | 2.89E-08 | 0.84703811 |
| Arteriolosclerosis (any | CD4Control | Tab | -9.44E-09 | 3.29E-08 | 0.612930288 |
| Atherosclerosis | CD4Control | Tab | -2.62E-09 | 2.25E-08 | 0.546313189 |
| Atherosclerosis (any/r | CD4Control | Tab | -1.64E-08 | 2.61E-08 | 0.734842301 |
| B Score | CD4Control | Tab | 1.35E-08 | 1.91E-08 | 0.239477284 |
| CAA | CD4Control | Tab | 4.73E-09 | 2.38E-08 | 0.42111686 |
| CAA (any/none) | CD4Control | Tab | 4.72E-09 | 2.60E-08 | 0.428046701 |
| CERAD (C Score) | CD4Control | Tab | -1.84E-08 | 1.90E-08 | 0.833047271 |
| Infarcts/lacunes | CD4Control | Tab | -1.47E-08 | 2.17E-08 | 0.750713621 |
| PD Braak | CD4Control | Tab | -4.26E-08 | 1.99E-08 | 0.984112857 |
| PD Braak (any/none) | CD4Control | Tab | -3.79E-08 | 2.01E-08 | 0.970068106 |
| TDP-43 (any/none) | CD4Control | Tab | 7.89E-09 | 5.51E-08 | 0.443089257 |
| Thal Phase | CD4Control | Tab | 4.93E-09 | 7.92E-08 | 0.475182077 |

| | | | | | |
|---|---|---|---|---|---|
| WMR (any/none) | CD4Control | Tab | -3.36E-09 | 3.21E-08 | 0.54175499 |
| #N/A | CD4Control | Tab | -1.56E-08 | 4.01E-08 | 0.651189771 |
| AD Braak | CD8.1h.LN | Tab | -1.87E-08 | 2.08E-08 | 0.816081299 |
| ADNC Score | CD8.1h.LN | Tab | 4.46E-08 | 5.87E-08 | 0.223699225 |
| AP (any/none) | CD8.1h.LN | Tab | 9.06E-10 | 2.40E-08 | 0.484936172 |
| Arteriolosclerosis | CD8.1h.LN | Tab | 2.02E-08 | 2.83E-08 | 0.237220938 |
| Arteriolosclerosis (any | CD8.1h.LN | Tab | -6.25E-10 | 3.10E-08 | 0.508028625 |
| Atherosclerosis | CD8.1h.LN | Tab | -7.68E-09 | 2.70E-08 | 0.611836747 |
| Atherosclerosis (any/r | CD8.1h.LN | Tab | -2.13E-08 | 2.52E-08 | 0.800950439 |
| B Score | CD8.1h.LN | Tab | -2.51E-08 | 2.06E-08 | 0.887832084 |
| CAA | CD8.1h.LN | Tab | -1.97E-09 | 2.42E-08 | 0.53240561 |
| CAA (any/none) | CD8.1h.LN | Tab | -9.93E-10 | 2.43E-08 | 0.516273203 |
| CERAD (C Score) | CD8.1h.LN | Tab | -8.95E-09 | 1.93E-08 | 0.67847721 |
| Infarcts/lacunes | CD8.1h.LN | Tab | -2.93E-09 | 2.56E-08 | 0.54549806 |
| PD Braak | CD8.1h.LN | Tab | 1.82E-09 | 1.91E-08 | 0.461955258 |
| PD Braak (any/none) | CD8.1h.LN | Tab | -6.46E-09 | 2.05E-08 | 0.623968763 |
| TDP-43 (any/none) | CD8.1h.LN | Tab | 1.28E-08 | 6.26E-08 | 0.419039358 |
| Thal Phase | CD8.1h.LN | Tab | 8.88E-09 | 8.43E-08 | 0.458039938 |
| WMR (any/none) | CD8.1h.LN | Tab | -6.35E-08 | 3.68E-08 | 0.958059369 |
| #N/A | CD8.1h.LN | Tab | -7.27E-08 | 4.04E-08 | 0.963931165 |
| AD Braak | CD8.24h.LN | Tab | 8.29E-09 | 2.19E-08 | 0.352394658 |
| ADNC Score | CD8.24h.LN | Tab | 4.63E-08 | 6.49E-08 | 0.237933062 |
| AP (any/none) | CD8.24h.LN | Tab | 1.97E-08 | 2.46E-08 | 0.212265112 |
| Arteriolosclerosis | CD8.24h.LN | Tab | -1.89E-08 | 3.00E-08 | 0.735733831 |
| Arteriolosclerosis (any | CD8.24h.LN | Tab | -4.15E-08 | 3.40E-08 | 0.888479033 |
| Atherosclerosis | CD8.24h.LN | Tab | 8.05E-09 | 2.88E-08 | 0.390042939 |
| Atherosclerosis (any/r | CD8.24h.LN | Tab | -7.57E-09 | 2.64E-08 | 0.612646714 |
| B Score | CD8.24h.LN | Tab | 1.74E-08 | 2.31E-08 | 0.225461601 |
| CAA | CD8.24h.LN | Tab | 1.38E-08 | 2.74E-08 | 0.307062939 |
| CAA (any/none) | CD8.24h.LN | Tab | -2.81E-09 | 2.79E-08 | 0.540100218 |
| CERAD (C Score) | CD8.24h.LN | Tab | 2.53E-08 | 2.24E-08 | 0.128498601 |
| Infarcts/lacunes | CD8.24h.LN | Tab | -3.89E-08 | 2.56E-08 | 0.935593995 |
| PD Braak | CD8.24h.LN | Tab | 4.34E-08 | 2.56E-08 | 0.045072517 |
| PD Braak (any/none) | CD8.24h.LN | Tab | 3.02E-08 | 2.41E-08 | 0.105538933 |

| | | | | | |
|---|---|---|---|---|---|
| TDP-43 (any/none) | CD8.24h.LN | Tab | 5.29E-09 | 5.71E-08 | 0.463042698 |
| Thal Phase | CD8.24h.LN | Tab | -9.48E-08 | 8.36E-08 | 0.871680127 |
| WMR (any/none) | CD8.24h.LN | Tab | 2.27E-08 | 3.89E-08 | 0.279584132 |
| #N/A | CD8.24h.LN | Tab | -2.85E-09 | 5.32E-08 | 0.521331536 |
| AD Braak | CD8.48h.LN | Tab | 2.55E-08 | 2.30E-08 | 0.133682716 |
| ADNC Score | CD8.48h.LN | Tab | -5.83E-08 | 7.04E-08 | 0.796382136 |
| AP (any/none) | CD8.48h.LN | Tab | 4.32E-09 | 2.36E-08 | 0.427252234 |
| Arteriolosclerosis | CD8.48h.LN | Tab | -4.93E-08 | 2.89E-08 | 0.955682789 |
| Arteriolosclerosis (any | CD8.48h.LN | Tab | -2.39E-08 | 3.16E-08 | 0.775496088 |
| Atherosclerosis | CD8.48h.LN | Tab | 3.15E-08 | 2.76E-08 | 0.127301837 |
| Atherosclerosis (any/r | CD8.48h.LN | Tab | 3.03E-08 | 2.57E-08 | 0.119551049 |
| B Score | CD8.48h.LN | Tab | 2.62E-08 | 2.20E-08 | 0.115957972 |
| CAA | CD8.48h.LN | Tab | 1.89E-08 | 2.41E-08 | 0.216852803 |
| CAA (any/none) | CD8.48h.LN | Tab | 5.11E-09 | 2.65E-08 | 0.423715625 |
| CERAD (C Score) | CD8.48h.LN | Tab | 3.50E-08 | 2.14E-08 | 0.051326128 |
| Infarcts/lacunes | CD8.48h.LN | Tab | 3.30E-08 | 2.51E-08 | 0.093877123 |
| PD Braak | CD8.48h.LN | Tab | -1.01E-08 | 2.51E-08 | 0.656434383 |
| PD Braak (any/none) | CD8.48h.LN | Tab | -1.33E-08 | 2.56E-08 | 0.698447385 |
| TDP-43 (any/none) | CD8.48h.LN | Tab | -2.61E-08 | 5.92E-08 | 0.670720155 |
| Thal Phase | CD8.48h.LN | Tab | -9.61E-08 | 8.80E-08 | 0.862576448 |
| WMR (any/none) | CD8.48h.LN | Tab | -2.40E-08 | 3.79E-08 | 0.736656522 |
| #N/A | CD8.48h.LN | Tab | -2.10E-08 | 4.95E-08 | 0.664427005 |
| AD Braak | CD8.5h.LN | Tab | -6.91E-09 | 2.21E-08 | 0.622934346 |
| ADNC Score | CD8.5h.LN | Tab | -2.86E-08 | 6.80E-08 | 0.663138588 |
| AP (any/none) | CD8.5h.LN | Tab | -4.11E-09 | 2.64E-08 | 0.561861764 |
| Arteriolosclerosis | CD8.5h.LN | Tab | -3.71E-08 | 3.31E-08 | 0.86899799 |
| Arteriolosclerosis (any | CD8.5h.LN | Tab | -4.12E-08 | 3.63E-08 | 0.871360581 |
| Atherosclerosis | CD8.5h.LN | Tab | 1.14E-08 | 2.94E-08 | 0.349503961 |
| Atherosclerosis (any/r | CD8.5h.LN | Tab | -5.41E-09 | 2.80E-08 | 0.57661425 |
| B Score | CD8.5h.LN | Tab | -1.32E-08 | 2.35E-08 | 0.713014584 |
| CAA | CD8.5h.LN | Tab | -1.31E-09 | 2.60E-08 | 0.519991223 |
| CAA (any/none) | CD8.5h.LN | Tab | -4.71E-09 | 2.60E-08 | 0.571846424 |
| CERAD (C Score) | CD8.5h.LN | Tab | 2.19E-08 | 2.18E-08 | 0.157459278 |
| Infarcts/lacunes | CD8.5h.LN | Tab | 2.73E-09 | 2.79E-08 | 0.461111082 |

**Table S15: MAGMA pathway analysis results**

| VARIABLE | PHENOTYPE | NGENES | BETA | BETA_STD | SE | P | FULL_NAME |
|---|---|---|---|---|---|---|---|
| GOBP_REGULATION_OF_VERY_LOW_...1 | LEWY_full | 4 | 3.4085 | 0.049082 | 0.68947 | 3.88E-07 | GOBP_REGULATION_OF_VERY_LOW_DENSITY_LIPOPROTEIN_PARTICLE_CLEARANCE |
| REACTOME_ACTIVATION_OF_RAS_I... | AMY_THAL | 5 | 1.7976 | 0.028932 | 0.37028 | 6.10E-07 | REACTOME_ACTIVATION_OF_RAS_IN_B_CELLS |
| GOBP_REGULATION_OF_VERY_LOW_...1 | LEWY_ANY | 4 | 2.8378 | 0.040865 | 0.59881 | 1.08E-06 | GOBP_REGULATION_OF_VERY_LOW_DENSITY_LIPOPROTEIN_PARTICLE_CLEARANCE |
| GOBP_RESPONSE_TO_FORSKOLIN | HS_S | 13 | 0.99675 | 0.025865 | 0.21855 | 2.57E-06 | GOBP_RESPONSE_TO_FORSKOLIN |
| GOBP_NEGATIVE_REGULATION_OF_...401 | AMY_THAL | 4 | 1.6462 | 0.023698 | 0.36754 | 3.78E-06 | GOBP_NEGATIVE_REGULATION_OF_METALLOENDOPEPTIDASE_ACTIVITY |
| BIOCARTA_PLC_PATHWAY | B_SCORE | 9 | 1.3115 | 0.028317 | 0.29531 | 4.51E-06 | BIOCARTA_PLC_PATHWAY |
| GOBP_REGULATION_OF_VERY_LOW_...1 | LEWY_grp | 4 | 3.072 | 0.044236 | 0.69724 | 5.31E-06 | GOBP_REGULATION_OF_VERY_LOW_DENSITY_LIPOPROTEIN_PARTICLE_CLEARANCE |
| GOBP_REGULATION_OF_NATURAL_K...4 | TDP_3 | 10 | 1.0397 | 0.02366 | 0.23948 | 7.13E-06 | GOBP_REGULATION_OF_NATURAL_KILLER_CELL_MEDIATED_IMMUNE_RESPONSE_TO_TUMOR_CELL |
| GOBP_POSITIVE_REGULATION_OF_...551 | AMY_THAL | 26 | 0.62416 | 0.022895 | 0.14431 | 7.68E-06 | GOBP_POSITIVE_REGULATION_OF_NATURAL_KILLER_CELL_MEDIATED_CYTOTOXICITY |
| GOBP_POSITIVE_REGULATION_OF_...550 | AMY_THAL | 8 | 1.2652 | 0.025754 | 0.29306 | 7.96E-06 | GOBP_POSITIVE_REGULATION_OF_NATURAL_KILLER_CELL_DIFFERENTIATION |
| REACTOME_HYALURONAN_BIOSYNTH... | CVD_ART | 5 | 1.3637 | 0.021948 | 0.31747 | 8.78E-06 | REACTOME_HYALURONAN_BIOSYNTHESIS_AND_EXPORT |
| GOBP_POSITIVE_REGULATION_OF_...553 | AMY_THAL | 31 | 0.57483 | 0.023021 | 0.13599 | 1.19E-05 | GOBP_POSITIVE_REGULATION_OF_NATURAL_KILLER_CELL_MEDIATED_IMMUNITY |
| GOBP_MYOBLAST_MIGRATION | MICR | 13 | 0.92358 | 0.023964 | 0.21853 | 1.20E-05 | GOBP_MYOBLAST_MIGRATION |
| GOBP_REGULATION_OF_RIBOSOME_... | CVD_ART | 5 | 1.4863 | 0.02392 | 0.35171 | 1.20E-05 | GOBP_REGULATION_OF_RIBOSOME_BIOGENESIS |
| WP_COVID19_AND_ENDOTHELIAL_C... | LEWY_ANY | 5 | 1.4848 | 0.023904 | 0.35142 | 1.20E-05 | WP_COVID19_AND_ENDOTHELIAL_CELL_SENESCENCE |
| GOBP_POSITIVE_REGULATION_OF_...552 | TDP_3 | 7 | 1.0593 | 0.020171 | 0.25158 | 1.28E-05 | GOBP_POSITIVE_REGULATION_OF_NATURAL_KILLER_CELL_MEDIATED_IMMUNE_RESPONSE_TO_TUMOR_CELL |
| GOBP_PARAXIAL_MESODERM_MORPH... | INFA | 10 | 0.99374 | 0.022616 | 0.23604 | 1.29E-05 | GOBP_PARAXIAL_MESODERM_MORPHOGENESIS |
| GOBP_MEMBRANE_FUSION_INVOLVE... | CVD_S | 5 | 1.539 | 0.024771 | 0.3671 | 1.39E-05 | GOBP_MEMBRANE_FUSION_INVOLVED_IN_VIRAL_ENTRY_INTO_HOST_CELL |
| WP_2Q211_COPY_NUMBER_VARIATI... | AMY_THAL | 17 | 0.88668 | 0.026305 | 0.21158 | 1.40E-05 | WP_2Q211_COPY_NUMBER_VARIATION_SYNDROME |
| PID_GMCSF_PATHWAY | BRAA | 35 | 0.57962 | 0.024668 | 0.1392 | 1.57E-05 | PID_GMCSF_PATHWAY |
| WP_MONOAMINE_TRANSPORT | HS_S | 31 | 0.61562 | 0.024657 | 0.14971 | 1.97E-05 | WP_MONOAMINE_TRANSPORT |
| GOBP_NATURAL_KILLER_CELL_MED...1 | TDP_3 | 11 | 0.95572 | 0.022811 | 0.23286 | 2.04E-05 | GOBP_NATURAL_KILLER_CELL_MEDIATED_IMMUNE_RESPONSE_TO_TUMOR_CELL |
| GOBP_REGULATION_OF_HEART_RAT...1 | LEWY_grp | 40 | 0.53812 | 0.024481 | 0.13166 | 2.20E-05 | GOBP_REGULATION_OF_HEART_RATE_BY_CARDIAC_CONDUCTION |
| GOBP_POSITIVE_REGULATION_OF_...401 | LEWY_ANY | 7 | 1.2741 | 0.02427 | 0.31202 | 2.23E-05 | GOBP_POSITIVE_REGULATION_OF_INTERLEUKIN_23_PRODUCTION |
| GOMF_ARACHIDONATE_COA_LIGASE... | TDP_3 | 10 | 1.0188 | 0.023186 | 0.24988 | 2.29E-05 | GOMF_ARACHIDONATE_COA_LIGASE_ACTIVITY |
| BIOCARTA_TRKA_PATHWAY | B_SCORE | 14 | 1.1577 | 0.03117 | 0.28403 | 2.31E-05 | BIOCARTA_TRKA_PATHWAY |
| MAGRANGEAS_MULTIPLE_MYELOMA_...3 | LEWY_full | 23 | 0.81955 | 0.028285 | 0.20292 | 2.70E-05 | MAGRANGEAS_MULTIPLE_MYELOMA_IGLL_VS_IGLK_DN |
| GOBP_PARAXIAL_MESODERM_DEVEL... | INFA | 17 | 0.78777 | 0.023372 | 0.19564 | 2.84E-05 | GOBP_PARAXIAL_MESODERM_DEVELOPMENT |
| GOBP_POSITIVE_REGULATION_OF_...678 | BRAA | 85 | 0.33723 | 0.022337 | 0.084071 | 3.04E-05 | GOBP_POSITIVE_REGULATION_OF_PROTEIN_LOCALIZATION_TO_NUCLEUS |
| GOBP_POSITIVE_REGULATION_OF_...374 | HS_S | 3 | 1.8225 | 0.022725 | 0.45442 | 3.04E-05 | GOBP_POSITIVE_REGULATION_OF_HYDROGEN_PEROXIDE_MEDIATED_PROGRAMMED_CELL_DEATH |
| GOBP_RETROGRADE_VESICLE_MEDI... | TDP43 | 49 | 0.47382 | 0.02385 | 0.1183 | 3.11E-05 | GOBP_RETROGRADE_VESICLE_MEDIATED_TRANSPORT_GOLGI_TO_ENDOPLASMIC_RETICULUM |
| GOBP_NEGATIVE_REGULATION_OF_...583 | TDP43 | 20 | 0.71585 | 0.023037 | 0.17887 | 3.16E-05 | GOBP_NEGATIVE_REGULATION_OF_RESPONSE_TO_OXIDATIVE_STRESS |
| GOBP_REGULATION_OF_HEART_RAT...1 | LEWY_full | 40 | 0.51647 | 0.023496 | 0.12933 | 3.28E-05 | GOBP_REGULATION_OF_HEART_RATE_BY_CARDIAC_CONDUCTION |
| REACTOME_ANDROGEN_BIOSYNTHES... | WBVD | 10 | 0.95042 | 0.021631 | 0.23819 | 3.32E-05 | REACTOME_ANDROGEN_BIOSYNTHESIS |
| GOBP_CEREBELLAR_GRANULAR_LAY...2 | CVD_ART_ANY | 6 | 1.0387 | 0.018315 | 0.2609 | 3.45E-05 | GOBP_CEREBELLAR_GRANULAR_LAYER_FORMATION |
| GOBP_PROTEIN_LOCALIZATION_TO...38 | BRAA | 295 | 0.18464 | 0.022659 | 0.046424 | 3.51E-05 | GOBP_PROTEIN_LOCALIZATION_TO_NUCLEUS |
| GOBP_NEGATIVE_REGULATION_OF_...561 | CVD_CAA_ANY | 56 | 0.38043 | 0.020466 | 0.095799 | 3.60E-05 | GOBP_NEGATIVE_REGULATION_OF_PROTEOLYSIS_INVOLVED_IN_PROTEIN_CATABOLIC_PROCESS |
| GOBP_REGULATION_OF_ENDOCYTOS... | LEWY_grp | 203 | 0.23358 | 0.023838 | 0.059067 | 3.86E-05 | GOBP_REGULATION_OF_ENDOCYTOSIS |
| GOMF_GLYCEROPHOSPHOLIPID_FLI... | WMRCAL | 12 | 0.85315 | 0.021267 | 0.21604 | 3.94E-05 | GOMF_GLYCEROPHOSPHOLIPID_FLIPPASE_ACTIVITY |
| GOBP_PHOSPHATIDYLSERINE_EXPO... | AMY_A | 7 | 1.134 | 0.021593 | 0.2894 | 4.48E-05 | GOBP_PHOSPHATIDYLSERINE_EXPOSURE_ON_APOPTOTIC_CELL_SURFACE |
| GOBP_POSITIVE_REGULATION_OF_...429 | B_SCORE | 34 | 0.49419 | 0.020726 | 0.12661 | 4.77E-05 | GOBP_POSITIVE_REGULATION_OF_LAMELLIPODIUM_ORGANIZATION |
| CHESLER_BRAIN_HIGHEST_GENETI... | CVD_CAA_ANY | 35 | 0.54657 | 0.023259 | 0.14004 | 4.77E-05 | CHESLER_BRAIN_HIGHEST_GENETIC_VARIANCE |
| PID_VEGFR1_PATHWAY | B_SCORE | 26 | 0.57473 | 0.021082 | 0.1474 | 4.85E-05 | PID_VEGFR1_PATHWAY |
| POMEROY_MEDULLOBLASTOMA_DESM...1 | HEMOR | 54 | 0.43122 | 0.022791 | 0.11105 | 5.18E-05 | POMEROY_MEDULLOBLASTOMA_DESMOPLASIC_VS_CLASSIC_DN |
| GOBP_REGULATION_OF_EPINEPHRI... | LEWY_grp | 6 | 1.3359 | 0.02356 | 0.3445 | 5.29E-05 | GOBP_REGULATION_OF_EPINEPHRINE_SECRETION |
| GOBP_NEGATIVE_REGULATION_OF_...320 | B_SCORE | 5 | 1.4752 | 0.023743 | 0.38051 | 5.32E-05 | GOBP_NEGATIVE_REGULATION_OF_INTERLEUKIN_1_MEDIATED_SIGNALING_PATHWAY |
| GOBP_MEMBRANE_RAFT_ASSEMBLY | WMRCAL | 9 | 1.2065 | 0.026049 | 0.31129 | 5.34E-05 | GOBP_MEMBRANE_RAFT_ASSEMBLY |
| REACTOME_NECTIN_NECL_TRANS_H... | HS_S | 7 | 1.272 | 0.024225 | 0.32844 | 5.40E-05 | REACTOME_NECTIN_NECL_TRANS_HETERODIMERIZATION |
| GOCC_CLATHRIN_SCULPTED_MONOA... | TDP_3 | 5 | 1.7301 | 0.027844 | 0.44728 | 5.51E-05 | GOCC_CLATHRIN_SCULPTED_MONOAMINE_TRANSPORT_VESICLE |
| GOBP_SEROTONIN_TRANSPORT | MICR | 19 | 0.72047 | 0.022597 | 0.1868 | 5.77E-05 | GOBP_SEROTONIN_TRANSPORT |
| GOBP_T_HELPER_CELL_LINEAGE_C... | WMR | 18 | 0.68959 | 0.02105 | 0.17951 | 6.15E-05 | GOBP_T_HELPER_CELL_LINEAGE_COMMITMENT |
| GOBP_REGULATION_OF_CARDIAC_M...2 | LEWY_grp | 28 | 0.60726 | 0.023122 | 0.15855 | 6.44E-05 | GOBP_REGULATION_OF_CARDIAC_MUSCLE_CELL_ACTION_POTENTIAL |
| GOBP_CARBOHYDRATE_HOMEOSTASIS | CVD_CAA_ANY | 231 | 0.20309 | 0.022089 | 0.053105 | 6.59E-05 | GOBP_CARBOHYDRATE_HOMEOSTASIS |
| GOBP_T_HELPER_17_CELL_LINEAG... | WMR | 14 | 0.73693 | 0.019841 | 0.19284 | 6.67E-05 | GOBP_T_HELPER_17_CELL_LINEAGE_COMMITMENT |
| GOBP_POSITIVE_REGULATION_OF_...427 | B_SCORE | 26 | 0.55897 | 0.020504 | 0.14636 | 6.73E-05 | GOBP_POSITIVE_REGULATION_OF_LAMELLIPODIUM_ASSEMBLY |
| GOBP_GAMMA_DELTA_T_CELL_DIFF... | BRAA | 13 | 0.89742 | 0.02329 | 0.23503 | 6.75E-05 | GOBP_GAMMA_DELTA_T_CELL_DIFFERENTIATION |
| GOBP_POSITIVE_REGULATION_OF_...840 | CVD_CAA | 101 | 0.30581 | 0.022069 | 0.080321 | 7.06E-05 | GOBP_POSITIVE_REGULATION_OF_TUMOR_NECROSIS_FACTOR_SUPERFAMILY_CYTOKINE_PRODUCTION |
| GOBP_REGULATION_OF_CARDIAC_M...2 | LEWY_ANY | 28 | 0.64038 | 0.024382 | 0.16835 | 7.16E-05 | GOBP_REGULATION_OF_CARDIAC_MUSCLE_CELL_ACTION_POTENTIAL |
| GOMF_S_METHYLTRANSFERASE_ACT... | MICR | 9 | 1.0108 | 0.021824 | 0.2661 | 7.31E-05 | GOMF_S_METHYLTRANSFERASE_ACTIVITY |
| GOBP_REGULATION_OF_METALLOEN... | AMY_THAL | 7 | 1.1255 | 0.021432 | 0.29652 | 7.40E-05 | GOBP_REGULATION_OF_METALLOENDOPEPTIDASE_ACTIVITY |
| PID_MYC_ACTIV_PATHWAY | CVD_S | 77 | 0.35172 | 0.022174 | 0.093147 | 8.01E-05 | PID_MYC_ACTIV_PATHWAY |
| GOBP_INTERLEUKIN_23_PRODUCTI... | LEWY_ANY | 9 | 1.0622 | 0.022941 | 0.28185 | 8.24E-05 | GOBP_INTERLEUKIN_23_PRODUCTION |
| WP_FAMILIAL_HYPERLIPIDEMIA_T...1 | B_SCORE | 17 | 0.81486 | 0.024175 | 0.2163 | 8.29E-05 | WP_FAMILIAL_HYPERLIPIDEMIA_TYPE_1 |
| GOBP_POSITIVE_REGULATION_OF_...621 | CVD_ART | 27 | 0.5835 | 0.02181 | 0.15497 | 8.36E-05 | GOBP_POSITIVE_REGULATION_OF_PEPTIDYL_THREONINE_PHOSPHORYLATION |
| GOBP_POSITIVE_REGULATION_OF_...555 | LEWY_ANY | 14 | 0.68348 | 0.018408 | 0.18219 | 8.83E-05 | GOBP_POSITIVE_REGULATION_OF_NECROTIC_CELL_DEATH |
| REACTOME_DAP12_INTERACTIONS | AMY_CERAD | 38 | 0.49012 | 0.021731 | 0.13146 | 9.68E-05 | REACTOME_DAP12_INTERACTIONS |
| GOMF_CORECEPTOR_ACTIVITY | TDP43 | 45 | 0.40891 | 0.019726 | 0.10977 | 9.80E-05 | GOMF_CORECEPTOR_ACTIVITY |
| GOBP_PROLACTIN_SECRETION | LEWY_grp | 5 | 1.4326 | 0.023064 | 0.38608 | 0.000104 | GOBP_PROLACTIN_SECRETION |
| GOBP_REGULATION_OF_ENDOTHELI...1 | TDP_3 | 19 | 0.71044 | 0.02228 | 0.19152 | 0.000104 | GOBP_REGULATION_OF_ENDOTHELIAL_CELL_CHEMOTAXIS |
| REACTOME_PHASE_2_PLATEAU_PHA... | LEWY_full | 14 | 0.79574 | 0.021432 | 0.2146 | 0.000105 | REACTOME_PHASE_2_PLATEAU_PHASE |
| GOBP_NEGATIVE_REGULATION_OF_...531 | CVD_CAA_ANY | 100 | 0.28226 | 0.020268 | 0.076239 | 0.000107 | GOBP_NEGATIVE_REGULATION_OF_PROTEIN_CATABOLIC_PROCESS |
| GOBP_REGULATION_OF_ENDOCYTOS... | LEWY_ANY | 203 | 0.21629 | 0.022073 | 0.058518 | 0.00011 | GOBP_REGULATION_OF_ENDOCYTOSIS |
| GOBP_CEREBELLAR_GRANULAR_LAY...3 | CVD_ART_ANY | 8 | 0.84631 | 0.01723 | 0.22916 | 0.000111 | GOBP_CEREBELLAR_GRANULAR_LAYER_MORPHOGENESIS |
| GOCC_AP_1_ADAPTOR_COMPLEX | INFA | 10 | 0.94973 | 0.021614 | 0.25774 | 0.000115 | GOCC_AP_1_ADAPTOR_COMPLEX |
| GOBP_PHOSPHATIDYLSERINE_EXPO... | AMY_THAL | 7 | 1.116 | 0.021252 | 0.30297 | 0.000115 | GOBP_PHOSPHATIDYLSERINE_EXPOSURE_ON_APOPTOTIC_CELL_SURFACE |
| GOMF_NUCLEAR_EXPORT_SIGNAL_R... | BRAA | 11 | 0.80076 | 0.019117 | 0.21778 | 0.000119 | GOMF_NUCLEAR_EXPORT_SIGNAL_RECEPTOR_ACTIVITY |
| REACTOME_PHASE_2_PLATEAU_PHA... | LEWY_grp | 14 | 0.81457 | 0.021939 | 0.22205 | 0.000123 | REACTOME_PHASE_2_PLATEAU_PHASE |
| REACTOME_SPERM_MOTILITY_AND_... | INFA | 9 | 1.1826 | 0.025533 | 0.32381 | 0.000131 | REACTOME_SPERM_MOTILITY_AND_TAXES |
| GOBP_SUBSTANTIA_NIGRA_DEVELO... | WMR | 40 | 0.43544 | 0.019803 | 0.11934 | 0.000132 | GOBP_SUBSTANTIA_NIGRA_DEVELOPMENT |
| GOBP_NEURAL_NUCLEUS_DEVELOPM... | WMR | 60 | 0.36574 | 0.020361 | 0.10026 | 0.000133 | GOBP_NEURAL_NUCLEUS_DEVELOPMENT |
| GOBP_NEUROBLAST_DIVISION | INFA | 13 | 0.80459 | 0.020876 | 0.22063 | 0.000133 | GOBP_NEUROBLAST_DIVISION |
| GOBP_POSITIVE_REGULATION_OF_...664 | BRAA | 38 | 0.41985 | 0.018617 | 0.11542 | 0.000138 | GOBP_POSITIVE_REGULATION_OF_PROTEIN_IMPORT_INTO_NUCLEUS |
| WP_STEROID_BIOSYNTHESIS | WBVD | 10 | 0.80217 | 0.018257 | 0.22086 | 0.000141 | WP_STEROID_BIOSYNTHESIS |
| REACTOME_METABOLISM_OF_STERO...2 | WBVD | 34 | 0.59559 | 0.02498 | 0.16401 | 0.000141 | REACTOME_METABOLISM_OF_STEROID_HORMONES |
| GOBP_NEGATIVE_REGULATION_OF_...633 | LEWY_ANY | 13 | 1.0091 | 0.02619 | 0.27806 | 0.000143 | GOBP_NEGATIVE_REGULATION_OF_STEROL_TRANSPORT |
| GOBP_NEGATIVE_REGULATION_OF_...526 | CVD_CAA_ANY | 42 | 0.40483 | 0.018868 | 0.11181 | 0.000147 | GOBP_NEGATIVE_REGULATION_OF_PROTEASOMAL_PROTEIN_CATABOLIC_PROCESS |
| GOBP_NEGATIVE_REGULATION_OF_...623 | TDP_3 | 14 | 0.87145 | 0.023463 | 0.2408 | 0.000148 | GOBP_NEGATIVE_REGULATION_OF_SODIUM_ION_TRANSMEMBRANE_TRANSPORT |
| GOBP_REGULATION_OF_LAMELLIPO...1 | B_SCORE | 35 | 0.47718 | 0.020304 | 0.1319 | 0.000149 | GOBP_REGULATION_OF_LAMELLIPODIUM_ASSEMBLY |
| GOBP_APOPTOTIC_PROCESS_INVOL...2 | AMY_A | 39 | 0.49758 | 0.022346 | 0.13767 | 0.000151 | GOBP_APOPTOTIC_PROCESS_INVOLVED_IN_DEVELOPMENT |
| GOMF_WATER_CHANNEL_ACTIVITY | HS_S | 13 | 0.94123 | 0.024424 | 0.26109 | 0.000157 | GOMF_WATER_CHANNEL_ACTIVITY |
| GOBP_B_CELL_MEDIATED_IMMUNITY | AMY_A | 139 | 0.27143 | 0.022953 | 0.075381 | 0.000159 | GOBP_B_CELL_MEDIATED_IMMUNITY |
| GOCC_ENDOSOME | LEWY_full | 981 | 0.094603 | 0.020787 | 0.026274 | 0.000159 | GOCC_ENDOSOME |
| GOBP_TRANS_SYNAPTIC_SIGNALIN...3 | TDP_3 | 12 | 0.84163 | 0.02098 | 0.23377 | 0.00016 | GOBP_TRANS_SYNAPTIC_SIGNALING_MODULATING_SYNAPTIC_TRANSMISSION |
| GOBP_RESPONSE_TO_EPINEPHRINE | HS_S | 14 | 0.83118 | 0.022382 | 0.23108 | 0.000162 | GOBP_RESPONSE_TO_EPINEPHRINE |

**Table S16: MAGMA tissue-specific associations**

| VARIABLE | PHENOTYPE | NGENES | BETA | BETA_STD | SE | P | FULL_NAME |
|---|---|---|---|---|---|---|---|
| Spleen | B_SCORE | 17388 | 0.029995 | 0.059493 | 0.007031 | 1.00E-05 | Spleen |
| Lung | BRAA | 17385 | 0.032442 | 0.062793 | 0.008753 | 0.000106 | Lung |
| Spleen | BRAA | 17385 | 0.023626 | 0.046862 | 0.007052 | 0.000405 | Spleen |
| Lung | B_SCORE | 17388 | 0.028451 | 0.055071 | 0.008733 | 0.000563 | Lung |
| Whole_Blood | B_SCORE | 17388 | 0.015938 | 0.028808 | 0.005427 | 0.001662 | Whole_Blood |
| Adipose_Subcutaneous | INFA | 17386 | 0.028935 | 0.057801 | 0.00992 | 0.00177 | Adipose_Subcutaneous |
| Breast_Mammary_Tissue | CVD_ART | 17388 | 0.032823 | 0.063188 | 0.012033 | 0.003193 | Breast_Mammary_Tissue |
| Pituitary | LEWY_grp | 17385 | 0.023638 | 0.04412 | 0.008681 | 0.003239 | Pituitary |
| Whole_Blood | WMRCAL | 17388 | 0.014084 | 0.025457 | 0.005365 | 0.004335 | Whole_Blood |
| Thyroid | CVD_ART | 17388 | 0.023585 | 0.046756 | 0.009468 | 0.006377 | Thyroid |
| Whole_Blood | BRAA | 17385 | 0.013414 | 0.024247 | 0.005458 | 0.007002 | Whole_Blood |
| Breast_Mammary_Tissue | INFA | 17386 | 0.027665 | 0.053262 | 0.011541 | 0.00827 | Breast_Mammary_Tissue |
| Lung | AMY_CERAD | 17386 | 0.020671 | 0.040011 | 0.008763 | 0.00917 | Lung |
| Liver | WMRCAL | 17388 | 0.014031 | 0.025278 | 0.005993 | 0.009619 | Liver |
| Pituitary | LEWY_full | 17385 | 0.02022 | 0.037741 | 0.008643 | 0.009659 | Pituitary |
| Thyroid | TDP_3 | 17388 | 0.021672 | 0.042964 | 0.009375 | 0.010405 | Thyroid |
| Cells_Cultured_fibroblasts | TDP43 | 17386 | 0.01496 | 0.032072 | 0.006502 | 0.010708 | Cells_Cultured_fibroblasts |
| Thyroid | B_SCORE | 17388 | 0.021212 | 0.042052 | 0.009262 | 0.011009 | Thyroid |
| Artery_Tibial | TDP43 | 17386 | 0.021782 | 0.044877 | 0.00971 | 0.012451 | Artery_Tibial |
| Nerve_Tibial | TDP43 | 17386 | 0.022105 | 0.044543 | 0.009935 | 0.013053 | Nerve_Tibial |
| Adipose_Visceral_Omentum | ADNC | 17388 | 0.023228 | 0.045157 | 0.010523 | 0.013656 | Adipose_Visceral_Omentum |
| Cervix_Ectocervix | TDP43 | 17386 | 0.025262 | 0.049565 | 0.011726 | 0.015615 | Cervix_Ectocervix |
| Lung | TDP_3 | 17388 | 0.01872 | 0.036234 | 0.008754 | 0.016251 | Lung |
| Colon_Transverse | WMR | 17388 | 0.023018 | 0.042366 | 0.010892 | 0.017296 | Colon_Transverse |
| Nerve_Tibial | TDP_3 | 17388 | 0.020048 | 0.040396 | 0.009677 | 0.019159 | Nerve_Tibial |
| Lung | CVD_ATH_ANY | 17386 | 0.017648 | 0.034159 | 0.008643 | 0.020592 | Lung |
| Whole_Blood | AMY_CERAD | 17386 | 0.011255 | 0.020344 | 0.005516 | 0.020659 | Whole_Blood |
| Breast_Mammary_Tissue | WMR | 17388 | 0.023983 | 0.046171 | 0.011803 | 0.021087 | Breast_Mammary_Tissue |
| Adipose_Subcutaneous | CVD_ART | 17388 | 0.020853 | 0.041656 | 0.010271 | 0.021179 | Adipose_Subcutaneous |
| Cervix_Ectocervix | HEMOR | 17378 | 0.022759 | 0.044653 | 0.011509 | 0.024007 | Cervix_Ectocervix |
| Adrenal_Gland | AMY_A | 17388 | 0.018408 | 0.035566 | 0.009421 | 0.025371 | Adrenal_Gland |
| Bladder | CVD_ATH | 17388 | 0.023688 | 0.046221 | 0.012325 | 0.027322 | Bladder |
| Thyroid | TDP43 | 17386 | 0.018035 | 0.035753 | 0.009443 | 0.028082 | Thyroid |

| | | | | | | | |
|---|---|---|---|---|---|---|---|
| Lung | CVD_ATH | 17388 | 0.016215 | 0.031385 | 0.008514 | 0.028437 | Lung |
| Esophagus_Muscularis | AMY_CERAD | 17386 | 0.022351 | 0.044128 | 0.01174 | 0.028473 | Esophagus_Muscularis |
| Prostate | AMY_CERAD | 17386 | 0.022071 | 0.041925 | 0.011602 | 0.02857 | Prostate |
| Liver | CVD_ART | 17388 | 0.011644 | 0.020978 | 0.006137 | 0.028907 | Liver |
| Uterus | HEMOR | 17378 | 0.019597 | 0.039966 | 0.010339 | 0.02903 | Uterus |
| Brain_Cerebellar_Hemisphere | CVD_ART_ANY | 17387 | 0.010914 | 0.021916 | 0.005803 | 0.030012 | Brain_Cerebellar_Hemisphere |
| Thyroid | BRAA | 17385 | 0.016934 | 0.033571 | 0.009237 | 0.033393 | Thyroid |
| Adipose_Visceral_Omentum | INFA | 17386 | 0.018789 | 0.036529 | 0.010293 | 0.033978 | Adipose_Visceral_Omentum |
| Bladder | BRAA | 17385 | 0.022721 | 0.044334 | 0.012451 | 0.034027 | Bladder |
| Lung | CVD_ART | 17388 | 0.015954 | 0.030881 | 0.008768 | 0.034429 | Lung |
| Adipose_Visceral_Omentum | WMR | 17388 | 0.019047 | 0.037029 | 0.010584 | 0.035976 | Adipose_Visceral_Omentum |
| Cervix_Ectocervix | CVD_ATH | 17388 | 0.02063 | 0.040475 | 0.011472 | 0.036085 | Cervix_Ectocervix |
| Artery_Aorta | TDP43 | 17386 | 0.017908 | 0.036574 | 0.01002 | 0.036969 | Artery_Aorta |
| Adrenal_Gland | INFA | 17386 | 0.016308 | 0.03151 | 0.009135 | 0.037114 | Adrenal_Gland |
| Artery_Tibial | INFA | 17386 | 0.016395 | 0.033778 | 0.00919 | 0.037226 | Artery_Tibial |
| Ovary | ADNC | 17388 | 0.016068 | 0.0326 | 0.009216 | 0.04064 | Ovary |
| Skin_Not_Sun_Exposed_Suprapu... | TDP43 | 17386 | 0.013002 | 0.0251 | 0.007553 | 0.042593 | Skin_Not_Sun_Exposed_Suprapubic |
| Small_Intestine_Terminal_Ile... | BRAA | 17385 | 0.015701 | 0.028908 | 0.009126 | 0.042684 | Small_Intestine_Terminal_Ileum |
| Cells_Cultured_fibroblasts | CVD_ATH_ANY | 17386 | 0.010711 | 0.022962 | 0.006231 | 0.042825 | Cells_Cultured_fibroblasts |
| Cervix_Endocervix | HEMOR | 17378 | 0.018911 | 0.037478 | 0.011014 | 0.043 | Cervix_Endocervix |
| Skin_Sun_Exposed_Lower_leg | TDP43 | 17386 | 0.012953 | 0.025161 | 0.007559 | 0.04332 | Skin_Sun_Exposed_Lower_leg |
| Skin_Sun_Exposed_Lower_leg | HS_S | 17386 | 0.012556 | 0.02439 | 0.007363 | 0.044078 | Skin_Sun_Exposed_Lower_leg |
| Nerve_Tibial | MICR | 17385 | 0.01568 | 0.031594 | 0.009362 | 0.046991 | Nerve_Tibial |
| Brain_Cerebellum | CVD_ART_ANY | 17387 | 0.010023 | 0.019912 | 0.006002 | 0.04748 | Brain_Cerebellum |
| Testis | CVD_S | 17387 | 0.008957 | 0.015483 | 0.005377 | 0.047893 | Testis |
| Spleen | CVD_ATH_ANY | 17386 | 0.011642 | 0.023092 | 0.007017 | 0.048563 | Spleen |
| Thyroid | INFA | 17386 | 0.01497 | 0.029677 | 0.009037 | 0.048823 | Thyroid |
| Whole_Blood | CVD_ATH_ANY | 17386 | 0.008938 | 0.016156 | 0.005401 | 0.048983 | Whole_Blood |
| Artery_Coronary | ADNC | 17388 | 0.018008 | 0.035879 | 0.011176 | 0.053574 | Artery_Coronary |
| Artery_Coronary | TDP43 | 17386 | 0.018625 | 0.037109 | 0.01163 | 0.054662 | Artery_Coronary |
| Brain_Hypothalamus | LEWY_grp | 17385 | 0.012674 | 0.02184 | 0.007968 | 0.055858 | Brain_Hypothalamus |
| Fallopian_Tube | ADNC | 17388 | 0.018086 | 0.035129 | 0.011387 | 0.056112 | Fallopian_Tube |
| Colon_Sigmoid | AMY_THAL | 17388 | 0.018949 | 0.037108 | 0.011953 | 0.056457 | Colon_Sigmoid |
| Artery_Aorta | TDP_3 | 17388 | 0.015534 | 0.031725 | 0.009876 | 0.057889 | Artery_Aorta |
| Artery_Tibial | ADNC | 17388 | 0.014846 | 0.030587 | 0.009462 | 0.058343 | Artery_Tibial |
| Esophagus_Gastroesophageal_J... | AMY_CERAD | 17386 | 0.018996 | 0.037428 | 0.012112 | 0.058419 | Esophagus_Gastroesophageal_Junction |

| | | | | | | | |
|---|---|---|---|---|---|---|---|
| Spleen | WMRCAL | 17388 | 0.010887 | 0.021594 | 0.006964 | 0.059004 | Spleen |
| Ovary | CVD_CAA_ANY | 17386 | 0.014362 | 0.029139 | 0.009244 | 0.060145 | Ovary |
| Skin_Not_Sun_Exposed_Suprapu... | HS_S | 17386 | 0.011412 | 0.022031 | 0.007353 | 0.060327 | Skin_Not_Sun_Exposed_Suprapubic |
| Skin_Sun_Exposed_Lower_leg | INFA | 17386 | 0.011132 | 0.021624 | 0.007208 | 0.061259 | Skin_Sun_Exposed_Lower_leg |
| Minor_Salivary_Gland | CVD_ART | 17388 | 0.014298 | 0.026637 | 0.009262 | 0.061343 | Minor_Salivary_Gland |
| Cervix_Endocervix | TDP43 | 17386 | 0.017293 | 0.034273 | 0.011217 | 0.06159 | Cervix_Endocervix |
| Ovary | CVD_ATH_ANY | 17386 | 0.014134 | 0.028676 | 0.009219 | 0.062634 | Ovary |
| Adrenal_Gland | CVD_ART | 17388 | 0.014419 | 0.027858 | 0.009409 | 0.062713 | Adrenal_Gland |
| Artery_Aorta | CVD_ATH | 17388 | 0.014683 | 0.029988 | 0.009634 | 0.063753 | Artery_Aorta |
| Minor_Salivary_Gland | WMRCAL | 17388 | 0.013937 | 0.025963 | 0.009193 | 0.064762 | Minor_Salivary_Gland |
| Brain_Cortex | LEWY_grp | 17385 | 0.010624 | 0.019393 | 0.007021 | 0.06512 | Brain_Cortex |
| Brain_Hypothalamus | LEWY_full | 17385 | 0.011706 | 0.020172 | 0.007829 | 0.067455 | Brain_Hypothalamus |
| Adrenal_Gland | CVD_CAA | 17385 | 0.014171 | 0.02738 | 0.009489 | 0.06768 | Adrenal_Gland |
| Adipose_Visceral_Omentum | CVD_ART | 17388 | 0.015755 | 0.03063 | 0.010566 | 0.06798 | Adipose_Visceral_Omentum |
| Colon_Sigmoid | ADNC | 17388 | 0.017412 | 0.034098 | 0.011726 | 0.068789 | Colon_Sigmoid |
| Skin_Sun_Exposed_Lower_leg | AMY_ANY | 17377 | 0.011023 | 0.021411 | 0.007472 | 0.07009 | Skin_Sun_Exposed_Lower_leg |
| Brain_Cortex | LEWY_full | 17385 | 0.010163 | 0.018551 | 0.006899 | 0.07039 | Brain_Cortex |
| Minor_Salivary_Gland | INFA | 17386 | 0.013016 | 0.024249 | 0.008856 | 0.070821 | Minor_Salivary_Gland |
| Skin_Not_Sun_Exposed_Suprapu... | AMY_ANY | 17377 | 0.01095 | 0.021137 | 0.00747 | 0.071349 | Skin_Not_Sun_Exposed_Suprapubic |
| Lung | ADNC | 17388 | 0.012652 | 0.024489 | 0.008631 | 0.071361 | Lung |
| Brain_Cerebellar_Hemisphere | HEMOR | 17378 | 0.00836 | 0.01679 | 0.005748 | 0.072911 | Brain_Cerebellar_Hemisphere |
| Cervix_Endocervix | CVD_ATH_ANY | 17386 | 0.015982 | 0.031675 | 0.010996 | 0.073061 | Cervix_Endocervix |
| Kidney_Cortex | BRAA | 17385 | 0.012338 | 0.0217 | 0.008491 | 0.07311 | Kidney_Cortex |
| Cervix_Endocervix | CVD_ATH | 17388 | 0.015876 | 0.031465 | 0.010972 | 0.073956 | Cervix_Endocervix |
| Cervix_Endocervix | B_SCORE | 17388 | 0.015779 | 0.031272 | 0.011002 | 0.075772 | Cervix_Endocervix |
| Stomach | ADNC | 17388 | 0.016032 | 0.029155 | 0.011245 | 0.076987 | Stomach |
| Spleen | CVD_ART | 17388 | 0.010119 | 0.020071 | 0.007124 | 0.077745 | Spleen |
| Brain_Cerebellum | HEMOR | 17378 | 0.008365 | 0.016621 | 0.005947 | 0.079786 | Brain_Cerebellum |
| Artery_Aorta | CVD_ART | 17388 | 0.013751 | 0.028084 | 0.009801 | 0.080314 | Artery_Aorta |
| Artery_Coronary | INFA | 17386 | 0.015363 | 0.030611 | 0.010958 | 0.080456 | Artery_Coronary |
| Testis | CVD_ART | 17388 | 0.007533 | 0.013021 | 0.005388 | 0.081063 | Testis |
| Uterus | CVD_ATH_ANY | 17386 | 0.014393 | 0.029352 | 0.010305 | 0.081277 | Uterus |
| Kidney_Medulla | BRAA | 17385 | 0.012272 | 0.022717 | 0.008803 | 0.081655 | Kidney_Medulla |
| Adipose_Subcutaneous | WMR | 17388 | 0.014259 | 0.028485 | 0.01023 | 0.081694 | Adipose_Subcutaneous |
| Brain_Cortex | CVD_CAA_ANY | 17386 | 0.009575 | 0.017477 | 0.006897 | 0.082545 | Brain_Cortex |
| Fallopian_Tube | B_SCORE | 17388 | 0.015477 | 0.030062 | 0.011179 | 0.083109 | Fallopian_Tube |

| | | | | | | | |
|---|---|---|---|---|---|---|---|
| Vagina | CVD_ATH | 17388 | 0.014521 | 0.027799 | 0.01052 | 0.083756 | Vagina |
| Brain_Caudate_basal_ganglia | AMY_ANY | 17377 | 0.010544 | 0.018231 | 0.007674 | 0.084749 | Brain_Caudate_basal_ganglia |
| Uterus | LEWY_full | 17385 | 0.014297 | 0.029156 | 0.010433 | 0.085299 | Uterus |
| Fallopian_Tube | CVD_S | 17387 | 0.015494 | 0.030093 | 0.011329 | 0.08573 | Fallopian_Tube |
| Prostate | WMRCAL | 17388 | 0.015553 | 0.029544 | 0.011384 | 0.085937 | Prostate |
| Esophagus_Mucosa | HS_S | 17386 | 0.009999 | 0.019347 | 0.007377 | 0.087664 | Esophagus_Mucosa |
| Small_Intestine_Terminal_Ile... | WMR | 17388 | 0.012108 | 0.022291 | 0.008979 | 0.088766 | Small_Intestine_Terminal_Ileum |
| Brain_Cortex | WBVD | 17388 | 0.009084 | 0.016581 | 0.006752 | 0.089262 | Brain_Cortex |
| Artery_Tibial | TDP_3 | 17388 | 0.01285 | 0.026475 | 0.009572 | 0.089727 | Artery_Tibial |
| Adrenal_Gland | TDP43 | 17386 | 0.013084 | 0.025281 | 0.00976 | 0.09003 | Adrenal_Gland |
| Pituitary | LEWY_ANY | 17385 | 0.011634 | 0.021715 | 0.008702 | 0.090622 | Pituitary |
| Vagina | HS_S | 17386 | 0.014163 | 0.027114 | 0.010596 | 0.090686 | Vagina |
| Skin_Not_Sun_Exposed_Suprapu... | WMR | 17388 | 0.009972 | 0.019249 | 0.007462 | 0.09073 | Skin_Not_Sun_Exposed_Suprapubic |
| Lung | INFA | 17386 | 0.011336 | 0.021941 | 0.008497 | 0.091097 | Lung |
| Brain_Amygdala | LEWY_grp | 17385 | 0.010309 | 0.01758 | 0.007805 | 0.093288 | Brain_Amygdala |
| Esophagus_Muscularis | AMY_THAL | 17388 | 0.015516 | 0.030633 | 0.011769 | 0.093703 | Esophagus_Muscularis |
| Muscle_Skeletal | AMY_A | 17388 | 0.008731 | 0.016521 | 0.006662 | 0.094997 | Muscle_Skeletal |
| Spleen | WBVD | 17388 | 0.009214 | 0.018276 | 0.007031 | 0.095025 | Spleen |
| Skin_Not_Sun_Exposed_Suprapu... | WMRCAL | 17388 | 0.009593 | 0.018519 | 0.007344 | 0.095733 | Skin_Not_Sun_Exposed_Suprapubic |
| Brain_Nucleus_accumbens_basa... | LEWY_grp | 17385 | 0.009795 | 0.017011 | 0.0075 | 0.095783 | Brain_Nucleus_accumbens_basal_ganglia |
| Brain_Amygdala | LEWY_full | 17385 | 0.009939 | 0.016948 | 0.007617 | 0.095994 | Brain_Amygdala |
| Adipose_Subcutaneous | ADNC | 17388 | 0.013223 | 0.026414 | 0.010144 | 0.096204 | Adipose_Subcutaneous |
| Bladder | AMY_CERAD | 17386 | 0.016306 | 0.031817 | 0.012531 | 0.096588 | Bladder |
| Brain_Nucleus_accumbens_basa... | AMY_ANY | 17377 | 0.009618 | 0.016704 | 0.00742 | 0.097455 | Brain_Nucleus_accumbens_basal_ganglia |
| Spleen | ADNC | 17388 | 0.008998 | 0.017847 | 0.006997 | 0.099237 | Spleen |
| Bladder | HS_S | 17386 | 0.015914 | 0.031052 | 0.012398 | 0.099659 | Bladder |
| Kidney_Cortex | AMY_CERAD | 17386 | 0.011029 | 0.019398 | 0.00861 | 0.10012 | Kidney_Cortex |
| Brain_Hypothalamus | LEWY_ANY | 17385 | 0.009976 | 0.017192 | 0.00781 | 0.10073 | Brain_Hypothalamus |
| Pituitary | B_SCORE | 17388 | 0.010812 | 0.02018 | 0.008472 | 0.10096 | Pituitary |
| Kidney_Cortex | CVD_CAA_ANY | 17386 | 0.011053 | 0.01944 | 0.008669 | 0.10116 | Kidney_Cortex |
| Spleen | CVD_ATH | 17388 | 0.008881 | 0.017616 | 0.007031 | 0.10326 | Spleen |
| Testis | ADNC | 17388 | 0.006721 | 0.011618 | 0.005339 | 0.10405 | Testis |
| Uterus | B_SCORE | 17388 | 0.012964 | 0.026437 | 0.010349 | 0.10518 | Uterus |
| Brain_Frontal_Cortex_BA9 | CVD_CAA_ANY | 17386 | 0.008314 | 0.015334 | 0.00666 | 0.10599 | Brain_Frontal_Cortex_BA9 |
| Adipose_Subcutaneous | MICR | 17385 | 0.012547 | 0.025064 | 0.010054 | 0.10603 | Adipose_Subcutaneous |
| Fallopian_Tube | HEMOR | 17378 | 0.014027 | 0.027245 | 0.011312 | 0.10751 | Fallopian_Tube |

| | | | | | | | |
|---|---|---|---|---|---|---|---|
| Heart_Atrial_Appendage | LEWY_ANY | 17385 | 0.011467 | 0.020603 | 0.009269 | 0.10802 | Heart_Atrial_Appendage |
| Uterus | ADNC | 17388 | 0.012955 | 0.02642 | 0.010501 | 0.10867 | Uterus |
| Small_Intestine_Terminal_Ile... | AMY_A | 17388 | 0.011254 | 0.020719 | 0.009126 | 0.10877 | Small_Intestine_Terminal_Ileum |
| Colon_Transverse | CVD_CAA | 17385 | 0.01346 | 0.024774 | 0.010927 | 0.10902 | Colon_Transverse |
| Vagina | BRAA | 17385 | 0.013181 | 0.025232 | 0.01071 | 0.10922 | Vagina |
| Skin_Not_Sun_Exposed_Suprapu... | INFA | 17386 | 0.008856 | 0.017097 | 0.007204 | 0.10947 | Skin_Not_Sun_Exposed_Suprapubic |
| Brain_Hippocampus | LEWY_grp | 17385 | 0.009679 | 0.016406 | 0.007883 | 0.10977 | Brain_Hippocampus |
| Brain_Hippocampus | LEWY_full | 17385 | 0.00945 | 0.016018 | 0.007711 | 0.11019 | Brain_Hippocampus |
| Brain_Frontal_Cortex_BA9 | WBVD | 17388 | 0.007988 | 0.014732 | 0.006518 | 0.11019 | Brain_Frontal_Cortex_BA9 |
| Brain_Putamen_basal_ganglia | AMY_ANY | 17377 | 0.009487 | 0.016158 | 0.007753 | 0.11055 | Brain_Putamen_basal_ganglia |
| Kidney_Cortex | AMY_A | 17388 | 0.010457 | 0.018393 | 0.008554 | 0.11077 | Kidney_Cortex |
| Adipose_Visceral_Omentum | AMY_CERAD | 17386 | 0.012844 | 0.024971 | 0.010596 | 0.11274 | Adipose_Visceral_Omentum |
| Brain_Cerebellum | WBVD | 17388 | 0.007211 | 0.014326 | 0.005958 | 0.11311 | Brain_Cerebellum |
| Artery_Tibial | CVD_ATH | 17388 | 0.011403 | 0.023493 | 0.009453 | 0.11388 | Artery_Tibial |
| Artery_Tibial | CVD_ART | 17388 | 0.011485 | 0.023663 | 0.009555 | 0.11468 | Artery_Tibial |
| Thyroid | AMY_ANY | 17377 | 0.011341 | 0.022484 | 0.00946 | 0.11529 | Thyroid |
| Small_Intestine_Terminal_Ile... | CVD_ART_ANY | 17387 | 0.010859 | 0.019992 | 0.009096 | 0.11628 | Small_Intestine_Terminal_Ileum |
| Minor_Salivary_Gland | WMR | 17388 | 0.010843 | 0.020201 | 0.009113 | 0.11706 | Minor_Salivary_Gland |
| Adrenal_Gland | CVD_S | 17387 | 0.011163 | 0.021568 | 0.009384 | 0.11711 | Adrenal_Gland |
| Brain_Nucleus_accumbens_basa... | WBVD | 17388 | 0.008597 | 0.014931 | 0.007236 | 0.11741 | Brain_Nucleus_accumbens_basal_ganglia |
| Adrenal_Gland | TDP_3 | 17388 | 0.011401 | 0.022028 | 0.009602 | 0.11755 | Adrenal_Gland |
| Brain_Anterior_cingulate_cor... | LEWY_grp | 17385 | 0.00849 | 0.01501 | 0.007163 | 0.11799 | Brain_Anterior_cingulate_cortex_BA24 |
| Uterus | MICR | 17385 | 0.012229 | 0.02494 | 0.010364 | 0.11902 | Uterus |
| Minor_Salivary_Gland | BRAA | 17385 | 0.010733 | 0.019994 | 0.009102 | 0.11919 | Minor_Salivary_Gland |
| Prostate | WMR | 17388 | 0.013472 | 0.02559 | 0.011471 | 0.12013 | Prostate |
| Nerve_Tibial | B_SCORE | 17388 | 0.011214 | 0.022597 | 0.009564 | 0.12048 | Nerve_Tibial |
| Ovary | LEWY_ANY | 17385 | 0.010793 | 0.021898 | 0.009245 | 0.12152 | Ovary |
| Colon_Transverse | BRAA | 17385 | 0.012825 | 0.023606 | 0.010989 | 0.12159 | Colon_Transverse |
| Heart_Atrial_Appendage | WBVD | 17388 | 0.010472 | 0.018814 | 0.008995 | 0.1222 | Heart_Atrial_Appendage |
| Colon_Sigmoid | TDP_3 | 17388 | 0.013745 | 0.026917 | 0.011861 | 0.12327 | Colon_Sigmoid |
| Brain_Cortex | AMY_ANY | 17377 | 0.008024 | 0.014647 | 0.006927 | 0.12338 | Brain_Cortex |
| Brain_Cortex | LEWY_ANY | 17385 | 0.007931 | 0.014477 | 0.006872 | 0.12425 | Brain_Cortex |
| Esophagus_Gastroesophageal_J... | ADNC | 17388 | 0.013848 | 0.027284 | 0.012 | 0.12426 | Esophagus_Gastroesophageal_Junction |
| Brain_Frontal_Cortex_BA9 | LEWY_grp | 17385 | 0.007824 | 0.014431 | 0.006781 | 0.12429 | Brain_Frontal_Cortex_BA9 |
| Cells_EBV-transformed_lympho... | CVD_ART_ANY | 17387 | 0.005291 | 0.011546 | 0.004595 | 0.12479 | Cells_EBV-transformed_lymphocytes |
| Adipose_Visceral_Omentum | TDP_3 | 17388 | 0.012285 | 0.023883 | 0.010728 | 0.12609 | Adipose_Visceral_Omentum |

| | | | | | | | |
|---|---|---|---|---|---|---|---|
| Brain_Amygdala | LEWY_ANY | 17385 | 0.008716 | 0.014863 | 0.007612 | 0.12611 | Brain_Amygdala |
| Artery_Aorta | INFA | 17386 | 0.010774 | 0.022004 | 0.009414 | 0.12623 | Artery_Aorta |
| Brain_Cerebellar_Hemisphere | WBVD | 17388 | 0.006597 | 0.013249 | 0.005766 | 0.12627 | Brain_Cerebellar_Hemisphere |
| Pituitary | AMY_ANY | 17377 | 0.010106 | 0.018863 | 0.008841 | 0.1265 | Pituitary |
| Cells_Cultured_fibroblasts | CVD_CAA_ANY | 17386 | 0.0072 | 0.015435 | 0.006366 | 0.12905 | Cells_Cultured_fibroblasts |
| Esophagus_Muscularis | ADNC | 17388 | 0.013145 | 0.025951 | 0.01163 | 0.12921 | Esophagus_Muscularis |
| Brain_Nucleus_accumbens_basa... | LEWY_ANY | 17385 | 0.008296 | 0.014408 | 0.007347 | 0.12941 | Brain_Nucleus_accumbens_basal_ganglia |
| Brain_Frontal_Cortex_BA9 | LEWY_full | 17385 | 0.007511 | 0.013853 | 0.006657 | 0.12962 | Brain_Frontal_Cortex_BA9 |
| Adrenal_Gland | CVD_ATH_ANY | 17386 | 0.010464 | 0.020219 | 0.009321 | 0.13079 | Adrenal_Gland |
| Brain_Anterior_cingulate_cor... | LEWY_full | 17385 | 0.007872 | 0.013918 | 0.007016 | 0.13096 | Brain_Anterior_cingulate_cortex_BA24 |
| Adrenal_Gland | AMY_CERAD | 17386 | 0.010778 | 0.020825 | 0.009627 | 0.13144 | Adrenal_Gland |
| Cells_Cultured_fibroblasts | CVD_S | 17387 | 0.00701 | 0.015027 | 0.006307 | 0.1332 | Cells_Cultured_fibroblasts |
| Cervix_Ectocervix | LEWY_full | 17385 | 0.012957 | 0.025422 | 0.01166 | 0.13325 | Cervix_Ectocervix |
| Ovary | CVD_S | 17387 | 0.010226 | 0.020746 | 0.00921 | 0.13345 | Ovary |
| Prostate | B_SCORE | 17388 | 0.012748 | 0.024215 | 0.011508 | 0.134 | Prostate |
| Bladder | WMR | 17388 | 0.013544 | 0.026428 | 0.012227 | 0.134 | Bladder |
| Pituitary | HEMOR | 17378 | 0.00948 | 0.017694 | 0.008588 | 0.13484 | Pituitary |
| Pancreas | AMY_A | 17388 | 0.008888 | 0.014872 | 0.008062 | 0.13516 | Pancreas |
| Spleen | AMY_A | 17388 | 0.007786 | 0.015443 | 0.00707 | 0.13541 | Spleen |
| Esophagus_Muscularis | AMY_A | 17388 | 0.01267 | 0.025014 | 0.011507 | 0.13544 | Esophagus_Muscularis |
| Prostate | BRAA | 17385 | 0.012766 | 0.02425 | 0.011594 | 0.13544 | Prostate |
| Skin_Sun_Exposed_Lower_leg | WMRCAL | 17388 | 0.008085 | 0.015705 | 0.007345 | 0.13551 | Skin_Sun_Exposed_Lower_leg |
| Esophagus_Gastroesophageal_J... | AMY_THAL | 17388 | 0.013363 | 0.026329 | 0.012152 | 0.13576 | Esophagus_Gastroesophageal_Junction |
| Brain_Anterior_cingulate_cor... | WBVD | 17388 | 0.00755 | 0.013348 | 0.006867 | 0.1358 | Brain_Anterior_cingulate_cortex_BA24 |
| Adipose_Subcutaneous | TDP_3 | 17388 | 0.01123 | 0.022433 | 0.010225 | 0.13605 | Adipose_Subcutaneous |
| Stomach | AMY_A | 17388 | 0.012439 | 0.022622 | 0.011365 | 0.13687 | Stomach |
| Fallopian_Tube | CVD_ATH_ANY | 17386 | 0.01215 | 0.023599 | 0.011147 | 0.13788 | Fallopian_Tube |
| Colon_Transverse | HS_S | 17386 | 0.011952 | 0.021999 | 0.010999 | 0.13861 | Colon_Transverse |
| Fallopian_Tube | CVD_ART | 17388 | 0.012355 | 0.023997 | 0.011402 | 0.13928 | Fallopian_Tube |
| Pancreas | INFA | 17386 | 0.008491 | 0.014209 | 0.007852 | 0.13977 | Pancreas |
| Adipose_Subcutaneous | TDP43 | 17386 | 0.01124 | 0.022452 | 0.010447 | 0.14101 | Adipose_Subcutaneous |
| Pituitary | CVD_ART_ANY | 17387 | 0.009256 | 0.017276 | 0.008662 | 0.14263 | Pituitary |
| Brain_Cortex | CVD_CAA | 17385 | 0.007236 | 0.013207 | 0.00679 | 0.14332 | Brain_Cortex |
| Fallopian_Tube | AMY_CERAD | 17386 | 0.012238 | 0.02377 | 0.011489 | 0.1434 | Fallopian_Tube |
| Fallopian_Tube | TDP43 | 17386 | 0.012216 | 0.023728 | 0.011485 | 0.14375 | Fallopian_Tube |
| Brain_Hippocampus | LEWY_ANY | 17385 | 0.008189 | 0.013882 | 0.007715 | 0.14424 | Brain_Hippocampus |

| | | | | | | | |
|---|---|---|---|---|---|---|---|
| Breast_Mammary_Tissue | AMY_CERAD | 17386 | 0.012751 | 0.024548 | 0.012036 | 0.14472 | Breast_Mammary_Tissue |
| Fallopian_Tube | TDP_3 | 17388 | 0.012074 | 0.023452 | 0.011424 | 0.14528 | Fallopian_Tube |
| Colon_Sigmoid | TDP43 | 17386 | 0.012671 | 0.024813 | 0.012006 | 0.14564 | Colon_Sigmoid |
| Cells_Cultured_fibroblasts | CVD_ATH | 17388 | 0.006568 | 0.01408 | 0.00624 | 0.14628 | Cells_Cultured_fibroblasts |
| Brain_Nucleus_accumbens_basa... | AMY_THAL | 17388 | 0.007677 | 0.013333 | 0.007298 | 0.14642 | Brain_Nucleus_accumbens_basal_ganglia |
| Artery_Aorta | BRAA | 17385 | 0.010241 | 0.020917 | 0.009748 | 0.14673 | Artery_Aorta |
| Uterus | CVD_ATH | 17388 | 0.010893 | 0.022214 | 0.010372 | 0.14682 | Uterus |
| Vagina | HEMOR | 17378 | 0.010973 | 0.021007 | 0.010546 | 0.14906 | Vagina |
| Adrenal_Gland | ADNC | 17388 | 0.009763 | 0.018863 | 0.009423 | 0.1501 | Adrenal_Gland |
| Fallopian_Tube | CVD_ATH | 17388 | 0.011666 | 0.022659 | 0.011262 | 0.15014 | Fallopian_Tube |
| Testis | B_SCORE | 17388 | 0.005546 | 0.009586 | 0.005355 | 0.15019 | Testis |
| Ovary | LEWY_full | 17385 | 0.009596 | 0.019468 | 0.009287 | 0.15076 | Ovary |
| Brain_Caudate_basal_ganglia | LEWY_grp | 17385 | 0.008021 | 0.013868 | 0.007765 | 0.15084 | Brain_Caudate_basal_ganglia |
| Cervix_Ectocervix | CVD_ATH_ANY | 17386 | 0.011867 | 0.023283 | 0.011514 | 0.15137 | Cervix_Ectocervix |
| Brain_Frontal_Cortex_BA9 | AMY_ANY | 17377 | 0.006874 | 0.012678 | 0.006671 | 0.15142 | Brain_Frontal_Cortex_BA9 |
| Minor_Salivary_Gland | AMY_CERAD | 17386 | 0.009369 | 0.017455 | 0.009144 | 0.15278 | Minor_Salivary_Gland |
| Whole_Blood | WBVD | 17388 | 0.005519 | 0.009976 | 0.005389 | 0.1529 | Whole_Blood |
| Nerve_Tibial | INFA | 17386 | 0.009485 | 0.019112 | 0.009267 | 0.15303 | Nerve_Tibial |
| Vagina | AMY_ANY | 17377 | 0.010951 | 0.020964 | 0.010758 | 0.15435 | Vagina |
| Heart_Left_Ventricle | WBVD | 17388 | 0.008746 | 0.014607 | 0.008671 | 0.1566 | Heart_Left_Ventricle |
| Muscle_Skeletal | ADNC | 17388 | 0.006697 | 0.012671 | 0.006645 | 0.15678 | Muscle_Skeletal |
| Whole_Blood | AMY_A | 17388 | 0.005452 | 0.009855 | 0.005473 | 0.15957 | Whole_Blood |
| Vagina | TDP43 | 17386 | 0.010797 | 0.02067 | 0.01087 | 0.16028 | Vagina |
| Heart_Atrial_Appendage | ADNC | 17388 | 0.008875 | 0.015945 | 0.008973 | 0.16133 | Heart_Atrial_Appendage |
| Brain_Frontal_Cortex_BA9 | CVD_CAA | 17385 | 0.006369 | 0.011747 | 0.006553 | 0.16554 | Brain_Frontal_Cortex_BA9 |
| Prostate | HS_S | 17386 | 0.011145 | 0.021171 | 0.011522 | 0.16671 | Prostate |
| Small_Intestine_Terminal_Ile... | CVD_S | 17387 | 0.008807 | 0.016214 | 0.009109 | 0.16682 | Small_Intestine_Terminal_Ileum |
| Brain_Anterior_cingulate_cor... | CVD_CAA_ANY | 17386 | 0.006783 | 0.011993 | 0.00702 | 0.16695 | Brain_Anterior_cingulate_cortex_BA24 |
| Brain_Anterior_cingulate_cor... | LEWY_ANY | 17385 | 0.006723 | 0.011886 | 0.007001 | 0.16848 | Brain_Anterior_cingulate_cortex_BA24 |
| Colon_Transverse | TDP_3 | 17388 | 0.010575 | 0.019465 | 0.01106 | 0.16951 | Colon_Transverse |
| Thyroid | CVD_CAA | 17385 | 0.008683 | 0.017214 | 0.009176 | 0.17201 | Thyroid |
| Uterus | INFA | 17386 | 0.009549 | 0.019474 | 0.01011 | 0.17245 | Uterus |
| Cells_EBV-transformed_lympho... | BRAA | 17385 | 0.004389 | 0.009578 | 0.004677 | 0.17405 | Cells_EBV-transformed_lymphocytes |
| Skin_Not_Sun_Exposed_Suprapu... | WBVD | 17388 | 0.006926 | 0.013369 | 0.007389 | 0.17433 | Skin_Not_Sun_Exposed_Suprapubic |
| Brain_Nucleus_accumbens_basa... | LEWY_full | 17385 | 0.006878 | 0.011944 | 0.007366 | 0.17525 | Brain_Nucleus_accumbens_basal_ganglia |
| Spleen | AMY_CERAD | 17386 | 0.006613 | 0.013116 | 0.007093 | 0.17559 | Spleen |

| | | | | | | | |
|---|---|---|---|---|---|---|---|
| Cervix_Ectocervix | MICR | 17385 | 0.010663 | 0.02092 | 0.011481 | 0.17652 | Cervix_Ectocervix |
| Brain_Caudate_basal_ganglia | AMY_THAL | 17388 | 0.007004 | 0.01211 | 0.007546 | 0.17669 | Brain_Caudate_basal_ganglia |
| Artery_Aorta | ADNC | 17388 | 0.009022 | 0.018425 | 0.009757 | 0.17758 | Artery_Aorta |
| Stomach | TDP_3 | 17388 | 0.0106 | 0.019278 | 0.011473 | 0.17777 | Stomach |
| Cervix_Endocervix | LEWY_full | 17385 | 0.010222 | 0.020258 | 0.01109 | 0.17835 | Cervix_Endocervix |
| Skin_Not_Sun_Exposed_Suprapu... | AMY_CERAD | 17386 | 0.006944 | 0.013405 | 0.007535 | 0.17839 | Skin_Not_Sun_Exposed_Suprapubic |
| Small_Intestine_Terminal_Ile... | AMY_THAL | 17388 | 0.008464 | 0.015582 | 0.00921 | 0.17907 | Small_Intestine_Terminal_Ileum |
| Cervix_Endocervix | MICR | 17385 | 0.009914 | 0.019647 | 0.010919 | 0.18198 | Cervix_Endocervix |
| Thyroid | CVD_ART_ANY | 17387 | 0.008512 | 0.016873 | 0.009382 | 0.18213 | Thyroid |
| Esophagus_Mucosa | BRAA | 17385 | 0.006784 | 0.013126 | 0.007483 | 0.18233 | Esophagus_Mucosa |
| Uterus | LEWY_ANY | 17385 | 0.009436 | 0.019243 | 0.010528 | 0.18507 | Uterus |
| Cells_Cultured_fibroblasts | CVD_CAA | 17385 | 0.005633 | 0.012076 | 0.006301 | 0.18566 | Cells_Cultured_fibroblasts |
| Thyroid | ADNC | 17388 | 0.008257 | 0.016369 | 0.009278 | 0.18674 | Thyroid |
| Spleen | CVD_ART_ANY | 17387 | 0.00624 | 0.012377 | 0.007022 | 0.18709 | Spleen |
| Artery_Aorta | B_SCORE | 17388 | 0.008551 | 0.017463 | 0.009736 | 0.18992 | Artery_Aorta |
| Skin_Sun_Exposed_Lower_leg | WMR | 17388 | 0.00655 | 0.012723 | 0.007474 | 0.19042 | Skin_Sun_Exposed_Lower_leg |
| Cervix_Ectocervix | WMR | 17388 | 0.010013 | 0.019645 | 0.011519 | 0.19236 | Cervix_Ectocervix |
| Esophagus_Gastroesophageal_J... | AMY_A | 17388 | 0.010342 | 0.020377 | 0.0119 | 0.19242 | Esophagus_Gastroesophageal_Junction |
| Heart_Left_Ventricle | LEWY_ANY | 17385 | 0.007752 | 0.012948 | 0.008942 | 0.19298 | Heart_Left_Ventricle |
| Heart_Left_Ventricle | ADNC | 17388 | 0.007601 | 0.012695 | 0.008777 | 0.19327 | Heart_Left_Ventricle |
| Brain_Caudate_basal_ganglia | LEWY_full | 17385 | 0.006556 | 0.011336 | 0.007626 | 0.19498 | Brain_Caudate_basal_ganglia |
| Brain_Frontal_Cortex_BA9 | LEWY_ANY | 17385 | 0.005694 | 0.010501 | 0.006633 | 0.19536 | Brain_Frontal_Cortex_BA9 |
| Nerve_Tibial | BRAA | 17385 | 0.008164 | 0.016451 | 0.009563 | 0.19663 | Nerve_Tibial |
| Pituitary | HS_S | 17386 | 0.007361 | 0.013739 | 0.00868 | 0.19824 | Pituitary |
| Stomach | AMY_CERAD | 17386 | 0.009797 | 0.017817 | 0.011559 | 0.19834 | Stomach |
| Lung | TDP43 | 17386 | 0.007475 | 0.014469 | 0.008915 | 0.20089 | Lung |
| Esophagus_Gastroesophageal_J... | TDP_3 | 17388 | 0.010204 | 0.020105 | 0.012176 | 0.20102 | Esophagus_Gastroesophageal_Junction |
| Esophagus_Gastroesophageal_J... | HEMOR | 17378 | 0.009998 | 0.0197 | 0.011987 | 0.20213 | Esophagus_Gastroesophageal_Junction |
| Artery_Coronary | TDP_3 | 17388 | 0.00952 | 0.018967 | 0.011444 | 0.20275 | Artery_Coronary |
| Brain_Putamen_basal_ganglia | LEWY_grp | 17385 | 0.006506 | 0.01108 | 0.007837 | 0.20325 | Brain_Putamen_basal_ganglia |
| Uterus | LEWY_grp | 17385 | 0.008768 | 0.017881 | 0.010627 | 0.20469 | Uterus |
| Lung | WMR | 17388 | 0.007056 | 0.013657 | 0.008605 | 0.20614 | Lung |
| Artery_Coronary | CVD_ATH | 17388 | 0.009166 | 0.018263 | 0.01119 | 0.20637 | Artery_Coronary |
| Brain_Caudate_basal_ganglia | WBVD | 17388 | 0.006101 | 0.010549 | 0.00747 | 0.20704 | Brain_Caudate_basal_ganglia |
| Brain_Anterior_cingulate_cor... | AMY_ANY | 17377 | 0.005734 | 0.010138 | 0.007052 | 0.20809 | Brain_Anterior_cingulate_cortex_BA24 |
| Colon_Sigmoid | AMY_CERAD | 17386 | 0.00967 | 0.018936 | 0.011928 | 0.20879 | Colon_Sigmoid |

| | | | | | | | |
|---|---|---|---|---|---|---|---|
| Skin_Sun_Exposed_Lower_leg | AMY_CERAD | 17386 | 0.006114 | 0.011876 | 0.007542 | 0.20882 | Skin_Sun_Exposed_Lower_leg |
| Artery_Aorta | AMY_CERAD | 17386 | 0.00796 | 0.016256 | 0.009842 | 0.20935 | Artery_Aorta |
| Thyroid | AMY_CERAD | 17386 | 0.00765 | 0.015166 | 0.009477 | 0.20977 | Thyroid |
| Adrenal_Gland | AMY_THAL | 17388 | 0.007531 | 0.014551 | 0.009403 | 0.21159 | Adrenal_Gland |
| Adipose_Visceral_Omentum | CVD_ATH_ANY | 17386 | 0.008342 | 0.016217 | 0.0105 | 0.21349 | Adipose_Visceral_Omentum |
| Prostate | AMY_ANY | 17377 | 0.009168 | 0.017415 | 0.011603 | 0.21473 | Prostate |
| Brain_Frontal_Cortex_BA9 | MICR | 17385 | 0.005124 | 0.009451 | 0.006488 | 0.21483 | Brain_Frontal_Cortex_BA9 |
| Muscle_Skeletal | AMY_THAL | 17388 | 0.00525 | 0.009934 | 0.006649 | 0.21488 | Muscle_Skeletal |
| Brain_Anterior_cingulate_cor... | CVD_CAA | 17385 | 0.005454 | 0.009642 | 0.006924 | 0.21547 | Brain_Anterior_cingulate_cortex_BA24 |
| Brain_Cortex | MICR | 17385 | 0.005283 | 0.009644 | 0.006728 | 0.21615 | Brain_Cortex |
| Small_Intestine_Terminal_Ile... | AMY_CERAD | 17386 | 0.007234 | 0.013319 | 0.009216 | 0.21626 | Small_Intestine_Terminal_Ileum |
| Adipose_Subcutaneous | AMY_CERAD | 17386 | 0.008087 | 0.016154 | 0.010332 | 0.2169 | Adipose_Subcutaneous |
| Colon_Transverse | AMY_A | 17388 | 0.008604 | 0.015836 | 0.011016 | 0.21741 | Colon_Transverse |
| Stomach | INFA | 17386 | 0.008669 | 0.015765 | 0.011172 | 0.21889 | Stomach |
| Stomach | BRAA | 17385 | 0.008768 | 0.015945 | 0.011373 | 0.22037 | Stomach |
| Cervix_Ectocervix | B_SCORE | 17388 | 0.00892 | 0.0175 | 0.011576 | 0.22051 | Cervix_Ectocervix |
| Esophagus_Muscularis | TDP_3 | 17388 | 0.009096 | 0.017959 | 0.011811 | 0.2206 | Esophagus_Muscularis |
| Cells_Cultured_fibroblasts | CVD_ART_ANY | 17387 | 0.004801 | 0.010291 | 0.006237 | 0.22076 | Cells_Cultured_fibroblasts |
| Ovary | LEWY_grp | 17385 | 0.007249 | 0.014707 | 0.009434 | 0.22114 | Ovary |
| Vagina | AMY_CERAD | 17386 | 0.008166 | 0.015634 | 0.01069 | 0.22246 | Vagina |
| Esophagus_Mucosa | AMY_THAL | 17388 | 0.005622 | 0.010877 | 0.007359 | 0.22247 | Esophagus_Mucosa |
| Skin_Not_Sun_Exposed_Suprapu... | CVD_S | 17387 | 0.005596 | 0.010803 | 0.00734 | 0.22291 | Skin_Not_Sun_Exposed_Suprapubic |
| Liver | HEMOR | 17378 | 0.004637 | 0.008354 | 0.006089 | 0.2232 | Liver |
| Testis | CVD_ART_ANY | 17387 | 0.004061 | 0.00702 | 0.005339 | 0.22343 | Testis |
| Colon_Transverse | TDP43 | 17386 | 0.008599 | 0.015828 | 0.011306 | 0.22346 | Colon_Transverse |
| Artery_Aorta | MICR | 17385 | 0.00729 | 0.014889 | 0.009593 | 0.22365 | Artery_Aorta |
| Breast_Mammary_Tissue | MICR | 17385 | 0.008899 | 0.017132 | 0.011796 | 0.22531 | Breast_Mammary_Tissue |
| Lung | AMY_A | 17388 | 0.006619 | 0.012812 | 0.008786 | 0.22563 | Lung |
| Uterus | CVD_CAA_ANY | 17386 | 0.007884 | 0.016078 | 0.010488 | 0.22613 | Uterus |
| Esophagus_Muscularis | HEMOR | 17378 | 0.008701 | 0.017178 | 0.011637 | 0.22734 | Esophagus_Muscularis |
| Testis | HS_S | 17386 | 0.003966 | 0.006855 | 0.005322 | 0.22809 | Testis |
| Cells_EBV-transformed_lympho... | CVD_S | 17387 | 0.003453 | 0.007537 | 0.004638 | 0.22829 | Cells_EBV-transformed_lymphocytes |
| Minor_Salivary_Gland | AMY_A | 17388 | 0.006739 | 0.012555 | 0.009061 | 0.22852 | Minor_Salivary_Gland |
| Nerve_Tibial | HEMOR | 17378 | 0.007001 | 0.014108 | 0.009529 | 0.23126 | Nerve_Tibial |
| Esophagus_Mucosa | AMY_CERAD | 17386 | 0.005461 | 0.010566 | 0.007471 | 0.23241 | Esophagus_Mucosa |
| Prostate | CVD_CAA_ANY | 17386 | 0.008533 | 0.016208 | 0.011675 | 0.23244 | Prostate |

| | | | | | | | |
|---|---|---|---|---|---|---|---|
| Brain_Anterior_cingulate_cor... | MICR | 17385 | 0.004973 | 0.008792 | 0.006842 | 0.23368 | Brain_Anterior_cingulate_cortex_BA24 |
| Adipose_Visceral_Omentum | CVD_ATH | 17388 | 0.007598 | 0.014771 | 0.01047 | 0.23403 | Adipose_Visceral_Omentum |
| Brain_Caudate_basal_ganglia | LEWY_ANY | 17385 | 0.005452 | 0.009427 | 0.007601 | 0.23662 | Brain_Caudate_basal_ganglia |
| Spleen | WMR | 17388 | 0.005063 | 0.010042 | 0.007059 | 0.23662 | Spleen |
| Fallopian_Tube | INFA | 17386 | 0.007809 | 0.015167 | 0.010957 | 0.23804 | Fallopian_Tube |
| Whole_Blood | ADNC | 17388 | 0.003902 | 0.007053 | 0.005478 | 0.23815 | Whole_Blood |
| Brain_Caudate_basal_ganglia | CVD_CAA_ANY | 17386 | 0.005419 | 0.00937 | 0.007623 | 0.23858 | Brain_Caudate_basal_ganglia |
| Stomach | CVD_ART | 17388 | 0.00818 | 0.014876 | 0.011541 | 0.23924 | Stomach |
| Brain_Putamen_basal_ganglia | AMY_THAL | 17388 | 0.005378 | 0.00916 | 0.007618 | 0.24008 | Brain_Putamen_basal_ganglia |
| Liver | CVD_CAA | 17385 | 0.004299 | 0.007745 | 0.006118 | 0.24112 | Liver |
| Brain_Putamen_basal_ganglia | LEWY_ANY | 17385 | 0.005392 | 0.009182 | 0.007685 | 0.24148 | Brain_Putamen_basal_ganglia |
| Cells_Cultured_fibroblasts | HEMOR | 17378 | 0.00437 | 0.009369 | 0.006241 | 0.2419 | Cells_Cultured_fibroblasts |
| Skin_Sun_Exposed_Lower_leg | WBVD | 17388 | 0.005178 | 0.010058 | 0.007404 | 0.24217 | Skin_Sun_Exposed_Lower_leg |
| Uterus | CVD_CAA | 17385 | 0.007192 | 0.014668 | 0.010444 | 0.24552 | Uterus |
| Esophagus_Mucosa | WMR | 17388 | 0.005082 | 0.009833 | 0.007382 | 0.2456 | Esophagus_Mucosa |
| Adipose_Visceral_Omentum | TDP43 | 17386 | 0.007384 | 0.014356 | 0.010801 | 0.24711 | Adipose_Visceral_Omentum |
| Thyroid | WMR | 17388 | 0.006323 | 0.012535 | 0.009255 | 0.24725 | Thyroid |
| Brain_Nucleus_accumbens_basa... | CVD_CAA_ANY | 17386 | 0.005028 | 0.008732 | 0.00736 | 0.24726 | Brain_Nucleus_accumbens_basal_ganglia |
| Adipose_Visceral_Omentum | AMY_A | 17388 | 0.007218 | 0.014033 | 0.010572 | 0.24738 | Adipose_Visceral_Omentum |
| Cervix_Endocervix | CVD_ART_ANY | 17387 | 0.007583 | 0.015027 | 0.011113 | 0.24752 | Cervix_Endocervix |
| Brain_Spinal_cord_cervical_c... | LEWY_full | 17385 | 0.005574 | 0.010029 | 0.008176 | 0.24771 | Brain_Spinal_cord_cervical_c-1 |
| Brain_Putamen_basal_ganglia | CVD_CAA_ANY | 17386 | 0.00523 | 0.008908 | 0.007709 | 0.24875 | Brain_Putamen_basal_ganglia |
| Brain_Caudate_basal_ganglia | CVD_CAA | 17385 | 0.005068 | 0.008763 | 0.007526 | 0.25033 | Brain_Caudate_basal_ganglia |
| Adrenal_Gland | LEWY_ANY | 17385 | 0.006357 | 0.012282 | 0.009459 | 0.25079 | Adrenal_Gland |
| Spleen | TDP43 | 17386 | 0.004828 | 0.009577 | 0.007187 | 0.25086 | Spleen |
| Testis | INFA | 17386 | 0.003501 | 0.006052 | 0.005223 | 0.25136 | Testis |
| Brain_Caudate_basal_ganglia | HS_S | 17386 | 0.005043 | 0.008719 | 0.007534 | 0.25165 | Brain_Caudate_basal_ganglia |
| Artery_Coronary | CVD_ART | 17388 | 0.007588 | 0.015119 | 0.011376 | 0.25239 | Artery_Coronary |
| Cervix_Endocervix | WMR | 17388 | 0.007278 | 0.014424 | 0.011029 | 0.25466 | Cervix_Endocervix |
| Fallopian_Tube | WMR | 17388 | 0.007489 | 0.014545 | 0.011351 | 0.25472 | Fallopian_Tube |
| Kidney_Medulla | AMY_A | 17388 | 0.005796 | 0.010728 | 0.00879 | 0.25485 | Kidney_Medulla |
| Cervix_Ectocervix | CVD_ART_ANY | 17387 | 0.007685 | 0.015078 | 0.01167 | 0.2551 | Cervix_Ectocervix |
| Brain_Putamen_basal_ganglia | HS_S | 17386 | 0.005012 | 0.008536 | 0.007617 | 0.25528 | Brain_Putamen_basal_ganglia |
| Cells_EBV-transformed_lympho... | CVD_ATH | 17388 | 0.002999 | 0.006544 | 0.004583 | 0.25644 | Cells_EBV-transformed_lymphocytes |
| Muscle_Skeletal | CVD_CAA_ANY | 17386 | 0.004343 | 0.008219 | 0.00664 | 0.25653 | Muscle_Skeletal |
| Brain_Putamen_basal_ganglia | LEWY_full | 17385 | 0.005023 | 0.008554 | 0.007698 | 0.25705 | Brain_Putamen_basal_ganglia |

| | | | | | | | |
|---|---|---|---|---|---|---|---|
| Cells_Cultured_fibroblasts | TDP_3 | 17388 | 0.004161 | 0.00892 | 0.006389 | 0.25744 | Cells_Cultured_fibroblasts |
| Lung | CVD_S | 17387 | 0.005639 | 0.010913 | 0.008669 | 0.25772 | Lung |
| Brain_Amygdala | WBVD | 17388 | 0.004839 | 0.008252 | 0.00745 | 0.258 | Brain_Amygdala |
| Adipose_Subcutaneous | CVD_ATH_ANY | 17386 | 0.006533 | 0.01305 | 0.010083 | 0.25853 | Adipose_Subcutaneous |
| Liver | CVD_CAA_ANY | 17386 | 0.003953 | 0.007121 | 0.006141 | 0.25991 | Liver |
| Colon_Sigmoid | CVD_S | 17387 | 0.007527 | 0.01474 | 0.011782 | 0.26146 | Colon_Sigmoid |
| Artery_Tibial | AMY_THAL | 17388 | 0.006083 | 0.012532 | 0.009524 | 0.26153 | Artery_Tibial |
| Nerve_Tibial | CVD_ATH | 17388 | 0.005954 | 0.011996 | 0.00933 | 0.2617 | Nerve_Tibial |
| Lung | WMRCAL | 17388 | 0.005464 | 0.010575 | 0.008608 | 0.2628 | Lung |
| Brain_Spinal_cord_cervical_c... | B_SCORE | 17388 | 0.005105 | 0.009184 | 0.008049 | 0.26299 | Brain_Spinal_cord_cervical_c-1 |
| Brain_Hippocampus | AMY_THAL | 17388 | 0.004861 | 0.00824 | 0.007671 | 0.26314 | Brain_Hippocampus |
| Pituitary | CVD_CAA | 17385 | 0.005476 | 0.010221 | 0.008747 | 0.26564 | Pituitary |
| Esophagus_Gastroesophageal_J... | MICR | 17385 | 0.007497 | 0.014771 | 0.011988 | 0.26588 | Esophagus_Gastroesophageal_Junction |
| Testis | CVD_ATH_ANY | 17386 | 0.003354 | 0.005797 | 0.005376 | 0.26637 | Testis |
| Brain_Cerebellum | LEWY_full | 17385 | 0.003752 | 0.007455 | 0.006018 | 0.2665 | Brain_Cerebellum |
| Cells_EBV-transformed_lympho... | CVD_ATH_ANY | 17386 | 0.002873 | 0.00627 | 0.004613 | 0.26673 | Cells_EBV-transformed_lymphocytes |
| Bladder | TDP_3 | 17388 | 0.007757 | 0.015135 | 0.012479 | 0.26712 | Bladder |
| Vagina | B_SCORE | 17388 | 0.006513 | 0.012468 | 0.010548 | 0.26849 | Vagina |
| Brain_Anterior_cingulate_cor... | AMY_THAL | 17388 | 0.0043 | 0.007603 | 0.006979 | 0.2689 | Brain_Anterior_cingulate_cortex_BA24 |
| Brain_Putamen_basal_ganglia | WBVD | 17388 | 0.004623 | 0.007873 | 0.007562 | 0.27049 | Brain_Putamen_basal_ganglia |
| Kidney_Medulla | CVD_ART | 17388 | 0.005379 | 0.009958 | 0.008878 | 0.27229 | Kidney_Medulla |
| Whole_Blood | AMY_THAL | 17388 | 0.003329 | 0.006018 | 0.005517 | 0.27309 | Whole_Blood |
| Esophagus_Muscularis | CVD_S | 17387 | 0.006999 | 0.013818 | 0.011714 | 0.2751 | Esophagus_Muscularis |
| Cells_EBV-transformed_lympho... | B_SCORE | 17388 | 0.002746 | 0.005993 | 0.004597 | 0.27513 | Cells_EBV-transformed_lymphocytes |
| Brain_Substantia_nigra | LEWY_full | 17385 | 0.004914 | 0.008467 | 0.008231 | 0.27526 | Brain_Substantia_nigra |
| Brain_Amygdala | CVD_CAA_ANY | 17386 | 0.004534 | 0.007732 | 0.007624 | 0.27603 | Brain_Amygdala |
| Artery_Coronary | LEWY_full | 17385 | 0.006792 | 0.013533 | 0.011449 | 0.2765 | Artery_Coronary |
| Prostate | MICR | 17385 | 0.006664 | 0.012658 | 0.011313 | 0.27792 | Prostate |
| Small_Intestine_Terminal_Ile... | HEMOR | 17378 | 0.005303 | 0.009764 | 0.00903 | 0.27851 | Small_Intestine_Terminal_Ileum |
| Brain_Amygdala | AMY_THAL | 17388 | 0.004432 | 0.007558 | 0.007582 | 0.27943 | Brain_Amygdala |
| Brain_Cerebellum | AMY_THAL | 17388 | 0.003494 | 0.006942 | 0.005987 | 0.27974 | Brain_Cerebellum |
| Esophagus_Gastroesophageal_J... | TDP43 | 17386 | 0.007145 | 0.014078 | 0.012322 | 0.28101 | Esophagus_Gastroesophageal_Junction |
| Cervix_Endocervix | CVD_ART | 17388 | 0.006447 | 0.012778 | 0.011125 | 0.28112 | Cervix_Endocervix |
| Muscle_Skeletal | HEMOR | 17378 | 0.003833 | 0.007252 | 0.006624 | 0.28144 | Muscle_Skeletal |
| Brain_Cerebellum | LEWY_grp | 17385 | 0.003516 | 0.006985 | 0.006129 | 0.28314 | Brain_Cerebellum |
| Brain_Nucleus_accumbens_basa... | CVD_CAA | 17385 | 0.004138 | 0.007186 | 0.007255 | 0.28422 | Brain_Nucleus_accumbens_basal_ganglia |

| | | | | | | | |
|---|---|---|---|---|---|---|---|
| Artery_Aorta | CVD_ART_ANY | 17387 | 0.005537 | 0.011308 | 0.00972 | 0.28445 | Artery_Aorta |
| Colon_Sigmoid | CVD_CAA | 17385 | 0.00666 | 0.013043 | 0.011718 | 0.2849 | Colon_Sigmoid |
| Adipose_Subcutaneous | AMY_A | 17388 | 0.005775 | 0.011537 | 0.010192 | 0.28548 | Adipose_Subcutaneous |
| Colon_Transverse | CVD_CAA_ANY | 17386 | 0.006277 | 0.011553 | 0.011107 | 0.28601 | Colon_Transverse |
| Cervix_Endocervix | CVD_CAA_ANY | 17386 | 0.006247 | 0.01238 | 0.01109 | 0.28663 | Cervix_Endocervix |
| Small_Intestine_Terminal_Ile... | CVD_ATH_ANY | 17386 | 0.005106 | 0.009401 | 0.009087 | 0.28709 | Small_Intestine_Terminal_Ileum |
| Heart_Atrial_Appendage | CVD_ATH | 17388 | 0.00509 | 0.009145 | 0.009063 | 0.28719 | Heart_Atrial_Appendage |
| Brain_Putamen_basal_ganglia | CVD_CAA | 17385 | 0.004237 | 0.007216 | 0.007608 | 0.28881 | Brain_Putamen_basal_ganglia |
| Cervix_Endocervix | INFA | 17386 | 0.005987 | 0.011866 | 0.010766 | 0.28907 | Cervix_Endocervix |
| Cells_Cultured_fibroblasts | WMR | 17388 | 0.003507 | 0.007517 | 0.006319 | 0.28947 | Cells_Cultured_fibroblasts |
| Brain_Frontal_Cortex_BA9 | AMY_THAL | 17388 | 0.003655 | 0.006741 | 0.006613 | 0.29025 | Brain_Frontal_Cortex_BA9 |
| Esophagus_Gastroesophageal_J... | CVD_S | 17387 | 0.006673 | 0.013147 | 0.012078 | 0.29032 | Esophagus_Gastroesophageal_Junction |
| Esophagus_Mucosa | CVD_S | 17387 | 0.004063 | 0.007862 | 0.007374 | 0.29082 | Esophagus_Mucosa |
| Small_Intestine_Terminal_Ile... | CVD_CAA_ANY | 17386 | 0.005031 | 0.009262 | 0.009201 | 0.29228 | Small_Intestine_Terminal_Ileum |
| Thyroid | AMY_A | 17388 | 0.005066 | 0.010043 | 0.009313 | 0.29323 | Thyroid |
| Kidney_Medulla | TDP_3 | 17388 | 0.004901 | 0.009073 | 0.009029 | 0.29363 | Kidney_Medulla |
| Adipose_Subcutaneous | CVD_ATH | 17388 | 0.005464 | 0.010914 | 0.010098 | 0.29423 | Adipose_Subcutaneous |
| Stomach | WMR | 17388 | 0.006149 | 0.011182 | 0.011413 | 0.29504 | Stomach |
| Brain_Cerebellar_Hemisphere | LEWY_full | 17385 | 0.003127 | 0.00628 | 0.005814 | 0.29534 | Brain_Cerebellar_Hemisphere |
| Kidney_Medulla | HS_S | 17386 | 0.004743 | 0.00878 | 0.008821 | 0.29541 | Kidney_Medulla |
| Whole_Blood | CVD_ART | 17388 | 0.002949 | 0.005331 | 0.005486 | 0.29544 | Whole_Blood |
| Brain_Amygdala | AMY_ANY | 17377 | 0.004108 | 0.007006 | 0.007692 | 0.29665 | Brain_Amygdala |
| Ovary | CVD_ATH | 17388 | 0.004864 | 0.009868 | 0.009166 | 0.29784 | Ovary |
| Whole_Blood | WMR | 17388 | 0.002888 | 0.005221 | 0.00545 | 0.29806 | Whole_Blood |
| Brain_Cortex | AMY_THAL | 17388 | 0.003617 | 0.006602 | 0.006837 | 0.29842 | Brain_Cortex |
| Artery_Tibial | MICR | 17385 | 0.004958 | 0.010214 | 0.009396 | 0.29888 | Artery_Tibial |
| Lung | AMY_ANY | 17377 | 0.004635 | 0.008972 | 0.008814 | 0.29949 | Lung |
| Colon_Transverse | AMY_THAL | 17388 | 0.00573 | 0.010547 | 0.011061 | 0.30222 | Colon_Transverse |
| Cervix_Ectocervix | LEWY_ANY | 17385 | 0.006054 | 0.011878 | 0.011747 | 0.30315 | Cervix_Ectocervix |
| Brain_Spinal_cord_cervical_c... | LEWY_grp | 17385 | 0.00429 | 0.007719 | 0.008344 | 0.30357 | Brain_Spinal_cord_cervical_c-1 |
| Brain_Hippocampus | AMY_ANY | 17377 | 0.003972 | 0.006733 | 0.007782 | 0.30489 | Brain_Hippocampus |
| Stomach | HEMOR | 17378 | 0.005653 | 0.010281 | 0.011121 | 0.30563 | Stomach |
| Brain_Substantia_nigra | LEWY_grp | 17385 | 0.004268 | 0.007354 | 0.008403 | 0.30576 | Brain_Substantia_nigra |
| Skin_Not_Sun_Exposed_Suprapu... | TDP_3 | 17388 | 0.003782 | 0.007302 | 0.007493 | 0.30685 | Skin_Not_Sun_Exposed_Suprapubic |
| Prostate | CVD_ART | 17388 | 0.005895 | 0.011198 | 0.011692 | 0.30706 | Prostate |
| Brain_Substantia_nigra | LEWY_ANY | 17385 | 0.004168 | 0.007182 | 0.008268 | 0.30708 | Brain_Substantia_nigra |

| | | | | | | | |
|---|---|---|---|---|---|---|---|
| Pancreas | CVD_ART | 17388 | 0.004089 | 0.006842 | 0.008124 | 0.30737 | Pancreas |
| Breast_Mammary_Tissue | TDP43 | 17386 | 0.00613 | 0.011802 | 0.012179 | 0.30737 | Breast_Mammary_Tissue |
| Cervix_Ectocervix | INFA | 17386 | 0.00566 | 0.011105 | 0.011247 | 0.3074 | Cervix_Ectocervix |
| Esophagus_Gastroesophageal_J... | CVD_ATH | 17388 | 0.005988 | 0.011798 | 0.011913 | 0.30761 | Esophagus_Gastroesophageal_Junction |
| Brain_Cerebellar_Hemisphere | AMY_THAL | 17388 | 0.002914 | 0.005852 | 0.005798 | 0.30763 | Brain_Cerebellar_Hemisphere |
| Cells_EBV-transformed_lympho... | WBVD | 17388 | 0.002307 | 0.005034 | 0.004595 | 0.30784 | Cells_EBV-transformed_lymphocytes |
| Spleen | AMY_THAL | 17388 | 0.00353 | 0.007001 | 0.00706 | 0.30857 | Spleen |
| Fallopian_Tube | CVD_CAA_ANY | 17386 | 0.005645 | 0.010964 | 0.011302 | 0.30873 | Fallopian_Tube |
| Whole_Blood | CVD_ATH | 17388 | 0.002682 | 0.004848 | 0.005379 | 0.30903 | Whole_Blood |
| Skin_Sun_Exposed_Lower_leg | TDP_3 | 17388 | 0.003728 | 0.007241 | 0.007503 | 0.30966 | Skin_Sun_Exposed_Lower_leg |
| Brain_Cerebellar_Hemisphere | LEWY_grp | 17385 | 0.002941 | 0.005907 | 0.005925 | 0.30983 | Brain_Cerebellar_Hemisphere |
| Brain_Cortex | HS_S | 17386 | 0.003317 | 0.006055 | 0.006795 | 0.31272 | Brain_Cortex |
| Colon_Transverse | HEMOR | 17378 | 0.005292 | 0.009742 | 0.010866 | 0.31311 | Colon_Transverse |
| Esophagus_Muscularis | BRAA | 17385 | 0.005528 | 0.010913 | 0.011601 | 0.31688 | Esophagus_Muscularis |
| Pituitary | CVD_S | 17387 | 0.004105 | 0.007662 | 0.00862 | 0.31696 | Pituitary |
| Breast_Mammary_Tissue | LEWY_full | 17385 | 0.005689 | 0.010953 | 0.011963 | 0.31719 | Breast_Mammary_Tissue |
| Artery_Tibial | CVD_CAA_ANY | 17386 | 0.004533 | 0.009339 | 0.009544 | 0.31742 | Artery_Tibial |
| Brain_Hypothalamus | CVD_S | 17387 | 0.003651 | 0.006292 | 0.007708 | 0.31785 | Brain_Hypothalamus |
| Colon_Transverse | ADNC | 17388 | 0.005217 | 0.009602 | 0.011074 | 0.31879 | Colon_Transverse |
| Pancreas | WMRCAL | 17388 | 0.003758 | 0.006288 | 0.00799 | 0.31905 | Pancreas |
| Stomach | TDP43 | 17386 | 0.0055 | 0.010003 | 0.011809 | 0.32069 | Stomach |
| Brain_Cerebellar_Hemisphere | CVD_CAA | 17385 | 0.002688 | 0.005397 | 0.005788 | 0.32122 | Brain_Cerebellar_Hemisphere |
| Brain_Nucleus_accumbens_basa... | MICR | 17385 | 0.003346 | 0.005811 | 0.007217 | 0.32148 | Brain_Nucleus_accumbens_basal_ganglia |
| Ovary | CVD_ART | 17388 | 0.004314 | 0.008753 | 0.009312 | 0.32156 | Ovary |
| Artery_Coronary | CVD_ATH_ANY | 17386 | 0.005158 | 0.010278 | 0.011158 | 0.32194 | Artery_Coronary |
| Brain_Amygdala | CVD_CAA | 17385 | 0.003469 | 0.005915 | 0.007531 | 0.32255 | Brain_Amygdala |
| Colon_Transverse | AMY_CERAD | 17386 | 0.005091 | 0.009371 | 0.011087 | 0.32304 | Colon_Transverse |
| Muscle_Skeletal | WMRCAL | 17388 | 0.003004 | 0.005684 | 0.006568 | 0.32369 | Muscle_Skeletal |
| Pituitary | CVD_ART | 17388 | 0.003972 | 0.007415 | 0.00871 | 0.32417 | Pituitary |
| Brain_Cerebellum | CVD_CAA_ANY | 17386 | 0.002729 | 0.005423 | 0.006027 | 0.32533 | Brain_Cerebellum |
| Kidney_Cortex | B_SCORE | 17388 | 0.003781 | 0.006651 | 0.008352 | 0.32537 | Kidney_Cortex |
| Stomach | CVD_ATH | 17388 | 0.005135 | 0.009339 | 0.011369 | 0.32576 | Stomach |
| Adrenal_Gland | LEWY_grp | 17385 | 0.004334 | 0.008374 | 0.009608 | 0.32597 | Adrenal_Gland |
| Ovary | HEMOR | 17378 | 0.004082 | 0.008283 | 0.009122 | 0.32726 | Ovary |
| Brain_Substantia_nigra | MICR | 17385 | 0.003602 | 0.006207 | 0.008067 | 0.32763 | Brain_Substantia_nigra |
| Esophagus_Mucosa | AMY_A | 17388 | 0.003315 | 0.006415 | 0.007449 | 0.32814 | Esophagus_Mucosa |

| | | | | | | | |
|---|---|---|---|---|---|---|---|
| Brain_Cerebellar_Hemisphere | CVD_CAA_ANY | 17386 | 0.002573 | 0.005168 | 0.005827 | 0.32938 | Brain_Cerebellar_Hemisphere |
| Adrenal_Gland | CVD_CAA_ANY | 17386 | 0.004158 | 0.008033 | 0.009423 | 0.32954 | Adrenal_Gland |
| Skin_Sun_Exposed_Lower_leg | MICR | 17385 | 0.003238 | 0.00629 | 0.007381 | 0.33045 | Skin_Sun_Exposed_Lower_leg |
| Artery_Coronary | BRAA | 17385 | 0.004928 | 0.009819 | 0.011285 | 0.33118 | Artery_Coronary |
| Artery_Tibial | CVD_CAA | 17385 | 0.004124 | 0.008497 | 0.009448 | 0.33124 | Artery_Tibial |
| Adrenal_Gland | CVD_ATH | 17388 | 0.004066 | 0.007855 | 0.009314 | 0.33125 | Adrenal_Gland |
| Skin_Sun_Exposed_Lower_leg | CVD_S | 17387 | 0.003157 | 0.006133 | 0.007348 | 0.33372 | Skin_Sun_Exposed_Lower_leg |
| Brain_Cerebellum | B_SCORE | 17388 | 0.002553 | 0.005073 | 0.005955 | 0.33406 | Brain_Cerebellum |
| Colon_Sigmoid | HEMOR | 17378 | 0.005062 | 0.009912 | 0.01181 | 0.33412 | Colon_Sigmoid |
| Brain_Hippocampus | WBVD | 17388 | 0.003226 | 0.005468 | 0.007553 | 0.33466 | Brain_Hippocampus |
| Breast_Mammary_Tissue | ADNC | 17388 | 0.00504 | 0.009703 | 0.011916 | 0.33615 | Breast_Mammary_Tissue |
| Kidney_Medulla | AMY_ANY | 17377 | 0.003776 | 0.006989 | 0.008926 | 0.33616 | Kidney_Medulla |
| Adipose_Visceral_Omentum | WBVD | 17388 | 0.004397 | 0.008549 | 0.010409 | 0.33636 | Adipose_Visceral_Omentum |
| Esophagus_Muscularis | TDP43 | 17386 | 0.005031 | 0.009933 | 0.011942 | 0.33678 | Esophagus_Muscularis |
| Adrenal_Gland | AMY_ANY | 17377 | 0.004018 | 0.007763 | 0.009579 | 0.33744 | Adrenal_Gland |
| Pancreas | CVD_ATH_ANY | 17386 | 0.003333 | 0.005577 | 0.008034 | 0.33914 | Pancreas |
| Brain_Hippocampus | CVD_CAA_ANY | 17386 | 0.003189 | 0.005406 | 0.007728 | 0.33995 | Brain_Hippocampus |
| Brain_Hypothalamus | AMY_ANY | 17377 | 0.003211 | 0.005533 | 0.00787 | 0.34165 | Brain_Hypothalamus |
| Bladder | B_SCORE | 17388 | 0.004929 | 0.009619 | 0.012212 | 0.34323 | Bladder |
| Brain_Hypothalamus | HS_S | 17386 | 0.003125 | 0.005386 | 0.007762 | 0.34361 | Brain_Hypothalamus |
| Kidney_Cortex | CVD_ART | 17388 | 0.003438 | 0.006046 | 0.008548 | 0.34379 | Kidney_Cortex |
| Artery_Tibial | CVD_ATH_ANY | 17386 | 0.003773 | 0.007774 | 0.009392 | 0.34393 | Artery_Tibial |
| Whole_Blood | AMY_ANY | 17377 | 0.00218 | 0.003941 | 0.00546 | 0.34486 | Whole_Blood |
| Brain_Substantia_nigra | AMY_ANY | 17377 | 0.003335 | 0.005746 | 0.008356 | 0.34494 | Brain_Substantia_nigra |
| Minor_Salivary_Gland | HEMOR | 17378 | 0.003589 | 0.006686 | 0.009038 | 0.34567 | Minor_Salivary_Gland |
| Uterus | TDP_3 | 17388 | 0.00415 | 0.008463 | 0.010454 | 0.3457 | Uterus |
| Cells_EBV-transformed_lympho... | WMRCAL | 17388 | 0.001802 | 0.003933 | 0.004546 | 0.34588 | Cells_EBV-transformed_lymphocytes |
| Liver | WMR | 17388 | 0.002418 | 0.004356 | 0.006104 | 0.34601 | Liver |
| Cells_EBV-transformed_lympho... | WMR | 17388 | 0.001837 | 0.004008 | 0.004655 | 0.34659 | Cells_EBV-transformed_lymphocytes |
| Small_Intestine_Terminal_Ile... | ADNC | 17388 | 0.003591 | 0.006612 | 0.009143 | 0.34723 | Small_Intestine_Terminal_Ileum |
| Uterus | TDP43 | 17386 | 0.004137 | 0.008437 | 0.010585 | 0.34796 | Uterus |
| Brain_Cerebellum | CVD_CAA | 17385 | 0.002332 | 0.004634 | 0.005974 | 0.34812 | Brain_Cerebellum |
| Vagina | WMR | 17388 | 0.004117 | 0.007882 | 0.010547 | 0.34814 | Vagina |
| Pituitary | WMRCAL | 17388 | 0.003285 | 0.006131 | 0.008459 | 0.34891 | Pituitary |
| Brain_Nucleus_accumbens_basa... | HS_S | 17386 | 0.002799 | 0.004862 | 0.007269 | 0.35009 | Brain_Nucleus_accumbens_basal_ganglia |
| Cervix_Endocervix | CVD_CAA | 17385 | 0.004278 | 0.008478 | 0.011116 | 0.35018 | Cervix_Endocervix |

| | | | | | | | |
|---|---|---|---|---|---|---|---|
| Breast_Mammary_Tissue | AMY_A | 17388 | 0.004557 | 0.008773 | 0.011927 | 0.3512 | Breast_Mammary_Tissue |
| Lung | CVD_CAA | 17385 | 0.003318 | 0.006423 | 0.008695 | 0.35138 | Lung |
| Esophagus_Muscularis | CVD_ATH | 17388 | 0.004385 | 0.008657 | 0.011554 | 0.35216 | Esophagus_Muscularis |
| Breast_Mammary_Tissue | WBVD | 17388 | 0.004476 | 0.008616 | 0.011831 | 0.35261 | Breast_Mammary_Tissue |
| Brain_Frontal_Cortex_BA9 | HS_S | 17386 | 0.002471 | 0.004557 | 0.00657 | 0.35345 | Brain_Frontal_Cortex_BA9 |
| Cervix_Ectocervix | CVD_S | 17387 | 0.004356 | 0.008546 | 0.011625 | 0.35395 | Cervix_Ectocervix |
| Artery_Tibial | HEMOR | 17378 | 0.003546 | 0.007305 | 0.009463 | 0.35395 | Artery_Tibial |
| Brain_Amygdala | HS_S | 17386 | 0.002805 | 0.004784 | 0.007527 | 0.35468 | Brain_Amygdala |
| Skin_Not_Sun_Exposed_Suprapu... | LEWY_ANY | 17385 | 0.002786 | 0.005378 | 0.007593 | 0.35685 | Skin_Not_Sun_Exposed_Suprapubic |
| Whole_Blood | MICR | 17385 | 0.001966 | 0.003554 | 0.005359 | 0.35686 | Whole_Blood |
| Cervix_Endocervix | CVD_S | 17387 | 0.004053 | 0.008032 | 0.011064 | 0.35707 | Cervix_Endocervix |
| Colon_Sigmoid | AMY_A | 17388 | 0.004349 | 0.008517 | 0.011903 | 0.35741 | Colon_Sigmoid |
| Artery_Tibial | AMY_A | 17388 | 0.003443 | 0.007093 | 0.00945 | 0.35783 | Artery_Tibial |
| Cervix_Ectocervix | TDP_3 | 17388 | 0.004214 | 0.008268 | 0.011667 | 0.35898 | Cervix_Ectocervix |
| Bladder | CVD_ATH_ANY | 17386 | 0.00447 | 0.008723 | 0.012437 | 0.35963 | Bladder |
| Esophagus_Muscularis | MICR | 17385 | 0.004107 | 0.008109 | 0.01161 | 0.36176 | Esophagus_Muscularis |
| Lung | WBVD | 17388 | 0.003068 | 0.005938 | 0.008673 | 0.36178 | Lung |
| Prostate | CVD_CAA | 17385 | 0.004038 | 0.00767 | 0.011524 | 0.36303 | Prostate |
| Skin_Not_Sun_Exposed_Suprapu... | MICR | 17385 | 0.002552 | 0.004926 | 0.007363 | 0.36446 | Skin_Not_Sun_Exposed_Suprapubic |
| Cervix_Endocervix | AMY_CERAD | 17386 | 0.003889 | 0.007707 | 0.011255 | 0.36487 | Cervix_Endocervix |
| Fallopian_Tube | MICR | 17385 | 0.003869 | 0.007515 | 0.011212 | 0.36501 | Fallopian_Tube |
| Skin_Sun_Exposed_Lower_leg | HEMOR | 17378 | 0.00254 | 0.004934 | 0.007363 | 0.36505 | Skin_Sun_Exposed_Lower_leg |
| Esophagus_Mucosa | CVD_ATH | 17388 | 0.00253 | 0.004895 | 0.00735 | 0.36535 | Esophagus_Mucosa |
| Colon_Sigmoid | BRAA | 17385 | 0.004012 | 0.007856 | 0.011796 | 0.36689 | Colon_Sigmoid |
| Artery_Aorta | CVD_ATH_ANY | 17386 | 0.003259 | 0.006657 | 0.00962 | 0.36738 | Artery_Aorta |
| Brain_Hippocampus | CVD_CAA | 17385 | 0.002573 | 0.004361 | 0.007603 | 0.36754 | Brain_Hippocampus |
| Small_Intestine_Terminal_Ile... | CVD_CAA | 17385 | 0.003035 | 0.005588 | 0.008993 | 0.36787 | Small_Intestine_Terminal_Ileum |
| Breast_Mammary_Tissue | TDP_3 | 17388 | 0.004021 | 0.00774 | 0.011929 | 0.36804 | Breast_Mammary_Tissue |
| Cervix_Endocervix | LEWY_grp | 17385 | 0.003817 | 0.007565 | 0.011325 | 0.36805 | Cervix_Endocervix |
| Thyroid | CVD_ATH | 17388 | 0.003109 | 0.006163 | 0.009349 | 0.36974 | Thyroid |
| Uterus | AMY_CERAD | 17386 | 0.003512 | 0.007162 | 0.010635 | 0.37062 | Uterus |
| Adipose_Subcutaneous | CVD_ART_ANY | 17387 | 0.003343 | 0.006678 | 0.010147 | 0.3709 | Adipose_Subcutaneous |
| Skin_Not_Sun_Exposed_Suprapu... | CVD_ART | 17388 | 0.00247 | 0.004769 | 0.007503 | 0.37099 | Skin_Not_Sun_Exposed_Suprapubic |
| Adrenal_Gland | CVD_ART_ANY | 17387 | 0.003048 | 0.005888 | 0.009339 | 0.3721 | Adrenal_Gland |
| Brain_Cerebellar_Hemisphere | B_SCORE | 17388 | 0.001872 | 0.00376 | 0.005759 | 0.37257 | Brain_Cerebellar_Hemisphere |
| Artery_Tibial | CVD_ART_ANY | 17387 | 0.003084 | 0.006353 | 0.009531 | 0.37315 | Artery_Tibial |

| | | | | | | | |
|---|---|---|---|---|---|---|---|
| Nerve_Tibial | AMY_CERAD | 17386 | 0.003134 | 0.006316 | 0.009736 | 0.37375 | Nerve_Tibial |
| Whole_Blood | LEWY_ANY | 17385 | 0.001759 | 0.00318 | 0.005536 | 0.37533 | Whole_Blood |
| Ovary | TDP43 | 17386 | 0.00296 | 0.006005 | 0.009449 | 0.37706 | Ovary |
| Cells_EBV-transformed_lympho... | AMY_THAL | 17388 | 0.001452 | 0.003169 | 0.00465 | 0.37742 | Cells_EBV-transformed_lymphocytes |
| Cervix_Ectocervix | HS_S | 17386 | 0.003599 | 0.007061 | 0.011541 | 0.37758 | Cervix_Ectocervix |
| Brain_Hippocampus | HS_S | 17386 | 0.002334 | 0.003956 | 0.007623 | 0.37976 | Brain_Hippocampus |
| Heart_Atrial_Appendage | LEWY_grp | 17385 | 0.002869 | 0.005155 | 0.009381 | 0.37986 | Heart_Atrial_Appendage |
| Thyroid | CVD_CAA_ANY | 17386 | 0.002841 | 0.005632 | 0.009371 | 0.38089 | Thyroid |
| Muscle_Skeletal | CVD_ATH | 17388 | 0.001979 | 0.003745 | 0.006575 | 0.38169 | Muscle_Skeletal |
| Artery_Coronary | LEWY_grp | 17385 | 0.003487 | 0.006947 | 0.011609 | 0.38196 | Artery_Coronary |
| Cells_EBV-transformed_lympho... | ADNC | 17388 | 0.001383 | 0.003018 | 0.004626 | 0.3825 | Cells_EBV-transformed_lymphocytes |
| Kidney_Medulla | HEMOR | 17378 | 0.002574 | 0.004765 | 0.008639 | 0.38289 | Kidney_Medulla |
| Small_Intestine_Terminal_Ile... | B_SCORE | 17388 | 0.002693 | 0.004958 | 0.009145 | 0.38419 | Small_Intestine_Terminal_Ileum |
| Cells_EBV-transformed_lympho... | AMY_A | 17388 | 0.00136 | 0.002968 | 0.004623 | 0.38431 | Cells_EBV-transformed_lymphocytes |
| Cervix_Ectocervix | LEWY_grp | 17385 | 0.003438 | 0.006745 | 0.0119 | 0.38633 | Cervix_Ectocervix |
| Adipose_Visceral_Omentum | CVD_ART_ANY | 17387 | 0.003006 | 0.005843 | 0.01045 | 0.38682 | Adipose_Visceral_Omentum |
| Ovary | AMY_THAL | 17388 | 0.002638 | 0.005351 | 0.009203 | 0.38721 | Ovary |
| Cervix_Ectocervix | CVD_CAA | 17385 | 0.003278 | 0.006432 | 0.011609 | 0.38883 | Cervix_Ectocervix |
| Adipose_Visceral_Omentum | CVD_S | 17387 | 0.002947 | 0.00573 | 0.010438 | 0.38883 | Adipose_Visceral_Omentum |
| Cervix_Ectocervix | AMY_ANY | 17377 | 0.003264 | 0.006403 | 0.011749 | 0.39059 | Cervix_Ectocervix |
| Small_Intestine_Terminal_Ile... | TDP_3 | 17388 | 0.002549 | 0.004692 | 0.009268 | 0.39167 | Small_Intestine_Terminal_Ileum |
| Heart_Left_Ventricle | CVD_ATH | 17388 | 0.002373 | 0.003964 | 0.008662 | 0.39205 | Heart_Left_Ventricle |
| Whole_Blood | INFA | 17386 | 0.001443 | 0.002609 | 0.005291 | 0.3925 | Whole_Blood |
| Uterus | CVD_S | 17387 | 0.002823 | 0.005758 | 0.010372 | 0.39273 | Uterus |
| Prostate | CVD_ATH | 17388 | 0.003068 | 0.005828 | 0.011467 | 0.39452 | Prostate |
| Bladder | TDP43 | 17386 | 0.003388 | 0.006612 | 0.012681 | 0.39466 | Bladder |
| Cervix_Ectocervix | BRAA | 17385 | 0.003082 | 0.006046 | 0.011646 | 0.39566 | Cervix_Ectocervix |
| Kidney_Cortex | CVD_CAA | 17385 | 0.002187 | 0.003846 | 0.008326 | 0.39642 | Kidney_Cortex |
| Brain_Putamen_basal_ganglia | MICR | 17385 | 0.001968 | 0.003352 | 0.007515 | 0.39671 | Brain_Putamen_basal_ganglia |
| Cervix_Endocervix | WMRCAL | 17388 | 0.002845 | 0.005638 | 0.010989 | 0.39786 | Cervix_Endocervix |
| Thyroid | WMRCAL | 17388 | 0.002361 | 0.004681 | 0.009148 | 0.39816 | Thyroid |
| Ovary | CVD_ART_ANY | 17387 | 0.002327 | 0.004721 | 0.009209 | 0.40026 | Ovary |
| Muscle_Skeletal | WBVD | 17388 | 0.001659 | 0.00314 | 0.006625 | 0.40112 | Muscle_Skeletal |
| Testis | AMY_THAL | 17388 | 0.001357 | 0.002345 | 0.005421 | 0.40121 | Testis |
| Brain_Hypothalamus | AMY_THAL | 17388 | 0.001922 | 0.003311 | 0.007762 | 0.40224 | Brain_Hypothalamus |
| Skin_Sun_Exposed_Lower_leg | CVD_ART | 17388 | 0.00186 | 0.003613 | 0.007514 | 0.40224 | Skin_Sun_Exposed_Lower_leg |

| | | | | | | | |
|---|---|---|---|---|---|---|---|
| Minor_Salivary_Gland | TDP43 | 17386 | 0.002329 | 0.004338 | 0.009467 | 0.40286 | Minor_Salivary_Gland |
| Skin_Not_Sun_Exposed_Suprapu... | HEMOR | 17378 | 0.001786 | 0.003448 | 0.00735 | 0.40402 | Skin_Not_Sun_Exposed_Suprapubic |
| Fallopian_Tube | BRAA | 17385 | 0.002753 | 0.005348 | 0.011387 | 0.40446 | Fallopian_Tube |
| Adipose_Visceral_Omentum | B_SCORE | 17388 | 0.002474 | 0.00481 | 0.010506 | 0.40692 | Adipose_Visceral_Omentum |
| Minor_Salivary_Gland | CVD_ART_ANY | 17387 | 0.002137 | 0.00398 | 0.009088 | 0.40707 | Minor_Salivary_Gland |
| Artery_Coronary | MICR | 17385 | 0.002568 | 0.005117 | 0.011124 | 0.40871 | Artery_Coronary |
| Brain_Amygdala | MICR | 17385 | 0.001713 | 0.002921 | 0.007437 | 0.4089 | Brain_Amygdala |
| Whole_Blood | CVD_ART_ANY | 17387 | 0.001251 | 0.002261 | 0.005434 | 0.40897 | Whole_Blood |
| Cervix_Endocervix | LEWY_ANY | 17385 | 0.002546 | 0.005046 | 0.011213 | 0.41019 | Cervix_Endocervix |
| Spleen | TDP_3 | 17388 | 0.001607 | 0.003188 | 0.007126 | 0.41078 | Spleen |
| Esophagus_Mucosa | INFA | 17386 | 0.001603 | 0.003101 | 0.00712 | 0.41096 | Esophagus_Mucosa |
| Artery_Tibial | CVD_S | 17387 | 0.00213 | 0.004388 | 0.009495 | 0.41125 | Artery_Tibial |
| Brain_Cerebellum | MICR | 17385 | 0.001309 | 0.0026 | 0.005913 | 0.41242 | Brain_Cerebellum |
| Adipose_Visceral_Omentum | BRAA | 17385 | 0.002305 | 0.004482 | 0.010526 | 0.41333 | Adipose_Visceral_Omentum |
| Small_Intestine_Terminal_Ile... | HS_S | 17386 | 0.001974 | 0.003634 | 0.009049 | 0.41367 | Small_Intestine_Terminal_Ileum |
| Pancreas | HEMOR | 17378 | 0.001751 | 0.00293 | 0.008034 | 0.41375 | Pancreas |
| Ovary | MICR | 17385 | 0.001986 | 0.00403 | 0.009138 | 0.41396 | Ovary |
| Kidney_Cortex | AMY_ANY | 17377 | 0.001817 | 0.003196 | 0.008562 | 0.41596 | Kidney_Cortex |
| Artery_Aorta | LEWY_full | 17385 | 0.002069 | 0.004226 | 0.009839 | 0.41671 | Artery_Aorta |
| Minor_Salivary_Gland | WBVD | 17388 | 0.0019 | 0.00354 | 0.009102 | 0.41731 | Minor_Salivary_Gland |
| Colon_Sigmoid | CVD_ATH | 17388 | 0.002397 | 0.004694 | 0.011739 | 0.4191 | Colon_Sigmoid |
| Pancreas | TDP_3 | 17388 | 0.001666 | 0.002788 | 0.008222 | 0.41971 | Pancreas |
| Cervix_Ectocervix | WMRCAL | 17388 | 0.002308 | 0.004528 | 0.011602 | 0.42117 | Cervix_Ectocervix |
| Artery_Coronary | CVD_ART_ANY | 17387 | 0.002237 | 0.004457 | 0.011265 | 0.42129 | Artery_Coronary |
| Colon_Transverse | CVD_S | 17387 | 0.002126 | 0.003913 | 0.010898 | 0.42268 | Colon_Transverse |
| Pancreas | ADNC | 17388 | 0.001588 | 0.002656 | 0.008153 | 0.42281 | Pancreas |
| Brain_Cerebellar_Hemisphere | MICR | 17385 | 0.001108 | 0.002226 | 0.005713 | 0.4231 | Brain_Cerebellar_Hemisphere |
| Artery_Coronary | WMR | 17388 | 0.002183 | 0.00435 | 0.011295 | 0.42337 | Artery_Coronary |
| Artery_Coronary | LEWY_ANY | 17385 | 0.002162 | 0.004307 | 0.011394 | 0.42477 | Artery_Coronary |
| Liver | HS_S | 17386 | 0.001158 | 0.002086 | 0.006108 | 0.42483 | Liver |
| Nerve_Tibial | CVD_ART | 17388 | 0.001826 | 0.00368 | 0.009637 | 0.42484 | Nerve_Tibial |
| Fallopian_Tube | AMY_ANY | 17377 | 0.002157 | 0.004189 | 0.011435 | 0.4252 | Fallopian_Tube |
| Bladder | INFA | 17386 | 0.002267 | 0.004424 | 0.012044 | 0.42534 | Bladder |
| Adipose_Subcutaneous | WBVD | 17388 | 0.001896 | 0.003788 | 0.010106 | 0.42559 | Adipose_Subcutaneous |
| Artery_Coronary | AMY_CERAD | 17386 | 0.002116 | 0.004216 | 0.011381 | 0.42625 | Artery_Coronary |
| Esophagus_Mucosa | TDP43 | 17386 | 0.001419 | 0.002746 | 0.007643 | 0.42635 | Esophagus_Mucosa |

| | | | | | | | |
|---|---|---|---|---|---|---|---|
| Skin_Not_Sun_Exposed_Suprapu... | LEWY_full | 17385 | 0.001389 | 0.002681 | 0.007507 | 0.42662 | Skin_Not_Sun_Exposed_Suprapubic |
| Heart_Left_Ventricle | AMY_A | 17388 | 0.00161 | 0.00269 | 0.008761 | 0.42708 | Heart_Left_Ventricle |
| Nerve_Tibial | CVD_ART_ANY | 17387 | 0.001696 | 0.003418 | 0.009509 | 0.42921 | Nerve_Tibial |
| Artery_Coronary | AMY_A | 17388 | 0.001994 | 0.003972 | 0.011256 | 0.42971 | Artery_Coronary |
| Esophagus_Gastroesophageal_J... | BRAA | 17385 | 0.002081 | 0.004101 | 0.011988 | 0.43109 | Esophagus_Gastroesophageal_Junction |
| Brain_Substantia_nigra | CVD_S | 17387 | 0.001399 | 0.00241 | 0.00811 | 0.43154 | Brain_Substantia_nigra |
| Brain_Spinal_cord_cervical_c... | BRAA | 17385 | 0.001404 | 0.002526 | 0.008147 | 0.43158 | Brain_Spinal_cord_cervical_c-1 |
| Brain_Anterior_cingulate_cor... | HS_S | 17386 | 0.001185 | 0.002096 | 0.006923 | 0.43203 | Brain_Anterior_cingulate_cortex_BA24 |
| Vagina | CVD_ATH_ANY | 17386 | 0.001793 | 0.003433 | 0.010562 | 0.43259 | Vagina |
| Esophagus_Muscularis | CVD_ART_ANY | 17387 | 0.001942 | 0.003834 | 0.011684 | 0.43399 | Esophagus_Muscularis |
| Ovary | AMY_A | 17388 | 0.0015 | 0.003044 | 0.009243 | 0.43552 | Ovary |
| Vagina | INFA | 17386 | 0.001592 | 0.003047 | 0.010223 | 0.43814 | Vagina |
| Vagina | WMRCAL | 17388 | 0.001661 | 0.003179 | 0.010705 | 0.43836 | Vagina |
| Artery_Aorta | WMR | 17388 | 0.001496 | 0.003056 | 0.0097 | 0.4387 | Artery_Aorta |
| Cervix_Ectocervix | CVD_CAA_ANY | 17386 | 0.00178 | 0.003492 | 0.0117 | 0.43955 | Cervix_Ectocervix |
| Liver | LEWY_ANY | 17385 | 0.000949 | 0.001709 | 0.00626 | 0.43977 | Liver |
| Brain_Hippocampus | MICR | 17385 | 0.00113 | 0.001916 | 0.00755 | 0.44051 | Brain_Hippocampus |
| Cells_Cultured_fibroblasts | HS_S | 17386 | 0.000912 | 0.001956 | 0.006293 | 0.44237 | Cells_Cultured_fibroblasts |
| Adipose_Visceral_Omentum | AMY_THAL | 17388 | 0.001507 | 0.00293 | 0.010512 | 0.44301 | Adipose_Visceral_Omentum |
| Kidney_Medulla | CVD_CAA_ANY | 17386 | 0.001276 | 0.002362 | 0.009004 | 0.44367 | Kidney_Medulla |
| Fallopian_Tube | WMRCAL | 17388 | 0.001585 | 0.003079 | 0.011233 | 0.44389 | Fallopian_Tube |
| Artery_Tibial | WBVD | 17388 | 0.001323 | 0.002726 | 0.009399 | 0.44403 | Artery_Tibial |
| Adipose_Visceral_Omentum | MICR | 17385 | 0.001447 | 0.002812 | 0.010459 | 0.445 | Adipose_Visceral_Omentum |
| Heart_Atrial_Appendage | AMY_A | 17388 | 0.001248 | 0.002243 | 0.009055 | 0.44518 | Heart_Atrial_Appendage |
| Vagina | CVD_S | 17387 | 0.001457 | 0.002789 | 0.010638 | 0.44553 | Vagina |
| Cells_Cultured_fibroblasts | LEWY_ANY | 17385 | 0.000844 | 0.00181 | 0.006392 | 0.44748 | Cells_Cultured_fibroblasts |
| Brain_Cerebellar_Hemisphere | AMY_ANY | 17377 | 0.000756 | 0.001519 | 0.005846 | 0.44853 | Brain_Cerebellar_Hemisphere |
| Nerve_Tibial | ADNC | 17388 | 0.001223 | 0.002465 | 0.009604 | 0.44933 | Nerve_Tibial |
| Brain_Cerebellum | AMY_ANY | 17377 | 0.000766 | 0.001523 | 0.006053 | 0.44962 | Brain_Cerebellum |
| Liver | AMY_A | 17388 | 0.00076 | 0.00137 | 0.006146 | 0.45077 | Liver |
| Esophagus_Mucosa | HEMOR | 17378 | 0.0009 | 0.001741 | 0.007341 | 0.45122 | Esophagus_Mucosa |
| Prostate | HEMOR | 17378 | 0.001381 | 0.002623 | 0.011268 | 0.45123 | Prostate |
| Skin_Sun_Exposed_Lower_leg | LEWY_full | 17385 | 0.000908 | 0.001764 | 0.00751 | 0.45187 | Skin_Sun_Exposed_Lower_leg |
| Brain_Nucleus_accumbens_basa... | AMY_A | 17388 | 0.000866 | 0.001503 | 0.007325 | 0.45297 | Brain_Nucleus_accumbens_basal_ganglia |
| Muscle_Skeletal | CVD_ART_ANY | 17387 | 0.000766 | 0.00145 | 0.00657 | 0.45359 | Muscle_Skeletal |
| Artery_Coronary | B_SCORE | 17388 | 0.001292 | 0.002574 | 0.011237 | 0.45423 | Artery_Coronary |

| | | | | | | | |
|---|---|---|---|---|---|---|---|
| Cells_EBV-transformed_lympho... | TDP43 | 17386 | 0.000527 | 0.001149 | 0.004761 | 0.45596 | Cells_EBV-transformed_lymphocytes |
| Pancreas | CVD_CAA | 17385 | 0.000899 | 0.001505 | 0.008134 | 0.45599 | Pancreas |
| Kidney_Cortex | WMRCAL | 17388 | 0.000896 | 0.001576 | 0.008289 | 0.45697 | Kidney_Cortex |
| Brain_Substantia_nigra | HS_S | 17386 | 0.000883 | 0.001522 | 0.008175 | 0.45698 | Brain_Substantia_nigra |
| Small_Intestine_Terminal_Ile... | CVD_ATH | 17388 | 0.000979 | 0.001803 | 0.009096 | 0.45714 | Small_Intestine_Terminal_Ileum |
| Brain_Hypothalamus | MICR | 17385 | 0.00078 | 0.001344 | 0.007621 | 0.45926 | Brain_Hypothalamus |
| Cells_EBV-transformed_lympho... | MICR | 17385 | 0.00045 | 0.000982 | 0.004592 | 0.46097 | Cells_EBV-transformed_lymphocytes |
| Pancreas | CVD_CAA_ANY | 17386 | 0.000787 | 0.001317 | 0.008232 | 0.46193 | Pancreas |
| Cells_Cultured_fibroblasts | LEWY_full | 17385 | 0.000594 | 0.001274 | 0.00643 | 0.4632 | Cells_Cultured_fibroblasts |
| Cells_EBV-transformed_lympho... | CVD_ART | 17388 | 0.000421 | 0.000919 | 0.004629 | 0.46375 | Cells_EBV-transformed_lymphocytes |
| Esophagus_Gastroesophageal_J... | WMR | 17388 | 0.001085 | 0.002137 | 0.011932 | 0.46379 | Esophagus_Gastroesophageal_Junction |
| Uterus | WMR | 17388 | 0.000945 | 0.001927 | 0.010407 | 0.46382 | Uterus |
| Pituitary | AMY_A | 17388 | 0.000783 | 0.001461 | 0.008629 | 0.46386 | Pituitary |
| Prostate | TDP_3 | 17388 | 0.001005 | 0.001909 | 0.011539 | 0.46531 | Prostate |
| Brain_Caudate_basal_ganglia | AMY_A | 17388 | 0.000638 | 0.001104 | 0.007596 | 0.46652 | Brain_Caudate_basal_ganglia |
| Pituitary | WMR | 17388 | 0.000735 | 0.001371 | 0.00875 | 0.46655 | Pituitary |
| Artery_Coronary | CVD_CAA | 17385 | 0.000936 | 0.001866 | 0.011195 | 0.46667 | Artery_Coronary |
| Adrenal_Gland | HS_S | 17386 | 0.000781 | 0.001509 | 0.009411 | 0.46693 | Adrenal_Gland |
| Breast_Mammary_Tissue | CVD_ATH_ANY | 17386 | 0.000976 | 0.001879 | 0.011771 | 0.46695 | Breast_Mammary_Tissue |
| Adrenal_Gland | WBVD | 17388 | 0.000769 | 0.001486 | 0.009358 | 0.46724 | Adrenal_Gland |
| Pituitary | TDP_3 | 17388 | 0.000707 | 0.001319 | 0.008808 | 0.46803 | Pituitary |
| Nerve_Tibial | CVD_ATH_ANY | 17386 | 0.000743 | 0.001498 | 0.009354 | 0.46833 | Nerve_Tibial |
| Skin_Sun_Exposed_Lower_leg | LEWY_ANY | 17385 | 0.000597 | 0.00116 | 0.007609 | 0.46873 | Skin_Sun_Exposed_Lower_leg |
| Vagina | TDP_3 | 17388 | 0.000806 | 0.001543 | 0.010612 | 0.46974 | Vagina |
| Brain_Putamen_basal_ganglia | AMY_A | 17388 | 0.000572 | 0.000974 | 0.007695 | 0.47037 | Brain_Putamen_basal_ganglia |
| Brain_Cerebellar_Hemisphere | CVD_ATH | 17388 | 0.000425 | 0.000853 | 0.005753 | 0.47058 | Brain_Cerebellar_Hemisphere |
| Cervix_Endocervix | TDP_3 | 17388 | 0.000818 | 0.001622 | 0.011101 | 0.47062 | Cervix_Endocervix |
| Cervix_Ectocervix | ADNC | 17388 | 0.00081 | 0.00159 | 0.011692 | 0.47238 | Cervix_Ectocervix |
| Thyroid | HEMOR | 17378 | 0.000618 | 0.001226 | 0.009147 | 0.47306 | Thyroid |
| Colon_Sigmoid | LEWY_grp | 17385 | 0.000812 | 0.001591 | 0.012024 | 0.47307 | Colon_Sigmoid |
| Adrenal_Gland | LEWY_full | 17385 | 0.000645 | 0.001247 | 0.00957 | 0.47312 | Adrenal_Gland |
| Kidney_Cortex | ADNC | 17388 | 0.000573 | 0.001009 | 0.008592 | 0.47339 | Kidney_Cortex |
| Heart_Atrial_Appendage | TDP_3 | 17388 | 0.000569 | 0.001023 | 0.009127 | 0.47513 | Heart_Atrial_Appendage |
| Pancreas | WMR | 17388 | 0.000503 | 0.000841 | 0.008067 | 0.47515 | Pancreas |
| Breast_Mammary_Tissue | CVD_ART_ANY | 17387 | 0.000681 | 0.00131 | 0.011855 | 0.47711 | Breast_Mammary_Tissue |
| Fallopian_Tube | AMY_THAL | 17388 | 0.000644 | 0.001252 | 0.011387 | 0.47744 | Fallopian_Tube |

| | | | | | | | |
|---|---|---|---|---|---|---|---|
| Artery_Coronary | WBVD | 17388 | 0.000622 | 0.00124 | 0.011142 | 0.47774 | Artery_Coronary |
| Breast_Mammary_Tissue | LEWY_ANY | 17385 | 0.000658 | 0.001266 | 0.012072 | 0.47828 | Breast_Mammary_Tissue |
| Esophagus_Mucosa | WMRCAL | 17388 | 0.000401 | 0.000776 | 0.007376 | 0.47831 | Esophagus_Mucosa |
| Spleen | MICR | 17385 | 0.000315 | 0.000624 | 0.006895 | 0.48181 | Spleen |
| Skin_Not_Sun_Exposed_Suprapu... | LEWY_grp | 17385 | 0.000324 | 0.000625 | 0.007565 | 0.48293 | Skin_Not_Sun_Exposed_Suprapubic |
| Brain_Caudate_basal_ganglia | MICR | 17385 | 0.000315 | 0.000544 | 0.007448 | 0.48315 | Brain_Caudate_basal_ganglia |
| Bladder | CVD_CAA | 17385 | 0.000521 | 0.001016 | 0.012417 | 0.48327 | Bladder |
| Skin_Sun_Exposed_Lower_leg | CVD_CAA | 17385 | 0.000278 | 0.00054 | 0.007496 | 0.48521 | Skin_Sun_Exposed_Lower_leg |
| Cells_EBV-transformed_lympho... | TDP_3 | 17388 | 0.00017 | 0.000371 | 0.004689 | 0.48553 | Cells_EBV-transformed_lymphocytes |
| Pituitary | AMY_CERAD | 17386 | 0.000285 | 0.000532 | 0.008731 | 0.48698 | Pituitary |
| Kidney_Cortex | HEMOR | 17378 | 0.00027 | 0.000476 | 0.008406 | 0.48717 | Kidney_Cortex |
| Fallopian_Tube | CVD_CAA | 17385 | 0.000332 | 0.000645 | 0.011351 | 0.48833 | Fallopian_Tube |
| Whole_Blood | CVD_S | 17387 | 0.000123 | 0.000222 | 0.005465 | 0.49103 | Whole_Blood |
| Minor_Salivary_Gland | AMY_ANY | 17377 | 0.000203 | 0.000377 | 0.009095 | 0.49111 | Minor_Salivary_Gland |
| Heart_Atrial_Appendage | AMY_THAL | 17388 | 0.000193 | 0.000348 | 0.009037 | 0.49146 | Heart_Atrial_Appendage |
| Pancreas | AMY_CERAD | 17386 | 0.000122 | 0.000204 | 0.008313 | 0.49416 | Pancreas |
| Small_Intestine_Terminal_Ile... | CVD_ART | 17388 | 0.000131 | 0.000242 | 0.009255 | 0.49434 | Small_Intestine_Terminal_Ileum |
| Brain_Spinal_cord_cervical_c... | AMY_ANY | 17377 | 0.000112 | 0.000202 | 0.008312 | 0.49462 | Brain_Spinal_cord_cervical_c-1 |
| Spleen | HS_S | 17386 | 7.83E-05 | 0.000155 | 0.007086 | 0.49559 | Spleen |
| Cervix_Endocervix | AMY_ANY | 17377 | 0.000104 | 0.000206 | 0.011222 | 0.4963 | Cervix_Endocervix |
| Brain_Nucleus_accumbens_basa... | CVD_S | 17387 | 6.60E-05 | 0.000115 | 0.007271 | 0.49638 | Brain_Nucleus_accumbens_basal_ganglia |
| Small_Intestine_Terminal_Ile... | WMRCAL | 17388 | 5.19E-05 | 9.55E-05 | 0.009071 | 0.49772 | Small_Intestine_Terminal_Ileum |
| Minor_Salivary_Gland | CVD_ATH | 17388 | 5.05E-05 | 9.40E-05 | 0.009112 | 0.49779 | Minor_Salivary_Gland |
| Esophagus_Gastroesophageal_J... | CVD_ART_ANY | 17387 | -3.95E-06 | -7.78E-06 | 0.012067 | 0.50013 | Esophagus_Gastroesophageal_Junction |
| Kidney_Cortex | CVD_S | 17387 | -1.12E-05 | -1.98E-05 | 0.008439 | 0.50053 | Kidney_Cortex |
| Brain_Cerebellum | CVD_ATH | 17388 | -1.27E-05 | -2.51E-05 | 0.00596 | 0.50085 | Brain_Cerebellum |
| Ovary | CVD_CAA | 17385 | -4.57E-05 | -9.26E-05 | 0.009132 | 0.50199 | Ovary |
| Cells_Cultured_fibroblasts | CVD_ART | 17388 | -3.22E-05 | -6.90E-05 | 0.006311 | 0.50203 | Cells_Cultured_fibroblasts |
| Colon_Sigmoid | LEWY_ANY | 17385 | -6.78E-05 | -0.00013 | 0.012072 | 0.50224 | Colon_Sigmoid |
| Spleen | LEWY_ANY | 17385 | -5.05E-05 | -0.0001 | 0.007067 | 0.50285 | Spleen |
| Bladder | CVD_ART | 17388 | -0.00015 | -0.00029 | 0.012565 | 0.50468 | Bladder |
| Colon_Transverse | WMRCAL | 17388 | -0.00014 | -0.00025 | 0.011025 | 0.50489 | Colon_Transverse |
| Whole_Blood | LEWY_grp | 17385 | -7.42E-05 | -0.00013 | 0.00558 | 0.50531 | Whole_Blood |
| Skin_Not_Sun_Exposed_Suprapu... | CVD_CAA | 17385 | -0.00011 | -0.00022 | 0.007476 | 0.50602 | Skin_Not_Sun_Exposed_Suprapubic |
| Uterus | CVD_ART | 17388 | -0.00018 | -0.00038 | 0.01057 | 0.50697 | Uterus |
| Cells_EBV-transformed_lympho... | AMY_ANY | 17377 | -8.81E-05 | -0.00019 | 0.004734 | 0.50743 | Cells_EBV-transformed_lymphocytes |

| | | | | | | | |
|---|---|---|---|---|---|---|---|
| Lung | HS_S | 17386 | -0.00019 | -0.00037 | 0.008689 | 0.50871 | Lung |
| Esophagus_Muscularis | WMRCAL | 17388 | -0.00039 | -0.00076 | 0.011505 | 0.51342 | Esophagus_Muscularis |
| Brain_Cerebellar_Hemisphere | TDP43 | 17386 | -0.00021 | -0.00042 | 0.005969 | 0.51395 | Brain_Cerebellar_Hemisphere |
| Adipose_Subcutaneous | HEMOR | 17378 | -0.00037 | -0.00074 | 0.010184 | 0.51445 | Adipose_Subcutaneous |
| Vagina | LEWY_full | 17385 | -0.00044 | -0.00084 | 0.010732 | 0.51639 | Vagina |
| Stomach | CVD_CAA_ANY | 17386 | -0.00049 | -0.0009 | 0.011522 | 0.51704 | Stomach |
| Kidney_Medulla | AMY_CERAD | 17386 | -0.0004 | -0.00074 | 0.008962 | 0.51788 | Kidney_Medulla |
| Stomach | CVD_S | 17387 | -0.00054 | -0.00098 | 0.01125 | 0.51902 | Stomach |
| Lung | CVD_ART_ANY | 17387 | -0.00042 | -0.00081 | 0.008661 | 0.51935 | Lung |
| Artery_Coronary | HEMOR | 17378 | -0.00056 | -0.00112 | 0.011211 | 0.52008 | Artery_Coronary |
| Heart_Atrial_Appendage | LEWY_full | 17385 | -0.0005 | -0.0009 | 0.009216 | 0.52162 | Heart_Atrial_Appendage |
| Brain_Spinal_cord_cervical_c... | LEWY_ANY | 17385 | -0.00052 | -0.00093 | 0.008247 | 0.52511 | Brain_Spinal_cord_cervical_c-1 |
| Kidney_Medulla | B_SCORE | 17388 | -0.00058 | -0.00107 | 0.008727 | 0.52632 | Kidney_Medulla |
| Brain_Substantia_nigra | WBVD | 17388 | -0.00056 | -0.00096 | 0.008048 | 0.52751 | Brain_Substantia_nigra |
| Brain_Substantia_nigra | B_SCORE | 17388 | -0.00057 | -0.00097 | 0.008104 | 0.52782 | Brain_Substantia_nigra |
| Stomach | CVD_CAA | 17385 | -0.0008 | -0.00145 | 0.011082 | 0.52868 | Stomach |
| Cells_Cultured_fibroblasts | WBVD | 17388 | -0.00045 | -0.00097 | 0.006218 | 0.52915 | Cells_Cultured_fibroblasts |
| Thyroid | MICR | 17385 | -0.00068 | -0.00136 | 0.009215 | 0.52956 | Thyroid |
| Testis | LEWY_ANY | 17385 | -0.00041 | -0.00071 | 0.005437 | 0.53032 | Testis |
| Lung | AMY_THAL | 17388 | -0.00067 | -0.00129 | 0.008747 | 0.53046 | Lung |
| Stomach | MICR | 17385 | -0.00088 | -0.00159 | 0.011287 | 0.53096 | Stomach |
| Cells_EBV-transformed_lympho... | INFA | 17386 | -0.00035 | -0.00077 | 0.004527 | 0.53104 | Cells_EBV-transformed_lymphocytes |
| Cells_Cultured_fibroblasts | LEWY_grp | 17385 | -0.00051 | -0.00109 | 0.006466 | 0.53143 | Cells_Cultured_fibroblasts |
| Brain_Putamen_basal_ganglia | WMRCAL | 17388 | -0.00064 | -0.00108 | 0.007584 | 0.53337 | Brain_Putamen_basal_ganglia |
| Vagina | LEWY_ANY | 17385 | -0.00091 | -0.00175 | 0.010864 | 0.53355 | Vagina |
| Ovary | TDP_3 | 17388 | -0.00079 | -0.00161 | 0.009345 | 0.53387 | Ovary |
| Kidney_Medulla | CVD_S | 17387 | -0.00074 | -0.00137 | 0.008676 | 0.53398 | Kidney_Medulla |
| Vagina | CVD_ART_ANY | 17387 | -0.00091 | -0.00174 | 0.010649 | 0.53404 | Vagina |
| Esophagus_Gastroesophageal_J... | INFA | 17386 | -0.00105 | -0.00206 | 0.011719 | 0.5356 | Esophagus_Gastroesophageal_Junction |
| Bladder | AMY_ANY | 17377 | -0.00114 | -0.00222 | 0.012682 | 0.53579 | Bladder |
| Colon_Sigmoid | LEWY_full | 17385 | -0.00109 | -0.00213 | 0.011919 | 0.53638 | Colon_Sigmoid |
| Cervix_Ectocervix | AMY_CERAD | 17386 | -0.00108 | -0.00211 | 0.011781 | 0.5364 | Cervix_Ectocervix |
| Prostate | TDP43 | 17386 | -0.0011 | -0.00209 | 0.011771 | 0.53722 | Prostate |
| Cervix_Endocervix | BRAA | 17385 | -0.00105 | -0.00209 | 0.011102 | 0.53777 | Cervix_Endocervix |
| Brain_Cerebellum | LEWY_ANY | 17385 | -0.00059 | -0.00116 | 0.006039 | 0.53864 | Brain_Cerebellum |
| Esophagus_Gastroesophageal_J... | WMRCAL | 17388 | -0.00122 | -0.0024 | 0.011922 | 0.54074 | Esophagus_Gastroesophageal_Junction |

| | | | | | | | |
|---|---|---|---|---|---|---|---|
| Artery_Coronary | CVD_S | 17387 | -0.00115 | -0.0023 | 0.01121 | 0.54102 | Artery_Coronary |
| Esophagus_Mucosa | WBVD | 17388 | -0.00076 | -0.00148 | 0.007392 | 0.54113 | Esophagus_Mucosa |
| Cells_EBV-transformed_lympho... | HEMOR | 17378 | -0.00048 | -0.00105 | 0.0046 | 0.54152 | Cells_EBV-transformed_lymphocytes |
| Colon_Sigmoid | HS_S | 17386 | -0.00125 | -0.00245 | 0.011909 | 0.54191 | Colon_Sigmoid |
| Skin_Sun_Exposed_Lower_leg | LEWY_grp | 17385 | -0.0008 | -0.00156 | 0.007575 | 0.54215 | Skin_Sun_Exposed_Lower_leg |
| Spleen | LEWY_grp | 17385 | -0.00077 | -0.00153 | 0.007253 | 0.54239 | Spleen |
| Pituitary | WBVD | 17388 | -0.00093 | -0.00174 | 0.008612 | 0.543 | Pituitary |
| Cells_EBV-transformed_lympho... | AMY_CERAD | 17386 | -0.00052 | -0.00113 | 0.004666 | 0.54415 | Cells_EBV-transformed_lymphocytes |
| Breast_Mammary_Tissue | LEWY_grp | 17385 | -0.00138 | -0.00265 | 0.012229 | 0.54477 | Breast_Mammary_Tissue |
| Spleen | INFA | 17386 | -0.00077 | -0.00152 | 0.006811 | 0.54478 | Spleen |
| Brain_Hypothalamus | WBVD | 17388 | -0.00087 | -0.00151 | 0.007679 | 0.54535 | Brain_Hypothalamus |
| Esophagus_Gastroesophageal_J... | B_SCORE | 17388 | -0.0014 | -0.00275 | 0.012007 | 0.54626 | Esophagus_Gastroesophageal_Junction |
| Brain_Amygdala | AMY_A | 17388 | -0.00091 | -0.00155 | 0.00763 | 0.54744 | Brain_Amygdala |
| Brain_Nucleus_accumbens_basa... | WMRCAL | 17388 | -0.00087 | -0.00151 | 0.007265 | 0.54758 | Brain_Nucleus_accumbens_basal_ganglia |
| Small_Intestine_Terminal_Ile... | MICR | 17385 | -0.00108 | -0.00199 | 0.008912 | 0.54829 | Small_Intestine_Terminal_Ileum |
| Colon_Transverse | INFA | 17386 | -0.00135 | -0.00248 | 0.010655 | 0.55031 | Colon_Transverse |
| Esophagus_Muscularis | B_SCORE | 17388 | -0.00148 | -0.00292 | 0.011632 | 0.5505 | Esophagus_Muscularis |
| Brain_Hypothalamus | CVD_CAA_ANY | 17386 | -0.00101 | -0.00173 | 0.007836 | 0.5511 | Brain_Hypothalamus |
| Skin_Not_Sun_Exposed_Suprapu... | CVD_CAA_ANY | 17386 | -0.00097 | -0.00187 | 0.007461 | 0.55152 | Skin_Not_Sun_Exposed_Suprapubic |
| Fallopian_Tube | LEWY_full | 17385 | -0.00148 | -0.00288 | 0.011288 | 0.55231 | Fallopian_Tube |
| Cervix_Endocervix | ADNC | 17388 | -0.00152 | -0.00302 | 0.011179 | 0.55422 | Cervix_Endocervix |
| Uterus | CVD_ART_ANY | 17387 | -0.00143 | -0.00292 | 0.01044 | 0.55455 | Uterus |
| Brain_Hypothalamus | CVD_ATH_ANY | 17386 | -0.00105 | -0.00181 | 0.007635 | 0.55461 | Brain_Hypothalamus |
| Adipose_Subcutaneous | LEWY_full | 17385 | -0.00142 | -0.00283 | 0.010294 | 0.55473 | Adipose_Subcutaneous |
| Adipose_Visceral_Omentum | WMRCAL | 17388 | -0.00143 | -0.00279 | 0.01042 | 0.55474 | Adipose_Visceral_Omentum |
| Testis | HEMOR | 17378 | -0.00074 | -0.00128 | 0.005323 | 0.55518 | Testis |
| Colon_Sigmoid | B_SCORE | 17388 | -0.00164 | -0.00322 | 0.011775 | 0.55549 | Colon_Sigmoid |
| Testis | AMY_CERAD | 17386 | -0.00077 | -0.00132 | 0.005477 | 0.55563 | Testis |
| Adrenal_Gland | WMRCAL | 17388 | -0.00132 | -0.00254 | 0.009336 | 0.55602 | Adrenal_Gland |
| Bladder | ADNC | 17388 | -0.00179 | -0.00348 | 0.012426 | 0.55714 | Bladder |
| Thyroid | AMY_THAL | 17388 | -0.00136 | -0.0027 | 0.009154 | 0.55906 | Thyroid |
| Colon_Transverse | CVD_ATH_ANY | 17386 | -0.00165 | -0.00303 | 0.011035 | 0.55936 | Colon_Transverse |
| Brain_Caudate_basal_ganglia | WMRCAL | 17388 | -0.00114 | -0.00196 | 0.007518 | 0.56003 | Brain_Caudate_basal_ganglia |
| Pituitary | AMY_THAL | 17388 | -0.00132 | -0.00247 | 0.008613 | 0.56102 | Pituitary |
| Brain_Cerebellar_Hemisphere | LEWY_ANY | 17385 | -0.00091 | -0.00184 | 0.005839 | 0.56223 | Brain_Cerebellar_Hemisphere |
| Kidney_Cortex | INFA | 17386 | -0.00131 | -0.00231 | 0.008173 | 0.56383 | Kidney_Cortex |

| | | | | | | | |
|---|---|---|---|---|---|---|---|
| Heart_Left_Ventricle | AMY_THAL | 17388 | -0.00142 | -0.00236 | 0.008771 | 0.5641 | Heart_Left_Ventricle |
| Adipose_Subcutaneous | HS_S | 17386 | -0.00164 | -0.00328 | 0.010124 | 0.56448 | Adipose_Subcutaneous |
| Small_Intestine_Terminal_Ile... | INFA | 17386 | -0.00146 | -0.00268 | 0.008821 | 0.56554 | Small_Intestine_Terminal_Ileum |
| Testis | LEWY_full | 17385 | -0.00089 | -0.00154 | 0.005372 | 0.56587 | Testis |
| Artery_Aorta | LEWY_ANY | 17385 | -0.00164 | -0.00335 | 0.009816 | 0.56636 | Artery_Aorta |
| Brain_Substantia_nigra | AMY_THAL | 17388 | -0.0014 | -0.00242 | 0.008169 | 0.56816 | Brain_Substantia_nigra |
| Cells_Cultured_fibroblasts | WMRCAL | 17388 | -0.0011 | -0.00235 | 0.006211 | 0.57007 | Cells_Cultured_fibroblasts |
| Artery_Coronary | AMY_THAL | 17388 | -0.002 | -0.00399 | 0.011313 | 0.5703 | Artery_Coronary |
| Cells_Cultured_fibroblasts | MICR | 17385 | -0.00112 | -0.0024 | 0.00621 | 0.57158 | Cells_Cultured_fibroblasts |
| Heart_Atrial_Appendage | AMY_CERAD | 17386 | -0.00169 | -0.00303 | 0.00927 | 0.57222 | Heart_Atrial_Appendage |
| Artery_Aorta | HEMOR | 17378 | -0.00177 | -0.00361 | 0.009609 | 0.573 | Artery_Aorta |
| Artery_Aorta | LEWY_grp | 17385 | -0.00185 | -0.00377 | 0.00993 | 0.57376 | Artery_Aorta |
| Breast_Mammary_Tissue | CVD_ATH | 17388 | -0.00225 | -0.00434 | 0.011837 | 0.57546 | Breast_Mammary_Tissue |
| Brain_Frontal_Cortex_BA9 | CVD_ART_ANY | 17387 | -0.00127 | -0.00235 | 0.006615 | 0.57641 | Brain_Frontal_Cortex_BA9 |
| Breast_Mammary_Tissue | AMY_ANY | 17377 | -0.00232 | -0.00447 | 0.012028 | 0.57654 | Breast_Mammary_Tissue |
| Adipose_Visceral_Omentum | HEMOR | 17378 | -0.00206 | -0.00401 | 0.010556 | 0.57741 | Adipose_Visceral_Omentum |
| Thyroid | CVD_ATH_ANY | 17386 | -0.0018 | -0.00357 | 0.009163 | 0.57786 | Thyroid |
| Brain_Hypothalamus | CVD_ART_ANY | 17387 | -0.00152 | -0.00262 | 0.007725 | 0.57815 | Brain_Hypothalamus |
| Kidney_Medulla | INFA | 17386 | -0.00167 | -0.00309 | 0.008454 | 0.57827 | Kidney_Medulla |
| Bladder | HEMOR | 17378 | -0.00243 | -0.00474 | 0.012292 | 0.57837 | Bladder |
| Kidney_Medulla | CVD_ATH | 17388 | -0.00175 | -0.00324 | 0.008828 | 0.57855 | Kidney_Medulla |
| Thyroid | CVD_S | 17387 | -0.00185 | -0.00367 | 0.009233 | 0.57942 | Thyroid |
| Kidney_Cortex | HS_S | 17386 | -0.00172 | -0.00302 | 0.008536 | 0.57962 | Kidney_Cortex |
| Stomach | LEWY_full | 17385 | -0.00232 | -0.00422 | 0.011487 | 0.58001 | Stomach |
| Brain_Cerebellum | INFA | 17386 | -0.00119 | -0.00236 | 0.005799 | 0.58097 | Brain_Cerebellum |
| Testis | CVD_CAA_ANY | 17386 | -0.00111 | -0.00192 | 0.005408 | 0.58143 | Testis |
| Colon_Transverse | AMY_ANY | 17377 | -0.00231 | -0.00425 | 0.011183 | 0.58186 | Colon_Transverse |
| Skin_Sun_Exposed_Lower_leg | BRAA | 17385 | -0.00156 | -0.00304 | 0.007528 | 0.58225 | Skin_Sun_Exposed_Lower_leg |
| Nerve_Tibial | AMY_ANY | 17377 | -0.00202 | -0.00407 | 0.009678 | 0.58262 | Nerve_Tibial |
| Whole_Blood | HS_S | 17386 | -0.00114 | -0.00207 | 0.005457 | 0.58294 | Whole_Blood |
| Heart_Left_Ventricle | LEWY_grp | 17385 | -0.0019 | -0.00317 | 0.009033 | 0.58315 | Heart_Left_Ventricle |
| Brain_Spinal_cord_cervical_c... | AMY_THAL | 17388 | -0.00174 | -0.00313 | 0.008139 | 0.58471 | Brain_Spinal_cord_cervical_c-1 |
| Spleen | LEWY_full | 17385 | -0.00155 | -0.00308 | 0.007174 | 0.5856 | Spleen |
| Skin_Sun_Exposed_Lower_leg | CVD_ART_ANY | 17387 | -0.00161 | -0.00312 | 0.007411 | 0.58592 | Skin_Sun_Exposed_Lower_leg |
| Fallopian_Tube | LEWY_grp | 17385 | -0.00251 | -0.00487 | 0.011531 | 0.586 | Fallopian_Tube |
| Artery_Aorta | WBVD | 17388 | -0.00213 | -0.00435 | 0.009618 | 0.58772 | Artery_Aorta |

| | | | | | | | |
|---|---|---|---|---|---|---|---|
| Brain_Hypothalamus | WMRCAL | 17388 | -0.00171 | -0.00295 | 0.007681 | 0.5881 | Brain_Hypothalamus |
| Fallopian_Tube | LEWY_ANY | 17385 | -0.00258 | -0.00501 | 0.011407 | 0.58939 | Fallopian_Tube |
| Nerve_Tibial | WBVD | 17388 | -0.00219 | -0.00441 | 0.00961 | 0.59007 | Nerve_Tibial |
| Liver | ADNC | 17388 | -0.0014 | -0.00252 | 0.00613 | 0.59014 | Liver |
| Testis | AMY_A | 17388 | -0.00125 | -0.00216 | 0.005353 | 0.59209 | Testis |
| Ovary | B_SCORE | 17388 | -0.00215 | -0.00435 | 0.009171 | 0.59253 | Ovary |
| Skin_Not_Sun_Exposed_Suprapu... | CVD_ART_ANY | 17387 | -0.00175 | -0.00337 | 0.0074 | 0.59331 | Skin_Not_Sun_Exposed_Suprapubic |
| Thyroid | LEWY_full | 17385 | -0.00224 | -0.00444 | 0.009334 | 0.5949 | Thyroid |
| Esophagus_Muscularis | WMR | 17388 | -0.0028 | -0.00554 | 0.011567 | 0.59579 | Esophagus_Muscularis |
| Brain_Spinal_cord_cervical_c... | HS_S | 17386 | -0.00202 | -0.00364 | 0.008122 | 0.59824 | Brain_Spinal_cord_cervical_c-1 |
| Whole_Blood | CVD_CAA_ANY | 17386 | -0.00137 | -0.00248 | 0.005507 | 0.59842 | Whole_Blood |
| Esophagus_Mucosa | AMY_ANY | 17377 | -0.00189 | -0.00365 | 0.007515 | 0.59919 | Esophagus_Mucosa |
| Small_Intestine_Terminal_Ile... | AMY_ANY | 17377 | -0.0023 | -0.00424 | 0.009158 | 0.59925 | Small_Intestine_Terminal_Ileum |
| Adipose_Visceral_Omentum | LEWY_ANY | 17385 | -0.00266 | -0.00518 | 0.010586 | 0.59935 | Adipose_Visceral_Omentum |
| Heart_Left_Ventricle | CVD_CAA | 17385 | -0.0022 | -0.00368 | 0.008669 | 0.60032 | Heart_Left_Ventricle |
| Skin_Sun_Exposed_Lower_leg | CVD_CAA_ANY | 17386 | -0.00194 | -0.00377 | 0.007463 | 0.60267 | Skin_Sun_Exposed_Lower_leg |
| Artery_Tibial | LEWY_full | 17385 | -0.00249 | -0.00513 | 0.009506 | 0.60337 | Artery_Tibial |
| Brain_Spinal_cord_cervical_c... | HEMOR | 17378 | -0.00215 | -0.00386 | 0.008164 | 0.6037 | Brain_Spinal_cord_cervical_c-1 |
| Brain_Cerebellar_Hemisphere | INFA | 17386 | -0.00148 | -0.00297 | 0.005606 | 0.60397 | Brain_Cerebellar_Hemisphere |
| Adipose_Subcutaneous | CVD_S | 17387 | -0.0027 | -0.00539 | 0.010121 | 0.60519 | Adipose_Subcutaneous |
| Esophagus_Mucosa | CVD_ART | 17388 | -0.00199 | -0.00384 | 0.00741 | 0.60571 | Esophagus_Mucosa |
| Skin_Not_Sun_Exposed_Suprapu... | BRAA | 17385 | -0.00203 | -0.00391 | 0.007525 | 0.6061 | Skin_Not_Sun_Exposed_Suprapubic |
| Brain_Hippocampus | AMY_A | 17388 | -0.0021 | -0.00356 | 0.007736 | 0.60706 | Brain_Hippocampus |
| Stomach | AMY_THAL | 17388 | -0.00307 | -0.00558 | 0.011286 | 0.60716 | Stomach |
| Heart_Left_Ventricle | CVD_ATH_ANY | 17386 | -0.00238 | -0.00397 | 0.008703 | 0.6076 | Heart_Left_Ventricle |
| Muscle_Skeletal | TDP43 | 17386 | -0.0019 | -0.0036 | 0.006851 | 0.60932 | Muscle_Skeletal |
| Esophagus_Gastroesophageal_J... | CVD_ATH_ANY | 17386 | -0.00334 | -0.00657 | 0.011988 | 0.60957 | Esophagus_Gastroesophageal_Junction |
| Brain_Spinal_cord_cervical_c... | MICR | 17385 | -0.00231 | -0.00416 | 0.008071 | 0.6126 | Brain_Spinal_cord_cervical_c-1 |
| Stomach | B_SCORE | 17388 | -0.00321 | -0.00584 | 0.011168 | 0.61324 | Stomach |
| Liver | TDP43 | 17386 | -0.00181 | -0.00327 | 0.006273 | 0.61379 | Liver |
| Colon_Transverse | CVD_ATH | 17388 | -0.00323 | -0.00595 | 0.01111 | 0.61436 | Colon_Transverse |
| Kidney_Cortex | TDP_3 | 17388 | -0.00255 | -0.00449 | 0.008754 | 0.61474 | Kidney_Cortex |
| Muscle_Skeletal | CVD_CAA | 17385 | -0.00195 | -0.00368 | 0.006648 | 0.61513 | Muscle_Skeletal |
| Spleen | CVD_S | 17387 | -0.00206 | -0.00408 | 0.007017 | 0.6153 | Spleen |
| Brain_Amygdala | WMRCAL | 17388 | -0.00221 | -0.00378 | 0.007519 | 0.61579 | Brain_Amygdala |
| Brain_Amygdala | CVD_S | 17387 | -0.00223 | -0.0038 | 0.007538 | 0.61628 | Brain_Amygdala |

| | | | | | | | |
|---|---|---|---|---|---|---|---|
| Cells_EBV-transformed_lympho... | CVD_CAA_ANY | 17386 | -0.00139 | -0.00303 | 0.004669 | 0.61679 | Cells_EBV-transformed_lymphocytes |
| Pituitary | CVD_CAA_ANY | 17386 | -0.00261 | -0.00487 | 0.008769 | 0.61692 | Pituitary |
| Brain_Cerebellum | TDP43 | 17386 | -0.00184 | -0.00366 | 0.006177 | 0.61729 | Brain_Cerebellum |
| Brain_Cortex | WMRCAL | 17388 | -0.00204 | -0.00372 | 0.006793 | 0.61802 | Brain_Cortex |
| Small_Intestine_Terminal_Ile... | LEWY_grp | 17385 | -0.00282 | -0.00519 | 0.009312 | 0.61892 | Small_Intestine_Terminal_Ileum |
| Esophagus_Mucosa | CVD_ATH_ANY | 17386 | -0.00223 | -0.00432 | 0.007375 | 0.61894 | Esophagus_Mucosa |
| Brain_Spinal_cord_cervical_c... | WBVD | 17388 | -0.00244 | -0.0044 | 0.008016 | 0.61978 | Brain_Spinal_cord_cervical_c-1 |
| Cervix_Ectocervix | CVD_ART | 17388 | -0.00362 | -0.00709 | 0.011618 | 0.62217 | Cervix_Ectocervix |
| Stomach | LEWY_ANY | 17385 | -0.00361 | -0.00656 | 0.011483 | 0.62337 | Stomach |
| Brain_Cerebellar_Hemisphere | CVD_ATH_ANY | 17386 | -0.00183 | -0.00368 | 0.005733 | 0.62535 | Brain_Cerebellar_Hemisphere |
| Heart_Atrial_Appendage | WMRCAL | 17388 | -0.00288 | -0.00518 | 0.008983 | 0.62577 | Heart_Atrial_Appendage |
| Testis | WMR | 17388 | -0.00171 | -0.00296 | 0.005307 | 0.62644 | Testis |
| Fallopian_Tube | HS_S | 17386 | -0.00366 | -0.0071 | 0.011228 | 0.6277 | Fallopian_Tube |
| Colon_Sigmoid | MICR | 17385 | -0.00387 | -0.00758 | 0.01177 | 0.62893 | Colon_Sigmoid |
| Brain_Hippocampus | WMRCAL | 17388 | -0.00252 | -0.00426 | 0.007636 | 0.62907 | Brain_Hippocampus |
| Lung | MICR | 17385 | -0.00283 | -0.00549 | 0.008587 | 0.62932 | Lung |
| Pancreas | HS_S | 17386 | -0.00271 | -0.00454 | 0.008153 | 0.63031 | Pancreas |
| Adipose_Visceral_Omentum | HS_S | 17386 | -0.00357 | -0.00694 | 0.010546 | 0.63246 | Adipose_Visceral_Omentum |
| Esophagus_Muscularis | CVD_CAA | 17385 | -0.00394 | -0.00778 | 0.011553 | 0.63345 | Esophagus_Muscularis |
| Stomach | WMRCAL | 17388 | -0.0039 | -0.00709 | 0.011401 | 0.63387 | Stomach |
| Brain_Substantia_nigra | AMY_A | 17388 | -0.00285 | -0.00491 | 0.008233 | 0.63548 | Brain_Substantia_nigra |
| Brain_Hypothalamus | AMY_A | 17388 | -0.00272 | -0.00468 | 0.007813 | 0.63597 | Brain_Hypothalamus |
| Brain_Caudate_basal_ganglia | CVD_S | 17387 | -0.00263 | -0.00454 | 0.007518 | 0.63664 | Brain_Caudate_basal_ganglia |
| Adipose_Visceral_Omentum | LEWY_full | 17385 | -0.00372 | -0.00722 | 0.010596 | 0.63708 | Adipose_Visceral_Omentum |
| Prostate | INFA | 17386 | -0.00386 | -0.00734 | 0.010992 | 0.63731 | Prostate |
| Bladder | WMRCAL | 17388 | -0.00453 | -0.00885 | 0.012483 | 0.64177 | Bladder |
| Ovary | WMR | 17388 | -0.00335 | -0.00681 | 0.009133 | 0.64331 | Ovary |
| Thyroid | WBVD | 17388 | -0.00339 | -0.00672 | 0.00922 | 0.64342 | Thyroid |
| Liver | CVD_S | 17387 | -0.00224 | -0.00403 | 0.006076 | 0.64356 | Liver |
| Adipose_Subcutaneous | BRAA | 17385 | -0.00374 | -0.00746 | 0.010138 | 0.64372 | Adipose_Subcutaneous |
| Brain_Hypothalamus | CVD_CAA | 17385 | -0.00284 | -0.0049 | 0.007712 | 0.6438 | Brain_Hypothalamus |
| Artery_Aorta | CVD_S | 17387 | -0.00362 | -0.00738 | 0.009695 | 0.64539 | Artery_Aorta |
| Artery_Aorta | AMY_ANY | 17377 | -0.00368 | -0.00751 | 0.009819 | 0.64606 | Artery_Aorta |
| Heart_Left_Ventricle | INFA | 17386 | -0.00322 | -0.00538 | 0.008587 | 0.6462 | Heart_Left_Ventricle |
| Adipose_Subcutaneous | LEWY_ANY | 17385 | -0.00391 | -0.00782 | 0.010303 | 0.64791 | Adipose_Subcutaneous |
| Nerve_Tibial | LEWY_full | 17385 | -0.00369 | -0.00744 | 0.009694 | 0.64838 | Nerve_Tibial |

| | | | | | | | |
|---|---|---|---|---|---|---|---|
| Stomach | LEWY_grp | 17385 | -0.00444 | -0.00807 | 0.011593 | 0.649 | Stomach |
| Brain_Anterior_cingulate_cor... | AMY_A | 17388 | -0.00268 | -0.00475 | 0.006999 | 0.64933 | Brain_Anterior_cingulate_cortex_BA24 |
| Brain_Spinal_cord_cervical_c... | AMY_A | 17388 | -0.00318 | -0.00572 | 0.008218 | 0.65051 | Brain_Spinal_cord_cervical_c-1 |
| Artery_Tibial | AMY_CERAD | 17386 | -0.00376 | -0.00774 | 0.009588 | 0.65247 | Artery_Tibial |
| Thyroid | LEWY_ANY | 17385 | -0.00374 | -0.00741 | 0.0095 | 0.65292 | Thyroid |
| Brain_Cortex | CVD_ART_ANY | 17387 | -0.0027 | -0.00493 | 0.006853 | 0.65319 | Brain_Cortex |
| Esophagus_Mucosa | B_SCORE | 17388 | -0.0029 | -0.00562 | 0.007338 | 0.65384 | Esophagus_Mucosa |
| Cells_Cultured_fibroblasts | INFA | 17386 | -0.00244 | -0.00523 | 0.006127 | 0.65461 | Cells_Cultured_fibroblasts |
| Fallopian_Tube | CVD_ART_ANY | 17387 | -0.00451 | -0.00877 | 0.011313 | 0.65503 | Fallopian_Tube |
| Nerve_Tibial | CVD_CAA | 17385 | -0.00381 | -0.00767 | 0.009476 | 0.65601 | Nerve_Tibial |
| Brain_Hippocampus | CVD_ART_ANY | 17387 | -0.0031 | -0.00526 | 0.007675 | 0.65706 | Brain_Hippocampus |
| Brain_Frontal_Cortex_BA9 | CVD_ATH_ANY | 17386 | -0.00265 | -0.00489 | 0.006517 | 0.65779 | Brain_Frontal_Cortex_BA9 |
| Vagina | LEWY_grp | 17385 | -0.00446 | -0.00855 | 0.010826 | 0.65998 | Vagina |
| Heart_Atrial_Appendage | CVD_ATH_ANY | 17386 | -0.00373 | -0.00671 | 0.009022 | 0.66052 | Heart_Atrial_Appendage |
| Heart_Atrial_Appendage | CVD_S | 17387 | -0.00379 | -0.0068 | 0.009128 | 0.6609 | Heart_Atrial_Appendage |
| Brain_Substantia_nigra | WMRCAL | 17388 | -0.00337 | -0.0058 | 0.008109 | 0.66104 | Brain_Substantia_nigra |
| Adrenal_Gland | B_SCORE | 17388 | -0.00392 | -0.00757 | 0.009392 | 0.66177 | Adrenal_Gland |
| Heart_Left_Ventricle | WMRCAL | 17388 | -0.00364 | -0.00607 | 0.0087 | 0.66197 | Heart_Left_Ventricle |
| Colon_Sigmoid | WMR | 17388 | -0.00495 | -0.00969 | 0.011835 | 0.66201 | Colon_Sigmoid |
| Pituitary | BRAA | 17385 | -0.00364 | -0.0068 | 0.008703 | 0.6622 | Pituitary |
| Esophagus_Mucosa | LEWY_ANY | 17385 | -0.00316 | -0.00612 | 0.007553 | 0.6623 | Esophagus_Mucosa |
| Liver | MICR | 17385 | -0.00255 | -0.0046 | 0.006066 | 0.66306 | Liver |
| Brain_Nucleus_accumbens_basa... | TDP_3 | 17388 | -0.00311 | -0.00539 | 0.007359 | 0.66347 | Brain_Nucleus_accumbens_basal_ganglia |
| Esophagus_Mucosa | TDP_3 | 17388 | -0.00316 | -0.00611 | 0.007424 | 0.66472 | Esophagus_Mucosa |
| Liver | AMY_CERAD | 17386 | -0.00263 | -0.00475 | 0.006178 | 0.66509 | Liver |
| Skin_Not_Sun_Exposed_Suprapu... | AMY_A | 17388 | -0.00322 | -0.00622 | 0.007465 | 0.66692 | Skin_Not_Sun_Exposed_Suprapubic |
| Heart_Left_Ventricle | TDP43 | 17386 | -0.00389 | -0.0065 | 0.00901 | 0.66705 | Heart_Left_Ventricle |
| Testis | LEWY_grp | 17385 | -0.00234 | -0.00404 | 0.005395 | 0.66767 | Testis |
| Kidney_Medulla | WMR | 17388 | -0.00386 | -0.00714 | 0.008788 | 0.66955 | Kidney_Medulla |
| Small_Intestine_Terminal_Ile... | TDP43 | 17386 | -0.00412 | -0.00758 | 0.009361 | 0.66988 | Small_Intestine_Terminal_Ileum |
| Minor_Salivary_Gland | CVD_CAA_ANY | 17386 | -0.00407 | -0.00758 | 0.009231 | 0.67023 | Minor_Salivary_Gland |
| Artery_Aorta | CVD_CAA | 17385 | -0.00433 | -0.00884 | 0.009668 | 0.67286 | Artery_Aorta |
| Adrenal_Gland | HEMOR | 17378 | -0.00422 | -0.00816 | 0.0094 | 0.67328 | Adrenal_Gland |
| Nerve_Tibial | LEWY_grp | 17385 | -0.00438 | -0.00882 | 0.009752 | 0.6733 | Nerve_Tibial |
| Pancreas | CVD_ATH | 17388 | -0.00359 | -0.006 | 0.007971 | 0.67366 | Pancreas |
| Brain_Cortex | B_SCORE | 17388 | -0.00305 | -0.00556 | 0.006752 | 0.67408 | Brain_Cortex |

| | | | | | | | |
|---|---|---|---|---|---|---|---|
| Brain_Putamen_basal_ganglia | CVD_S | 17387 | -0.00343 | -0.00584 | 0.007597 | 0.67413 | Brain_Putamen_basal_ganglia |
| Breast_Mammary_Tissue | HEMOR | 17378 | -0.00536 | -0.01031 | 0.011842 | 0.67448 | Breast_Mammary_Tissue |
| Liver | LEWY_grp | 17385 | -0.00281 | -0.00507 | 0.006187 | 0.67525 | Liver |
| Testis | CVD_ATH | 17388 | -0.00241 | -0.00417 | 0.005276 | 0.6763 | Testis |
| Colon_Sigmoid | CVD_ATH_ANY | 17386 | -0.00543 | -0.01064 | 0.011812 | 0.67715 | Colon_Sigmoid |
| Esophagus_Muscularis | INFA | 17386 | -0.00524 | -0.01035 | 0.011329 | 0.67816 | Esophagus_Muscularis |
| Muscle_Skeletal | CVD_S | 17387 | -0.00309 | -0.00585 | 0.006647 | 0.67919 | Muscle_Skeletal |
| Bladder | CVD_S | 17387 | -0.00576 | -0.01125 | 0.012349 | 0.67964 | Bladder |
| Uterus | BRAA | 17385 | -0.00488 | -0.00996 | 0.010425 | 0.68028 | Uterus |
| Stomach | CVD_ATH_ANY | 17386 | -0.00542 | -0.00986 | 0.011448 | 0.68206 | Stomach |
| Brain_Cortex | CVD_ATH_ANY | 17386 | -0.0032 | -0.00584 | 0.006758 | 0.68211 | Brain_Cortex |
| Adipose_Subcutaneous | B_SCORE | 17388 | -0.00478 | -0.00955 | 0.010093 | 0.6822 | Adipose_Subcutaneous |
| Vagina | MICR | 17385 | -0.00502 | -0.00962 | 0.010487 | 0.68402 | Vagina |
| Brain_Hippocampus | CVD_S | 17387 | -0.00366 | -0.0062 | 0.007619 | 0.68451 | Brain_Hippocampus |
| Cells_Cultured_fibroblasts | AMY_CERAD | 17386 | -0.00309 | -0.00663 | 0.006421 | 0.68498 | Cells_Cultured_fibroblasts |
| Heart_Left_Ventricle | AMY_CERAD | 17386 | -0.00431 | -0.0072 | 0.008921 | 0.68541 | Heart_Left_Ventricle |
| Brain_Cerebellar_Hemisphere | HS_S | 17386 | -0.0028 | -0.00562 | 0.005784 | 0.68567 | Brain_Cerebellar_Hemisphere |
| Brain_Nucleus_accumbens_basa... | CVD_ATH_ANY | 17386 | -0.0035 | -0.00607 | 0.007213 | 0.6861 | Brain_Nucleus_accumbens_basal_ganglia |
| Cervix_Endocervix | HS_S | 17386 | -0.00536 | -0.01062 | 0.010984 | 0.68717 | Cervix_Endocervix |
| Brain_Cerebellum | CVD_ATH_ANY | 17386 | -0.0029 | -0.00576 | 0.005929 | 0.68767 | Brain_Cerebellum |
| Brain_Anterior_cingulate_cor... | CVD_ART_ANY | 17387 | -0.00344 | -0.00608 | 0.006985 | 0.68864 | Brain_Anterior_cingulate_cortex_BA24 |
| Brain_Anterior_cingulate_cor... | WMRCAL | 17388 | -0.00341 | -0.00603 | 0.006922 | 0.68883 | Brain_Anterior_cingulate_cortex_BA24 |
| Brain_Substantia_nigra | CVD_ATH_ANY | 17386 | -0.00401 | -0.0069 | 0.008111 | 0.6893 | Brain_Substantia_nigra |
| Kidney_Medulla | CVD_ATH_ANY | 17386 | -0.00426 | -0.00788 | 0.008588 | 0.68997 | Kidney_Medulla |
| Prostate | CVD_ART_ANY | 17387 | -0.00569 | -0.01081 | 0.011447 | 0.69048 | Prostate |
| Bladder | CVD_ART_ANY | 17387 | -0.00618 | -0.01206 | 0.012414 | 0.69071 | Bladder |
| Brain_Spinal_cord_cervical_c... | CVD_ATH_ANY | 17386 | -0.00402 | -0.00724 | 0.008077 | 0.69085 | Brain_Spinal_cord_cervical_c-1 |
| Brain_Cerebellar_Hemisphere | CVD_ART | 17388 | -0.00292 | -0.00586 | 0.005852 | 0.69092 | Brain_Cerebellar_Hemisphere |
| Artery_Tibial | BRAA | 17385 | -0.00477 | -0.00983 | 0.009511 | 0.69195 | Artery_Tibial |
| Brain_Cerebellum | HS_S | 17386 | -0.003 | -0.00597 | 0.005979 | 0.69222 | Brain_Cerebellum |
| Brain_Amygdala | WMR | 17388 | -0.00373 | -0.00637 | 0.007405 | 0.69296 | Brain_Amygdala |
| Cells_EBV-transformed_lympho... | CVD_CAA | 17385 | -0.00234 | -0.00511 | 0.00463 | 0.69331 | Cells_EBV-transformed_lymphocytes |
| Pancreas | WBVD | 17388 | -0.00413 | -0.00692 | 0.008176 | 0.69343 | Pancreas |
| Brain_Cortex | WMR | 17388 | -0.00342 | -0.00624 | 0.006732 | 0.69406 | Brain_Cortex |
| Heart_Atrial_Appendage | INFA | 17386 | -0.00451 | -0.0081 | 0.008852 | 0.69483 | Heart_Atrial_Appendage |
| Esophagus_Gastroesophageal_J... | CVD_CAA | 17385 | -0.00607 | -0.01196 | 0.011897 | 0.69505 | Esophagus_Gastroesophageal_Junction |

| | | | | | | | |
|---|---|---|---|---|---|---|---|
| Minor_Salivary_Gland | MICR | 17385 | -0.00467 | -0.00871 | 0.009113 | 0.69596 | Minor_Salivary_Gland |
| Breast_Mammary_Tissue | BRAA | 17385 | -0.0061 | -0.01173 | 0.011881 | 0.69603 | Breast_Mammary_Tissue |
| Esophagus_Mucosa | LEWY_grp | 17385 | -0.00394 | -0.00762 | 0.007604 | 0.69769 | Esophagus_Mucosa |
| Vagina | AMY_THAL | 17388 | -0.00549 | -0.01051 | 0.010598 | 0.69785 | Vagina |
| Liver | BRAA | 17385 | -0.00323 | -0.00582 | 0.006221 | 0.69817 | Liver |
| Brain_Cerebellar_Hemisphere | BRAA | 17385 | -0.00301 | -0.00604 | 0.005778 | 0.69866 | Brain_Cerebellar_Hemisphere |
| Kidney_Cortex | WMR | 17388 | -0.00442 | -0.00777 | 0.00845 | 0.69952 | Kidney_Cortex |
| Bladder | AMY_THAL | 17388 | -0.00652 | -0.01272 | 0.012437 | 0.69987 | Bladder |
| Brain_Spinal_cord_cervical_c... | CVD_ART | 17388 | -0.00431 | -0.00775 | 0.008167 | 0.70095 | Brain_Spinal_cord_cervical_c-1 |
| Esophagus_Muscularis | AMY_ANY | 17377 | -0.0063 | -0.01243 | 0.011878 | 0.70203 | Esophagus_Muscularis |
| Colon_Sigmoid | CVD_ART_ANY | 17387 | -0.0063 | -0.01234 | 0.011847 | 0.70267 | Colon_Sigmoid |
| Heart_Left_Ventricle | MICR | 17385 | -0.00461 | -0.00769 | 0.008639 | 0.703 | Heart_Left_Ventricle |
| Skin_Sun_Exposed_Lower_leg | AMY_A | 17388 | -0.00398 | -0.00773 | 0.007453 | 0.70327 | Skin_Sun_Exposed_Lower_leg |
| Brain_Anterior_cingulate_cor... | CVD_S | 17387 | -0.00371 | -0.00656 | 0.006943 | 0.70333 | Brain_Anterior_cingulate_cortex_BA24 |
| Small_Intestine_Terminal_Ile... | LEWY_full | 17385 | -0.00492 | -0.00905 | 0.009196 | 0.70357 | Small_Intestine_Terminal_Ileum |
| Heart_Atrial_Appendage | HS_S | 17386 | -0.00477 | -0.00858 | 0.008924 | 0.70366 | Heart_Atrial_Appendage |
| Muscle_Skeletal | TDP_3 | 17388 | -0.00357 | -0.00676 | 0.006664 | 0.70391 | Muscle_Skeletal |
| Minor_Salivary_Gland | CVD_ATH_ANY | 17386 | -0.00492 | -0.00916 | 0.009157 | 0.70441 | Minor_Salivary_Gland |
| Brain_Cortex | AMY_A | 17388 | -0.00369 | -0.00673 | 0.006859 | 0.70451 | Brain_Cortex |
| Esophagus_Muscularis | CVD_ATH_ANY | 17386 | -0.00638 | -0.0126 | 0.011628 | 0.70839 | Esophagus_Muscularis |
| Stomach | AMY_ANY | 17377 | -0.00642 | -0.01167 | 0.011648 | 0.70909 | Stomach |
| Brain_Cerebellar_Hemisphere | TDP_3 | 17388 | -0.00329 | -0.00661 | 0.005905 | 0.71141 | Brain_Cerebellar_Hemisphere |
| Heart_Left_Ventricle | CVD_S | 17387 | -0.00492 | -0.00821 | 0.008808 | 0.71163 | Heart_Left_Ventricle |
| Uterus | AMY_ANY | 17377 | -0.00589 | -0.01202 | 0.010523 | 0.71228 | Uterus |
| Adipose_Subcutaneous | LEWY_grp | 17385 | -0.00588 | -0.01174 | 0.010454 | 0.71294 | Adipose_Subcutaneous |
| Skin_Sun_Exposed_Lower_leg | CVD_ATH | 17388 | -0.00415 | -0.00806 | 0.007386 | 0.71298 | Skin_Sun_Exposed_Lower_leg |
| Thyroid | LEWY_grp | 17385 | -0.0053 | -0.01051 | 0.009429 | 0.71308 | Thyroid |
| Artery_Aorta | CVD_CAA_ANY | 17386 | -0.00554 | -0.01132 | 0.009807 | 0.71399 | Artery_Aorta |
| Heart_Left_Ventricle | HS_S | 17386 | -0.00495 | -0.00827 | 0.008731 | 0.71465 | Heart_Left_Ventricle |
| Artery_Tibial | WMR | 17388 | -0.00537 | -0.01106 | 0.009461 | 0.7147 | Artery_Tibial |
| Pituitary | TDP43 | 17386 | -0.00505 | -0.00943 | 0.008766 | 0.71783 | Pituitary |
| Artery_Tibial | B_SCORE | 17388 | -0.00551 | -0.01135 | 0.009506 | 0.7188 | Artery_Tibial |
| Nerve_Tibial | CVD_S | 17387 | -0.00554 | -0.01116 | 0.009453 | 0.72106 | Nerve_Tibial |
| Brain_Caudate_basal_ganglia | CVD_ART_ANY | 17387 | -0.00444 | -0.00768 | 0.007551 | 0.72186 | Brain_Caudate_basal_ganglia |
| Brain_Amygdala | CVD_ART_ANY | 17387 | -0.00447 | -0.00762 | 0.007577 | 0.72233 | Brain_Amygdala |
| Whole_Blood | LEWY_full | 17385 | -0.00324 | -0.00585 | 0.005484 | 0.72247 | Whole_Blood |

| | | | | | | | |
|---|---|---|---|---|---|---|---|
| Nerve_Tibial | LEWY_ANY | 17385 | -0.00579 | -0.01166 | 0.009778 | 0.72305 | Nerve_Tibial |
| Minor_Salivary_Gland | HS_S | 17386 | -0.00545 | -0.01016 | 0.009175 | 0.72378 | Minor_Salivary_Gland |
| Brain_Spinal_cord_cervical_c... | CVD_ART_ANY | 17387 | -0.00486 | -0.00873 | 0.008126 | 0.72491 | Brain_Spinal_cord_cervical_c-1 |
| Breast_Mammary_Tissue | WMRCAL | 17388 | -0.00703 | -0.01353 | 0.011752 | 0.72507 | Breast_Mammary_Tissue |
| Liver | TDP_3 | 17388 | -0.0037 | -0.00666 | 0.006163 | 0.72563 | Liver |
| Colon_Transverse | MICR | 17385 | -0.00652 | -0.012 | 0.010838 | 0.72618 | Colon_Transverse |
| Spleen | AMY_ANY | 17377 | -0.00427 | -0.00847 | 0.007096 | 0.7263 | Spleen |
| Esophagus_Muscularis | HS_S | 17386 | -0.00709 | -0.014 | 0.01168 | 0.72813 | Esophagus_Muscularis |
| Esophagus_Gastroesophageal_J... | AMY_ANY | 17377 | -0.00747 | -0.01471 | 0.012253 | 0.72883 | Esophagus_Gastroesophageal_Junction |
| Cervix_Ectocervix | AMY_THAL | 17388 | -0.00711 | -0.01394 | 0.011605 | 0.72984 | Cervix_Ectocervix |
| Brain_Substantia_nigra | BRAA | 17385 | -0.00502 | -0.00865 | 0.008178 | 0.73027 | Brain_Substantia_nigra |
| Thyroid | HS_S | 17386 | -0.00569 | -0.01129 | 0.009269 | 0.73048 | Thyroid |
| Brain_Hippocampus | WMR | 17388 | -0.00465 | -0.00788 | 0.007517 | 0.73184 | Brain_Hippocampus |
| Pituitary | INFA | 17386 | -0.00526 | -0.00981 | 0.008479 | 0.73229 | Pituitary |
| Heart_Atrial_Appendage | TDP43 | 17386 | -0.00579 | -0.0104 | 0.009297 | 0.73329 | Heart_Atrial_Appendage |
| Breast_Mammary_Tissue | B_SCORE | 17388 | -0.00737 | -0.01419 | 0.011775 | 0.73435 | Breast_Mammary_Tissue |
| Brain_Cerebellum | BRAA | 17385 | -0.00375 | -0.00745 | 0.005981 | 0.73452 | Brain_Cerebellum |
| Lung | LEWY_full | 17385 | -0.00548 | -0.01061 | 0.008722 | 0.73507 | Lung |
| Brain_Frontal_Cortex_BA9 | AMY_A | 17388 | -0.00417 | -0.00769 | 0.006615 | 0.73576 | Brain_Frontal_Cortex_BA9 |
| Brain_Spinal_cord_cervical_c... | ADNC | 17388 | -0.00514 | -0.00925 | 0.00815 | 0.73597 | Brain_Spinal_cord_cervical_c-1 |
| Esophagus_Mucosa | CVD_ART_ANY | 17387 | -0.00468 | -0.00905 | 0.007381 | 0.73681 | Esophagus_Mucosa |
| Kidney_Medulla | WMRCAL | 17388 | -0.00552 | -0.01021 | 0.008621 | 0.73882 | Kidney_Medulla |
| Colon_Sigmoid | INFA | 17386 | -0.00744 | -0.01457 | 0.01159 | 0.73957 | Colon_Sigmoid |
| Uterus | WMRCAL | 17388 | -0.00668 | -0.01361 | 0.010387 | 0.73977 | Uterus |
| Fallopian_Tube | AMY_A | 17388 | -0.00739 | -0.01436 | 0.011455 | 0.74066 | Fallopian_Tube |
| Esophagus_Muscularis | CVD_ART | 17388 | -0.00753 | -0.01487 | 0.01166 | 0.74085 | Esophagus_Muscularis |
| Adipose_Visceral_Omentum | CVD_CAA | 17385 | -0.00674 | -0.0131 | 0.010423 | 0.74098 | Adipose_Visceral_Omentum |
| Brain_Cerebellum | WMR | 17388 | -0.00387 | -0.00769 | 0.005981 | 0.74121 | Brain_Cerebellum |
| Ovary | HS_S | 17386 | -0.00603 | -0.01224 | 0.009322 | 0.74125 | Ovary |
| Brain_Cerebellum | CVD_ART | 17388 | -0.00393 | -0.00781 | 0.006072 | 0.74143 | Brain_Cerebellum |
| Lung | HEMOR | 17378 | -0.00556 | -0.01077 | 0.008584 | 0.7415 | Lung |
| Stomach | WBVD | 17388 | -0.0073 | -0.01327 | 0.011257 | 0.74153 | Stomach |
| Breast_Mammary_Tissue | CVD_CAA_ANY | 17386 | -0.00775 | -0.01492 | 0.011942 | 0.74183 | Breast_Mammary_Tissue |
| Lung | LEWY_grp | 17385 | -0.00581 | -0.01124 | 0.00884 | 0.74443 | Lung |
| Ovary | WMRCAL | 17388 | -0.006 | -0.01217 | 0.009127 | 0.74453 | Ovary |
| Colon_Transverse | CVD_ART_ANY | 17387 | -0.00733 | -0.0135 | 0.011014 | 0.74724 | Colon_Transverse |

| | | | | | | | |
|---|---|---|---|---|---|---|---|
| Brain_Frontal_Cortex_BA9 | B_SCORE | 17388 | -0.00433 | -0.00799 | 0.006508 | 0.74725 | Brain_Frontal_Cortex_BA9 |
| Esophagus_Gastroesophageal_J... | CVD_ART | 17388 | -0.00803 | -0.01582 | 0.012017 | 0.74793 | Esophagus_Gastroesophageal_Junction |
| Brain_Spinal_cord_cervical_c... | WMR | 17388 | -0.00539 | -0.0097 | 0.008047 | 0.74852 | Brain_Spinal_cord_cervical_c-1 |
| Brain_Substantia_nigra | CVD_CAA_ANY | 17386 | -0.00556 | -0.00958 | 0.008267 | 0.74931 | Brain_Substantia_nigra |
| Artery_Aorta | AMY_A | 17388 | -0.00652 | -0.01332 | 0.0097 | 0.74938 | Artery_Aorta |
| Pancreas | CVD_S | 17387 | -0.00548 | -0.00918 | 0.008135 | 0.74986 | Pancreas |
| Brain_Caudate_basal_ganglia | CVD_ATH_ANY | 17386 | -0.00505 | -0.00874 | 0.007479 | 0.75042 | Brain_Caudate_basal_ganglia |
| Skin_Not_Sun_Exposed_Suprapu... | CVD_ATH | 17388 | -0.00501 | -0.00967 | 0.007382 | 0.75123 | Skin_Not_Sun_Exposed_Suprapubic |
| Brain_Amygdala | TDP_3 | 17388 | -0.00524 | -0.00894 | 0.00766 | 0.75314 | Brain_Amygdala |
| Brain_Nucleus_accumbens_basa... | WMR | 17388 | -0.00492 | -0.00855 | 0.007187 | 0.75327 | Brain_Nucleus_accumbens_basal_ganglia |
| Breast_Mammary_Tissue | CVD_S | 17387 | -0.00816 | -0.01572 | 0.011889 | 0.75385 | Breast_Mammary_Tissue |
| Adipose_Visceral_Omentum | LEWY_grp | 17385 | -0.00739 | -0.01437 | 0.010756 | 0.75394 | Adipose_Visceral_Omentum |
| Whole_Blood | TDP_3 | 17388 | -0.00383 | -0.00692 | 0.005449 | 0.75873 | Whole_Blood |
| Ovary | INFA | 17386 | -0.00632 | -0.01282 | 0.00898 | 0.75916 | Ovary |
| Adipose_Visceral_Omentum | AMY_ANY | 17377 | -0.00763 | -0.01484 | 0.010713 | 0.76194 | Adipose_Visceral_Omentum |
| Small_Intestine_Terminal_Ile... | WBVD | 17388 | -0.00645 | -0.01188 | 0.009053 | 0.76197 | Small_Intestine_Terminal_Ileum |
| Brain_Putamen_basal_ganglia | CVD_ATH_ANY | 17386 | -0.00539 | -0.00917 | 0.007542 | 0.76238 | Brain_Putamen_basal_ganglia |
| Brain_Nucleus_accumbens_basa... | CVD_ART_ANY | 17387 | -0.00522 | -0.00906 | 0.007294 | 0.76269 | Brain_Nucleus_accumbens_basal_ganglia |
| Pituitary | CVD_ATH_ANY | 17386 | -0.00612 | -0.01143 | 0.008557 | 0.76284 | Pituitary |
| Artery_Tibial | LEWY_grp | 17385 | -0.00693 | -0.01429 | 0.009638 | 0.7641 | Artery_Tibial |
| Artery_Aorta | WMRCAL | 17388 | -0.00696 | -0.01421 | 0.009656 | 0.76434 | Artery_Aorta |
| Kidney_Medulla | CVD_CAA | 17385 | -0.00627 | -0.01161 | 0.008692 | 0.7647 | Kidney_Medulla |
| Stomach | HS_S | 17386 | -0.00824 | -0.01498 | 0.011411 | 0.76479 | Stomach |
| Cells_EBV-transformed_lympho... | HS_S | 17386 | -0.00338 | -0.00739 | 0.004631 | 0.76754 | Cells_EBV-transformed_lymphocytes |
| Minor_Salivary_Gland | TDP_3 | 17388 | -0.00668 | -0.01244 | 0.009131 | 0.76766 | Minor_Salivary_Gland |
| Heart_Atrial_Appendage | CVD_ART_ANY | 17387 | -0.0066 | -0.01187 | 0.008999 | 0.76851 | Heart_Atrial_Appendage |
| Brain_Putamen_basal_ganglia | CVD_ART_ANY | 17387 | -0.0056 | -0.00954 | 0.007614 | 0.76898 | Brain_Putamen_basal_ganglia |
| Kidney_Cortex | CVD_ATH_ANY | 17386 | -0.00614 | -0.01079 | 0.008342 | 0.76898 | Kidney_Cortex |
| Brain_Cerebellum | TDP_3 | 17388 | -0.00454 | -0.00902 | 0.006112 | 0.77124 | Brain_Cerebellum |
| Kidney_Medulla | ADNC | 17388 | -0.00657 | -0.01217 | 0.008835 | 0.77153 | Kidney_Medulla |
| Bladder | WBVD | 17388 | -0.00916 | -0.01788 | 0.01231 | 0.77167 | Bladder |
| Heart_Atrial_Appendage | CVD_CAA | 17385 | -0.00665 | -0.01195 | 0.008926 | 0.77185 | Heart_Atrial_Appendage |
| Artery_Tibial | LEWY_ANY | 17385 | -0.00713 | -0.01469 | 0.009569 | 0.77187 | Artery_Tibial |
| Brain_Frontal_Cortex_BA9 | WMRCAL | 17388 | -0.00488 | -0.00901 | 0.00655 | 0.77205 | Brain_Frontal_Cortex_BA9 |
| Brain_Hippocampus | CVD_ATH_ANY | 17386 | -0.00564 | -0.00956 | 0.007562 | 0.77216 | Brain_Hippocampus |
| Muscle_Skeletal | LEWY_ANY | 17385 | -0.00504 | -0.00954 | 0.006753 | 0.77222 | Muscle_Skeletal |

| | | | | | | | |
|---|---|---|---|---|---|---|---|
| Brain_Hypothalamus | B_SCORE | 17388 | -0.00573 | -0.00987 | 0.007655 | 0.7728 | Brain_Hypothalamus |
| Colon_Sigmoid | AMY_ANY | 17377 | -0.0091 | -0.01783 | 0.012145 | 0.77324 | Colon_Sigmoid |
| Brain_Substantia_nigra | CVD_ART_ANY | 17387 | -0.00616 | -0.01061 | 0.008195 | 0.77376 | Brain_Substantia_nigra |
| Heart_Left_Ventricle | TDP_3 | 17388 | -0.00657 | -0.01097 | 0.008737 | 0.7739 | Heart_Left_Ventricle |
| Muscle_Skeletal | INFA | 17386 | -0.00489 | -0.00925 | 0.006482 | 0.7746 | Muscle_Skeletal |
| Brain_Amygdala | BRAA | 17385 | -0.00568 | -0.00969 | 0.007537 | 0.77465 | Brain_Amygdala |
| Brain_Spinal_cord_cervical_c... | CVD_S | 17387 | -0.00611 | -0.011 | 0.008084 | 0.77525 | Brain_Spinal_cord_cervical_c-1 |
| Brain_Anterior_cingulate_cor... | CVD_ATH_ANY | 17386 | -0.00522 | -0.00924 | 0.006862 | 0.77679 | Brain_Anterior_cingulate_cortex_BA24 |
| Pituitary | ADNC | 17388 | -0.00657 | -0.01226 | 0.008622 | 0.77692 | Pituitary |
| Adipose_Subcutaneous | WMRCAL | 17388 | -0.00763 | -0.01524 | 0.010012 | 0.77695 | Adipose_Subcutaneous |
| Testis | WMRCAL | 17388 | -0.00403 | -0.00696 | 0.005279 | 0.77719 | Testis |
| Muscle_Skeletal | CVD_ATH_ANY | 17386 | -0.00494 | -0.00935 | 0.006472 | 0.7775 | Muscle_Skeletal |
| Brain_Spinal_cord_cervical_c... | WMRCAL | 17388 | -0.00614 | -0.01105 | 0.008035 | 0.77758 | Brain_Spinal_cord_cervical_c-1 |
| Brain_Frontal_Cortex_BA9 | TDP43 | 17386 | -0.00515 | -0.00951 | 0.006743 | 0.77767 | Brain_Frontal_Cortex_BA9 |
| Artery_Coronary | CVD_CAA_ANY | 17386 | -0.00868 | -0.01729 | 0.011341 | 0.77785 | Artery_Coronary |
| Breast_Mammary_Tissue | HS_S | 17386 | -0.00907 | -0.01747 | 0.011849 | 0.7781 | Breast_Mammary_Tissue |
| Brain_Hypothalamus | HEMOR | 17378 | -0.00588 | -0.01014 | 0.007672 | 0.77832 | Brain_Hypothalamus |
| Brain_Spinal_cord_cervical_c... | CVD_ATH | 17388 | -0.00616 | -0.01108 | 0.008026 | 0.77858 | Brain_Spinal_cord_cervical_c-1 |
| Brain_Anterior_cingulate_cor... | WMR | 17388 | -0.00528 | -0.00933 | 0.006847 | 0.77951 | Brain_Anterior_cingulate_cortex_BA24 |
| Kidney_Medulla | TDP43 | 17386 | -0.00695 | -0.01287 | 0.009024 | 0.77954 | Kidney_Medulla |
| Prostate | CVD_ATH_ANY | 17386 | -0.00889 | -0.01689 | 0.011473 | 0.78086 | Prostate |
| Heart_Left_Ventricle | LEWY_full | 17385 | -0.00692 | -0.01155 | 0.008886 | 0.78177 | Heart_Left_Ventricle |
| Adipose_Subcutaneous | AMY_THAL | 17388 | -0.00799 | -0.01596 | 0.010235 | 0.78245 | Adipose_Subcutaneous |
| Brain_Cerebellum | WMRCAL | 17388 | -0.00462 | -0.00918 | 0.005918 | 0.78261 | Brain_Cerebellum |
| Skin_Sun_Exposed_Lower_leg | B_SCORE | 17388 | -0.00586 | -0.01137 | 0.007435 | 0.78452 | Skin_Sun_Exposed_Lower_leg |
| Brain_Caudate_basal_ganglia | TDP_3 | 17388 | -0.006 | -0.01038 | 0.007621 | 0.7846 | Brain_Caudate_basal_ganglia |
| Brain_Hypothalamus | WMR | 17388 | -0.00601 | -0.01035 | 0.00762 | 0.78479 | Brain_Hypothalamus |
| Small_Intestine_Terminal_Ile... | LEWY_ANY | 17385 | -0.00729 | -0.01343 | 0.009238 | 0.78507 | Small_Intestine_Terminal_Ileum |
| Prostate | CVD_S | 17387 | -0.00903 | -0.01715 | 0.011405 | 0.78576 | Prostate |
| Minor_Salivary_Gland | AMY_THAL | 17388 | -0.00717 | -0.01337 | 0.00902 | 0.7868 | Minor_Salivary_Gland |
| Liver | CVD_ATH | 17388 | -0.00489 | -0.0088 | 0.006116 | 0.78784 | Liver |
| Prostate | LEWY_full | 17385 | -0.00925 | -0.01756 | 0.011517 | 0.78893 | Prostate |
| Skin_Not_Sun_Exposed_Suprapu... | B_SCORE | 17388 | -0.006 | -0.01158 | 0.007426 | 0.79041 | Skin_Not_Sun_Exposed_Suprapubic |
| Brain_Nucleus_accumbens_basa... | CVD_ART | 17388 | -0.00596 | -0.01035 | 0.007374 | 0.79059 | Brain_Nucleus_accumbens_basal_ganglia |
| Nerve_Tibial | WMR | 17388 | -0.00764 | -0.0154 | 0.009412 | 0.79156 | Nerve_Tibial |
| Esophagus_Gastroesophageal_J... | HS_S | 17386 | -0.0098 | -0.01931 | 0.012045 | 0.79204 | Esophagus_Gastroesophageal_Junction |

| | | | | | | | |
|---|---|---|---|---|---|---|---|
| Brain_Cerebellar_Hemisphere | WMR | 17388 | -0.00472 | -0.00947 | 0.005766 | 0.79328 | Brain_Cerebellar_Hemisphere |
| Brain_Cerebellar_Hemisphere | AMY_CERAD | 17386 | -0.0048 | -0.00964 | 0.005849 | 0.79404 | Brain_Cerebellar_Hemisphere |
| Esophagus_Gastroesophageal_J... | LEWY_full | 17385 | -0.00996 | -0.01962 | 0.0121 | 0.79466 | Esophagus_Gastroesophageal_Junction |
| Bladder | LEWY_full | 17385 | -0.01042 | -0.02033 | 0.012571 | 0.79643 | Bladder |
| Esophagus_Muscularis | CVD_CAA_ANY | 17386 | -0.00975 | -0.01926 | 0.011754 | 0.79669 | Esophagus_Muscularis |
| Artery_Tibial | HS_S | 17386 | -0.0079 | -0.01627 | 0.009432 | 0.79874 | Artery_Tibial |
| Brain_Frontal_Cortex_BA9 | CVD_S | 17387 | -0.0055 | -0.01015 | 0.00657 | 0.79888 | Brain_Frontal_Cortex_BA9 |
| Esophagus_Mucosa | LEWY_full | 17385 | -0.00634 | -0.01226 | 0.007542 | 0.79955 | Esophagus_Mucosa |
| Liver | LEWY_full | 17385 | -0.00512 | -0.00923 | 0.006088 | 0.79987 | Liver |
| Muscle_Skeletal | BRAA | 17385 | -0.00559 | -0.01059 | 0.006649 | 0.79995 | Muscle_Skeletal |
| Brain_Frontal_Cortex_BA9 | TDP_3 | 17388 | -0.00565 | -0.01041 | 0.006692 | 0.80054 | Brain_Frontal_Cortex_BA9 |
| Brain_Cortex | TDP_3 | 17388 | -0.00587 | -0.01072 | 0.006936 | 0.80129 | Brain_Cortex |
| Vagina | CVD_CAA | 17385 | -0.00904 | -0.0173 | 0.010668 | 0.80157 | Vagina |
| Pancreas | LEWY_grp | 17385 | -0.00708 | -0.01186 | 0.008354 | 0.80178 | Pancreas |
| Liver | AMY_THAL | 17388 | -0.00521 | -0.00938 | 0.006138 | 0.80184 | Liver |
| Minor_Salivary_Gland | CVD_CAA | 17385 | -0.00777 | -0.01447 | 0.009111 | 0.80301 | Minor_Salivary_Gland |
| Pancreas | LEWY_ANY | 17385 | -0.00705 | -0.0118 | 0.008272 | 0.80301 | Pancreas |
| Brain_Amygdala | CVD_ATH_ANY | 17386 | -0.00639 | -0.0109 | 0.00744 | 0.80494 | Brain_Amygdala |
| Kidney_Medulla | WBVD | 17388 | -0.00747 | -0.01383 | 0.008694 | 0.80495 | Kidney_Medulla |
| Pancreas | LEWY_full | 17385 | -0.00716 | -0.01198 | 0.008324 | 0.80511 | Pancreas |
| Heart_Left_Ventricle | CVD_ART_ANY | 17387 | -0.00742 | -0.0124 | 0.008622 | 0.80531 | Heart_Left_Ventricle |
| Brain_Cerebellar_Hemisphere | ADNC | 17388 | -0.00499 | -0.01001 | 0.005788 | 0.80548 | Brain_Cerebellar_Hemisphere |
| Artery_Aorta | AMY_THAL | 17388 | -0.00843 | -0.01721 | 0.009767 | 0.80583 | Artery_Aorta |
| Brain_Caudate_basal_ganglia | B_SCORE | 17388 | -0.00645 | -0.01116 | 0.007459 | 0.80652 | Brain_Caudate_basal_ganglia |
| Kidney_Cortex | AMY_THAL | 17388 | -0.00732 | -0.01288 | 0.008432 | 0.80738 | Kidney_Cortex |
| Esophagus_Gastroesophageal_J... | CVD_CAA_ANY | 17386 | -0.0106 | -0.02088 | 0.012133 | 0.8088 | Esophagus_Gastroesophageal_Junction |
| Stomach | CVD_ART_ANY | 17387 | -0.00997 | -0.01813 | 0.011378 | 0.8095 | Stomach |
| Brain_Cerebellum | ADNC | 17388 | -0.00527 | -0.01047 | 0.005983 | 0.81075 | Brain_Cerebellum |
| Brain_Frontal_Cortex_BA9 | WMR | 17388 | -0.00572 | -0.01056 | 0.006491 | 0.81108 | Brain_Frontal_Cortex_BA9 |
| Testis | TDP43 | 17386 | -0.00486 | -0.0084 | 0.005472 | 0.81268 | Testis |
| Skin_Not_Sun_Exposed_Suprapu... | AMY_THAL | 17388 | -0.00658 | -0.0127 | 0.007411 | 0.81269 | Skin_Not_Sun_Exposed_Suprapubic |
| Kidney_Medulla | MICR | 17385 | -0.00762 | -0.01411 | 0.008585 | 0.81273 | Kidney_Medulla |
| Brain_Frontal_Cortex_BA9 | INFA | 17386 | -0.00569 | -0.01049 | 0.006401 | 0.81292 | Brain_Frontal_Cortex_BA9 |
| Muscle_Skeletal | AMY_CERAD | 17386 | -0.00607 | -0.01148 | 0.006789 | 0.81427 | Muscle_Skeletal |
| Pancreas | TDP43 | 17386 | -0.00751 | -0.01257 | 0.008336 | 0.81625 | Pancreas |
| Brain_Anterior_cingulate_cor... | B_SCORE | 17388 | -0.00618 | -0.01093 | 0.006861 | 0.81629 | Brain_Anterior_cingulate_cortex_BA24 |

| | | | | | | | |
|---|---|---|---|---|---|---|---|
| Adrenal_Gland | MICR | 17385 | -0.0084 | -0.01623 | 0.009305 | 0.81662 | Adrenal_Gland |
| Brain_Cortex | INFA | 17386 | -0.00602 | -0.01099 | 0.006657 | 0.81715 | Brain_Cortex |
| Colon_Sigmoid | CVD_CAA_ANY | 17386 | -0.01088 | -0.0213 | 0.011997 | 0.81766 | Colon_Sigmoid |
| Brain_Cerebellar_Hemisphere | WMRCAL | 17388 | -0.00519 | -0.01043 | 0.005729 | 0.81769 | Brain_Cerebellar_Hemisphere |
| Esophagus_Mucosa | MICR | 17385 | -0.00667 | -0.01291 | 0.007311 | 0.81926 | Esophagus_Mucosa |
| Muscle_Skeletal | HS_S | 17386 | -0.00606 | -0.01147 | 0.006627 | 0.81986 | Muscle_Skeletal |
| Brain_Amygdala | CVD_ART | 17388 | -0.00699 | -0.01192 | 0.007616 | 0.82066 | Brain_Amygdala |
| Testis | CVD_CAA | 17385 | -0.00499 | -0.00862 | 0.005396 | 0.82232 | Testis |
| Testis | TDP_3 | 17388 | -0.00508 | -0.00877 | 0.005462 | 0.82364 | Testis |
| Nerve_Tibial | CVD_CAA_ANY | 17386 | -0.00907 | -0.01828 | 0.009727 | 0.82443 | Nerve_Tibial |
| Vagina | ADNC | 17388 | -0.00989 | -0.01893 | 0.010571 | 0.82521 | Vagina |
| Kidney_Cortex | CVD_ATH | 17388 | -0.00803 | -0.01413 | 0.008567 | 0.82578 | Kidney_Cortex |
| Liver | WBVD | 17388 | -0.00575 | -0.01036 | 0.006108 | 0.82678 | Liver |
| Brain_Cortex | CVD_S | 17387 | -0.00641 | -0.0117 | 0.006809 | 0.82679 | Brain_Cortex |
| Brain_Substantia_nigra | CVD_CAA | 17385 | -0.00776 | -0.01337 | 0.008179 | 0.82867 | Brain_Substantia_nigra |
| Ovary | AMY_CERAD | 17386 | -0.00883 | -0.01792 | 0.009294 | 0.82898 | Ovary |
| Brain_Hypothalamus | ADNC | 17388 | -0.00738 | -0.01271 | 0.007722 | 0.83024 | Brain_Hypothalamus |
| Brain_Nucleus_accumbens_basa... | B_SCORE | 17388 | -0.00692 | -0.01202 | 0.007206 | 0.83154 | Brain_Nucleus_accumbens_basal_ganglia |
| Bladder | CVD_CAA_ANY | 17386 | -0.01193 | -0.02328 | 0.012412 | 0.83178 | Bladder |
| Breast_Mammary_Tissue | AMY_THAL | 17388 | -0.01147 | -0.02209 | 0.011913 | 0.83224 | Breast_Mammary_Tissue |
| Vagina | WBVD | 17388 | -0.01025 | -0.01961 | 0.010624 | 0.83255 | Vagina |
| Brain_Anterior_cingulate_cor... | INFA | 17386 | -0.00652 | -0.01154 | 0.006763 | 0.83266 | Brain_Anterior_cingulate_cortex_BA24 |
| Kidney_Cortex | TDP43 | 17386 | -0.00839 | -0.01476 | 0.008675 | 0.83331 | Kidney_Cortex |
| Bladder | MICR | 17385 | -0.01196 | -0.02334 | 0.012345 | 0.83367 | Bladder |
| Brain_Caudate_basal_ganglia | INFA | 17386 | -0.00716 | -0.01239 | 0.007358 | 0.83482 | Brain_Caudate_basal_ganglia |
| Brain_Cerebellum | AMY_CERAD | 17386 | -0.0059 | -0.01172 | 0.006057 | 0.83493 | Brain_Cerebellum |
| Whole_Blood | HEMOR | 17378 | -0.00529 | -0.00957 | 0.005426 | 0.83537 | Whole_Blood |
| Artery_Coronary | AMY_ANY | 17377 | -0.01116 | -0.02224 | 0.011419 | 0.83582 | Artery_Coronary |
| Pancreas | MICR | 17385 | -0.00792 | -0.01326 | 0.008054 | 0.8374 | Pancreas |
| Lung | LEWY_ANY | 17385 | -0.00857 | -0.01659 | 0.00871 | 0.83749 | Lung |
| Uterus | AMY_THAL | 17388 | -0.01034 | -0.02108 | 0.010501 | 0.83755 | Uterus |
| Brain_Nucleus_accumbens_basa... | ADNC | 17388 | -0.00716 | -0.01244 | 0.007258 | 0.83808 | Brain_Nucleus_accumbens_basal_ganglia |
| Brain_Hippocampus | HEMOR | 17378 | -0.00754 | -0.01278 | 0.007623 | 0.83873 | Brain_Hippocampus |
| Brain_Nucleus_accumbens_basa... | HEMOR | 17378 | -0.0072 | -0.0125 | 0.007246 | 0.83972 | Brain_Nucleus_accumbens_basal_ganglia |
| Brain_Putamen_basal_ganglia | TDP_3 | 17388 | -0.00764 | -0.01302 | 0.007692 | 0.83982 | Brain_Putamen_basal_ganglia |
| Brain_Amygdala | B_SCORE | 17388 | -0.00742 | -0.01265 | 0.00745 | 0.84023 | Brain_Amygdala |

| | | | | | | | |
|---|---|---|---|---|---|---|---|
| Bladder | LEWY_ANY | 17385 | -0.01267 | -0.02472 | 0.012683 | 0.84105 | Bladder |
| Brain_Anterior_cingulate_cor... | TDP_3 | 17388 | -0.00708 | -0.01252 | 0.007067 | 0.84189 | Brain_Anterior_cingulate_cortex_BA24 |
| Brain_Substantia_nigra | WMR | 17388 | -0.00813 | -0.014 | 0.008036 | 0.84404 | Brain_Substantia_nigra |
| Nerve_Tibial | AMY_A | 17388 | -0.00974 | -0.01962 | 0.00961 | 0.84456 | Nerve_Tibial |
| Spleen | CVD_CAA | 17385 | -0.00719 | -0.01427 | 0.007063 | 0.84574 | Spleen |
| Uterus | WBVD | 17388 | -0.0105 | -0.02142 | 0.010305 | 0.84592 | Uterus |
| Colon_Transverse | B_SCORE | 17388 | -0.01109 | -0.02041 | 0.010876 | 0.84604 | Colon_Transverse |
| Cells_Cultured_fibroblasts | AMY_THAL | 17388 | -0.00644 | -0.01381 | 0.00627 | 0.8479 | Cells_Cultured_fibroblasts |
| Cells_EBV-transformed_lympho... | LEWY_ANY | 17385 | -0.00484 | -0.01057 | 0.004705 | 0.84827 | Cells_EBV-transformed_lymphocytes |
| Esophagus_Muscularis | LEWY_full | 17385 | -0.01208 | -0.02384 | 0.011711 | 0.84876 | Esophagus_Muscularis |
| Prostate | LEWY_ANY | 17385 | -0.01205 | -0.02289 | 0.011682 | 0.84888 | Prostate |
| Adipose_Subcutaneous | AMY_ANY | 17377 | -0.01074 | -0.02144 | 0.010321 | 0.85083 | Adipose_Subcutaneous |
| Brain_Putamen_basal_ganglia | INFA | 17386 | -0.00773 | -0.01316 | 0.007422 | 0.85104 | Brain_Putamen_basal_ganglia |
| Testis | MICR | 17385 | -0.00558 | -0.00964 | 0.005346 | 0.8516 | Testis |
| Esophagus_Mucosa | CVD_CAA | 17385 | -0.00779 | -0.01506 | 0.007453 | 0.85189 | Esophagus_Mucosa |
| Brain_Hippocampus | BRAA | 17385 | -0.00797 | -0.01352 | 0.007615 | 0.85247 | Brain_Hippocampus |
| Brain_Frontal_Cortex_BA9 | CVD_ART | 17388 | -0.00697 | -0.01285 | 0.006632 | 0.85327 | Brain_Frontal_Cortex_BA9 |
| Colon_Sigmoid | WMRCAL | 17388 | -0.01233 | -0.02414 | 0.011729 | 0.85335 | Colon_Sigmoid |
| Heart_Left_Ventricle | HEMOR | 17378 | -0.00923 | -0.01542 | 0.008762 | 0.85397 | Heart_Left_Ventricle |
| Cells_Cultured_fibroblasts | AMY_A | 17388 | -0.00663 | -0.01421 | 0.006287 | 0.85409 | Cells_Cultured_fibroblasts |
| Skin_Sun_Exposed_Lower_leg | AMY_THAL | 17388 | -0.00779 | -0.01514 | 0.007392 | 0.85414 | Skin_Sun_Exposed_Lower_leg |
| Brain_Frontal_Cortex_BA9 | HEMOR | 17378 | -0.00694 | -0.01279 | 0.00654 | 0.85552 | Brain_Frontal_Cortex_BA9 |
| Brain_Caudate_basal_ganglia | ADNC | 17388 | -0.00798 | -0.01379 | 0.007514 | 0.85579 | Brain_Caudate_basal_ganglia |
| Brain_Putamen_basal_ganglia | WMR | 17388 | -0.00799 | -0.01361 | 0.00751 | 0.85631 | Brain_Putamen_basal_ganglia |
| Brain_Nucleus_accumbens_basa... | INFA | 17386 | -0.00758 | -0.01317 | 0.007104 | 0.85717 | Brain_Nucleus_accumbens_basal_ganglia |
| Brain_Nucleus_accumbens_basa... | TDP43 | 17386 | -0.00795 | -0.01381 | 0.007433 | 0.8577 | Brain_Nucleus_accumbens_basal_ganglia |
| Cervix_Endocervix | AMY_THAL | 17388 | -0.01193 | -0.02364 | 0.011095 | 0.85886 | Cervix_Endocervix |
| Brain_Anterior_cingulate_cor... | BRAA | 17385 | -0.00744 | -0.01315 | 0.006917 | 0.8589 | Brain_Anterior_cingulate_cortex_BA24 |
| Brain_Caudate_basal_ganglia | WMR | 17388 | -0.008 | -0.01383 | 0.007439 | 0.8589 | Brain_Caudate_basal_ganglia |
| Heart_Left_Ventricle | CVD_CAA_ANY | 17386 | -0.00934 | -0.0156 | 0.008643 | 0.86007 | Heart_Left_Ventricle |
| Brain_Hippocampus | TDP_3 | 17388 | -0.00839 | -0.01422 | 0.007742 | 0.86061 | Brain_Hippocampus |
| Colon_Transverse | LEWY_grp | 17385 | -0.01219 | -0.02243 | 0.01123 | 0.86104 | Colon_Transverse |
| Brain_Cortex | TDP43 | 17386 | -0.00759 | -0.01385 | 0.006993 | 0.86112 | Brain_Cortex |
| Brain_Hippocampus | B_SCORE | 17388 | -0.00817 | -0.01386 | 0.007524 | 0.86135 | Brain_Hippocampus |
| Vagina | CVD_ART | 17388 | -0.01152 | -0.02206 | 0.010575 | 0.862 | Vagina |
| Brain_Cerebellum | CVD_S | 17387 | -0.00644 | -0.01279 | 0.005908 | 0.8621 | Brain_Cerebellum |

| | | | | | | | |
|---|---|---|---|---|---|---|---|
| Liver | CVD_ART_ANY | 17387 | -0.00671 | -0.01209 | 0.00615 | 0.86238 | Liver |
| Brain_Hypothalamus | BRAA | 17385 | -0.00841 | -0.0145 | 0.007709 | 0.86249 | Brain_Hypothalamus |
| Brain_Putamen_basal_ganglia | B_SCORE | 17388 | -0.00823 | -0.01402 | 0.007534 | 0.86273 | Brain_Putamen_basal_ganglia |
| Heart_Atrial_Appendage | BRAA | 17385 | -0.00993 | -0.01784 | 0.00908 | 0.86291 | Heart_Atrial_Appendage |
| Spleen | HEMOR | 17378 | -0.00774 | -0.01536 | 0.007061 | 0.86361 | Spleen |
| Cells_Cultured_fibroblasts | ADNC | 17388 | -0.00692 | -0.01483 | 0.006306 | 0.86374 | Cells_Cultured_fibroblasts |
| Brain_Anterior_cingulate_cor... | HEMOR | 17378 | -0.00759 | -0.01342 | 0.006909 | 0.86404 | Brain_Anterior_cingulate_cortex_BA24 |
| Skin_Sun_Exposed_Lower_leg | ADNC | 17388 | -0.00813 | -0.01579 | 0.007383 | 0.86459 | Skin_Sun_Exposed_Lower_leg |
| Pancreas | AMY_THAL | 17388 | -0.009 | -0.01507 | 0.008166 | 0.8649 | Pancreas |
| Prostate | WBVD | 17388 | -0.01271 | -0.02415 | 0.011509 | 0.86531 | Prostate |
| Brain_Cortex | HEMOR | 17378 | -0.00755 | -0.01377 | 0.00679 | 0.86674 | Brain_Cortex |
| Brain_Caudate_basal_ganglia | AMY_CERAD | 17386 | -0.00848 | -0.01466 | 0.007628 | 0.86681 | Brain_Caudate_basal_ganglia |
| Heart_Atrial_Appendage | CVD_CAA_ANY | 17386 | -0.00998 | -0.01794 | 0.008978 | 0.86691 | Heart_Atrial_Appendage |
| Brain_Nucleus_accumbens_basa... | BRAA | 17385 | -0.0081 | -0.01407 | 0.007256 | 0.86786 | Brain_Nucleus_accumbens_basal_ganglia |
| Brain_Anterior_cingulate_cor... | CVD_ART | 17388 | -0.00785 | -0.01388 | 0.007001 | 0.86886 | Brain_Anterior_cingulate_cortex_BA24 |
| Brain_Hypothalamus | CVD_ATH | 17388 | -0.00859 | -0.0148 | 0.007658 | 0.86901 | Brain_Hypothalamus |
| Brain_Putamen_basal_ganglia | AMY_CERAD | 17386 | -0.00866 | -0.01475 | 0.007717 | 0.86906 | Brain_Putamen_basal_ganglia |
| Brain_Substantia_nigra | ADNC | 17388 | -0.00926 | -0.01596 | 0.00821 | 0.87034 | Brain_Substantia_nigra |
| Brain_Nucleus_accumbens_basa... | AMY_CERAD | 17386 | -0.00834 | -0.01449 | 0.007372 | 0.87108 | Brain_Nucleus_accumbens_basal_ganglia |
| Cells_EBV-transformed_lympho... | LEWY_grp | 17385 | -0.00537 | -0.01173 | 0.004738 | 0.87171 | Cells_EBV-transformed_lymphocytes |
| Brain_Substantia_nigra | HEMOR | 17378 | -0.00926 | -0.01596 | 0.00816 | 0.87182 | Brain_Substantia_nigra |
| Brain_Anterior_cingulate_cor... | TDP43 | 17386 | -0.00809 | -0.0143 | 0.007113 | 0.87216 | Brain_Anterior_cingulate_cortex_BA24 |
| Brain_Amygdala | HEMOR | 17378 | -0.00858 | -0.01463 | 0.007525 | 0.87278 | Brain_Amygdala |
| Pituitary | MICR | 17385 | -0.0098 | -0.0183 | 0.00859 | 0.87308 | Pituitary |
| Brain_Cortex | CVD_ATH | 17388 | -0.00774 | -0.01412 | 0.006759 | 0.87379 | Brain_Cortex |
| Brain_Putamen_basal_ganglia | ADNC | 17388 | -0.00873 | -0.01487 | 0.007606 | 0.87448 | Brain_Putamen_basal_ganglia |
| Brain_Hippocampus | CVD_ART | 17388 | -0.00886 | -0.01502 | 0.007721 | 0.87449 | Brain_Hippocampus |
| Artery_Coronary | WMRCAL | 17388 | -0.01284 | -0.02557 | 0.011156 | 0.87504 | Artery_Coronary |
| Brain_Cortex | CVD_ART | 17388 | -0.00794 | -0.0145 | 0.006882 | 0.8758 | Brain_Cortex |
| Muscle_Skeletal | WMR | 17388 | -0.00766 | -0.0145 | 0.0066 | 0.87722 | Muscle_Skeletal |
| Ovary | AMY_ANY | 17377 | -0.01078 | -0.02187 | 0.009279 | 0.87726 | Ovary |
| Esophagus_Mucosa | ADNC | 17388 | -0.00854 | -0.01653 | 0.007356 | 0.87727 | Esophagus_Mucosa |
| Brain_Cerebellar_Hemisphere | CVD_S | 17387 | -0.00671 | -0.01348 | 0.005722 | 0.87955 | Brain_Cerebellar_Hemisphere |
| Brain_Caudate_basal_ganglia | HEMOR | 17378 | -0.00885 | -0.0153 | 0.007509 | 0.88066 | Brain_Caudate_basal_ganglia |
| Skin_Not_Sun_Exposed_Suprapu... | ADNC | 17388 | -0.00873 | -0.01685 | 0.007389 | 0.88129 | Skin_Not_Sun_Exposed_Suprapubic |
| Cervix_Ectocervix | WBVD | 17388 | -0.0136 | -0.02669 | 0.011504 | 0.88148 | Cervix_Ectocervix |

| | | | | | | | |
|---|---|---|---|---|---|---|---|
| Brain_Nucleus_accumbens_basa... | CVD_ATH | 17388 | -0.00852 | -0.01479 | 0.007199 | 0.88162 | Brain_Nucleus_accumbens_basal_ganglia |
| Brain_Amygdala | ADNC | 17388 | -0.00899 | -0.01534 | 0.007566 | 0.88271 | Brain_Amygdala |
| Cells_Cultured_fibroblasts | AMY_ANY | 17377 | -0.00764 | -0.01639 | 0.006424 | 0.88295 | Cells_Cultured_fibroblasts |
| Brain_Hippocampus | ADNC | 17388 | -0.00912 | -0.01546 | 0.007656 | 0.88314 | Brain_Hippocampus |
| Bladder | LEWY_grp | 17385 | -0.0152 | -0.02966 | 0.012751 | 0.88335 | Bladder |
| Brain_Caudate_basal_ganglia | BRAA | 17385 | -0.009 | -0.01555 | 0.007547 | 0.88336 | Brain_Caudate_basal_ganglia |
| Muscle_Skeletal | AMY_ANY | 17377 | -0.00823 | -0.01558 | 0.006889 | 0.88396 | Muscle_Skeletal |
| Adipose_Subcutaneous | CVD_CAA | 17385 | -0.01219 | -0.02435 | 0.010116 | 0.88585 | Adipose_Subcutaneous |
| Brain_Cerebellum | AMY_A | 17388 | -0.00722 | -0.01435 | 0.005985 | 0.88622 | Brain_Cerebellum |
| Heart_Atrial_Appendage | MICR | 17385 | -0.01093 | -0.01963 | 0.009031 | 0.88685 | Heart_Atrial_Appendage |
| Vagina | CVD_CAA_ANY | 17386 | -0.01303 | -0.02494 | 0.010758 | 0.88704 | Vagina |
| Fallopian_Tube | WBVD | 17388 | -0.01356 | -0.02634 | 0.011182 | 0.88741 | Fallopian_Tube |
| Ovary | BRAA | 17385 | -0.01129 | -0.02291 | 0.009283 | 0.88805 | Ovary |
| Bladder | AMY_A | 17388 | -0.01528 | -0.02981 | 0.012495 | 0.88928 | Bladder |
| Kidney_Cortex | WBVD | 17388 | -0.01021 | -0.01796 | 0.008348 | 0.88929 | Kidney_Cortex |
| Brain_Cortex | ADNC | 17388 | -0.00831 | -0.01517 | 0.006785 | 0.88967 | Brain_Cortex |
| Vagina | AMY_A | 17388 | -0.01314 | -0.02515 | 0.010707 | 0.89009 | Vagina |
| Brain_Substantia_nigra | CVD_ATH | 17388 | -0.00996 | -0.01717 | 0.00807 | 0.89151 | Brain_Substantia_nigra |
| Minor_Salivary_Gland | LEWY_grp | 17385 | -0.01161 | -0.02163 | 0.009391 | 0.89177 | Minor_Salivary_Gland |
| Muscle_Skeletal | LEWY_grp | 17385 | -0.00835 | -0.01581 | 0.006755 | 0.8919 | Muscle_Skeletal |
| Brain_Cerebellar_Hemisphere | AMY_A | 17388 | -0.00716 | -0.01438 | 0.005789 | 0.89199 | Brain_Cerebellar_Hemisphere |
| Uterus | HS_S | 17386 | -0.01275 | -0.026 | 0.010278 | 0.89258 | Uterus |
| Adrenal_Gland | WMR | 17388 | -0.01166 | -0.02253 | 0.0094 | 0.89265 | Adrenal_Gland |
| Kidney_Medulla | AMY_THAL | 17388 | -0.01091 | -0.0202 | 0.008754 | 0.89363 | Kidney_Medulla |
| Brain_Frontal_Cortex_BA9 | CVD_ATH | 17388 | -0.00819 | -0.01511 | 0.006518 | 0.89553 | Brain_Frontal_Cortex_BA9 |
| Heart_Atrial_Appendage | AMY_ANY | 17377 | -0.01164 | -0.02091 | 0.009243 | 0.89595 | Heart_Atrial_Appendage |
| Brain_Amygdala | TDP43 | 17386 | -0.00974 | -0.01661 | 0.007714 | 0.89664 | Brain_Amygdala |
| Adipose_Visceral_Omentum | CVD_CAA_ANY | 17386 | -0.01331 | -0.02588 | 0.01054 | 0.89665 | Adipose_Visceral_Omentum |
| Kidney_Medulla | CVD_ART_ANY | 17387 | -0.01102 | -0.0204 | 0.008718 | 0.89691 | Kidney_Medulla |
| Esophagus_Gastroesophageal_J... | LEWY_grp | 17385 | -0.01549 | -0.03053 | 0.012237 | 0.89723 | Esophagus_Gastroesophageal_Junction |
| Brain_Caudate_basal_ganglia | CVD_ATH | 17388 | -0.00945 | -0.01635 | 0.007462 | 0.89741 | Brain_Caudate_basal_ganglia |
| Prostate | LEWY_grp | 17385 | -0.0148 | -0.02812 | 0.011681 | 0.89746 | Prostate |
| Breast_Mammary_Tissue | CVD_CAA | 17385 | -0.01498 | -0.02884 | 0.011802 | 0.89782 | Breast_Mammary_Tissue |
| Minor_Salivary_Gland | ADNC | 17388 | -0.01147 | -0.02137 | 0.009038 | 0.89784 | Minor_Salivary_Gland |
| Pancreas | CVD_ART_ANY | 17387 | -0.01019 | -0.01705 | 0.007985 | 0.89902 | Pancreas |
| Testis | AMY_ANY | 17377 | -0.00694 | -0.012 | 0.005435 | 0.8993 | Testis |

| | | | | | | | |
|---|---|---|---|---|---|---|---|
| Brain_Amygdala | INFA | 17386 | -0.0094 | -0.01602 | 0.007348 | 0.89947 | Brain_Amygdala |
| Brain_Frontal_Cortex_BA9 | ADNC | 17388 | -0.0084 | -0.01549 | 0.006556 | 0.89995 | Brain_Frontal_Cortex_BA9 |
| Brain_Frontal_Cortex_BA9 | BRAA | 17385 | -0.00841 | -0.01551 | 0.006544 | 0.90053 | Brain_Frontal_Cortex_BA9 |
| Brain_Cortex | BRAA | 17385 | -0.00872 | -0.01592 | 0.006785 | 0.90061 | Brain_Cortex |
| Brain_Anterior_cingulate_cor... | ADNC | 17388 | -0.00896 | -0.01585 | 0.006938 | 0.90178 | Brain_Anterior_cingulate_cortex_BA24 |
| Brain_Anterior_cingulate_cor... | CVD_ATH | 17388 | -0.00891 | -0.01575 | 0.006878 | 0.9023 | Brain_Anterior_cingulate_cortex_BA24 |
| Adrenal_Gland | BRAA | 17385 | -0.01239 | -0.02393 | 0.009543 | 0.90285 | Adrenal_Gland |
| Artery_Tibial | AMY_ANY | 17377 | -0.01255 | -0.02585 | 0.009625 | 0.90376 | Artery_Tibial |
| Minor_Salivary_Gland | B_SCORE | 17388 | -0.01175 | -0.02189 | 0.008982 | 0.90458 | Minor_Salivary_Gland |
| Brain_Putamen_basal_ganglia | HEMOR | 17378 | -0.00994 | -0.01693 | 0.007599 | 0.9046 | Brain_Putamen_basal_ganglia |
| Artery_Aorta | HS_S | 17386 | -0.01269 | -0.02591 | 0.009683 | 0.90491 | Artery_Aorta |
| Lung | CVD_CAA_ANY | 17386 | -0.01151 | -0.02228 | 0.008777 | 0.90514 | Lung |
| Whole_Blood | CVD_CAA | 17385 | -0.00721 | -0.01304 | 0.005447 | 0.90733 | Whole_Blood |
| Brain_Putamen_basal_ganglia | BRAA | 17385 | -0.01011 | -0.01722 | 0.007629 | 0.90751 | Brain_Putamen_basal_ganglia |
| Spleen | CVD_CAA_ANY | 17386 | -0.00941 | -0.01867 | 0.007099 | 0.90751 | Spleen |
| Whole_Blood | TDP43 | 17386 | -0.00738 | -0.01334 | 0.005559 | 0.90788 | Whole_Blood |
| Pancreas | BRAA | 17385 | -0.01091 | -0.01826 | 0.00821 | 0.90805 | Pancreas |
| Prostate | ADNC | 17388 | -0.01515 | -0.02878 | 0.011372 | 0.90863 | Prostate |
| Heart_Atrial_Appendage | HEMOR | 17378 | -0.01214 | -0.02181 | 0.009093 | 0.909 | Heart_Atrial_Appendage |
| Brain_Caudate_basal_ganglia | CVD_ART | 17388 | -0.01023 | -0.01769 | 0.007633 | 0.90993 | Brain_Caudate_basal_ganglia |
| Esophagus_Gastroesophageal_J... | LEWY_ANY | 17385 | -0.01634 | -0.0322 | 0.012185 | 0.91006 | Esophagus_Gastroesophageal_Junction |
| Brain_Spinal_cord_cervical_c... | CVD_CAA | 17385 | -0.01095 | -0.0197 | 0.008155 | 0.91031 | Brain_Spinal_cord_cervical_c-1 |
| Pancreas | AMY_ANY | 17377 | -0.01114 | -0.01864 | 0.008277 | 0.91084 | Pancreas |
| Brain_Putamen_basal_ganglia | CVD_ATH | 17388 | -0.01018 | -0.01734 | 0.007531 | 0.91175 | Brain_Putamen_basal_ganglia |
| Liver | B_SCORE | 17388 | -0.00828 | -0.01492 | 0.006027 | 0.91531 | Liver |
| Testis | BRAA | 17385 | -0.00746 | -0.01289 | 0.005423 | 0.91538 | Testis |
| Nerve_Tibial | HS_S | 17386 | -0.01326 | -0.02673 | 0.009611 | 0.91621 | Nerve_Tibial |
| Minor_Salivary_Gland | LEWY_full | 17385 | -0.01276 | -0.02378 | 0.009244 | 0.91629 | Minor_Salivary_Gland |
| Kidney_Cortex | MICR | 17385 | -0.0116 | -0.0204 | 0.008364 | 0.9172 | Kidney_Cortex |
| Brain_Hippocampus | INFA | 17386 | -0.01035 | -0.01754 | 0.007441 | 0.91778 | Brain_Hippocampus |
| Brain_Caudate_basal_ganglia | TDP43 | 17386 | -0.01075 | -0.01859 | 0.007689 | 0.91896 | Brain_Caudate_basal_ganglia |
| Brain_Amygdala | CVD_ATH | 17388 | -0.0105 | -0.0179 | 0.007469 | 0.92009 | Brain_Amygdala |
| Brain_Spinal_cord_cervical_c... | CVD_CAA_ANY | 17386 | -0.01158 | -0.02083 | 0.008227 | 0.92034 | Brain_Spinal_cord_cervical_c-1 |
| Liver | CVD_ATH_ANY | 17386 | -0.00865 | -0.01559 | 0.006091 | 0.92231 | Liver |
| Esophagus_Mucosa | CVD_CAA_ANY | 17386 | -0.01064 | -0.02059 | 0.007472 | 0.9228 | Esophagus_Mucosa |
| Brain_Hippocampus | TDP43 | 17386 | -0.01116 | -0.01892 | 0.007822 | 0.92317 | Brain_Hippocampus |

| | | | | | | | |
|---|---|---|---|---|---|---|---|
| Heart_Left_Ventricle | BRAA | 17385 | -0.0126 | -0.02105 | 0.0088 | 0.92389 | Heart_Left_Ventricle |
| Brain_Hypothalamus | CVD_ART | 17388 | -0.01121 | -0.01931 | 0.007801 | 0.92455 | Brain_Hypothalamus |
| Esophagus_Muscularis | LEWY_grp | 17385 | -0.017 | -0.03356 | 0.011831 | 0.92462 | Esophagus_Muscularis |
| Brain_Putamen_basal_ganglia | CVD_ART | 17388 | -0.01111 | -0.01891 | 0.007709 | 0.92515 | Brain_Putamen_basal_ganglia |
| Brain_Hypothalamus | AMY_CERAD | 17386 | -0.01133 | -0.01953 | 0.007853 | 0.92544 | Brain_Hypothalamus |
| Cervix_Endocervix | WBVD | 17388 | -0.01586 | -0.03144 | 0.010971 | 0.92586 | Cervix_Endocervix |
| Kidney_Cortex | LEWY_ANY | 17385 | -0.01255 | -0.02207 | 0.008646 | 0.92661 | Kidney_Cortex |
| Brain_Spinal_cord_cervical_c... | AMY_CERAD | 17386 | -0.01209 | -0.02176 | 0.008299 | 0.92747 | Brain_Spinal_cord_cervical_c-1 |
| Brain_Hippocampus | AMY_CERAD | 17386 | -0.01137 | -0.01928 | 0.007766 | 0.92841 | Brain_Hippocampus |
| Colon_Transverse | WBVD | 17388 | -0.016 | -0.02945 | 0.010887 | 0.92912 | Colon_Transverse |
| Pituitary | CVD_ATH | 17388 | -0.0127 | -0.0237 | 0.008569 | 0.9308 | Pituitary |
| Brain_Hypothalamus | TDP_3 | 17388 | -0.01166 | -0.02009 | 0.007839 | 0.93154 | Brain_Hypothalamus |
| Minor_Salivary_Gland | CVD_S | 17387 | -0.01345 | -0.02505 | 0.00902 | 0.93195 | Minor_Salivary_Gland |
| Brain_Hippocampus | CVD_ATH | 17388 | -0.0113 | -0.01915 | 0.007562 | 0.93239 | Brain_Hippocampus |
| Colon_Transverse | CVD_ART | 17388 | -0.01666 | -0.03067 | 0.011108 | 0.93318 | Colon_Transverse |
| Colon_Transverse | LEWY_full | 17385 | -0.0167 | -0.03074 | 0.011107 | 0.93366 | Colon_Transverse |
| Colon_Transverse | LEWY_ANY | 17385 | -0.01682 | -0.03096 | 0.011183 | 0.93368 | Colon_Transverse |
| Heart_Left_Ventricle | WMR | 17388 | -0.01324 | -0.02211 | 0.008783 | 0.93413 | Heart_Left_Ventricle |
| Brain_Substantia_nigra | AMY_CERAD | 17386 | -0.0127 | -0.02188 | 0.008369 | 0.93537 | Brain_Substantia_nigra |
| Cervix_Ectocervix | AMY_A | 17388 | -0.01765 | -0.03463 | 0.011632 | 0.93542 | Cervix_Ectocervix |
| Liver | AMY_ANY | 17377 | -0.00959 | -0.01728 | 0.006262 | 0.93715 | Liver |
| Esophagus_Muscularis | LEWY_ANY | 17385 | -0.01816 | -0.03586 | 0.011787 | 0.93833 | Esophagus_Muscularis |
| Uterus | AMY_A | 17388 | -0.01609 | -0.03282 | 0.010434 | 0.9385 | Uterus |
| Kidney_Cortex | CVD_ART_ANY | 17387 | -0.01313 | -0.02309 | 0.008508 | 0.93853 | Kidney_Cortex |
| Ovary | WBVD | 17388 | -0.01437 | -0.02915 | 0.009299 | 0.93886 | Ovary |
| Heart_Left_Ventricle | AMY_ANY | 17377 | -0.014 | -0.02338 | 0.008962 | 0.94085 | Heart_Left_Ventricle |
| Heart_Atrial_Appendage | WMR | 17388 | -0.01419 | -0.02549 | 0.009074 | 0.94102 | Heart_Atrial_Appendage |
| Adipose_Subcutaneous | CVD_CAA_ANY | 17386 | -0.016 | -0.03197 | 0.010144 | 0.94265 | Adipose_Subcutaneous |
| Esophagus_Gastroesophageal_J... | WBVD | 17388 | -0.01894 | -0.03731 | 0.011979 | 0.94301 | Esophagus_Gastroesophageal_Junction |
| Muscle_Skeletal | MICR | 17385 | -0.0103 | -0.01949 | 0.006481 | 0.94399 | Muscle_Skeletal |
| Cervix_Endocervix | AMY_A | 17388 | -0.01784 | -0.03536 | 0.011133 | 0.94549 | Cervix_Endocervix |
| Skin_Sun_Exposed_Lower_leg | CVD_ATH_ANY | 17386 | -0.01191 | -0.02313 | 0.007386 | 0.9465 | Skin_Sun_Exposed_Lower_leg |
| Heart_Atrial_Appendage | B_SCORE | 17388 | -0.01481 | -0.02661 | 0.009026 | 0.94956 | Heart_Atrial_Appendage |
| Brain_Cortex | AMY_CERAD | 17386 | -0.01147 | -0.02093 | 0.006906 | 0.95155 | Brain_Cortex |
| Brain_Amygdala | AMY_CERAD | 17386 | -0.0128 | -0.02183 | 0.007676 | 0.9523 | Brain_Amygdala |
| Nerve_Tibial | AMY_THAL | 17388 | -0.01615 | -0.03255 | 0.009679 | 0.95239 | Nerve_Tibial |

| | | | | | | | |
|---|---|---|---|---|---|---|---|
| Brain_Substantia_nigra | CVD_ART | 17388 | -0.01372 | -0.02365 | 0.008211 | 0.95266 | Brain_Substantia_nigra |
| Brain_Substantia_nigra | TDP_3 | 17388 | -0.01384 | -0.02384 | 0.008271 | 0.95282 | Brain_Substantia_nigra |
| Colon_Sigmoid | CVD_ART | 17388 | -0.02037 | -0.03989 | 0.011857 | 0.95708 | Colon_Sigmoid |
| Minor_Salivary_Gland | LEWY_ANY | 17385 | -0.01591 | -0.02963 | 0.009242 | 0.95735 | Minor_Salivary_Gland |
| Testis | WBVD | 17388 | -0.00918 | -0.01587 | 0.005294 | 0.95854 | Testis |
| Artery_Tibial | WMRCAL | 17388 | -0.01624 | -0.03346 | 0.009319 | 0.9593 | Artery_Tibial |
| Kidney_Cortex | LEWY_grp | 17385 | -0.01503 | -0.02644 | 0.008622 | 0.95938 | Kidney_Cortex |
| Cells_EBV-transformed_lympho... | LEWY_full | 17385 | -0.00817 | -0.01783 | 0.004668 | 0.95997 | Cells_EBV-transformed_lymphocytes |
| Muscle_Skeletal | LEWY_full | 17385 | -0.01175 | -0.02224 | 0.006706 | 0.96015 | Muscle_Skeletal |
| Pancreas | B_SCORE | 17388 | -0.01432 | -0.02396 | 0.008102 | 0.96141 | Pancreas |
| Heart_Atrial_Appendage | CVD_ART | 17388 | -0.01638 | -0.02944 | 0.009199 | 0.96254 | Heart_Atrial_Appendage |
| Brain_Hypothalamus | INFA | 17386 | -0.0134 | -0.02309 | 0.007516 | 0.96263 | Brain_Hypothalamus |
| Brain_Putamen_basal_ganglia | TDP43 | 17386 | -0.01384 | -0.02357 | 0.007759 | 0.96275 | Brain_Putamen_basal_ganglia |
| Brain_Anterior_cingulate_cor... | AMY_CERAD | 17386 | -0.01263 | -0.02232 | 0.007051 | 0.96331 | Brain_Anterior_cingulate_cortex_BA24 |
| Brain_Spinal_cord_cervical_c... | TDP_3 | 17388 | -0.01502 | -0.02702 | 0.008263 | 0.96544 | Brain_Spinal_cord_cervical_c-1 |
| Nerve_Tibial | WMRCAL | 17388 | -0.01747 | -0.03521 | 0.009384 | 0.96868 | Nerve_Tibial |
| Cells_Cultured_fibroblasts | B_SCORE | 17388 | -0.0118 | -0.02529 | 0.006317 | 0.96907 | Cells_Cultured_fibroblasts |
| Brain_Frontal_Cortex_BA9 | AMY_CERAD | 17386 | -0.01279 | -0.02358 | 0.006663 | 0.9725 | Brain_Frontal_Cortex_BA9 |
| Esophagus_Muscularis | WBVD | 17388 | -0.02233 | -0.04408 | 0.011625 | 0.97259 | Esophagus_Muscularis |
| Cells_Cultured_fibroblasts | BRAA | 17385 | -0.01216 | -0.02606 | 0.006329 | 0.97261 | Cells_Cultured_fibroblasts |
| Kidney_Cortex | LEWY_full | 17385 | -0.01643 | -0.0289 | 0.008549 | 0.97266 | Kidney_Cortex |
| Muscle_Skeletal | CVD_ART | 17388 | -0.01293 | -0.02446 | 0.006683 | 0.97345 | Muscle_Skeletal |
| Skin_Not_Sun_Exposed_Suprapu... | CVD_ATH_ANY | 17386 | -0.01427 | -0.02756 | 0.007366 | 0.97366 | Skin_Not_Sun_Exposed_Suprapubic |
| Brain_Substantia_nigra | INFA | 17386 | -0.01541 | -0.02656 | 0.007951 | 0.9737 | Brain_Substantia_nigra |
| Brain_Spinal_cord_cervical_c... | INFA | 17386 | -0.01561 | -0.02808 | 0.007952 | 0.97516 | Brain_Spinal_cord_cervical_c-1 |
| Liver | INFA | 17386 | -0.0118 | -0.02127 | 0.005928 | 0.97675 | Liver |
| Heart_Left_Ventricle | B_SCORE | 17388 | -0.01787 | -0.02985 | 0.008687 | 0.98015 | Heart_Left_Ventricle |
| Kidney_Medulla | LEWY_ANY | 17385 | -0.01909 | -0.03533 | 0.008949 | 0.98352 | Kidney_Medulla |
| Kidney_Medulla | LEWY_grp | 17385 | -0.01922 | -0.03557 | 0.008964 | 0.98397 | Kidney_Medulla |
| Kidney_Medulla | LEWY_full | 17385 | -0.01928 | -0.03568 | 0.008874 | 0.98507 | Kidney_Medulla |
| Brain_Hypothalamus | TDP43 | 17386 | -0.01743 | -0.03004 | 0.007947 | 0.98585 | Brain_Hypothalamus |
| Artery_Coronary | HS_S | 17386 | -0.02511 | -0.05003 | 0.011233 | 0.9873 | Artery_Coronary |
| Brain_Spinal_cord_cervical_c... | TDP43 | 17386 | -0.01955 | -0.03517 | 0.008286 | 0.99083 | Brain_Spinal_cord_cervical_c-1 |
| Brain_Substantia_nigra | TDP43 | 17386 | -0.02006 | -0.03457 | 0.008353 | 0.99182 | Brain_Substantia_nigra |
| Muscle_Skeletal | B_SCORE | 17388 | -0.01588 | -0.03005 | 0.006592 | 0.99199 | Muscle_Skeletal |
| Heart_Left_Ventricle | CVD_ART | 17388 | -0.0228 | -0.03809 | 0.008826 | 0.99511 | Heart_Left_Ventricle |

| | | | | | | | |
|---|---|---|---|---|---|---|---|
| Colon_Sigmoid | WBVD | 17388 | -0.03112 | -0.06094 | 0.011828 | 0.99574 | Colon_Sigmoid |
| Prostate | AMY_A | 17388 | -0.03154 | -0.05991 | 0.011434 | 0.99709 | Prostate |
| Prostate | AMY_THAL | 17388 | -0.03269 | -0.0621 | 0.011354 | 0.998 | Prostate |

**Table S17: MAGMA tissue-specific associations, general tissues**

| VARIABLE | PHENOTYPE | NGENES | BETA | BETA_STD | SE | P |
|---|---|---|---|---|---|---|
| Adipose_Tissue | ADNC | 17388 | 0.017029 | 0.033465 | 0.012582 | 0.08797 |
| Ovary | ADNC | 17388 | 0.013038 | 0.026451 | 0.009803 | 0.091781 |
| Fallopian_Tube | ADNC | 17388 | 0.014953 | 0.029044 | 0.013363 | 0.13157 |
| Testis | ADNC | 17388 | 0.005889 | 0.010179 | 0.005355 | 0.13574 |
| Lung | ADNC | 17388 | 0.010746 | 0.020799 | 0.009875 | 0.13828 |
| Stomach | ADNC | 17388 | 0.012181 | 0.022151 | 0.012687 | 0.16852 |
| Spleen | ADNC | 17388 | 0.007153 | 0.014187 | 0.007477 | 0.16937 |
| Blood_Vessel | ADNC | 17388 | 0.01064 | 0.021606 | 0.011209 | 0.17126 |
| Muscle | ADNC | 17388 | 0.004907 | 0.009285 | 0.006683 | 0.23139 |
| Uterus | ADNC | 17388 | 0.008327 | 0.016982 | 0.011577 | 0.23599 |
| Adrenal_Gland | ADNC | 17388 | 0.005855 | 0.011313 | 0.009723 | 0.27351 |
| Heart | ADNC | 17388 | 0.00552 | 0.009505 | 0.00925 | 0.27535 |
| Colon | ADNC | 17388 | 0.007272 | 0.013626 | 0.014767 | 0.31121 |
| Blood | ADNC | 17388 | 0.002763 | 0.004963 | 0.006018 | 0.32309 |
| Thyroid | ADNC | 17388 | 0.004228 | 0.008381 | 0.010433 | 0.34265 |
| Small_Intestine | ADNC | 17388 | -0.00118 | -0.00216 | 0.01024 | 0.5457 |
| Pancreas | ADNC | 17388 | -0.00146 | -0.00245 | 0.008528 | 0.56807 |
| Breast | ADNC | 17388 | -0.00477 | -0.00918 | 0.014773 | 0.62654 |
| Nerve | ADNC | 17388 | -0.00362 | -0.00729 | 0.009905 | 0.64261 |
| Kidney | ADNC | 17388 | -0.0034 | -0.00599 | 0.009038 | 0.64665 |
| Liver | ADNC | 17388 | -0.00332 | -0.00598 | 0.00633 | 0.70012 |
| Cervix_Uteri | ADNC | 17388 | -0.01091 | -0.02146 | 0.013652 | 0.78789 |
| Esophagus | ADNC | 17388 | -0.01407 | -0.02653 | 0.016388 | 0.80471 |
| Brain | ADNC | 17388 | -0.00624 | -0.01085 | 0.006611 | 0.82729 |
| Pituitary | ADNC | 17388 | -0.00766 | -0.01429 | 0.008044 | 0.82945 |
| Bladder | ADNC | 17388 | -0.01579 | -0.03081 | 0.015212 | 0.85033 |
| Vagina | ADNC | 17388 | -0.0213 | -0.04078 | 0.012163 | 0.96005 |
| Salivary_Gland | ADNC | 17388 | -0.02044 | -0.03807 | 0.010202 | 0.97741 |
| Skin | ADNC | 17388 | -0.0199 | -0.03799 | 0.009691 | 0.97998 |
| Prostate | ADNC | 17388 | -0.0294 | -0.05585 | 0.013118 | 0.9875 |
| Adrenal_Gland | AMY_A | 17388 | 0.01769 | 0.034178 | 0.009744 | 0.034742 |

| | | | | | | |
|---|---|---|---|---|---|---|
| Muscle | AMY_A | 17388 | 0.007966 | 0.015074 | 0.006672 | 0.11624 |
| Small_Intestine | AMY_A | 17388 | 0.011586 | 0.02133 | 0.010227 | 0.12864 |
| Kidney | AMY_A | 17388 | 0.009678 | 0.017045 | 0.008984 | 0.14071 |
| Spleen | AMY_A | 17388 | 0.00749 | 0.014856 | 0.007568 | 0.16117 |
| Pancreas | AMY_A | 17388 | 0.008296 | 0.013882 | 0.008407 | 0.16188 |
| Stomach | AMY_A | 17388 | 0.012179 | 0.022148 | 0.012865 | 0.17192 |
| Esophagus | AMY_A | 17388 | 0.01426 | 0.026891 | 0.016348 | 0.19154 |
| Blood | AMY_A | 17388 | 0.005121 | 0.009201 | 0.006017 | 0.19735 |
| Lung | AMY_A | 17388 | 0.006015 | 0.011642 | 0.010095 | 0.27566 |
| Salivary_Gland | AMY_A | 17388 | 0.00595 | 0.011084 | 0.010198 | 0.27982 |
| Colon | AMY_A | 17388 | 0.006779 | 0.012702 | 0.015104 | 0.32679 |
| Adipose_Tissue | AMY_A | 17388 | 0.00546 | 0.010729 | 0.012636 | 0.33284 |
| Thyroid | AMY_A | 17388 | 0.003604 | 0.007145 | 0.010425 | 0.36477 |
| Breast | AMY_A | 17388 | 0.001419 | 0.002732 | 0.014753 | 0.46168 |
| Heart | AMY_A | 17388 | 7.06E-05 | 0.000122 | 0.009278 | 0.49696 |
| Liver | AMY_A | 17388 | -2.26E-05 | -4.08E-05 | 0.006361 | 0.50142 |
| Pituitary | AMY_A | 17388 | -0.00013 | -0.00024 | 0.008044 | 0.50647 |
| Ovary | AMY_A | 17388 | -0.00058 | -0.00117 | 0.009829 | 0.52337 |
| Blood_Vessel | AMY_A | 17388 | -0.003 | -0.00609 | 0.01115 | 0.6061 |
| Testis | AMY_A | 17388 | -0.00162 | -0.0028 | 0.005362 | 0.61879 |
| Brain | AMY_A | 17388 | -0.00255 | -0.00443 | 0.006647 | 0.64912 |
| Fallopian_Tube | AMY_A | 17388 | -0.01471 | -0.02857 | 0.013465 | 0.86269 |
| Skin | AMY_A | 17388 | -0.01122 | -0.02141 | 0.009798 | 0.87383 |
| Nerve | AMY_A | 17388 | -0.01257 | -0.02533 | 0.009933 | 0.89714 |
| Vagina | AMY_A | 17388 | -0.02137 | -0.0409 | 0.012384 | 0.95775 |
| Bladder | AMY_A | 17388 | -0.02879 | -0.05618 | 0.0153 | 0.97004 |
| Uterus | AMY_A | 17388 | -0.02273 | -0.04634 | 0.011486 | 0.97605 |
| Cervix_Uteri | AMY_A | 17388 | -0.02981 | -0.05862 | 0.013566 | 0.98598 |
| Prostate | AMY_A | 17388 | -0.04538 | -0.0862 | 0.013087 | 0.99974 |
| Thyroid | AMY_ANY | 17377 | 0.017024 | 0.03375 | 0.010609 | 0.054296 |
| Vagina | AMY_ANY | 17377 | 0.01836 | 0.035147 | 0.012411 | 0.069532 |
| Pituitary | AMY_ANY | 17377 | 0.011194 | 0.020894 | 0.008219 | 0.086603 |
| Prostate | AMY_ANY | 17377 | 0.016585 | 0.031504 | 0.013339 | 0.10688 |
| Skin | AMY_ANY | 17377 | 0.011132 | 0.021248 | 0.009793 | 0.12783 |

| | | | | | | |
|---|---|---|---|---|---|---|
| Brain | AMY_ANY | 17377 | 0.007099 | 0.012343 | 0.006721 | 0.14545 |
| Lung | AMY_ANY | 17377 | 0.008239 | 0.015947 | 0.01014 | 0.20826 |
| Adrenal_Gland | AMY_ANY | 17377 | 0.007109 | 0.013736 | 0.009868 | 0.23564 |
| Fallopian_Tube | AMY_ANY | 17377 | 0.007595 | 0.014751 | 0.013483 | 0.28663 |
| Cervix_Uteri | AMY_ANY | 17377 | 0.00734 | 0.014435 | 0.01379 | 0.29727 |
| Blood | AMY_ANY | 17377 | 0.002795 | 0.005021 | 0.006015 | 0.3211 |
| Kidney | AMY_ANY | 17377 | 0.003962 | 0.006978 | 0.009017 | 0.3302 |
| Bladder | AMY_ANY | 17377 | 0.004608 | 0.008992 | 0.015578 | 0.38369 |
| Salivary_Gland | AMY_ANY | 17377 | 0.002481 | 0.004621 | 0.010219 | 0.40411 |
| Breast | AMY_ANY | 17377 | 0.001819 | 0.003502 | 0.015028 | 0.45184 |
| Nerve | AMY_ANY | 17377 | 0.00092 | 0.001855 | 0.01007 | 0.46359 |
| Small_Intestine | AMY_ANY | 17377 | -0.00073 | -0.00134 | 0.010251 | 0.52836 |
| Colon | AMY_ANY | 17377 | -0.0026 | -0.00488 | 0.015285 | 0.56765 |
| Esophagus | AMY_ANY | 17377 | -0.00384 | -0.00724 | 0.016711 | 0.59081 |
| Uterus | AMY_ANY | 17377 | -0.00331 | -0.00675 | 0.011632 | 0.61204 |
| Stomach | AMY_ANY | 17377 | -0.00405 | -0.00736 | 0.013211 | 0.62028 |
| Spleen | AMY_ANY | 17377 | -0.00369 | -0.00732 | 0.007613 | 0.68614 |
| Blood_Vessel | AMY_ANY | 17377 | -0.00835 | -0.01695 | 0.011343 | 0.76909 |
| Adipose_Tissue | AMY_ANY | 17377 | -0.01007 | -0.01979 | 0.012948 | 0.78162 |
| Ovary | AMY_ANY | 17377 | -0.00945 | -0.01918 | 0.009902 | 0.83008 |
| Muscle | AMY_ANY | 17377 | -0.00689 | -0.01305 | 0.006891 | 0.84144 |
| Testis | AMY_ANY | 17377 | -0.0064 | -0.01106 | 0.005447 | 0.87985 |
| Heart | AMY_ANY | 17377 | -0.0114 | -0.01964 | 0.009446 | 0.88629 |
| Pancreas | AMY_ANY | 17377 | -0.01057 | -0.0177 | 0.008627 | 0.88982 |
| Liver | AMY_ANY | 17377 | -0.00952 | -0.01715 | 0.006478 | 0.92916 |
| Lung | AMY_CERAD | 17386 | 0.020993 | 0.040634 | 0.01009 | 0.018743 |
| Blood | AMY_CERAD | 17386 | 0.009127 | 0.016397 | 0.006037 | 0.065308 |
| Prostate | AMY_CERAD | 17386 | 0.018399 | 0.034949 | 0.013331 | 0.083792 |
| Esophagus | AMY_CERAD | 17386 | 0.018509 | 0.034904 | 0.016435 | 0.13005 |
| Kidney | AMY_CERAD | 17386 | 0.007304 | 0.012864 | 0.009042 | 0.20961 |
| Bladder | AMY_CERAD | 17386 | 0.009493 | 0.018524 | 0.015385 | 0.2686 |
| Adrenal_Gland | AMY_CERAD | 17386 | 0.00571 | 0.011033 | 0.009903 | 0.2821 |
| Spleen | AMY_CERAD | 17386 | 0.004125 | 0.008182 | 0.00759 | 0.29339 |
| Salivary_Gland | AMY_CERAD | 17386 | 0.005551 | 0.010342 | 0.010274 | 0.2945 |

| | | | | | | |
|---|---|---|---|---|---|---|
| Fallopian_Tube | AMY_CERAD | 17386 | 0.005487 | 0.010657 | 0.013559 | 0.34287 |
| Adipose_Tissue | AMY_CERAD | 17386 | 0.004799 | 0.009431 | 0.012859 | 0.3545 |
| Breast | AMY_CERAD | 17386 | 0.005558 | 0.0107 | 0.014978 | 0.3553 |
| Small_Intestine | AMY_CERAD | 17386 | 0.002869 | 0.005281 | 0.010354 | 0.39088 |
| Thyroid | AMY_CERAD | 17386 | 0.002506 | 0.004967 | 0.010627 | 0.40681 |
| Stomach | AMY_CERAD | 17386 | 0.002671 | 0.004858 | 0.013107 | 0.41925 |
| Vagina | AMY_CERAD | 17386 | 0.00146 | 0.002796 | 0.012332 | 0.45287 |
| Skin | AMY_CERAD | 17386 | 0.000223 | 0.000426 | 0.009831 | 0.49095 |
| Colon | AMY_CERAD | 17386 | -0.00113 | -0.00211 | 0.015017 | 0.52995 |
| Nerve | AMY_CERAD | 17386 | -0.00289 | -0.00583 | 0.010083 | 0.61295 |
| Testis | AMY_CERAD | 17386 | -0.00191 | -0.0033 | 0.005482 | 0.63602 |
| Pituitary | AMY_CERAD | 17386 | -0.00291 | -0.00543 | 0.008112 | 0.64017 |
| Uterus | AMY_CERAD | 17386 | -0.00466 | -0.0095 | 0.011787 | 0.6537 |
| Pancreas | AMY_CERAD | 17386 | -0.00355 | -0.00595 | 0.008665 | 0.65915 |
| Blood_Vessel | AMY_CERAD | 17386 | -0.0071 | -0.01442 | 0.011392 | 0.73352 |
| Cervix_Uteri | AMY_CERAD | 17386 | -0.00983 | -0.01934 | 0.013907 | 0.76025 |
| Liver | AMY_CERAD | 17386 | -0.00481 | -0.00867 | 0.006385 | 0.77454 |
| Heart | AMY_CERAD | 17386 | -0.00785 | -0.01352 | 0.00947 | 0.79649 |
| Muscle | AMY_CERAD | 17386 | -0.00851 | -0.0161 | 0.006808 | 0.89436 |
| Brain | AMY_CERAD | 17386 | -0.00982 | -0.01707 | 0.006707 | 0.92835 |
| Ovary | AMY_CERAD | 17386 | -0.01622 | -0.03292 | 0.009904 | 0.94929 |
| Esophagus | AMY_THAL | 17388 | 0.033892 | 0.063911 | 0.016377 | 0.01926 |
| Colon | AMY_THAL | 17388 | 0.025407 | 0.047608 | 0.015108 | 0.046329 |
| Small_Intestine | AMY_THAL | 17388 | 0.01253 | 0.023068 | 0.010375 | 0.11361 |
| Adrenal_Gland | AMY_THAL | 17388 | 0.010338 | 0.019974 | 0.009714 | 0.14361 |
| Muscle | AMY_THAL | 17388 | 0.006347 | 0.012009 | 0.006678 | 0.17098 |
| Brain | AMY_THAL | 17388 | 0.00569 | 0.009893 | 0.006609 | 0.19463 |
| Blood | AMY_THAL | 17388 | 0.004314 | 0.007751 | 0.006069 | 0.23858 |
| Spleen | AMY_THAL | 17388 | 0.005049 | 0.010015 | 0.00757 | 0.25238 |
| Ovary | AMY_THAL | 17388 | 0.00526 | 0.010671 | 0.009751 | 0.29481 |
| Fallopian_Tube | AMY_THAL | 17388 | 0.004621 | 0.008976 | 0.013392 | 0.36502 |
| Testis | AMY_THAL | 17388 | 0.001855 | 0.003206 | 0.005428 | 0.36627 |
| Blood_Vessel | AMY_THAL | 17388 | 0.003425 | 0.006955 | 0.011302 | 0.38093 |
| Heart | AMY_THAL | 17388 | 0.00131 | 0.002257 | 0.009265 | 0.44376 |

| | | | | | | |
|---|---|---|---|---|---|---|
| Pituitary | AMY_THAL | 17388 | 0.000894 | 0.001668 | 0.00802 | 0.45564 |
| Lung | AMY_THAL | 17388 | 0.000773 | 0.001496 | 0.010083 | 0.46945 |
| Thyroid | AMY_THAL | 17388 | 0.000348 | 0.00069 | 0.010293 | 0.48651 |
| Stomach | AMY_THAL | 17388 | -0.00074 | -0.00134 | 0.01283 | 0.52294 |
| Adipose_Tissue | AMY_THAL | 17388 | -0.00237 | -0.00465 | 0.012662 | 0.57413 |
| Bladder | AMY_THAL | 17388 | -0.0049 | -0.00955 | 0.015284 | 0.62562 |
| Vagina | AMY_THAL | 17388 | -0.00431 | -0.00824 | 0.012189 | 0.63805 |
| Salivary_Gland | AMY_THAL | 17388 | -0.00735 | -0.0137 | 0.010156 | 0.76543 |
| Cervix_Uteri | AMY_THAL | 17388 | -0.00979 | -0.01926 | 0.013513 | 0.76568 |
| Kidney | AMY_THAL | 17388 | -0.00675 | -0.01188 | 0.008877 | 0.77638 |
| Liver | AMY_THAL | 17388 | -0.00501 | -0.00903 | 0.006358 | 0.78478 |
| Uterus | AMY_THAL | 17388 | -0.00935 | -0.01907 | 0.011574 | 0.79046 |
| Breast | AMY_THAL | 17388 | -0.0132 | -0.02542 | 0.01472 | 0.81514 |
| Pancreas | AMY_THAL | 17388 | -0.00864 | -0.01446 | 0.008545 | 0.84411 |
| Skin | AMY_THAL | 17388 | -0.01128 | -0.02153 | 0.009692 | 0.87771 |
| Nerve | AMY_THAL | 17388 | -0.01473 | -0.02969 | 0.01003 | 0.92905 |
| Prostate | AMY_THAL | 17388 | -0.03972 | -0.07544 | 0.013051 | 0.99883 |
| Spleen | B_SCORE | 17388 | 0.03217 | 0.063807 | 0.007539 | 9.98E-06 |
| Lung | B_SCORE | 17388 | 0.033489 | 0.064821 | 0.010075 | 0.000445 |
| Blood | B_SCORE | 17388 | 0.015367 | 0.027608 | 0.005954 | 0.004934 |
| Thyroid | B_SCORE | 17388 | 0.022144 | 0.043899 | 0.010401 | 0.016635 |
| Fallopian_Tube | B_SCORE | 17388 | 0.014152 | 0.027487 | 0.013178 | 0.14144 |
| Pituitary | B_SCORE | 17388 | 0.007535 | 0.014064 | 0.007899 | 0.17009 |
| Testis | B_SCORE | 17388 | 0.004867 | 0.008412 | 0.005371 | 0.18246 |
| Uterus | B_SCORE | 17388 | 0.010315 | 0.021035 | 0.011505 | 0.185 |
| Nerve | B_SCORE | 17388 | 0.007992 | 0.016103 | 0.009863 | 0.2089 |
| Cervix_Uteri | B_SCORE | 17388 | 0.010688 | 0.021019 | 0.013598 | 0.21594 |
| Prostate | B_SCORE | 17388 | 0.009879 | 0.018766 | 0.013324 | 0.2292 |
| Vagina | B_SCORE | 17388 | 0.002518 | 0.00482 | 0.012173 | 0.41808 |
| Kidney | B_SCORE | 17388 | 0.001268 | 0.002233 | 0.008787 | 0.44265 |
| Small_Intestine | B_SCORE | 17388 | -0.0008 | -0.00148 | 0.010285 | 0.5311 |
| Bladder | B_SCORE | 17388 | -0.00223 | -0.00435 | 0.015048 | 0.55889 |
| Blood_Vessel | B_SCORE | 17388 | -0.00527 | -0.01071 | 0.011251 | 0.6804 |
| Brain | B_SCORE | 17388 | -0.00348 | -0.00606 | 0.006544 | 0.70281 |

| | | | | | | |
|---|---|---|---|---|---|---|
| Ovary | B_SCORE | 17388 | -0.00641 | -0.01301 | 0.009757 | 0.74438 |
| Adipose_Tissue | B_SCORE | 17388 | -0.00944 | -0.01855 | 0.012646 | 0.77233 |
| Adrenal_Gland | B_SCORE | 17388 | -0.00782 | -0.01511 | 0.00972 | 0.78948 |
| Stomach | B_SCORE | 17388 | -0.01033 | -0.01879 | 0.012639 | 0.79311 |
| Esophagus | B_SCORE | 17388 | -0.01759 | -0.03317 | 0.016187 | 0.86139 |
| Colon | B_SCORE | 17388 | -0.02036 | -0.03815 | 0.014737 | 0.91643 |
| Breast | B_SCORE | 17388 | -0.02061 | -0.03968 | 0.014653 | 0.92022 |
| Liver | B_SCORE | 17388 | -0.01016 | -0.0183 | 0.006231 | 0.94842 |
| Salivary_Gland | B_SCORE | 17388 | -0.01915 | -0.03568 | 0.010136 | 0.97058 |
| Skin | B_SCORE | 17388 | -0.01877 | -0.03583 | 0.009706 | 0.97342 |
| Pancreas | B_SCORE | 17388 | -0.01787 | -0.02991 | 0.008439 | 0.9829 |
| Heart | B_SCORE | 17388 | -0.02053 | -0.03535 | 0.009207 | 0.98711 |
| Muscle | B_SCORE | 17388 | -0.01751 | -0.03313 | 0.00662 | 0.99591 |
| Lung | BRAA | 17385 | 0.038412 | 0.07435 | 0.010099 | 7.16E-05 |
| Spleen | BRAA | 17385 | 0.024606 | 0.048807 | 0.007561 | 0.00057 |
| Blood | BRAA | 17385 | 0.013268 | 0.023838 | 0.005992 | 0.013417 |
| Thyroid | BRAA | 17385 | 0.016413 | 0.032538 | 0.0104 | 0.057265 |
| Bladder | BRAA | 17385 | 0.023582 | 0.046013 | 0.015403 | 0.062896 |
| Small_Intestine | BRAA | 17385 | 0.015297 | 0.028164 | 0.010276 | 0.0683 |
| Kidney | BRAA | 17385 | 0.010553 | 0.018586 | 0.008957 | 0.11937 |
| Salivary_Gland | BRAA | 17385 | 0.008981 | 0.016731 | 0.010268 | 0.19089 |
| Vagina | BRAA | 17385 | 0.010607 | 0.020305 | 0.012379 | 0.19577 |
| Prostate | BRAA | 17385 | 0.009022 | 0.017137 | 0.013406 | 0.25049 |
| Nerve | BRAA | 17385 | 0.004327 | 0.008718 | 0.009899 | 0.33104 |
| Colon | BRAA | 17385 | 0.005694 | 0.010669 | 0.014926 | 0.35144 |
| Esophagus | BRAA | 17385 | 0.005457 | 0.010289 | 0.016313 | 0.36901 |
| Stomach | BRAA | 17385 | 0.004122 | 0.007497 | 0.012835 | 0.37404 |
| Fallopian_Tube | BRAA | 17385 | -0.00459 | -0.00891 | 0.013473 | 0.63328 |
| Blood_Vessel | BRAA | 17385 | -0.004 | -0.00812 | 0.011301 | 0.63824 |
| Cervix_Uteri | BRAA | 17385 | -0.00754 | -0.01483 | 0.01367 | 0.7094 |
| Pituitary | BRAA | 17385 | -0.00539 | -0.01005 | 0.008063 | 0.74791 |
| Adipose_Tissue | BRAA | 17385 | -0.00929 | -0.01826 | 0.012649 | 0.76865 |
| Liver | BRAA | 17385 | -0.00496 | -0.00894 | 0.00643 | 0.77974 |
| Brain | BRAA | 17385 | -0.00662 | -0.01151 | 0.00658 | 0.84275 |

| | | | | | | |
|---|---|---|---|---|---|---|
| Uterus | BRAA | 17385 | -0.01236 | -0.02521 | 0.01156 | 0.85751 |
| Muscle | BRAA | 17385 | -0.00736 | -0.01392 | 0.006685 | 0.8644 |
| Breast | BRAA | 17385 | -0.01975 | -0.03801 | 0.014804 | 0.90885 |
| Skin | BRAA | 17385 | -0.01474 | -0.02813 | 0.00988 | 0.93209 |
| Testis | BRAA | 17385 | -0.00829 | -0.01433 | 0.005433 | 0.93648 |
| Heart | BRAA | 17385 | -0.01561 | -0.02689 | 0.009343 | 0.95264 |
| Pancreas | BRAA | 17385 | -0.0145 | -0.02426 | 0.008551 | 0.95501 |
| Ovary | BRAA | 17385 | -0.01727 | -0.03503 | 0.009867 | 0.95992 |
| Adrenal_Gland | BRAA | 17385 | -0.01744 | -0.0337 | 0.009872 | 0.96136 |
| Breast | CVD_ART | 17388 | 0.036911 | 0.071058 | 0.014992 | 0.006915 |
| Thyroid | CVD_ART | 17388 | 0.022756 | 0.045112 | 0.010634 | 0.016191 |
| Liver | CVD_ART | 17388 | 0.010442 | 0.018812 | 0.006328 | 0.049474 |
| Lung | CVD_ART | 17388 | 0.014771 | 0.028591 | 0.010082 | 0.071451 |
| Adipose_Tissue | CVD_ART | 17388 | 0.017914 | 0.035204 | 0.012819 | 0.081154 |
| Testis | CVD_ART | 17388 | 0.006516 | 0.011263 | 0.005398 | 0.11369 |
| Salivary_Gland | CVD_ART | 17388 | 0.011718 | 0.02183 | 0.010429 | 0.1306 |
| Spleen | CVD_ART | 17388 | 0.008147 | 0.01616 | 0.007623 | 0.1426 |
| Adrenal_Gland | CVD_ART | 17388 | 0.009986 | 0.019293 | 0.009701 | 0.15167 |
| Blood_Vessel | CVD_ART | 17388 | 0.007282 | 0.014787 | 0.011366 | 0.26087 |
| Fallopian_Tube | CVD_ART | 17388 | 0.006023 | 0.011698 | 0.013475 | 0.32746 |
| Blood | CVD_ART | 17388 | 0.001377 | 0.002475 | 0.006 | 0.40921 |
| Pancreas | CVD_ART | 17388 | 0.000966 | 0.001616 | 0.008462 | 0.45456 |
| Stomach | CVD_ART | 17388 | 0.000771 | 0.001402 | 0.013058 | 0.47647 |
| Pituitary | CVD_ART | 17388 | 0.000383 | 0.000714 | 0.008115 | 0.48119 |
| Kidney | CVD_ART | 17388 | -0.00028 | -0.0005 | 0.008944 | 0.51266 |
| Ovary | CVD_ART | 17388 | -0.00116 | -0.00235 | 0.009923 | 0.54638 |
| Prostate | CVD_ART | 17388 | -0.003 | -0.0057 | 0.013516 | 0.58792 |
| Skin | CVD_ART | 17388 | -0.003 | -0.00573 | 0.009823 | 0.61998 |
| Nerve | CVD_ART | 17388 | -0.00408 | -0.00821 | 0.009996 | 0.65824 |
| Small_Intestine | CVD_ART | 17388 | -0.0061 | -0.01123 | 0.010403 | 0.72113 |
| Cervix_Uteri | CVD_ART | 17388 | -0.00894 | -0.01759 | 0.013712 | 0.74289 |
| Uterus | CVD_ART | 17388 | -0.00898 | -0.01832 | 0.011741 | 0.77789 |
| Bladder | CVD_ART | 17388 | -0.01537 | -0.02999 | 0.015532 | 0.83882 |
| Brain | CVD_ART | 17388 | -0.00764 | -0.01328 | 0.006682 | 0.87352 |

| | | | | | | |
|---|---|---|---|---|---|---|
| Esophagus | CVD_ART | 17388 | -0.02912 | -0.05491 | 0.016464 | 0.96152 |
| Vagina | CVD_ART | 17388 | -0.02434 | -0.0466 | 0.012181 | 0.97715 |
| Muscle | CVD_ART | 17388 | -0.01522 | -0.0288 | 0.006688 | 0.98855 |
| Heart | CVD_ART | 17388 | -0.02565 | -0.04416 | 0.009357 | 0.99693 |
| Colon | CVD_ART | 17388 | -0.04485 | -0.08403 | 0.015049 | 0.99856 |
| Small_Intestine | CVD_ART_ANY | 17387 | 0.01329 | 0.024467 | 0.01028 | 0.098039 |
| Pituitary | CVD_ART_ANY | 17387 | 0.008416 | 0.015708 | 0.008083 | 0.14891 |
| Thyroid | CVD_ART_ANY | 17387 | 0.010244 | 0.020306 | 0.01053 | 0.16535 |
| Spleen | CVD_ART_ANY | 17387 | 0.006874 | 0.013634 | 0.00753 | 0.18067 |
| Testis | CVD_ART_ANY | 17387 | 0.004076 | 0.007045 | 0.005349 | 0.22302 |
| Cervix_Uteri | CVD_ART_ANY | 17387 | 0.01038 | 0.020413 | 0.013624 | 0.22305 |
| Blood | CVD_ART_ANY | 17387 | 0.00279 | 0.005013 | 0.005954 | 0.31966 |
| Blood_Vessel | CVD_ART_ANY | 17387 | 0.004708 | 0.00956 | 0.011261 | 0.33794 |
| Adrenal_Gland | CVD_ART_ANY | 17387 | 0.003164 | 0.006113 | 0.009624 | 0.37118 |
| Adipose_Tissue | CVD_ART_ANY | 17387 | 0.004107 | 0.00807 | 0.012541 | 0.37165 |
| Ovary | CVD_ART_ANY | 17387 | 0.002426 | 0.004922 | 0.00977 | 0.40194 |
| Salivary_Gland | CVD_ART_ANY | 17387 | 0.00216 | 0.004023 | 0.010273 | 0.41675 |
| Nerve | CVD_ART_ANY | 17387 | 0.001739 | 0.003503 | 0.009847 | 0.42992 |
| Muscle | CVD_ART_ANY | 17387 | 0.000787 | 0.001489 | 0.006582 | 0.45243 |
| Brain | CVD_ART_ANY | 17387 | 0.000737 | 0.001281 | 0.006629 | 0.45574 |
| Skin | CVD_ART_ANY | 17387 | 0.000885 | 0.00169 | 0.009667 | 0.46351 |
| Breast | CVD_ART_ANY | 17387 | 5.71E-05 | 0.00011 | 0.014673 | 0.49845 |
| Lung | CVD_ART_ANY | 17387 | -0.00121 | -0.00234 | 0.009971 | 0.54819 |
| Vagina | CVD_ART_ANY | 17387 | -0.00182 | -0.00348 | 0.012266 | 0.55883 |
| Uterus | CVD_ART_ANY | 17387 | -0.00208 | -0.00425 | 0.01151 | 0.57185 |
| Esophagus | CVD_ART_ANY | 17387 | -0.00791 | -0.01492 | 0.016278 | 0.68652 |
| Fallopian_Tube | CVD_ART_ANY | 17387 | -0.00687 | -0.01334 | 0.013253 | 0.6979 |
| Prostate | CVD_ART_ANY | 17387 | -0.00817 | -0.01552 | 0.013193 | 0.73209 |
| Bladder | CVD_ART_ANY | 17387 | -0.01012 | -0.01975 | 0.015183 | 0.74751 |
| Colon | CVD_ART_ANY | 17387 | -0.01218 | -0.02282 | 0.014937 | 0.79256 |
| Heart | CVD_ART_ANY | 17387 | -0.00757 | -0.01304 | 0.009159 | 0.79588 |
| Stomach | CVD_ART_ANY | 17387 | -0.01362 | -0.02476 | 0.013006 | 0.85245 |
| Liver | CVD_ART_ANY | 17387 | -0.00735 | -0.01323 | 0.006361 | 0.8759 |
| Pancreas | CVD_ART_ANY | 17387 | -0.01126 | -0.01885 | 0.008323 | 0.91199 |

| | | | | | | |
|---|---|---|---|---|---|---|
| Kidney | CVD_ART_ANY | 17387 | -0.01457 | -0.02567 | 0.00895 | 0.94827 |
| Lung | CVD_ATH | 17388 | 0.018327 | 0.035473 | 0.009784 | 0.030536 |
| Bladder | CVD_ATH | 17388 | 0.027734 | 0.054116 | 0.015188 | 0.033933 |
| Cervix_Uteri | CVD_ATH | 17388 | 0.019612 | 0.038569 | 0.0135 | 0.073164 |
| Vagina | CVD_ATH | 17388 | 0.014324 | 0.027421 | 0.012159 | 0.11941 |
| Spleen | CVD_ATH | 17388 | 0.008478 | 0.016816 | 0.007566 | 0.13126 |
| Blood_Vessel | CVD_ATH | 17388 | 0.011235 | 0.022815 | 0.011195 | 0.1578 |
| Fallopian_Tube | CVD_ATH | 17388 | 0.010048 | 0.019517 | 0.013275 | 0.22455 |
| Uterus | CVD_ATH | 17388 | 0.008415 | 0.017162 | 0.011485 | 0.23186 |
| Blood | CVD_ATH | 17388 | 0.002649 | 0.004758 | 0.005897 | 0.32667 |
| Adipose_Tissue | CVD_ATH | 17388 | 0.004008 | 0.007876 | 0.012641 | 0.37562 |
| Nerve | CVD_ATH | 17388 | 0.00289 | 0.005823 | 0.009666 | 0.38248 |
| Ovary | CVD_ATH | 17388 | 0.001987 | 0.004032 | 0.009761 | 0.41934 |
| Heart | CVD_ATH | 17388 | 0.001216 | 0.002094 | 0.009204 | 0.44746 |
| Adrenal_Gland | CVD_ATH | 17388 | 0.001029 | 0.001987 | 0.009598 | 0.45733 |
| Stomach | CVD_ATH | 17388 | 0.001141 | 0.002076 | 0.012908 | 0.46477 |
| Muscle | CVD_ATH | 17388 | 0.000544 | 0.001029 | 0.006564 | 0.46698 |
| Esophagus | CVD_ATH | 17388 | 0.001333 | 0.002513 | 0.016333 | 0.46748 |
| Thyroid | CVD_ATH | 17388 | 0.000113 | 0.000223 | 0.010555 | 0.49575 |
| Prostate | CVD_ATH | 17388 | -0.00193 | -0.00367 | 0.013259 | 0.55786 |
| Small_Intestine | CVD_ATH | 17388 | -0.00202 | -0.00372 | 0.010283 | 0.57791 |
| Salivary_Gland | CVD_ATH | 17388 | -0.00331 | -0.00616 | 0.010273 | 0.6263 |
| Skin | CVD_ATH | 17388 | -0.00439 | -0.00838 | 0.009637 | 0.67559 |
| Testis | CVD_ATH | 17388 | -0.00307 | -0.00531 | 0.005283 | 0.71966 |
| Colon | CVD_ATH | 17388 | -0.00914 | -0.01713 | 0.014996 | 0.72892 |
| Pancreas | CVD_ATH | 17388 | -0.00583 | -0.00976 | 0.008298 | 0.75896 |
| Breast | CVD_ATH | 17388 | -0.01105 | -0.02127 | 0.014763 | 0.77292 |
| Liver | CVD_ATH | 17388 | -0.00627 | -0.01131 | 0.006312 | 0.8399 |
| Brain | CVD_ATH | 17388 | -0.00777 | -0.01351 | 0.00656 | 0.88186 |
| Kidney | CVD_ATH | 17388 | -0.01105 | -0.01945 | 0.008996 | 0.89023 |
| Pituitary | CVD_ATH | 17388 | -0.01342 | -0.02504 | 0.008007 | 0.95307 |
| Lung | CVD_ATH_ANY | 17386 | 0.020897 | 0.040448 | 0.009929 | 0.017674 |
| Spleen | CVD_ATH_ANY | 17386 | 0.012025 | 0.023851 | 0.007527 | 0.055086 |
| Blood | CVD_ATH_ANY | 17386 | 0.008971 | 0.016118 | 0.005943 | 0.065579 |

| | | | | | | |
|---|---|---|---|---|---|---|
| Ovary | CVD_ATH_ANY | 17386 | 0.013444 | 0.027275 | 0.009892 | 0.087085 |
| Uterus | CVD_ATH_ANY | 17386 | 0.013818 | 0.028181 | 0.011403 | 0.11281 |
| Cervix_Uteri | CVD_ATH_ANY | 17386 | 0.015461 | 0.030407 | 0.013567 | 0.12724 |
| Fallopian_Tube | CVD_ATH_ANY | 17386 | 0.012351 | 0.023989 | 0.013193 | 0.17461 |
| Adrenal_Gland | CVD_ATH_ANY | 17386 | 0.008577 | 0.016571 | 0.009647 | 0.18699 |
| Adipose_Tissue | CVD_ATH_ANY | 17386 | 0.006713 | 0.013193 | 0.012633 | 0.29757 |
| Testis | CVD_ATH_ANY | 17386 | 0.002824 | 0.004882 | 0.005388 | 0.30008 |
| Small_Intestine | CVD_ATH_ANY | 17386 | 0.004016 | 0.007394 | 0.010229 | 0.34731 |
| Pancreas | CVD_ATH_ANY | 17386 | 0.002104 | 0.003521 | 0.008388 | 0.40097 |
| Blood_Vessel | CVD_ATH_ANY | 17386 | 0.001254 | 0.002546 | 0.011118 | 0.4551 |
| Bladder | CVD_ATH_ANY | 17386 | 0.000285 | 0.000556 | 0.015321 | 0.49258 |
| Vagina | CVD_ATH_ANY | 17386 | -0.00153 | -0.00293 | 0.012158 | 0.55012 |
| Nerve | CVD_ATH_ANY | 17386 | -0.00201 | -0.00405 | 0.009697 | 0.58204 |
| Breast | CVD_ATH_ANY | 17386 | -0.00418 | -0.00805 | 0.014666 | 0.61222 |
| Thyroid | CVD_ATH_ANY | 17386 | -0.00508 | -0.01008 | 0.0103 | 0.68924 |
| Heart | CVD_ATH_ANY | 17386 | -0.00549 | -0.00945 | 0.009214 | 0.7242 |
| Brain | CVD_ATH_ANY | 17386 | -0.00491 | -0.00853 | 0.006535 | 0.77364 |
| Colon | CVD_ATH_ANY | 17386 | -0.012 | -0.02248 | 0.014942 | 0.78901 |
| Salivary_Gland | CVD_ATH_ANY | 17386 | -0.00887 | -0.01652 | 0.010344 | 0.80435 |
| Stomach | CVD_ATH_ANY | 17386 | -0.01129 | -0.02053 | 0.012988 | 0.80758 |
| Pituitary | CVD_ATH_ANY | 17386 | -0.00726 | -0.01355 | 0.007966 | 0.81894 |
| Muscle | CVD_ATH_ANY | 17386 | -0.00609 | -0.01152 | 0.00649 | 0.82588 |
| Kidney | CVD_ATH_ANY | 17386 | -0.00854 | -0.01503 | 0.008786 | 0.83435 |
| Esophagus | CVD_ATH_ANY | 17386 | -0.01772 | -0.03342 | 0.016296 | 0.86159 |
| Skin | CVD_ATH_ANY | 17386 | -0.01144 | -0.02183 | 0.009679 | 0.88129 |
| Prostate | CVD_ATH_ANY | 17386 | -0.01643 | -0.03121 | 0.013209 | 0.89319 |
| Liver | CVD_ATH_ANY | 17386 | -0.01008 | -0.01816 | 0.006309 | 0.94493 |
| Adrenal_Gland | CVD_CAA | 17385 | 0.015884 | 0.03069 | 0.009789 | 0.052348 |
| Colon | CVD_CAA | 17385 | 0.019791 | 0.037084 | 0.014787 | 0.090395 |
| Thyroid | CVD_CAA | 17385 | 0.01181 | 0.023413 | 0.010271 | 0.12512 |
| Uterus | CVD_CAA | 17385 | 0.010012 | 0.020417 | 0.011542 | 0.19286 |
| Liver | CVD_CAA | 17385 | 0.004886 | 0.008802 | 0.006334 | 0.22024 |
| Pituitary | CVD_CAA | 17385 | 0.005175 | 0.009659 | 0.008089 | 0.26117 |
| Lung | CVD_CAA | 17385 | 0.005316 | 0.010289 | 0.010054 | 0.2985 |

| | | | | | | |
|---|---|---|---|---|---|---|
| Cervix_Uteri | CVD_CAA | 17385 | 0.007151 | 0.014062 | 0.013594 | 0.29944 |
| Prostate | CVD_CAA | 17385 | 0.006888 | 0.013084 | 0.013294 | 0.30219 |
| Skin | CVD_CAA | 17385 | 0.004715 | 0.009 | 0.00979 | 0.31505 |
| Small_Intestine | CVD_CAA | 17385 | 0.004706 | 0.008665 | 0.010148 | 0.32141 |
| Brain | CVD_CAA | 17385 | 0.002671 | 0.004644 | 0.006591 | 0.34267 |
| Kidney | CVD_CAA | 17385 | 0.002599 | 0.004577 | 0.008759 | 0.38335 |
| Blood_Vessel | CVD_CAA | 17385 | 0.002307 | 0.004685 | 0.011168 | 0.41818 |
| Bladder | CVD_CAA | 17385 | 0.002898 | 0.005655 | 0.015307 | 0.42492 |
| Pancreas | CVD_CAA | 17385 | 0.001465 | 0.002451 | 0.008456 | 0.43124 |
| Fallopian_Tube | CVD_CAA | 17385 | 0.002053 | 0.003988 | 0.013398 | 0.4391 |
| Ovary | CVD_CAA | 17385 | 0.0008 | 0.001623 | 0.009735 | 0.46725 |
| Stomach | CVD_CAA | 17385 | 0.000269 | 0.000489 | 0.012547 | 0.49145 |
| Muscle | CVD_CAA | 17385 | -0.00162 | -0.00306 | 0.006663 | 0.59587 |
| Nerve | CVD_CAA | 17385 | -0.00322 | -0.00649 | 0.009813 | 0.62868 |
| Heart | CVD_CAA | 17385 | -0.00408 | -0.00703 | 0.009135 | 0.67252 |
| Vagina | CVD_CAA | 17385 | -0.01072 | -0.02051 | 0.012308 | 0.80802 |
| Salivary_Gland | CVD_CAA | 17385 | -0.00896 | -0.01669 | 0.010252 | 0.80896 |
| Testis | CVD_CAA | 17385 | -0.00484 | -0.00837 | 0.005403 | 0.81482 |
| Spleen | CVD_CAA | 17385 | -0.00783 | -0.01553 | 0.007593 | 0.84868 |
| Adipose_Tissue | CVD_CAA | 17385 | -0.01352 | -0.02658 | 0.012659 | 0.85729 |
| Esophagus | CVD_CAA | 17385 | -0.0177 | -0.03338 | 0.016277 | 0.86161 |
| Blood | CVD_CAA | 17385 | -0.0076 | -0.01365 | 0.005994 | 0.89754 |
| Breast | CVD_CAA | 17385 | -0.02152 | -0.04143 | 0.01477 | 0.92743 |
| Ovary | CVD_CAA_ANY | 17386 | 0.017196 | 0.034889 | 0.009834 | 0.04019 |
| Kidney | CVD_CAA_ANY | 17386 | 0.012395 | 0.021831 | 0.009118 | 0.087011 |
| Prostate | CVD_CAA_ANY | 17386 | 0.012703 | 0.02413 | 0.013383 | 0.17126 |
| Uterus | CVD_CAA_ANY | 17386 | 0.011054 | 0.022543 | 0.011676 | 0.17191 |
| Muscle | CVD_CAA_ANY | 17386 | 0.004814 | 0.00911 | 0.006665 | 0.23506 |
| Liver | CVD_CAA_ANY | 17386 | 0.004439 | 0.007997 | 0.006344 | 0.24207 |
| Fallopian_Tube | CVD_CAA_ANY | 17386 | 0.00928 | 0.018024 | 0.013339 | 0.24331 |
| Small_Intestine | CVD_CAA_ANY | 17386 | 0.007028 | 0.012939 | 0.010353 | 0.24863 |
| Brain | CVD_CAA_ANY | 17386 | 0.004388 | 0.007629 | 0.006687 | 0.25585 |
| Cervix_Uteri | CVD_CAA_ANY | 17386 | 0.007846 | 0.015431 | 0.013727 | 0.2838 |
| Adrenal_Gland | CVD_CAA_ANY | 17386 | 0.005387 | 0.010408 | 0.009735 | 0.29002 |

| | | | | | | |
|---|---|---|---|---|---|---|
| Thyroid | CVD_CAA_ANY | 17386 | 0.004413 | 0.008748 | 0.010479 | 0.33686 |
| Skin | CVD_CAA_ANY | 17386 | 0.00367 | 0.007005 | 0.009743 | 0.35321 |
| Pancreas | CVD_CAA_ANY | 17386 | 0.001357 | 0.00227 | 0.008552 | 0.43698 |
| Stomach | CVD_CAA_ANY | 17386 | 0.000718 | 0.001305 | 0.013057 | 0.47808 |
| Blood_Vessel | CVD_CAA_ANY | 17386 | 0.000523 | 0.001063 | 0.011346 | 0.48161 |
| Colon | CVD_CAA_ANY | 17386 | -2.46E-05 | -4.60E-05 | 0.015133 | 0.50065 |
| Pituitary | CVD_CAA_ANY | 17386 | -0.0013 | -0.00243 | 0.008143 | 0.56348 |
| Testis | CVD_CAA_ANY | 17386 | -0.00091 | -0.00158 | 0.005416 | 0.5669 |
| Blood | CVD_CAA_ANY | 17386 | -0.00155 | -0.00278 | 0.006039 | 0.60124 |
| Salivary_Gland | CVD_CAA_ANY | 17386 | -0.00443 | -0.00825 | 0.010371 | 0.66533 |
| Breast | CVD_CAA_ANY | 17386 | -0.0102 | -0.01964 | 0.014778 | 0.755 |
| Nerve | CVD_CAA_ANY | 17386 | -0.00861 | -0.01734 | 0.010063 | 0.8038 |
| Bladder | CVD_CAA_ANY | 17386 | -0.01601 | -0.03123 | 0.015217 | 0.85354 |
| Heart | CVD_CAA_ANY | 17386 | -0.00967 | -0.01665 | 0.009181 | 0.85383 |
| Vagina | CVD_CAA_ANY | 17386 | -0.01607 | -0.03075 | 0.012386 | 0.90267 |
| Spleen | CVD_CAA_ANY | 17386 | -0.01042 | -0.02068 | 0.007604 | 0.91479 |
| Lung | CVD_CAA_ANY | 17386 | -0.01458 | -0.02822 | 0.010078 | 0.92599 |
| Adipose_Tissue | CVD_CAA_ANY | 17386 | -0.02107 | -0.04141 | 0.012661 | 0.95197 |
| Esophagus | CVD_CAA_ANY | 17386 | -0.02928 | -0.05522 | 0.016497 | 0.96205 |
| Testis | CVD_S | 17387 | 0.008748 | 0.015121 | 0.00538 | 0.051999 |
| Fallopian_Tube | CVD_S | 17387 | 0.018753 | 0.036422 | 0.013419 | 0.08115 |
| Adrenal_Gland | CVD_S | 17387 | 0.010664 | 0.020604 | 0.009686 | 0.13545 |
| Ovary | CVD_S | 17387 | 0.01016 | 0.020612 | 0.009819 | 0.1504 |
| Small_Intestine | CVD_S | 17387 | 0.009481 | 0.017455 | 0.010303 | 0.17872 |
| Skin | CVD_S | 17387 | 0.008471 | 0.016168 | 0.009617 | 0.18922 |
| Esophagus | CVD_S | 17387 | 0.011999 | 0.022626 | 0.016428 | 0.23259 |
| Lung | CVD_S | 17387 | 0.005648 | 0.010931 | 0.010005 | 0.28621 |
| Pituitary | CVD_S | 17387 | 0.003173 | 0.005921 | 0.008007 | 0.34598 |
| Colon | CVD_S | 17387 | 0.004702 | 0.008811 | 0.014856 | 0.37581 |
| Cervix_Uteri | CVD_S | 17387 | 0.003093 | 0.006083 | 0.013599 | 0.41003 |
| Blood | CVD_S | 17387 | 0.000766 | 0.001376 | 0.006018 | 0.44937 |
| Uterus | CVD_S | 17387 | 0.001364 | 0.002781 | 0.011458 | 0.45264 |
| Vagina | CVD_S | 17387 | -0.00052 | -0.001 | 0.012271 | 0.51692 |
| Kidney | CVD_S | 17387 | -0.00111 | -0.00196 | 0.008926 | 0.5495 |

| | | | | | | |
|---|---|---|---|---|---|---|
| Blood_Vessel | CVD_S | 17387 | -0.00219 | -0.00445 | 0.011205 | 0.57749 |
| Adipose_Tissue | CVD_S | 17387 | -0.00302 | -0.00594 | 0.012528 | 0.59526 |
| Stomach | CVD_S | 17387 | -0.00321 | -0.00584 | 0.012862 | 0.59864 |
| Thyroid | CVD_S | 17387 | -0.00416 | -0.00825 | 0.010375 | 0.65587 |
| Brain | CVD_S | 17387 | -0.00291 | -0.00507 | 0.006577 | 0.67115 |
| Spleen | CVD_S | 17387 | -0.00336 | -0.00667 | 0.007547 | 0.6721 |
| Liver | CVD_S | 17387 | -0.00295 | -0.00531 | 0.006288 | 0.68037 |
| Muscle | CVD_S | 17387 | -0.00357 | -0.00675 | 0.006653 | 0.70414 |
| Heart | CVD_S | 17387 | -0.00564 | -0.00971 | 0.009334 | 0.72716 |
| Nerve | CVD_S | 17387 | -0.00718 | -0.01447 | 0.009747 | 0.76941 |
| Pancreas | CVD_S | 17387 | -0.00691 | -0.01156 | 0.008502 | 0.79182 |
| Bladder | CVD_S | 17387 | -0.01259 | -0.02456 | 0.015206 | 0.7961 |
| Breast | CVD_S | 17387 | -0.01632 | -0.03141 | 0.014767 | 0.86538 |
| Prostate | CVD_S | 17387 | -0.01474 | -0.028 | 0.013162 | 0.86863 |
| Salivary_Gland | CVD_S | 17387 | -0.0188 | -0.03502 | 0.010159 | 0.96785 |
| Cervix_Uteri | HEMOR | 17378 | 0.025163 | 0.049486 | 0.013477 | 0.03095 |
| Uterus | HEMOR | 17378 | 0.020452 | 0.04171 | 0.011372 | 0.03606 |
| Fallopian_Tube | HEMOR | 17378 | 0.014986 | 0.029108 | 0.013308 | 0.13008 |
| Vagina | HEMOR | 17378 | 0.010984 | 0.021027 | 0.012189 | 0.18376 |
| Pituitary | HEMOR | 17378 | 0.007171 | 0.013386 | 0.007987 | 0.18462 |
| Liver | HEMOR | 17378 | 0.004157 | 0.00749 | 0.006295 | 0.25451 |
| Nerve | HEMOR | 17378 | 0.00521 | 0.010499 | 0.009855 | 0.29851 |
| Muscle | HEMOR | 17378 | 0.00299 | 0.005656 | 0.006616 | 0.32569 |
| Small_Intestine | HEMOR | 17378 | 0.004252 | 0.007829 | 0.010185 | 0.33815 |
| Esophagus | HEMOR | 17378 | 0.00637 | 0.012012 | 0.016303 | 0.34801 |
| Skin | HEMOR | 17378 | 0.003441 | 0.006568 | 0.009618 | 0.36027 |
| Stomach | HEMOR | 17378 | 0.003599 | 0.006544 | 0.012601 | 0.38761 |
| Colon | HEMOR | 17378 | 0.003938 | 0.007379 | 0.014864 | 0.39553 |
| Ovary | HEMOR | 17378 | 0.002313 | 0.004693 | 0.009669 | 0.40547 |
| Salivary_Gland | HEMOR | 17378 | 0.002045 | 0.003811 | 0.010218 | 0.42067 |
| Pancreas | HEMOR | 17378 | 0.000525 | 0.000879 | 0.008393 | 0.47505 |
| Blood_Vessel | HEMOR | 17378 | -0.00148 | -0.00301 | 0.011121 | 0.55307 |
| Kidney | HEMOR | 17378 | -0.00121 | -0.00213 | 0.008843 | 0.55434 |
| Prostate | HEMOR | 17378 | -0.00224 | -0.00425 | 0.013015 | 0.56825 |

| | | | | | | |
|---|---|---|---|---|---|---|
| Thyroid | HEMOR | 17378 | -0.00189 | -0.00375 | 0.010276 | 0.57293 |
| Testis | HEMOR | 17378 | -0.00114 | -0.00197 | 0.005336 | 0.5846 |
| Adipose_Tissue | HEMOR | 17378 | -0.00576 | -0.01133 | 0.012692 | 0.67515 |
| Brain | HEMOR | 17378 | -0.00333 | -0.00579 | 0.006576 | 0.69358 |
| Bladder | HEMOR | 17378 | -0.00956 | -0.01865 | 0.015178 | 0.73553 |
| Adrenal_Gland | HEMOR | 17378 | -0.00655 | -0.01266 | 0.009678 | 0.75085 |
| Breast | HEMOR | 17378 | -0.01383 | -0.02662 | 0.014741 | 0.82585 |
| Lung | HEMOR | 17378 | -0.00997 | -0.0193 | 0.009907 | 0.84291 |
| Blood | HEMOR | 17378 | -0.00604 | -0.01085 | 0.00598 | 0.84372 |
| Spleen | HEMOR | 17378 | -0.01037 | -0.02057 | 0.007589 | 0.91407 |
| Heart | HEMOR | 17378 | -0.01295 | -0.02231 | 0.009231 | 0.91972 |
| Bladder | HS_S | 17386 | 0.025416 | 0.049593 | 0.015209 | 0.047358 |
| Skin | HS_S | 17386 | 0.016044 | 0.030625 | 0.009627 | 0.04781 |
| Vagina | HS_S | 17386 | 0.019663 | 0.037643 | 0.012207 | 0.053617 |
| Prostate | HS_S | 17386 | 0.015777 | 0.029969 | 0.013218 | 0.11634 |
| Pituitary | HS_S | 17386 | 0.007135 | 0.013318 | 0.008044 | 0.18754 |
| Colon | HS_S | 17386 | 0.012392 | 0.02322 | 0.015026 | 0.20478 |
| Testis | HS_S | 17386 | 0.00414 | 0.007157 | 0.005332 | 0.21875 |
| Esophagus | HS_S | 17386 | 0.011283 | 0.021277 | 0.016388 | 0.24558 |
| Brain | HS_S | 17386 | 0.002192 | 0.003811 | 0.006588 | 0.3697 |
| Small_Intestine | HS_S | 17386 | 0.002951 | 0.005433 | 0.010214 | 0.38632 |
| Liver | HS_S | 17386 | 0.001396 | 0.002516 | 0.006309 | 0.41243 |
| Adrenal_Gland | HS_S | 17386 | 0.001601 | 0.003093 | 0.009725 | 0.43463 |
| Spleen | HS_S | 17386 | 0.000349 | 0.000693 | 0.007625 | 0.48174 |
| Lung | HS_S | 17386 | 0.000137 | 0.000266 | 0.010014 | 0.49453 |
| Cervix_Uteri | HS_S | 17386 | -0.00071 | -0.00139 | 0.013542 | 0.52086 |
| Kidney | HS_S | 17386 | -0.00111 | -0.00196 | 0.008997 | 0.54918 |
| Adipose_Tissue | HS_S | 17386 | -0.00278 | -0.00547 | 0.012697 | 0.58671 |
| Pancreas | HS_S | 17386 | -0.00256 | -0.00428 | 0.008481 | 0.61854 |
| Fallopian_Tube | HS_S | 17386 | -0.00404 | -0.00785 | 0.013255 | 0.61976 |
| Blood | HS_S | 17386 | -0.00202 | -0.00362 | 0.005994 | 0.63168 |
| Heart | HS_S | 17386 | -0.00458 | -0.00788 | 0.009177 | 0.69097 |
| Ovary | HS_S | 17386 | -0.00607 | -0.01232 | 0.009921 | 0.72969 |
| Salivary_Gland | HS_S | 17386 | -0.00643 | -0.01198 | 0.010338 | 0.7331 |

| | | | | | | |
|---|---|---|---|---|---|---|
| Thyroid | HS_S | 17386 | -0.00652 | -0.01293 | 0.010383 | 0.7351 |
| Stomach | HS_S | 17386 | -0.00964 | -0.01754 | 0.012972 | 0.77136 |
| Muscle | HS_S | 17386 | -0.00574 | -0.01086 | 0.006647 | 0.806 |
| Breast | HS_S | 17386 | -0.0128 | -0.02463 | 0.01472 | 0.80763 |
| Blood_Vessel | HS_S | 17386 | -0.01442 | -0.02929 | 0.011173 | 0.90164 |
| Uterus | HS_S | 17386 | -0.01474 | -0.03005 | 0.011402 | 0.90187 |
| Nerve | HS_S | 17386 | -0.01326 | -0.02671 | 0.009917 | 0.90932 |
| Adipose_Tissue | INFA | 17386 | 0.030768 | 0.060464 | 0.012367 | 0.00643 |
| Breast | INFA | 17386 | 0.034589 | 0.066591 | 0.0143 | 0.007794 |
| Thyroid | INFA | 17386 | 0.014855 | 0.02945 | 0.010137 | 0.071417 |
| Adrenal_Gland | INFA | 17386 | 0.013307 | 0.025711 | 0.009412 | 0.078721 |
| Salivary_Gland | INFA | 17386 | 0.013187 | 0.024567 | 0.009992 | 0.093474 |
| Blood_Vessel | INFA | 17386 | 0.01325 | 0.026906 | 0.010865 | 0.11133 |
| Lung | INFA | 17386 | 0.011812 | 0.022864 | 0.009768 | 0.11329 |
| Pancreas | INFA | 17386 | 0.00711 | 0.011898 | 0.008203 | 0.19304 |
| Skin | INFA | 17386 | 0.007294 | 0.013924 | 0.009448 | 0.22005 |
| Nerve | INFA | 17386 | 0.005714 | 0.011514 | 0.009568 | 0.27519 |
| Uterus | INFA | 17386 | 0.00599 | 0.012215 | 0.011133 | 0.29529 |
| Testis | INFA | 17386 | 0.002678 | 0.004629 | 0.005234 | 0.30446 |
| Stomach | INFA | 17386 | 0.005153 | 0.009371 | 0.012736 | 0.34289 |
| Fallopian_Tube | INFA | 17386 | 0.004199 | 0.008156 | 0.012916 | 0.37255 |
| Cervix_Uteri | INFA | 17386 | 0.00105 | 0.002065 | 0.013174 | 0.46824 |
| Blood | INFA | 17386 | 0.000289 | 0.00052 | 0.005799 | 0.4801 |
| Vagina | INFA | 17386 | -0.00324 | -0.0062 | 0.011751 | 0.60851 |
| Spleen | INFA | 17386 | -0.00271 | -0.00537 | 0.007301 | 0.6445 |
| Bladder | INFA | 17386 | -0.0055 | -0.01073 | 0.014787 | 0.64503 |
| Kidney | INFA | 17386 | -0.00412 | -0.00726 | 0.008609 | 0.68387 |
| Small_Intestine | INFA | 17386 | -0.00505 | -0.0093 | 0.00992 | 0.69469 |
| Esophagus | INFA | 17386 | -0.01205 | -0.02271 | 0.015901 | 0.77561 |
| Heart | INFA | 17386 | -0.00764 | -0.01316 | 0.009066 | 0.80038 |
| Prostate | INFA | 17386 | -0.0113 | -0.02146 | 0.012594 | 0.81512 |
| Pituitary | INFA | 17386 | -0.00799 | -0.01491 | 0.007897 | 0.84405 |
| Muscle | INFA | 17386 | -0.00674 | -0.01275 | 0.006502 | 0.84991 |
| Colon | INFA | 17386 | -0.01626 | -0.03047 | 0.014604 | 0.86718 |

| | | | | | | |
|---|---|---|---|---|---|---|
| Ovary | INFA | 17386 | -0.01128 | -0.02288 | 0.009549 | 0.8812 |
| Brain | INFA | 17386 | -0.00957 | -0.01663 | 0.00643 | 0.93157 |
| Liver | INFA | 17386 | -0.01368 | -0.02465 | 0.006132 | 0.98715 |
| Pituitary | LEWY_ANY | 17385 | 0.011941 | 0.022287 | 0.008101 | 0.07026 |
| Ovary | LEWY_ANY | 17385 | 0.01435 | 0.029114 | 0.009829 | 0.072167 |
| Heart | LEWY_ANY | 17385 | 0.012202 | 0.021012 | 0.009452 | 0.09836 |
| Uterus | LEWY_ANY | 17385 | 0.01454 | 0.029653 | 0.011643 | 0.10588 |
| Brain | LEWY_ANY | 17385 | 0.006248 | 0.010863 | 0.006657 | 0.17398 |
| Adrenal_Gland | LEWY_ANY | 17385 | 0.008912 | 0.01722 | 0.009766 | 0.18073 |
| Cervix_Uteri | LEWY_ANY | 17385 | 0.009895 | 0.019459 | 0.01376 | 0.23604 |
| Skin | LEWY_ANY | 17385 | 0.00454 | 0.008666 | 0.009976 | 0.32454 |
| Breast | LEWY_ANY | 17385 | 0.005272 | 0.01015 | 0.0151 | 0.36349 |
| Liver | LEWY_ANY | 17385 | 0.001587 | 0.00286 | 0.006472 | 0.40313 |
| Vagina | LEWY_ANY | 17385 | 0.001765 | 0.003379 | 0.012551 | 0.44409 |
| Blood | LEWY_ANY | 17385 | 0.000751 | 0.001349 | 0.00608 | 0.45086 |
| Spleen | LEWY_ANY | 17385 | 0.000872 | 0.001729 | 0.007572 | 0.45417 |
| Testis | LEWY_ANY | 17385 | 2.94E-05 | 5.08E-05 | 0.005448 | 0.49785 |
| Fallopian_Tube | LEWY_ANY | 17385 | -7.18E-05 | -0.00014 | 0.01348 | 0.50212 |
| Stomach | LEWY_ANY | 17385 | -0.00147 | -0.00267 | 0.01298 | 0.54504 |
| Adipose_Tissue | LEWY_ANY | 17385 | -0.00186 | -0.00366 | 0.012801 | 0.55787 |
| Blood_Vessel | LEWY_ANY | 17385 | -0.00233 | -0.00474 | 0.01135 | 0.58139 |
| Thyroid | LEWY_ANY | 17385 | -0.00257 | -0.0051 | 0.010701 | 0.59499 |
| Nerve | LEWY_ANY | 17385 | -0.00377 | -0.0076 | 0.010139 | 0.64507 |
| Muscle | LEWY_ANY | 17385 | -0.00409 | -0.00774 | 0.006777 | 0.72687 |
| Small_Intestine | LEWY_ANY | 17385 | -0.00758 | -0.01397 | 0.010428 | 0.76648 |
| Colon | LEWY_ANY | 17385 | -0.0114 | -0.02136 | 0.0153 | 0.77184 |
| Pancreas | LEWY_ANY | 17385 | -0.00648 | -0.01084 | 0.008625 | 0.77372 |
| Bladder | LEWY_ANY | 17385 | -0.01448 | -0.02825 | 0.015695 | 0.82184 |
| Prostate | LEWY_ANY | 17385 | -0.01263 | -0.02399 | 0.013526 | 0.82477 |
| Lung | LEWY_ANY | 17385 | -0.00978 | -0.01892 | 0.010017 | 0.83543 |
| Esophagus | LEWY_ANY | 17385 | -0.02048 | -0.03863 | 0.01654 | 0.89221 |
| Kidney | LEWY_ANY | 17385 | -0.01269 | -0.02234 | 0.009106 | 0.91819 |
| Salivary_Gland | LEWY_ANY | 17385 | -0.01857 | -0.03459 | 0.010452 | 0.96215 |
| Pituitary | LEWY_full | 17385 | 0.020117 | 0.037549 | 0.008054 | 0.006255 |

| | | | | | | |
|---|---|---|---|---|---|---|
| Uterus | LEWY_full | 17385 | 0.021898 | 0.044658 | 0.011567 | 0.029184 |
| Cervix_Uteri | LEWY_full | 17385 | 0.022246 | 0.043748 | 0.013646 | 0.051537 |
| Ovary | LEWY_full | 17385 | 0.014044 | 0.028492 | 0.00988 | 0.077601 |
| Brain | LEWY_full | 17385 | 0.008689 | 0.015107 | 0.006681 | 0.096722 |
| Breast | LEWY_full | 17385 | 0.015031 | 0.028938 | 0.01491 | 0.15671 |
| Skin | LEWY_full | 17385 | 0.004631 | 0.008839 | 0.009863 | 0.31936 |
| Blood_Vessel | LEWY_full | 17385 | 0.004643 | 0.009428 | 0.011334 | 0.34104 |
| Adrenal_Gland | LEWY_full | 17385 | 0.003863 | 0.007463 | 0.009864 | 0.34767 |
| Vagina | LEWY_full | 17385 | 0.003739 | 0.007157 | 0.012378 | 0.38131 |
| Fallopian_Tube | LEWY_full | 17385 | 0.003041 | 0.005907 | 0.013351 | 0.40991 |
| Stomach | LEWY_full | 17385 | 0.001714 | 0.003116 | 0.012957 | 0.4474 |
| Adipose_Tissue | LEWY_full | 17385 | 0.001139 | 0.002239 | 0.012828 | 0.46462 |
| Thyroid | LEWY_full | 17385 | 0.000248 | 0.000492 | 0.010506 | 0.49058 |
| Spleen | LEWY_full | 17385 | -0.00028 | -0.00056 | 0.007682 | 0.51467 |
| Nerve | LEWY_full | 17385 | -0.00046 | -0.00093 | 0.010005 | 0.51844 |
| Testis | LEWY_full | 17385 | -0.00026 | -0.00045 | 0.005379 | 0.51945 |
| Heart | LEWY_full | 17385 | -0.00144 | -0.00248 | 0.009405 | 0.56095 |
| Small_Intestine | LEWY_full | 17385 | -0.00364 | -0.00671 | 0.010357 | 0.63752 |
| Lung | LEWY_full | 17385 | -0.00482 | -0.00932 | 0.010017 | 0.68471 |
| Prostate | LEWY_full | 17385 | -0.0073 | -0.01386 | 0.013312 | 0.70816 |
| Bladder | LEWY_full | 17385 | -0.00866 | -0.0169 | 0.01548 | 0.71209 |
| Colon | LEWY_full | 17385 | -0.00953 | -0.01785 | 0.014963 | 0.73785 |
| Pancreas | LEWY_full | 17385 | -0.00595 | -0.00996 | 0.008659 | 0.75405 |
| Liver | LEWY_full | 17385 | -0.00461 | -0.0083 | 0.006272 | 0.76864 |
| Blood | LEWY_full | 17385 | -0.00491 | -0.00883 | 0.006016 | 0.79301 |
| Esophagus | LEWY_full | 17385 | -0.01626 | -0.03067 | 0.016439 | 0.83874 |
| Salivary_Gland | LEWY_full | 17385 | -0.01362 | -0.02537 | 0.010458 | 0.90356 |
| Muscle | LEWY_full | 17385 | -0.01047 | -0.01982 | 0.006737 | 0.93995 |
| Kidney | LEWY_full | 17385 | -0.01615 | -0.02844 | 0.009002 | 0.96354 |
| Pituitary | LEWY_grp | 17385 | 0.023208 | 0.043318 | 0.008108 | 0.002105 |
| Brain | LEWY_grp | 17385 | 0.00924 | 0.016065 | 0.006812 | 0.087489 |
| Uterus | LEWY_grp | 17385 | 0.01529 | 0.031182 | 0.01175 | 0.096589 |
| Ovary | LEWY_grp | 17385 | 0.011517 | 0.023365 | 0.010017 | 0.12515 |
| Cervix_Uteri | LEWY_grp | 17385 | 0.011378 | 0.022376 | 0.013954 | 0.20742 |

| | | | | | | |
|---|---|---|---|---|---|---|
| Adrenal_Gland | LEWY_grp | 17385 | 0.007825 | 0.015118 | 0.009896 | 0.21457 |
| Heart | LEWY_grp | 17385 | 0.003242 | 0.005582 | 0.009578 | 0.36752 |
| Breast | LEWY_grp | 17385 | 0.004498 | 0.00866 | 0.015197 | 0.38362 |
| Skin | LEWY_grp | 17385 | 0.002246 | 0.004288 | 0.009965 | 0.41083 |
| Fallopian_Tube | LEWY_grp | 17385 | 0.001981 | 0.003847 | 0.013628 | 0.44222 |
| Spleen | LEWY_grp | 17385 | 0.000679 | 0.001347 | 0.00776 | 0.46513 |
| Blood_Vessel | LEWY_grp | 17385 | -0.00056 | -0.00113 | 0.011446 | 0.51941 |
| Stomach | LEWY_grp | 17385 | -0.00081 | -0.00147 | 0.01307 | 0.5247 |
| Small_Intestine | LEWY_grp | 17385 | -0.00085 | -0.00156 | 0.01047 | 0.53229 |
| Nerve | LEWY_grp | 17385 | -0.00111 | -0.00223 | 0.010066 | 0.54386 |
| Vagina | LEWY_grp | 17385 | -0.00145 | -0.00278 | 0.012463 | 0.54634 |
| Blood | LEWY_grp | 17385 | -0.0009 | -0.00162 | 0.006118 | 0.55868 |
| Colon | LEWY_grp | 17385 | -0.00357 | -0.00669 | 0.015124 | 0.59328 |
| Testis | LEWY_grp | 17385 | -0.00169 | -0.00293 | 0.0054 | 0.62309 |
| Thyroid | LEWY_grp | 17385 | -0.00345 | -0.00683 | 0.010565 | 0.62781 |
| Liver | LEWY_grp | 17385 | -0.00213 | -0.00384 | 0.006384 | 0.63079 |
| Adipose_Tissue | LEWY_grp | 17385 | -0.0048 | -0.00942 | 0.012985 | 0.64406 |
| Lung | LEWY_grp | 17385 | -0.00508 | -0.00982 | 0.010138 | 0.69168 |
| Pancreas | LEWY_grp | 17385 | -0.00585 | -0.00979 | 0.008694 | 0.74942 |
| Bladder | LEWY_grp | 17385 | -0.0154 | -0.03006 | 0.015634 | 0.83774 |
| Muscle | LEWY_grp | 17385 | -0.00699 | -0.01323 | 0.00678 | 0.84885 |
| Prostate | LEWY_grp | 17385 | -0.01434 | -0.02724 | 0.013451 | 0.85677 |
| Esophagus | LEWY_grp | 17385 | -0.01772 | -0.03342 | 0.016589 | 0.8573 |
| Salivary_Gland | LEWY_grp | 17385 | -0.01192 | -0.02221 | 0.010601 | 0.86964 |
| Kidney | LEWY_grp | 17385 | -0.01455 | -0.02562 | 0.009064 | 0.94576 |
| Nerve | MICR | 17385 | 0.019029 | 0.038344 | 0.009721 | 0.02515 |
| Uterus | MICR | 17385 | 0.018017 | 0.036743 | 0.011475 | 0.058212 |
| Cervix_Uteri | MICR | 17385 | 0.01901 | 0.037385 | 0.013495 | 0.079469 |
| Breast | MICR | 17385 | 0.018559 | 0.035729 | 0.014695 | 0.10331 |
| Adipose_Tissue | MICR | 17385 | 0.015211 | 0.029892 | 0.012541 | 0.11259 |
| Prostate | MICR | 17385 | 0.012408 | 0.023568 | 0.013008 | 0.1701 |
| Blood_Vessel | MICR | 17385 | 0.009887 | 0.020077 | 0.011099 | 0.18654 |
| Fallopian_Tube | MICR | 17385 | 0.009241 | 0.017948 | 0.01324 | 0.24261 |
| Skin | MICR | 17385 | 0.004826 | 0.009213 | 0.009666 | 0.30878 |

| | | | | | | |
|---|---|---|---|---|---|---|
| Ovary | MICR | 17385 | 0.004388 | 0.008903 | 0.009734 | 0.32606 |
| Blood | MICR | 17385 | 0.002635 | 0.004735 | 0.005893 | 0.32736 |
| Brain | MICR | 17385 | 0.002805 | 0.004876 | 0.006524 | 0.33365 |
| Spleen | MICR | 17385 | 0.001487 | 0.00295 | 0.007392 | 0.42027 |
| Stomach | MICR | 17385 | 0.002207 | 0.004014 | 0.012781 | 0.43145 |
| Thyroid | MICR | 17385 | 0.001469 | 0.002912 | 0.01032 | 0.4434 |
| Small_Intestine | MICR | 17385 | 0.00064 | 0.001179 | 0.01002 | 0.47453 |
| Esophagus | MICR | 17385 | -0.00049 | -0.00093 | 0.016327 | 0.51203 |
| Lung | MICR | 17385 | -0.00168 | -0.00325 | 0.009877 | 0.56748 |
| Vagina | MICR | 17385 | -0.00355 | -0.0068 | 0.012123 | 0.6153 |
| Colon | MICR | 17385 | -0.0046 | -0.00861 | 0.014836 | 0.62168 |
| Liver | MICR | 17385 | -0.00206 | -0.00371 | 0.00628 | 0.62865 |
| Salivary_Gland | MICR | 17385 | -0.00379 | -0.00707 | 0.010297 | 0.64373 |
| Heart | MICR | 17385 | -0.00659 | -0.01136 | 0.009146 | 0.76454 |
| Adrenal_Gland | MICR | 17385 | -0.00697 | -0.01346 | 0.009591 | 0.76619 |
| Bladder | MICR | 17385 | -0.01304 | -0.02545 | 0.015244 | 0.80387 |
| Pancreas | MICR | 17385 | -0.00738 | -0.01235 | 0.008395 | 0.81034 |
| Pituitary | MICR | 17385 | -0.00724 | -0.01351 | 0.007993 | 0.81737 |
| Testis | MICR | 17385 | -0.0052 | -0.00898 | 0.005355 | 0.83407 |
| Kidney | MICR | 17385 | -0.01115 | -0.01964 | 0.008794 | 0.89763 |
| Muscle | MICR | 17385 | -0.00951 | -0.01799 | 0.006487 | 0.92855 |
| Thyroid | TDP_3 | 17388 | 0.023633 | 0.04685 | 0.01053 | 0.012417 |
| Lung | TDP_3 | 17388 | 0.021589 | 0.041788 | 0.010105 | 0.016334 |
| Nerve | TDP_3 | 17388 | 0.018064 | 0.036399 | 0.010022 | 0.035756 |
| Blood_Vessel | TDP_3 | 17388 | 0.012745 | 0.02588 | 0.01134 | 0.13054 |
| Adipose_Tissue | TDP_3 | 17388 | 0.011992 | 0.023566 | 0.012838 | 0.17514 |
| Adrenal_Gland | TDP_3 | 17388 | 0.008906 | 0.017207 | 0.009883 | 0.18377 |
| Colon | TDP_3 | 17388 | 0.013149 | 0.024639 | 0.014903 | 0.18882 |
| Fallopian_Tube | TDP_3 | 17388 | 0.010711 | 0.020804 | 0.013494 | 0.21368 |
| Stomach | TDP_3 | 17388 | 0.008252 | 0.015006 | 0.012937 | 0.26181 |
| Skin | TDP_3 | 17388 | 0.004307 | 0.008222 | 0.009837 | 0.33075 |
| Bladder | TDP_3 | 17388 | 0.003626 | 0.007075 | 0.015256 | 0.40606 |
| Uterus | TDP_3 | 17388 | 0.000381 | 0.000776 | 0.011533 | 0.48684 |
| Spleen | TDP_3 | 17388 | -9.69E-06 | -1.92E-05 | 0.007627 | 0.50051 |

| | | | | | | |
|---|---|---|---|---|---|---|
| Small_Intestine | TDP_3 | 17388 | -0.00014 | -0.00026 | 0.010445 | 0.50541 |
| Pancreas | TDP_3 | 17388 | -0.00014 | -0.00024 | 0.008552 | 0.50672 |
| Breast | TDP_3 | 17388 | -0.00119 | -0.00229 | 0.014856 | 0.53189 |
| Pituitary | TDP_3 | 17388 | -0.00137 | -0.00255 | 0.008221 | 0.56594 |
| Cervix_Uteri | TDP_3 | 17388 | -0.00295 | -0.0058 | 0.013602 | 0.58586 |
| Esophagus | TDP_3 | 17388 | -0.00379 | -0.00715 | 0.016477 | 0.59097 |
| Vagina | TDP_3 | 17388 | -0.00384 | -0.00735 | 0.012196 | 0.62351 |
| Prostate | TDP_3 | 17388 | -0.00441 | -0.00837 | 0.013282 | 0.62993 |
| Ovary | TDP_3 | 17388 | -0.00432 | -0.00877 | 0.009968 | 0.6677 |
| Kidney | TDP_3 | 17388 | -0.0049 | -0.00864 | 0.009198 | 0.70307 |
| Heart | TDP_3 | 17388 | -0.00585 | -0.01008 | 0.009257 | 0.73637 |
| Muscle | TDP_3 | 17388 | -0.0049 | -0.00927 | 0.006684 | 0.76822 |
| Blood | TDP_3 | 17388 | -0.0046 | -0.00826 | 0.005989 | 0.77863 |
| Liver | TDP_3 | 17388 | -0.005 | -0.00902 | 0.006366 | 0.78409 |
| Brain | TDP_3 | 17388 | -0.00692 | -0.01203 | 0.006701 | 0.849 |
| Testis | TDP_3 | 17388 | -0.00573 | -0.0099 | 0.005472 | 0.85227 |
| Salivary_Gland | TDP_3 | 17388 | -0.01176 | -0.02191 | 0.010273 | 0.87387 |
| Skin | TDP43 | 17386 | 0.022785 | 0.043492 | 0.009965 | 0.01112 |
| Nerve | TDP43 | 17386 | 0.020251 | 0.040806 | 0.010335 | 0.025042 |
| Blood_Vessel | TDP43 | 17386 | 0.021568 | 0.043796 | 0.011551 | 0.030946 |
| Thyroid | TDP43 | 17386 | 0.018813 | 0.037296 | 0.01061 | 0.038121 |
| Cervix_Uteri | TDP43 | 17386 | 0.023643 | 0.046497 | 0.0138 | 0.043345 |
| Adrenal_Gland | TDP43 | 17386 | 0.010632 | 0.020543 | 0.010052 | 0.14511 |
| Fallopian_Tube | TDP43 | 17386 | 0.010527 | 0.020447 | 0.013524 | 0.21818 |
| Vagina | TDP43 | 17386 | 0.00898 | 0.017192 | 0.012561 | 0.23734 |
| Colon | TDP43 | 17386 | 0.010191 | 0.019096 | 0.015167 | 0.25083 |
| Adipose_Tissue | TDP43 | 17386 | 0.008324 | 0.016358 | 0.012994 | 0.2609 |
| Lung | TDP43 | 17386 | 0.006166 | 0.011935 | 0.010242 | 0.27358 |
| Spleen | TDP43 | 17386 | 0.003543 | 0.007028 | 0.007688 | 0.32245 |
| Stomach | TDP43 | 17386 | 0.001281 | 0.00233 | 0.013437 | 0.46203 |
| Breast | TDP43 | 17386 | 0.001354 | 0.002606 | 0.015118 | 0.46433 |
| Uterus | TDP43 | 17386 | 0.000135 | 0.000276 | 0.011766 | 0.49541 |
| Esophagus | TDP43 | 17386 | 0.000165 | 0.000312 | 0.016676 | 0.49604 |
| Ovary | TDP43 | 17386 | -0.00019 | -0.00038 | 0.010097 | 0.50749 |

| | | | | | | |
|---|---|---|---|---|---|---|
| Salivary_Gland | TDP43 | 17386 | -0.00086 | -0.0016 | 0.010715 | 0.53186 |
| Bladder | TDP43 | 17386 | -0.00349 | -0.0068 | 0.01558 | 0.58853 |
| Muscle | TDP43 | 17386 | -0.00324 | -0.00614 | 0.006851 | 0.68205 |
| Liver | TDP43 | 17386 | -0.00308 | -0.00556 | 0.006473 | 0.68316 |
| Prostate | TDP43 | 17386 | -0.00769 | -0.0146 | 0.01358 | 0.71435 |
| Pituitary | TDP43 | 17386 | -0.00615 | -0.01148 | 0.008163 | 0.77434 |
| Heart | TDP43 | 17386 | -0.00773 | -0.01331 | 0.009512 | 0.79168 |
| Small_Intestine | TDP43 | 17386 | -0.00881 | -0.01623 | 0.010515 | 0.79902 |
| Testis | TDP43 | 17386 | -0.00549 | -0.00948 | 0.005481 | 0.84158 |
| Pancreas | TDP43 | 17386 | -0.01024 | -0.01713 | 0.008687 | 0.88068 |
| Brain | TDP43 | 17386 | -0.00824 | -0.01433 | 0.006777 | 0.88803 |
| Kidney | TDP43 | 17386 | -0.0116 | -0.02043 | 0.009115 | 0.89844 |
| Blood | TDP43 | 17386 | -0.00805 | -0.01447 | 0.006113 | 0.90614 |
| Spleen | WBVD | 17388 | 0.012755 | 0.0253 | 0.007538 | 0.045321 |
| Heart | WBVD | 17388 | 0.013 | 0.022386 | 0.009185 | 0.078509 |
| Blood | WBVD | 17388 | 0.007125 | 0.012802 | 0.00591 | 0.114 |
| Skin | WBVD | 17388 | 0.010529 | 0.020097 | 0.009665 | 0.138 |
| Breast | WBVD | 17388 | 0.015583 | 0.03 | 0.01473 | 0.14506 |
| Brain | WBVD | 17388 | 0.005684 | 0.009883 | 0.006542 | 0.19247 |
| Adipose_Tissue | WBVD | 17388 | 0.01096 | 0.021537 | 0.012619 | 0.19257 |
| Lung | WBVD | 17388 | 0.007993 | 0.01547 | 0.009964 | 0.21124 |
| Salivary_Gland | WBVD | 17388 | 0.006334 | 0.011799 | 0.010232 | 0.26797 |
| Muscle | WBVD | 17388 | 0.00308 | 0.005827 | 0.006636 | 0.3213 |
| Blood_Vessel | WBVD | 17388 | 0.005082 | 0.01032 | 0.011146 | 0.32421 |
| Adrenal_Gland | WBVD | 17388 | 0.004144 | 0.008006 | 0.009613 | 0.33323 |
| Nerve | WBVD | 17388 | 0.001433 | 0.002888 | 0.009933 | 0.44263 |
| Pituitary | WBVD | 17388 | 0.00107 | 0.001997 | 0.008 | 0.4468 |
| Thyroid | WBVD | 17388 | -3.07E-05 | -6.08E-05 | 0.010325 | 0.50118 |
| Pancreas | WBVD | 17388 | -0.00226 | -0.00378 | 0.008508 | 0.60464 |
| Stomach | WBVD | 17388 | -0.00346 | -0.0063 | 0.012803 | 0.6066 |
| Bladder | WBVD | 17388 | -0.00471 | -0.00919 | 0.015202 | 0.62166 |
| Small_Intestine | WBVD | 17388 | -0.00435 | -0.00801 | 0.010213 | 0.6649 |
| Vagina | WBVD | 17388 | -0.00782 | -0.01496 | 0.012263 | 0.73805 |
| Uterus | WBVD | 17388 | -0.00766 | -0.01563 | 0.01146 | 0.74815 |

| | | | | | | |
|---|---|---|---|---|---|---|
| Prostate | WBVD | 17388 | -0.0102 | -0.01938 | 0.01325 | 0.77928 |
| Liver | WBVD | 17388 | -0.00491 | -0.00884 | 0.00632 | 0.7812 |
| Fallopian_Tube | WBVD | 17388 | -0.01203 | -0.02337 | 0.013209 | 0.8188 |
| Kidney | WBVD | 17388 | -0.00863 | -0.01521 | 0.008806 | 0.83658 |
| Esophagus | WBVD | 17388 | -0.01636 | -0.03085 | 0.016406 | 0.84066 |
| Cervix_Uteri | WBVD | 17388 | -0.0143 | -0.02812 | 0.013503 | 0.85515 |
| Ovary | WBVD | 17388 | -0.01243 | -0.02522 | 0.0099 | 0.89541 |
| Testis | WBVD | 17388 | -0.00854 | -0.01477 | 0.005296 | 0.94661 |
| Colon | WBVD | 17388 | -0.03128 | -0.05861 | 0.014959 | 0.98172 |
| Breast | WMR | 17388 | 0.032071 | 0.061741 | 0.014896 | 0.015669 |
| Adipose_Tissue | WMR | 17388 | 0.020565 | 0.040413 | 0.012943 | 0.05606 |
| Small_Intestine | WMR | 17388 | 0.012823 | 0.023608 | 0.010097 | 0.10206 |
| Salivary_Gland | WMR | 17388 | 0.011338 | 0.021123 | 0.010361 | 0.1369 |
| Skin | WMR | 17388 | 0.009758 | 0.018625 | 0.009788 | 0.15942 |
| Prostate | WMR | 17388 | 0.013057 | 0.024803 | 0.013254 | 0.16227 |
| Bladder | WMR | 17388 | 0.014159 | 0.027628 | 0.015142 | 0.17488 |
| Colon | WMR | 17388 | 0.012039 | 0.022559 | 0.014895 | 0.20947 |
| Lung | WMR | 17388 | 0.00695 | 0.013452 | 0.009948 | 0.24242 |
| Spleen | WMR | 17388 | 0.004375 | 0.008678 | 0.007572 | 0.28171 |
| Cervix_Uteri | WMR | 17388 | 0.006992 | 0.013751 | 0.013624 | 0.3039 |
| Thyroid | WMR | 17388 | 0.00498 | 0.009872 | 0.010417 | 0.31633 |
| Blood | WMR | 17388 | 0.002639 | 0.004741 | 0.005992 | 0.32983 |
| Fallopian_Tube | WMR | 17388 | 0.005386 | 0.010461 | 0.013487 | 0.34483 |
| Liver | WMR | 17388 | 0.001734 | 0.003124 | 0.006311 | 0.39175 |
| Stomach | WMR | 17388 | 0.00347 | 0.00631 | 0.013058 | 0.39524 |
| Vagina | WMR | 17388 | 0.001446 | 0.002767 | 0.012288 | 0.45318 |
| Esophagus | WMR | 17388 | 0.000147 | 0.000277 | 0.016317 | 0.49641 |
| Pancreas | WMR | 17388 | -0.00109 | -0.00182 | 0.008438 | 0.55116 |
| Pituitary | WMR | 17388 | -0.00163 | -0.00305 | 0.008108 | 0.57981 |
| Uterus | WMR | 17388 | -0.00298 | -0.00609 | 0.01154 | 0.60203 |
| Testis | WMR | 17388 | -0.00229 | -0.00396 | 0.005315 | 0.66663 |
| Blood_Vessel | WMR | 17388 | -0.00672 | -0.01365 | 0.011187 | 0.72604 |
| Ovary | WMR | 17388 | -0.00672 | -0.01363 | 0.009697 | 0.75582 |
| Kidney | WMR | 17388 | -0.0069 | -0.01216 | 0.008923 | 0.78036 |

| | | | | | | |
|---|---|---|---|---|---|---|
| Brain | WMR | 17388 | -0.00662 | -0.01151 | 0.006517 | 0.84519 |
| Nerve | WMR | 17388 | -0.01113 | -0.02242 | 0.009684 | 0.87467 |
| Muscle | WMR | 17388 | -0.00894 | -0.01692 | 0.006608 | 0.91202 |
| Adrenal_Gland | WMR | 17388 | -0.01522 | -0.02941 | 0.009665 | 0.94232 |
| Heart | WMR | 17388 | -0.01715 | -0.02952 | 0.009249 | 0.9681 |
| Blood | WMRCAL | 17388 | 0.013536 | 0.024319 | 0.005869 | 0.010556 |
| Liver | WMRCAL | 17388 | 0.01416 | 0.02551 | 0.006193 | 0.011129 |
| Spleen | WMRCAL | 17388 | 0.011082 | 0.021979 | 0.007473 | 0.069069 |
| Salivary_Gland | WMRCAL | 17388 | 0.014899 | 0.027756 | 0.010357 | 0.075155 |
| Prostate | WMRCAL | 17388 | 0.015696 | 0.029814 | 0.013051 | 0.11457 |
| Skin | WMRCAL | 17388 | 0.007587 | 0.014482 | 0.00962 | 0.21517 |
| Lung | WMRCAL | 17388 | 0.004632 | 0.008966 | 0.009901 | 0.31995 |
| Pancreas | WMRCAL | 17388 | 0.002521 | 0.004218 | 0.008345 | 0.38131 |
| Muscle | WMRCAL | 17388 | 0.001868 | 0.003534 | 0.006597 | 0.38854 |
| Pituitary | WMRCAL | 17388 | 0.001006 | 0.001879 | 0.007898 | 0.4493 |
| Thyroid | WMRCAL | 17388 | 1.82E-06 | 3.61E-06 | 0.010242 | 0.49993 |
| Cervix_Uteri | WMRCAL | 17388 | -0.00162 | -0.00319 | 0.013464 | 0.54793 |
| Kidney | WMRCAL | 17388 | -0.00121 | -0.00212 | 0.00873 | 0.55494 |
| Vagina | WMRCAL | 17388 | -0.00205 | -0.00392 | 0.012307 | 0.56599 |
| Fallopian_Tube | WMRCAL | 17388 | -0.0028 | -0.00543 | 0.013194 | 0.5839 |
| Small_Intestine | WMRCAL | 17388 | -0.00257 | -0.00474 | 0.010189 | 0.59965 |
| Adrenal_Gland | WMRCAL | 17388 | -0.0041 | -0.00792 | 0.009619 | 0.66506 |
| Esophagus | WMRCAL | 17388 | -0.00778 | -0.01467 | 0.016228 | 0.68418 |
| Heart | WMRCAL | 17388 | -0.00578 | -0.00996 | 0.009218 | 0.73479 |
| Brain | WMRCAL | 17388 | -0.00424 | -0.00736 | 0.006604 | 0.73936 |
| Stomach | WMRCAL | 17388 | -0.00947 | -0.01723 | 0.012912 | 0.76846 |
| Testis | WMRCAL | 17388 | -0.00457 | -0.0079 | 0.00529 | 0.80623 |
| Bladder | WMRCAL | 17388 | -0.01348 | -0.0263 | 0.015208 | 0.81225 |
| Adipose_Tissue | WMRCAL | 17388 | -0.01202 | -0.02363 | 0.012509 | 0.83174 |
| Ovary | WMRCAL | 17388 | -0.00966 | -0.01959 | 0.009722 | 0.83969 |
| Uterus | WMRCAL | 17388 | -0.01207 | -0.02462 | 0.011423 | 0.8547 |
| Colon | WMRCAL | 17388 | -0.01566 | -0.02934 | 0.0147 | 0.85662 |
| Breast | WMRCAL | 17388 | -0.01683 | -0.0324 | 0.014584 | 0.87575 |
| Blood_Vessel | WMRCAL | 17388 | -0.02026 | -0.04115 | 0.011022 | 0.96699 |

| | | | | | | |
|---|---|---|---|---|---|---|
| Nerve | WMRCAL | 17388 | -0.02151 | -0.04335 | 0.009686 | 0.98682 |

**Table S18: Genotype Imputation Quality (R2) by batch, for genome-wide associated variants**

| Pathology | Type | Chr | Position | Top SNP | Average | Minimum | ACT1 | ACT2 | ADC1 | ADC2 | ADC3 | ADC4 | ADC5 | ADC6 | ADC7 | ADC8 | ADC9 | ADC10 | ADC11 | ADC12 | ADC13 | ADC14 | ADC15 | IDIBAPS | NIA-LOAD | TGEN | MAYO | ROSMAP1 | ROSMAP2 | UM1_A | UM1_B | UM1_C | UM2 | UPITT |
|---|---|---|---|---|---|---|---|---|---|---|---|---|---|---|---|---|---|---|---|---|---|---|---|---|---|---|---|---|---|---|---|---|---|---|
| B Score | Ordinal | 2 | 127,135,234 | rs6733839 | 0.973099 | 0.91 | 0.97203 | 0.99972 | 0.97065 | 0.97445 | 0.95866 | 0.96376 | 0.96906 | 0.96866 | 0.96909 | 0.96743 | 0.99895 | 0.99895 | 0.99867 | 0.9993 | 0.99884 | 0.99912 | 0.99888 | 0.999 | 0.97364 | 0.92225 | 0.93831 | 0.94293 | 0.95914 | 0.98097 | 0.97887 | 0.91 | 0.96651 | 0.96894 |
| Braak Stage (AD) | Ordinal | 2 | 127,135,234 | rs6733839 | 0.973099 | 0.91 | 0.97203 | 0.99972 | 0.97065 | 0.97445 | 0.95866 | 0.96376 | 0.96906 | 0.96866 | 0.96909 | 0.96743 | 0.99895 | 0.99895 | 0.99867 | 0.9993 | 0.99884 | 0.99912 | 0.99888 | 0.999 | 0.97364 | 0.92225 | 0.93831 | 0.94293 | 0.95914 | 0.98097 | 0.97887 | 0.91 | 0.96651 | 0.96894 |
| CERAD (C score) | Ordinal | 2 | 127,135,234 | rs6733839 | 0.973099 | 0.91 | 0.97203 | 0.99972 | 0.97065 | 0.97445 | 0.95866 | 0.96376 | 0.96906 | 0.96866 | 0.96909 | 0.96743 | 0.99895 | 0.99895 | 0.99867 | 0.9993 | 0.99884 | 0.99912 | 0.99888 | 0.999 | 0.97364 | 0.92225 | 0.93831 | 0.94293 | 0.95914 | 0.98097 | 0.97887 | 0.91 | 0.96651 | 0.96894 |
| CERAD (C score) | Ordinal | 11 | 86,157,598 | rs3851179 | 0.998067 | 0.98352 | 0.99951 | 0.99995 | 0.99908 | 0.99979 | 0.99957 | 0.99901 | 0.9987 | 0.99998 | 0.99991 | 0.99982 | 0.99999 | 0.99964 | 0.99979 | 0.99978 | 0.99998 | 0.99979 | 0.99999 | 0.994 | 0.99949 | 0.98974 | 0.99934 | 0.98352 | 0.99998 | 0.99927 | 1 | 0.98875 | 0.99839 | 0.99911 |
| Any amyloid | Binary | 4 | 65,028,134 | rs1448239 | 0.923166 | 0.856 | 0.92129 | 0.91983 | 0.90375 | 0.92121 | 0.94133 | 0.962 | 0.93813 | 0.94592 | 0.9436 | 0.94881 | 0.94124 | 0.91895 | 0.91258 | 0.92846 | 0.90581 | 0.93649 | 0.94275 | 0.856 | 0.91034 | 0.87733 | 0.92668 | 0.86959 | 0.91939 | 0.91527 | 0.95941 | 0.94455 | 0.92845 | 0.90949 |
| Any amyloid | Binary | 21 | 39,094,313 | rs2836880 | 0.976457 | 0.953 | 0.9782 | 0.97968 | 0.97951 | 0.9816 | 0.98564 | 0.98035 | 0.97667 | 0.97615 | 0.98126 | 0.9784 | 0.98143 | 0.97601 | 0.98202 | 0.97551 | 0.97174 | 0.97841 | 0.96838 | 0.953 | 0.97849 | 0.97229 | 0.97002 | 0.96391 | 0.98334 | 0.97919 | 0.98616 | 0.96867 | 0.9773 | 0.97746 |
| Atherosclerosis | Binary | 2 | 13,906,464 | rs4144605 | 0.992734 | 0.978 | 0.99678 | 0.9962 | 0.99657 | 0.99612 | 0.99682 | 0.99747 | 0.99704 | 0.99663 | 0.99731 | 0.99687 | 0.99658 | 0.9966 | 0.99662 | 0.99642 | 0.99616 | 0.99693 | 0.9968 | 0.978 | 0.98701 | 0.98825 | 0.98033 | 0.98548 | 0.98773 | 0.98811 | 0.99107 | 0.99095 | 0.99015 | 0.98554 |
| Atherosclerosis | Binary | 18 | 10,284,176 | rs206499 | 0.978746 | 0.906 | 0.97322 | 0.97117 | 0.97611 | 0.97414 | 0.9999 | 0.99971 | 0.99997 | 0.99998 | 0.99998 | 0.99942 | 0.96434 | 0.96031 | 0.9636 | 0.96762 | 0.96297 | 0.96785 | 0.96107 | 0.906 | 0.97567 | 0.9989 | 0.92477 | 0.99872 | 0.99991 | 0.96991 | 0.98981 | 0.99994 | 0.99999 | 0.9999 |
| Any CVD | Binary | 4 | 180,633,067 | rs1129924 | 0.960119 | 0.89111 | 0.94245 | 0.96421 | 0.94468 | 0.95177 | 0.95055 | 0.96425 | 0.9617 | 0.94569 | 0.96218 | 0.95551 | 0.99127 | 0.98882 | 0.96701 | 0.94842 | 0.97861 | 0.98007 | 0.9625 | 0.947 | 0.95438 | 0.97838 | 0.93059 | 0.97381 | 0.94078 | 0.94888 | 0.99353 | 0.99879 | 0.9664 | 0.89111 |
| Vascular infarcts | Binary | 7 | 111,843,829 | rs6976029 | 0.984537 | 0.93959 | 0.98388 | 0.99287 | 0.98508 | 0.98388 | 0.98398 | 0.98687 | 0.98567 | 0.98401 | 0.98534 | 0.98654 | 0.99085 | 0.9908 | 0.99313 | 0.99084 | 0.9914 | 0.99231 | 0.99336 | 0.97 | 0.98479 | 0.98059 | 0.93959 | 0.98047 | 0.98254 | 0.98447 | 0.9894 | 0.98619 | 0.98522 | 0.98296 |
| WMR | Ordinal | 4 | 48,443,972 | rs4346719 | 0.988207 | 0.97345 | 0.98863 | 0.98764 | 0.98938 | 0.98745 | 0.99213 | 0.99103 | 0.99094 | 0.99037 | 0.99127 | 0.99114 | 0.99014 | 0.98991 | 0.99042 | 0.98776 | 0.99011 | 0.98912 | 0.991 | 0.984 | 0.98864 | 0.98056 | 0.97969 | 0.98213 | 0.99183 | 0.9874 | 0.99312 | 0.97345 | 0.99206 | 0.98848 |
| Lewy bodies | Binary | 4 | 89,838,405 | rs3806789 | 0.991935 | 0.98279 | 0.99233 | 0.98511 | 0.99194 | 0.99257 | 0.99636 | 0.99502 | 0.99456 | 0.99457 | 0.99637 | 0.99671 | 0.98653 | 0.98842 | 0.98955 | 0.98279 | 0.98658 | 0.98908 | 0.98684 | 0.993 | 0.99501 | 0.99501 | 0.98953 | 0.99574 | 0.98623 | 0.99365 | 0.99684 | 0.99378 | 0.99537 | 0.99468 |
| Lewy bodies | Ordinal, 3 | 4 | 89,838,405 | rs3806789 | 0.991935 | 0.98279 | 0.99233 | 0.98511 | 0.99194 | 0.99257 | 0.99636 | 0.99502 | 0.99456 | 0.99457 | 0.99637 | 0.99671 | 0.98653 | 0.98842 | 0.98955 | 0.98279 | 0.98658 | 0.98908 | 0.98684 | 0.993 | 0.99501 | 0.99501 | 0.98953 | 0.99574 | 0.98623 | 0.99365 | 0.99684 | 0.99378 | 0.99537 | 0.99468 |
| Lewy bodies | Ordinal, 5 | 4 | 89,838,405 | rs3806789 | 0.991935 | 0.98279 | 0.99233 | 0.98511 | 0.99194 | 0.99257 | 0.99636 | 0.99502 | 0.99456 | 0.99457 | 0.99637 | 0.99671 | 0.98653 | 0.98842 | 0.98955 | 0.98279 | 0.98658 | 0.98908 | 0.98684 | 0.993 | 0.99501 | 0.99501 | 0.98953 | 0.99574 | 0.98623 | 0.99365 | 0.99684 | 0.99378 | 0.99537 | 0.99468 |
| Hippocampal sclerosis | Binary | 7 | 12,227,630 | rs4721058 | 0.997891 | 0.993 | 0.99833 | 0.99745 | 0.99957 | 0.99897 | 0.99949 | 0.9996 | 0.99959 | 0.99942 | 0.9994 | 0.99961 | 0.99919 | 0.99664 | 0.99718 | 0.99774 | 0.99683 | 0.99717 | 0.99668 | 0.993 | 0.99806 | 0.99648 | 0.99696 | 0.99729 | 0.9973 | 0.99776 | 0.99883 | 0.99811 | 0.99687 | 0.99744 |
| Hippocampal sclerosis | Binary | 17 | 44,352,876 | rs5848 | 0.973779 | 0.92994 | 0.96218 | 0.96495 | 0.95888 | 0.95907 | 0.99906 | 0.99818 | 0.99867 | 0.99802 | 0.99903 | 0.99889 | 0.97027 | 0.96828 | 0.96737 | 0.97521 | 0.96674 | 0.97247 | 0.96671 | 0.997 | 0.96062 | 0.92994 | 0.95104 | 0.9381 | 0.99869 | 0.95762 | 0.9984 | 0.94589 | 0.99983 | 0.9647 |
| TDP-43 proteinopathy | Binary | 7 | 12,235,882 | rsl468804 | 0.996902 | 0.99186 | 0.99798 | 0.99533 | 0.99823 | 0.99668 | 0.99787 | 0.99845 | 0.99802 | 0.99822 | 0.99782 | 0.99802 | 0.99512 | 0.99376 | 0.99562 | 0.9964 | 0.99186 | 0.99398 | 0.99313 | 0.993 | 0.99806 | 0.99971 | 0.99696 | 0.99987 | 0.99738 | 0.99768 | 0.99877 | 0.99999 | 0.99796 | 0.99739 |
| TDP-43 proteinopathy | Ordinal | 7 | 12,235,882 | rsl468804 | 0.996902 | 0.99186 | 0.99798 | 0.99533 | 0.99823 | 0.99668 | 0.99787 | 0.99845 | 0.99802 | 0.99822 | 0.99782 | 0.99802 | 0.99512 | 0.99376 | 0.99562 | 0.9964 | 0.99186 | 0.99398 | 0.99313 | 0.993 | 0.99806 | 0.99971 | 0.99696 | 0.99987 | 0.99738 | 0.99768 | 0.99877 | 0.99999 | 0.99796 | 0.99739 |

**Table S19: Neuropathology phenotype coding**

| Category | Phenotype | Coding | Type | N indiv. | N sets |
|---|---|---|---|---|---|
| AD | Braak Stage | Braak stage, 0-VI | Ordinal | 11,541 | 17 |
| | Thal Phase | Thal phase, 0-5 | Ordinal | 3,610 | 9 |
| | A Score | 0 = none<br>1 = Thal phase 1 or 2<br>2 = Thal phase 3<br>3 = Thal phase 4 or 5 | Ordinal | 8,524 | 10 |
| | B Score | 0 = none<br>1 = Braak stage I-II<br>2 = Braak stage III-IV<br>3 = Braak stage V-VI | Ordinal | 11,684 | 17 |
| | C Score (CERAD) | 0 = none<br>1 = sparse<br>2 = moderate<br>3 = frequent | Ordinal | 11,067 | 17 |
| | NIA-Reagan Criteria | 0 = none<br>1 = low<br>2 = intermediate<br>3 = high | Ordinal | 7,332 | 8 |
| | Amyloid, any | 0 = none; 1 = any | Binary | 10,825 | 17 |
| | ADNC/ABC Score | 0 = not AD<br>1 = low ADNC<br>2 = intermediate ADNC<br>3 = high ADNC | Ordinal | 8,667 | 10 |
| | CVD, any | 0 = none; 1 = any | Binary | 10,071 | 13 |
| | Arteriolosclerosis, any | 0 = none; 1 = any | Binary | 7,933 | 10 |
| | Arteriolosclerosis severity | 0 = none<br>1 = mild<br>2 = moderate | Ordinal | 7,933 | 10 |

| | | | | | |
|---|---|---|---|---|---|
| | | 3 = severe | | | |
| CVD | Atherosclerosis, any | 0 = none; 1 = any | Binary | 8,705 | 11 |
| | Atherosclerosis severity | 0 = none<br>1 = mild<br>2 = moderate<br>3 = severe | Ordinal | 8,705 | 11 |
| | CAA, any | 0 = none; 1 = any | Binary | 9,722 | 13 |
| | CAA severity | 0 = none<br>1 = mild<br>2 = moderate<br>3 = severe | Ordinal | 8,771 | 11 |
| LB | Lewy bodies, any | 0 = none; 1 = any | Binary | 10,454 | 15 |
| | Lewy bodies, three groups | 0 = none<br>1 = brainstem predominant<br>2 = limbic/neocortical | Ordinal | 10,454 | 15 |
| | Lewy bodies, five groups | 0 = none<br>1 = olfactory or unspecified<br>2 = brainstem predominant<br>3 = limbic<br>4 = neocortical | Ordinal | 10,454 | 15 |
| HS | Hippocampal sclerosis | 0 = none; 1 = any | Binary | 9,509 | 13 |
| VBI | VBI, any | 0 = none; 1 = any | Binary | 10,063 | 13 |
| | WMR, any | 0 = none; 1 = any | Binary | 6,568 | 6 |
| | WMR severity | 0 = none<br>1 = mild<br>2 = moderate<br>3 = severe | Ordinal | 2,260 | 6 |
| | Microinfarcts, any | 0 = none; 1 = any | Binary | 8,850 | 10 |
| | Infarcts, any | 0 = none; 1 = any | Binary | 8,755 | 10 |
| TDP-43 proteinopathy | TDP-43 proteinopathy, any | 0 = none; 1 = any | Binary | 4,669 | 11 |
| | [illegible] | 0 = none<br>1 = amygdala | | | |

| | | | | | |
|---|---|---|---|---|---|
| **TDP-43 proteinopathy** | TDP-43 proteinopathy severity | 2 = hippocampus or entorhinal<br>3 = neocortex | Ordinal | 3,840 | 10 |
